# Genome-wide association studies identify 77 loci for suicidality and provide novel biological insights

**DOI:** 10.1101/2025.10.22.25338076

**Authors:** Sarah MC Colbert, Maria Koromina, Alexander S Hatoum, Mallory Stephenson, Alexis C Edwards, Emma C Johnson, Xuejun Qin, Andrey A Shabalin, Lucas T Ito, Kevin S O’Connell, Arvid Harder, Jens Hjerling-Leffler, Min Ji Kim, Ikuo Otsuka, Laura Vilar-Ribó, Arpana Agrawal, Martin Alda, Lars Alfredsson, Fazil Aliev, Till FM Andlauer, Celso Arango, Arnoud Arntz, Swapnil Awasthi, Olatunde O Ayinde, Silviu-Alin Bacanu, Peter B Barr, Claiton HD Bau, Bernhard T Baune, Jean C Beckham, Cosmin A Bejan, Sintia Belangero, Klaus Berger, Joanna M Biernacka, Ryan Bogdan, Dorret I Boomsma, Anders D Børglum, Kyle J Bourassa, David L Braff, Alice Braun, Rodrigo A Bressan, Tanja M Brueckl, Monika Budde, Brenda Cabrera-Mendoza, Bernardo Carpiniello, Zuriel Ceja, Jorge A Cervilla, Chiao-Erh Chang, Boris Chaumette, Myeong Jae Cheon, Sven Cichon, Jonathan RI Coleman, Hilary Coon, William E Copeland, Darina Czamara, Nina Dalkner, Udo Dannlowski, Friederike S David, Ditte Demontis, Arianna Di Florio, Carmen C Diaconu, Danielle M Dick, Dimitris Dikeos, Srdjan Djurovic, Howard J Edenberg, Annette Erlangsen, Sebastian Euler, Peter Falkai, Giuseppe Fanelli, Frederike T Fellendorf, Panagiotis Ferentinos, Fernando Fernandez-Aranda, Andreas J Forstner, Oleksandr Frei, Gabriel R Fries, Janice M Fullerton, Marie E Gaine, Hanga C Galfalvy, Marco Galimberti, Judith Garcia-Aymerich, Melanie E Garrett, Micha Gawlik, Joel Gelernter, Katherine Gordon-Smith, Aaron J Gorelik, Philip Gorwood, Hans J Grabe, Melissa J Green, Maria Grigoroiu-Serbanescu, Priya Gupta, Blanca Gutiérrez, Jose Guzman-Parra, Seonggyun Han, Marit Haram, Elizabeth R Hauser, Urs Heilbronner, Sabine C Herpertz, Jesús Herrera-Imbroda, Victor M Hesselbrock, Akitoyo Hishimoto, Bharath Holla, Anastasia Izotova, Yoonjeong Jang, Susana Jimenez-Murcia, Lisa A Jones, Lina Jonsson, JooEun Kang, Joon Ho Kang, Pamela K Keel, Jaeyoung Kim, Dongjun Kim, Tilo Kircher, George Kirov, Julia Kraft, John Kramer, Henry R Kranzler, Po-Hsiu Kuo, Siim Kurvits, Dongbing Lai, Marilyn T Lake, Mikael Landén, Séverine Lannoy, Matthew H Law, Byung-Chul Lee, Young Kee Lee, Kelli Lehto, Daniel F Levey, Cathryn M Lewis, Qingqin S Li, Calwing Liao, Penelope A Lind, Christine Lochner, Adriana Lori, Hermine HM Maes, Jayant Mahadevan, Mirko Manchia, Becky Mars, Nicholas G Martin, Lourdes Martorell, Andrew M McIntosh, Shelley F McMain, Andrew McQuillin, Sarah E Medland, Philip B Mitchell, Esther Molina, Eric T Monson, Mary S Mufford, Gerard Muntané, Richard Musil, Woojae Myung, Ana Iulia Neagu, Trine T Nielsen, Markus M Nöthen, Yaira Z Nunez, John I Nurnberger, Satoshi Okazaki, Catherine M Olsen, Roel A Ophoff, Michael J Owen, Pedro M Pan, Sergi Papiol, Juan C Pascual, George P Patrinos, Joanna M Pawlak, Brenda WJH Penninx, Ana M Pérez-Gutiérrez, Nader Perroud, Roseann E Peterson, Claudia Pisanu, Giorgio Pistis, Bernice Porjesz, Danielle Posthuma, Abigail Powers, Martin Preisig, Meera Purushottam, Andreas Reif, Eva Z Reininghaus, Miguel E Rentería, Stephan Ripke, Michael A Ripperger, Margarita Rivera, Emily K Roberts, Gloria Roberts, Linn Rødevand, Stefan Roepke, Diego L Rovaris, Giovanni A Salum, Alan R Sanders, Marcos L Santoro, Chelsea Sawyers, Stephen W Scherer, Claudia Schilling, Christian Schmahl, Peter R Schofield, Thomas G Schulze, Laura J Scott, Alessandro Serretti, Alexey Shadrin, Toshiyuki Shirai, Olav B Smeland, Jordan W Smoller, Marcus Sokolowski, Edmund J Sonuga-Barke, Alessio Squassina, Anna Starnawska, Nils Eiel Steen, Dan J Stein, Frederike Stein, Murray B Stein, Fabian Streit, Reeteka Sud, Patrick F Sullivan, Chikashi Terao, Claudio Toma, Leonardo Tondo, Gustavo Turecki, Rudolf Uher, Robert J Ursano, Sandra Van der Auwera, Marquis P Vawter, Alja Videtic Paska, Elisabet Vilella, John B Vincent, Biju Viswanath, Vladimir Vladimirov, Danuta E Wasserman, Thomas W Weickert, David C Whiteman, Virginia L Willour, Erik D Wiström, Stephanie H Witt, Hong-Hee Won, Robyn E Wootton, Clement C Zai, Jian Zhang, Lea Zillich, CVEDA and MGL, Genoplan Research Team, International Borderline Genomics Consortium, Ole A Andreassen, Abraham A Palmer, Sandra Sanchez-Roige, J John Mann, Nathan A Kimbrel, Allison E Ashley-Koch, Douglas M Ruderfer, Anna R Docherty, Niamh Mullins

## Abstract

Suicidality contributes substantially to global morbidity and mortality, yet despite its heritability, its biological etiology remains largely elusive. We conducted multi-ancestry genome-wide association study meta-analyses of suicidal ideation (259,747 cases), suicide attempt (64,993 cases), suicide death (9,197 cases), and suicidal behavior (suicide attempt/ death, 75,300 cases), across 54 cohorts (e.g., the Psychiatric Genomics Consortium, Million Veteran Program, UK Biobank). We identified 77 significant loci across meta-analyses, including 59 previously unreported for suicidality. SNP-based heritability ranged from 2.0-6.7% and there were strong, yet incomplete, genetic correlations between suicidality phenotypes (0.70-0.88). Fine-mapping prioritized putative causal SNPs and 20 credible genes. Enrichment analyses implicated synaptic pathways and neuronal populations predominantly in subcortical brain regions (e.g., amygdala excitatory, medium spiny, hippocampal CA1-3). Together, these findings establish suicidality as a polygenic set of traits with both shared and distinct genetic influences, providing a foundation for future studies of suicide biology and etiology.

## MAIN

Over 700,000 individuals die by suicide each year, making suicide a leading cause of death worldwide^1–3^. Beyond mortality, many more think about or attempt suicide^2,4,5^, and suicide deaths themselves are associated with significant morbidity among suicide-loss survivors^6^.

Suicidality encompasses a phenotypically heterogenous spectrum of thoughts, behaviors, and outcomes^7–12^, ranging from suicidal ideation (SI) to suicide attempt (SA) and suicide death (SD). SI reflects thoughts of engaging in suicide-related behavior, SA involves non-fatal self-directed potentially injurious behavior with any intent to die, and SD represents death caused by self-injurious behavior with any intent to die^13^.

Twin and family studies suggest that genetic factors influence suicidality, with heritability estimates ranging from 30-55%^14^. While suicidality often co-occurs with psychiatric disorders^15–17^, the heritability of suicidality phenotypes remains significant even when accounting for comorbid psychiatric disorders, supporting the notion that while there is partial genetic overlap, suicidality also involves unique genetic influences not entirely explained by psychiatric disorders^18–20^. Genome-wide association studies (GWAS) have begun to uncover the polygenic architecture of suicidality phenotypes, previously identifying five loci for SI, 12 for SA (in some studies combining SA and SD), and two for SD^21–23^. However, their genetic architecture and underlying biology remain incompletely understood, largely due to modest sample sizes and insufficient ancestral diversity.

To address these gaps, the Psychiatric Genomics Consortium Suicide Working Group (PGC SUI) reports here the largest and most ancestrally diverse GWAS meta-analyses of SI (N_Cases_ = 259,747), SA (N_Cases_ = 64,993), and SD (N_Cases_ = 9,197) to date. In addition to these individual phenotypes, we also analyzed an aggregated phenotype, suicidal behavior (SB; N_Cases_ = 75,300), defined here to include SA or SD as per previous GWAS^19,22^ to maximize sample size. Across these four suicidality phenotypes, we identify 77 genome-wide significant (GWS) loci, 59 of which are previously unreported, and further characterize the biological underpinnings of suicidality, providing insights to inform future prevention and intervention efforts.

## RESULTS

### GWAS meta-analyses reveal heritability and loci associated with suicidality

We conducted GWAS meta-analyses for SI, SA, SD, and SB across 54 studies worldwide. Cases and controls were defined using our published PGC SUI phenotyping protocol from Colbert et al.^7^ which details the specific criteria and rationale and is briefly described in Table 1 and Supplementary Figure 1. Cohort-specific phenotype definitions are available in the Supplementary Note.

**Table 1:** Suicidality phenotype definitions and case and control criteria summarized from PGC SUI Phenotyping Protocol. A flowchart visualizing the phenotyping process is provided in Supplementary Figure 1.

|  |  |  |  |  |
| --- | --- | --- | --- | --- |
| **Phenotype (Abbreviation), Definition** | **Case criteria** | **Control criteria** | **Aggregated phenotype (Abbreviation)** |  |
| Suicidal Ideation (SI), Thoughts of engaging in suicide-related behavior. | Evidence of lifetime SI specifically (SA/SD do not automatically meet criteria). SI must be distinguished from thoughts of self-harm. Psychiatric disorder may/ may not be present. | No history of SI. Psychiatric disorder may be present. Exclusion criteria: history of SA or SD; missing SI information AND a psychiatric diagnosis is present. | Suicidality |  |
| Suicide Attempt (SA), A non-fatal self-directed potentially injurious behavior with any intent to die. May/ may not result in injury. | Evidence of lifetime SA specifically (SD does not automatically meet criteria). SA must be distinguished from non-suicidal self-injury. Psychiatric disorder may/ may not be present. | No history of SA. Psychiatric disorder or SI may be present. Exclusion criteria: SD; missing SA information AND a psychiatric diagnosis is present. |  | Suicidal Behavior (SB) |
| Suicide Death (SD), Death caused by self-directed injurious behavior with any intent to die as a result of the behavior. | Evidence of SD. Psychiatric disorder may/ may not be present. | No SD, SA (since most controls are living) or deaths of undetermined intent. Psychiatric disorder or SI may be present. Exclusion criteria: missing SA information and a psychiatric diagnosis is present. |  |  |

Inverse-variance-weighted fixed-effects meta-analyses were performed both within and across genetic ancestry or population groups (AFR = African, SAS = South Asian, EAS = East Asian, EUR = European, and LA = Latin American). Table 2 provides sample sizes for each meta-analysis, and Supplementary Table 1 details cohort-specific sample sizes.

**Table 2:** Sample sizes and numbers of cohorts for each suicidality phenotype by genetic ancestry or population.

|  | Suicidal ideation (SI) |  |  | Suicide attempt (SA) |  |  | Suicide death (SD) |  |  | Suicidal behavior (SB)* |  |  |
| --- | --- | --- | --- | --- | --- | --- | --- | --- | --- | --- | --- | --- |
| **Genetic Ancestry or Population** | N Cohorts | N Cases | N Controls<sup>†</sup> | N Cohorts | N Cases | N Controls<sup>†</sup> | N Cohorts | N Cases | N Controls<sup>†</sup> | N Cohorts | N Cases | N Controls<sup>†</sup> |
| African | 7 | 52,615 | 188,733 | 8 | 6,710 | 128,960 | - | - | - | 8 | 6,753 | 128,960 |
| South Asian | 1 | 243 | 1,471 | 1 | 82 | 1,632 | - | - | - | 1 | 82 | 1,632 |
| East Asian | 5 | 8,026 | 30,999 | 5 | 2,169 | 23,519 | 1 | 746 | 14,049 | 6 | 2,918 | 37,568 |
| European | 20 | 176,147 | 1,010,300 | 29 | 53,919 | 1,063,988 | 5 | 7,584 | 652,070 | 30 | 62,533 | 1,090,754 |
| Latin American | 4 | 22,716 | 78,440 | 3 | 2,113 | 50,938 | 1 | 867 | 2,043 | 4 | 3,014 | 52,981 |
| Multi-ancestry | 37 | 259,747 | 1,309,943 | 46 | 64,993 | 1,269,037 | 7 | 9,197 | 668,162 | 49 | 75,300 | 1,311,895 |

\*The SB phenotype enabled inclusion of cohorts where both SA and SD data were available but the number of SD cases alone was insufficient to contribute to the SD only GWAS meta-analysis. As a result, the total SB case count does not equal the simple sum of SA and SD cases.
<sup>†</sup> Control sample sizes vary across suicidality phenotypes due to differences in cohort composition and phenotype availability. Availability of SI, SA, and SD data did not fully overlap across cohorts; therefore, inclusion of a cohort in one analysis did not guarantee inclusion in others. Differences in control counts reflect cohort participation rather than differences in underlying prevalence of the suicidality phenotype.
\*<sup>†</sup> More details are provided in the Methods and Supplementary Table 1.

All meta-analyses were assessed for inflation using linkage disequilibrium score regression (LDSC) with ancestry/population-matched reference panels^24^. LDSC intercepts ranged from 0.99-1.06, indicating little to no test statistic inflation (Supplementary Table 2). We observed significant liability scale SNP-heritability (*h^2^*; p < 0.0029) in the EUR GWAS for SA (6.6%), SI (3.0%), SD (5.1%), and SB (6.7%) (Supplementary Table 2). *h^2^* estimates in the AFR, EAS, and LA GWAS were similar, but showed larger standard errors due to limited sample sizes (Supplementary Table 2).

Across all GWAS meta-analyses (Supplementary Tables 3-10), we identified 77 unique genome-wide significant loci (p < 5×10^-8^) associated with at least one suicidality phenotype (Fig. 1, Supplementary Table 11). These included 18 loci previously reported for suicidality and 59 loci not overlapping with any locus previously reported as GWS for any suicidality phenotype^19–23,25,26^. We checked the 59 previously unreported loci for consistent directions of effect with, and whether they reached replication significance (p < 0.05/77) in, the most recent previously published GWAS of the corresponding phenotype (Supplementary Table 12). All loci showed consistent directions of effect, and 49 reached replication significance.

**Figure 1.**
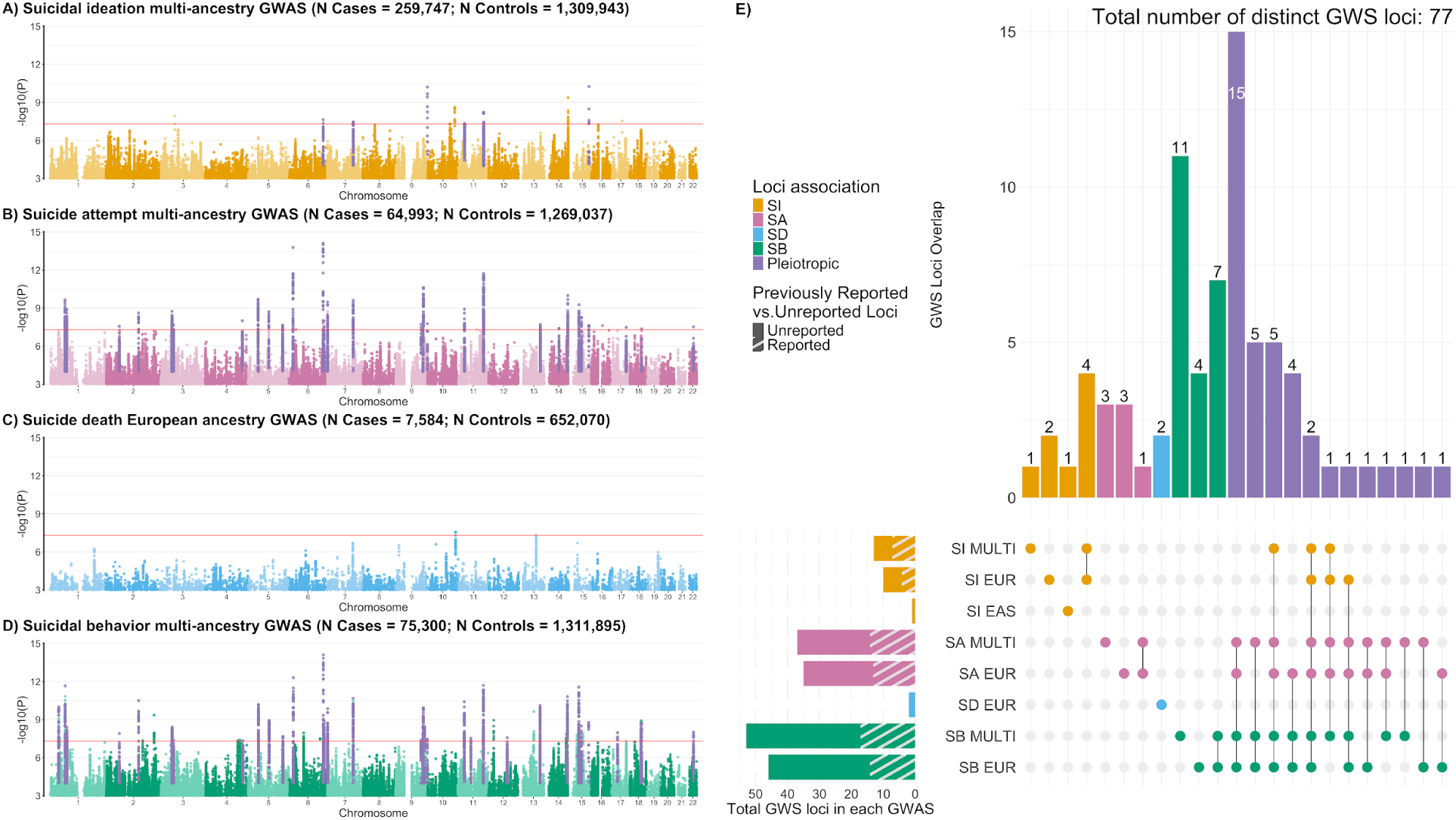
Manhattan and UpSet plots for GWAS meta-analyses of each suicidality phenotype. In panels A-D, the x-axis shows chromosomal position and the y-axis shows significance of the association as -log10(P). The red line indicates the genome-wide significance threshold (p = 5 x 10^-8^). Purple peaks indicate pleiotropic loci (i.e., loci significantly associated with more than one phenotype). **A)** Manhattan plot for the multi-ancestry GWAS of suicidal ideation (SI, yellow). **B)** Manhattan plot for the multi-ancestry (MULTI) GWAS of suicide attempt (SA, pink). **C)** Manhattan plot for the European (EUR) ancestry GWAS of suicide death (SD, blue). **D)** Manhattan plot for the multi-ancestry GWAS of suicidal behavior (SB, green). **E)** UpSet plot depicting the 77 distinct GWAS/phenotype-specific (SI = yellow, SA = pink, SD = blue, SB = green) and pleiotropic (purple) genome-wide significant (GWS) loci associated with the various suicidality GWAS. Vertical bars represent the number of loci associated with each combination of GWAS, as indicated in the intersection matrix below. For example, 11 loci were identified only in the SB multi-ancestry GWAS, while 15 loci overlapped between the SB multi-ancestry, SB EUR, SA multi-ancestry, and SA EUR GWAS. Horizontal bars represent the number of loci identified in the respective GWAS and full shading represents previously unreported loci, while diagonal shading represents previously reported loci. For example, the SD EUR GWAS identified two genome-wide significant loci, both of which have not been reported in previous suicidality GWAS. Full information for each locus and its overlap is provided in Supplementary Table 11. Results for genome-wide significant loci in each GWAS are reported in Supplementary Tables 3-10 and forest and area plots of the lead SNPs at GWS loci are provided in Supplementary Data 1 and 2, respectively. Ancestry/population-specific Manhattan plots are presented in Extended Data Figs. 1-4.

The multi-ancestry SI GWAS identified 13 GWS loci (Supplementary Table 3, Fig. 1A). The EUR SI GWAS detected seven of these and three additional loci (Supplementary Table 4, Extended Data Fig. 1C). The EAS SI GWAS identified one additional previously unreported locus (Supplementary Table 5, Extended Data Fig. 1B). No GWS loci were identified in the AFR or LA SI GWAS (Extended Data Fig. 1).

For SA, we identified 37 GWS loci in the multi-ancestry GWAS (Supplementary Table 6, Fig. 1B) and 35 loci in the EUR GWAS (Supplementary Table 7, Extended Data Fig. 2C), with 27 overlapping between the two (Fig. 1E). Of the 45 total loci associated with SA, nine overlapped with loci identified in the SI GWAS (Fig. 1E, Supplementary Table 11). No GWS loci were detected in the AFR, EAS, or LA GWAS of SA (Extended Data Fig. 2).

For SD, no loci reached genome-wide significance in the multi-ancestry GWAS (Extended Data Fig. 3). However, the EUR GWAS identified two loci (Supplementary Table 8, Fig. 1C) not previously associated with any published suicidality GWAS, and not identified in the current SI, SA, or SB GWAS. These loci included one in the *ARMS2* gene on chromosome 10 and one intergenic locus on chromosome 13.

In the SB GWAS, we detected 53 GWS loci in the multi-ancestry analysis (Supplementary Table 9, Fig. 1D) and 46 loci in the EUR analysis (Supplementary Table 10, Extended Data Fig. 4), with 39 of these loci overlapping between the two analyses (Fig. 1E, Supplementary Table 11). Notably, the SB GWAS identified 22 additional loci which did not reach genome-wide significance in the SA GWAS. Furthermore, nine of the SB loci overlapped with SI GWS loci. No GWS loci were detected in the AFR, EAS, or LA GWAS of SB (Extended Data Fig. 4).

Although no GWS loci were identified for SI, SA, and SB in AFR and LA GWAS, or for SA and SB in EAS GWAS, several lead SNPs from the multi-ancestry GWAS showed concordant effect directions in these groups (Supplementary Tables 13-15).

### Shared and distinct genetic architectures of suicidality and related phenotypes

Unless otherwise specified, all post-GWAS analyses were based on multi-ancestry GWAS results to maximize statistical power. We estimated genome-wide genetic correlations (r_g_) between the suicidality phenotypes using LDSC^27^. SA and SB showed near-complete genetic correlation (r_g_ = 1.02, se = 0.003, false discovery rate (FDR) p = 3.96×10^-307^; Supplementary Table 16). Given their substantial sample overlap and resulting genetic similarity, we prioritized the SB GWAS over the SA GWAS for downstream analyses because it had greater power (larger sample size, higher mean χ^2^, and more significant hits). Among SI, SD, and SB, the strongest genetic correlation was observed between SI and SB (r_g_ = 0.88), followed by lower correlations between SD and SB (r_g_ = 0.73) and between SD and SI (r_g_ = 0.70) (all FDR p < 0.05) (Fig. 2, Supplementary Table 16). All three genetic correlations were significantly below 1 (all p < 8.01×10^-5^ for tests for r_g_ < 1), indicating partially distinct genetic architectures among these phenotypes. Each phenotype also displayed a moderate genetic correlation with non-suicidal self-injury (r_g_ = 0.39-0.56, all FDR p < 0.05), indicating a potentially unique genetic component specific to suicidal intent.

**Figure 2.**
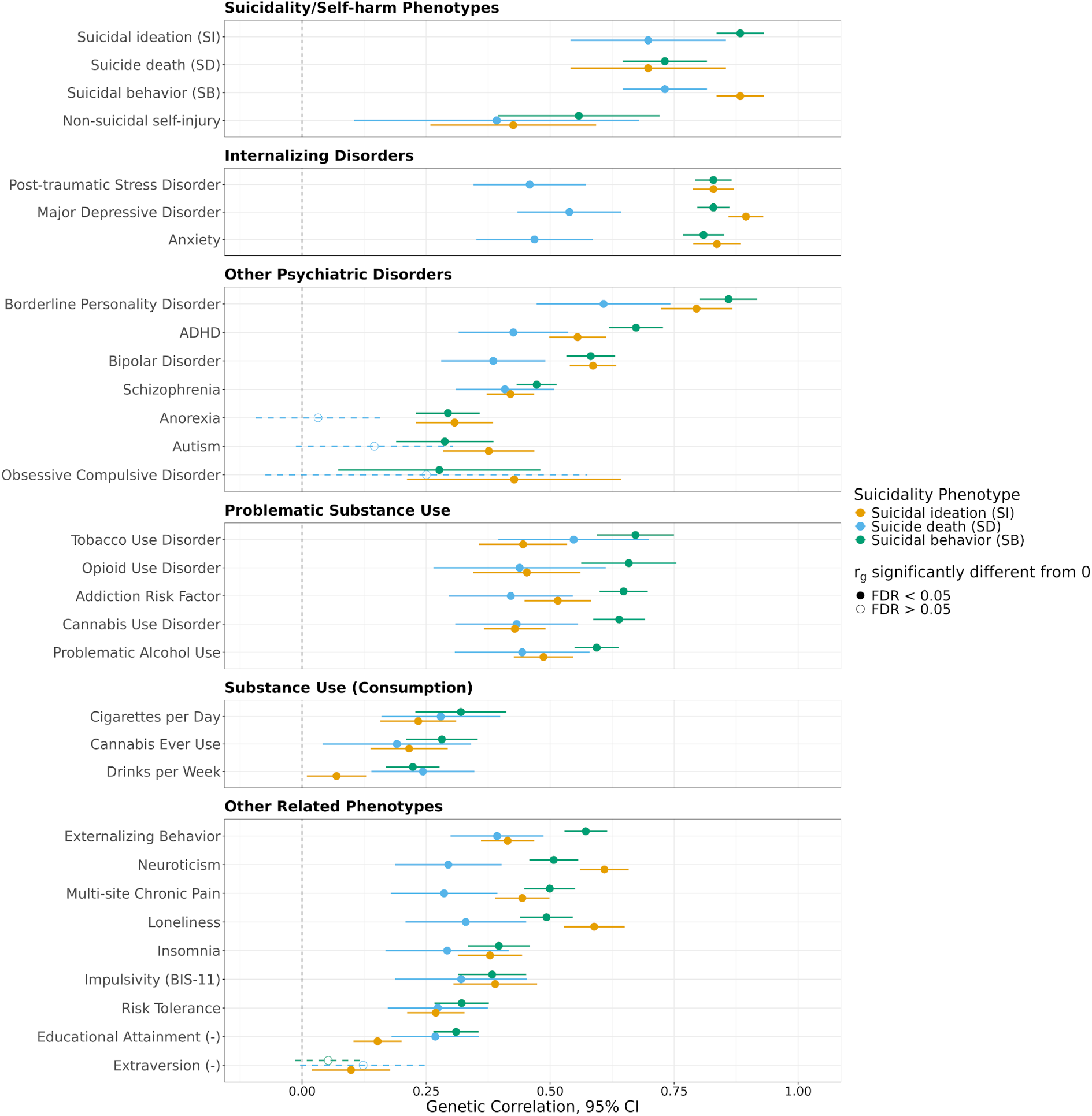
Genetic correlations (r_g_) between suicidality, psychiatric, and behavioral phenotypes. The x-axis shows genetic correlation estimates (points) with 95% confidence intervals (error bars). Points are colored according to suicidality phenotype (yellow = suicidal ideation, blue = suicide death, green = suicidal behavior). Shaded points indicate an r_g_ significantly different from zero (false discovery rate (FDR) < 5%). (-) indicates reverse-coded phenotypes. Additional related phenotypes tested but not shown on the figure are available in Supplementary Table 16.

We also calculated the genetic correlation of SI, SD, and SB with relevant psychiatric and behavioral phenotypes (Supplementary Table 16). Among psychiatric disorders, suicidality phenotypes had the strongest genetic correlations with borderline personality disorder (r_g_ = 0.61-0.86) and internalizing disorders, including post-traumatic stress disorder (PTSD: r_g_ = 0.46-0.83), anxiety (r_g_ = 0.47-0.84), and major depressive disorder (MDD: r_g_ = 0.54-0.90) (all FDR p < 0.05) (Fig. 2, Supplementary Table 16). Across most psychiatric disorders, and especially for internalizing disorders, genetic correlations with SI and SB were consistently higher than those with SD (Fig. 2). Genetic correlations with substance use revealed further distinctions. All suicidality phenotypes showed generally modest genetic correlations with consumption-related substance use, whereas genetic correlations with problematic substance use were significantly higher (all tests for significant differences FDR p<0.05; Supplementary Tables 16-17). SB exhibited significantly higher genetic correlations than SI with externalizing behavior (r_g_ = 0.57 vs. 0.41), problematic substance use phenotypes (e.g., addiction risk factor: r_g_ = 0.65 vs. 0.52), and attention-deficit/hyperactivity disorder (ADHD: r_g_ = 0.67 vs. 0.55) (all tests for significant differences FDR p<0.05; Supplementary Table 16, Supplementary Table 18), indicating that externalizing liability potentially contributes specifically to the etiology of and progression to SB. Interestingly, however, SI and SB did not differ in their genetic correlations with impulsivity (Fig. 2, Supplementary Table 16).

### Polygenic risk scores capture phenotypic variance in suicidality

Using PRS-CSx^28^, we generated multi-ancestry polygenic risk scores (PRS) for suicidality phenotypes in 25 cohorts, each excluded in turn from the discovery GWAS. Within each cohort, we evaluated PRS performance using variance explained on the liability scale (*R*^2^), odds ratios (ORs) comparing individuals in the top versus middle PRS quintiles, and area under the receiver operating characteristic curve (AUC) values (Supplementary Tables 19-23 and Supplementary Figs. 2-4). Then, we calculated effective sample size (N_eff_)-weighted averages of these metrics within and across the AFR, EAS, EUR, and LA groups.

Across all cohorts, the weighted average liability scale variance in SI explained by the SI PRS was 0.49% (95% confidence interval (CI) = 0.37-0.62%) (Fig. 3). Individuals in the top quintile of the SI PRS had 1.23 times higher odds (95% CI = 1.16-1.30) of SI compared with those in the middle quintile (Fig. 3). The corresponding AUC was 0.54 (95% CI = 0.53-0.54).

**Figure 3.**
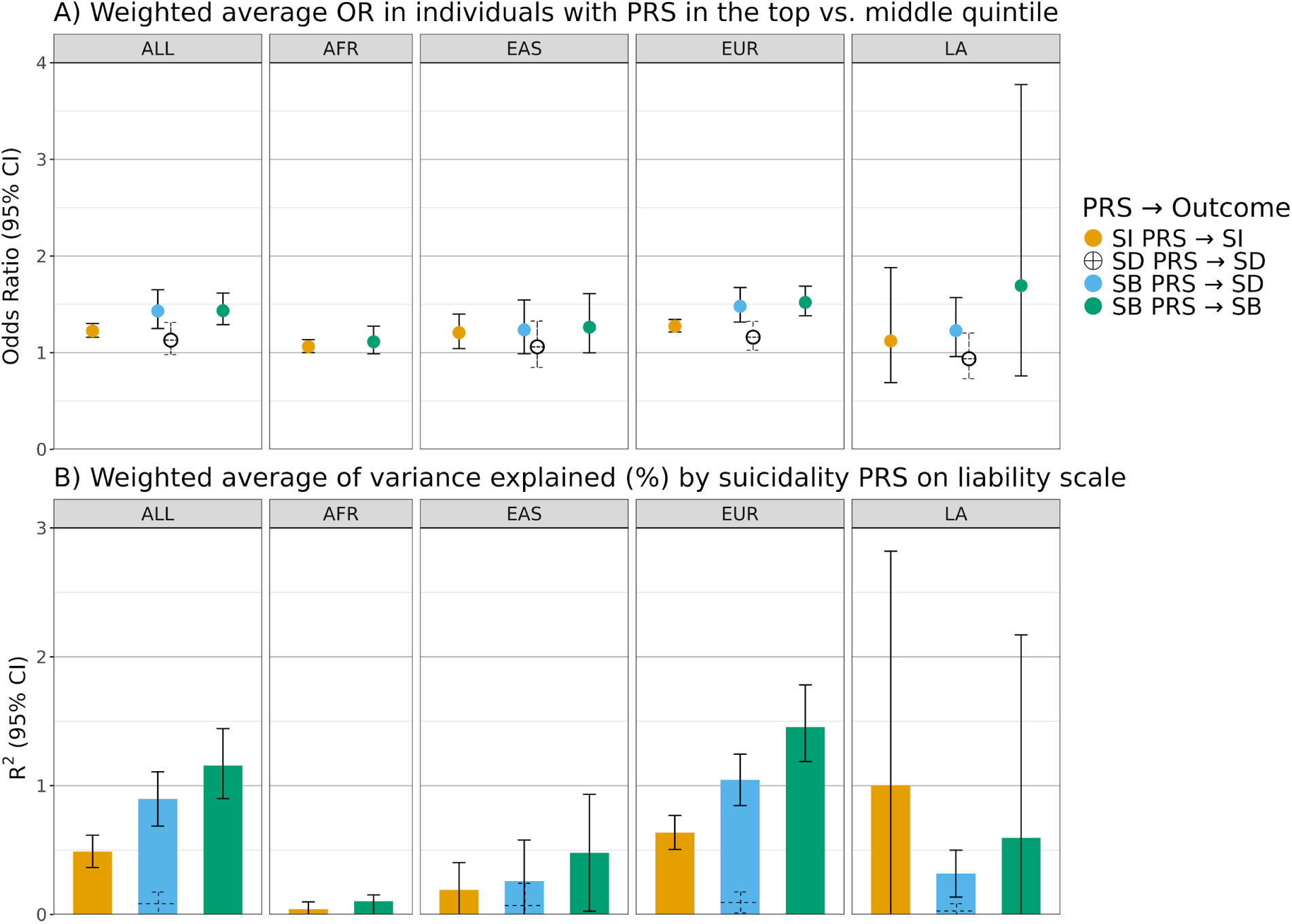
Polygenic risk score (PRS) prediction of suicidality phenotypes across genetic ancestries/populations. **A)** Effective sample size N_eff_ weighted average odds ratios (OR) with 95% confidence intervals (CIs) for the association between suicidality PRS and the corresponding outcome phenotype. ORs compare individuals in the top PRS quintile to those in the middle quintile, stratified by genetic ancestry/population group: all cohorts combined (ALL), African (AFR), East Asian (EAS), European (EUR), and Latin American (LA). **B)** N_eff_ weighted average variance explained (*R*^2^, %) by suicidality PRS on the liability scale, with 95% CIs, across the same genetic ancestry/population groups described above. Colors indicate PRS-phenotype associations: suicidal behavior (SB) PRS predicting SB in green, SB PRS predicting suicide death (SD) in blue, and suicidal ideation (SI) PRS predicting SI in yellow. Cohort-specific results are provided in Supplementary Tables 19-23 and Supplementary Figs. 2-4.

Consistent with the lower power of the discovery GWAS, the SD PRS had limited performance (Fig. 3). The SB PRS explained 1.16% (95% CI = 0.90-1.44%) of the variance in SB and individuals in the top PRS quintile had 1.43 times higher odds (95% CI = 1.29-1.62) of SB than those in the middle quintile (Fig. 3). The corresponding AUC was 0.57 (95% CI = 0.56-0.58).

The SB PRS also explained 0.90% (95% CI = 0.69-1.11%) of the variance in SD (Fig. 3). The SI PRS explained 0.66% (95% CI = 0.32 - 1.00%) of the variance in SB (Supplementary Table 23). The performance of each suicidality PRS was generally higher in EUR target cohorts than in other cohorts, consistent with the more limited power of the AFR, EAS, and LA discovery GWAS to contribute to the multi-ancestry PRS (Fig. 3).

Furthermore, examination of PRS performance across individual cohorts did not reveal systematic differences due to phenotyping method (Supplementary Tables 19-23 and Supplementary Figs. 2-4). Cohorts with more extensive phenotyping did not have improved PRS performance. For example, among EUR cohorts, the variance explained in SI by SI PRS ranged from 0.14-1.81% in cohorts using only interview data, compared to 0.69-1.16% in cohorts using self-report and/or interview data, and was 0.57% in the MVP cohort which combined interview, self-report and ICD code data. Similarly, for SB, the variance explained in SB by SB PRS ranged from 0.55-2.88% in cohorts using only interview data, 1.09-1.41% in cohorts using self-report and/or interview, and 1.60% in the MVP cohort. In further support of minimal bias due to heterogeneity in phenotyping methods, we stratified cohorts by phenotyping method and conducted GWAS within each group. The resulting post-hoc GWAS and their genetic correlations indicated no significant differences across phenotyping methods (see Supplementary Note).

### Fine-mapping and gene prioritization implicate putative causal SNPs and credible genes for SI and SB

Fine-mapping was performed on GWS loci for SI, SD (EUR) and SB using GWAS summary statistics by applying complementary methods: SuSiE^29^ (statistical fine-mapping), PolyFun+SuSiE^30^ (functional fine-mapping), and SuSiEx^31^ (multi-ancestry fine-mapping). At a posterior inclusion probability (PIP) threshold >0.5, we identified nine putative causal SNPs for SI, none for SD, and 18 for SB (Supplementary Table 24). These SNPs were mapped to 14 genes using Variant Effect Predictor, based on their genomic position relative to annotated transcripts and regulatory features (Fig. 4).

**Figure 4.**
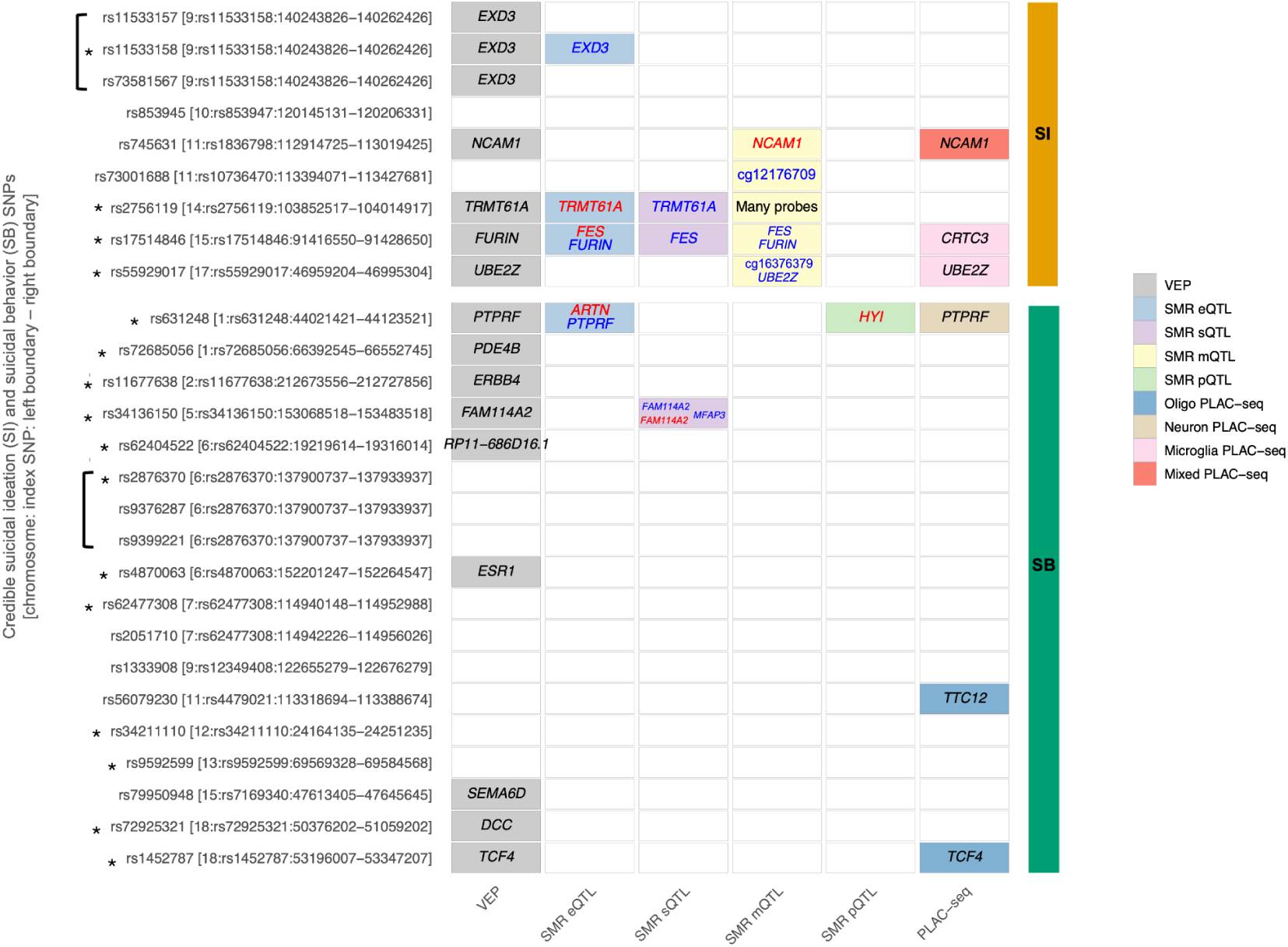
Gene prioritization results for suicidal ideation and suicidal behavior. The y-axis displays credible SNPs for suicidal ideation (SI) and suicidal behavior (SB), followed by the genome-wide significant locus labelled as [chromosome number:index SNP:left range: right range], with ranges denoting the fine-mapping windows. The asterisk symbol (*) indicates credible SNPs that are also the index SNP in the locus. Multiple credible SNPs from the same locus are enclosed in a square bracket. The x-axis displays the results from various gene prioritization analyses, including VEP annotations, SMR analyses of brain eQTL, sQTL, mQTL and pQTLs, and the integration of PLAC-seq data. Methylation probe IDs were mapped to genes using the EWAS atlas. For SMR analyses, gene and probe names are color coded as blue or red based on the effect direction of the risk allele on the molecular feature. The label ‘Many probes’ is assigned to credible SNPs for which multiple methylation probes are implicated via SMR analyses. The label ‘Mixed PLAC-seq’ indicates cases where evidence for a SNP came from more than one PLAC-seq cell type (Neuron and Oligo PLAC-seq).

Additionally, we applied an integrative quantitative trait locus (QTL)-based strategy, leveraging fine-mapped SNPs for SI and SB as instruments in Summary data-based Mendelian randomization (SMR) analyses^32^, including brain QTLs for expression (eQTLs)^33^, splicing (sQTLs)^33^, methylation (mQTLs)^34^ and protein abundance (pQTLs)^35^. This approach identified 11 credible genes (Fig. 4, Supplementary Table 25). We also mapped SNPs to target genes using PLAC-seq chromatin interaction data from brain cell-types^36^, which additionally prioritized *TTC12* and *CRTC3* (Fig. 4). The credible genes identified have roles in neurodevelopmental and signaling processes, among other functions (Supplementary Table 26). We investigated whether the protein products of credible genes are drug targets using the Drug-Gene Interaction Database v4^37^ and Connectivity Map^38^. For SI, we found several drug-gene pairings, including *NCAM1*, a target of the serotonin-norepinephrine reuptake inhibitor duloxetine, *KLC1 and FES*, both targets of cancer therapies, and *FURIN*, which is targeted by the anti-inflammatory drug pirfenidone (Supplementary Table 27). For SB, we found three main drug targets: *PDE4B* (an anti-inflammatory target), *ERBB4* (a tyrosine kinase receptor often targeted by cancer drugs), and *ESR1* (an estrogen receptor target) (Supplementary Table 27).

### Enrichment analyses reveal neurobiological underpinnings of suicidality

Gene set enrichment analyses were conducted using MAGMA^39^ and GSA-MiXeR^40^ to identify biological pathways associated with suicidality phenotypes. No gene sets reached significance for SI or SD. MAGMA analysis of SB identified six significantly enriched gene sets, primarily pointing to synaptic and neuronal structure and function (Supplementary Table 28).

GSA-MiXeR results indicated 13 gene sets enriched for SB (Akaike information criterion (AIC) > 0; Supplementary Table 29), although leave-one-gene-out analyses revealed that almost all were driven by single, highly enriched genes, suggesting limited contributions from the remaining genes in the set (Supplementary Table 29). Amongst these single highly enriched genes driving associations were *ESR1* and *PDE4B*, both of which were also identified as credible genes and drug targets.

MAGMA and GSA-MiXeR did not identify any of the same gene sets, with MAGMA-implicated gene sets being larger and broader and GSA-MiXeR-implicated gene sets being smaller and more specific, consistent with prior studies^40^. However, there was functional convergence in the gene sets implicated between the methods. For instance, the dopamine neurotransmitter receptor activity gene set identified by GSA-MiXeR represents a molecular function that occurs in the postsynapse, synaptic membrane, and postsynaptic membrane cellular component gene sets identified by MAGMA, according to Gene Ontology (https://geneontology.org/).

We also performed tissue-specific gene expression enrichment analyses using MAGMA via FUMA^41^. Among 30 general GTEx tissue types^42^, significant enrichment was observed for SI and SB in brain and pituitary tissues, while SD showed enrichment only in brain tissue (all p < 0.00167) (Supplementary Table 30). Similar analysis of 54 specific GTEx tissue types revealed widespread enrichment across brain regions (Supplementary Table 31).

To identify cell-type specific enrichment by brain region, we performed stratified LDSC^43^ to estimate *h^2^*enrichment in the top decile of expression proportion (TDEP) genes across 31 brain cell type superclusters and 461 brain cell type clusters, as previously described^44,45^. SI showed significant heritability enrichment in four neuronal cell-type superclusters: amygdala excitatory, medium spiny neuron, miscellaneous, and thalamic excitatory (Fig. 5B, Supplementary Table 32). No cell-type superclusters showed significant enrichment in the analysis of SD. For SB, four neuronal superclusters were enriched: amygdala excitatory, caudal ganglionic eminence (CGE) interneuron, eccentric medium spiny neuron, and hippocampal CA1-3 (Fig. 5, Supplementary Table 32).

**Figure 5.**
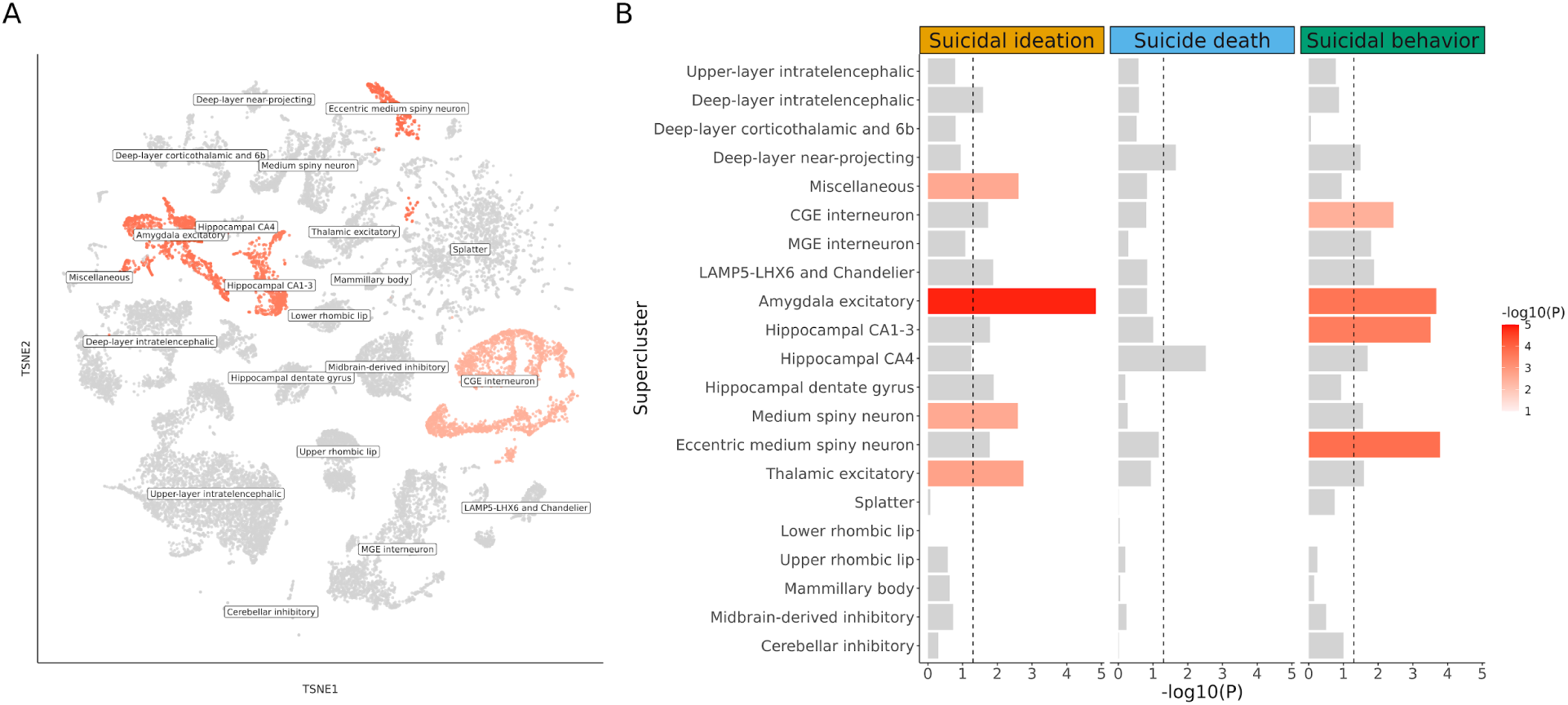
Supercluster-level *h^2^* enrichment for suicidality phenotypes. **A)** t-Distributed Stochastic Neighbour Embedding (tSNE) plot for suicidal behavior (naming convention and source of the single-cell data from Siletti et al.^45^) colored by -log10(P) enrichment values. Grey indicates non-significantly enriched superclusters (false discovery rate > 0.05). **B)** Bar plot showing enrichment significance for 21 neuronal superclusters for suicidal ideation (yellow), suicide death (blue), and suicidal behavior (green). For all phenotypes, no enrichment was observed in non-neuronal cell clusters (Supplementary Table 32).

## DISCUSSION

This study represents the largest and most ancestrally diverse GWAS of suicidality phenotypes to date. We identified 77 unique loci, including 59 previously unreported, representing a marked expansion of the known genetic architecture for suicidality. Consistent with prior literature^19,22^, *h^2^* was highest for SA (6.6%) and SB (6.7%). We also observed significant *h^2^* for SI (3.0%) and SD (5.1%), reinforcing their polygenic nature. Notably, *h^2^*estimates were not significant in the AFR, EAS, or LA GWAS, and low mean χ^2^ estimates reflect persistent disparities in power, likely due to sample size imbalances across these groups and the limited performance of existing reference panels for admixed populations.

Our results demonstrate both shared and distinct genetic etiologies across suicidality phenotypes. Many GWS loci overlapped between the SA and SB meta-analyses (Fig. 1E) and indeed they were based on mostly similar component GWAS. Nonetheless, the SB GWAS uncovered 22 additional loci not identified in SA, highlighting the potential for additional discovery through larger sample sizes^46^. The largest previous SB GWAS identified 12 GWS loci^22^, of which nine were GWS, two were near GWS (p < 1.35×10^-6^), and one had a minimum p-value of 0.005 in the current European or multi-ancestry SB GWAS. The SI GWAS yielded fewer GWS loci, but still revealed partial overlap with loci implicated in SA and SB. In contrast, we identified only two GWS loci in the EUR SD GWAS, neither of which has been reported in previous suicidality GWAS, nor emerged in the current GWAS of SI, SA, or SB. While a previous SD GWAS^23^ identified six variants across two loci on chromosomes 13 and 15, these loci were not associated with SD in the current study (p>0.01). This discrepancy may reflect greater heterogeneity introduced by combining multiple cohorts in meta-analysis, and heterogeneity among SD cases, given recent evidence for distinct genetic profiles in those with versus without prior suicidality^47^. While the limited detection of GWS loci in the SD GWAS is likely a result of limited statistical power due to smaller case numbers, the specificity of these loci may also suggest distinct genetic risk factors for SD.

Further characterizing the genetic architecture of suicidality, we observed strong genetic correlations among SI, SD, and SB, although all were significantly below 1, supporting prior evidence that these phenotypes, while related, are genetically distinguishable and may reflect different stages or pathways in the suicidality continuum^11,48–50^. Notably, while the genetic correlations of SD with SI and SB were high, both were lower than the correlation between SI and SB, reinforcing the interpretation of genetic specificity in SD. This pattern further supports the possibility that SD captures additional, partially distinct liability pathways (potentially including genetic influences related to physical health and non-psychiatric conditions) that are less central to non-fatal suicidality^12,51^. These findings also indicate that, although SI and SB share some genetic risk factors, their biological etiology may be partially distinct, as evidenced by differences in *h^2^_SNP_*and GWS loci between them, along with the genetic correlation significantly < 1. For SI, the GWAS results may additionally be influenced by the complexity and heterogeneity of the phenotype, which represents an internal cognitive process that ranges in severity and duration.

Complementary fine-mapping and gene mapping approaches prioritized seven and 13 credible genes for SI and SB, respectively. None of the GWS loci that overlapped between SI and SB were successfully fine-mapped for both, and thus we could not determine if the two phenotypes share putative causal SNPs or credible genes. Ten prioritized genes had been previously associated with suicidality phenotypes^19,21,22,25^, for example *ESR1* and *FURIN*. The ten previously unreported genes included *CRTC3*, *NCAM1*, *TRMT61A*, and *UBE2Z* for SI and *ERBB4*, *FAM114A2*, *HYI*, *MFAP3*, *PTPRF,* and *TCF4* for SB, several of which have been associated with other psychiatric and behavioral phenotypes (Supplementary Table 26). For example, *NCAM1*, *PTPRF*, and *TCF4* were identified in GWAS of impulsivity-related phenotypes^52^, and *NCAM1* and *EXD3* have been implicated in externalizing behavior^53,54^ and borderline personality disorder^55^, respectively. Additionally, *ESR1* and *FURIN* were credibly associated with bipolar disorder and MDD in similar gene prioritization analyses^56,57^.

To further contextualize the biological underpinnings of suicidality, we conducted enrichment analyses spanning gene set, tissue and cell-type expression. Collectively, these analyses converged on neuronal pathways and brain-expressed genes as key contributors to suicidality phenotypes. Gene set enrichment analyses using MAGMA implicated broad synaptic gene sets (consistent with interactome evidence^58^) in SB, while GSA-MiXeR highlighted more targeted processes, such as dopaminergic signaling. Many of the GSA-MiXeR-results appeared to be driven by a small number of highly associated genes, and should be interpreted with caution, as the enrichment may reflect a single gene rather than involvement of the entire pathway.

SI and SB both showed significant enrichment in genes highly expressed in pituitary and brain tissues, and more granular analyses of brain cell-types revealed heritability enrichment across multiple neuronal populations. Enrichment was absent in non-neuronal superclusters, consistent with results for other psychiatric phenotypes^44^. For example, SI showed enrichment in the amygdala excitatory, medium spiny neuron, and thalamic excitatory superclusters, while SB also showed enrichment in amygdala excitatory, along with the CGE interneuron, eccentric medium spiny neuron, and hippocampal CA1-3 superclusters. Excitatory and medium spiny neurons are closely aligned with glutamatergic and dopaminergic signaling pathways, respectively, which have long been central to psychopharmacology^59,60^. The hippocampal CA1-3 enrichment may additionally point toward involvement of neuronal populations critical for memory formation. Memory processes may be especially relevant to suicide risk; epigenetic alterations in the hippocampus of suicide decedents have been linked to disrupted expression of genes involved in memory^61^, which may also impair processes like problem-solving and stress regulation that protect against suicidal behavior^62^. Furthermore, neuroimaging studies of suicidal ideation have repeatedly implicated limbic regions, including the hippocampus and amygdala^63^.

Prior work has shown that many of the neuronal populations implicated here are concentrated in specific brain regions^44^, such that cell-type enrichment may also reflect brain region involvement. Most of the enriched cell types are primarily found in subcortical structures, such as the limbic system (i.e., hippocampus, amygdala, and hypothalamus), basal forebrain, thalamus, and midbrain^44^. These cell types are not unique to suicidality and are also enriched across multiple psychiatric phenotypes. Instead, suicidality phenotypes appear distinguished by a relative absence of enrichment in cell type superclusters primarily located in the cortex that are otherwise broadly implicated in psychiatric disorders^44^. This pattern may indicate differences in neurobiological mechanisms underlying suicidality and suggest that dysregulation in processes more closely tied to subcortical circuity – e.g., impulsivity, emotion processing, stress reactivity, and threat detection – may play a more critical role in suicidal crises^64–66^ than cortical processes broadly associated with psychiatric liability.

Polygenic prediction of SI has historically been limited, with SI PRS explaining just 0.2% of phenotypic variance^21^. Here, using multi-ancestry PRS derived from our better-powered GWAS, we observe modest improvement, with SI PRS explaining ∼0.5% of the variance (Fig. 3). PRS generally performed better for SB, with an average *R*^2^ of ∼1.2% and a maximum of 2.9% in a single cohort (previous maximum *R*^2^ ∼1.1%)^22^. These improvements likely reflect larger discovery GWAS and the advantages of multi-ancestry PRS methods^28^. In contrast, the weaker performance of the SD PRS is likely due to smaller discovery GWAS sizes and limited ancestral diversity. Notably, these PRS alone are not adequate for individual-level clinical prediction or risk stratification. However, with future advances, they may provide clinical utility when combined with complementary measures of risk derived from electronic health records^67^ and other data modalities. At present, the greatest utility of PRS lies in research to identify genetic contributions to specific facets of suicidality^47,68^, or examine associations between genetics and other risk factors^69,70^ or changes in brain morphology^71^.

We also examined genetic correlations between suicidality phenotypes and a broad set of psychiatric and behavioral traits. Compared to the genetic correlations between SI, SD, and SB, we observed even lower correlations with non-suicidal self-injury, pointing to a genetic component potentially unique to suicidal intent. Consistent with previous evidence that internalizing disorders are key contributors to suicidality at both the phenotypic and genetic levels^10,19,72,73^, MDD, anxiety, and PTSD showed some of the strongest genetic correlations with suicidality. SB, in particular, showed stronger correlations with externalizing behavior^53,54^ and associated traits, such as problematic substance use and ADHD, than SI. This is consistent with the role of behavioral disinhibition in the transition from ideation to action^74–77^. We did not exclude cases or controls based on psychiatric diagnoses to avoid bias from comorbidities; however, individuals missing suicidality history were excluded if they had a psychiatric disorder, to avoid misclassification. While this approach maximizes inclusivity and power, it may also inflate the observed genetic correlations with psychiatric disorders. Still, no genetic correlations with psychiatric disorders suggested total genetic sharing, reinforcing the view that genetic risk underlying suicidality cannot be fully explained by genetic liability to psychiatric disorders^19,78^. Several other limitations of this study should be acknowledged. First, we analyzed SI, SA, SD, and SB as separate phenotypes, despite their shared features and the increased power that analyzing these phenotypes together might provide. However, this was a deliberate design choice aimed at generating high-quality, specific GWAS that will provide a valuable resource to the broader psychiatric genetics community^7^. Future work from PGC SUI will involve meta-analytic and multivariate methods, like genomic structural equation modeling^79^, to formally test the extent of genetic overlap and specificity both across suicidality phenotypes and between suicidality phenotypes and psychiatric disorders. Second, we were unable to perform “nested” GWAS and thus cannot fully disentangle the genetic contributions to progression between phenotypes. For example, a GWAS of SB restricted to individuals with SI, which would enable comparisons between those who experience SI without acting on it, and those who progress to action. Similarly, we also could not parse potential differences in the pathways that lead to SA versus SD. Third, we did not perform age- or sex-stratified GWAS; however, these analyses are of interest to PGC SUI and will be examined in future work. Fourth, as is common in large-scale meta-analyses, cohorts included in this study differed in data collection strategies. While this enables larger and more inclusive GWAS, it also has the potential to introduce phenotypic variability. However, examination of PRS performance and genetic correlations across different phenotyping methods did not reveal notable differences, suggesting that variance in phenotyping method did not meaningfully influence the primary findings. Finally, although fine-mapping and enrichment analyses identified credible genes associated with suicidality risk and promising biological pathways, these findings should be investigated in functional experiments to validate mechanisms.

In conclusion, this study represents the largest and most ancestrally diverse GWAS of suicidality phenotypes to date. Fine-mapping and enrichment analyses highlight neuron-specific expression patterns and implicate subcortical brain regions and synaptic signaling pathways in the etiology of suicidality. Differences in heritability, genetic correlation, and predictive performance across SI, SA, SD and SB support both shared and phenotype-specific genetic influences. These findings provide a foundational resource for future functional, translational, and cross-trait studies aimed at understanding the biological factors underlying suicide risk. Ultimately, biological insights represent just one component of suicidality risk, and their integration with clinical and environmental factors will be critical for informing and developing prevention efforts.

## METHODS

### Cohorts and phenotype definitions

For each cohort, we provide detailed information on sample size, genetic ancestry/population, ascertainment methods, phenotyping approach, type of data received, and ethical approvals summarized in Supplementary Table 1 and described further in the Supplementary Note. To support harmonization across cohorts, each study followed a previously published phenotyping protocol by PGC SUI^7^ to guide the construction of case and control definitions. We classified suicidality phenotypes into three primary categories based on standard definitions^13^: 1) SI, wherein an individual contemplates ending their own life with or without a specific plan; 2) SA, wherein an individual takes action to cause harm to themselves with at least some intent to die; 3) SD, or death caused by intentional action to take one’s own life.

Inclusion and exclusion criteria used for cases and controls in each GWAS were designed to maximize phenotypic specificity, while minimizing potential misclassification (Table 1, Supplementary Figure 1). A full description of specific case/control criteria and the rationale behind these decisions can be found in the PGC SUI phenotyping protocol^7^. Briefly, cases were required to be assessed for the specific suicidality phenotype being studied, rather than inferred by proxy based on a more severe suicidality phenotype to preserve phenotypic specificity in each GWAS (Table 1). The SI GWAS included SI cases (some of whom may also meet criteria for SA or SD) and controls without evidence of SI, and without evidence of SA, or SD as a safeguard against potential misclassified controls due to undocumented SI. The SA GWAS comprised non-fatal SA cases (some of whom may later have died by suicide) and controls who did not have evidence of SA or SD, but who may have reported SI. Our definition of SA required at least some intent to die, such that non-suicidal self-injury was not classified as SA. The SD GWAS included SD cases and controls who did not die by suicide or have evidence of SA, as a safeguard against misclassification, since most controls were still living. In addition to these individual phenotypes, we also analyzed an aggregated phenotype, SB, defined here to include SA or SD. Thus, the SB GWAS combined SA and SD cases and used controls without evidence of either. Psychiatric diagnoses were not used as exclusion criteria for cases or controls in any GWAS to avoid bias related to comorbid psychiatric conditions. As an exception, if an individual was missing information on the suicidality phenotype of interest and also had a known psychiatric diagnosis, they were removed from the study to minimize bias from phenotype misclassification. In the Supplementary Note, we provide examples to demonstrate that additionally screening for psychiatric disorders in individuals not directly assessed for the suicidality phenotype of interest improves the informativeness of the study. SI and SA data were obtained through a variety of sources, including structured psychiatric interviews, self-report questionnaires, and International Classification of Diseases (ICD) codes, or combinations of these sources (Supplementary Table 1). Because differences in phenotyping methods across cohorts could introduce heterogeneity, we conducted sensitivity GWAS meta-analyses of SI and SA stratified by phenotyping method in EUR samples and tested whether genetic correlations between them were significantly < 1. We observed no significant differences across phenotyping methods and results are presented in the Supplementary Note and Supplementary Note Table 4. SDs were identified using cause of death codes, Coroners’ reports, or Medical Examiner’s reports (Supplementary Table 1).

If a study included individuals from multiple genetic ancestries or populations, individuals were grouped according to the ancestry or population they were most genetically similar to, and here are referred to as distinct analytic cohorts. Groups were determined in each cohort using principal component analysis and reflects which population (from the reference panel used by each cohort) an individual was most genetically similar to. In total, the analyses included 37 SI cohorts (N_Cases_ = 259,747 and N_Controls_ = 1,309,943), 46 SA cohorts (N_Cases_ = 64,993 and N_Controls_ = 1,269,037), 49 SB cohorts (N_Cases_ = 75,300 and N_Controls_ = 1,311,895), and 7 SD cohorts (N_Cases_ = 9,197 and N_Controls_ = 668,162) (Table 2). The current SB meta-analysis represents an increase of 31,492 cases not included in the previous GWAS meta-analysis of SB^22^.

### Genotyping, quality control, imputation, and analysis

Studies either securely transferred individual-level genotype and phenotype data to the PGC server for central analysis or shared GWAS summary statistic data (Supplementary Table 1). All samples underwent standard genotyping performed by the collaborating research teams, the details of which are provided in the Supplementary Note. Where individual-level genotype data were provided, quality control (QC) and imputation were performed using the Rapid Imputation for COnsortias PIpeLIne (RICOPILI) developed by the PGC and described in detail previously^80^. In summary, QC parameters for retaining SNPs and subjects were: SNP missingness < 0.05 (before sample removal), subject missingness < 0.02, autosomal heterozygosity deviation (F_het_) < 0.2, SNP missingness < 0.02 (after sample removal), difference in SNP missingness between cases and controls < 0.02 and SNP Hardy-Weinberg equilibrium (*P* > 10^−10^ in cases, *P* > 10^−6^ in controls). When sample overlap or relatedness was suspected, relatedness was calculated within or across cohorts using identity by descent, and one of each pair of related individuals (pi_hat > 0.2) was excluded, with a preference for retaining cases. Genotype imputation was performed using the prephasing/imputation stepwise approach implemented in Eagle (v2.3.5; https://alkesgroup.broadinstitute.org/Eagle/)^81^ and Minimac3 (v2.0.1; https://genome.sph.umich.edu/wiki/Minimac3)^82^ to the Haplotype Reference Consortium (HRC) reference panel (v1.1)^83^. GWAS were then performed separately in each cohort and genetic ancestry or population group using PLINK 1.9 (v1.90b6.4)^84^ by comparing imputed marker dosages under an additive logistic regression model between cases and controls, covarying for the first five ancestry principal components (PCs) and only additional PCs if they were associated with case-control status. Studies contributing GWAS summary statistics performed QC, imputation, and GWAS locally using comparable procedures (see the Supplementary Note for details). Studies submitting GWAS summary statistics sometimes strayed from the standard analytic procedure to account for unique components of their study design. For example, some family-based cohorts used GWAS methods that account for relatedness within the model, such that related individuals were retained and analyzed in the sample (see the Supplementary Note for details).

### GWAS quality control and meta-analysis

GWAS summary statistics from all cohorts underwent standard QC procedures and data cleaning after being transferred to the PGC SUI analytic team. First, SNPs were filtered from the GWAS summary statistics from each cohort using sample minor allele frequency (MAF) < 1% and sample MAF corresponding to a minor allele count (MAC) < 10 in cases or controls (whichever had smaller N), in order to control test statistic inflation at low MAFs from small cohorts. We then checked summary statistics files for missingness, duplicate SNPs, correct build (hg19), and test statistic values being reported on the appropriate scale. Any errors or inconsistencies found were remedied before moving on to allele frequency checks. SNP allele frequencies from the GWAS summary statistics were compared against allele frequencies from the (appropriate ancestry/population-specific) 1000 Genomes Project (1KG)^85^ and HRC reference panels^83^. If a SNP had an allele frequency difference > 0.20 between the summary statistics and both of the reference panels, it was removed. Finally, we used DENTIST^86^ to identify and remove SNPs with errors in the test statistics or heterogeneity between the GWAS and reference.

After each individual GWAS passed cleaning and QC, we conducted multi-ancestry meta-analyses of each phenotype using an inverse variance-weighted fixed-effects model in METAL^87^, implemented using RICOPILI^80^. We also conducted ancestry/population-specific meta-analyses when GWAS summary statistics for a specific ancestry/population and phenotype were available from more than one cohort. We defined GWS loci as regions surrounding SNPs with p-values < 5 × 10^-8^ with LD *r^2^* > 0.1, within a 3,000 kb window based on the LD structure from the HRC reference panel, using the EUR panel for multi-ancestry meta-analyses and the ancestry/population-matched reference panel for ancestry/population-specific meta-analyses.

To determine if a GWS locus identified in the current analysis was previously associated with suicidality, or was overlapping amongst suicidality phenotypes, we used the intersect function from bedtools (v2.31.1)^88^ to check for overlapping regions. To determine which GWS loci in the current analysis were previously unreported, we checked for overlap with GWS loci identified in seven different previous GWAS of various suicidality phenotypes^19–23,25,26^.

For all post-GWAS analyses, unless otherwise specified, we used the multi-ancestry GWAS of SB, SI, and SD with a filter applied to remove SNPs with a per-SNP effective sample size (N_eff_) more than 3 standard deviations below the mean per-SNP N_eff_. For follow-up analyses requiring an LD reference panel, we used a EUR LD reference panel with multi-ancestry GWAS. To assess the appropriateness of this reference panel, we followed the approach outlined by Yengo et al. (2022)^89^, which involves evaluating various population stratification metrics.

Specifically, we examined heterogeneity (Cochran’s Q-statistic), the fixation index of genetic differentiation (*F*_ST_) across SNPs^90,91^, and the LDSC attenuation ratio^24^. For all phenotypes, the mean Q-statistic across SNPs in the multi-ancestry GWAS indicated only moderate heterogeneity across ancestry and population groups (5-38%) (Supplementary Table 33). The mean *F*_ST_ between the EUR and multi-ancestry GWAS SNPs was low for all phenotypes (0.0003-0.003), consistent with values observed between populations of the same genetic ancestry^89^. Finally, all LDSC attenuation ratios calculated using the 1KG EUR LD scores were below 20%, indicating a good match between the EUR LD reference and the multi-ancestry GWAS^24^ (Supplementary Table 33). Taken together, these results supported the use of the EUR reference as an approximation for analyses involving the multi-ancestry GWAS.

### Linkage Disequilibrium Score Regression

We used LDSC^24^ to estimate the *h^2^* of SI, SA, SD, and SB from ancestry/population-specific and multi-ancestry GWAS using ancestry matched LD scores (Supplementary Table 16). We converted *h^2^* to the liability scale assuming lifetime prevalence of 0.2% for SD^23^, 2% for SA/SB (median of the range reported worldwide)^92^, and 9% for SI^92^. We acknowledge that prevalence may vary across populations, and as such *h^2^*estimates on the liability scale could be over- or underestimated, although the significance of the estimate is unaffected. We also used LDSC to estimate genome-wide genetic correlations^27^ between the suicidality phenotypes and other phenotypes previously shown to have genetic relationships with suicidality (described in detail in Supplementary Table 16). For the non-suicidality phenotypes, we used multi-ancestry GWAS when available and when the LDSC statistics indicated a good match to the EUR LD reference panel (as described above). For all other non-suicidality phenotypes, we used European ancestry GWAS. The genetic ancestry of the non-suicidality phenotypes used are provided in Supplementary Table 16. To account for multiple testing, we applied the Benjamini-Hochberg procedure, considering correlations with FDR p < 0.05 as statistically significant; this threshold was applied as the maximum acceptable FDR across all genetic correlation analyses. To test for a significant difference between two genetic correlations, we followed the method from Coleman et al.^93^ which applies a block jackknife.

### Polygenic Risk Scoring

We used PRS-CSx (version from Nov 21, 2024)^28^ to compute multi-ancestry meta-analyzed posterior effect size estimates and generate multi-ancestry PRS for suicidality phenotypes in 25 target cohorts. For each target cohort, we used a leave-one-out approach, excluding that cohort from the discovery GWAS used to calculate the PRS. PRS-CSx jointly models ancestry/population-specific GWAS summary statistics using a shared continuous shrinkage prior, enabling correlated yet varying effect sizes across ancestries/populations. Thus, for discovery GWAS we used ancestry/population-specific GWAS for as many genetic ancestry/population groups as available for each phenotype. PRS-CSx improves effect size estimation through sharing information between GWAS, leveraging LD diversity, and accounting for population-specific allele frequencies. We used the “--auto” option to automatically infer model parameters, and then combined population-specific posterior effect size estimates using the“--meta” option^28^. Associations between PRS and suicidality phenotypes were tested separately within each cohort using logistic regression (via the glm() function in R^94^), with the PRS standardized (mean = 0, standard deviation = 1) and the same covariates used in that cohort’s GWAS. We converted the variance explained by the PRS (*R*^2^) to the liability scale using the method from Lee et al. (2012)^95^ to account for the proportion of cases in the target cohort and the population prevalences from above. We calculated 95% CIs for the liability scale *R*^2^ using non-parametric bootstrapping (10,000 resamples) via the boot package in R^96,97^. We also calculated ORs comparing individuals in the top quintile/decile of the PRS distribution to those in the middle quintile/decile, using standardized residuals of the PRS (adjusted for the same covariates used in the logistic regression). We calculated AUC statistics using the pROC package in R^98^ with models including only the scaled PRS as a predictor following standard PGC procedures^80^. To ensure consistency across cohorts, PRS weights were centrally generated on the PGC server and subsequently distributed to study analysts along with standardized analysis scripts (https://github.com/sarahcolbert/PGC_SUI_PRS). Finally, the *R*^2^s, ORs, AUCs, and their CIs were meta-analyzed using effective sample size weighting to calculate N_eff_-weighted averages within and across genetic ancestries/populations.

### Fine-mapping

We conducted statistical and functional fine-mapping of GWS loci for SI, SB, and SD (EUR). The fine-mapping windows were defined as regions surrounding SNPs with p-values < 5 × 10^-8^ with LD *r^2^* > 0.6, within a 3,000 kb window based on the LD structure from the HRC EUR reference panel (v1.1)^83^; and loci in the MHC (chromosome 6: 28,000,000–34,000,000, GRCh37) were excluded due to the complexity of the LD structure in this region. After these filters, we fine-mapped 12 GWS loci from SI multi-ancestry, 51 loci from SB multi-ancestry, and two loci from SD EUR. We employed statistical fine-mapping using SuSiE^29^ and functional fine-mapping with Polyfun+SuSiE^30^, both implemented via the SAFFARI pipeline^99^, to the multi-ancestry SI and SB results and to the EUR ancestry SD results. LD estimates were obtained from the HRC EUR reference panel, and the functional annotations for Polyfun+SuSiE were from the baseline-LF2.2 UK Biobank model^100^. We restricted the maximum number of causal variants per fine-mapped region to the default of five (K=5)^29^, and performed single variant fine-mapping (no LD), assuming one causal variant in each locus (K=1), as this approach does not require population-accurate estimation of LD. Additionally, we applied SuSiEx^31^, a multi-ancestry fine-mapping method, leveraging ancestry/population-specific meta-analysis results and the corresponding ancestry/population LD reference panel from the HRC. For both SI and SB, the SuSiEx analyses included summary statistics from EUR, LA, AFR, EAS, and SAS GWAS. Putative causal SNPs were defined as those with PIP > 0.5 and included in the 95% credible set for the multivariant analyses (K=5) and as those with PIP > 0.5 for the single variant analysis.

### Gene prioritization analyses

Fine-mapped SNPs were mapped to genes through variant annotation using the Ensemble Variant Effect Predictor (VEP; GRCh37, Ensembl release 114, https://www.ensembl.org/info/docs/tools/vep/index.html). Furthermore, we integrated brain quantitative trait loci (QTL) data to link fine-mapped variants to molecular features and elucidate potential molecular mechanisms by which they increase risk of SB or SI. Data on eQTLs and sQTLs were obtained from the BrainMeta study (version 2), which comprised RNA-sequencing data of 2,865 brain cortex samples from 2,443 unrelated individuals of EUR ancestry with genome-wide SNP data^33^. Data on mQTLs were obtained from the BrainMeta study^34^, a meta-analysis of adult cortex or fetal brain samples, comprising 1,160 individuals with methylation levels measured using the Illumina HumanMethylation450K array. We also used pQTLs from a study of brain proteomes of 1,277 individuals of EUR ancestry primarily from the dorsolateral prefrontal cortex^35^. Summary data-based Mendelian randomization (SMR; v1.3; https://yanglab.westlake.edu.cn/software/smr/)^32^ with subsequent heterogeneity in dependent instruments (HEIDI) tests was performed for each QTL dataset using the fine-mapped SNPs as the QTL instruments. Using the multi-ancestry SB and SI GWAS and QTL summary statistics, each putative causal SNP was analyzed as the target SNP for probes within a 2-Mb window on either side using the --extract-target-snp-probe option in SMR. The EUR HRC LD reference panel was used for the analyses of the multi-ancestry meta-analysis. SNP-QTL probe combinations were filtered according to the following criteria: *P_QTL_* < 5×10^-8^ and the significance threshold for the HEIDI test was *P_HEIDI_* ≥ 0.01. To improve the robustness of our SMR findings, we repeated our SMR analysis using the European GWAS meta-analyses for SI and SB, while keeping the remaining inputs and filtering criteria unaltered. This analysis further solidified our findings while showing cross-ancestry generalizability (Supplementary Table 25).

Furthermore, we annotated SNPs to genes using PLAC-seq data from brain cell-types^36^, providing an additional layer of chromatin interaction-based evidence. Specifically, genes were annotated when either end of a chromatin interaction loop was linked to them, using previously published epigenomic data from purified bulk H3K27ac and H3K4me3 ChIP–seq of brain cell-types^36^.

### Enrichment analyses

#### Gene, gene set, and tissue enrichment analyses

We conducted gene, gene set and tissue-level enrichment analyses to explore the biological underpinnings of suicidality phenotypes. Gene and gene set analyses were conducted following previously established methods^39,40^. Briefly, associations between each suicidality phenotype and genes or gene sets were assessed using a SNP-wise mean model with a ± 10-kb window implemented in MAGMA^39^. Gene sets were obtained from Molecular Signature Database (MsigDB) v7.0^101^ and 7,167 gene sets were tested after removing those with < 10 genes. Furthermore, we also used GSA-MiXeR^40^ to assess gene set enrichment for 10,475 gene sets from the Gene Ontology^102^ and SynGO^103^ databases. A positive AIC value indicates that the full MiXeR model (with gene-specific contributions to the heritability) has a better fit than the baseline model (in which individual genes do not contribute) for that gene set. Tissue enrichment analyses were also conducted using MAGMA via FUMA^41^, testing for enrichment of association signals in genes expressed across 30 broad and 54 specific tissue types from GTEx v8^42^. Bonferroni correction was applied to account for multiple testing in all enrichment analyses and thresholds for specific analyses are reported in the Supplementary Tables.

#### Brain cell-type enrichment analysis

Following a previously published pipeline^44^, we applied stratified LDSC (s-LDSC)^43^ to estimate partitioned *h^2^* for the TDEP genes across brain cell types. In summary, cell types were defined using the Human Brain Atlas single-nucleus RNA sequencing dataset^45^, which includes over 3 million nuclei collected from post-mortem adult human brains, samples across 106 anatomical regions within 10 major brain areas. Cell types were organized into 461 clusters and 31 superclusters, for each of which we identified the top 10% of genes ranked by expression proportion. These gene sets were then used as the functional annotations in s-LDSC. Multiple testing was addressed using a FDR threshold of 5%.

#### Drug target analysis

After identifying plausibly associated genes (see ‘Gene prioritization analyses’), we linked them to known drug targets following a previously published pipeline that demonstrated enrichment across psychiatric disorders^104^. Briefly, drug targets were obtained from gene-drug interaction database v4^37^ and connectivity map^38^. We then constructed a Sankey network diagram based on genes that linked to targets of specific pharmaceuticals. The Sankey network is a two dimensional graph network where drugs that relate to more discovered genes or vice versa are shown to have larger edges in the overall model. Those edges are then plotted in two dimensions with the connections between gene and drug visualized as a graph. It should be noted that this approach does not evaluate whether the actions of these agents align with or counteract the effects associated with genetic liability.

## CODE AVAILABILITY

All software used is publicly available at the URLs or references cited. Code used to test PRS associations is available on GitHub (https://github.com/sarahcolbert/PGC_SUI_PRS).

## Data Availability

Summary statistics from these GWAS are available by application through the Psychiatric Genomics Consortium data access portal: (https://pgc.unc.edu/for-researchers/data-access-committee/data-access-portal/). Full summary statistics corresponding to the GWAS reported in this study are available for the SI and SA GWAS. For the SB and SD GWAS, the summary statistics available through the PGC portal exclude data from the Utah cohorts. Data from the Utah site will be available upon request from the administrative manager of the Utah Suicide Mortality Research Study (https://rge.utah.edu/usmrs.php) and will be contingent upon the permissions required by institutional regulations and state statutes.

## ACKNOWLEDGEMENTS

We thank the participants who donated their time, life experiences, and DNA to this research, and the clinical and scientific teams that worked with them.

This material is based upon work supported by the National Science Foundation Graduate Research Fellowship Program under Grant No. 1842169 (PI Colbert) and the National Institute of Mental Health R01 MH132733 (PI Mullins). The PGC is supported by grants to UCSD (R01MH124847), UNC (R01MH124871), MGH (R01MH124851), Mount Sinai School of Medicine (R01MH124839), Cardiff University (R01MH124873), Trinity College Dublin (R01MH124875) and Washington University St. Louis (R01DA054869). The content is solely the responsibility of the authors and does not necessarily represent the official views of the US National Institutes of Health.

We thank SURF (www.surf.nl) for the support in using the National Supercomputer Snellius. Statistical analyses were carried out on the NL Genetic Cluster Computer (http://www.geneticcluster.org) hosted by SURFsara and the Mount Sinai high performance computing cluster (http://hpc.mssm.edu), which is supported by the Office of Research Infrastructure of the National Institutes of Health (Grant Nos. <u>S10OD018522</u> and <u>S10OD026880</u>).

This research has been conducted using the UK Biobank Resource under Application Number 82087 (PI: J Coleman). This work uses data provided by patients and collected by the NHS as part of their care and support. This research was funded by the UK National Institute for Health and Care Research (NIHR) Maudsley Biomedical Research Centre (BRC). The views expressed are those of the authors and not necessarily those of the NIHR or the Department of Health and Social Care. This study included some publicly available datasets accessed through dbGaP (Psychiatric Genomics Consortium [PGC] bundle phs001254) and the Haplotype Reference Consortium reference panel v.1.0 (http://www.haplotype-reference-consortium.org/home).

Individual and cohort-specific funding acknowledgements are detailed in the Supplementary Note.

## AUTHOR CONTRIBUTIONS

N.M., S.M.C.C., A.R.D., and D.M.R. were responsible for overall study design and conceptualization with additional input from A.S.H. and E.T.M. The main GWAS analyses presented in the paper were conducted by S.M.C.C. with supervision from N.M. Fine-mapping was conducted by M.K. and N.M. Gene set enrichment analyses were done by K.S.O. and cell-type enrichment analyses were done by A. Harder and J.H.-L. Gene-drug pairing was performed by A.S.H. PRS analyses were led by S.M.C.C. with contributions from M. Stephenson, A.C.E., E.C.J., X.Q., M.J.K., L.T.I., A.E.A.-K., A.S.H., R.B., and A.A.S. Genetic correlation analyses were led by S.M.C.C. with contributions from S.S.-R., A.P., and L.V.-R. S.M.C.C. drafted the manuscript with supervision from N.M. M.K. and A.S.H. contributed substantially to the writing of the manuscript and many co-authors provided revisions and edits, including: L.A., O.A.A., A.E.A.-K., O.O.A., S.-A.B., C.H.D.B., B.T.B., A.D.B., D.L.B., M.B., B.C.-M., Z.C., B. Chaumette, H.C., D. Demontis, S.D., A.R.D., H.J.E., A.C.E., A.E., G.F., F.F.-A., A.J.F., G.R.F., J.M.F., M.E. Gaine, M. Galimberti, J.G.-A., M.E. Garrett, A.J.G., M.J.G., M.G.-S., J.G.-P., U.H., J.H.-I., L.T.I., A.I., S.J.-M., E.C.J., L.A.J., P.K.K., N.A.K., T.K., H.R.K., S.K., M.L., S.L., M.H.L., C. Liao, M.M., L.M., A.M.M., E.T.M., W.M., J.I.N., M.J.O., A.A.P., P.M.P., J.M.P., C.P., A.P., A.R., E.K.R., G.R., L.R., D.L.R., D.M.R., S.S.-R., A.R.S., C. Sawyers, S.W.S., P.R.S., J.W.S., M. Sokolowski, E.J.S.-B., A. Squassina, A. Starnawska, N.E.S., D.J.S., M. Stephenson, F. Streit, R.S., P.F.S., C. Toma, G.T., R.J.U., M.P.V., A.V.P., B.V., V.V., D.E.W., V.L.W., S.H.W., C.C.Z., and L.Z. Many individuals contributed to the individual cohorts included in this study. Cohort PIs: A. Agrawal, M.A., O.A.A., C.A., A. Arntz, J.C.B., S.B., K.B., J.M.B., R.B., D.I.B., A.D.B., R.A.B., J.A.C., S.C., J.R.I.C., H.C., W.E.C., U.D., D. Demontis, A.D.F., D.M.D., A.R.D., H.J.E., S.E., P. Falkai, P. Ferentinos, F.F.-A., J.M.F., M. Gawlik, J.G., P. Gorwood, H.J.G., M.J.G., M.G.-S., B.G., J.G.-P., E.R.H., S.C.H., V.M.H., A. Hishimoto, B.H., L.A.J., N.A.K., T.K., G.K., P.-H.K., M.L., M.H.L., K.L., C.M.L., Q.S.L., C. Lochner, H.H.M.M., M.M., B.M., N.G.M., L.M., A.M.M., S.F.M., A.M., S.E.M., P.B.M., R.M., W.M., M.M.N., J.I.N., R.A.O., I.O., M.J.O., P.M.P., J.C.P., G.P.P., B.W.J.H.P., N.P., R.E.P., B.P., D.P., M. Preisig, A.R., E.Z.R., M.E.R., S. Ripke, M.R., S. Roepke, D.M.R., G.A.S., A.R.S., C. Schilling, P.R.S., T.G.S., L.J.S., A. Serretti, T.S., M. Sokolowski, A. Squassina, D.J.S., M.B.S., F. Streit, G.T., R.U., E.V., J.B.V., B.V., D.E.W., T.W.W., D.C.W., S.H.W., H.-H.W., and C.C.Z. Cohort collection/genotyping: A. Agrawal, M.A., A. Arntz, C.A.B., S.B., J.M.B., K.J.B., A.B., R.A.B., T.M.B., M.B., B.C.-M., B. Carpiniello, J.A.C., M.J.C., S.C., H.C., W.E.C., N.D., C.C.D., D.M.D., D. Dikeos, S.D., A.R.D., H.J.E., S.E., P. Falkai, F.T.F., A.J.F., O.F., J.M.F., M. Gawlik, J.G., K.G.-S., P. Gorwood, H.J.G., M.J.G., M.G.-S., B.G., J.G.-P., M.H., U.H., J.H.-I., V.M.H., B.H., S.J.-M., L.A.J., J.H.K., D.K., N.A.K., T.K., G.K., J. Kraft, J. Kramer, H.R.K., P.-H.K., M.L., M.H.L., Y.K.L., Q.S.L., C. Liao, P.A.L., C. Lochner, H.H.M.M., J.M., M.M., J.J.M., B.M., N.G.M., L.M., A.M., S.E.M., E.M., G.M., W.M., A.I.N., M.M.N., Y.Z.N., J.I.N., S.O., C.M.O., R.A.O., M.J.O., P.M.P., G.P.P., J.M.P., B.W.J.H.P., R.E.P., B.P., D.P., M. Purushottam, A.R., E.Z.R., M.R., L.R., G.A.S., A.R.S., M.L.S., C. Schilling, C. Schmahl, P.R.S., T.G.S., O.B.S., M. Sokolowski, N.E.S., F. Stein, M.B.S., F. Streit, C. Terao, L.T., E.V., J.B.V., B.V., T.W.W., D.C.W., E.D.W., S.H.W., and C.C.Z. Data processing for a specific cohort: M.A., F.A., T.F.M.A., A.E.A.-K., S.A., P.B.B., C.A.B., S.B., K.B., J.M.B., R.B., A.D.B., K.J.B., A.B., R.A.B., M.B., B.C.-M., B. Carpiniello, Z.C., J.A.C., C.-E.C., M.J.C., J.R.I.C., H.C., W.E.C., D.C., U.D., F.S.D., D. Demontis, A.R.D., A.C.E., S.E., G.F., O.F., J.M.F., H.C.G., M.E. Garrett, K.G.-S., A.J.G., M.J.G., M.G.-S., P. Gupta, B.G., J.G.-P., S.H., A.S.H., U.H., V.M.H., B.H., L.T.I., Y.J., S.J.-M., E.C.J., L.A.J., L.J., J. Kang, J.H.K., M.J.K., J. Kim, D.K., G.K., S.K., D.L., M.T.L., M.L., S.L., M.H.L., B.-C.L., Y.K.L., D.F.L., Q.S.L., P.A.L., A.L., H.H.M.M., J.J.M., B.M., S.E.M., E.M., M.S.M., G.M., T.T.N., M.M.N., Y.Z.N., K.S.O., S.P., J.M.P., A.M.P.-G., R.E.P., C.P., G.P., X.Q., M.A.R., M.R., D.M.R., A.R.S., M.L.S., L.J.S., A. Shadrin, M. Sokolowski, A. Starnawska, F. Stein, M. Stephenson, F. Streit, C. Toma, S.V.d.A, T.W.W., S.H.W., H.-H.W., R.E.W., C.C.Z., J.Z., and L.Z. N.M., A.R.D., and D.M.R. supervised the overall study, with assistance from A.E.A.-K. and A.S.H. N.M., A.R.D., D.M.R. and A.E.A.-K. obtained funding for the Psychiatric Genomics Consortium Suicide Working Group to conduct this study.

## EXTENDED DATA

**Extended Data Figure 1.**
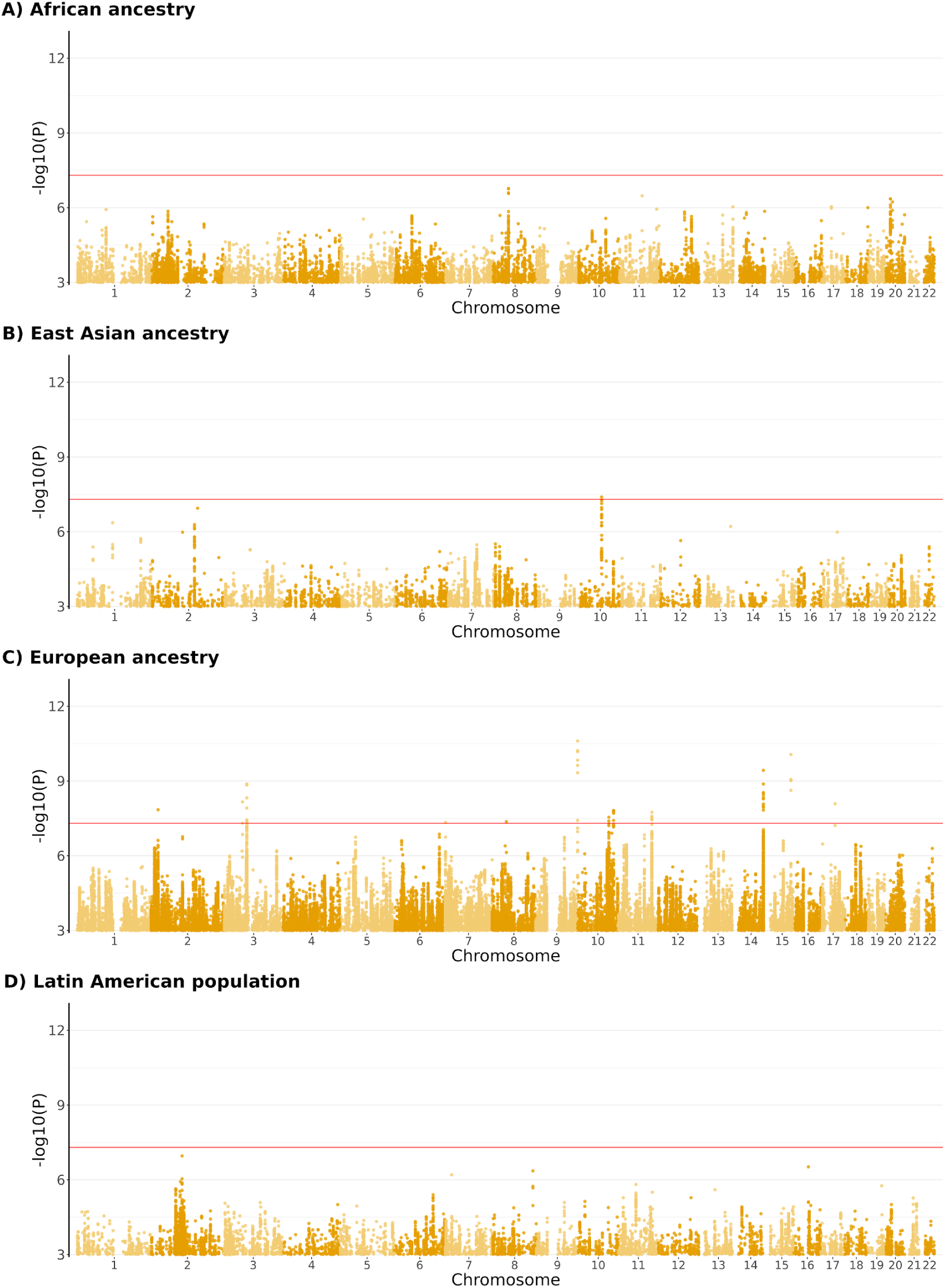
Manhattan plots of ancestry/population-specific suicidal ideation GWAS meta-analyses. The x-axis shows chromosomal position and the y-axis shows significance of the association as -log10(P). The red line shows the genome-wide significance threshold (p = 5×10^-8^).

**Extended Data Figure 2.**
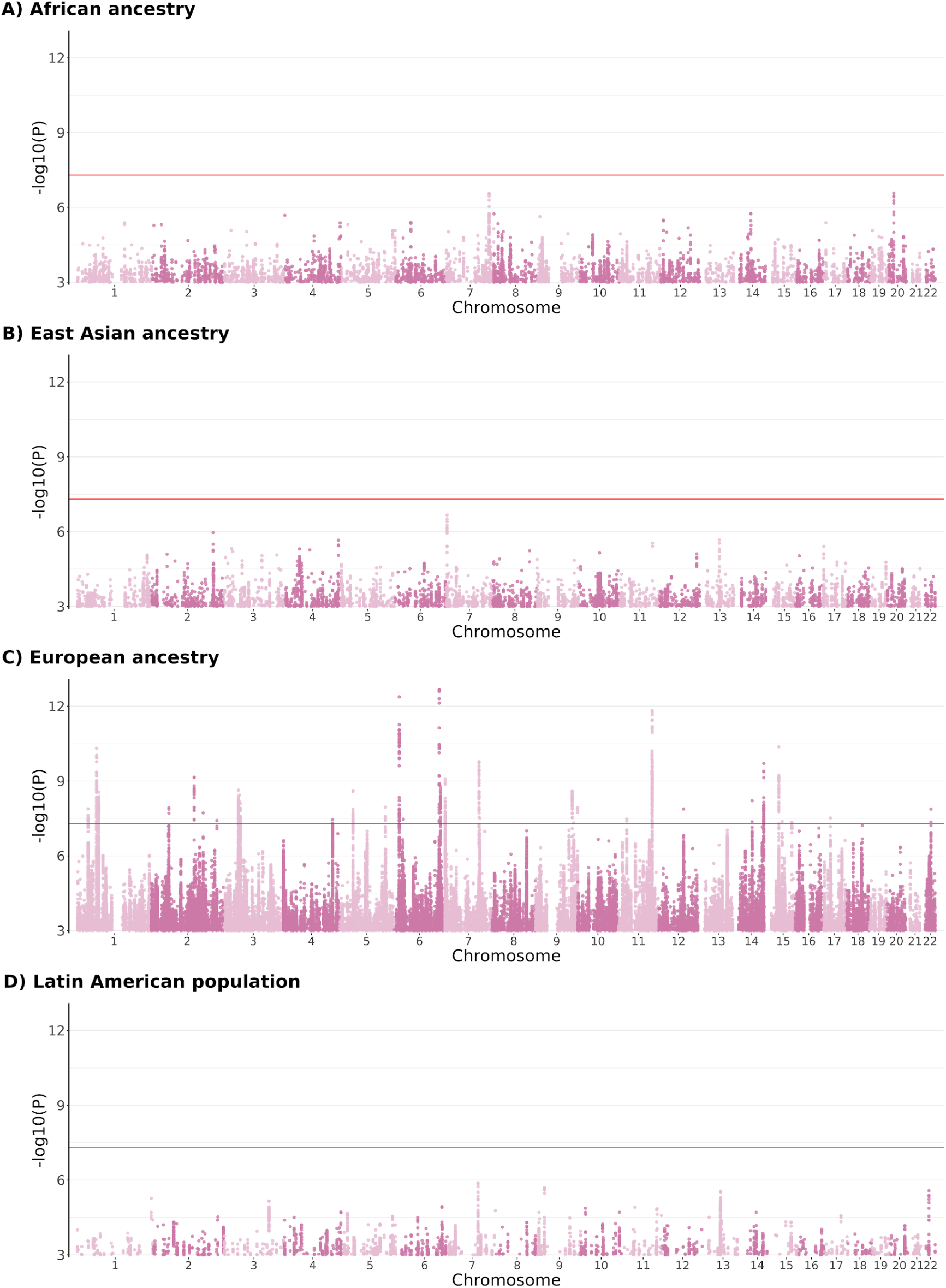
Manhattan plots of ancestry/population-specific suicide attempt GWAS meta-analyses. The x-axis shows chromosomal position and the y-axis shows significance of the association as -log10(P). The red line shows the genome-wide significance threshold (p = 5×10^-8^).

**Extended Data Figure 3.**
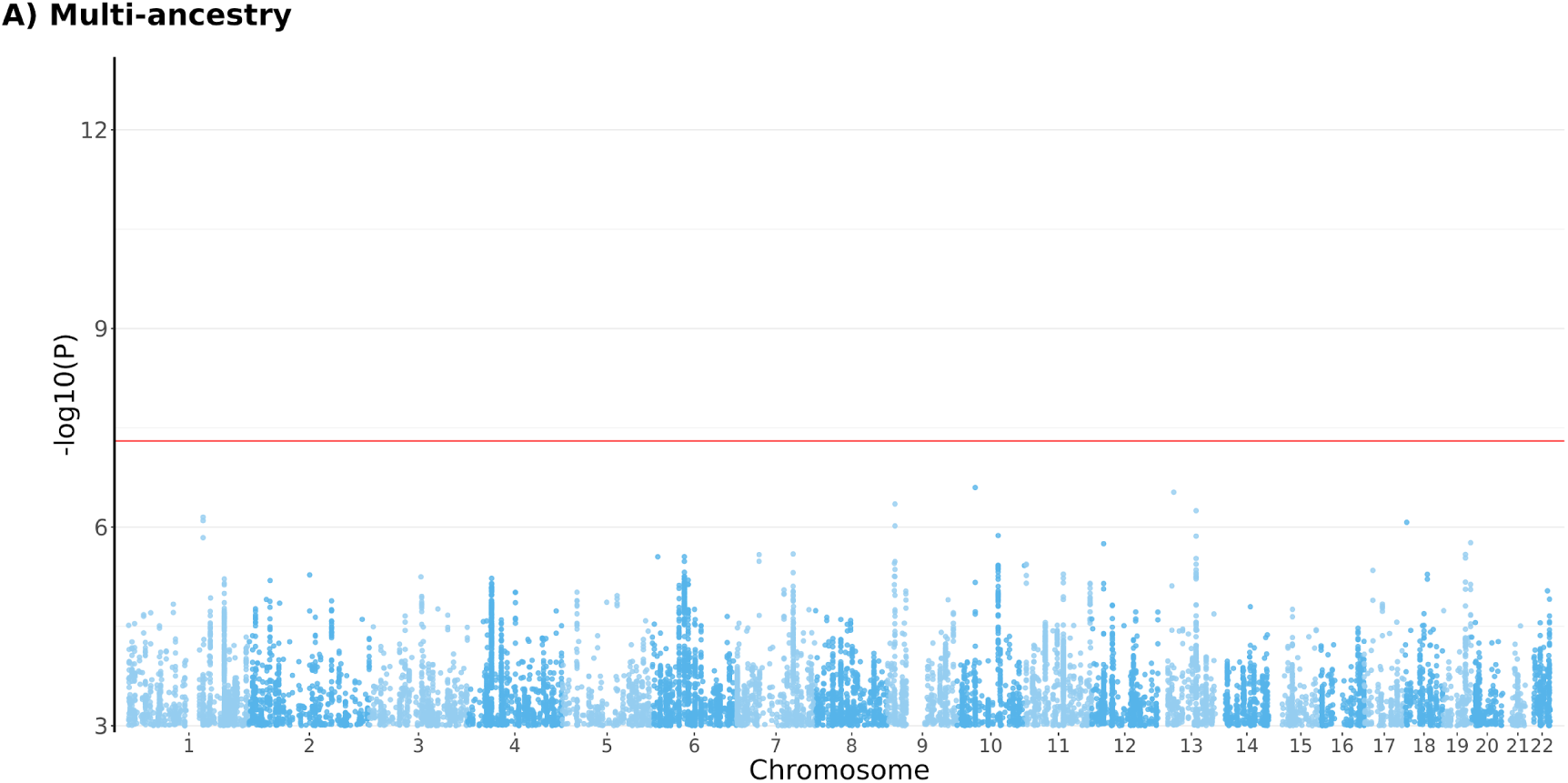
Manhattan plot of multi-ancestry suicide death GWAS meta-analysis. The x-axis shows chromosomal position and the y-axis shows significance of the association as -log10(P). The red line shows the genome-wide significance threshold (p = 5×10^-8^).

**Extended Data Figure 4.**
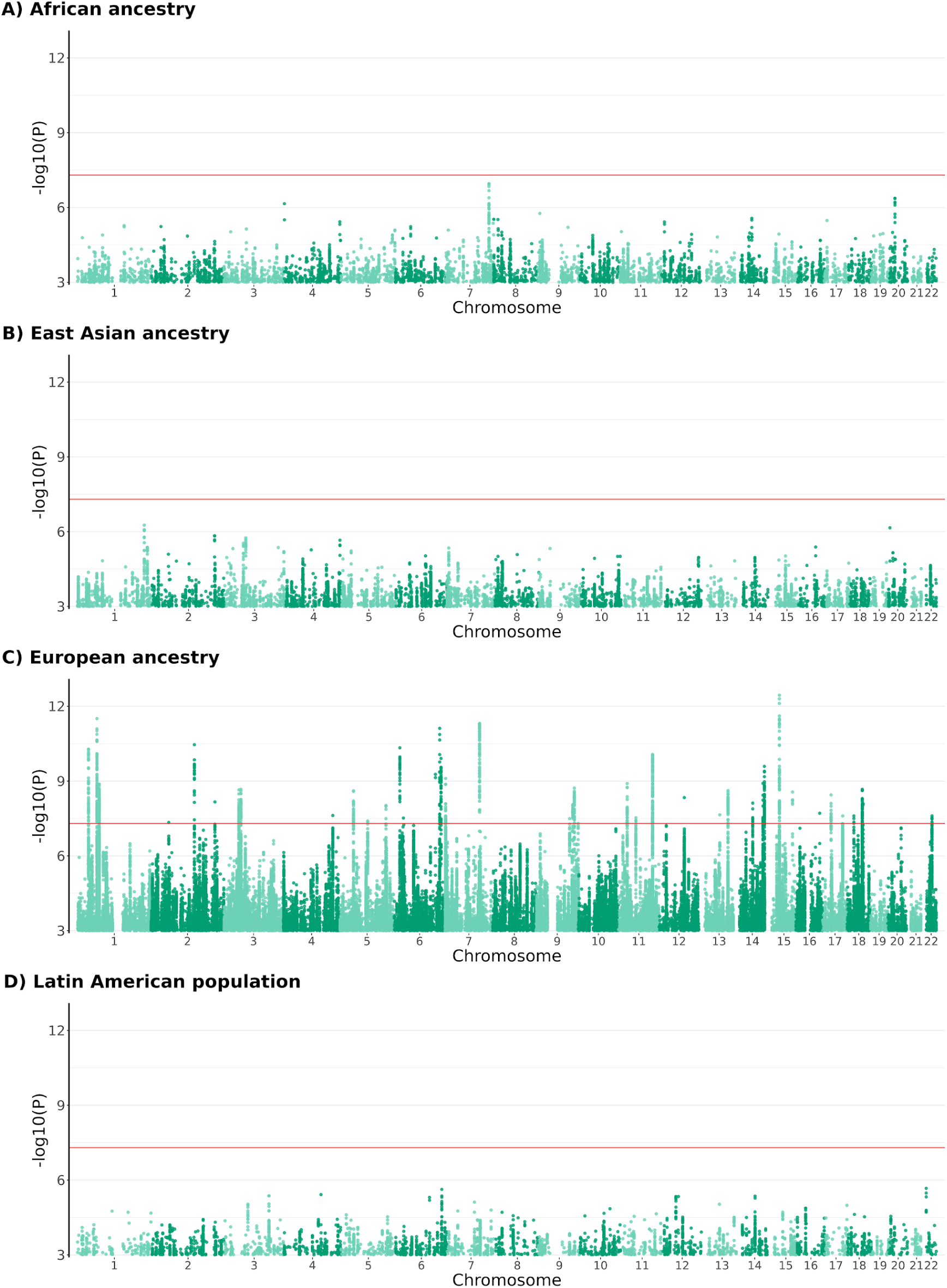
Manhattan plots of ancestry/population-specific suicidal behavior GWAS meta-analyses. The x-axis shows chromosomal position and the y-axis shows significance of the association as -log10(P). The red line shows the genome-wide significance threshold (p = 5×10^-8^).

