## Supplementary Information for "Genome-wide association studies identify 77 loci for suicidality and provide novel biological insights"

### SUPPLEMENTARY FIGURES

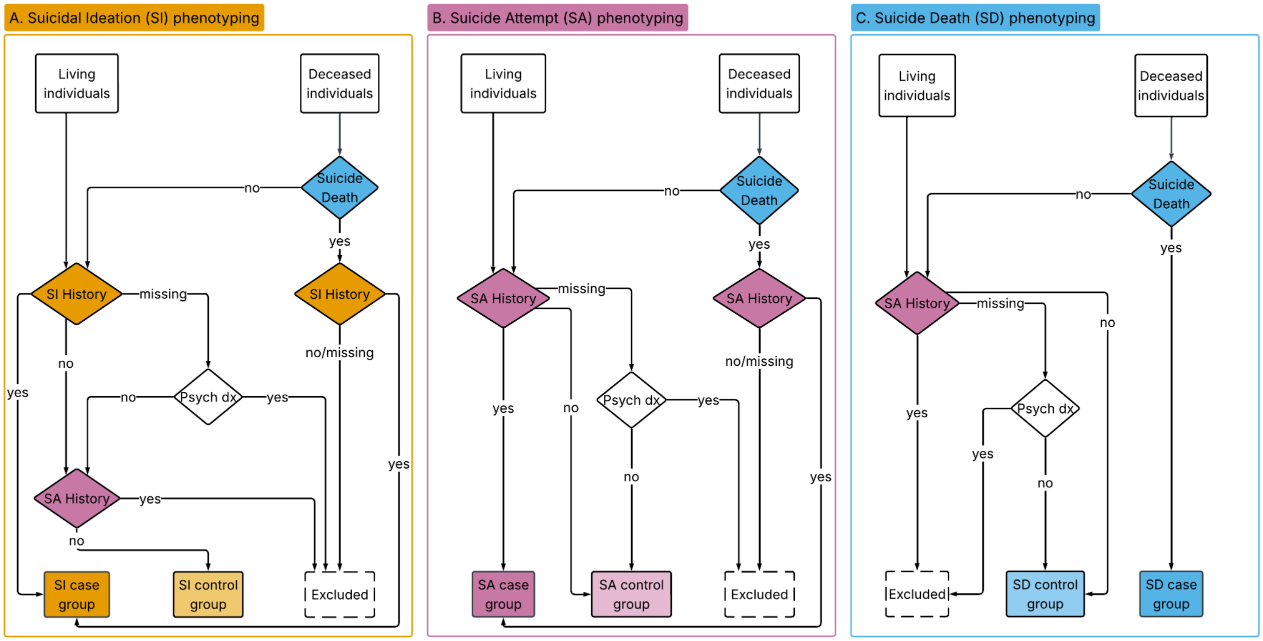

**Supplementary Figure 1: Flow diagrams illustrating case and control definitions for (A) suicidal ideation (SI), (B) suicide attempt (SA), and (C) suicide death (SD).** Individuals were first stratified according to whether they were living or deceased at the time of data collection. Colored boxes denote phenotype-specific case and control groups (SI, yellow; SA, pink; SD, blue) and dashed boxes indicate individuals excluded from the analysis of the specific phenotype. For all phenotypes, individuals missing the specific suicidality phenotype were assessed for the presence of a psychiatric diagnosis or a more severe suicidality phenotype, and then excluded if one was present. Diamonds indicate decision nodes, e.g., suicidality history (colored according to the suicidality phenotype being assessed) or psychiatric diagnosis (white diamonds).

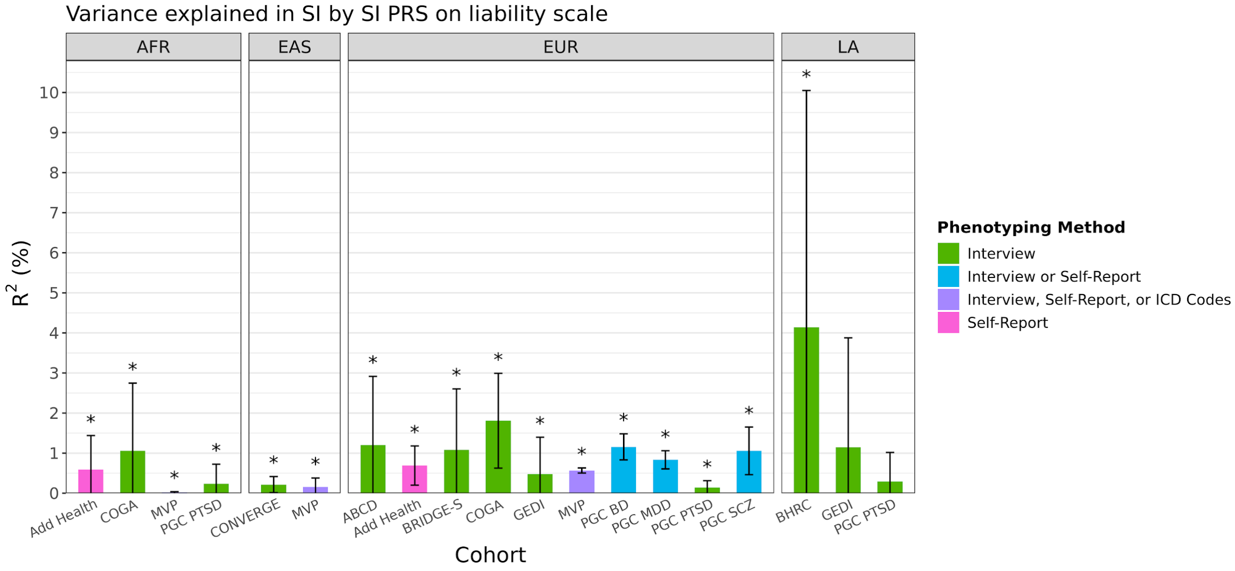

**Supplementary Figure 2: Cohort-specific estimates for the variance explained ($R^2$) in suicidal ideation by suicidal ideation PRS on the liability scale.**
Cohorts are grouped according to genetic ancestry/population (AFR = African, EAS = East Asian, EUR = European, LA = Latin American) and colored according to phenotyping method. * indicates significant p-values after correction for 5% false discovery rate. Exact estimates and sample sizes for each cohort can be found in Supplementary Table 19.

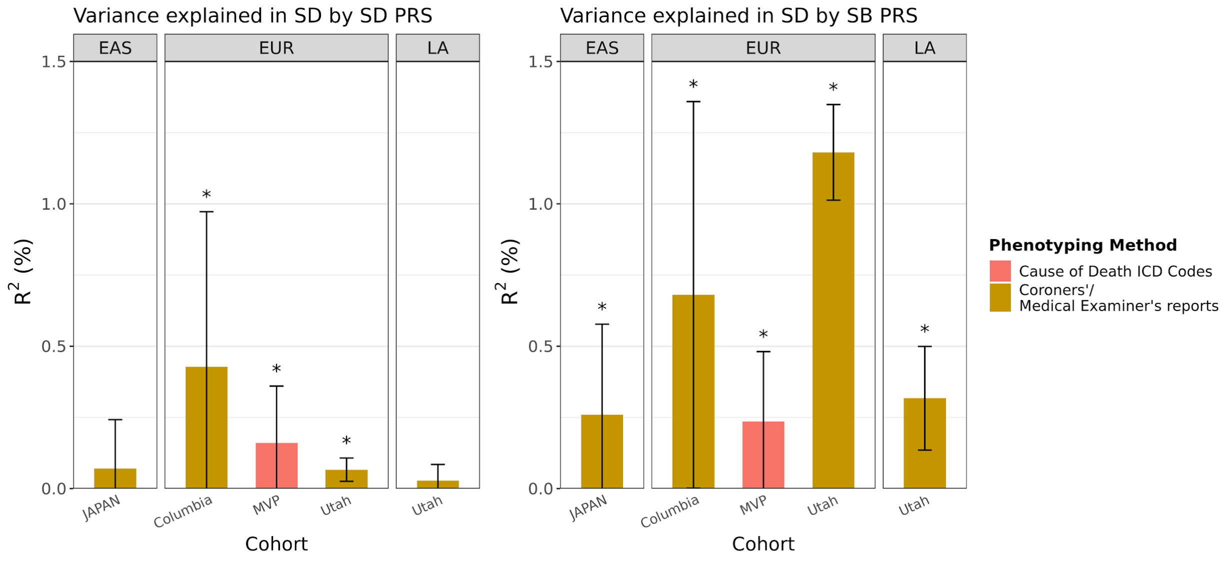

**Supplementary Figure 3: Cohort-specific estimates for the variance explained ($R^2$) in SD by the SD and SB PRS on the liability scale.**

Cohorts are grouped according to genetic ancestry/population (AFR = African, EAS = East Asian, EUR = European, LA = Latin American) and colored according to phenotyping method. * indicates significant p-values after correction for 5% false discovery rate. Exact estimates and sample sizes for each cohort can be found in Supplementary Tables 20 and 22.

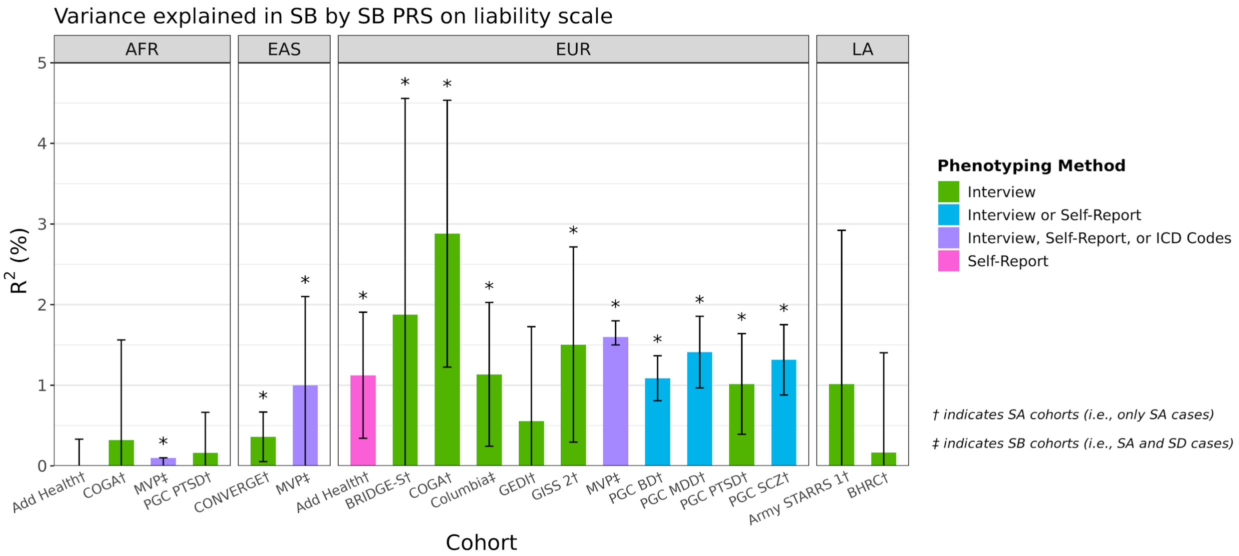

**Supplementary Figure 4: Cohort-specific estimates for the variance explained ($R^2$) in SB by SB PRS on the liability scale.**

Cohorts are grouped according to genetic ancestry/population (AFR = African, EAS = East Asian, EUR = European, LA = Latin American) and colored according to phenotyping method. * indicates significant p-values after correction for 5% false discovery rate. Exact estimates and sample sizes for each cohort can be found in Supplementary Table 21.

### SUPPLEMENTARY NOTE

#### COHORT ASCERTAINMENT, CASE AND CONTROL DEFINITIONS

##### The Adolescent Brain and Cognitive Development SM Study (ABCD Study®)

The ongoing longitudinal Adolescent Brain and Cognitive DevelopmentSM Study (ABCD Study®) recruited 11,879 children ages 9-11 (born 2005 - 2009) at baseline from 21 research sites across the United States to study health and development from middle childhood to early adulthood [1,2]. It includes a family-based component in which twin (n=2,108), triplet (n=30), non-twin siblings (n=1,589), and singletons (n=8,148) were recruited. Further details on the ABCD Study® are available elsewhere. We used the 5.1 release of the ABCD Study® data which includes data from ages 9-13. Suicidality was assessed using the KSADS-5 (Kiddie Schedule for Affective Disorders and Schizophrenia), a semi-structured interview of children psychopathology that corresponds to DSM disorder criteria. The assessment is administered to both the child and a parent/caregiver; endorsement by either informant is classified as a positive case.

##### Australian Genetics of Bipolar Disorder Study + Australian Genetics of Depression Study (GBP+AGDS)

This dataset includes from the Australian Genetics of Depression Study (AGDS)[3] and the Australian Genetics of Bipolar Disorder Study (GBP)[4] cohorts. Both studies aim to provide a comprehensive understanding of the genetic and environmental factors influencing major depression and bipolar disorder in Australia. Participants were recruited using two separate approaches: (1) for AGDS, recruitment was based on nationwide, pharmaceutical prescription history in the last 4.5 years for any of the 10 most commonly prescribed antidepressant medications (single medication or a combination) and a media publicity campaign throughout Australia. Recruitment for the GBP study was similar, but the ascertainment was based on previous Lithium treatment prescriptions instead of antidepressants. SA and SI were assessed using the suicidal ideation attributes scales (SIDAS).

For this project, QSkin participants serve as unscreened population controls, providing a population baseline sample against which case cohorts can be compared. The QSkin Sun and Health Study invited participants from Queensland, Australia at random from the compulsory Australian Electoral Roll in 2011, allowing recruitment of a representative sample. The final cohort included 43,794 men and women aged between 40 and 69 years[5]. As QSkin was established to better understand skin cancer, self-reported surveys collected detailed demographics, medical history and a range of skin cancer risk factors (e.g. skin colour and tanning ability). Further medical, treatment and cancer data was collated via consent to link to electronic health records, cancer registries, pathology clinics and the Australian Pharmaceutical Benefits Scheme.

#### Avon Longitudinal Study of Parents and Children (ALSPAC)

ALSPAC is a prospective longitudinal birth cohort that recruited pregnant women resident in Avon, UK with expected dates of delivery between 1st April 1991 and 31st December 1992 [6,7]. The initial number of pregnancies enrolled was 14,541 with 13,988 children alive at 1 year of age. Parents and offspring have been followed up regularly via self-report questionnaires and research clinics. The study website contains details of all the data that is available through a fully searchable data dictionary and variable search tool: http://www.bristol.ac.uk/alspac/researchers/our-data. Ethical approval for the study was obtained from the ALSPAC Ethics and Law Committee and the Local Research Ethics Committees. Informed consent for the use of all data collected was obtained from participants following the recommendations of the ALSPAC Ethics and Law Committee at the time. Participants can contact the study team at any time to retrospectively withdraw consent for their data to be used. Study participation is voluntary and during all data collection sweeps, information was provided on the intended use of data. Consent for biological samples has been collected in accordance with the Human Tissue Act (2004). Study data were collected and managed using REDCap electronic data capture tools hosted at the University of Bristol [8]. REDCap (Research Electronic Data Capture) is a secure, web-based software platform designed to support data capture for research studies. Data on suicide attempts was assessed using self-report questionnaire. Suicide attempts were assessed on 11 occasions in ALSPAC mothers and on 12 occasions in offspring (between age 11 and age 29). Participants were included as a case if they completed at least one assessment and reported having attempted suicide on at least one occasion. The sample was restricted to unrelated individuals (if a mother was included as a case then her child was excluded). Participants were included as a control if they completed at least one assessment and never reported having attempted suicide.
Note on data availability: The informed consent obtained from ALSPAC (Avon Longitudinal Study of Parents and Children) participants does not allow the data to be made available through any third party maintained public repository. Supporting data are available from ALSPAC on request under the approved proposal number, B4089. Full instructions for applying for data access can be found here: http://www.bristol.ac.uk/alspac/researchers/access/.

#### The Berlin Research Initiative for Diagnostics, Genetic and Environmental Factors of Schizophrenia (BRIDGE-S)

BRIDGE-S is a cross-sectional study aiming i) to explore genetic and environmental factors that mediate risk and resilience towards schizophrenia (SCZ) and ii) to facilitate genotype-informed disease subtyping through the collection of a densely phenotyped case-control sample in collected in Berlin, Germany. Detailed study procedures are published elsewhere [9].

The case sample comprises subjects who met diagnostic criteria for schizophrenia (ICD-10: F20) or schizoaffective disorder (ICD-10: F25) at any point during their life. Diagnosis is ascertained upon referral by clinical staff or access to medical records or provision of a physician's letter. Control subjects are eligible if they have never been diagnosed with Schizophrenia, Schizoaffective Disorder or bipolar disorder (ICD-10: F31), mental disorders other than that are not an exclusion criterion. Participants were recruited at multiple clinical sites and from the general population. Within the scope of the study all subjects donate a biological sample for genetic analyses and complete a broad questionnaire including measures of suicidality. Wave 7 included 438 cases with schizophrenia/schizoaffective disorder and 582 controls of European ancestry with available data on suicidality.

Current and lifetime suicidality including ideation and attempts is primarily captured with the Suicidal Behavior Questionnaire Revised (SBQ-R). In addition, age at first suicidal thoughts and first suicide attempt is recorded. Item 1 of the SBQ-R which assesses lifetime suicide ideation and attempts is used to stratify subjects into a suicide attempt (SA) group ("*I have attempted to kill myself and really hoped to die*" [4b]), suicide ideation (SI) group ([>=2]) and control group without a history of SI ("*Never*" [1]) and SA ([<=3b]). A subset of participants completed yes/no questions for SA ("*Have you ever attempted to take your own life?"*) and SI ("*Have you ever thought about taking your own life?"*), which were used to identify additional SA/SI cases and controls.

#### Brazilian High Risk Cohort for Mental Health Conditions (BHRC)

The BHRC is an accelerated school-based longitudinal study of child and adolescent brain development and mental health, conducted in Porto Alegre and São Paulo, Brazil[10,11]. Screening took place in 2009–2010 across 57 public schools, with caregivers of 9,937 children aged 6–12 years providing information on family psychiatric history, demographics, perinatal events, and exposure to stressful life events. From this screening sample, 2,511 children were selected for baseline evaluation (2010–2011, Wave 0), including 1,553 at high risk of mental illness based on family history and current symptoms, and 958 randomly selected participants. Follow-up assessments have been conducted every 2–4 years, encompassing Waves 1 (2013–2014), 2 (2018–2019), and 3 (2023–2025), with approximately 80% retention. Across waves, multimodal assessments included diagnostic interviews, behavioral questionnaires, cognitive and educational testing, neuroimaging, genomics, and other biomarker analyses. Psychiatric evaluations incorporated multiple informants (children and parents) and standardized instruments, including the Development and Well-Being Behavior Assessment (DAWBA) for children and the Mini-International Neuropsychiatric Interview (MINI) for the main caregiver. In the present study, suicidality phenotypes were derived exclusively from the parents' own baseline psychiatric assessment, using the suicidality module of the MINI v5.0.0, which includes standardized items on suicidal ideation and suicide attempt.

#### China, Oxford and VCU Experimental Research on Genetic Epidemiology (CONVERGE)

MDD cases were recruited from 58 provincial mental health centers and psychiatric departments within general hospitals, from 23 provinces in China[12]. Controls were recruited from patients undergoing minor surgery at general hospitals or local community centers. All subjects were Han Chinese women with four Han Chinese grandparents. Cases were excluded if they had a history of bipolar disorder, psychosis, or mental retardation. Cases were between ages 30-60 and had at least two episodes of MDD based on DSM-IV criteria, and with the first episode occurring between ages 14-50. They could not have abused drugs or alcohol prior to their first depressive episode. All subjects were interviewed using a computerized assessment program. The MDD diagnosis was determined using the Composite International Diagnostic Interview (WHO lifetime version 2.1; Chinese version). Instruments used to assess suicidality in the sample have been described in previous publications[13,14]. Cases were asked whether they had contemplated suicide during their worst depressive episode, and if so, whether they made a plan. Those who endorsed making a plan were asked whether they had attempted suicide. Controls were asked whether they had thought a lot about death or harming themselves and excluded if they responded in the affirmative after all disorders).

#### Collaborative Study on the Genetics of Alcoholism (COGA)

Probands with alcohol dependence were recruited from alcohol use disorder inpatient and outpatient treatment facilities and their family members were recruited from the seven participating COGA sites[15,16]. Comparison individuals and their families were ascertained from the community in the same seven areas. Institutional review boards at all sites approved the study and all participants provided informed consent. The Semi-Structured Assessment for the Genetics of Alcoholism (SSAGA) interview and an adolescent version of the SSAGA were administered to adults and individuals age 17 or under, respectively. The SSAGA collects information not only related to alcohol use disorders, but other substance use disorders and psychopathology. Lifetime SA was assessed with the question “Have you ever tried to kill yourself?” and lifetime SI was assessed with the question “Have you ever thought about killing yourself?”.

#### Columbia University (CUINT)

The cohort consists of healthy volunteers, MDD nonattempters based on SCID and Columbia Suicide history, MDD attempters based on SCID and Columbia Suicide history, suicide decedents based on medical examiner and our expert review, and sudden death controls based on medical examiner and our expert review. Cases consisted of those subjects who either died by suicide or attempted suicide, where a suicide attempt was defined as a self-injurious act during which the individual had, at least, partial intent to end his/her life. The number, method, and medical damage of past suicide attempts for live subjects were recorded on the Columbia Suicide History Form. Suicidal ideation for the USA and Canadian subjects was measured using the Scale for Suicidal Ideation. Diagnosis of major psychiatric disorders in suicides was determined using the SCID I by means of a validated psychological autopsy method, as previously described[17]. Controls from the Munich site were randomly selected from the general population of Munich, Germany, and were contacted by mail. Controls from the New York and Montreal sites were solicited through advertising. The Montreal sample was composed of French Canadians, whereas the New York sample was composed of Europeans of any origin. Controls were assessed by psychiatrists or clinical psychologists and evaluated using the SCID

NP version and the SCID II. In this study, we included depressed controls and non-psychiatric controls. The latter were free of axis I diagnoses, cluster B personality disorder, substance use disorder and lifetime history of a suicide attempt. Depressed controls were individuals who did not have histories of suicide attempts, but met criteria for MDD.

#### The Consortium on Vulnerability to Externalizing Disorders and Addictions (cVEDA) and Molecular Genetics Lab (MGL) cohort

The cVEDA cohort includes individuals aged 6–23 from seven Indian sites, recruited from mental health facilities, local communities, and educational institutions[18,19]. Detailed phenotypic assessments are conducted using instruments translated into seven local languages and age-appropriately tailored to gather comprehensive data ranging from socio-demographics to psychiatric morbidity. Participants with psychiatric disorders, specifically Bipolar Disorder (BD), are identified and diagnosed at NIMHANS, Bengaluru, with confirmations by two independent experts through clinical interviews and medical record reviews. Biological samples are stored in a biobank at NIMHANS to support research by consortium partners and collaborators on various aspects of mental and physical health, and cross-cultural studies. In the cVEDA cohort, suicidality phenotypes were assessed using specific items from the MINI v5.0.0, focusing on both suicide attempts (Item C9) and ideation (Items C2, C4, C5, C6). For individuals diagnosed with bipolar disorder (BD), relevant questions from the SCID-5-CV covered suicidal ideation and attempts. Control subjects were volunteers from the general population, free from any lifetime psychiatric diagnoses or history of suicidal ideations and attempts.

#### Estonian Biobank (EstBB)

Estonian Biobank (EstBB) is a volunteer-based cohort of 212,000 adults (20% of adult population in Estonia) with a rich variety of phenotypic and health-related information collected for 212,000 adult individuals[20]. At recruitment, participants signed a broad consent form allowing follow-up linkage of their electronic health records, thereby providing a longitudinal collection of their phenotypic information. The EstBB database includes health records from the National Health Insurance Fund Treatment Bills (from 2004), Tartu University Hospital (from 2008), and North Estonia Medical Center (from 2005), and data from national quality registries (causes of death, cancer, etc.). In addition, questionnaire data has been collected for lifestyle, mental health and wellbeing, medication side effects and personality. SD cases were ascertained from cause of death "suicide" recorded in the national Cause of Death Registry. SA cases were ascertained from ICD-10 diagnoses as per the lists provided by PGC SUI[21] OR answered “Yes” to self-report questionnaire item “Have you tried to take your own life?”. SI cases were ascertained from ICD-10 diagnosis R45.8 OR answered “Yes” to self-report questionnaire item “Have you felt that life is not worth living?”. For SA, SB and SD analyses, controls were all EstBB participants without any ICD-10 F diagnosis. For the SI analysis, controls were all EstBB participants without any ICD-10 F diagnosis OR individuals with ICD-10 F diagnosis AND without questionnaire-based self-reported suicide ideation. The activities of the EstBB are regulated by the Human Genes Research Act, which was adopted in 2000 specifically for the operations of the EstBB. Individual level data analysis in the EstBB was carried out under ethical approval "1.1-12/3093" from the Estonian Committee on Bioethics and Human Research (Estonian Ministry of Social Affairs), using data according to release application 6-7/GI/23635 from the Estonian Biobank.

#### FinnGen

The SA analysis using FinnGen data release 6 (R6) from the FinnGen Study (https://www.finngen.fi/en/about) included 4098 individuals with SA history, defined as the presence of SA International Classification of Diseases codes, and 247,898 individuals without the relevant codes. Ascertainment, case, and control definitions for this cohort have been previously published and described in full[22].

#### Genetic Investigation of Suicide and SA Wave 1 (GISS 1)

Sample recruitment, selection criteria, demographics, ancestry and psychiatric diagnoses have been described previously[23–26]. Briefly, lifetime SA was the main outcome ascertained in the offspring of nuclear family trios (all complete with both biological parents and one SA offspring per trio; n = 660). Trios were collected in Ukraine by first recruiting offspring from emergency care due to a severe SA, defined as a score of = 2 on the Medical Damage Rating Scale (MDS)[27], which represented the primary ascertainment criteria for inclusion. Persons who have engaged in suicidal thoughts without actual behavior would not be included. Other exclusion criteria were subject adopted, mental retardation, organic mental disorder, or other chronic medical illness involving the central nervous system. The SA were verified independently by both parents, the suicide attempter and by examining medical records. The suicidal intent of the SA was assessed by using both objective (levels of precaution) and subjective (intent to die) aspects [28]. Previous life-time SA was documented, as well as the history of suicides in family and relatives. Secondary outcomes included ICD-10 diagnoses according to the Composite International Diagnostic Interview (CIDI), personality traits according to the NEO personality inventory (NEO-PI-R), levels of anger, Beck's depression inventory, the WHO well-being index and the Global assessment of functioning (GAF) scale. Exposures to lifetime stressful and traumatic life-events (SLEs) were also assessed. Overall, the SA offspring included 51.1% males (n=337)/48.9% females (n=323), with mean ages of 24.6 (S.D. ± 7.3)/23.8 (S.D. ± 7.1) years, and 94.4% (n=318)/93.2% (n=301) of the SA subjects had = 3 Ukrainian or Russian grandparents, respectively. Overall, n=498 SA subjects did not have any of the major psychiatric diagnoses, e.g. schizophrenia (ICD-10 code F20), schizoaffective disorder (F25) or moderate / severe depression diagnoses (F32-33). The collection of research subjects followed the code of ethics of the World Medical Association (Declaration of Helsinki), and written consent was obtained. The study was approved by the Research Ethics Committee at the Karolinska Institute (Dnr 97–188) and by the Ministry of Health in Ukraine.

#### Genetic Investigation of Suicide and SA Wave 2 (GISS 2)

The GISS2 sample was the result of the same sample collection used for generating the GISS1 family trios, but consists of suicide attempt (SA) cases without parents and an additional sample of non-SA healthy volunteer (HV) controls.
The n=564 SA cases GISS2 data had been collected by 29 interviewers located across the entire Ukraine, near or in the following 16 cities (# SA cases): Chernihiv (1), Dnipro (11), Donetsk (24), Kharkiv (20), Kyiv (32), Kirovohrad (21), Kryvyj Rih (1), Lutsk (3), Lviv (75), Mykolaiv (68), Odesa (217), Poltava (23), Symferopil (38), Ternopil (1), Vinnytsia (3) and Zaporizhzhia (25). Collection was on-going at an even rate during years 2002-2006. As for GISS1, the main ascertained outcome was self-inflicted injuries which had required a certain level of medical treatment for recovery (as rated by Medical Damage Rating Scale [MDRS] score of >1; [27]), further characterized with regard to the method used (ICD-10 codes X60-X84), the suicidal intent, number of previous lifetime attempts, lifetime suicide ideation (79%, n=446 SA), use of drugs/alcohol at SA and age of onset. The mean (S.D.) age of the SA cases in GISS2 were 26.1 (9.0) and 29.4 (13.5), for the 47.5% (268) males and 52.5% (296) females, respectively. Secondary features assessed were lifetime ICD-10 diagnoses (e.g. posttraumatic stress disorder [F43.1], n=54 SA, moderate or severe depression diagnoses [F32-33], n=61 SA, past-year harmful use of dependence of alcohol [F10.1 or F10.2; n=52 SA], Schizophrenia or Schizoaffective disorder [F20G or F25], n=21 SA, anxiety disorders, n=115), NEO personality inventory (NEO-PI-R; n=402 SA), lifetime stressful and traumatic life-events (SLEs), trait anger scale (TAS; n=547 SA), past feelings and acts of violence (PFAV; n=552 SA), Beck's depression inventory (BDI-I; n=552 SA), the WHO well-being index (WBI; n=552 SA), Beck hopelessness (BH; n=552 SA) and the Global assessment of functioning (GAF; n=564 SA) scales.

The n=513 HVs have been used previously in candidate gene association studies [26]. The HV data had been collected by one interviewer in the city of Odesa, during years 2003-2005. Exclusion criteria was any previous lifetime SA, or being below 18 years of age, known relationship to any previously collected SA subjects, somatic / psychiatric diseases, alcohol abuse/dependence or homelessness. The mean (S.D.) age of the HVs were 35.7 (16.2) and 34.6 (14.9), for the 46.2% (237) males and 53.8% (276) females, respectively. 78% of HVs were employed covering a large variety of different jobs, 13.5% were students, 6.6% were retired and 0.4% of HVs were unemployed. Nevertheless, 6.8% (35) of HVs had experienced lifetime suicide ideation (BDI-I item 9 > 0). However, only 2.5% (13) had BDI-I scores >20 (i.e. at least moderate levels of current depression) at the time of interview, compared to 38% (215) among the GISS2 SA cases. As for the SA cases, other secondary features assessed were NEO-PI-R (n=430 HVs), as well as lifetime SLEs, TAS, PFAV, WBI, BH and GAF scales (for all n=513 HVs).

#### The Genes, Environment, and Development Initiative (GEDI)/Great Smoky Mountains Study (GSMS) & Virginia Twin Study of Adolescent Behavioral Development (VTSABD)

The Great Smoky Mountains Study is a longitudinal, representative study of 1420 children in 11 predominantly rural counties in Southeastern United States[29]. Annual assessments on psychopathology and associated factors were completed on the 1420 children until age 16 (6674 observations of 1420 individuals; 1993 to 2000) and then again at ages 19, 21, 25, and 30 (4556 observations of 1336 participants; 1999 to 2015) for a total of 11,230 total assessments. The study protocol and consent consent/assent forms were approved by the Duke University Medical Center Institutional Review Board. Participants in all the studies gave consent for their DNA to be genotyped. Suicidality phenotypes were assessed via items in the structured diagnostic Child and Adolescent Psychiatric Assessment interview.
The Virginia Twin Study of Adolescent Behavioral Development study[30] is a population-based multi-wave, cohort-sequential twin study of adolescent psychopathology and its risk factors, and has two follow-up studies, the Young Adult Follow Up (YAFU)[31] and the Transitions to Substance Abuse (TSA) study[32]. Up to four assessments on psychopathology and associated factors were completed on adolescent twins under age 18 in 1412 families (6331 observations of 2775 individual twins; 1990 to 1998), and at mean ages of 21 (YAFU) and 26 (TSA)(3410 observations of 2326 participants; 1998 to 2006) as well as a GEDI genotypic data collection of 913 individuals for a total of 10,654 total assessments. The study protocol and consent consent/assent forms were approved by VCU's Institutional Review Board. Participants in all the studies gave consent for their DNA to be genotyped. Suicidality phenotypes were assessed via items in the structured diagnostic Child and Adolescent Psychiatric Assessment interview.

#### Genomic Research and Epidemiological Studies for Affective Disorders in Taiwan (GREAT)

Participants were drawn from the database of Genomic Research and Epidemiological Studies for Affective Disorders in Taiwan (GREAT)[33], which consisted of both family-based and case-control study designs. Patients diagnosed with bipolar or major depressive disorder according to the criteria of the DSM-5 were consecutively referred by psychiatrists in several collaborating hospitals in Taiwan. Subjects aged between 18 and 70 years were recruited between the year 2008 and 2023 from hospitals and control participants are drawn from the same geographic area as case participants. All participants were undergone a face-to-face interview by well-trained lay interviewers using a modified Chinese version of the Composite International Diagnostic Interview (CIDI) or the modified Schedule of Affective Disorder and Schizophrenia- Lifetime (SADS-L) to collect demographic characteristics and suicidal behaviours. The interview was used to assess clinical features of mood disorders and to collect information about lifetime suicidality, such as 'Have you ever had a serious thought about attempting suicide or wanting to die?' (SI) and 'Have you ever non-fatal deliberate self-harm with at least some intent to die' (SA), with a yes/no dichotomous answer.

#### Grady Trauma Project (GTP)

The subjects for this study were part of a larger investigation of genetic and environmental factors that predict the response to stressful life events in a predominantly African American, urban population of low socioeconomic status. Participants were approached while in the waiting rooms of primary care, diabetes, or obstetrical-gynecological clinics of Grady Memorial Hospital in Atlanta, Georgia. Screen interviews, including participants' demographic information (e.g., self-identified race, sex, and age), prior hospitalization for psychiatric diseases, and psychiatric symptoms including Posttraumatic Stress Disorder (PTSD), depression, schizophrenia, and bipolar disorder, were completed on site. Suicide attempt history was assessed based on self-report (yes/no) when obtaining demographic information. Further details regarding the GTP dataset can be found in Gillespie et al[34]. Written and verbal informed consent was obtained for all participants and all procedures in this study were approved by the institutional review boards of Emory University School of Medicine and Grady Memorial Hospital, Atlanta, Georgia. The primary GWAS of SA included 669 cases and 4473 controls.

#### The Individualized Medicine: Pharmacogenetics Assessment and Clinical Treatment Study (IMPACT)

The IMPACT study is a naturalistic longitudinal pharmacogenetic study in which psychiatric patients were followed for up to 8 weeks following pharmacogenetic testing[35]. The study included patients with diverse psychiatric diagnoses who were 7 years of age or older and were on or about to be prescribed psychotropic medications. For this study, we included participants of genetically European ancestry with depression based on referral forms and available Beck Depression Inventory (BDI) scores at baseline. Participants who endorsed BDI item 9 (score of at least 1) were categorized as cases and those who scored 0 on this item were categorized as controls.

#### International Borderline Genomics Consortium - Central European subset (IBGC CE)

Subjects were part of the central European subsample of an international GWAS of Borderline Personality Disorder[36]. The diagnosis of Borderline Personality Disorder was assigned according to DSM-IV criteria on the basis of structured clinical interviews (IPDE; SCID-I; SCID-II; SCID-IV) and SCID II interviews[37–41], adapted for proxy-based interviews for the post-mortem subsample[42]. Life-time attempt of suicide were documented. Diagnostic interviews were conducted by trained and experienced raters. For the RCT sample[37], the Borderline Personality Disorder Severity Index (BPDSI) was applied at the beginning and at the end of the baseline assessment. For each timepoint, the occurence of suicide attempts and suicide ideation during the previous three month period was assessed. Subjects positive for SA or SI in the BPDSI were included as cases for the respective analysis, subjects negative for SA and SI in the BPDSI were excluded from analysis, as this does not exclude life-time occurrence. For the Montreal sample[42], a post-mortem study using tissue from the Douglas Bell-Canada Brain Bank, cases were only included to the SB model including SD. Controls were only included from the subsamples, which excluded controls with a history of mental illness[37,39,40] due to missing SI/SA information.

#### International Borderline Genomics Consortium - German Borderline Genomics Consortium subset (GBGC)

Subjects were part of a German subsample of an international GWAS of Borderline Personality Disorder[36,43]. The selected subjects consist of cases recruited in Berlin and Mannheim, and controls recruited in Mainz and from a sample of blood donors recruited in Mannheim, Germany. The diagnosis of Borderline Personality Disorder was assigned according to DSM-IV criteria on the basis of structured clinical interviews (either IPDE or SCIDII). Diagnostic interviews were conducted by trained and experienced raters. Life-time attempt of suicide and, in the case of a positive answer, the number of attempts were documented. Diagnostic interviews were conducted by trained and experienced raters. Controls from Mannheim were blood donors who filled out a questionnaire including questions on mental and somatic health. For the current study the following information was used to exclude blood donors with a history of mental disorders: self-report of psychiatric disorders, self-report of diagnosis of psychiatric disorder by a healthcare professional, and a questionnaire version of the SCID items for depression criteria A1–A9. The subgroup of subjects affirming at least one of the two SCID depression screening items were asked for their lifetime history of suicide attempts. Control subjects with a history of suicide attempt were included as suicide attempt cases in the respective model. Controls from Mainz missing information on SA data were excluded in the case of history of a psychiatric disorder (panic disorder, agoraphobia, social phobia, specific phobia, generalized anxiety disorder, PTSD, obsessive-compulsive disorder, major depression, dysthymia, mania, hypochondriacal disorder, somatoform disorder, pain, conversion disorders, anorexia nervosa, bulimia nervosa, harmful alcohol use, alcoholism, harmful drug use, drug addiction schizophrenia, schizotypal disorders).

#### International Borderline Genomics Consortium - Spanish subset (IBGC SPAIN)

Subjects were part of the central Spanish subsample of an international GWAS of Borderline Personality Disorder [36]. The diagnosis of Borderline Personality Disorder was assigned according to DSM-IV criteria on the basis of structured clinical interviews (SCID-I; DIB-R). Life-time SA was documented. Diagnostic interviews were conducted by trained and experienced raters. Healthy controls (CTL) were recruited from the local population and controls missing information on SI/SA were excluded in the case of a history of mental illness, drug use or treatment with psychotropic medication.

#### Japan

For the Japanese cohort, we used data from 746 suicide decedents (386 suicides who died between June 1996 and July 2012 in the 1st set and 360 suicides who died between August 2012 and February 2017 in the 2nd set)[44]. Autopsies on suicides were performed and the decision of assigning the status “suicide” was made through discussion with the Medical Examiner’s Office of the Hyogo Prefecture and the Division of Legal Medicine in the Kobe University Graduate School of Medicine. For non-suicide controls, we used genome-wide genotype data from 14,049 subjects (7,458 controls in the 1st set and 6,591 controls in the 2nd set) in the Biobank Japan project who had been genotyped as case subjects for non-psychiatric disorders and healthy volunteers.

#### Janssen Waves 1 + 2

The Janssen samples consist of suicide attempt cases and controls of European ancestry and were drawn from 12 clinical trial samples (NCT00044681, NCT00397033, NCT00412373, NCT00334126, NCT01193153, NCT02497287, NCT02422186, NCT01627782, NCT00253162, NCT00257075, NCT01515423, and NCT01529515) conducted by Janssen Research & Development, LLC.. Ascertainment, case, and control definitions for these cohorts have been previously published and described in full [22].

#### The Lundbeck Foundation Initiative for Integrative Psychiatric Research (iPSYCH)

All individuals included in this study were a part of the Danish iPSYCH 2012 population-based case-control cohort[45]. SA cases were identified according to information available from the Danish Psychiatric Central Research Register and the National Registry of Patients both complete until December 31, 2016. SA cases were identified as individuals with ICD-10 diagnoses of SA (ICD-10: X60-X84, equivalent to intentional self-harm), with SA indicated as ‘reason for contact’ in the registers, and with a main diagnosis of poisoning (ICD-10: T39, T42, T43, and T58). The SA case group also included individuals with a diagnosis in the ICD- 10: F chapter as main diagnosis and report of poisoning by drugs or other substances (ICD-10: T36–T50, T52–T60) or injuries to hand, wrist, and forearm (ICD-10: S51, S55, S59, S61, S65, S69). Individuals who died by suicide according to Cause of Death Register available until December 31, 2015 were classified as SD cases for use in the SB GWAS. Only contacts starting at age 10 years old or older were considered to be reliably reported SA cases. Individuals not fulfilling any of the above case criteria were considered to be controls. The study was approved by the regional Danish ethics committee and the Danish Data Protection Agency.

#### Mental Illness Research Education and Clinical Center (MIRECC)

A description of the participants in this cohort have been described previously[46,47]. Briefly, participants are comprised of Iraq and Afghanistan-era veterans collected through the Mid-Atlantic Mental Illness Research, Education, and Clinical Center (VISN 6 MIRECC) at four VA medical centers: Durham VA, Salisbury VA, Hampton VA, and Richmond VA.  Informed consent was obtained from all study participants whereupon questionnaires and blood samples were collected. The sample was comprised of both African-American (51.3%; n=1,329) and European-American (48.7%; n=1,320) veterans. Women veterans were also well represented

(25.3%; n=685). History of suicide attempts was assessed with item #20 on the BSS. Suicidal ideation was defined as any endorsement of suicidal thinking on the BSS, BDI-II, or SCL-90-R.

#### Million Veteran Program (MVP)

The MVP cohort included 633 778 US military veterans of African, Asian, European, or Hispanic ancestry. MVP study procedures included providing informed consent, donating a blood sample, and agreeing to have one's genetic information linked with one's electronic health record data within the MVP biorepository. Ascertainment, case, and control definitions for the SI analyses in MVP have been previously published and described in full[48]. Several different data sources were used to identify SA cases in the MVP Cohort, including International Classification of Diseases, Ninth Revision (ICD-9) and Tenth Revision (ICD-10) codes from the electronic health record, suicide behavior reports from the VA's Suicide Prevention Applications Network database, mental health survey responses from the VA's Mental Health Assistant database[49]. Cause of death codes from the National Death Index were used to identify SD cases. For the SA, SB, and SD GWAS, participants were classified as control individuals if they had no documented lifetime history of suicide attempt or suicide death.

#### National Longitudinal Study of Adolescent to Adult Health (Add Health)

The National Longitudinal Study of Adolescent to Adult Health (Add Health) is a nationally representative study of more than 20,000 adolescents in the United States[50]. Interviews were conducted in 1994 (Wave I), 1996 (Wave II), 2001-2002 (Wave III), 2008-2009 (Wave IV), and 2016-2018 (Wave V). Add Health has collected demographic, social, familial, socioeconomic, behavioral, psychosocial, cognitive, and health survey data from participants and their parents; a vast array of contextual data from participants' schools, neighborhoods, and geographies of residence; and in-home physical and biological data from participants, including genetic markers, blood-based assays, anthropometric measures, and medications.

To evaluate suicidal ideation, participants were asked, "During the past 12 months, did you ever seriously think about committing suicide?" Participants who responded positively to this question at any assessment were considered SI cases. To evaluate suicide attempt, participants were asked, "During the past 12 months, how many times did you actually attempt suicide?". At Waves I-III, this item was administered only to individuals who endorsed SI, whereas at Waves IV and V it was administered to all participants. Participants who responded positively to this question at any assessment were considered SA cases. Participants who responded to the suicide-related items at least once and did not endorse suicidal thoughts or behaviors at any assessment were treated as controls.

#### Predictors For ECT study (PREFECT)

The Predictors For ECT (PREFECT) study enrolled individuals from the Swedish National Quality Register for ECT (Q-ECT) between 2013 and 2017. All Swedish hospitals administering ECT report to Q-ECT, which records clinical and demographic information including indication for the current ECT series and self-rated Montgomery–Åsberg Depression Rating Scale (MADRS-S) scores. PREFECT participants were included if the indication for ECT was a major depressive episode (MDE). Blood samples were collected either retrospectively (via mailed kits following telephone consent) or prospectively (prior to first ECT session in planned series of ≥6 treatments) at seven participating psychiatric hospitals in Sweden., Information about whether the patient had ever made a suicide attempt were based on a structured interview with a trained nurse (no, 1–2 times, 3 or more times, missing) . Information about completed suicide was extracted from the the Swedish national cause of death register (X60-X84, Y10-Y34).
The PREFECT study has been described previously[51,52]. Controls (n = 3290) were obtained from the Swedish arm of the Anorexia Nervosa Genetics Initiative (ANGI) [53], either population-based (n = 1035) or from the LifeGene study (n = 3000). For this study, controls were excluded if they self-reported lifetime history of MDD, bipolar disorder, schizophrenia, or schizoaffective disorder.

#### Psychiatric Genomics Consortium Bipolar Disorder (PGC BD)

Subjects were drawn from 35 bipolar disorder (BD) case-control cohorts in the Psychiatric Genomics Consortium (PGC)[54]: amq1, bmau, bmpo, bmrom, bmsp, bonn, dub1, dutch, fat2, fran, gain, germ1, graza, greek, gsk1, hal2, ital1, may1, mich, neuc1, norgs, noroe, rom3, rom4, spsp3, st2c, stp1, swa2, tgco2, top7, top8, uclo, ukwa1, ume4, wtcc. Structured psychiatric interviews and self-report questionnaires were used to diagnose BD and ascertain information on SI and SA. Patients with BD endorsing SI or SA were included as cases in the SI and SA GWAS, respectively. The controls included individuals with BD who did not endorse the case phenotype as well as healthy controls. BD cases who were missing information on SI or SA were excluded from the relevant GWAS. The healthy controls from PGC BD were screened for the absence of lifetime psychiatric disorders. The source, inclusion and exclusion criteria for each individual PGC BD cohort have been reported in detail previously [54].

#### Psychiatric Genomics Consortium Eating Disorders (PGC ED)

Subjects originated from 3 anorexia nervosa (AN) case-control cohorts in PGC, where information on SA had been collected. The ascertainment, phenotype measurement, and inclusion and exclusion criteria have been described previously for these cohorts[55,56]. The cohorts were the Children's Hospital of Philadelphia/Price Foundation Collaborative Group (CHOP/PFCG) case-control cohort, and the France and Spain case cohorts from the Genetic Consortium for Anorexia Nervosa/Wellcome Trust Case Control Consortium-3 (GCAN/WTCCC-3) with controls sourced as described in Duncan et al[56]. Control cohorts from a similar geographic location and genotyping platform were preferentially sought. PGC AN cases had DSM-III-R or DSM-IV diagnoses of AN or EDNOS-AN (i.e., without the requirement of amenorrhea) based on structured diagnostic interviews. Controls had not been screened for AN but prevalence of lifetime AN is rare (~1%), nor had they been screened for SA. The same procedures described for the PGC MDD cohorts were used to define cases and controls.

#### Psychiatric Genomics Consortium Major Depressive Disorder (PGC MDD)

Subjects were drawn from 29 major depressive disorder (MDD) case-control cohorts in PGC[57]: bidi1, boma, cof3, col3, formm, gens, gep3, grdg, grnd, gsk2, gsrdf, gsrdg, gsrdi, gsrdp, mmi2, mmo4, mrive, nes1, qi3c, qi6c, qio2, rad3, rage, rau2, rde4, rot4, shp0, stm2, twg2. As described for the PGC BD cohorts, structured psychiatric interviews and self-report questionnaires were used to diagnose MDD and ascertain information on SA and SI. Cases and controls were defined in the same way as for the PGC BD sample. The healthy controls from most PGC MDD cohorts were screened for the absence of lifetime psychiatric disorders. The source, inclusion and exclusion criteria for each individual PGC MDD cohort have been reported in detail previously[57].

#### Psychiatric Genomics Consortium Post-Traumatic Stress Disorder (PGC PTSD) and Army Study to Assess Risk and Resilience in Servicemembers - Wave 1 Latin American subset (ARMY STARRS)

Subjects were drawn from 4 post-traumatic stress disorder (PTSD) case-control cohorts in the PGC[58]: safr, saf2, nss1 (ARMY STARRS wave 1), nss2. The same procedures were used to ascertain information on SI and SA and define cases and controls as described previously for the other PGC studies. The source, inclusion and exclusion criteria for each individual PGC PTSD cohort have been reported in detail previously[58].

#### Psychiatric Genomics Consortium Schizophrenia (PGC SCZ)

Subjects were drawn from 12 schizophrenia (SCZ) case-control cohorts in the PGC[59]: butr, cgs1c, celso, xboco, xcou3, xdenm, xmgs2, xmunc, xport, xtop8, xucla, xuclo. The same procedures were used to make psychiatric diagnoses, ascertain information on SA and define cases and controls, as described previously for PGC MDD and BD studies. The source, inclusion and exclusion criteria for each individual PGC SCZ cohort have been reported in detail previously[59].

#### PsyCourse

The PsyCourse Study is a multi-site, transdiagnostic, observational, longitudinal study conducted in Germany and Austria[60] within the frameworks of the Clinical Research Group 241 (KFO241 consortium; www.kfo241.de) and the PsyCourse consortium (www.psycourse.de). The official period of data collection was from January 2012 through December 2019. Participants of the PsyCourse Study are either clinical participants with a diagnosis from the affective-to-psychotic spectrum or neurotypic (control) participants. Twenty study centers contributed to the project, recruiting former and current in- and outpatients of their respective psychiatric clinics. Diagnoses of clinical participants were made according to DSM-IV, a small subset of patients with schizophrenia was diagnosed according to ICD-10. Neurotypic participants were drawn from the same geographic area as clinical participants. They were contacted via registration offices and public notices in three of the 20 study centers and screened for the absence of the target diagnoses for study inclusion. Within the PsyCourse Study, participants had up to four evenly spaced study visits across a span of 18 months. Each study visit involved an extensive interview, neurocognitive tests and self-rating scales as well as the blood sampling. Details on the scales used can be found here: https://data.ub.uni-muenchen.de/390/1/230614_PsyCourse_v6.0.html. For clinical participants, relevant sections of the Structured Clinical Interview for DSM-IV (SCID I) were assessed at baseline, including a section on suicidal ideations and attempts (lifetime assessment). At the follow-up visits, these questions were asked again referring to the period of time since the last interview. Clinical participants were rated as cases for the analysis on suicide attempt, if they reported at least one suicide attempt at any of the study visits. Accordingly, they were rated as cases for the analysis on suicidal ideation, if they reported suicidal ideations at any of the study visits. Clinical participants were rated as controls for the analysis on suicide attempt, if they did not report any suicide attempts across all study visits available. Accordingly, they were rated as controls for the analysis on suicidal ideation if they reported no suicidal ideations across all study visits available. According to the phenotyping protocol of the PGC Suicide Working Group, since PsyCourse control participants were not assessed for SI/SA, they were only considered controls for the analyses on suicide attempts and suicidal ideations in the absence of psychiatric illness. One German study center (Münster) recruited participants for both PsyCourse and FOR2107. To avoid overlap, participants from Münster were excluded from the PsyCourse sample for these analyses.

#### Seoul National University Bundang Hospital PsyGen Cohort Waves 1+2 (SNUBH)

Participants were recruited from Seoul National University Bundang Hospital (SNUBH), consisting of individuals diagnosed with psychiatric disorders such as major depression, bipolar disorder, schizophrenia, anxiety disorder, PTSD, ADHD, alcohol use disorder (AUD), and obsessive-compulsive disorder (OCD). These participants were recruited from both inpatient and outpatient settings. Diagnoses were confirmed by board-certified psychiatrists using structured interviews, a comprehensive review of case records, and other available clinical data, following DSM-5 criteria. Information on suicidal ideation and suicide attempts was obtained using the suicidality module of the Mini-International Neuropsychiatric Interview (MINI), and all responses were reviewed in conjunction with medical charts and other relevant clinical records to ensure accurate case classification.

#### UK Biobank (UKB)

The UK Biobank is a prospective cohort study of approximately 500,000 individuals, recruited from 23 centres across the United Kingdom [61]. Extensive phenotypic data are available. For the current study, data was drawn from the death register (Resource 115559 on http://biobank.ctsu.ox.ac.uk) and questionnaires, including two online follow-up questionnaires focussing on mental health (Resources 22 and 2800 on http://biobank.ctsu.ox.ac.uk) [62,63]. Participants provided responses to questions on self harm during two online mental health follow-up questionnaires. In data released in 2016, 157,297 participants were asked "Have you deliberately harmed yourself, whether or not you meant to end your life?" (Data-Field 20480), of which 6861 responded "yes". In further data released in 2023, 169,810 participants were asked the same question (Data-Field 29111), of which 7805 responded "yes". Data from 2023 overlap with data from 2016. Participants provided responses to questions on suicide attempts during two online mental health follow-up questionnaires. In data released in 2016, the 6861 participants who reported deliberate self-harm were asked "Have you harmed yourself with the intention to end your life?" (Data-Field 20483), of which 3558 responded "yes". In further data released in 2023, the 7805 participants who reported deliberate self-harm were asked the same question (Data-Field 29116), of which 4111 responded "yes". Individuals who endorsed this question in the 2023 data were classified as cases. Individuals who endorsed this question in 2016 and did not answer the question in 2023 were additionally defined as cases. Individuals who gave contradictory answers, that is those who endorsed this question in 2016 and then answered but did not endorse this question in 2023, were excluded from analysis. Primary and secondary ICD10 codes related to causes of death were extracted from death registers (data updated September 2023) and individuals with ICD10 codes X60-X84, Y87.0, or U03 were classified as cases for the suicide death phenotype. Individuals who a) answered but did not endorse the question pertaining to self-harm in 2016 and in 2023, or b) endorsed the question pertaining to self-harm and then answered but did not endorse the question pertaining to suicide attempt were considered as controls, and were included as controls if they were not in the death register in September 2023 (that is, they were still alive), or if they were in the death register and did not meet criteria for suicide death.

#### University of Utah - European sample (UTAH EUR)

Ascertainment, case, and control definitions are described in a previously published GWAS of SD in the Utah cohort[64]. The same procedures were used to include additional case and control samples not available for the previous GWAS. All Utah SD samples arise from the Utah Office of the Medical Examiner (OME), which obtained postmortem samples from all “likely suicides” based on initial judgements, and subsequent death investigations result in final death certificate determinations. Briefly, suicide cause-of-death determination results from a detailed investigation, done by the centralized Utah State Office of the Medical Examiner, of the scene of the death and circumstances of death, determination of medical conditions by full autopsy, review of medical and other public records concerning the case, interviews with survivors, in addition to standard toxicology workups. Suicide determination is traditionally made quite conservatively due to its impact on surviving relatives, and all suicides (87%) and likely suicides (13%) were included as SD cases here. DNA from suicide deaths was extracted from whole blood using the Qiagen Autopure LS automated DNA extractor (www.qiagen.com). Ancestry matched controls reflect a general population sample obtained from the Generation Scotland Scottish Family Health Study[65] and the UK10K Rare Genetic Variants in Health and Disease Project[66], as described in Docherty et al.[64]. The Generation Scotland Scottish Family Health

Study is a population-based sample and only founders were used as controls to remove confounding resulting from intra-dataset relatedness. The UK10K controls included population-based samples as well as individuals with selected health phenotypes, as the UK10K study was designed to investigate a range of complex and rare disorders, including obesity, neurodevelopmental and psychiatric conditions, and cardiovascular and metabolic diseases. All controls were living at the time of sample collection.

#### University of Utah - Latin American sample (UTAH LAT)

A total of 867 genotyped cases from the population-based UTAH suicide death cohort (above) were selected for significant AMR ancestry admixture based on ancestry PCA. Controls for this GWAS were 2,043 ancestry-matched individuals from central Mexico drawn from a screened subset of the MxGDAR (Mexican Genomic Database for Addiction Research) cohort. MxGDAR is an epidemiological cohort composed of 3,393 healthy individuals recruited in Mexico from the 2016 National Survey of Drug, Alcohol, and Tobacco Use. MxGDAR was performed in accordance with the Declaration of Helsinki and approved by the Ethics and Research Committee at the National Institute of Genomic Medicine and the National Institute of Psychiatry in Mexico (INMEGEN and INPRFM).

#### Vanderbilt University Medical Center (VUMC)

Deidentified clinical data were extracted from VUMC's Synthetic Derivative, which stores EHR data from over 3.2 million patients receiving care at Vanderbilt University Medical Center (VUMC)[67]. VUMC is an academic medical center in Nashville Tennessee that manages over 2 million patient visits every year across Tennessee and its neighboring states. Genetic data came from VUMC's biobank BioVU which includes over 300,000 DNA samples.

Cases of suicidal ideation (SI) were ascertained using three sources of structured data (International Classification of Diseases, 9th/10th Revision, Clinical Modification diagnostic codes), semi-structured data (psychiatric hospital screening questionnaire response) and unstructured data (NLP on clinical notes). Both ICD-9/10-CM of suicidal ideation (ICD9: V62.84, ICD10: R45.851) were used as prior studies in suicide attempt ascertainment have shown improvement of PPV using ICD-10 (PPV 85% using ICD-10 compared to PPV 58.6% using ICD-9). The PPV of ICD10CM code for SI was 96% in a prior study, while the PPV of ICD-9 code for SI in a different study was 55%. Despite lower PPV of the ICD-9 codes, the decision to include cases was made to increase the overall case number for genetic studies. The psychiatric hospital screening questionnaire given to patients at the Vanderbilt Psychiatric Hospital included a binary question of whether a patient has suicidal ideation. The NLP method for extracting SI cases from EHR notes was described in Bejan et al[68], where Google's word2vec method was used to generate a list of seed words that describe suicidal ideation in notes (also called query terms), and an information retrieval approach was employed to rank patients based on the similarity between their notes and the suicidal ideation query vector. The NLP approach also proposed a probabilistic model to compute the precision of the top K highest ranked patients (P@K) for any K value. In this study, from the ranked list of patients extracted by the NLP method, we selected as SI cases the top K highest ranked patients such that all patients in the list above the cutoff value K have a precision of at least 80% (i.e,. P@K=80%). SI cases were defined using an 80% PPV cutoff.

Individuals with any evidence of SI among the three sources listed above (screeners, ICD-9/10-CM code, NLP above PPV cutoff) were considered a SI case to maximize power for further genetic analyses.

Controls were defined as individuals who matched all the following criteria: 1) negative or absence of positive assertions to SI in psychiatric forms of suicide assessment, 2) absence of SI ICD codes, or 3) individuals that were not included in the 80% PPV cutoff for NLP ascertainment of SI. Individuals below the 80% PPV cutoff without ICD9/10 codes, manual validation, or forms were excluded from the study to avoid including those who may not be true controls due to insufficient evidence to be a SI case.

##### Yale-Penn

Participants for the discovery GWAS in this study were recruited from five sites in the eastern United States, for studies of the genetics of drug or alcohol dependence - the Yale-Penn study[69–71]. All participants were interviewed using the Semi-Structured Assessment for Drug Dependence and Alcoholism (SSADDA), which contains several items relevant to suicidal behavior, as discussed below. Participants provided written informed consent and the study was approved by the institutional review board at each participating site (Yale Human Research Protection Program, VA CT HSC, University of Pennsylvania Institutional Review Board, University of Connecticut Human Subjects Protection Program, Medical University of South Carolina Institutional Review Board for Human Research, and the McLean Hospital Institutional Review Board). Section N of the SSADDA includes 12 items that assess suicidal behaviors, specifically evaluating history of suicidal ideation, planning, and attempts. Participants reporting suicidal ideation were screened for prior suicide attempts. Individuals who endorsed a past attempt were excluded from both ideation case and control groups. For the suicide attempt phenotype, all individuals who reported a suicide attempt were retained as cases, while controls were required to have no history of suicide attempt. Additionally, data on death by suicide were extracted from the National Death Index. Eight individuals identified as having died by suicide were excluded from all analyses.

#### COHORT GENOTYPING, QC, IMPUTATION AND ANALYSIS

##### The Adolescent Brain and Cognitive Development $^{SM}$ Study (ABCD Study®)

Genotypes are from the Phase 3.0 release of the ABCD Study®. Saliva samples were genotyped on the Smokescreen array [72] by the Rutgers University Cell and DNA Repository (now SAMPLED; https://sampled.com/). Genotyped calls were aligned to GRC37 (hg19). The Rapid Imputation and COmputational PIpeLIne for Genome-Wide Association Studies

(RICOPILI) [73] was used to perform quality control (QC) on the 11,099 individuals with available ABCD Study® phase 3.0 genotypic data, using RICOPILI's default parameters. The 10,585 individuals who passed QC checks were matched to broad caregiver-reported racial groups using the ABCD Study® Parent Demographics Survey (i.e., caregiver responding to the survey question "What race do you consider the child to be?"). 6,787 caregivers indicated that their child's race was "White," and 5,561 of those individuals did not endorse any Hispanic ethnicity. Principal component analysis (PCA) in RICOPILI was used to confirm the genetic ancestry of these individuals by mapping onto the 1000 Genomes reference panel, resulting in a PCA-selected European-ancestry subset of 5,556 individuals.The TOPMed imputation reference panel was used for imputation [74]. Imputation dosages were converted to best-guess hard-called genotypes, and only SNPs with Rsq > 0.8 and MAF > 0.01 were kept for PRS analyses. GWAS was performed using an additive logistic regression adjusting for 5 ancestry principal components (PLINK 1.9).

#### Australian Genetics of Bipolar Disorder Study + Australian Genetics of Depression Study (GBP+AGDS)

Genotyping for both cohorts was performed across six batches using the Illumina Global Screening Array (GSA). The first three batches were primarily from the Australian Genetics of Depression Study (AGDS), while the subsequent batches included a combination of AGDS and the GBP samples, with the latter predominating. Depending on the batch, samples were genotyped on either the GSA v1.0 or v3.0 arrays (v2.0 was not used).

QSkin samples were genotyped using Illumina GSA v1.0 arrays as a sixth, separate batch. Quality control (QC) was conducted within each batch, after which datasets were merged at markers passing QC across all batches. Ancestry inference was conducted by projecting study samples onto principal components from the 1000 Genomes Project, retaining individuals within six standard deviations of the European centroid. Imputation was performed using the Michigan Imputation Server on the combined dataset following QC.

#### Avon Longitudinal Study of Parents and Children (ALSPAC)

ALSPAC children were genotyped using the Illumina HumanHap550 quad chip genotyping platforms by 23andme subcontracting the Wellcome Trust Sanger Institute, Cambridge, UK and the Laboratory Corporation of America, Burlington, NC, US. The resulting raw genomewide data were subjected to standard quality control methods on 9,915 subjects and 550,000 SNPs. Individuals were excluded on the basis of gender mismatches; minimal or excessive heterozygosity; disproportionate levels of individual missingness (>3%) and insufficient sample replication (IBD < 0.8). Population stratification was assessed by multidimensional scaling analysis and compared with Hapmap II (release 22) European descent (CEU), Han Chinese, Japanese and Yoruba reference populations; all individuals with non-European ancestry were removed by removing samples that clustered outside the CEU HapMap2 population using this multidimensional scaling of genome-wide IBS pairwise distances. SNPs with a minor allele frequency of < 1%, a call rate of < 95% or evidence for violations of HardyWeinberg equilibrium

(P < 5E-7) were removed. Cryptic relatedness was measured as proportion of identity by descent (IBD > 0.1). Related subjects that passed all other quality control thresholds were retained during subsequent phasing and imputation. 9,115 subjects and 500,527 SNPs passed these quality control filters.
ALSPAC mothers were genotyped using the Illumina human660W-quad array at Centre National de Genotypage (CNG) and genotypes were called with Illumina GenomeStudio. PLINK (v1.07) was used to carry out quality control measures on an initial set of 10,015 subjects and 557,124 directly genotyped SNPs. SNPs were removed if they displayed more than 5% missingness or a Hardy-Weinberg equilibrium P value of less than 1.0e-06. Additionally, SNPs with a minor allele frequency of less than 1% were removed. Samples were excluded if they displayed more than 5% missingness, had indeterminate X chromosome heterozygosity or extreme autosomal heterozygosity. Samples showing evidence of population stratification were identified by multidimensional scaling of genomewide identity by state pairwise distances using the four HapMap populations as a reference, and then excluded. Cryptic relatedness was assessed using an IBD estimate of more than 0.125 which is expected to correspond to roughly 12.5% alleles shared IBD or a relatedness at the first cousin level. Related subjects that passed all other quality control thresholds were retained during subsequent phasing and imputation. 9,048 subjects and 526,688 SNPs passed these quality control filters.

#### The Berlin Research Initiative for Diagnostics, Genetic and Environmental Factors of Schizophrenia (BRIDGE-S)

DNA samples either derived from whole-blood EDTA samples or saliva samples collected using 1.0 ml OraGene (Genotek, Ottawa, Ontario, Canada) saliva DNA-Self-Collection kits. All samples were genotyped using the Illumina Infinium GSA MD v1-3 at the Genomics Core Facility, Erasmus MC, University Medical Center Rotterdam, The Netherlands. Standardized quality control and imputation and analyses were performed centrally using RICOPILI (Rapid Imputation for COnsortias PIpeLIne) [73]. Briefly, the quality control parameters for retaining SNPs and subjects were: SNP missingness < 0.05 (before sample removal), subject missingness < 0.02, autosomal heterozygosity deviation (Fhet < 0.2), SNP missingness < 0.02 (after sample removal), difference in SNP missingness between psychiatric cases and healthy controls < 0.02 and SNP Hardy-Weinberg equilibrium ($P > 10^{-10}$ in psychiatric cases, $P > 10^{-6}$ in healthy controls). Genotype imputation was performed using the prephasing/imputation stepwise approach implemented in Eagle (v2.3.5; https://alkesgroup.broadinstitute.org/Eagle/) [75] and Minimac3 (https://genome.sph.umich.edu/wiki/Minimac3) [76] to the Haplotype Reference Consortium (HRC) reference panel (v1.0) [77]. SI and SA GWAS were performed using PLINK 1.9 by comparing imputed marker dosages under an additive logistic regression model between cases and controls [78]. Principal components (PCs) generated using EIGENSTRAT were used as covariates in all GWAS as required, to control for population stratification [79].

#### Brazilian High Risk Cohort for Mental Health Conditions (BHRC)

Samples were genotyped at the Broad Institute of MIT and Harvard using the Global Screening Array (GSA-MD v1.0). Standardized quality control was performed centrally using RICOPILI

(Rapid Imputation for COnsortias PIpeLIne) [73]. Briefly, the quality control parameters for retaining SNPs and subjects were: SNP missingness < 0.05 (before sample removal), subject missingness < 0.02, autosomal heterozygosity deviation (Fhet < 0.2), SNP missingness < 0.02 (after sample removal), difference in SNP missingness between cases and controls < 0.02 and SNP Hardy-Weinberg equilibrium (P > 10−10 in cases, P > 10−6 in controls). Phasing was performed with SHAPEIT5 [80], using information from both the family structure (trios and duos) and the TGP+HGDP reference panel [81]. Imputation was performed on the TOPMed server [74]. Since admixture is known to exist in Brazilian populations, we ran ADMIXTURE [82] with cross-validation for K values up to 4. CV errors indicated that a tri-hybrid admixture model between AFR, LAT and EUR fit the population best. We used a 5% ancestry inclusion threshold to restrict to a set of 3-way admixed individuals. The total sample of admixed individuals included 75 cases and 561 controls for the SA analysis and 60 cases and 522 controls for the SI analysis. GWAS of each ancestry component of the sample was conducted using the separate GWAS version of TRACTOR [83]. To calculate approximate sample sizes for the GWAS in each ancestry tract, we distributed the total sample size for the cohort according to the ancestry fractions. To do so, for cases and controls separately, we counted the number of alleles observed in each ancestry tract at each SNP (e.g., if there were 10 100% EUR individuals, there would be 20 alleles observed at each SNP in the EUR tract), then divided that by the total number of alleles present at that SNP for the entire sample (e.g., if the sample included 40 individuals, that would be 80 alleles assuming no missingness). Those proportions were averaged across all SNPs to represent the ancestry fractions in all the cases and all the controls. The total N cases and N controls were then multiplied by those fractions to get Ns for each ancestry. For example, the average proportion of alleles observed at SNPs in SI controls (N = 522) for each ancestry were 29.3% for AFR, 56.9% for EUR, and 13.8% for LAT. Therefore, we calculated N controls for SI to be 153 for AFR, 297 for EUR, and 72 for LAT.

#### China, Oxford and VCU Experimental Research on Genetic Epidemiology (CONVERGE)

DNA sequencing, variant calling, and imputation have been previously described [12]. Briefly, sequencing reads were aligned to GRCh37.p5 with Stampy (c.10.17) [84] using default parameters after filtering out reads of poor quality. Variant discovery and genotyping at all SNPs in the 1000 Genomes Phase 1 East Asian (ASN) [85] was performed using the GATK's UnifiedGenotyper (version 2.7-2-g6bda569). Imputation was performed using BEAGLE (version 3.3.2) [86]. GWAS were performed using PLINK 1.9 by comparing imputed marker dosages under an additive logistic regression model between cases and controls. Based on prior studies, the first two principal components were included as covariates; these were derived from an eigen-decomposition of the genetic relatedness matrix [12,87]. Variants were excluded from analysis if they had an INFO score <0.3, minor allele frequency <0.001, or HWE p<1e-7.

#### Collaborative Study on the Genetics of Alcoholism (COGA)

COGA participants were genotyped on four different genotyping arrays: the Illumina 1 M, Illumina OmniExpress 12VI, and Illumina 2.5 M (Illumina, San Diego, CA),and Smokescreen (BioRealm LLC, Walnut, CA) [88]. EUR individuals were imputed to Haplotype Reference Consortium (HRC) imputation panel, for AFR individuals Consortium on Asthma among African-ancestry Populations in the Americas (CAAPA) imputation panel was used. Only variants with non-A/T or C/G alleles, missing rates <5%, MAF >3%, and HWE P-values >.0001 were used for imputation. Imputed variants with R2 < .30 were excluded, and genotype probabilities were converted to genotypes if probabilities ≥.90. Mendelian inconsistencies cleaned and genotyped and imputed variants with missing rates <5%, MAF ≥1% and HWE P-values >1E-6 were included in analyses. GWAS was performed using the SAIGE method [89] to take into account family-based structure. First five ancestral PCs used as covariates.

#### Columbia University (CUINT)

The genotype data were cleaned using the quality control steps suggested by PGC-sui consortium protocol. Markers were retained if they had a minor allele frequency (MAF) of 1% or more, a call rate ≥ 95%, and no significant departures from Hardy–Weinberg Equilibrium (HWE p value ≥ 1e-06). Samples with sex violations, genotyping call completeness <95 heterozygosity rate deviating more than 3 SD from the mean, and duplicated individuals were excluded. PCA in PLINK, and comparison to 1000 genome populations were used to exclude individuals of non-European ancestry. The majority of samples from all three sites were found to be superimposed on the European population and outliers were removed. Subsequently, the genotype data were imputed using the Michigan Imputation Server. 1000 genome population were chosen as reference panels, and phasing and imputation were performed using Eagle v2.4 and Minimac4, respectively. Among the imputed genotypes, variants whose MAF <0.01 and R2<0.3 were excluded. Further, the GWAS was run with PLINK using logistic regression adjusting for the top five PC scores.

#### The Consortium on Vulnerability to Externalizing Disorders and Addictions (CVEDA)

Initial QC followed the standardized quality control implemented in RICOPILI. Briefly, the quality control parameters for retaining SNPs and subjects were: SNP missingness < 0.05 (before sample removal), subject missingness < 0.02, autosomal heterozygosity deviation ($F_{het}$ < 0.2), SNP missingness < 0.02 (after sample removal), MAF ≥ 0.01, difference in SNP missingness between cases and healthy controls < 0.02 and SNP Hardy-Weinberg equilibrium ($P > 10^{-10}$ in cases, $P > 10^{-6}$ in healthy controls). Phasing was performed using Eagle v2.4 and imputation was performed using Minimac4 with Haplotype Reference Consortium (HRC) Version r1.1 2016. “Hard call” genotypes were filtered for an imputation info score (2) > 0.3 and a minor allele frequency > 0.01. Ancestry principal component analysis (PCA) was performed using PLINK

1.9. GWAS were performed using an additive logistic regression adjusting for the first five ancestry PCs implemented in PLINK 2.

#### Estonian Biobank (ESTBB)

Genotyping of DNA samples from the Estonian Biobank was done at the Core Genotyping Lab of the Institute of Genomics, University of Tartu using the Illumina Global Screening Arrays (GSAv1.0, GSAv2.0, and GSAv2.0_EST). Altogether 206,448 samples were genotyped and then PLINK format files were created using Illumina GenomeStudio v2.0.4. During the quality control all individuals with call-rate < 95% or mismatching sex that was defined based on the heterozygosity of X chromosome and sex in the phenotype data, were excluded from the analysis. Variants were filtered by call-rate < 95% and HWE p-value < 1e-4 (autosomal variants only). Variant positions were updated to Genome Reference Consortium Human Build 37 and all variants were changed to be from TOP strand using reference information provided by Dr. Will Rayner from the University of Oxford (https://www.well.ox.ac.uk/~wrayner/strand/). After QC the dataset contained 202,910 samples for imputation. Before imputation variants with MAF<1% and Indels were removed. Prephasing was done using the Eagle v2.3 software [75] (number of conditioning haplotypes Eagle2 uses when phasing each sample was set to: --Kpbwt=20000) and imputation was carried out using Beagle v.18May20.d20 [86,90] with an effective population size ne=20,000. As a reference, Estonian population specific imputation reference of 2297 WGS samples was used [91]. Further, EstBB samples were combined with the 1000 genomes phase 3 dataset for ancestry analysis. Genetic principal components were calculated using a subset of quality controlled and pruned genotyped SNPs. This was further used to identify and remove samples that deviated from the main cluster. GWAS were performed using REGENIE [92] to account for the unbalanced case-control ratios in the study.

#### FinnGen

Genotyping, QC, imputation, and analysis for this cohort have been previously published and described in full [22].

#### Genetic Investigation of Suicide and SA Wave 1 (GISS 1)

SNP genotyping was done using the HumanOmni1-Quad_v1 chip (Illumina Inc.) at the SNP&SEQ Technology Platform facility (snpseq.medsci.uu.se), assaying ~1 million SNPs with each trio plated consecutively. For the raw data, 96.7% of SNPs had call rate >99%, >99.99% of calls were reproducible, >99.99% of family-wize calls had no mendelian errors, and duplicated individuals could be ruled out. SNPs were filtered to obtain call rates >= 95%, hardy weinberg equilibrium (HWE) exakt P = 10-6, minor allele frequency (MAF) = 0.01 and no mendelian errors, whereby 739,780 autosomal- and 17,501 X- chromosomal SNPs remained. Phased reference panels (1000 genomes, phase 1; filtered for 1.00<MAF<0.005), BEAGLE v.3.3.2 and utils were downloaded (faculty.washington.edu/browning) [93]. SNPs were checked against the phased EUR individuals in the 1000 genomes reference-panel, for inconsistencies in SNP-strands, -positions, -names, MAFs, linkage disequilibrium (LD) and number of alleles, using the available check_strands python routines. 729,956 autosomal SNPs remained for imputation using ~9 million reference panel SNPs (1.00 > MAF > 0.005). The X-chromosome was not imputed. Phasing (nsamples=2) and imputation (nsamples=1) were executed separately, running one chromosome at a time in low-memory mode on a desktop PC. Only SNPs imputed with Beagle allelic R2 = 0.7 were retained. ~5.5% of SNPs had a rare frequency (MAF < 0.01). The net imputation SNP gain after accounting for LD with r2-threshold < 0.8 pruning and MAF > 0.01, was from 450,348 autosomal SNPs pre- imputation to 1,035,345 autosomal SNPs post-imputation, i.e. ~2.3 fold. Quantiles vs quantiles (QQ) plots showed that observed SNP P-values followed the uniform null (genomic inflation = 1.002), as previously described [24]. For this analysis, the ~6.8 million post-imputation SNP data was converted into a case-control sample by use of --tucc command in plink v.1.07 (660 cases and 660 controls; each control is a non-SA pseudo-sib, matched to a case on all other features), followed by analysis with --assoc --ci 0.95 in plink v.1.9.

#### Genetic Investigation of Suicide and SA Wave 2 (GISS 2)

SNP genotyping was performed in the year 2018 by using the Global screening array MD (BeadChip GSAMD-24v2-0_20024620_A1; Illumina Inc.) at the SNP&SEQ Technology Platform facility (snpseq.medsci.uu.se), assaying 760k markers (mapped with build 37) for suicide attempt (SA) cases and healthy volunteer (HV) controls, plated interchangeably. For the raw data of a total of 1883 genotyped subjects, 96.15% of SNPs had call rate >98%, >99.99% of calls were reproducible, and for the available family-data (~300 nuclear or one-parent trios), 99% of calls showed correct inheritances. Subsequently, the GWAS QC results from the genotyping facility (misclassified parenthood or sex, any unexplained duplicates / excess genetic relationships, or other suspect genotyping performances) was complemented with our own sample quality-control procedures on demographic (cross check verifications of data entries and various reasonability checks of entries) and psychometric data (nay-saying, random responding, excess entry missingness or inter-subject response-duplicates). Further removal of all parent data, resulted in a final GWAS genotype “GISS2” sample consisting of n=564 SA cases and n=513 non-SA healthy volunteer (HV) controls. ~2.2k markers with HapMap CEPH control subject discordances were removed and a mean of 175 markers per subject showing XY errors were set to missing, resulting in a genotyping rate of 99.1% for 757.7k markers in the final set of 1077 case and control subjects shared with PGC SUI. The data shared with PGC SUI then underwent the standard QC, imputation and analysis implemented in RICOPILI as described previously for other cohorts.

#### The Genes, Environment, and Development Initiative (GEDI)/Great Smoky Mountains Study (GSMS) & Virginia Twin Study of Adolescent Behavioral Development (VTSABD)

GSMS and VTSABD were genotyped using Illumina Human660W-Quad v1 and imputation was performed on the TOPMed server [74]. Liftover to hg19 was performed using CrossMap [94] and conversion-unstable positions were removed [95]. Samples were filtered using Mahalanobis distance applied to the top 10 population stratification principal component scores, to ensure EA, using 1000 Genomes samples as a population structure reference panel. GWAS were performed separately in each genetic ancestry group using an additive logistic regression adjusting for PCs 1-5 and 7 in EUR samples and PCs 1-6 in LAT samples.

#### Genomic Research and Epidemiological Studies for Affective Disorders in Taiwan (GREAT)

The GREAT samples were genotyped at genomics research center in Academia Sinica in Taiwan, and genomic variants were obtained using the Affymetrix Axiom Genome-Wide CHB Array, Affymetrix Axiom Genome-Wide TWB 1.0 Array, and Axiom Genome-Wide TWB 2.0 Array, totaling of 8,733,528 markers initially. The reference genome version was GRCh38/hg38. Imputation was conducted using diverse reference panel information from 97,256 deeply sequenced human genomes (NHLBI Trans-Omics for Precision Medicine). We implemented a series of quality control processes before and after imputation; we excluded SNPs with imputation information < 0.7, data missing in > 2% of the sample, minor allele frequency < 0.1%, or deviated from Hardy-Weinberg equilibrium ($P < 1 \times 10$-6), with 5,727,556 markers remained in GWAS analysis using PLINK.

#### Grady Trauma Project (GTP)

Genotyping for the Grady Trauma Project was performed using the Omni-Quad 1M Bead Chip. Quality control and imputation (1000 Genomes Phase 3-hg19) were performed by using the Psychiatric Genomics Consortium PTSD Workgroup guidelines [96]. Only individuals with African American ancestry based on SNPweights software17 were included in the models. Principal components for ancestry were calculated according to the PGC guidelines in each separate ancestry group [96]. For each model, GWAS was performed using an additive logistic regression adjusting for 5 ancestry principal components (PLINK 1.9).

#### The Individualized Medicine: Pharmacogenetics Assessment and Clinical Treatment Study (IMPACT)

DNA for the IMPACT study was collected using Oragene saliva kits (Genetech) and purified using Chemagen. The samples were run on Infinium Omni 2.5 chips (Illumina). Quality control steps were performed similar to those described previously [97]. SNPs with minor allele frequency of <0.01, genotyping rate of >0.05, or significant deviation from Hardy-Weinberg Equilibrium (p<1e-6) were excluded, and participants with genotyping missingness >0.05, outlying heterozygosity (>3SD from mean), and relatedness/duplicates (keeping one participant from each set of related individuals) were removed. Participants who were genetically non-Europeans or self-reported non-Europeans or were population outliers (>6SD from mean) were also removed. Whole-genome imputation was carried out using Minimac4 executed on the Michigan Imputation Server with haplotype phasing (Eagle v2.4) and the 1000 Genomes Phase 3 reference panel. After imputation, SNPs with R-squared values of less than 0.9 were excluded. Binary suicidal ideation status (985 cases, 689 controls) was analyzed using glm firth-fallback in PLINK2 with the first five principal components included as covariates.

#### International Borderline Genomics Consortium - Central European subset (IBGC CE)

Details on a subset of the sample have been published previously [98]. Samples were genotyped using the Infinium PsychArray-24 Bead Chip (Illumina, San Diego, CA, USA). Quality control and imputation were carried out using the RICOPILI GWAS pipeline [73] (see below). Briefly, the exclusion criteria for SNPs and subjects in the first round of quality control were: genotyping call rate for given SNPs or individuals < 98%, difference in SNP genotyping call rate between cases and controls > 2%, deviation of autosomal heterozygosity from the mean (|Fhet|>0.2), or a deviation from Hardy-Weinberg equilibrium ($p < 1 \times 10^{-10}$ in cases; $p < 1 \times 10^{-6}$ in controls). Imputation was conducted using a publicly available reference panel consisting of 54,330 phased haplotypes with 36,678,882 variants from the haplotype reference consortium (EGAD00001002729) and the prephasing/imputation stepwise approach in EAGLE/MINIMAC3 (default parameters and a variable chunk size of 132 genomic chunks) [99]. Relatedness testing and population structure analysis were performed using a subset of 55,001 SNPs that fulfilled strict quality criteria after imputation (INFO > 0.8, missingness < 1%, minor allele frequency > 0.05), and which had been subjected to LD pruning ($r^2 > 0.02$) in the second round of quality control. In the case of cryptically related subjects with pi-hat > 0.2, one member of each pair was removed at random following the preferential retention of cases over controls. Principal components (PCs) were estimated from the quality-controlled genotypic data, and phenotype association was tested using logistic regression.

#### International Borderline Genomics Consortium - German Borderline Genomics Consortium subset (GBGC)

Details on the sample have been published previously [43]. Present analyses in the present manuscript are based on an updated quality control and imputation carried out using the RICOPILI GWAS pipeline [73] (see below), and have been reported before [100,101]. DNA extraction was carried out using the chemagic Magnetic Separation Module I (Chemagen Biopolymer-Technologie, Baesweiler, Germany) and samples were genotyped using the Infinium PsychArray-24 Bead Chip (Illumina, San Diego, CA, USA). Individuals and SNPs were removed if they met any of the following exclusion criteria in the first round of quality control: genotyping call rate for given SNPs or individuals <98%, difference in SNP genotyping call rate between cases and controls >2%, deviation for the autosomal heterozygosity from the mean (|Fhet|>0.2), or a deviation from Hardy-Weinberg equilibrium ($p<1 \times 10^{-10}$ in cases; $p<1 \times 10^{-6}$ in controls). Genotype data were imputed using a publicly available reference panel consisting of 54,330 phased haplotypes with 36,678,882 variants from the haplotype reference consortium (EGAD00001002729) with the pre-phasing/imputation stepwise approach in EAGLE/MINIMAC3 (default parameters and a variable chunk size of 132 genomic chunks) [99]. In the second round of quality control, relatedness testing and population structure analysis were performed using a SNP subset that fulfilled strict quality criteria after imputation (INFO >0.8, missingness <1%, minor allele frequency >0.05), and which had been subjected to LD pruning ($r^2$>0.02). This subset comprised 66,240 SNPs. For cryptic relatives with pi-hat >0.2, one member of each pair was removed at random following the preferential retention of cases over controls. The thresholds for exclusion of genetic outliers on the first four principle components were determined via visual inspection.

#### International Borderline Genomics Consortium - Spanish subset (IBGC SPAIN)

Samples were genotyped using the Infinium PsychArray-24 Bead Chip (Illumina, San Diego, CA, USA). Quality control and imputation were carried out using the RICOPILI GWAS pipeline [73] (see below). Briefly, the exclusion criteria for SNPs and subjects in the first round of quality control were: genotyping call rate for given SNPs or individuals < 98%, difference in SNP genotyping call rate between cases and controls > 2%, deviation of autosomal heterozygosity from the mean (|Fhet|>0.2), or a deviation from Hardy-Weinberg equilibrium ($p < 1 \times 10^{-10}$ in cases; $p < 1 \times 10^{-6}$ in controls). Imputation was conducted using a publicly available reference panel consisting of 54,330 phased haplotypes with 36,678,882 variants from the haplotype reference consortium (EGAD00001002729) and the prephasing/imputation stepwise approach in EAGLE/MINIMAC3 (default parameters and a variable chunk size of 132 genomic chunks) [76,99]. Relatedness testing and population structure analysis were performed using a subset of 55,001 SNPs that fulfilled strict quality criteria after imputation (INFO > 0.8, missingness < 1%, minor allele frequency > 0.05), and which had been subjected to LD pruning ($r^2$ > 0.02) in the second round of quality control. In the case of cryptically related subjects with pi-hat > 0.2, one member of each pair was removed at random following the preferential retention of cases over controls. Principal components (PCs) were estimated from the quality-controlled genotypic data, and phenotype association was tested using logistic regression.

#### Japan

The details of genotyping, QC and imputation are reported previously [44]. Briefly, samples were genotyped using Illumina HumanOmniExpress and HumanOmniExpressExome BeadChips for the 1st and 2nd set of samples ascertained, respectively. We performed QC using PLINK 1.9. Firstly, for each set, we excluded SNPs with a call rate < 0.98 and MAF < 0.01, and those with $p < 1.0 \times 10^{-6}$ for HWE in controls. Related individuals were excluded (PI_HAT ≥ 0.175). We performed PCA, and confirmed all of the above subjects were in the Japanese cluster. After estimating haplotypes using SHAPEIT2 (v2.r778), we performed genotype imputation by Minimac3 (1.0.13) using ALL samples in the 1000 Genomes Project phase 3v5 as a reference. In order to finalize the summary statistics of imputed data which consist of 746 suicide decedents and 14,049 controls, we combined the summary statistics of imputed variants of the 1st and 2nd control sets as implemented in Rvtests software, performing meta-analysis with METAL software using a fixed effects model with inverse-variance weighted approach, with adjustment for 10 PCs.

#### Janssen Waves 1 + 2

Genotyping, QC, imputation, and analysis for these cohorts have been previously published and described in full [22].

#### The Lundbeck Foundation Initiative for Integrative Psychiatric Research (iPSYCH)

Genotyping, QC and imputation procedures for iPSYCH 2012 cohort were conducted in the same manner as described for previous GWAS [102–104]. Genotyping waves with less than 50 SB cases were removed from the analysis followed by removal of related individuals, duplicated samples, and restricting individuals to European population and Danish origin only. After the filtering of genotyping data 7,003 SB cases and 52,227 non-SB controls were identified. The GWAS analysis was adjusted for sex, the first 10 principal components of genetic ancestry and genotyping batch. Association analyses were performed and are reported only for variants for which P-value was calculated and for variants with MAF ≥ 1% or ≤ 99% in the control group. The GWAS analysis of non-fatal SA (i.e., excluding SD cases) has been published previously [105].

#### Mental Illness Research Education and Clinical Center (MIRECC)

This study utilizes genome-wide imputed genotypes; the QC pipeline and procedures used have been previously described in detail [46,47]. DNA was extracted from whole blood and genotyped in three different batches on three different Illumina BeadChips: HumanHap650 BeadChip, Human1M-Duo BeadChip, and the HumanOmni2.5 BeadChip (Illumina, San Diego, CA). Data from the three batches was merged to produce the largest overlapping set and imputed using a global reference panel from 1000Genomes. Imputed probes were required to have 90% certainty or better and were removed altogether if the call rate across all samples was < 97%. Additionally, imputed probes were removed if Hardy-Weinberg Equilibrium (HWE) was < $10^{-6}$ in the controls or if minor allele frequency (MAF) was < 1%.

#### Million Veteran Program (MVP)

Genotyping, QC, imputation, and genetic ancestry assignment procedures for the MVP sample were conducted in the same manner as described for previous suicidality GWAS in MVP [48,49]. The SI GWAS summary statistics are from a published study that has been previously described in full [48]. For the SA and SB GWAS, ancestry-specific GWAS were performed with PLINK2 [78], covarying for genetic PCs. The SD GWAS was performed using the SAIGE method [89], implemented in SAIGEgds [106], to account for the unbalanced case-control ratio.

#### National Longitudinal Study of Adolescent to Adult Health (Add Health)

Genotyping, quality control, and imputation has been reported previously for this cohort [107].

#### Predictors For ECT study (PREFECT)

Genotyping for PREFECT [51,52] was performed on the Illumina GSA-MD array at Life & Brain GmbH (Bonn, Germany). Quality control followed the PGC Ricopili pipeline, excluding samples with genotype missingness >0.02, sex discrepancies, heterozygosity outliers, relatedness (IBD >0.2), or non-European ancestry (>3 SD from 1000 Genomes EUR on PC1/PC2), and SNPs with call rate <0.99, case–control missingness difference >0.005, MAF <0.01, or Hardy–Weinberg $p<1\times10^{-6}$. Post-QC data were imputed to the HRC r1.1 reference panel using the Sanger Imputation Service with Eagle2 for phasing and PBWT for imputation. GWAS was conducted using PLINK1.9 adjusting for five PCs.

#### Psychiatric Genomics Consortium Bipolar Disorder (PGC BD)

Cohorts were genotyped following their local protocols, after which standardized quality control and imputation and analyses were performed centrally using RICOPILI (Rapid Imputation for COnsortias PIpeLIne), for each cohort separately [73]. These procedures have been described in detail previously [54]. Briefly, the quality control parameters for retaining SNPs and subjects were: SNP missingness < 0.05 (before sample removal), subject missingness < 0.02, autosomal heterozygosity deviation (Fhet < 0.2), SNP missingness < 0.02 (after sample removal), difference in SNP missingness between psychiatric cases and healthy controls < 0.02 and SNP Hardy-Weinberg equilibrium ($P > 10^{-10}$ in psychiatric cases, $P > 10^{-6}$ in healthy controls). Genotype imputation was performed using the prephasing/imputation stepwise approach implemented in Eagle (v2.3.5; https://alkesgroup.broadinstitute.org/Eagle/) [75] and Minimac3 (https://genome.sph.umich.edu/wiki/Minimac3) [76] to the Haplotype Reference Consortium (HRC) reference panel (v1.0) [77]. Relatedness between subjects was calculated using identity by descent and one of each pair of related individuals (pi_hat > 0.2) was excluded. Relatedness with subjects in all other PGC disorder group samples was also calculated and one of each pair of relatives (pi_hat > 0.2) was excluded across all of the samples.

GWAS were performed using PLINK 1.9 by comparing imputed marker dosages under an additive logistic regression model between cases and controls in each of the cohorts separately [78]. Principal components (PCs) generated using EIGENSTRAT were used as covariates in all GWAS as required, to control for population stratification [79]. SNPs were filtered from the GWAS summary statistics from each cohort using sample minor allele frequency (MAF) < 1% and sample MAF corresponding to a minor allele count (MAC) < 10 in cases or controls (whichever had smaller N), in order to control test statistic inflation at low MAFs from small cohorts. Meta-analyses were then performed across cohorts using an inverse variance-weighted fixed effects model in METAL, to obtain results for the GWAS of SI and SA [108].

#### Psychiatric Genomics Consortium Eating Disorders (PGC ED)

Genotyping has been described previously for these cohorts [55,56]. Quality control, principal components analysis to identify and remove ancestry outliers and generate covariates, and imputation to the 1000 Genomes Phase 3 reference panel were performed within PGC's GWAS pipeline RICOPILI [73] as described in full previously [55]. The first 5 PCs were included as covariates and GWASs were performed within RICOPILI using imputed variant dosages and an additive model. Identical individuals between PGC ED cohorts and PGC MDD, BIP and SCZ cohorts were detected using genotype-based checksums (https://personal.broadinstitute.org/sripke/share_links/zpXkV8INxUg9bayDpLToG4g58TMtjN_PGC_SCZ _w3.0718d.76). The PGC ED cohorts were meta-analyzed using an inverse-variance weighted fixed effects model in METAL [108].

#### Psychiatric Genomics Consortium Major Depressive Disorder (PGC MDD)

Genotyping, QC imputation and analyses were conducted in the same manner as described for the PGC BD sample and have been described in full previously [57].

#### Psychiatric Genomics Consortium Post-Traumatic Stress Disorder (PGC PTSD) and Army Study to Assess Risk and Resilience in Servicemembers - Wave 1 Latin American subset (ARMY STARRS)

Genotyping, QC imputation and analyses were conducted in the same manner as described for the other PGC disorder samples and have been described in full previously [96]. As an exception, since there was only one PGC PTSD cohort that had information on SA in Latin American samples, it was analyzed individually and did not undergo meta-analysis. All other GWAS using PGC PTSD samples contained at least two cohorts and were therefore meta-analyzed.

#### Psychiatric Genomics Consortium Schizophrenia (PGC SCZ)

Genotyping, QC imputation and analyses were conducted in the same manner as described for the other PGC disorder samples and have been described in full previously [59].

#### PsyCourse (PSYCR)

Genotyping, QC and imputation procedures in this sample have been described elsewhere [109]. Briefly, individuals were genotyped with the Illumina Infinium Global Screening Array-24 Kit (GSA Array, version 1 and 3; Illumina, San Diego, CA, USA). Single-nucleotide polymorphisms (SNPs) were excluded if they had a missing call rate greater than 2 %, had a Minor Allele Frequency (MAF) <0.5 %, or deviated from Hardy-Weinberg equilibrium with $p < 0.0001$. Individuals were excluded if they had a missing call rate greater than 2 %, if the phenotypic sex of the individual did not match the genotypic sex, if they were duplicated samples according to the pairwise identity by descent or had a large deviation in their heterozygosity value (abs(SD) > 4.17). Additionally, we excluded individuals that had non-European ancestry according to a Multidimensional Scaling (MDS) analysis. Likewise, palindromic SNPs and SNPs with a large MAF deviation (>10 %) with respect to 1000 Genomes European reference populations were also removed. Imputation was performed using the Haplotype Reference Consortium panel [77] in the Michigan Imputation Server [76]. A post-imputation QC was carried out to exclude SNPs that had an imputation quality score of $R2 <0.3$ or had a MAF <1 %.

#### Seoul National University Bundang Hospital PsyGen Cohort Waves 1+2 (SNUBH)

Blood samples were processed into buffy coat and serum fractions and stored at temperatures below -80°C for future genetic and molecular analyses. DNA was genotyped using either the Illumina Asian Screening Array or the Affymetrix Axiom Korea Biobank Array. Stringent QC procedures were applied separately for the SNUBH Wave 1 and Wave 2 cohorts.
For Wave 1, at the variant level, we excluded variants with a missing call rate >5%, a Hardy-Weinberg equilibrium (HWE) P-value <1e-06, or minor allele frequency (MAF) <1%. At the sample level, we excluded one sample from each pair of genetically related individuals (second-degree relatedness; kinship coefficient 0.0884), individuals with missingness >5%, outliers out of three standard deviations from the mean for the first and second principal components of genetic ancestry, or heterozygosity rate beyond three standard deviations from the mean. Prior to imputation, additional quality checks were performed for strand, alleles, position, and reference and alternative allele assignments, as well as allele frequency differences, using tools provided by the McCarthy group (www.well.ox.ac.uk/~wrayner/tools/). Genotype imputation was performed using the 1000 Genomes Project Phase 3 reference panel, with Eagle v2.4.1 for phasing and Minimac4 for imputation. Variants with imputation quality Rsq <0.3 or MAF <1% were removed.
For Wave 2, at the variant level, we excluded duplicate variants and those with a missing call rate >1%, MAF <1%, or HWE P-value <1e-06. At the sample level, we excluded one sample from each pair of genetically related individuals (second-degree relatedness; kinship coefficient 0.0884), samples with missingness >5%, heterozygosity rates beyond five standard deviations from the mean, or sex mismatches. Genotype imputation was performed on the Michigan Imputation Server using the 1aplotype Reference Consortium reference panel, with phasing performed using Eagle v2.4 and imputation using Minimac4. Variants with imputation quality Rsq <0.8 or MAF <1% were removed. The GWAS were performed for both Wave 1 and Wave 2 cohorts using Regenie by adjusting for the top 5 PCs.

#### UK Biobank (UKB)

Data was centrally QC'd by UK Biobank [110]. Additional quality control was performed restricting to individuals in the largest cluster from 4 means clustering of the top two principal components from high quality genome-wide genotypes (these individuals primarily report White British and Irish ethnicities, as well as broader European ethnicities). Individuals recommended for exclusion by UK Biobank due to excess heterozygosity, low genotyping call rate, sex chromosome aneuploidy, and having excessive numbers of relatives in the dataset (N=1056; Resource 531 on http://biobank.ctsu.ox.ac.uk) were excluded. A further 2504 individuals were excluded due to low genotype call rate (<98% call rate), and 466 due to disagreement between NHS recorded or self-reported sex and sex inferred from X chromosome heterozygosity (female Fx >= 0.5, male Fx <= 0.9). A final sample of 105,907 participants was available for analyses – note that this includes related individuals. GWAS analyses were run using REGENIE and covarying for 6 PCs, genotyping batch, and site.

#### University of Utah - European sample (UTAH EUR)

Genotyping, QC, imputation, and analysis procedures for the sample were conducted in the same manner as described for previously published SD GWAS in the Utah cohort [64], except that the GWAS was updated to include additional case and control samples (described above in Cohort ascertainment, case and control definitions).

#### University of Utah - Latin American sample (UTAH LAT)

660 of the 867 suicide cases were genotyped using Illumina Infinium PsychArray platform measuring 593,260 single nucleotide polymorphisms (SNPs), and 207 were genotyped using the Illumina Global Diversity Array + PsychArray content. Control genotyping was performed with the Infinium PsychArray (Illumina, USA) in the high-technology microarray unit of the Instituto Nacional de Medicina Genómica. Control variants were extracted to match the 291,011 available QC'd hard-called variants in the suicide cases. Genotypes were subsequently imputed jointly in all cases and controls (details of imputation are presented in Analytics, below). Cryptic relatedness was modeled via the derivation of genomic relatedness matrices. Genotyping quality control was performed using SNP clustering in Illumina Genome Studio. SNPs were retained if the GenTrain score was > 0.5 and the Cluster separation score was > 0.4. SNPs were converted to HG19 plus strand, and SNPs with >5% missing genotypes were removed. Samples with a call rate < 95% were removed.

GWAS cases were selected based on their estimated genetic ancestry. Genetic ancestry was estimated as a composition of 5 super-populations in the 1000 Genomes reference panel (EUR, AFR, NAT, EAS, and SAS). Case samples were included in the GWAS if the estimated component of AMR ancestral admixtures exceeded 5% and the total of non-EUR and non-AMR ancestries did not exceed 10%. Pre-imputation genotypes were processed using McCarthy Group Tools [https://www.chg.ox.ac.uk/~wrayner/tools/index.html] scripts for HRC Imputation preparation and checking. These scripts check SNPs against the HRC reference panel and exclude 1) SNPs with allele frequencies differing by 0.2 from the reference, and 2) palindromic SNPs with MAF > 0.4. After imputation using the HRC reference panel, SNPs with minor allele frequency below 0.01 or imputation $R^2$ < 0.5 were also excluded. Genomic data were handled using PLINK and PLINK2 [78]. Final GWAS analysis was performed on 6,920,683 variants passing quality control. Analyses were run using a Linear Mixed Model (LMM) algorithm in GEMMA [111] with follow-up examination of significant effects for linkage disequilibrium and gene set enrichment. The first five ancestry PCs and any additional PCs 1-30 that were significantly associated with the outcome were included as covariates in the mixed model to reduce lambda inflation, per RICOPILI methods. Sex was not included as a covariate in GWAS analyses due to the significant association of suicide with sex status in the U.S. at a ratio of approximately 3:1 males to females.

#### Vanderbilt University Medical Center (VUMC)

Standard quality control procedures were applied to the genotype data of BioVU individuals genotyped by the BioVU Infinium expanded multi-ethnic genotyping array (MEGAEX) containing more than two million markers. Ancestry was determined with 1000 Genomes phase 3 (1000GP3) data. A subset of SNPs in linkage disequilibrium was used to generate principal components (PCs) using flashpca version 2.0. Only individuals of European ancestry were included for genetic analyses. SNPs with MAF < 0.005 and Hardy-Weinberg equilibrium test P value < 1 × 10-10 were excluded. SI GWAS was conducted using the SI cases ascertained based on psychiatric forms for suicide assessment, ICD-9/10 codes, and NLP. Firth regression of the binary SI phenotype was performed on SI cases and controls using Regenie v2.2, with age, sex and genetic ancestry-informative principal components 1-20 as covariates (22 covariates) to account for population stratification. Default settings of block size 200 and 20 threads were used. Variants with minor allele frequency < 0.01 were excluded.

#### Yale-Penn

DNA from participants in the Yale-Penn cohort was genotyped using three different Illumina microarrays: Yale-Penn 1 on the HumanOmni1-Quad v1.0 (OMNI), Yale-Penn 2 on the Infinium Human Core Exome (HCE), and Yale-Penn 3 on the Multi-ethnic Global Array (MEGA). Genotype quality control (QC) procedures followed established methods [112]. Specifically, individuals and single nucleotide polymorphisms (SNPs) with call rates below 98% were removed, as were variants with a minor allele frequency (MAF) less than 1%. To assess genetic relatedness, pairwise identity-by-descent (IBD) estimates were calculated in PLINK, and individuals with an IBD > 25% were grouped into the same family. Only a single individual from each family was retained for analysis. If there was discrepancy within families with regard to phenotype, the eldest affected individual was retained while the others were excluded from further analysis. Sex discrepancies were identified by examining X chromosome heterozygosity: self-reported males with heterozygosity > 20% and self-reported females with heterozygosity < 20% were excluded. Genotype imputation was performed separately in African American (AA) and European American (EA) participants using IMPUTE2 and the March 2012 1000 Genomes reference panel (1,000 Genomes Project, 2012; http://www.1000genomes.org/), as implemented on the Michigan Imputation Server (https://imputationserver.sph.umich.edu). GWAS was performed within each stratified cohort and ancestry (6 subsets) using logistic regression in Plink 1.9.

### JUSTIFICATION FOR EXCLUDING INDIVIDUALS WITH PSYCHIATRIC DIAGNOSES AND MISSING SUICIDALITY INFORMATION FROM SUICIDALITY GWAS

#### REMOVAL OF POTENTIALLY MISCLASSIFIED CONTROLS IMPROVES INFORMATIVENESS IN SUICIDALITY GWAS

For controls, the key requirement was confidence that the suicidality phenotype of interest was truly absent. Individuals were retained as controls if they endorsed no prior suicidality, including those who had a psychiatric diagnosis. However, as a safeguard, individuals who were not screened for suicidality were excluded from the control group if they also had a psychiatric diagnosis. Excluding these individuals prevents potential misclassification of suicidality cases as controls, so that genetic signal is not attenuated. Importantly, individuals that were specifically assessed for the suicidality phenotype and reported no history of the suicidality phenotype, but who did have a psychiatric diagnosis, were still retained in the control group, thus this does not imply that cases and controls are drawn from different genetic backgrounds with respect to psychiatric liability. Instead, it reflects differential certainty of phenotype classification.

We also considered the alternative: allowing individuals with unknown SUI status and a known psychiatric diagnosis to remain in the control group. However, we determined that this would introduce greater bias by inflating phenotype misclassification, because the prevalence of suicidality phenotypes in individuals with psychiatric disorders is much higher than the general population average.

In support of this, we calculated the impact of leaving potentially misclassified controls in the control group, versus our approach of removing them, on the ability of our GWAS to detect an association between an allele and case status, using a method established by Hodge et al. (2012)[113]. Briefly, Hodge et al. developed an informativeness measure of a study's ability to identify real differences between cases and controls and examined the measure's behavior when there are no misclassified controls, when there are misclassified controls, and when misclassified controls are removed from the study. These calculations involve allele frequency in cases and controls, case sample size, control:case ratio and misclassification rate of controls. Hodge et al. concluded that removing misclassified controls from the control group is better than leaving them in, even though doing so reduces total sample size, and we confirmed this in our data by performing the same calculations.

As a demonstration, here we use the PGC MDD cohort, the SI phenotype, and the equations from Hodge et al. (2012), to calculate $\chi^2$ values for three scenarios:

(1) allowing potentially misclassified controls to remain in the sample ($\chi^2_{MC}$)
(2) removing potentially misclassified controls ($\chi^2_{R}$), as we have done in the current study
(3) a perfect sample in which all controls were actually screened for SI and are correctly classified ($\chi^2_{CC}$)

First, we assume that in MDD cases the prevalence of suicidal ideation is 56% [114]. Then, using the raw phenotype data for the PGC MDD cohort (before any genetic QC was performed), we identify the following phenotype groups:

| | Endorsed a history of SI | Endorsed no history of SI | Missing SI history | Total |
|---|---|---|---|---|
| MDD case | 8,964 | 8,231 | 1,673 | 18,868 |
| MDD control | 218 | 14,092 | 8,451 | 22,761 |
| Total | 9,182 | 22,323 | 10,124 | 41,629 |

Of those missing SI history, the 8,451 MDD controls were classified as SI controls and then the 1,673 MDD cases were treated according to each of the first two scenarios above:

(1) they were classified as SI controls, resulting in:

| SI cases | SI controls |
|---|---|
| 9,182 | 32,447 |

Or (2) they were removed from the PGC MDD cohort entirely, resulting in:

| SI cases | SI controls |
|---|---|
| 9,182 | 30,774 |

Under scenario (1), assuming that 56% of the 1,673 MDD cases missing SI history experienced SI, approximately 937 (=1673*56%) controls would be misclassified. This represents a misclassification rate of 2.89% (=937/32,447) in the entire control sample in scenario 1. For this example, we use the top SNP from the GWAS of SI in the reduced PGC MDD cohort, rs111471812, which has a C allele frequency of 0.917 in cases and 0.925 in controls, to calculate $\chi^2$ statistics for each scenario. The informativeness measure  is calculated as the $\chi^2$ statistic for that scenario divided by the $\chi^2$ statistic for the perfect sample described in scenario (3). For the PGC MDD SI GWAS, we calculated the following $\chi^2$ statistics and informativeness measures:

| Scenario | Misclassification rate | $\chi^2$ statistic | Informativeness versus scenario (3) |
|---|---|---|---|
| (1) allowing potentially misclassified controls to remain in the sample ($\chi^2_{MC}$) | 2.89% | 6.082 | 94.1% |
| (2) removing potentially misclassified controls ($\chi^2_{R}$), as we have done in the current study | 0% | 6.418 | 99.3% |
| (3) a perfect sample in which all controls were actually screened for SI and are correctly classified ($\chi^2_{CC}$) | 0% | 6.463 | - |

As a result, we see that scenario (1) where misclassified controls are allowed to remain in the sample has the lowest $\chi^2$ statistic and informativeness versus $\chi^2_{CC}$. The informativeness of the SI GWAS in the PGC MDD cohort is reduced to only 99.3% when removing potentially misclassified controls, despite the decrease in sample size (scenario 2), but to 94.1% when allowing potentially misclassified controls to remain.

We performed the same calculations for the SA sample in PGC MDD, and also the SI and SA samples in the PGC BD and PGC SCZ cohorts, the results of which are presented in **Supplementary Note Table 1**. We see that in all cohort and phenotype combinations examined, the study's informativeness (assumed to be 100% in a sample with only correctly classified controls) is reduced less when potentially misclassified controls are removed compared to when they are allowed to remain in the study. While the reduction in informativeness varies, we show that in some cases (i.e., the GWAS of SI in the PGC SCZ cohort), the study's informativeness can be reduced to as much as 80.1% when potentially misclassified controls are allowed to remain, versus 98.6% when they are removed. Thus our approach of removing individuals with psychiatric disorders who were missing the SUI phenotype as a safeguard against potential misclassification amongst controls, can produce major benefits to maintaining informativeness for our GWAS of suicidality phenotypes.

**Supplementary Note Table 1.** Tests for power to detect a SNP association in GWAS of suicidal ideation (SI) and suicide attempt (SA) when potentially misclassified controls are allowed to remain in the sample (MC) versus when potentially misclassified controls are removed from the sample (R). $\chi^2_{MC}$ = $\chi^2$ value when potentially misclassified controls are allowed to remain in the sample; $\chi^2_R$ = $\chi^2$ value when potentially misclassified controls are removed from the sample as we have done in the current study; $\chi^2_{CC}$ = $\chi^2$ value when a perfect sample is used and all controls are correctly classified; K = prevalence of the suicidality phenotype (either SI or SA) in individuals with the disorder the cohort was ascertained for (BD = bipolar disorder, MDD = major depressive disorder, SCZ = schizophrenia); A1= effect allele at the tested SNP; AF = frequency of A1.

| Cohort | GWAS | SNP tested | A1 | AF in cases | AF in controls | K (%) | $\chi^2_{MC}$ (p-value) | $\chi^2_R$ (p-value) | $\chi^2_{CC}$ (p-value) | including misclassified controls ratio (%) | removing misclassified controls ratio (%) |
|---|---|---|---|---|---|---|---|---|---|---|---|
| PGC BD | SI | rs17156675 | C | 0.68 | 0.709 | 54 | 17.955 (2.26e-05) | 19.828 (8.48e-06) | 20.046 (7.56e-06) | 89.57 | 98.91 |
| PGC BD | SA | rs4149 | C | 0.941 | 0.956 | 34 | 19.74 (8.87e-06) | 21.365 (3.80e-06) | 21.461 (3.61e-06) | 91.98 | 99.56 |
| PGC MDD | SI | rs111471812 | C | 0.917 | 0.925 | 56 | 6.082 (0.014) | 6.418 (0.011) | 6.463 (0.011) | 94.11 | 99.30 |
| PGC MDD | SA | rs77802938 | A | 0.938 | 0.959 | 31 | 21.681 (3.22e-06) | 22.422 (2.19e-06) | 22.454 (2.15e-06) | 96.56 | 99.86 |
| PGC SCZ | SI | rs147388136 | G | 0.938 | 0.965 | 35 | 21.211 (4.11e-06) | 26.114 (3.22e-07) | 26.49 (2.65e-07) | 80.07 | 98.58 |
| PGC SCZ | SA | rs140630769 | T | 0.971 | 0.987 | 27 | 29.32 (6.13e-08) | 29.382 (5.94e-08) | 29.387 (5.93e-08) | 99.77 | 99.98 |

#### EVALUATING THE IMPACT OF COHORT-LEVEL EXCLUSION OF INDIVIDUALS WITH PSYCHIATRIC DIAGNOSES AND MISSING SUICIDALITY INFORMATION USING SENSITIVITY META-ANALYSES

Furthermore, the exclusion of individuals with psychiatric disorders who were missing the SUI phenotype was only necessary in six of the 37 cohorts contributing to the SI GWAS (PGC BD, PGC MDD, PGC SCZ, PsyCourse, IBGC CE, and Estonian Biobank), representing 10.6% of the total SI GWAS sample, and ten of the 46 SA and 49 SB cohorts (PGC BD, PGC ED, PGC MDD, PGC SCZ, PsyCourse, IBGC CE, IBGC Spain, GBGC, PREFECT, and Estonian Biobank), representing 14.0% and 13.4% of the total SA and SB GWAS samples, respectively. As such, this impacted only a small portion of the total sample.

Still, we performed a sensitivity analysis in which we have run new GWAS meta-analyses of SI and SB leaving out the cohorts that used this approach (which we refer to as the sensitivity meta-analyses). In **Supplementary Note Table 2**, we show that compared to the primary meta-analysis, the sensitivity meta-analysis showed a slight significant (p=0.003) reduction in liability scale SNP-heritability ($h^2_{SNP}$) for SI, but no significant difference for SB (p=0.238). Additionally, the primary and sensitivity meta-analyses did not show significant differences in their genetic correlations with any external phenotypes (**Supplementary Note Figure 1**), thus including cohorts which used this phenotyping approach is unlikely to have introduced any substantial bias in genetic correlations with other phenotypes.

**Supplementary Note Table 2.** Liability scale SNP-heritability ($h^2_{SNP}$) estimates in the primary and sensitivity meta-analyses of suicidal ideation (SI) and suicidal behavior (SB).

| Phenotype | Meta-analysis | N cohorts | N cases<br>N controls | Sum of $N_{Eff}$ | Liability scale $h^2_{SNP}$ (SE) | $h^2_{SNP}$ P |
|---|---|---|---|---|---|---|
| SI | primary | 37 | 259,747<br>1,309,943 | 840,700 | 0.020 (0.001) | 2.15e-93 |
| SI | sensitivity | 31 | 230,674<br>1,172,164 | 748,358 | 0.017 (0.001) | 3.48e-62 |
| SB | primary | 49 | 75,300<br>1,311,895 | 268,258 | 0.057 (0.003) | 6.29e-99 |
| SB | sensitivity | 39 | 60,118<br>1,140,709 | 215,066 | 0.052 (0.003) | 2.74e-77 |

**Supplementary Note Figure 1. Genetic correlations ($r_g$) between the primary and sensitivity SI/SB meta-analyses and related phenotypes.** The x-axis shows genetic correlation estimates (points) with 95% confidence intervals (error bars). A) shows genetic correlation estimates with the SI primary GWAS (yellow points) and SI sensitivity GWAS (gray points) and B) shows genetic correlation estimates with the SB primary GWAS (green points) and SB sensitivity GWAS (gray points).

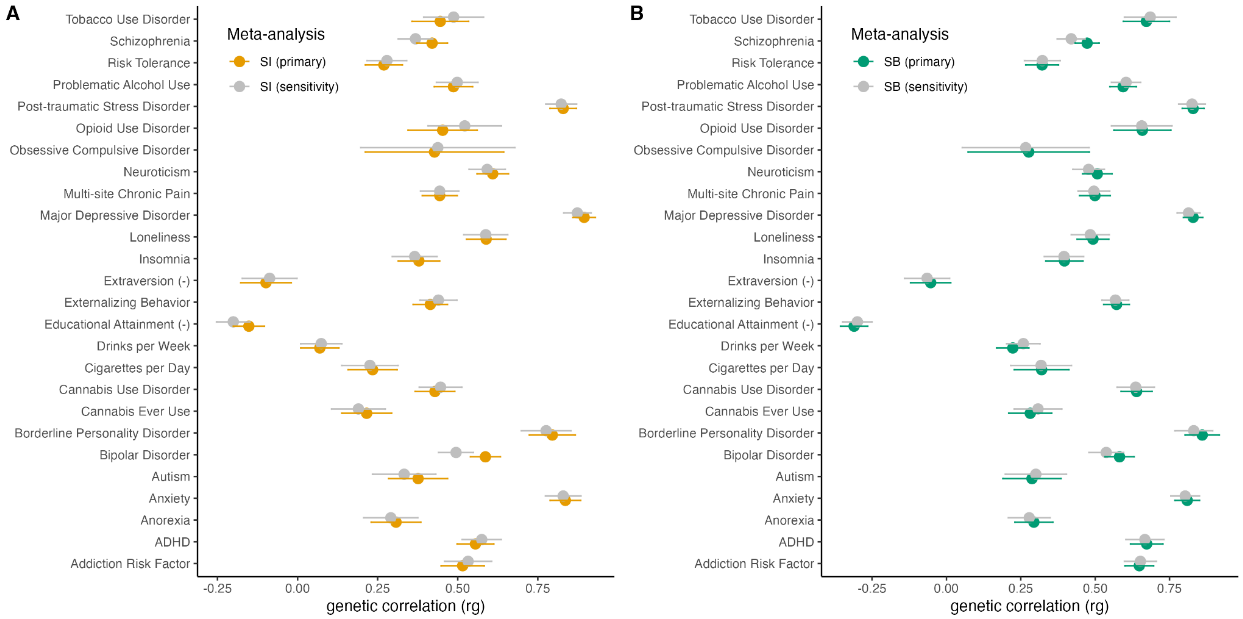

### POST-HOC GWAS META-ANALYSES STRATIFIED BY PHENOTYPING METHOD INDICATE STRONG GENETIC SIMILARITY ACROSS METHODS

#### METHODS

To determine whether phenotypes differed genetically depending on the phenotyping method used, we conducted post-hoc GWAS meta-analyses of SI and SA stratified by phenotyping method. We stratified according to two phenotyping methods for SI (interview and self-report) and three for SA (interview, self-report, and ICD codes) according to the availability of EUR cohorts using a single phenotyping method. We used only EUR samples for this analysis to ensure genetic ancestry would not contribute to any potential heterogeneity. The number of cohorts and sample sizes included in each post-hoc GWAS meta-analysis are presented in Supplementary Note Table 3, below. The resulting GWAS were used to estimate genetic correlations between: (1) interview-based and self-report-based SI, (2) interview-based and self-report-based SA, (3) interview-based and ICD code-based SA, and (4) self-report-based and ICD code-based SA. We then tested whether any genetic correlations were significantly less than 1 after correcting for multiple tests ($p < 0.05/4 = 0.0125$). The post-hoc GWAS meta-analyses and LDSC genetic correlation analyses described here were conducted in the same manner as described in the methods in the main text.

**Supplementary Note Table 3.** Sample size information for the post-hoc GWAS meta-analyses stratified by phenotyping method.

| Phenotype | Phenotyping Method | N cohorts | N cases | N controls |
|---|---|---:|---:|---:|
| SI | interview | 11 | 9,329 | 22,401 |
| SI | self-report | 3 | 12,580 | 31,416 |
| SA | interview | 15 | 6,187 | 31,364 |
| SA | self-report | 4 | 11,734 | 136,436 |
| SA | ICD codes | 2 | 10,122 | 292,138 |

#### RESULTS

Genetic correlations ranged from $r_g = 0.72$ (se = 0.15) between the interview-based and ICD code-based SA meta-analyses and $r_g = 0.86$ (se = 0.11) between the interview-based and ICD code-based SA meta-analyses (**Supplementary Note Table 4**). No genetic correlations were significantly < 1. Thus, our results indicated no significant differences between GWAS using the different phenotyping methods.

**Supplementary Note Table 4.** Genetic correlations between post-hoc GWAS meta-analyses stratified by phenotyping method.

| Phenotype | Method 1 | Method 2 | $r_g$ | Standard Error | P-value for $r_g \neq 0$ | P-value for $r_g < 1$ |
|---|---|---|---|---|---|---|
| SI | interview | self-report | 0.7259 | 0.3390 | 0.0322 | 0.2094 |
| SA | interview | self-report | 0.8559 | 0.1124 | 2.59E-14 | 0.0999 |
| SA | interview | ICD codes | 0.7197 | 0.1525 | 2.38E-06 | 0.0330 |
| SA | self-report | ICD codes | 0.8267 | 0.1064 | 7.82E-15 | 0.0517 |

### FUNDING & ACKNOWLEDGEMENTS

##### General Acknowledgements

Only a brief list of acknowledgements was possible in the main manuscript. The full list of acknowledgements is provided here. We thank the participants who donated their time, life experiences and DNA to this research, and the clinical and scientific teams that worked with them.

##### Cohort Acknowledgements

**ABCD Study®**

Data used in the preparation of this article were obtained from the Adolescent Brain Cognitive Development℠ (ABCD) Study (https://abcdstudy.org), held in the NIMH Data Archive (NDA). This is a multisite, longitudinal study designed to recruit more than 10,000 children age 9-10 and follow them over 10 years into early adulthood. The ABCD Study® is supported by the National Institutes of Health and additional federal partners under award numbers U01DA041048, U01DA050989, U01DA051016, U01DA041022, U01DA051018, U01DA051037, U01DA050987, U01DA041174, U01DA041106, U01DA041117, U01DA041028, U01DA041134, U01DA050988, U01DA051039, U01DA041156, U01DA041025, U01DA041120, U01DA051038, U01DA041148, U01DA041093, U01DA041089, U24DA041123, U24DA041147. A full list of supporters is available at https://abcdstudy.org/federal-partners.html. A listing of participating sites and a complete listing of the study investigators can be found at https://abcdstudy.org/consortium_members/. ABCD consortium investigators designed and implemented the study and/or provided data but did not necessarily participate in the analysis or writing of this report. This manuscript reflects the views of the authors and may not reflect the opinions or views of the NIH or ABCD consortium investigators.The ABCD data repository grows and changes over time. The ABCD data used in this report came from NIMH Data Archive. DOIs can be found at https://dx.doi.org/10.15154/z563-zd24.

**ALSPAC**

We are extremely grateful to all the families who took part in this study, the midwives for their help in recruiting them, and the whole ALSPAC team, which includes data collection staff, data and administrations staff, technical managers and the technical staff with the Bristol Bioresource

Laboratory, based within the University of Bristol. The UK Medical Research Council and Wellcome (Grant ref: MR/Z505924/1) and the University of Bristol provide core support for ALSPAC. This publication is the work of the authors and B. Mars and R. Wootton will serve as guarantors for the contents of this paper. Genomewide genotyping data was generated by Sample Logistics and Genotyping Facilities at Wellcome Sanger Institute and LabCorp (Laboratory Corporation of America) using support from 23andMe. A comprehensive list of grants funding is available on the ALSPAC website (http://www.bristol.ac.uk/alspac/external/documents/grant-acknowledgements.pdf).

**Australian Genetics of Bipolar Disorder Study + Australian Genetics of Depression Study (GBP+AGDS)**

We thank the participants for giving their time and support for this project. We acknowledge and thank M. Steffens for her generous donations and fundraising support for the GBP study. Data collection was funded by the Australian National Health and Medical Research Council (NHMRC) (No. APP1138514, APP1172917 and APP2025674) to S.E.M. S.E.M. is supported by a NHMRC Investigator Grant (No. APP2025674). MER thanks support from the Rebecca L Cooper Medical Research Foundation through an Al & Val Rosenstrauss Fellowship (F20231230). Z.C. is supported by funding from the National Institute of Mental Health (R01MH129356 awarded to Alexis C Edwards). The QSkin Study is supported by a Clinical Trials and Cohort Grant [APP1185416] from the National Health and Medical Research Council of Australia (NHMRC), and genotyping was supported by NHMRC APP1185416. D.C.W. is supported by NHMRC Investigator Grant APP2026567.

**Brazilian High-Risk Cohort for Mental Health Conditions (BHRC)**

The study was funded by Conselho Nacional de Desenvolvimento Científico e Tecnológico (CNPq grant numbers 573974/2008-0 and 465550/2014-2), Fundação de Amparo à Pesquisa do Estado de São Paulo (FAPESP grant numbers: 2008/57896-8, 2013/08531-5, 2014/50917-0, 2020/06172-1, 2021/05332-8, 2021/12901-9, 2023/00437-1, 2023/05560-6,), Coordenação de Aperfeiçoamento de Pessoal de Nível Superior (CAPES: code #001), European Research Council (ERC grant numbers: 337673 and 101057390), UK Medical Research Council (MRC grant number: MR/R022763/1), Ministério da Saúde (Decit/SECTICS/MS Grant number: 888379/2019 - Portaria Nº 1.949, 04/08/2020) and Banco Industrial do Brasil S/A (CISM grant). Collaboration between the BHRC and other cohorts has been funded by the National Institutes of Health (NIMH grant number: R01MH120482-01). Involvement of NIMH Intramural investigators has been funded by NIMH-Intramural Research Program Project MH 002782. Additional support was provided by the National Institute of Developmental Psychiatry for Children and Adolescents (INPD), AWS/Brasil - Healthy & Biological Science Division, and DataRain Consulting – Cloud Computing Services.

**BRIDGE-S**

BRIDGE-S was supported by a research grant of the Deutsche Forschungsgemeinschaft (DFG) awarded to Stephan Ripke (Project Number: 445050869).

**COGA**

The Collaborative Study on the Genetics of Alcoholism (COGA), Principal Investigators B. Porjesz, V. Hesselbrock, A. Agrawal; Scientific Director, A. Agrawal; Translational Director, D. Dick, includes ten different centers: University of Connecticut (V. Hesselbrock); Indiana University (H.J. Edenberg, T. Foroud, Y. Liu, M.H. Plawecki); University of Iowa Carver College of Medicine (S. Kuperman, A. Anderson); SUNY Downstate Health Sciences University (B. Porjesz, J. Meyers); Washington University in St. Louis (L. Bierut, A. Agrawal, S. Hartz); University of California at San Diego (M. Schuckit); Rutgers University (D. Dick, R. Hart, J. Salvatore, J. Tischfield); The Children's Hospital of Philadelphia, University of Pennsylvania (L. Almasy); Icahn School of Medicine at Mount Sinai (A. Goate, P. Slesinger); and Howard University (D. Scott). Other COGA collaborators include: C. Holzhauer, M. Hesselbrock (University of Connecticut); D. Lai, J. Nurnberger Jr., L. Wetherill, X., Xuei, S. O'Connor, (Indiana University); J. Kramer (University of Iowa), G. Chan (University of Iowa; University of Connecticut); C. Kamarajan, A. Pandey, D.B. Chorlian, P. Barr, S. Kinreich, G. Pandey, Z. Neale, S., C. Chatzinakos, J. Zhang, Saenz deViteri, R. Christian, A. Bingly (SUNY Downstate); G. Pathak (Icahn School of Medicine at Mount Sinai); A. Anokhin, K. Bucholz, F. Dong, A. Hatoum, E. Johnson, V. McCutcheon, J. Rice, S. Saccone (Washington University); F. Aliev, Z. Pang, S. Kuo, S. Brislin, J. Moore (Rutgers University); A. Merikangas (The Children's Hospital of Philadelphia and University of Pennsylvania); M. Gitik, NIAAA Staff Collaborator. We continue to be inspired by our memories of Henri Begleiter and Theodore Reich, founding PI and Co-PI of COGA, and also owe a debt of gratitude to other past organizers of COGA, including Ting- Kai Li, P. Michael Conneally, Raymond Crowe, and Wendy Reich, for their critical contributions. This national collaborative study is supported by NIH Grant U10AA008401 from the National Institute on Alcohol Abuse and Alcoholism (NIAAA) and the National Institute on Drug Abuse (NIDA).

**CONVERGE**

This study was funded by the Wellcome Trust (WT090532/Z/09/Z, WT083573/Z/07/Z, WT089269/Z/09/Z) and by NIH grant MH100549. MH129356 provided support for the current analyses. The CONVERGE consortium gratefully acknowledges the support of all partners in hospitals across China. Special thanks to all the CONVERGE collaborators and patients who made this work possible. CONVERGE Consortium: Na Cai, Tim B. Bigdeli, Warren Kretzschmar, Yihan Li, Jieqin Liang, Li Song, Jingchu Hu, Qibin Li, Wei Jin, Zhenfei Hu, Guangbiao Wang, Linmao Wang, Puyi Qian, Yuan Liu, Tao Jiang, Yao Lu, Xiuqing Zhang, Ye Yin, Yingrui Li, Xun Xu, Jingfang Gao, Mark Reimers, Todd Webb, Brien Riley, Silviu Bacanu, Roseann E. Peterson, Yiping Chen, Hui Zhong, Zhengrong Liu, Gang Wang, Jing Sun, Hong Sang, Guoqing Jiang, Xiaoyan Zhou, Yi Li, Yi Li, Wei Zhang, Xueyi Wang, Xiang Fang, Runde Pan, Guodong Miao, Qiwen Zhang, Jian Hu, Fengyu Yu, Bo Du, Wenhua Sang, Keqing Li, Guibing Chen, Min Cai, Lijun Yang, Donglin Yang, Baowei Ha, Xiaohong Hong, Hong Deng, Gongying Li, Kan Li, Yan Song, Shugui Gao, Jinbei Zhang, Zhaoyu Gan, Huaqing Meng, Jiyang Pan, Chengge Gao, Kerang Zhang, Ning Sun, Youhui Li, Qihui Niu, Yutang Zhang, Tieqiao Liu, Chunmei Hu, Zhen Zhang, Luxian Lv, Jicheng Dong, Xiaoping Wang, Ming Tao, Xumei Wang, Jing Xia, Han Rong, Qiang He, Tiebang Liu, Guoping Huang, Qiyi Mei, Zhenming Shen, Ying Liu, Jianhua Shen, Tian Tian, Xiaojuan Liu, Wenyuan Wu, Danhua Gu, Guangyi Fu, Jianguo Shi, Yunchun Chen, Xiangchao Gan, Lanfen Liu, Lina Wang, Fuzhong Yang, Enzhao Cong, Jonathan Marchini,

Huanming Yang, Jian Wang, Shenxun Shi, Richard Mott, Qi Xu, Jun Wang, Kenneth S. Kendler, and Jonathan Flint.

**CVEDA and MGL cohort**
Funding: DBT/Wellcome Trust India Alliance: Intermediate Clinical Fellowship (IA/CPHI/20/1/505266), Scientific Knowledge for Ageing and Neurological Ailments (SKAN) trust: (SKAN/002/208/2021/014), ADBS program funded by the Department of Biotechnology and the Pratiksha Trust (BT/PR17316/MED/31/326/2015), Centre for Brain and Mind grant of the Rohini Nilekani Philanthropies, Department of Science and Technology/INSPIRE/04/2021/003250, MQ grant 2023-MPSIP-54, MQ: Transforming Mental Health fellowship MQF22/20, Newton-Bhabha Grant for the cVEDA study, jointly funded by the Medical Research Council, UK (MR/N000390/1) and the Indian Council of Medical Research (ICMR/MRC-UK/3/M/2015-NCD-I).

**EstBB**
We thank the Estonian Biobank research team for their support and contributions. In particular, we acknowledge Andres Metspalu, Lili Milani, Tõnu Esko, and Mait Metspalu (Estonian Genome Centre, Institute of Genomics, University of Tartu) for their leadership in data collection, genotyping, quality control, and imputation.

This research was supported by the Estonian Research Council (grant nr PSG615) and the Estonian Centre of Excellence for Well-Being Sciences, funded by grant TK218 from the Estonian Ministry of Education and Research. This research was conducted using the Estonian Biobank, which has been supported by the Estonian Center of Genomics/Roadmap II and funded by the Estonian Research Council (project number TT17).
Data analysis was carried out in part in the High-Performance Computing Center of the University of Tartu.

**FinnGen**
We want to acknowledge the participants and investigators of the FinnGen study. The FinnGen project is funded by two grants from Business Finland (HUS 4685/31/2016 and UH 4386/31/2016) and the following industry partners: AbbVie Inc., AstraZeneca UK Ltd, Biogen MA Inc., Bristol Myers Squibb (and Celgene Corporation & Celgene International II Sàrl), Genentech Inc., Merck Sharp & Dohme Corp, Pfizer Inc., GlaxoSmithKline Intellectual Property Development Ltd., Sanofi US Services Inc., Maze Therapeutics Inc., Janssen Biotech Inc, Novartis AG, and Boehringer Ingelheim. Following biobanks are acknowledged for delivering biobank samples to FinnGen: Auria Biobank (www.auria.fi/biopankki), THL Biobank (www.thl.fi/biobank), Helsinki Biobank (www.helsinginbiopankki.fi), Biobank Borealis of Northern Finland (https://www.ppshp.fi/Tutkimus-ja-opetus/Biopankki/Pages/Biobank-Borealis-briefly-in-English.aspx), Finnish Clinical Biobank Tampere (www.tays.fi/en-US/Research_and_development/Finnish_Clinical_Biobank_Tampere), Biobank of Eastern Finland (www.ita-suomenbiopankki.fi/en), Central Finland Biobank (www.ksshp.fi/fi-FI/Potilaalle/Biopankki), Finnish Red Cross Blood Service Biobank

(www.veripalvelu.fi/verenluovutus/biopankkitoiminta), and Terveystalo Biobank (www.terveystalo.com/fi/Yritystietoa/Terveystalo-Biopankki/Biopankki/). All Finnish Biobanks are members of the BBMRI.fi infrastructure (www.bbmri.fi). Finnish Biobank Cooperative – FINBB (https://finbb.fi/) is the coordinator of BBMRI-ERIC operations in Finland. The Finnish biobank data can be accessed through the Fingenious® services (https://site.fingenious.fi/en/) managed by FINBB.

**GEDI/VTSABD**
The VTSABD Study has been supported by the National Institutes of Health grants R01MH045268, R01MH068521, U01DA024413 and R01DA054313.

**MIRECC**
This research was supported by a VA Senior Research Career Scientist Award to Dr. Beckham (#IK6BX003777) from the Clinical Science and Research and Development (CSR&D) Service of Department of Veterans Affairs Office of Research and Development (VA ORD), a VA Research Career Scientist Award to Dr. Kimbrel (#IK6BX006523) from the Biomedical Laboratory Research & Development (BLR&D) Service of VA ORD, and a VA Career Development Award to Dr. Bourassa (#IK2CX002694) from the CSR&D Service of VA ORD. This work was also supported by the VA Mid-Atlantic Mental Illness Research, Education, and Clinical Center (MIRECC). The views expressed in this article are those of the authors and do not necessarily reflect the position or policy of the VA, the U.S. government, Duke University, or any other affiliated institution.

VA Mid-Atlantic MIRECC Workgroup Members: The VA Mid-Atlantic MIRECC Workgroup contributors include: Patrick S. Calhoun, PhD, Eric Dedert, PhD, Eric B. Elbogen, PhD, Robin A. Hurley, MD, Jason D. Kilts, PhD, Angela Kirby, MS, Scott D. McDonald, PhD, Sarah L. Martindale, Ph.D, Christine E. Marx, MD, MS, Scott D. Moore, MD, PhD, Rajendra A. Morey, MD, MS, Jared A. Rowland, PhD, Robert D. Shura, PsyD, Cindy Swinkels, PhD, H. Ryan Wagner, PhD.

**MVP**
This research is based on data from the Million Veteran Program (MVP), Office of Research and Development (ORD), Veterans Health Administration (VA), and was supported by MVP000 as well as VA Merit Award #I01BX005881 from the Biomedical Laboratory Research and Development (BLR&D) Service of VA ORD to Drs. Kimbrel and Beckham. This research was also supported by a VA Research Career Scientist Award to Dr. Kimbrel (#IK6BX006523) from the BLR&D Service of VA ORD and a VA Senior Research Career Scientist Award to Dr. Beckham (#IK6BX003777) from the Clinical Science and Research and Development Service of VA ORD. Dr. Bourassa was supported by a VA Career Development Award (#IK2CX002694) from the CSR&D Service of VA ORD. The views expressed in this article are those of the authors and do not necessarily reflect the position or policy of the VA, the U.S. government, Duke University, or any other affiliated institution.

VA Million Veteran Program (MVP) Core Acknowledgements: MVP Program Office - Sumitra Muralidhar, Ph.D., Program Director US Department of Veterans Affairs, 810 Vermont Avenue NW, Washington, DC 20420 - Jennifer Moser, Ph.D., Associate Director, Scientific Programs US Department of Veterans Affairs, 810 Vermont Avenue NW, Washington, DC 20420 - Jennifer E. Deen, B.S., Associate Director, Cohort & Public Relations US Department of Veterans Affairs, 810 Vermont Avenue NW, Washington, DC 20420 MVP Executive Committee - Co-Chair: Philip S. Tsao, Ph.D. VA Palo Alto Health Care System, 3801 Miranda Avenue, Palo Alto, CA 94304 - Co-Chair: Sumitra Muralidhar, Ph.D. US Department of Veterans Affairs, 810 Vermont Avenue NW, Washington, DC 20420 - J. Michael Gaziano, M.D., M.P.H. VA Boston Healthcare System, 150 S. Huntington Avenue, Boston, MA 02130 - Elizabeth Hauser, Ph.D. Durham VA Medical Center, 508 Fulton Street, Durham, NC 27705 - Amy Kilbourne, Ph.D., M.P.H. VA HSR&D, 2215 Fuller Road, Ann Arbor, MI 48105 - Michael Matheny, M.D., M.S., M.P.H. VA Tennessee Valley Healthcare System, 1310 24th Ave. South, Nashville, TN 37212 - Dave Oslin, M.D. Philadelphia VA Medical Center, 3900 Woodland Avenue, Philadelphia, PA 19104 - Deepak Voora, MD Durham VA Medical Center, 508 Fulton Street, Durham, NC 27705 MVP Co-Principal Investigators - J. Michael Gaziano, M.D., M.P.H. VA Boston Healthcare System, 150 S. Huntington Avenue, Boston, MA 02130 - Philip S. Tsao, Ph.D. VA Palo Alto Health Care System, 3801 Miranda Avenue, Palo Alto, CA 94304 MVP Core Operations - Jessica V. Brewer, M.P.H., Director, MVP Cohort Operations VA Boston Healthcare System, 150 S. Huntington Avenue, Boston, MA 02130 - Mary T. Brophy M.D., M.P.H., Director, VA Central Biorepository VA Boston Healthcare System, 150 S. Huntington Avenue, Boston, MA 02130 - Kelly Cho, M.P.H, Ph.D., Director, MVP Phenomics MVP Core Acknowledgements for Publications_June 2025 VA Boston Healthcare System, 150 S. Huntington Avenue, Boston, MA 02130 - Lori Churby, B.S., Director, MVP Regulatory Affairs VA Palo Alto Health Care System, 3801 Miranda Avenue, Palo Alto, CA 94304 - Scott L. DuVall, Ph.D., Director, VA Informatics and Computing Infrastructure (VINCI) VA Salt Lake City Health Care System, 500 Foothill Drive, Salt Lake City, UT 84148 - Saiju Pyarajan Ph.D., Director, Data and Computational Sciences VA Boston Healthcare System, 150 S. Huntington Avenue, Boston, MA 02130 - Robert Ringer, Pharm.D., Director, VA Albuquerque Central Biorepository New Mexico VA Health Care System, 1501 San Pedro Drive SE, Albuquerque, NM 87108 - Luis E. Selva, Ph.D., Director, MVP Biorepository Coordination VA Boston Healthcare System, 150 S. Huntington Avenue, Boston, MA 02130 - Shahpoor (Alex) Shayan, M.S., Director, MVP PRE Informatics VA Boston Healthcare System, 150 S. Huntington Avenue, Boston, MA 02130 - Brady Stephens, M.S., Principal Investigator, MVP Information Center Canandaigua VA Medical Center, 400 Fort Hill Avenue, Canandaigua, NY 14424 - Stacey B. Whitbourne, Ph.D., Director, MVP Cohort Development and Management VA Boston Healthcare System, 150 S. Huntington Avenue, Boston, MA 0213

**PGCMD_FORMM; FOR2107**

Tilo Kircher receives funding from the German Research Foundation (DFG) FOR 2107, SFB/TRR 393 (“Trajectories of Affective Disorders”, project grant no 521379614), and the Germany’s Excellence Strategy (EXC 3066/1 “The Adaptive Mind”, Project No. 533717223), as well as the DYNAMIC center, funded by the LOEWE program of the Hessian Ministry of Science and Arts (grant number: LOEWE1/16/519/03/09.001(0009)/98). Frederike Stein receives funding from the German Research Foundation (DFG) CRC/TRR 393 (“Trajectories of Affective

Disorders”, project grant no 521379614). Biosamples and corresponding data were sampled, processed and stored in the Marburg Biobank CBBMR.

**International Borderline Genomics Consortium (IBGC)**

We thank all research participants and all researchers and clinicians who collected, generated, or processed the data used in this study. Fabian Streit is supported by a 2023 NARSAD Young Investigator Grant (#31537) from the Brain & Behavior Research Foundation with support from the Families for Borderline Personality Disorder Research. This research was supported by the Hector foundation II and was endorsed by the German Center for Mental Health (DZPG).

**MultiRCT (in IBGC CE)**

Funding/Support: The sites in the Netherlands were supported by ZonMW grant 80-82310-97-12142 from the Netherlands Organization for Health Research and Development (Dr Arntz) and grant 2008 6350 from the Netherlands Foundation for Mental Health (Dr Arntz). The sites in Germany were supported by Else Kröner-Fresenius-Stiftung (Dr Jacob). The sites in Australia were supported by Australian Rotary Health (Dr Lee). The site in Greece was supported by the Greek Society of Schema Therapy, the First Department of Psychiatry of the Medical School of the University of Athens, and the Institut für Verhaltenstherapie Ausbildung Hamburg. The site in London, UK, was supported by the South London and Maudsley NHS Foundation Trust and by Research Center Experimental Psychopathology, Maastricht University. The site in Bradford, UK, was supported by the Bradford District Care NHS Foundation Trust. The site in Basel, Switzerland, was supported by the Research Pool of the Psychiatric University Hospital.

**CANADA - Centre for Addiction and Mental Health, Toronto. FASTER study and DBT vs GPM study cohorts (in IBGC CE)**

The FASTER Dialectical Behaviour Therapy study of borderline personality disorder was funded by the Canadian Institutes of Health Research (CIHR FRN 133428). The Dialectical Behaviour Therapy vs General Psychiatric Management study for borderline personality disorder was supported by Canadian Institutes of Health Research grant 200204MCT-101123. We want to acknowledge and thank all the study investigators including: Guimond, T., Streiner, D. L., Cardish, R. J., & Links, P. S., Gnam, W., Korman, L., (DBT vs GPM study) and Chapman, A. L., (Co-Principal Investigator) Kuo, J. (Co-Principle investigator) R., Dixon-Gordon, K. L., Guimond, T. H., Labrish, C., Isaranuwatchai, W, & Streiner, D. L ( FASTER study). We also wish to acknowledge the support of the Centre for Addiction and Mental Health through the CIHR Postdoctoral Fellowship awarded to A. Lisoway.

**GREAT**

Sample collection, genotyping, and GWAS analysis was supported in part by National Science and Technology Council (NSTC 108-2314-B-002-136-MY3, 110-2314-B-002-067-MY3, NSTC 113-2314-B-002 -168-MY3), the National Taiwan University Career Development Project (109L7860), and the Population Health Research Center from Featured Areas Research Center Program within the framework of the Higher Education Sprout Project by the Ministry of Education in Taiwan (grant number NTU-112L9004).

**Japan**

Sample collection, genotyping, and GWAS of DNA from suicide decedents and non-suicide controls in Japan and the current collaboration was supported, in part, by JSPS KAKENHI (Grant Number 25K19058, 24K10732, 24K10710, 24K02383, 20KK0194, 20H00462, 21H02852, 21H02854, and 21K15712), JST Moonshot Research and Development Program (JPMJMS239F), Japan Agency for Medical Research and Development grants (22dk0307111, 21ek0109555, 21tm0424220, 21ck0106642, 22wm0425008, 23ek0410114, and 23tm0424225), SENSHIN Medical Research Foundation, the Smoking Research Foundation, the Biobank Japan, Takeda Hosho Grants for Research in Medicine, and the Rotary Club of OsakaMidosuji District 2660 Rotary International in Japan.

**The Lundbeck Foundation Initiative for Integrative Psychiatric Research (iPSYCH)**
The iPSYCH team was supported by grants from the Lundbeck Foundation (R102-A9118, R155-2014-1724, and R248-2017-2003), NIH/NIMH (1R01MH124851-01 to Anders Dupont Børglum), and the Universities and University Hospitals of Aarhus and Copenhagen. The Danish National Biobank resource was supported by the Novo Nordisk Foundation. High-performance computer capacity for handling and statistical analysis of iPSYCH data on the GenomeDK HPC facility was provided by the Center for Genomics and Personalized Medicine and the Centre for Integrative Sequencing, iSEQ, Aarhus University, Denmark (grant to Anders Dupont Børglum).

**Predictors For ECT study (PREFECT)**
The authors thank the study participants for their contribution to this research. They also thank the staff at ECT units throughout Sweden and the Swedish National Quality Register for ECT (Q-ECT) for collection and sharing of data. The study was funded by grants from the Swedish Research Council (2018-02653), the Swedish Foundation for Strategic Research (KF10-0039 to Dr. Landén), and the Swedish state under the agreement between the Swedish government and the county councils, the ALF agreement (ALFGBG-716801).

**PGC BD**
**bmrom, rom3, rom4**
The DNA extraction was performed by Dr. Carmen C. Diaconu and Dr. Ana Iulia Neagu at the “Stefan Nicolau” Institute of Virology, Bucharest, Romania. The Romanian samples were funded by UEFISCDI, Bucharest, Romania through several grants to Maria Grigoroiu-Serbanescu.

**neuc1, bmau**
These sample collections were supported by several grants from the Australian National Health and Medical Research Council [grant numbers: 1037196 (PBM, PRS), 1066177 (JMF, JIN), 1063960 (JMF, PRS), 1200428 (JMF, PRS, MJG, CT), 1176716 (PRS), 1177991 (PBM), 1117079 (CSW), 1021970 (CSW), 630471 (MJG), 1081603 (MJG), 1061875 (MJG)] the NSW Ministry of Health (Office of Health and Medical Research). We thank the Lansdowne Foundation, The Aberdeen Fund directors, Janette M. O'Neil and Betty C. Lynch OAM (dec) for their support.

**st2c, stp1**

We thank the late Prof. Pamela Sklar MD PhD from the Icahn School of Medicine at Mount Sinai for the establishment of the Systematic Treatment Enhancement Program for Bipolar Disorder cohorts.

**PGC MDD**

**BiDirect-Study (“Establishing the links between depression and subclinical arteriosclerosis”) (bidi1)**

BiDirect was supported by grants of the German Ministry of Research and Education (BMBF) to the University of Muenster (01ER0816 and 01ER1506) to the University of Münster, Germany.

**FOR2107 (formm)**

Tilo Kircher receives funding from the German Research Foundation (DFG) FOR 2107, SFB/TRR 393 (“Trajectories of Affective Disorders”, project grant no 521379614), and the Germany’s Excellence Strategy (EXC 3066/1 “The Adaptive Mind”, Project No. 533717223), as well as the DYNAMIC center, funded by the LOEWE program of the Hessian Ministry of Science and Arts (grant number: LOEWE1/16/519/03/09.001(0009)/98). Frederike Stein receives funding from the German Research Foundation (DFG) CRC/TRR 393 (“Trajectories of Affective Disorders”, project grant no 521379614). Andreas J. Forstner also receives funding from the German Research Foundation (DFG) CRC/TRR 393 (“Trajectories of Affective Disorders”, project grant no 521379614). Biosamples and corresponding data were sampled, processed and stored in the Marburg Biobank CBBMR.

**Hal2**

Hal2 cohort has been supported by grants from Dalhousie Medical Research Foundation and by Canadian Institutes of Health Research(#166098) to MA.

**SHIP (shp0)**

SHIP is part of the Community Medicine Research net of the University of Greifswald, Germany, which is funded by the Federal Ministry of Education and Research (grants no. 01ZZ9603, 01ZZ0103, and 01ZZ0403), the Ministry of Cultural Affairs and the Social Ministry of the Federal State of Mecklenburg-West Pomerania. Genome-wide SNP typing in SHIP has been supported by a joint grant from Siemens Healthineers, Erlangen, Germany and the Federal State of Mecklenburg-West Pomerania. This study was further supported by the German Research Foundation (GR 1912/5-1).

**PGC PTSD**

**Drakenstein Child Health Study (safr and saf2)**

The Drakenstein Child Health Study (DCHS) was funded by the Bill and Melinda Gates Foundation (OPP1017641 and OPP1017579), the National Institute of Mental Health (1R21MH098662-01), the NIH/H3Africa (1U01AI110466-01A1), the National Research Foundation, the South African Medical Research Council, and the Wellcome Trust (221372/Z/20/Z).

**PGC SCZ**

**HUIPM Reus (celso)**

We are grateful to the study participants and researchers for their time and commitment to this project, which was supported by grant 2021SGR01065 from the Generalitat de Catalunya.

**UK Biobank**
This research has been conducted using the UK Biobank Resource under Application Number 82087 (PI: J Coleman). This work uses data provided by patients and collected by the NHS as part of their care and support. This research was funded by the UK National Institute for Health and Care Research (NIHR) Maudsley Biomedical Research Centre (BRC). The views expressed are those of the authors and not necessarily those of the NIHR or the Department of Health and Social Care.

**PsyCourse (PSYCR)**
Thomas G. Schulze and Peter Falkai were supported by the German Research Foundation (Deutsche Forschungsgemeinschaft [DFG]) within the framework of the projects KFO241 (www.kfo241.de) and PsyCourse (www.PsyCourse.de) (SCHU 1603/4-1, 5-1, 7-1, FA241/16-1 ). Thomas G. Schulze was also supported by the Dr. Lisa Oehler Foundation (Kassel, Germany). The study was endorsed by the Federal Ministry of Education and Research (Bundesministerium für Bildung und Forschung [BMBF]) within the initial phase of the German Center for Mental Health (DZPG) (grant: 01EE2503A, 01EE2503F to PF, TGS).

**UTAH EUR and LAT**
Utah computing infrastructure, analytic pipelines, and genetic data were developed with funding from the National Institute of Mental Health (R01MH123619, K01MH109765, R01MH123489), with funding from Janssen, and with significant additional funding to ARD and AAS from the University of Utah Department of Psychiatry and the Huntsman Mental Health Institute. UTAH cohorts were also supported by the National Institute of Environmental Health Sciences (R01ES032028), by the Utah Department of Health and Human Services, and by the University of Utah. The support and resources from the Center for High Performance Computing at the University of Utah and from the Cellular Translational Research Core of the University of Utah Clinical and Translational Science Institute (UM1TR004409, PI Hess) are also gratefully acknowledged. Sample collection and processing was funded by the National Institute of Mental Health (R01MH123619, R01MH122412, R01MH099134).

#### LIST OF CONSORTIUM MEMBERS FOR BYLINE CONSORTIUM AUTHORSHIP

**CVEDA and MGL cohort collaborators**
Jon Heron 1, Debashish Basu 2, Subodh Bhagyalakshmi Nanjayya 2, Rajkumar Lenin Singh 3, Roshan Lourembam 4, Kalayanaraman Kumaran 5, Murali Krishna 6, Rebecca Kuriyan 7, Sunita Simon Kurpad 8, Kamakshi Kartik 9, Kartik Kalyanram 9, Sylvane Desrivieres 10, Gareth J Barker 11, Dimitri Papadopoulos Orfanos 12, Mireille Toledano 13, Rose Dawn Bharath 14, Pratima Murthy 15, Eesha Sharma 16, Nilakshi Vaidya 17, Amit Chakrabarti 18, Gunter Schumann 17, Jayant Mahadevan 15, Bhagyalakshmi Shankarappa 15, Ashitha Siddappa

Niranjana Murthy 15, Pradip Paul 15, Reeteka Sud 15, Suhas Ganesh 15, Vivek Benegal 15, Sanjeev Jain 15.

1 – Population Health Sciences, Bristol Medical School, University of Bristol, United Kingdom
2 – Department of Psychiatry, Post Graduate Institute of Medical Education and Research, Chandigarh, India
3 – Department of Psychiatry, Regional Institute of Medical Sciences, Imphal, India
4 – Department of Psychology, Regional Institute of Medical Sciences, Imphal, India
5 – MRC Lifecourse Epidemiology Unit, University of Southampton, United Kingdom & Epidemiology Research Unit, CSI Holdsworth Memorial Hospital, Mysuru, India
6 – Institute of Public Health, Banashankari, Bengaluru, India
7 – Division of Nutrition, St John's Research Institute, Bengaluru, India
8 – Department of Psychiatry & Department of Medical Ethics, St. John's Medical College & Hospital, Bengaluru, India
9 – Rishi Valley Rural Health Centre, Madanapalle, Chittoor, India
10 – Centre for Population Neuroscience and Precision Medicine, Institute of Psychology, Psychiatry & Neuroscience, MRC SGDP Centre, King's College London, London, United Kingdom
11 – Department of Neuroimaging, Institute of Psychology, Psychiatry & Neuroscience, King's College London, London, United Kingdom
12 – NeuroSpin, CEA, Universite Paris-Saclay, Paris, France
13 – MRC Centre for Environment and Health, School of Public Health, Imperial College, London, United Kingdom
14 – Department of Neuroimaging and Interventional Radiology, National Institute of Mental Health and Neurosciences, Bengaluru, India
15 – Department of Psychiatry, National Institute of Mental Health and Neurosciences, Bangalore, India
16 – Department of Child and Adolescent Psychiatry, National Institute of Mental Health and Neurosciences, Bangalore, India
17 – Centre for Population Neuroscience and Precision Medicine, Charite Mental Health, Dept. of Psychiatry and Psychotherapy, Charite Universitaetsmedizin Berlin, Germany; Centre for Population Neuroscience and Precision Medicine, Institute for Science and Technology of Brain-Inspired Intelligence, Fudan University, Shanghai, China
18 – Indian Council of Medical Research–Centre for Ageing and Mental Health, Kolkata, India

**Members of the Genoplan Research Team**

Byung-Chul Lee 1, Ji-Woong Kim 1, Young Kee Lee 1, Joon Ho Kang 1, Myeong Jae Cheon 1, Dong Jun Kim 1.

1 – Genoplan RnD Division, Genoplan Korea, Seoul, Republic of Korea

**Members of the International Borderline Genomics Consortium**

Fabian Streit 1, 2, 3, 4, Swapnil Awasthi 5, 6, Alisha SM Hall 7, 8, Alice Braun 5, 6, Maria Niarchou 9, Eirini Marouli 10, Oladapo Babajide 10, Josef Frank 3, Lea Zillich 3, 4, 11, 12,

Carolin M Callies 3, 13, Diana Avetyan 3, Eric Zillich 3, Joonas Naamanka 1, 2, 14, Jean Gonzalez 15, Arvid Harder 16, Yi Lu 16, Zouhair Aherrahrou 17, 18, 19, Zain-Ul-Abideen Ahmad 20, Helga Ask 21, 22, 23, Anthony Batzler 24, Michael E Benros 25, 26, Odette M Brand-de Wilde 27, Søren Brunak 28, Mie T Bruun 29, Lea AN Christoffersen 30, 31, Lucía Colodro-Conde 32, 33, Brandon J Coombes 24, Elizabeth C Corfield 21, 34, Norbert Dahmen 35, Maria Didriksen 36, 37, Khoa M Dinh 37, 38, Srdjan Djurovic 39, 40, Joseph Dowsett 37, Ole Kristian Drange 41, 42, Helene Dukal 3, Susanne Edelmann 43, 44, Christian Erikstrup 7, 38, Mariana K Espinola 3, Eva Fassbinder 45, Annika Faucon 9, Diana S Ferreira de Sá 46, Jerome C Foo 3, 47, 48, 49, Maria Gilles 2, Alfonso Gutiérrez-Zotes 50, 51, Thomas F Hansen 52, 53, Magnus Haraldsson 54, R. Patrick Harper 55, Alexandra Havdahl 21, 23, 34, Urs Heilbronner 56, Stefan Herms 57, 58, Henrik Hjalgrim 26, 59, Christopher Hübel 60, 61, 62, 63, Gitta A Jacob 64, Bitten Aagaard 65, Anders Jorgensen 26, 66, Martin Jungkunz 67, 68, Nikolaus Kleindienst 69, Nora Knoblich 43, Stefanie Koglin 70, Julia Kraft 5, Kristi Krebs 71, Christopher W Lee 72, Yuhao Lin 60, 61, Stefanie Lis 69, 73, Amanda Lisoway 74, 75, Ioannis A Malogiannis 76, Amy Martinsen 77, 78, 79, Tolou Maslahati 70, Katharina Merz 80, Andreas Meyer-Lindenberg 2, Susan Mikkelsen 38, Christina Mikkelsen 37, Arian Mobascher 35, Gerard Muntané 50, 51, 81, Asmundur Oddsson 82, Sisse R Ostrowski 26, 37, Teemu Palviainen 83, Ole BV Pedersen 26, 30, Geir Pedersen 84, 85, Liam Quinn 30, Matthias A Reinhard 80, 86, Florian A Ruths 87, Björn H Schott 88, 89, 90, 91, Michael Schredl 92, Emanuel Schwarz 1, 2, 4, Cornelia E Schwarze 93, Michael Schwinn 37, Tabea Send 2, Engilbert Sigurdsson 54, 94, Katja Simon-Keller 3, Astros T Skuladottir 82, 95, Joaquim Soler 96, 97, 98, Anne Sonley 99, 100, Erik Sørensen 37, Hreinn Stefansson 82, Peter Straub 9, Jaana Suvisaari 101, Martin Tesli 102, 103, Jacob Træholt 37, Henrik Ullum 104, Maja P Völker 3, G Bragi Walters 82, Rujia Wang 60, 61, Christian C Witt 105, 105, Gerhard Zarbock 106, Peter Zill 80, John-Anker Zwart 77, 78, 79, DBDS Genomic Consortium 107, Estonian Biobank Research Team 108, the GLAD Study 109, HUNT All-In Psychiatry 110, Ole A Andreassen 111, 112, 113, Arnoud Arntz 114, Joanna M Biernacka 115, Martin Bohus 69, Gerome Breen 60, 61, Alexander L Chapman 116, 117, Sven Cichon 118, 119, 120, Lea K Davis 9, 121, Michael Deuschle 2, 4, Sebastian Euler 122, Sabine C Herpertz 4, 123, Benjamin Hummelen 84, Andrea Jobst 80, Jaakko Kaprio 83, James L Kennedy 74, 75, 100, Kelli Lehto 71, Klaus Lieb 35, Lourdes Martorell 50, 51, Shelley McMain 100, 124, Richard Musil 80, 125, Vanessa Nieratschker 43, 44, Markus M Nöthen 126, Frank Padberg 80, 86, Aarno Palotie 83, 127, 128, Juan C Pascual 96, 97, 129, Nader Perroud 130, Josep A Ramos-Quiroga 51, 131, 132, 133, Ted Reichborn-Kjennerud 21, 134, Marta Ribases 51, 131, 132, 135, Stefan Roepke 70, 136, Ina Giegling 137, Dan Rujescu 138, Sandra Sanchez-Roige 9, 139, 140, Claudia Schilling 92, Christian Schmahl 4, 69, Kari Stefansson 82, 95, Thorgeir E Thorgeirsson 82, Gustavo Turecki 141, Elisabet Vilella 50, 51, Thomas Werge 26, 31, Bendik S Winsvold 77, 79, 142, Johannes Wrege 143, Marcella Rietschel 3, Stephan Ripke 5, 6, 144, Stephanie H Witt 3, 4

1, Hector Institute for Artificial Intelligence in Psychiatry, Central Institute of Mental Health, Medical Faculty Mannheim, Heidelberg University, Mannheim, Germany
2, Department of Psychiatry and Psychotherapy, Central Institute of Mental Health, Medical Faculty Mannheim, Heidelberg University, Mannheim, Germany

3, Department of Genetic Epidemiology in Psychiatry, Central Institute of Mental Health, Medical Faculty Mannheim, Heidelberg University, Mannheim, Germany
4, German Center for Mental Health (DZPG), Partner Site Mannheim - Heidelberg - Ulm, Germany
5, Department of Psychiatry and Psychotherapy, Charité Universitätsmedizin Berlin, Berlin, Germany
6, Stanley Center for Psychiatric Research, Broad Institute of MIT and Harvard, Cambridge, MA, USA
7, Department of Clinical Medicine, Aarhus University, Aarhus, Denmark
8, Department of Affective Disorders, Aarhus University Hospital - Psychiatry, Aarhus, Denmark
9, Department of Medicine, Division of Genetic Medicine, Vanderbilt University Medical Center, Nashville, TN, USA
10, William Harvey Research Institute, Faculty of Medicine and Dentistry, Queen Mary University London, London, UK
11, Hector Institute for Translational Brain Research, Central Institute of Mental Health, Medical Faculty Mannheim, Heidelberg University, Mannheim, Germany
12, Department of Psychiatry and Psychotherapy, Medical Center - University of Freiburg, Faculty of Medicine, University of Freiburg, Freiburg, Germany
13, Health Psychology, School of Social Sciences, University of Mannheim, Mannheim, Germany
14, SleepWell Research Program, Faculty of Medicine, University of Helsinki, Helsinki, Finland
15, Department of Psychiatry, University of California San Diego, La Jolla, USA
16, Department of Medical Epidemiology and Biostatistics, Karolinska Institute, Stockholm, Sweden
17, Institute for Cardiogenetics, University of Lübeck, Lübeck, Germany
18, DZHK (German Research Centre for Cardiovascular Research), partner site Hamburg/Lübeck/Kiel, Lübeck, Germany
19, University Heart Center Lübeck, Lübeck, Germany
20, Social, Genetic and Developmental Psychiatry Centre, Institute of Psychology, Institute of Psychology, Psychiatry and Neuroscience, King's College London, London, UK
21, PsychGen Centre for Genetic Epidemiology and Mental Health, Norwegian Institute of Public Health, Oslo, Norway
22, Department of Child Health and Development, Norwegian Institute of Public Health, Oslo, Norway
23, PROMENTA Research Center, Department of Psychology, University of Oslo, Oslo, Norway
24, Department of Quantitative Health Sciences, Division of Computational Biology, Mayo Clinic, Rochester, MN, USA
25, Copenhagen Research Centre for Biological and Precision Psychiatry, Mental Health Centre Copenhagen, Copenhagen University Hospital, Copenhagen, Denmark
26, Department of Clinical Medicine, Faculty of Health and Medical Sciences, University of Copenhagen, Copenhagen, Denmark
27, Netherlands Institute for Personality Disorders, PO Box 15933, Halsteren, Netherlands
28, Department of Public Health & Novo Nordisk Foundation Center for Protein Research, Faculty of Health and Medical Sciences, University of Copenhagen, Copenhagen, Denmark

29, Clinical Immunology Research Unit, Department of Clinical Immunology, Odense University Hospital, Odense, Denmark
30, Department of Clinical Immunology, Zealand University Hospital, Køge, Denmark
31, Institute of Biological Psychiatry, Mental Health Services, University of Copenhagen, Copenhagen, Denmark
32, Brain and Mental Health Research Program, QIMR Berghofer, Brisbane, QLD, Australia
33, School of Psychology, The University of Queensland, Brisbane, QLD, Australia
34, Nic Waals Institute, Lovisenberg Diaconal Hospital, Oslo, Norway
35, Department of Psychiatry and Psychotherapy, University Medical Center, University of Mainz, Mainz, Germany
36, Department of Neuroscience, Faculty of Health and Medical Sciences, University of Copenhagen, Copenhagen, Denmark
37, Department of Clinical Immunology, Copenhagen University Hospital, Rigshospitalet, Copenhagen, Denmark
38, Department of Clinical Immunology, Aarhus University Hospital, Aarhus, Denmark
39, Department of Medical Genetics, Oslo University Hospital, Oslo, Norway
40, Department of Clinical Science, University of Bergen, Bergen, Norway
41, NORMENT Centre, Institute of Clinical Medicine, University of Oslo, Oslo, Oslo, Norway
42, Department of psychiatry, Sørlandet hospital, Kristiansand, Agder, Norway
43, Department of Psychiatry and Psychotherapy, University Hospital Tübingen, Tübingen, Germany
44, German Center for Mental Health (DZPG), partner site Tübingen, Tübingen, Germany
45, Department of Psychiatry and Psychotherapy, Christian-Albrechts-Universität zu Kiel, Kiel, Germany
46, Department of Psychology, Division of Clinical Psychology and Psychotherapy, Saarland University, Saarbrücken, Germany
47, Institute for Psychopharmacology, Central Institute of Mental Health, Medical Faculty Mannheim, Heidelberg University, Mannheim, Germany
48, Department of Psychiatry, College of Health Sciences, University of Alberta, Edmonton, AB, Canada
49, Neuroscience and Mental Health Institute, University of Alberta, Edmonton, AB, Canada
50, Hospital Universitari Institut Pere Mata (HUIPM), Institut d'Investigació Sanitària Pere Virgili (IISPV-CERCA), Universitat Rovira i Virgili (URV), Reus, Spain
51, Biomedical Network Research Centre on Mental Health (CIBERSAM), Instituto de Salud Carlos III, Madrid, Spain
52, Neurogenomic, Translational Research Centre, Copenhagen University Hospital, Glostrup, Denmark
53, Danish Headache Center, Copenhagen University Hospital, Rigshospitalet, Copenhagen, Denmark
54, Landspitali University Hospital, Reykjavik, Iceland
55, Bradford District Care NHS Foundation Trust, Bradford, UK
56, Institute of Psychiatric Phenomics and Genomics (IPPG), LMU University Hospital, LMU Munich, Munich, Germany

57, Human Genomics Research Group, Department of Biomedicine, University of Basel, Basel, Switzerland
58, Institute of Human Genetics, University Hospital Bonn & University of Bonn, Bonn, Germany
59, Danish Cancer Institute, Denmark
60, Social, Genetic and Developmental Psychiatry Centre, Institute of Psychology, Psychiatry and Neuroscience, King’s College London, London, UK
61, UK National Institute for Health Research (NIHR) Biomedical Research Centre, South London and Maudsley Hospital and King’s College London, London, UK
62, National Centre for Register-based Research, Aarhus University, Aarhus, Denmark
63, Clinic for Child and Adolescent Psychiatry, German Red Cross Hospitals Westend, Berlin, Germany
64, Institute for Psychology, Department of Clinical Psychology and Psychotherapy, University of Freiburg, University of Freiburg, Germany
65, Department of Clinical Immunology, Aalborg University Hospital, Aalborg, Denmark
66, Psychiatric Center Copenhagen, Denmark
67, Institute for Medical and Data Ethics, Heidelberg University, Faculty of Medicine, Heidelberg, Germany
68, German Cancer Research Center (DKFZ), Heidelberg, Germany
69, Department of Psychosomatic Medicine and Psychotherapy, Central Institute of Mental Health, Medical Faculty Mannheim, Heidelberg University, Mannheim, Germany
70, Department of Psychiatry and Neurosciences, Charité – Universitätsmedizin Berlin, corporate member of Freie Universität Berlin and Humboldt- Universität zu Berlin, Campus Benjamin Franklin, Berlin, Germany
71, Estonian Genome Centre, Institute of Genomics, University of Tartu, Tartu, Estonia
72, Faculty of Health and Medical Sciences, University of Western Australia, Perth, Western Australia, Australia
73, Department of Clinical Psychology, Central Institute of Mental Health, Medical Faculty Mannheim, Heidelberg University, Mannheim, Germany
74, Molecular Brain Science, Campbell Family Mental Health Research Institute, Centre for Addiction and Mental Health, Toronto, Ontario, Canada
75, Institute of Medical Science, University of Toronto, Toronto, Canada
76, First Department of Psychiatry, Eginition Hospital Medical School, National and Kapodistrian University of Athens, Athens, Greece
77, Department of Research and Innovation, Division of Clinical Neuroscience, Oslo University Hospital, Oslo, Norway
78, Institute of Clinical Medicine, Faculty of Medicine, University of Oslo, Oslo, Norway
79, HUNT Center for Molecular and Clinical Epidemiology, Department of Public Health and Nursing, Faculty of Medicine and Health Sciences, Norwegian University of Science and Technology, Trondheim, Norway
80, Department of Psychiatry and Psychotherapy, LMU University Hospital Munich, Ludwig Maximilian University Munich, Munich, Germany
81, Institut de Biologia Evolutiva (UPF-CSIC), Department of Medicine and Life Sciences, Universitat Pompeu Fabra, Parc de Recerca Biomèdica de Barcelona, Barcelona, Spain
82, deCODE genetics / AMGEN, Reykjavik, Iceland

83, Institute for Molecular Medicine Finland (FIMM), HiLIFE, University of Helsinki, Helsinki, Finland
84, Department of Research and Innovation, Division of Mental Health and Addiction, Oslo University Hospital, Oslo, Norway
85, Institute of Basic Medical Sciences, Faculty of Medicine, University of Oslo, Oslo, Norway
86, German Center for Mental Health (DZPG), Partner Site Munich-Augsburg, Germany
87, South London and Maudsley NHS Foundation Trust, London, UK
88, Department of Psychiatry and Psychotherapy, University Medical Center Göttingen, Göttingen, Germany
89, German Center for Neurodegenerative Diseases (DZNE), Göttingen, Germany
90, Leibniz Institute for Neurobiology, Magdeburg, Germany
91, Department of Psychiatry and Psychotherapy, Otto von Guericke University, Magdeburg, Germany
92, Department of Psychiatry and Psychotherapy, Sleep Laboratory, Central Institute of Mental Health, Medical Faculty Mannheim, Heidelberg University, Mannheim, Germany
93, Department of Clinical Psychology and Psychotherapy, Charlotte Fresenius University, Wiesbaden, Germany
94, Faculty of Medicine, Department of Psychiatry, School of Health Sciences, University of Iceland, Reykjavik, Iceland
95, Faculty of Medicine, University of Iceland, Reykjavik, Iceland
96, Centro de Investigación Biomédica en Red de Salud Mental (CIBERSAM), Institut de Recerca Biomèdica Sant Pau (IIB-Sant Pau), Barcelona, Spain
97, Department of Psychiatry, Hospital de la Santa Creu i Sant Pau, Barcelona, Spain
98, Department of Psychiatry and Forensic Medicine & Institute of Neurosciences, Universitat Autònoma de Barcelona, Bellaterra, Spain
99, Borderline Personality Disorder Clinic, Centre for Addiction and Mental Health, Toronto, Ontario, Canada
100, Department of Psychiatry, University of Toronto, Toronto, Canada
101, Finnish Institute for Health and Welfare (THL), Helsinki, Finland
102, Mental health and suicide, Norwegian Institute of Public Health, Oslo, Norway
103, Division of Mental Health and Addiction, Oslo University Hospital, Oslo, Norway
104, Statens Serum Institut, Copenhagen, Denmark
105, Department of Anaesthesiology and Operative Intensive Care, University Hospital Mannheim, Medical Faculty Mannheim/Heidelberg University, Mannheim, Germany
106, IVAH, Institut für Verhaltenstherapie-Ausbildung Hamburg, gemeinnützige GmbH, Hamburg, Germany
107, We thank the members of the DBDS Genomic Consortium are listed in the Supplementary Methods
108, We thank the members of the Estonian Biobank Research Team are listed in the Supplementary Methods, Estonian Genome Centre, Institute of Genomics, University of Tartu, Tartu, Estonia
109, We thank the members the GLAD Study are listed in the Supplementary Methods
110, Members of HUNT All-In Psychiatry are listed in the Supplementary Methods

111, NORMENT Centre, Division of Mental Health and Addiction, Oslo University Hospital, Oslo, Norway
112, KG Jebsen Centre for Neurodevelopmental Disorders, University of Oslo, Oslo, Norway
113, NORMENT Centre, Institute of Clinical Medicine, University of Oslo, Oslo, Norway
114, Department of Clinical Psychology, University of Amsterdam, Amsterdam, Netherlands
115, Department of Quantitative Health Sciences, Department of Psychiatry & Psychology, Division of Computational Biology, Mayo Clinic, Rochester, MN, USA
116, Department of Psychology, Faculty of Arts and Social Sciences, Simon Fraser University, Vancouver, British Columbia, Canada
117, DBT Centre of Vancouver, Vancouver, British Columbia, Canada
118, Department of Biomedicine, University of Basel, Basel, Switzerland
119, Medical Genetics, Institute of Medical Genetics and Pathology, University Hospital Basel, Basel, Switzerland
120, Institute of Neuroscience and Medicine (INM-1), Research Center Juelich, Juelich, Germany
121, Vanderbilt Genetics Institute, Vanderbilt University Medical Center, Nashville, TN, USA
122, Department of Consultation Psychiatry and Psychosomatics, University Hospital Zurich, Zurich, Switzerland
123, Department of General Psychiatry, University Hospital Heidelberg, Heidelberg University, Heidelberg, Germany
124, Borderline Personality Disorder Clinic, General Adult Psychiatry and Health Systems Division, Centre for Addiction and Mental Health, Toronto, Ontario, Canada
125, Oberberg Fachkliniken for Psychiatry, Psychosomatics and Psychotherapy, Bad Tölz, Germany
126, Insitute of Human Genetics, University Hospital Bonn & University of Bonn, Bonn, Germany
127, Analytic and Translational Genetics Unit, Department of Medicine, Department of Neurology and Department of Psychiatry, Massachusetts General Hospital, Boston, MA, USA
128, The Stanley Center for Psychiatric Research and Program in Medical and Population Genetics, The Broad Institute of MIT and Harvard, Cambridge, MA, USA
129, Mental Health and Psychiatry Department, Vic Hospital Consortium, Spain
130, Department of Psychiatric, University Hospitals of Geneva, Geneva, Switzerland
131, Psychiatric Genetics Unit, Group of Psychiatry, Mental Health and Addiction, Vall d'Hebron Research Institute (VHIR), Universitat Autònoma de Barcelona, Barcelona, Spain
132, Department of Psychiatry, Hospital Universitari Vall d'Hebron, Barcelona, Spain
133, Department of Psychiatry and Forensic Medicine, Universitat Autònoma de Barcelona (UAB), Barcelona, Spain
134, Institute of Clinical Medicine, University of Oslo, Oslo, Norway
135, Department of Genetics, Microbiology, and Statistics, Faculty of Biology, Universitat de Barcelona, Barcelona, Spain
136, Department of Psychiatry, Oberberg Fachkliniken for Psychiatry, Psychosomatics and Psychotherapy, Wendisch Rietz, Germany
137, Medical University of Vienna, Department of Neurology, Vienna, Austria
138, Department of Psychiatry, University of Halle, Halle, Germany

139, Institute for Genomic Medicine, University of California San Diego, La Jolla, CA, USA
140, Department of Psychiatry, University of California San Diego, La Jolla, CA, USA
141, Douglas Institute, Department of Psychiatry, McGill University, Montreal, Canada
142, Department of Neurology, Oslo University Hospital, Oslo, Norway
143, Medical Faculty, University of Basel, Basel, Switzerland
144, Analytic and Translational Genetics Unit, Massachusetts General Hospital, Boston, MA, USA
