## Supplementary Data 1 for "Genome-wide association studies identify 77 loci for suicidality and provide novel biological insights"

**Supplementary Data 1A: Forest plots of lead SNPs at the 13 genome-wide significant loci from the multi-ancestry GWAS meta-analysis of suicidal ideation.**
Each box represents the log odds ratio (OR) from an individual contributing cohort, with horizontal lines indicating the 95% confidence interval (CI). The diamond represents the overall meta-analytic estimate across studies.

**rs10736470 G/A 11:113418371**

| Cohort | P | ln(OR) | SE |
|---|---|---|---|
| ABCD3_EUR | 0.2398 | −0.132 | 0.113 |
| ADHEA_AFR | 0.7395 | −0.029 | 0.088 |
| ADHEA_EUR | 0.908 | 0.005 | 0.045 |
| ALSPC_EUR | 0.7712 | −0.011 | 0.039 |
| BEPS7_EUR | 0.2602 | −0.134 | 0.119 |
| BHRCM_AFR | 0.0479 | 0.439 | 0.222 |
| BHRCM_EUR | 0.5045 | −0.116 | 0.174 |
| BOR2C_EUR | 0.8747 | −0.02 | 0.129 |
| CAMHI_EUR | 0.7341 | 0.025 | 0.073 |
| CNVRG_EAS | 0.353 | 0.082 | 0.088 |
| COGA1_EUR | 0.7782 | 0.012 | 0.042 |
| CVEDA_CSA | 0.8183 | 0.024 | 0.106 |
| ESTB2_EUR | 0.2073 | −0.028 | 0.022 |
| GEDIS_EUR | 0.0022 | −0.297 | 0.097 |
| GEDIS_LAT | 0.6236 | 0.107 | 0.218 |
| MVPXQ_AFR | 0.0389 | −0.025 | 0.012 |
| MVPXQ_EAS | 0.9357 | −0.007 | 0.082 |
| MVPXQ_EUR | 1e−04 | −0.025 | 0.007 |
| MVPXQ_LAT | 0.1641 | −0.024 | 0.017 |
| PGCBD_EUR | 0.173 | −0.034 | 0.025 |
| PGCMD_EUR | 0.0679 | −0.038 | 0.02 |
| PGCPT_AFR | 0.4179 | −0.062 | 0.076 |
| PGCPT_EUR | 0.8637 | −0.006 | 0.034 |
| PGCPT_LAT | 0.5439 | 0.062 | 0.102 |
| PGCSZ_EUR | 0.0567 | −0.098 | 0.051 |
| PSYCR_EUR | 0.0521 | 0.16 | 0.082 |
| QIMRB_EUR | 0.0802 | −0.041 | 0.024 |
| SNUBH−ASA_EAS | 0.2735 | −0.222 | 0.203 |
| VUMC1_EUR | 0.4623 | −0.024 | 0.033 |
| YPENN_EUR | 0.859 | −0.013 | 0.073 |
| **meta** | **3.67e−08** | **−0.026** | **0.005** |

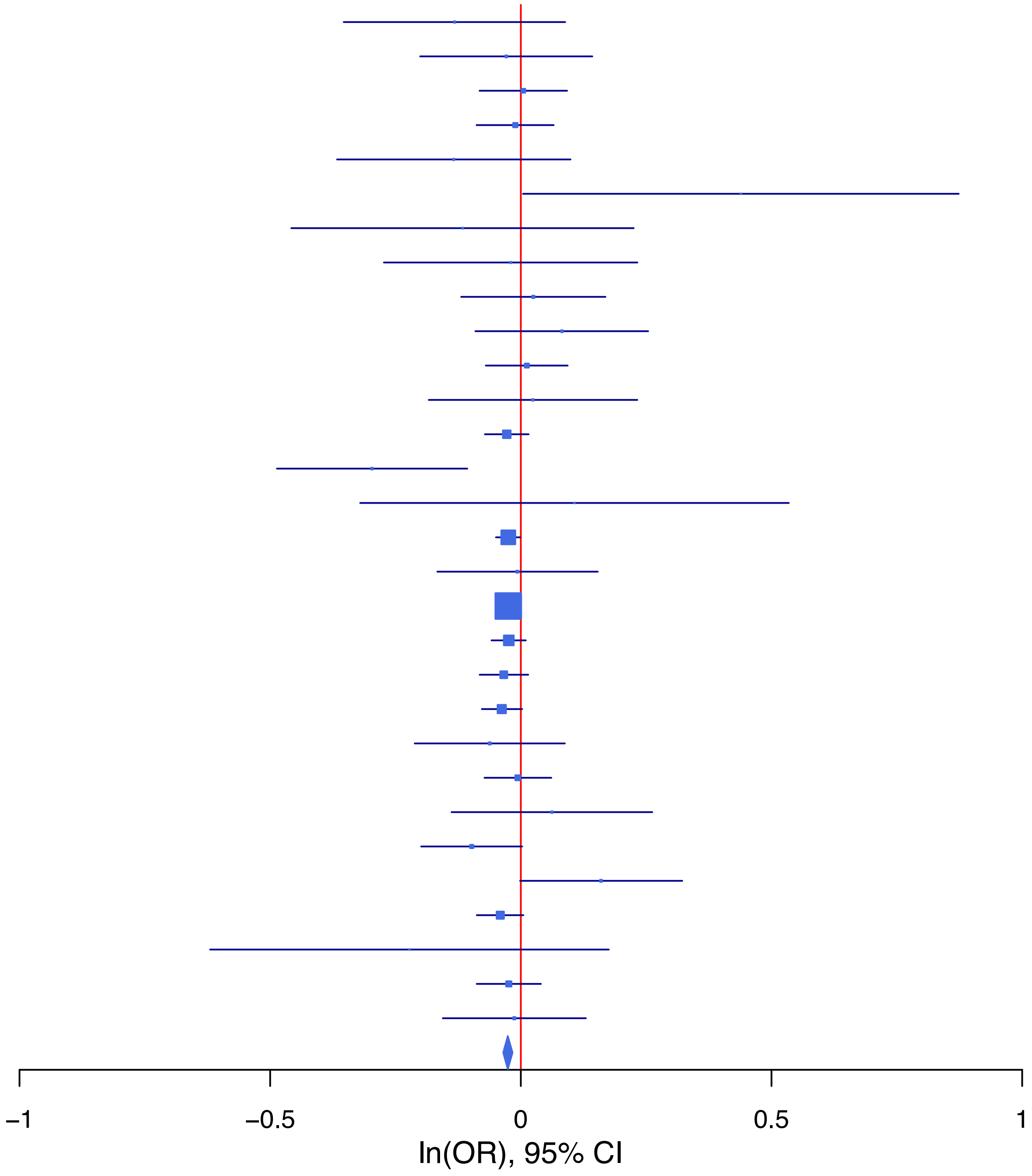

**rs10835373 C/T 11:28643942**

| Cohort | P | ln(OR) | SE |
|---|---|---|---|
| ABCD3_EUR | 0.7389 | 0.035 | 0.107 |
| ALSPC_EUR | 0.8964 | −0.005 | 0.039 |
| BEPS7_EUR | 0.312 | −0.114 | 0.113 |
| BHRCM_AFR | 0.4454 | 0.199 | 0.261 |
| BHRCM_EUR | 0.3629 | 0.152 | 0.167 |
| BHRCM_LAT | 0.2617 | −0.358 | 0.318 |
| BOR2C_EUR | 0.7246 | 0.044 | 0.124 |
| CAMHI_EUR | 0.6005 | 0.038 | 0.072 |
| CNVRG_EAS | 0.0711 | −0.062 | 0.034 |
| COGA1_AFR | 0.4906 | −0.05 | 0.073 |
| COGA1_EUR | 0.504 | −0.027 | 0.041 |
| CVEDA_CSA | 0.76 | 0.032 | 0.106 |
| ESTB2_EUR | 0.6596 | −0.009 | 0.02 |
| GEDIS_EUR | 0.0789 | −0.169 | 0.096 |
| GEDIS_LAT | 0.6207 | −0.101 | 0.204 |
| GREAT_EAS | 0.5268 | −0.056 | 0.089 |
| MIREC_AFR | 0.8365 | 0.02 | 0.097 |
| MIREC_EUR | 0.857 | 0.017 | 0.095 |
| MVPXQ_AFR | 0.0379 | −0.023 | 0.011 |
| MVPXQ_EAS | 0.0134 | −0.116 | 0.047 |
| MVPXQ_EUR | 4.23e−05 | −0.027 | 0.007 |
| MVPXQ_LAT | 0.7905 | −0.004 | 0.016 |
| PGCBD_EUR | 0.2998 | −0.025 | 0.024 |
| PGCMD_EUR | 0.1227 | −0.031 | 0.02 |
| PGCPT_AFR | 0.286 | −0.074 | 0.069 |
| PGCPT_EUR | 0.6085 | −0.017 | 0.034 |
| PGCPT_LAT | 0.1071 | 0.155 | 0.096 |
| PGCSZ_EUR | 0.1713 | −0.068 | 0.05 |
| PSYCR_EUR | 0.6649 | −0.035 | 0.081 |
| QIMRB_EUR | 0.3796 | −0.02 | 0.023 |
| SNUBH−ASA_EAS | 0.2647 | 0.096 | 0.086 |
| SNUBH−KCHIP_EAS | 0.1123 | −0.209 | 0.132 |
| YPENN_EUR | 0.1716 | −0.101 | 0.074 |
| **meta** | **4.70e−08** | **−0.025** | **0.004** |

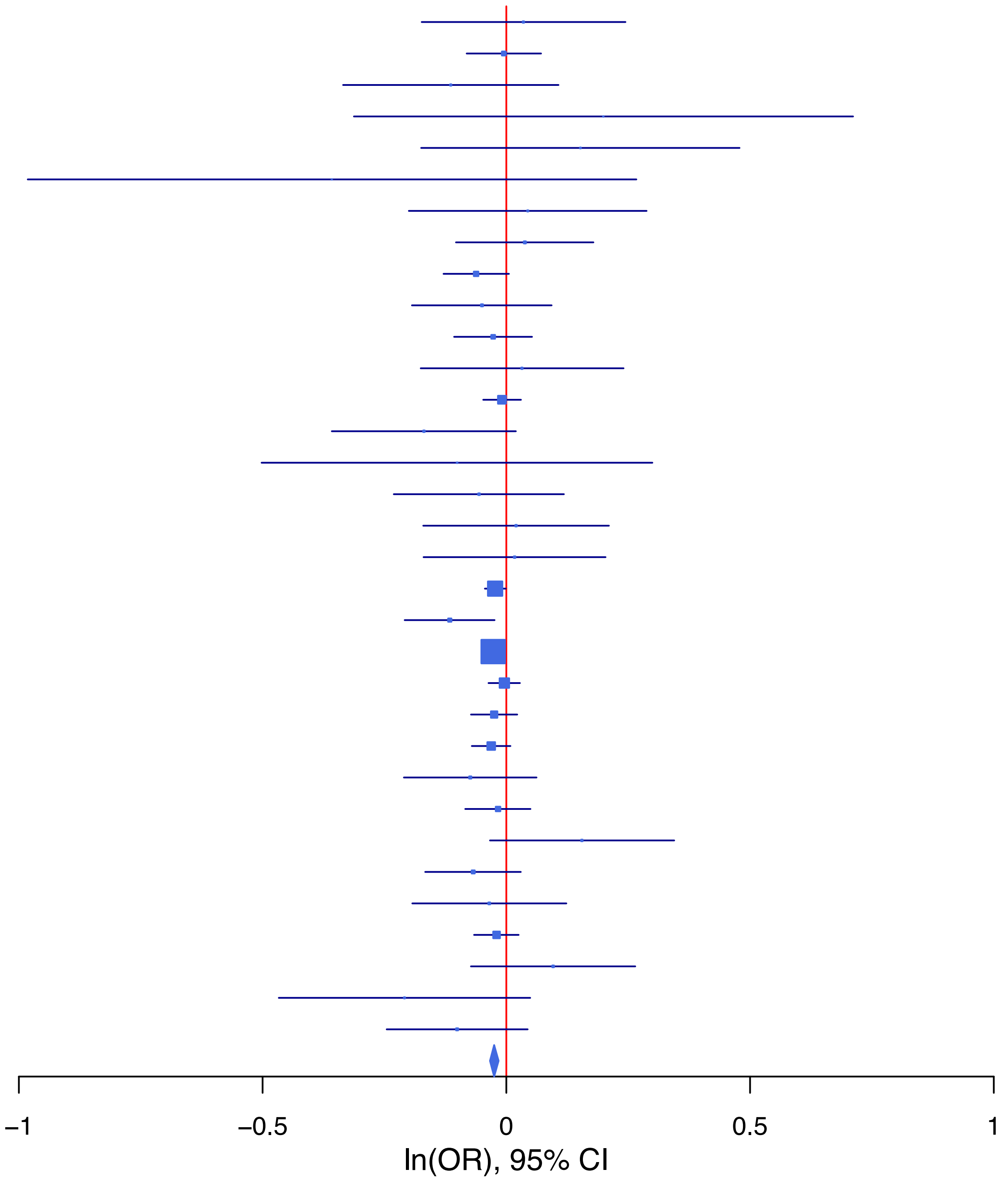

**rs1116313 A/G 11:113296107**

| Cohort | P | ln(OR) | SE |
|---|---|---|---|
| ABCD3_EUR | 0.127 | 0.16 | 0.105 |
| ALSPC_EUR | 0.3824 | 0.033 | 0.038 |
| BEPS7_EUR | 0.0732 | 0.205 | 0.114 |
| BHRCM_AFR | 0.9211 | 0.03 | 0.307 |
| BHRCM_EUR | 0.577 | −0.104 | 0.186 |
| BOR2C_EUR | 0.7845 | −0.035 | 0.127 |
| CAMHI_EUR | 0.5894 | 0.039 | 0.073 |
| CNVRG_EAS | 0.803 | 0.016 | 0.065 |
| COGA1_AFR | 0.9318 | −0.008 | 0.09 |
| COGA1_EUR | 0.869 | −0.007 | 0.04 |
| CVEDA_CSA | 0.4425 | 0.081 | 0.105 |
| ESTB2_EUR | 0.2544 | 0.022 | 0.02 |
| GEDIS_EUR | 0.0033 | 0.268 | 0.091 |
| GEDIS_LAT | 0.2608 | −0.252 | 0.224 |
| GREAT_EAS | 0.1468 | −0.24 | 0.165 |
| MVPXQ_AFR | 0.2516 | 0.015 | 0.013 |
| MVPXQ_EAS | 0.7868 | −0.019 | 0.07 |
| MVPXQ_EUR | 1.64e−05 | 0.028 | 0.006 |
| MVPXQ_LAT | 0.0035 | 0.048 | 0.016 |
| PGCBD_EUR | 0.4571 | 0.018 | 0.024 |
| PGCMD_EUR | 0.0839 | 0.035 | 0.02 |
| PGCPT_AFR | 0.5356 | −0.053 | 0.086 |
| PGCPT_EUR | 0.0506 | −0.065 | 0.033 |
| PGCPT_LAT | 0.924 | 0.009 | 0.098 |
| PGCSZ_EUR | 0.3141 | 0.049 | 0.049 |
| PSYCR_EUR | 0.0475 | −0.155 | 0.078 |
| QIMRB_EUR | 0.2685 | 0.025 | 0.023 |
| SNUBH−ASA_EAS | 0.4752 | 0.114 | 0.16 |
| SNUBH−KCHIP_EAS | 0.2277 | 0.302 | 0.251 |
| VUMC1_EUR | 0.4332 | 0.025 | 0.032 |
| YPENN_AFR | 0.7221 | −0.042 | 0.119 |
| YPENN_EUR | 0.7039 | 0.027 | 0.072 |
| **meta** | **4.33e−08** | **0.025** | **0.004** |

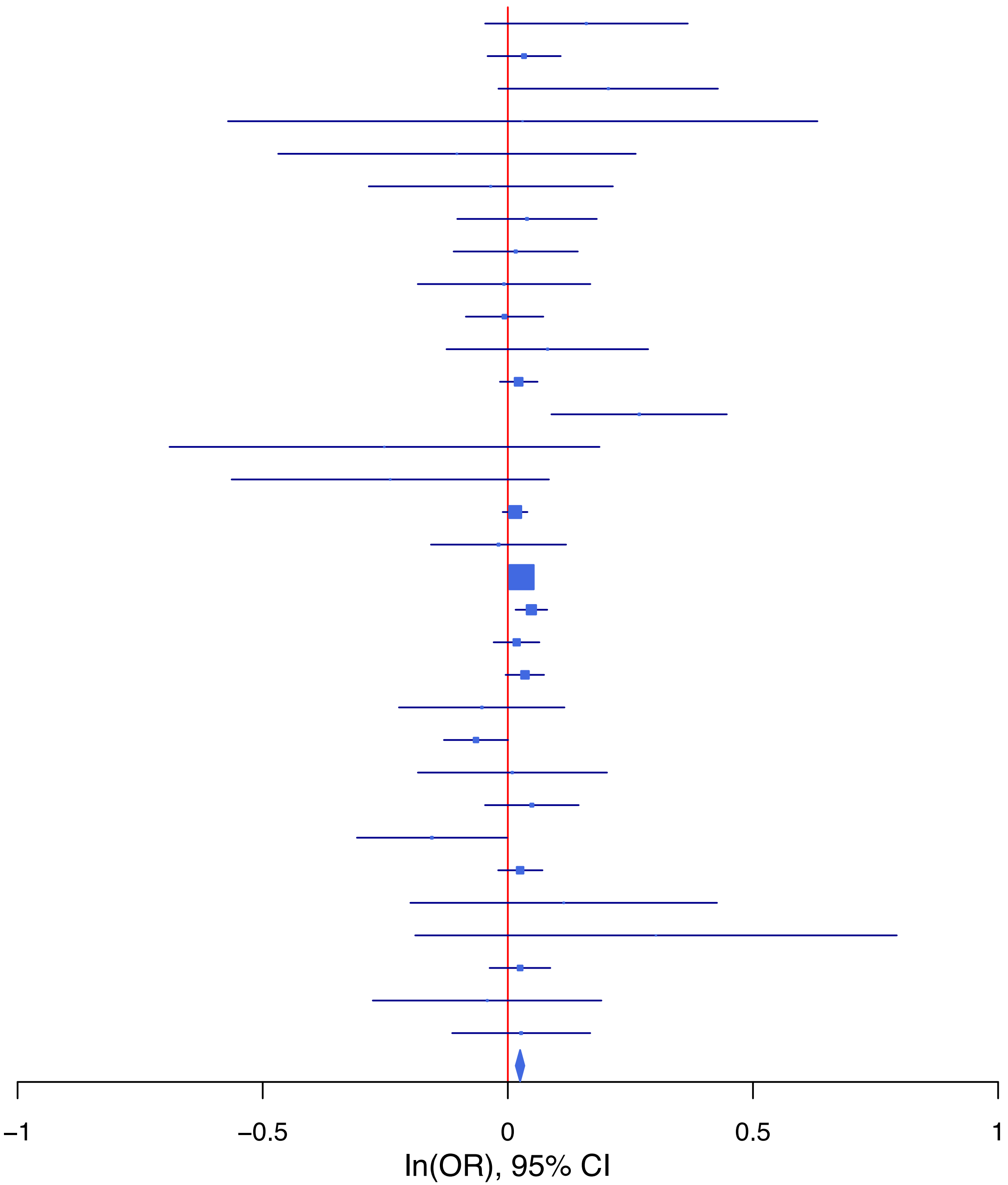

**rs114164545 A/G 3:61820545**

| Cohort | P | ln(OR) | SE |
|---|---|---|---|
| ABCD3_EUR | 0.8821 | 0.046 | 0.311 |
| ALSPC_EUR | 0.5157 | −0.08 | 0.123 |
| BEPS7_EUR | 0.1649 | −0.508 | 0.366 |
| BOR2C_EUR | 0.4389 | −0.271 | 0.35 |
| CAMHI_EUR | 0.7638 | 0.064 | 0.211 |
| COGA1_EUR | 0.8778 | 0.019 | 0.123 |
| ESTB2_EUR | 0.1961 | −0.058 | 0.045 |
| GEDIS_EUR | 0.6374 | −0.133 | 0.283 |
| MVPXQ_EUR | 1.44e−06 | −0.096 | 0.02 |
| MVPXQ_LAT | 0.7767 | −0.019 | 0.066 |
| PGCBD_EUR | 0.7732 | −0.026 | 0.09 |
| PGCMD_EUR | 0.0204 | −0.16 | 0.069 |
| PGCPT_EUR | 0.1707 | −0.144 | 0.105 |
| PGCSZ_EUR | 0.8274 | 0.042 | 0.193 |
| PSYCR_EUR | 0.964 | −0.01 | 0.228 |
| QIMRB_EUR | 0.0337 | −0.156 | 0.073 |
| VUMC1_EUR | 0.6236 | −0.048 | 0.098 |
| YPENN_EUR | 0.5709 | −0.133 | 0.234 |
| **meta** | **1.19e−08** | **−0.088** | **0.015** |

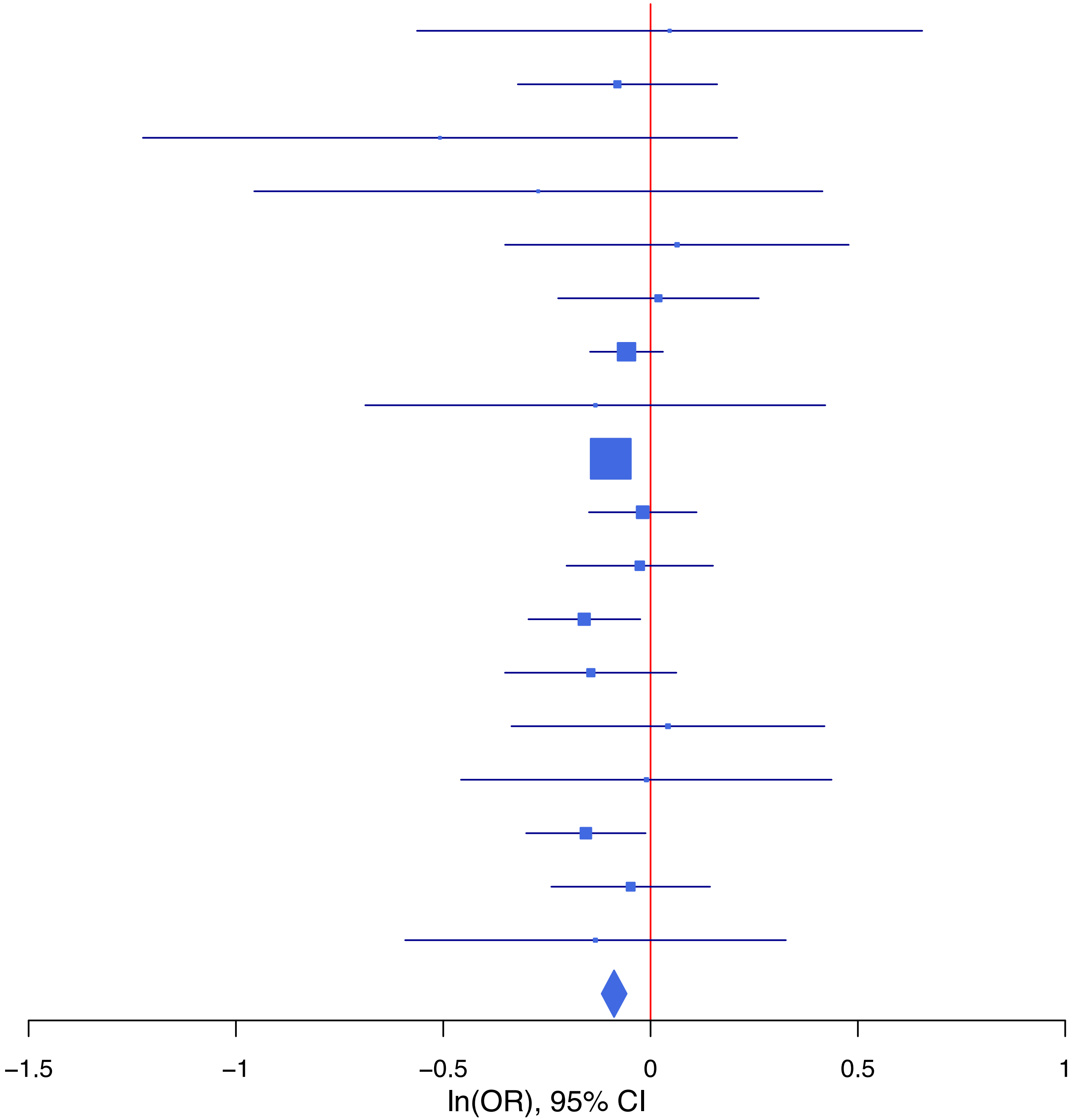

**rs1147600 C/T 10:98981643**

| Cohort | P | ln(OR) | SE |
|---|---|---|---|
| ABCD3_EUR | 0.5137 | 0.071 | 0.109 |
| ADHEA_AFR | 0.0766 | 0.15 | 0.085 |
| ADHEA_EUR | 0.4261 | 0.036 | 0.046 |
| ALSPC_EUR | 0.619 | 0.02 | 0.04 |
| BEPS7_EUR | 0.7846 | 0.033 | 0.122 |
| BHRCM_AFR | 0.2919 | 0.225 | 0.214 |
| BHRCM_EUR | 0.3388 | −0.195 | 0.204 |
| BHRCM_LAT | 0.0437 | 0.876 | 0.434 |
| BOR2C_EUR | 0.1091 | 0.211 | 0.132 |
| CAMHI_EUR | 0.5293 | −0.047 | 0.075 |
| CNVRG_EAS | 0.4874 | 0.031 | 0.044 |
| COGA1_AFR | 0.2528 | 0.094 | 0.082 |
| COGA1_EUR | 0.9868 | −0.001 | 0.042 |
| CVEDA_CSA | 0.5496 | 0.066 | 0.11 |
| ESTB2_EUR | 0.2131 | 0.025 | 0.02 |
| GEDIS_EUR | 0.2013 | −0.124 | 0.097 |
| GEDIS_LAT | 0.6014 | 0.108 | 0.207 |
| GREAT_EAS | 0.1232 | 0.142 | 0.092 |
| MIREC_AFR | 0.6894 | 0.041 | 0.104 |
| MIREC_EUR | 0.5073 | 0.065 | 0.098 |
| MVPXQ_AFR | 0.0129 | 0.029 | 0.012 |
| MVPXQ_EAS | 0.6831 | −0.024 | 0.058 |
| MVPXQ_EUR | 0.0079 | 0.018 | 0.007 |
| MVPXQ_LAT | 0.359 | 0.017 | 0.018 |
| PGCBD_EUR | 0.5701 | 0.014 | 0.025 |
| PGCMD_EUR | 0.0071 | 0.056 | 0.021 |
| PGCPT_AFR | 0.7134 | −0.028 | 0.075 |
| PGCPT_EUR | 0.5317 | 0.022 | 0.035 |
| PGCPT_LAT | 0.0806 | 0.183 | 0.104 |
| PGCSZ_EUR | 0.5395 | 0.032 | 0.051 |
| PSYCR_EUR | 0.2553 | 0.093 | 0.081 |
| QIMRB_EUR | 0.1494 | 0.035 | 0.024 |
| SNUBH−ASA_EAS | 0.6495 | 0.057 | 0.125 |
| SNUBH−KCHIP_EAS | 0.7516 | −0.059 | 0.185 |
| VUMC1_EUR | 0.028 | 0.073 | 0.033 |
| YPENN_AFR | 0.8345 | −0.024 | 0.115 |
| YPENN_EUR | 0.1655 | 0.104 | 0.075 |
| **meta** | **4.71e−08** | **0.025** | **0.005** |

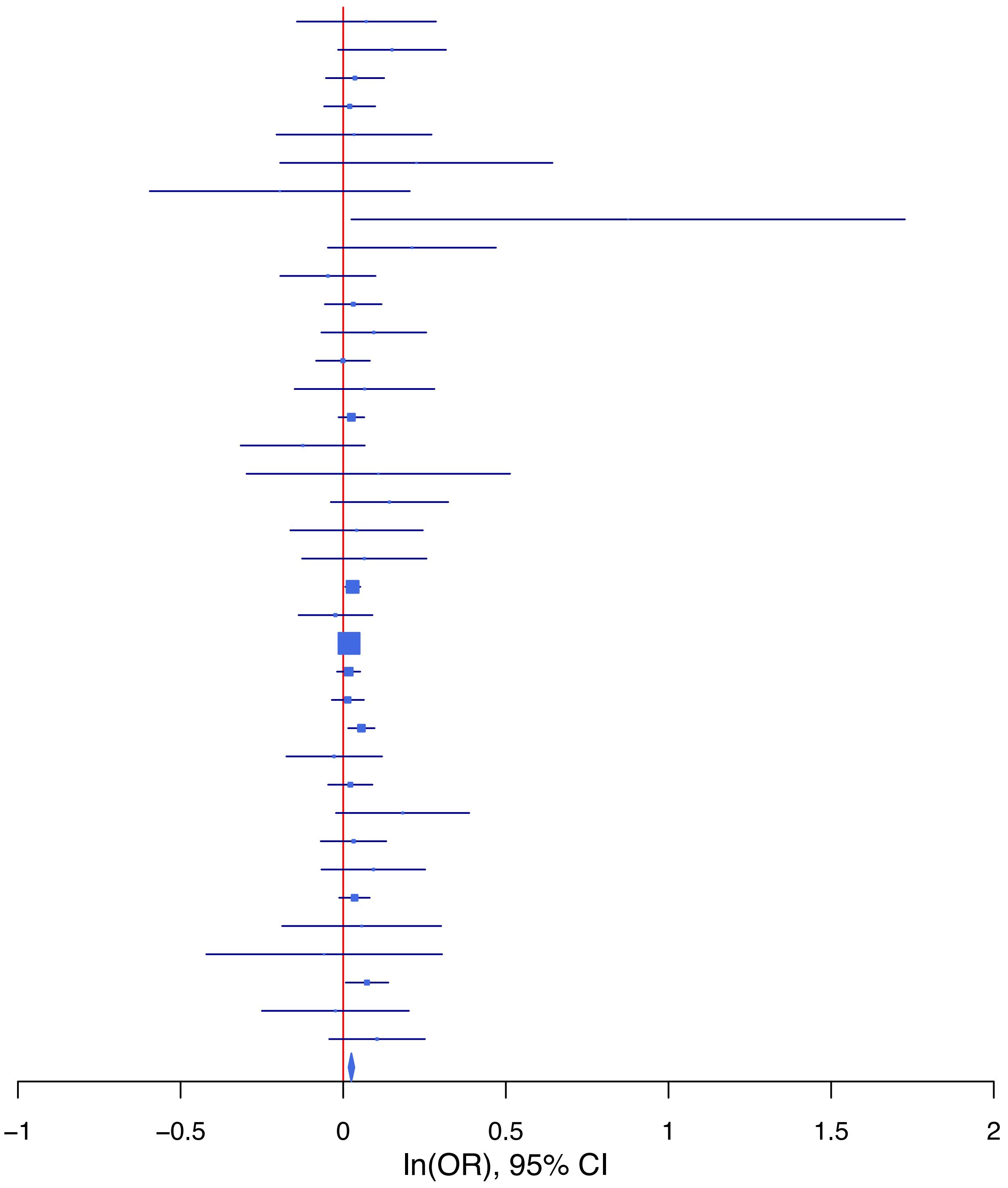

**rs11533158 C/T 9:140262426**

| Cohort | P | ln(OR) | SE |
|---|---|---|---|
| BEPS7_EUR | 0.0393 | 0.273 | 0.133 |
| BHRCM_AFR | 0.173 | 0.292 | 0.214 |
| BHRCM_EUR | 0.0381 | 0.36 | 0.174 |
| BHRCM_LAT | 0.3223 | 0.27 | 0.273 |
| BOR2C_EUR | 0.8306 | 0.031 | 0.146 |
| CAMHI_EUR | 0.8005 | −0.019 | 0.077 |
| CNVRG_EAS | 0.6338 | 0.02 | 0.041 |
| COGA1_AFR | 0.2079 | 0.091 | 0.072 |
| COGA1_EUR | 0.8553 | −0.008 | 0.043 |
| CVEDA_CSA | 0.7531 | 0.038 | 0.12 |
| ESTB2_EUR | 0.0276 | 0.048 | 0.022 |
| GREAT_EAS | 0.6352 | 0.051 | 0.107 |
| MVPXQ_AFR | 0.1973 | 0.013 | 0.01 |
| MVPXQ_EAS | 0.5641 | 0.031 | 0.054 |
| MVPXQ_EUR | 5.99e−08 | 0.036 | 0.007 |
| MVPXQ_LAT | 0.2733 | 0.019 | 0.017 |
| PGCBD_EUR | 0.5116 | −0.024 | 0.037 |
| PGCMD_EUR | 0.1656 | 0.038 | 0.027 |
| PGCPT_AFR | 0.7653 | 0.021 | 0.069 |
| PGCPT_EUR | 0.1184 | 0.054 | 0.034 |
| PGCPT_LAT | 0.5173 | 0.064 | 0.098 |
| PGCSZ_EUR | 0.5569 | 0.041 | 0.07 |
| PSYCR_EUR | 0.5665 | 0.052 | 0.09 |
| QIMRB_EUR | 0.0052 | 0.073 | 0.026 |
| SNUBH−ASA_EAS | 0.2717 | −0.105 | 0.096 |
| SNUBH−KCHIP_EAS | 0.6084 | 0.079 | 0.154 |
| **meta** | **6.20e−11** | **0.031** | **0.005** |

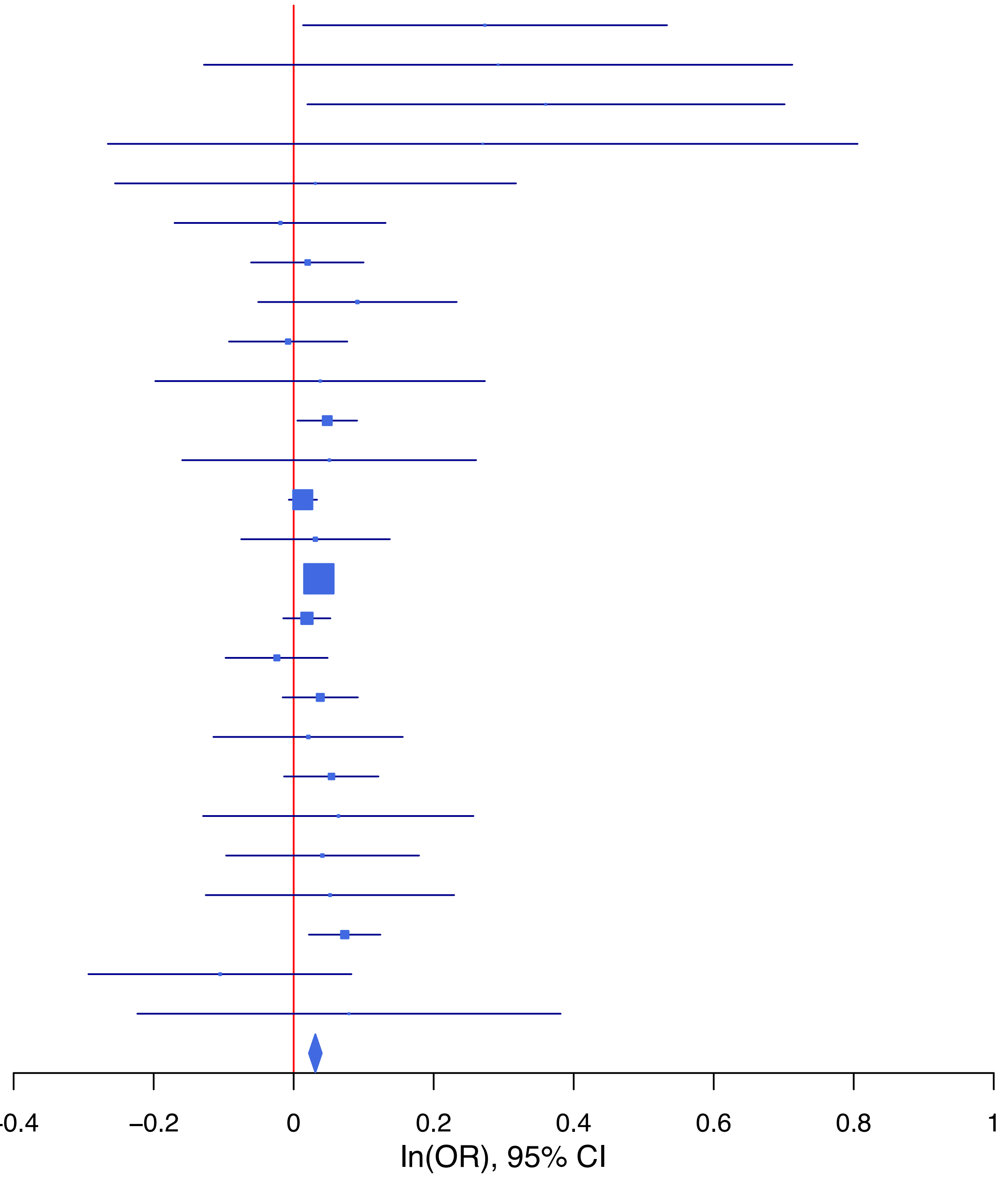

**rs17514846 A/C 15:91416550**

| Cohort | P | ln(OR) | SE |
|---|---|---|---|
| ABCD3_EUR | 0.8171 | 0.025 | 0.105 |
| ALSPC_EUR | 0.2034 | −0.049 | 0.039 |
| BEPS7_EUR | 0.7626 | 0.033 | 0.109 |
| BHRCM_AFR | 0.2542 | 0.501 | 0.439 |
| BHRCM_EUR | 0.1561 | −0.266 | 0.187 |
| BOR2C_EUR | 0.4059 | −0.104 | 0.125 |
| CAMHI_EUR | 0.1985 | −0.088 | 0.069 |
| CNVRG_EAS | 0.2134 | −0.054 | 0.043 |
| COGA1_AFR | 0.5172 | 0.057 | 0.088 |
| COGA1_EUR | 0.1295 | −0.06 | 0.04 |
| CVEDA_CSA | 0.7 | −0.039 | 0.102 |
| GEDIS_EUR | 0.3009 | −0.094 | 0.091 |
| GEDIS_LAT | 0.282 | −0.256 | 0.238 |
| GREAT_EAS | 0.4196 | 0.087 | 0.108 |
| MVPXQ_AFR | 0.6586 | −0.006 | 0.014 |
| MVPXQ_EAS | 0.6139 | −0.027 | 0.053 |
| MVPXQ_EUR | 1e−04 | −0.025 | 0.007 |
| MVPXQ_LAT | 0.082 | −0.03 | 0.017 |
| PGCBD_EUR | 1e−04 | −0.098 | 0.025 |
| PGCMD_EUR | 0.0574 | −0.038 | 0.02 |
| PGCPT_AFR | 0.4569 | −0.065 | 0.088 |
| PGCPT_EUR | 0.539 | −0.02 | 0.033 |
| PGCPT_LAT | 0.7318 | 0.034 | 0.098 |
| PGCSZ_EUR | 0.487 | 0.035 | 0.051 |
| PSYCR_EUR | 0.9564 | −0.004 | 0.077 |
| QIMRB_EUR | 4e−04 | −0.081 | 0.023 |
| SNUBH−ASA_EAS | 0.3518 | −0.103 | 0.11 |
| SNUBH−KCHIP_EAS | 0.6146 | 0.087 | 0.172 |
| VUMC1_EUR | 0.0659 | −0.058 | 0.031 |
| YPENN_AFR | 0.3491 | −0.111 | 0.118 |
| YPENN_EUR | 0.0313 | −0.154 | 0.071 |
| **meta** | **5.59e−11** | **−0.031** | **0.005** |

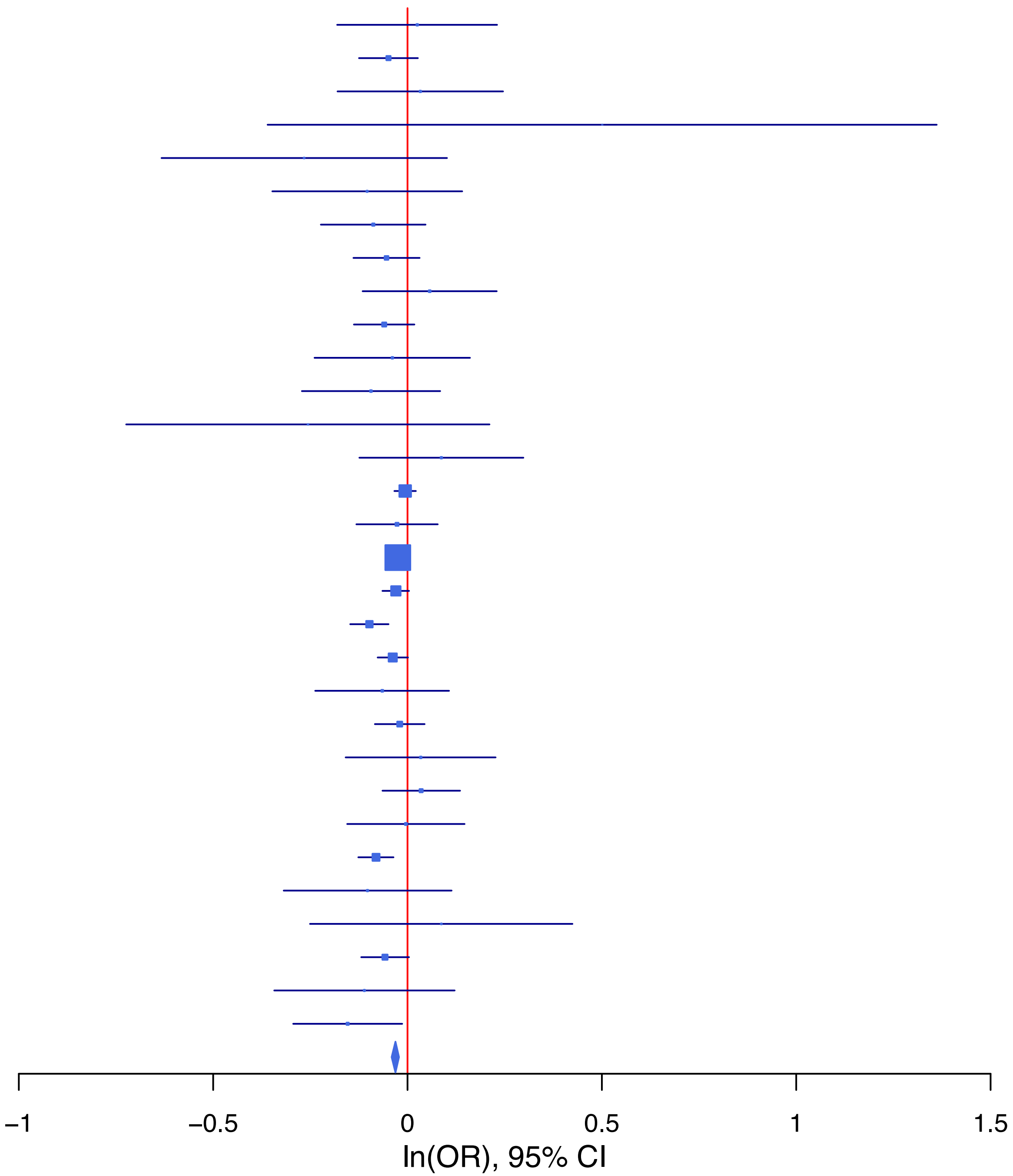

**rs1836798 G/A 11:112931625**

| Cohort | P | ln(OR) | SE |
|---|---|---|---|
| ABCD3_EUR | 0.8175 | −0.03 | 0.13 |
| ADHEA_AFR | 0.5403 | −0.05 | 0.081 |
| ADHEA_EUR | 0.3338 | 0.05 | 0.051 |
| ALSPC_EUR | 0.0278 | −0.103 | 0.047 |
| BEPS7_EUR | 0.1364 | 0.217 | 0.146 |
| BHRCM_AFR | 0.6544 | −0.094 | 0.211 |
| BHRCM_EUR | 0.6784 | 0.091 | 0.218 |
| BHRCM_LAT | 0.0471 | 0.659 | 0.332 |
| BOR2C_EUR | 0.3482 | 0.143 | 0.153 |
| CAMHI_EUR | 0.6818 | −0.036 | 0.088 |
| CNVRG_EAS | 0.1924 | −0.04 | 0.031 |
| COGA1_AFR | 0.3572 | 0.071 | 0.077 |
| COGA1_EUR | 0.1699 | 0.066 | 0.048 |
| CVEDA_CSA | 0.3198 | 0.115 | 0.116 |
| ESTB2_EUR | 0.0092 | 0.054 | 0.021 |
| GEDIS_LAT | 0.7239 | 0.081 | 0.229 |
| GREAT_EAS | 0.7139 | 0.023 | 0.063 |
| MVPXQ_AFR | 0.007 | 0.03 | 0.011 |
| MVPXQ_EUR | 2e−04 | 0.03 | 0.008 |
| MVPXQ_LAT | 0.0207 | 0.039 | 0.017 |
| PGCBD_EUR | 0.0564 | 0.055 | 0.029 |
| PGCMD_EUR | 0.0117 | 0.061 | 0.024 |
| PGCPT_EUR | 0.3631 | 0.036 | 0.04 |
| PGCPT_LAT | 0.4168 | −0.08 | 0.099 |
| PGCSZ_EUR | 0.6826 | 0.024 | 0.058 |
| PSYCR_EUR | 0.7842 | −0.027 | 0.097 |
| QIMRB_EUR | 0.6178 | −0.014 | 0.028 |
| SNUBH−ASA_EAS | 0.9774 | 0.002 | 0.077 |
| SNUBH−KCHIP_EAS | 0.4686 | 0.089 | 0.122 |
| VUMC1_EUR | 0.8678 | 0.006 | 0.038 |
| YPENN_AFR | 0.0287 | −0.228 | 0.104 |
| YPENN_EUR | 0.1226 | 0.13 | 0.084 |
| **meta** | **6.06e−09** | **0.029** | **0.005** |

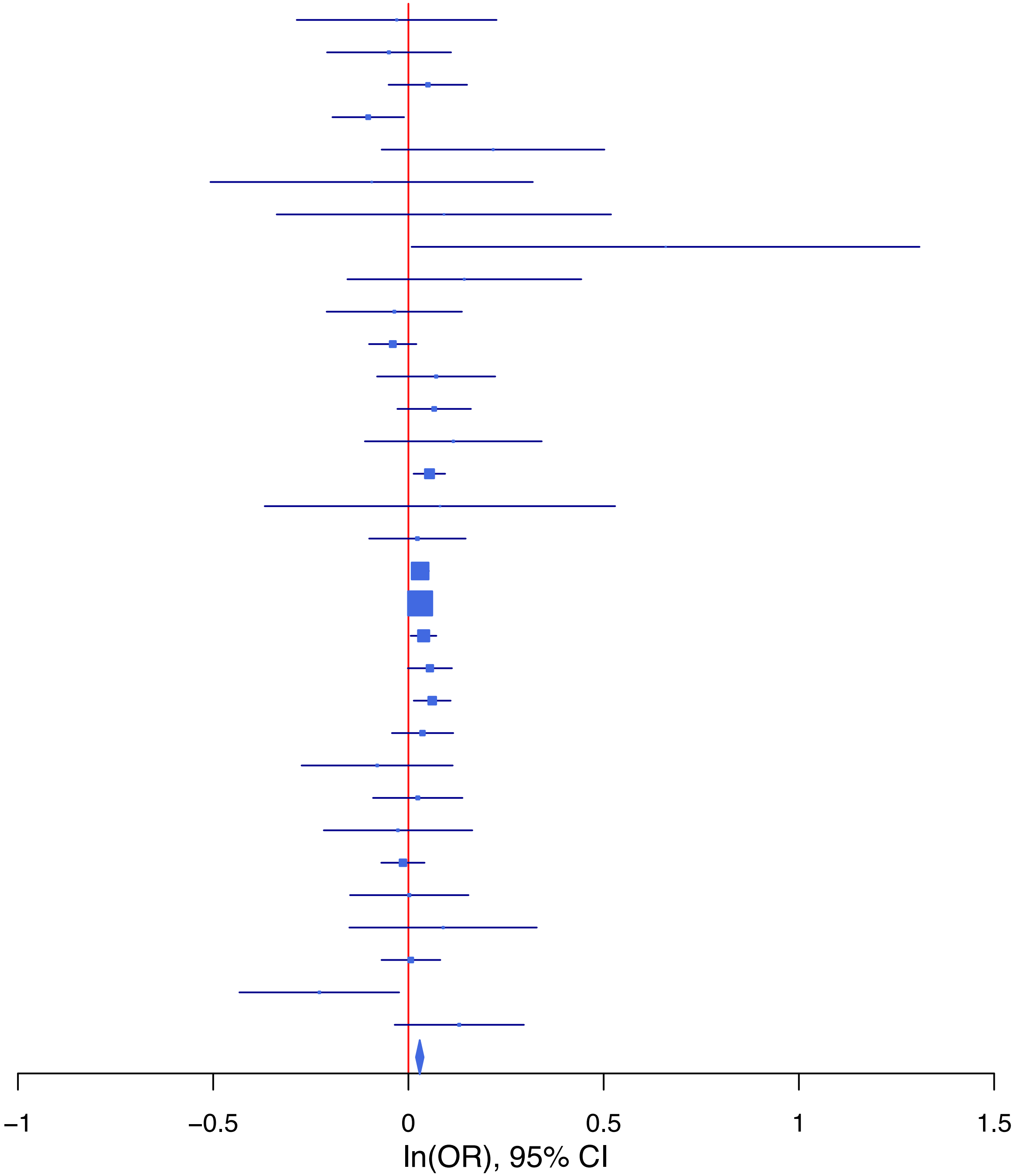

**rs2756119   A/G   14:104001517**

| Cohort | P | ln(OR) | SE |
|---|---|---|---|
| ABCD3_EUR | 0.05 | −0.219 | 0.112 |
| ADHEA_AFR | 0.3147 | −0.082 | 0.081 |
| ADHEA_EUR | 0.4633 | −0.033 | 0.045 |
| ALSPC_EUR | 0.0269 | −0.086 | 0.039 |
| BEPS7_EUR | 0.5688 | 0.07 | 0.122 |
| BHRCM_AFR | 0.4662 | −0.17 | 0.233 |
| BHRCM_EUR | 0.7673 | 0.054 | 0.186 |
| BHRCM_LAT | 0.3773 | −0.219 | 0.248 |
| BOR2C_EUR | 0.9222 | 0.013 | 0.136 |
| CAMHI_EUR | 0.8809 | −0.011 | 0.073 |
| CNVRG_EAS | 0.5936 | −0.016 | 0.03 |
| COGA1_EUR | 0.0329 | −0.088 | 0.041 |
| CVEDA_CSA | 0.4855 | −0.068 | 0.098 |
| ESTB2_EUR | 0.3914 | −0.017 | 0.02 |
| GEDIS_EUR | 0.2528 | −0.111 | 0.097 |
| GEDIS_LAT | 0.8983 | 0.026 | 0.198 |
| GREAT_EAS | 0.0446 | 0.141 | 0.07 |
| MVPXQ_AFR | 0.1578 | −0.016 | 0.012 |
| MVPXQ_EAS | 0.7295 | −0.016 | 0.046 |
| MVPXQ_EUR | 4.23e−06 | −0.032 | 0.007 |
| PGCBD_EUR | 0.0332 | −0.053 | 0.025 |
| PGCMD_EUR | 0.4903 | −0.014 | 0.021 |
| PGCPT_AFR | 0.4295 | −0.057 | 0.072 |
| PGCPT_EUR | 0.2762 | −0.037 | 0.034 |
| PGCPT_LAT | 0.0697 | 0.169 | 0.093 |
| PGCSZ_EUR | 0.3234 | −0.05 | 0.051 |
| PSYCR_EUR | 0.9274 | −0.007 | 0.08 |
| QIMRB_EUR | 0.0132 | −0.059 | 0.024 |
| SNUBH−ASA_EAS | 0.8824 | 0.012 | 0.08 |
| SNUBH−KCHIP_EAS | 0.0029 | −0.364 | 0.12 |
| **meta** | **4.25e−10** | **−0.03** | **0.005** |

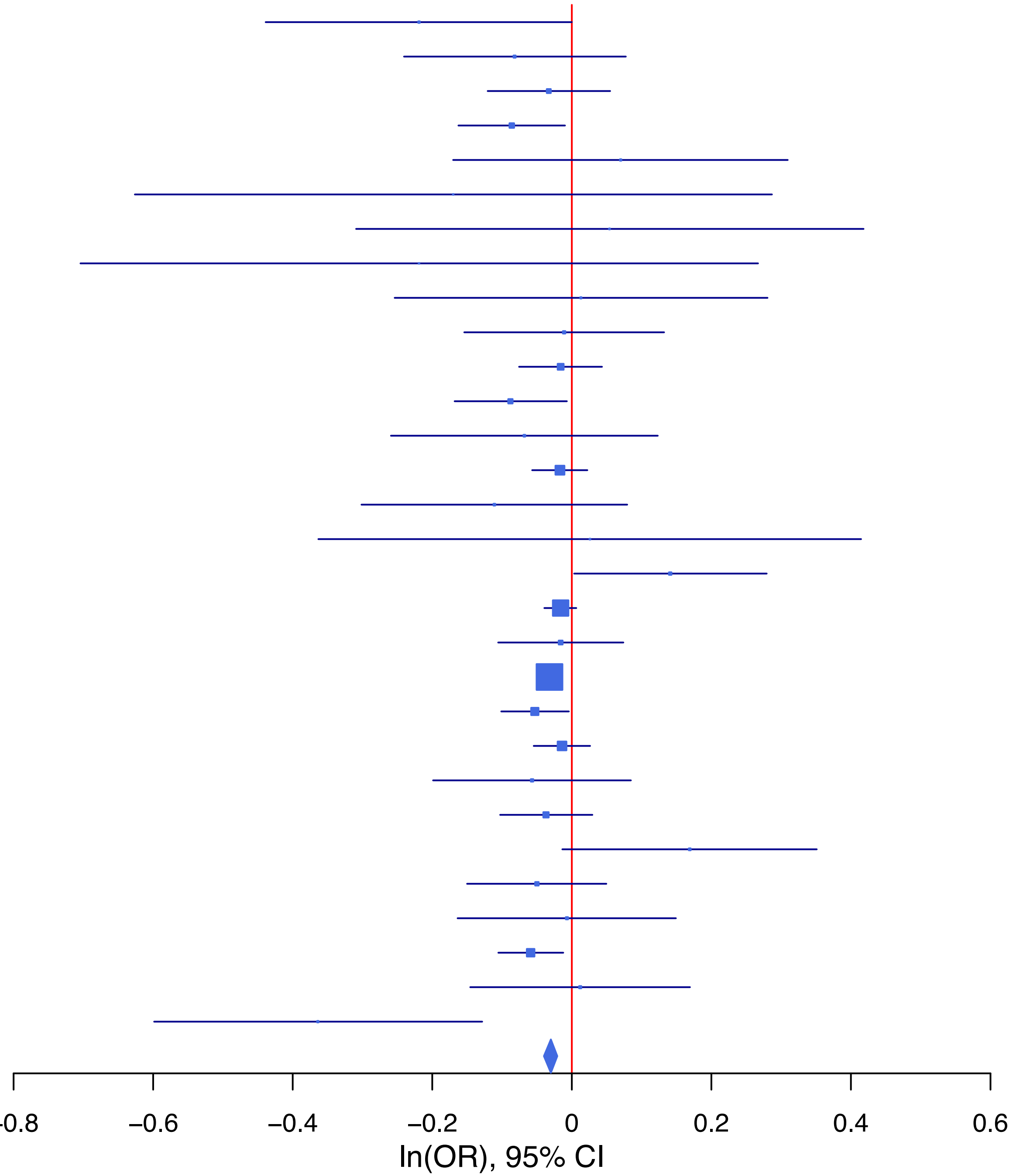

## rs4386905 C/T 7:115076026

| Cohort | P | ln(OR) | SE |
|---|---|---|---|
| ABCD3_EUR | 0.4485 | 0.08 | 0.105 |
| ADHEA_AFR | 0.0721 | −0.165 | 0.092 |
| ADHEA_EUR | 0.3124 | −0.043 | 0.043 |
| ALSPC_EUR | 0.5157 | −0.024 | 0.037 |
| BEPS7_EUR | 0.5087 | −0.073 | 0.111 |
| BHRCM_AFR | 0.3009 | −0.387 | 0.374 |
| BHRCM_EUR | 0.2907 | 0.188 | 0.178 |
| BHRCM_LAT | 0.6398 | −0.142 | 0.303 |
| BOR2C_EUR | 0.2474 | 0.143 | 0.124 |
| CNVRG_EAS | 0.7425 | −0.01 | 0.031 |
| COGA1_AFR | 0.4814 | −0.062 | 0.087 |
| COGA1_EUR | 0.2476 | −0.046 | 0.04 |
| CVEDA_CSA | 0.9361 | −0.008 | 0.099 |
| ESTB2_EUR | 0.0951 | −0.032 | 0.019 |
| GEDIS_EUR | 0.0038 | −0.261 | 0.09 |
| GEDIS_LAT | 0.3656 | −0.178 | 0.197 |
| GREAT_EAS | 0.6614 | −0.031 | 0.07 |
| MIREC_AFR | 0.6313 | 0.054 | 0.113 |
| MIREC_EUR | 0.352 | 0.086 | 0.092 |
| MVPXQ_AFR | 0.4942 | −0.01 | 0.014 |
| MVPXQ_EAS | 0.9912 | 0.001 | 0.048 |
| MVPXQ_EUR | 0.0046 | −0.02 | 0.007 |
| MVPXQ_LAT | 2e−04 | −0.065 | 0.018 |
| PGCBD_EUR | 0.3955 | −0.02 | 0.023 |
| PGCMD_EUR | 0.2658 | −0.022 | 0.02 |
| PGCPT_EUR | 0.7889 | 0.009 | 0.032 |
| PGCPT_LAT | 0.6574 | 0.041 | 0.092 |
| PGCSZ_EUR | 0.3369 | 0.046 | 0.048 |
| PSYCR_EUR | 0.3722 | −0.068 | 0.077 |
| QIMRB_EUR | 6e−04 | −0.078 | 0.023 |
| SNUBH−ASA_EAS | 0.2505 | −0.09 | 0.078 |
| SNUBH−KCHIP_EAS | 0.4478 | −0.095 | 0.125 |
| VUMC1_EUR | 0.1198 | −0.049 | 0.031 |
| YPENN_AFR | 0.2351 | −0.147 | 0.124 |
| YPENN_EUR | 0.9006 | 0.009 | 0.07 |
| **meta** | **3.51e−08** | **−0.026** | **0.005** |

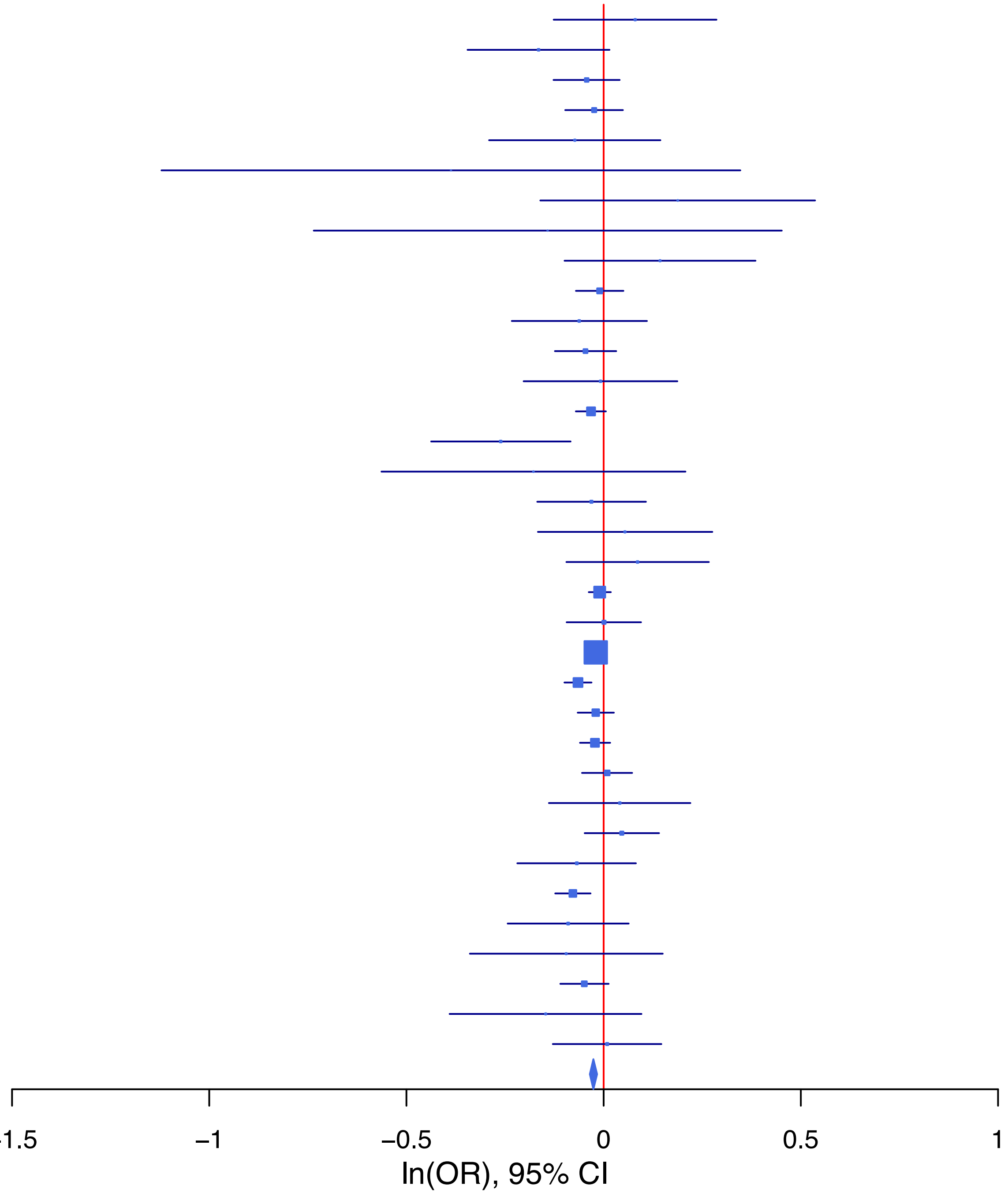

**rs55929017   A/T   17:46995304**

| Cohort | P | ln(OR) | SE |
|---|---|---|---|
| ABCD3_EUR | 0.1368 | −0.218 | 0.146 |
| ALSPC_EUR | 0.0935 | −0.089 | 0.053 |
| BEPS7_EUR | 0.2637 | 0.174 | 0.156 |
| BHRCM_AFR | 0.7662 | 0.114 | 0.383 |
| BHRCM_EUR | 0.9159 | 0.025 | 0.237 |
| BOR2C_EUR | 0.1527 | 0.248 | 0.173 |
| COGA1_EUR | 0.1792 | −0.071 | 0.053 |
| CVEDA_CSA | 0.2177 | −0.21 | 0.17 |
| ESTB2_EUR | 0.4656 | −0.017 | 0.023 |
| GEDIS_EUR | 0.9133 | 0.013 | 0.123 |
| GEDIS_LAT | 0.3484 | −0.269 | 0.287 |
| MVPXQ_AFR | 0.9769 | 0.001 | 0.025 |
| MVPXQ_EUR | 2.67e−06 | −0.042 | 0.009 |
| MVPXQ_LAT | 0.7206 | −0.011 | 0.03 |
| PGCBD_EUR | 0.0083 | −0.087 | 0.033 |
| PGCMD_EUR | 0.4045 | −0.023 | 0.027 |
| PGCPT_EUR | 0.6399 | −0.021 | 0.045 |
| PGCPT_LAT | 0.6283 | −0.078 | 0.161 |
| PGCSZ_EUR | 0.0349 | −0.141 | 0.067 |
| PSYCR_EUR | 0.2106 | −0.133 | 0.106 |
| QIMRB_EUR | 0.3221 | −0.03 | 0.03 |
| **meta** | **2.92e−08** | **−0.037** | **0.007** |

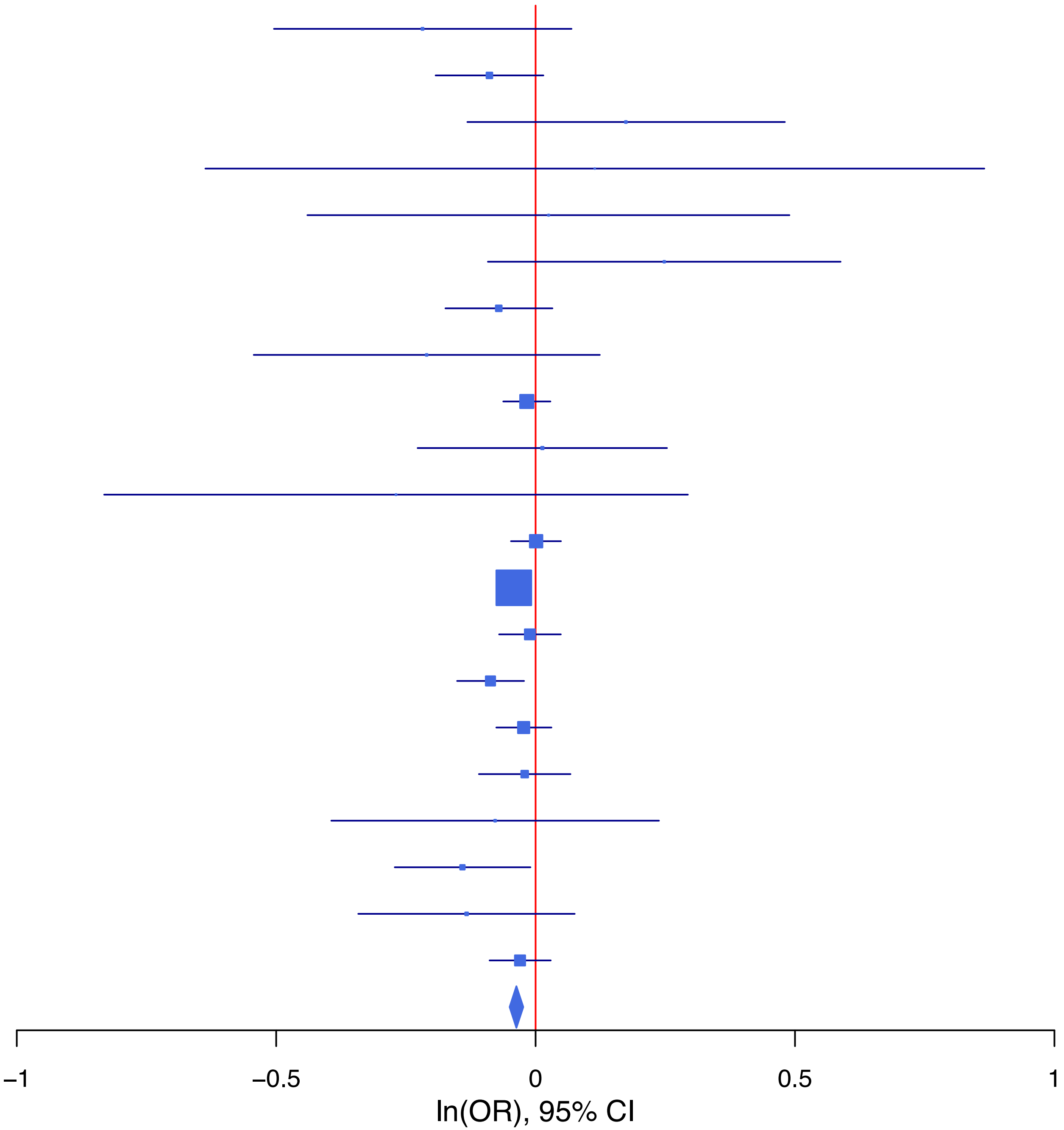

**rs6557168 C/T 6:152201201**

| Cohort | P | ln(OR) | SE |
|---|---|---|---|
| ABCD3_EUR | 0.5679 | −0.063 | 0.11 |
| ALSPC_EUR | 0.7812 | −0.011 | 0.04 |
| BEPS7_EUR | 0.4611 | −0.088 | 0.12 |
| BHRCM_AFR | 0.7394 | −0.081 | 0.243 |
| BHRCM_EUR | 0.8162 | −0.044 | 0.189 |
| BHRCM_LAT | 0.5078 | −0.207 | 0.313 |
| BOR2C_EUR | 0.5334 | 0.082 | 0.131 |
| CAMHI_EUR | 0.1274 | −0.112 | 0.073 |
| CNVRG_EAS | 0.9624 | −0.002 | 0.033 |
| COGA1_AFR | 0.5228 | −0.049 | 0.077 |
| COGA1_EUR | 0.3808 | 0.036 | 0.041 |
| CVEDA_CSA | 0.889 | 0.014 | 0.101 |
| ESTB2_EUR | 0.4313 | 0.017 | 0.021 |
| GEDIS_EUR | 0.7185 | 0.034 | 0.095 |
| GEDIS_LAT | 0.3833 | 0.179 | 0.206 |
| GREAT_EAS | 0.3546 | 0.068 | 0.073 |
| MVPXQ_AFR | 0.0123 | 0.031 | 0.013 |
| MVPXQ_EAS | 0.7932 | 0.013 | 0.051 |
| MVPXQ_EUR | 3.64e−07 | 0.035 | 0.007 |
| MVPXQ_LAT | 0.32 | 0.017 | 0.017 |
| PGCBD_EUR | 0.0252 | 0.056 | 0.025 |
| PGCMD_EUR | 0.7508 | 0.007 | 0.021 |
| PGCPT_AFR | 0.3431 | −0.076 | 0.08 |
| PGCPT_EUR | 0.3186 | −0.034 | 0.034 |
| PGCPT_LAT | 0.3234 | 0.095 | 0.096 |
| PGCSZ_EUR | 0.4318 | 0.04 | 0.051 |
| PSYCR_EUR | 0.2723 | 0.09 | 0.082 |
| QIMRB_EUR | 0.7879 | 0.006 | 0.024 |
| SNUBH-ASA_EAS | 0.2844 | 0.087 | 0.081 |
| SNUBH-KCHIP_EAS | 0.2346 | −0.154 | 0.13 |
| YPENN_EUR | 0.051 | 0.144 | 0.074 |
| **meta** | **2.32e−08** | **0.027** | **0.005** |

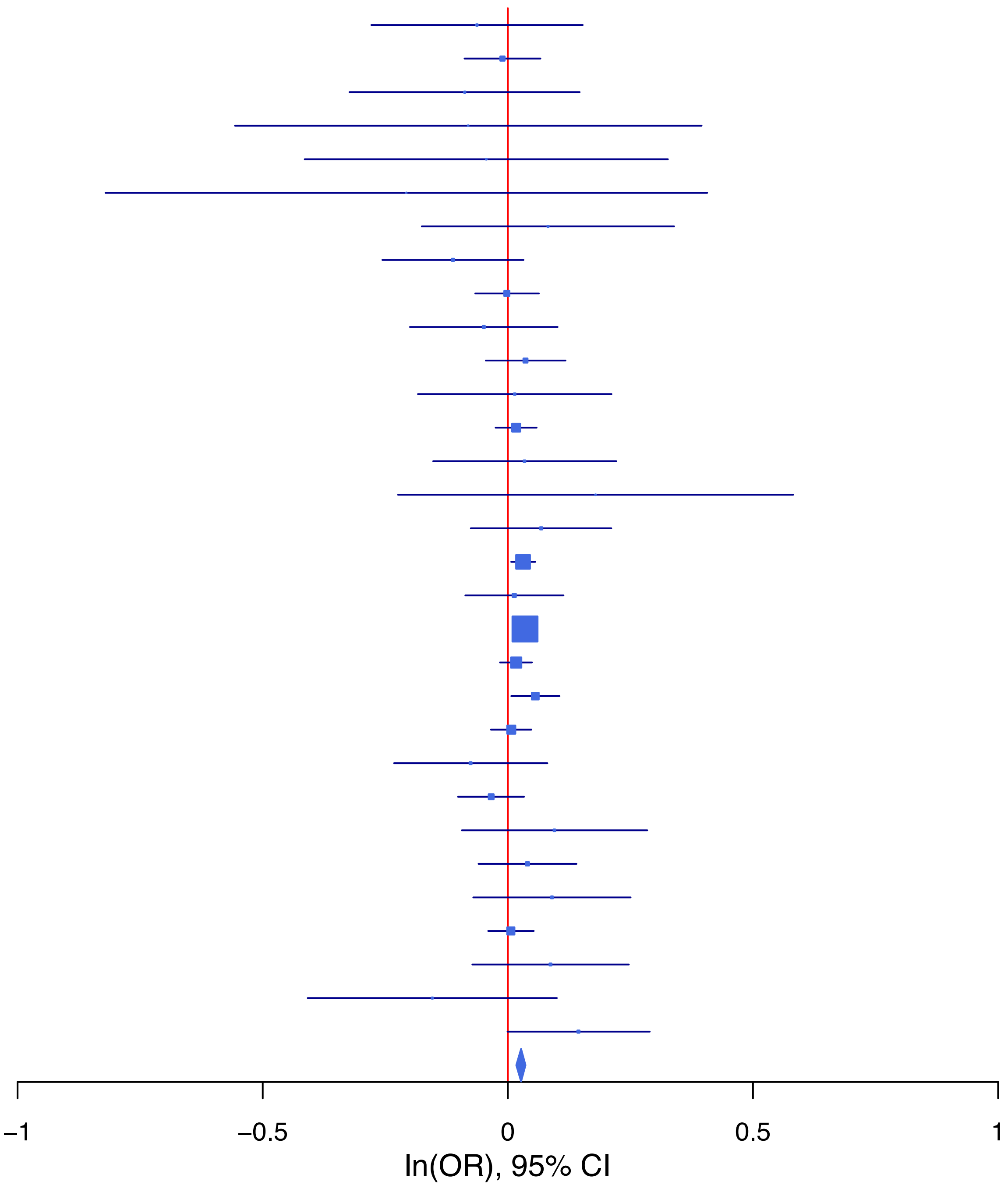

**rs853947 C/T 10:120166631**

| Cohort | P | ln(OR) | SE |
|---|---|---|---|
| ABCD3_EUR | 0.6665 | −0.064 | 0.149 |
| ADHEA_AFR | 0.0037 | −0.443 | 0.153 |
| ADHEA_EUR | 0.8178 | −0.014 | 0.06 |
| ALSPC_EUR | 0.034 | −0.114 | 0.054 |
| BEPS7_EUR | 0.7462 | 0.058 | 0.178 |
| BHRCM_AFR | 0.3907 | 0.409 | 0.476 |
| BHRCM_EUR | 0.1397 | −0.577 | 0.391 |
| BOR2C_EUR | 0.2355 | −0.208 | 0.176 |
| CAMHI_EUR | 0.6053 | −0.051 | 0.099 |
| COGA1_AFR | 0.6677 | −0.056 | 0.131 |
| COGA1_EUR | 0.8152 | −0.013 | 0.056 |
| CVEDA_CSA | 0.3918 | −0.187 | 0.219 |
| ESTB2_EUR | 0.0188 | −0.076 | 0.032 |
| GEDIS_EUR | 0.7834 | −0.036 | 0.13 |
| GEDIS_LAT | 0.5567 | −0.181 | 0.307 |
| MIREC_AFR | 0.3359 | −0.159 | 0.165 |
| MIREC_EUR | 0.1164 | −0.214 | 0.136 |
| MVPXQ_AFR | 0.1745 | −0.027 | 0.02 |
| MVPXQ_EUR | 1e−04 | −0.037 | 0.009 |
| MVPXQ_LAT | 0.4869 | −0.018 | 0.027 |
| PGCBD_EUR | 0.3414 | −0.032 | 0.034 |
| PGCMD_EUR | 0.013 | −0.071 | 0.028 |
| PGCPT_AFR | 0.7617 | −0.036 | 0.118 |
| PGCPT_EUR | 0.6003 | 0.024 | 0.046 |
| PGCPT_LAT | 0.2215 | −0.186 | 0.152 |
| PGCSZ_EUR | 0.4914 | 0.047 | 0.068 |
| PSYCR_EUR | 0.35 | 0.109 | 0.117 |
| QIMRB_EUR | 0.0147 | −0.076 | 0.031 |
| VUMC1_EUR | 0.8849 | −0.006 | 0.044 |
| YPENN_AFR | 0.4359 | 0.148 | 0.19 |
| YPENN_EUR | 0.0373 | −0.224 | 0.107 |
| **meta** | **2.41e−09** | **−0.04** | **0.007** |

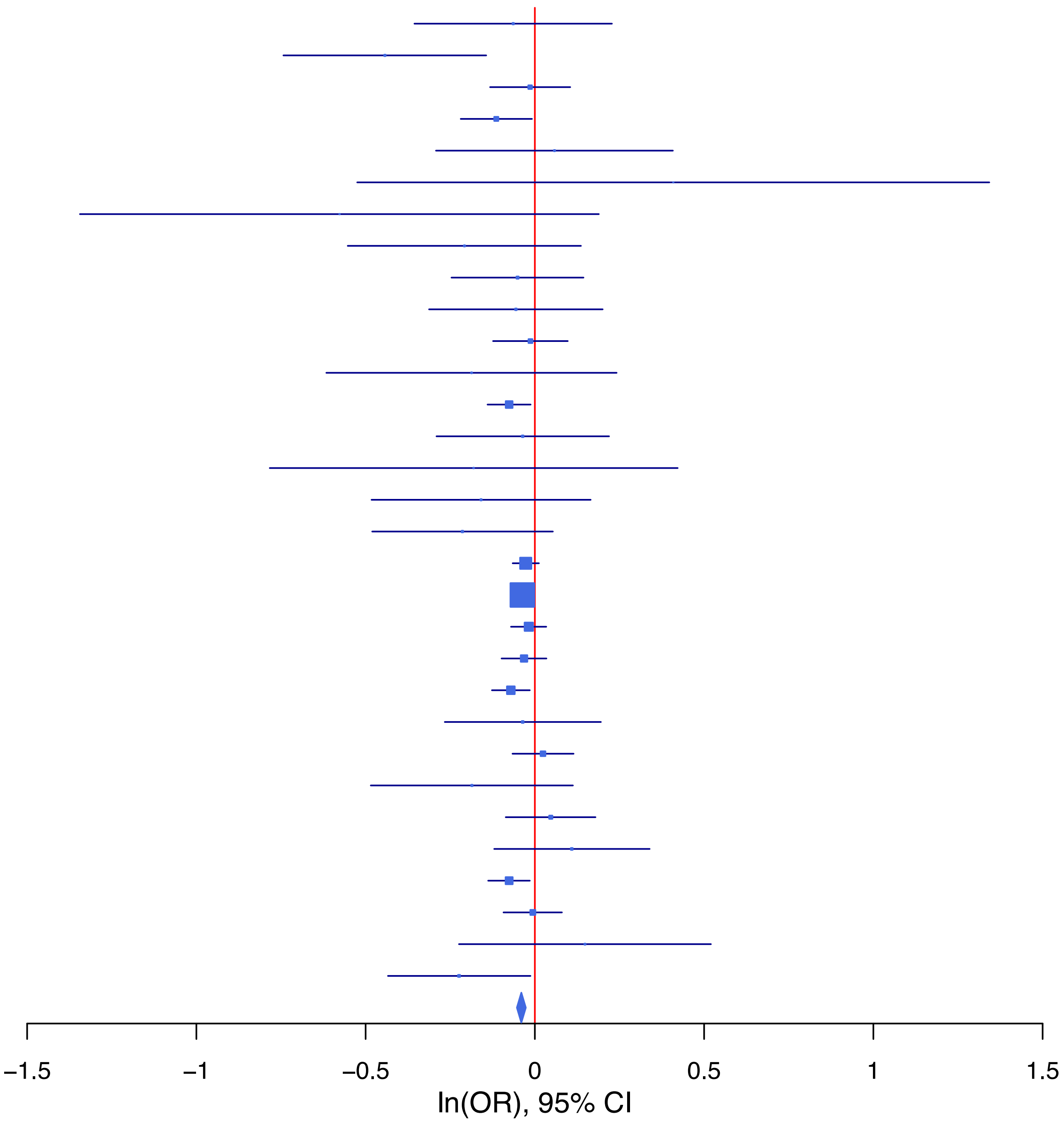

**Supplementary Data 1B: Forest plots of lead SNPs at the 10 genome-wide significant loci from the GWAS meta-analysis of suicidal ideation in European ancestry samples.**
Each box represents the log odds ratio (OR) from an individual contributing cohort, with horizontal lines indicating the 95% confidence interval (CI). The diamond represents the overall meta-analytic estimate across studies.

**rs114164545 A/G 3:61820545**

| Cohort | P | ln(OR) | SE |
|---|---|---|---|
| ABCD3_EUR | 0.8821 | 0.046 | 0.311 |
| ALSPC_EUR | 0.5157 | −0.08 | 0.123 |
| BEPS7_EUR | 0.1649 | −0.508 | 0.366 |
| BOR2C_EUR | 0.4389 | −0.271 | 0.35 |
| CAMHI_EUR | 0.7638 | 0.064 | 0.211 |
| COGA1_EUR | 0.8778 | 0.019 | 0.123 |
| ESTB2_EUR | 0.1961 | −0.058 | 0.045 |
| GEDIS_EUR | 0.6374 | −0.133 | 0.283 |
| MVPXQ_EUR | 1.44e−06 | −0.096 | 0.02 |
| PGCBD_EUR | 0.7732 | −0.026 | 0.09 |
| PGCMD_EUR | 0.0204 | −0.16 | 0.069 |
| PGCPT_EUR | 0.1707 | −0.144 | 0.105 |
| PGCSZ_EUR | 0.8274 | 0.042 | 0.193 |
| PSYCR_EUR | 0.964 | −0.01 | 0.228 |
| QIMRB_EUR | 0.0337 | −0.156 | 0.073 |
| VUMC1_EUR | 0.6236 | −0.048 | 0.098 |
| YPENN_EUR | 0.5709 | −0.133 | 0.234 |
| **meta** | **6.86e−09** | **−0.092** | **0.016** |

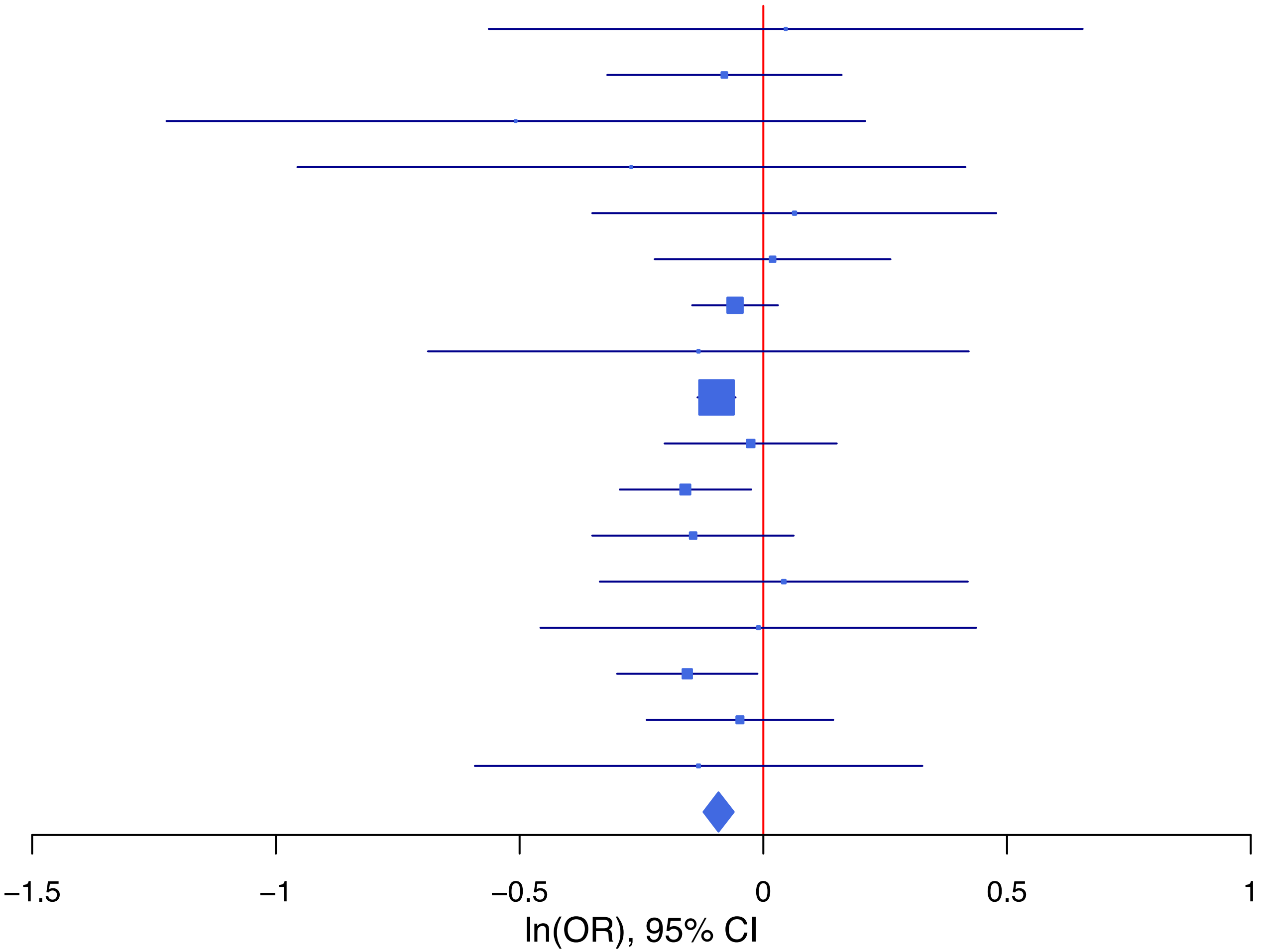

## rs11591571 A/G 10:104342804

| Cohort | P | ln(OR) | SE |
|---|---|---|---|
| ABCD3_EUR | 0.1293 | 0.165 | 0.108 |
| ADHEA_EUR | 0.4983 | 0.031 | 0.045 |
| ALSPC_EUR | 0.0237 | 0.09 | 0.04 |
| BEPS7_EUR | 0.1127 | −0.199 | 0.125 |
| BHRCM_EUR | 0.3714 | 0.164 | 0.184 |
| BOR2C_EUR | 0.8616 | −0.023 | 0.131 |
| CAMHI_EUR | 0.4783 | 0.053 | 0.075 |
| COGA1_EUR | 0.6193 | 0.021 | 0.042 |
| ESTB2_EUR | 0.0953 | 0.037 | 0.022 |
| GEDIS_EUR | 0.0293 | 0.206 | 0.095 |
| MIREC_EUR | 0.9781 | −0.003 | 0.098 |
| MVPXQ_EUR | 4.43e−06 | 0.031 | 0.007 |
| PGCBD_EUR | 0.0697 | 0.045 | 0.025 |
| PGCMD_EUR | 0.6783 | 0.009 | 0.021 |
| PGCPT_EUR | 0.8475 | −0.007 | 0.034 |
| PGCSZ_EUR | 0.0754 | 0.09 | 0.051 |
| PSYCR_EUR | 0.2754 | −0.091 | 0.084 |
| QIMRB_EUR | 0.21 | 0.03 | 0.024 |
| VUMC1_EUR | 0.7205 | −0.012 | 0.033 |
| **meta** | **2.84e−08** | **0.03** | **0.005** |

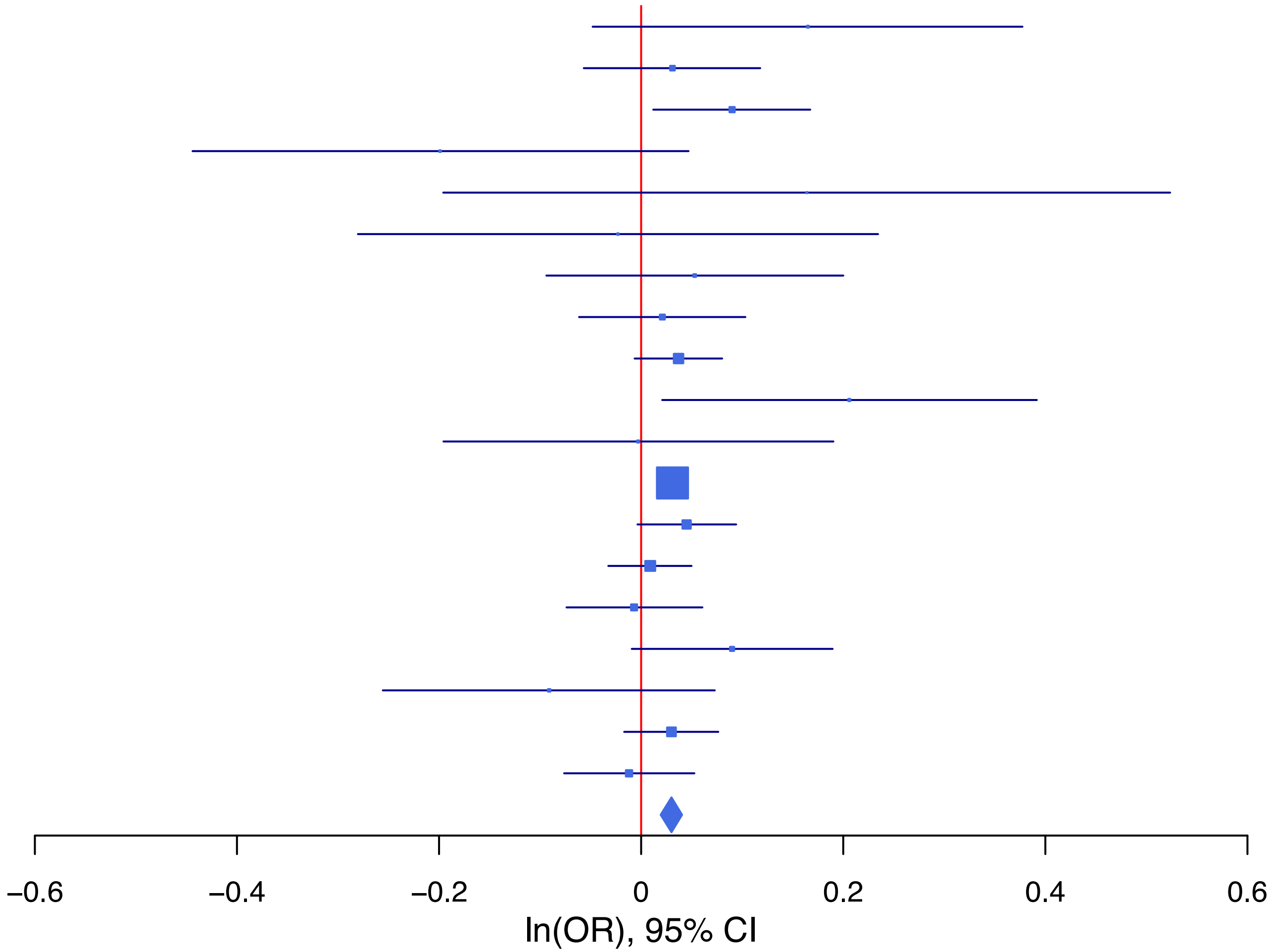

**rs17514846 A/C 15:91416550**

| Cohort | P | ln(OR) | SE |
|---|---|---|---|
| ABCD3_EUR | 0.8171 | 0.025 | 0.105 |
| ALSPC_EUR | 0.2034 | −0.049 | 0.039 |
| BEPS7_EUR | 0.7626 | 0.033 | 0.109 |
| BHRCM_EUR | 0.1561 | −0.266 | 0.187 |
| BOR2C_EUR | 0.4059 | −0.104 | 0.125 |
| CAMHI_EUR | 0.1985 | −0.088 | 0.069 |
| COGA1_EUR | 0.1295 | −0.06 | 0.04 |
| GEDIS_EUR | 0.3009 | −0.094 | 0.091 |
| MVPXQ_EUR | 1e−04 | −0.025 | 0.007 |
| PGCBD_EUR | 1e−04 | −0.098 | 0.025 |
| PGCMD_EUR | 0.0574 | −0.038 | 0.02 |
| PGCPT_EUR | 0.539 | −0.02 | 0.033 |
| PGCSZ_EUR | 0.487 | 0.035 | 0.051 |
| PSYCR_EUR | 0.9564 | −0.004 | 0.077 |
| QIMRB_EUR | 4e−04 | −0.081 | 0.023 |
| VUMC1_EUR | 0.0659 | −0.058 | 0.031 |
| YPENN_EUR | 0.0313 | −0.154 | 0.071 |
| **meta** | **8.60e−11** | **−0.035** | **0.005** |

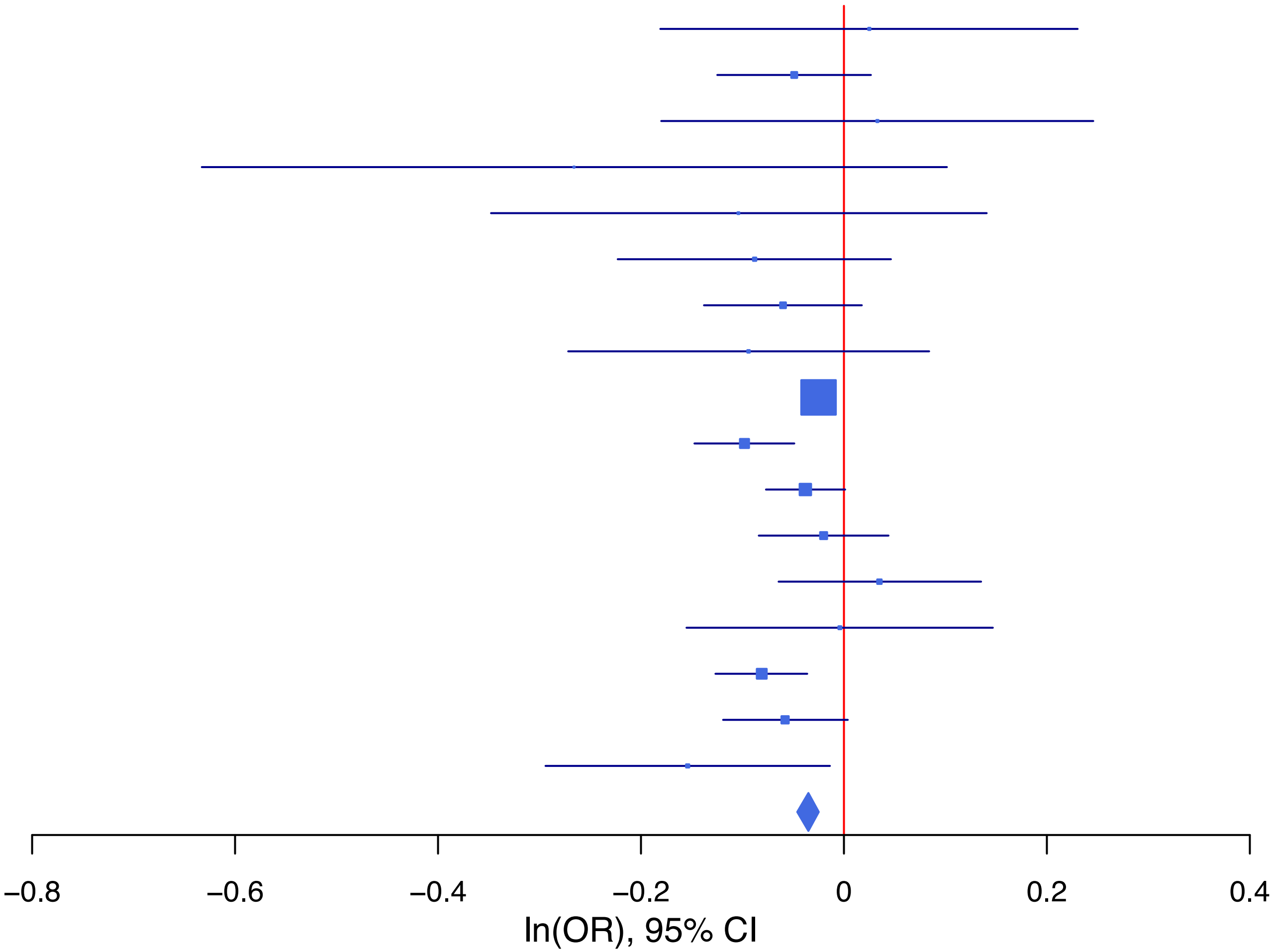

**rs2514218 T/C 11:113392994**

| Cohort | P | ln(OR) | SE |
|---|---|---|---|
| ABCD3_EUR | 0.155 | −0.161 | 0.113 |
| ADHEA_EUR | 0.151 | −0.065 | 0.045 |
| BEPS7_EUR | 0.7561 | −0.038 | 0.122 |
| BHRCM_EUR | 0.2214 | −0.222 | 0.182 |
| BOR2C_EUR | 0.5106 | 0.086 | 0.13 |
| CAMHI_EUR | 0.5142 | −0.048 | 0.073 |
| COGA1_EUR | 0.796 | −0.011 | 0.041 |
| ESTB2_EUR | 0.3238 | −0.022 | 0.022 |
| GEDIS_EUR | 0.0341 | −0.205 | 0.097 |
| MVPXQ_EUR | 3.49e−05 | −0.027 | 0.007 |
| PGCBD_EUR | 0.0432 | −0.05 | 0.025 |
| PGCMD_EUR | 0.0192 | −0.048 | 0.02 |
| PGCPT_EUR | 0.6063 | 0.017 | 0.034 |
| PGCSZ_EUR | 0.0226 | −0.117 | 0.051 |
| PSYCR_EUR | 7e−04 | 0.274 | 0.081 |
| QIMRB_EUR | 0.2559 | −0.027 | 0.023 |
| VUMC1_EUR | 0.0784 | −0.058 | 0.033 |
| YPENN_EUR | 0.4879 | −0.051 | 0.074 |
| **meta** | **1.80e−08** | **−0.03** | **0.005** |

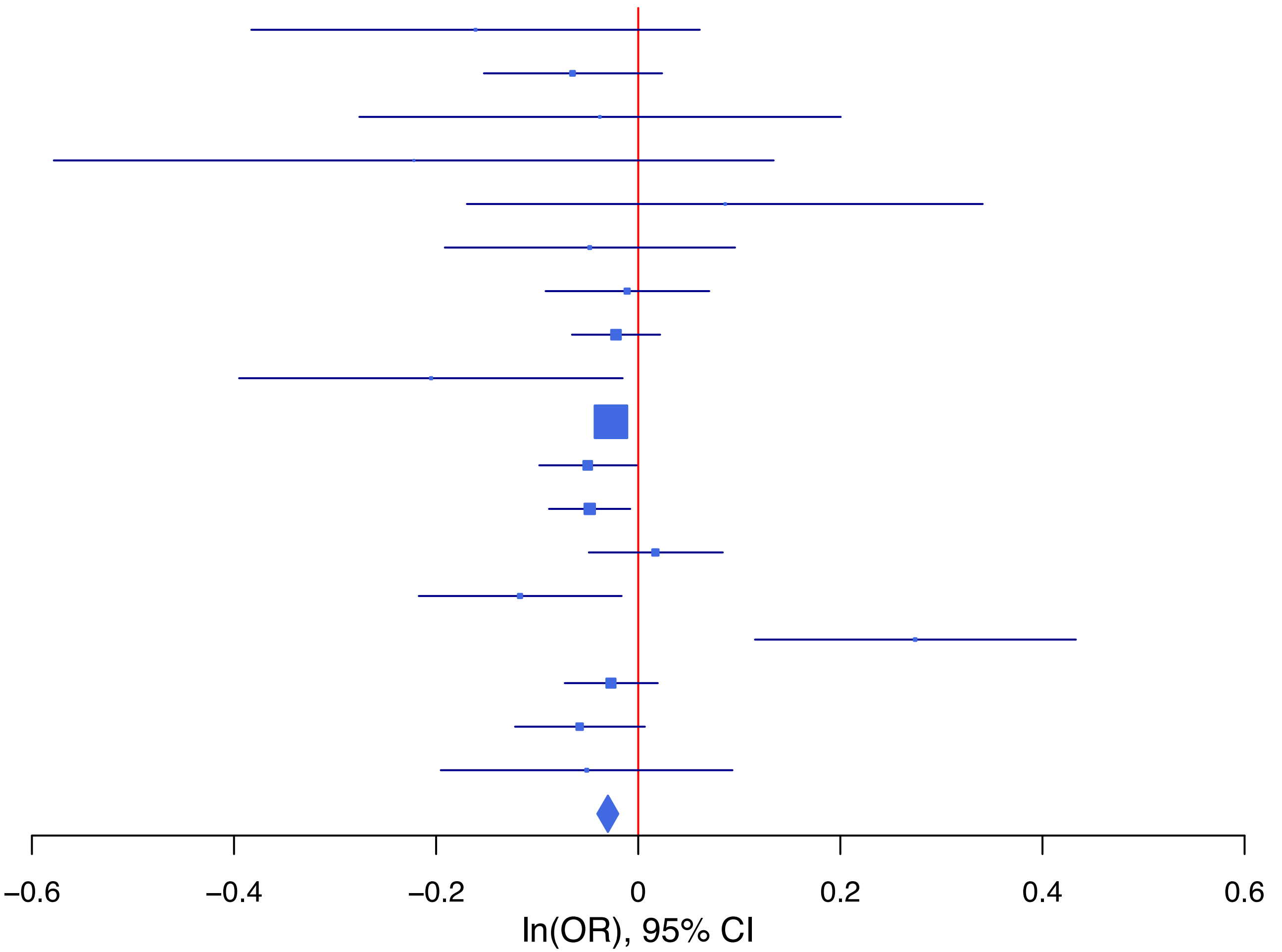

**rs2756119 A/G 14:104001517**

| Cohort | P | ln(OR) | SE |
|---|---|---|---|
| ABCD3_EUR | 0.05 | −0.219 | 0.112 |
| ADHEA_EUR | 0.4633 | −0.033 | 0.045 |
| ALSPC_EUR | 0.0269 | −0.086 | 0.039 |
| BEPS7_EUR | 0.5688 | 0.07 | 0.122 |
| BHRCM_EUR | 0.7673 | 0.054 | 0.186 |
| BOR2C_EUR | 0.9222 | 0.013 | 0.136 |
| CAMHI_EUR | 0.8809 | −0.011 | 0.073 |
| COGA1_EUR | 0.0329 | −0.088 | 0.041 |
| ESTB2_EUR | 0.3914 | −0.017 | 0.02 |
| GEDIS_EUR | 0.2528 | −0.111 | 0.097 |
| MVPXQ_EUR | 4.23e−06 | −0.032 | 0.007 |
| PGCBD_EUR | 0.0332 | −0.053 | 0.025 |
| PGCMD_EUR | 0.4903 | −0.014 | 0.021 |
| PGCPT_EUR | 0.2762 | −0.037 | 0.034 |
| PGCSZ_EUR | 0.3234 | −0.05 | 0.051 |
| PSYCR_EUR | 0.9274 | −0.007 | 0.08 |
| QIMRB_EUR | 0.0132 | −0.059 | 0.024 |
| **meta** | **3.74e−10** | **−0.035** | **0.006** |

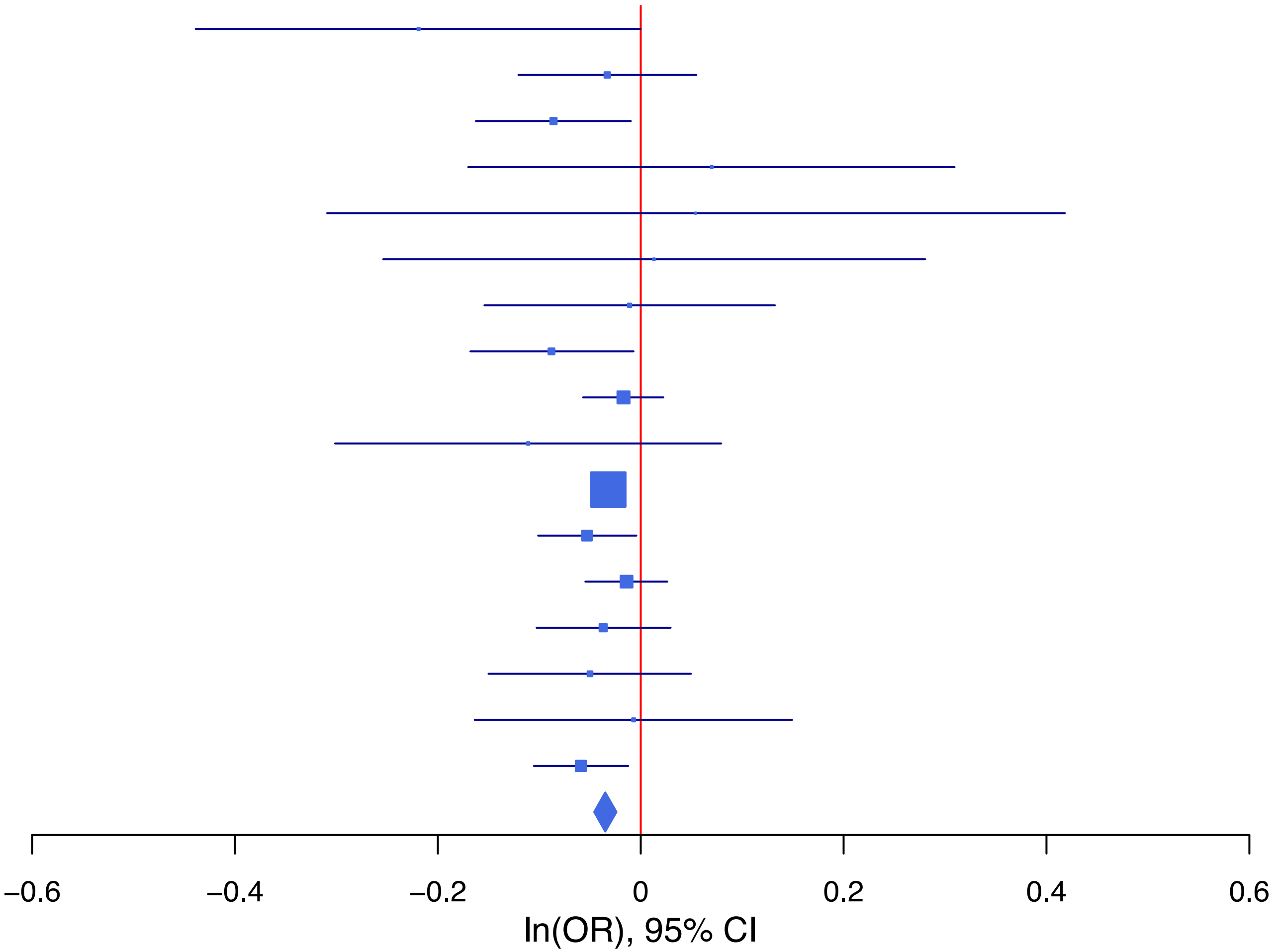

## rs35161231 G/C 3:76220819

| Cohort | P | ln(OR) | SE |
|---|---|---|---|
| ABCD3_EUR | 0.0017 | −0.357 | 0.114 |
| ADHEA_EUR | 0.8511 | −0.008 | 0.044 |
| ALSPC_EUR | 0.1289 | 0.059 | 0.039 |
| BEPS7_EUR | 0.9723 | −0.004 | 0.118 |
| BHRCM_EUR | 0.2548 | 0.218 | 0.191 |
| BOR2C_EUR | 0.2017 | 0.165 | 0.129 |
| CAMHI_EUR | 0.4411 | 0.056 | 0.073 |
| COGA1_EUR | 0.0827 | 0.072 | 0.042 |
| ESTB2_EUR | 0.16 | 0.028 | 0.02 |
| GEDIS_EUR | 0.0534 | 0.181 | 0.094 |
| MVPXQ_EUR | 1e−04 | 0.025 | 0.006 |
| PGCBD_EUR | 0.126 | 0.038 | 0.024 |
| PGCMD_EUR | 0.0162 | 0.048 | 0.02 |
| PGCPT_EUR | 0.5736 | 0.019 | 0.034 |
| PGCSZ_EUR | 0.095 | 0.082 | 0.049 |
| PSYCR_EUR | 0.3717 | −0.07 | 0.078 |
| QIMRB_EUR | 0.0798 | 0.041 | 0.023 |
| VUMC1_EUR | 0.003 | 0.095 | 0.032 |
| YPENN_EUR | 0.4194 | 0.059 | 0.073 |
| **meta** | **1.33e−09** | **0.031** | **0.005** |

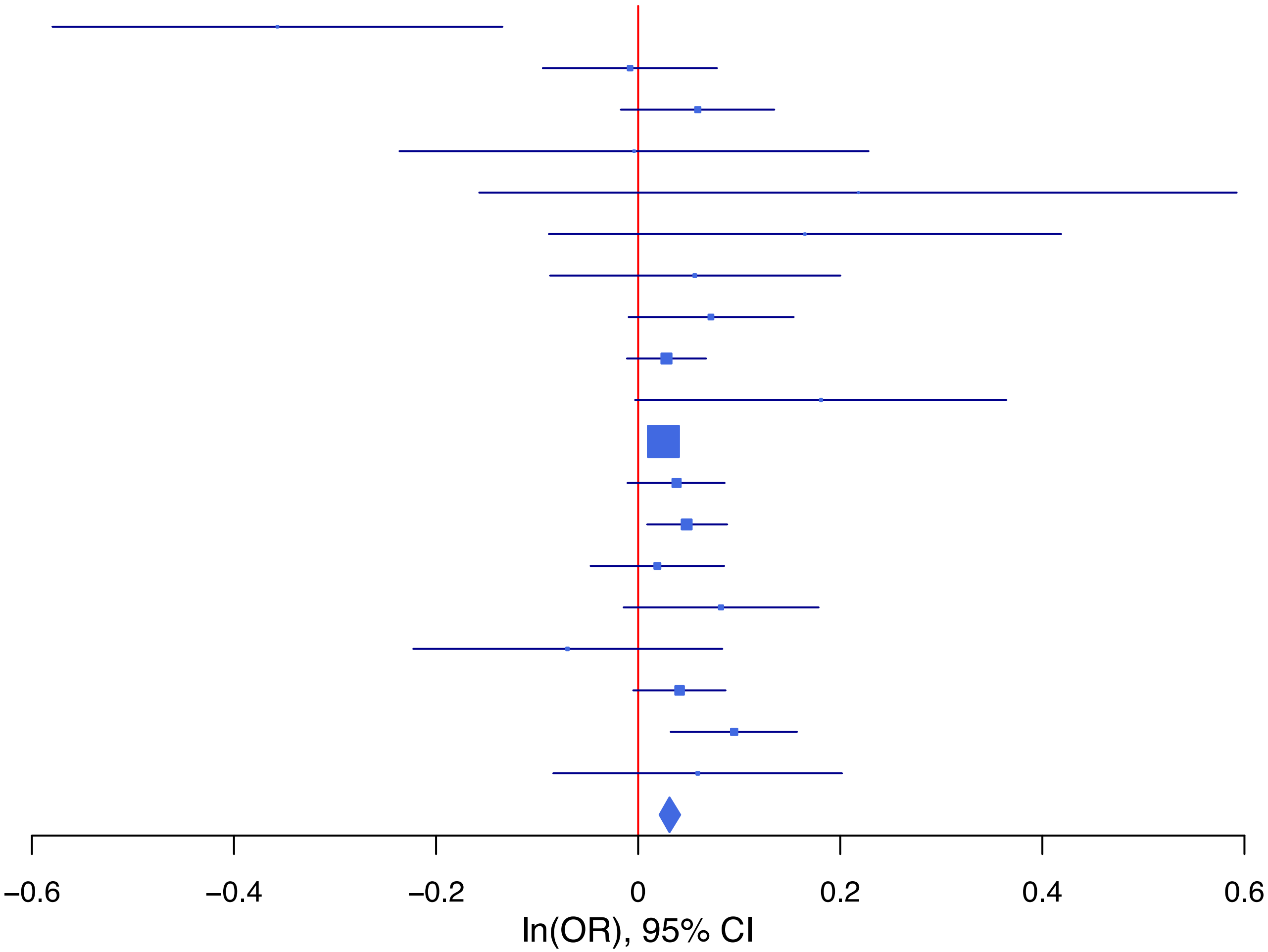

**rs3823624 C/T 7:2110346**

| Cohort | P | ln(OR) | SE |
|---|---|---|---|
| ABCD3_EUR | 0.3276 | 0.124 | 0.127 |
| ADHEA_EUR | 0.4838 | −0.038 | 0.055 |
| BEPS7_EUR | 0.0543 | −0.254 | 0.132 |
| BHRCM_EUR | 0.1282 | −0.414 | 0.272 |
| BOR2C_EUR | 0.2966 | −0.159 | 0.153 |
| CAMHI_EUR | 0.0253 | −0.195 | 0.087 |
| COGA1_EUR | 0.9875 | −0.001 | 0.05 |
| ESTB2_EUR | 0.0552 | −0.042 | 0.022 |
| GEDIS_EUR | 0.9911 | −0.001 | 0.115 |
| MVPXQ_EUR | 5.85e−07 | −0.041 | 0.008 |
| PGCMD_EUR | 0.5358 | 0.015 | 0.024 |
| PGCPT_EUR | 0.2377 | −0.049 | 0.042 |
| PGCSZ_EUR | 0.6612 | 0.027 | 0.061 |
| PSYCR_EUR | 0.9462 | −0.006 | 0.087 |
| QIMRB_EUR | 0.1109 | −0.046 | 0.029 |
| VUMC1_EUR | 0.7959 | 0.01 | 0.04 |
| YPENN_EUR | 0.0402 | −0.181 | 0.088 |
| **meta** | **4.62e−08** | **−0.036** | **0.007** |

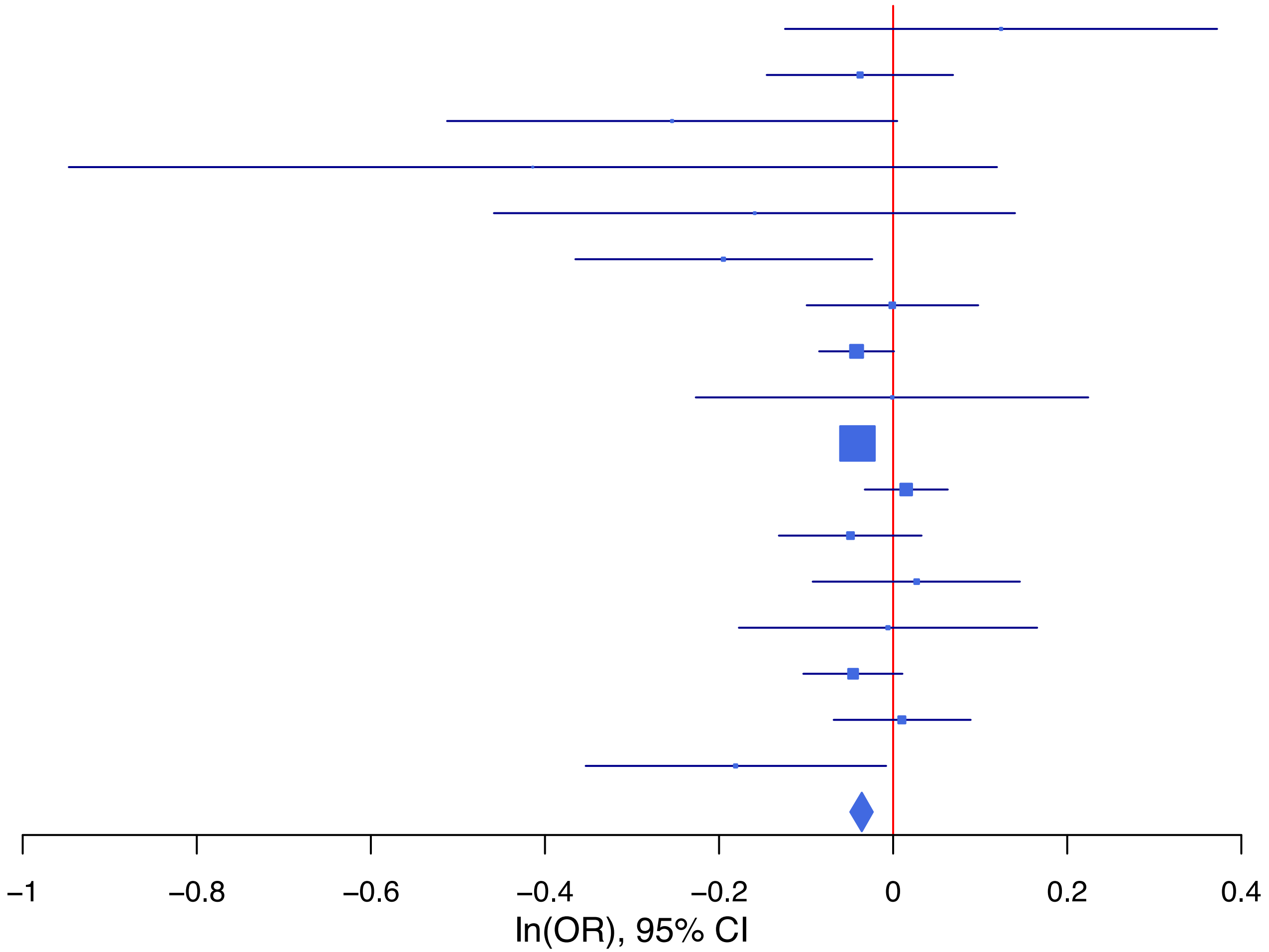

**rs55929017 A/T 17:46995304**

| Cohort | P | ln(OR) | SE |
|---|---|---|---|
| ABCD3_EUR | 0.1368 | −0.218 | 0.146 |
| ALSPC_EUR | 0.0935 | −0.089 | 0.053 |
| BEPS7_EUR | 0.2637 | 0.174 | 0.156 |
| BHRCM_EUR | 0.9159 | 0.025 | 0.237 |
| BOR2C_EUR | 0.1527 | 0.248 | 0.173 |
| COGA1_EUR | 0.1792 | −0.071 | 0.053 |
| ESTB2_EUR | 0.4656 | −0.017 | 0.023 |
| GEDIS_EUR | 0.9133 | 0.013 | 0.123 |
| MVPXQ_EUR | 2.67e−06 | −0.042 | 0.009 |
| PGCBD_EUR | 0.0083 | −0.087 | 0.033 |
| PGCMD_EUR | 0.4045 | −0.023 | 0.027 |
| PGCPT_EUR | 0.6399 | −0.021 | 0.045 |
| PGCSZ_EUR | 0.0349 | −0.141 | 0.067 |
| PSYCR_EUR | 0.2106 | −0.133 | 0.106 |
| QIMRB_EUR | 0.3221 | −0.03 | 0.03 |
| **meta** | **8.15e−09** | **−0.041** | **0.007** |

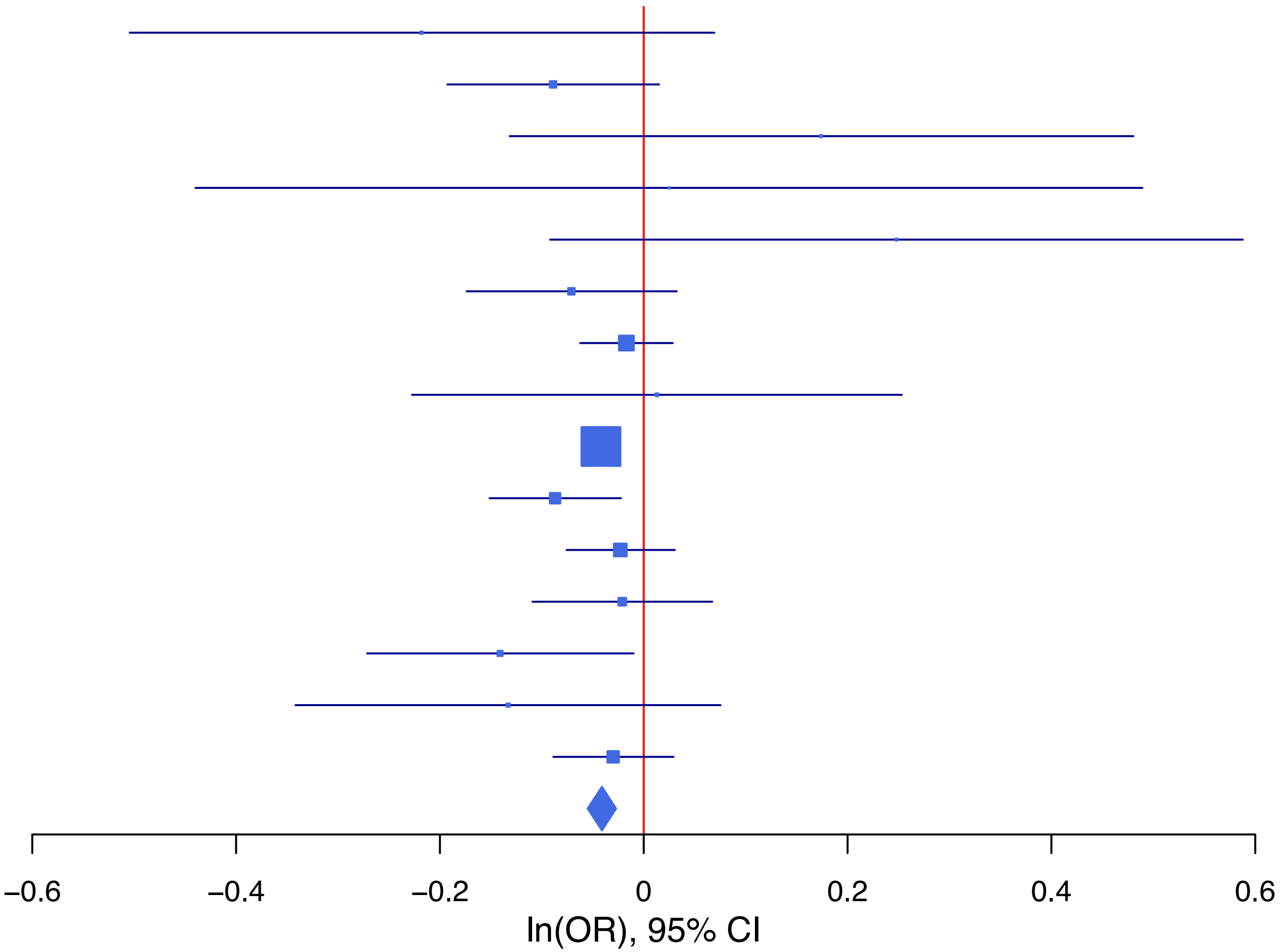

**rs73581567 T/C 9:140247759**

| Cohort | P | ln(OR) | SE |
|---|---|---|---|
| ABCD3_EUR | 0.4696 | 0.105 | 0.145 |
| BEPS7_EUR | 0.0023 | 0.501 | 0.164 |
| BHRCM_EUR | 0.9021 | −0.036 | 0.29 |
| BOR2C_EUR | 0.2698 | 0.193 | 0.175 |
| CAMHI_EUR | 0.5826 | 0.057 | 0.104 |
| ESTB2_EUR | 0.0164 | 0.063 | 0.026 |
| MVPXQ_EUR | 1.82e−07 | 0.049 | 0.009 |
| PGCBD_EUR | 0.4279 | 0.04 | 0.05 |
| PGCMD_EUR | 0.4095 | 0.03 | 0.037 |
| PGCPT_EUR | 0.1453 | 0.063 | 0.043 |
| PGCSZ_EUR | 0.5959 | 0.054 | 0.101 |
| PSYCR_EUR | 0.9988 | 0 | 0.108 |
| QIMRB_EUR | 0.0075 | 0.086 | 0.032 |
| YPENN_EUR | 0.9564 | 0.006 | 0.108 |
| **meta** | **2.50e−11** | **0.053** | **0.008** |

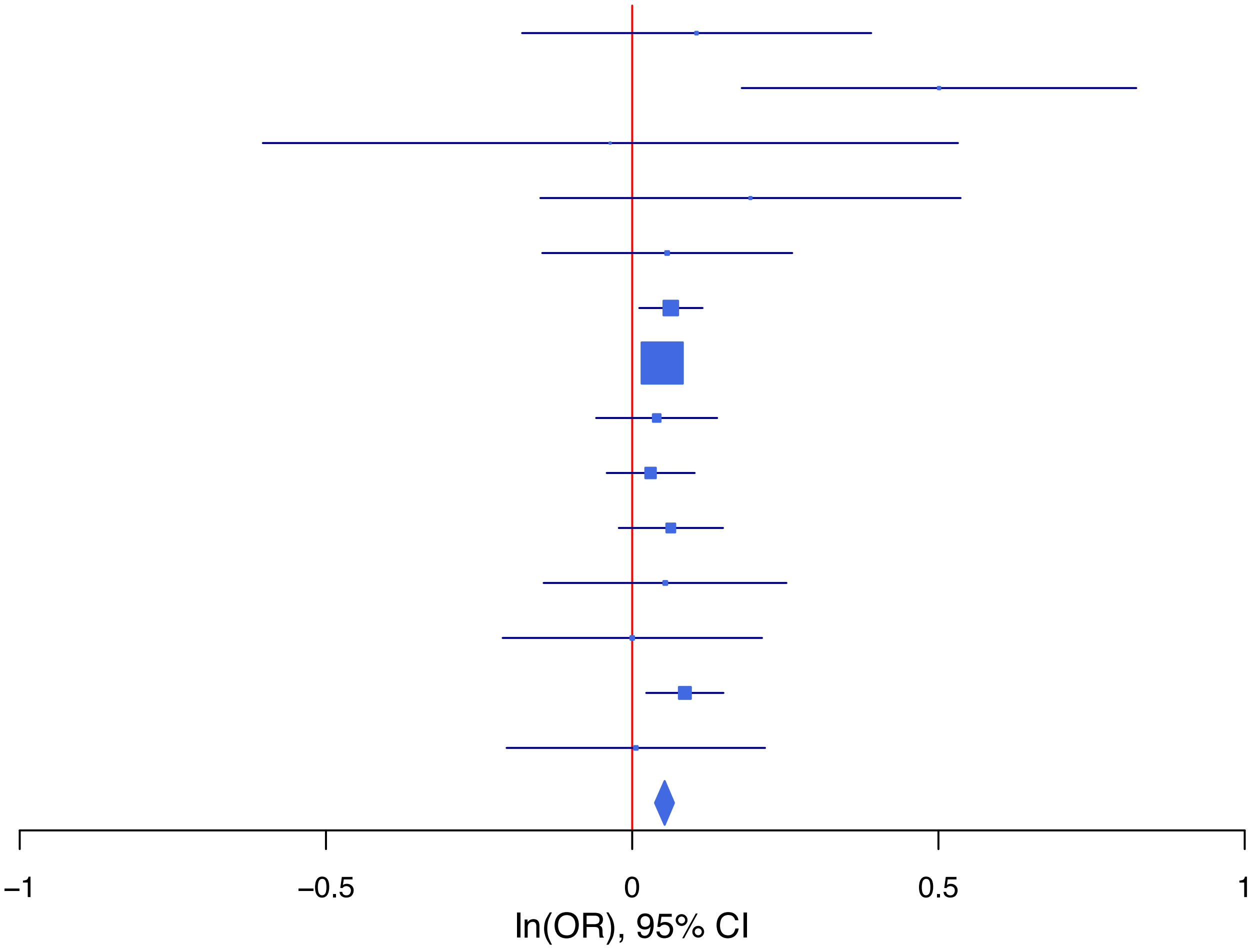

**rs853937 C/T 10:120162568**

| Cohort | P | ln(OR) | SE |
|---|---|---|---|
| ABCD3_EUR | 0.6598 | −0.065 | 0.149 |
| ADHEA_EUR | 0.9013 | −0.007 | 0.06 |
| ALSPC_EUR | 0.0391 | −0.11 | 0.054 |
| BEPS7_EUR | 0.7061 | 0.067 | 0.178 |
| BHRCM_EUR | 0.1397 | −0.577 | 0.391 |
| BOR2C_EUR | 0.2439 | −0.205 | 0.176 |
| CAMHI_EUR | 0.5267 | −0.063 | 0.099 |
| COGA1_EUR | 0.7016 | −0.022 | 0.056 |
| ESTB2_EUR | 0.0186 | −0.076 | 0.032 |
| GEDIS_EUR | 0.8243 | −0.029 | 0.13 |
| MIREC_EUR | 0.1079 | −0.219 | 0.136 |
| MVPXQ_EUR | 1e−04 | −0.037 | 0.009 |
| PGCBD_EUR | 0.3828 | −0.029 | 0.034 |
| PGCMD_EUR | 0.0139 | −0.07 | 0.028 |
| PGCPT_EUR | 0.6302 | 0.022 | 0.046 |
| PGCSZ_EUR | 0.5156 | 0.044 | 0.068 |
| PSYCR_EUR | 0.336 | 0.113 | 0.117 |
| QIMRB_EUR | 0.0162 | −0.075 | 0.031 |
| VUMC1_EUR | 0.9322 | −0.004 | 0.044 |
| YPENN_EUR | 0.0281 | −0.236 | 0.107 |
| **meta** | **1.53e−08** | **−0.042** | **0.007** |

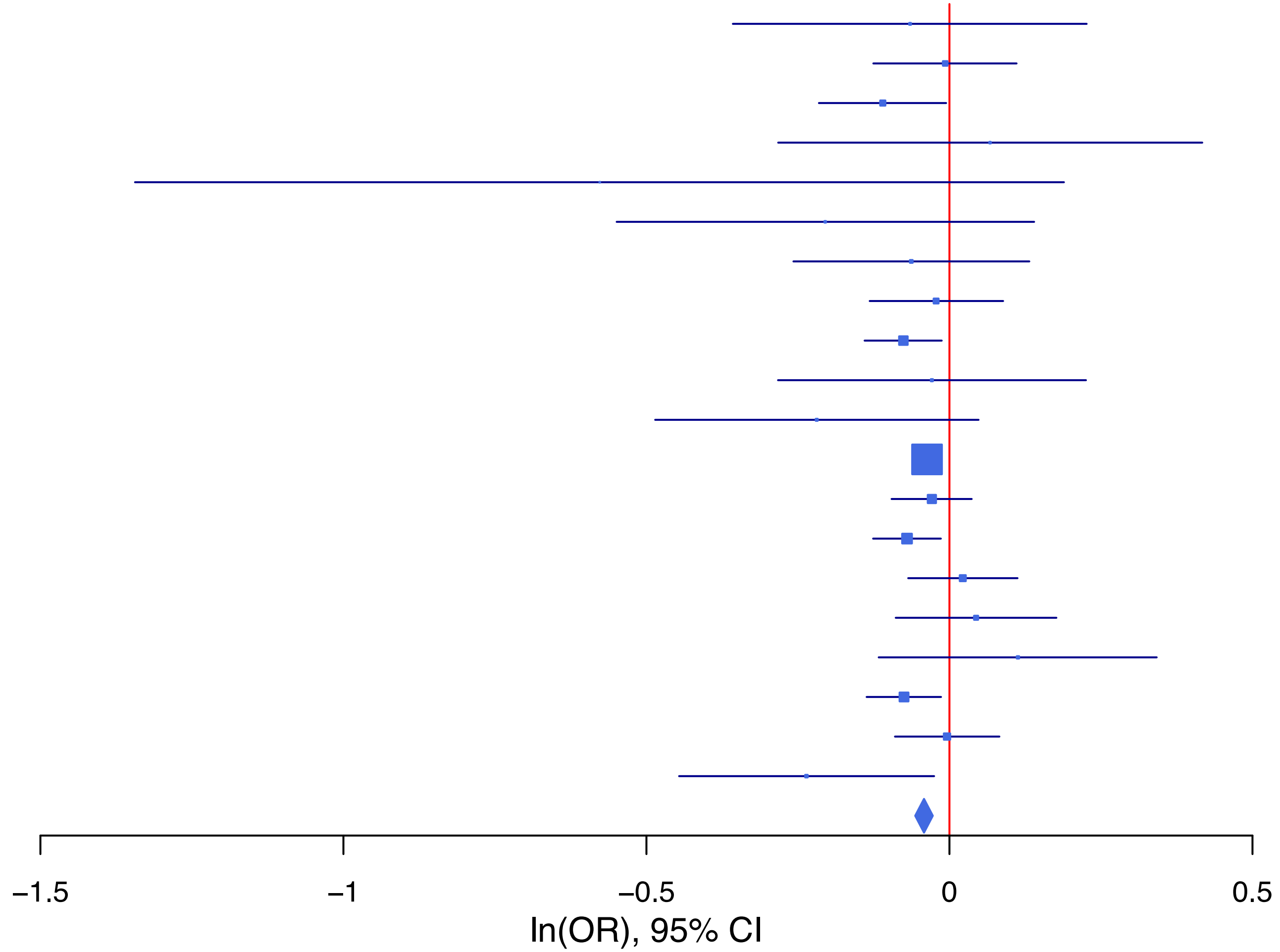

**Supplementary Data 1C: Forest plot of the lead SNP at the genome-wide significant locus from the GWAS meta-analysis of suicidal ideation in East Asian ancestry samples.**
Each box represents the log odds ratio (OR) from an individual contributing cohort, with horizontal lines indicating the 95% confidence interval (CI). The diamond represents the overall meta-analytic estimate across studies.

**rs60008145   T/G   10:69715297**

| Cohort | P | ln(OR) | SE |
|---|---|---|---|
| CNVRG_EAS | 4.81e−07 | −0.165 | 0.033 |
| MVPXQ_EAS | 0.0858 | −0.098 | 0.057 |
| SNUBH−ASA_EAS | 0.462 | −0.062 | 0.085 |
| SNUBH−KCHIP_EAS | 0.0501 | −0.252 | 0.126 |
| **meta** | **4.02e−08** | **−0.145** | **0.026** |

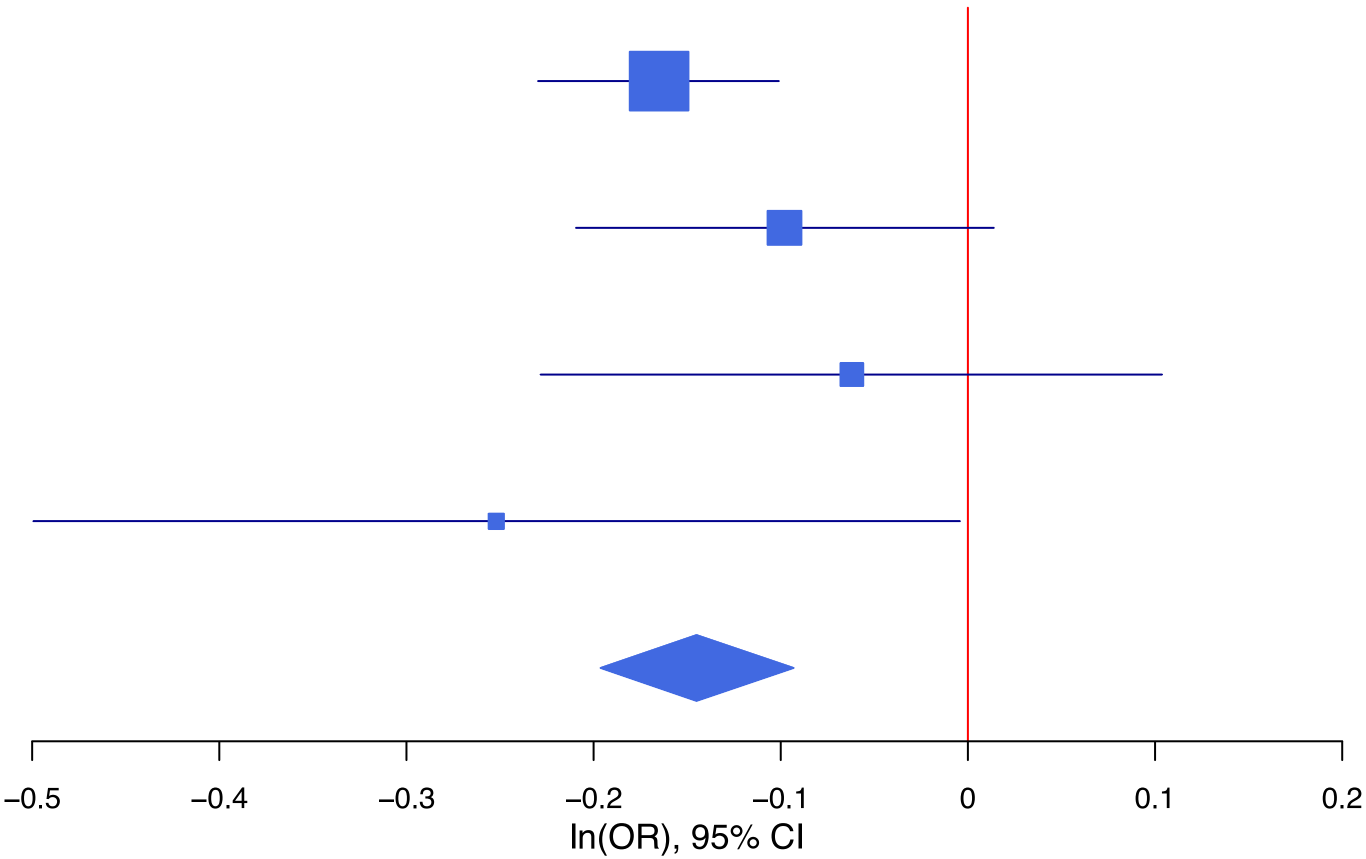

**Supplementary Data 1D: Forest plots of lead SNPs at the 37 genome-wide significant loci from the multi-ancestry GWAS meta-analysis of suicide attempt.**

Each box represents the log odds ratio (OR) from an individual contributing cohort, with horizontal lines indicating the 95% confidence interval (CI). The diamond represents the overall meta-analytic estimate across studies.

**rs10759942 T/C 9:120515918**

| Cohort | P | ln(OR) | SE |
|---|---|---|---|
| ADHEA_AFR | 0.4321 | 0.093 | 0.117 |
| ADHEA_EUR | 0.5906 | 0.038 | 0.07 |
| BEPS7_EUR | 0.7092 | −0.07 | 0.188 |
| BHRCM_AFR | 0.3731 | 0.171 | 0.193 |
| BHRCM_EUR | 0.562 | 0.094 | 0.162 |
| BHRCM_LAT | 0.1855 | −0.497 | 0.375 |
| BOR17_EUR | 0.6807 | 0.037 | 0.089 |
| BOR2C_EUR | 0.4101 | 0.074 | 0.09 |
| BOR2E_EUR | 0.713 | 0.067 | 0.182 |
| CNVRG_EAS | 0.7268 | 0.072 | 0.206 |
| COGA1_AFR | 0.0025 | 0.31 | 0.102 |
| COGA1_EUR | 0.2949 | 0.065 | 0.062 |
| CUINT_EUR | 0.514 | 0.069 | 0.105 |
| CVEDA_CSA | 0.4955 | 0.127 | 0.187 |
| ESTB2_EUR | 4e−04 | 0.085 | 0.024 |
| FINNG_EUR | 0.2279 | 0.029 | 0.024 |
| GEDIS_EUR | 0.7796 | 0.038 | 0.136 |
| GISS1_EUR | 0.8338 | −0.018 | 0.084 |
| GISS2_EUR | 0.1968 | 0.126 | 0.098 |
| GTPRJ_AFR | 0.1243 | 0.092 | 0.06 |
| IPSYC_EUR | 0.1084 | 0.033 | 0.021 |
| JANS3_EUR | 0.3065 | −0.116 | 0.113 |
| JANS4_EUR | 0.2676 | 0.134 | 0.121 |
| MIREC_EUR | 0.8037 | −0.039 | 0.158 |
| MVPXQ_AFR | 0.1598 | 0.031 | 0.022 |
| MVPXQ_EAS | 0.8232 | −0.05 | 0.224 |
| MVPXQ_EUR | 4e−04 | 0.047 | 0.013 |
| MVPXQ_LAT | 0.2843 | 0.037 | 0.035 |
| PGCBD_EUR | 0.9462 | 0.002 | 0.027 |
| PGCED_EUR | 0.0561 | −0.243 | 0.127 |
| PGCMD_EUR | 0.6722 | 0.015 | 0.035 |
| PGCPT_AFR | 0.6206 | −0.051 | 0.104 |
| PGCPT_EUR | 0.8958 | −0.008 | 0.06 |
| PGCSZ_EUR | 0.6246 | −0.019 | 0.039 |
| PRFCT_EUR | 0.8184 | −0.011 | 0.046 |
| PSYCR_EUR | 0.9349 | 0.008 | 0.092 |
| QIMRB_EUR | 0.0298 | 0.062 | 0.028 |
| STRR1_LAT | 0.6592 | 0.081 | 0.185 |
| UKBJC_EUR | 0.5779 | 0.012 | 0.021 |
| YPENN_EUR | 0.3954 | 0.112 | 0.132 |
| **meta** | **3.03e−08** | **0.036** | **0.006** |

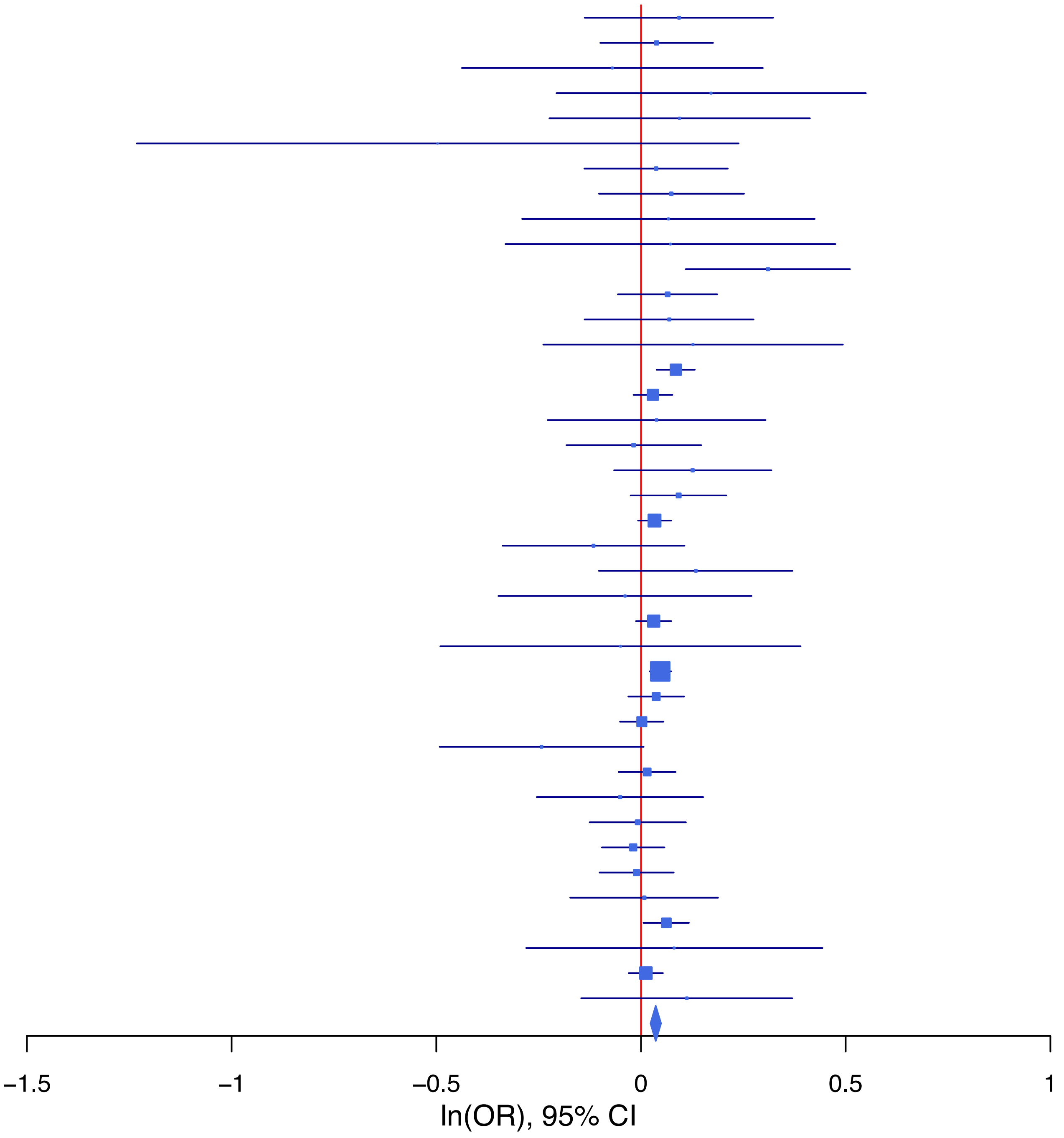

**rs10835368 T/C 11:28643299**

| Cohort | P | ln(OR) | SE |
|---|---|---|---|
| ALSPC_EUR | 0.8448 | −0.011 | 0.056 |
| BEPS7_EUR | 0.2723 | 0.194 | 0.176 |
| BHRCM_AFR | 0.4531 | 0.18 | 0.24 |
| BHRCM_EUR | 0.3065 | −0.17 | 0.167 |
| BHRCM_LAT | 0.0363 | −0.702 | 0.335 |
| BOR17_EUR | 0.6208 | 0.044 | 0.09 |
| BOR2C_EUR | 0.4136 | −0.073 | 0.089 |
| BOR2E_EUR | 0.004 | −0.554 | 0.193 |
| CNVRG_EAS | 0.3747 | −0.045 | 0.051 |
| COGA1_AFR | 0.4636 | −0.08 | 0.109 |
| COGA1_EUR | 0.937 | −0.005 | 0.06 |
| CUINT_EUR | 0.0987 | −0.18 | 0.109 |
| CVEDA_CSA | 0.6367 | −0.08 | 0.17 |
| ESTB2_EUR | 0.0643 | −0.043 | 0.023 |
| FINNG_EUR | 0.752 | 0.007 | 0.024 |
| GEDIS_EUR | 0.7667 | −0.041 | 0.138 |
| GISS1_EUR | 0.9374 | 0.006 | 0.079 |
| GISS2_EUR | 0.8025 | −0.023 | 0.093 |
| GREAT_EAS | 0.3331 | −0.103 | 0.106 |
| GTPRJ_AFR | 0.4078 | 0.057 | 0.068 |
| IPSYC_EUR | 0.9133 | 0.002 | 0.021 |
| JANS3_EUR | 0.456 | −0.081 | 0.108 |
| JANS4_EUR | 0.2779 | −0.134 | 0.124 |
| MVPXQ_AFR | 0.0144 | −0.057 | 0.023 |
| MVPXQ_EAS | 0.1436 | −0.178 | 0.122 |
| MVPXQ_EUR | 1e−04 | −0.05 | 0.013 |
| MVPXQ_LAT | 0.5006 | −0.023 | 0.034 |
| PGCBD_EUR | 0.074 | −0.048 | 0.027 |
| PGCED_EUR | 0.5728 | −0.066 | 0.117 |
| PGCMD_EUR | 0.0044 | −0.101 | 0.036 |
| PGCPT_AFR | 0.6549 | 0.052 | 0.115 |
| PGCPT_EUR | 0.3135 | 0.06 | 0.06 |
| PGCSZ_EUR | 0.2504 | −0.044 | 0.038 |
| PRFCT_EUR | 0.3877 | −0.04 | 0.046 |
| PSYCR_EUR | 0.062 | −0.171 | 0.092 |
| QIMRB_EUR | 0.0073 | −0.075 | 0.028 |
| SNUBH-ASA_EAS | 0.5625 | 0.081 | 0.14 |
| SNUBH-KCHIP_EAS | 0.1914 | −0.235 | 0.18 |
| STRR1_LAT | 0.8458 | 0.036 | 0.183 |
| UKBJC_EUR | 0.0999 | −0.034 | 0.021 |
| YPENN_EUR | 0.9898 | 0.002 | 0.132 |
| **meta** | **1.20e−09** | **−0.039** | **0.006** |

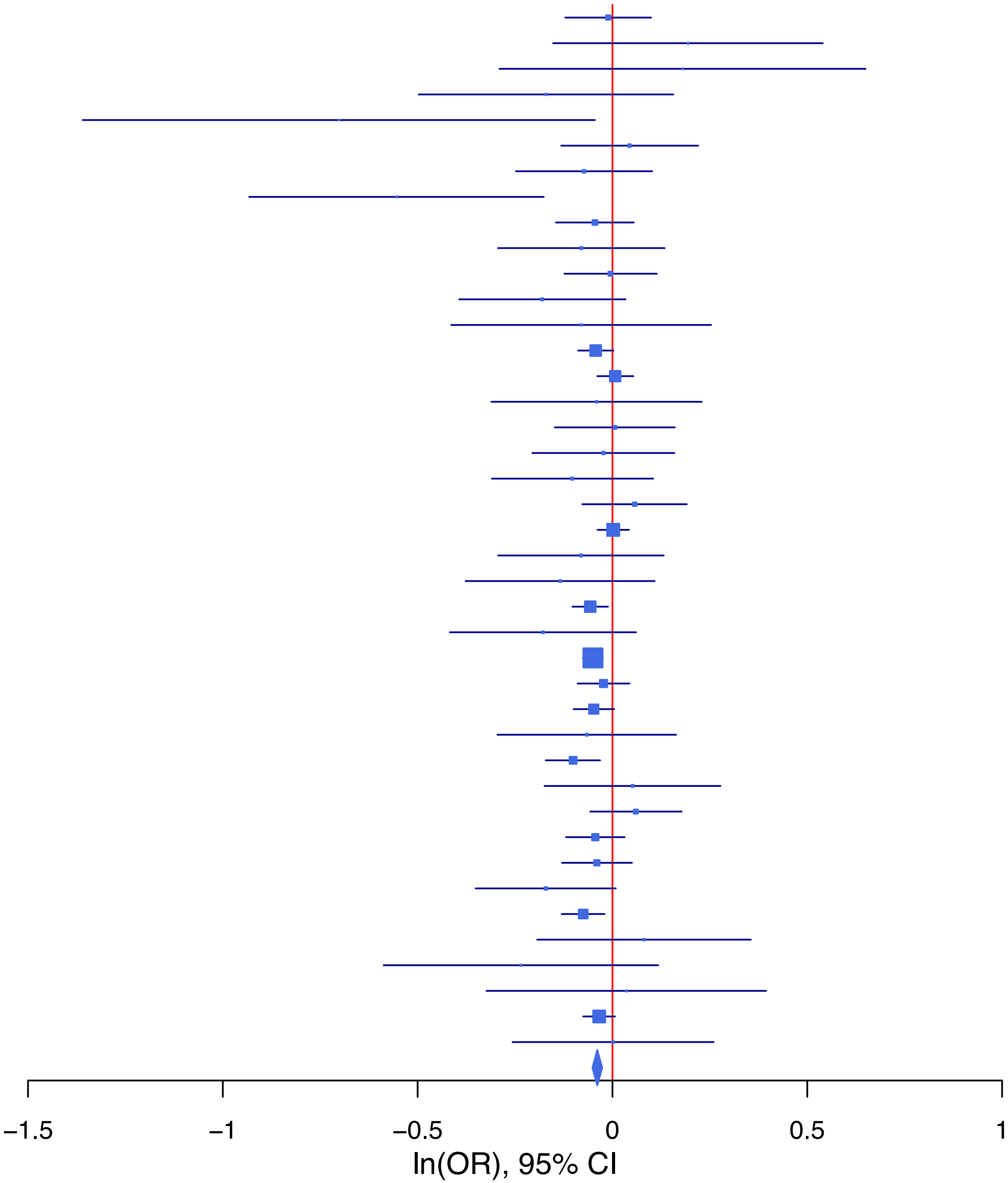

**rs10890034 A/G 1:73824279**

| Cohort | P | ln(OR) | SE |
|---|---|---|---|
| ADHEA_AFR | 0.2419 | −0.134 | 0.114 |
| ADHEA_EUR | 0.5094 | −0.045 | 0.069 |
| BEPS7_EUR | 0.8635 | −0.031 | 0.179 |
| BHRCM_AFR | 0.4428 | 0.148 | 0.193 |
| BHRCM_EUR | 0.0226 | −0.364 | 0.16 |
| BHRCM_LAT | 0.6177 | −0.165 | 0.331 |
| BOR17_EUR | 0.8765 | 0.013 | 0.085 |
| BOR2C_EUR | 0.3753 | 0.076 | 0.086 |
| BOR2E_EUR | 0.6122 | −0.09 | 0.178 |
| CNVRG_EAS | 0.3132 | 0.057 | 0.056 |
| COGA1_AFR | 0.4412 | −0.076 | 0.099 |
| COGA1_EUR | 0.175 | −0.082 | 0.06 |
| CUINT_EUR | 0.2906 | −0.108 | 0.102 |
| CVEDA_CSA | 0.8122 | −0.039 | 0.163 |
| ESTB2_EUR | 0.6305 | −0.011 | 0.024 |
| FINNG_EUR | 0.2309 | −0.028 | 0.024 |
| GEDIS_EUR | 0.0059 | −0.363 | 0.132 |
| GISS1_EUR | 0.9362 | 0.006 | 0.08 |
| GISS2_EUR | 0.6311 | −0.045 | 0.093 |
| GREAT_EAS | 0.1538 | −0.145 | 0.102 |
| GTPRJ_AFR | 0.1194 | −0.091 | 0.059 |
| IPSYC_EUR | 0.0931 | −0.033 | 0.02 |
| JANS3_EUR | 0.0964 | 0.18 | 0.108 |
| JANS4_EUR | 0.0697 | 0.225 | 0.124 |
| MIREC_AFR | 0.4886 | −0.097 | 0.14 |
| MIREC_EUR | 0.8527 | 0.028 | 0.152 |
| MVPXQ_AFR | 0.0721 | −0.037 | 0.02 |
| MVPXQ_EAS | 0.7292 | 0.038 | 0.11 |
| MVPXQ_EUR | 1e−04 | −0.051 | 0.013 |
| PGCBD_EUR | 0.9461 | −0.002 | 0.026 |
| PGCED_EUR | 0.3244 | −0.114 | 0.115 |
| PGCMD_EUR | 0.1532 | −0.049 | 0.034 |
| PGCPT_AFR | 0.539 | −0.062 | 0.102 |
| PGCPT_EUR | 0.4994 | 0.04 | 0.059 |
| PGCSZ_EUR | 0.323 | −0.037 | 0.038 |
| PRFCT_EUR | 0.1292 | −0.067 | 0.044 |
| PSYCR_EUR | 0.2045 | 0.116 | 0.091 |
| QIMRB_EUR | 0.1013 | −0.046 | 0.028 |
| SNUBH−ASA_EAS | 0.498 | −0.099 | 0.145 |
| SNUBH−KCHIP_EAS | 0.8552 | 0.037 | 0.205 |
| STRR1_LAT | 0.809 | 0.043 | 0.177 |
| UKBJC_EUR | 8e−04 | −0.069 | 0.021 |
| YPENN_EUR | 0.3911 | −0.112 | 0.13 |
| **meta** | **1.27e−09** | **−0.038** | **0.006** |

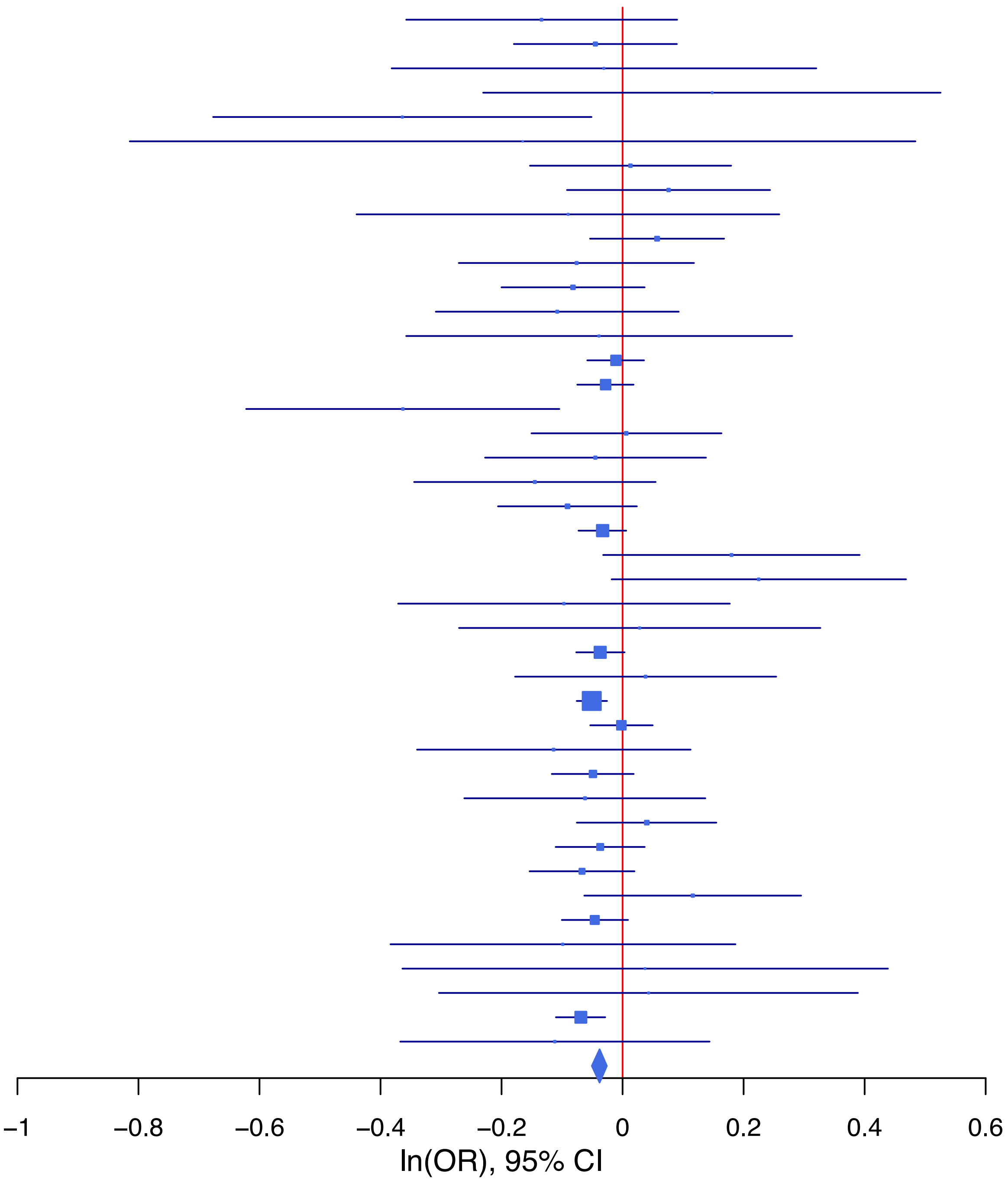

**rs10891549 T/C 11:113278447**

| Cohort | P | ln(OR) | SE |
|---|---|---|---|
| ALSPC_EUR | 0.6629 | 0.023 | 0.054 |
| BEPS7_EUR | 0.7576 | 0.055 | 0.177 |
| BHRCM_AFR | 0.6836 | 0.14 | 0.344 |
| BHRCM_EUR | 0.5741 | −0.094 | 0.167 |
| BOR17_EUR | 0.8463 | −0.016 | 0.085 |
| BOR2C_EUR | 0.5024 | 0.059 | 0.088 |
| BOR2E_EUR | 0.0026 | −0.566 | 0.188 |
| CNVRG_EAS | 0.6029 | 0.052 | 0.1 |
| COGA1_AFR | 0.2428 | 0.152 | 0.13 |
| COGA1_EUR | 0.9268 | −0.005 | 0.059 |
| CUINT_EUR | 0.0268 | 0.221 | 0.1 |
| CVEDA_CSA | 0.1186 | 0.283 | 0.181 |
| ESTB2_EUR | 0.0513 | 0.044 | 0.023 |
| FINNG_EUR | 0.0937 | 0.038 | 0.023 |
| GEDIS_EUR | 0.1502 | 0.188 | 0.131 |
| GISS1_EUR | 0.2758 | 0.085 | 0.078 |
| GISS2_EUR | 0.6356 | −0.043 | 0.09 |
| GREAT_EAS | 0.5283 | 0.129 | 0.205 |
| GTPRJ_AFR | 0.4814 | 0.058 | 0.082 |
| IPSYC_EUR | 0.0098 | 0.051 | 0.02 |
| JANS3_EUR | 0.0773 | 0.181 | 0.103 |
| JANS4_EUR | 0.292 | −0.12 | 0.114 |
| MIREC_AFR | 0.9815 | −0.004 | 0.189 |
| MIREC_EUR | 0.5472 | 0.092 | 0.152 |
| MVPXQ_AFR | 0.0562 | 0.053 | 0.028 |
| MVPXQ_EAS | 0.3218 | −0.153 | 0.154 |
| MVPXQ_EUR | 5e−04 | 0.044 | 0.013 |
| MVPXQ_LAT | 0.5027 | 0.026 | 0.039 |
| PGCBD_EUR | 0.6511 | 0.012 | 0.026 |
| PGCED_EUR | 0.7676 | 0.033 | 0.113 |
| PGCMD_EUR | 0.0441 | 0.069 | 0.034 |
| PGCPT_AFR | 0.4673 | 0.12 | 0.165 |
| PGCPT_EUR | 0.6906 | −0.023 | 0.058 |
| PGCSZ_EUR | 0.5795 | 0.02 | 0.037 |
| PRFCT_EUR | 0.3538 | 0.04 | 0.043 |
| PSYCR_EUR | 0.8133 | 0.02 | 0.087 |
| QIMRB_EUR | 0.9112 | 0.003 | 0.027 |
| SNUBH−ASA_EAS | 0.3829 | −0.236 | 0.27 |
| STRR1_LAT | 0.9604 | −0.009 | 0.189 |
| UKBJC_EUR | 0.0066 | 0.055 | 0.02 |
| YPENN_AFR | 0.6352 | 0.091 | 0.191 |
| YPENN_EUR | 0.847 | −0.024 | 0.125 |
| **meta** | **5.22e−10** | **0.039** | **0.006** |

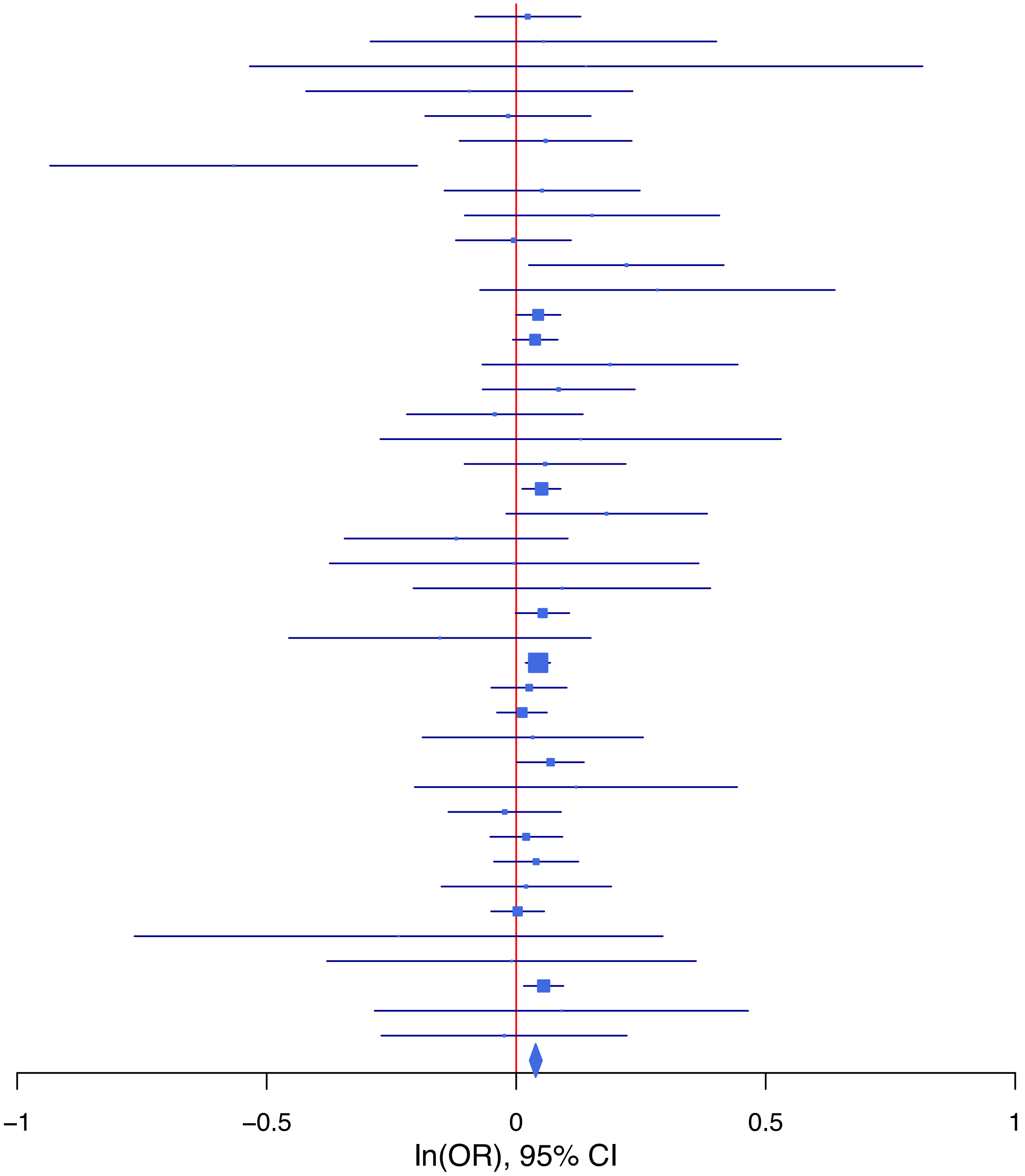

**rs1113737   A/G   7:1993078**

| Cohort | P | ln(OR) | SE |
|---|---|---|---|
| ADHEA_AFR | 0.6623 | 0.068 | 0.154 |
| ADHEA_EUR | 0.0781 | 0.126 | 0.072 |
| ALSPC_EUR | 0.3205 | 0.06 | 0.061 |
| BEPS7_EUR | 0.0729 | 0.333 | 0.186 |
| BHRCM_AFR | 0.2654 | −0.421 | 0.378 |
| BHRCM_EUR | 0.8866 | −0.025 | 0.175 |
| BOR17_EUR | 0.8724 | −0.015 | 0.091 |
| BOR2C_EUR | 0.1798 | 0.122 | 0.091 |
| BOR2E_EUR | 0.759 | −0.061 | 0.198 |
| CNVRG_EAS | 0.6674 | 0.021 | 0.048 |
| COGA1_AFR | 0.8057 | 0.035 | 0.142 |
| COGA1_EUR | 0.766 | 0.019 | 0.063 |
| CUINT_EUR | 0.7428 | −0.036 | 0.108 |
| CVEDA_CSA | 0.6593 | 0.088 | 0.2 |
| ESTB2_EUR | 0.8247 | 0.005 | 0.025 |
| FINNG_EUR | 0.075 | 0.044 | 0.024 |
| GEDIS_EUR | 0.5061 | 0.09 | 0.135 |
| GISS1_EUR | 0.7049 | −0.032 | 0.084 |
| GISS2_EUR | 0.8481 | 0.019 | 0.098 |
| GREAT_EAS | 0.1576 | 0.135 | 0.095 |
| GTPRJ_AFR | 0.1675 | −0.121 | 0.088 |
| IPSYC_EUR | 0.0635 | 0.04 | 0.022 |
| JANS3_EUR | 0.5766 | 0.064 | 0.115 |
| JANS4_EUR | 0.0472 | 0.245 | 0.124 |
| MVPXQ_AFR | 0.9722 | 0.001 | 0.029 |
| MVPXQ_EAS | 0.9845 | 0.002 | 0.104 |
| MVPXQ_EUR | 0.0131 | 0.035 | 0.014 |
| MVPXQ_LAT | 0.0586 | 0.079 | 0.042 |
| PGCBD_EUR | 0.0872 | 0.048 | 0.028 |
| PGCED_EUR | 0.5785 | −0.074 | 0.133 |
| PGCMD_EUR | 0.3326 | −0.036 | 0.038 |
| PGCPT_AFR | 0.5055 | 0.099 | 0.148 |
| PGCPT_EUR | 0.4513 | 0.046 | 0.061 |
| PGCSZ_EUR | 0.1731 | 0.055 | 0.04 |
| PRFCT_EUR | 0.9609 | 0.002 | 0.047 |
| PSYCR_EUR | 0.5746 | 0.052 | 0.092 |
| QIMRB_EUR | 0.1434 | 0.043 | 0.029 |
| SNUBH−ASA_EAS | 0.5457 | −0.078 | 0.128 |
| SNUBH−KCHIP_EAS | 0.1567 | 0.255 | 0.18 |
| STRR1_LAT | 0.2681 | 0.225 | 0.203 |
| UKBJC_EUR | 1e−04 | 0.083 | 0.022 |
| YPENN_EUR | 0.3288 | 0.133 | 0.136 |
| **meta** | **2.44e−08** | **0.038** | **0.007** |

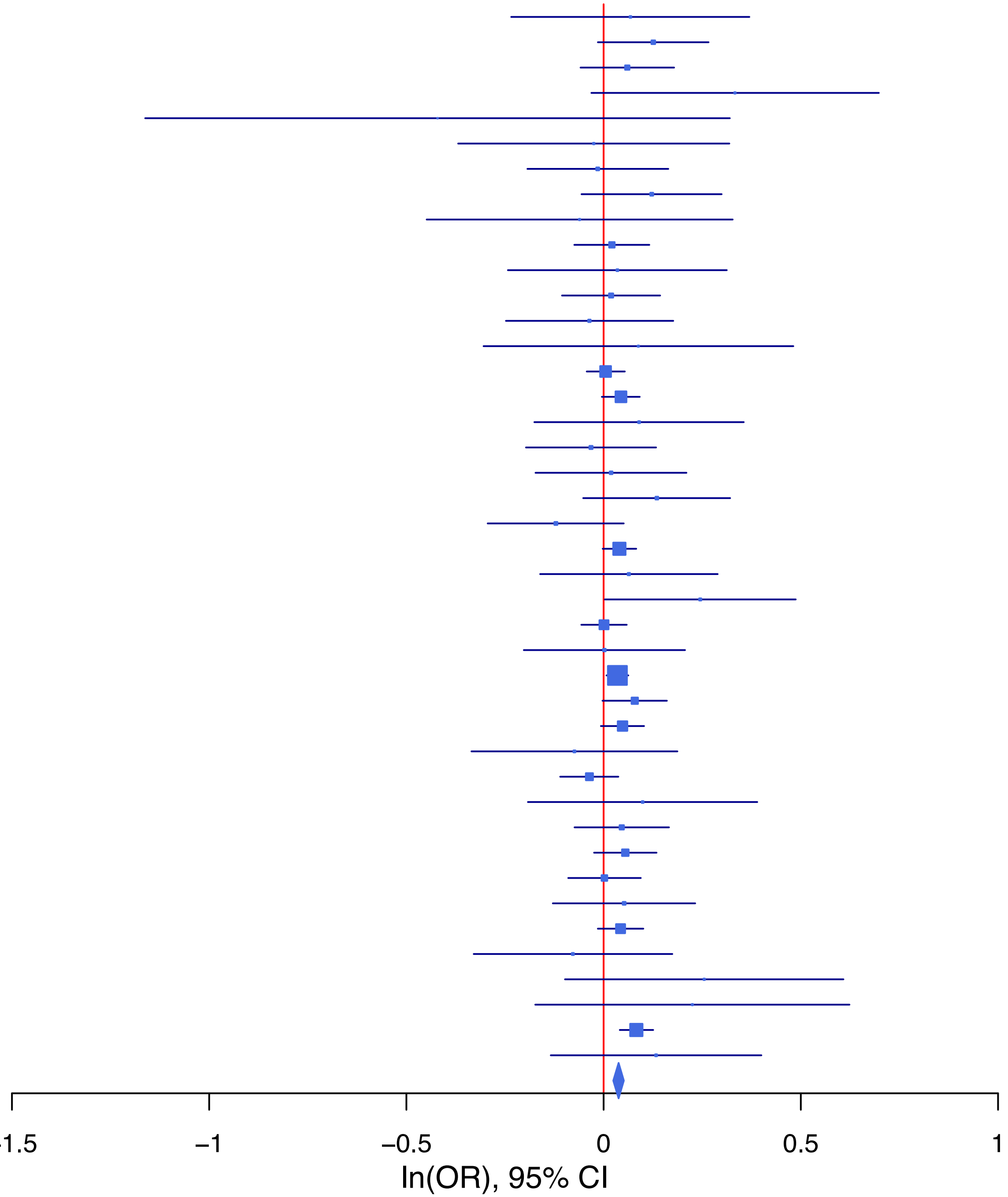

**rs11233675 C/T 11:83274215**

| Cohort | P | ln(OR) | SE |
|---|---|---|---|
| ADHEA_AFR | 0.1356 | 0.258 | 0.172 |
| ADHEA_EUR | 0.6787 | −0.069 | 0.168 |
| ALSPC_EUR | 0.5511 | 0.079 | 0.132 |
| BHRCM_AFR | 0.305 | 0.26 | 0.253 |
| BHRCM_EUR | 0.7689 | 0.077 | 0.263 |
| BHRCM_LAT | 0.2802 | 0.369 | 0.342 |
| BOR17_EUR | 0.691 | 0.088 | 0.222 |
| BOR2C_EUR | 0.2031 | 0.252 | 0.198 |
| BOR2E_EUR | 0.0717 | 0.739 | 0.41 |
| CNVRG_EAS | 0.2482 | 0.095 | 0.082 |
| COGA1_AFR | 0.73 | 0.067 | 0.194 |
| COGA1_EUR | 0.1648 | 0.197 | 0.142 |
| CUINT_EUR | 0.1301 | −0.353 | 0.233 |
| CVEDA_CSA | 0.8363 | 0.048 | 0.232 |
| ESTB2_EUR | 0.2615 | 0.073 | 0.065 |
| FINNG_EUR | 0.1576 | 0.099 | 0.07 |
| GEDIS_EUR | 0.2091 | 0.365 | 0.291 |
| GISS1_EUR | 0.3134 | 0.185 | 0.184 |
| GISS2_EUR | 0.4689 | −0.16 | 0.222 |
| GREAT_EAS | 0.5219 | −0.091 | 0.143 |
| GTPRJ_AFR | 0.5948 | 0.052 | 0.097 |
| IPSYC_EUR | 0.0327 | 0.102 | 0.048 |
| JANS3_EUR | 0.7196 | −0.088 | 0.246 |
| JANS4_EUR | 0.7399 | −0.086 | 0.26 |
| MVPXQ_AFR | 0.0663 | 0.065 | 0.035 |
| MVPXQ_EAS | 0.8685 | −0.029 | 0.175 |
| MVPXQ_LAT | 0.5806 | 0.037 | 0.068 |
| PGCBD_EUR | 0.2016 | 0.096 | 0.075 |
| PGCMD_EUR | 0.0091 | 0.238 | 0.091 |
| PGCPT_AFR | 0.1179 | 0.245 | 0.157 |
| PGCPT_EUR | 0.6852 | 0.056 | 0.138 |
| PGCSZ_EUR | 0.0975 | 0.163 | 0.098 |
| PRFCT_EUR | 0.7187 | −0.047 | 0.13 |
| PSYCR_EUR | 0.1999 | 0.252 | 0.196 |
| QIMRB_EUR | 0.0193 | 0.162 | 0.069 |
| SNUBH−ASA_EAS | 0.0909 | 0.441 | 0.261 |
| SNUBH−KCHIP_EAS | 0.8987 | −0.043 | 0.341 |
| UKBJC_EUR | 0.4072 | 0.044 | 0.053 |
| YPENN_AFR | 0.3468 | 0.24 | 0.255 |
| YPENN_EUR | 0.4197 | 0.226 | 0.279 |
| **meta** | **4.78e−08** | **0.088** | **0.016** |

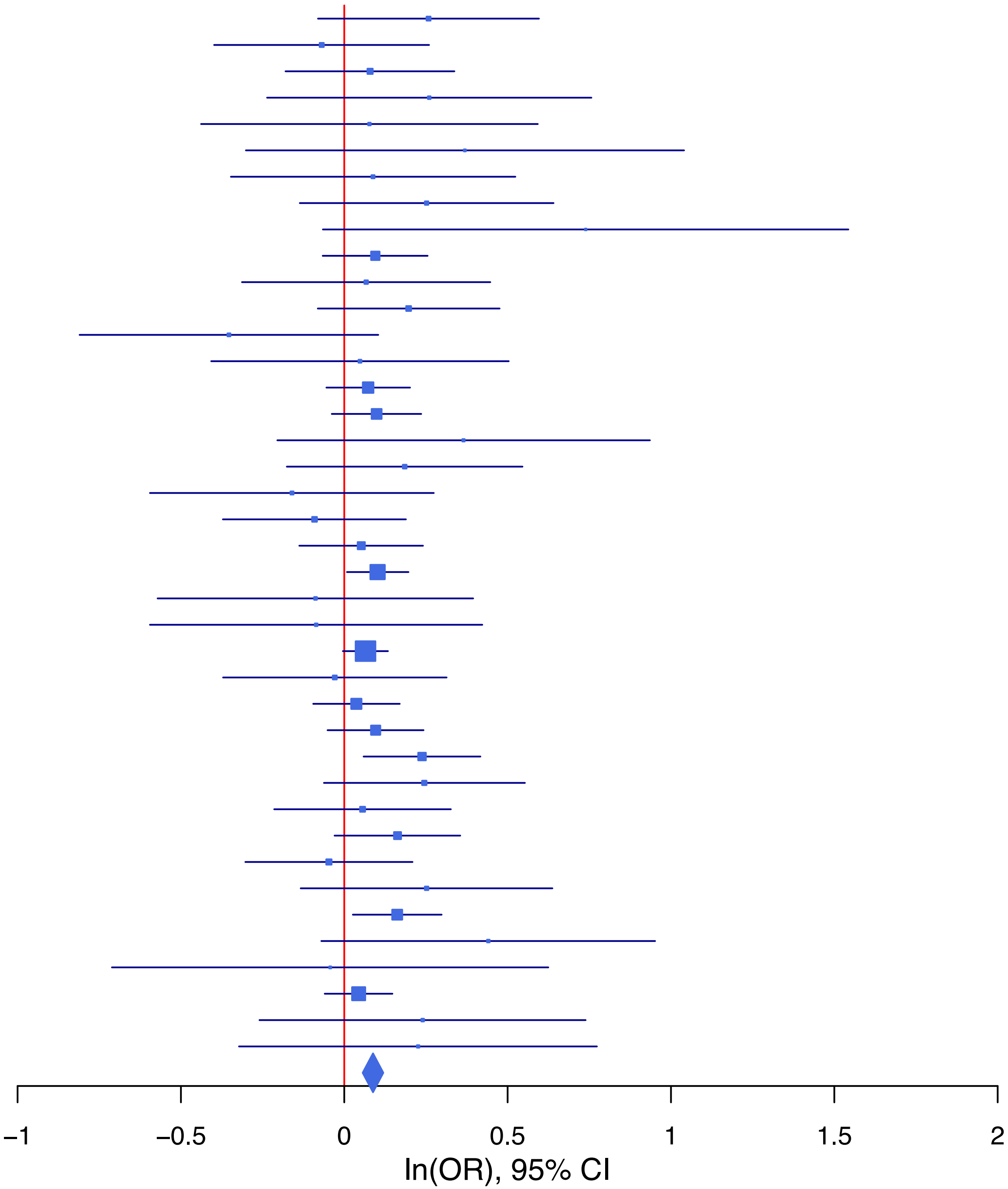

**rs114470015   A/T   5:92520938**

| Cohort | P | ln(OR) | SE |
|---|---|---|---|
| ADHEA_AFR | 0.3075 | 0.123 | 0.121 |
| ADHEA_EUR | 0.7372 | 0.035 | 0.105 |
| ALSPC_EUR | 0.444 | −0.062 | 0.081 |
| BEPS7_EUR | 0.1425 | −0.505 | 0.344 |
| BHRCM_AFR | 0.3264 | −0.21 | 0.214 |
| BHRCM_EUR | 0.8404 | 0.044 | 0.219 |
| BOR17_EUR | 0.3101 | 0.134 | 0.132 |
| BOR2C_EUR | 0.7543 | −0.044 | 0.142 |
| BOR2E_EUR | 0.862 | −0.044 | 0.255 |
| COGA1_AFR | 0.4631 | −0.083 | 0.113 |
| COGA1_EUR | 0.0053 | −0.245 | 0.088 |
| CUINT_EUR | 0.3121 | −0.165 | 0.164 |
| ESTB2_EUR | 0.469 | −0.028 | 0.038 |
| GEDIS_EUR | 0.5011 | 0.13 | 0.194 |
| GISS1_EUR | 0.8797 | −0.023 | 0.151 |
| GISS2_EUR | 0.3576 | 0.165 | 0.179 |
| GTPRJ_AFR | 0.5868 | 0.035 | 0.064 |
| IPSYC_EUR | 0.0061 | −0.083 | 0.03 |
| JANS3_EUR | 0.213 | −0.207 | 0.166 |
| JANS4_EUR | 0.5271 | −0.127 | 0.201 |
| MIREC_AFR | 0.6449 | −0.069 | 0.149 |
| MIREC_EUR | 0.5431 | 0.137 | 0.226 |
| MVPXQ_AFR | 0.004 | −0.069 | 0.024 |
| MVPXQ_EUR | 0.1112 | −0.031 | 0.019 |
| MVPXQ_LAT | 0.7473 | −0.017 | 0.052 |
| PGCBD_EUR | 0.0031 | −0.121 | 0.041 |
| PGCED_EUR | 0.004 | −0.602 | 0.209 |
| PGCMD_EUR | 0.0538 | −0.111 | 0.058 |
| PGCPT_AFR | 0.0823 | −0.198 | 0.114 |
| PGCPT_EUR | 0.0313 | −0.199 | 0.093 |
| PGCSZ_EUR | 0.06 | −0.11 | 0.058 |
| PRFCT_EUR | 0.1308 | −0.108 | 0.072 |
| PSYCR_EUR | 0.8127 | −0.034 | 0.142 |
| QIMRB_EUR | 0.3227 | −0.041 | 0.042 |
| STRR1_LAT | 0.0984 | −0.608 | 0.368 |
| UKBJC_EUR | 0.3075 | −0.03 | 0.03 |
| YPENN_AFR | 0.8459 | −0.032 | 0.164 |
| YPENN_EUR | 0.9964 | 0.001 | 0.198 |
| **meta** | **1.92e−09** | **−0.057** | **0.01** |

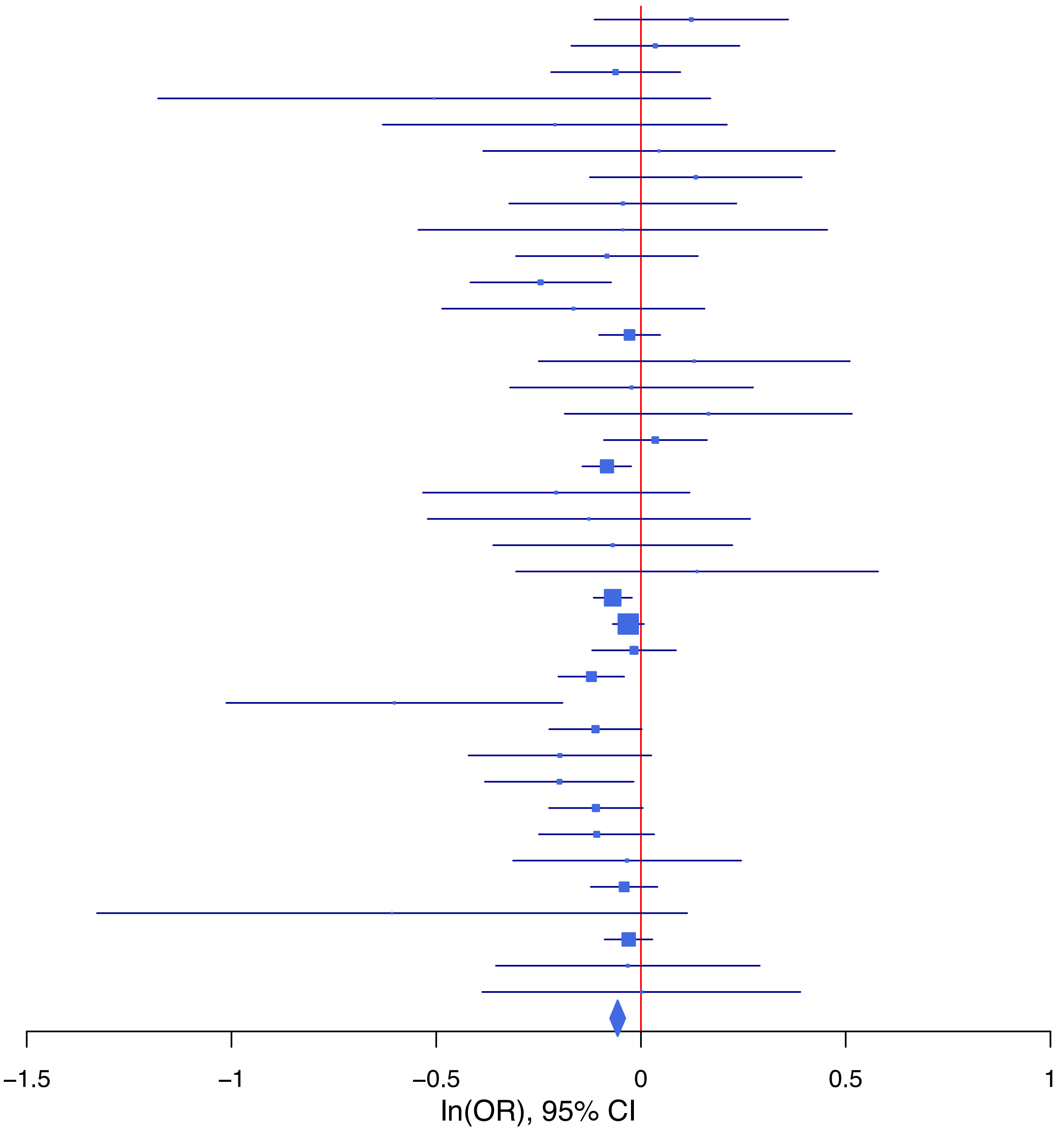

**rs11958069 A/G 5:153435497**

| Cohort | P | ln(OR) | SE |
|---|---|---|---|
| ADHEA_AFR | 0.5912 | 0.09 | 0.166 |
| ADHEA_EUR | 0.0653 | 0.186 | 0.101 |
| ALSPC_EUR | 0.291 | 0.086 | 0.081 |
| BEPS7_EUR | 0.2963 | 0.289 | 0.277 |
| BHRCM_AFR | 0.5412 | 0.154 | 0.253 |
| BHRCM_EUR | 0.1179 | 0.322 | 0.206 |
| BOR17_EUR | 0.2446 | 0.155 | 0.134 |
| BOR2C_EUR | 0.0834 | 0.229 | 0.132 |
| BOR2E_EUR | 0.2969 | 0.274 | 0.263 |
| CNVRG_EAS | 0.51 | −0.091 | 0.139 |
| COGA1_AFR | 0.4933 | 0.097 | 0.141 |
| COGA1_EUR | 0.143 | 0.134 | 0.091 |
| CUINT_EUR | 0.6536 | −0.07 | 0.156 |
| ESTB2_EUR | 0.5565 | 0.02 | 0.034 |
| FINNG_EUR | 0.0055 | 0.09 | 0.032 |
| GEDIS_EUR | 0.8127 | −0.048 | 0.2 |
| GISS1_EUR | 0.3569 | −0.113 | 0.123 |
| GISS2_EUR | 0.9276 | −0.014 | 0.149 |
| GTPRJ_AFR | 0.1827 | 0.103 | 0.078 |
| IPSYC_EUR | 0.0116 | 0.075 | 0.03 |
| JANS3_EUR | 0.5243 | −0.101 | 0.159 |
| JANS4_EUR | 0.207 | 0.213 | 0.169 |
| MIREC_AFR | 0.0953 | −0.35 | 0.21 |
| MIREC_EUR | 0.2394 | 0.248 | 0.211 |
| MVPXQ_EUR | 0.0103 | 0.049 | 0.019 |
| MVPXQ_LAT | 0.1699 | 0.077 | 0.056 |
| PGCBD_EUR | 0.7678 | 0.012 | 0.04 |
| PGCED_EUR | 0.3904 | 0.161 | 0.187 |
| PGCMD_EUR | 0.2751 | 0.062 | 0.057 |
| PGCPT_AFR | 0.4965 | 0.111 | 0.163 |
| PGCPT_EUR | 0.1339 | 0.127 | 0.085 |
| PGCSZ_EUR | 0.6667 | 0.025 | 0.058 |
| PRFCT_EUR | 0.6111 | 0.034 | 0.067 |
| PSYCR_EUR | 0.8036 | 0.035 | 0.141 |
| QIMRB_EUR | 0.553 | 0.024 | 0.041 |
| SNUBH−ASA_EAS | 0.6973 | 0.134 | 0.346 |
| STRR1_LAT | 0.2779 | 0.323 | 0.298 |
| UKBJC_EUR | 0.2143 | 0.038 | 0.031 |
| YPENN_AFR | 0.2944 | −0.228 | 0.218 |
| YPENN_EUR | 0.1464 | 0.257 | 0.177 |
| **meta** | **2.21e−08** | **0.054** | **0.01** |

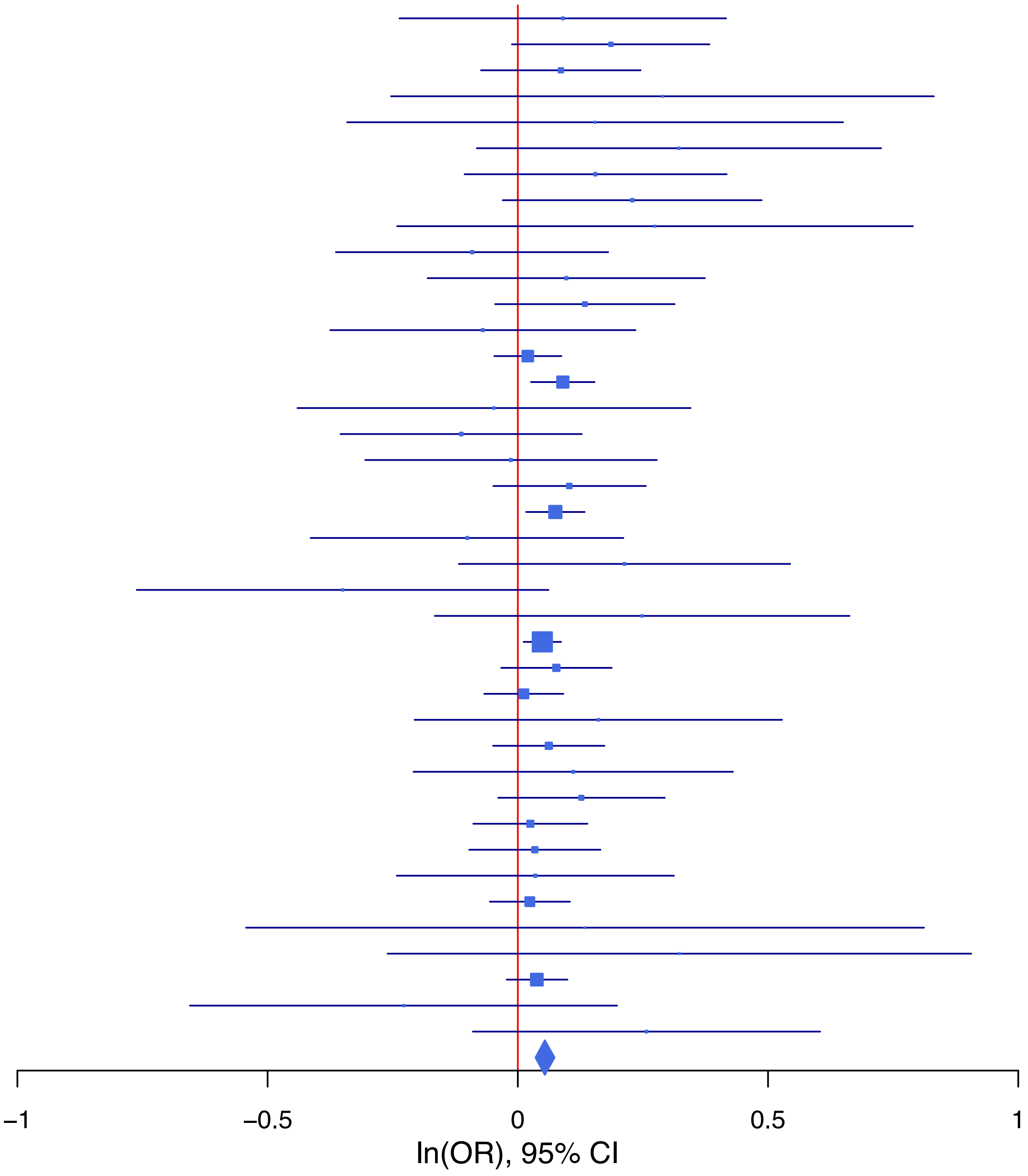

**rs12665582 C/T 6:156439031**

| Cohort | P | ln(OR) | SE |
|---|---|---|---|
| ADHEA_AFR | 0.7666 | 0.039 | 0.132 |
| ADHEA_EUR | 0.8029 | −0.028 | 0.113 |
| ALSPC_EUR | 0.6536 | 0.04 | 0.088 |
| BEPS7_EUR | 0.9973 | −0.001 | 0.298 |
| BHRCM_AFR | 0.4573 | 0.153 | 0.206 |
| BHRCM_EUR | 0.3352 | 0.223 | 0.232 |
| BOR17_EUR | 0.1536 | −0.213 | 0.149 |
| BOR2C_EUR | 0.9094 | −0.017 | 0.148 |
| BOR2E_EUR | 0.5525 | −0.178 | 0.3 |
| CNVRG_EAS | 0.8326 | 0.01 | 0.048 |
| COGA1_AFR | 0.2202 | 0.145 | 0.118 |
| COGA1_EUR | 0.0608 | 0.179 | 0.096 |
| CUINT_EUR | 0.5416 | 0.103 | 0.168 |
| CVEDA_CSA | 0.1984 | −0.378 | 0.294 |
| ESTB2_EUR | 0.8505 | 0.008 | 0.043 |
| FINNG_EUR | 0.0956 | 0.081 | 0.049 |
| GEDIS_EUR | 0.2996 | −0.256 | 0.247 |
| GISS1_EUR | 0.096 | −0.214 | 0.129 |
| GISS2_EUR | 0.7238 | −0.055 | 0.155 |
| GREAT_EAS | 0.2851 | 0.084 | 0.079 |
| GTPRJ_AFR | 0.5987 | 0.035 | 0.068 |
| IPSYC_EUR | 0.3267 | 0.033 | 0.034 |
| JANS3_EUR | 0.0457 | 0.318 | 0.159 |
| JANS4_EUR | 0.969 | −0.008 | 0.197 |
| MVPXQ_EUR | 0.0018 | 0.073 | 0.023 |
| PGCBD_EUR | 4.70e−05 | 0.177 | 0.044 |
| PGCED_EUR | 0.4966 | 0.137 | 0.201 |
| PGCMD_EUR | 0.7051 | 0.024 | 0.062 |
| PGCPT_AFR | 0.2423 | 0.138 | 0.118 |
| PGCPT_EUR | 0.4921 | 0.065 | 0.094 |
| PGCSZ_EUR | 0.8006 | 0.016 | 0.064 |
| PRFCT_EUR | 0.0371 | 0.155 | 0.074 |
| PSYCR_EUR | 0.6277 | −0.071 | 0.146 |
| QIMRB_EUR | 0.4203 | 0.036 | 0.045 |
| SNUBH−ASA_EAS | 0.9167 | −0.014 | 0.133 |
| SNUBH−KCHIP_EAS | 0.2876 | 0.189 | 0.178 |
| STRR1_LAT | 0.3966 | −0.183 | 0.216 |
| UKBJC_EUR | 0.0022 | 0.103 | 0.034 |
| YPENN_AFR | 0.2423 | 0.192 | 0.164 |
| YPENN_EUR | 0.1783 | 0.263 | 0.196 |
| **meta** | **1.05e−08** | **0.063** | **0.011** |

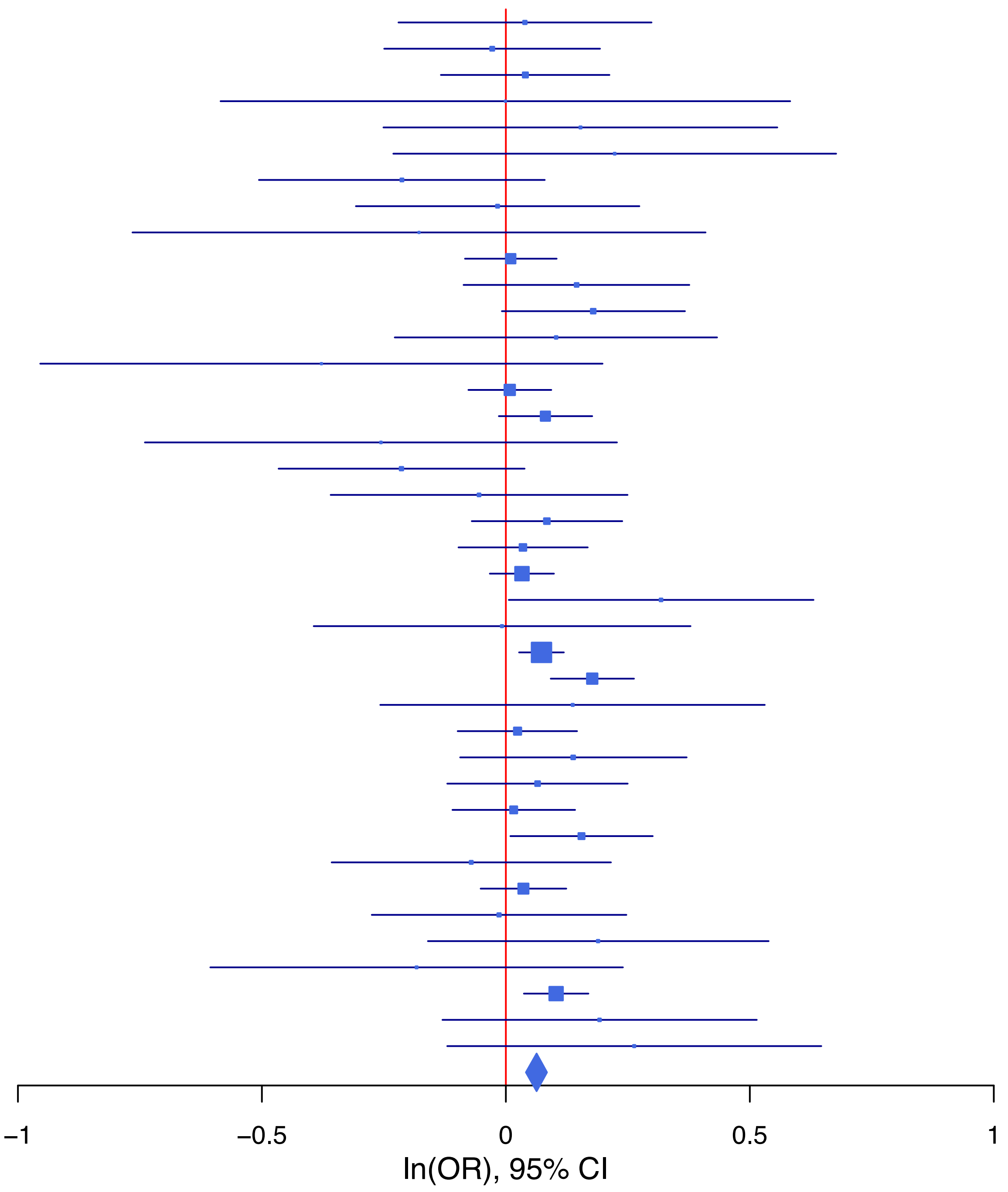

**rs12666306 G/A 7:115082406**

| Cohort | P | ln(OR) | SE |
|---|---|---|---|
| ADHEA_AFR | 0.6546 | −0.062 | 0.138 |
| ADHEA_EUR | 0.8373 | −0.014 | 0.068 |
| ALSPC_EUR | 0.3336 | −0.052 | 0.054 |
| BEPS7_EUR | 0.3269 | −0.168 | 0.171 |
| BHRCM_AFR | 0.8577 | −0.054 | 0.299 |
| BHRCM_EUR | 0.189 | −0.209 | 0.159 |
| BHRCM_LAT | 0.4167 | −0.222 | 0.273 |
| BOR17_EUR | 0.6436 | 0.039 | 0.084 |
| BOR2C_EUR | 0.4588 | 0.063 | 0.085 |
| BOR2E_EUR | 0.2291 | −0.215 | 0.179 |
| CNVRG_EAS | 0.9044 | 0.006 | 0.046 |
| COGA1_AFR | 0.8957 | −0.016 | 0.123 |
| COGA1_EUR | 0.8843 | −0.009 | 0.059 |
| CUINT_EUR | 0.377 | −0.088 | 0.1 |
| CVEDA_CSA | 0.0655 | −0.297 | 0.161 |
| ESTB2_EUR | 0.0025 | −0.069 | 0.023 |
| FINNG_EUR | 0.1191 | −0.036 | 0.023 |
| GEDIS_EUR | 0.0023 | −0.401 | 0.131 |
| GISS1_EUR | 0.4832 | −0.054 | 0.078 |
| GISS2_EUR | 0.8279 | 0.02 | 0.092 |
| GREAT_EAS | 0.6684 | 0.037 | 0.087 |
| GTPRJ_AFR | 0.9007 | −0.009 | 0.075 |
| IPSYC_EUR | 0.0136 | −0.05 | 0.02 |
| JANS3_EUR | 0.9763 | −0.003 | 0.104 |
| JANS4_EUR | 0.1695 | −0.164 | 0.119 |
| MIREC_AFR | 0.8787 | −0.026 | 0.172 |
| MIREC_EUR | 0.1038 | 0.248 | 0.153 |
| PGCBD_EUR | 0.0081 | −0.068 | 0.026 |
| PGCED_EUR | 0.525 | 0.072 | 0.114 |
| PGCMD_EUR | 0.0345 | −0.072 | 0.034 |
| PGCPT_AFR | 0.9105 | −0.015 | 0.131 |
| PGCPT_EUR | 0.4953 | −0.039 | 0.057 |
| PGCSZ_EUR | 0.2807 | −0.039 | 0.037 |
| PRFCT_EUR | 0.4794 | −0.031 | 0.044 |
| PSYCR_EUR | 0.1969 | 0.111 | 0.086 |
| QIMRB_EUR | 6e−04 | −0.094 | 0.027 |
| SNUBH−ASA_EAS | 0.7981 | 0.032 | 0.123 |
| SNUBH−KCHIP_EAS | 0.4231 | −0.132 | 0.165 |
| STRR1_LAT | 0.9538 | 0.01 | 0.174 |
| UKBJC_EUR | 0.0681 | −0.037 | 0.02 |
| YPENN_AFR | 0.3912 | −0.161 | 0.188 |
| YPENN_EUR | 0.9232 | −0.012 | 0.127 |
| **meta** | **2.48e−10** | **−0.048** | **0.008** |

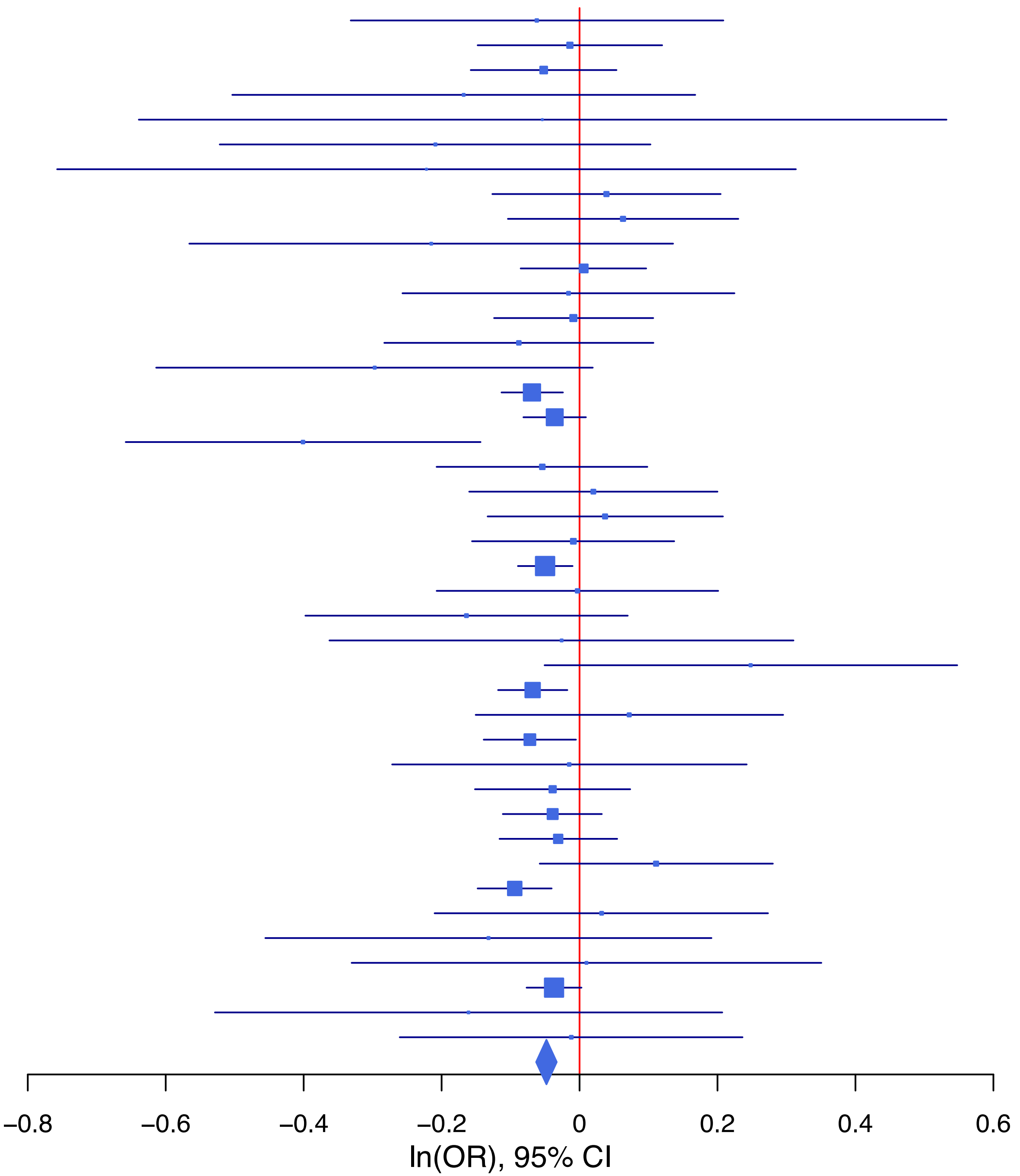

**rs12715437 T/C 3:49751856**

| Cohort | P | ln(OR) | SE |
|---|---|---|---|
| ADHEA_AFR | 0.9869 | −0.002 | 0.13 |
| ADHEA_EUR | 0.2272 | −0.107 | 0.089 |
| ALSPC_EUR | 0.8086 | 0.016 | 0.067 |
| BEPS7_EUR | 0.456 | 0.167 | 0.224 |
| BHRCM_AFR | 0.2049 | 0.248 | 0.196 |
| BHRCM_EUR | 0.6114 | 0.099 | 0.194 |
| BOR17_EUR | 0.2523 | 0.127 | 0.111 |
| BOR2C_EUR | 0.0667 | 0.208 | 0.113 |
| BOR2E_EUR | 0.1276 | 0.349 | 0.229 |
| COGA1_AFR | 0.35 | 0.106 | 0.114 |
| COGA1_EUR | 0.0559 | 0.144 | 0.075 |
| CUINT_EUR | 0.8749 | 0.02 | 0.124 |
| ESTB2_EUR | 0.5115 | 0.022 | 0.034 |
| FINNG_EUR | 0.3842 | 0.032 | 0.037 |
| GEDIS_EUR | 0.6841 | 0.066 | 0.162 |
| GISS1_EUR | 0.1556 | −0.155 | 0.109 |
| GISS2_EUR | 0.376 | 0.112 | 0.126 |
| GTPRJ_AFR | 0.8484 | 0.012 | 0.065 |
| IPSYC_EUR | 0.0067 | 0.068 | 0.025 |
| JANS3_EUR | 0.7885 | −0.036 | 0.133 |
| MIREC_AFR | 0.1974 | 0.197 | 0.153 |
| MIREC_EUR | 0.7495 | −0.062 | 0.193 |
| MVPXQ_EUR | 0.004 | 0.047 | 0.016 |
| PGCBD_EUR | 0.0053 | 0.092 | 0.033 |
| PGCED_EUR | 0.559 | 0.084 | 0.144 |
| PGCMD_EUR | 0.5599 | 0.026 | 0.045 |
| PGCPT_AFR | 0.2687 | 0.121 | 0.109 |
| PGCPT_EUR | 0.005 | 0.196 | 0.07 |
| PGCSZ_EUR | 0.3843 | 0.041 | 0.047 |
| PRFCT_EUR | 0.761 | 0.017 | 0.056 |
| PSYCR_EUR | 0.3176 | −0.111 | 0.112 |
| QIMRB_EUR | 0.1113 | 0.056 | 0.035 |
| STRR1_LAT | 0.7597 | 0.078 | 0.256 |
| UKBJC_EUR | 0.0177 | 0.062 | 0.026 |
| YPENN_AFR | 0.943 | −0.012 | 0.164 |
| YPENN_EUR | 0.1219 | −0.273 | 0.176 |
| **meta** | **1.80e−09** | **0.052** | **0.009** |

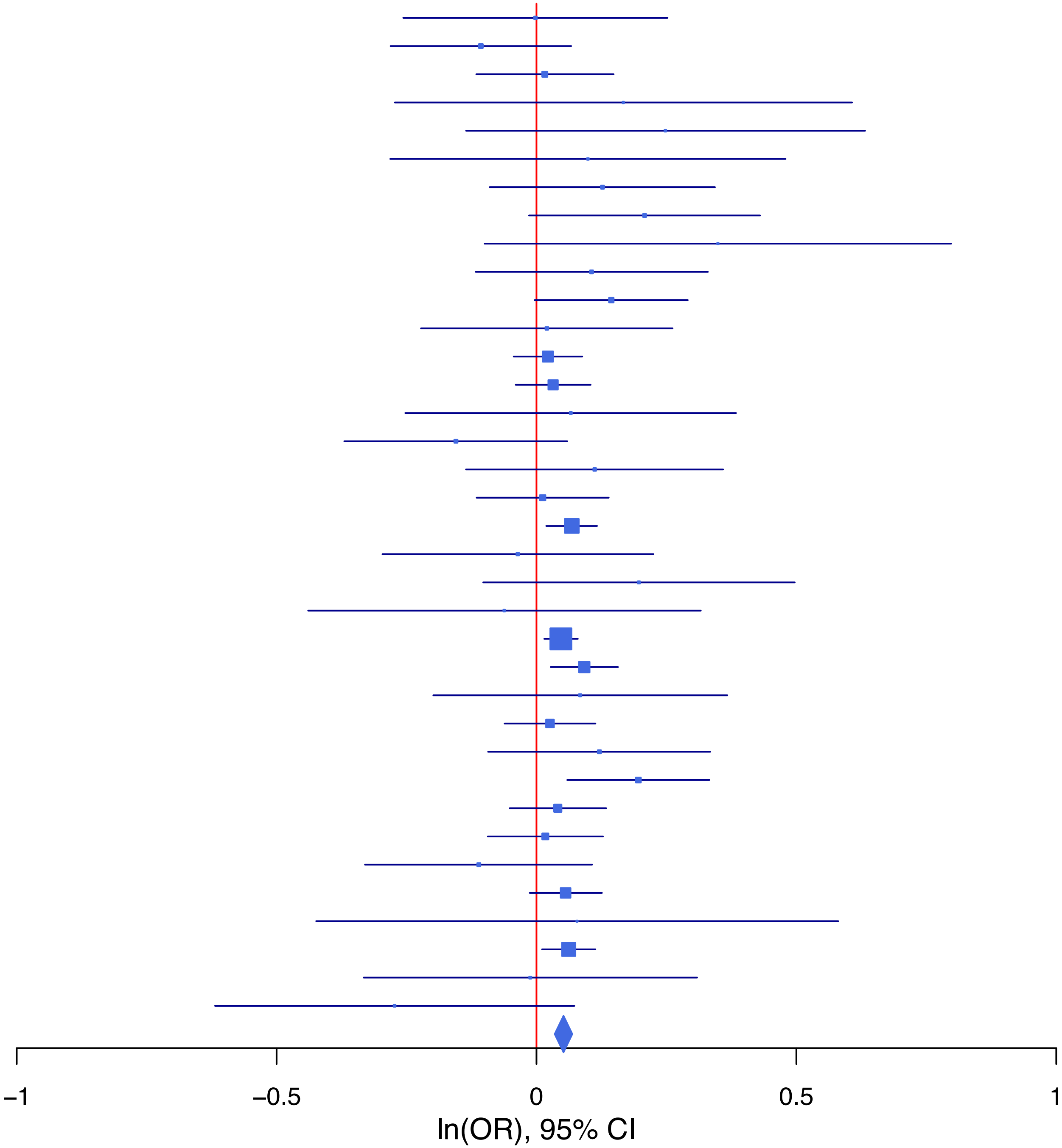

## rs12925759 A/G 16:51189544

| Cohort | P | ln(OR) | SE |
|---|---|---|---|
| ADHEA_AFR | 0.8008 | −0.043 | 0.169 |
| ADHEA_EUR | 0.7342 | 0.028 | 0.082 |
| ALSPC_EUR | 0.9163 | −0.007 | 0.067 |
| BEPS7_EUR | 0.0663 | −0.421 | 0.229 |
| BHRCM_AFR | 0.508 | −0.224 | 0.338 |
| BHRCM_EUR | 0.6587 | 0.083 | 0.187 |
| BHRCM_LAT | 0.7127 | 0.221 | 0.599 |
| BOR17_EUR | 0.7812 | −0.03 | 0.109 |
| BOR2C_EUR | 0.1351 | 0.153 | 0.102 |
| BOR2E_EUR | 0.0078 | −0.583 | 0.219 |
| COGA1_AFR | 0.1542 | 0.208 | 0.146 |
| COGA1_EUR | 0.8712 | 0.012 | 0.072 |
| CVEDA_CSA | 0.309 | 0.247 | 0.242 |
| ESTB2_EUR | 0.2635 | 0.029 | 0.026 |
| GEDIS_EUR | 0.6429 | −0.077 | 0.166 |
| GISS1_EUR | 1 | 0 | 0.092 |
| GISS2_EUR | 0.2454 | 0.123 | 0.106 |
| GREAT_EAS | 0.3886 | 0.143 | 0.166 |
| GTPRJ_AFR | 0.5371 | 0.054 | 0.086 |
| JANS3_EUR | 0.1614 | 0.169 | 0.121 |
| JANS4_EUR | 0.7801 | −0.04 | 0.145 |
| MVPXQ_AFR | 0.0311 | 0.072 | 0.033 |
| MVPXQ_EAS | 0.5708 | −0.14 | 0.246 |
| MVPXQ_LAT | 0.743 | −0.015 | 0.045 |
| PGCBD_EUR | 3e−04 | 0.118 | 0.032 |
| PGCED_EUR | 0.0124 | −0.389 | 0.155 |
| PGCMD_EUR | 3e−04 | 0.152 | 0.042 |
| PGCPT_AFR | 0.4698 | 0.102 | 0.142 |
| PGCPT_EUR | 0.6189 | 0.035 | 0.07 |
| PGCSZ_EUR | 0.2348 | 0.055 | 0.046 |
| PRFCT_EUR | 0.301 | 0.055 | 0.053 |
| PSYCR_EUR | 0.9287 | −0.009 | 0.105 |
| QIMRB_EUR | 0.316 | 0.034 | 0.034 |
| SNUBH−ASA_EAS | 0.9549 | −0.023 | 0.408 |
| STRR1_LAT | 0.144 | 0.322 | 0.22 |
| UKBJC_EUR | 0.0014 | 0.08 | 0.025 |
| YPENN_AFR | 0.1946 | 0.263 | 0.203 |
| YPENN_EUR | 0.8868 | −0.022 | 0.152 |
| **meta** | **2.39e−08** | **0.057** | **0.01** |

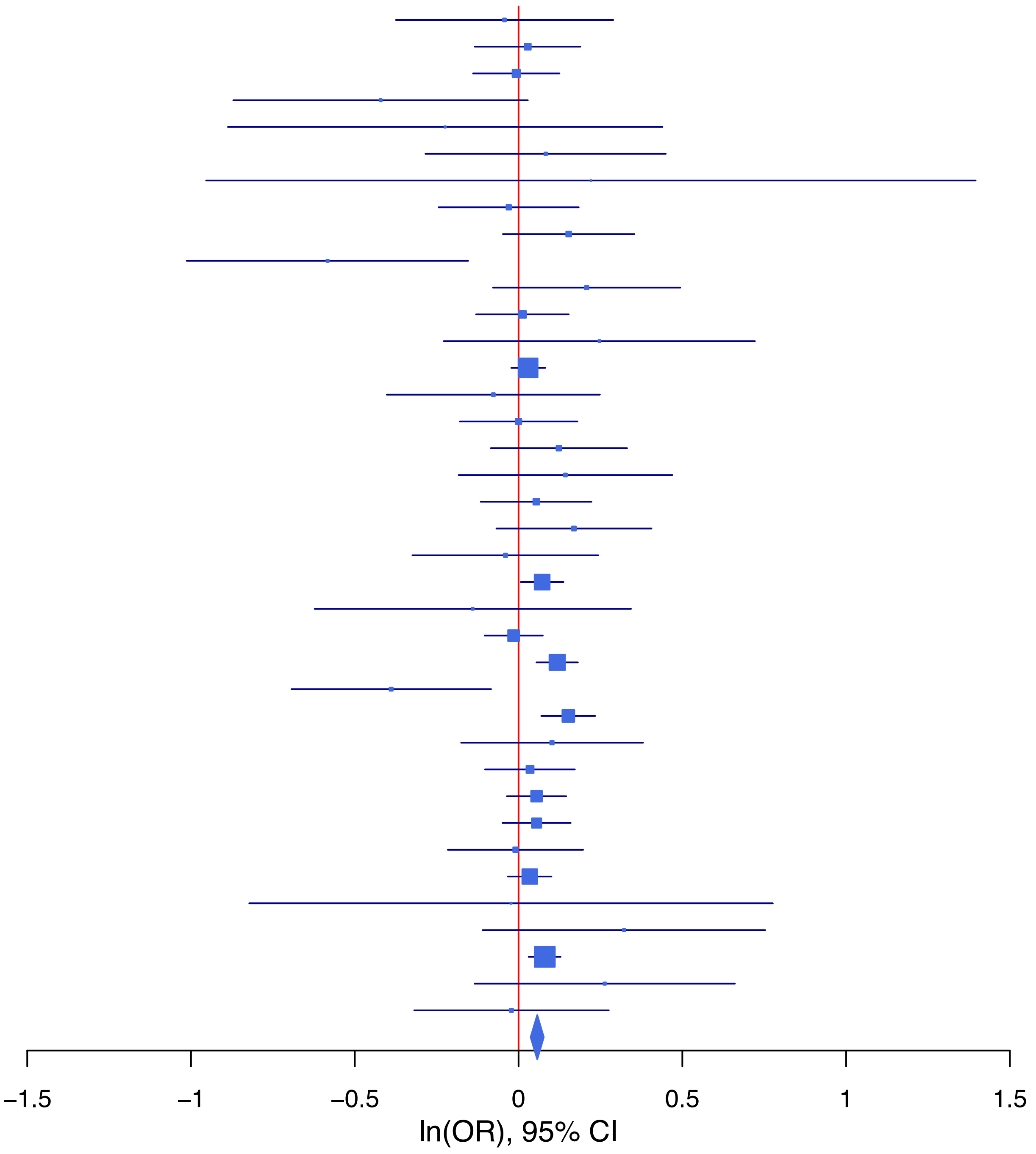

**rs13296973   A/C   9:122659318**

| Cohort | P | ln(OR) | SE |
|---|---|---|---|
| ADHEA_AFR | 0.7297 | −0.042 | 0.121 |
| ADHEA_EUR | 0.2819 | −0.077 | 0.071 |
| ALSPC_EUR | 0.6318 | −0.026 | 0.055 |
| BEPS7_EUR | 0.0255 | −0.429 | 0.192 |
| BHRCM_AFR | 0.4342 | 0.167 | 0.214 |
| BHRCM_EUR | 0.2237 | 0.195 | 0.16 |
| BHRCM_LAT | 0.7393 | 0.091 | 0.274 |
| BOR17_EUR | 0.4589 | 0.066 | 0.089 |
| BOR2C_EUR | 0.7292 | 0.031 | 0.088 |
| BOR2E_EUR | 0.2302 | 0.215 | 0.179 |
| CNVRG_EAS | 0.5517 | −0.031 | 0.052 |
| COGA1_AFR | 0.1671 | −0.145 | 0.105 |
| COGA1_EUR | 0.2874 | −0.063 | 0.059 |
| CUINT_EUR | 0.163 | −0.142 | 0.102 |
| CVEDA_CSA | 0.7744 | −0.046 | 0.159 |
| ESTB2_EUR | 0.0777 | −0.042 | 0.024 |
| FINNG_EUR | 0.0227 | −0.054 | 0.024 |
| GEDIS_EUR | 0.5031 | 0.09 | 0.134 |
| GISS1_EUR | 0.4948 | −0.055 | 0.08 |
| GISS2_EUR | 0.7371 | 0.033 | 0.098 |
| GREAT_EAS | 0.3999 | −0.07 | 0.084 |
| GTPRJ_AFR | 0.6883 | 0.025 | 0.063 |
| IPSYC_EUR | 0.0032 | −0.06 | 0.02 |
| JANS3_EUR | 0.1797 | −0.148 | 0.11 |
| JANS4_EUR | 0.9002 | −0.015 | 0.122 |
| MVPXQ_AFR | 0.0319 | −0.047 | 0.022 |
| MVPXQ_EAS | 0.0339 | −0.225 | 0.106 |
| MVPXQ_EUR | 0.0155 | −0.031 | 0.013 |
| MVPXQ_LAT | 0.0199 | −0.077 | 0.033 |
| PGCBD_EUR | 0.0533 | −0.051 | 0.027 |
| PGCED_EUR | 0.8554 | −0.022 | 0.121 |
| PGCMD_EUR | 0.0435 | −0.071 | 0.035 |
| PGCPT_AFR | 0.7244 | 0.038 | 0.106 |
| PGCPT_EUR | 0.3425 | −0.056 | 0.059 |
| PGCSZ_EUR | 0.9527 | −0.002 | 0.038 |
| PRFCT_EUR | 0.3008 | −0.047 | 0.045 |
| PSYCR_EUR | 0.9531 | 0.005 | 0.09 |
| QIMRB_EUR | 0.5324 | −0.018 | 0.028 |
| SNUBH−ASA_EAS | 0.4077 | −0.117 | 0.142 |
| STRR1_LAT | 0.9715 | 0.006 | 0.176 |
| UKBJC_EUR | 0.046 | −0.041 | 0.021 |
| YPENN_EUR | 0.8299 | −0.028 | 0.13 |
| **meta** | **2.39e−11** | **−0.042** | **0.006** |

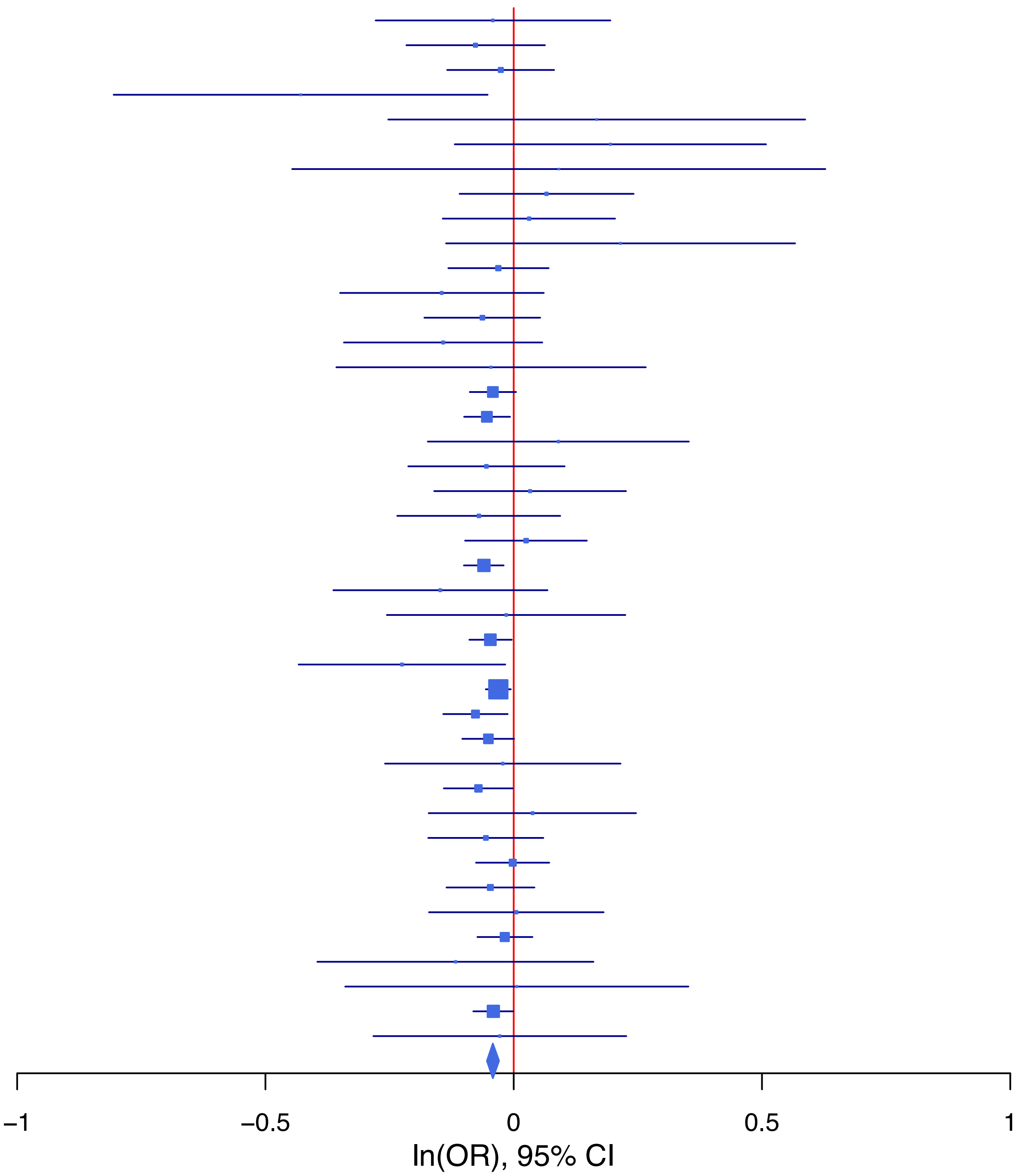

**rs1452787 G/A 18:53207207**

| Cohort | P | ln(OR) | SE |
|---|---|---|---|
| ADHEA_AFR | 0.5636 | 0.109 | 0.189 |
| ADHEA_EUR | 0.3651 | 0.066 | 0.073 |
| ALSPC_EUR | 0.6703 | −0.026 | 0.061 |
| BEPS7_EUR | 0.3116 | 0.194 | 0.192 |
| BHRCM_AFR | 0.3893 | −0.396 | 0.46 |
| BHRCM_EUR | 0.5528 | −0.103 | 0.173 |
| BHRCM_LAT | 0.3291 | 0.262 | 0.269 |
| BOR17_EUR | 0.9278 | −0.008 | 0.094 |
| BOR2C_EUR | 0.1167 | −0.152 | 0.097 |
| BOR2E_EUR | 0.4452 | −0.152 | 0.199 |
| CNVRG_EAS | 0.236 | 0.055 | 0.046 |
| COGA1_AFR | 0.4578 | −0.124 | 0.167 |
| COGA1_EUR | 0.4333 | 0.05 | 0.064 |
| CUINT_EUR | 0.0914 | −0.194 | 0.115 |
| CVEDA_CSA | 0.3156 | −0.169 | 0.169 |
| ESTB2_EUR | 0.2277 | 0.03 | 0.025 |
| FINNG_EUR | 0.1266 | 0.038 | 0.025 |
| GEDIS_EUR | 0.5074 | −0.097 | 0.146 |
| GISS1_EUR | 0.0353 | 0.174 | 0.083 |
| GISS2_EUR | 0.5508 | −0.057 | 0.096 |
| GTPRJ_AFR | 0.0391 | 0.208 | 0.101 |
| IPSYC_EUR | 0.0338 | 0.047 | 0.022 |
| JANS3_EUR | 0.0944 | −0.198 | 0.118 |
| JANS4_EUR | 0.6287 | −0.063 | 0.13 |
| MIREC_AFR | 0.5286 | −0.15 | 0.238 |
| MIREC_EUR | 0.5564 | −0.099 | 0.168 |
| MVPXQ_AFR | 0.5423 | 0.021 | 0.034 |
| MVPXQ_EUR | 2.45e−05 | 0.058 | 0.014 |
| MVPXQ_LAT | 0.3206 | −0.034 | 0.034 |
| PGCBD_EUR | 0.7559 | −0.009 | 0.029 |
| PGCED_EUR | 0.5108 | −0.083 | 0.127 |
| PGCMD_EUR | 0.8691 | −0.006 | 0.038 |
| PGCPT_EUR | 0.984 | −0.001 | 0.063 |
| PGCSZ_EUR | 0.0079 | 0.107 | 0.04 |
| PRFCT_EUR | 0.1114 | 0.078 | 0.049 |
| PSYCR_EUR | 0.5973 | 0.05 | 0.094 |
| QIMRB_EUR | 0.3472 | 0.028 | 0.03 |
| SNUBH−ASA_EAS | 0.0676 | 0.23 | 0.126 |
| SNUBH−KCHIP_EAS | 0.0987 | 0.277 | 0.168 |
| STRR1_LAT | 0.5123 | 0.116 | 0.178 |
| UKBJC_EUR | 0.0034 | 0.065 | 0.022 |
| YPENN_AFR | 0.5252 | 0.141 | 0.222 |
| YPENN_EUR | 0.4574 | 0.099 | 0.133 |
| **meta** | **4.34e−08** | **0.038** | **0.007** |

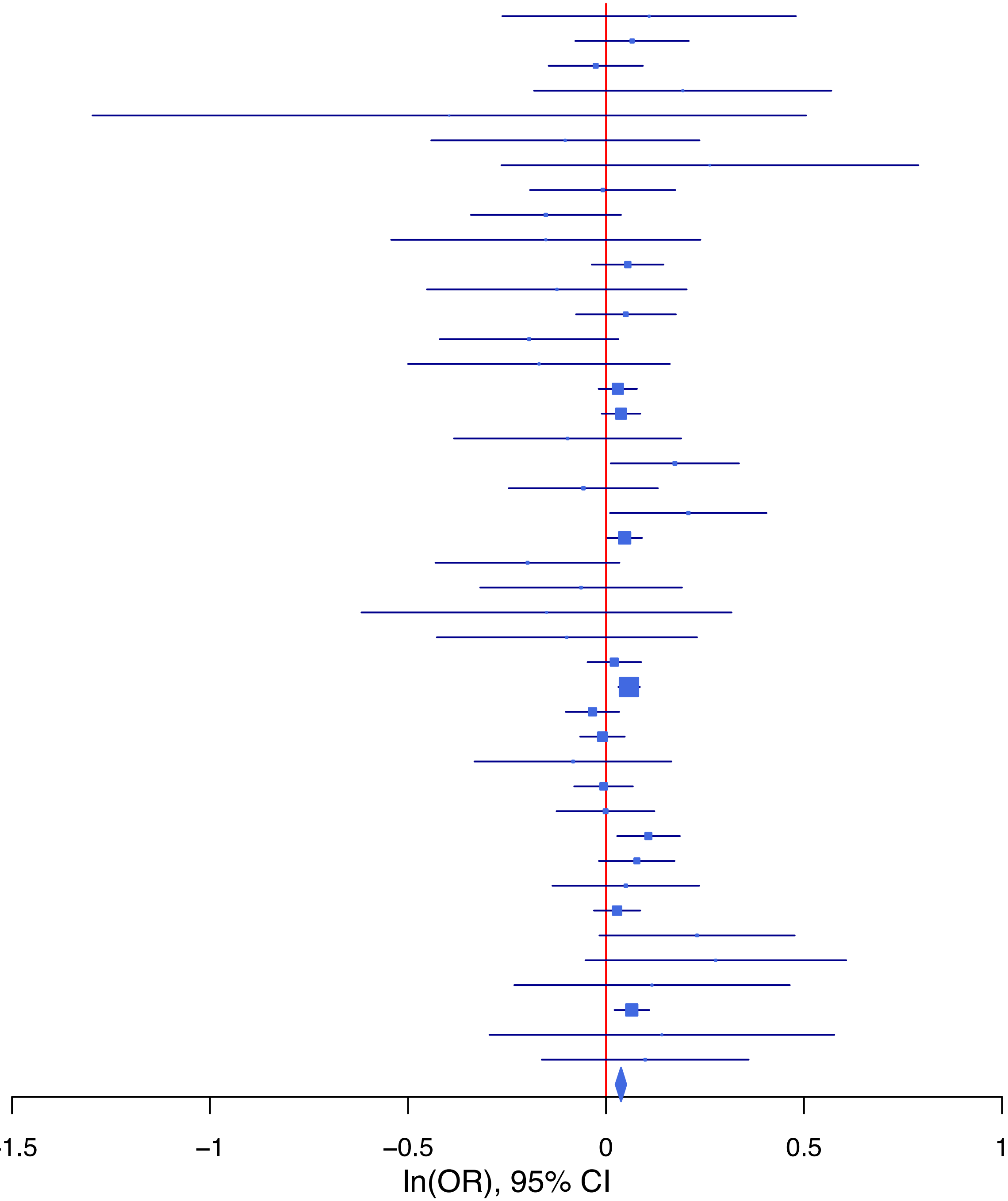

**rs17602038 C/T 11:113364691**

| Cohort | P | ln(OR) | SE |
|---|---|---|---|
| ADHEA_AFR | 0.2165 | −0.241 | 0.195 |
| ADHEA_EUR | 0.2342 | −0.085 | 0.071 |
| ALSPC_EUR | 0.6781 | −0.023 | 0.055 |
| BEPS7_EUR | 0.8831 | 0.028 | 0.193 |
| BHRCM_AFR | 0.3445 | 0.31 | 0.328 |
| BHRCM_EUR | 0.8547 | −0.03 | 0.163 |
| BOR17_EUR | 0.6915 | 0.034 | 0.086 |
| BOR2C_EUR | 0.3625 | −0.081 | 0.09 |
| BOR2E_EUR | 0.4234 | 0.154 | 0.193 |
| CNVRG_EAS | 0.4303 | 0.108 | 0.137 |
| COGA1_AFR | 0.5432 | 0.087 | 0.143 |
| COGA1_EUR | 0.4442 | −0.047 | 0.061 |
| CUINT_EUR | 0.9354 | −0.008 | 0.103 |
| CVEDA_CSA | 0.7048 | 0.074 | 0.196 |
| ESTB2_EUR | 0.0077 | −0.069 | 0.026 |
| FINNG_EUR | 0.0415 | −0.058 | 0.028 |
| GEDIS_EUR | 0.13 | −0.21 | 0.139 |
| GISS1_EUR | 0.9008 | −0.01 | 0.083 |
| GISS2_EUR | 0.9638 | 0.004 | 0.094 |
| GTPRJ_AFR | 0.8486 | 0.018 | 0.091 |
| IPSYC_EUR | 0.002 | −0.064 | 0.021 |
| JANS3_EUR | 0.1025 | −0.177 | 0.108 |
| JANS4_EUR | 0.1201 | 0.186 | 0.12 |
| MIREC_AFR | 0.1123 | −0.379 | 0.238 |
| MIREC_EUR | 0.016 | −0.405 | 0.168 |
| MVPXQ_AFR | 0.0261 | −0.068 | 0.031 |
| MVPXQ_EAS | 0.0979 | 0.312 | 0.188 |
| MVPXQ_EUR | 7.54e−08 | −0.07 | 0.013 |
| PGCBD_EUR | 0.192 | −0.035 | 0.027 |
| PGCED_EUR | 0.323 | −0.115 | 0.116 |
| PGCMD_EUR | 0.6273 | 0.017 | 0.035 |
| PGCPT_AFR | 0.7706 | 0.052 | 0.179 |
| PGCPT_EUR | 0.8734 | −0.009 | 0.059 |
| PGCSZ_EUR | 0.0775 | −0.068 | 0.038 |
| PRFCT_EUR | 0.7379 | −0.016 | 0.047 |
| PSYCR_EUR | 0.0211 | 0.204 | 0.088 |
| QIMRB_EUR | 0.0976 | −0.046 | 0.028 |
| SNUBH−ASA_EAS | 0.9773 | −0.01 | 0.357 |
| STRR1_LAT | 0.0182 | 0.427 | 0.181 |
| UKBJC_EUR | 0.2132 | −0.026 | 0.021 |
| YPENN_AFR | 0.2933 | 0.213 | 0.203 |
| YPENN_EUR | 0.9499 | −0.008 | 0.132 |
| **meta** | **1.84e−12** | **−0.048** | **0.007** |

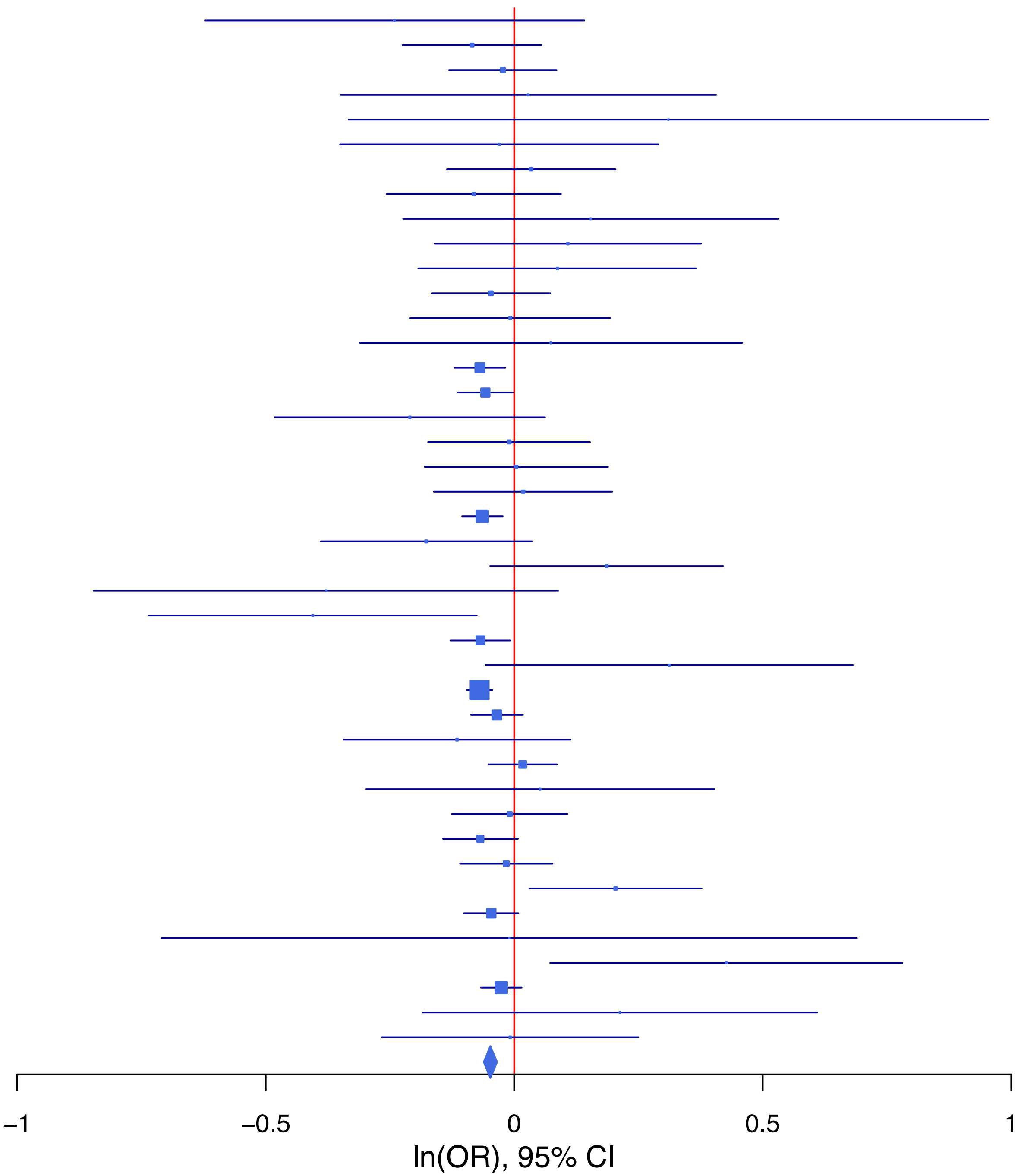

**rs185782836 A/G 14:64520663**

| Cohort | P | ln(OR) | SE |
|---|---|---|---|
| ALSPC_EUR | 0.1242 | 0.272 | 0.177 |
| BHRCM_EUR | 0.1301 | 0.751 | 0.496 |
| BOR17_EUR | 0.2941 | 0.309 | 0.294 |
| BOR2C_EUR | 0.7057 | 0.114 | 0.301 |
| ESTB2_EUR | 0.014 | 0.128 | 0.052 |
| FINNG_EUR | 0.0281 | 0.235 | 0.107 |
| GISS2_EUR | 0.8451 | 0.051 | 0.259 |
| IPSYC_EUR | 1.08e−05 | 0.332 | 0.075 |
| MVPXQ_EUR | 0.0081 | 0.153 | 0.058 |
| MVPXQ_LAT | 0.1695 | −0.216 | 0.157 |
| PGCPT_EUR | 0.2954 | −0.287 | 0.274 |
| PGCSZ_EUR | 0.7751 | −0.053 | 0.187 |
| PRFCT_EUR | 0.5743 | 0.088 | 0.156 |
| PSYCR_EUR | 0.4005 | 0.245 | 0.292 |
| QIMRB_EUR | 0.4612 | 0.077 | 0.104 |
| UKBJC_EUR | 0.189 | 0.1 | 0.076 |
| **meta** | **3.91e−08** | **0.147** | **0.027** |

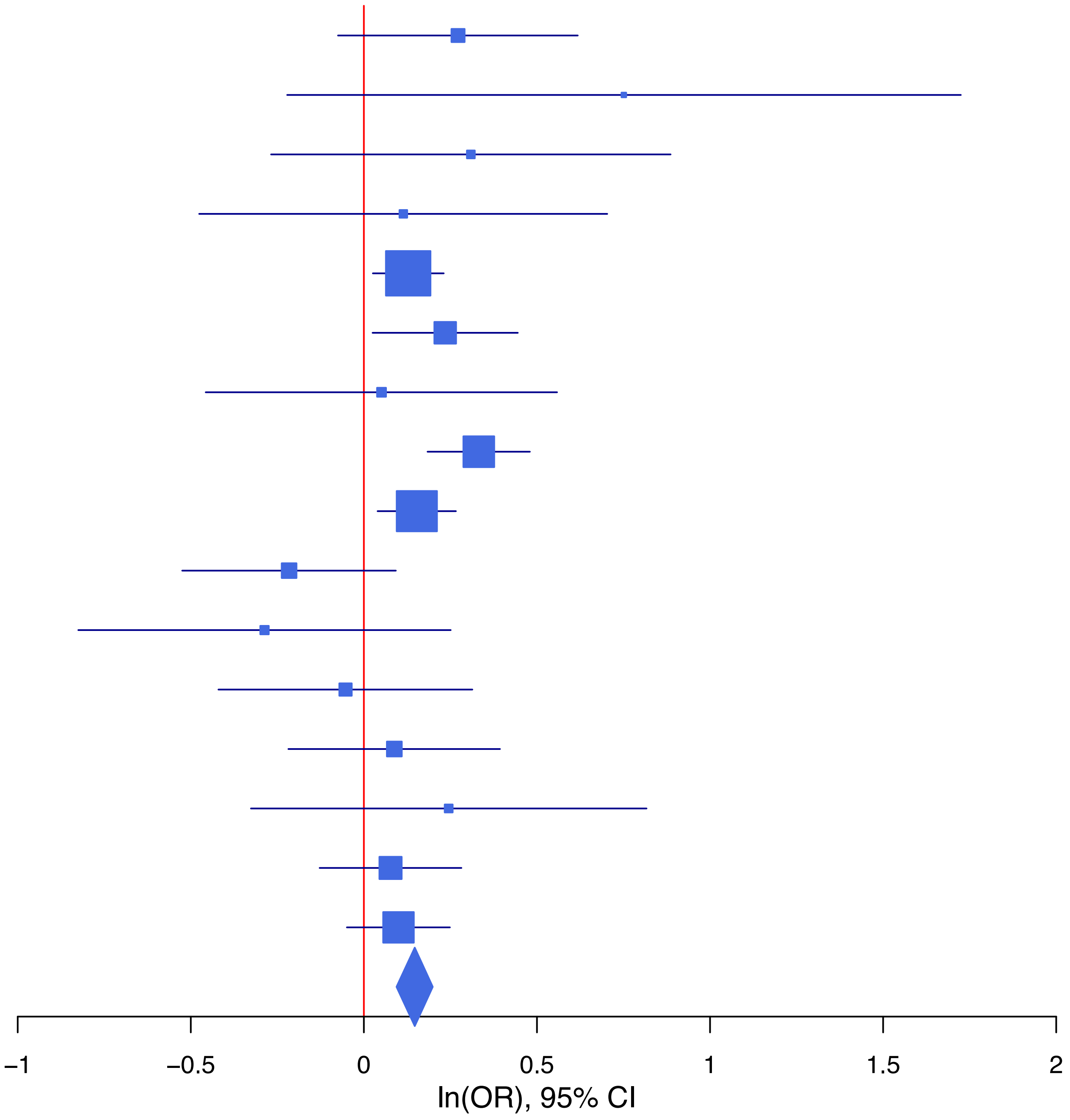

**rs28855509 C/T 17:65998167**

| Cohort | P | ln(OR) | SE |
|---|---|---|---|
| ALSPC_EUR | 0.9074 | 0.008 | 0.071 |
| BEPS7_EUR | 0.747 | 0.07 | 0.216 |
| BOR17_EUR | 0.9058 | −0.012 | 0.105 |
| BOR2C_EUR | 0.0931 | 0.173 | 0.103 |
| BOR2E_EUR | 0.6925 | −0.093 | 0.234 |
| CNVRG_EAS | 0.7463 | −0.015 | 0.047 |
| COGA1_EUR | 0.7715 | −0.02 | 0.07 |
| CVEDA_CSA | 0.9988 | 0 | 0.184 |
| GISS1_EUR | 0.7694 | −0.029 | 0.098 |
| GISS2_EUR | 0.0159 | 0.277 | 0.115 |
| GTPRJ_AFR | 0.4946 | 0.067 | 0.098 |
| IPSYC_EUR | 0.1079 | 0.038 | 0.024 |
| JANS3_EUR | 0.5546 | −0.076 | 0.128 |
| JANS4_EUR | 0.3256 | 0.135 | 0.138 |
| MVPXQ_AFR | 0.0014 | 0.116 | 0.036 |
| MVPXQ_EAS | 0.8002 | 0.027 | 0.107 |
| MVPXQ_EUR | 9e−04 | 0.051 | 0.015 |
| PGCBD_EUR | 0.6338 | 0.015 | 0.032 |
| PGCED_EUR | 0.3694 | 0.141 | 0.157 |
| PGCMD_EUR | 0.3323 | 0.041 | 0.043 |
| PGCPT_EUR | 0.3311 | 0.066 | 0.068 |
| PGCSZ_EUR | 0.8312 | 0.01 | 0.046 |
| PRFCT_EUR | 0.031 | 0.11 | 0.051 |
| PSYCR_EUR | 0.5011 | 0.068 | 0.101 |
| QIMRB_EUR | 0.4704 | 0.024 | 0.033 |
| SNUBH-ASA_EAS | 0.3566 | 0.117 | 0.127 |
| SNUBH-KCHIP_EAS | 0.9257 | 0.016 | 0.171 |
| STRR1_LAT | 0.1295 | 0.288 | 0.19 |
| UKBJC_EUR | 0.0168 | 0.058 | 0.024 |
| YPENN_EUR | 0.6706 | −0.065 | 0.153 |
| **meta** | **3.32e−08** | **0.047** | **0.009** |

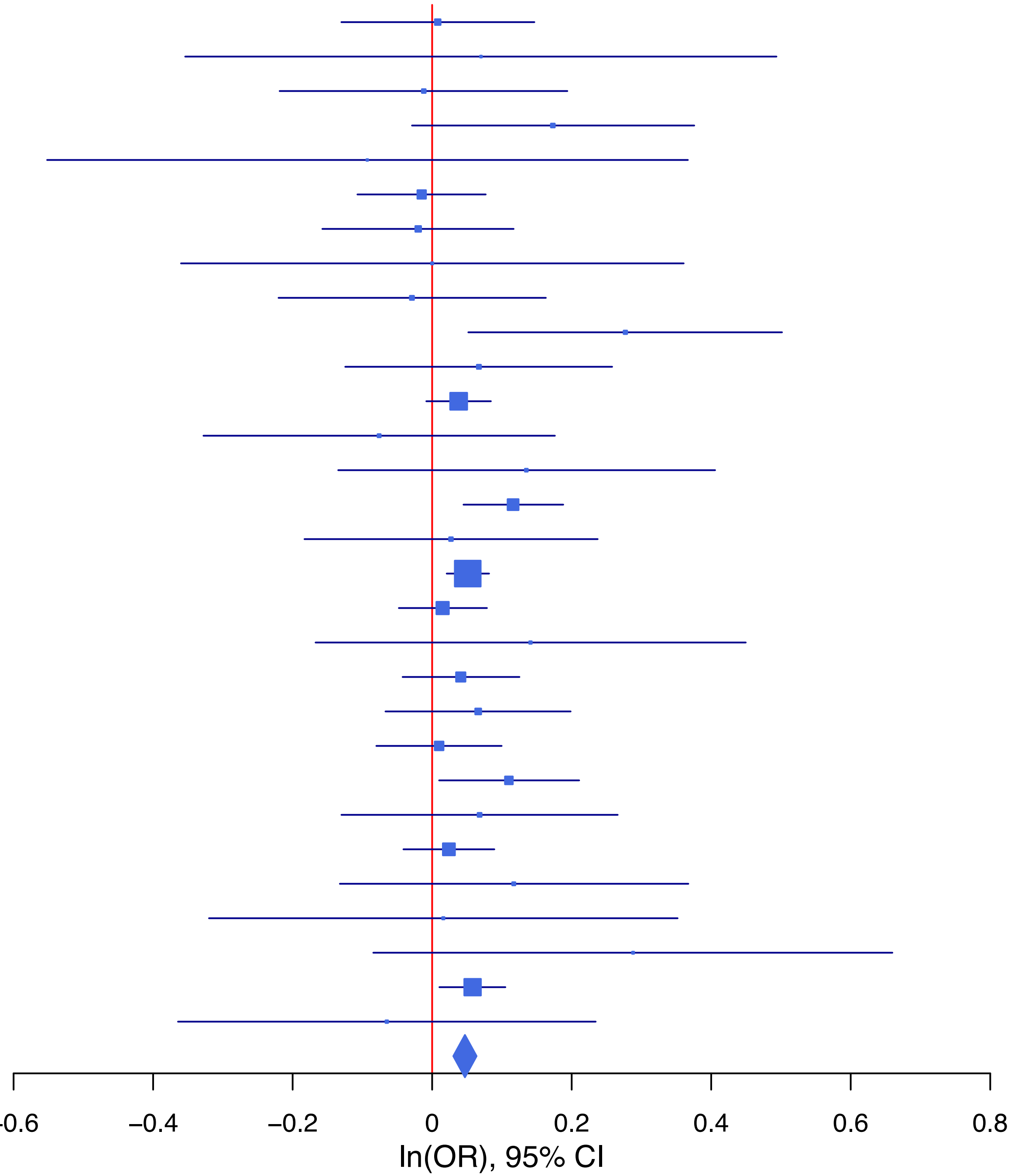

**rs35942385 T/G 2:144208523**

| Cohort | P | ln(OR) | SE |
|---|---|---|---|
| ALSPC_EUR | 0.3846 | 0.048 | 0.055 |
| BEPS7_EUR | 0.6335 | 0.09 | 0.188 |
| BHRCM_AFR | 0.0142 | 0.674 | 0.275 |
| BHRCM_EUR | 0.7833 | −0.046 | 0.167 |
| BOR17_EUR | 0.2627 | −0.099 | 0.088 |
| BOR2C_EUR | 0.5127 | −0.058 | 0.088 |
| BOR2E_EUR | 0.5254 | −0.118 | 0.186 |
| COGA1_EUR | 0.4845 | −0.043 | 0.061 |
| CUINT_EUR | 0.4602 | 0.076 | 0.103 |
| CVEDA_CSA | 0.481 | 0.162 | 0.229 |
| ESTB2_EUR | 0.0589 | −0.047 | 0.025 |
| FINNG_EUR | 0.0218 | −0.056 | 0.024 |
| GEDIS_EUR | 7e−04 | −0.496 | 0.147 |
| GISS1_EUR | 0.0458 | −0.17 | 0.085 |
| GISS2_EUR | 0.8264 | 0.022 | 0.101 |
| GTPRJ_AFR | 0.8496 | 0.018 | 0.094 |
| IPSYC_EUR | 0.0102 | −0.053 | 0.02 |
| JANS3_EUR | 0.0385 | 0.223 | 0.108 |
| MVPXQ_AFR | 0.7944 | −0.008 | 0.032 |
| MVPXQ_EAS | 0.595 | 0.127 | 0.239 |
| MVPXQ_EUR | 9.56e−07 | −0.064 | 0.013 |
| MVPXQ_LAT | 0.2902 | −0.04 | 0.038 |
| PGCBD_EUR | 0.3855 | 0.023 | 0.027 |
| PGCED_EUR | 0.7154 | 0.043 | 0.118 |
| PGCMD_EUR | 0.0738 | −0.064 | 0.036 |
| PGCPT_AFR | 0.9191 | 0.015 | 0.151 |
| PGCPT_EUR | 0.5448 | 0.036 | 0.06 |
| PGCSZ_EUR | 0.1958 | −0.05 | 0.039 |
| PRFCT_EUR | 0.0297 | −0.1 | 0.046 |
| PSYCR_EUR | 0.6664 | 0.04 | 0.092 |
| QIMRB_EUR | 0.1037 | −0.046 | 0.028 |
| STRR1_LAT | 0.1037 | 0.303 | 0.186 |
| UKBJC_EUR | 0.4793 | −0.015 | 0.021 |
| YPENN_EUR | 0.8745 | −0.021 | 0.132 |
| **meta** | **2.45e−09** | **−0.04** | **0.007** |

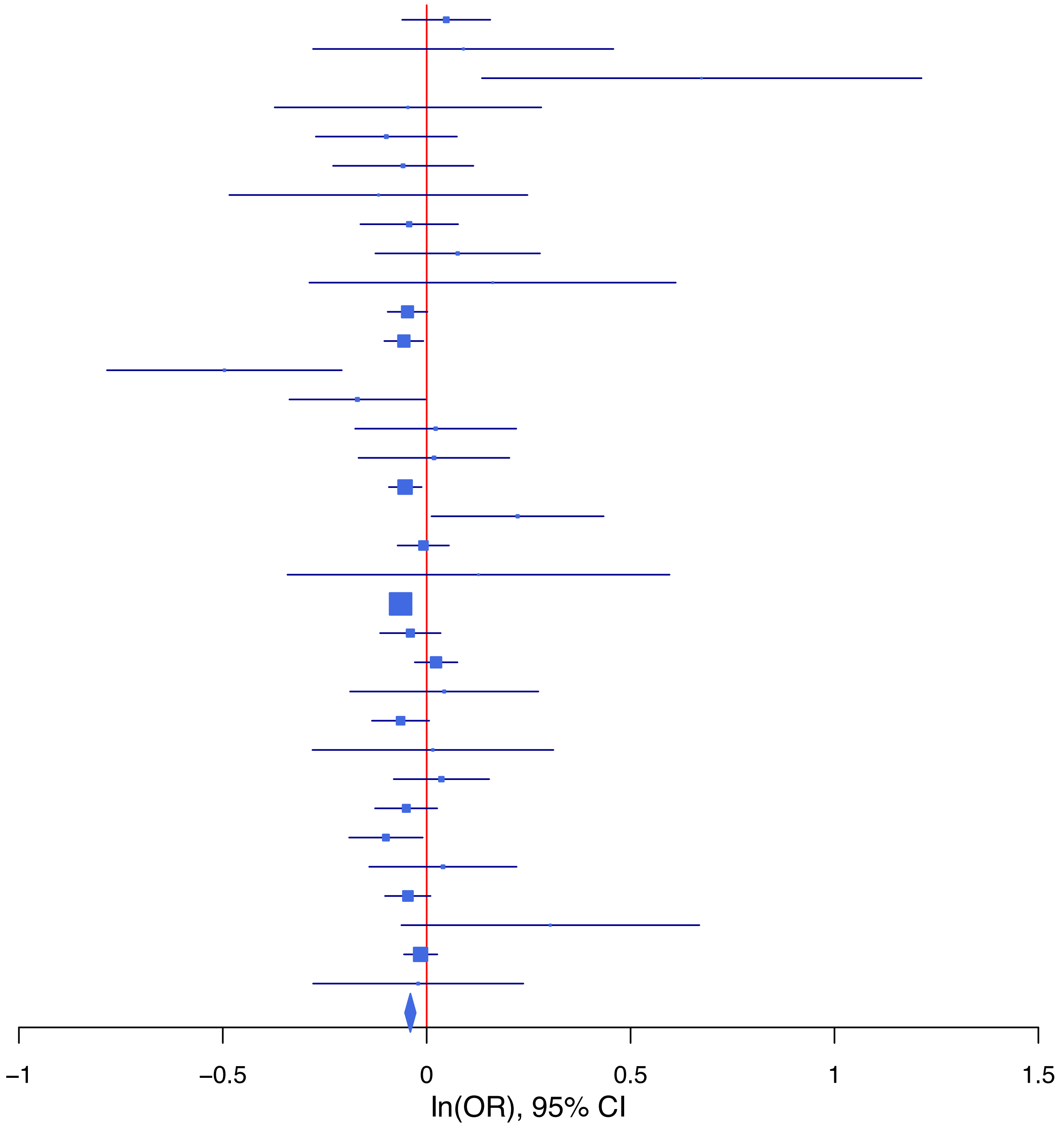

**rs36065861 T/C 4:166063595**

| Cohort | P | ln(OR) | SE |
|---|---|---|---|
| ADHEA_AFR | 0.5189 | −0.138 | 0.214 |
| ADHEA_EUR | 0.6919 | −0.041 | 0.103 |
| ALSPC_EUR | 0.5565 | 0.046 | 0.078 |
| BEPS7_EUR | 0.7954 | 0.064 | 0.247 |
| BHRCM_AFR | 0.221 | −0.515 | 0.421 |
| BHRCM_EUR | 0.7173 | −0.105 | 0.289 |
| BHRCM_LAT | 0.4666 | 0.214 | 0.294 |
| BOR17_EUR | 0.6737 | −0.056 | 0.133 |
| BOR2C_EUR | 0.0213 | −0.299 | 0.13 |
| BOR2E_EUR | 0.663 | −0.135 | 0.31 |
| CNVRG_EAS | 0.1551 | 0.173 | 0.122 |
| COGA1_AFR | 0.0347 | −0.377 | 0.178 |
| COGA1_EUR | 0.0214 | −0.208 | 0.09 |
| CUINT_EUR | 0.5095 | 0.101 | 0.153 |
| CVEDA_CSA | 0.8066 | 0.041 | 0.168 |
| ESTB2_EUR | 0.2099 | −0.041 | 0.033 |
| FINNG_EUR | 0.0216 | −0.068 | 0.03 |
| GEDIS_EUR | 0.7703 | −0.057 | 0.196 |
| GISS1_EUR | 0.102 | −0.191 | 0.117 |
| GISS2_EUR | 0.9691 | 0.005 | 0.133 |
| GREAT_EAS | 0.5317 | −0.149 | 0.238 |
| GTPRJ_AFR | 0.6418 | 0.049 | 0.105 |
| IPSYC_EUR | 0.1735 | −0.039 | 0.029 |
| JANS3_EUR | 0.1186 | −0.26 | 0.167 |
| JANS4_EUR | 0.8559 | 0.035 | 0.191 |
| MVPXQ_AFR | 0.0611 | −0.072 | 0.038 |
| MVPXQ_EAS | 0.5241 | −0.137 | 0.214 |
| MVPXQ_EUR | 0.0125 | −0.048 | 0.019 |
| PGCBD_EUR | 0.1435 | −0.058 | 0.04 |
| PGCED_EUR | 0.8423 | 0.038 | 0.19 |
| PGCMD_EUR | 0.8295 | −0.011 | 0.053 |
| PGCPT_EUR | 0.6573 | −0.039 | 0.088 |
| PGCSZ_EUR | 0.5533 | −0.033 | 0.056 |
| PRFCT_EUR | 0.0181 | −0.142 | 0.06 |
| PSYCR_EUR | 0.0108 | −0.369 | 0.145 |
| QIMRB_EUR | 0.0843 | −0.07 | 0.04 |
| SNUBH−ASA_EAS | 0.0315 | 0.728 | 0.338 |
| STRR1_LAT | 0.2571 | −0.285 | 0.252 |
| UKBJC_EUR | 0.167 | −0.04 | 0.029 |
| YPENN_EUR | 0.6326 | −0.091 | 0.19 |
| **meta** | **9.93e−09** | **−0.054** | **0.009** |

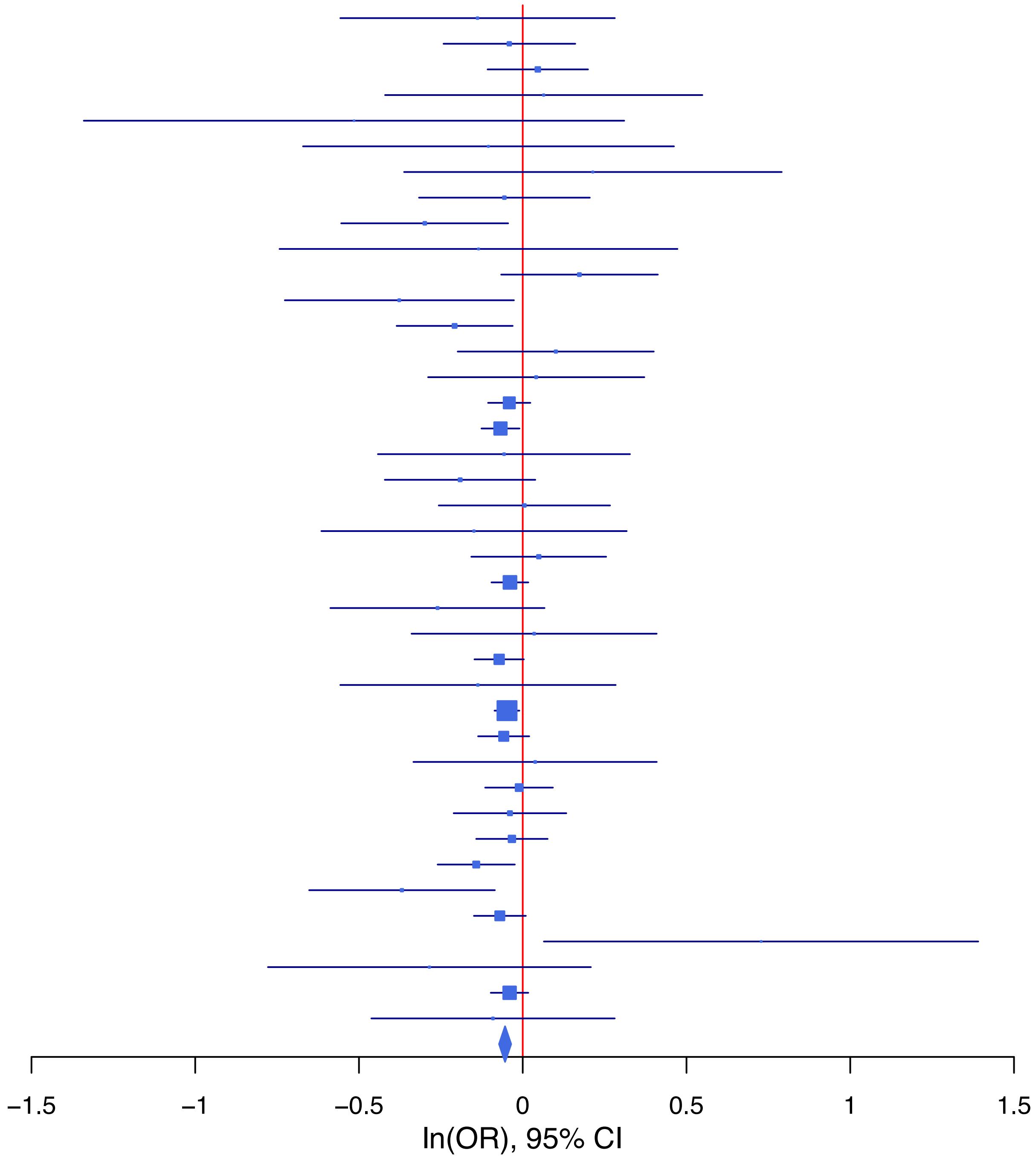

**rs4534184 C/G 9:112055493**

| Cohort | P | ln(OR) | SE |
|---|---|---|---|
| ADHEA_AFR | 0.4186 | 0.152 | 0.188 |
| ADHEA_EUR | 0.0334 | 0.212 | 0.1 |
| ALSPC_EUR | 0.5469 | −0.05 | 0.083 |
| BEPS7_EUR | 0.7567 | 0.089 | 0.287 |
| BHRCM_AFR | 0.185 | −0.655 | 0.494 |
| BHRCM_EUR | 0.6075 | −0.165 | 0.322 |
| BOR17_EUR | 0.1896 | 0.198 | 0.151 |
| BOR2C_EUR | 0.5886 | 0.079 | 0.146 |
| BOR2E_EUR | 0.4231 | 0.24 | 0.299 |
| CNVRG_EAS | 0.2593 | 0.112 | 0.099 |
| COGA1_AFR | 0.0593 | 0.333 | 0.176 |
| COGA1_EUR | 0.4472 | 0.071 | 0.093 |
| CUINT_EUR | 0.5925 | −0.094 | 0.176 |
| CVEDA_CSA | 0.9325 | 0.026 | 0.312 |
| ESTB2_EUR | 0.0074 | 0.101 | 0.038 |
| FINNG_EUR | 0.398 | 0.034 | 0.04 |
| GEDIS_EUR | 0.0901 | 0.323 | 0.19 |
| GISS1_EUR | 0.5613 | 0.075 | 0.129 |
| GISS2_EUR | 0.0483 | 0.306 | 0.155 |
| GTPRJ_AFR | 0.6639 | 0.044 | 0.1 |
| IPSYC_EUR | 0.3031 | 0.032 | 0.032 |
| JANS3_EUR | 0.6825 | −0.067 | 0.163 |
| JANS4_EUR | 0.1026 | 0.307 | 0.188 |
| PGCBD_EUR | 0.0431 | 0.086 | 0.042 |
| PGCED_EUR | 0.9266 | 0.019 | 0.208 |
| PGCMD_EUR | 0.9982 | 0 | 0.059 |
| PGCPT_AFR | 0.9642 | 0.01 | 0.218 |
| PGCPT_EUR | 0.3251 | 0.087 | 0.088 |
| PGCSZ_EUR | 0.0074 | 0.155 | 0.058 |
| PRFCT_EUR | 0.5681 | −0.041 | 0.072 |
| PSYCR_EUR | 0.9368 | 0.012 | 0.147 |
| QIMRB_EUR | 0.1255 | 0.065 | 0.042 |
| UKBJC_EUR | 0.0037 | 0.088 | 0.03 |
| YPENN_AFR | 0.2349 | 0.288 | 0.242 |
| YPENN_EUR | 0.8337 | 0.043 | 0.204 |
| **meta** | **1.88e−08** | **0.069** | **0.012** |

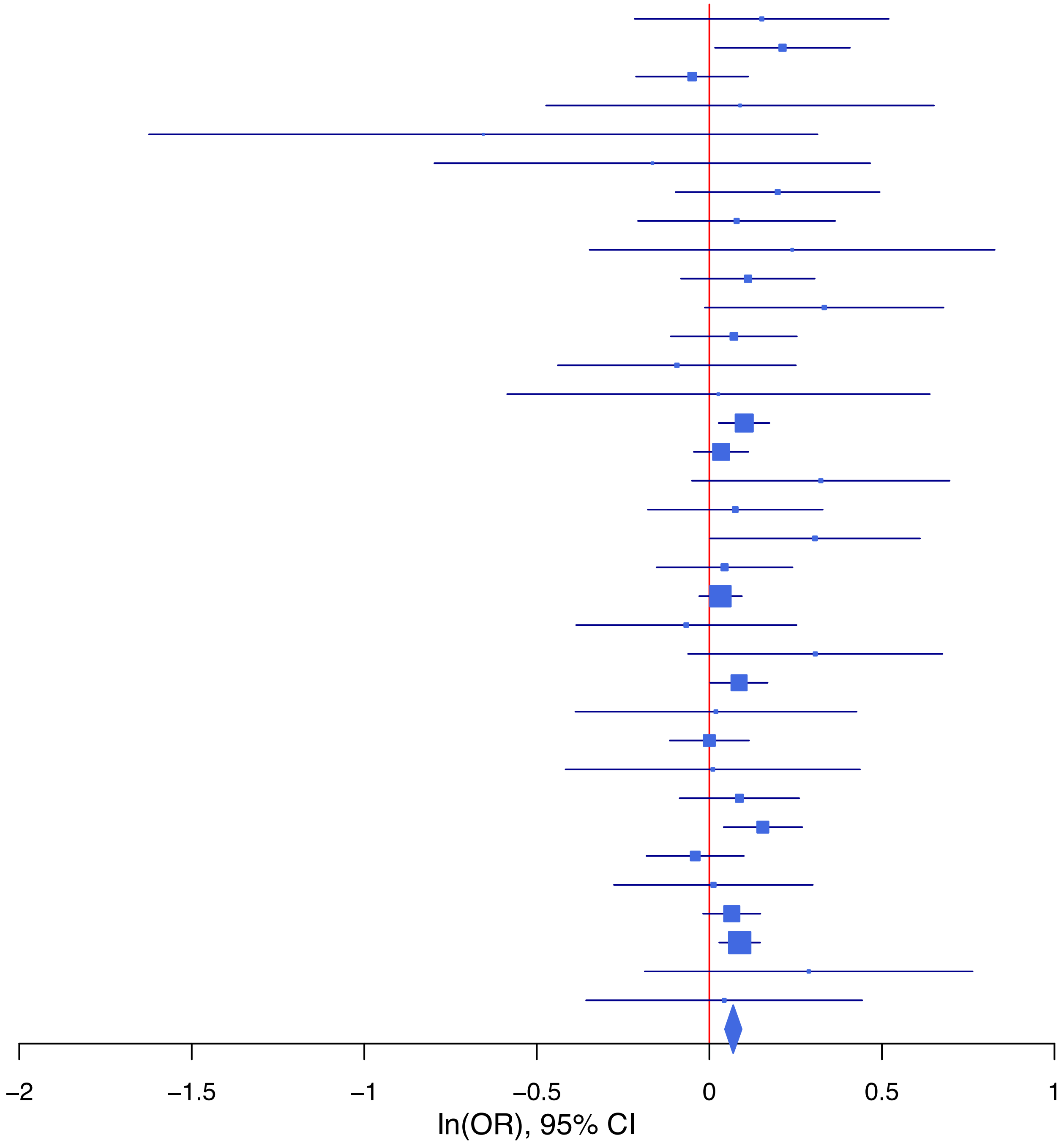

**rs4870063 C/A 6:152244647**

| Cohort | P | ln(OR) | SE |
|---|---|---|---|
| BEPS7_EUR | 0.4874 | 0.129 | 0.186 |
| BHRCM_AFR | 0.9257 | 0.022 | 0.235 |
| BHRCM_EUR | 0.8904 | 0.025 | 0.176 |
| BHRCM_LAT | 0.1375 | −0.381 | 0.257 |
| BOR17_EUR | 0.2816 | 0.098 | 0.091 |
| BOR2C_EUR | 0.4433 | 0.07 | 0.091 |
| BOR2E_EUR | 0.1557 | 0.267 | 0.188 |
| CNVRG_EAS | 0.2714 | 0.064 | 0.058 |
| COGA1_AFR | 0.9477 | −0.007 | 0.109 |
| COGA1_EUR | 0.2317 | 0.074 | 0.062 |
| CUINT_EUR | 0.3426 | −0.101 | 0.106 |
| CVEDA_CSA | 0.9159 | 0.018 | 0.166 |
| ESTB2_EUR | 8e−04 | 0.083 | 0.025 |
| FINNG_EUR | 0.3476 | 0.023 | 0.024 |
| GEDIS_EUR | 0.2779 | 0.146 | 0.134 |
| GISS1_EUR | 0.1173 | −0.14 | 0.09 |
| GISS2_EUR | 0.9328 | −0.008 | 0.099 |
| GREAT_EAS | 0.0171 | 0.233 | 0.098 |
| GTPRJ_AFR | 0.3077 | 0.07 | 0.069 |
| IPSYC_EUR | 0.5516 | 0.012 | 0.021 |
| JANS3_EUR | 0.1333 | −0.168 | 0.112 |
| JANS4_EUR | 0.7978 | −0.033 | 0.129 |
| MVPXQ_EUR | 2.13e−05 | 0.057 | 0.013 |
| MVPXQ_LAT | 0.0032 | 0.099 | 0.034 |
| PGCBD_EUR | 0.1681 | 0.038 | 0.027 |
| PGCED_EUR | 0.0473 | −0.252 | 0.127 |
| PGCMD_EUR | 0.123 | 0.056 | 0.036 |
| PGCPT_AFR | 0.0613 | 0.227 | 0.121 |
| PGCPT_EUR | 0.3291 | 0.059 | 0.06 |
| PGCSZ_EUR | 0.5284 | 0.024 | 0.039 |
| PRFCT_EUR | 0.0018 | 0.145 | 0.046 |
| PSYCR_EUR | 0.0842 | 0.156 | 0.09 |
| QIMRB_EUR | 0.1849 | 0.038 | 0.028 |
| SNUBH−ASA_EAS | 0.2648 | 0.169 | 0.151 |
| SNUBH−KCHIP_EAS | 0.6746 | −0.084 | 0.2 |
| STRR1_LAT | 0.0061 | 0.498 | 0.182 |
| UKBJC_EUR | 1e−04 | 0.079 | 0.021 |
| YPENN_AFR | 0.5527 | 0.103 | 0.173 |
| YPENN_EUR | 0.8904 | −0.018 | 0.132 |
| **meta** | **7.58e−15** | **0.053** | **0.007** |

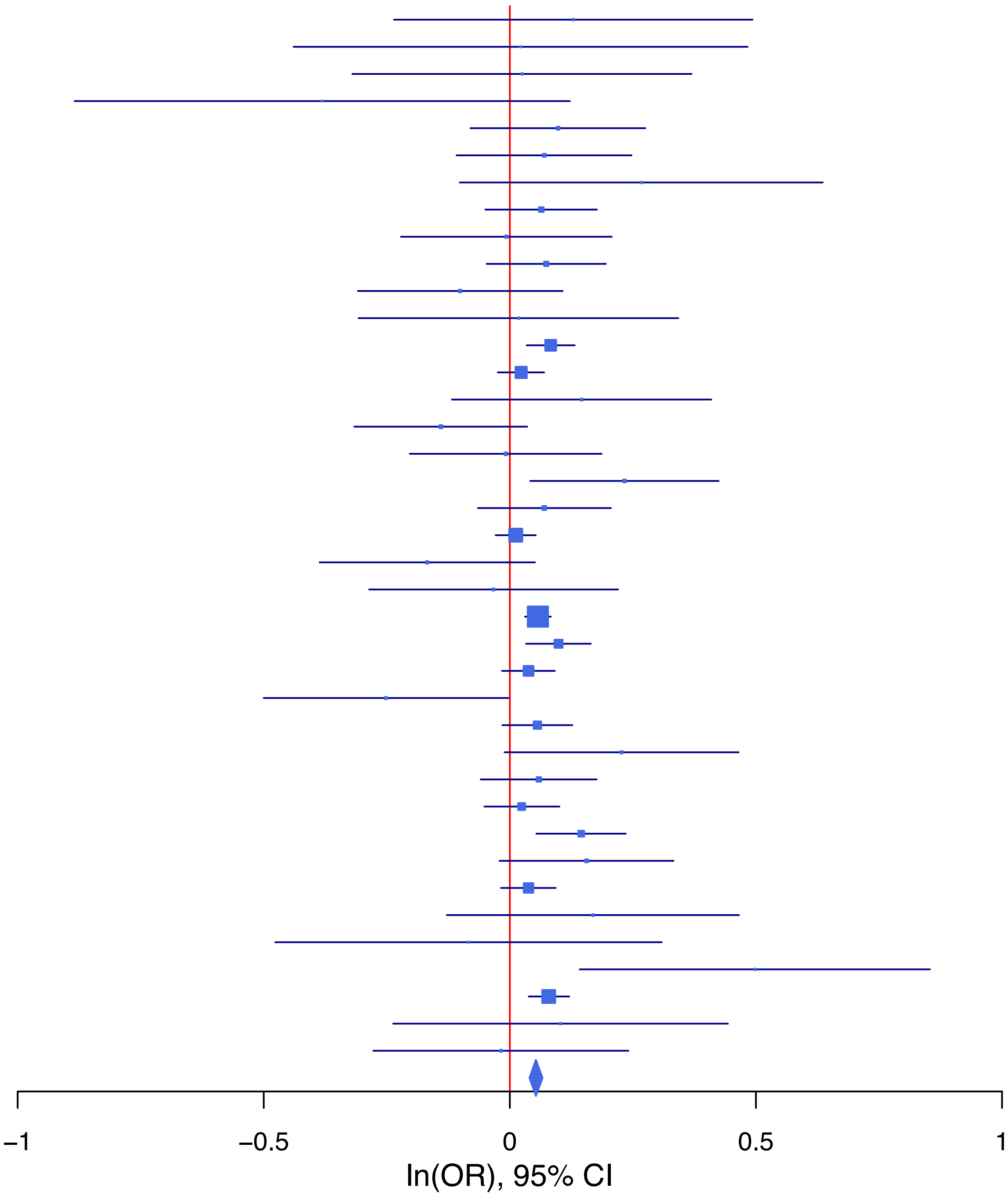

**rs55995895 T/C 7:1862183**

| Cohort | P | ln(OR) | SE |
|---|---|---|---|
| ADHEA_AFR | 0.6235 | 0.114 | 0.234 |
| ADHEA_EUR | 0.2072 | −0.121 | 0.096 |
| ALSPC_EUR | 0.7761 | 0.022 | 0.077 |
| BEPS7_EUR | 0.2763 | −0.254 | 0.233 |
| BOR17_EUR | 0.8961 | 0.014 | 0.11 |
| BOR2C_EUR | 0.324 | −0.11 | 0.111 |
| BOR2E_EUR | 0.4332 | −0.183 | 0.234 |
| CNVRG_EAS | 0.6745 | 0.032 | 0.077 |
| COGA1_EUR | 0.1027 | −0.132 | 0.081 |
| CUINT_EUR | 0.4381 | 0.1 | 0.129 |
| CVEDA_CSA | 0.9291 | 0.016 | 0.185 |
| ESTB2_EUR | 0.0063 | −0.074 | 0.027 |
| FINNG_EUR | 8e−04 | −0.09 | 0.027 |
| GISS1_EUR | 0.7848 | 0.025 | 0.091 |
| GISS2_EUR | 0.7959 | −0.027 | 0.103 |
| GREAT_EAS | 0.4212 | −0.113 | 0.141 |
| GTPRJ_AFR | 0.1375 | −0.217 | 0.146 |
| IPSYC_EUR | 0.004 | −0.081 | 0.028 |
| JANS3_EUR | 0.277 | −0.157 | 0.144 |
| JANS4_EUR | 0.0214 | −0.376 | 0.163 |
| MIREC_AFR | 0.408 | 0.257 | 0.311 |
| MIREC_EUR | 0.7999 | −0.053 | 0.208 |
| MVPXQ_AFR | 0.5758 | −0.027 | 0.048 |
| MVPXQ_EAS | 0.7762 | −0.045 | 0.159 |
| MVPXQ_EUR | 7e−04 | −0.06 | 0.018 |
| MVPXQ_LAT | 0.2092 | −0.06 | 0.048 |
| PGCBD_EUR | 0.1966 | 0.044 | 0.034 |
| PGCED_EUR | 0.5168 | −0.108 | 0.167 |
| PGCMD_EUR | 0.2842 | 0.049 | 0.046 |
| PGCPT_EUR | 0.2804 | −0.086 | 0.079 |
| PGCSZ_EUR | 0.3063 | 0.05 | 0.049 |
| PRFCT_EUR | 0.6729 | −0.024 | 0.057 |
| PSYCR_EUR | 0.0621 | −0.204 | 0.109 |
| QIMRB_EUR | 0.0018 | −0.115 | 0.037 |
| SNUBH−ASA_EAS | 0.2376 | −0.276 | 0.234 |
| SNUBH−KCHIP_EAS | 0.4724 | −0.228 | 0.317 |
| STRR1_LAT | 0.9055 | 0.029 | 0.247 |
| UKBJC_EUR | 0.1022 | −0.046 | 0.028 |
| YPENN_EUR | 0.4864 | −0.123 | 0.177 |
| **meta** | **3.32e−10** | **−0.053** | **0.009** |

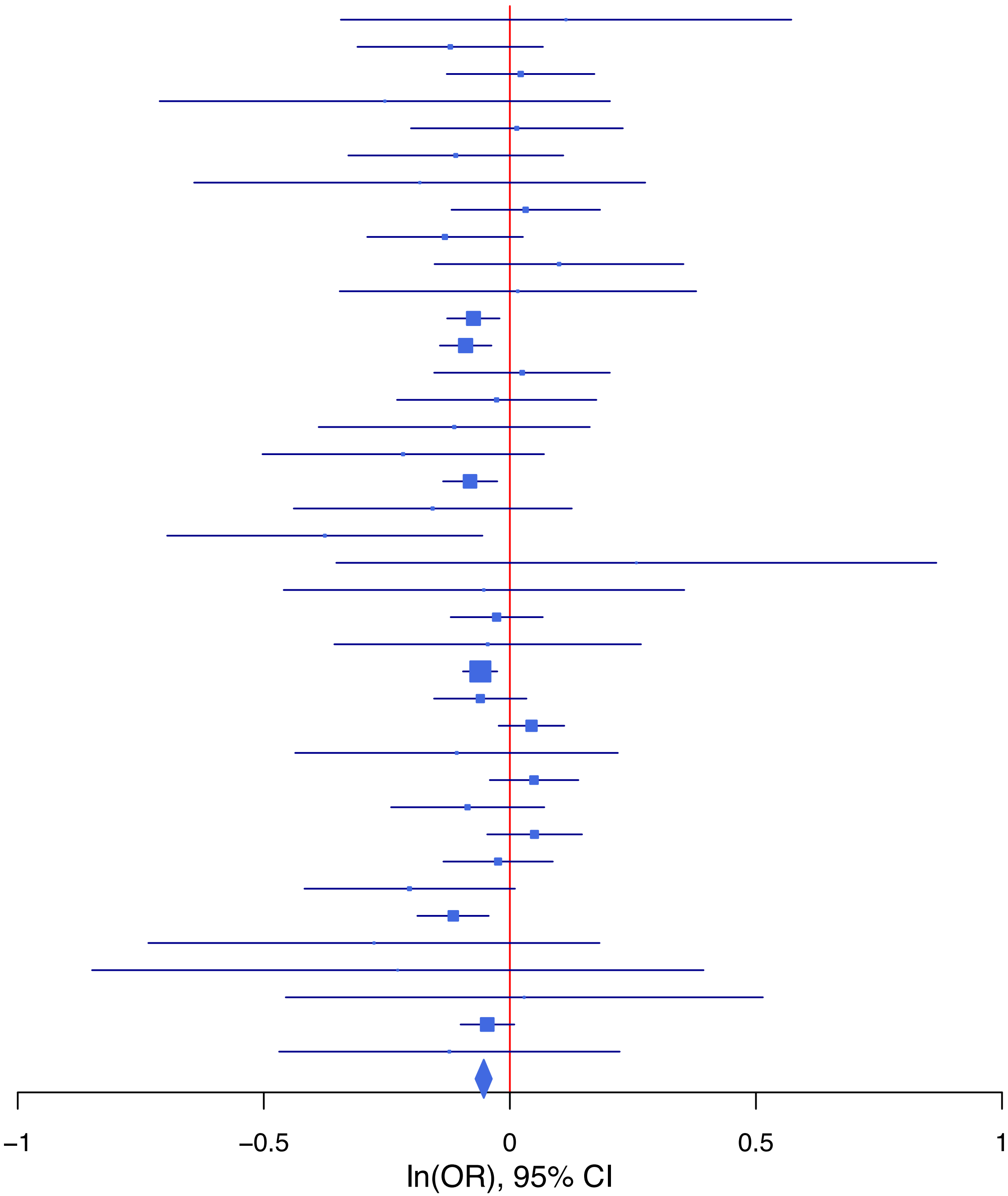

## rs6224 T/G 15:91423543

| Cohort | P | ln(OR) | SE |
|---|---|---|---|
| ALSPC_EUR | 0.2823 | −0.059 | 0.055 |
| BEPS7_EUR | 0.0632 | −0.315 | 0.17 |
| BHRCM_AFR | 0.5094 | −0.176 | 0.267 |
| BHRCM_EUR | 0.7509 | −0.049 | 0.154 |
| BOR17_EUR | 0.1344 | −0.127 | 0.085 |
| BOR2C_EUR | 0.9255 | 0.008 | 0.087 |
| BOR2E_EUR | 0.0685 | 0.317 | 0.174 |
| CNVRG_EAS | 0.4111 | −0.053 | 0.065 |
| COGA1_AFR | 0.2039 | 0.156 | 0.123 |
| COGA1_EUR | 0.4692 | −0.043 | 0.059 |
| CUINT_EUR | 0.938 | 0.008 | 0.099 |
| CVEDA_CSA | 0.8628 | −0.029 | 0.167 |
| ESTB2_EUR | 0.5981 | 0.012 | 0.023 |
| FINNG_EUR | 0.4387 | −0.018 | 0.023 |
| GEDIS_EUR | 0.7646 | 0.039 | 0.13 |
| GISS1_EUR | 0.0565 | −0.149 | 0.078 |
| GISS2_EUR | 0.3829 | −0.081 | 0.092 |
| GREAT_EAS | 0.8593 | −0.023 | 0.128 |
| GTPRJ_AFR | 0.9038 | 0.01 | 0.08 |
| IPSYC_EUR | 0.325 | −0.02 | 0.02 |
| JANS3_EUR | 0.3432 | 0.096 | 0.102 |
| JANS4_EUR | 0.2522 | −0.138 | 0.12 |
| MVPXQ_EAS | 0.7554 | −0.038 | 0.123 |
| MVPXQ_EUR | 0.0082 | −0.034 | 0.013 |
| MVPXQ_LAT | 0.13 | −0.055 | 0.036 |
| PGCBD_EUR | 0.3367 | −0.025 | 0.026 |
| PGCED_EUR | 0.2005 | −0.149 | 0.117 |
| PGCMD_EUR | 0.0288 | −0.077 | 0.035 |
| PGCPT_AFR | 0.897 | −0.019 | 0.144 |
| PGCPT_EUR | 0.4166 | −0.047 | 0.058 |
| PGCSZ_EUR | 0.3361 | −0.037 | 0.038 |
| PRFCT_EUR | 0.1755 | −0.06 | 0.044 |
| PSYCR_EUR | 0.7078 | 0.034 | 0.09 |
| QIMRB_EUR | 0.0039 | −0.079 | 0.027 |
| SNUBH−ASA_EAS | 0.0606 | −0.318 | 0.17 |
| SNUBH−KCHIP_EAS | 0.1091 | 0.383 | 0.239 |
| STRR1_LAT | 0.0734 | 0.33 | 0.184 |
| UKBJC_EUR | 4e−04 | −0.072 | 0.02 |
| YPENN_EUR | 0.7827 | −0.036 | 0.129 |
| **meta** | **2.37e−08** | **−0.037** | **0.007** |

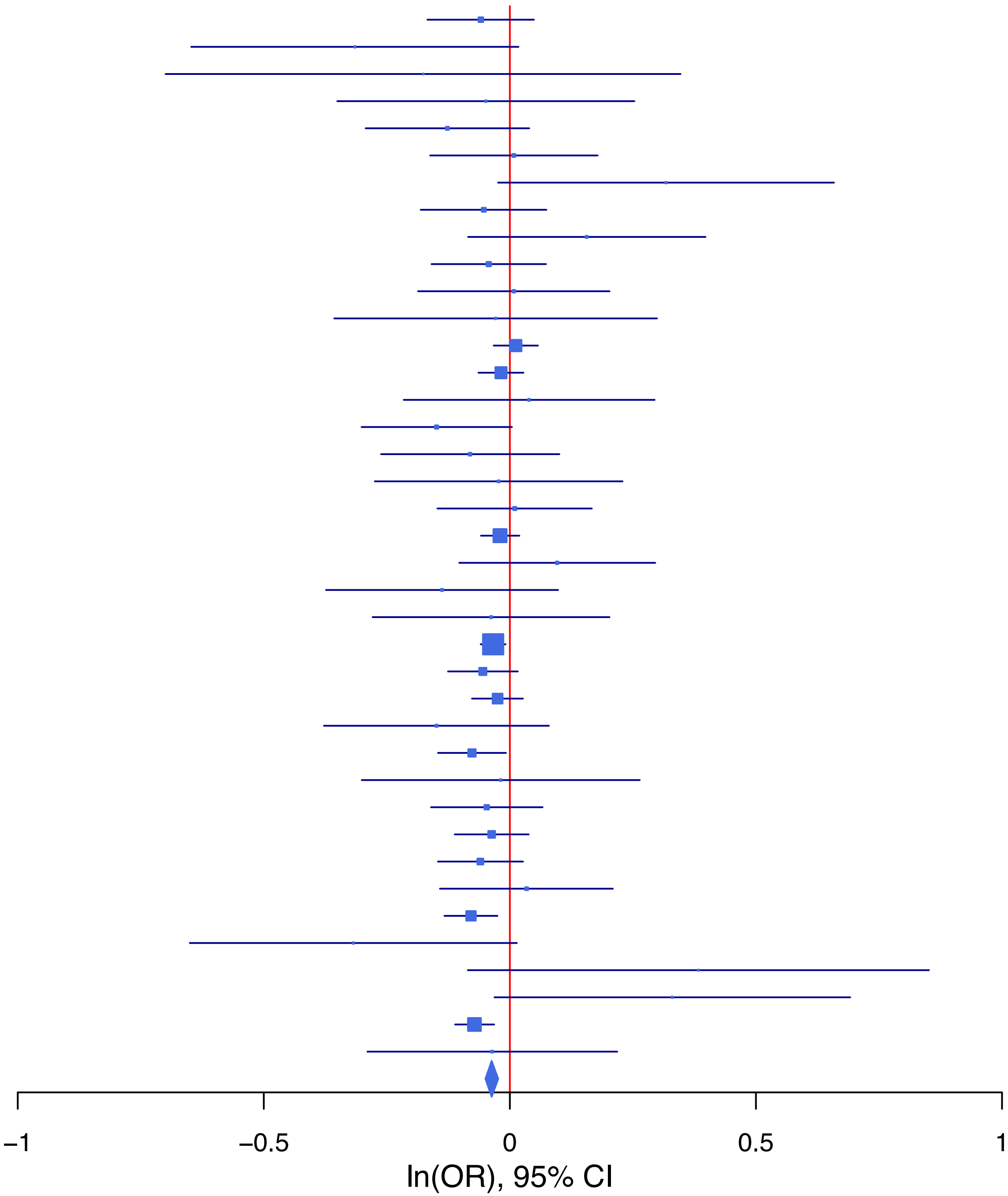

**rs62367522   C/A   5:45280212**

| Cohort | P | ln(OR) | SE |
|---|---|---|---|
| ALSPC_EUR | 0.1126 | −0.113 | 0.071 |
| BEPS7_EUR | 0.9049 | −0.027 | 0.226 |
| BHRCM_AFR | 0.3948 | 0.364 | 0.427 |
| BHRCM_LAT | 0.4087 | 0.223 | 0.27 |
| BOR17_EUR | 0.4751 | 0.078 | 0.11 |
| BOR2C_EUR | 0.2621 | −0.124 | 0.111 |
| BOR2E_EUR | 0.9648 | 0.01 | 0.228 |
| CNVRG_EAS | 0.9831 | 0.002 | 0.072 |
| COGA1_AFR | 0.9697 | 0.008 | 0.21 |
| COGA1_EUR | 0.8662 | 0.013 | 0.075 |
| CUINT_EUR | 0.5204 | −0.082 | 0.128 |
| CVEDA_CSA | 0.7873 | −0.054 | 0.199 |
| ESTB2_EUR | 0.0024 | −0.096 | 0.032 |
| FINNG_EUR | 0.4942 | −0.022 | 0.032 |
| GEDIS_EUR | 0.8482 | 0.033 | 0.173 |
| GISS1_EUR | 0.4336 | −0.077 | 0.098 |
| GISS2_EUR | 0.691 | 0.047 | 0.118 |
| GREAT_EAS | 0.5511 | −0.081 | 0.135 |
| GTPRJ_AFR | 0.5596 | −0.075 | 0.128 |
| IPSYC_EUR | 0.132 | −0.04 | 0.027 |
| JANS3_EUR | 0.8927 | −0.018 | 0.132 |
| JANS4_EUR | 0.4105 | 0.122 | 0.148 |
| MIREC_AFR | 0.1098 | −0.557 | 0.348 |
| MIREC_EUR | 0.8345 | 0.041 | 0.195 |
| MVPXQ_AFR | 0.5459 | −0.026 | 0.044 |
| MVPXQ_EAS | 0.8217 | 0.039 | 0.175 |
| MVPXQ_EUR | 9e−04 | −0.057 | 0.017 |
| MVPXQ_LAT | 0.0032 | −0.121 | 0.041 |
| PGCBD_EUR | 0.4479 | −0.025 | 0.034 |
| PGCED_EUR | 0.369 | 0.125 | 0.14 |
| PGCMD_EUR | 0.2249 | −0.056 | 0.046 |
| PGCPT_EUR | 0.6207 | −0.038 | 0.077 |
| PGCSZ_EUR | 0.9696 | 0.002 | 0.048 |
| PRFCT_EUR | 0.0507 | −0.117 | 0.06 |
| PSYCR_EUR | 0.8606 | 0.019 | 0.109 |
| QIMRB_EUR | 0.0191 | −0.081 | 0.035 |
| SNUBH-ASA_EAS | 0.1585 | 0.264 | 0.187 |
| SNUBH-KCHIP_EAS | 0.8831 | −0.035 | 0.239 |
| STRR1_LAT | 0.7769 | −0.06 | 0.213 |
| UKBJC_EUR | 0.0014 | −0.084 | 0.026 |
| YPENN_EUR | 0.5228 | −0.104 | 0.163 |
| **meta** | **2.05e−10** | **−0.054** | **0.008** |

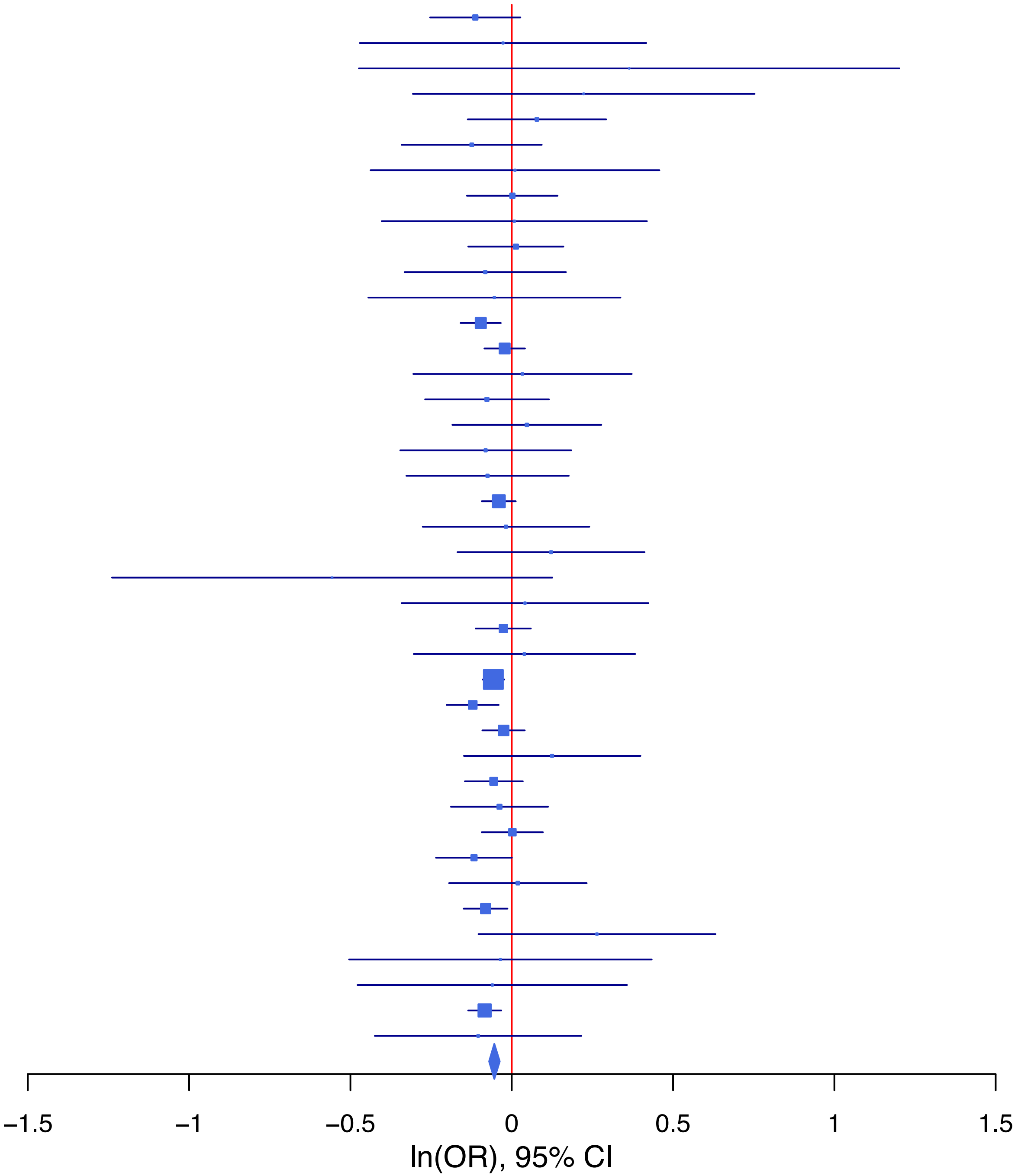

**rs62404522 C/T 6:19307114**

| Cohort | P | ln(OR) | SE |
|---|---|---|---|
| ADHEA_AFR | 0.1805 | −0.289 | 0.216 |
| ADHEA_EUR | 0.1413 | −0.154 | 0.105 |
| ALSPC_EUR | 0.2246 | 0.093 | 0.077 |
| BEPS7_EUR | 0.7783 | 0.07 | 0.25 |
| BHRCM_AFR | 0.9811 | 0.008 | 0.327 |
| BHRCM_EUR | 0.5224 | −0.166 | 0.26 |
| BHRCM_LAT | 0.9721 | −0.01 | 0.296 |
| BOR17_EUR | 0.9851 | −0.002 | 0.126 |
| BOR2C_EUR | 0.0556 | 0.245 | 0.128 |
| BOR2E_EUR | 0.3309 | −0.281 | 0.29 |
| CNVRG_EAS | 0.7083 | −0.025 | 0.066 |
| COGA1_AFR | 0.0497 | 0.329 | 0.168 |
| COGA1_EUR | 0.1278 | 0.133 | 0.087 |
| CUINT_EUR | 0.5468 | 0.089 | 0.147 |
| CVEDA_CSA | 0.5702 | 0.101 | 0.177 |
| ESTB2_EUR | 0.2609 | 0.038 | 0.034 |
| FINNG_EUR | 0.0012 | 0.11 | 0.034 |
| GEDIS_EUR | 0.1439 | 0.257 | 0.176 |
| GISS1_EUR | 0.728 | 0.04 | 0.116 |
| GISS2_EUR | 0.0643 | 0.272 | 0.147 |
| GREAT_EAS | 0.6291 | −0.054 | 0.111 |
| GTPRJ_AFR | 0.0903 | 0.161 | 0.095 |
| IPSYC_EUR | 0.0489 | 0.055 | 0.028 |
| JANS3_EUR | 0.2348 | 0.171 | 0.144 |
| JANS4_EUR | 0.5364 | −0.116 | 0.187 |
| MVPXQ_AFR | 0.0293 | 0.081 | 0.037 |
| MVPXQ_EAS | 0.5215 | −0.095 | 0.149 |
| MVPXQ_EUR | 1.26e−05 | 0.083 | 0.019 |
| MVPXQ_LAT | 0.0447 | 0.083 | 0.041 |
| PGCBD_EUR | 1e−04 | 0.146 | 0.037 |
| PGCPT_AFR | 0.3129 | −0.193 | 0.191 |
| PGCPT_EUR | 0.4313 | 0.063 | 0.08 |
| PGCSZ_EUR | 0.7464 | 0.018 | 0.054 |
| PRFCT_EUR | 0.0094 | 0.158 | 0.061 |
| PSYCR_EUR | 0.081 | −0.227 | 0.13 |
| QIMRB_EUR | 0.0417 | 0.081 | 0.04 |
| SNUBH-ASA_EAS | 0.9686 | −0.008 | 0.204 |
| SNUBH-KCHIP_EAS | 0.707 | 0.101 | 0.269 |
| STRR1_LAT | 0.9538 | −0.012 | 0.212 |
| UKBJC_EUR | 0.4223 | 0.024 | 0.03 |
| YPENN_EUR | 0.8099 | 0.045 | 0.188 |
| **meta** | **1.65e−14** | **0.07** | **0.009** |

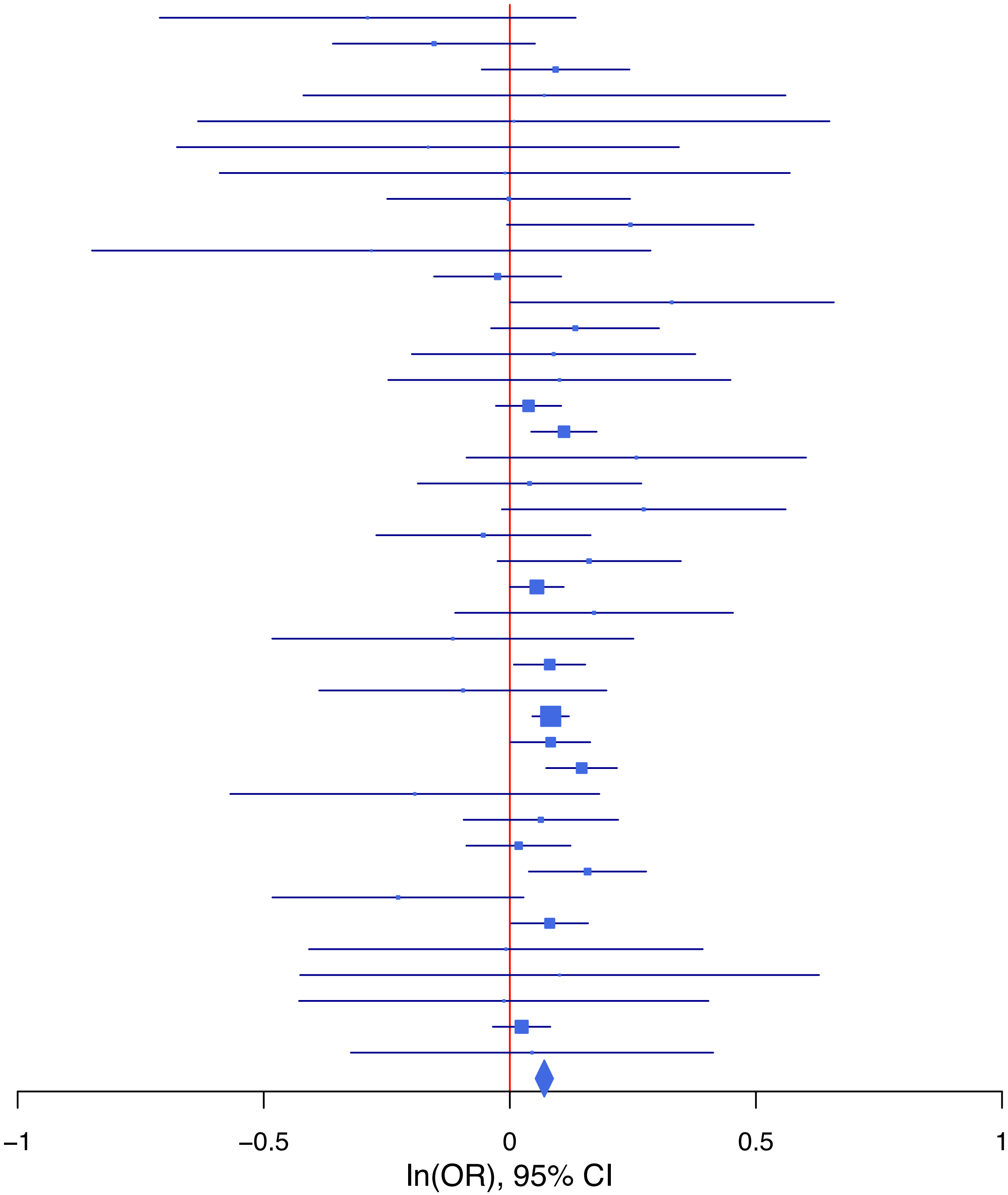

**rs640704 G/A 15:59049021**

| Cohort | P | ln(OR) | SE |
|---|---|---|---|
| ALSPC_EUR | 0.1963 | −0.077 | 0.06 |
| BEPS7_EUR | 0.8101 | 0.046 | 0.192 |
| BHRCM_AFR | 0.196 | −0.28 | 0.216 |
| BHRCM_EUR | 0.9895 | 0.002 | 0.185 |
| BOR17_EUR | 0.5758 | −0.053 | 0.094 |
| BOR2C_EUR | 0.8381 | 0.019 | 0.095 |
| BOR2E_EUR | 0.8457 | −0.041 | 0.211 |
| CNVRG_EAS | 0.1097 | −0.103 | 0.064 |
| COGA1_AFR | 0.8185 | 0.025 | 0.111 |
| CUINT_EUR | 0.5594 | −0.065 | 0.111 |
| CVEDA_CSA | 0.6399 | −0.078 | 0.166 |
| ESTB2_EUR | 0.0142 | −0.06 | 0.025 |
| GEDIS_EUR | 0.6767 | 0.059 | 0.141 |
| GISS1_EUR | 0.8981 | −0.011 | 0.085 |
| GISS2_EUR | 0.2417 | −0.118 | 0.101 |
| GREAT_EAS | 0.7992 | −0.032 | 0.129 |
| GTPRJ_AFR | 0.2012 | 0.086 | 0.067 |
| IPSYC_EUR | 0.0079 | −0.056 | 0.021 |
| JANS3_EUR | 0.8377 | −0.024 | 0.116 |
| JANS4_EUR | 0.4256 | 0.11 | 0.138 |
| MVPXQ_EAS | 0.889 | −0.018 | 0.131 |
| MVPXQ_EUR | 0.0076 | −0.038 | 0.014 |
| MVPXQ_LAT | 0.0979 | −0.06 | 0.036 |
| PGCBD_EUR | 0.0498 | −0.055 | 0.028 |
| PGCED_EUR | 0.526 | 0.079 | 0.125 |
| PGCMD_EUR | 0.9331 | −0.003 | 0.037 |
| PGCPT_AFR | 0.4327 | −0.09 | 0.115 |
| PGCPT_EUR | 0.9585 | 0.003 | 0.063 |
| PGCSZ_EUR | 0.0048 | −0.115 | 0.041 |
| PRFCT_EUR | 0.4335 | 0.036 | 0.047 |
| PSYCR_EUR | 0.6788 | 0.04 | 0.096 |
| QIMRB_EUR | 0.0729 | −0.053 | 0.03 |
| SNUBH−ASA_EAS | 0.3433 | −0.165 | 0.174 |
| STRR1_LAT | 0.1863 | −0.241 | 0.183 |
| UKBJC_EUR | 0.06 | −0.041 | 0.022 |
| YPENN_EUR | 0.8707 | 0.023 | 0.14 |
| **meta** | **5.47e−09** | **−0.043** | **0.007** |

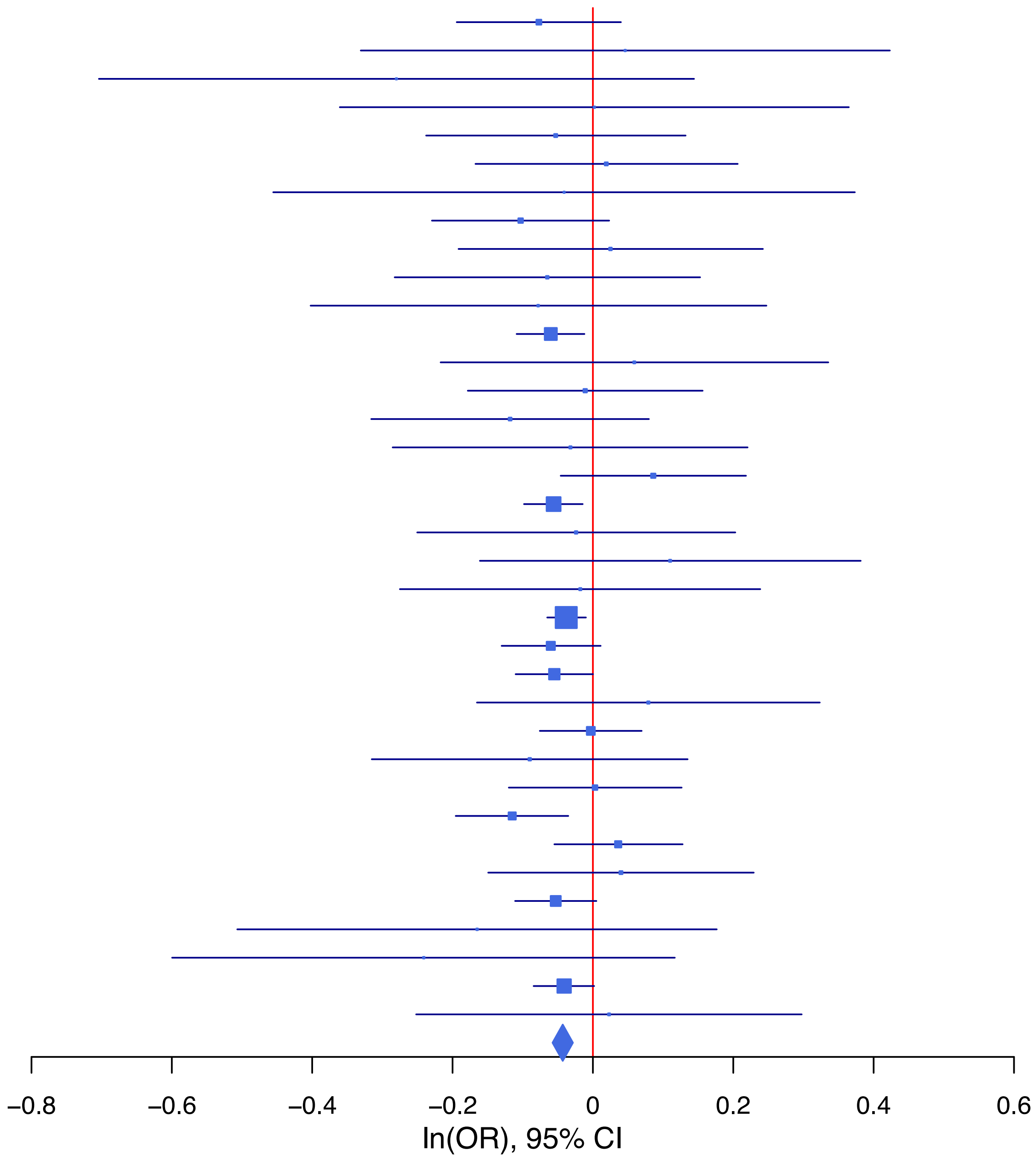

**rs6765986 C/A 3:56362210**

| Cohort | P | ln(OR) | SE |
|---|---|---|---|
| ADHEA_AFR | 0.5757 | 0.11 | 0.196 |
| ADHEA_EUR | 0.8898 | 0.01 | 0.074 |
| ALSPC_EUR | 0.2383 | 0.069 | 0.059 |
| BEPS7_EUR | 0.8414 | −0.041 | 0.203 |
| BHRCM_AFR | 0.9442 | −0.045 | 0.639 |
| BHRCM_EUR | 0.3315 | −0.195 | 0.201 |
| BHRCM_LAT | 0.6529 | 0.133 | 0.294 |
| BOR17_EUR | 0.1426 | 0.143 | 0.098 |
| BOR2C_EUR | 0.8039 | −0.024 | 0.098 |
| BOR2E_EUR | 0.1003 | 0.324 | 0.197 |
| CNVRG_EAS | 0.8965 | −0.01 | 0.074 |
| COGA1_AFR | 0.0683 | 0.317 | 0.174 |
| COGA1_EUR | 0.7055 | 0.024 | 0.064 |
| CUINT_EUR | 0.404 | 0.095 | 0.114 |
| CVEDA_CSA | 0.3933 | −0.194 | 0.227 |
| ESTB2_EUR | 0.0963 | 0.044 | 0.026 |
| FINNG_EUR | 0.458 | 0.018 | 0.025 |
| GEDIS_EUR | 0.262 | 0.154 | 0.137 |
| GISS1_EUR | 0.4913 | 0.063 | 0.092 |
| GISS2_EUR | 0.9727 | 0.004 | 0.108 |
| GREAT_EAS | 0.6709 | 0.045 | 0.106 |
| IPSYC_EUR | 9e−04 | 0.07 | 0.021 |
| JANS3_EUR | 0.6639 | −0.05 | 0.115 |
| JANS4_EUR | 0.5332 | 0.081 | 0.131 |
| MIREC_AFR | 0.47 | −0.191 | 0.265 |
| MIREC_EUR | 0.1953 | −0.226 | 0.174 |
| MVPXQ_AFR | 0.1519 | 0.054 | 0.037 |
| MVPXQ_EUR | 0.0602 | 0.026 | 0.014 |
| PGCBD_EUR | 0.2753 | 0.031 | 0.028 |
| PGCED_EUR | 0.8014 | −0.033 | 0.13 |
| PGCMD_EUR | 0.1313 | 0.057 | 0.038 |
| PGCPT_EUR | 0.6445 | −0.03 | 0.064 |
| PGCSZ_EUR | 0.4847 | 0.029 | 0.041 |
| PRFCT_EUR | 0.2434 | 0.055 | 0.048 |
| PSYCR_EUR | 0.8253 | 0.021 | 0.096 |
| QIMRB_EUR | 0.0552 | 0.058 | 0.03 |
| SNUBH−ASA_EAS | 0.6361 | −0.103 | 0.217 |
| SNUBH−KCHIP_EAS | 0.9293 | 0.026 | 0.29 |
| STRR1_LAT | 0.6082 | 0.1 | 0.196 |
| UKBJC_EUR | 0.0109 | 0.056 | 0.022 |
| YPENN_AFR | 0.9584 | 0.014 | 0.26 |
| YPENN_EUR | 0.8868 | −0.021 | 0.146 |
| **meta** | **2.17e−08** | **0.04** | **0.007** |

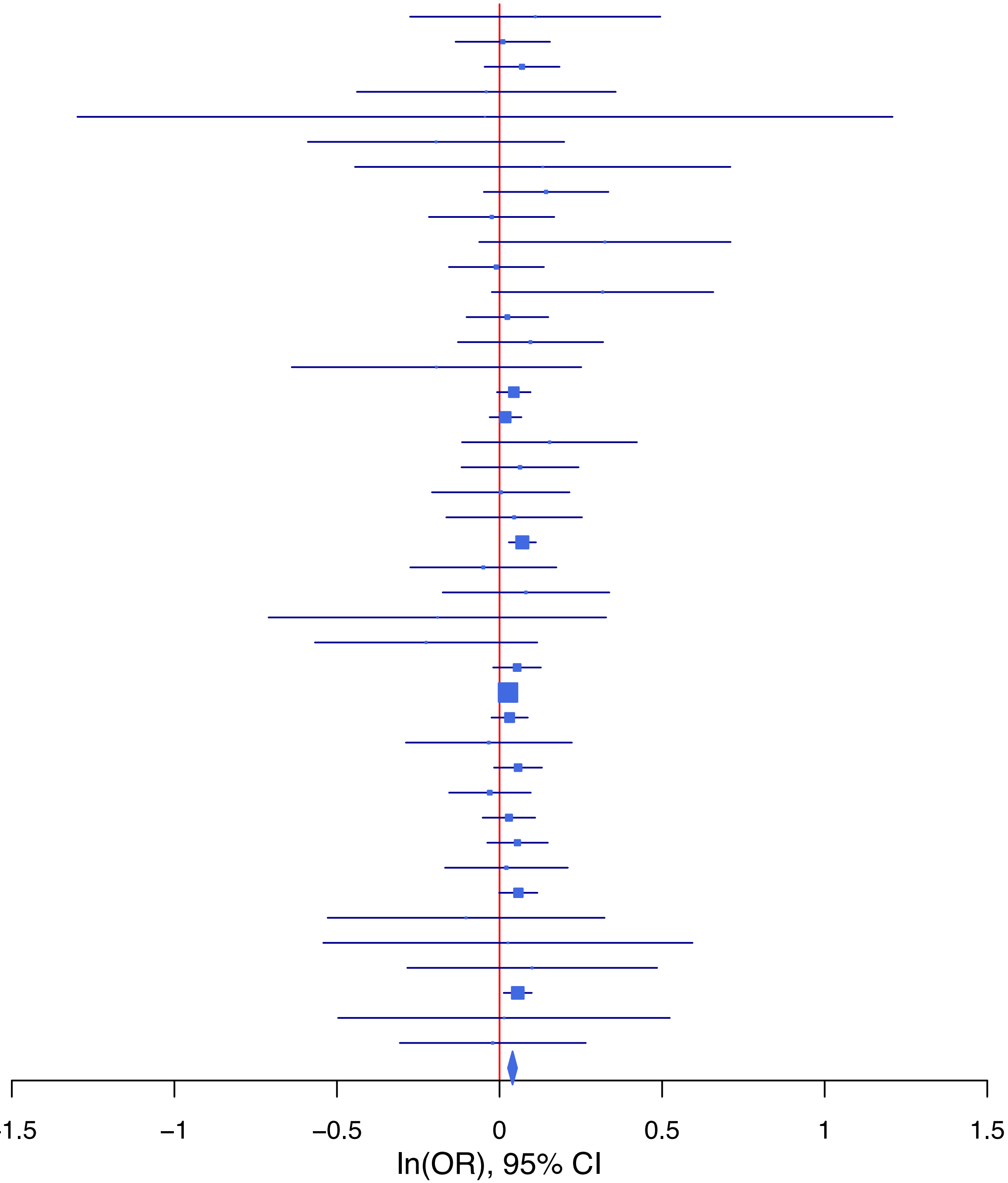

**rs7174904 T/C 15:47676110**

| Cohort | P | ln(OR) | SE |
|---|---|---|---|
| ADHEA_AFR | 0.5332 | −0.081 | 0.13 |
| ADHEA_EUR | 0.4549 | 0.06 | 0.08 |
| ALSPC_EUR | 0.5444 | 0.039 | 0.065 |
| BEPS7_EUR | 0.762 | 0.065 | 0.215 |
| BHRCM_AFR | 0.4377 | −0.172 | 0.221 |
| BHRCM_EUR | 0.5326 | −0.115 | 0.184 |
| BOR17_EUR | 0.2716 | 0.107 | 0.098 |
| BOR2C_EUR | 0.815 | −0.024 | 0.101 |
| BOR2E_EUR | 0.2723 | 0.217 | 0.197 |
| CNVRG_EAS | 0.2922 | 0.118 | 0.112 |
| COGA1_AFR | 0.7614 | 0.033 | 0.11 |
| COGA1_EUR | 0.4087 | 0.058 | 0.07 |
| CVEDA_CSA | 0.774 | −0.092 | 0.322 |
| ESTB2_EUR | 0.033 | 0.063 | 0.029 |
| GISS1_EUR | 0.7613 | −0.031 | 0.101 |
| GISS2_EUR | 0.0084 | 0.311 | 0.118 |
| GREAT_EAS | 0.3263 | −0.263 | 0.268 |
| GTPRJ_AFR | 0.2218 | 0.08 | 0.065 |
| IPSYC_EUR | 0.0153 | 0.057 | 0.023 |
| JANS3_EUR | 0.7426 | 0.04 | 0.123 |
| JANS4_EUR | 0.5265 | −0.094 | 0.149 |
| MIREC_AFR | 0.0174 | −0.395 | 0.166 |
| MIREC_EUR | 0.014 | 0.416 | 0.169 |
| MVPXQ_AFR | 0.4379 | 0.018 | 0.023 |
| MVPXQ_EAS | 0.3162 | −0.185 | 0.185 |
| MVPXQ_EUR | 0.0682 | 0.028 | 0.015 |
| MVPXQ_LAT | 0.5714 | −0.023 | 0.04 |
| PGCBD_EUR | 0.0144 | 0.075 | 0.031 |
| PGCED_EUR | 0.1337 | 0.195 | 0.13 |
| PGCMD_EUR | 0.0327 | 0.086 | 0.04 |
| PGCPT_AFR | 0.1093 | 0.172 | 0.108 |
| PGCPT_EUR | 0.1523 | −0.1 | 0.07 |
| PGCSZ_EUR | 7e−04 | 0.148 | 0.043 |
| PRFCT_EUR | 0.1093 | −0.084 | 0.053 |
| PSYCR_EUR | 0.8528 | 0.019 | 0.105 |
| QIMRB_EUR | 4e−04 | 0.117 | 0.033 |
| STRR1_LAT | 0.7217 | 0.078 | 0.218 |
| UKBJC_EUR | 0.0012 | 0.079 | 0.024 |
| YPENN_EUR | 0.8397 | −0.032 | 0.157 |
| **meta** | **5.31e−10** | **0.047** | **0.008** |

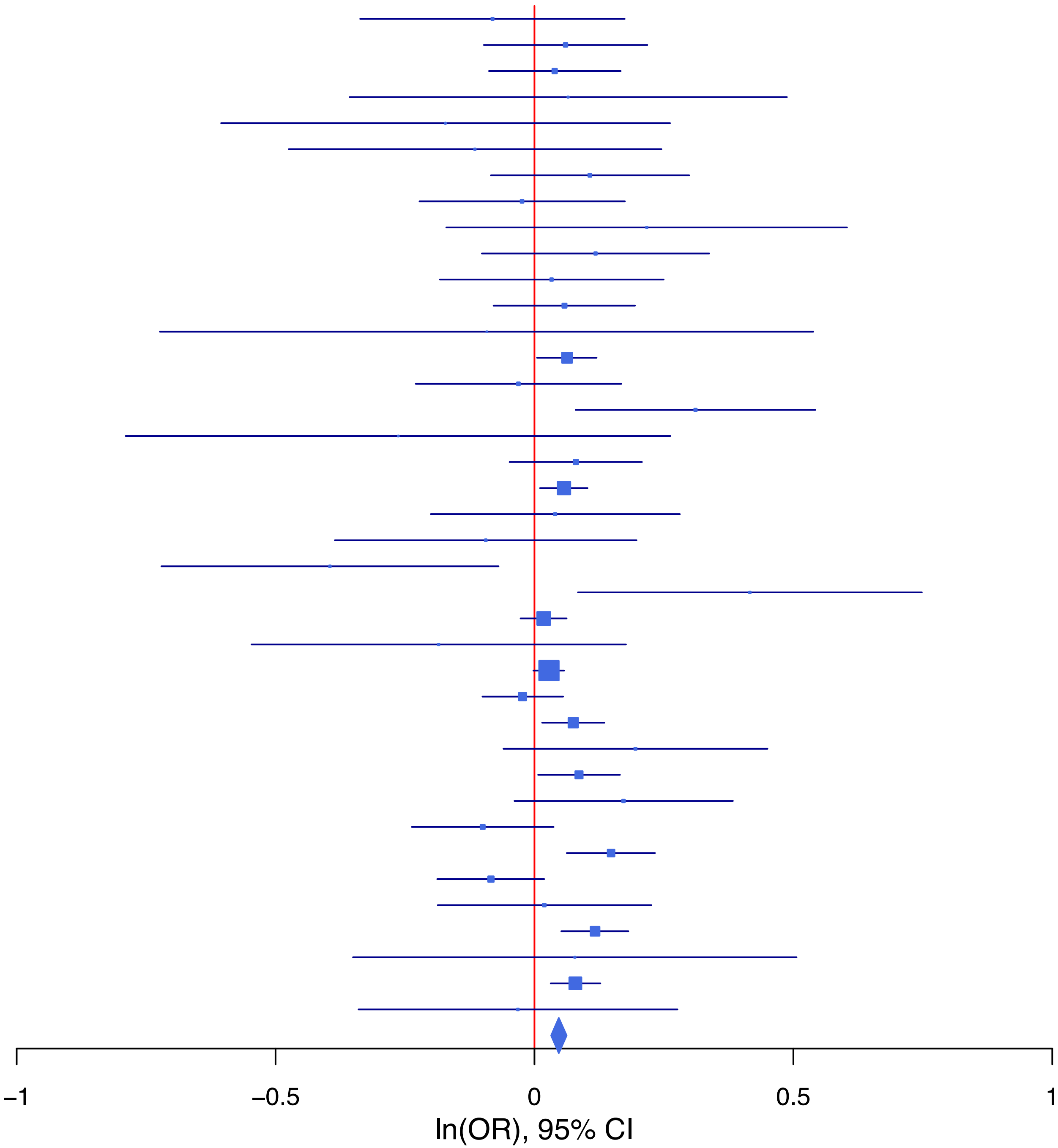

**rs720023 T/A 11:112838867**

| Cohort | P | ln(OR) | SE |
|---|---|---|---|
| ADHEA_AFR | 0.3805 | 0.099 | 0.113 |
| ADHEA_EUR | 0.4464 | 0.053 | 0.069 |
| ALSPC_EUR | 0.6248 | −0.027 | 0.055 |
| BEPS7_EUR | 0.5792 | 0.107 | 0.194 |
| BHRCM_AFR | 0.3664 | 0.163 | 0.18 |
| BHRCM_EUR | 0.9287 | 0.015 | 0.17 |
| BHRCM_LAT | 0.8843 | −0.037 | 0.257 |
| BOR17_EUR | 0.7325 | 0.03 | 0.087 |
| BOR2C_EUR | 0.091 | 0.153 | 0.09 |
| BOR2E_EUR | 0.3849 | −0.177 | 0.204 |
| CNVRG_EAS | 0.6436 | 0.027 | 0.058 |
| COGA1_AFR | 0.9138 | −0.011 | 0.102 |
| COGA1_EUR | 0.4085 | 0.049 | 0.059 |
| CUINT_EUR | 0.0547 | 0.192 | 0.1 |
| CVEDA_CSA | 0.5212 | 0.106 | 0.164 |
| ESTB2_EUR | 0.0661 | 0.042 | 0.023 |
| FINNG_EUR | 0.1466 | 0.034 | 0.024 |
| GEDIS_EUR | 0.8247 | −0.03 | 0.136 |
| GISS1_EUR | 0.4595 | −0.058 | 0.078 |
| GISS2_EUR | 0.0716 | 0.173 | 0.096 |
| GREAT_EAS | 0.6277 | −0.054 | 0.112 |
| GTPRJ_AFR | 0.7037 | 0.023 | 0.059 |
| IPSYC_EUR | 0.0397 | 0.042 | 0.021 |
| JANS3_EUR | 0.0797 | 0.185 | 0.106 |
| JANS4_EUR | 0.4795 | 0.087 | 0.123 |
| MVPXQ_EUR | 3.13e−06 | 0.062 | 0.013 |
| MVPXQ_LAT | 0.2565 | 0.043 | 0.038 |
| PGCBD_EUR | 0.0811 | 0.046 | 0.026 |
| PGCED_EUR | 0.2971 | 0.12 | 0.115 |
| PGCMD_EUR | 0.5445 | 0.021 | 0.035 |
| PGCPT_AFR | 0.1724 | 0.142 | 0.104 |
| PGCPT_EUR | 0.3729 | −0.052 | 0.059 |
| PGCSZ_EUR | 0.1041 | 0.06 | 0.037 |
| PRFCT_EUR | 0.6032 | −0.023 | 0.044 |
| PSYCR_EUR | 0.3994 | 0.076 | 0.091 |
| QIMRB_EUR | 0.1819 | 0.038 | 0.029 |
| SNUBH-ASA_EAS | 0.1487 | −0.207 | 0.144 |
| SNUBH-KCHIP_EAS | 0.9086 | 0.025 | 0.215 |
| STRR1_LAT | 0.9836 | 0.004 | 0.175 |
| UKBJC_EUR | 0.0474 | 0.041 | 0.021 |
| YPENN_EUR | 0.9861 | −0.002 | 0.128 |
| **meta** | **3.87e−11** | **0.044** | **0.007** |

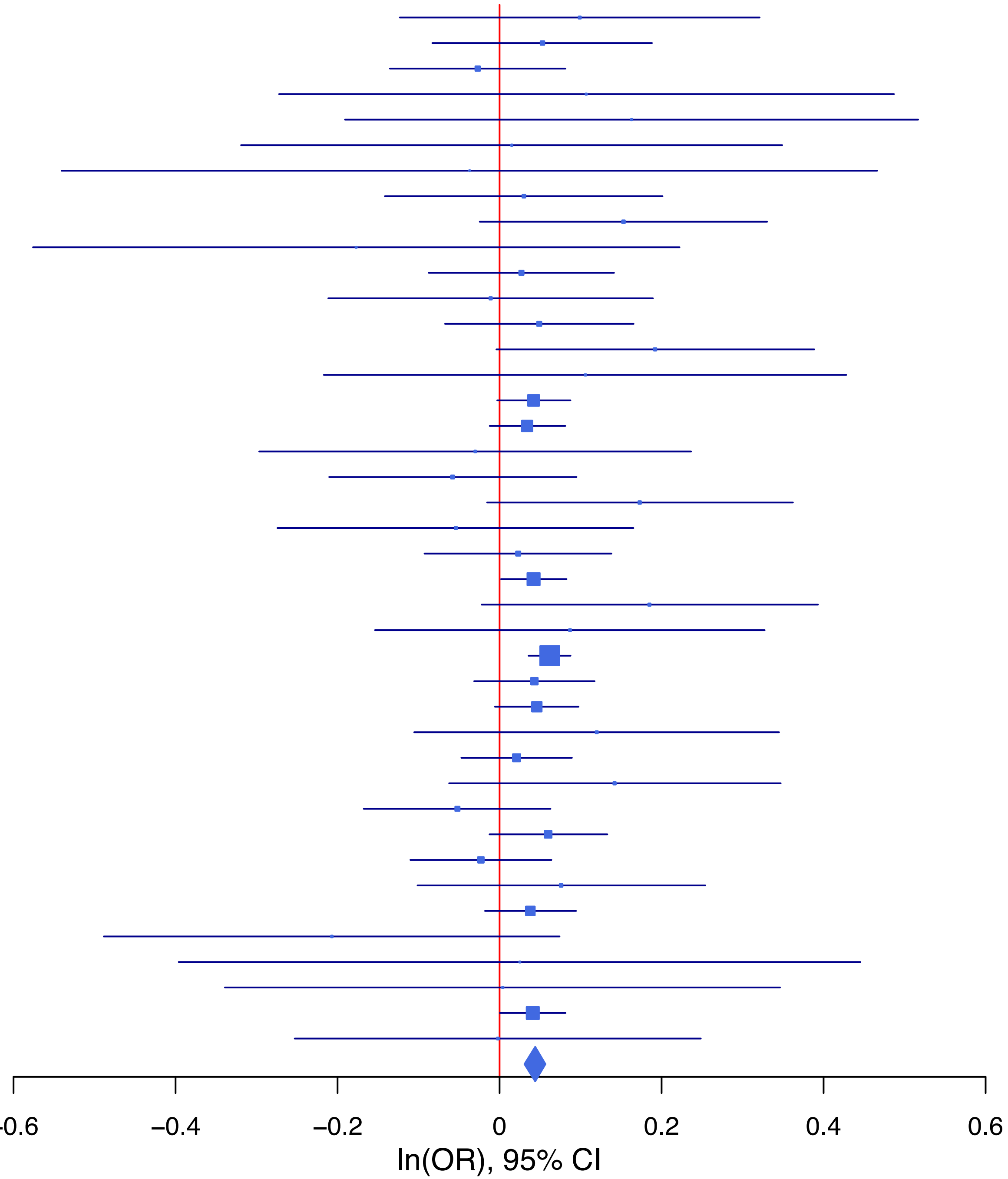

**rs73581580 G/A 9:140251458**

| Cohort | P | ln(OR) | SE |
|---|---|---|---|
| BEPS7_EUR | 0.4242 | 0.218 | 0.273 |
| BHRCM_AFR | 0.2967 | −0.257 | 0.246 |
| BHRCM_EUR | 0.2504 | 0.417 | 0.362 |
| BOR17_EUR | 0.1072 | −0.2 | 0.124 |
| BOR2C_EUR | 0.348 | −0.124 | 0.132 |
| BOR2E_EUR | 0.3079 | −0.294 | 0.288 |
| CNVRG_EAS | 0.3738 | −0.101 | 0.114 |
| COGA1_EUR | 0.1899 | −0.12 | 0.092 |
| ESTB2_EUR | 2e−04 | −0.121 | 0.032 |
| FINNG_EUR | 0.1796 | −0.04 | 0.03 |
| GISS1_EUR | 0.1752 | −0.176 | 0.13 |
| GISS2_EUR | 0.545 | −0.095 | 0.157 |
| GTPRJ_AFR | 0.5309 | −0.053 | 0.084 |
| JANS3_EUR | 0.1097 | 0.369 | 0.23 |
| JANS4_EUR | 0.6193 | 0.105 | 0.212 |
| MVPXQ_EUR | 0.0629 | −0.041 | 0.022 |
| MVPXQ_LAT | 0.7778 | −0.021 | 0.076 |
| PGCBD_EUR | 0.9134 | 0.006 | 0.059 |
| PGCMD_EUR | 0.4458 | −0.059 | 0.078 |
| PGCPT_AFR | 0.2239 | 0.175 | 0.144 |
| PGCPT_EUR | 0.3355 | −0.079 | 0.082 |
| PGCSZ_EUR | 0.5747 | 0.051 | 0.091 |
| PRFCT_EUR | 0.9844 | −0.001 | 0.06 |
| PSYCR_EUR | 0.3494 | −0.117 | 0.125 |
| QIMRB_EUR | 0.0291 | −0.09 | 0.041 |
| SNUBH−ASA_EAS | 0.759 | 0.109 | 0.354 |
| STRR1_LAT | 0.5432 | 0.209 | 0.344 |
| UKBJC_EUR | 1e−04 | −0.118 | 0.03 |
| YPENN_AFR | 0.3277 | −0.202 | 0.206 |
| YPENN_EUR | 0.3871 | 0.186 | 0.215 |
| **meta** | **1.72e−08** | **−0.064** | **0.011** |

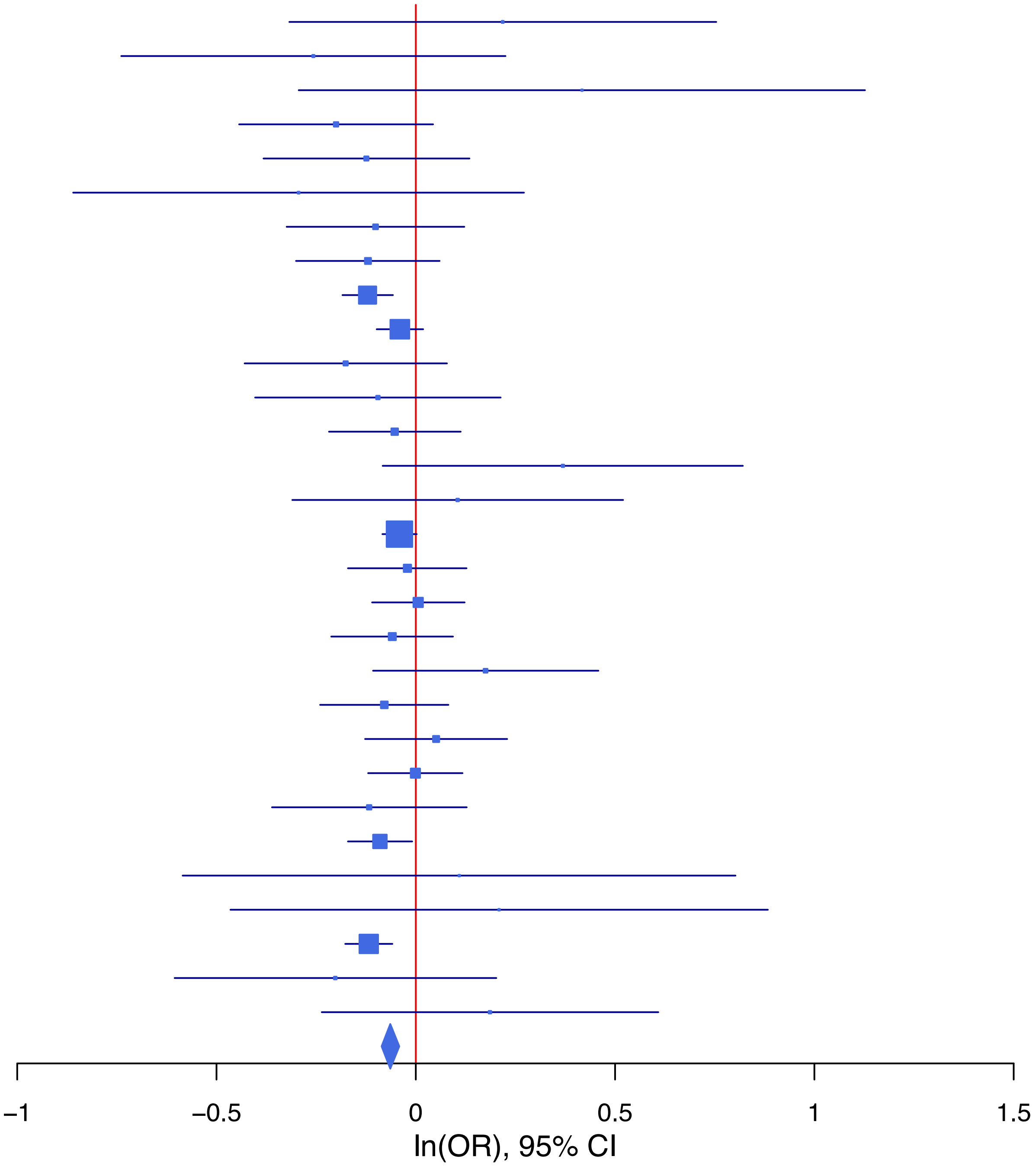

**rs7534143 G/T 1:66470379**

| Cohort | P | ln(OR) | SE |
|---|---|---|---|
| ADHEA_AFR | 0.6915 | 0.048 | 0.12 |
| ADHEA_EUR | 0.4575 | 0.05 | 0.068 |
| ALSPC_EUR | 0.6252 | 0.027 | 0.055 |
| BEPS7_EUR | 0.6307 | 0.085 | 0.176 |
| BHRCM_EUR | 0.4344 | 0.124 | 0.159 |
| BHRCM_LAT | 0.5339 | 0.202 | 0.324 |
| BOR17_EUR | 0.1322 | −0.127 | 0.084 |
| BOR2C_EUR | 0.069 | 0.159 | 0.087 |
| BOR2E_EUR | 0.4863 | −0.123 | 0.176 |
| CNVRG_EAS | 0.7347 | −0.018 | 0.052 |
| COGA1_AFR | 0.898 | 0.014 | 0.112 |
| COGA1_EUR | 0.993 | −0.001 | 0.059 |
| CUINT_EUR | 0.8914 | 0.014 | 0.099 |
| CVEDA_CSA | 0.8865 | −0.026 | 0.183 |
| ESTB2_EUR | 0.0854 | 0.039 | 0.023 |
| FINNG_EUR | 0.083 | 0.04 | 0.023 |
| GEDIS_EUR | 0.5672 | 0.073 | 0.127 |
| GISS1_EUR | 0.2748 | −0.085 | 0.078 |
| GISS2_EUR | 0.1579 | −0.129 | 0.091 |
| GREAT_EAS | 0.9769 | 0.003 | 0.094 |
| GTPRJ_AFR | 0.8987 | 0.008 | 0.064 |
| IPSYC_EUR | 4.24e−06 | 0.092 | 0.02 |
| JANS4_EUR | 0.8988 | 0.014 | 0.112 |
| MIREC_EUR | 0.5374 | −0.093 | 0.151 |
| MVPXQ_EUR | 0.0024 | 0.04 | 0.013 |
| PGCBD_EUR | 0.0734 | 0.046 | 0.026 |
| PGCED_EUR | 0.5243 | 0.074 | 0.116 |
| PGCMD_EUR | 0.0849 | 0.059 | 0.034 |
| PGCPT_AFR | 0.5388 | 0.066 | 0.107 |
| PGCPT_EUR | 0.7423 | 0.019 | 0.058 |
| PGCSZ_EUR | 0.2049 | 0.047 | 0.037 |
| PRFCT_EUR | 0.0111 | 0.111 | 0.044 |
| PSYCR_EUR | 0.925 | 0.008 | 0.088 |
| QIMRB_EUR | 0.8411 | 0.006 | 0.027 |
| SNUBH−ASA_EAS | 0.4818 | 0.106 | 0.15 |
| SNUBH−KCHIP_EAS | 0.9455 | −0.014 | 0.2 |
| STRR1_LAT | 0.3854 | −0.156 | 0.179 |
| UKBJC_EUR | 0.0141 | 0.05 | 0.02 |
| YPENN_EUR | 0.1157 | −0.202 | 0.128 |
| **meta** | **2.28e−10** | **0.042** | **0.007** |

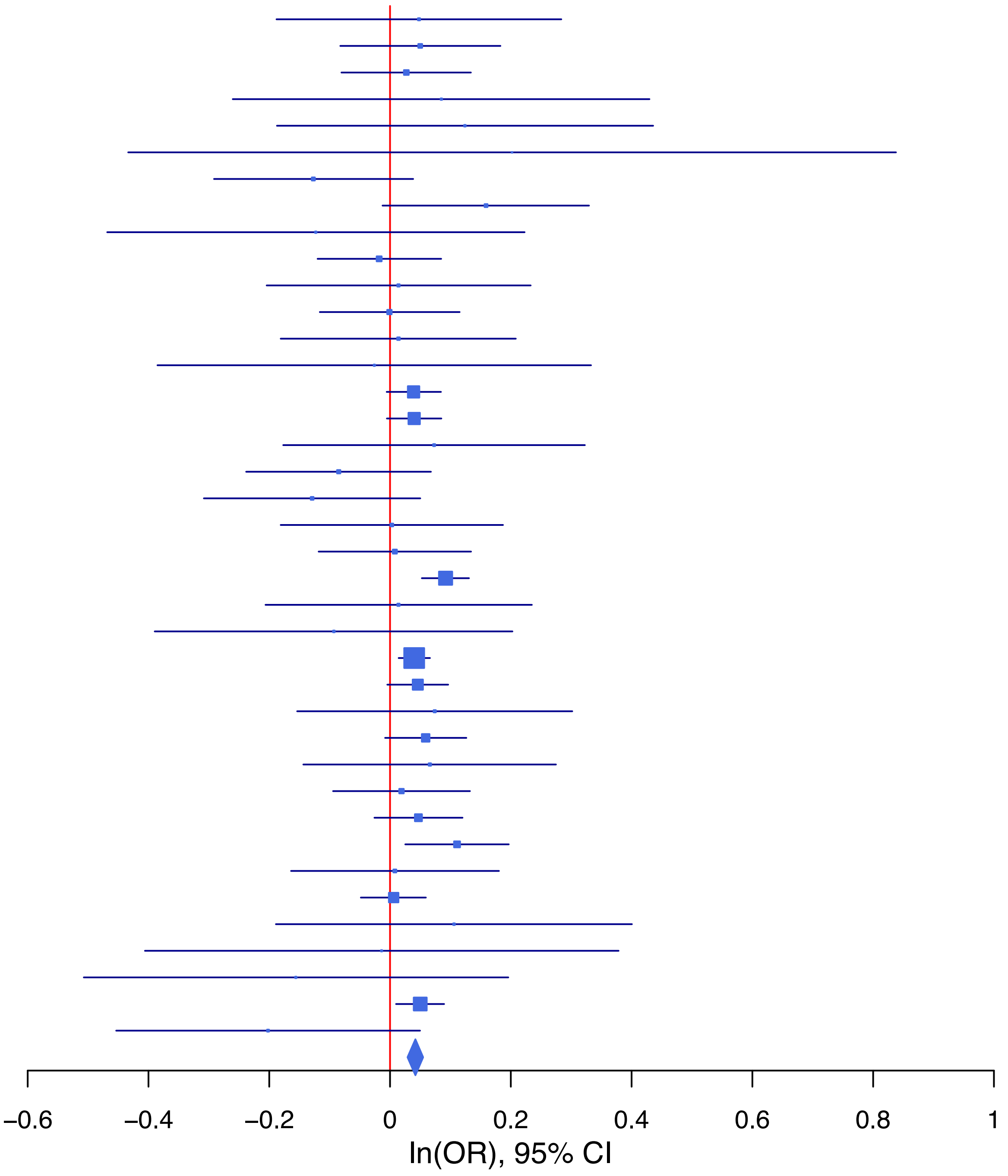

**rs78940908 G/C 2:58921049**

| Cohort | P | ln(OR) | SE |
|---|---|---|---|
| ADHEA_AFR | 0.3614 | −0.108 | 0.119 |
| ADHEA_EUR | 0.0089 | −0.183 | 0.07 |
| BEPS7_EUR | 0.9368 | −0.015 | 0.185 |
| BHRCM_AFR | 0.062 | −0.382 | 0.204 |
| BHRCM_LAT | 0.8783 | 0.037 | 0.241 |
| BOR17_EUR | 0.5683 | −0.05 | 0.087 |
| BOR2C_EUR | 0.6002 | −0.046 | 0.087 |
| BOR2E_EUR | 0.3827 | −0.156 | 0.179 |
| CNVRG_EAS | 0.5883 | 0.026 | 0.048 |
| COGA1_AFR | 0.1021 | −0.167 | 0.102 |
| COGA1_EUR | 0.5856 | −0.032 | 0.059 |
| CUINT_EUR | 0.768 | 0.031 | 0.104 |
| ESTB2_EUR | 0.1372 | −0.035 | 0.023 |
| FINNG_EUR | 0.7245 | 0.008 | 0.024 |
| GEDIS_EUR | 0.5801 | 0.071 | 0.129 |
| GISS1_EUR | 0.6598 | 0.035 | 0.08 |
| GISS2_EUR | 0.4138 | −0.076 | 0.093 |
| GREAT_EAS | 0.9599 | −0.004 | 0.076 |
| GTPRJ_AFR | 0.4787 | 0.042 | 0.059 |
| IPSYC_EUR | 0.0058 | −0.055 | 0.02 |
| JANS3_EUR | 0.7527 | 0.034 | 0.106 |
| JANS4_EUR | 0.9779 | −0.003 | 0.118 |
| MVPXQ_EUR | 0.004 | −0.038 | 0.013 |
| PGCBD_EUR | 0.239 | −0.031 | 0.026 |
| PGCED_EUR | 0.4613 | 0.083 | 0.113 |
| PGCMD_EUR | 0.5107 | −0.023 | 0.035 |
| PGCPT_AFR | 0.0962 | −0.171 | 0.103 |
| PGCPT_EUR | 0.264 | −0.065 | 0.058 |
| PGCSZ_EUR | 0.4735 | −0.027 | 0.037 |
| PRFCT_EUR | 0.5112 | −0.029 | 0.044 |
| PSYCR_EUR | 0.7609 | −0.026 | 0.087 |
| QIMRB_EUR | 0.0193 | −0.064 | 0.028 |
| SNUBH−ASA_EAS | 0.0603 | −0.243 | 0.13 |
| SNUBH−KCHIP_EAS | 0.3454 | −0.16 | 0.17 |
| STRR1_LAT | 0.2594 | 0.197 | 0.174 |
| UKBJC_EUR | 0.0079 | −0.054 | 0.021 |
| YPENN_AFR | 0.1816 | −0.197 | 0.148 |
| YPENN_EUR | 0.4275 | 0.103 | 0.13 |
| **meta** | **2.70e−08** | **−0.037** | **0.007** |

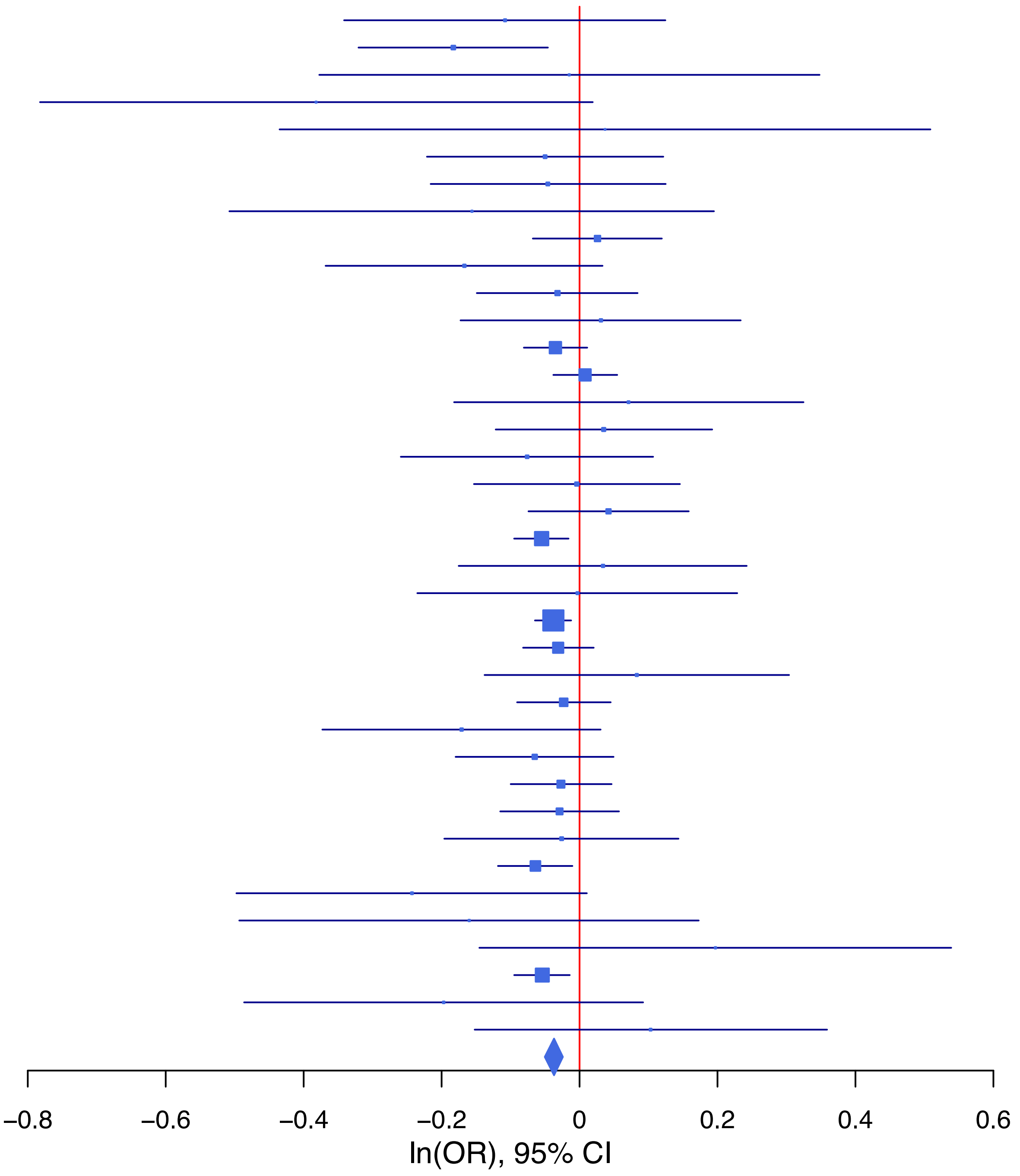

**rs8002865 A/G 13:96895532**

| Cohort | P | ln(OR) | SE |
|---|---|---|---|
| ADHEA_AFR | 0.0833 | −0.205 | 0.118 |
| ADHEA_EUR | 0.1755 | −0.092 | 0.068 |
| ALSPC_EUR | 0.8616 | 0.009 | 0.054 |
| BEPS7_EUR | 0.2936 | 0.183 | 0.174 |
| BHRCM_AFR | 0.9807 | 0.005 | 0.199 |
| BHRCM_EUR | 0.6314 | −0.074 | 0.155 |
| BOR17_EUR | 0.5151 | −0.055 | 0.084 |
| BOR2C_EUR | 0.9306 | 0.008 | 0.088 |
| BOR2E_EUR | 0.0252 | 0.418 | 0.187 |
| CNVRG_EAS | 0.0204 | −0.107 | 0.046 |
| COGA1_AFR | 0.4875 | 0.069 | 0.099 |
| COGA1_EUR | 0.9416 | 0.004 | 0.058 |
| CUINT_EUR | 0.712 | 0.037 | 0.101 |
| CVEDA_CSA | 0.5186 | −0.102 | 0.157 |
| ESTB2_EUR | 0.1099 | −0.036 | 0.023 |
| FINNG_EUR | 0.2239 | −0.028 | 0.023 |
| GEDIS_EUR | 0.9637 | 0.006 | 0.134 |
| GISS1_EUR | 0.3102 | 0.079 | 0.078 |
| GISS2_EUR | 0.5043 | 0.062 | 0.093 |
| GREAT_EAS | 0.1434 | 0.11 | 0.075 |
| IPSYC_EUR | 0.0025 | −0.06 | 0.02 |
| JANS3_EUR | 0.8893 | −0.014 | 0.102 |
| JANS4_EUR | 0.3797 | 0.104 | 0.119 |
| MVPXQ_AFR | 0.2692 | −0.023 | 0.02 |
| MVPXQ_EAS | 0.9531 | −0.006 | 0.096 |
| MVPXQ_EUR | 0.1049 | −0.02 | 0.013 |
| MVPXQ_LAT | 0.3007 | −0.036 | 0.035 |
| PGCBD_EUR | 0.0636 | −0.048 | 0.026 |
| PGCED_EUR | 0.1578 | 0.16 | 0.113 |
| PGCMD_EUR | 0.5543 | −0.02 | 0.034 |
| PGCPT_EUR | 0.1111 | −0.092 | 0.057 |
| PGCSZ_EUR | 0.8732 | −0.006 | 0.037 |
| PRFCT_EUR | 0.1133 | −0.07 | 0.044 |
| PSYCR_EUR | 0.6498 | 0.04 | 0.088 |
| QIMRB_EUR | 0.0012 | −0.089 | 0.027 |
| SNUBH−ASA_EAS | 0.1762 | −0.167 | 0.123 |
| SNUBH−KCHIP_EAS | 0.2072 | −0.209 | 0.166 |
| STRR1_LAT | 0.1309 | −0.281 | 0.186 |
| UKBJC_EUR | 0.0165 | −0.049 | 0.02 |
| YPENN_AFR | 0.5594 | −0.087 | 0.149 |
| YPENN_EUR | 0.5569 | −0.076 | 0.129 |
| **meta** | **1.95e−08** | **−0.034** | **0.006** |

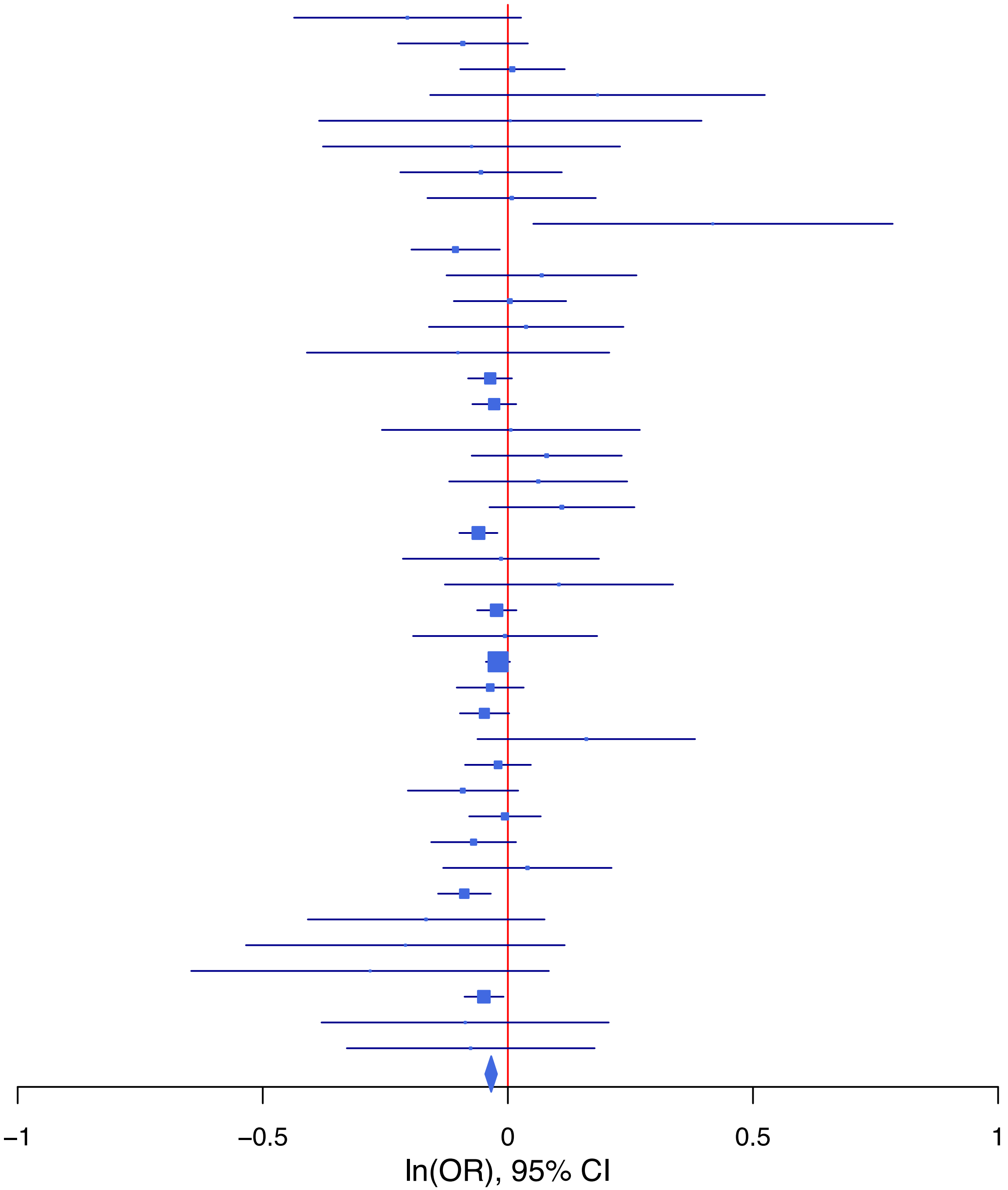

**rs8013266 C/T 14:103326780**

| Cohort | P | ln(OR) | SE |
|---|---|---|---|
| ALSPC_EUR | 0.009 | 0.169 | 0.065 |
| BEPS7_EUR | 0.8619 | 0.039 | 0.226 |
| BHRCM_AFR | 0.3574 | 0.241 | 0.261 |
| BHRCM_EUR | 0.8759 | 0.031 | 0.204 |
| BHRCM_LAT | 0.0641 | 0.434 | 0.234 |
| BOR17_EUR | 0.6035 | 0.056 | 0.109 |
| BOR2C_EUR | 0.4965 | −0.074 | 0.108 |
| BOR2E_EUR | 0.4177 | −0.168 | 0.208 |
| CNVRG_EAS | 0.0635 | 0.086 | 0.046 |
| COGA1_AFR | 0.5206 | −0.069 | 0.107 |
| CUINT_EUR | 0.9156 | −0.013 | 0.124 |
| CVEDA_CSA | 0.5384 | −0.157 | 0.255 |
| ESTB2_EUR | 0.053 | 0.057 | 0.029 |
| FINNG_EUR | 0.0081 | 0.071 | 0.027 |
| GEDIS_EUR | 0.7571 | −0.049 | 0.16 |
| GISS1_EUR | 0.3859 | −0.089 | 0.102 |
| GISS2_EUR | 0.7295 | 0.04 | 0.116 |
| GREAT_EAS | 0.9233 | −0.008 | 0.078 |
| IPSYC_EUR | 0.1962 | 0.032 | 0.024 |
| JANS3_EUR | 0.9404 | −0.01 | 0.132 |
| JANS4_EUR | 0.1947 | −0.191 | 0.147 |
| MIREC_AFR | 0.346 | 0.138 | 0.146 |
| MIREC_EUR | 0.9825 | −0.004 | 0.19 |
| MVPXQ_EAS | 0.717 | −0.038 | 0.106 |
| MVPXQ_EUR | 2.26e−06 | 0.076 | 0.016 |
| MVPXQ_LAT | 0.4401 | 0.027 | 0.035 |
| PGCBD_EUR | 0.5336 | −0.02 | 0.033 |
| PGCMD_EUR | 0.9094 | 0.005 | 0.044 |
| PGCPT_AFR | 0.8451 | 0.022 | 0.111 |
| PGCPT_EUR | 0.2249 | 0.086 | 0.07 |
| PGCSZ_EUR | 0.4226 | 0.037 | 0.046 |
| PRFCT_EUR | 0.0553 | 0.1 | 0.052 |
| PSYCR_EUR | 0.1717 | 0.154 | 0.113 |
| QIMRB_EUR | 0.6146 | 0.017 | 0.035 |
| SNUBH−ASA_EAS | 0.3091 | 0.127 | 0.125 |
| SNUBH−KCHIP_EAS | 0.6814 | −0.067 | 0.164 |
| STRR1_LAT | 0.2085 | −0.246 | 0.196 |
| UKBJC_EUR | 0.001 | 0.083 | 0.025 |
| YPENN_AFR | 0.1448 | 0.232 | 0.159 |
| YPENN_EUR | 0.6367 | −0.075 | 0.159 |
| **meta** | **1.02e−10** | **0.051** | **0.008** |

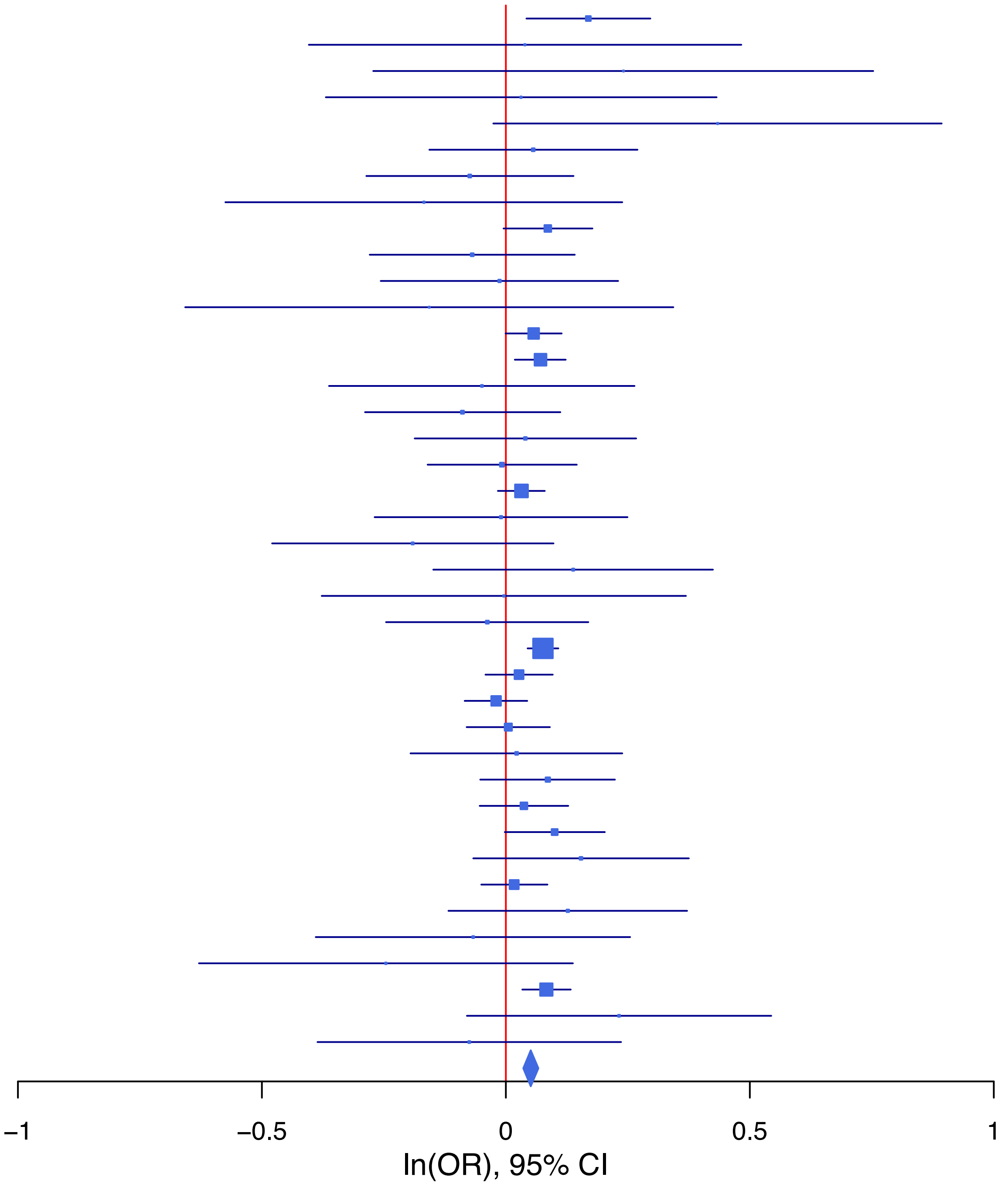

**rs9276627 T/C 6:32743835**

| Cohort | P | ln(OR) | SE |
|---|---|---|---|
| ADHEA_EUR | 0.0549 | 0.221 | 0.115 |
| ALSPC_EUR | 0.6818 | 0.04 | 0.098 |
| BEPS7_EUR | 0.6116 | −0.173 | 0.342 |
| BHRCM_EUR | 0.6243 | −0.151 | 0.308 |
| BHRCM_LAT | 0.772 | 0.119 | 0.41 |
| BOR17_EUR | 0.0168 | 0.356 | 0.149 |
| BOR2C_EUR | 0.7379 | −0.051 | 0.153 |
| BOR2E_EUR | 0.8894 | −0.047 | 0.337 |
| CNVRG_EAS | 0.6894 | −0.037 | 0.092 |
| COGA1_EUR | 0.7615 | −0.032 | 0.105 |
| CUINT_EUR | 0.6687 | 0.077 | 0.179 |
| CVEDA_CSA | 0.2286 | 0.306 | 0.254 |
| ESTB2_EUR | 0.0668 | 0.078 | 0.043 |
| FINNG_EUR | 0.1316 | 0.057 | 0.038 |
| GEDIS_EUR | 0.88 | −0.034 | 0.223 |
| GISS2_EUR | 0.3619 | 0.156 | 0.171 |
| GTPRJ_AFR | 0.7421 | 0.072 | 0.221 |
| IPSYC_EUR | 0.4554 | 0.024 | 0.032 |
| JANS3_EUR | 0.7185 | −0.07 | 0.194 |
| JANS4_EUR | 0.729 | 0.076 | 0.22 |
| MVPXQ_AFR | 0.6426 | −0.038 | 0.081 |
| MVPXQ_EAS | 0.6598 | −0.091 | 0.206 |
| MVPXQ_EUR | 0.0051 | 0.066 | 0.023 |
| MVPXQ_LAT | 0.2295 | 0.065 | 0.054 |
| PGCBD_EUR | 0.0712 | 0.089 | 0.05 |
| PGCMD_EUR | 0.2438 | 0.08 | 0.069 |
| PGCPT_EUR | 0.4887 | −0.073 | 0.105 |
| PRFCT_EUR | 0.7163 | 0.026 | 0.073 |
| PSYCR_EUR | 0.8964 | −0.021 | 0.161 |
| QIMRB_EUR | 0.0124 | 0.122 | 0.049 |
| SNUBH−ASA_EAS | 0.203 | 0.247 | 0.194 |
| SNUBH−KCHIP_EAS | 0.1507 | −0.369 | 0.257 |
| STRR1_LAT | 0.9098 | 0.031 | 0.271 |
| UKBJC_EUR | 8e−04 | 0.12 | 0.036 |
| YPENN_EUR | 0.2775 | −0.282 | 0.259 |
| **meta** | **5.00e−08** | **0.063** | **0.012** |

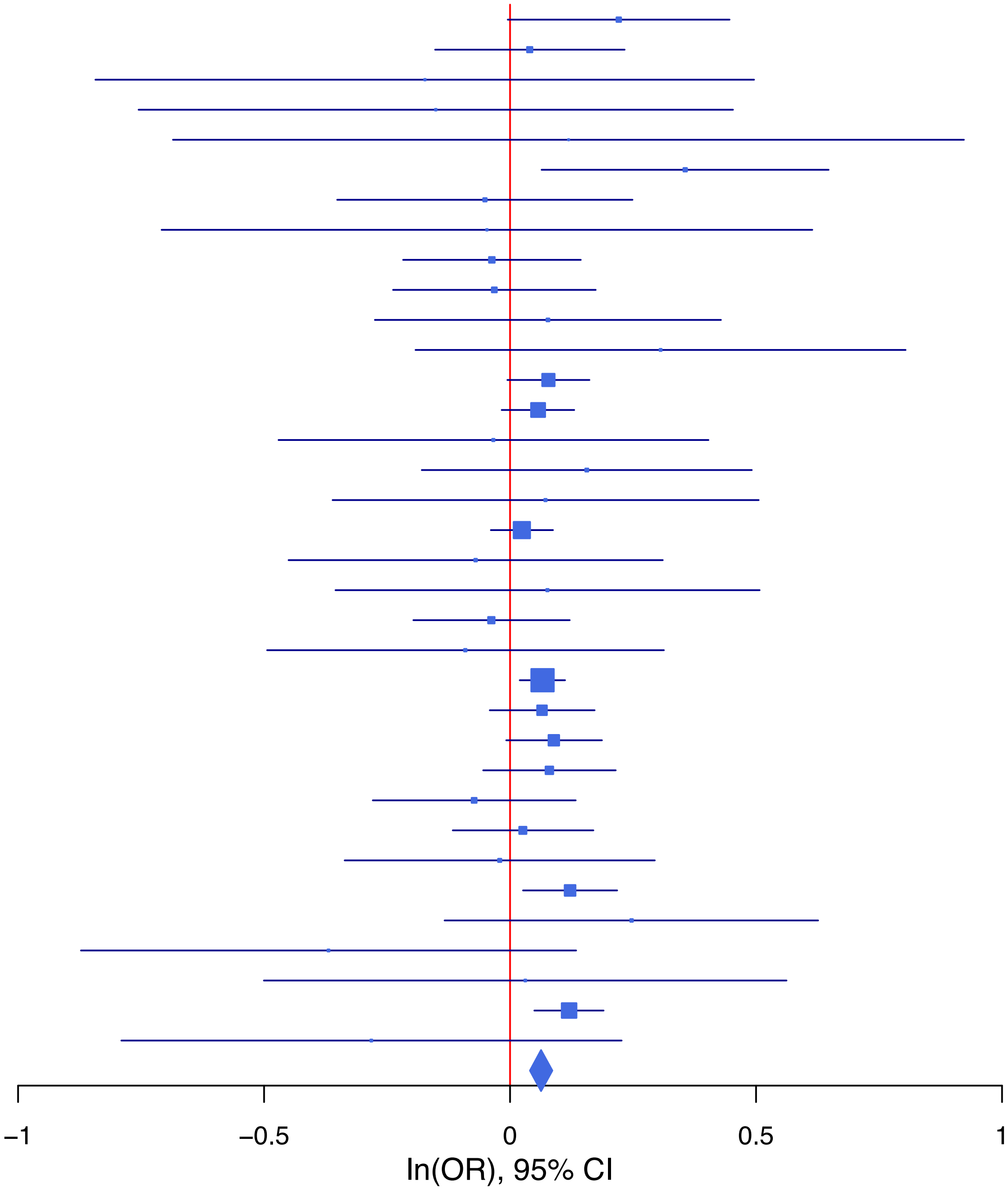

**rs9306311   G/C   22:37050675**

| Cohort | P | ln(OR) | SE |
|---|---|---|---|
| ALSPC_EUR | 0.9088 | 0.007 | 0.06 |
| BEPS7_EUR | 0.939 | −0.017 | 0.226 |
| BOR17_EUR | 0.9511 | −0.006 | 0.103 |
| BOR2C_EUR | 0.8632 | −0.018 | 0.103 |
| BOR2E_EUR | 0.1125 | −0.349 | 0.22 |
| CNVRG_EAS | 0.1465 | −0.09 | 0.062 |
| COGA1_AFR | 0.4346 | 0.107 | 0.138 |
| COGA1_EUR | 0.461 | −0.048 | 0.066 |
| CVEDA_CSA | 0.1567 | 0.238 | 0.168 |
| ESTB2_EUR | 0.1903 | −0.036 | 0.028 |
| FINNG_EUR | 0.8793 | −0.004 | 0.027 |
| GEDIS_EUR | 0.7627 | −0.043 | 0.143 |
| GISS1_EUR | 0.964 | −0.004 | 0.09 |
| GISS2_EUR | 0.3267 | −0.121 | 0.123 |
| GREAT_EAS | 0.4946 | −0.07 | 0.103 |
| GTPRJ_AFR | 0.1643 | −0.126 | 0.091 |
| JANS3_EUR | 0.1971 | −0.163 | 0.127 |
| JANS4_EUR | 0.7904 | 0.038 | 0.143 |
| MVPXQ_AFR | 0.6978 | 0.011 | 0.029 |
| MVPXQ_EUR | 5e−04 | −0.049 | 0.014 |
| MVPXQ_LAT | 0.463 | −0.028 | 0.039 |
| PGCBD_EUR | 0.0205 | −0.07 | 0.03 |
| PGCED_EUR | 0.5029 | −0.084 | 0.126 |
| PGCMD_EUR | 0.9855 | −0.001 | 0.039 |
| PGCPT_AFR | 0.5497 | 0.088 | 0.147 |
| PGCPT_EUR | 0.1725 | −0.092 | 0.068 |
| PGCSZ_EUR | 0.6643 | −0.019 | 0.043 |
| PRFCT_EUR | 0.0084 | −0.145 | 0.055 |
| PSYCR_EUR | 0.9842 | −0.002 | 0.109 |
| QIMRB_EUR | 0.1081 | −0.055 | 0.034 |
| SNUBH−ASA_EAS | 0.2409 | −0.197 | 0.168 |
| STRR1_LAT | 0.6922 | 0.083 | 0.211 |
| UKBJC_EUR | 0.0014 | −0.073 | 0.023 |
| **meta** | **3.04e−08** | **−0.042** | **0.008** |

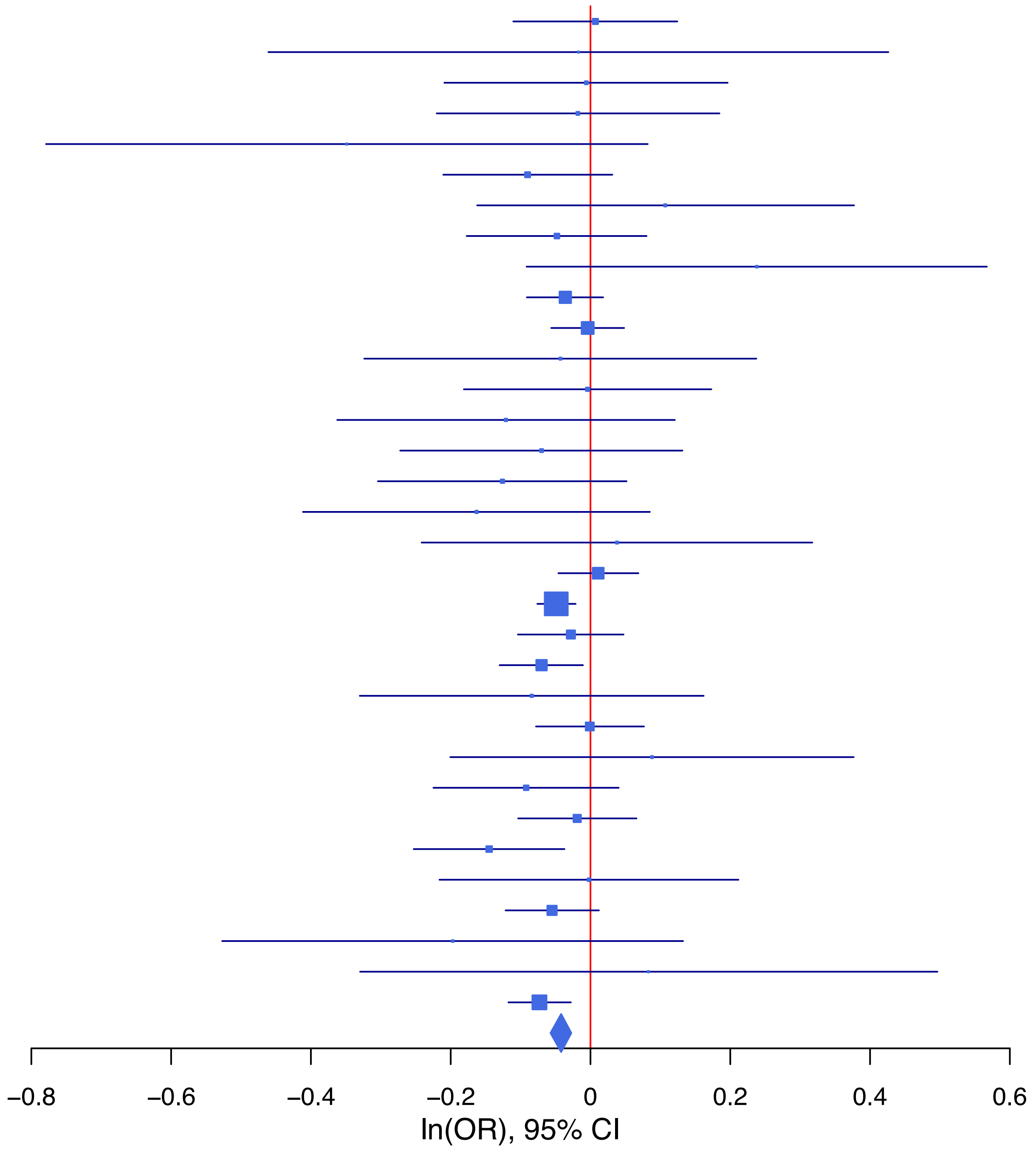

**rs959922 A/G 5:92450649**

| Cohort | P | ln(OR) | SE |
|---|---|---|---|
| ADHEA_AFR | 0.2029 | 0.156 | 0.123 |
| ADHEA_EUR | 0.7728 | 0.02 | 0.068 |
| ALSPC_EUR | 0.0389 | 0.113 | 0.055 |
| BEPS7_EUR | 0.2582 | 0.198 | 0.175 |
| BHRCM_AFR | 0.1373 | 0.331 | 0.222 |
| BOR17_EUR | 0.0299 | 0.191 | 0.088 |
| BOR2C_EUR | 0.8821 | 0.013 | 0.09 |
| BOR2E_EUR | 0.771 | 0.052 | 0.177 |
| CNVRG_EAS | 0.752 | −0.016 | 0.051 |
| COGA1_AFR | 0.8394 | −0.022 | 0.108 |
| COGA1_EUR | 0.0137 | 0.146 | 0.059 |
| CVEDA_CSA | 0.9974 | 0.001 | 0.164 |
| ESTB2_EUR | 0.043 | 0.047 | 0.023 |
| FINNG_EUR | 0.6227 | −0.011 | 0.023 |
| GEDIS_EUR | 0.5339 | 0.081 | 0.131 |
| GISS1_EUR | 0.533 | 0.049 | 0.078 |
| GISS2_EUR | 0.9308 | −0.008 | 0.091 |
| GREAT_EAS | 0.4843 | 0.067 | 0.095 |
| GTPRJ_AFR | 0.5036 | 0.043 | 0.064 |
| IPSYC_EUR | 0.1148 | 0.031 | 0.02 |
| JANS3_EUR | 0.6079 | −0.052 | 0.102 |
| JANS4_EUR | 0.558 | −0.069 | 0.117 |
| MIREC_AFR | 0.0526 | 0.28 | 0.145 |
| MIREC_EUR | 0.6001 | −0.08 | 0.152 |
| MVPXQ_AFR | 0.013 | 0.055 | 0.022 |
| MVPXQ_EAS | 0.7283 | −0.039 | 0.111 |
| MVPXQ_EUR | 0.0014 | 0.04 | 0.013 |
| PGCBD_EUR | 0.2893 | 0.027 | 0.026 |
| PGCED_EUR | 0.229 | 0.139 | 0.116 |
| PGCMD_EUR | 0.8042 | 0.009 | 0.034 |
| PGCPT_AFR | 0.2991 | 0.116 | 0.112 |
| PGCPT_EUR | 0.7946 | −0.015 | 0.057 |
| PGCSZ_EUR | 0.6342 | 0.017 | 0.037 |
| PRFCT_EUR | 0.8248 | −0.01 | 0.044 |
| PSYCR_EUR | 0.7011 | −0.034 | 0.089 |
| QIMRB_EUR | 0.2949 | 0.029 | 0.028 |
| SNUBH−ASA_EAS | 0.9264 | 0.013 | 0.145 |
| SNUBH−KCHIP_EAS | 0.6921 | 0.081 | 0.205 |
| STRR1_LAT | 0.8085 | 0.045 | 0.184 |
| UKBJC_EUR | 0.0195 | 0.047 | 0.02 |
| YPENN_AFR | 0.1075 | 0.246 | 0.153 |
| YPENN_EUR | 0.5315 | −0.079 | 0.127 |
| **meta** | **2.08e−08** | **0.035** | **0.006** |

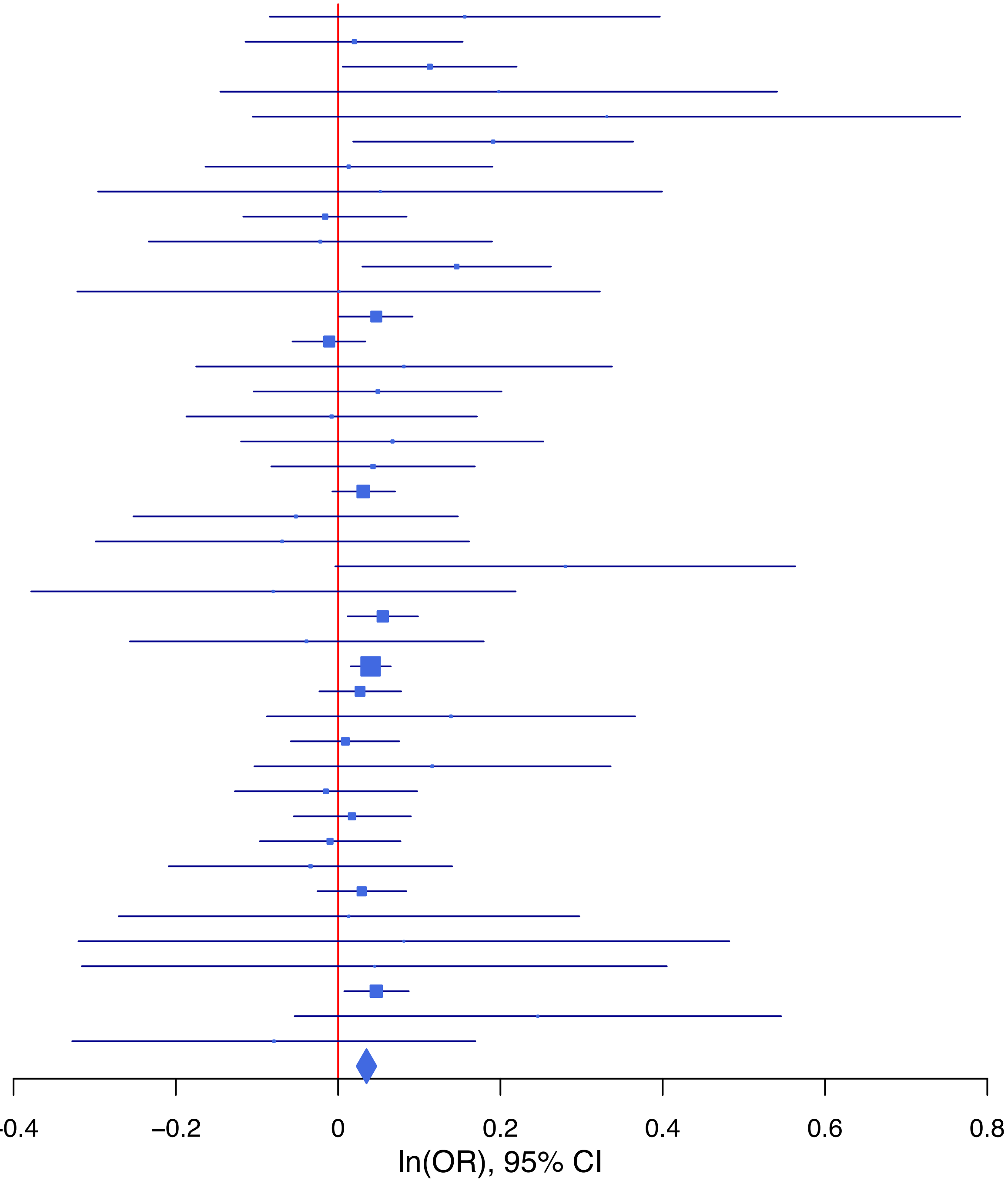

**Supplementary Data 1E: Forest plots of lead SNPs at the 35 genome-wide significant loci from the GWAS meta-analysis of suicide attempt in European ancestry samples.**

Each box represents the log odds ratio (OR) from an individual contributing cohort, with horizontal lines indicating the 95% confidence interval (CI). The diamond represents the overall meta-analytic estimate across studies.

**rs10890032 T/C 1:73789324**

| Cohort | P | ln(OR) | SE |
|---|---|---|---|
| ALSPC_EUR | 0.22 | 0.069 | 0.056 |
| BEPS7_EUR | 0.6099 | 0.091 | 0.178 |
| BHRCM_EUR | 0.0201 | 0.367 | 0.158 |
| BOR17_EUR | 0.6077 | −0.044 | 0.085 |
| BOR2C_EUR | 0.2634 | −0.097 | 0.087 |
| BOR2E_EUR | 0.3088 | 0.177 | 0.174 |
| COGA1_EUR | 0.3066 | 0.062 | 0.061 |
| CUINT_EUR | 0.329 | 0.099 | 0.101 |
| ESTB2_EUR | 0.6117 | 0.012 | 0.024 |
| FINNG_EUR | 0.1903 | 0.031 | 0.024 |
| GEDIS_EUR | 0.0067 | 0.36 | 0.133 |
| GISS1_EUR | 0.9358 | −0.007 | 0.081 |
| GISS2_EUR | 0.7562 | 0.029 | 0.093 |
| IPSYC_EUR | 0.1975 | 0.027 | 0.02 |
| JANS3_EUR | 0.0641 | −0.204 | 0.11 |
| JANS4_EUR | 0.1772 | −0.169 | 0.126 |
| MIREC_EUR | 0.7961 | −0.041 | 0.157 |
| MVPXQ_EUR | 3.73e−05 | 0.053 | 0.013 |
| PGCBD_EUR | 0.9846 | −0.001 | 0.027 |
| PGCED_EUR | 0.3467 | 0.112 | 0.119 |
| PGCMD_EUR | 0.0511 | 0.068 | 0.035 |
| PGCPT_EUR | 0.9279 | 0.005 | 0.059 |
| PGCSZ_EUR | 0.2004 | 0.049 | 0.038 |
| PRFCT_EUR | 0.1231 | 0.068 | 0.044 |
| PSYCR_EUR | 0.3669 | −0.082 | 0.091 |
| QIMRB_EUR | 0.0562 | 0.054 | 0.028 |
| UKBJC_EUR | 0.0011 | 0.069 | 0.021 |
| YPENN_EUR | 0.2547 | 0.147 | 0.129 |
| **meta** | **2.77e−09** | **0.041** | **0.007** |

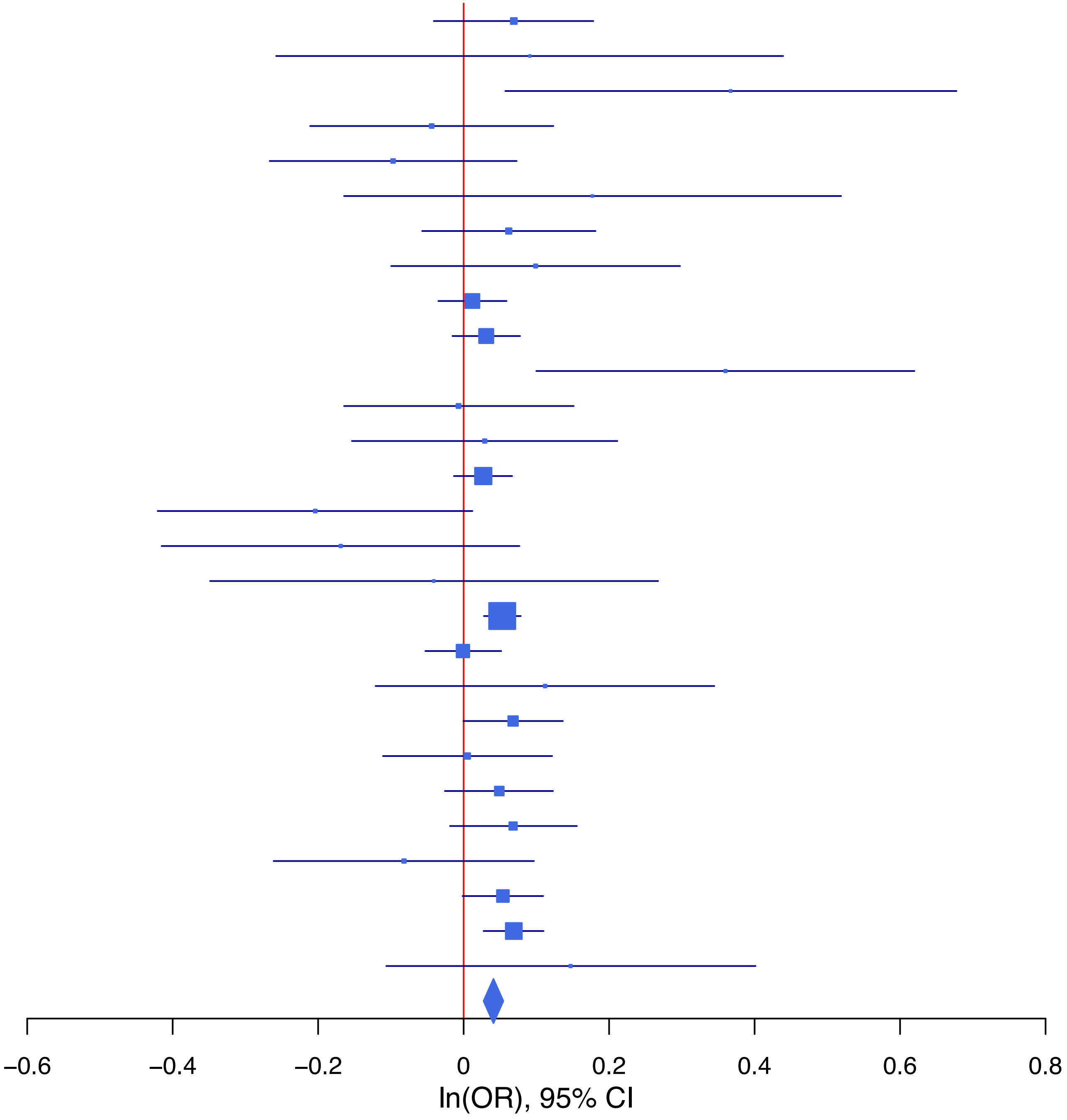

**rs1125394 C/T 11:113297185**

| Cohort | P | ln(OR) | SE |
|---|---|---|---|
| ADHEA_EUR | 0.0415 | 0.181 | 0.088 |
| ALSPC_EUR | 0.776 | 0.021 | 0.074 |
| BEPS7_EUR | 0.8993 | −0.031 | 0.244 |
| BHRCM_EUR | 0.2864 | 0.249 | 0.234 |
| BOR17_EUR | 0.3507 | −0.111 | 0.118 |
| BOR2C_EUR | 0.3726 | 0.107 | 0.121 |
| BOR2E_EUR | 0.4488 | −0.218 | 0.288 |
| COGA1_EUR | 0.4013 | 0.069 | 0.083 |
| CUINT_EUR | 0.3503 | 0.133 | 0.142 |
| ESTB2_EUR | 0.2743 | 0.032 | 0.029 |
| FINNG_EUR | 0.1936 | 0.037 | 0.029 |
| GEDIS_EUR | 0.4641 | −0.136 | 0.186 |
| GISS1_EUR | 0.2708 | 0.111 | 0.1 |
| GISS2_EUR | 0.6007 | −0.062 | 0.119 |
| IPSYC_EUR | 9e−04 | 0.09 | 0.027 |
| JANS3_EUR | 0.1086 | 0.216 | 0.134 |
| JANS4_EUR | 0.5088 | −0.108 | 0.164 |
| MIREC_EUR | 0.2544 | 0.225 | 0.197 |
| MVPXQ_EUR | 3.16e−05 | 0.071 | 0.017 |
| PGCBD_EUR | 0.3129 | 0.036 | 0.035 |
| PGCED_EUR | 0.6335 | 0.081 | 0.171 |
| PGCMD_EUR | 0.0445 | 0.097 | 0.048 |
| PGCPT_EUR | 0.2192 | 0.093 | 0.076 |
| PGCSZ_EUR | 0.9428 | 0.004 | 0.051 |
| PRFCT_EUR | 0.0836 | 0.102 | 0.059 |
| PSYCR_EUR | 0.4611 | 0.087 | 0.118 |
| QIMRB_EUR | 0.7337 | 0.012 | 0.037 |
| UKBJC_EUR | 0.1779 | 0.038 | 0.028 |
| YPENN_EUR | 0.4093 | −0.149 | 0.18 |
| **meta** | **6.08e−10** | **0.056** | **0.009** |

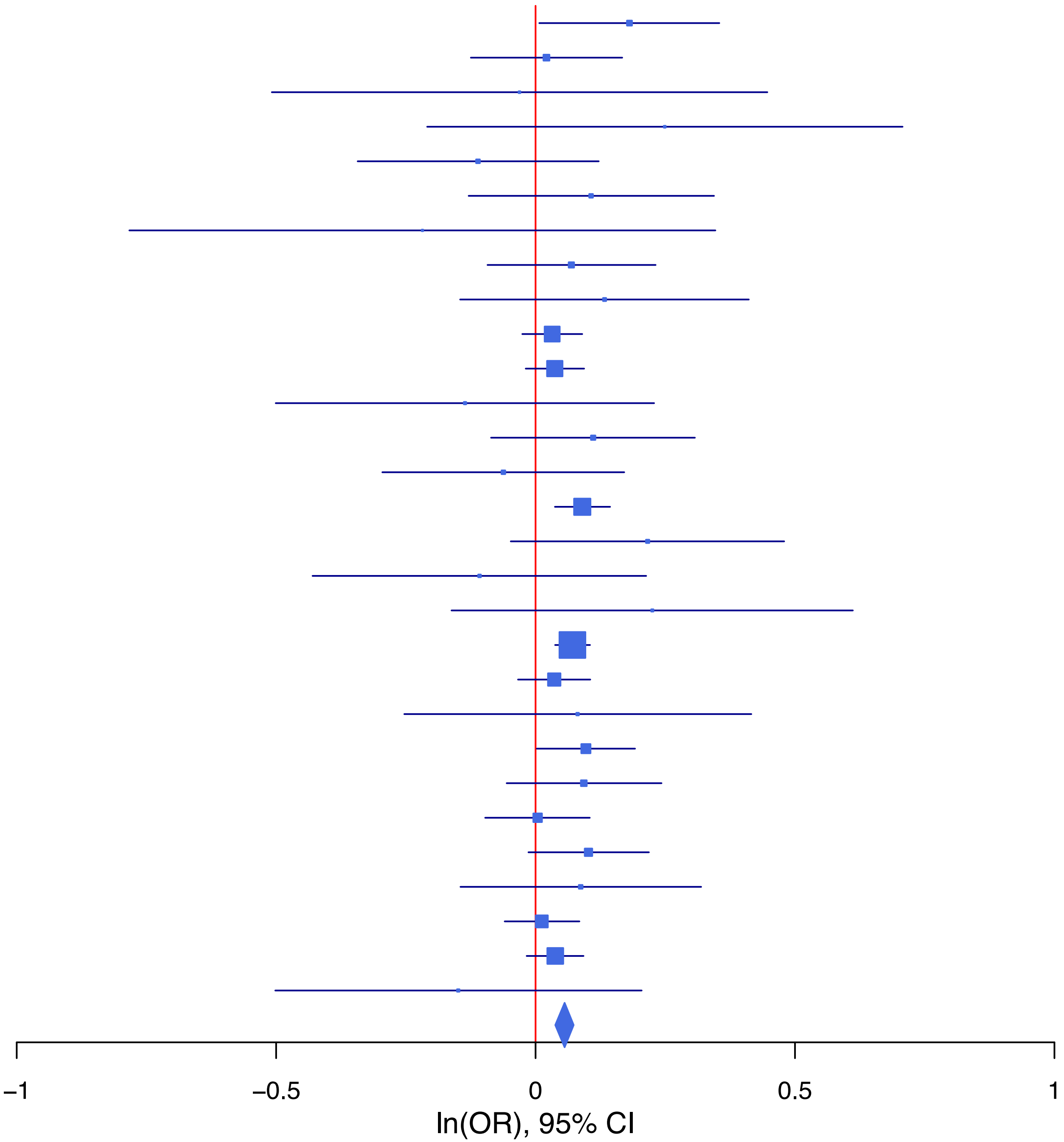

**rs12378745 G/A 9:122672307**

| Cohort | P | ln(OR) | SE |
|---|---|---|---|
| ADHEA_EUR | 0.2172 | −0.088 | 0.071 |
| ALSPC_EUR | 0.6704 | −0.023 | 0.055 |
| BEPS7_EUR | 0.0141 | −0.475 | 0.194 |
| BOR17_EUR | 0.413 | 0.073 | 0.089 |
| BOR2C_EUR | 0.7945 | 0.023 | 0.089 |
| BOR2E_EUR | 0.1522 | 0.256 | 0.179 |
| COGA1_EUR | 0.1816 | −0.079 | 0.059 |
| CUINT_EUR | 0.1221 | −0.158 | 0.102 |
| ESTB2_EUR | 0.0922 | −0.04 | 0.024 |
| FINNG_EUR | 0.0252 | −0.053 | 0.024 |
| GEDIS_EUR | 0.4912 | 0.093 | 0.134 |
| GISS1_EUR | 0.4702 | −0.058 | 0.08 |
| GISS2_EUR | 0.8043 | 0.024 | 0.099 |
| IPSYC_EUR | 0.005 | −0.058 | 0.02 |
| JANS3_EUR | 0.2861 | −0.117 | 0.11 |
| JANS4_EUR | 0.7929 | −0.032 | 0.124 |
| MVPXQ_EUR | 0.0071 | −0.035 | 0.013 |
| PGCBD_EUR | 0.0608 | −0.05 | 0.026 |
| PGCED_EUR | 0.9052 | −0.014 | 0.118 |
| PGCMD_EUR | 0.0554 | −0.068 | 0.035 |
| PGCPT_EUR | 0.3097 | −0.06 | 0.059 |
| PGCSZ_EUR | 0.9618 | −0.002 | 0.037 |
| PRFCT_EUR | 0.3194 | −0.045 | 0.045 |
| PSYCR_EUR | 0.9782 | 0.002 | 0.09 |
| QIMRB_EUR | 0.4957 | −0.019 | 0.028 |
| UKBJC_EUR | 0.0382 | −0.043 | 0.021 |
| YPENN_EUR | 0.7876 | 0.035 | 0.13 |
| **meta** | **2.49e−09** | **−0.041** | **0.007** |

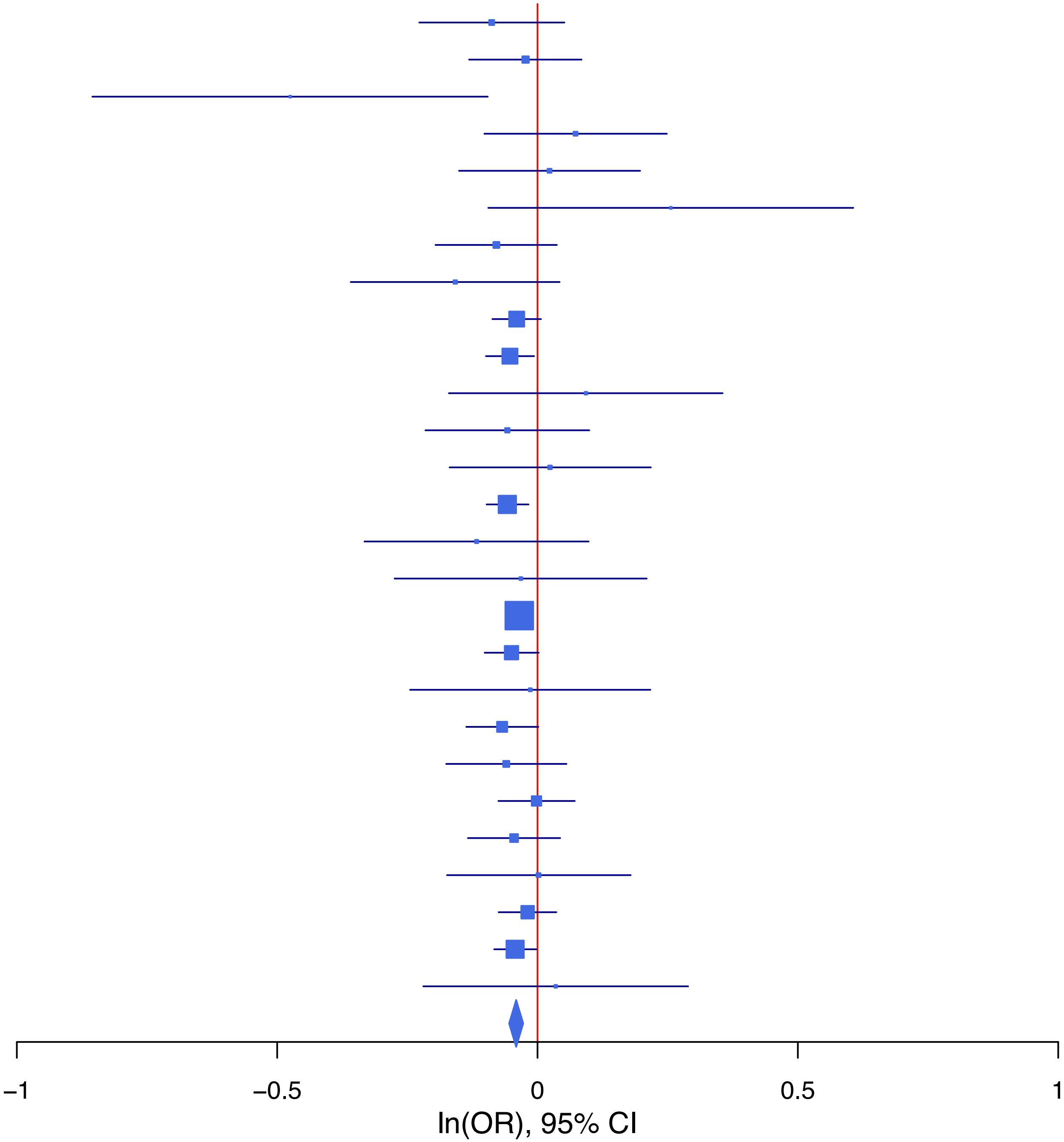

**rs12453624 A/G 17:27426153**

| Cohort | P | ln(OR) | SE |
|---|---|---|---|
| ALSPC_EUR | 0.6734 | 0.024 | 0.056 |
| BEPS7_EUR | 0.8028 | 0.049 | 0.194 |
| BHRCM_EUR | 0.6113 | 0.081 | 0.159 |
| BOR17_EUR | 0.4605 | 0.067 | 0.091 |
| BOR2C_EUR | 0.5602 | 0.054 | 0.093 |
| BOR2E_EUR | 0.7724 | −0.054 | 0.186 |
| COGA1_EUR | 0.2762 | 0.067 | 0.061 |
| CUINT_EUR | 0.1265 | 0.16 | 0.105 |
| ESTB2_EUR | 0.0649 | 0.046 | 0.025 |
| FINNG_EUR | 0.0427 | 0.049 | 0.024 |
| GEDIS_EUR | 0.1773 | 0.181 | 0.135 |
| GISS1_EUR | 0.7991 | 0.022 | 0.085 |
| GISS2_EUR | 0.299 | 0.104 | 0.1 |
| IPSYC_EUR | 0.033 | 0.045 | 0.021 |
| JANS3_EUR | 0.4492 | 0.082 | 0.108 |
| JANS4_EUR | 0.4648 | −0.09 | 0.124 |
| MIREC_EUR | 0.3624 | 0.141 | 0.154 |
| MVPXQ_EUR | 0.0492 | 0.026 | 0.013 |
| PGCBD_EUR | 0.003 | 0.081 | 0.027 |
| PGCED_EUR | 0.8302 | 0.026 | 0.119 |
| PGCMD_EUR | 0.1354 | 0.054 | 0.036 |
| PGCPT_EUR | 0.9285 | −0.005 | 0.061 |
| PGCSZ_EUR | 0.2833 | 0.042 | 0.039 |
| PRFCT_EUR | 0.9786 | 0.001 | 0.046 |
| PSYCR_EUR | 0.0583 | 0.172 | 0.091 |
| QIMRB_EUR | 0.1054 | 0.046 | 0.029 |
| YPENN_EUR | 0.0482 | −0.287 | 0.145 |
| **meta** | **3.01e−08** | **0.042** | **0.007** |

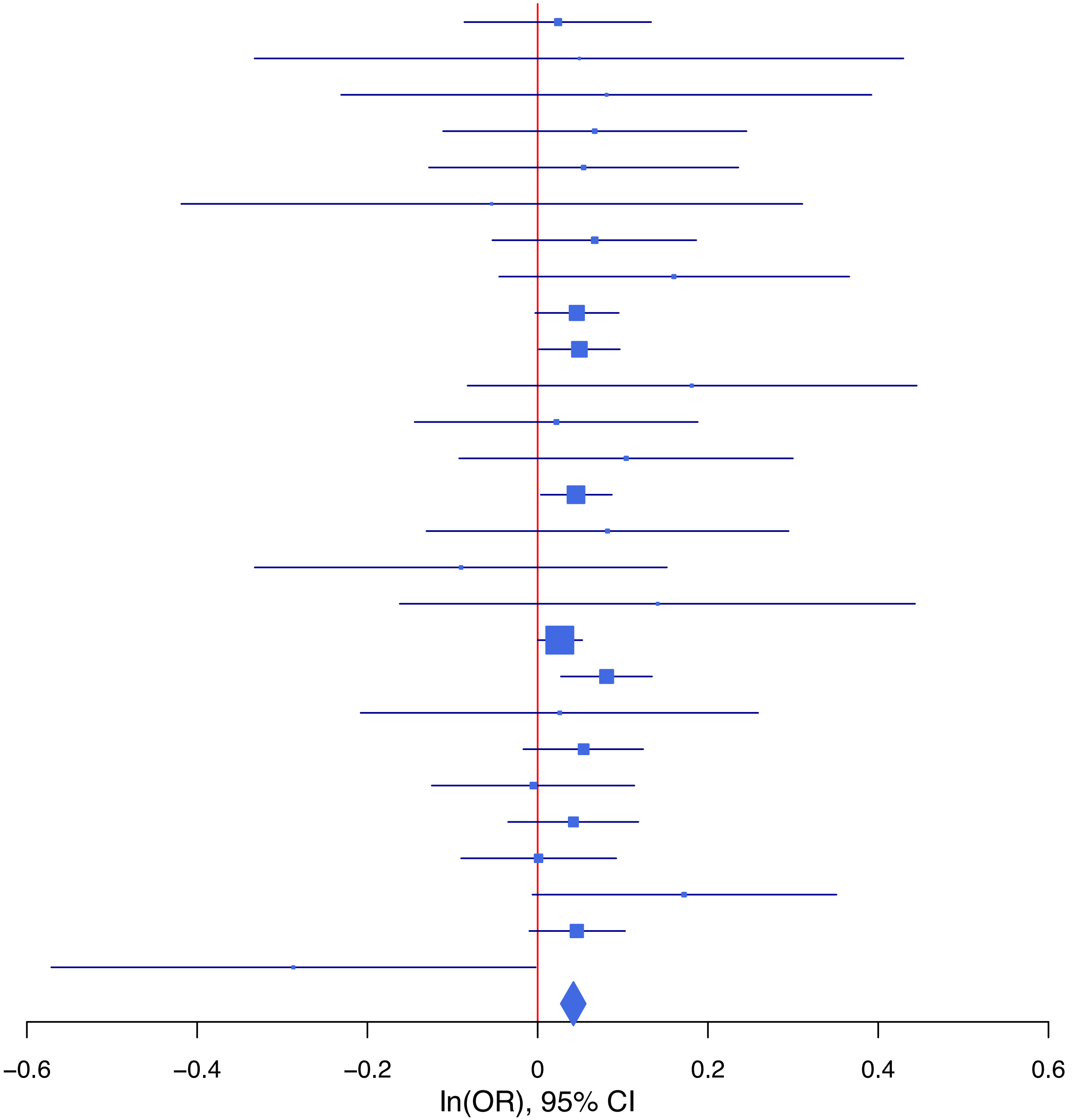

**rs12666306 G/A 7:115082406**

| Cohort | P | ln(OR) | SE |
|---|---|---|---|
| ADHEA_EUR | 0.8373 | −0.014 | 0.068 |
| ALSPC_EUR | 0.3336 | −0.052 | 0.054 |
| BEPS7_EUR | 0.3269 | −0.168 | 0.171 |
| BHRCM_EUR | 0.189 | −0.209 | 0.159 |
| BOR17_EUR | 0.6436 | 0.039 | 0.084 |
| BOR2C_EUR | 0.4588 | 0.063 | 0.085 |
| BOR2E_EUR | 0.2291 | −0.215 | 0.179 |
| COGA1_EUR | 0.8843 | −0.009 | 0.059 |
| CUINT_EUR | 0.377 | −0.088 | 0.1 |
| ESTB2_EUR | 0.0025 | −0.069 | 0.023 |
| FINNG_EUR | 0.1191 | −0.036 | 0.023 |
| GEDIS_EUR | 0.0023 | −0.401 | 0.131 |
| GISS1_EUR | 0.4832 | −0.054 | 0.078 |
| GISS2_EUR | 0.8279 | 0.02 | 0.092 |
| IPSYC_EUR | 0.0136 | −0.05 | 0.02 |
| JANS3_EUR | 0.9763 | −0.003 | 0.104 |
| JANS4_EUR | 0.1695 | −0.164 | 0.119 |
| MIREC_EUR | 0.1038 | 0.248 | 0.153 |
| PGCBD_EUR | 0.0081 | −0.068 | 0.026 |
| PGCED_EUR | 0.525 | 0.072 | 0.114 |
| PGCMD_EUR | 0.0345 | −0.072 | 0.034 |
| PGCPT_EUR | 0.4953 | −0.039 | 0.057 |
| PGCSZ_EUR | 0.2807 | −0.039 | 0.037 |
| PRFCT_EUR | 0.4794 | −0.031 | 0.044 |
| PSYCR_EUR | 0.1969 | 0.111 | 0.086 |
| QIMRB_EUR | 6e−04 | −0.094 | 0.027 |
| UKBJC_EUR | 0.0681 | −0.037 | 0.02 |
| YPENN_EUR | 0.9232 | −0.012 | 0.127 |
| **meta** | **1.69e−10** | **−0.05** | **0.008** |

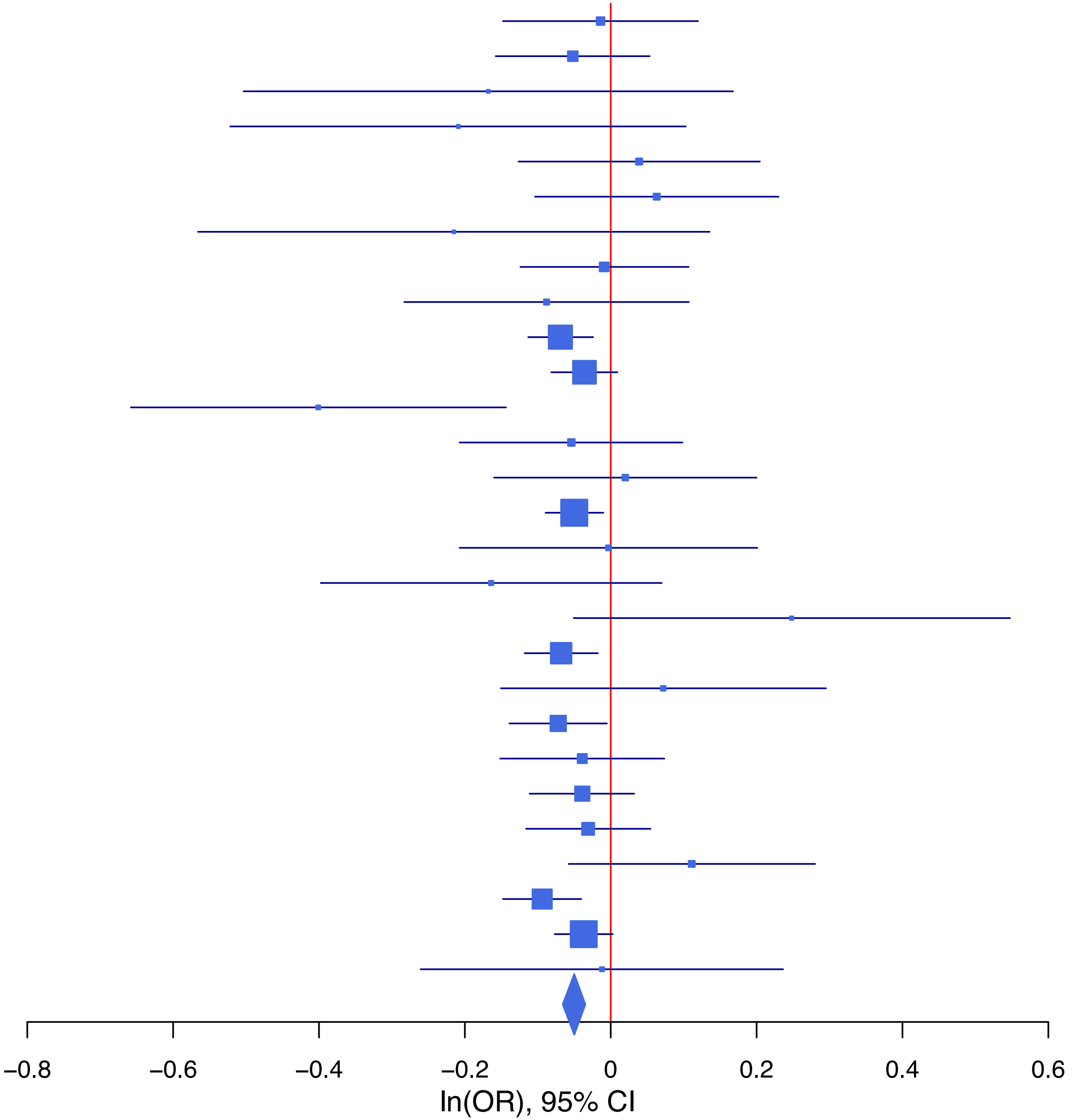

**rs17115481 A/G 5:153358226**

| Cohort | P | ln(OR) | SE |
|---|---|---|---|
| ADHEA_EUR | 0.4654 | 0.055 | 0.076 |
| ALSPC_EUR | 0.1243 | 0.092 | 0.06 |
| BEPS7_EUR | 0.5916 | 0.107 | 0.2 |
| BHRCM_EUR | 0.5657 | 0.094 | 0.164 |
| BOR17_EUR | 0.605 | 0.048 | 0.093 |
| BOR2C_EUR | 0.3727 | 0.083 | 0.093 |
| BOR2E_EUR | 0.766 | −0.056 | 0.188 |
| COGA1_EUR | 0.0638 | 0.122 | 0.066 |
| CUINT_EUR | 0.1325 | 0.16 | 0.106 |
| ESTB2_EUR | 0.0307 | 0.056 | 0.026 |
| FINNG_EUR | 0.1376 | 0.037 | 0.025 |
| GEDIS_EUR | 0.4559 | −0.109 | 0.146 |
| GISS1_EUR | 0.8284 | −0.019 | 0.087 |
| GISS2_EUR | 0.3542 | 0.098 | 0.106 |
| IPSYC_EUR | 2e−04 | 0.084 | 0.022 |
| JANS3_EUR | 0.5913 | 0.06 | 0.113 |
| JANS4_EUR | 0.6286 | 0.062 | 0.127 |
| MIREC_EUR | 0.412 | 0.132 | 0.161 |
| MVPXQ_EUR | 0.1143 | 0.023 | 0.014 |
| PGCBD_EUR | 0.4015 | 0.024 | 0.029 |
| PGCED_EUR | 0.8718 | −0.02 | 0.126 |
| PGCMD_EUR | 0.02 | 0.088 | 0.038 |
| PGCPT_EUR | 0.8404 | 0.013 | 0.065 |
| PGCSZ_EUR | 0.1372 | 0.061 | 0.041 |
| PRFCT_EUR | 0.6357 | −0.024 | 0.051 |
| PSYCR_EUR | 0.8576 | −0.018 | 0.101 |
| QIMRB_EUR | 0.0912 | 0.052 | 0.031 |
| UKBJC_EUR | 0.2659 | 0.025 | 0.023 |
| YPENN_EUR | 0.3021 | 0.144 | 0.139 |
| **meta** | **1.12e−08** | **0.043** | **0.007** |

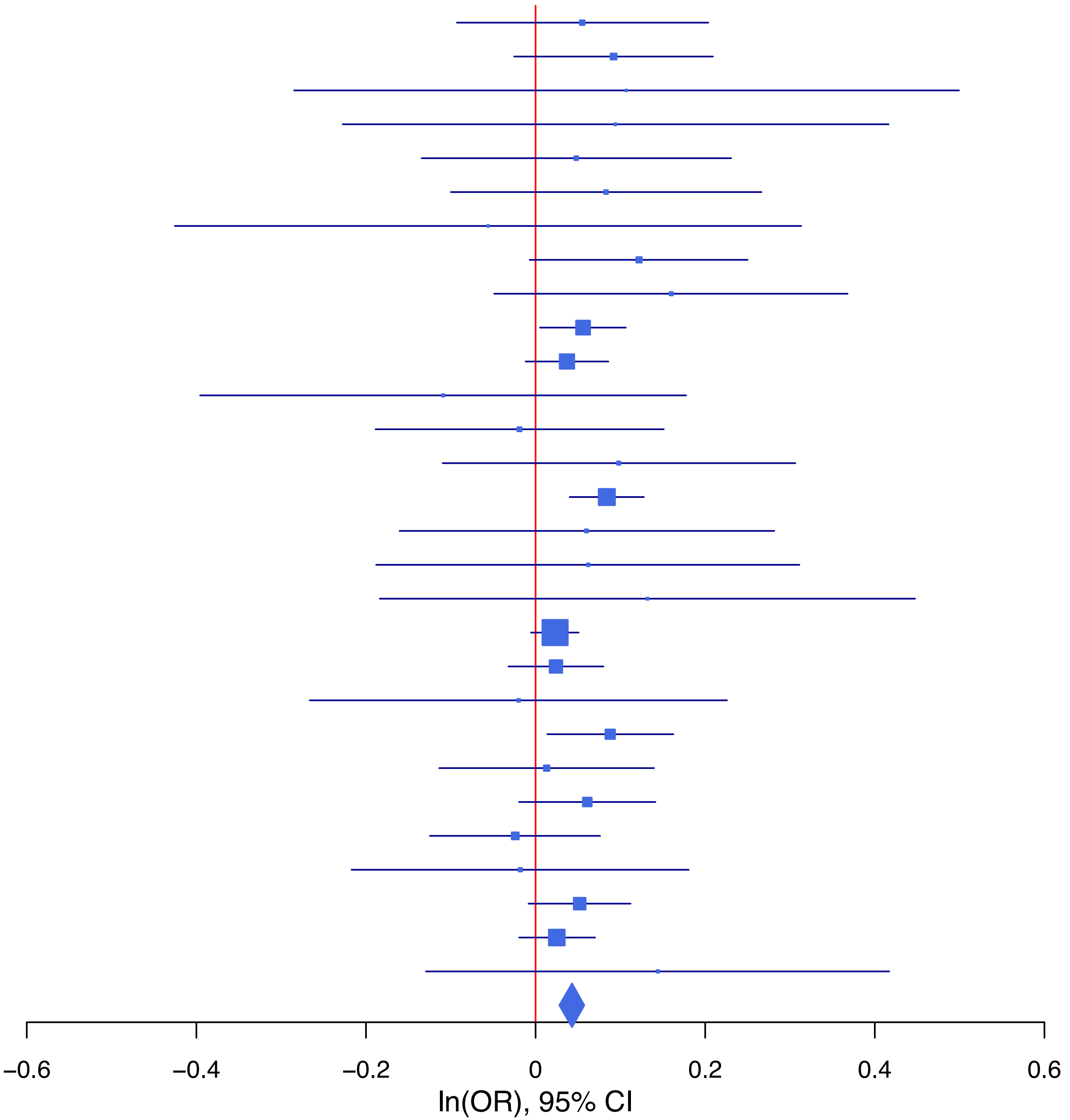

**rs185782836 A/G 14:64520663**

| Cohort | P | ln(OR) | SE |
|---|---|---|---|
| ALSPC_EUR | 0.1242 | 0.272 | 0.177 |
| BHRCM_EUR | 0.1301 | 0.751 | 0.496 |
| BOR17_EUR | 0.2941 | 0.309 | 0.294 |
| BOR2C_EUR | 0.7057 | 0.114 | 0.301 |
| ESTB2_EUR | 0.014 | 0.128 | 0.052 |
| FINNG_EUR | 0.0281 | 0.235 | 0.107 |
| GISS2_EUR | 0.8451 | 0.051 | 0.259 |
| IPSYC_EUR | 1.08e−05 | 0.332 | 0.075 |
| MVPXQ_EUR | 0.0081 | 0.153 | 0.058 |
| PGCPT_EUR | 0.2954 | −0.287 | 0.274 |
| PGCSZ_EUR | 0.7751 | −0.053 | 0.187 |
| PRFCT_EUR | 0.5743 | 0.088 | 0.156 |
| PSYCR_EUR | 0.4005 | 0.245 | 0.292 |
| QIMRB_EUR | 0.4612 | 0.077 | 0.104 |
| UKBJC_EUR | 0.189 | 0.1 | 0.076 |
| **meta** | **6.18e−09** | **0.157** | **0.027** |

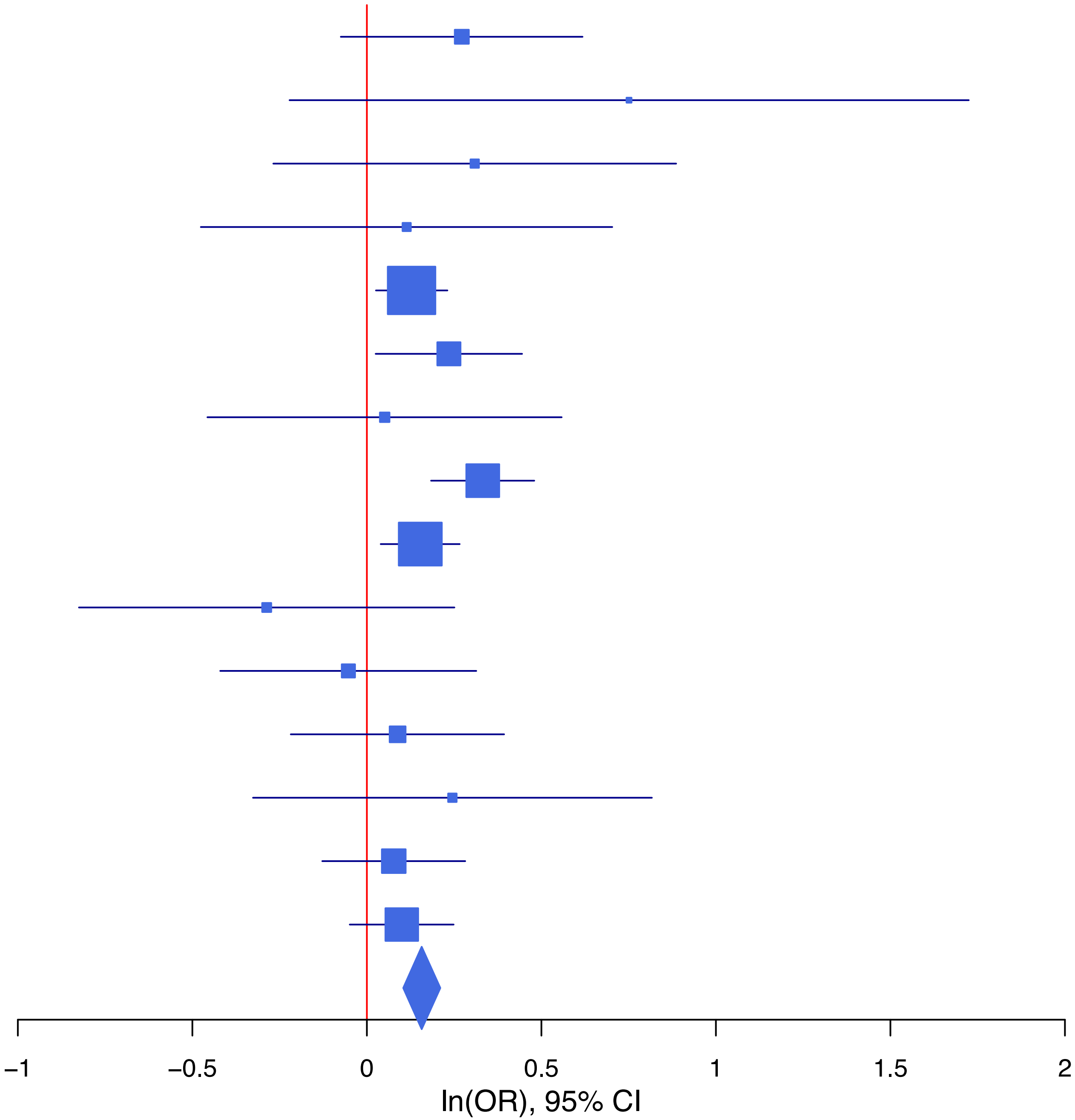

**rs1894401 G/A 15:91429042**

| Cohort | P | ln(OR) | SE |
|---|---|---|---|
| ALSPC_EUR | 0.218 | −0.068 | 0.055 |
| BEPS7_EUR | 0.0995 | −0.278 | 0.169 |
| BHRCM_EUR | 0.917 | 0.016 | 0.153 |
| BOR17_EUR | 0.2113 | −0.106 | 0.085 |
| BOR2C_EUR | 0.8694 | 0.014 | 0.086 |
| BOR2E_EUR | 0.187 | 0.232 | 0.176 |
| COGA1_EUR | 0.5051 | −0.04 | 0.059 |
| CUINT_EUR | 0.8835 | −0.014 | 0.099 |
| ESTB2_EUR | 0.6696 | 0.01 | 0.023 |
| FINNG_EUR | 0.1938 | −0.03 | 0.023 |
| GEDIS_EUR | 0.4488 | 0.098 | 0.129 |
| GISS1_EUR | 0.1021 | −0.127 | 0.078 |
| GISS2_EUR | 0.4938 | −0.063 | 0.092 |
| IPSYC_EUR | 0.3358 | −0.019 | 0.02 |
| JANS3_EUR | 0.3362 | 0.098 | 0.102 |
| JANS4_EUR | 0.2652 | −0.135 | 0.121 |
| MVPXQ_EUR | 0.0042 | −0.037 | 0.013 |
| PGCBD_EUR | 0.2614 | −0.03 | 0.026 |
| PGCMD_EUR | 0.0367 | −0.074 | 0.035 |
| PGCPT_EUR | 0.4035 | −0.048 | 0.058 |
| PGCSZ_EUR | 0.3317 | −0.037 | 0.038 |
| PRFCT_EUR | 0.3514 | −0.041 | 0.044 |
| PSYCR_EUR | 0.8819 | 0.013 | 0.087 |
| QIMRB_EUR | 0.0081 | −0.071 | 0.027 |
| UKBJC_EUR | 0.001 | −0.067 | 0.02 |
| YPENN_EUR | 0.7638 | −0.038 | 0.128 |
| **meta** | **4.58e−08** | **−0.037** | **0.007** |

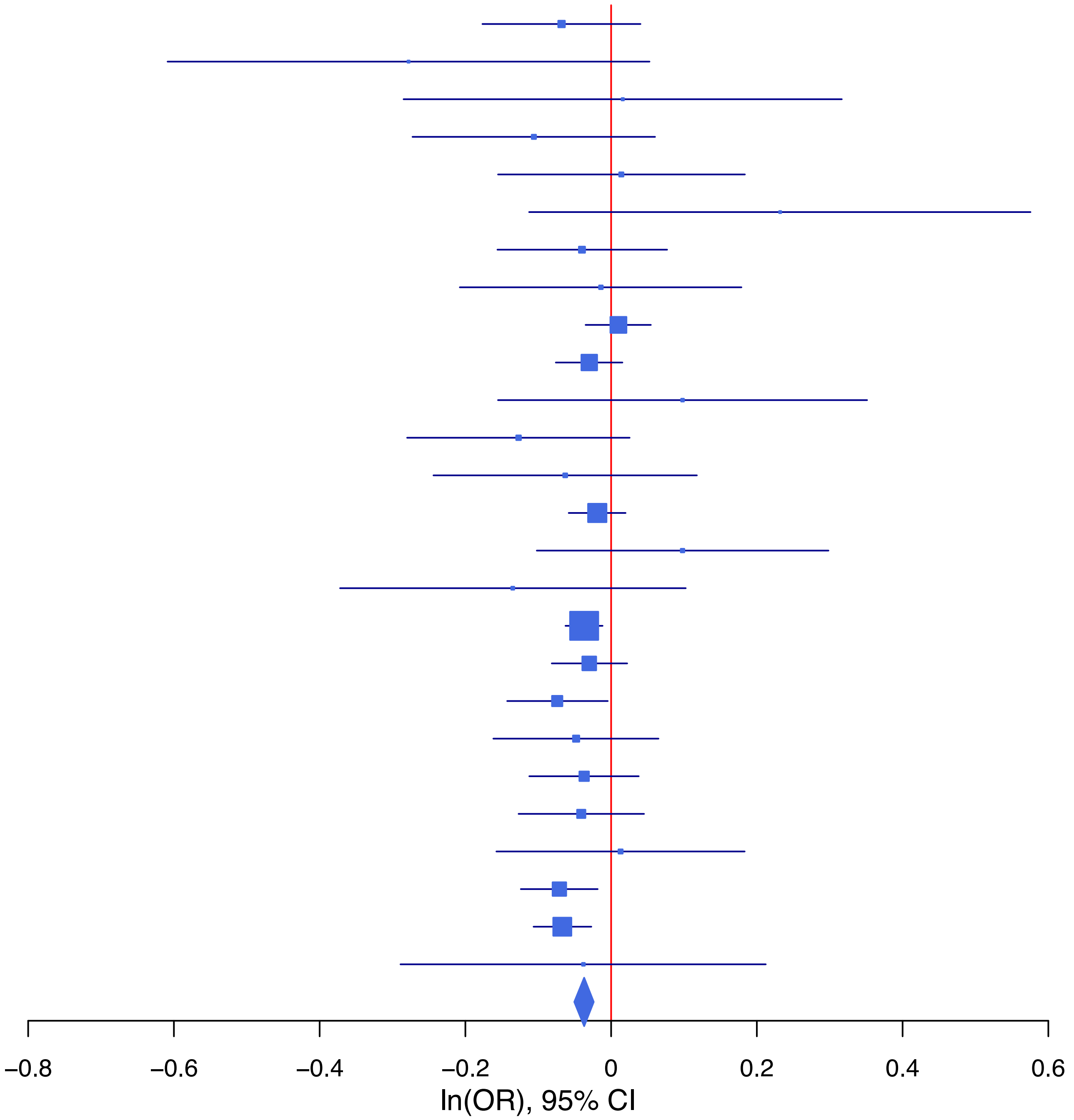

**rs2295402 A/T 14:103338324**

| Cohort | P | ln(OR) | SE |
|---|---|---|---|
| ALSPC_EUR | 0.0088 | 0.17 | 0.065 |
| BEPS7_EUR | 0.8371 | 0.046 | 0.226 |
| BHRCM_EUR | 0.9071 | 0.024 | 0.204 |
| BOR17_EUR | 0.6678 | 0.047 | 0.109 |
| BOR2C_EUR | 0.4137 | −0.088 | 0.108 |
| BOR2E_EUR | 0.4328 | −0.163 | 0.208 |
| COGA1_EUR | 0.7583 | −0.022 | 0.073 |
| CUINT_EUR | 0.9303 | −0.011 | 0.124 |
| ESTB2_EUR | 0.055 | 0.056 | 0.029 |
| FINNG_EUR | 0.006 | 0.073 | 0.027 |
| GEDIS_EUR | 0.7434 | −0.052 | 0.16 |
| GISS1_EUR | 0.4141 | −0.083 | 0.102 |
| GISS2_EUR | 0.7618 | 0.035 | 0.116 |
| IPSYC_EUR | 0.1648 | 0.034 | 0.024 |
| JANS3_EUR | 0.8992 | −0.017 | 0.132 |
| JANS4_EUR | 0.1983 | −0.189 | 0.147 |
| MIREC_EUR | 0.9735 | −0.006 | 0.19 |
| MVPXQ_EUR | 8.66e−07 | 0.078 | 0.016 |
| PGCBD_EUR | 0.5228 | −0.021 | 0.033 |
| PGCPT_EUR | 0.2403 | 0.083 | 0.07 |
| PGCSZ_EUR | 0.4012 | 0.039 | 0.046 |
| PRFCT_EUR | 0.0546 | 0.1 | 0.052 |
| PSYCR_EUR | 0.1992 | 0.145 | 0.113 |
| QIMRB_EUR | 0.6521 | 0.016 | 0.035 |
| UKBJC_EUR | 0.0015 | 0.08 | 0.025 |
| YPENN_EUR | 0.622 | −0.078 | 0.159 |
| **meta** | **1.95e−10** | **0.054** | **0.009** |

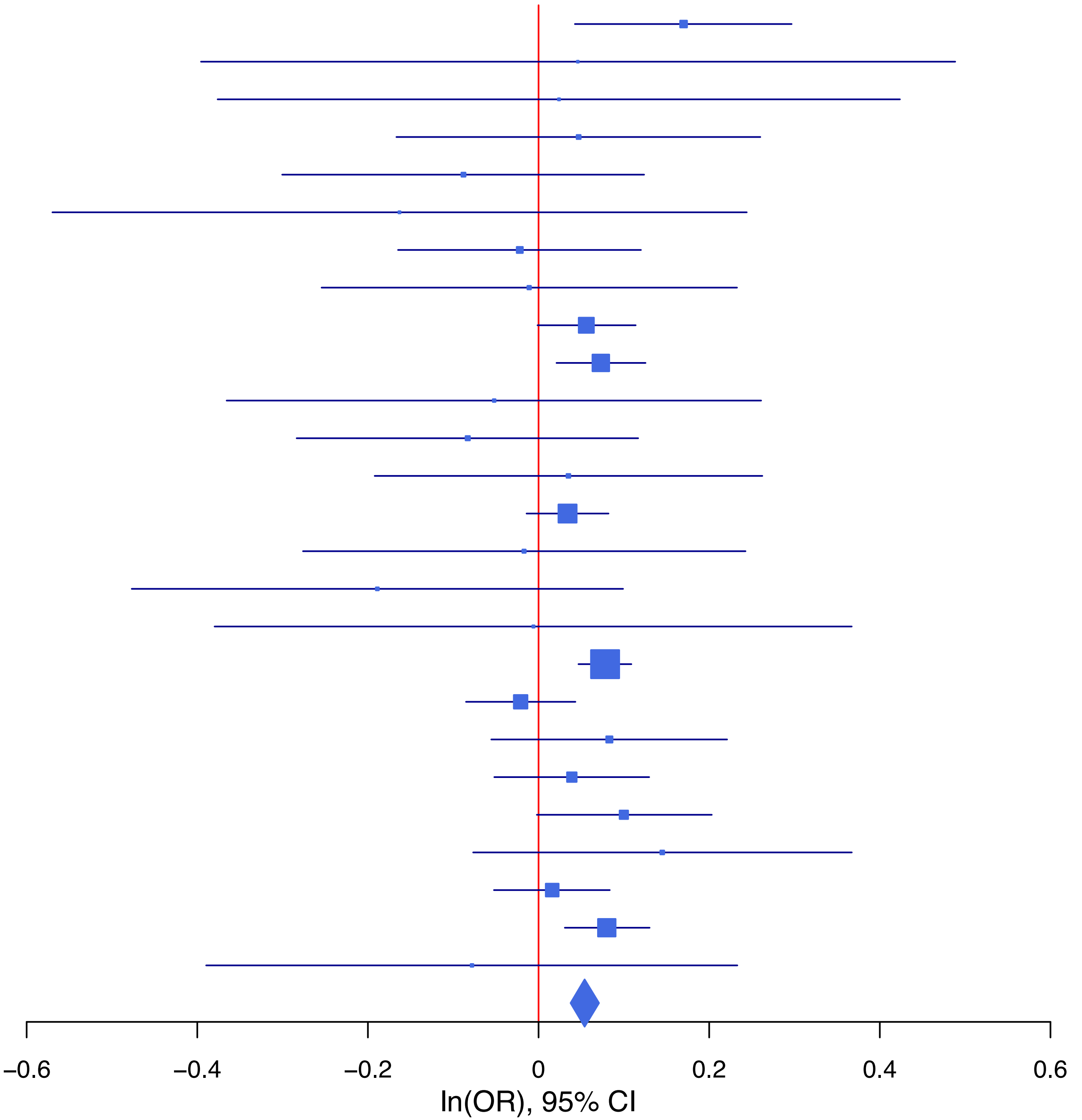

**rs2503185 G/A 1:66461401**

| Cohort | P | ln(OR) | SE |
|---|---|---|---|
| ALSPC_EUR | 0.525 | −0.034 | 0.054 |
| BEPS7_EUR | 0.7202 | −0.063 | 0.176 |
| BHRCM_EUR | 0.3888 | −0.136 | 0.158 |
| BOR17_EUR | 0.181 | 0.113 | 0.084 |
| BOR2C_EUR | 0.0615 | −0.163 | 0.087 |
| BOR2E_EUR | 0.5384 | 0.108 | 0.176 |
| COGA1_EUR | 0.9308 | −0.005 | 0.059 |
| CUINT_EUR | 0.8089 | −0.024 | 0.1 |
| ESTB2_EUR | 0.0973 | −0.038 | 0.023 |
| FINNG_EUR | 0.0782 | −0.04 | 0.023 |
| GEDIS_EUR | 0.5361 | −0.079 | 0.128 |
| GISS1_EUR | 0.258 | 0.088 | 0.078 |
| GISS2_EUR | 0.1551 | 0.13 | 0.091 |
| IPSYC_EUR | 4.03e−06 | −0.092 | 0.02 |
| JANS3_EUR | 0.8184 | −0.023 | 0.102 |
| JANS4_EUR | 0.9833 | −0.002 | 0.114 |
| MIREC_EUR | 0.525 | 0.096 | 0.151 |
| MVPXQ_EUR | 0.0018 | −0.041 | 0.013 |
| PGCBD_EUR | 0.0503 | −0.05 | 0.026 |
| PGCED_EUR | 0.4949 | −0.078 | 0.114 |
| PGCMD_EUR | 0.0648 | −0.063 | 0.034 |
| PGCPT_EUR | 0.642 | −0.027 | 0.058 |
| PGCSZ_EUR | 0.2226 | −0.045 | 0.037 |
| PRFCT_EUR | 0.0065 | −0.119 | 0.044 |
| PSYCR_EUR | 0.7537 | −0.027 | 0.087 |
| QIMRB_EUR | 0.8134 | −0.006 | 0.027 |
| UKBJC_EUR | 0.0189 | −0.048 | 0.02 |
| YPENN_EUR | 0.0715 | 0.229 | 0.127 |
| **meta** | **4.84e−11** | **−0.044** | **0.007** |

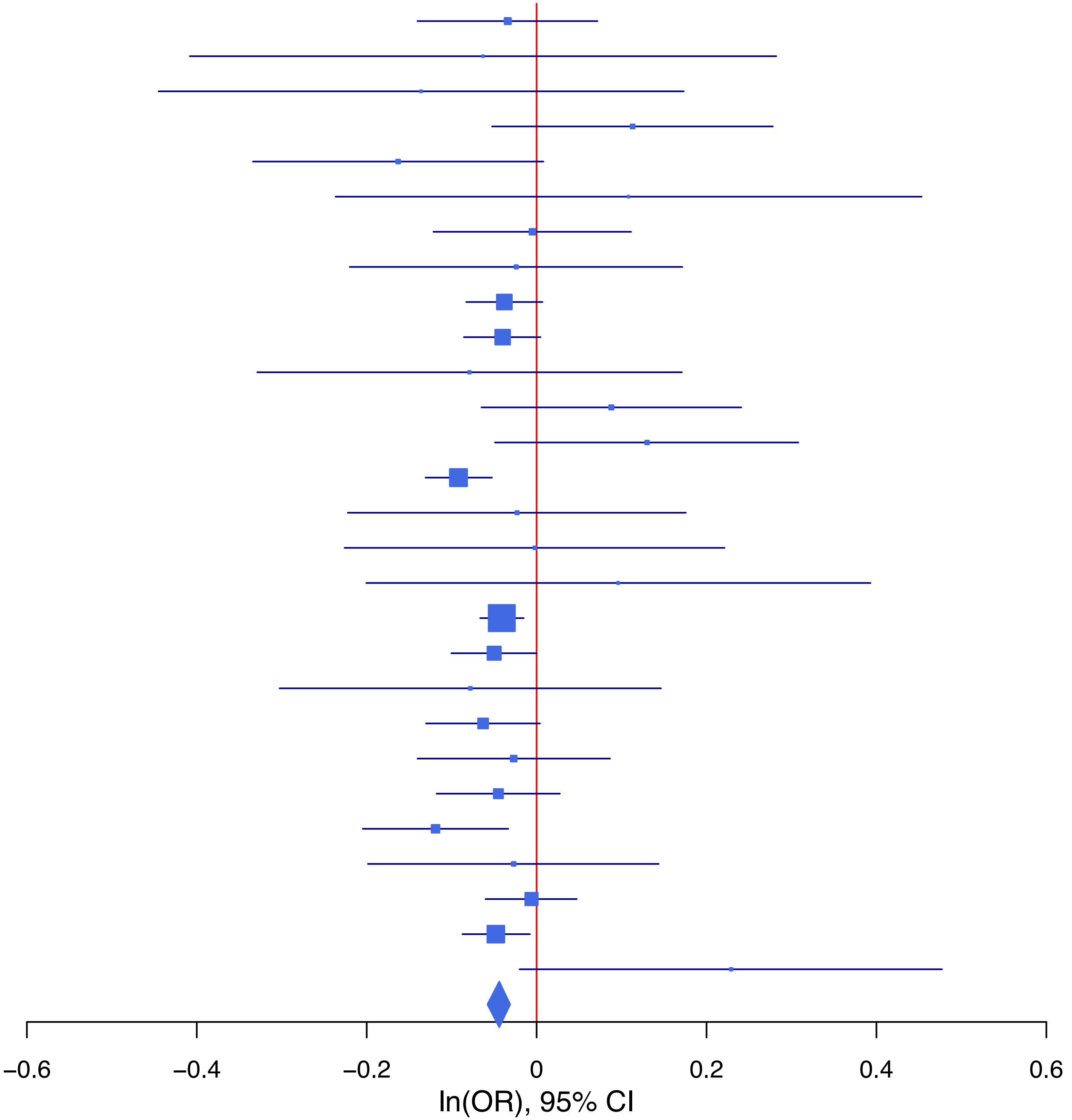

**rs35942385 T/G 2:144208523**

| Cohort | P | ln(OR) | SE |
|---|---|---|---|
| ALSPC_EUR | 0.3846 | 0.048 | 0.055 |
| BEPS7_EUR | 0.6335 | 0.09 | 0.188 |
| BHRCM_EUR | 0.7833 | −0.046 | 0.167 |
| BOR17_EUR | 0.2627 | −0.099 | 0.088 |
| BOR2C_EUR | 0.5127 | −0.058 | 0.088 |
| BOR2E_EUR | 0.5254 | −0.118 | 0.186 |
| COGA1_EUR | 0.4845 | −0.043 | 0.061 |
| CUINT_EUR | 0.4602 | 0.076 | 0.103 |
| ESTB2_EUR | 0.0589 | −0.047 | 0.025 |
| FINNG_EUR | 0.0218 | −0.056 | 0.024 |
| GEDIS_EUR | 7e−04 | −0.496 | 0.147 |
| GISS1_EUR | 0.0458 | −0.17 | 0.085 |
| GISS2_EUR | 0.8264 | 0.022 | 0.101 |
| IPSYC_EUR | 0.0102 | −0.053 | 0.02 |
| JANS3_EUR | 0.0385 | 0.223 | 0.108 |
| MVPXQ_EUR | 9.56e−07 | −0.064 | 0.013 |
| PGCBD_EUR | 0.3855 | 0.023 | 0.027 |
| PGCED_EUR | 0.7154 | 0.043 | 0.118 |
| PGCMD_EUR | 0.0738 | −0.064 | 0.036 |
| PGCPT_EUR | 0.5448 | 0.036 | 0.06 |
| PGCSZ_EUR | 0.1958 | −0.05 | 0.039 |
| PRFCT_EUR | 0.0297 | −0.1 | 0.046 |
| PSYCR_EUR | 0.6664 | 0.04 | 0.092 |
| QIMRB_EUR | 0.1037 | −0.046 | 0.028 |
| UKBJC_EUR | 0.4793 | −0.015 | 0.021 |
| YPENN_EUR | 0.8745 | −0.021 | 0.132 |
| **meta** | **7.08e−10** | **−0.043** | **0.007** |

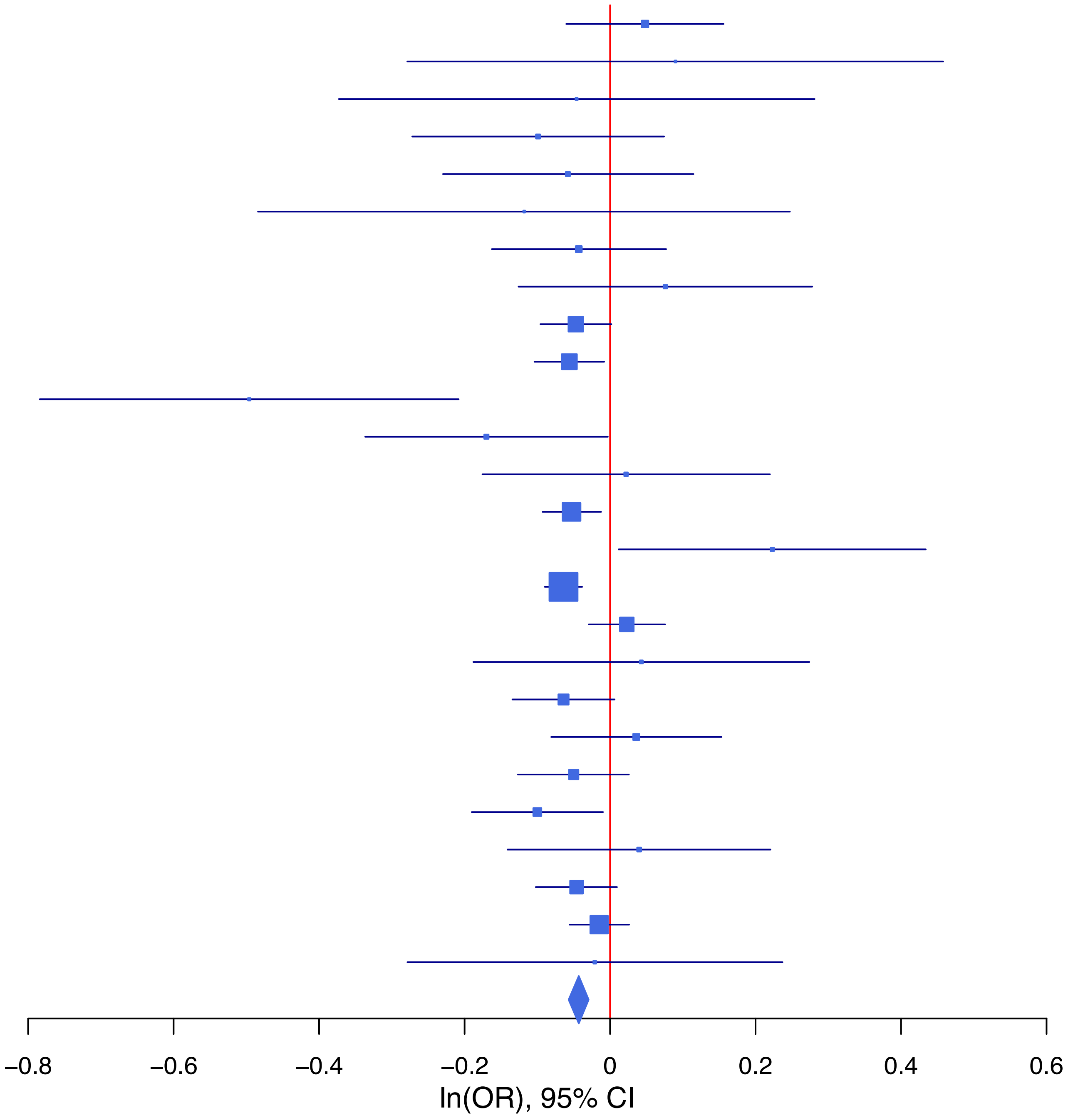

**rs36065861   T/C   4:166063595**

| Cohort | P | ln(OR) | SE |
|---|---|---|---|
| ADHEA_EUR | 0.6919 | −0.041 | 0.103 |
| ALSPC_EUR | 0.5565 | 0.046 | 0.078 |
| BEPS7_EUR | 0.7954 | 0.064 | 0.247 |
| BHRCM_EUR | 0.7173 | −0.105 | 0.289 |
| BOR17_EUR | 0.6737 | −0.056 | 0.133 |
| BOR2C_EUR | 0.0213 | −0.299 | 0.13 |
| BOR2E_EUR | 0.663 | −0.135 | 0.31 |
| COGA1_EUR | 0.0214 | −0.208 | 0.09 |
| CUINT_EUR | 0.5095 | 0.101 | 0.153 |
| ESTB2_EUR | 0.2099 | −0.041 | 0.033 |
| FINNG_EUR | 0.0216 | −0.068 | 0.03 |
| GEDIS_EUR | 0.7703 | −0.057 | 0.196 |
| GISS1_EUR | 0.102 | −0.191 | 0.117 |
| GISS2_EUR | 0.9691 | 0.005 | 0.133 |
| IPSYC_EUR | 0.1735 | −0.039 | 0.029 |
| JANS3_EUR | 0.1186 | −0.26 | 0.167 |
| JANS4_EUR | 0.8559 | 0.035 | 0.191 |
| MVPXQ_EUR | 0.0125 | −0.048 | 0.019 |
| PGCBD_EUR | 0.1435 | −0.058 | 0.04 |
| PGCED_EUR | 0.8423 | 0.038 | 0.19 |
| PGCMD_EUR | 0.8295 | −0.011 | 0.053 |
| PGCPT_EUR | 0.6573 | −0.039 | 0.088 |
| PGCSZ_EUR | 0.5533 | −0.033 | 0.056 |
| PRFCT_EUR | 0.0181 | −0.142 | 0.06 |
| PSYCR_EUR | 0.0108 | −0.369 | 0.145 |
| QIMRB_EUR | 0.0843 | −0.07 | 0.04 |
| UKBJC_EUR | 0.167 | −0.04 | 0.029 |
| YPENN_EUR | 0.6326 | −0.091 | 0.19 |
| **meta** | **3.59e−08** | **−0.054** | **0.01** |

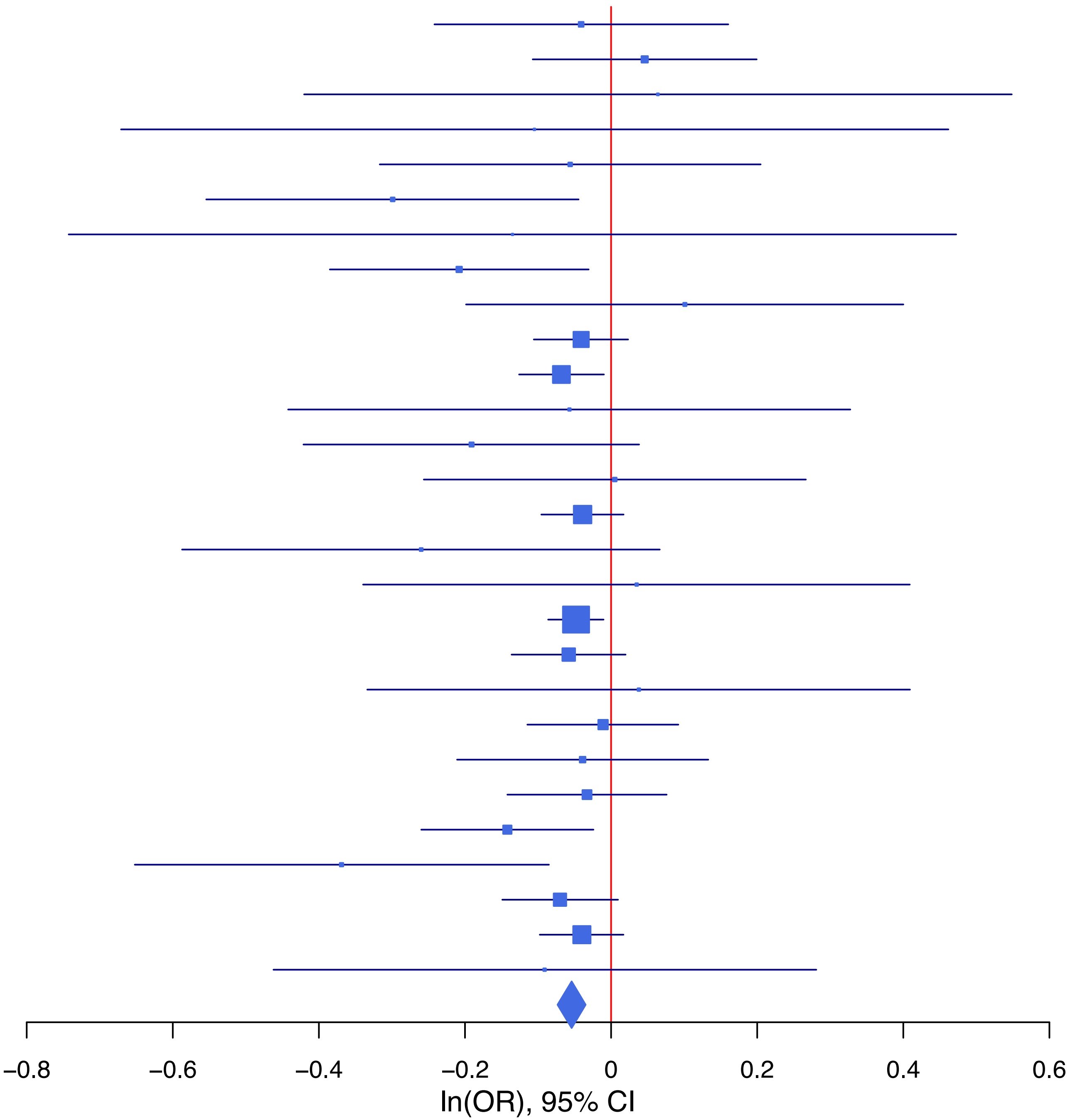

**rs4267058 C/T 11:28645584**

| Cohort | P | ln(OR) | SE |
|---|---|---|---|
| ALSPC_EUR | 0.8713 | −0.009 | 0.056 |
| BEPS7_EUR | 0.2638 | 0.197 | 0.176 |
| BHRCM_EUR | 0.3065 | −0.17 | 0.167 |
| BOR17_EUR | 0.6204 | 0.044 | 0.09 |
| BOR2C_EUR | 0.3906 | −0.076 | 0.089 |
| BOR2E_EUR | 0.004 | −0.555 | 0.193 |
| COGA1_EUR | 0.8856 | −0.009 | 0.06 |
| CUINT_EUR | 0.1547 | −0.155 | 0.109 |
| ESTB2_EUR | 0.0674 | −0.042 | 0.023 |
| FINNG_EUR | 0.7559 | 0.007 | 0.024 |
| GEDIS_EUR | 0.7645 | −0.041 | 0.138 |
| GISS1_EUR | 0.8443 | 0.016 | 0.079 |
| GISS2_EUR | 0.8226 | −0.021 | 0.093 |
| JANS3_EUR | 0.4795 | −0.076 | 0.108 |
| JANS4_EUR | 0.3092 | −0.125 | 0.122 |
| MIREC_EUR | 0.9978 | 0 | 0.154 |
| MVPXQ_EUR | 0.0012 | −0.043 | 0.013 |
| PGCBD_EUR | 0.0686 | −0.048 | 0.027 |
| PGCED_EUR | 0.5292 | −0.073 | 0.116 |
| PGCMD_EUR | 0.004 | −0.102 | 0.035 |
| PGCPT_EUR | 0.3367 | 0.057 | 0.06 |
| PGCSZ_EUR | 0.2552 | −0.044 | 0.038 |
| PRFCT_EUR | 0.4018 | −0.038 | 0.046 |
| PSYCR_EUR | 0.0595 | −0.172 | 0.092 |
| QIMRB_EUR | 0.0061 | −0.077 | 0.028 |
| UKBJC_EUR | 0.098 | −0.034 | 0.021 |
| YPENN_EUR | 0.9558 | −0.007 | 0.132 |
| **meta** | **3.33e−08** | **−0.041** | **0.007** |

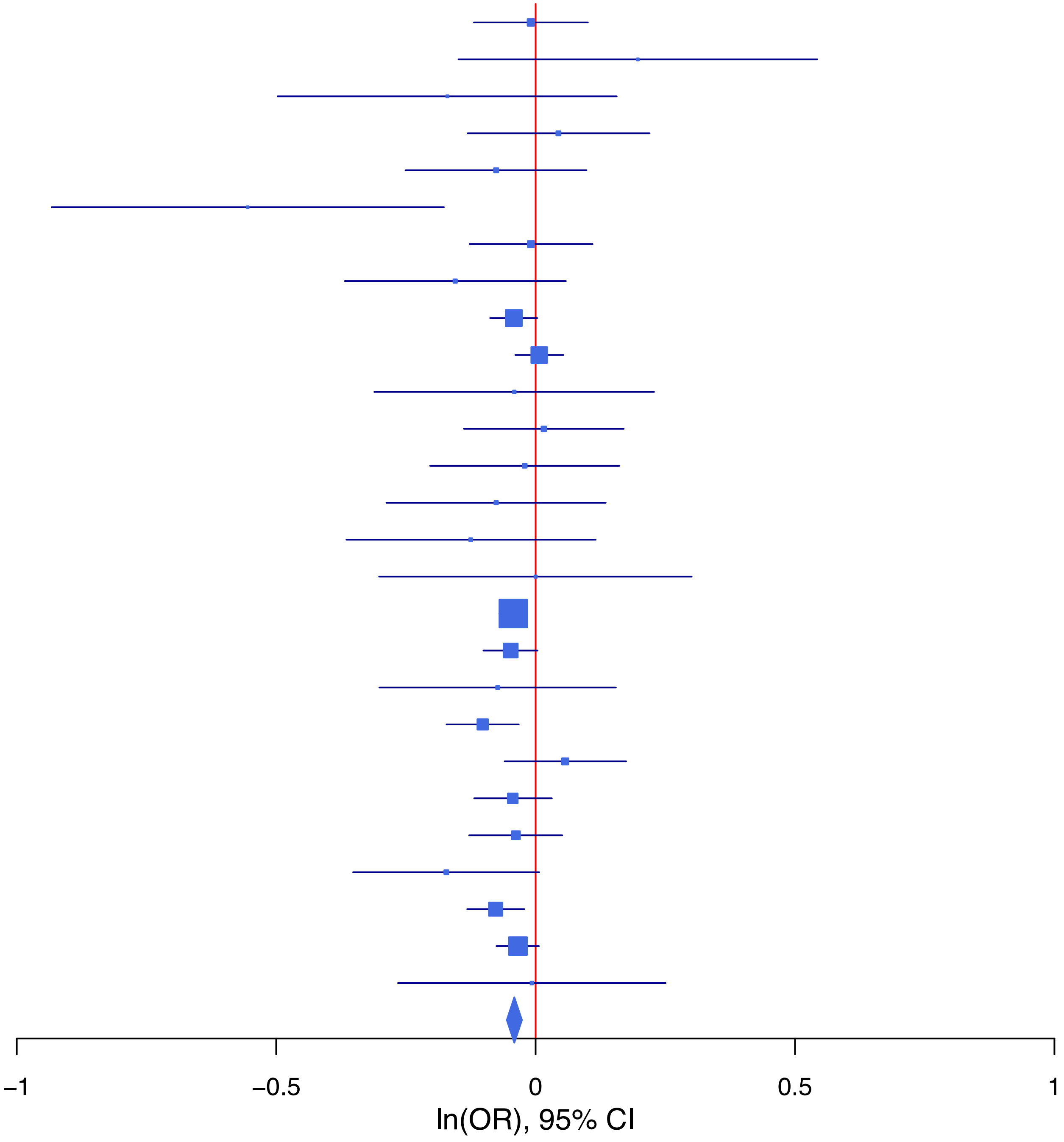

**rs4305732 A/G 6:152240448**

| Cohort | P | ln(OR) | SE |
|---|---|---|---|
| ALSPC_EUR | 0.6778 | −0.023 | 0.056 |
| BEPS7_EUR | 0.6492 | 0.085 | 0.187 |
| BHRCM_EUR | 0.5377 | −0.111 | 0.18 |
| BOR17_EUR | 0.1939 | 0.118 | 0.091 |
| BOR2C_EUR | 0.2707 | 0.1 | 0.091 |
| BOR2E_EUR | 0.1923 | 0.242 | 0.186 |
| COGA1_EUR | 0.1446 | 0.089 | 0.061 |
| CUINT_EUR | 0.3035 | −0.108 | 0.105 |
| ESTB2_EUR | 5e−04 | 0.085 | 0.025 |
| FINNG_EUR | 0.5498 | 0.014 | 0.024 |
| GEDIS_EUR | 0.8972 | 0.018 | 0.136 |
| GISS1_EUR | 0.4307 | −0.069 | 0.088 |
| GISS2_EUR | 0.8851 | 0.014 | 0.099 |
| IPSYC_EUR | 0.32 | 0.021 | 0.021 |
| JANS3_EUR | 0.4963 | −0.075 | 0.11 |
| JANS4_EUR | 0.7285 | 0.044 | 0.128 |
| MVPXQ_EUR | 8.99e−07 | 0.067 | 0.014 |
| PGCBD_EUR | 0.3649 | 0.025 | 0.027 |
| PGCED_EUR | 0.0702 | −0.227 | 0.125 |
| PGCMD_EUR | 0.233 | 0.043 | 0.036 |
| PGCPT_EUR | 0.1496 | 0.086 | 0.06 |
| PGCSZ_EUR | 0.419 | 0.031 | 0.039 |
| PRFCT_EUR | 0.0017 | 0.144 | 0.046 |
| PSYCR_EUR | 0.1498 | 0.132 | 0.092 |
| QIMRB_EUR | 0.2196 | 0.035 | 0.028 |
| UKBJC_EUR | 2.39e−05 | 0.088 | 0.021 |
| YPENN_EUR | 0.6556 | 0.058 | 0.131 |
| **meta** | **2.19e−13** | **0.052** | **0.007** |

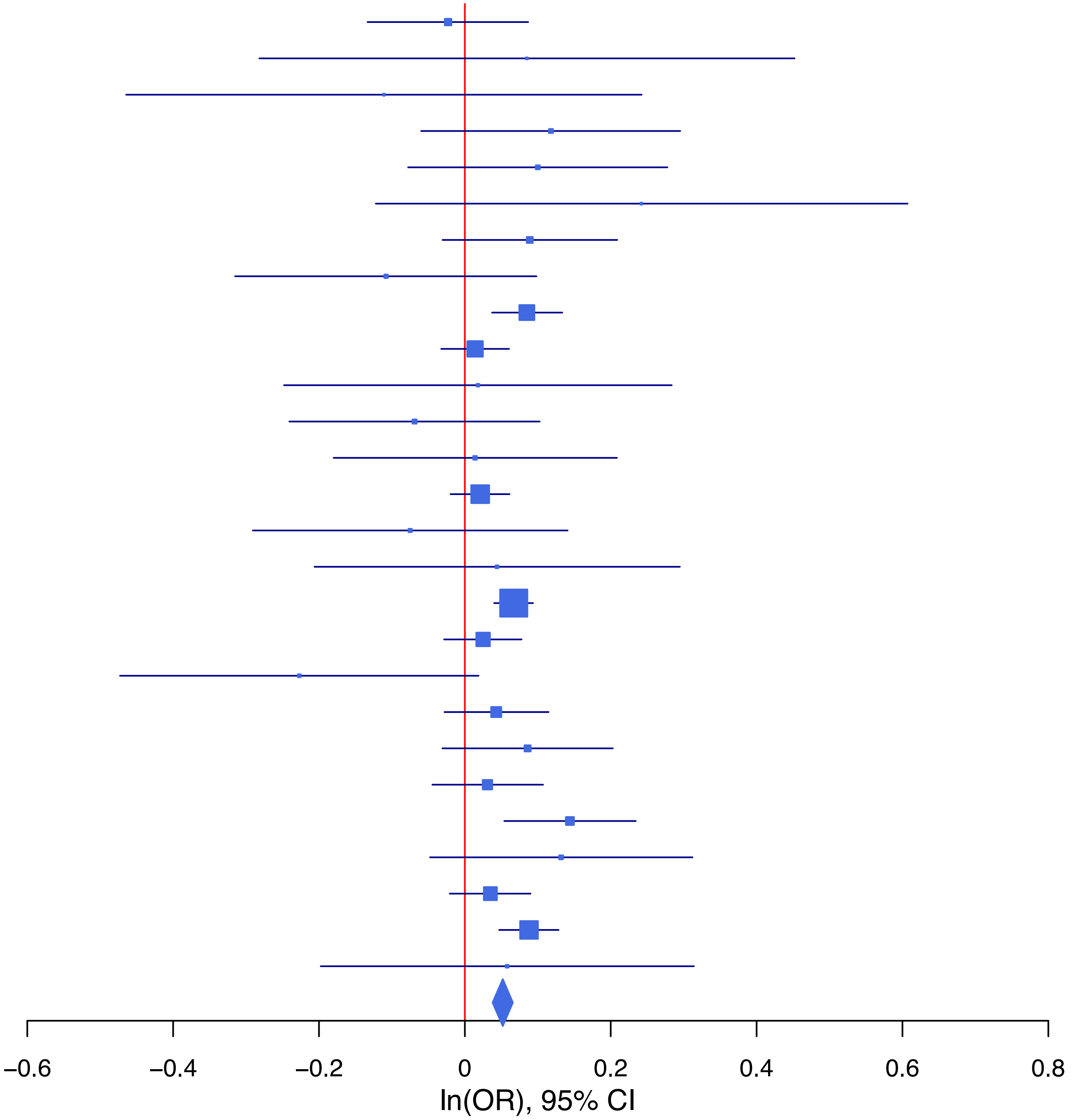

**rs4974203 T/C 3:56390992**

| Cohort | P | ln(OR) | SE |
|---|---|---|---|
| ADHEA_EUR | 0.97 | 0.003 | 0.075 |
| ALSPC_EUR | 0.1236 | 0.091 | 0.059 |
| BEPS7_EUR | 0.8633 | −0.036 | 0.207 |
| BHRCM_EUR | 0.3389 | −0.193 | 0.202 |
| BOR17_EUR | 0.0658 | 0.181 | 0.098 |
| BOR2C_EUR | 0.6499 | −0.045 | 0.1 |
| BOR2E_EUR | 0.1239 | 0.304 | 0.198 |
| COGA1_EUR | 0.8579 | 0.012 | 0.064 |
| CUINT_EUR | 0.3514 | 0.106 | 0.114 |
| ESTB2_EUR | 0.103 | 0.043 | 0.026 |
| FINNG_EUR | 0.4073 | 0.021 | 0.025 |
| GEDIS_EUR | 0.2394 | 0.166 | 0.141 |
| GISS1_EUR | 0.5787 | 0.052 | 0.093 |
| GISS2_EUR | 0.9132 | −0.012 | 0.109 |
| IPSYC_EUR | 8e−04 | 0.072 | 0.021 |
| JANS3_EUR | 0.5429 | −0.071 | 0.117 |
| JANS4_EUR | 0.661 | 0.058 | 0.132 |
| MIREC_EUR | 0.2058 | −0.223 | 0.176 |
| MVPXQ_EUR | 0.0199 | 0.033 | 0.014 |
| PGCBD_EUR | 0.2811 | 0.031 | 0.029 |
| PGCED_EUR | 0.7211 | −0.047 | 0.131 |
| PGCMD_EUR | 0.1431 | 0.056 | 0.038 |
| PGCPT_EUR | 0.5048 | −0.043 | 0.064 |
| PGCSZ_EUR | 0.4098 | 0.034 | 0.041 |
| PRFCT_EUR | 0.1975 | 0.062 | 0.048 |
| PSYCR_EUR | 0.9863 | −0.002 | 0.099 |
| QIMRB_EUR | 0.0315 | 0.066 | 0.031 |
| UKBJC_EUR | 0.0054 | 0.061 | 0.022 |
| YPENN_EUR | 0.7014 | −0.056 | 0.147 |
| **meta** | **8.10e−09** | **0.043** | **0.007** |

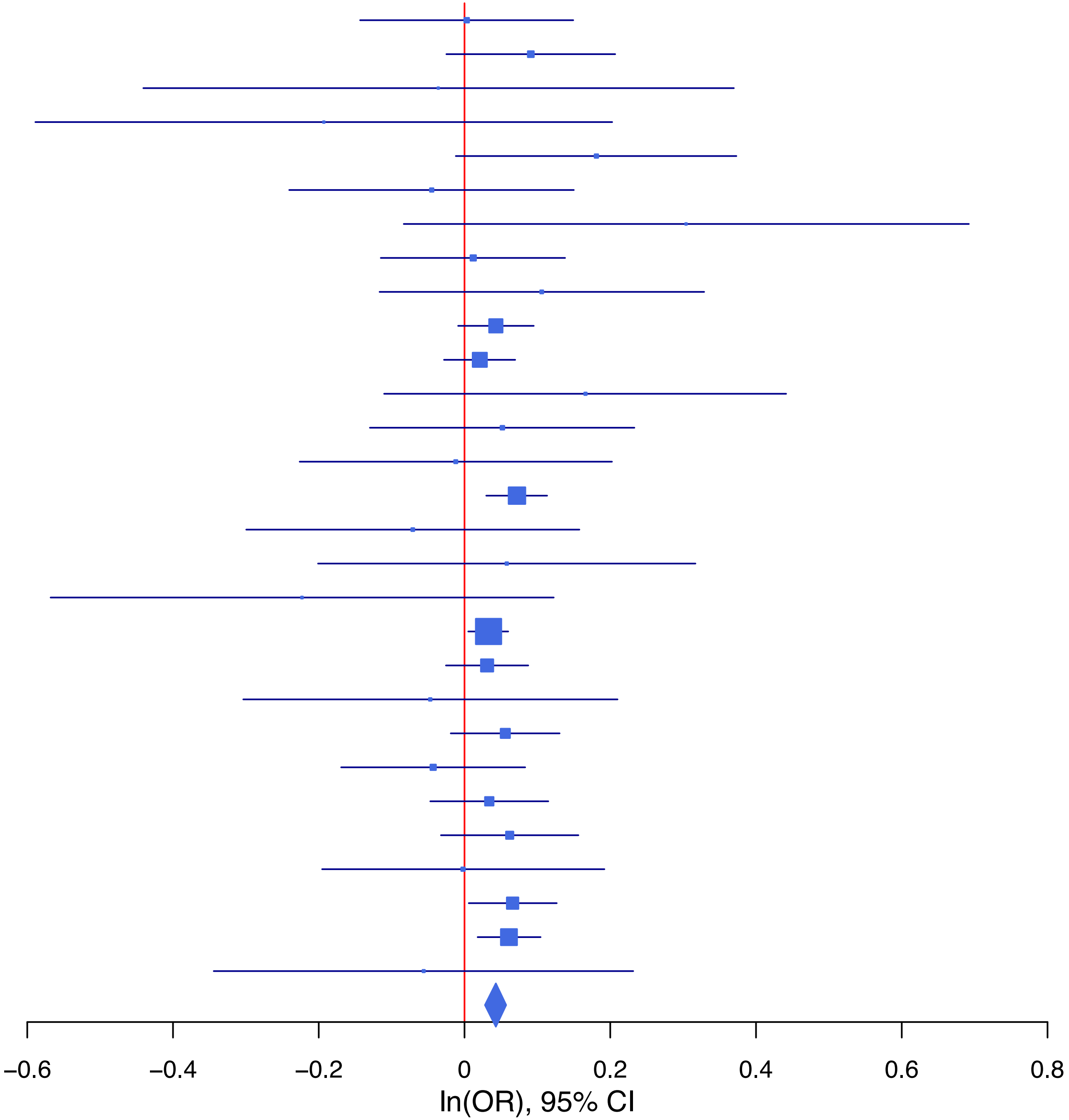

**rs499168 A/T 1:38228797**

| Cohort | P | ln(OR) | SE |
|---|---|---|---|
| ADHEA_EUR | 0.37 | −0.061 | 0.068 |
| ALSPC_EUR | 0.4812 | 0.038 | 0.054 |
| BEPS7_EUR | 0.257 | −0.202 | 0.178 |
| BHRCM_EUR | 0.6076 | −0.08 | 0.156 |
| BOR17_EUR | 0.4035 | −0.073 | 0.087 |
| BOR2C_EUR | 0.3955 | 0.075 | 0.088 |
| BOR2E_EUR | 0.7104 | 0.069 | 0.185 |
| COGA1_EUR | 0.4647 | 0.043 | 0.059 |
| CUINT_EUR | 0.4102 | 0.083 | 0.101 |
| ESTB2_EUR | 0.0752 | −0.041 | 0.023 |
| FINNG_EUR | 0.0425 | −0.046 | 0.023 |
| GEDIS_EUR | 0.6274 | −0.063 | 0.13 |
| GISS1_EUR | 0.1036 | −0.129 | 0.079 |
| GISS2_EUR | 0.4017 | 0.08 | 0.095 |
| IPSYC_EUR | 0.031 | −0.043 | 0.02 |
| JANS3_EUR | 0.8317 | −0.022 | 0.105 |
| JANS4_EUR | 0.6562 | 0.054 | 0.12 |
| MVPXQ_EUR | 0.001 | −0.042 | 0.013 |
| PGCBD_EUR | 0.1098 | −0.042 | 0.026 |
| PGCED_EUR | 0.3287 | −0.116 | 0.119 |
| PGCMD_EUR | 0.1233 | −0.055 | 0.035 |
| PGCPT_EUR | 0.8999 | −0.007 | 0.058 |
| PGCSZ_EUR | 0.7789 | 0.011 | 0.038 |
| PRFCT_EUR | 6e−04 | −0.152 | 0.044 |
| PSYCR_EUR | 0.1234 | −0.136 | 0.088 |
| QIMRB_EUR | 0.7763 | 0.008 | 0.027 |
| UKBJC_EUR | 0.0069 | −0.055 | 0.02 |
| YPENN_EUR | 0.0988 | 0.211 | 0.128 |
| **meta** | **1.31e−08** | **−0.038** | **0.007** |

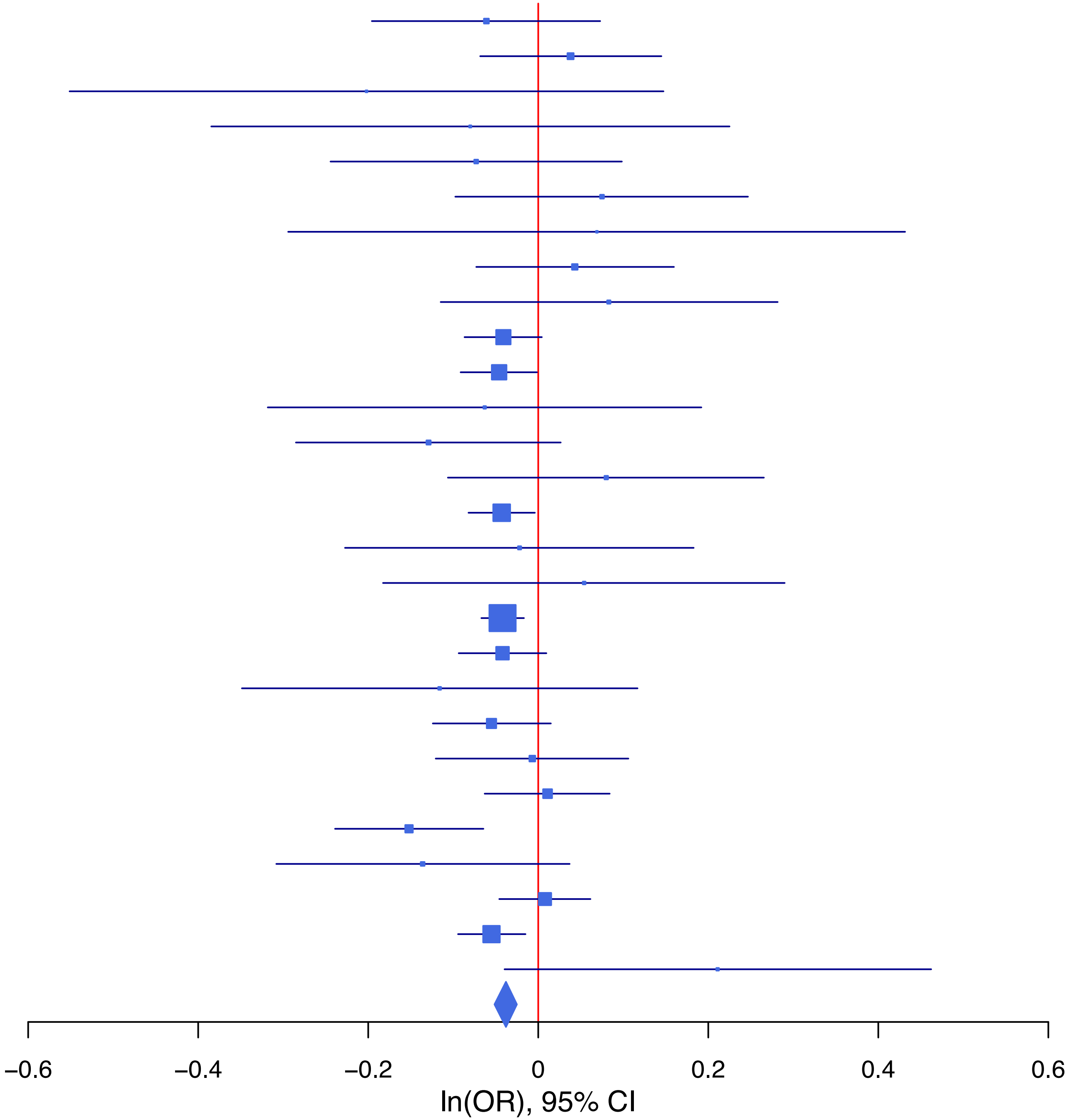

**rs55995895 T/C 7:1862183**

| Cohort | P | ln(OR) | SE |
|---|---|---|---|
| ADHEA_EUR | 0.2072 | −0.121 | 0.096 |
| ALSPC_EUR | 0.7761 | 0.022 | 0.077 |
| BEPS7_EUR | 0.2763 | −0.254 | 0.233 |
| BOR17_EUR | 0.8961 | 0.014 | 0.11 |
| BOR2C_EUR | 0.324 | −0.11 | 0.111 |
| BOR2E_EUR | 0.4332 | −0.183 | 0.234 |
| COGA1_EUR | 0.1027 | −0.132 | 0.081 |
| CUINT_EUR | 0.4381 | 0.1 | 0.129 |
| ESTB2_EUR | 0.0063 | −0.074 | 0.027 |
| FINNG_EUR | 8e−04 | −0.09 | 0.027 |
| GISS1_EUR | 0.7848 | 0.025 | 0.091 |
| GISS2_EUR | 0.7959 | −0.027 | 0.103 |
| IPSYC_EUR | 0.004 | −0.081 | 0.028 |
| JANS3_EUR | 0.277 | −0.157 | 0.144 |
| JANS4_EUR | 0.0214 | −0.376 | 0.163 |
| MIREC_EUR | 0.7999 | −0.053 | 0.208 |
| MVPXQ_EUR | 7e−04 | −0.06 | 0.018 |
| PGCBD_EUR | 0.1966 | 0.044 | 0.034 |
| PGCED_EUR | 0.5168 | −0.108 | 0.167 |
| PGCMD_EUR | 0.2842 | 0.049 | 0.046 |
| PGCPT_EUR | 0.2804 | −0.086 | 0.079 |
| PGCSZ_EUR | 0.3063 | 0.05 | 0.049 |
| PRFCT_EUR | 0.6729 | −0.024 | 0.057 |
| PSYCR_EUR | 0.0621 | −0.204 | 0.109 |
| QIMRB_EUR | 0.0018 | −0.115 | 0.037 |
| UKBJC_EUR | 0.1022 | −0.046 | 0.028 |
| YPENN_EUR | 0.4864 | −0.123 | 0.177 |
| **meta** | **8.58e−10** | **−0.055** | **0.009** |

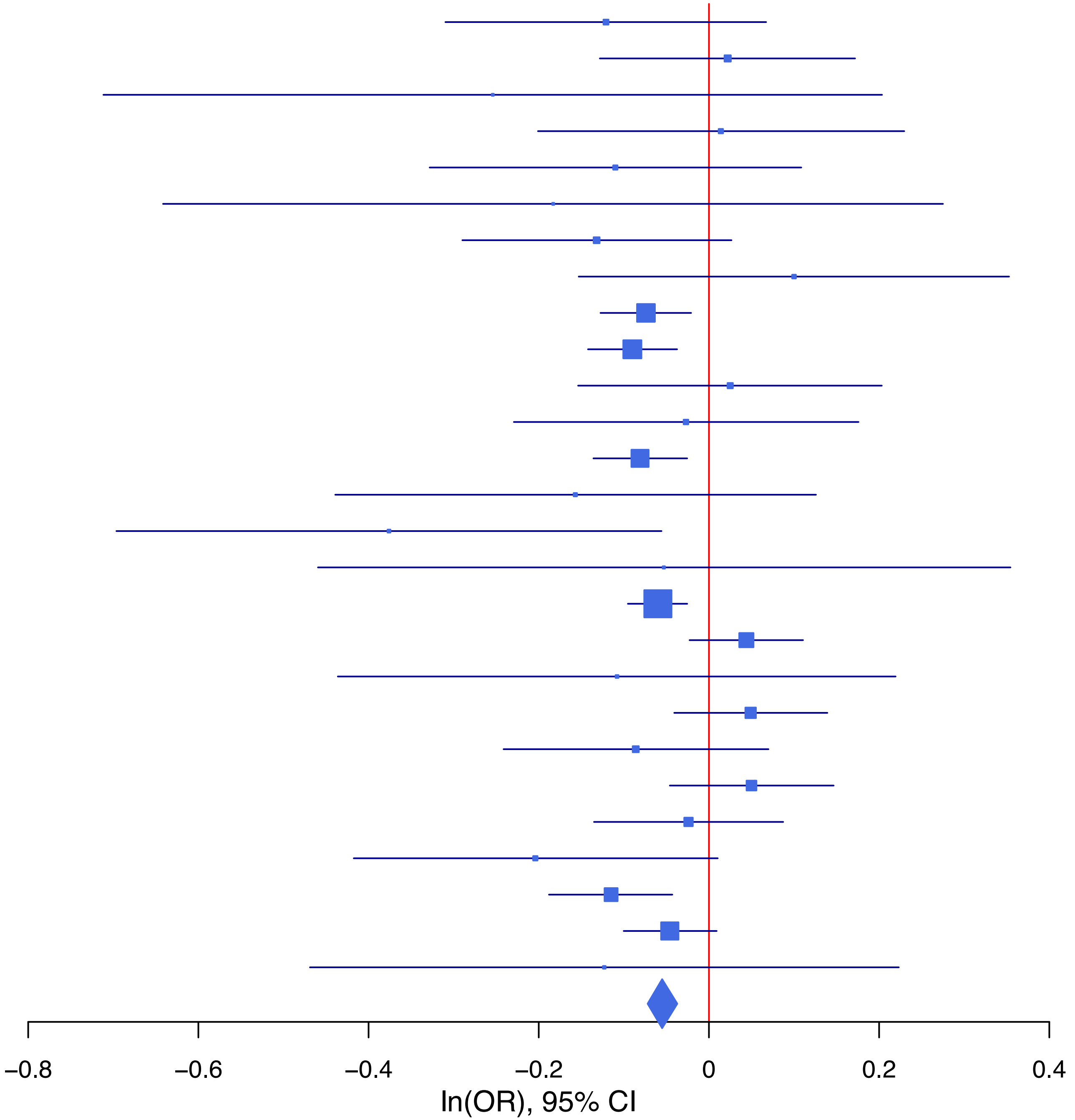

**rs58907748 G/T 6:156468874**

| Cohort | P | ln(OR) | SE |
|---|---|---|---|
| ADHEA_EUR | 0.3721 | 0.111 | 0.124 |
| ALSPC_EUR | 0.6451 | −0.05 | 0.108 |
| BEPS7_EUR | 0.6622 | −0.16 | 0.366 |
| BHRCM_EUR | 0.8506 | −0.057 | 0.302 |
| BOR17_EUR | 0.1115 | −0.273 | 0.172 |
| BOR2C_EUR | 0.1097 | 0.251 | 0.157 |
| BOR2E_EUR | 0.4477 | 0.247 | 0.326 |
| COGA1_EUR | 0.0856 | 0.192 | 0.112 |
| CUINT_EUR | 0.1638 | 0.246 | 0.177 |
| ESTB2_EUR | 0.2791 | 0.049 | 0.045 |
| FINNG_EUR | 0.0145 | 0.14 | 0.058 |
| GEDIS_EUR | 0.8416 | 0.05 | 0.247 |
| GISS1_EUR | 0.5684 | 0.082 | 0.143 |
| GISS2_EUR | 0.5235 | 0.108 | 0.169 |
| IPSYC_EUR | 0.4095 | 0.032 | 0.038 |
| JANS3_EUR | 0.1003 | 0.291 | 0.177 |
| JANS4_EUR | 0.7259 | −0.08 | 0.229 |
| MVPXQ_EUR | 0.0045 | 0.074 | 0.026 |
| PGCBD_EUR | 0.0069 | 0.148 | 0.055 |
| PGCMD_EUR | 0.3937 | 0.062 | 0.072 |
| PGCPT_EUR | 0.7354 | 0.037 | 0.11 |
| PGCSZ_EUR | 0.0683 | 0.134 | 0.074 |
| PRFCT_EUR | 0.0292 | 0.185 | 0.085 |
| PSYCR_EUR | 0.7352 | 0.054 | 0.161 |
| QIMRB_EUR | 0.521 | 0.034 | 0.053 |
| UKBJC_EUR | 0.0043 | 0.113 | 0.04 |
| YPENN_EUR | 0.5045 | 0.154 | 0.231 |
| **meta** | **1.68e−09** | **0.082** | **0.014** |

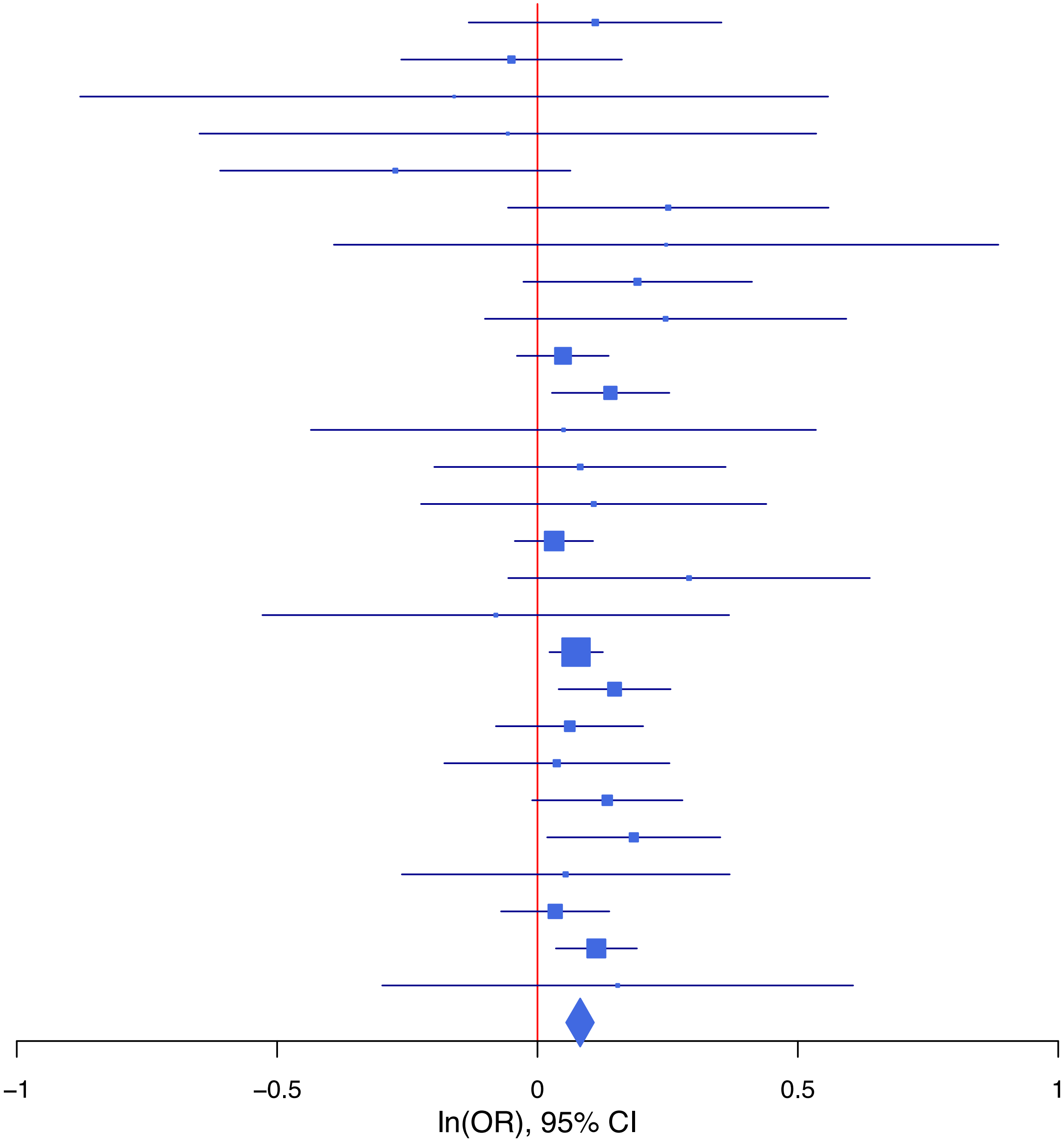

**rs62143649   A/G   2:60131553**

| Cohort | P | ln(OR) | SE |
|---|---|---|---|
| ADHEA_EUR | 0.2313 | 0.081 | 0.067 |
| ALSPC_EUR | 0.0246 | −0.121 | 0.054 |
| BEPS7_EUR | 0.7069 | 0.065 | 0.172 |
| BHRCM_EUR | 0.3371 | −0.156 | 0.163 |
| BOR17_EUR | 0.8769 | −0.013 | 0.086 |
| BOR2C_EUR | 0.84 | 0.017 | 0.085 |
| BOR2E_EUR | 0.1279 | −0.265 | 0.174 |
| COGA1_EUR | 0.033 | −0.124 | 0.058 |
| CUINT_EUR | 0.9713 | 0.004 | 0.098 |
| ESTB2_EUR | 0.1166 | −0.036 | 0.023 |
| FINNG_EUR | 0.3855 | −0.02 | 0.024 |
| GEDIS_EUR | 0.2377 | −0.157 | 0.132 |
| GISS1_EUR | 0.4804 | −0.055 | 0.078 |
| GISS2_EUR | 0.2162 | −0.11 | 0.089 |
| IPSYC_EUR | 0.005 | −0.056 | 0.02 |
| JANS3_EUR | 0.3444 | −0.098 | 0.104 |
| JANS4_EUR | 0.5229 | −0.074 | 0.116 |
| MVPXQ_EUR | 0.0719 | −0.023 | 0.013 |
| PGCBD_EUR | 0.0298 | −0.056 | 0.026 |
| PGCED_EUR | 0.2016 | −0.147 | 0.115 |
| PGCMD_EUR | 0.0973 | −0.057 | 0.034 |
| PGCPT_EUR | 0.4959 | −0.039 | 0.057 |
| PGCSZ_EUR | 0.319 | −0.037 | 0.037 |
| PRFCT_EUR | 0.646 | −0.02 | 0.043 |
| PSYCR_EUR | 0.9853 | 0.002 | 0.087 |
| QIMRB_EUR | 0.3348 | −0.026 | 0.027 |
| UKBJC_EUR | 0.0235 | −0.046 | 0.02 |
| YPENN_EUR | 0.7384 | −0.043 | 0.13 |
| **meta** | **1.18e−08** | **−0.039** | **0.007** |

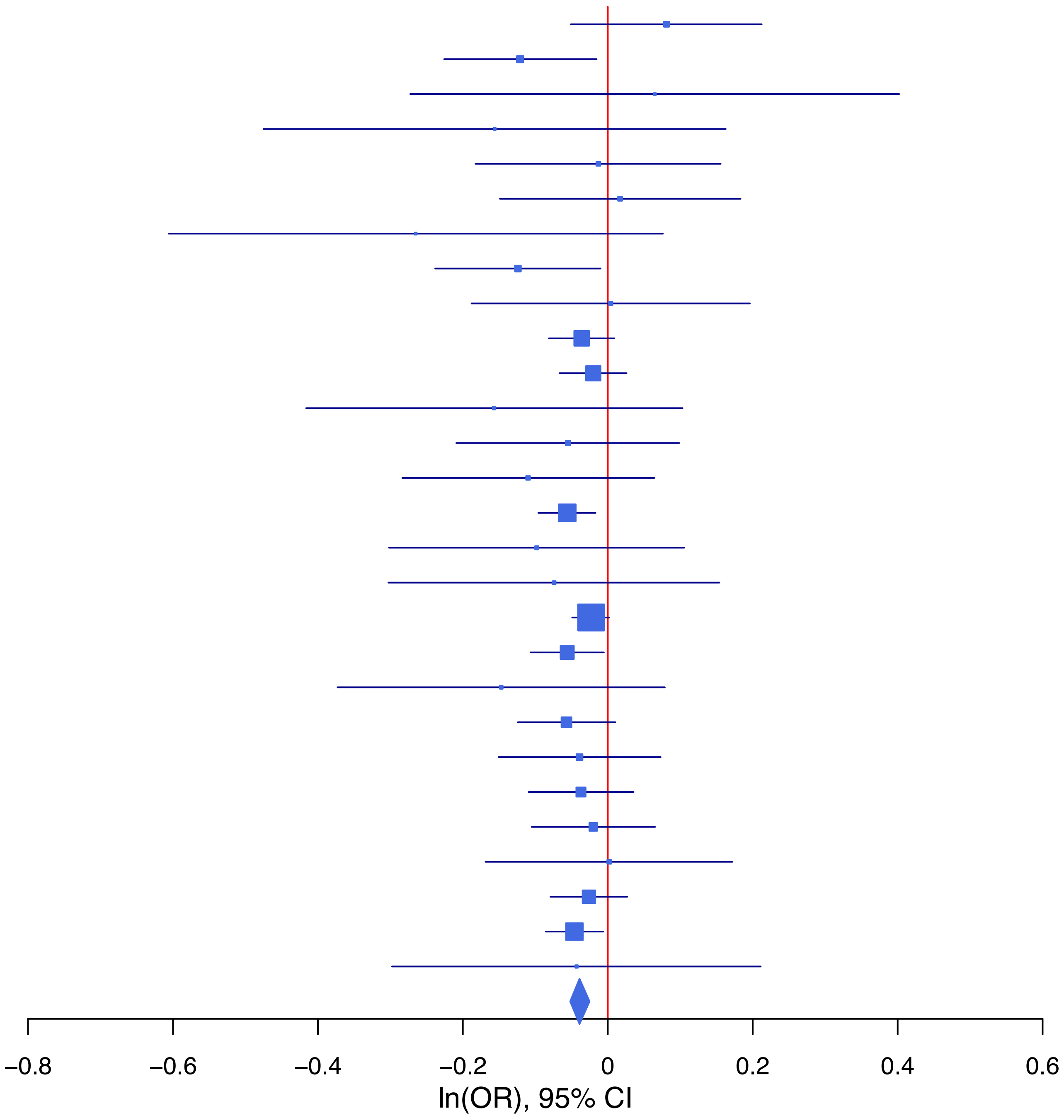

**rs62367522 C/A 5:45280212**

| Cohort | P | ln(OR) | SE |
|---|---|---|---|
| ALSPC_EUR | 0.1126 | −0.113 | 0.071 |
| BEPS7_EUR | 0.9049 | −0.027 | 0.226 |
| BOR17_EUR | 0.4751 | 0.078 | 0.11 |
| BOR2C_EUR | 0.2621 | −0.124 | 0.111 |
| BOR2E_EUR | 0.9648 | 0.01 | 0.228 |
| COGA1_EUR | 0.8662 | 0.013 | 0.075 |
| CUINT_EUR | 0.5204 | −0.082 | 0.128 |
| ESTB2_EUR | 0.0024 | −0.096 | 0.032 |
| FINNG_EUR | 0.4942 | −0.022 | 0.032 |
| GEDIS_EUR | 0.8482 | 0.033 | 0.173 |
| GISS1_EUR | 0.4336 | −0.077 | 0.098 |
| GISS2_EUR | 0.691 | 0.047 | 0.118 |
| IPSYC_EUR | 0.132 | −0.04 | 0.027 |
| JANS3_EUR | 0.8927 | −0.018 | 0.132 |
| JANS4_EUR | 0.4105 | 0.122 | 0.148 |
| MIREC_EUR | 0.8345 | 0.041 | 0.195 |
| MVPXQ_EUR | 9e−04 | −0.057 | 0.017 |
| PGCBD_EUR | 0.4479 | −0.025 | 0.034 |
| PGCED_EUR | 0.369 | 0.125 | 0.14 |
| PGCMD_EUR | 0.2249 | −0.056 | 0.046 |
| PGCPT_EUR | 0.6207 | −0.038 | 0.077 |
| PGCSZ_EUR | 0.9696 | 0.002 | 0.048 |
| PRFCT_EUR | 0.0507 | −0.117 | 0.06 |
| PSYCR_EUR | 0.8606 | 0.019 | 0.109 |
| QIMRB_EUR | 0.0191 | −0.081 | 0.035 |
| UKBJC_EUR | 0.0014 | −0.084 | 0.026 |
| YPENN_EUR | 0.5228 | −0.104 | 0.163 |
| **meta** | **2.43e−09** | **−0.053** | **0.009** |

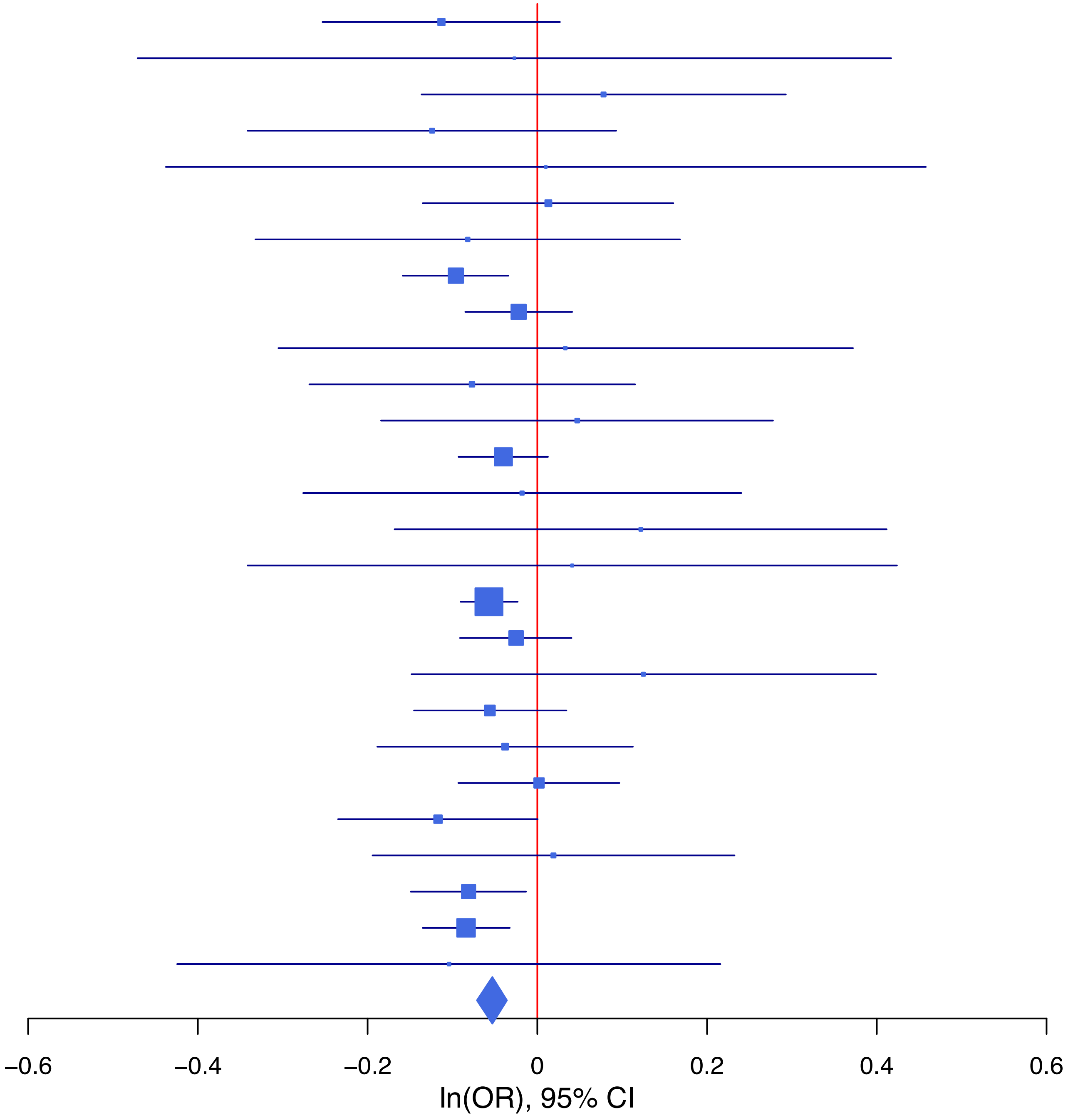

**rs62404522 C/T 6:19307114**

| Cohort | P | ln(OR) | SE |
|---|---|---|---|
| ADHEA_EUR | 0.1413 | −0.154 | 0.105 |
| ALSPC_EUR | 0.2246 | 0.093 | 0.077 |
| BEPS7_EUR | 0.7783 | 0.07 | 0.25 |
| BHRCM_EUR | 0.5224 | −0.166 | 0.26 |
| BOR17_EUR | 0.9851 | −0.002 | 0.126 |
| BOR2C_EUR | 0.0556 | 0.245 | 0.128 |
| BOR2E_EUR | 0.3309 | −0.281 | 0.29 |
| COGA1_EUR | 0.1278 | 0.133 | 0.087 |
| CUINT_EUR | 0.5468 | 0.089 | 0.147 |
| ESTB2_EUR | 0.2609 | 0.038 | 0.034 |
| FINNG_EUR | 0.0012 | 0.11 | 0.034 |
| GEDIS_EUR | 0.1439 | 0.257 | 0.176 |
| GISS1_EUR | 0.728 | 0.04 | 0.116 |
| GISS2_EUR | 0.0643 | 0.272 | 0.147 |
| IPSYC_EUR | 0.0489 | 0.055 | 0.028 |
| JANS3_EUR | 0.2348 | 0.171 | 0.144 |
| JANS4_EUR | 0.5364 | −0.116 | 0.187 |
| MVPXQ_EUR | 1.26e−05 | 0.083 | 0.019 |
| PGCBD_EUR | 1e−04 | 0.146 | 0.037 |
| PGCPT_EUR | 0.4313 | 0.063 | 0.08 |
| PGCSZ_EUR | 0.7464 | 0.018 | 0.054 |
| PRFCT_EUR | 0.0094 | 0.158 | 0.061 |
| PSYCR_EUR | 0.081 | −0.227 | 0.13 |
| QIMRB_EUR | 0.0417 | 0.081 | 0.04 |
| UKBJC_EUR | 0.4223 | 0.024 | 0.03 |
| YPENN_EUR | 0.8099 | 0.045 | 0.188 |
| **meta** | **4.25e−13** | **0.072** | **0.01** |

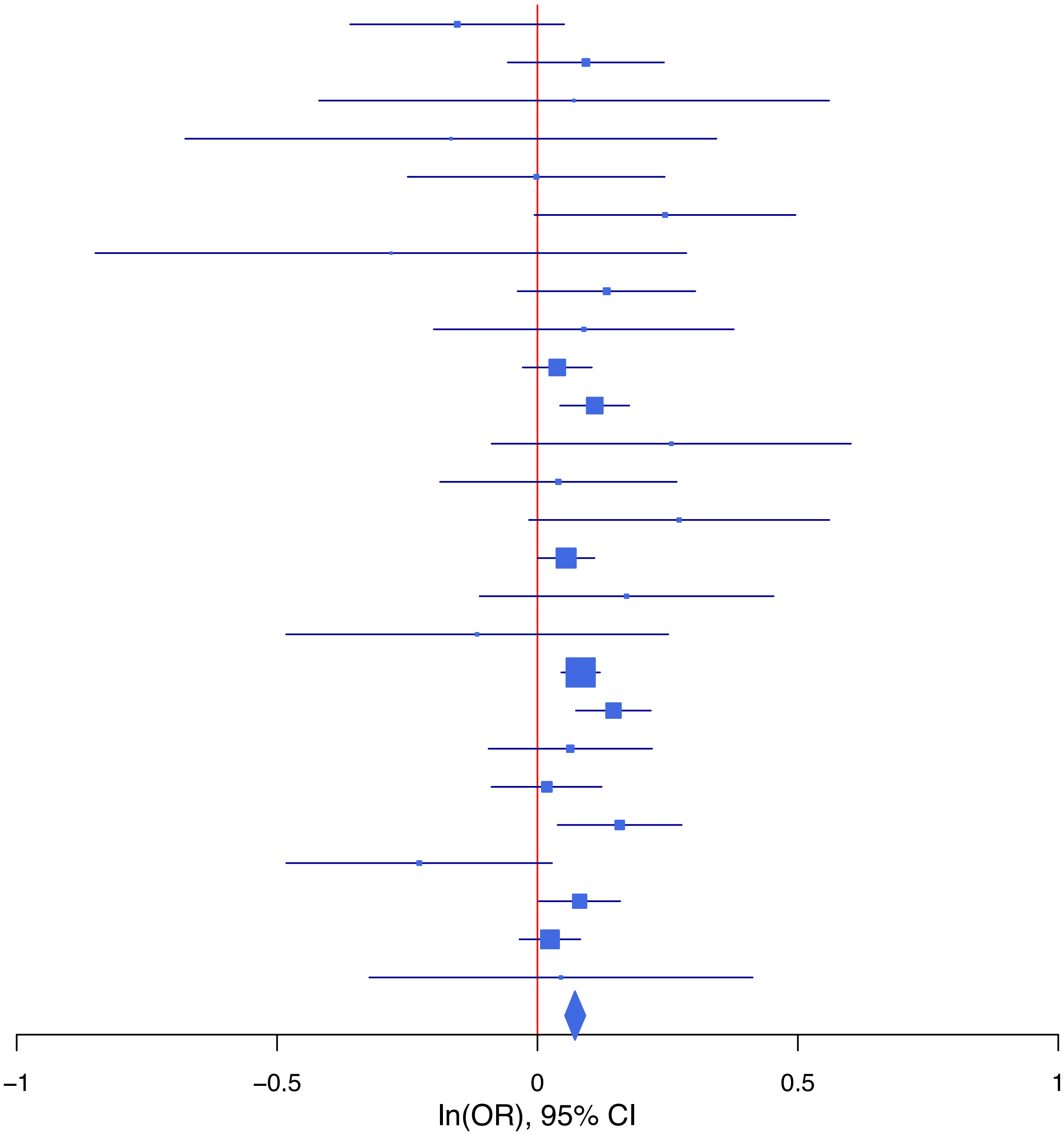

**rs640704 G/A 15:59049021**

| Cohort | P | ln(OR) | SE |
|---|---|---|---|
| ALSPC_EUR | 0.1963 | −0.077 | 0.06 |
| BEPS7_EUR | 0.8101 | 0.046 | 0.192 |
| BHRCM_EUR | 0.9895 | 0.002 | 0.185 |
| BOR17_EUR | 0.5758 | −0.053 | 0.094 |
| BOR2C_EUR | 0.8381 | 0.019 | 0.095 |
| BOR2E_EUR | 0.8457 | −0.041 | 0.211 |
| CUINT_EUR | 0.5594 | −0.065 | 0.111 |
| ESTB2_EUR | 0.0142 | −0.06 | 0.025 |
| GEDIS_EUR | 0.6767 | 0.059 | 0.141 |
| GISS1_EUR | 0.8981 | −0.011 | 0.085 |
| GISS2_EUR | 0.2417 | −0.118 | 0.101 |
| IPSYC_EUR | 0.0079 | −0.056 | 0.021 |
| JANS3_EUR | 0.8377 | −0.024 | 0.116 |
| JANS4_EUR | 0.4256 | 0.11 | 0.138 |
| MVPXQ_EUR | 0.0076 | −0.038 | 0.014 |
| PGCBD_EUR | 0.0498 | −0.055 | 0.028 |
| PGCED_EUR | 0.526 | 0.079 | 0.125 |
| PGCMD_EUR | 0.9331 | −0.003 | 0.037 |
| PGCPT_EUR | 0.9585 | 0.003 | 0.063 |
| PGCSZ_EUR | 0.0048 | −0.115 | 0.041 |
| PRFCT_EUR | 0.4335 | 0.036 | 0.047 |
| PSYCR_EUR | 0.6788 | 0.04 | 0.096 |
| QIMRB_EUR | 0.0729 | −0.053 | 0.03 |
| UKBJC_EUR | 0.06 | −0.041 | 0.022 |
| YPENN_EUR | 0.8707 | 0.023 | 0.14 |
| **meta** | **4.19e−08** | **−0.042** | **0.008** |

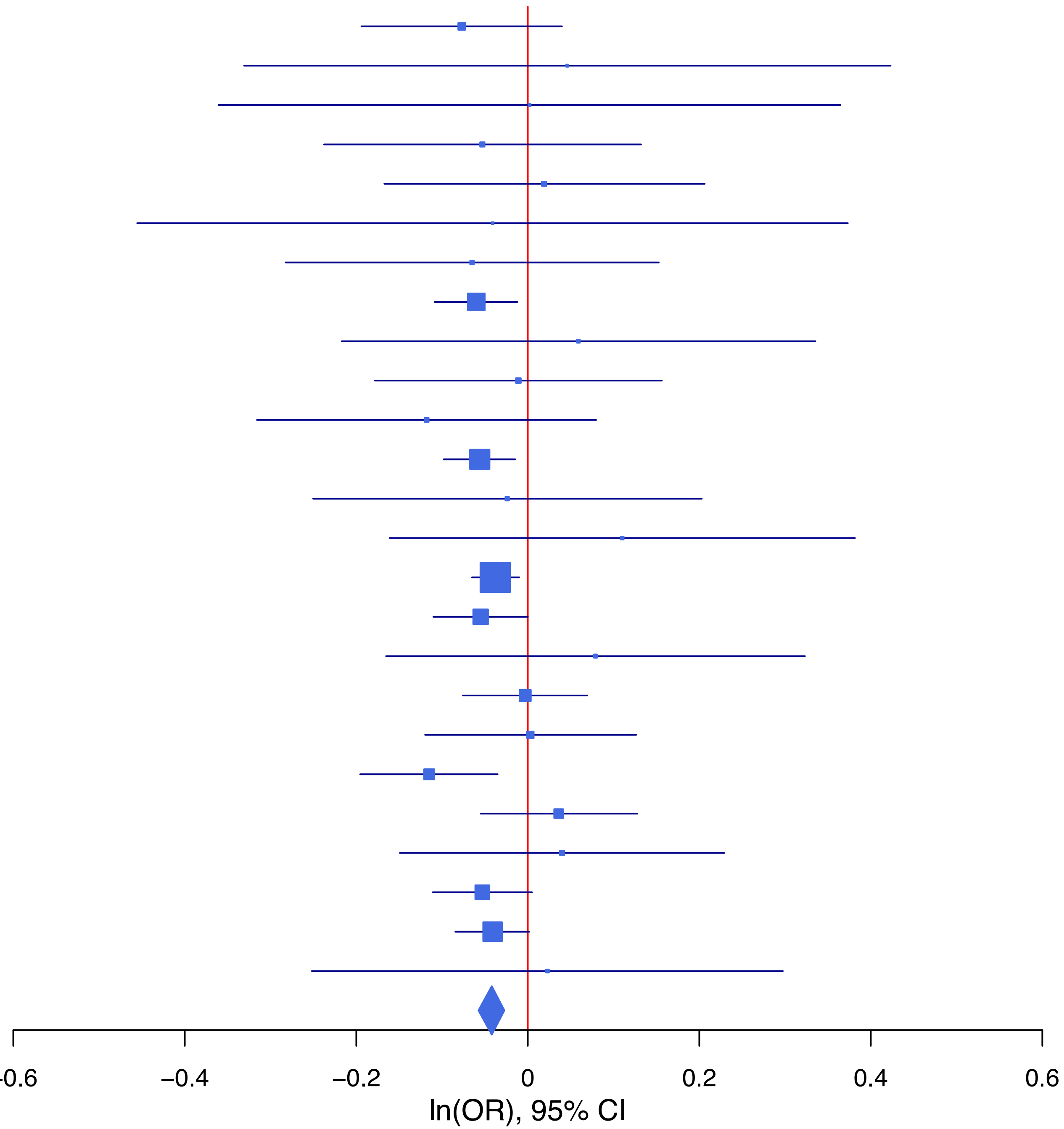

**rs6539788 T/G 12:84226327**

| Cohort | P | ln(OR) | SE |
|---|---|---|---|
| ADHEA_EUR | 0.2718 | −0.075 | 0.068 |
| ALSPC_EUR | 0.0531 | −0.106 | 0.055 |
| BEPS7_EUR | 0.3793 | −0.155 | 0.176 |
| BHRCM_EUR | 0.3214 | 0.15 | 0.152 |
| BOR17_EUR | 0.0508 | −0.17 | 0.087 |
| BOR2C_EUR | 0.3894 | −0.074 | 0.086 |
| BOR2E_EUR | 0.9028 | −0.022 | 0.184 |
| COGA1_EUR | 0.7323 | −0.02 | 0.059 |
| CUINT_EUR | 0.416 | −0.081 | 0.099 |
| ESTB2_EUR | 0.6158 | −0.012 | 0.023 |
| FINNG_EUR | 0.0239 | −0.051 | 0.023 |
| GEDIS_EUR | 0.9992 | 0 | 0.131 |
| GISS1_EUR | 0.7554 | 0.024 | 0.078 |
| GISS2_EUR | 0.049 | −0.176 | 0.09 |
| IPSYC_EUR | 0.8084 | −0.005 | 0.02 |
| JANS3_EUR | 0.1733 | 0.144 | 0.106 |
| JANS4_EUR | 0.9686 | −0.005 | 0.12 |
| MVPXQ_EUR | 8e−04 | −0.043 | 0.013 |
| PGCBD_EUR | 0.0035 | −0.075 | 0.026 |
| PGCED_EUR | 0.6692 | 0.05 | 0.117 |
| PGCMD_EUR | 0.237 | −0.041 | 0.034 |
| PGCPT_EUR | 0.9824 | 0.001 | 0.058 |
| PGCSZ_EUR | 0.9065 | 0.004 | 0.037 |
| PRFCT_EUR | 0.2457 | −0.051 | 0.044 |
| PSYCR_EUR | 0.5712 | −0.049 | 0.087 |
| QIMRB_EUR | 0.2897 | −0.029 | 0.027 |
| UKBJC_EUR | 0.012 | −0.051 | 0.02 |
| YPENN_EUR | 0.088 | −0.225 | 0.132 |
| **meta** | **1.33e−08** | **−0.038** | **0.007** |

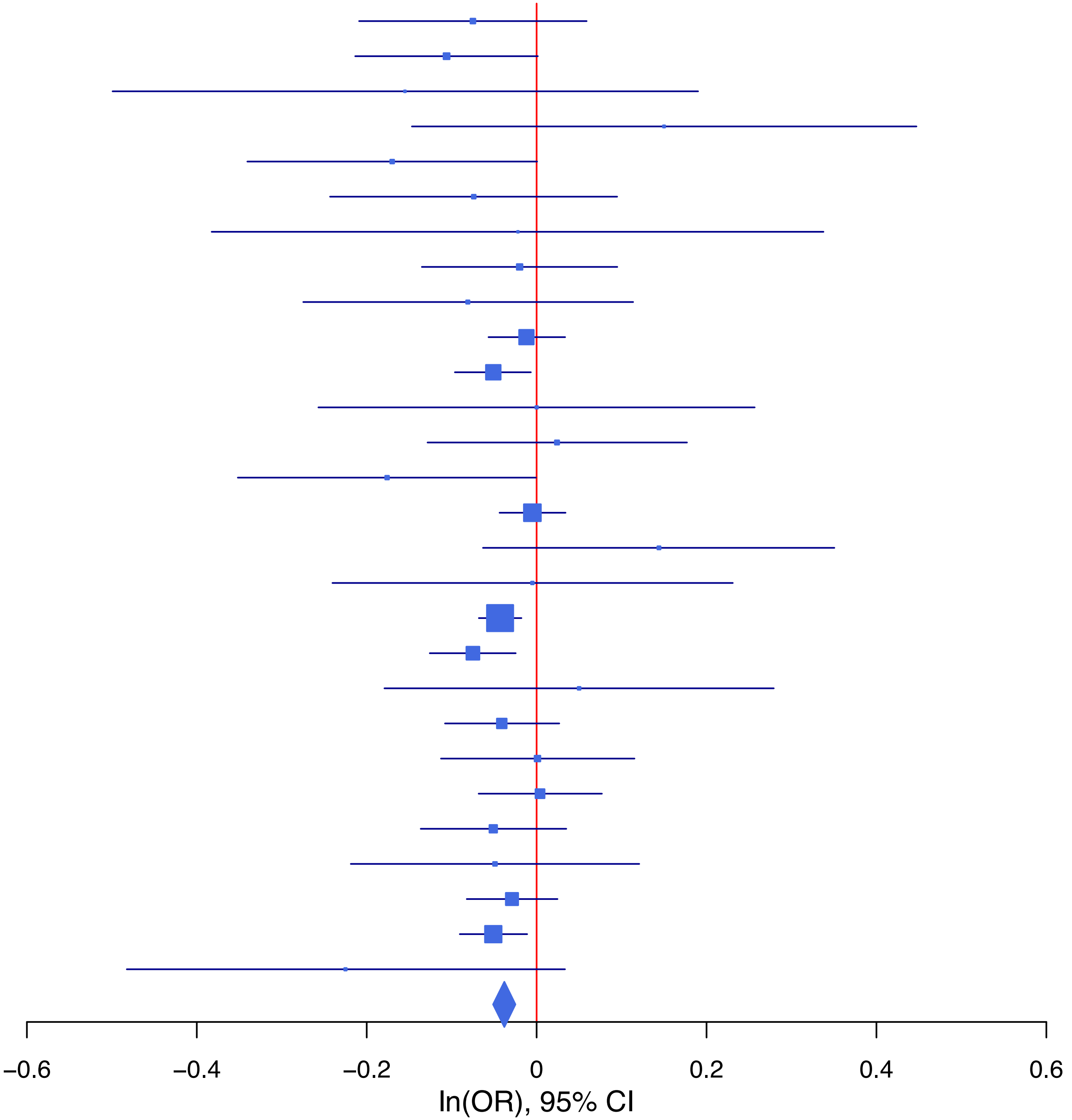

**rs6589377 G/A 11:113355736**

| Cohort | P | ln(OR) | SE |
|---|---|---|---|
| ADHEA_EUR | 0.2158 | −0.088 | 0.071 |
| ALSPC_EUR | 0.6826 | −0.023 | 0.055 |
| BEPS7_EUR | 0.8831 | 0.028 | 0.193 |
| BHRCM_EUR | 0.8435 | −0.032 | 0.163 |
| BOR17_EUR | 0.6963 | 0.034 | 0.086 |
| BOR2C_EUR | 0.3629 | −0.081 | 0.09 |
| BOR2E_EUR | 0.4234 | 0.154 | 0.193 |
| COGA1_EUR | 0.4617 | −0.045 | 0.061 |
| CUINT_EUR | 0.8864 | −0.015 | 0.103 |
| ESTB2_EUR | 0.0078 | −0.069 | 0.026 |
| FINNG_EUR | 0.0399 | −0.058 | 0.028 |
| GEDIS_EUR | 0.13 | −0.21 | 0.139 |
| GISS1_EUR | 0.8354 | −0.017 | 0.083 |
| GISS2_EUR | 0.9638 | 0.004 | 0.094 |
| IPSYC_EUR | 0.0015 | −0.066 | 0.021 |
| JANS3_EUR | 0.0964 | −0.18 | 0.108 |
| JANS4_EUR | 0.1245 | 0.183 | 0.119 |
| MIREC_EUR | 0.0158 | −0.406 | 0.168 |
| MVPXQ_EUR | 6.04e−07 | −0.067 | 0.013 |
| PGCBD_EUR | 0.1923 | −0.035 | 0.027 |
| PGCED_EUR | 0.3179 | −0.116 | 0.116 |
| PGCPT_EUR | 0.9283 | −0.005 | 0.059 |
| PGCSZ_EUR | 0.0784 | −0.068 | 0.038 |
| PRFCT_EUR | 0.7484 | −0.015 | 0.047 |
| PSYCR_EUR | 0.0211 | 0.204 | 0.088 |
| QIMRB_EUR | 0.1029 | −0.046 | 0.028 |
| UKBJC_EUR | 0.2071 | −0.026 | 0.021 |
| YPENN_EUR | 0.9107 | −0.015 | 0.132 |
| **meta** | **1.51e−12** | **−0.051** | **0.007** |

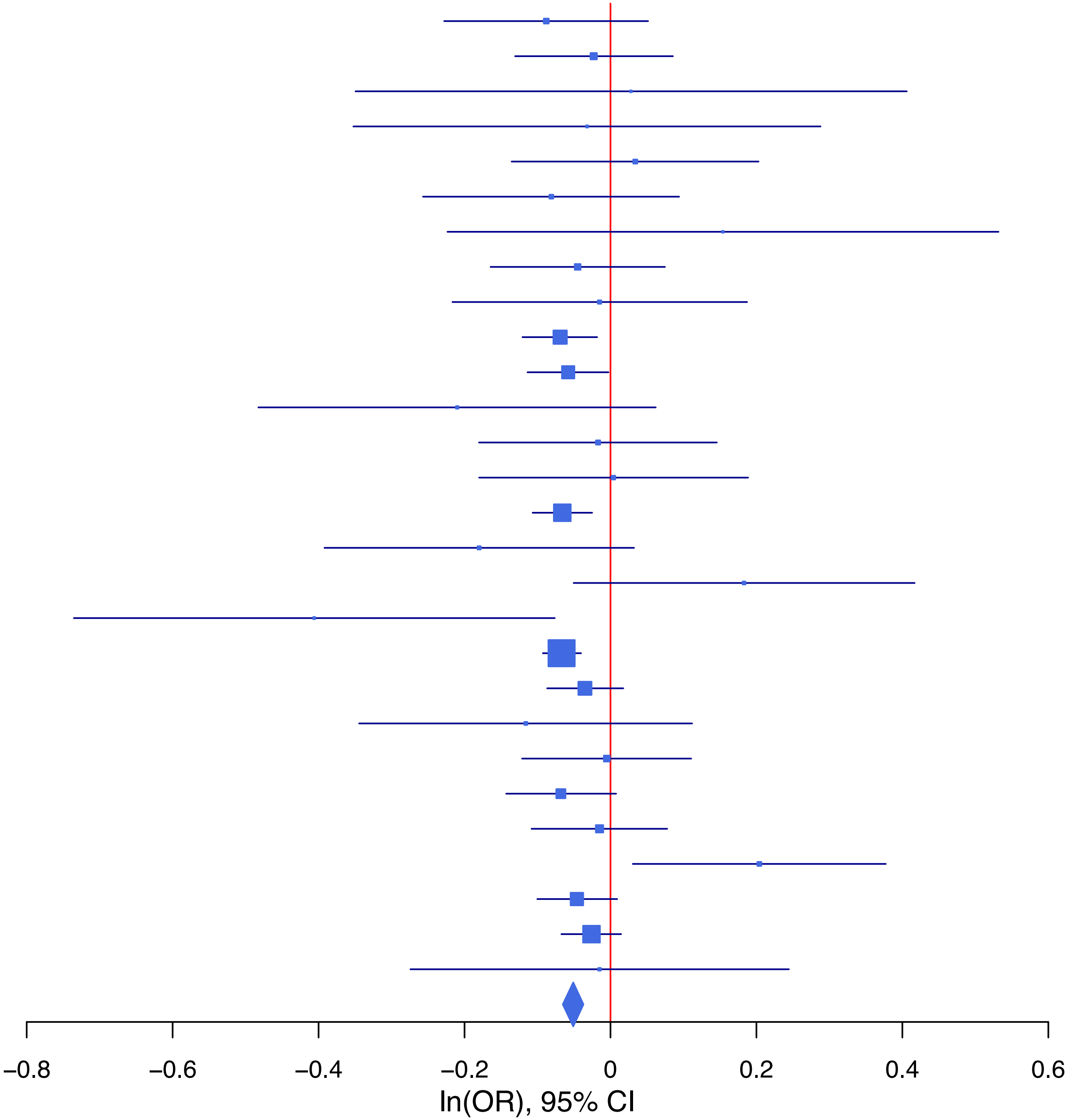

**rs687654 T/C 9:127833905**

| Cohort | P | ln(OR) | SE |
|---|---|---|---|
| ADHEA_EUR | 0.7646 | −0.022 | 0.072 |
| ALSPC_EUR | 0.0063 | −0.161 | 0.059 |
| BEPS7_EUR | 0.1771 | −0.267 | 0.198 |
| BHRCM_EUR | 0.5874 | 0.084 | 0.155 |
| BOR17_EUR | 0.5953 | −0.049 | 0.091 |
| BOR2C_EUR | 0.2119 | −0.118 | 0.094 |
| BOR2E_EUR | 0.9066 | −0.022 | 0.188 |
| COGA1_EUR | 0.023 | −0.144 | 0.063 |
| CUINT_EUR | 0.912 | −0.012 | 0.105 |
| ESTB2_EUR | 0.2078 | −0.033 | 0.026 |
| FINNG_EUR | 0.4888 | −0.02 | 0.029 |
| GEDIS_EUR | 0.8042 | −0.035 | 0.141 |
| GISS1_EUR | 0.581 | −0.047 | 0.085 |
| GISS2_EUR | 0.963 | −0.005 | 0.099 |
| IPSYC_EUR | 0.2679 | −0.024 | 0.021 |
| JANS3_EUR | 0.1476 | −0.169 | 0.117 |
| JANS4_EUR | 0.7399 | −0.041 | 0.122 |
| MIREC_EUR | 0.0221 | 0.369 | 0.161 |
| MVPXQ_EUR | 5.54e−06 | −0.062 | 0.014 |
| PGCBD_EUR | 0.1417 | −0.041 | 0.028 |
| PGCED_EUR | 0.793 | 0.032 | 0.123 |
| PGCMD_EUR | 0.2178 | −0.045 | 0.037 |
| PGCPT_EUR | 0.9065 | 0.007 | 0.061 |
| PGCSZ_EUR | 0.9877 | 0.001 | 0.04 |
| PRFCT_EUR | 0.9446 | 0.003 | 0.048 |
| PSYCR_EUR | 0.3911 | −0.08 | 0.093 |
| QIMRB_EUR | 0.3146 | −0.029 | 0.029 |
| YPENN_EUR | 0.8586 | −0.024 | 0.136 |
| **meta** | **4.79e−08** | **−0.042** | **0.008** |

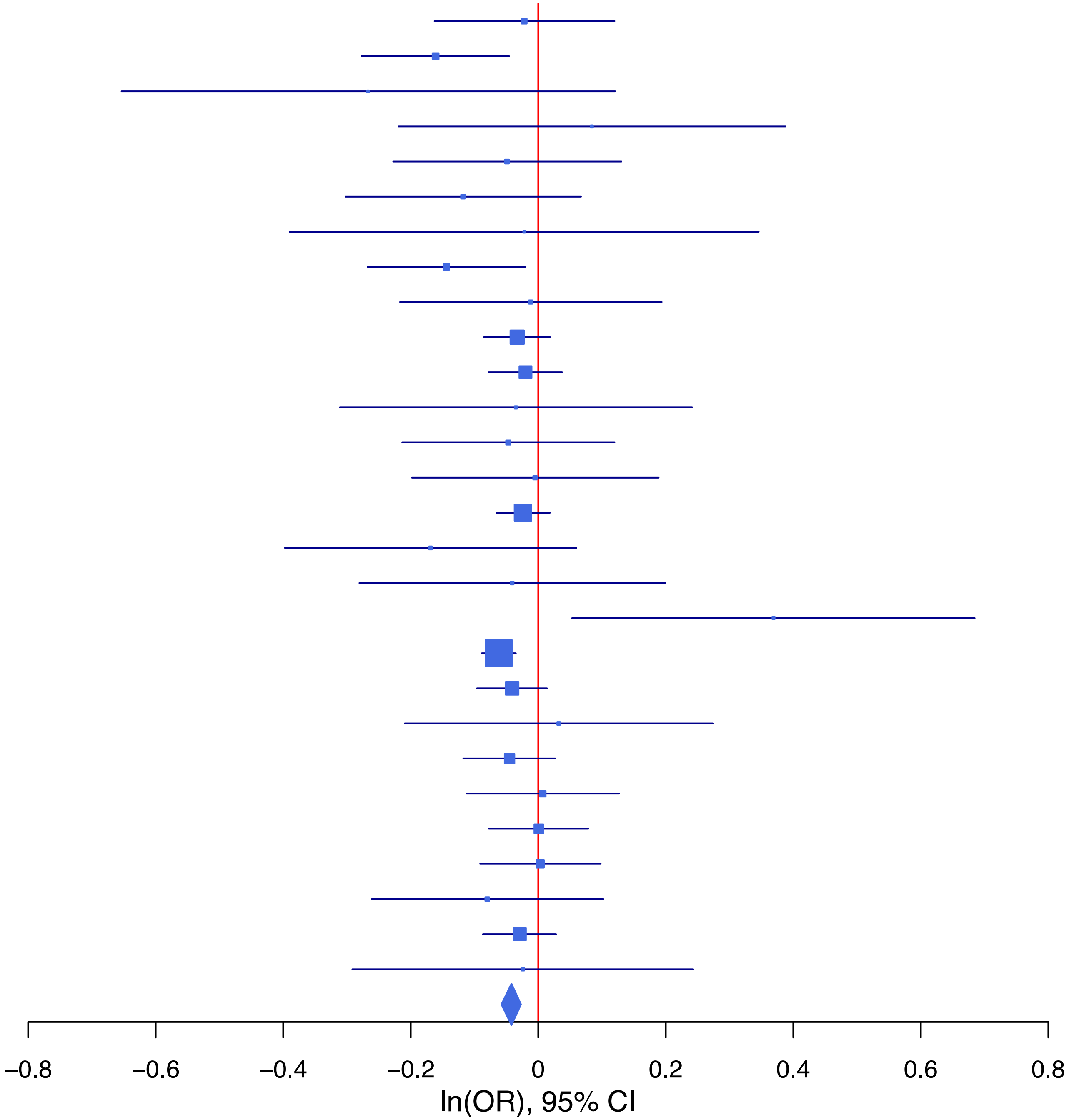

**rs6959688 G/A 7:1966831**

| Cohort | P | ln(OR) | SE |
|---|---|---|---|
| ADHEA_EUR | 0.3461 | 0.065 | 0.069 |
| ALSPC_EUR | 0.3065 | 0.057 | 0.056 |
| BEPS7_EUR | 0.0642 | 0.339 | 0.183 |
| BHRCM_EUR | 0.9133 | −0.018 | 0.166 |
| BOR17_EUR | 0.5783 | −0.048 | 0.087 |
| BOR2C_EUR | 0.0112 | 0.219 | 0.086 |
| BOR2E_EUR | 0.1128 | −0.298 | 0.188 |
| COGA1_EUR | 0.1911 | 0.079 | 0.06 |
| ESTB2_EUR | 0.9481 | 0.002 | 0.023 |
| FINNG_EUR | 0.0575 | 0.045 | 0.023 |
| GEDIS_EUR | 0.0838 | 0.221 | 0.128 |
| GISS1_EUR | 0.9665 | 0.004 | 0.084 |
| GISS2_EUR | 0.8088 | 0.023 | 0.095 |
| IPSYC_EUR | 0.0017 | 0.063 | 0.02 |
| JANS3_EUR | 0.2813 | 0.114 | 0.106 |
| JANS4_EUR | 0.1698 | 0.162 | 0.118 |
| MVPXQ_EUR | 0.0037 | 0.039 | 0.013 |
| PGCBD_EUR | 0.2113 | 0.033 | 0.026 |
| PGCED_EUR | 0.4801 | −0.085 | 0.12 |
| PGCMD_EUR | 0.1072 | −0.057 | 0.035 |
| PGCPT_EUR | 0.332 | 0.056 | 0.058 |
| PGCSZ_EUR | 0.6581 | 0.017 | 0.038 |
| PRFCT_EUR | 0.9823 | 0.001 | 0.045 |
| PSYCR_EUR | 0.3533 | 0.079 | 0.085 |
| QIMRB_EUR | 0.0542 | 0.053 | 0.027 |
| UKBJC_EUR | 0.0067 | 0.056 | 0.021 |
| YPENN_EUR | 0.4516 | 0.096 | 0.128 |
| **meta** | **1.79e−08** | **0.039** | **0.007** |

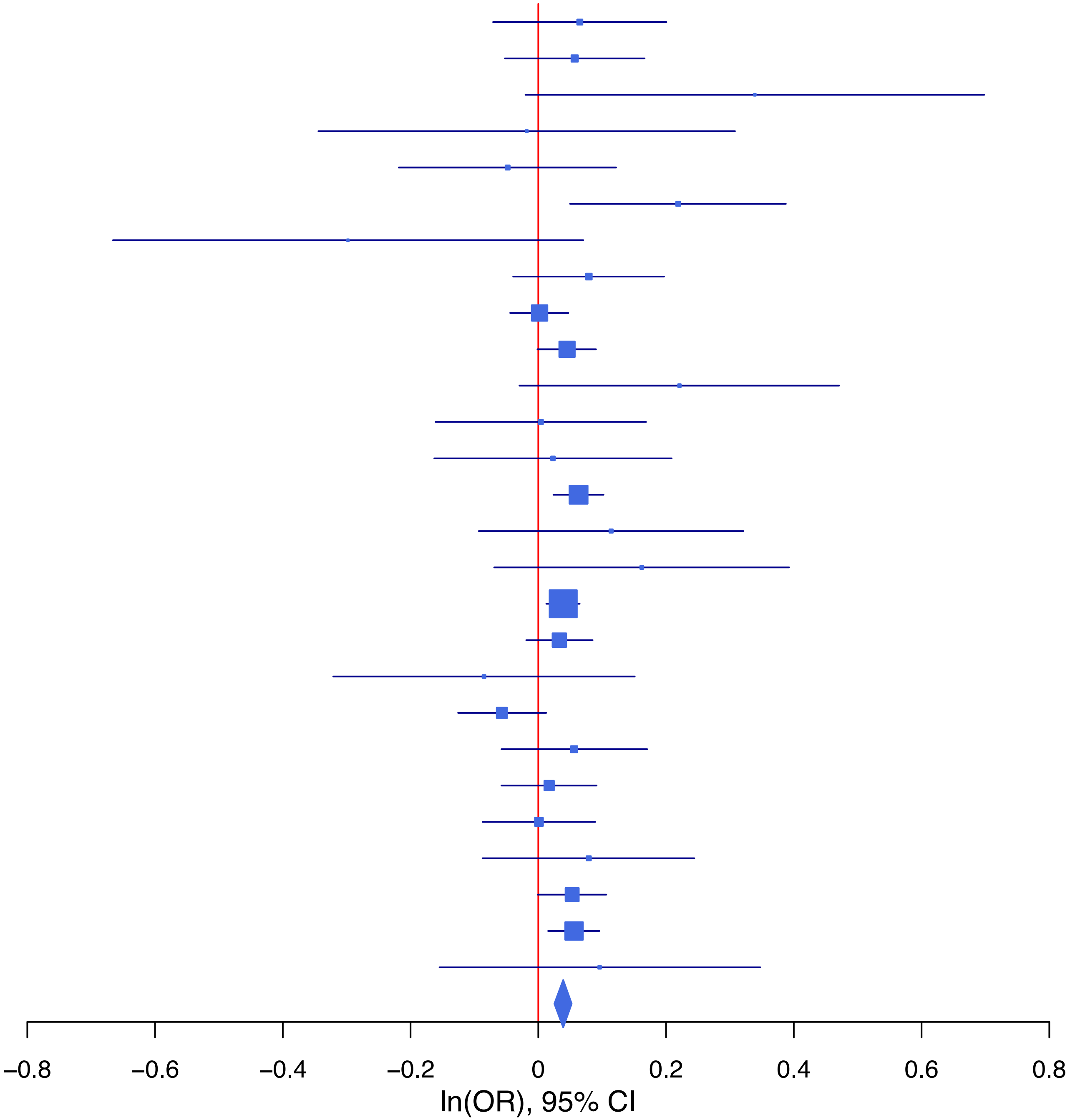

**rs7174904 T/C 15:47676110**

| Cohort | P | ln(OR) | SE |
|---|---|---|---|
| ADHEA_EUR | 0.4549 | 0.06 | 0.08 |
| ALSPC_EUR | 0.5444 | 0.039 | 0.065 |
| BEPS7_EUR | 0.762 | 0.065 | 0.215 |
| BHRCM_EUR | 0.5326 | −0.115 | 0.184 |
| BOR17_EUR | 0.2716 | 0.107 | 0.098 |
| BOR2C_EUR | 0.815 | −0.024 | 0.101 |
| BOR2E_EUR | 0.2723 | 0.217 | 0.197 |
| COGA1_EUR | 0.4087 | 0.058 | 0.07 |
| ESTB2_EUR | 0.033 | 0.063 | 0.029 |
| GISS1_EUR | 0.7613 | −0.031 | 0.101 |
| GISS2_EUR | 0.0084 | 0.311 | 0.118 |
| IPSYC_EUR | 0.0153 | 0.057 | 0.023 |
| JANS3_EUR | 0.7426 | 0.04 | 0.123 |
| JANS4_EUR | 0.5265 | −0.094 | 0.149 |
| MIREC_EUR | 0.014 | 0.416 | 0.169 |
| MVPXQ_EUR | 0.0682 | 0.028 | 0.015 |
| PGCBD_EUR | 0.0144 | 0.075 | 0.031 |
| PGCED_EUR | 0.1337 | 0.195 | 0.13 |
| PGCMD_EUR | 0.0327 | 0.086 | 0.04 |
| PGCPT_EUR | 0.1523 | −0.1 | 0.07 |
| PGCSZ_EUR | 7e−04 | 0.148 | 0.043 |
| PRFCT_EUR | 0.1093 | −0.084 | 0.053 |
| PSYCR_EUR | 0.8528 | 0.019 | 0.105 |
| QIMRB_EUR | 4e−04 | 0.117 | 0.033 |
| UKBJC_EUR | 0.0012 | 0.079 | 0.024 |
| YPENN_EUR | 0.8397 | −0.032 | 0.157 |
| **meta** | **4.26e−11** | **0.056** | **0.008** |

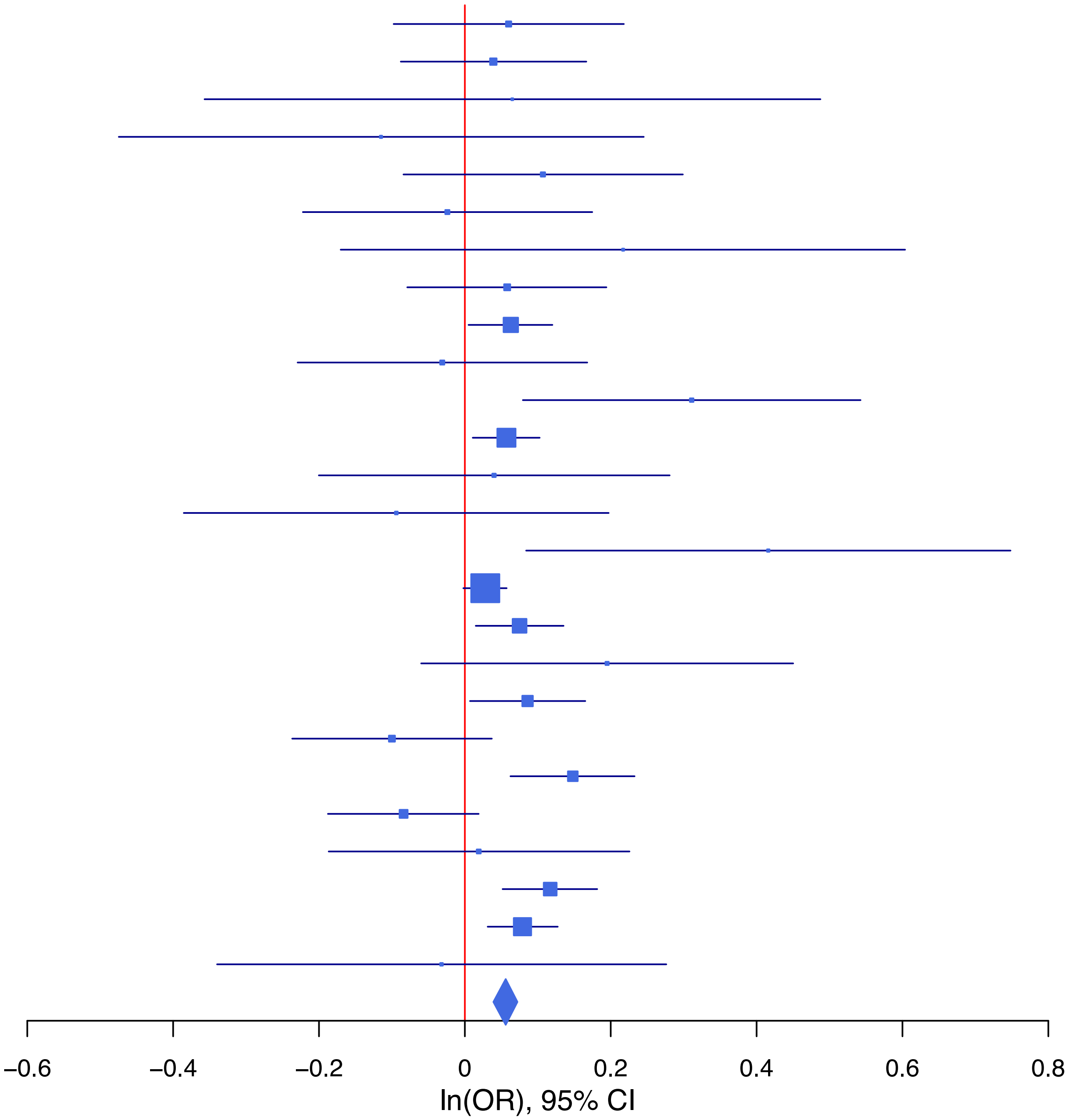

## rs7577690 A/G 2:220045035

| Cohort | P | ln(OR) | SE |
|---|---|---|---|
| ALSPC_EUR | 0.057 | 0.107 | 0.056 |
| BEPS7_EUR | 0.2678 | 0.208 | 0.187 |
| BHRCM_EUR | 0.6723 | 0.07 | 0.167 |
| BOR17_EUR | 0.3964 | 0.076 | 0.09 |
| BOR2C_EUR | 0.6068 | 0.049 | 0.096 |
| BOR2E_EUR | 0.5246 | 0.115 | 0.18 |
| COGA1_EUR | 0.6144 | 0.031 | 0.062 |
| CUINT_EUR | 0.5701 | 0.062 | 0.109 |
| ESTB2_EUR | 0.0642 | 0.045 | 0.024 |
| FINNG_EUR | 0.098 | 0.041 | 0.025 |
| GEDIS_EUR | 0.6098 | −0.07 | 0.136 |
| GISS1_EUR | 0.3801 | −0.073 | 0.084 |
| GISS2_EUR | 0.8447 | −0.02 | 0.103 |
| IPSYC_EUR | 0.0043 | 0.062 | 0.022 |
| JANS3_EUR | 0.1257 | 0.172 | 0.112 |
| JANS4_EUR | 0.1331 | −0.189 | 0.126 |
| MIREC_EUR | 0.2949 | −0.173 | 0.165 |
| MVPXQ_EUR | 0.007 | 0.037 | 0.014 |
| PGCBD_EUR | 0.043 | 0.055 | 0.027 |
| PGCMD_EUR | 0.3299 | 0.036 | 0.037 |
| PGCPT_EUR | 0.6463 | 0.028 | 0.061 |
| PGCSZ_EUR | 0.2976 | 0.041 | 0.04 |
| PRFCT_EUR | 0.8342 | 0.01 | 0.048 |
| PSYCR_EUR | 0.788 | −0.025 | 0.093 |
| QIMRB_EUR | 0.2036 | 0.038 | 0.03 |
| UKBJC_EUR | 0.2301 | 0.026 | 0.022 |
| YPENN_EUR | 0.8283 | 0.029 | 0.134 |
| **meta** | **3.81e−08** | **0.04** | **0.007** |

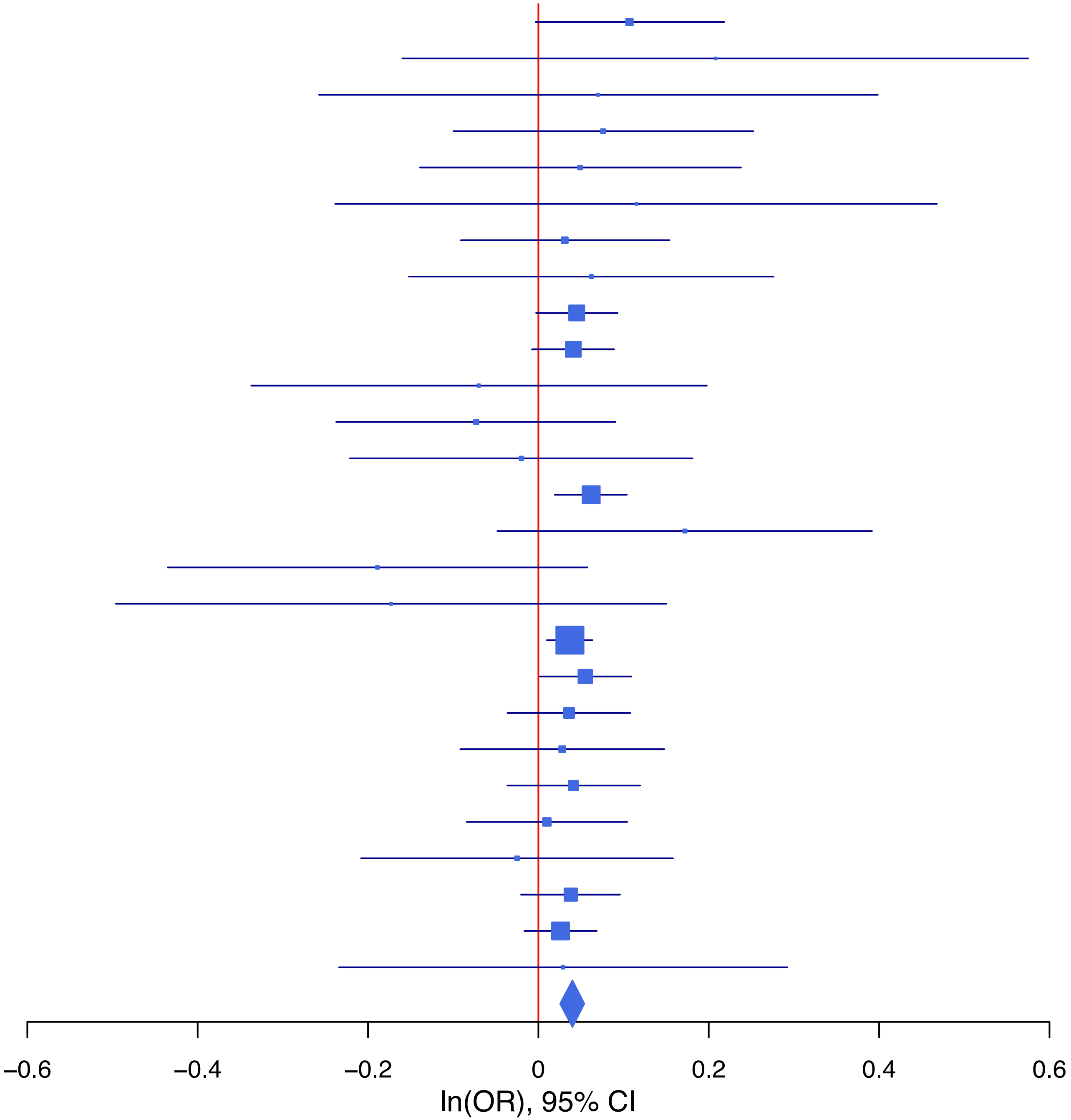

**rs77641763 C/T 9:140265782**

| Cohort | P | ln(OR) | SE |
|---|---|---|---|
| BEPS7_EUR | 0.5778 | 0.151 | 0.272 |
| BHRCM_EUR | 0.3104 | 0.365 | 0.36 |
| BOR17_EUR | 0.0923 | −0.21 | 0.125 |
| BOR2C_EUR | 0.303 | −0.138 | 0.134 |
| BOR2E_EUR | 0.3094 | −0.299 | 0.294 |
| COGA1_EUR | 0.2574 | −0.103 | 0.091 |
| ESTB2_EUR | 3e−04 | −0.119 | 0.033 |
| FINNG_EUR | 0.2663 | −0.033 | 0.03 |
| GISS2_EUR | 0.6224 | −0.077 | 0.157 |
| JANS3_EUR | 0.1574 | 0.333 | 0.236 |
| JANS4_EUR | 0.745 | 0.077 | 0.238 |
| MVPXQ_EUR | 0.0145 | −0.055 | 0.022 |
| PGCBD_EUR | 0.9596 | 0.003 | 0.059 |
| PGCMD_EUR | 0.3383 | −0.074 | 0.077 |
| PGCPT_EUR | 0.3959 | −0.07 | 0.083 |
| PGCSZ_EUR | 0.6073 | 0.047 | 0.09 |
| PRFCT_EUR | 0.9185 | 0.006 | 0.061 |
| PSYCR_EUR | 0.3353 | −0.125 | 0.129 |
| QIMRB_EUR | 0.0297 | −0.092 | 0.042 |
| UKBJC_EUR | 1e−04 | −0.115 | 0.03 |
| YPENN_EUR | 0.5377 | 0.133 | 0.216 |
| **meta** | **1.17e−08** | **−0.067** | **0.012** |

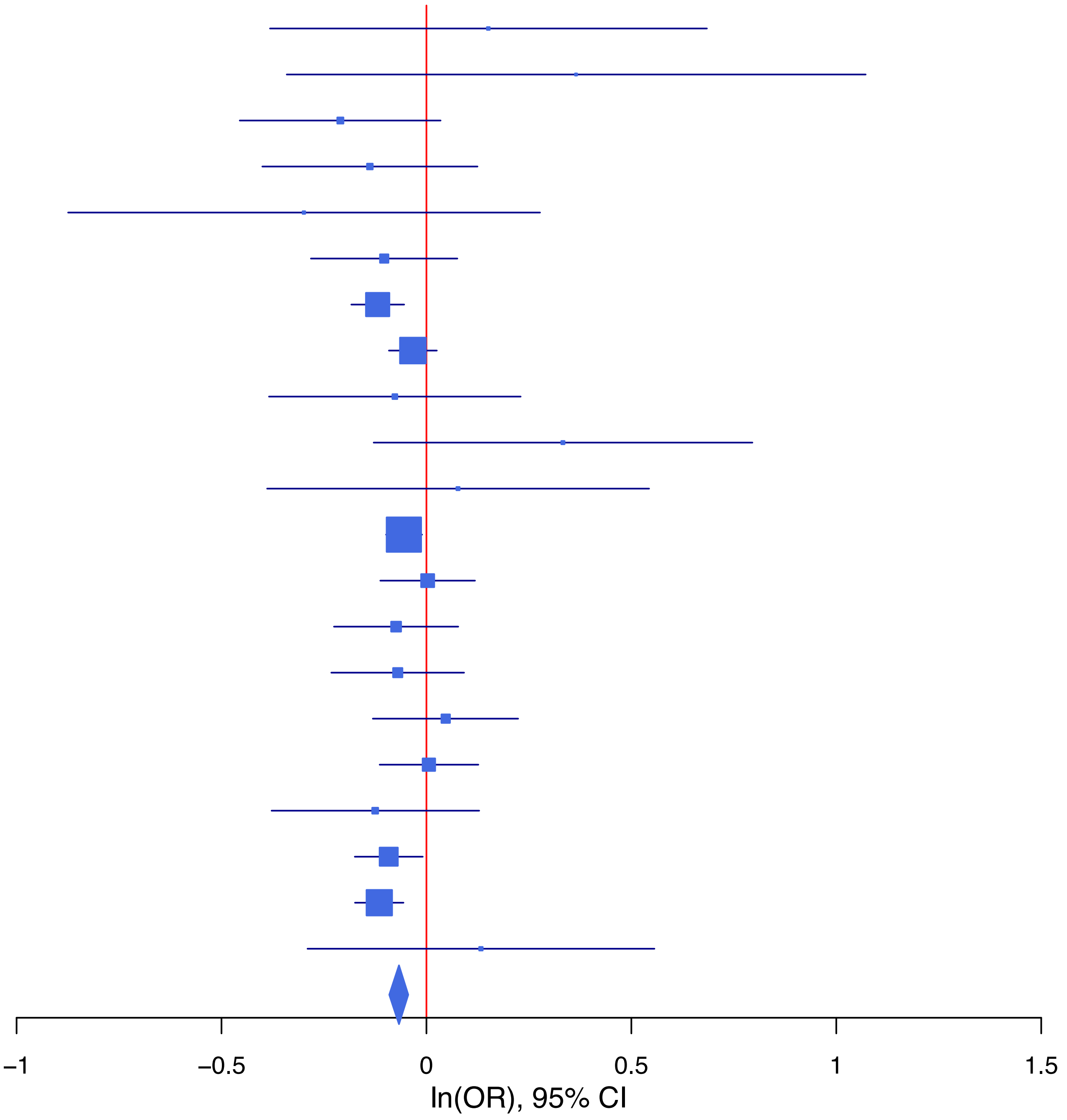

**rs7809993 C/G 7:117513664**

| Cohort | P | ln(OR) | SE |
|---|---|---|---|
| ADHEA_EUR | 0.414 | −0.056 | 0.068 |
| ALSPC_EUR | 0.4464 | 0.041 | 0.054 |
| BEPS7_EUR | 0.842 | −0.037 | 0.186 |
| BHRCM_EUR | 0.6107 | 0.079 | 0.154 |
| BOR17_EUR | 0.3899 | −0.075 | 0.087 |
| BOR2C_EUR | 0.1544 | −0.123 | 0.086 |
| BOR2E_EUR | 0.1065 | −0.289 | 0.179 |
| COGA1_EUR | 0.2936 | −0.063 | 0.06 |
| CUINT_EUR | 0.4099 | −0.083 | 0.101 |
| ESTB2_EUR | 1e−04 | −0.095 | 0.024 |
| GEDIS_EUR | 0.8481 | 0.026 | 0.132 |
| GISS2_EUR | 0.2371 | −0.111 | 0.094 |
| IPSYC_EUR | 0.001 | −0.067 | 0.02 |
| JANS3_EUR | 0.6726 | 0.044 | 0.105 |
| JANS4_EUR | 0.2607 | −0.133 | 0.118 |
| MVPXQ_EUR | 0.0165 | −0.032 | 0.013 |
| PGCBD_EUR | 0.2744 | 0.028 | 0.026 |
| PGCED_EUR | 0.6189 | 0.056 | 0.113 |
| PGCMD_EUR | 0.9436 | −0.002 | 0.034 |
| PGCPT_EUR | 0.7444 | 0.019 | 0.058 |
| PRFCT_EUR | 0.0133 | −0.112 | 0.045 |
| PSYCR_EUR | 0.9922 | −0.001 | 0.087 |
| QIMRB_EUR | 0.3676 | −0.025 | 0.027 |
| UKBJC_EUR | 0.0118 | −0.051 | 0.02 |
| YPENN_EUR | 0.0954 | −0.216 | 0.129 |
| **meta** | **3.08e−08** | **−0.04** | **0.007** |

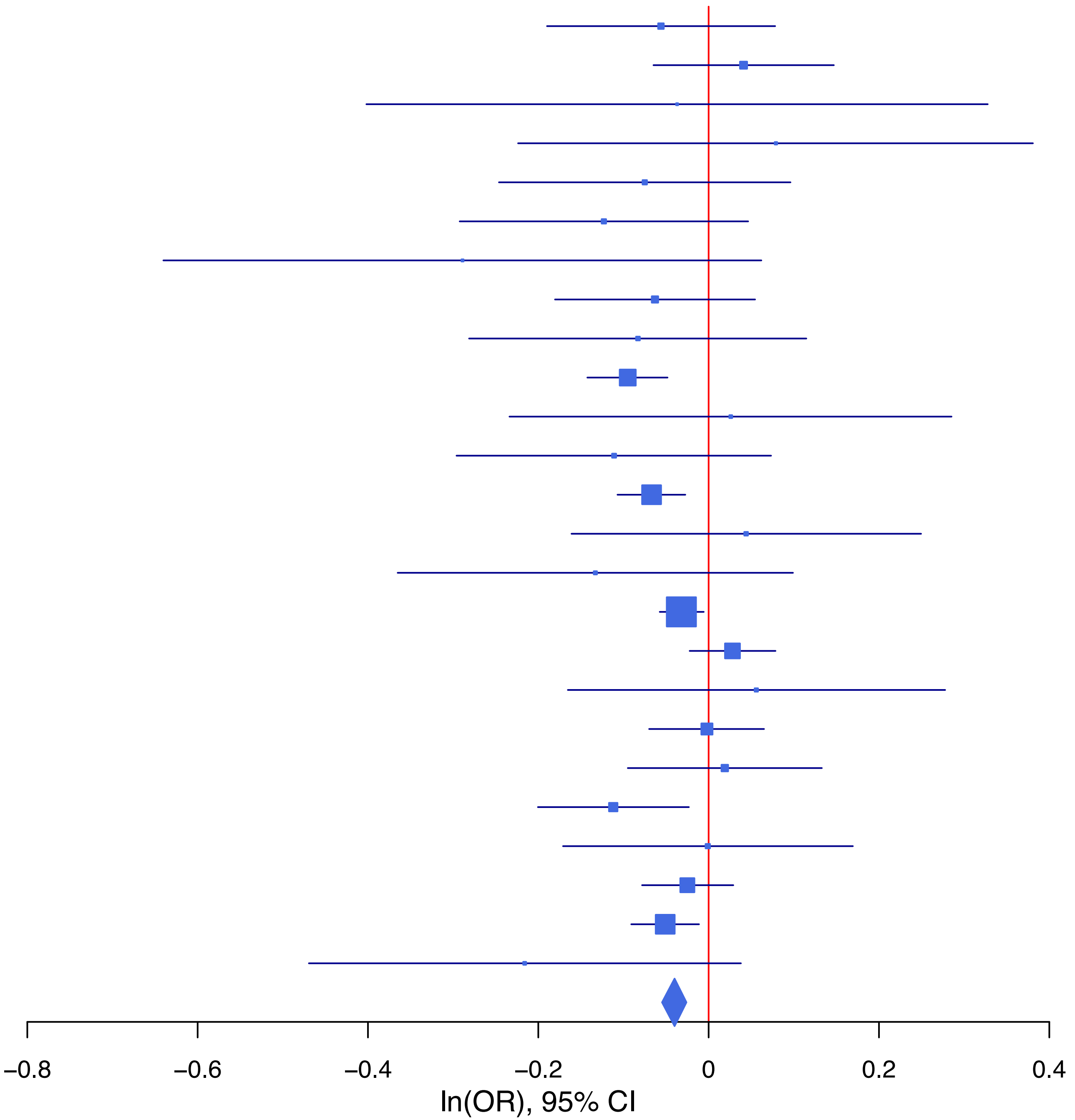

**rs7937151 G/T 11:112835024**

| Cohort | P | ln(OR) | SE |
|---|---|---|---|
| ADHEA_EUR | 0.4095 | 0.057 | 0.069 |
| ALSPC_EUR | 0.5907 | −0.03 | 0.055 |
| BEPS7_EUR | 0.5711 | 0.11 | 0.194 |
| BHRCM_EUR | 0.9451 | 0.012 | 0.171 |
| BOR17_EUR | 0.794 | 0.023 | 0.088 |
| BOR2C_EUR | 0.0941 | 0.152 | 0.091 |
| BOR2E_EUR | 0.3638 | −0.186 | 0.205 |
| COGA1_EUR | 0.377 | 0.052 | 0.059 |
| CUINT_EUR | 0.0547 | 0.192 | 0.1 |
| ESTB2_EUR | 0.069 | 0.042 | 0.023 |
| FINNG_EUR | 0.1464 | 0.034 | 0.024 |
| GEDIS_EUR | 0.8024 | −0.034 | 0.136 |
| GISS1_EUR | 0.4595 | −0.058 | 0.078 |
| GISS2_EUR | 0.07 | 0.175 | 0.096 |
| IPSYC_EUR | 0.041 | 0.042 | 0.021 |
| JANS3_EUR | 0.0845 | 0.182 | 0.106 |
| JANS4_EUR | 0.5029 | 0.083 | 0.124 |
| MVPXQ_EUR | 2.72e−06 | 0.062 | 0.013 |
| PGCBD_EUR | 0.0666 | 0.048 | 0.026 |
| PGCED_EUR | 0.3068 | 0.117 | 0.115 |
| PGCMD_EUR | 0.5069 | 0.023 | 0.035 |
| PGCPT_EUR | 0.4147 | −0.048 | 0.059 |
| PGCSZ_EUR | 0.1022 | 0.061 | 0.037 |
| PRFCT_EUR | 0.58 | −0.025 | 0.044 |
| PSYCR_EUR | 0.401 | 0.077 | 0.092 |
| QIMRB_EUR | 0.193 | 0.038 | 0.029 |
| UKBJC_EUR | 0.0364 | 0.043 | 0.021 |
| YPENN_EUR | 0.8752 | −0.02 | 0.128 |
| **meta** | **6.16e−11** | **0.045** | **0.007** |

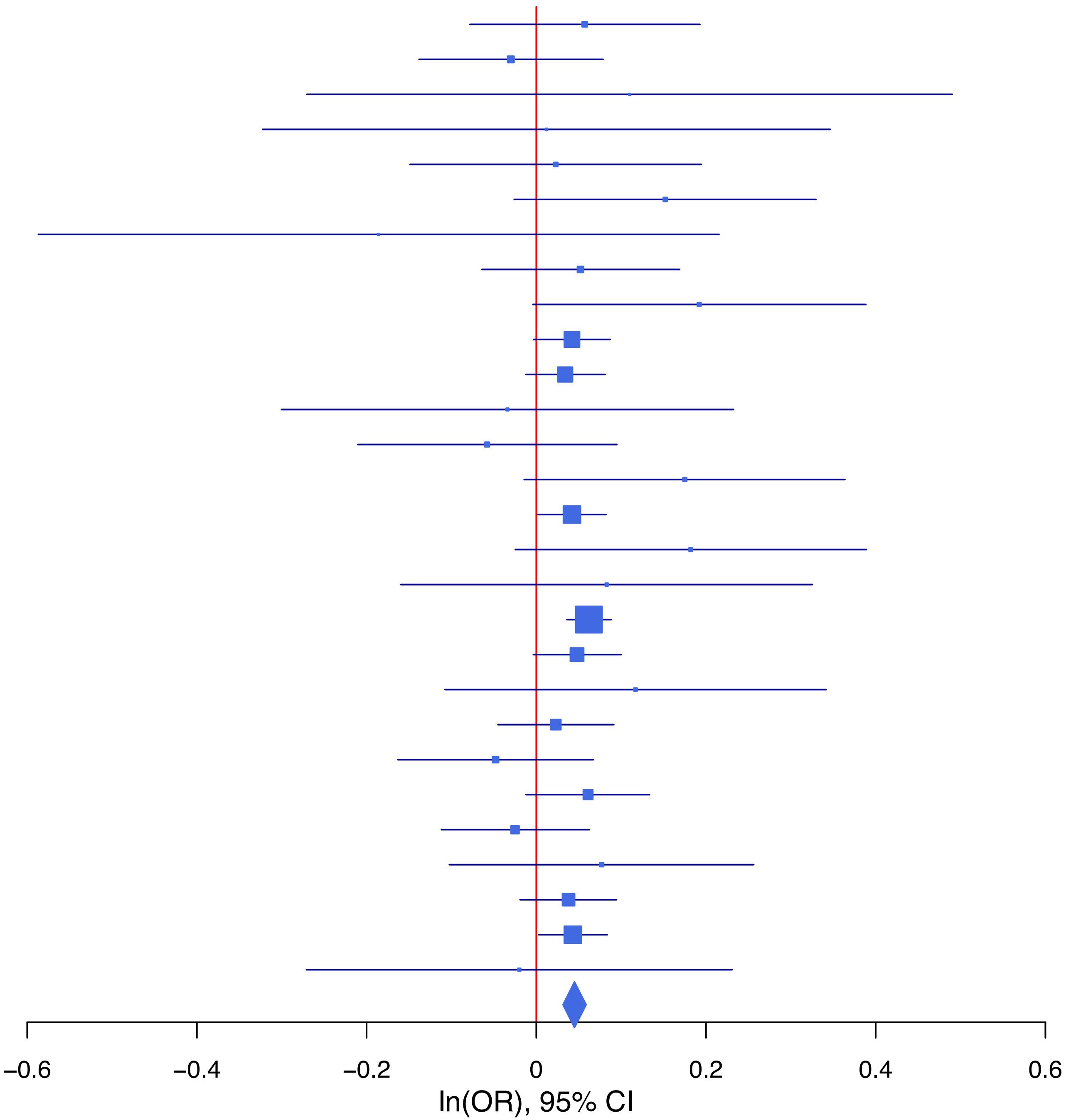

**rs9276627 T/C 6:32743835**

| Cohort | P | ln(OR) | SE |
|---|---|---|---|
| ADHEA_EUR | 0.0549 | 0.221 | 0.115 |
| ALSPC_EUR | 0.6818 | 0.04 | 0.098 |
| BEPS7_EUR | 0.6116 | −0.173 | 0.342 |
| BHRCM_EUR | 0.6243 | −0.151 | 0.308 |
| BOR17_EUR | 0.0168 | 0.356 | 0.149 |
| BOR2C_EUR | 0.7379 | −0.051 | 0.153 |
| BOR2E_EUR | 0.8894 | −0.047 | 0.337 |
| COGA1_EUR | 0.7615 | −0.032 | 0.105 |
| CUINT_EUR | 0.6687 | 0.077 | 0.179 |
| ESTB2_EUR | 0.0668 | 0.078 | 0.043 |
| FINNG_EUR | 0.1316 | 0.057 | 0.038 |
| GEDIS_EUR | 0.88 | −0.034 | 0.223 |
| GISS2_EUR | 0.3619 | 0.156 | 0.171 |
| IPSYC_EUR | 0.4554 | 0.024 | 0.032 |
| JANS3_EUR | 0.7185 | −0.07 | 0.194 |
| JANS4_EUR | 0.729 | 0.076 | 0.22 |
| MVPXQ_EUR | 0.0051 | 0.066 | 0.023 |
| PGCBD_EUR | 0.0712 | 0.089 | 0.05 |
| PGCMD_EUR | 0.2438 | 0.08 | 0.069 |
| PGCPT_EUR | 0.4887 | −0.073 | 0.105 |
| PRFCT_EUR | 0.7163 | 0.026 | 0.073 |
| PSYCR_EUR | 0.8964 | −0.021 | 0.161 |
| QIMRB_EUR | 0.0124 | 0.122 | 0.049 |
| UKBJC_EUR | 8e−04 | 0.12 | 0.036 |
| YPENN_EUR | 0.2775 | −0.282 | 0.259 |
| **meta** | **3.43e−08** | **0.067** | **0.012** |

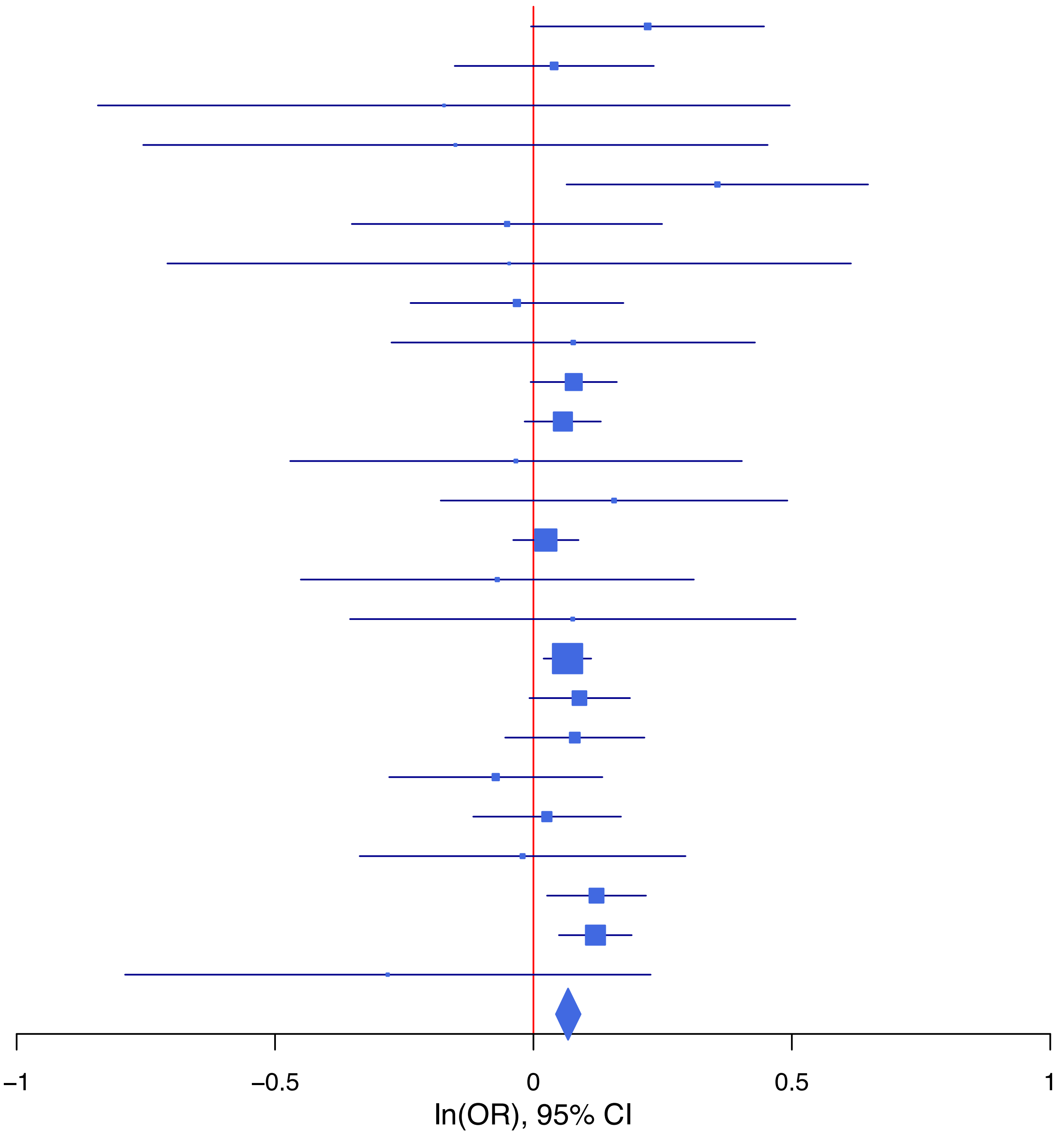

**rs9306311   G/C   22:37050675**

| Cohort | P | ln(OR) | SE |
|---|---|---|---|
| ALSPC_EUR | 0.9088 | 0.007 | 0.06 |
| BEPS7_EUR | 0.939 | −0.017 | 0.226 |
| BOR17_EUR | 0.9511 | −0.006 | 0.103 |
| BOR2C_EUR | 0.8632 | −0.018 | 0.103 |
| BOR2E_EUR | 0.1125 | −0.349 | 0.22 |
| COGA1_EUR | 0.461 | −0.048 | 0.066 |
| ESTB2_EUR | 0.1903 | −0.036 | 0.028 |
| FINNG_EUR | 0.8793 | −0.004 | 0.027 |
| GEDIS_EUR | 0.7627 | −0.043 | 0.143 |
| GISS1_EUR | 0.964 | −0.004 | 0.09 |
| GISS2_EUR | 0.3267 | −0.121 | 0.123 |
| JANS3_EUR | 0.1971 | −0.163 | 0.127 |
| JANS4_EUR | 0.7904 | 0.038 | 0.143 |
| MVPXQ_EUR | 5e−04 | −0.049 | 0.014 |
| PGCBD_EUR | 0.0205 | −0.07 | 0.03 |
| PGCED_EUR | 0.5029 | −0.084 | 0.126 |
| PGCMD_EUR | 0.9855 | −0.001 | 0.039 |
| PGCPT_EUR | 0.1725 | −0.092 | 0.068 |
| PGCSZ_EUR | 0.6643 | −0.019 | 0.043 |
| PRFCT_EUR | 0.0084 | −0.145 | 0.055 |
| PSYCR_EUR | 0.9842 | −0.002 | 0.109 |
| QIMRB_EUR | 0.1081 | −0.055 | 0.034 |
| UKBJC_EUR | 0.0014 | −0.073 | 0.023 |
| **meta** | **1.34e−08** | **−0.047** | **0.008** |

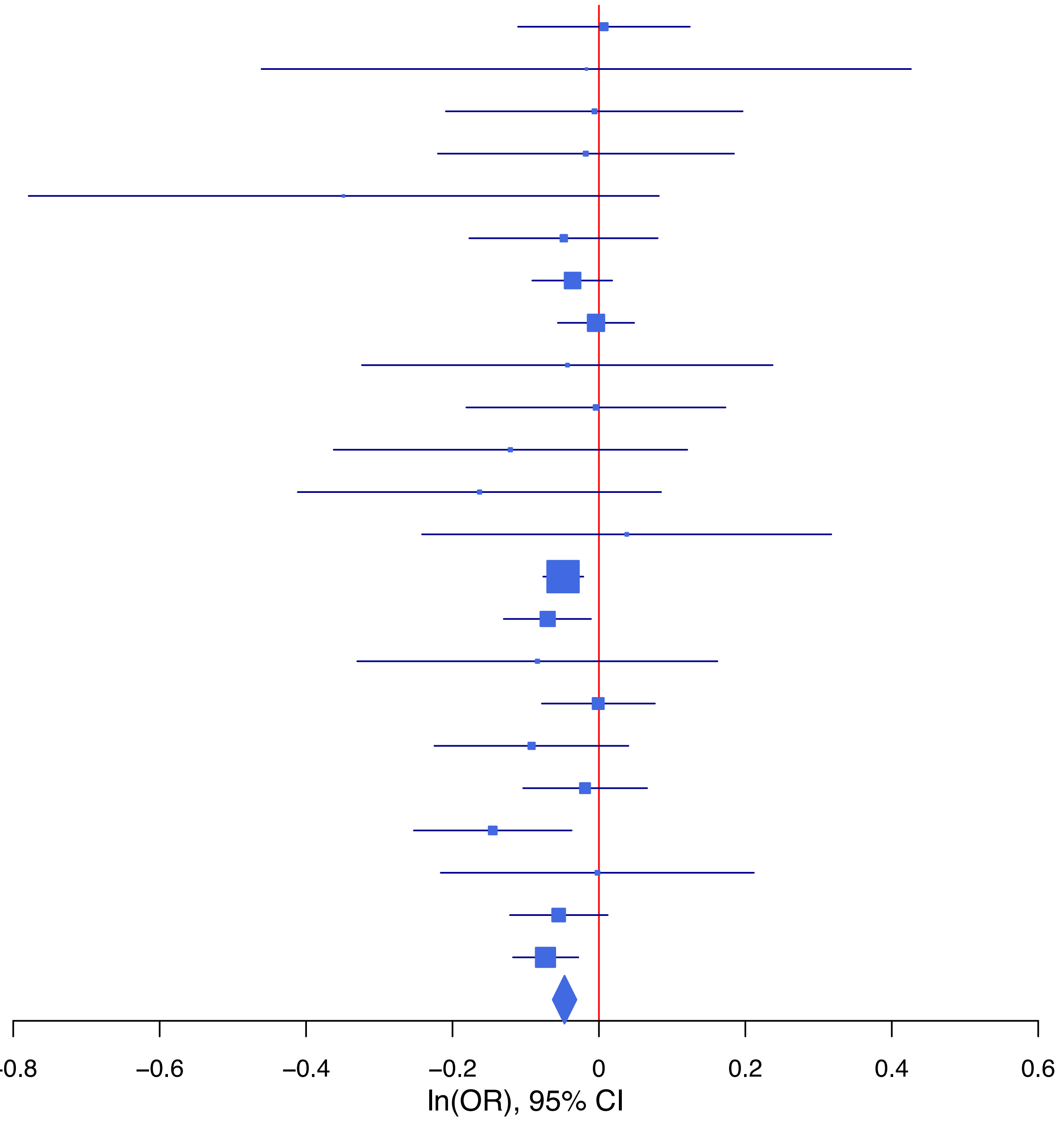

**rs9849038 A/G 3:49771990**

| Cohort | P | ln(OR) | SE |
|---|---|---|---|
| ADHEA_EUR | 0.2214 | −0.108 | 0.089 |
| ALSPC_EUR | 0.8127 | 0.016 | 0.067 |
| BEPS7_EUR | 0.4554 | 0.167 | 0.224 |
| BHRCM_EUR | 0.5565 | 0.112 | 0.191 |
| BOR17_EUR | 0.2631 | 0.124 | 0.111 |
| BOR2C_EUR | 0.0682 | 0.207 | 0.113 |
| BOR2E_EUR | 0.1136 | 0.363 | 0.229 |
| COGA1_EUR | 0.0646 | 0.139 | 0.075 |
| CUINT_EUR | 0.8817 | 0.019 | 0.124 |
| ESTB2_EUR | 0.5116 | 0.022 | 0.034 |
| FINNG_EUR | 0.3783 | 0.032 | 0.037 |
| GEDIS_EUR | 0.6841 | 0.066 | 0.162 |
| GISS2_EUR | 0.3998 | 0.106 | 0.126 |
| IPSYC_EUR | 0.0065 | 0.068 | 0.025 |
| JANS3_EUR | 0.7886 | −0.036 | 0.133 |
| JANS4_EUR | 0.8374 | −0.031 | 0.149 |
| MIREC_EUR | 0.755 | −0.06 | 0.193 |
| MVPXQ_EUR | 0.0047 | 0.047 | 0.016 |
| PGCBD_EUR | 0.0055 | 0.092 | 0.033 |
| PGCED_EUR | 0.5783 | 0.08 | 0.144 |
| PGCMD_EUR | 0.5155 | 0.029 | 0.045 |
| PGCPT_EUR | 0.0051 | 0.195 | 0.07 |
| PGCSZ_EUR | 0.4341 | 0.037 | 0.047 |
| PRFCT_EUR | 0.7851 | 0.015 | 0.056 |
| PSYCR_EUR | 0.3577 | −0.102 | 0.111 |
| QIMRB_EUR | 0.1086 | 0.057 | 0.035 |
| UKBJC_EUR | 0.0191 | 0.061 | 0.026 |
| **meta** | **2.32e−09** | **0.053** | **0.009** |

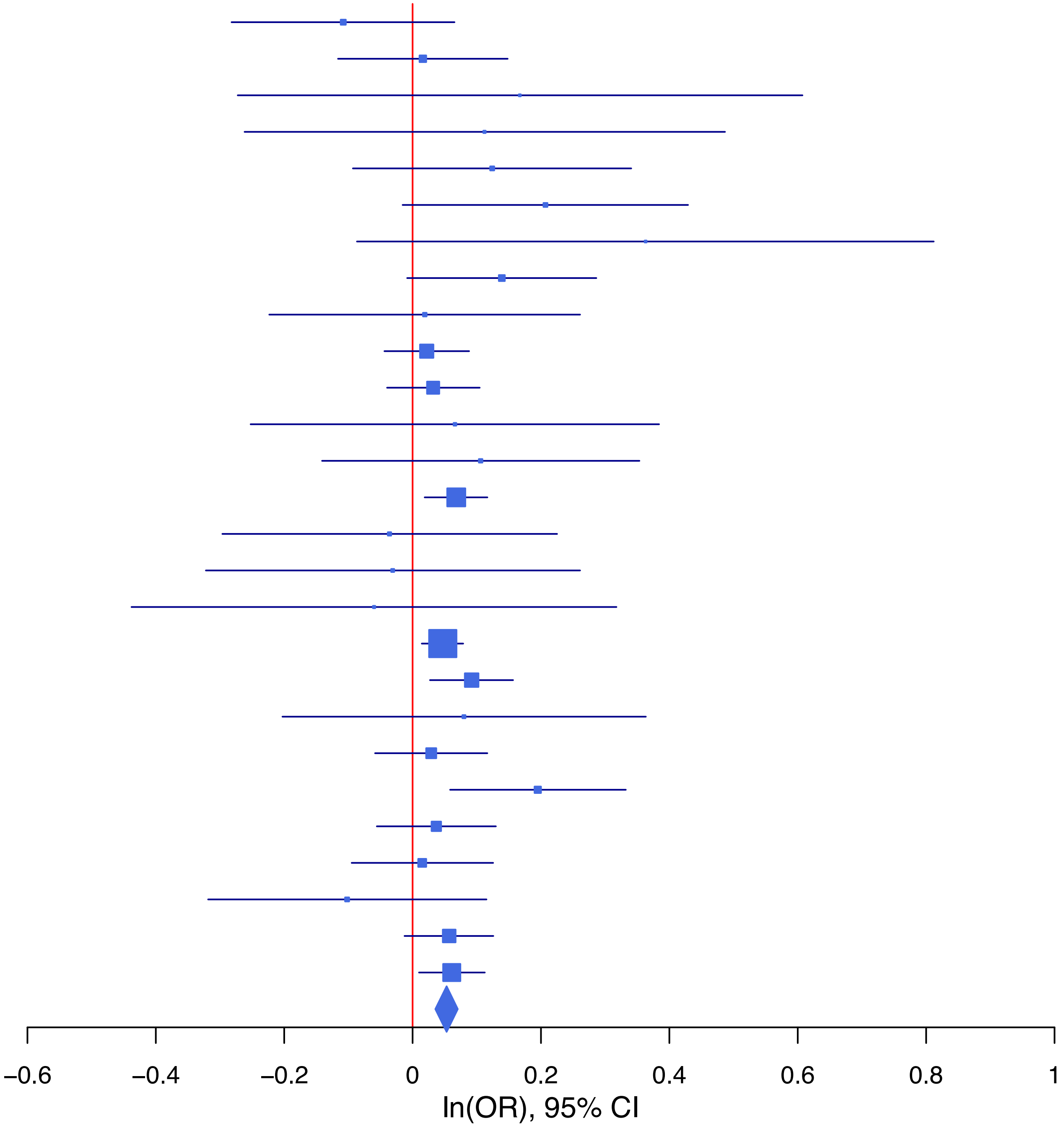

**rs9853056 T/C 3:52555957**

| Cohort | P | ln(OR) | SE |
|---|---|---|---|
| ADHEA_EUR | 0.5671 | 0.039 | 0.069 |
| ALSPC_EUR | 0.3922 | 0.047 | 0.054 |
| BEPS7_EUR | 0.9062 | −0.021 | 0.174 |
| BHRCM_EUR | 0.1986 | 0.205 | 0.16 |
| BOR17_EUR | 0.1935 | 0.11 | 0.085 |
| BOR2C_EUR | 0.0588 | 0.163 | 0.086 |
| BOR2E_EUR | 0.7324 | −0.065 | 0.19 |
| COGA1_EUR | 0.0158 | 0.144 | 0.06 |
| CUINT_EUR | 0.754 | 0.031 | 0.101 |
| ESTB2_EUR | 0.002 | 0.071 | 0.023 |
| FINNG_EUR | 0.5305 | 0.014 | 0.023 |
| GEDIS_EUR | 0.444 | 0.101 | 0.132 |
| GISS1_EUR | 0.2197 | 0.097 | 0.079 |
| GISS2_EUR | 0.6887 | −0.037 | 0.093 |
| IPSYC_EUR | 0.1294 | 0.031 | 0.02 |
| JANS3_EUR | 0.6169 | 0.051 | 0.103 |
| JANS4_EUR | 0.6327 | 0.056 | 0.118 |
| MVPXQ_EUR | 0.0105 | 0.049 | 0.019 |
| PGCBD_EUR | 0.0092 | 0.067 | 0.026 |
| PGCED_EUR | 0.5776 | 0.065 | 0.116 |
| PGCMD_EUR | 0.6359 | −0.016 | 0.035 |
| PGCPT_EUR | 0.5841 | 0.031 | 0.057 |
| PGCSZ_EUR | 0.0023 | 0.114 | 0.038 |
| PRFCT_EUR | 0.6694 | 0.019 | 0.044 |
| PSYCR_EUR | 0.0302 | 0.189 | 0.087 |
| QIMRB_EUR | 0.0333 | 0.058 | 0.027 |
| UKBJC_EUR | 0.9541 | −0.001 | 0.02 |
| YPENN_EUR | 0.4615 | 0.095 | 0.129 |
| **meta** | **3.69e−09** | **0.043** | **0.007** |

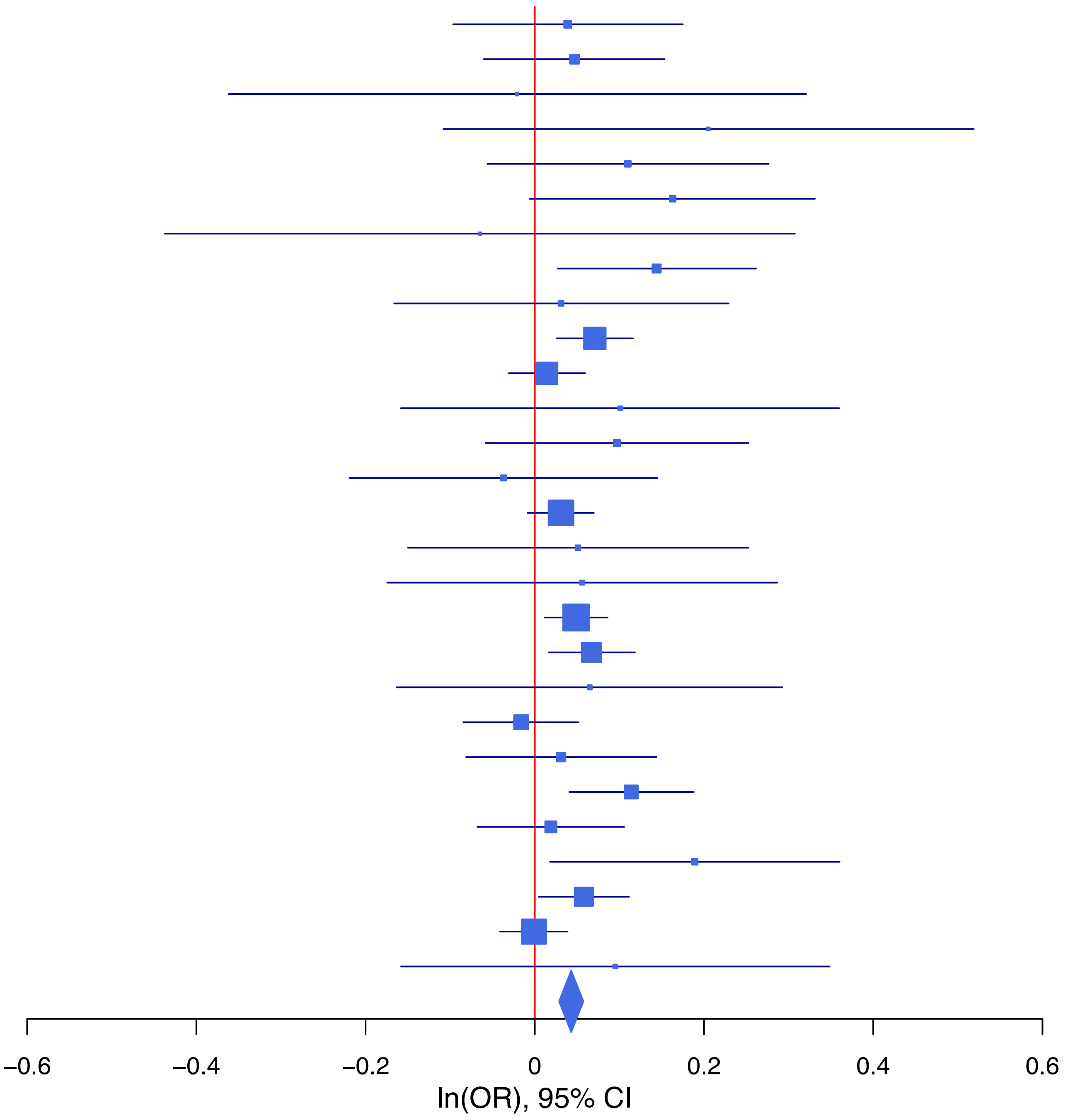

**Supplementary Data 1F: Forest plots of lead SNPs at the 2 genome-wide significant loci from the GWAS meta-analysis of suicide death in European ancestry samples.**
Each box represents the log odds ratio (OR) from an individual contributing cohort, with horizontal lines indicating the 95% confidence interval (CI). The diamond represents the overall meta-analytic estimate across studies.

**rs36212732 G/A 10:124215198**

| Cohort | P | ln(OR) | SE |
| --- | --- | --- | --- |
| CUINT_EUR | 0.1558 | −0.165 | 0.116 |
| ESTB2_EUR | 0.7787 | 0.039 | 0.139 |
| UKBJC_EUR | 0.3857 | −0.09 | 0.104 |
| UTAH2_EUR | 5.46e−08 | −0.121 | 0.022 |
| **meta** | **2.74e−08** | **−0.118** | **0.021** |

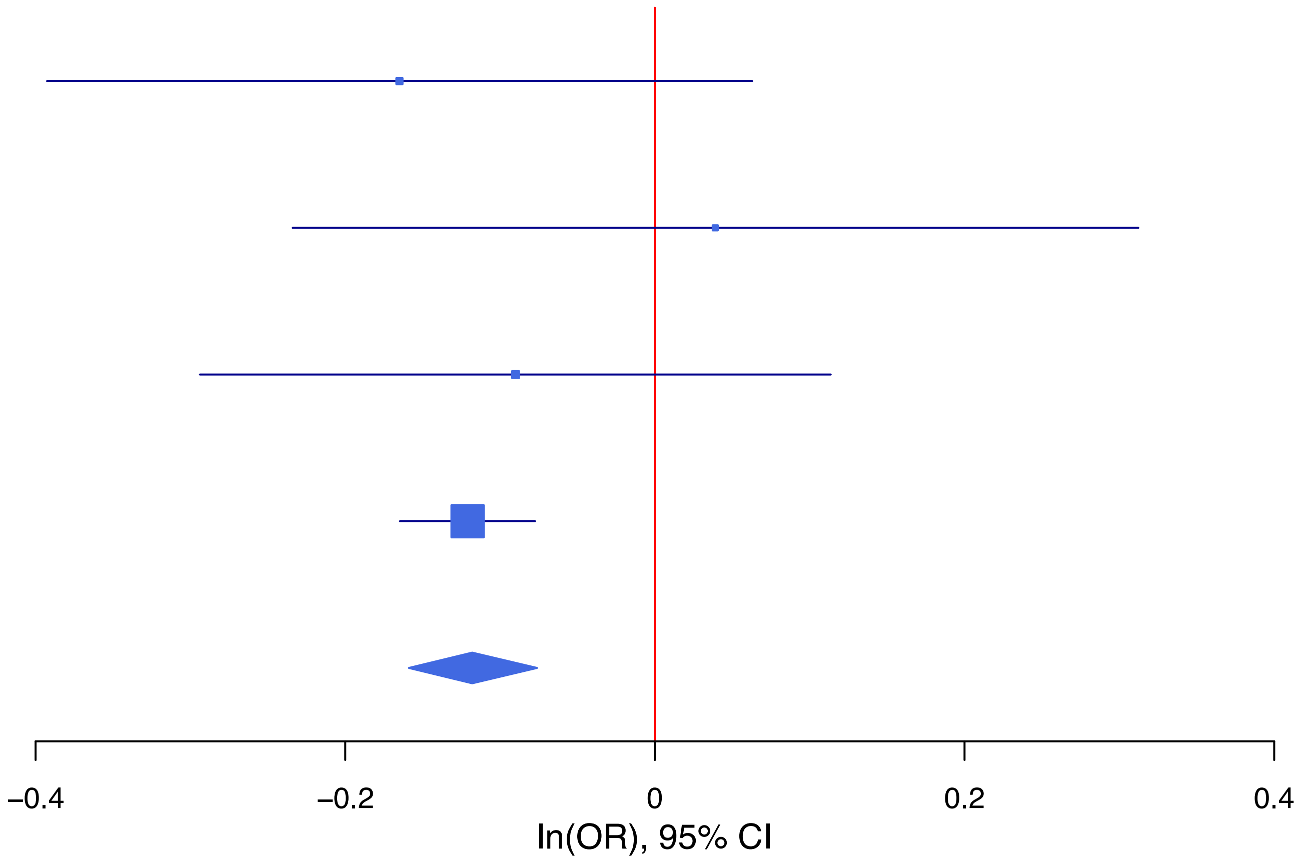

**rs7335526   A/G   13:76707220**

| Cohort | P | ln(OR) | SE |
|---|---|---|---|
| CUINT_EUR | 0.071 | −0.167 | 0.093 |
| ESTB2_EUR | 0.3321 | 0.123 | 0.127 |
| MVPXQ_EUR | 0.0429 | −0.115 | 0.057 |
| UKBJC_EUR | 0.0623 | −0.158 | 0.085 |
| UTAH2_EUR | 2.30e−06 | −0.089 | 0.019 |
| **meta** | **4.78e−08** | **−0.093** | **0.017** |

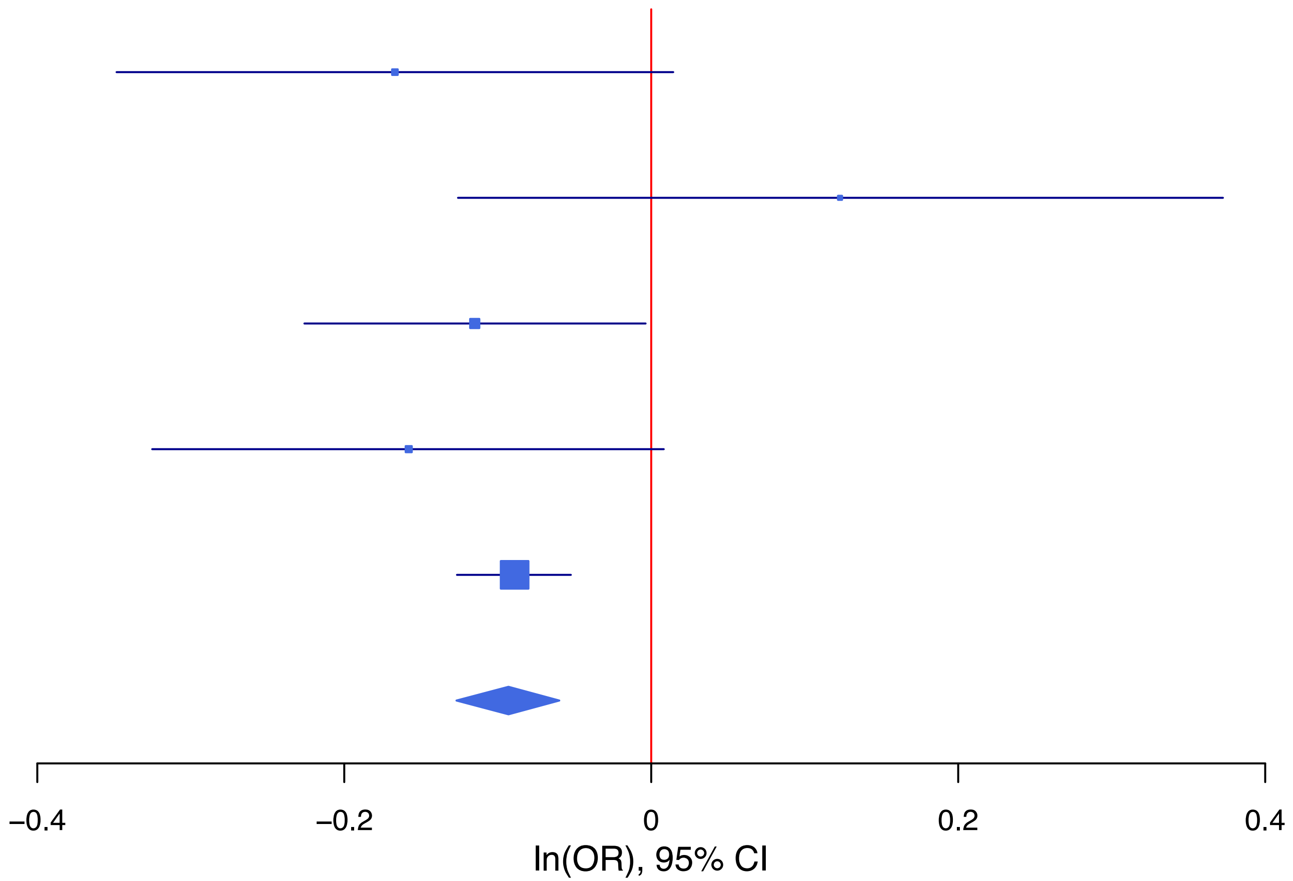

**Supplementary Data 1G: Forest plots of lead SNPs at the 53 genome-wide significant loci from the multi-ancestry GWAS meta-analysis of suicidal behavior.**

Each box represents the log odds ratio (OR) from an individual contributing cohort, with horizontal lines indicating the 95% confidence interval (CI). The diamond represents the overall meta-analytic estimate across studies.

**rs10759942 T/C 9:120515918**

| Cohort | P | ln(OR) | SE |
|---|---|---|---|
| ADHEA_AFR | 0.4321 | 0.093 | 0.117 |
| ADHEA_EUR | 0.5906 | 0.038 | 0.07 |
| BEPS7_EUR | 0.7092 | −0.07 | 0.188 |
| BHRCM_AFR | 0.3731 | 0.171 | 0.193 |
| BHRCM_EUR | 0.562 | 0.094 | 0.162 |
| BHRCM_LAT | 0.1855 | −0.497 | 0.375 |
| BOR17_EUR | 0.6807 | 0.037 | 0.089 |
| BOR2C_EUR | 0.3705 | 0.08 | 0.09 |
| BOR2E_EUR | 0.713 | 0.067 | 0.182 |
| CNVRG_EAS | 0.7268 | 0.072 | 0.206 |
| COGA1_AFR | 0.0025 | 0.31 | 0.102 |
| COGA1_EUR | 0.2949 | 0.065 | 0.062 |
| CUINT_EUR | 0.3567 | 0.071 | 0.078 |
| CVEDA_CSA | 0.4955 | 0.127 | 0.187 |
| ESTB2_EUR | 6e−04 | 0.08 | 0.023 |
| FINNG_EUR | 0.2279 | 0.029 | 0.024 |
| GEDIS_EUR | 0.7796 | 0.038 | 0.136 |
| GISS1_EUR | 0.8338 | −0.018 | 0.084 |
| GISS2_EUR | 0.1968 | 0.126 | 0.098 |
| GTPRJ_AFR | 0.1243 | 0.092 | 0.06 |
| IPSYC_EUR | 0.0179 | 0.045 | 0.019 |
| JANS3_EUR | 0.3065 | −0.116 | 0.113 |
| JANS4_EUR | 0.2676 | 0.134 | 0.121 |
| MIREC_EUR | 0.8037 | −0.039 | 0.158 |
| MVPXQ_AFR | 0.1489 | 0.031 | 0.022 |
| MVPXQ_EAS | 0.8029 | −0.056 | 0.223 |
| MVPXQ_EUR | 3e−04 | 0.048 | 0.013 |
| MVPXQ_LAT | 0.3362 | 0.033 | 0.035 |
| PGCBD_EUR | 0.9462 | 0.002 | 0.027 |
| PGCED_EUR | 0.0561 | −0.243 | 0.127 |
| PGCMD_EUR | 0.6722 | 0.015 | 0.035 |
| PGCPT_AFR | 0.6206 | −0.051 | 0.104 |
| PGCPT_EUR | 0.8958 | −0.008 | 0.06 |
| PGCSZ_EUR | 0.6246 | −0.019 | 0.039 |
| PRFCT_EUR | 0.8491 | −0.009 | 0.046 |
| PSYCR_EUR | 0.9349 | 0.008 | 0.092 |
| QIMRB_EUR | 0.0298 | 0.062 | 0.028 |
| STRR1_LAT | 0.6592 | 0.081 | 0.185 |
| UKBJC_EUR | 0.4289 | 0.016 | 0.021 |
| UTAH2_EUR | 0.3358 | 0.019 | 0.02 |
| UTAMR_LAT | 0.7863 | 0.01 | 0.037 |
| YPENN_EUR | 0.3954 | 0.112 | 0.132 |
| **meta** | **4.09e−09** | **0.035** | **0.006** |

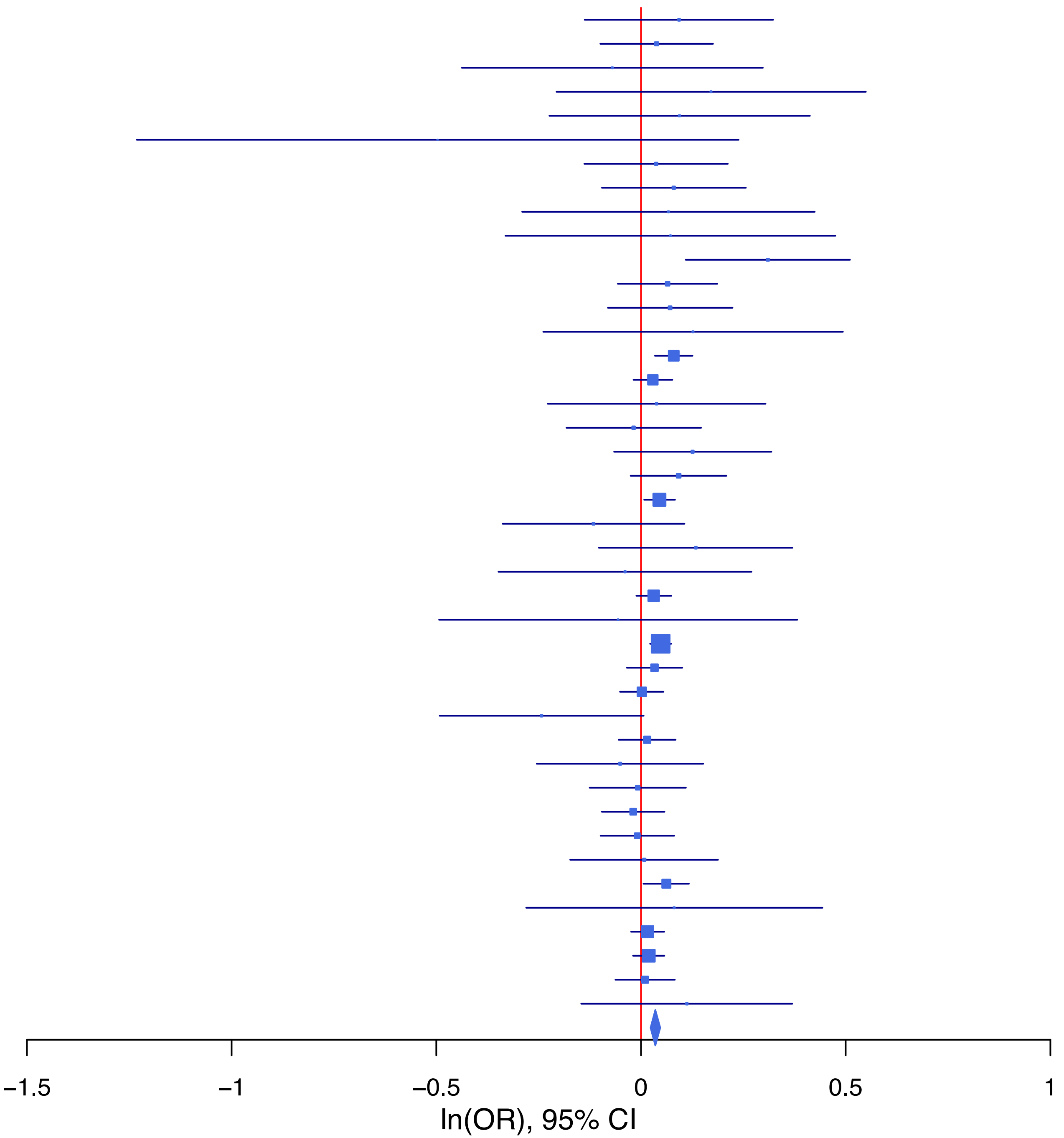

**rs10835368 T/C 11:28643299**

| Cohort | P | ln(OR) | SE |
|---|---|---|---|
| ALSPC_EUR | 0.8448 | −0.011 | 0.056 |
| BEPS7_EUR | 0.2723 | 0.194 | 0.176 |
| BHRCM_AFR | 0.4531 | 0.18 | 0.24 |
| BHRCM_EUR | 0.3065 | −0.17 | 0.167 |
| BHRCM_LAT | 0.0363 | −0.702 | 0.335 |
| BOR17_EUR | 0.6208 | 0.044 | 0.09 |
| BOR2C_EUR | 0.3551 | −0.082 | 0.088 |
| BOR2E_EUR | 0.004 | −0.554 | 0.193 |
| CNVRG_EAS | 0.3747 | −0.045 | 0.051 |
| COGA1_AFR | 0.4636 | −0.08 | 0.109 |
| COGA1_EUR | 0.937 | −0.005 | 0.06 |
| CUINT_EUR | 0.182 | −0.106 | 0.079 |
| CVEDA_CSA | 0.6367 | −0.08 | 0.17 |
| ESTB2_EUR | 0.0764 | −0.04 | 0.023 |
| FINNG_EUR | 0.752 | 0.007 | 0.024 |
| GEDIS_EUR | 0.7667 | −0.041 | 0.138 |
| GISS1_EUR | 0.9374 | 0.006 | 0.079 |
| GISS2_EUR | 0.8025 | −0.023 | 0.093 |
| GREAT_EAS | 0.3331 | −0.103 | 0.106 |
| GTPRJ_AFR | 0.4078 | 0.057 | 0.068 |
| IPSYC_EUR | 0.6252 | −0.009 | 0.019 |
| JANS3_EUR | 0.456 | −0.081 | 0.108 |
| JANS4_EUR | 0.2779 | −0.134 | 0.124 |
| MVPXQ_AFR | 0.0146 | −0.057 | 0.023 |
| MVPXQ_EAS | 0.0949 | −0.203 | 0.122 |
| MVPXQ_EUR | 1e−04 | −0.05 | 0.013 |
| MVPXQ_LAT | 0.6468 | −0.015 | 0.034 |
| PGCBD_EUR | 0.074 | −0.048 | 0.027 |
| PGCED_EUR | 0.5728 | −0.066 | 0.117 |
| PGCMD_EUR | 0.0044 | −0.101 | 0.036 |
| PGCPT_AFR | 0.6549 | 0.052 | 0.115 |
| PGCPT_EUR | 0.3135 | 0.06 | 0.06 |
| PGCSZ_EUR | 0.2504 | −0.044 | 0.038 |
| PRFCT_EUR | 0.3573 | −0.042 | 0.046 |
| PSYCR_EUR | 0.062 | −0.171 | 0.092 |
| QIMRB_EUR | 0.0073 | −0.075 | 0.028 |
| SNUBH−ASA_EAS | 0.5625 | 0.081 | 0.14 |
| SNUBH−KCHIP_EAS | 0.1914 | −0.235 | 0.18 |
| STRR1_LAT | 0.8458 | 0.036 | 0.183 |
| UKBJC_EUR | 0.0713 | −0.036 | 0.02 |
| UTAH2_EUR | 0.1691 | −0.027 | 0.02 |
| UTAMR_LAT | 0.0688 | −0.068 | 0.037 |
| YPENN_EUR | 0.9898 | 0.002 | 0.132 |
| **meta** | **3.88e−11** | **−0.039** | **0.006** |

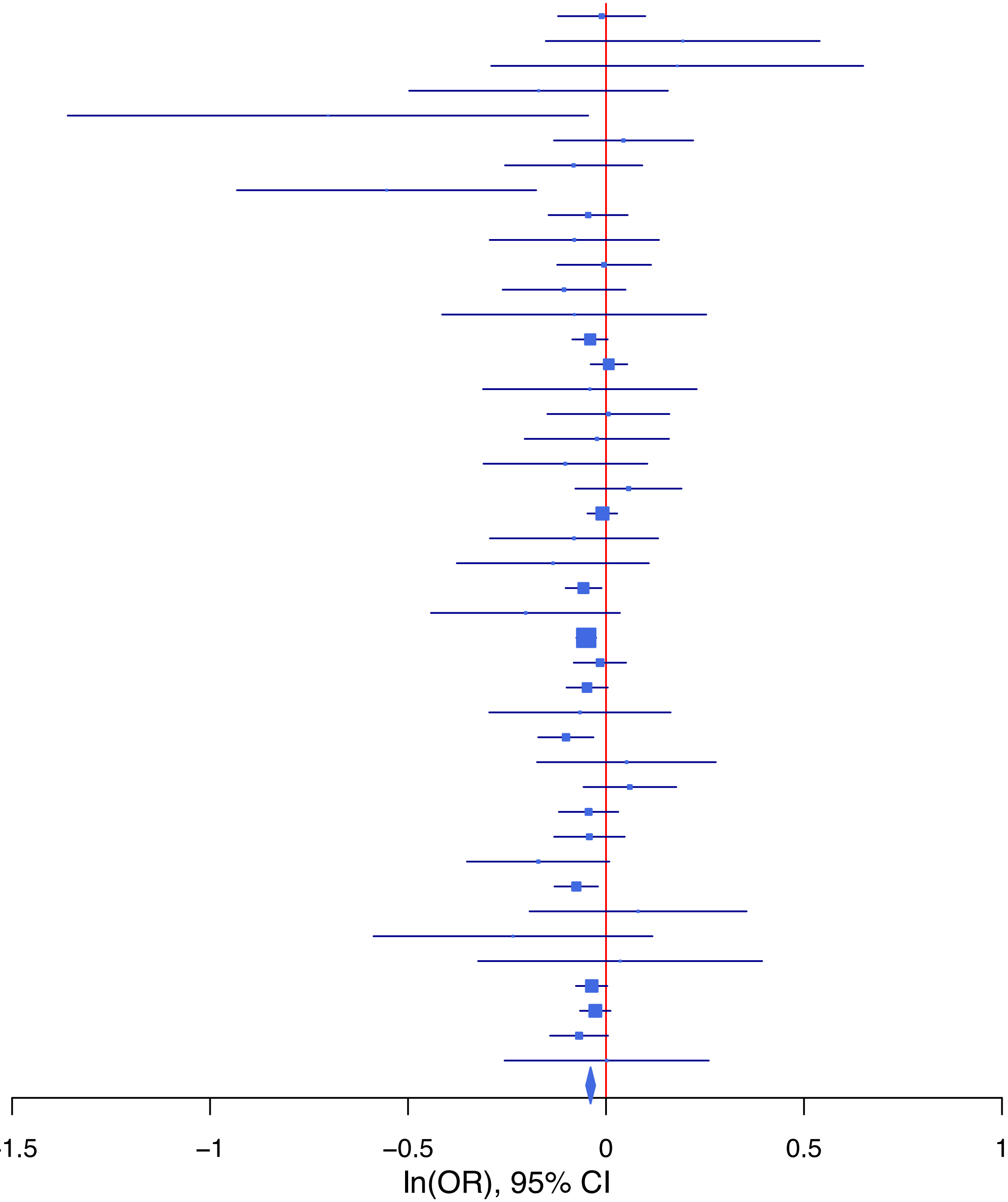

rs10896656 G/C 11:57668941

| Cohort | P | ln(OR) | SE |
|---|---|---|---|
| ADHEA_AFR | 0.9221 | −0.019 | 0.194 |
| ADHEA_EUR | 0.3708 | 0.065 | 0.072 |
| ALSPC_EUR | 0.59 | −0.031 | 0.058 |
| BEPS7_EUR | 0.8203 | −0.043 | 0.189 |
| BHRCM_AFR | 0.9896 | 0.008 | 0.605 |
| BHRCM_EUR | 0.0335 | 0.34 | 0.16 |
| BHRCM_LAT | 0.8543 | 0.103 | 0.565 |
| BOR17_EUR | 0.3861 | 0.081 | 0.093 |
| BOR2C_EUR | 0.4197 | 0.075 | 0.093 |
| BOR2E_EUR | 0.9638 | 0.009 | 0.192 |
| CNVRG_EAS | 0.4544 | 0.057 | 0.076 |
| COGA1_AFR | 0.8674 | −0.025 | 0.152 |
| COGA1_EUR | 0.7362 | 0.021 | 0.063 |
| CUINT_EUR | 0.6438 | −0.037 | 0.08 |
| CVEDA_CSA | 0.0678 | 0.307 | 0.168 |
| ESTB2_EUR | 4e−04 | 0.083 | 0.024 |
| FINNG_EUR | 0.7267 | 0.008 | 0.024 |
| GEDIS_EUR | 0.8009 | 0.035 | 0.14 |
| GISS1_EUR | 0.1667 | 0.116 | 0.084 |
| GISS2_EUR | 0.4959 | 0.068 | 0.1 |
| GTPRJ_AFR | 0.2297 | 0.108 | 0.09 |
| IPSYC_EUR | 0.0033 | 0.057 | 0.019 |
| JANS4_EUR | 0.1627 | 0.173 | 0.124 |
| JAPAN_EAS | 0.6082 | 0.06 | 0.117 |
| MIREC_AFR | 0.8713 | 0.036 | 0.223 |
| MIREC_EUR | 0.9443 | 0.012 | 0.168 |
| MVPXQ_EAS | 0.3817 | −0.152 | 0.173 |
| MVPXQ_EUR | 0.036 | 0.028 | 0.013 |
| MVPXQ_LAT | 0.3829 | −0.038 | 0.044 |
| PGCBD_EUR | 0.3043 | 0.029 | 0.028 |
| PGCED_EUR | 0.4669 | −0.095 | 0.13 |
| PGCMD_EUR | 0.3011 | 0.038 | 0.037 |
| PGCPT_AFR | 0.1636 | 0.25 | 0.179 |
| PGCPT_EUR | 0.2632 | 0.068 | 0.061 |
| PGCSZ_EUR | 0.8419 | 0.008 | 0.04 |
| PRFCT_EUR | 0.179 | −0.064 | 0.048 |
| PSYCR_EUR | 0.6816 | 0.04 | 0.097 |
| QIMRB_EUR | 0.0049 | 0.083 | 0.03 |
| SNUBH−KCHIP_EAS | 0.8412 | −0.062 | 0.309 |
| STRR1_LAT | 0.6361 | 0.101 | 0.213 |
| UKBJC_EUR | 0.3933 | 0.018 | 0.021 |
| UTAH2_EUR | 0.0904 | 0.034 | 0.02 |
| UTAMR_LAT | 0.5254 | 0.036 | 0.056 |
| YPENN_AFR | 0.5209 | 0.144 | 0.225 |
| YPENN_EUR | 0.1714 | 0.187 | 0.137 |
| **meta** | **2.90e−08** | **0.035** | **0.006** |

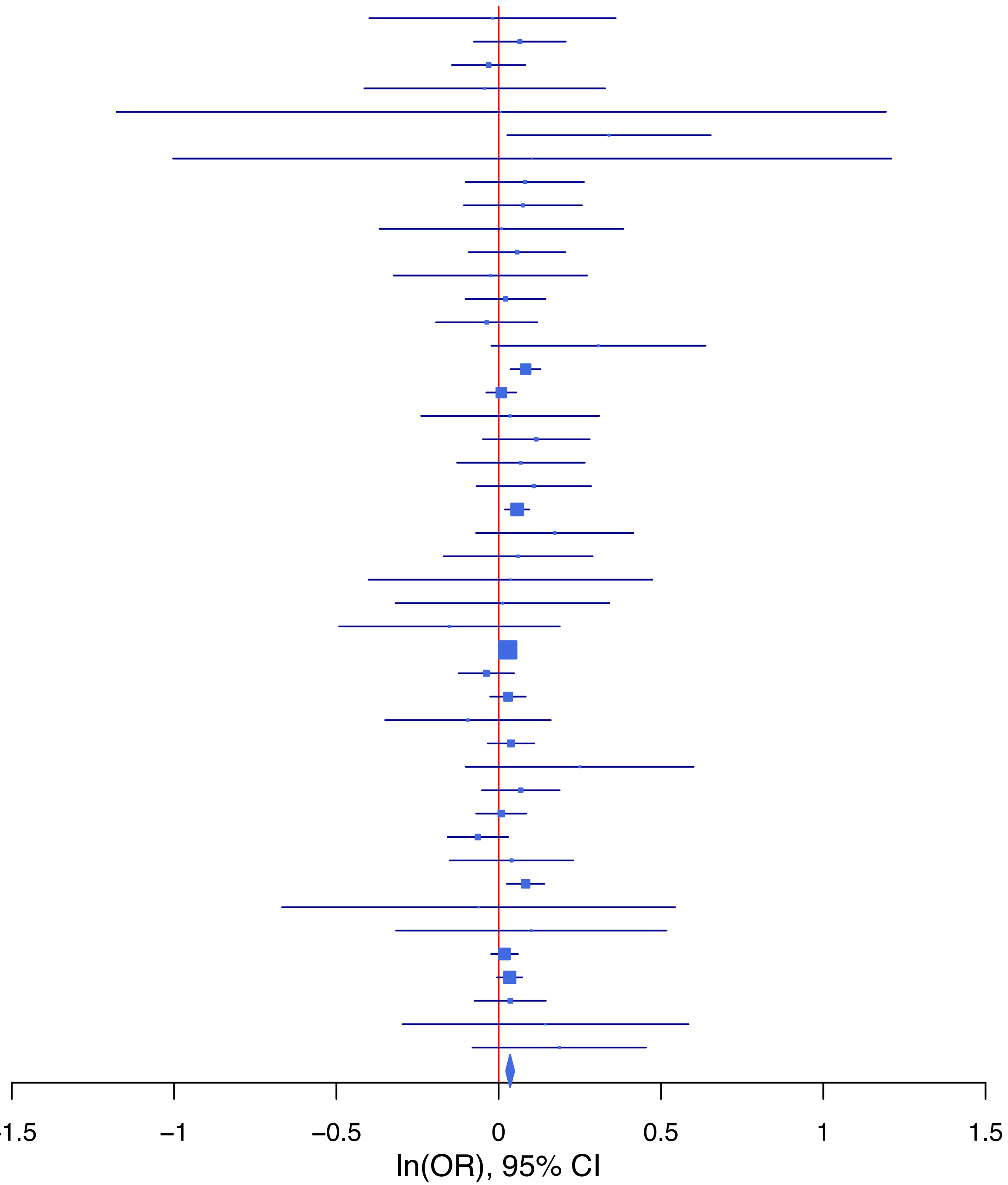

**rs10979816 G/A 9:112067488**

| Cohort | P | ln(OR) | SE |
|---|---|---|---|
| ADHEA_AFR | 0.0098 | 0.581 | 0.225 |
| ADHEA_EUR | 0.0356 | 0.212 | 0.101 |
| ALSPC_EUR | 0.508 | −0.057 | 0.086 |
| BEPS7_EUR | 0.9336 | 0.025 | 0.296 |
| BOR17_EUR | 0.2007 | 0.194 | 0.152 |
| BOR2C_EUR | 0.4763 | 0.105 | 0.147 |
| BOR2E_EUR | 0.4387 | 0.234 | 0.303 |
| COGA1_AFR | 0.8883 | 0.036 | 0.256 |
| COGA1_EUR | 0.4048 | 0.077 | 0.093 |
| ESTB2_EUR | 0.0063 | 0.1 | 0.037 |
| GISS1_EUR | 0.7888 | 0.036 | 0.134 |
| GISS2_EUR | 0.0677 | 0.282 | 0.154 |
| GTPRJ_AFR | 0.71 | 0.054 | 0.143 |
| IPSYC_EUR | 0.0776 | 0.052 | 0.03 |
| JANS3_EUR | 0.6483 | −0.077 | 0.17 |
| JANS4_EUR | 0.0913 | 0.322 | 0.19 |
| PGCBD_EUR | 0.0396 | 0.088 | 0.043 |
| PGCED_EUR | 0.9698 | 0.008 | 0.215 |
| PGCMD_EUR | 0.9144 | 0.006 | 0.059 |
| PGCPT_EUR | 0.3866 | 0.078 | 0.09 |
| PGCSZ_EUR | 0.0075 | 0.157 | 0.059 |
| PRFCT_EUR | 0.5306 | −0.045 | 0.072 |
| PSYCR_EUR | 0.9659 | 0.006 | 0.147 |
| QIMRB_EUR | 0.1555 | 0.06 | 0.042 |
| UKBJC_EUR | 0.0046 | 0.083 | 0.029 |
| UTAMR_LAT | 0.3312 | −0.086 | 0.089 |
| YPENN_EUR | 0.9838 | 0.004 | 0.213 |
| **meta** | **4.27e−08** | **0.071** | **0.013** |

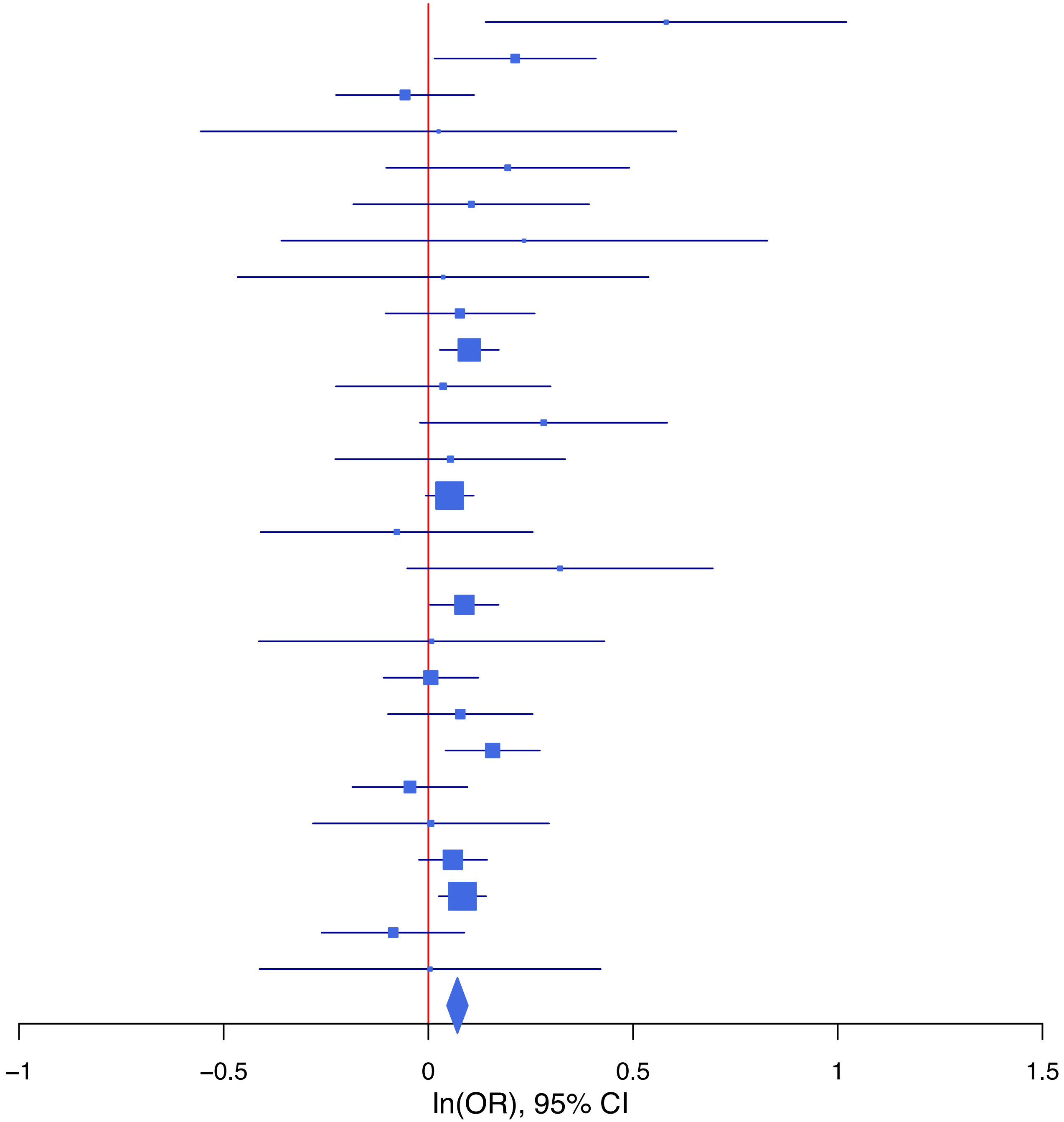

**rs113727800   G/A   9:140258802**

| Cohort | P | ln(OR) | SE |
|---|---|---|---|
| BEPS7_EUR | 0.4563 | 0.205 | 0.275 |
| BHRCM_AFR | 0.2762 | −0.269 | 0.248 |
| BHRCM_EUR | 0.2504 | 0.417 | 0.362 |
| BOR17_EUR | 0.1018 | −0.203 | 0.124 |
| BOR2C_EUR | 0.3952 | −0.113 | 0.133 |
| BOR2E_EUR | 0.3122 | −0.294 | 0.291 |
| CNVRG_EAS | 0.3556 | −0.105 | 0.113 |
| COGA1_AFR | 0.732 | 0.051 | 0.148 |
| COGA1_EUR | 0.231 | −0.11 | 0.091 |
| CUINT_EUR | 0.6597 | 0.058 | 0.133 |
| ESTB2_EUR | 5e−04 | −0.112 | 0.032 |
| FINNG_EUR | 0.1895 | −0.039 | 0.03 |
| GISS1_EUR | 0.1752 | −0.176 | 0.13 |
| GISS2_EUR | 0.5311 | −0.099 | 0.158 |
| GREAT_EAS | 0.6177 | 0.131 | 0.263 |
| GTPRJ_AFR | 0.5121 | −0.055 | 0.085 |
| IPSYC_EUR | 0.6343 | −0.017 | 0.037 |
| JANS3_EUR | 0.1141 | 0.365 | 0.231 |
| JANS4_EUR | 0.6457 | 0.097 | 0.211 |
| JAPAN_EAS | 0.0326 | −0.438 | 0.205 |
| MVPXQ_EUR | 0.0518 | −0.041 | 0.021 |
| MVPXQ_LAT | 0.4025 | −0.063 | 0.075 |
| PGCBD_EUR | 0.9474 | 0.004 | 0.059 |
| PGCMD_EUR | 0.4006 | −0.065 | 0.078 |
| PGCPT_AFR | 0.3334 | 0.14 | 0.145 |
| PGCPT_EUR | 0.3698 | −0.074 | 0.082 |
| PGCSZ_EUR | 0.5948 | 0.048 | 0.091 |
| PRFCT_EUR | 0.8886 | −0.008 | 0.06 |
| PSYCR_EUR | 0.3592 | −0.118 | 0.129 |
| QIMRB_EUR | 0.0261 | −0.094 | 0.042 |
| SNUBH−ASA_EAS | 0.8481 | 0.069 | 0.36 |
| STRR1_LAT | 0.5566 | 0.201 | 0.343 |
| UKBJC_EUR | 5e−04 | −0.102 | 0.029 |
| UTAH2_EUR | 0.2225 | −0.038 | 0.031 |
| YPENN_EUR | 0.4945 | 0.148 | 0.216 |
| **meta** | **3.49e−08** | **−0.055** | **0.01** |

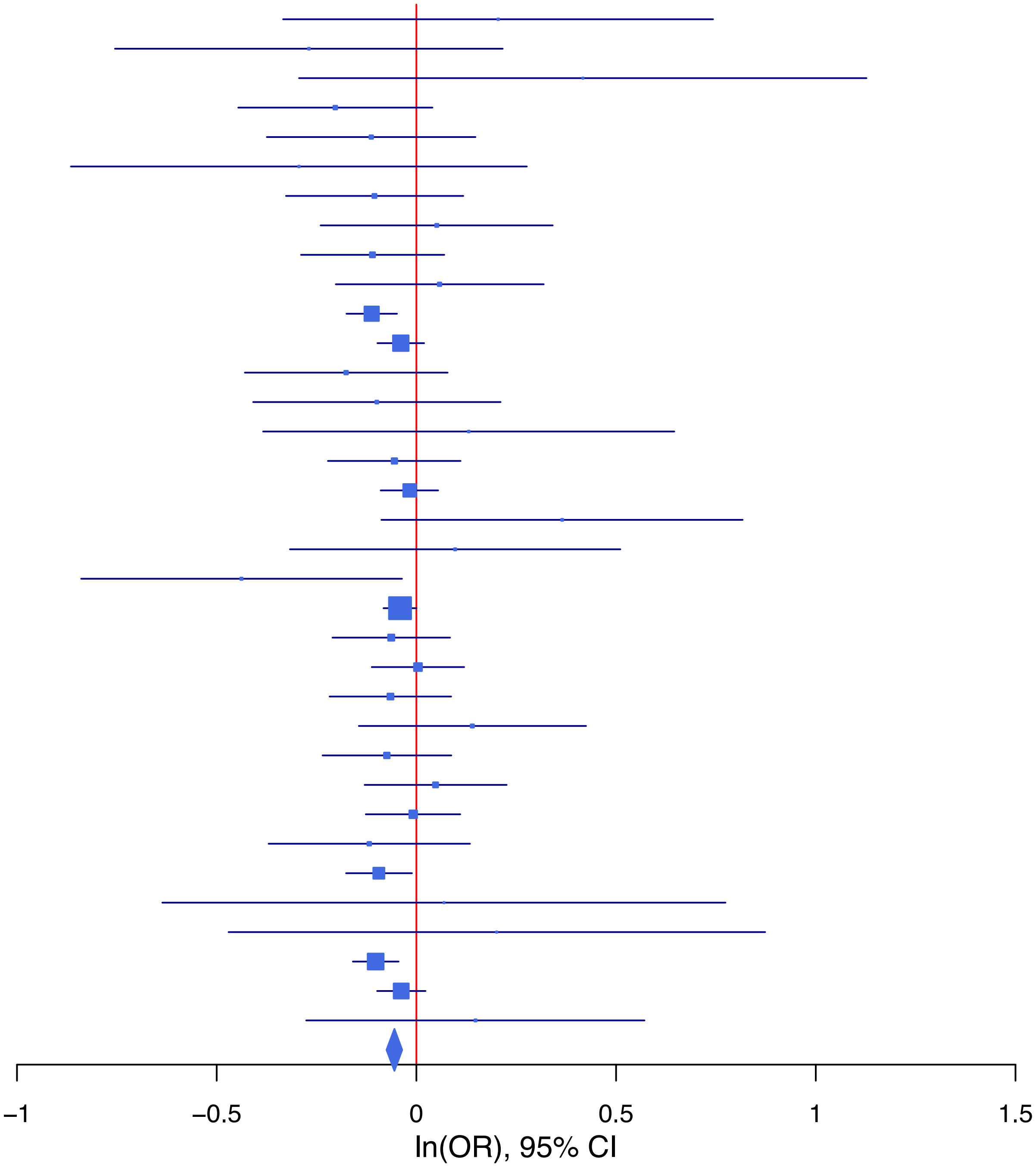

**rs11631591 C/T 15:38850262**

| Cohort | P | ln(OR) | SE |
|---|---|---|---|
| ALSPC_EUR | 0.8522 | 0.011 | 0.061 |
| BEPS7_EUR | 0.218 | 0.244 | 0.198 |
| BHRCM_AFR | 0.9833 | 0.005 | 0.229 |
| BHRCM_EUR | 0.0873 | −0.34 | 0.199 |
| BOR17_EUR | 0.6201 | 0.05 | 0.1 |
| BOR2C_EUR | 0.345 | 0.094 | 0.1 |
| BOR2E_EUR | 0.6728 | 0.087 | 0.206 |
| CNVRG_EAS | 0.4023 | 0.039 | 0.046 |
| COGA1_AFR | 0.1837 | 0.146 | 0.11 |
| COGA1_EUR | 0.5608 | −0.04 | 0.069 |
| CUINT_EUR | 0.463 | 0.063 | 0.086 |
| CVEDA_CSA | 0.7716 | 0.048 | 0.166 |
| ESTB2_EUR | 0.019 | 0.059 | 0.025 |
| FINNG_EUR | 0.3284 | 0.025 | 0.025 |
| GEDIS_EUR | 0.2517 | −0.171 | 0.149 |
| GISS1_EUR | 0.3538 | −0.082 | 0.088 |
| GISS2_EUR | 0.0509 | −0.2 | 0.102 |
| GREAT_EAS | 0.4275 | 0.06 | 0.076 |
| GTPRJ_AFR | 0.7344 | 0.023 | 0.066 |
| IPSYC_EUR | 0.0351 | 0.044 | 0.021 |
| JANS3_EUR | 0.0031 | −0.359 | 0.121 |
| JANS4_EUR | 0.4594 | −0.098 | 0.133 |
| JAPAN_EAS | 0.4131 | 0.046 | 0.056 |
| MVPXQ_AFR | 0.5392 | 0.014 | 0.022 |
| MVPXQ_EAS | 0.2861 | 0.106 | 0.099 |
| MVPXQ_EUR | 0.0582 | 0.026 | 0.014 |
| MVPXQ_LAT | 0.1008 | 0.054 | 0.033 |
| PGCBD_EUR | 0.0755 | 0.052 | 0.029 |
| PGCED_EUR | 0.3 | 0.136 | 0.131 |
| PGCMD_EUR | 0.6719 | 0.016 | 0.039 |
| PGCPT_AFR | 0.7494 | −0.037 | 0.116 |
| PGCPT_EUR | 0.7951 | −0.017 | 0.064 |
| PGCSZ_EUR | 0.4432 | 0.032 | 0.042 |
| PRFCT_EUR | 0.6269 | 0.024 | 0.049 |
| PSYCR_EUR | 0.2672 | 0.105 | 0.094 |
| QIMRB_EUR | 0.2446 | 0.036 | 0.031 |
| SNUBH−ASA_EAS | 0.5083 | 0.081 | 0.123 |
| SNUBH−KCHIP_EAS | 0.7355 | 0.057 | 0.169 |
| STRR1_LAT | 0.8686 | −0.03 | 0.182 |
| UKBJC_EUR | 0.0031 | 0.066 | 0.022 |
| UTAH2_EUR | 0.0197 | 0.049 | 0.021 |
| UTAMR_LAT | 0.3777 | 0.032 | 0.036 |
| YPENN_EUR | 0.0733 | 0.246 | 0.138 |
| **meta** | **1.63e−08** | **0.035** | **0.006** |

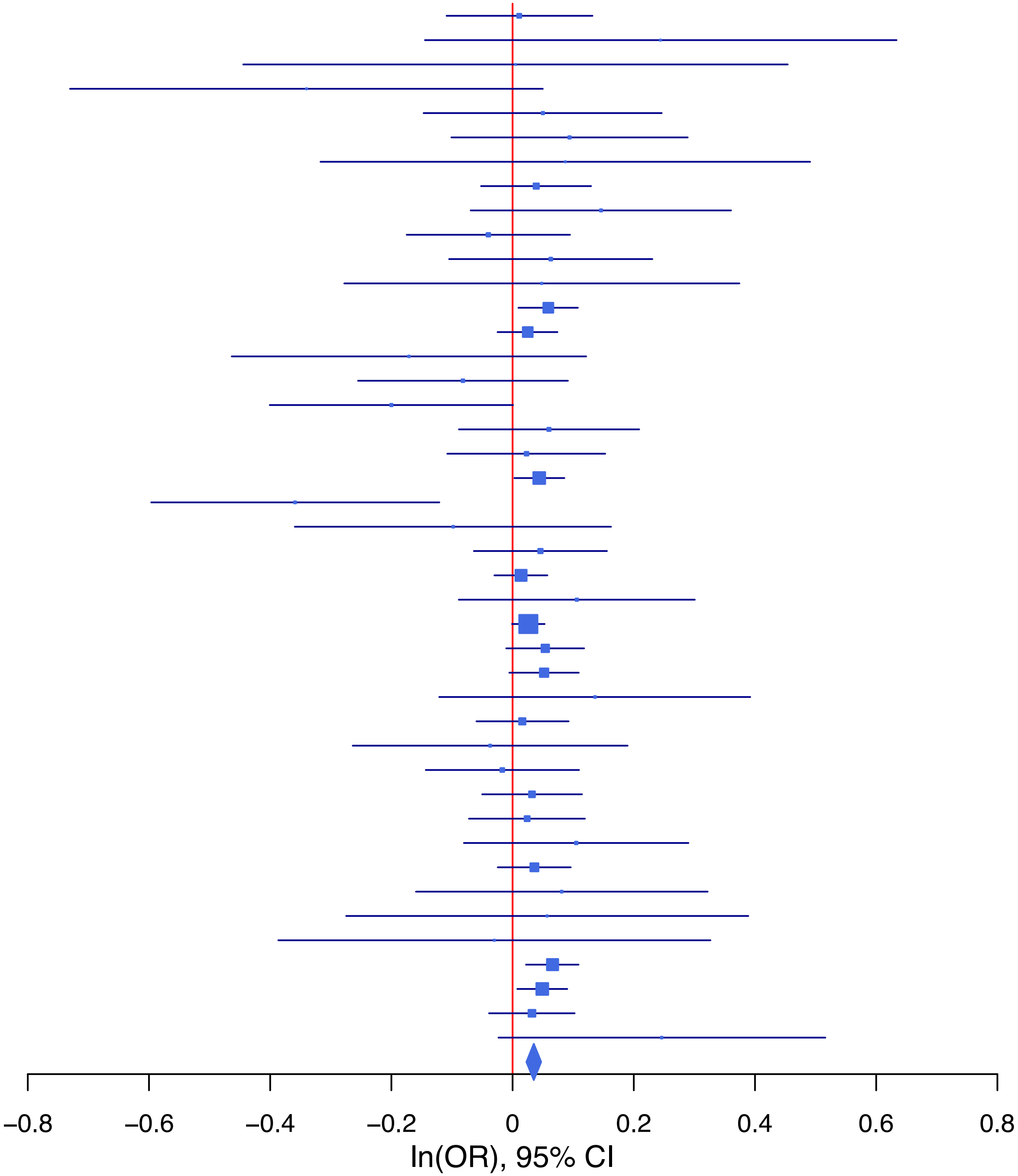

**rs11664320 C/T 18:50872623**

| Cohort | P | ln(OR) | SE |
|---|---|---|---|
| ADHEA_AFR | 0.949 | 0.009 | 0.135 |
| ADHEA_EUR | 0.6664 | −0.03 | 0.069 |
| ALSPC_EUR | 0.5822 | 0.03 | 0.054 |
| BEPS7_EUR | 0.224 | −0.22 | 0.181 |
| BHRCM_AFR | 0.4828 | 0.166 | 0.237 |
| BHRCM_EUR | 0.2295 | 0.184 | 0.153 |
| BOR17_EUR | 0.0021 | 0.257 | 0.084 |
| BOR2C_EUR | 0.0644 | 0.16 | 0.087 |
| BOR2E_EUR | 0.6889 | 0.072 | 0.181 |
| CNVRG_EAS | 0.007 | 0.225 | 0.083 |
| COGA1_AFR | 0.8392 | −0.024 | 0.116 |
| COGA1_EUR | 0.6742 | 0.025 | 0.059 |
| CUINT_EUR | 0.1151 | 0.116 | 0.074 |
| CVEDA_CSA | 0.8197 | 0.048 | 0.209 |
| ESTB2_EUR | 0.056 | 0.043 | 0.023 |
| FINNG_EUR | 0.9381 | 0.002 | 0.023 |
| GEDIS_EUR | 0.1647 | −0.188 | 0.135 |
| GISS1_EUR | 0.1856 | 0.103 | 0.078 |
| GISS2_EUR | 0.553 | −0.055 | 0.092 |
| GREAT_EAS | 0.4545 | −0.118 | 0.158 |
| GTPRJ_AFR | 0.8979 | 0.009 | 0.07 |
| IPSYC_EUR | 0.3317 | 0.018 | 0.018 |
| JANS3_EUR | 0.8628 | 0.018 | 0.105 |
| JAPAN_EAS | 0.446 | 0.095 | 0.124 |
| MIREC_AFR | 0.9008 | 0.02 | 0.16 |
| MIREC_EUR | 0.7129 | 0.056 | 0.154 |
| MVPXQ_AFR | 0.7929 | 0.006 | 0.024 |
| MVPXQ_EUR | 0.0336 | 0.027 | 0.013 |
| MVPXQ_LAT | 0.0766 | 0.066 | 0.037 |
| PGCBD_EUR | 0.572 | 0.015 | 0.026 |
| PGCED_EUR | 0.2447 | 0.132 | 0.114 |
| PGCMD_EUR | 0.1233 | 0.053 | 0.034 |
| PGCPT_AFR | 0.0538 | 0.219 | 0.113 |
| PGCPT_EUR | 0.6152 | 0.029 | 0.058 |
| PGCSZ_EUR | 0.3796 | 0.033 | 0.037 |
| PRFCT_EUR | 0.133 | 0.065 | 0.043 |
| PSYCR_EUR | 0.0933 | 0.142 | 0.085 |
| QIMRB_EUR | 0.5056 | 0.018 | 0.028 |
| SNUBH−ASA_EAS | 0.106 | 0.459 | 0.284 |
| STRR1_LAT | 0.6306 | 0.096 | 0.199 |
| UKBJC_EUR | 3e−04 | 0.07 | 0.02 |
| UTAH2_EUR | 0.0181 | 0.046 | 0.019 |
| UTAMR_LAT | 0.617 | 0.023 | 0.046 |
| YPENN_AFR | 0.6528 | −0.076 | 0.169 |
| YPENN_EUR | 0.1314 | 0.189 | 0.125 |
| **meta** | **1.21e−09** | **0.036** | **0.006** |

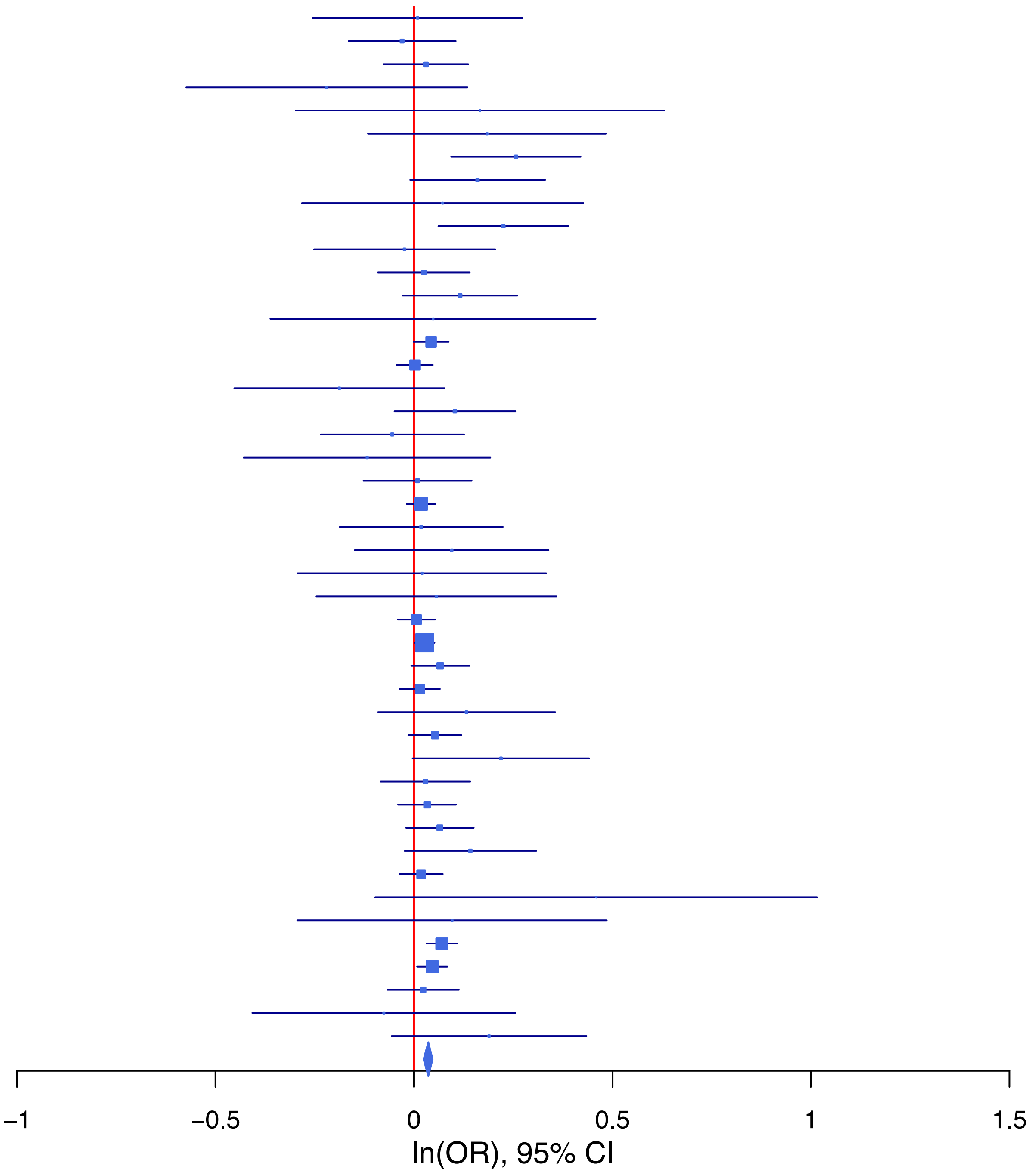

**rs11677638 G/A 2:212697656**

| Cohort | P | ln(OR) | SE |
|---|---|---|---|
| ALSPC_EUR | 0.2872 | −0.064 | 0.06 |
| BEPS7_EUR | 0.2876 | 0.221 | 0.208 |
| BHRCM_EUR | 0.1811 | −0.244 | 0.183 |
| BHRCM_LAT | 0.2578 | −0.364 | 0.322 |
| BOR17_EUR | 0.5403 | −0.058 | 0.094 |
| BOR2C_EUR | 0.9008 | −0.012 | 0.097 |
| BOR2E_EUR | 0.6199 | 0.097 | 0.195 |
| CNVRG_EAS | 0.9629 | 0.002 | 0.045 |
| COGA1_AFR | 0.9395 | 0.014 | 0.184 |
| COGA1_EUR | 0.1788 | −0.086 | 0.064 |
| CUINT_EUR | 0.061 | −0.148 | 0.079 |
| CVEDA_CSA | 0.3477 | −0.183 | 0.195 |
| ESTB2_EUR | 0.0082 | −0.069 | 0.026 |
| FINNG_EUR | 0.9317 | 0.002 | 0.026 |
| GEDIS_EUR | 0.2858 | −0.156 | 0.146 |
| GISS1_EUR | 0.5876 | −0.045 | 0.083 |
| GISS2_EUR | 0.1884 | −0.13 | 0.099 |
| GREAT_EAS | 0.6553 | −0.035 | 0.079 |
| GTPRJ_AFR | 0.0754 | −0.232 | 0.131 |
| JANS3_EUR | 0.4297 | −0.088 | 0.112 |
| JANS4_EUR | 0.8082 | 0.033 | 0.135 |
| JAPAN_EAS | 0.9134 | −0.006 | 0.056 |
| MVPXQ_AFR | 0.13 | −0.066 | 0.044 |
| PGCBD_EUR | 0.6045 | −0.015 | 0.028 |
| PGCED_EUR | 0.3601 | −0.113 | 0.124 |
| PGCMD_EUR | 0.6738 | −0.016 | 0.038 |
| PGCPT_EUR | 0.0121 | 0.155 | 0.062 |
| PGCSZ_EUR | 0.0659 | −0.074 | 0.04 |
| PRFCT_EUR | 0.0148 | −0.126 | 0.052 |
| PSYCR_EUR | 0.2327 | 0.123 | 0.103 |
| QIMRB_EUR | 2e−04 | −0.123 | 0.033 |
| SNUBH−KCHIP_EAS | 0.0938 | −0.286 | 0.171 |
| STRR1_LAT | 0.7079 | 0.068 | 0.183 |
| UKBJC_EUR | 0.0073 | −0.057 | 0.021 |
| UTAH2_EUR | 0.0022 | −0.064 | 0.021 |
| UTAMR_LAT | 0.1109 | −0.059 | 0.037 |
| **meta** | **4.43e−10** | **−0.05** | **0.008** |

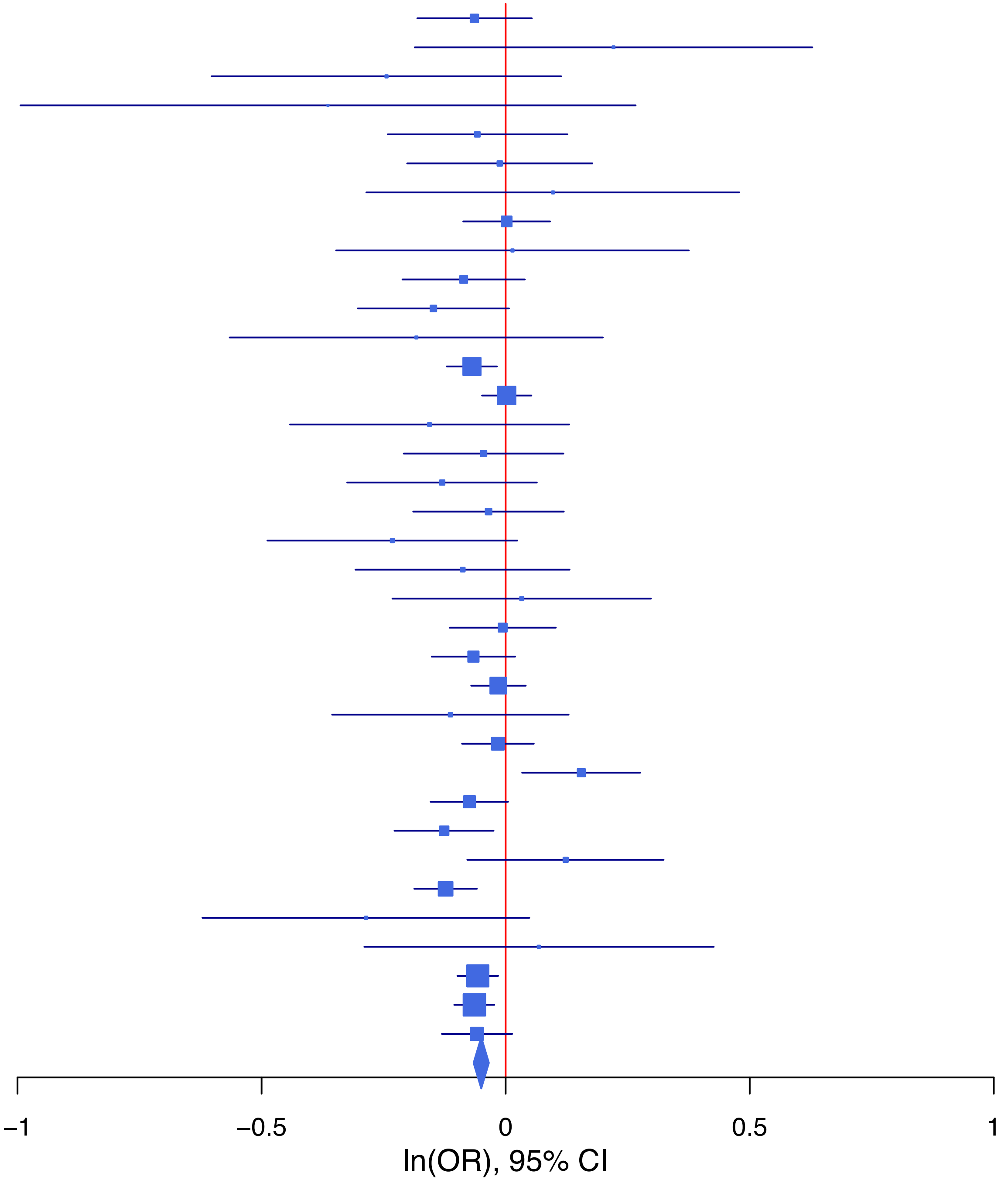

**rs12123415 T/G 1:66496545**

| Cohort | P | ln(OR) | SE |
|---|---|---|---|
| ADHEA_AFR | 0.5285 | 0.072 | 0.114 |
| ADHEA_EUR | 0.8435 | 0.013 | 0.067 |
| ALSPC_EUR | 0.6296 | −0.026 | 0.054 |
| BEPS7_EUR | 0.9317 | 0.016 | 0.182 |
| BHRCM_AFR | 0.9536 | −0.011 | 0.19 |
| BHRCM_EUR | 0.2136 | 0.202 | 0.163 |
| BHRCM_LAT | 0.7188 | 0.116 | 0.322 |
| BOR17_EUR | 0.1309 | −0.127 | 0.084 |
| BOR2C_EUR | 0.054 | 0.165 | 0.086 |
| BOR2E_EUR | 0.6556 | −0.078 | 0.174 |
| CNVRG_EAS | 0.4553 | 0.039 | 0.052 |
| COGA1_AFR | 0.5641 | 0.06 | 0.104 |
| COGA1_EUR | 0.493 | 0.04 | 0.059 |
| CUINT_EUR | 0.8062 | 0.018 | 0.075 |
| CVEDA_CSA | 0.6411 | 0.082 | 0.177 |
| ESTB2_EUR | 0.049 | 0.044 | 0.023 |
| FINNG_EUR | 0.0835 | 0.04 | 0.023 |
| GEDIS_EUR | 0.9837 | 0.003 | 0.126 |
| GISS1_EUR | 0.1855 | −0.103 | 0.078 |
| GISS2_EUR | 0.0836 | −0.158 | 0.091 |
| GREAT_EAS | 0.3399 | 0.091 | 0.095 |
| GTPRJ_AFR | 0.1106 | 0.097 | 0.061 |
| IPSYC_EUR | 2e−04 | 0.069 | 0.018 |
| JANS3_EUR | 0.705 | 0.039 | 0.103 |
| JANS4_EUR | 0.6192 | 0.057 | 0.114 |
| JAPAN_EAS | 0.1208 | 0.102 | 0.066 |
| MIREC_AFR | 0.7748 | −0.042 | 0.145 |
| MIREC_EUR | 0.2994 | −0.157 | 0.151 |
| MVPXQ_EUR | 6e−04 | 0.044 | 0.013 |
| PGCBD_EUR | 0.3162 | 0.026 | 0.025 |
| PGCED_EUR | 0.7339 | 0.039 | 0.115 |
| PGCMD_EUR | 0.3805 | 0.03 | 0.034 |
| PGCPT_AFR | 0.5289 | 0.065 | 0.103 |
| PGCPT_EUR | 0.211 | −0.072 | 0.057 |
| PGCSZ_EUR | 0.2644 | 0.041 | 0.037 |
| PRFCT_EUR | 0.0362 | 0.091 | 0.043 |
| PSYCR_EUR | 0.665 | 0.037 | 0.086 |
| QIMRB_EUR | 0.9192 | 0.003 | 0.027 |
| SNUBH−ASA_EAS | 0.7443 | 0.049 | 0.151 |
| SNUBH−KCHIP_EAS | 0.9421 | 0.015 | 0.201 |
| STRR1_LAT | 0.6374 | −0.085 | 0.181 |
| UKBJC_EUR | 0.005 | 0.055 | 0.02 |
| UTAH2_EUR | 0.0027 | 0.057 | 0.019 |
| UTAMR_LAT | 0.0958 | 0.06 | 0.036 |
| YPENN_AFR | 0.3938 | −0.13 | 0.152 |
| YPENN_EUR | 0.2972 | −0.131 | 0.126 |
| **meta** | **2.15e−12** | **0.042** | **0.006** |

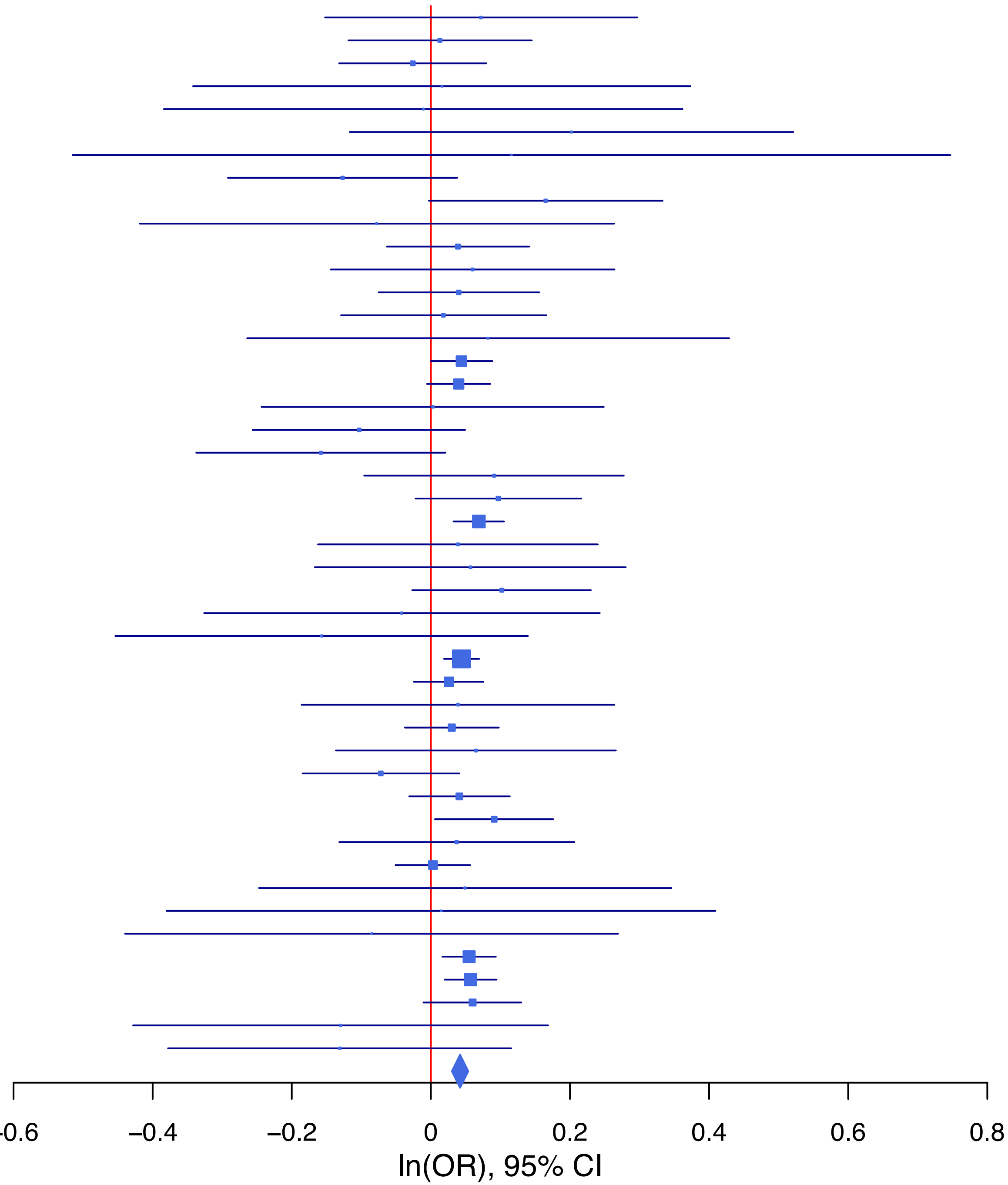

**rs12349408 A/G 9:122662679**

| Cohort | P | ln(OR) | SE |
|---|---|---|---|
| ADHEA_AFR | 0.6709 | −0.051 | 0.121 |
| ADHEA_EUR | 0.2761 | −0.078 | 0.071 |
| ALSPC_EUR | 0.6532 | −0.025 | 0.055 |
| BEPS7_EUR | 0.0247 | −0.433 | 0.193 |
| BHRCM_AFR | 0.6207 | 0.108 | 0.218 |
| BHRCM_EUR | 0.1277 | 0.241 | 0.158 |
| BHRCM_LAT | 0.7182 | 0.098 | 0.272 |
| BOR17_EUR | 0.4362 | 0.07 | 0.09 |
| BOR2C_EUR | 0.7639 | 0.027 | 0.088 |
| BOR2E_EUR | 0.259 | 0.203 | 0.179 |
| CNVRG_EAS | 0.3094 | −0.053 | 0.052 |
| COGA1_AFR | 0.1197 | −0.163 | 0.105 |
| COGA1_EUR | 0.2958 | −0.062 | 0.06 |
| CUINT_EUR | 0.6532 | −0.033 | 0.074 |
| CVEDA_CSA | 0.8886 | −0.022 | 0.159 |
| ESTB2_EUR | 0.07 | −0.042 | 0.023 |
| FINNG_EUR | 0.0235 | −0.053 | 0.024 |
| GEDIS_EUR | 0.51 | 0.088 | 0.134 |
| GISS1_EUR | 0.4948 | −0.055 | 0.08 |
| GISS2_EUR | 0.7495 | 0.032 | 0.099 |
| GTPRJ_AFR | 0.8259 | 0.014 | 0.063 |
| IPSYC_EUR | 2e−04 | −0.071 | 0.019 |
| JANS3_EUR | 0.2424 | −0.128 | 0.109 |
| JANS4_EUR | 0.9021 | −0.015 | 0.123 |
| JAPAN_EAS | 0.0014 | −0.195 | 0.061 |
| MVPXQ_AFR | 0.0471 | −0.043 | 0.022 |
| MVPXQ_EAS | 0.0551 | −0.201 | 0.105 |
| MVPXQ_EUR | 0.0152 | −0.031 | 0.013 |
| MVPXQ_LAT | 0.037 | −0.068 | 0.033 |
| PGCBD_EUR | 0.0547 | −0.051 | 0.027 |
| PGCED_EUR | 0.8268 | −0.026 | 0.12 |
| PGCMD_EUR | 0.0528 | −0.068 | 0.035 |
| PGCPT_AFR | 0.7798 | 0.03 | 0.107 |
| PGCPT_EUR | 0.3578 | −0.054 | 0.059 |
| PGCSZ_EUR | 0.9939 | 0 | 0.037 |
| PRFCT_EUR | 0.4005 | −0.038 | 0.045 |
| PSYCR_EUR | 0.9401 | 0.007 | 0.09 |
| QIMRB_EUR | 0.5061 | −0.019 | 0.028 |
| SNUBH-ASA_EAS | 0.3323 | −0.138 | 0.143 |
| STRR1_LAT | 0.9733 | 0.006 | 0.176 |
| UKBJC_EUR | 0.0857 | −0.034 | 0.02 |
| UTAH2_EUR | 0.9229 | −0.002 | 0.019 |
| UTAMR_LAT | 0.3189 | 0.035 | 0.035 |
| YPENN_EUR | 0.7445 | −0.042 | 0.13 |
| **meta** | **1.16e−10** | **−0.037** | **0.006** |

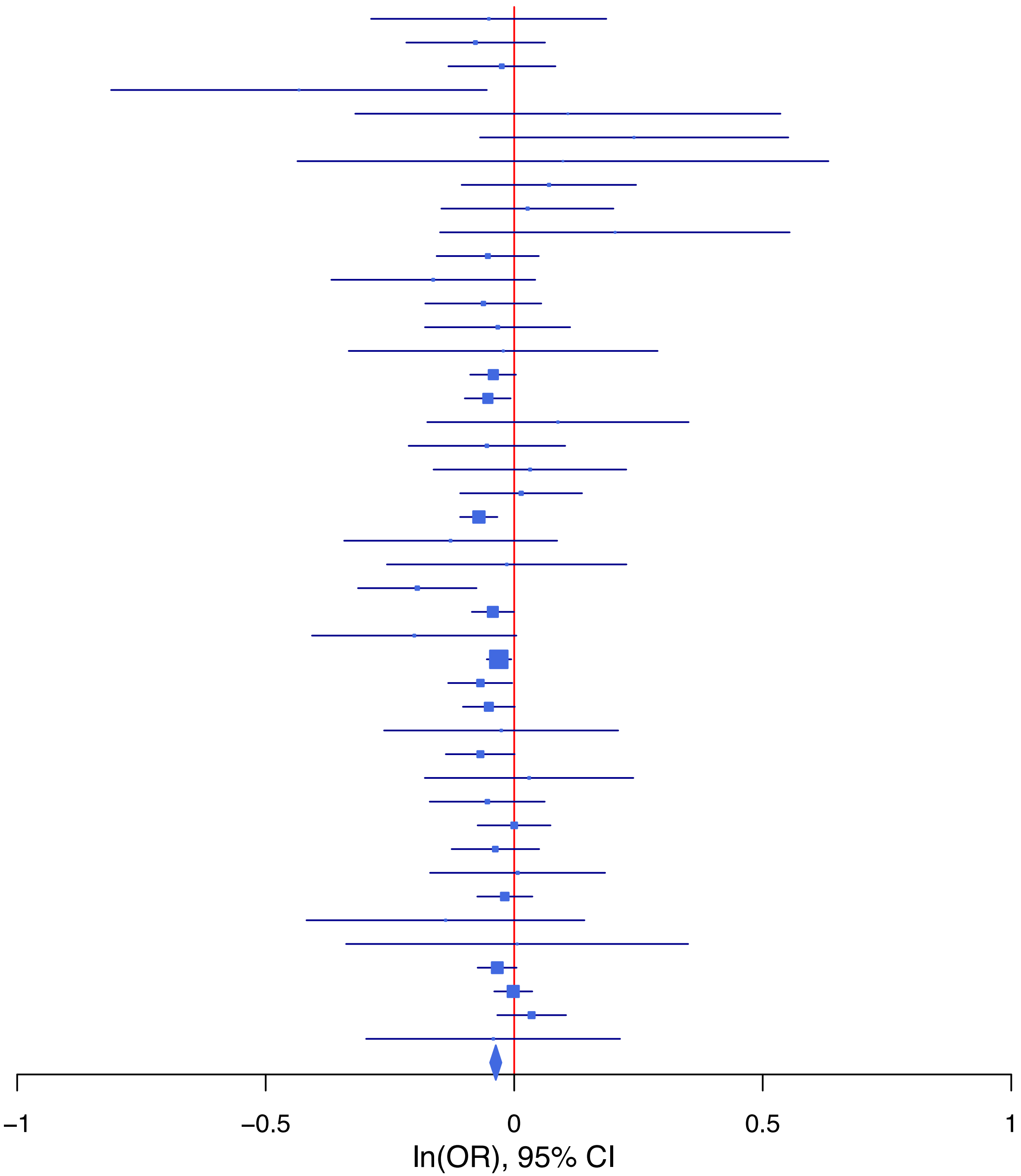

**rs12364051   A/G   11:113305314**

| Cohort | P | ln(OR) | SE |
|---|---|---|---|
| ALSPC_EUR | 0.7221 | 0.019 | 0.055 |
| BEPS7_EUR | 0.9292 | 0.016 | 0.181 |
| BHRCM_AFR | 0.9903 | −0.003 | 0.253 |
| BHRCM_EUR | 0.9317 | −0.015 | 0.174 |
| BOR17_EUR | 0.1551 | −0.126 | 0.089 |
| BOR2C_EUR | 0.4374 | 0.07 | 0.09 |
| BOR2E_EUR | 6e−04 | −0.69 | 0.201 |
| CNVRG_EAS | 0.5622 | 0.058 | 0.101 |
| COGA1_AFR | 0.3731 | −0.107 | 0.12 |
| COGA1_EUR | 0.6781 | −0.025 | 0.06 |
| CUINT_EUR | 0.6935 | 0.031 | 0.078 |
| CVEDA_CSA | 0.2417 | 0.206 | 0.176 |
| ESTB2_EUR | 0.0046 | 0.064 | 0.023 |
| FINNG_EUR | 0.0736 | 0.041 | 0.023 |
| GEDIS_EUR | 0.0381 | 0.27 | 0.13 |
| GISS2_EUR | 0.1754 | −0.125 | 0.092 |
| GREAT_EAS | 0.5454 | 0.125 | 0.206 |
| GTPRJ_AFR | 0.8475 | 0.014 | 0.075 |
| IPSYC_EUR | 0.0097 | 0.049 | 0.019 |
| JANS3_EUR | 0.1471 | 0.15 | 0.104 |
| JANS4_EUR | 0.0961 | −0.201 | 0.12 |
| MVPXQ_AFR | 0.0089 | 0.067 | 0.025 |
| MVPXQ_EUR | 1e−04 | 0.051 | 0.013 |
| MVPXQ_LAT | 0.0454 | 0.069 | 0.034 |
| PGCBD_EUR | 0.4513 | 0.02 | 0.026 |
| PGCED_EUR | 0.3426 | 0.111 | 0.117 |
| PGCMD_EUR | 0.097 | 0.058 | 0.035 |
| PGCPT_AFR | 0.4398 | 0.111 | 0.143 |
| PGCPT_EUR | 0.5193 | −0.038 | 0.059 |
| PGCSZ_EUR | 0.2005 | 0.048 | 0.038 |
| PRFCT_EUR | 0.5259 | 0.028 | 0.044 |
| PSYCR_EUR | 0.9296 | −0.008 | 0.088 |
| QIMRB_EUR | 0.2203 | 0.034 | 0.028 |
| SNUBH-ASA_EAS | 0.3616 | −0.247 | 0.271 |
| STRR1_LAT | 0.496 | −0.127 | 0.186 |
| UKBJC_EUR | 0.1078 | 0.032 | 0.02 |
| UTAH2_EUR | 0.0686 | 0.035 | 0.019 |
| UTAMR_LAT | 0.1324 | 0.062 | 0.041 |
| **meta** | **2.01e−12** | **0.042** | **0.006** |

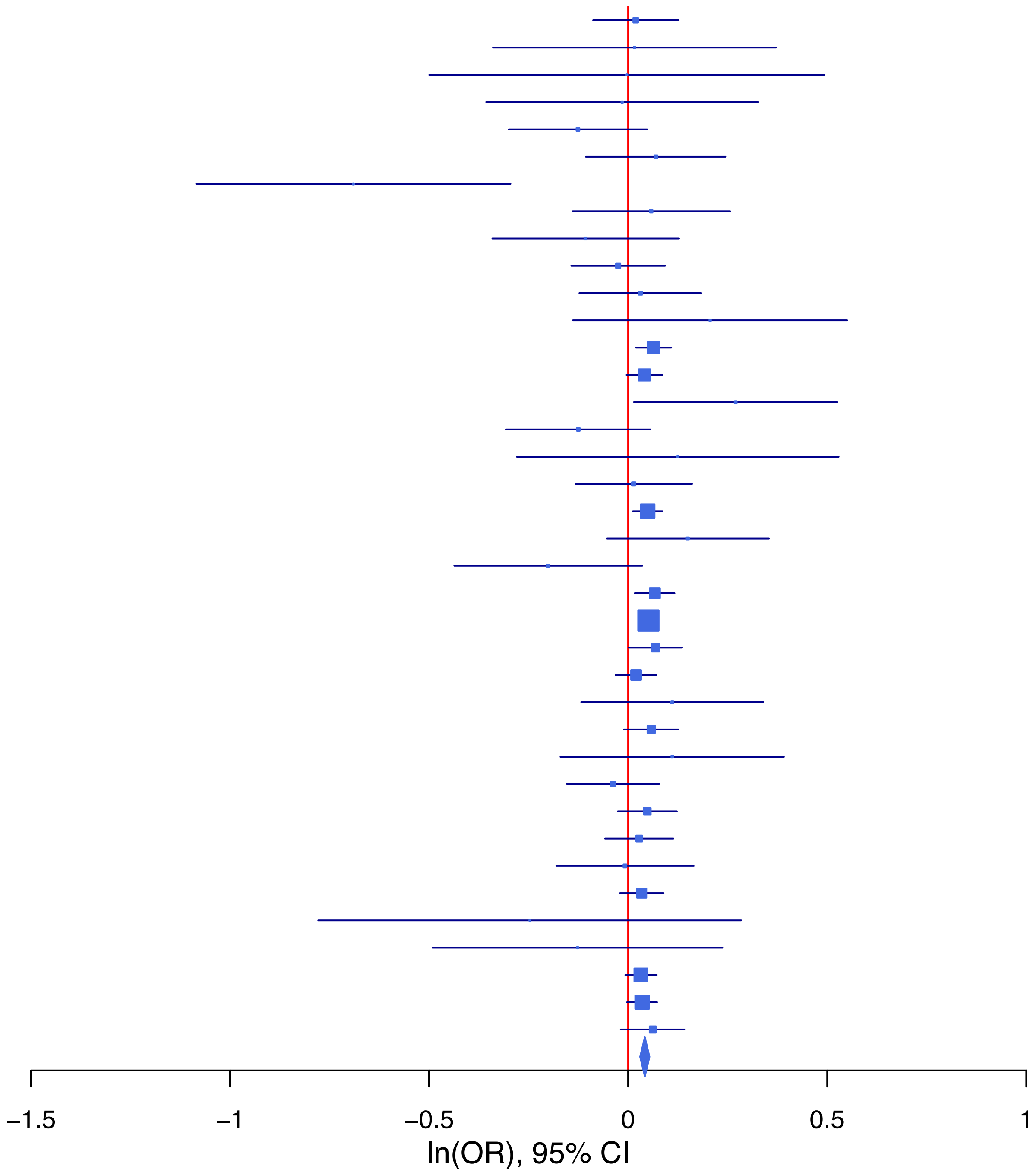

**rs12453624   A/G   17:27426153**

| Cohort | P | ln(OR) | SE |
|---|---|---|---|
| ALSPC_EUR | 0.6734 | 0.024 | 0.056 |
| BEPS7_EUR | 0.8028 | 0.049 | 0.194 |
| BHRCM_AFR | 0.9745 | −0.007 | 0.204 |
| BHRCM_EUR | 0.6113 | 0.081 | 0.159 |
| BHRCM_LAT | 0.7257 | −0.109 | 0.312 |
| BOR17_EUR | 0.4605 | 0.067 | 0.091 |
| BOR2C_EUR | 0.5768 | 0.051 | 0.092 |
| BOR2E_EUR | 0.7724 | −0.054 | 0.186 |
| CNVRG_EAS | 0.0901 | 0.097 | 0.057 |
| COGA1_AFR | 0.9137 | 0.011 | 0.103 |
| COGA1_EUR | 0.2762 | 0.067 | 0.061 |
| CUINT_EUR | 0.1329 | 0.118 | 0.079 |
| CVEDA_CSA | 0.1485 | 0.235 | 0.163 |
| ESTB2_EUR | 0.0324 | 0.052 | 0.025 |
| FINNG_EUR | 0.0427 | 0.049 | 0.024 |
| GEDIS_EUR | 0.1773 | 0.181 | 0.135 |
| GISS1_EUR | 0.7991 | 0.022 | 0.085 |
| GISS2_EUR | 0.299 | 0.104 | 0.1 |
| GREAT_EAS | 0.9924 | 0.001 | 0.125 |
| GTPRJ_AFR | 0.4387 | 0.047 | 0.06 |
| IPSYC_EUR | 0.0042 | 0.056 | 0.02 |
| JANS3_EUR | 0.4492 | 0.082 | 0.108 |
| JANS4_EUR | 0.4648 | −0.09 | 0.124 |
| MIREC_AFR | 0.045 | 0.29 | 0.144 |
| MIREC_EUR | 0.3624 | 0.141 | 0.154 |
| MVPXQ_EUR | 0.0732 | 0.023 | 0.013 |
| PGCBD_EUR | 0.003 | 0.081 | 0.027 |
| PGCED_EUR | 0.8302 | 0.026 | 0.119 |
| PGCMD_EUR | 0.1354 | 0.054 | 0.036 |
| PGCPT_EUR | 0.9285 | −0.005 | 0.061 |
| PGCSZ_EUR | 0.2833 | 0.042 | 0.039 |
| PRFCT_EUR | 0.9351 | −0.004 | 0.046 |
| PSYCR_EUR | 0.0583 | 0.172 | 0.091 |
| QIMRB_EUR | 0.1054 | 0.046 | 0.029 |
| SNUBH−KCHIP_EAS | 0.4476 | −0.159 | 0.209 |
| STRR1_LAT | 0.5309 | −0.117 | 0.187 |
| UKBJC_EUR | 0.8088 | −0.005 | 0.021 |
| UTAH2_EUR | 0.1417 | 0.029 | 0.02 |
| UTAMR_LAT | 0.9616 | −0.002 | 0.037 |
| YPENN_AFR | 0.7769 | −0.043 | 0.152 |
| YPENN_EUR | 0.0482 | −0.287 | 0.145 |
| **meta** | **1.06e−08** | **0.036** | **0.006** |

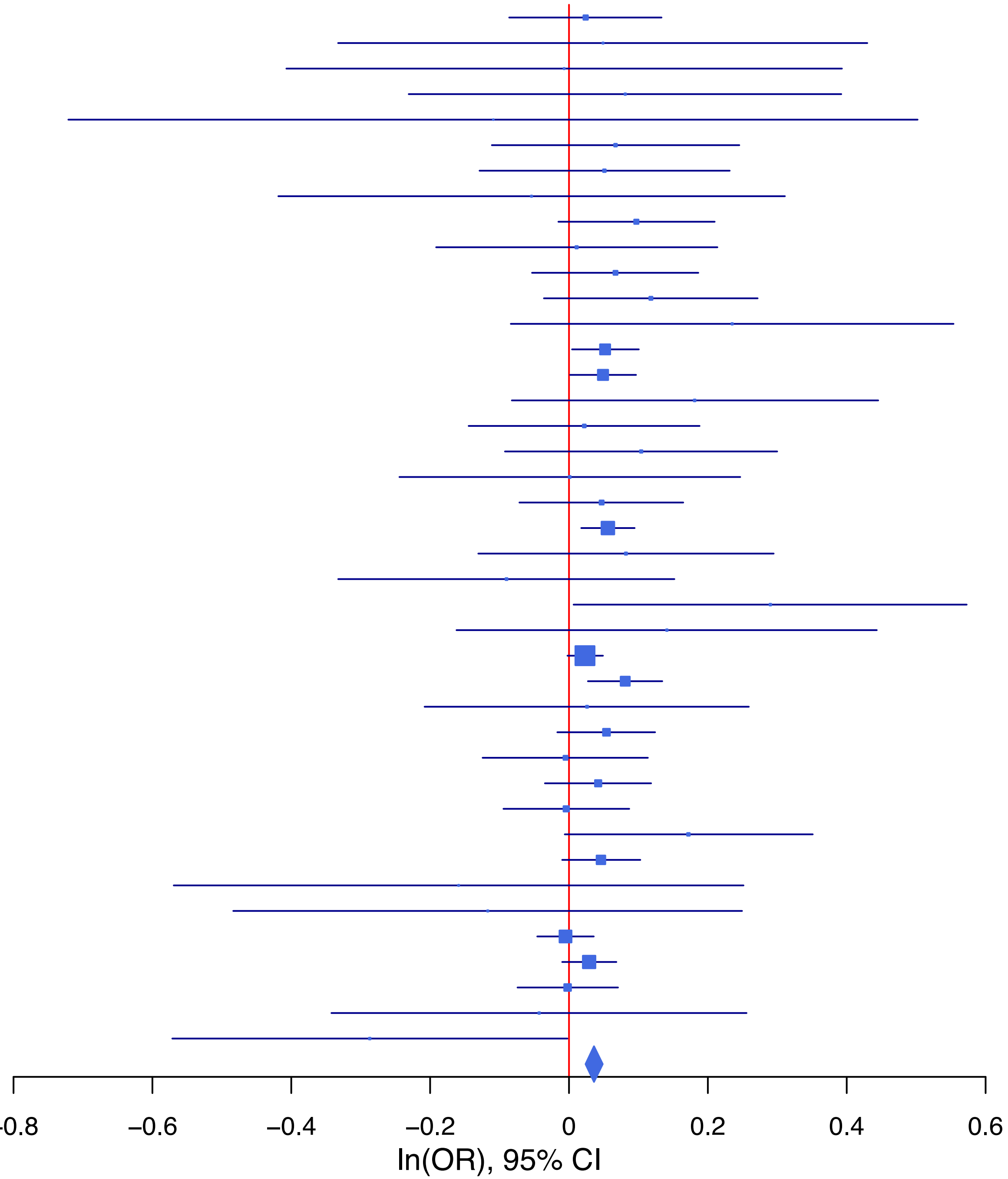

**rs12665582 C/T 6:156439031**

| Cohort | P | ln(OR) | SE |
|---|---|---|---|
| ADHEA_AFR | 0.7666 | 0.039 | 0.132 |
| ADHEA_EUR | 0.8029 | −0.028 | 0.113 |
| ALSPC_EUR | 0.6536 | 0.04 | 0.088 |
| BEPS7_EUR | 0.9973 | −0.001 | 0.298 |
| BHRCM_AFR | 0.4573 | 0.153 | 0.206 |
| BHRCM_EUR | 0.3352 | 0.223 | 0.232 |
| BOR17_EUR | 0.1536 | −0.213 | 0.149 |
| BOR2C_EUR | 0.9907 | 0.002 | 0.146 |
| BOR2E_EUR | 0.5525 | −0.178 | 0.3 |
| CNVRG_EAS | 0.8326 | 0.01 | 0.048 |
| COGA1_AFR | 0.2202 | 0.145 | 0.118 |
| COGA1_EUR | 0.0608 | 0.179 | 0.096 |
| CUINT_EUR | 0.1326 | 0.184 | 0.122 |
| CVEDA_CSA | 0.1984 | −0.378 | 0.294 |
| ESTB2_EUR | 0.5547 | 0.025 | 0.043 |
| FINNG_EUR | 0.0956 | 0.081 | 0.049 |
| GEDIS_EUR | 0.2996 | −0.256 | 0.247 |
| GISS1_EUR | 0.096 | −0.214 | 0.129 |
| GISS2_EUR | 0.7238 | −0.055 | 0.155 |
| GREAT_EAS | 0.2851 | 0.084 | 0.079 |
| GTPRJ_AFR | 0.5987 | 0.035 | 0.068 |
| IPSYC_EUR | 0.1471 | 0.045 | 0.031 |
| JANS3_EUR | 0.0457 | 0.318 | 0.159 |
| JANS4_EUR | 0.969 | −0.008 | 0.197 |
| JAPAN_EAS | 0.6985 | −0.023 | 0.06 |
| MVPXQ_EUR | 0.0032 | 0.067 | 0.023 |
| PGCBD_EUR | 4.70e−05 | 0.177 | 0.044 |
| PGCED_EUR | 0.4966 | 0.137 | 0.201 |
| PGCMD_EUR | 0.7051 | 0.024 | 0.062 |
| PGCPT_AFR | 0.2423 | 0.138 | 0.118 |
| PGCPT_EUR | 0.4921 | 0.065 | 0.094 |
| PGCSZ_EUR | 0.8006 | 0.016 | 0.064 |
| PRFCT_EUR | 0.0311 | 0.159 | 0.074 |
| PSYCR_EUR | 0.6277 | −0.071 | 0.146 |
| QIMRB_EUR | 0.4203 | 0.036 | 0.045 |
| SNUBH−ASA_EAS | 0.9167 | −0.014 | 0.133 |
| SNUBH−KCHIP_EAS | 0.2876 | 0.189 | 0.178 |
| STRR1_LAT | 0.3966 | −0.183 | 0.216 |
| UKBJC_EUR | 0.001 | 0.108 | 0.033 |
| UTAH2_EUR | 0.4604 | 0.024 | 0.032 |
| UTAMR_LAT | 0.1065 | 0.062 | 0.038 |
| YPENN_AFR | 0.2423 | 0.192 | 0.164 |
| YPENN_EUR | 0.1783 | 0.263 | 0.196 |
| **meta** | **1.28e−09** | **0.059** | **0.01** |

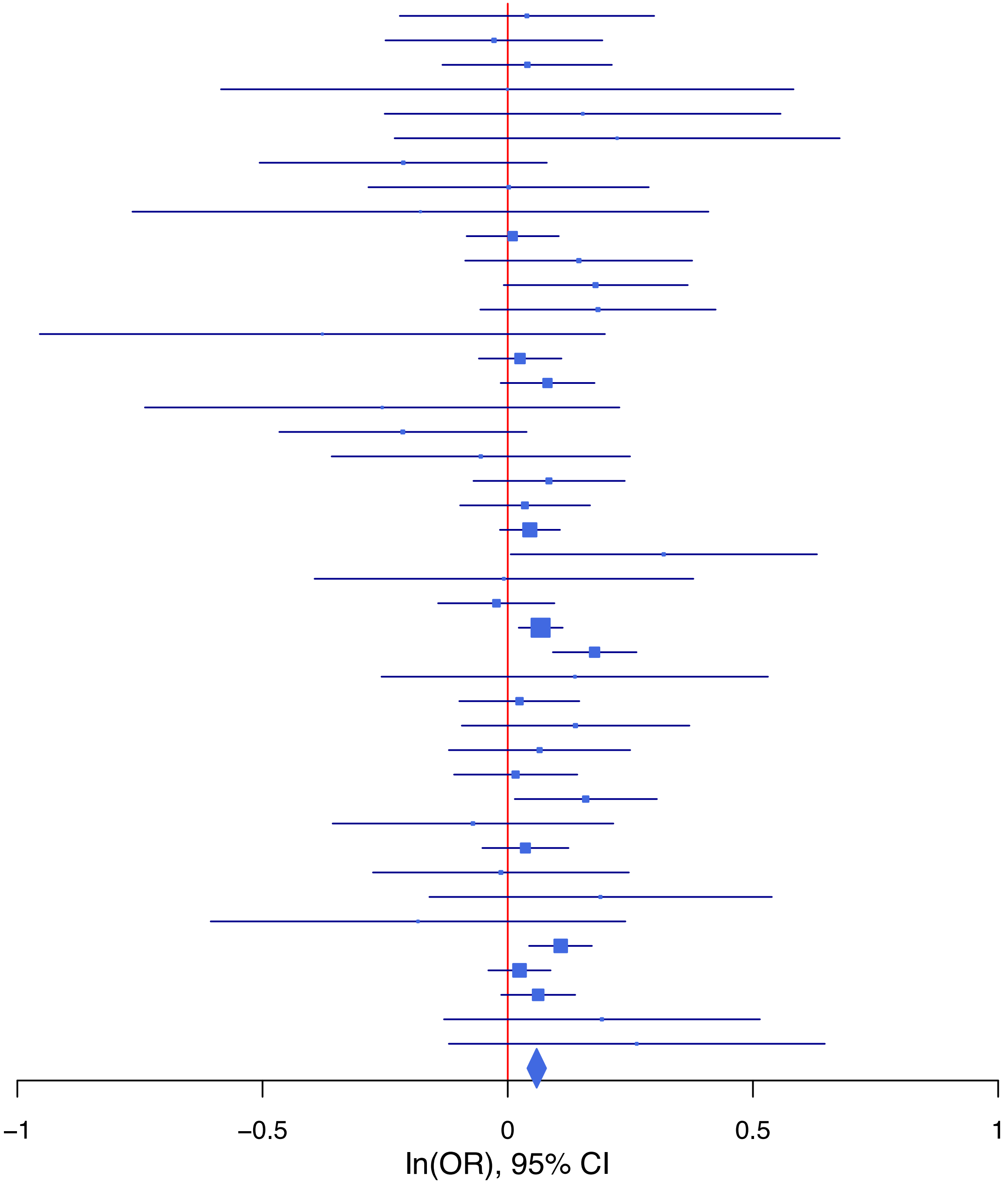

**rs1267063 A/T 2:162002267**

| Cohort | P | ln(OR) | SE |
|---|---|---|---|
| ADHEA_AFR | 0.4216 | 0.128 | 0.16 |
| ALSPC_EUR | 0.0775 | −0.107 | 0.06 |
| BEPS7_EUR | 0.6567 | −0.088 | 0.197 |
| BHRCM_AFR | 0.3744 | −0.262 | 0.295 |
| BOR17_EUR | 0.1673 | −0.126 | 0.091 |
| BOR2C_EUR | 0.5734 | 0.052 | 0.092 |
| BOR2E_EUR | 0.2655 | −0.236 | 0.212 |
| CNVRG_EAS | 0.67 | −0.02 | 0.048 |
| COGA1_AFR | 0.9117 | 0.016 | 0.149 |
| COGA1_EUR | 0.0237 | −0.145 | 0.064 |
| CUINT_EUR | 0.054 | −0.159 | 0.083 |
| CVEDA_CSA | 0.2884 | −0.204 | 0.193 |
| ESTB2_EUR | 0.238 | −0.029 | 0.024 |
| FINNG_EUR | 0.182 | −0.035 | 0.026 |
| GEDIS_EUR | 0.2978 | −0.154 | 0.148 |
| GISS1_EUR | 0.1967 | −0.108 | 0.083 |
| GISS2_EUR | 0.6134 | −0.049 | 0.097 |
| GTPRJ_AFR | 0.8437 | 0.017 | 0.087 |
| IPSYC_EUR | 0.1566 | −0.029 | 0.02 |
| JANS3_EUR | 0.0817 | −0.208 | 0.12 |
| JANS4_EUR | 0.1034 | −0.216 | 0.132 |
| JAPAN_EAS | 0.3799 | −0.055 | 0.062 |
| MIREC_AFR | 0.402 | −0.174 | 0.208 |
| MIREC_EUR | 0.2344 | 0.188 | 0.158 |
| MVPXQ_EUR | 0.1499 | −0.02 | 0.014 |
| MVPXQ_LAT | 0.2747 | −0.045 | 0.042 |
| PGCBD_EUR | 0.0248 | −0.064 | 0.028 |
| PGCED_EUR | 0.5436 | −0.075 | 0.123 |
| PGCMD_EUR | 0.963 | 0.002 | 0.037 |
| PGCPT_AFR | 0.3251 | 0.139 | 0.141 |
| PGCPT_EUR | 0.5879 | −0.034 | 0.064 |
| PGCSZ_EUR | 0.3295 | −0.04 | 0.041 |
| PRFCT_EUR | 0.8786 | 0.007 | 0.049 |
| PSYCR_EUR | 0.5303 | −0.059 | 0.094 |
| QIMRB_EUR | 0.0559 | −0.057 | 0.03 |
| SNUBH-ASA_EAS | 0.0211 | −0.308 | 0.134 |
| SNUBH-KCHIP_EAS | 0.1881 | 0.242 | 0.184 |
| STRR1_LAT | 0.5317 | −0.137 | 0.22 |
| UKBJC_EUR | 0.1941 | −0.028 | 0.021 |
| UTAH2_EUR | 0.0907 | −0.036 | 0.021 |
| UTAMR_LAT | 0.2534 | −0.054 | 0.047 |
| YPENN_AFR | 0.4577 | 0.147 | 0.198 |
| YPENN_EUR | 0.2831 | −0.152 | 0.142 |
| **meta** | **4.55e−08** | **−0.036** | **0.007** |

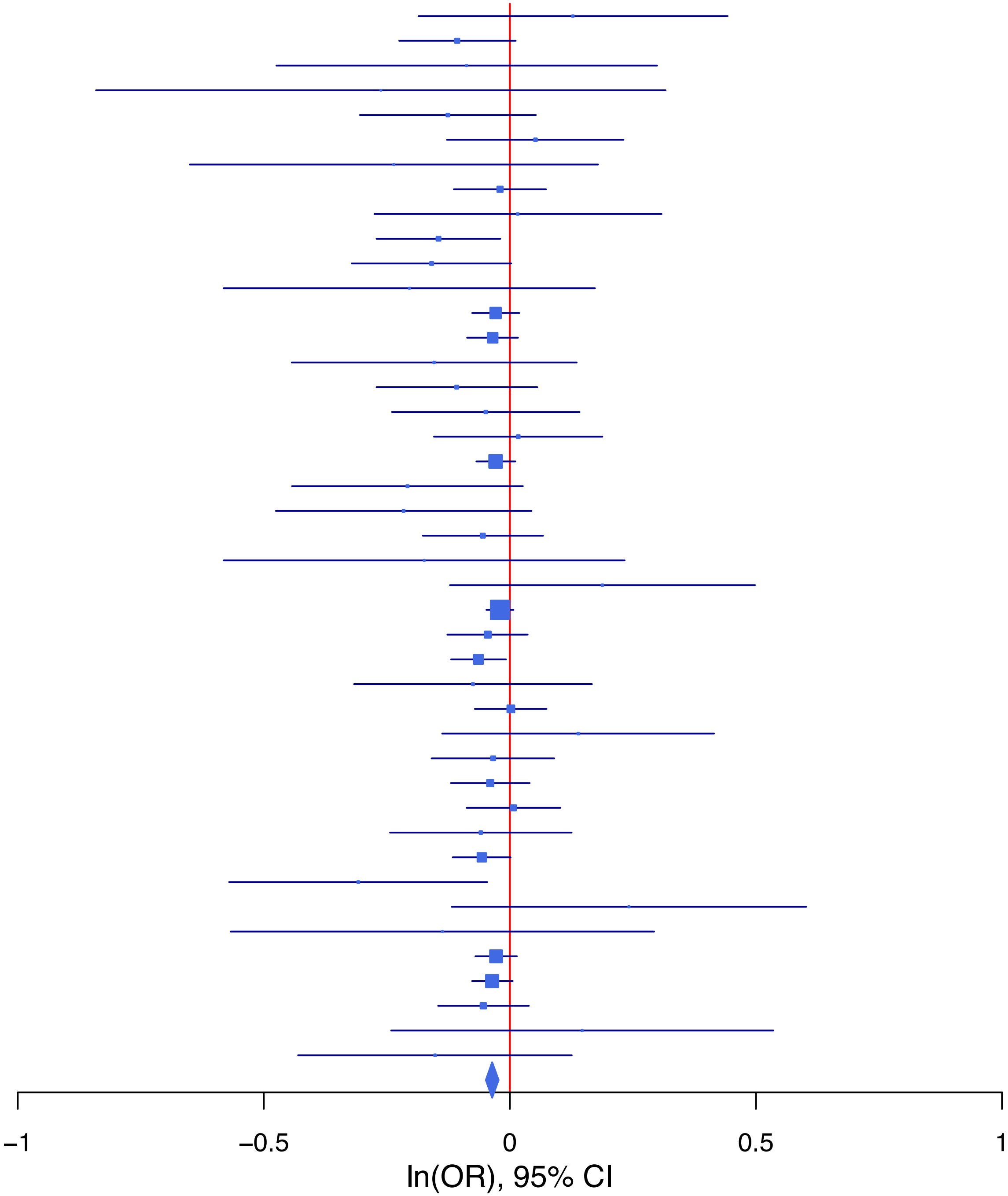

**rs13078566 A/G 3:56394837**

| Cohort | P | ln(OR) | SE |
|---|---|---|---|
| ADHEA_AFR | 0.2741 | 0.186 | 0.17 |
| ALSPC_EUR | 0.0649 | 0.107 | 0.058 |
| BEPS7_EUR | 0.9875 | −0.003 | 0.202 |
| BHRCM_AFR | 0.103 | 0.563 | 0.345 |
| BHRCM_EUR | 0.2882 | −0.215 | 0.203 |
| BHRCM_LAT | 0.6737 | 0.124 | 0.295 |
| BOR17_EUR | 0.117 | 0.151 | 0.096 |
| BOR2C_EUR | 0.7063 | −0.037 | 0.098 |
| BOR2E_EUR | 0.1054 | 0.32 | 0.198 |
| CNVRG_EAS | 0.6786 | −0.031 | 0.074 |
| COGA1_AFR | 0.3794 | 0.134 | 0.153 |
| COGA1_EUR | 0.7968 | 0.016 | 0.063 |
| CUINT_EUR | 0.772 | −0.025 | 0.085 |
| ESTB2_EUR | 0.1952 | 0.033 | 0.025 |
| FINNG_EUR | 0.3055 | 0.025 | 0.025 |
| GEDIS_EUR | 0.1686 | 0.191 | 0.138 |
| GISS1_EUR | 0.7553 | 0.028 | 0.089 |
| GISS2_EUR | 0.9512 | −0.006 | 0.106 |
| GREAT_EAS | 0.5753 | 0.059 | 0.106 |
| GTPRJ_AFR | 0.2034 | 0.119 | 0.093 |
| IPSYC_EUR | 0.0116 | 0.049 | 0.02 |
| JANS3_EUR | 0.7401 | −0.038 | 0.116 |
| JANS4_EUR | 0.6856 | 0.053 | 0.13 |
| JAPAN_EAS | 0.2618 | −0.108 | 0.097 |
| MIREC_AFR | 0.2836 | −0.26 | 0.242 |
| MIREC_EUR | 0.1078 | −0.282 | 0.175 |
| MVPXQ_AFR | 0.8989 | 0.004 | 0.034 |
| MVPXQ_EAS | 0.8485 | −0.027 | 0.144 |
| MVPXQ_EUR | 0.0274 | 0.03 | 0.014 |
| MVPXQ_LAT | 0.3051 | −0.037 | 0.036 |
| PGCBD_EUR | 0.1555 | 0.04 | 0.028 |
| PGCED_EUR | 0.7616 | −0.039 | 0.129 |
| PGCMD_EUR | 0.1155 | 0.059 | 0.037 |
| PGCPT_EUR | 0.839 | −0.013 | 0.063 |
| PGCSZ_EUR | 0.5565 | 0.024 | 0.041 |
| PRFCT_EUR | 0.1678 | 0.065 | 0.047 |
| PSYCR_EUR | 0.5444 | −0.059 | 0.097 |
| QIMRB_EUR | 0.0294 | 0.066 | 0.03 |
| SNUBH−ASA_EAS | 0.6034 | −0.113 | 0.218 |
| SNUBH−KCHIP_EAS | 0.8564 | 0.053 | 0.294 |
| STRR1_LAT | 0.6962 | 0.076 | 0.195 |
| UKBJC_EUR | 0.0105 | 0.054 | 0.021 |
| UTAH2_EUR | 0.0225 | 0.047 | 0.02 |
| YPENN_AFR | 0.9536 | 0.014 | 0.233 |
| YPENN_EUR | 0.7536 | −0.045 | 0.144 |
| **meta** | **4.32e−08** | **0.035** | **0.006** |

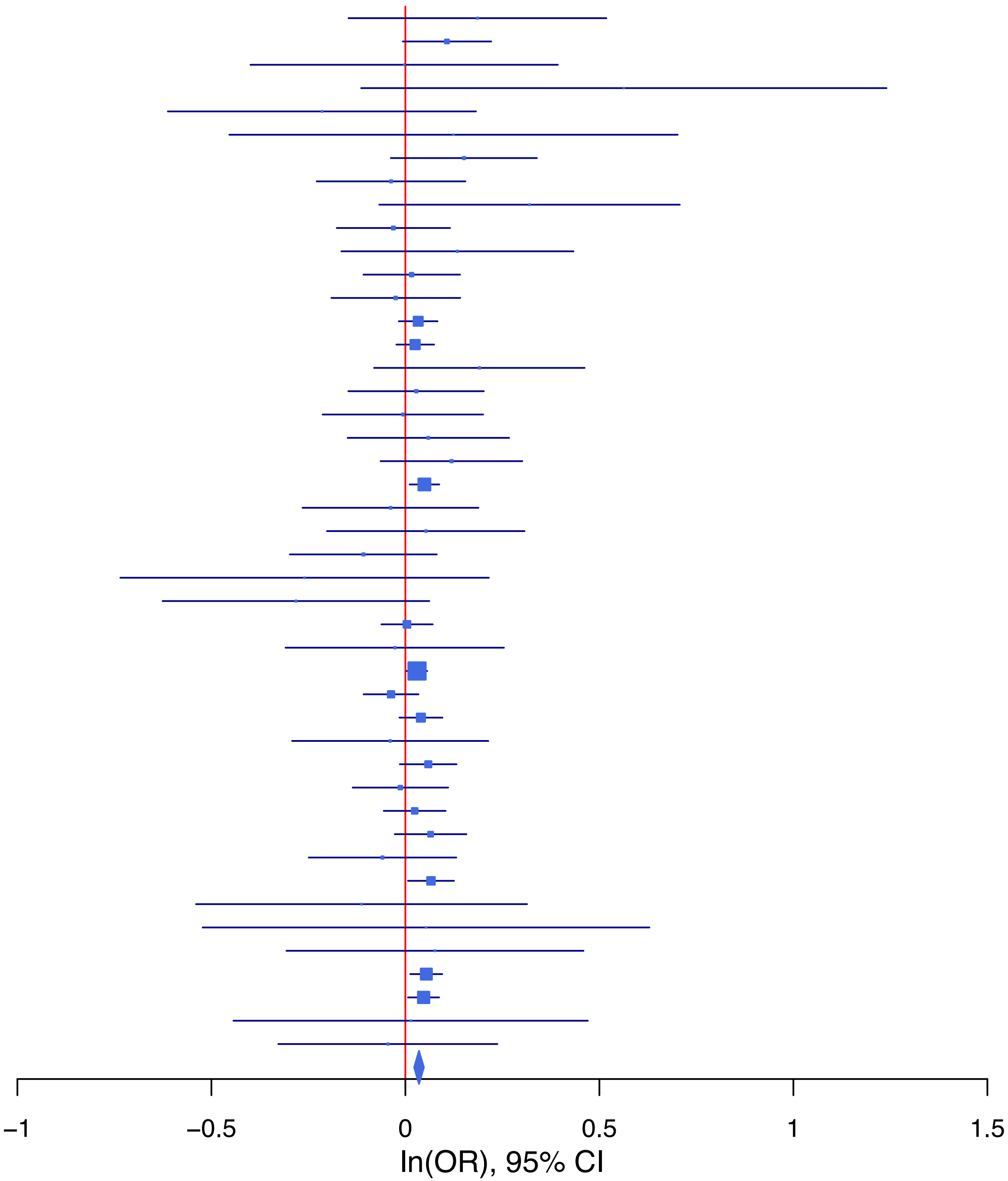

**rs13409451 G/A 2:144257639**

| Cohort | P | ln(OR) | SE |
|---|---|---|---|
| ALSPC_EUR | 0.5179 | 0.035 | 0.055 |
| BEPS7_EUR | 0.5857 | 0.102 | 0.186 |
| BHRCM_AFR | 0.0104 | 0.711 | 0.277 |
| BHRCM_EUR | 0.799 | −0.042 | 0.164 |
| BOR17_EUR | 0.2649 | −0.098 | 0.088 |
| BOR2C_EUR | 0.3117 | −0.087 | 0.086 |
| BOR2E_EUR | 0.7528 | −0.058 | 0.184 |
| COGA1_EUR | 0.5976 | −0.032 | 0.061 |
| CUINT_EUR | 0.7733 | 0.022 | 0.076 |
| CVEDA_CSA | 0.5508 | 0.137 | 0.229 |
| ESTB2_EUR | 0.0757 | −0.043 | 0.024 |
| FINNG_EUR | 0.0191 | −0.057 | 0.024 |
| GEDIS_EUR | 0.0027 | −0.421 | 0.141 |
| GISS1_EUR | 0.0631 | −0.157 | 0.085 |
| GISS2_EUR | 0.9151 | 0.011 | 0.1 |
| GTPRJ_AFR | 0.7629 | 0.029 | 0.095 |
| IPSYC_EUR | 0.013 | −0.047 | 0.019 |
| JANS3_EUR | 0.0638 | 0.198 | 0.107 |
| JANS4_EUR | 0.0708 | 0.223 | 0.123 |
| MVPXQ_AFR | 0.5034 | −0.022 | 0.033 |
| MVPXQ_EUR | 5.18e−06 | −0.061 | 0.013 |
| MVPXQ_LAT | 0.119 | −0.06 | 0.039 |
| PGCED_EUR | 0.7979 | −0.03 | 0.116 |
| PGCMD_EUR | 0.17 | −0.049 | 0.035 |
| PGCPT_AFR | 0.7888 | 0.039 | 0.147 |
| PGCPT_EUR | 0.5612 | 0.034 | 0.059 |
| PRFCT_EUR | 0.0167 | −0.109 | 0.045 |
| PSYCR_EUR | 0.809 | −0.022 | 0.092 |
| QIMRB_EUR | 0.0825 | −0.049 | 0.028 |
| UKBJC_EUR | 0.3595 | −0.018 | 0.02 |
| UTAH2_EUR | 0.0363 | −0.041 | 0.02 |
| UTAMR_LAT | 0.5554 | −0.026 | 0.044 |
| YPENN_AFR | 0.3838 | 0.174 | 0.2 |
| YPENN_EUR | 0.8171 | −0.031 | 0.132 |
| **meta** | **3.19e−11** | **−0.043** | **0.006** |

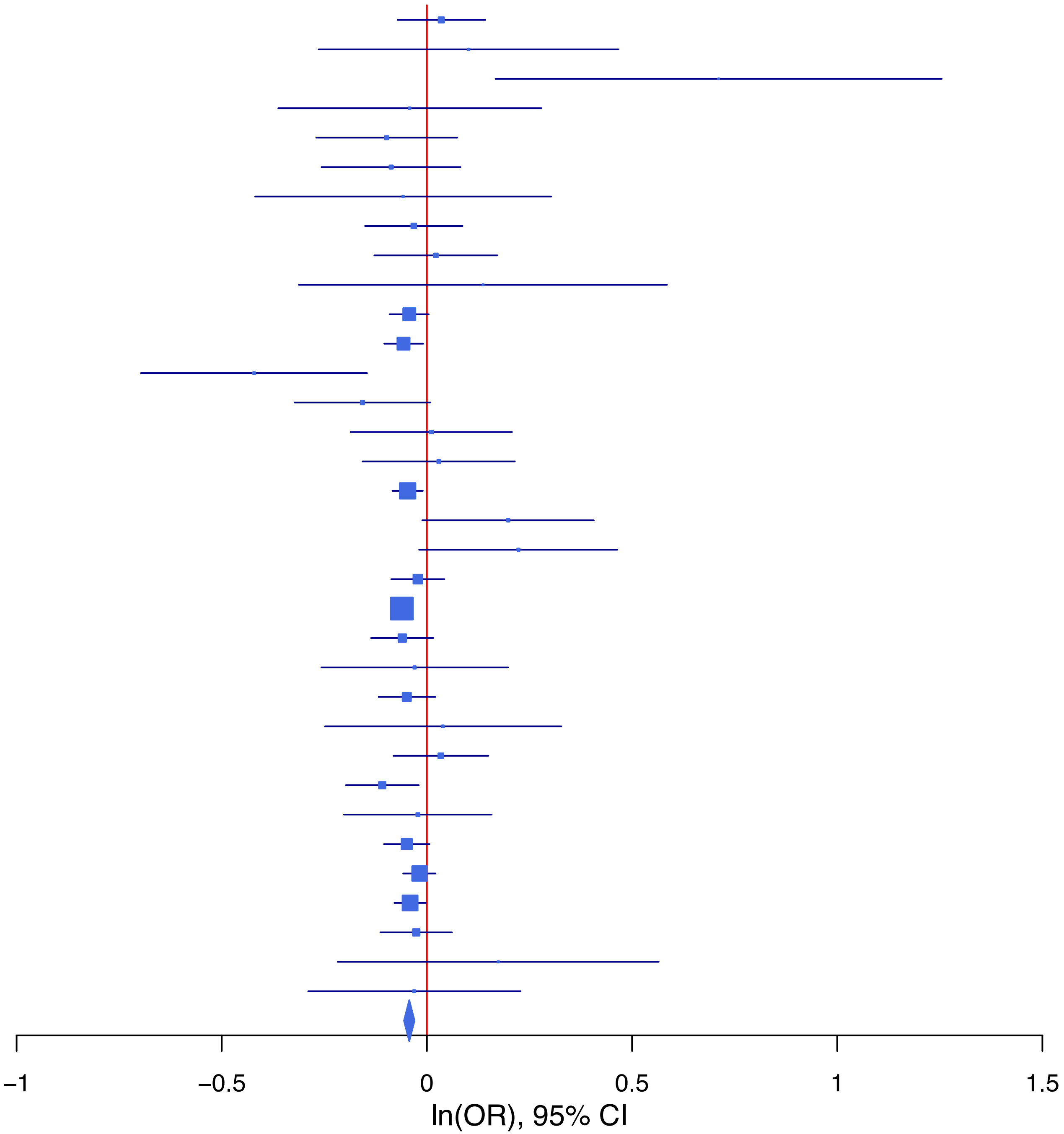

**rs1403174 A/T 7:2032865**

| Cohort | P | ln(OR) | SE |
|---|---|---|---|
| ALSPC_EUR | 0.3681 | 0.05 | 0.055 |
| BEPS7_EUR | 0.034 | 0.383 | 0.181 |
| BHRCM_AFR | 0.2078 | 0.435 | 0.345 |
| BHRCM_EUR | 0.3216 | −0.17 | 0.172 |
| BHRCM_LAT | 0.9352 | −0.02 | 0.247 |
| BOR17_EUR | 0.4994 | −0.058 | 0.086 |
| BOR2C_EUR | 0.0306 | 0.185 | 0.085 |
| BOR2E_EUR | 0.7202 | 0.065 | 0.182 |
| CNVRG_EAS | 0.7122 | 0.018 | 0.048 |
| COGA1_EUR | 0.08 | 0.105 | 0.06 |
| CUINT_EUR | 0.4134 | −0.061 | 0.075 |
| CVEDA_CSA | 0.4935 | −0.117 | 0.171 |
| ESTB2_EUR | 0.72 | 0.008 | 0.023 |
| FINNG_EUR | 0.0986 | 0.039 | 0.024 |
| GEDIS_EUR | 0.3574 | 0.118 | 0.128 |
| GISS1_EUR | 1 | 0 | 0.079 |
| GISS2_EUR | 0.6396 | 0.044 | 0.093 |
| GREAT_EAS | 0.1741 | 0.127 | 0.093 |
| IPSYC_EUR | 0.0176 | 0.044 | 0.019 |
| JANS3_EUR | 0.1217 | 0.159 | 0.103 |
| JANS4_EUR | 0.1556 | 0.17 | 0.12 |
| MIREC_AFR | 0.8264 | −0.036 | 0.166 |
| MIREC_EUR | 0.6955 | 0.061 | 0.155 |
| MVPXQ_AFR | 0.5516 | 0.014 | 0.024 |
| MVPXQ_EUR | 0.0025 | 0.038 | 0.012 |
| MVPXQ_LAT | 0.4371 | −0.025 | 0.033 |
| PGCBD_EUR | 0.1902 | 0.034 | 0.026 |
| PGCED_EUR | 0.743 | −0.039 | 0.117 |
| PGCMD_EUR | 0.0706 | −0.063 | 0.035 |
| PGCPT_AFR | 0.3503 | 0.128 | 0.138 |
| PGCPT_EUR | 0.529 | 0.037 | 0.058 |
| PGCSZ_EUR | 0.8273 | 0.008 | 0.037 |
| PRFCT_EUR | 0.6694 | 0.019 | 0.044 |
| PSYCR_EUR | 0.7589 | 0.026 | 0.086 |
| QIMRB_EUR | 0.0645 | 0.051 | 0.028 |
| SNUBH−ASA_EAS | 0.4592 | −0.094 | 0.127 |
| SNUBH−KCHIP_EAS | 0.0685 | 0.322 | 0.177 |
| STRR1_LAT | 0.2434 | 0.209 | 0.179 |
| UKBJC_EUR | 0.0019 | 0.061 | 0.02 |
| UTAH2_EUR | 0.1773 | 0.026 | 0.019 |
| UTAMR_LAT | 0.0892 | 0.062 | 0.036 |
| YPENN_AFR | 0.4693 | 0.126 | 0.174 |
| YPENN_EUR | 0.739 | 0.043 | 0.13 |
| **meta** | **1.78e−08** | **0.033** | **0.006** |

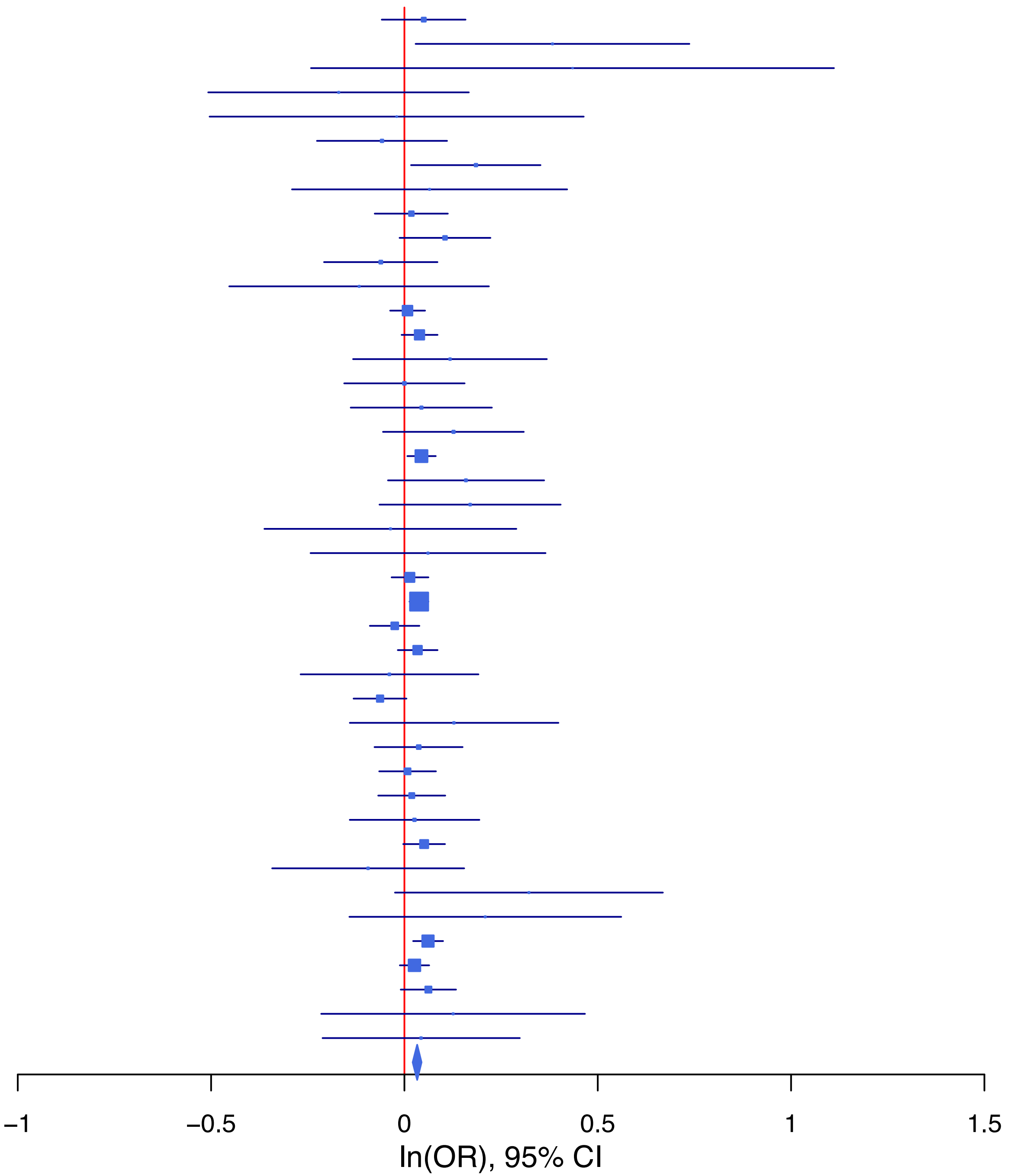

**rs1452787 G/A 18:53207207**

| Cohort | P | ln(OR) | SE |
|---|---|---|---|
| ADHEA_AFR | 0.5636 | 0.109 | 0.189 |
| ADHEA_EUR | 0.3651 | 0.066 | 0.073 |
| ALSPC_EUR | 0.6703 | −0.026 | 0.061 |
| BEPS7_EUR | 0.3116 | 0.194 | 0.192 |
| BHRCM_AFR | 0.3893 | −0.396 | 0.46 |
| BHRCM_EUR | 0.5528 | −0.103 | 0.173 |
| BHRCM_LAT | 0.3291 | 0.262 | 0.269 |
| BOR17_EUR | 0.9278 | −0.008 | 0.094 |
| BOR2C_EUR | 0.1103 | −0.153 | 0.096 |
| BOR2E_EUR | 0.4452 | −0.152 | 0.199 |
| CNVRG_EAS | 0.236 | 0.055 | 0.046 |
| COGA1_AFR | 0.4578 | −0.124 | 0.167 |
| COGA1_EUR | 0.4333 | 0.05 | 0.064 |
| CUINT_EUR | 0.5911 | 0.044 | 0.082 |
| CVEDA_CSA | 0.3156 | −0.169 | 0.169 |
| ESTB2_EUR | 0.1561 | 0.035 | 0.024 |
| FINNG_EUR | 0.1266 | 0.038 | 0.025 |
| GEDIS_EUR | 0.5074 | −0.097 | 0.146 |
| GISS1_EUR | 0.0353 | 0.174 | 0.083 |
| GISS2_EUR | 0.5508 | −0.057 | 0.096 |
| GTPRJ_AFR | 0.0391 | 0.208 | 0.101 |
| IPSYC_EUR | 0.0896 | 0.035 | 0.021 |
| JANS3_EUR | 0.0944 | −0.198 | 0.118 |
| JANS4_EUR | 0.6287 | −0.063 | 0.13 |
| JAPAN_EAS | 0.5863 | −0.03 | 0.056 |
| MIREC_AFR | 0.5286 | −0.15 | 0.238 |
| MIREC_EUR | 0.5564 | −0.099 | 0.168 |
| MVPXQ_AFR | 0.5978 | 0.018 | 0.034 |
| MVPXQ_EUR | 1e−04 | 0.052 | 0.014 |
| MVPXQ_LAT | 0.2712 | −0.037 | 0.034 |
| PGCBD_EUR | 0.7559 | −0.009 | 0.029 |
| PGCED_EUR | 0.5108 | −0.083 | 0.127 |
| PGCMD_EUR | 0.8691 | −0.006 | 0.038 |
| PGCPT_EUR | 0.984 | −0.001 | 0.063 |
| PGCSZ_EUR | 0.0079 | 0.107 | 0.04 |
| PRFCT_EUR | 0.1278 | 0.074 | 0.048 |
| PSYCR_EUR | 0.5973 | 0.05 | 0.094 |
| QIMRB_EUR | 0.3472 | 0.028 | 0.03 |
| SNUBH-ASA_EAS | 0.0676 | 0.23 | 0.126 |
| SNUBH-KCHIP_EAS | 0.0987 | 0.277 | 0.168 |
| STRR1_LAT | 0.5123 | 0.116 | 0.178 |
| UKBJC_EUR | 0.0034 | 0.063 | 0.022 |
| UTAH2_EUR | 0.0198 | 0.048 | 0.021 |
| UTAMR_LAT | 0.0237 | 0.08 | 0.035 |
| YPENN_AFR | 0.5252 | 0.141 | 0.222 |
| YPENN_EUR | 0.4574 | 0.099 | 0.133 |
| **meta** | **2.18e−09** | **0.037** | **0.006** |

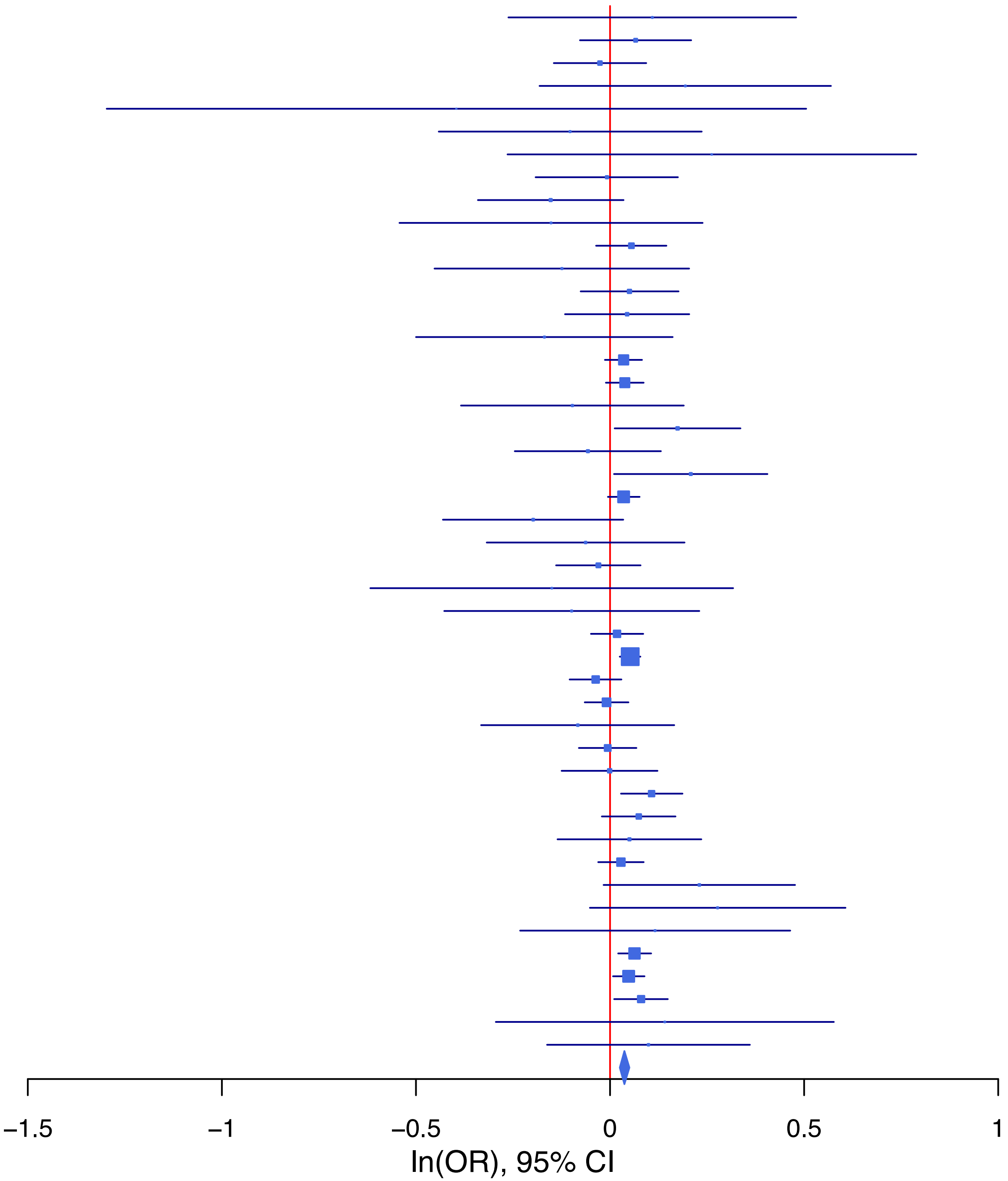

**rs1797235 C/G 15:47821612**

| Cohort | P | ln(OR) | SE |
|---|---|---|---|
| ALSPC_EUR | 0.8318 | −0.012 | 0.056 |
| BEPS7_EUR | 0.0208 | 0.429 | 0.185 |
| BHRCM_AFR | 0.7219 | −0.093 | 0.262 |
| BHRCM_EUR | 0.9407 | −0.012 | 0.156 |
| BHRCM_LAT | 0.1136 | 0.493 | 0.311 |
| BOR17_EUR | 0.7886 | 0.023 | 0.087 |
| BOR2C_EUR | 0.4423 | −0.07 | 0.091 |
| BOR2E_EUR | 0.2324 | −0.228 | 0.191 |
| CNVRG_EAS | 0.315 | 0.046 | 0.046 |
| COGA1_AFR | 0.7051 | 0.044 | 0.117 |
| COGA1_EUR | 0.1913 | 0.079 | 0.06 |
| CUINT_EUR | 0.0328 | −0.163 | 0.076 |
| CVEDA_CSA | 0.381 | −0.146 | 0.167 |
| ESTB2_EUR | 0.1723 | 0.032 | 0.024 |
| FINNG_EUR | 0.0721 | 0.043 | 0.024 |
| GEDIS_EUR | 0.9033 | −0.016 | 0.134 |
| GISS1_EUR | 0.9038 | −0.01 | 0.081 |
| GISS2_EUR | 0.0221 | 0.224 | 0.098 |
| GREAT_EAS | 0.0341 | 0.161 | 0.076 |
| GTPRJ_AFR | 0.162 | 0.097 | 0.069 |
| IPSYC_EUR | 0.1461 | 0.028 | 0.019 |
| JANS3_EUR | 0.1342 | 0.156 | 0.104 |
| JANS4_EUR | 0.939 | 0.009 | 0.12 |
| JAPAN_EAS | 0.0703 | 0.099 | 0.055 |
| PGCBD_EUR | 0.6509 | 0.012 | 0.026 |
| PGCED_EUR | 0.7598 | 0.036 | 0.117 |
| PGCMD_EUR | 0.0114 | 0.089 | 0.035 |
| PGCPT_AFR | 0.9004 | 0.015 | 0.118 |
| PGCPT_EUR | 0.4513 | −0.044 | 0.059 |
| PGCSZ_EUR | 0.0015 | 0.121 | 0.038 |
| PRFCT_EUR | 0.038 | −0.095 | 0.046 |
| PSYCR_EUR | 0.0563 | 0.17 | 0.089 |
| QIMRB_EUR | 0.2652 | 0.032 | 0.028 |
| SNUBH−ASA_EAS | 0.4821 | −0.087 | 0.123 |
| SNUBH−KCHIP_EAS | 0.0133 | −0.405 | 0.16 |
| STRR1_LAT | 0.5336 | 0.107 | 0.173 |
| UKBJC_EUR | 1e−04 | 0.078 | 0.02 |
| UTAH2_EUR | 2e−04 | 0.071 | 0.019 |
| UTAMR_LAT | 0.7264 | 0.013 | 0.036 |
| YPENN_AFR | 0.7714 | −0.048 | 0.165 |
| YPENN_EUR | 0.1261 | 0.195 | 0.128 |
| **meta** | **3.03e−10** | **0.044** | **0.007** |

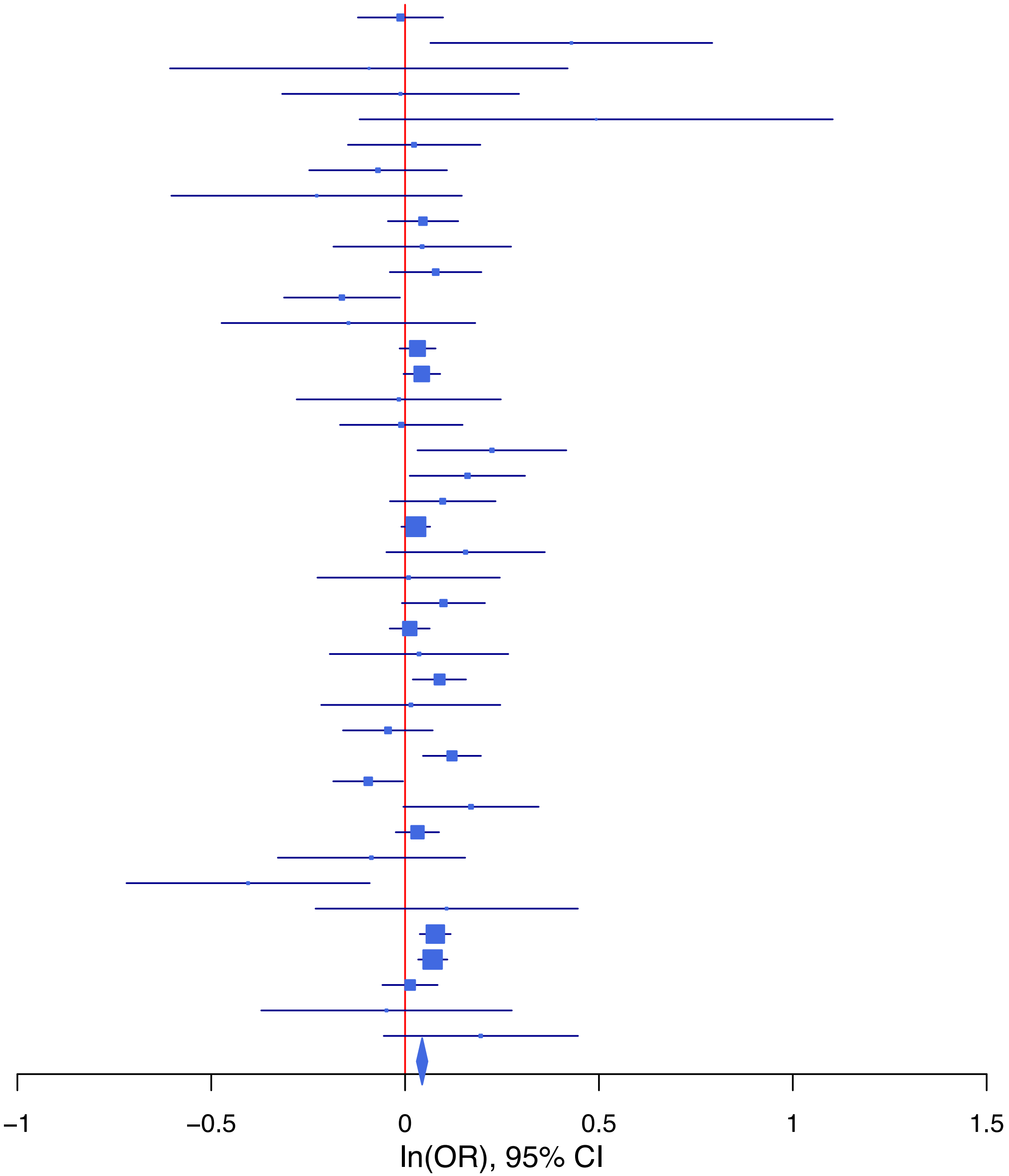

**rs2032465 G/A 22:37025819**

| Cohort | P | ln(OR) | SE |
|---|---|---|---|
| ALSPC_EUR | 0.5842 | 0.031 | 0.056 |
| BEPS7_EUR | 0.8381 | 0.038 | 0.185 |
| BOR17_EUR | 0.7839 | −0.025 | 0.09 |
| BOR2C_EUR | 0.1399 | −0.132 | 0.09 |
| BOR2E_EUR | 0.4344 | 0.147 | 0.188 |
| CNVRG_EAS | 0.132 | −0.07 | 0.046 |
| COGA1_AFR | 0.6393 | 0.052 | 0.11 |
| COGA1_EUR | 0.1607 | −0.086 | 0.062 |
| CUINT_EUR | 0.0142 | −0.188 | 0.077 |
| CVEDA_CSA | 0.1204 | 0.251 | 0.161 |
| ESTB2_EUR | 0.0051 | −0.066 | 0.024 |
| FINNG_EUR | 0.7784 | −0.007 | 0.024 |
| GEDIS_EUR | 0.4136 | 0.109 | 0.133 |
| GISS1_EUR | 0.9676 | 0.003 | 0.081 |
| GISS2_EUR | 0.1098 | −0.152 | 0.095 |
| GREAT_EAS | 0.4746 | −0.059 | 0.083 |
| GTPRJ_AFR | 0.8155 | −0.016 | 0.069 |
| IPSYC_EUR | 0.1851 | −0.026 | 0.02 |
| JANS3_EUR | 0.7321 | −0.037 | 0.108 |
| JANS4_EUR | 0.6453 | 0.055 | 0.119 |
| PGCBD_EUR | 0.0465 | −0.054 | 0.027 |
| PGCED_EUR | 0.4236 | −0.095 | 0.119 |
| PGCMD_EUR | 0.7909 | 0.01 | 0.036 |
| PGCPT_AFR | 0.9589 | 0.007 | 0.128 |
| PGCPT_EUR | 0.3345 | −0.058 | 0.06 |
| PGCSZ_EUR | 0.8725 | −0.006 | 0.039 |
| PRFCT_EUR | 0.0722 | −0.083 | 0.046 |
| PSYCR_EUR | 0.0701 | 0.172 | 0.095 |
| QIMRB_EUR | 0.3967 | −0.025 | 0.029 |
| SNUBH−ASA_EAS | 0.4353 | −0.098 | 0.126 |
| STRR1_LAT | 0.2313 | 0.219 | 0.183 |
| UKBJC_EUR | 2e−04 | −0.077 | 0.021 |
| UTAH2_EUR | 0.0158 | −0.047 | 0.02 |
| UTAMR_LAT | 0.0782 | −0.067 | 0.038 |
| YPENN_AFR | 0.6248 | 0.083 | 0.17 |
| YPENN_EUR | 0.108 | −0.223 | 0.139 |
| **meta** | **9.52e−09** | **−0.041** | **0.007** |

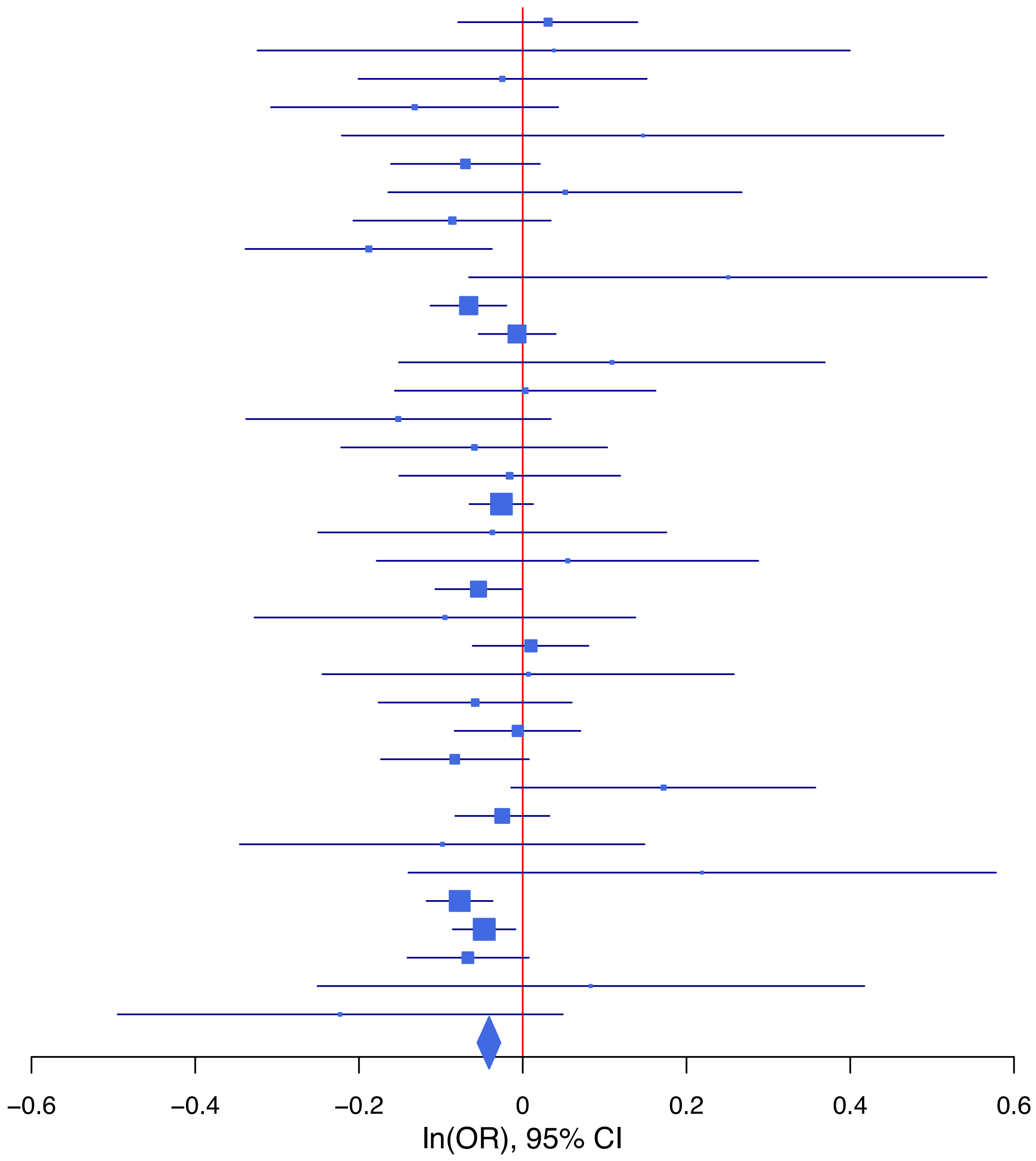

**rs2051710 T/G 7:114942226**

| Cohort | P | ln(OR) | SE |
|---|---|---|---|
| ADHEA_EUR | 0.8549 | 0.027 | 0.147 |
| ALSPC_EUR | 0.0513 | 0.214 | 0.11 |
| BHRCM_AFR | 0.4437 | −0.192 | 0.251 |
| BHRCM_EUR | 0.9199 | 0.031 | 0.317 |
| BOR17_EUR | 0.5539 | −0.108 | 0.183 |
| BOR2C_EUR | 0.0861 | −0.314 | 0.183 |
| BOR2E_EUR | 0.1138 | 0.55 | 0.348 |
| CNVRG_EAS | 0.6829 | 0.037 | 0.091 |
| COGA1_AFR | 0.3983 | 0.111 | 0.132 |
| COGA1_EUR | 0.1127 | 0.193 | 0.121 |
| CUINT_EUR | 0.3585 | 0.136 | 0.148 |
| CVEDA_CSA | 0.2233 | 0.316 | 0.259 |
| ESTB2_EUR | 0.0249 | 0.113 | 0.051 |
| FINNG_EUR | 0.0777 | 0.106 | 0.06 |
| GEDIS_EUR | 0.2868 | 0.263 | 0.247 |
| GISS1_EUR | 0.2573 | 0.198 | 0.175 |
| GISS2_EUR | 0.1139 | 0.337 | 0.213 |
| IPSYC_EUR | 0.0059 | 0.11 | 0.04 |
| JANS3_EUR | 0.7776 | −0.061 | 0.218 |
| JANS4_EUR | 0.3408 | 0.211 | 0.221 |
| JAPAN_EAS | 0.6876 | 0.051 | 0.127 |
| MVPXQ_EAS | 0.2733 | 0.268 | 0.244 |
| MVPXQ_EUR | 0.0626 | 0.049 | 0.027 |
| MVPXQ_LAT | 0.0239 | 0.167 | 0.074 |
| PGCBD_EUR | 0.1575 | 0.09 | 0.064 |
| PGCMD_EUR | 0.7819 | −0.023 | 0.082 |
| PGCPT_AFR | 0.8757 | 0.021 | 0.135 |
| PGCPT_EUR | 0.0174 | 0.265 | 0.111 |
| PGCSZ_EUR | 0.0443 | 0.165 | 0.082 |
| PRFCT_EUR | 0.7297 | −0.036 | 0.104 |
| PSYCR_EUR | 0.8073 | −0.046 | 0.191 |
| QIMRB_EUR | 0.2389 | 0.068 | 0.058 |
| SNUBH−ASA_EAS | 0.2893 | 0.311 | 0.293 |
| UKBJC_EUR | 0.4821 | −0.03 | 0.042 |
| UTAH2_EUR | 0.2407 | 0.047 | 0.04 |
| YPENN_AFR | 0.0629 | 0.371 | 0.199 |
| YPENN_EUR | 0.8064 | 0.066 | 0.269 |
| **meta** | **4.12e−08** | **0.071** | **0.013** |

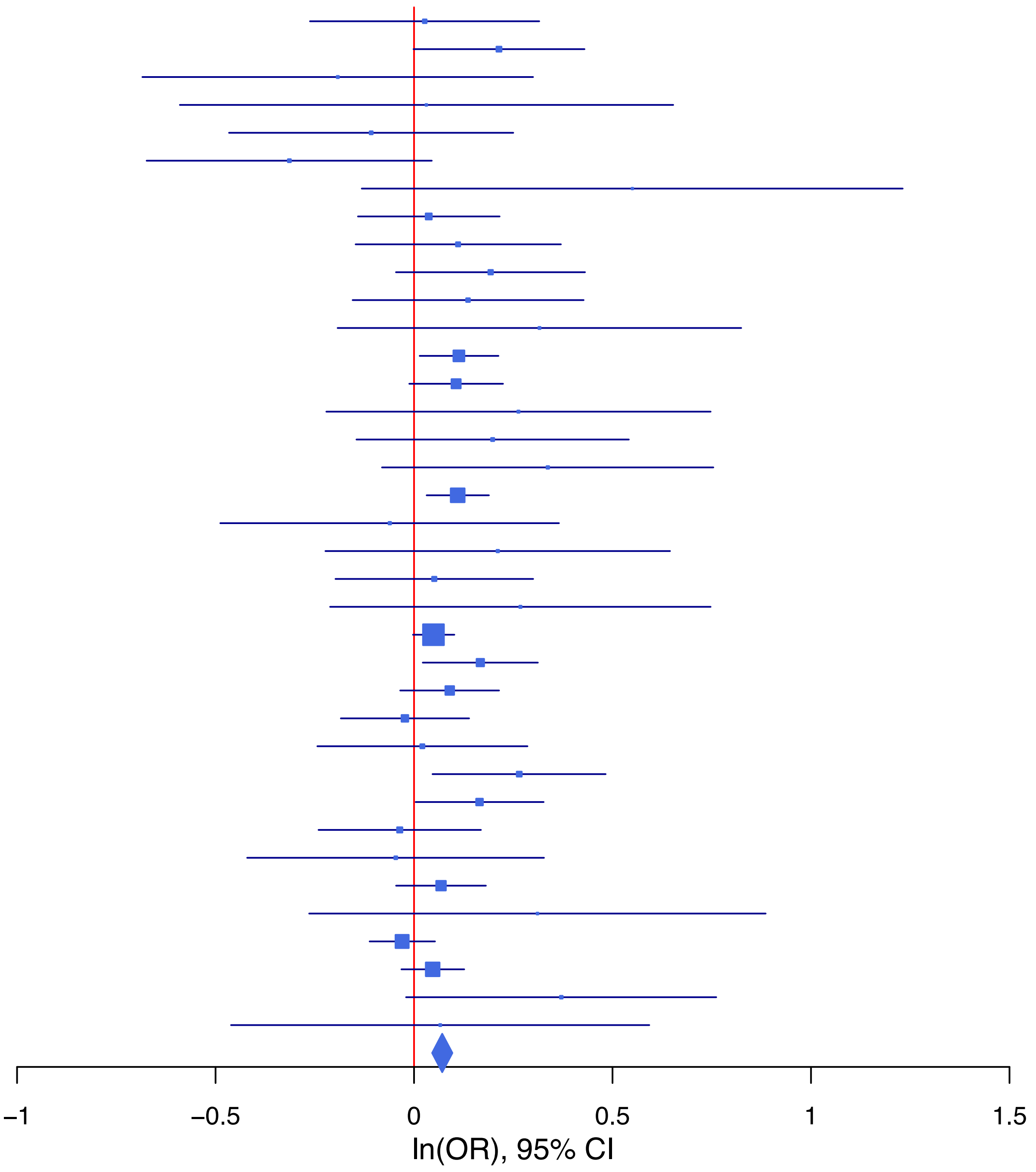

**rs2071382 T/C 15:91428197**

| Cohort | P | ln(OR) | SE |
|---|---|---|---|
| ALSPC_EUR | 0.2147 | −0.069 | 0.055 |
| BEPS7_EUR | 0.0924 | −0.284 | 0.169 |
| BHRCM_AFR | 0.1315 | 0.287 | 0.19 |
| BHRCM_EUR | 0.7667 | 0.045 | 0.153 |
| BHRCM_LAT | 0.6946 | 0.248 | 0.631 |
| BOR17_EUR | 0.1767 | −0.115 | 0.085 |
| BOR2C_EUR | 0.8866 | 0.012 | 0.086 |
| BOR2E_EUR | 0.1794 | 0.237 | 0.176 |
| CNVRG_EAS | 0.206 | −0.078 | 0.061 |
| COGA1_AFR | 0.2684 | −0.112 | 0.101 |
| COGA1_EUR | 0.4558 | −0.044 | 0.059 |
| CUINT_EUR | 0.5807 | −0.04 | 0.073 |
| CVEDA_CSA | 0.7058 | −0.064 | 0.168 |
| ESTB2_EUR | 0.8926 | 0.003 | 0.023 |
| FINNG_EUR | 0.3543 | −0.022 | 0.023 |
| GEDIS_EUR | 0.5086 | 0.085 | 0.129 |
| GISS1_EUR | 0.1021 | −0.127 | 0.078 |
| GISS2_EUR | 0.4894 | −0.064 | 0.092 |
| GREAT_EAS | 0.7413 | −0.039 | 0.119 |
| GTPRJ_AFR | 0.4637 | 0.044 | 0.061 |
| IPSYC_EUR | 0.0652 | −0.034 | 0.018 |
| JANS3_EUR | 0.2832 | 0.109 | 0.102 |
| JANS4_EUR | 0.2372 | −0.143 | 0.121 |
| JAPAN_EAS | 0.1608 | −0.117 | 0.083 |
| MVPXQ_EUR | 0.0086 | −0.034 | 0.013 |
| MVPXQ_LAT | 0.2442 | −0.044 | 0.038 |
| PGCBD_EUR | 0.3198 | −0.026 | 0.026 |
| PGCED_EUR | 0.2289 | −0.142 | 0.118 |
| PGCMD_EUR | 0.0437 | −0.071 | 0.035 |
| PGCPT_EUR | 0.4088 | −0.048 | 0.058 |
| PGCSZ_EUR | 0.3233 | −0.038 | 0.038 |
| PRFCT_EUR | 0.2479 | −0.051 | 0.044 |
| PSYCR_EUR | 0.8823 | 0.013 | 0.09 |
| QIMRB_EUR | 0.0069 | −0.075 | 0.028 |
| SNUBH−ASA_EAS | 0.0986 | −0.268 | 0.162 |
| STRR1_LAT | 0.1354 | 0.282 | 0.189 |
| UKBJC_EUR | 0.0015 | −0.062 | 0.02 |
| UTAH2_EUR | 0.1729 | −0.025 | 0.019 |
| UTAMR_LAT | 0.6574 | −0.016 | 0.037 |
| YPENN_EUR | 0.5948 | −0.068 | 0.129 |
| **meta** | **1.69e−09** | **−0.036** | **0.006** |

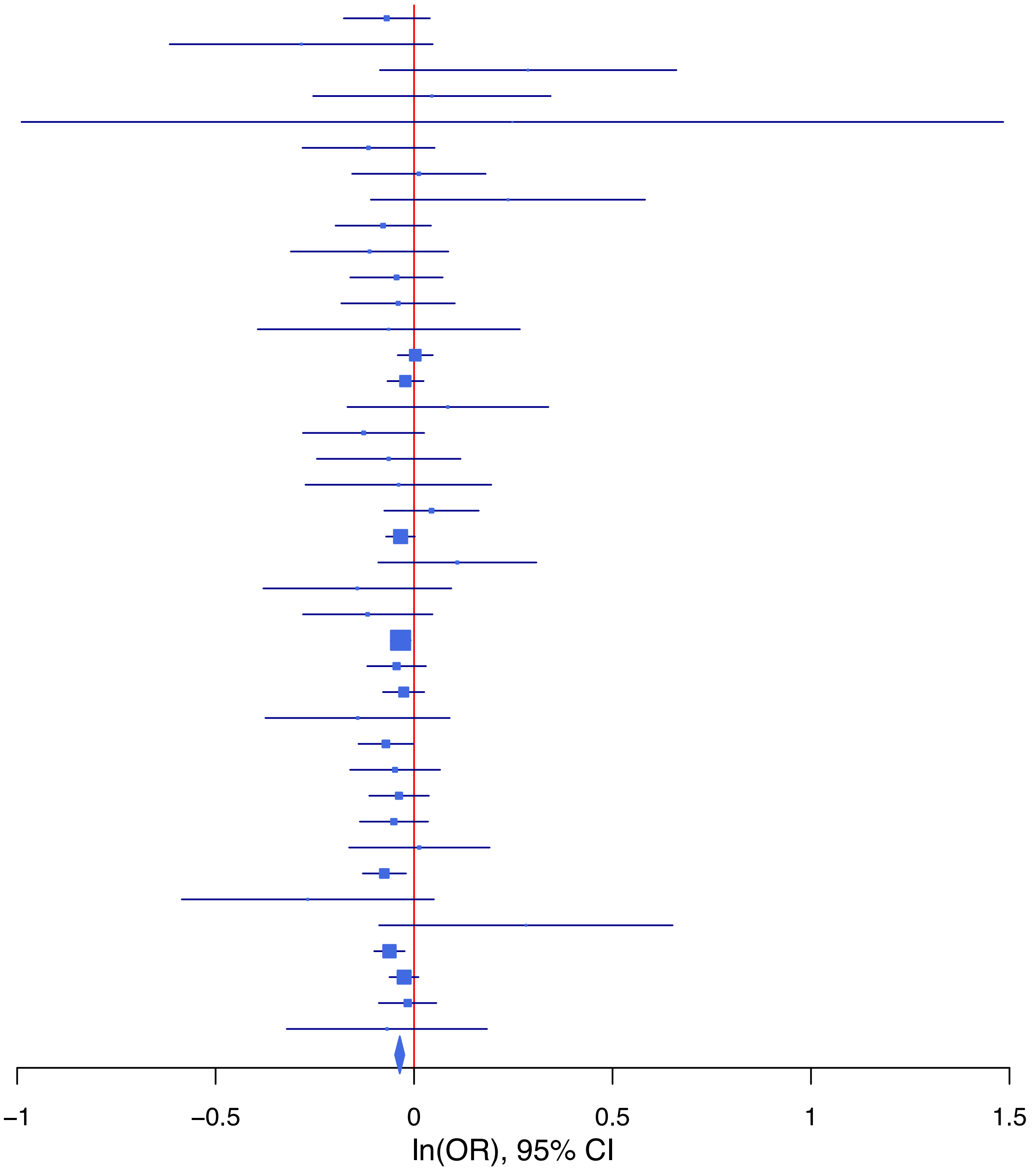

**rs2155281 A/G 11:112838338**

| Cohort | P | ln(OR) | SE |
|---|---|---|---|
| ADHEA_AFR | 0.3913 | 0.097 | 0.113 |
| ADHEA_EUR | 0.4464 | 0.053 | 0.069 |
| ALSPC_EUR | 0.6289 | −0.027 | 0.055 |
| BEPS7_EUR | 0.5775 | 0.108 | 0.194 |
| BHRCM_AFR | 0.3664 | 0.163 | 0.18 |
| BHRCM_EUR | 0.9287 | 0.015 | 0.17 |
| BHRCM_LAT | 0.8843 | −0.037 | 0.257 |
| BOR17_EUR | 0.7363 | 0.029 | 0.087 |
| BOR2C_EUR | 0.0787 | 0.158 | 0.09 |
| BOR2E_EUR | 0.3841 | −0.177 | 0.204 |
| CNVRG_EAS | 0.6436 | 0.027 | 0.058 |
| COGA1_AFR | 0.9138 | −0.011 | 0.102 |
| COGA1_EUR | 0.4002 | 0.05 | 0.059 |
| CUINT_EUR | 0.4539 | 0.056 | 0.075 |
| CVEDA_CSA | 0.5176 | 0.106 | 0.165 |
| ESTB2_EUR | 0.0862 | 0.039 | 0.023 |
| FINNG_EUR | 0.1467 | 0.034 | 0.024 |
| GEDIS_EUR | 0.8247 | −0.03 | 0.136 |
| GISS1_EUR | 0.4595 | −0.058 | 0.078 |
| GISS2_EUR | 0.0712 | 0.174 | 0.096 |
| GREAT_EAS | 0.6277 | −0.054 | 0.112 |
| GTPRJ_AFR | 0.7217 | 0.021 | 0.059 |
| IPSYC_EUR | 0.1056 | 0.031 | 0.019 |
| JANS3_EUR | 0.0781 | 0.187 | 0.106 |
| JANS4_EUR | 0.4831 | 0.086 | 0.123 |
| JAPAN_EAS | 0.6366 | −0.031 | 0.066 |
| MVPXQ_EUR | 1.72e−05 | 0.056 | 0.013 |
| MVPXQ_LAT | 0.2509 | 0.039 | 0.034 |
| PGCBD_EUR | 0.0797 | 0.046 | 0.026 |
| PGCED_EUR | 0.2986 | 0.12 | 0.115 |
| PGCMD_EUR | 0.5444 | 0.021 | 0.035 |
| PGCPT_AFR | 0.1678 | 0.144 | 0.104 |
| PGCPT_EUR | 0.3805 | −0.052 | 0.059 |
| PGCSZ_EUR | 0.1023 | 0.061 | 0.037 |
| PRFCT_EUR | 0.5988 | −0.023 | 0.044 |
| PSYCR_EUR | 0.387 | 0.078 | 0.09 |
| QIMRB_EUR | 0.1797 | 0.038 | 0.029 |
| SNUBH−ASA_EAS | 0.1447 | −0.209 | 0.144 |
| SNUBH−KCHIP_EAS | 0.9069 | 0.025 | 0.215 |
| STRR1_LAT | 0.9803 | 0.004 | 0.175 |
| UKBJC_EUR | 0.0513 | 0.039 | 0.02 |
| UTAH2_EUR | 0.9756 | 0.001 | 0.019 |
| UTAMR_LAT | 0.9488 | 0.002 | 0.038 |
| YPENN_EUR | 0.8922 | −0.017 | 0.128 |
| **meta** | **1.02e−08** | **0.034** | **0.006** |

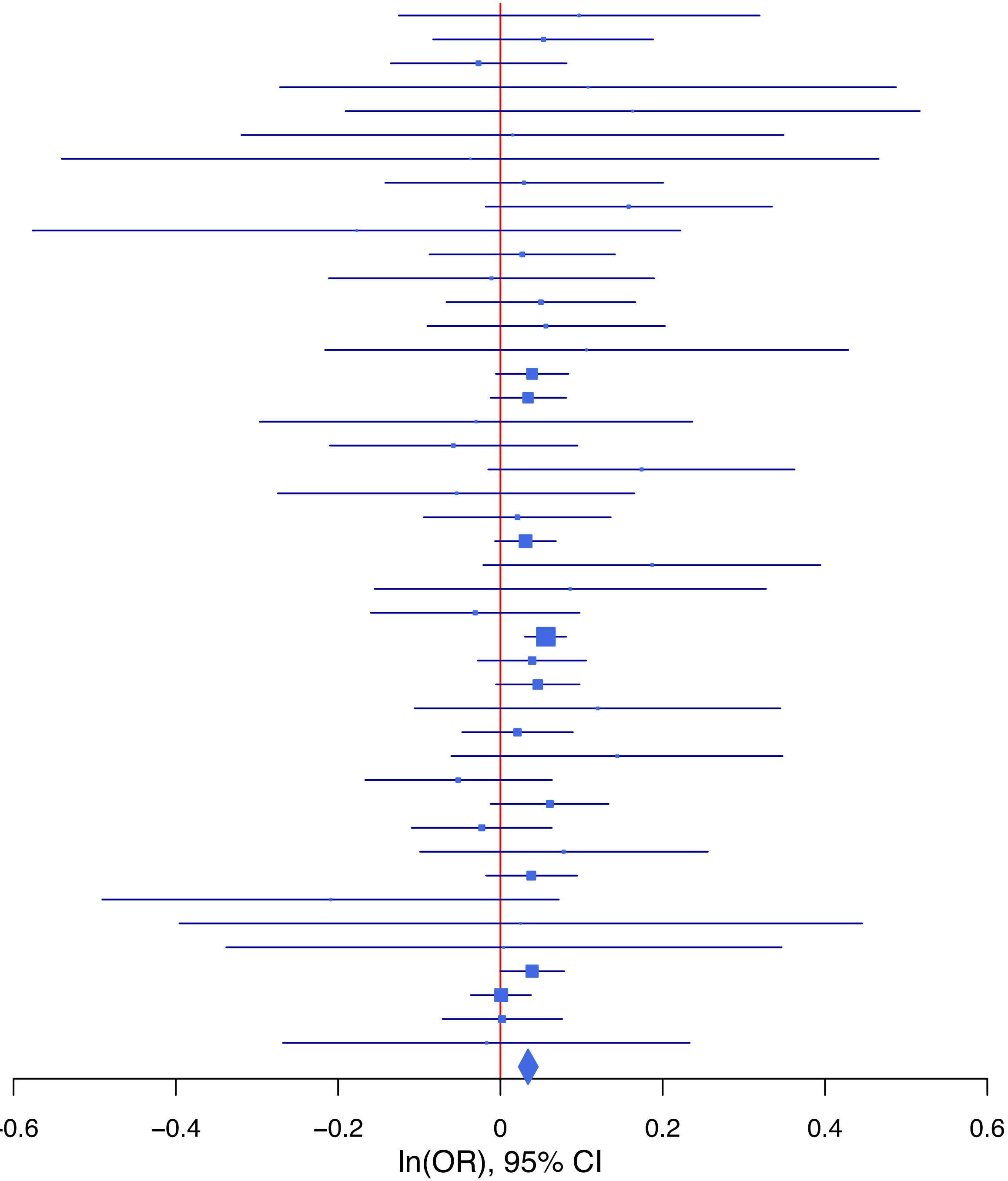

**rs2645977   A/G   12:84009188**

| Cohort | P | ln(OR) | SE |
|---|---|---|---|
| ADHEA_AFR | 0.8767 | −0.031 | 0.199 |
| ADHEA_EUR | 0.0112 | −0.186 | 0.073 |
| ALSPC_EUR | 0.886 | −0.008 | 0.056 |
| BEPS7_EUR | 0.0265 | 0.399 | 0.18 |
| BHRCM_AFR | 0.7251 | −0.191 | 0.543 |
| BHRCM_EUR | 0.9409 | −0.012 | 0.164 |
| BHRCM_LAT | 0.1352 | −0.616 | 0.412 |
| BOR17_EUR | 0.0227 | −0.201 | 0.088 |
| BOR2C_EUR | 0.3931 | 0.078 | 0.091 |
| BOR2E_EUR | 0.5303 | −0.125 | 0.199 |
| CNVRG_EAS | 0.3711 | −0.042 | 0.047 |
| COGA1_EUR | 0.3258 | −0.06 | 0.061 |
| CUINT_EUR | 0.2851 | 0.083 | 0.077 |
| CVEDA_CSA | 0.8629 | −0.031 | 0.18 |
| ESTB2_EUR | 0.8291 | −0.005 | 0.023 |
| FINNG_EUR | 0.0409 | −0.047 | 0.023 |
| GEDIS_EUR | 0.9746 | −0.004 | 0.131 |
| GISS1_EUR | 0.3456 | −0.074 | 0.079 |
| GISS2_EUR | 0.0221 | −0.213 | 0.093 |
| GREAT_EAS | 0.9913 | 0.001 | 0.078 |
| GTPRJ_AFR | 0.9006 | 0.013 | 0.107 |
| IPSYC_EUR | 0.0685 | −0.035 | 0.019 |
| JANS3_EUR | 0.1201 | 0.163 | 0.105 |
| JANS4_EUR | 0.3344 | 0.121 | 0.125 |
| JAPAN_EAS | 0.0212 | −0.135 | 0.058 |
| MIREC_EUR | 0.833 | 0.032 | 0.155 |
| MVPXQ_AFR | 0.981 | 0.001 | 0.037 |
| MVPXQ_LAT | 0.6482 | 0.017 | 0.037 |
| PGCBD_EUR | 0.7394 | 0.009 | 0.027 |
| PGCED_EUR | 0.148 | −0.177 | 0.123 |
| PGCMD_EUR | 0.2869 | −0.038 | 0.036 |
| PGCPT_EUR | 0.4523 | −0.046 | 0.061 |
| PGCSZ_EUR | 0.4729 | −0.028 | 0.039 |
| PRFCT_EUR | 0.0581 | −0.086 | 0.045 |
| PSYCR_EUR | 0.8488 | −0.017 | 0.09 |
| QIMRB_EUR | 0.1493 | −0.041 | 0.028 |
| SNUBH-ASA_EAS | 0.0048 | −0.363 | 0.13 |
| STRR1_LAT | 0.2929 | −0.207 | 0.197 |
| UKBJC_EUR | 0.0061 | −0.056 | 0.02 |
| UTAH2_EUR | 0.0012 | −0.064 | 0.02 |
| UTAMR_LAT | 0.1254 | −0.059 | 0.039 |
| YPENN_AFR | 0.3415 | −0.252 | 0.264 |
| YPENN_EUR | 0.342 | 0.126 | 0.132 |
| **meta** | **2.69e−08** | **−0.038** | **0.007** |

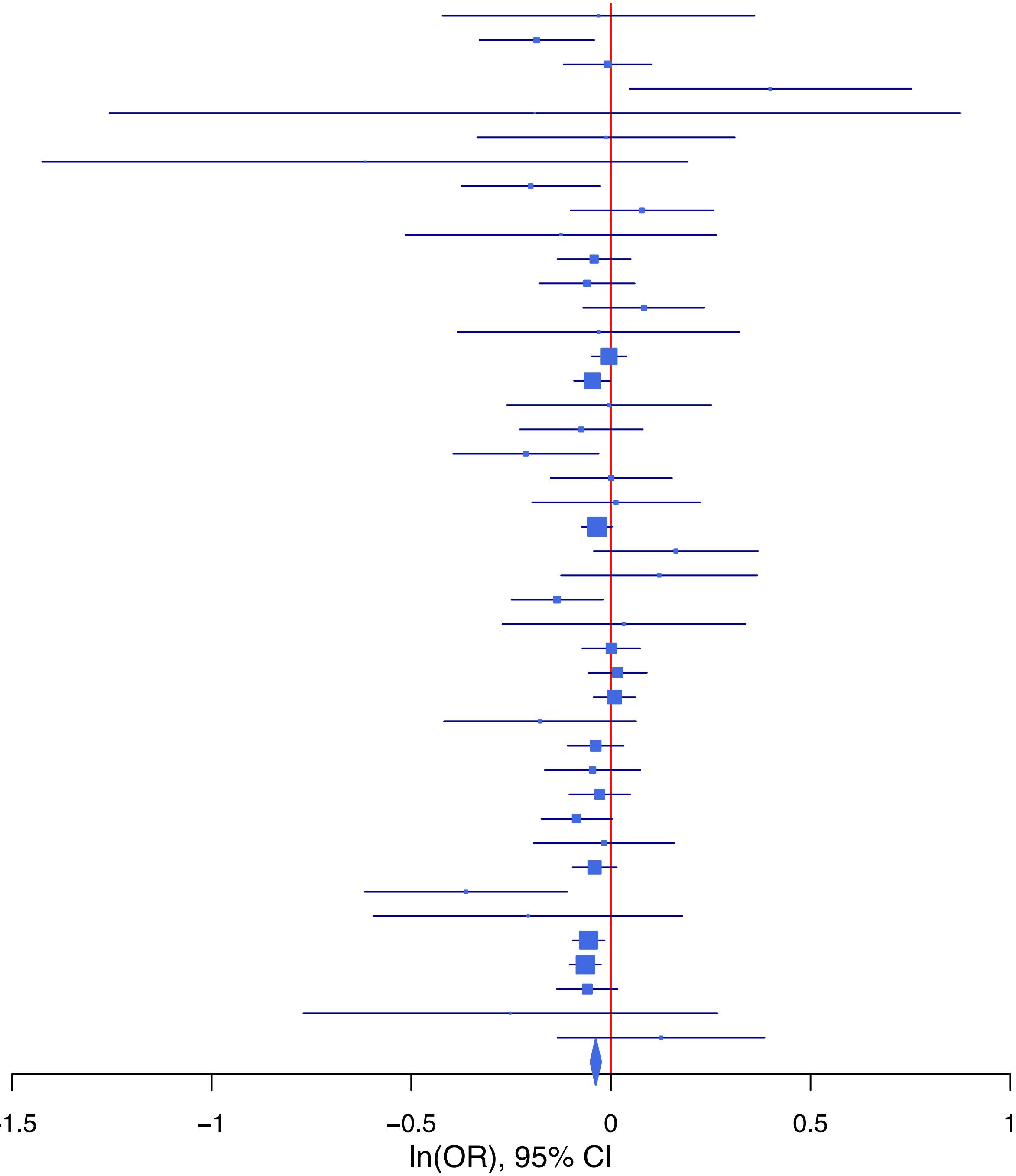

**rs2876370 T/A 6:137917237**

| Cohort | P | ln(OR) | SE |
|---|---|---|---|
| ADHEA_AFR | 0.1118 | 0.339 | 0.214 |
| ADHEA_EUR | 0.8042 | 0.022 | 0.086 |
| ALSPC_EUR | 0.8358 | −0.015 | 0.07 |
| BEPS7_EUR | 0.2285 | 0.259 | 0.215 |
| BHRCM_AFR | 0.0662 | 0.696 | 0.379 |
| BHRCM_EUR | 0.0864 | −0.361 | 0.21 |
| BHRCM_LAT | 0.1508 | 0.476 | 0.332 |
| BOR17_EUR | 0.6046 | −0.057 | 0.111 |
| BOR2C_EUR | 0.799 | −0.028 | 0.11 |
| BOR2E_EUR | 0.2867 | −0.227 | 0.213 |
| CNVRG_EAS | 0.5735 | 0.026 | 0.046 |
| COGA1_AFR | 0.6333 | 0.086 | 0.18 |
| COGA1_EUR | 0.242 | −0.09 | 0.077 |
| CVEDA_CSA | 0.0724 | 0.329 | 0.183 |
| ESTB2_EUR | 0.0512 | −0.054 | 0.028 |
| FINNG_EUR | 0.0018 | −0.078 | 0.025 |
| GEDIS_EUR | 0.8399 | −0.035 | 0.172 |
| GISS1_EUR | 0.4855 | −0.07 | 0.1 |
| GISS2_EUR | 0.402 | 0.099 | 0.118 |
| GREAT_EAS | 0.8591 | −0.014 | 0.078 |
| GTPRJ_AFR | 0.777 | 0.032 | 0.116 |
| IPSYC_EUR | 0.0664 | −0.042 | 0.023 |
| JANS3_EUR | 0.8131 | −0.033 | 0.138 |
| JANS4_EUR | 0.0525 | 0.278 | 0.144 |
| MVPXQ_AFR | 0.2652 | −0.046 | 0.041 |
| MVPXQ_EUR | 0.003 | −0.05 | 0.017 |
| PGCBD_EUR | 0.3128 | −0.034 | 0.033 |
| PGCED_EUR | 0.7759 | −0.043 | 0.15 |
| PGCMD_EUR | 0.0887 | −0.077 | 0.046 |
| PGCPT_EUR | 0.6282 | 0.035 | 0.073 |
| PGCSZ_EUR | 0.2866 | −0.052 | 0.048 |
| PRFCT_EUR | 0.6737 | −0.023 | 0.055 |
| PSYCR_EUR | 0.6454 | −0.052 | 0.113 |
| QIMRB_EUR | 0.2397 | −0.041 | 0.035 |
| SNUBH−ASA_EAS | 0.7757 | −0.035 | 0.124 |
| SNUBH−KCHIP_EAS | 0.0044 | −0.466 | 0.16 |
| STRR1_LAT | 0.0587 | 0.424 | 0.224 |
| UKBJC_EUR | 0.1672 | −0.035 | 0.025 |
| UTAH2_EUR | 0.0022 | −0.072 | 0.024 |
| UTAMR_LAT | 0.0836 | 0.082 | 0.047 |
| YPENN_AFR | 0.4198 | −0.26 | 0.322 |
| YPENN_EUR | 0.5128 | 0.103 | 0.157 |
| **meta** | **3.08e−08** | **−0.041** | **0.007** |

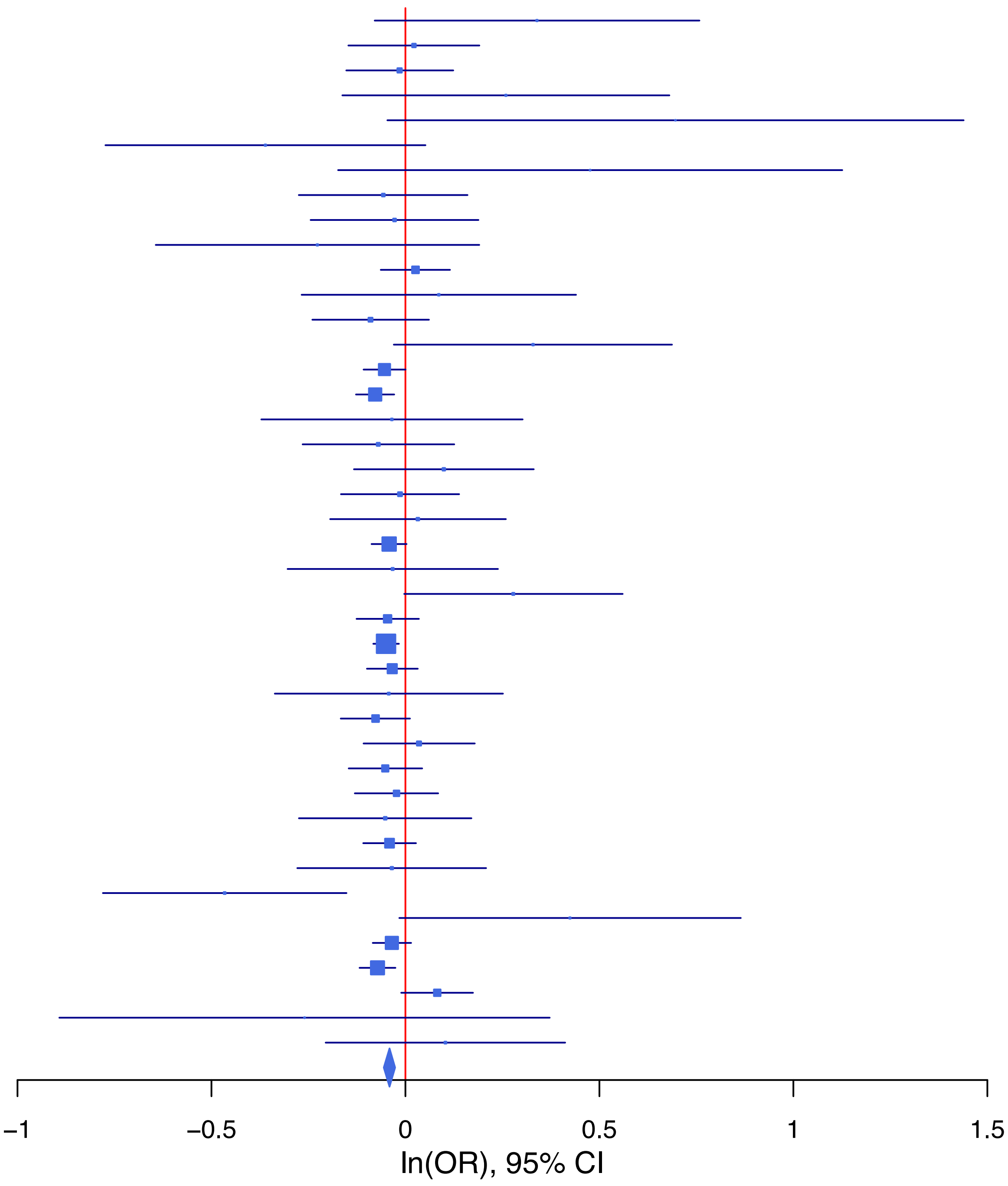

**rs34136150 A/G 5:153382518**

| Cohort | P | ln(OR) | SE |
|---|---|---|---|
| ADHEA_AFR | 0.5687 | 0.094 | 0.166 |
| ADHEA_EUR | 0.0653 | 0.186 | 0.101 |
| ALSPC_EUR | 0.301 | 0.084 | 0.081 |
| BEPS7_EUR | 0.2948 | 0.29 | 0.277 |
| BHRCM_AFR | 0.5211 | 0.162 | 0.252 |
| BHRCM_EUR | 0.1361 | 0.307 | 0.206 |
| BOR17_EUR | 0.2484 | 0.154 | 0.134 |
| BOR2C_EUR | 0.0994 | 0.218 | 0.132 |
| BOR2E_EUR | 0.2945 | 0.276 | 0.263 |
| CNVRG_EAS | 0.51 | −0.091 | 0.139 |
| COGA1_AFR | 0.4103 | 0.118 | 0.144 |
| COGA1_EUR | 0.1593 | 0.129 | 0.091 |
| CUINT_EUR | 0.6388 | −0.054 | 0.114 |
| FINNG_EUR | 0.0049 | 0.091 | 0.032 |
| GEDIS_EUR | 0.8127 | −0.048 | 0.2 |
| GISS1_EUR | 0.3906 | −0.105 | 0.123 |
| GISS2_EUR | 0.9778 | −0.004 | 0.149 |
| GREAT_EAS | 0.4746 | 0.189 | 0.264 |
| GTPRJ_AFR | 0.1413 | 0.117 | 0.079 |
| IPSYC_EUR | 0.0065 | 0.075 | 0.028 |
| JANS3_EUR | 0.5331 | −0.099 | 0.16 |
| JANS4_EUR | 0.2095 | 0.213 | 0.17 |
| JAPAN_EAS | 0.9044 | −0.028 | 0.233 |
| MIREC_AFR | 0.0705 | −0.387 | 0.214 |
| MIREC_EUR | 0.2394 | 0.248 | 0.211 |
| MVPXQ_AFR | 0.8369 | 0.006 | 0.028 |
| MVPXQ_EUR | 0.0029 | 0.056 | 0.019 |
| MVPXQ_LAT | 0.0934 | 0.095 | 0.056 |
| PGCBD_EUR | 0.7497 | 0.013 | 0.04 |
| PGCED_EUR | 0.3638 | 0.169 | 0.186 |
| PGCMD_EUR | 0.2945 | 0.059 | 0.057 |
| PGCPT_AFR | 0.4862 | 0.114 | 0.163 |
| PGCPT_EUR | 0.1332 | 0.128 | 0.085 |
| PGCSZ_EUR | 0.6949 | 0.023 | 0.058 |
| PRFCT_EUR | 0.5175 | 0.043 | 0.066 |
| PSYCR_EUR | 0.7954 | 0.037 | 0.142 |
| QIMRB_EUR | 0.5445 | 0.025 | 0.041 |
| SNUBH−ASA_EAS | 0.6973 | 0.134 | 0.346 |
| STRR1_LAT | 0.278 | 0.323 | 0.298 |
| UKBJC_EUR | 0.2361 | 0.035 | 0.03 |
| UTAH2_EUR | 0.3033 | 0.029 | 0.029 |
| UTAMR_LAT | 0.174 | −0.109 | 0.08 |
| YPENN_AFR | 0.3011 | −0.225 | 0.218 |
| YPENN_EUR | 0.1496 | 0.255 | 0.177 |
| **meta** | **2.13e−08** | **0.05** | **0.009** |

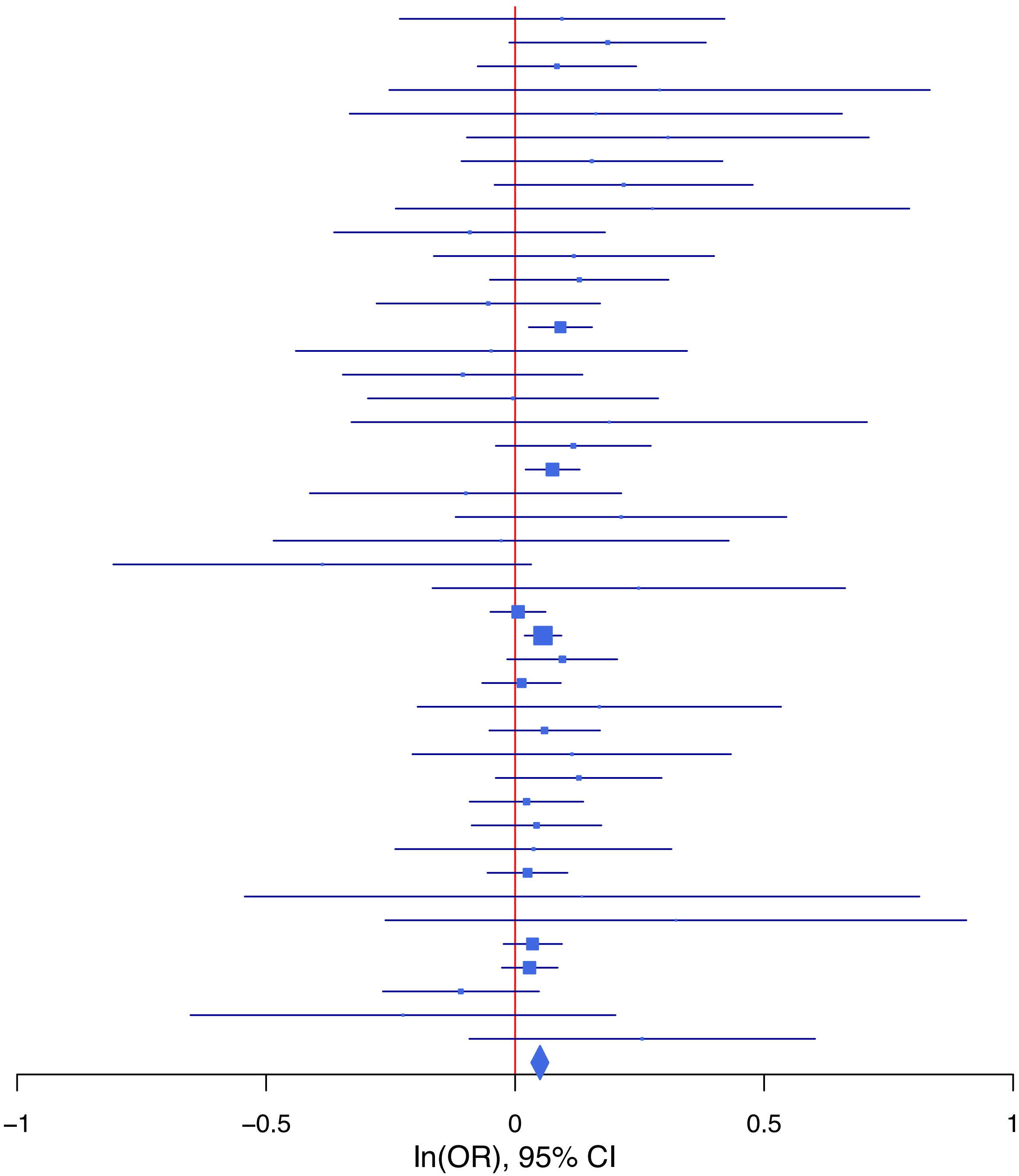

**rs34211110 A/G 12:24222335**

| Cohort | P | ln(OR) | SE |
|---|---|---|---|
| ADHEA_AFR | 0.3471 | −0.137 | 0.146 |
| ADHEA_EUR | 0.3218 | 0.074 | 0.075 |
| ALSPC_EUR | 0.3727 | −0.052 | 0.058 |
| BEPS7_EUR | 0.7888 | 0.053 | 0.199 |
| BHRCM_AFR | 0.1541 | −0.698 | 0.49 |
| BHRCM_EUR | 0.2756 | 0.184 | 0.169 |
| BHRCM_LAT | 0.1637 | 0.504 | 0.362 |
| BOR17_EUR | 0.2694 | −0.105 | 0.095 |
| BOR2C_EUR | 0.2904 | 0.099 | 0.094 |
| BOR2E_EUR | 0.5729 | −0.113 | 0.2 |
| COGA1_AFR | 0.9736 | −0.004 | 0.127 |
| COGA1_EUR | 0.9879 | −0.001 | 0.066 |
| CUINT_EUR | 0.6802 | −0.035 | 0.086 |
| CVEDA_CSA | 0.9703 | 0.01 | 0.275 |
| FINNG_EUR | 0.0078 | −0.071 | 0.027 |
| GEDIS_EUR | 0.9015 | −0.018 | 0.146 |
| GISS1_EUR | 0.0959 | −0.146 | 0.088 |
| GISS2_EUR | 0.2049 | 0.135 | 0.106 |
| GTPRJ_AFR | 0.8766 | −0.011 | 0.072 |
| IPSYC_EUR | 0.0609 | −0.038 | 0.02 |
| JANS3_EUR | 0.8217 | 0.026 | 0.114 |
| JANS4_EUR | 0.8847 | 0.019 | 0.13 |
| MVPXQ_AFR | 0.1554 | −0.035 | 0.025 |
| MVPXQ_EAS | 0.5377 | −0.173 | 0.281 |
| MVPXQ_EUR | 0.0321 | −0.029 | 0.014 |
| MVPXQ_LAT | 0.0134 | −0.101 | 0.041 |
| PGCBD_EUR | 0.0996 | −0.047 | 0.029 |
| PGCED_EUR | 0.2282 | −0.161 | 0.134 |
| PGCMD_EUR | 0.8994 | 0.005 | 0.038 |
| PGCPT_AFR | 0.9729 | −0.004 | 0.124 |
| PGCPT_EUR | 0.9939 | 0.001 | 0.064 |
| PGCSZ_EUR | 0.4436 | −0.032 | 0.041 |
| PRFCT_EUR | 0.5416 | −0.03 | 0.049 |
| PSYCR_EUR | 0.0895 | −0.162 | 0.096 |
| QIMRB_EUR | 0.2196 | −0.036 | 0.029 |
| STRR1_LAT | 0.8175 | 0.049 | 0.211 |
| UKBJC_EUR | 0.0594 | −0.04 | 0.021 |
| UTAH2_EUR | 8e−04 | −0.07 | 0.021 |
| UTAMR_LAT | 0.1719 | −0.063 | 0.046 |
| YPENN_AFR | 0.2356 | −0.225 | 0.19 |
| YPENN_EUR | 0.7853 | −0.039 | 0.143 |
| **meta** | **1.12e−09** | **−0.04** | **0.007** |

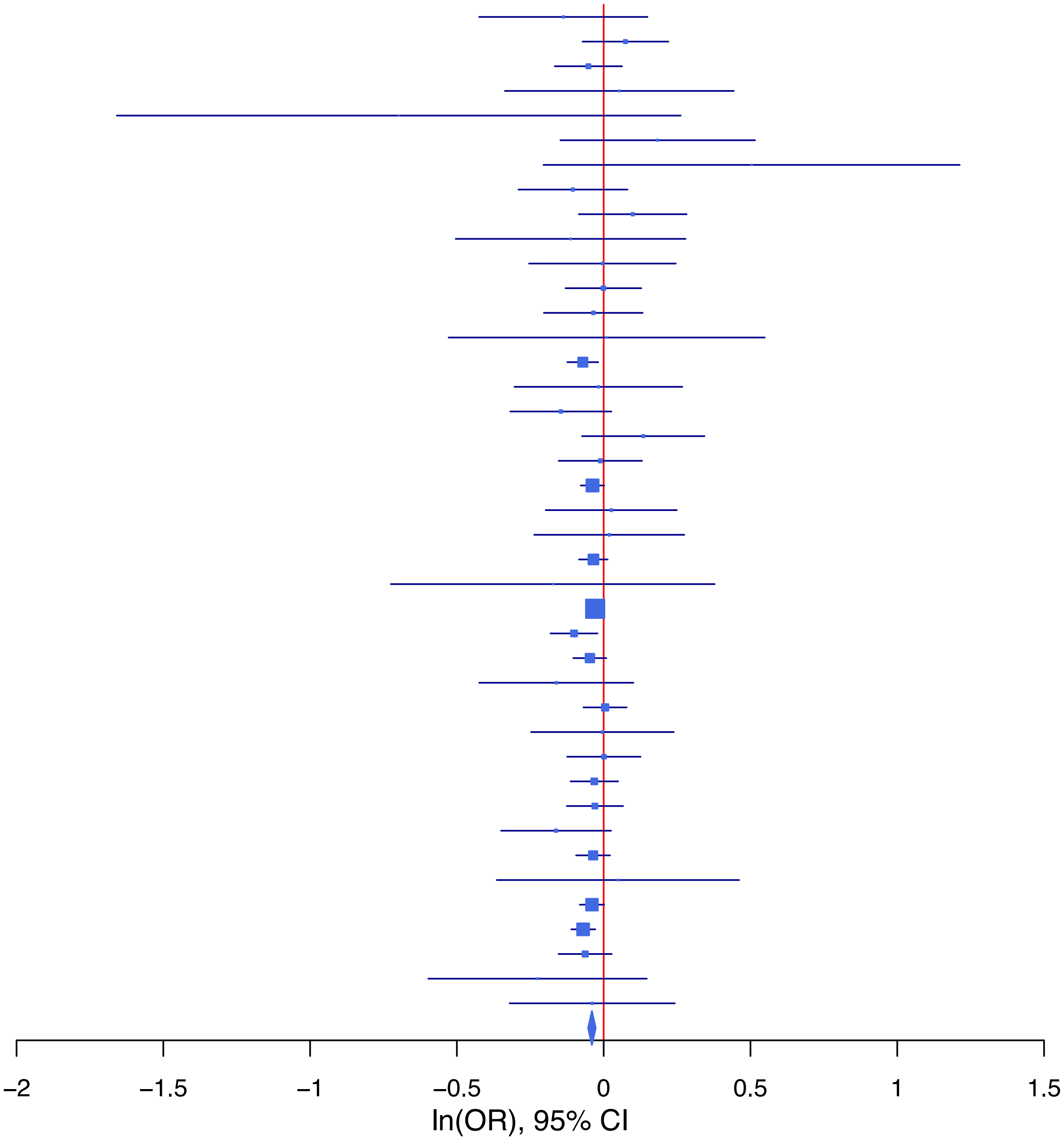

**rs35111904 A/C 4:153068837**

| Cohort | P | ln(OR) | SE |
|---|---|---|---|
| ALSPC_EUR | 0.1797 | −0.073 | 0.054 |
| BEPS7_EUR | 0.7972 | −0.046 | 0.177 |
| BHRCM_AFR | 0.3713 | 0.204 | 0.228 |
| BHRCM_EUR | 0.6853 | −0.063 | 0.156 |
| BOR17_EUR | 0.7017 | −0.032 | 0.084 |
| BOR2C_EUR | 0.9899 | −0.001 | 0.086 |
| BOR2E_EUR | 0.9561 | −0.01 | 0.176 |
| CNVRG_EAS | 0.2737 | −0.239 | 0.218 |
| COGA1_AFR | 0.5793 | 0.068 | 0.123 |
| COGA1_EUR | 0.4702 | −0.043 | 0.059 |
| CUINT_EUR | 0.6613 | 0.033 | 0.076 |
| CVEDA_CSA | 0.6854 | 0.073 | 0.18 |
| ESTB2_EUR | 0.1074 | 0.036 | 0.023 |
| FINNG_EUR | 0.7377 | −0.008 | 0.023 |
| GEDIS_EUR | 0.6143 | −0.066 | 0.131 |
| GISS1_EUR | 0.1105 | 0.128 | 0.08 |
| GISS2_EUR | 0.0305 | 0.204 | 0.094 |
| GTPRJ_AFR | 0.4119 | −0.061 | 0.074 |
| IPSYC_EUR | 0.0847 | 0.032 | 0.018 |
| JANS3_EUR | 0.4392 | 0.079 | 0.102 |
| JANS4_EUR | 0.6452 | −0.053 | 0.116 |
| MVPXQ_AFR | 0.1336 | 0.041 | 0.027 |
| MVPXQ_EAS | 0.8839 | 0.031 | 0.213 |
| MVPXQ_EUR | 0.0631 | 0.024 | 0.013 |
| MVPXQ_LAT | 0.0129 | 0.096 | 0.039 |
| PGCBD_EUR | 0.7059 | 0.01 | 0.026 |
| PGCED_EUR | 0.6926 | 0.046 | 0.116 |
| PGCMD_EUR | 0.6374 | 0.016 | 0.034 |
| PGCPT_AFR | 0.6622 | −0.056 | 0.128 |
| PGCPT_EUR | 0.5402 | 0.035 | 0.058 |
| PGCSZ_EUR | 0.246 | 0.043 | 0.037 |
| PRFCT_EUR | 0.0484 | 0.087 | 0.044 |
| PSYCR_EUR | 0.3296 | 0.085 | 0.087 |
| QIMRB_EUR | 0.0452 | 0.055 | 0.027 |
| STRR1_LAT | 0.8812 | −0.03 | 0.2 |
| UKBJC_EUR | 0.0085 | 0.052 | 0.02 |
| UTAH2_EUR | 0.0016 | 0.059 | 0.019 |
| UTAMR_LAT | 0.67 | 0.02 | 0.046 |
| YPENN_EUR | 0.8025 | 0.032 | 0.129 |
| **meta** | **4.06e−08** | **0.033** | **0.006** |

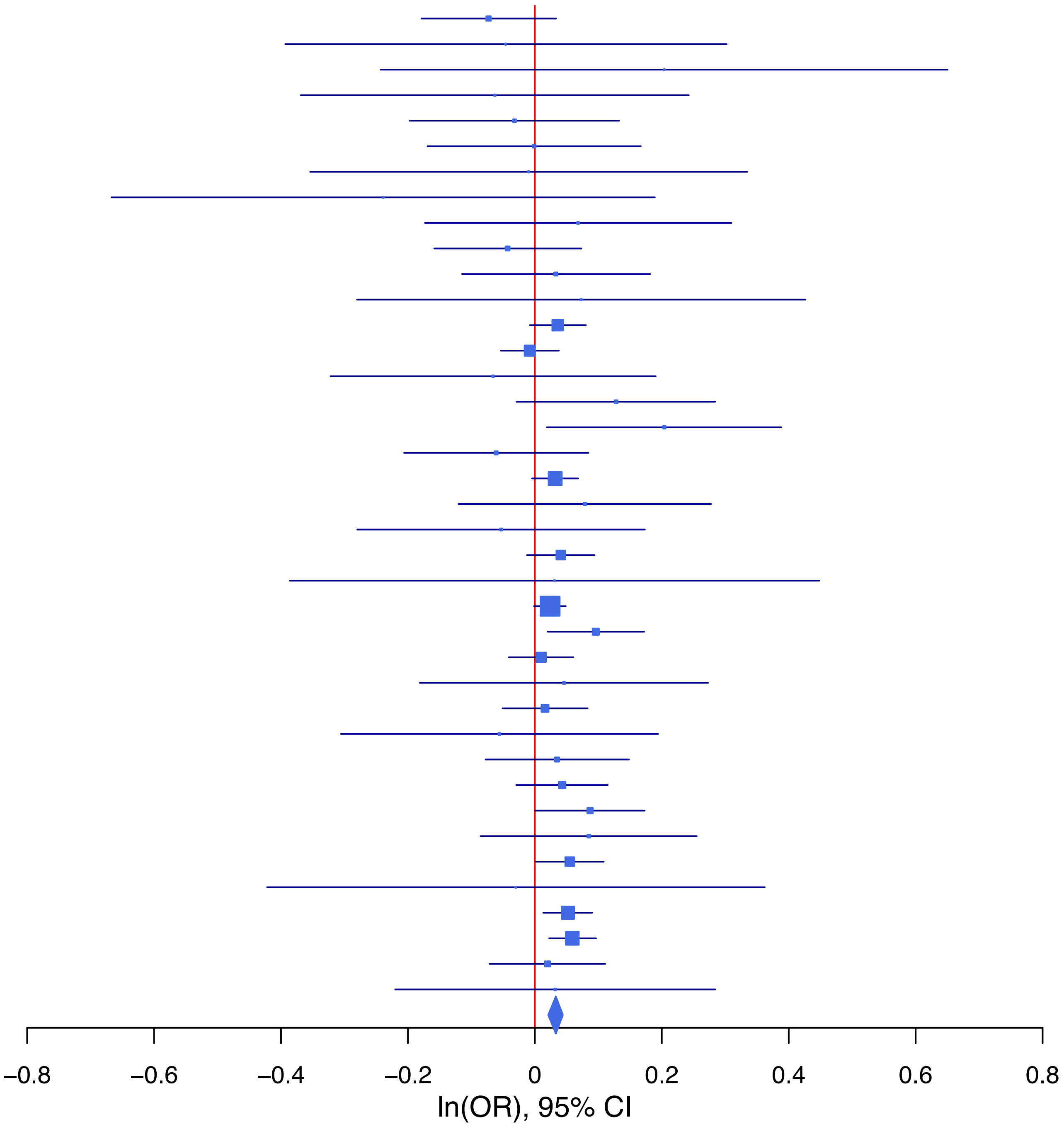

**rs4479021 A/G 11:113383394**

| Cohort | P | ln(OR) | SE |
|---|---|---|---|
| ADHEA_AFR | 0.9678 | 0.005 | 0.114 |
| ADHEA_EUR | 0.1202 | −0.108 | 0.069 |
| ALSPC_EUR | 0.3336 | −0.052 | 0.054 |
| BEPS7_EUR | 0.8427 | −0.036 | 0.181 |
| BHRCM_AFR | 0.7182 | −0.07 | 0.194 |
| BHRCM_EUR | 0.9541 | 0.009 | 0.162 |
| BOR17_EUR | 0.7436 | −0.028 | 0.085 |
| BOR2C_EUR | 0.2191 | −0.106 | 0.087 |
| BOR2E_EUR | 0.9429 | −0.013 | 0.185 |
| CNVRG_EAS | 0.9733 | 0.003 | 0.085 |
| COGA1_AFR | 0.6184 | −0.05 | 0.101 |
| COGA1_EUR | 0.0996 | −0.098 | 0.059 |
| CUINT_EUR | 0.6335 | 0.036 | 0.075 |
| CVEDA_CSA | 0.5093 | 0.119 | 0.18 |
| ESTB2_EUR | 0.0718 | −0.043 | 0.024 |
| FINNG_EUR | 0.0222 | −0.06 | 0.026 |
| GEDIS_EUR | 0.3271 | −0.13 | 0.133 |
| GISS1_EUR | 0.6603 | 0.035 | 0.08 |
| GISS2_EUR | 0.7219 | 0.033 | 0.092 |
| GREAT_EAS | 0.8846 | −0.022 | 0.149 |
| GTPRJ_AFR | 0.9723 | −0.002 | 0.059 |
| IPSYC_EUR | 0.0762 | −0.033 | 0.019 |
| JANS3_EUR | 0.3542 | −0.097 | 0.104 |
| JANS4_EUR | 0.4967 | 0.081 | 0.119 |
| JAPAN_EAS | 0.7217 | −0.03 | 0.084 |
| MVPXQ_AFR | 0.0087 | −0.056 | 0.021 |
| MVPXQ_EAS | 0.4108 | −0.116 | 0.141 |
| MVPXQ_EUR | 4.23e−05 | −0.051 | 0.012 |
| MVPXQ_LAT | 0.1174 | −0.052 | 0.033 |
| PGCBD_EUR | 0.0457 | −0.052 | 0.026 |
| PGCED_EUR | 0.0895 | −0.194 | 0.114 |
| PGCMD_EUR | 0.4777 | 0.024 | 0.034 |
| PGCPT_AFR | 0.2034 | 0.132 | 0.104 |
| PGCPT_EUR | 0.606 | −0.03 | 0.058 |
| PGCSZ_EUR | 0.1241 | −0.058 | 0.038 |
| PRFCT_EUR | 0.5056 | −0.03 | 0.045 |
| PSYCR_EUR | 0.0058 | 0.24 | 0.087 |
| QIMRB_EUR | 0.3692 | −0.025 | 0.027 |
| SNUBH−ASA_EAS | 0.8072 | −0.051 | 0.21 |
| SNUBH−KCHIP_EAS | 0.2332 | −0.329 | 0.276 |
| STRR1_LAT | 0.0103 | 0.446 | 0.174 |
| UKBJC_EUR | 0.2474 | −0.023 | 0.02 |
| UTAH2_EUR | 0.696 | −0.008 | 0.019 |
| UTAMR_LAT | 0.759 | −0.012 | 0.039 |
| YPENN_AFR | 0.3962 | 0.125 | 0.147 |
| YPENN_EUR | 0.4659 | −0.095 | 0.13 |
| **meta** | **1.11e−09** | **−0.035** | **0.006** |

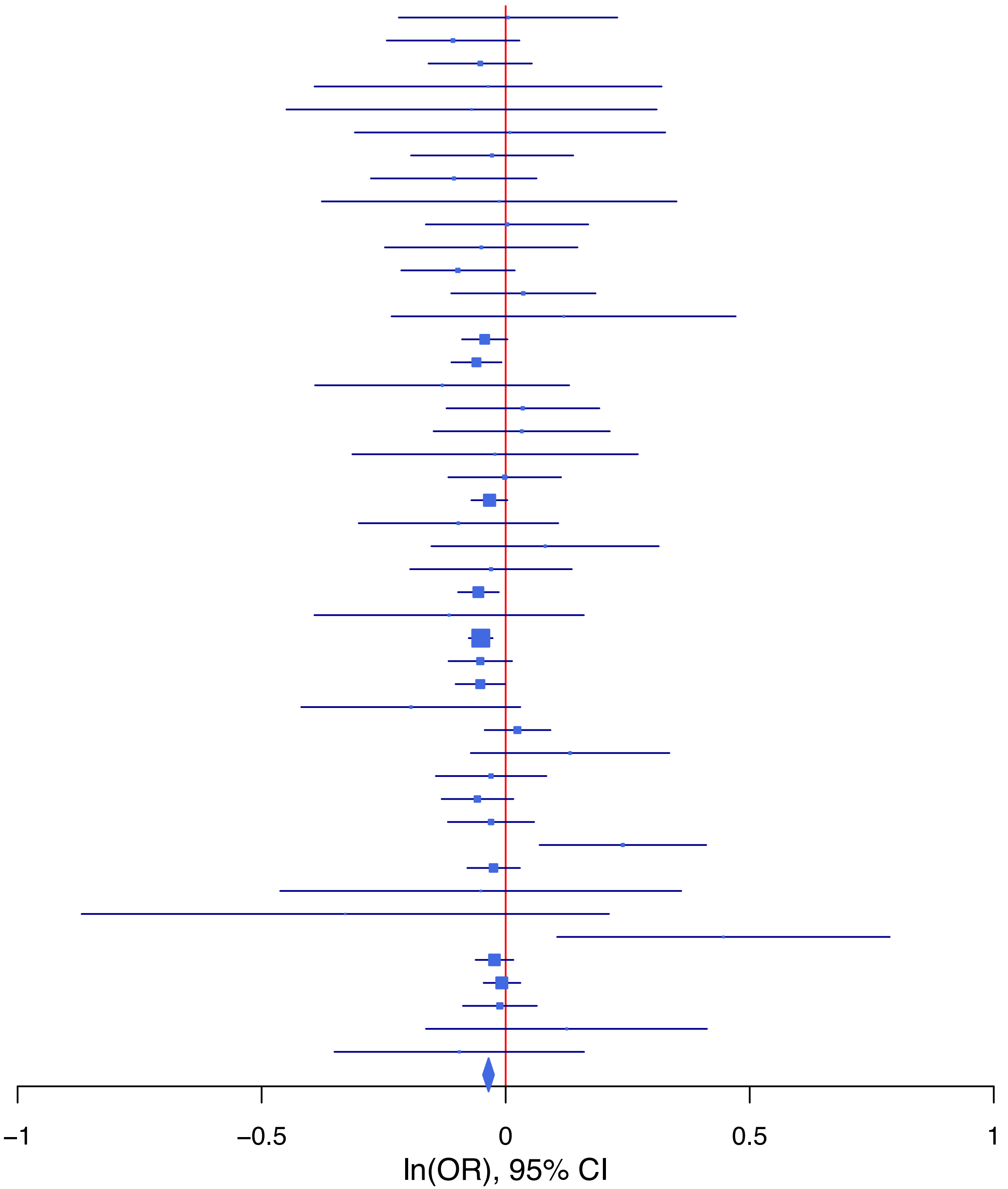

**rs4632782 G/T 5:92588151**

| Cohort | P | ln(OR) | SE |
|---|---|---|---|
| BEPS7_EUR | 0.0394 | 0.512 | 0.249 |
| BHRCM_AFR | 0.1556 | 0.298 | 0.21 |
| BHRCM_EUR | 0.7663 | 0.052 | 0.174 |
| BHRCM_LAT | 0.4086 | 0.428 | 0.518 |
| BOR17_EUR | 0.278 | −0.116 | 0.107 |
| BOR2C_EUR | 0.4194 | −0.089 | 0.111 |
| BOR2E_EUR | 0.4925 | −0.156 | 0.228 |
| CNVRG_EAS | 0.0975 | 0.148 | 0.09 |
| COGA1_EUR | 0.0456 | 0.133 | 0.066 |
| CVEDA_CSA | 0.5458 | −0.107 | 0.176 |
| ESTB2_EUR | 0.5479 | −0.016 | 0.026 |
| FINNG_EUR | 0.0246 | 0.055 | 0.024 |
| GEDIS_EUR | 0.4581 | −0.107 | 0.144 |
| GISS1_EUR | 0.5029 | 0.06 | 0.089 |
| GISS2_EUR | 0.5828 | 0.063 | 0.115 |
| GREAT_EAS | 0.5625 | −0.074 | 0.128 |
| GTPRJ_AFR | 0.5101 | 0.042 | 0.064 |
| IPSYC_EUR | 0.0141 | 0.057 | 0.023 |
| JANS3_EUR | 0.4352 | 0.103 | 0.133 |
| JANS4_EUR | 0.4314 | −0.125 | 0.159 |
| MVPXQ_AFR | 0.006 | 0.058 | 0.021 |
| MVPXQ_EAS | 0.1105 | 0.208 | 0.13 |
| MVPXQ_EUR | 0.0311 | 0.03 | 0.014 |
| MVPXQ_LAT | 0.2469 | 0.042 | 0.036 |
| PGCBD_EUR | 0.0013 | 0.102 | 0.032 |
| PGCED_EUR | 0.0036 | 0.472 | 0.162 |
| PGCMD_EUR | 0.002 | 0.134 | 0.043 |
| PGCPT_AFR | 0.558 | 0.066 | 0.113 |
| PGCPT_EUR | 0.1217 | 0.11 | 0.071 |
| PGCSZ_EUR | 0.473 | 0.032 | 0.045 |
| PRFCT_EUR | 0.8491 | 0.01 | 0.054 |
| PSYCR_EUR | 0.3965 | −0.095 | 0.112 |
| QIMRB_EUR | 0.1967 | 0.043 | 0.034 |
| SNUBH−ASA_EAS | 0.0389 | −0.443 | 0.215 |
| SNUBH−KCHIP_EAS | 0.8137 | 0.063 | 0.268 |
| UKBJC_EUR | 0.2796 | 0.024 | 0.022 |
| UTAH2_EUR | 0.088 | 0.036 | 0.021 |
| UTAMR_LAT | 0.2548 | −0.049 | 0.043 |
| **meta** | **1.14e−09** | **0.04** | **0.007** |

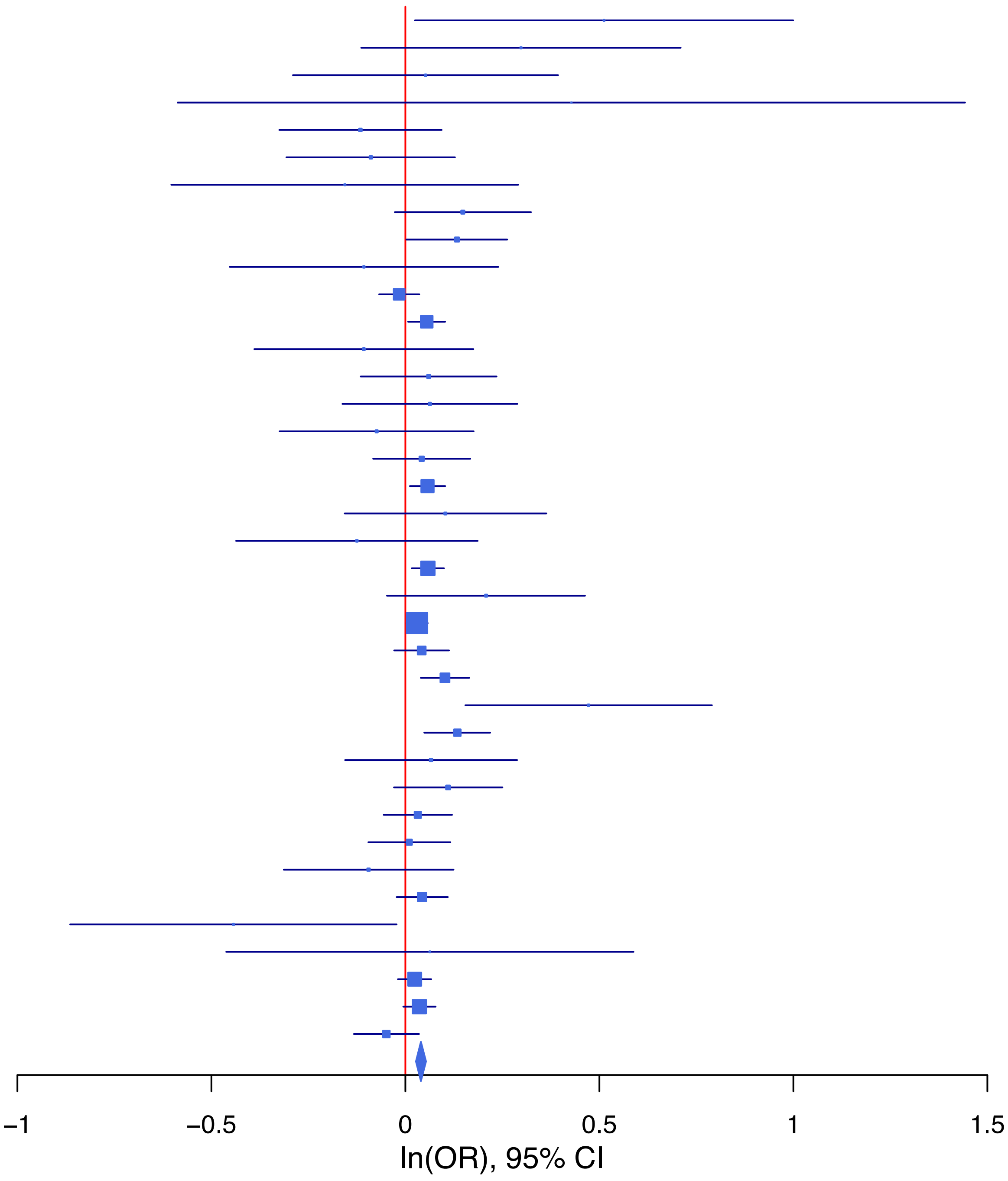

**rs4771936 A/G 13:97005377**

| Cohort | P | ln(OR) | SE |
|---|---|---|---|
| ADHEA_AFR | 0.0135 | 0.287 | 0.116 |
| ADHEA_EUR | 0.2897 | 0.075 | 0.071 |
| ALSPC_EUR | 0.3166 | −0.058 | 0.058 |
| BEPS7_EUR | 0.3375 | −0.174 | 0.182 |
| BHRCM_AFR | 0.8785 | −0.029 | 0.19 |
| BOR17_EUR | 0.0057 | 0.245 | 0.089 |
| BOR2C_EUR | 0.6238 | 0.044 | 0.09 |
| BOR2E_EUR | 0.1666 | −0.261 | 0.189 |
| CNVRG_EAS | 0.0385 | 0.097 | 0.047 |
| COGA1_AFR | 0.7481 | −0.032 | 0.1 |
| COGA1_EUR | 0.8334 | 0.013 | 0.061 |
| CUINT_EUR | 0.1653 | −0.11 | 0.079 |
| CVEDA_CSA | 0.3391 | 0.153 | 0.16 |
| ESTB2_EUR | 0.0018 | 0.072 | 0.023 |
| GISS2_EUR | 0.4909 | −0.065 | 0.095 |
| GREAT_EAS | 0.2224 | −0.099 | 0.082 |
| GTPRJ_AFR | 0.064 | −0.111 | 0.06 |
| IPSYC_EUR | 0.2137 | 0.024 | 0.02 |
| JANS3_EUR | 0.9891 | −0.001 | 0.109 |
| JANS4_EUR | 0.37 | −0.114 | 0.127 |
| JAPAN_EAS | 0.3577 | 0.051 | 0.055 |
| MVPXQ_EAS | 0.8788 | 0.015 | 0.101 |
| MVPXQ_EUR | 0.0267 | 0.029 | 0.013 |
| MVPXQ_LAT | 0.12 | 0.058 | 0.038 |
| PGCBD_EUR | 0.109 | 0.043 | 0.027 |
| PGCED_EUR | 0.2925 | −0.128 | 0.122 |
| PGCMD_EUR | 0.3761 | 0.032 | 0.036 |
| PGCPT_EUR | 0.0522 | 0.116 | 0.06 |
| PGCSZ_EUR | 0.4647 | 0.029 | 0.039 |
| PRFCT_EUR | 0.0632 | 0.086 | 0.046 |
| PSYCR_EUR | 0.9631 | 0.004 | 0.092 |
| QIMRB_EUR | 0.0432 | 0.059 | 0.029 |
| SNUBH−ASA_EAS | 0.1048 | 0.207 | 0.128 |
| SNUBH−KCHIP_EAS | 0.7728 | 0.048 | 0.167 |
| STRR1_LAT | 0.1259 | 0.3 | 0.196 |
| UKBJC_EUR | 0.002 | 0.064 | 0.021 |
| UTAH2_EUR | 0.0133 | 0.049 | 0.02 |
| UTAMR_LAT | 0.0289 | 0.107 | 0.049 |
| YPENN_EUR | 0.8211 | −0.031 | 0.135 |
| **meta** | **7.79e−11** | **0.042** | **0.006** |

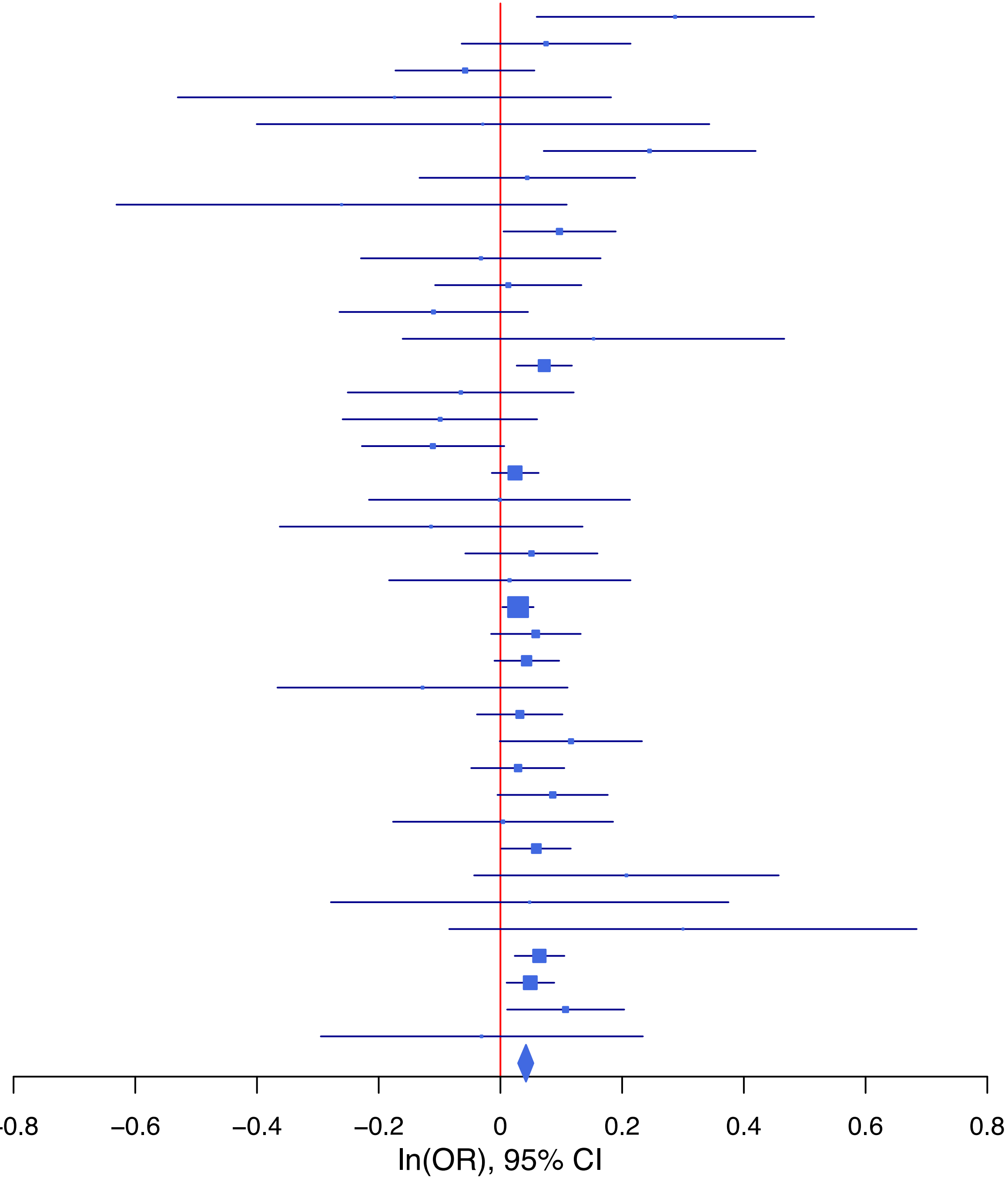

**rs4776718 G/C 15:66369961**

| Cohort | P | ln(OR) | SE |
|---|---|---|---|
| ADHEA_AFR | 0.9144 | −0.017 | 0.159 |
| ADHEA_EUR | 0.2049 | −0.11 | 0.087 |
| ALSPC_EUR | 0.1565 | 0.094 | 0.067 |
| BEPS7_EUR | 0.721 | 0.085 | 0.237 |
| BHRCM_AFR | 0.9229 | −0.026 | 0.272 |
| BOR17_EUR | 0.825 | −0.023 | 0.103 |
| BOR2C_EUR | 0.2275 | 0.133 | 0.11 |
| BOR2E_EUR | 0.7565 | −0.062 | 0.199 |
| CNVRG_EAS | 0.2683 | 0.051 | 0.046 |
| COGA1_AFR | 0.4068 | 0.115 | 0.138 |
| COGA1_EUR | 0.276 | 0.079 | 0.073 |
| CUINT_EUR | 0.5473 | 0.054 | 0.089 |
| CVEDA_CSA | 0.7718 | 0.058 | 0.202 |
| ESTB2_EUR | 0.1235 | 0.043 | 0.028 |
| FINNG_EUR | 0.4451 | −0.022 | 0.028 |
| GISS2_EUR | 0.2696 | 0.123 | 0.112 |
| GREAT_EAS | 0.061 | 0.169 | 0.09 |
| IPSYC_EUR | 0.3901 | 0.02 | 0.023 |
| JANS3_EUR | 0.12 | 0.193 | 0.124 |
| JANS4_EUR | 0.225 | −0.192 | 0.159 |
| JAPAN_EAS | 0.1704 | 0.076 | 0.055 |
| MVPXQ_AFR | 0.0512 | 0.056 | 0.029 |
| MVPXQ_EAS | 0.5345 | −0.068 | 0.11 |
| MVPXQ_EUR | 5e−04 | 0.054 | 0.015 |
| MVPXQ_LAT | 0.4005 | 0.035 | 0.041 |
| PGCBD_EUR | 0.3688 | 0.029 | 0.032 |
| PGCED_EUR | 0.1664 | 0.186 | 0.135 |
| PGCMD_EUR | 0.7978 | 0.011 | 0.043 |
| PGCPT_AFR | 0.7529 | −0.044 | 0.139 |
| PGCPT_EUR | 0.7264 | −0.025 | 0.071 |
| PGCSZ_EUR | 0.0187 | 0.106 | 0.045 |
| PRFCT_EUR | 0.0379 | 0.111 | 0.054 |
| PSYCR_EUR | 0.7862 | 0.03 | 0.112 |
| QIMRB_EUR | 0.3393 | 0.033 | 0.034 |
| SNUBH−ASA_EAS | 0.2981 | −0.132 | 0.126 |
| SNUBH−KCHIP_EAS | 0.7535 | −0.052 | 0.166 |
| STRR1_LAT | 0.9081 | −0.026 | 0.229 |
| UKBJC_EUR | 0.0191 | 0.057 | 0.024 |
| UTAH2_EUR | 0.1803 | 0.031 | 0.023 |
| UTAMR_LAT | 0.4611 | 0.034 | 0.047 |
| YPENN_EUR | 0.3411 | −0.161 | 0.169 |
| **meta** | **1.17e−08** | **0.04** | **0.007** |

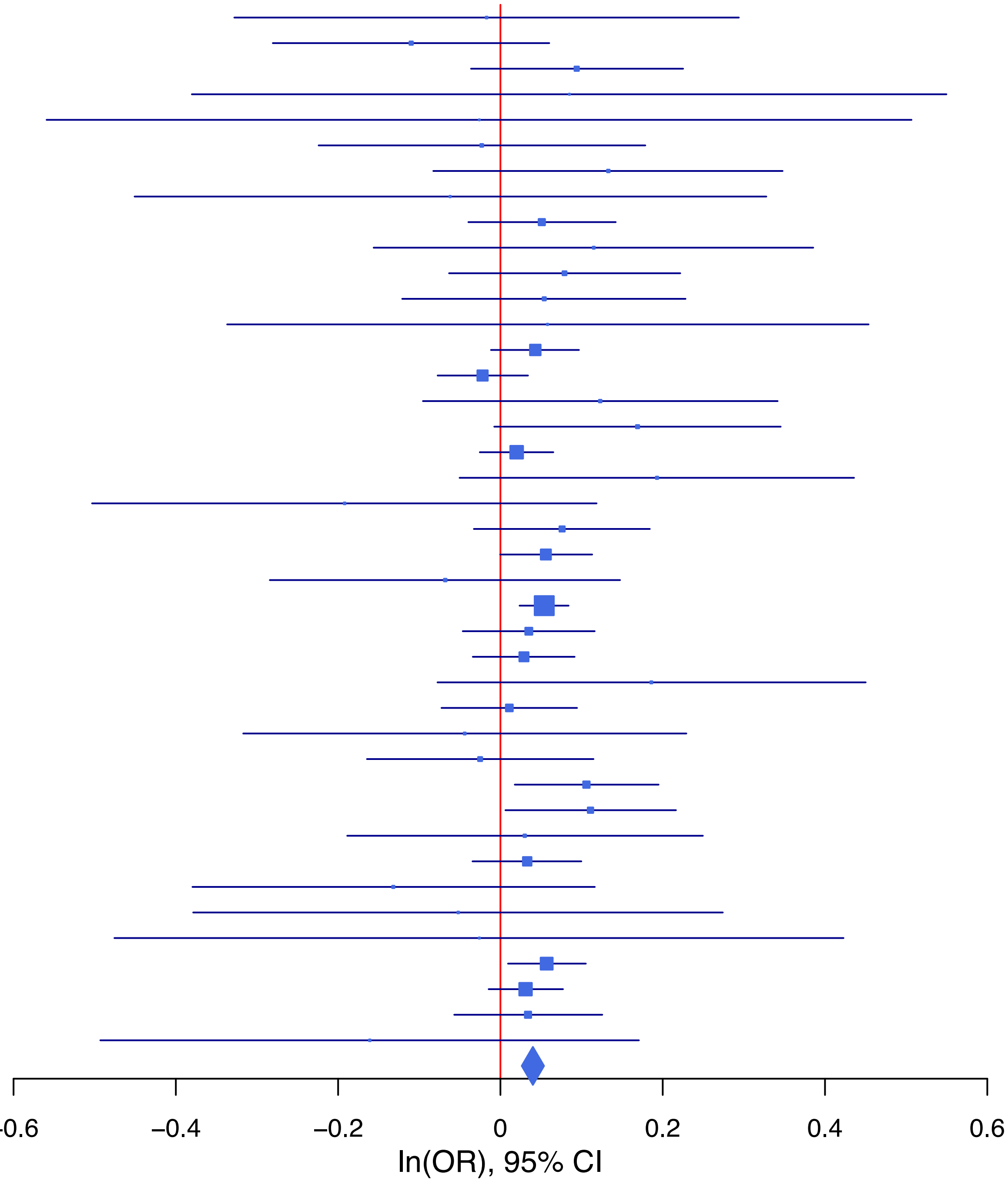

**rs4870063 C/A 6:152244647**

| Cohort | P | ln(OR) | SE |
|---|---|---|---|
| BEPS7_EUR | 0.4874 | 0.129 | 0.186 |
| BHRCM_AFR | 0.9257 | 0.022 | 0.235 |
| BHRCM_EUR | 0.8904 | 0.025 | 0.176 |
| BHRCM_LAT | 0.1375 | −0.381 | 0.257 |
| BOR17_EUR | 0.2816 | 0.098 | 0.091 |
| BOR2C_EUR | 0.5715 | 0.051 | 0.09 |
| BOR2E_EUR | 0.1557 | 0.267 | 0.188 |
| CNVRG_EAS | 0.2714 | 0.064 | 0.058 |
| COGA1_AFR | 0.9477 | −0.007 | 0.109 |
| COGA1_EUR | 0.2317 | 0.074 | 0.062 |
| CUINT_EUR | 0.1798 | −0.106 | 0.079 |
| CVEDA_CSA | 0.9159 | 0.018 | 0.166 |
| ESTB2_EUR | 0.0017 | 0.077 | 0.024 |
| FINNG_EUR | 0.3476 | 0.023 | 0.024 |
| GEDIS_EUR | 0.2779 | 0.146 | 0.134 |
| GISS1_EUR | 0.1173 | −0.14 | 0.09 |
| GISS2_EUR | 0.9328 | −0.008 | 0.099 |
| GREAT_EAS | 0.0171 | 0.233 | 0.098 |
| GTPRJ_AFR | 0.3077 | 0.07 | 0.069 |
| IPSYC_EUR | 0.9672 | 0.001 | 0.019 |
| JANS3_EUR | 0.1333 | −0.168 | 0.112 |
| JANS4_EUR | 0.7978 | −0.033 | 0.129 |
| MVPXQ_EUR | 2.11e−05 | 0.056 | 0.013 |
| MVPXQ_LAT | 0.0018 | 0.104 | 0.033 |
| PGCBD_EUR | 0.1681 | 0.038 | 0.027 |
| PGCED_EUR | 0.0473 | −0.252 | 0.127 |
| PGCMD_EUR | 0.123 | 0.056 | 0.036 |
| PGCPT_AFR | 0.0613 | 0.227 | 0.121 |
| PGCPT_EUR | 0.3291 | 0.059 | 0.06 |
| PGCSZ_EUR | 0.5284 | 0.024 | 0.039 |
| PRFCT_EUR | 0.0011 | 0.15 | 0.046 |
| PSYCR_EUR | 0.0842 | 0.156 | 0.09 |
| QIMRB_EUR | 0.1849 | 0.038 | 0.028 |
| SNUBH−ASA_EAS | 0.2648 | 0.169 | 0.151 |
| SNUBH−KCHIP_EAS | 0.6746 | −0.084 | 0.2 |
| STRR1_LAT | 0.0061 | 0.498 | 0.182 |
| UKBJC_EUR | 3.88e−05 | 0.083 | 0.02 |
| UTAH2_EUR | 0.4813 | 0.014 | 0.02 |
| UTAMR_LAT | 0.0058 | 0.104 | 0.038 |
| YPENN_AFR | 0.5527 | 0.103 | 0.173 |
| YPENN_EUR | 0.8904 | −0.018 | 0.132 |
| **meta** | **7.70e−15** | **0.049** | **0.006** |

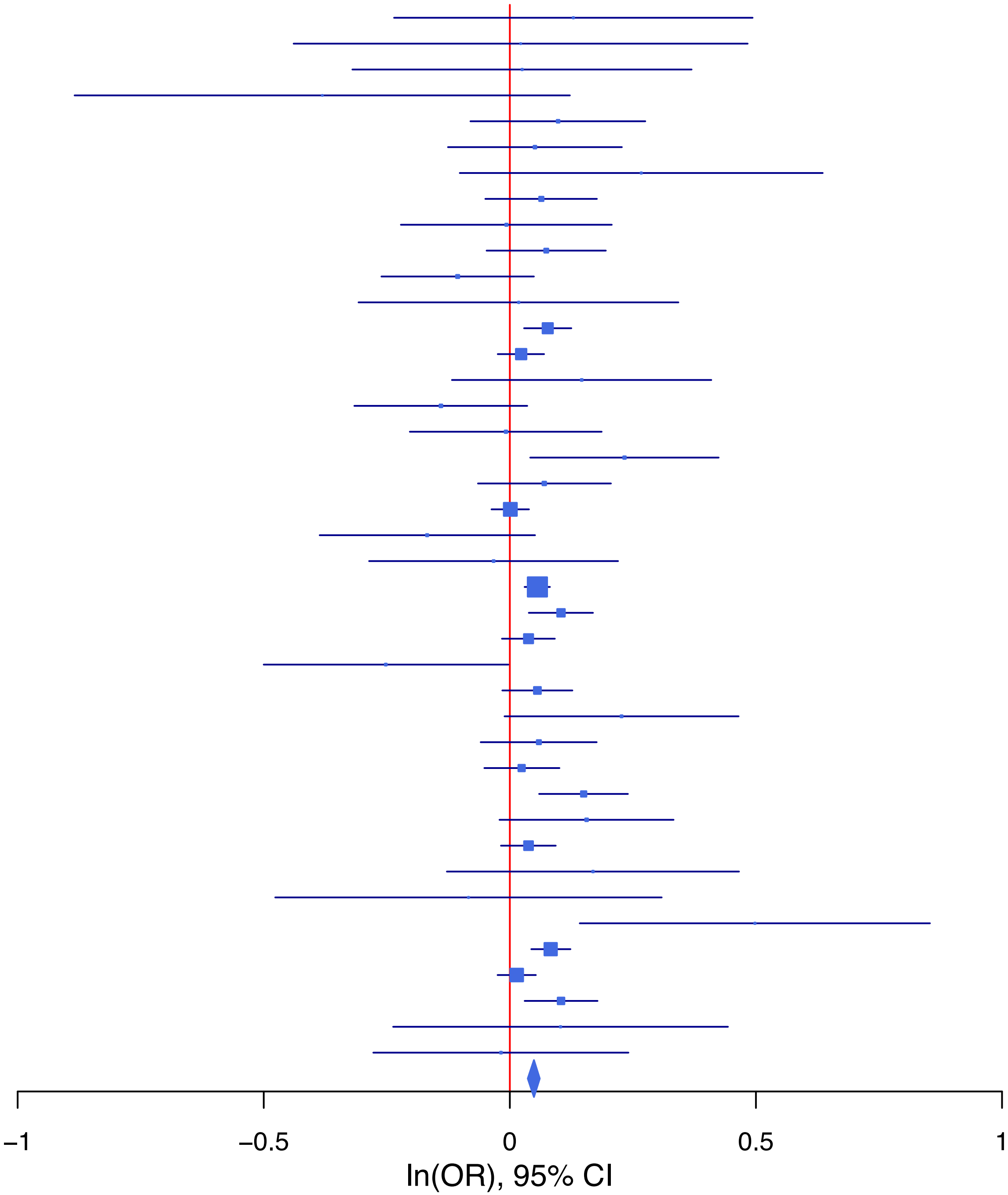

**rs502621 C/T 1:38236393**

| Cohort | P | ln(OR) | SE |
|---|---|---|---|
| ALSPC_EUR | 0.4069 | 0.044 | 0.054 |
| BEPS7_EUR | 0.4735 | −0.128 | 0.179 |
| BHRCM_AFR | 0.9077 | −0.037 | 0.323 |
| BHRCM_EUR | 0.8849 | −0.022 | 0.154 |
| BHRCM_LAT | 0.2938 | 0.268 | 0.255 |
| BOR17_EUR | 0.7616 | 0.026 | 0.087 |
| BOR2C_EUR | 0.6859 | 0.035 | 0.087 |
| BOR2E_EUR | 0.5939 | 0.096 | 0.18 |
| CNVRG_EAS | 0.4568 | −0.044 | 0.059 |
| COGA1_AFR | 0.7814 | 0.034 | 0.124 |
| COGA1_EUR | 0.5057 | 0.039 | 0.059 |
| CUINT_EUR | 0.4525 | 0.055 | 0.073 |
| CVEDA_CSA | 0.2054 | −0.236 | 0.187 |
| ESTB2_EUR | 0.1545 | −0.032 | 0.023 |
| FINNG_EUR | 0.031 | −0.049 | 0.023 |
| GEDIS_EUR | 0.5399 | −0.079 | 0.13 |
| GISS1_EUR | 0.4053 | −0.066 | 0.079 |
| GISS2_EUR | 0.3199 | 0.093 | 0.094 |
| GREAT_EAS | 0.6106 | 0.052 | 0.1 |
| GTPRJ_AFR | 0.0827 | 0.127 | 0.073 |
| IPSYC_EUR | 0.0316 | −0.04 | 0.018 |
| JANS3_EUR | 0.9576 | 0.006 | 0.104 |
| JANS4_EUR | 0.4872 | 0.084 | 0.12 |
| JAPAN_EAS | 0.042 | −0.138 | 0.068 |
| MIREC_AFR | 0.1009 | −0.307 | 0.187 |
| MIREC_EUR | 0.0683 | 0.28 | 0.154 |
| MVPXQ_AFR | 0.97 | 0.001 | 0.026 |
| MVPXQ_EAS | 0.3028 | −0.126 | 0.122 |
| MVPXQ_EUR | 1e−04 | −0.048 | 0.012 |
| MVPXQ_LAT | 0.1556 | −0.048 | 0.034 |
| PGCBD_EUR | 0.0187 | −0.061 | 0.026 |
| PGCED_EUR | 0.2044 | −0.144 | 0.114 |
| PGCMD_EUR | 0.2456 | −0.041 | 0.035 |
| PGCPT_AFR | 0.5538 | 0.088 | 0.149 |
| PGCPT_EUR | 0.7292 | 0.02 | 0.058 |
| PGCSZ_EUR | 0.9036 | −0.005 | 0.037 |
| PRFCT_EUR | 3e−04 | −0.158 | 0.044 |
| PSYCR_EUR | 0.216 | −0.109 | 0.088 |
| QIMRB_EUR | 0.4951 | −0.019 | 0.027 |
| SNUBH−ASA_EAS | 0.5868 | −0.08 | 0.147 |
| STRR1_LAT | 0.2487 | 0.2 | 0.173 |
| UKBJC_EUR | 0.0364 | −0.041 | 0.02 |
| UTAH2_EUR | 0.0013 | −0.061 | 0.019 |
| UTAMR_LAT | 0.7703 | −0.011 | 0.037 |
| YPENN_AFR | 0.9458 | 0.012 | 0.182 |
| YPENN_EUR | 0.4402 | 0.099 | 0.129 |
| **meta** | **9.78e−11** | **−0.037** | **0.006** |

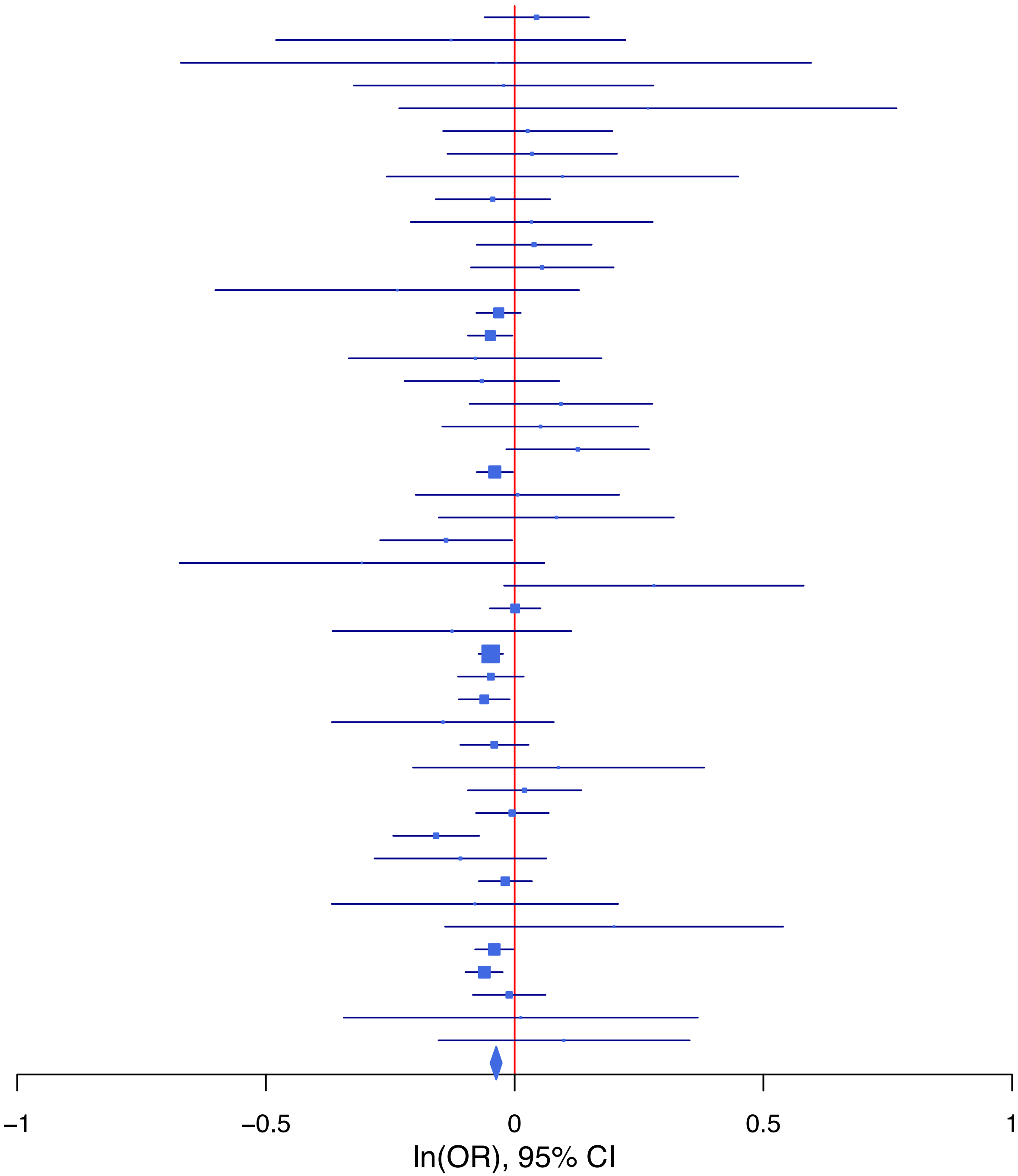

**rs55995895 T/C 7:1862183**

| Cohort | P | ln(OR) | SE |
|---|---|---|---|
| ADHEA_AFR | 0.6235 | 0.114 | 0.234 |
| ADHEA_EUR | 0.2072 | −0.121 | 0.096 |
| ALSPC_EUR | 0.7761 | 0.022 | 0.077 |
| BEPS7_EUR | 0.2763 | −0.254 | 0.233 |
| BOR17_EUR | 0.8961 | 0.014 | 0.11 |
| BOR2C_EUR | 0.4235 | −0.088 | 0.11 |
| BOR2E_EUR | 0.4332 | −0.183 | 0.234 |
| CNVRG_EAS | 0.6745 | 0.032 | 0.077 |
| COGA1_EUR | 0.1027 | −0.132 | 0.081 |
| CUINT_EUR | 0.626 | 0.048 | 0.098 |
| CVEDA_CSA | 0.9291 | 0.016 | 0.185 |
| ESTB2_EUR | 0.0091 | −0.07 | 0.027 |
| FINNG_EUR | 8e−04 | −0.09 | 0.027 |
| GISS1_EUR | 0.7848 | 0.025 | 0.091 |
| GISS2_EUR | 0.7959 | −0.027 | 0.103 |
| GREAT_EAS | 0.4212 | −0.113 | 0.141 |
| GTPRJ_AFR | 0.1375 | −0.217 | 0.146 |
| IPSYC_EUR | 0.0025 | −0.077 | 0.025 |
| JANS3_EUR | 0.277 | −0.157 | 0.144 |
| JANS4_EUR | 0.0214 | −0.376 | 0.163 |
| JAPAN_EAS | 0.8794 | −0.016 | 0.104 |
| MIREC_AFR | 0.408 | 0.257 | 0.311 |
| MIREC_EUR | 0.7999 | −0.053 | 0.208 |
| MVPXQ_AFR | 0.5418 | −0.029 | 0.048 |
| MVPXQ_EAS | 0.8282 | −0.034 | 0.158 |
| MVPXQ_EUR | 0.0014 | −0.056 | 0.017 |
| MVPXQ_LAT | 0.226 | −0.058 | 0.048 |
| PGCBD_EUR | 0.1966 | 0.044 | 0.034 |
| PGCED_EUR | 0.5168 | −0.108 | 0.167 |
| PGCMD_EUR | 0.2842 | 0.049 | 0.046 |
| PGCPT_EUR | 0.2804 | −0.086 | 0.079 |
| PGCSZ_EUR | 0.3063 | 0.05 | 0.049 |
| PRFCT_EUR | 0.57 | −0.032 | 0.056 |
| PSYCR_EUR | 0.0621 | −0.204 | 0.109 |
| QIMRB_EUR | 0.0018 | −0.115 | 0.037 |
| SNUBH−ASA_EAS | 0.2376 | −0.276 | 0.234 |
| SNUBH−KCHIP_EAS | 0.4724 | −0.228 | 0.317 |
| STRR1_LAT | 0.9055 | 0.029 | 0.247 |
| UKBJC_EUR | 0.0678 | −0.049 | 0.027 |
| UTAMR_LAT | 0.1818 | −0.072 | 0.054 |
| YPENN_EUR | 0.4864 | −0.123 | 0.177 |
| **meta** | **1.37e−10** | **−0.053** | **0.008** |

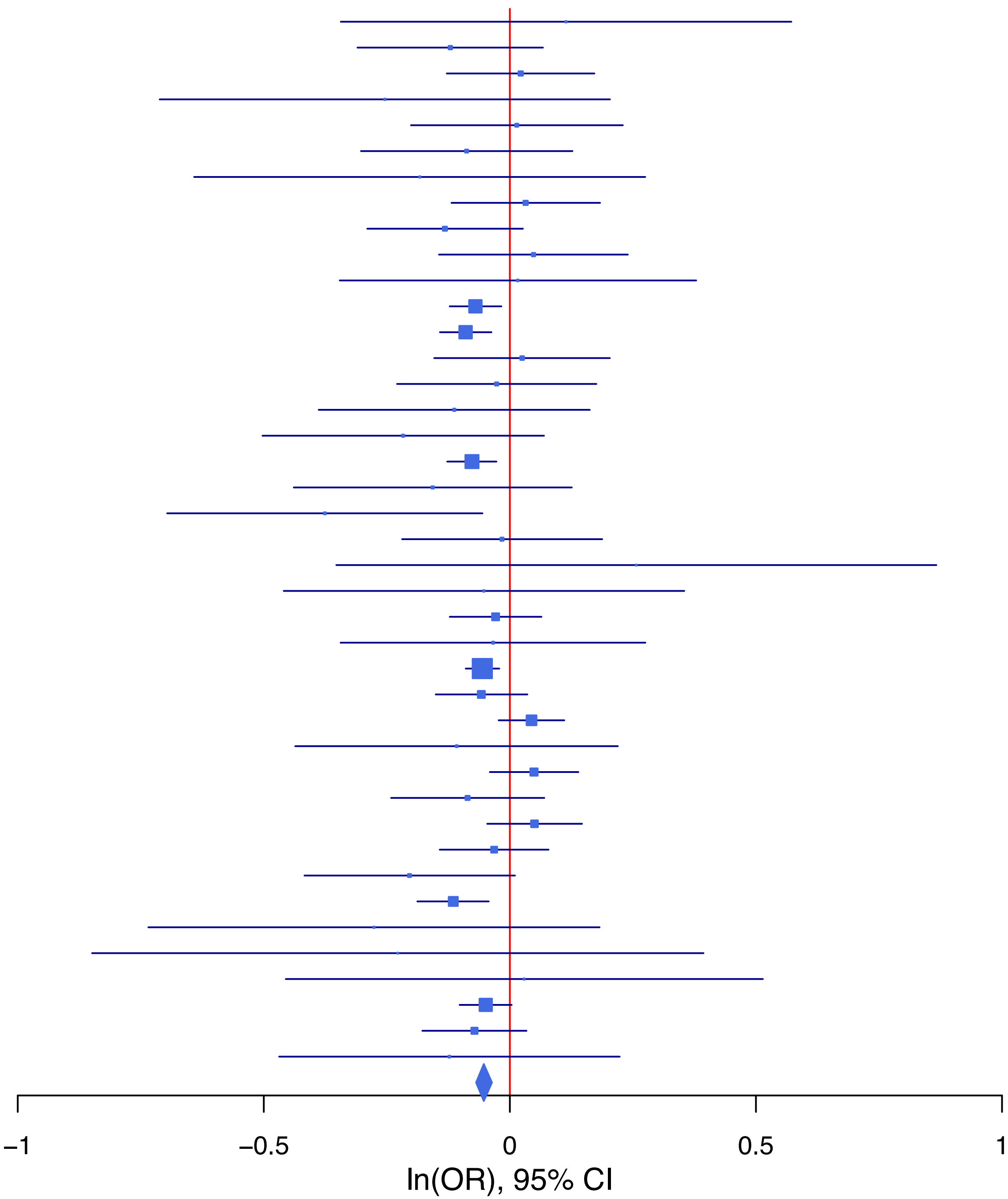

**rs62367520 C/A 5:45279858**

| Cohort | P | ln(OR) | SE |
|---|---|---|---|
| ALSPC_EUR | 0.1131 | −0.113 | 0.071 |
| BEPS7_EUR | 0.9055 | −0.027 | 0.226 |
| BHRCM_AFR | 0.3948 | 0.364 | 0.427 |
| BHRCM_LAT | 0.4087 | 0.223 | 0.27 |
| BOR17_EUR | 0.4756 | 0.078 | 0.11 |
| BOR2C_EUR | 0.2053 | −0.14 | 0.11 |
| BOR2E_EUR | 0.965 | 0.01 | 0.228 |
| CNVRG_EAS | 0.9831 | 0.002 | 0.072 |
| COGA1_AFR | 0.9697 | 0.008 | 0.21 |
| COGA1_EUR | 0.8662 | 0.013 | 0.075 |
| CUINT_EUR | 0.4997 | −0.063 | 0.094 |
| CVEDA_CSA | 0.7873 | −0.054 | 0.199 |
| ESTB2_EUR | 0.0027 | −0.094 | 0.031 |
| FINNG_EUR | 0.4943 | −0.022 | 0.032 |
| GEDIS_EUR | 0.8482 | 0.033 | 0.173 |
| GISS1_EUR | 0.4626 | −0.072 | 0.098 |
| GISS2_EUR | 0.6955 | 0.046 | 0.118 |
| GREAT_EAS | 0.5511 | −0.081 | 0.135 |
| GTPRJ_AFR | 0.5397 | −0.079 | 0.128 |
| IPSYC_EUR | 0.0788 | −0.044 | 0.025 |
| JANS3_EUR | 0.8867 | −0.019 | 0.132 |
| JANS4_EUR | 0.4113 | 0.121 | 0.148 |
| JAPAN_EAS | 0.1264 | −0.129 | 0.084 |
| MIREC_AFR | 0.1144 | −0.55 | 0.348 |
| MIREC_EUR | 0.8345 | 0.041 | 0.195 |
| MVPXQ_AFR | 0.4687 | −0.032 | 0.043 |
| MVPXQ_EAS | 0.8747 | 0.027 | 0.174 |
| MVPXQ_EUR | 0.002 | −0.052 | 0.017 |
| MVPXQ_LAT | 0.0028 | −0.122 | 0.041 |
| PGCBD_EUR | 0.4619 | −0.025 | 0.034 |
| PGCED_EUR | 0.3692 | 0.125 | 0.14 |
| PGCMD_EUR | 0.2234 | −0.056 | 0.046 |
| PGCPT_EUR | 0.6205 | −0.038 | 0.077 |
| PGCSZ_EUR | 0.9704 | 0.002 | 0.048 |
| PRFCT_EUR | 0.0543 | −0.114 | 0.06 |
| PSYCR_EUR | 0.8607 | 0.019 | 0.109 |
| QIMRB_EUR | 0.0188 | −0.081 | 0.035 |
| SNUBH−ASA_EAS | 0.1585 | 0.264 | 0.187 |
| SNUBH−KCHIP_EAS | 0.8831 | −0.035 | 0.239 |
| STRR1_LAT | 0.7765 | −0.061 | 0.213 |
| UKBJC_EUR | 6e−04 | −0.087 | 0.025 |
| UTAH2_EUR | 0.4114 | −0.02 | 0.025 |
| UTAMR_LAT | 0.414 | −0.033 | 0.04 |
| YPENN_EUR | 0.5228 | −0.104 | 0.163 |
| **meta** | **6.53e−11** | **−0.05** | **0.008** |

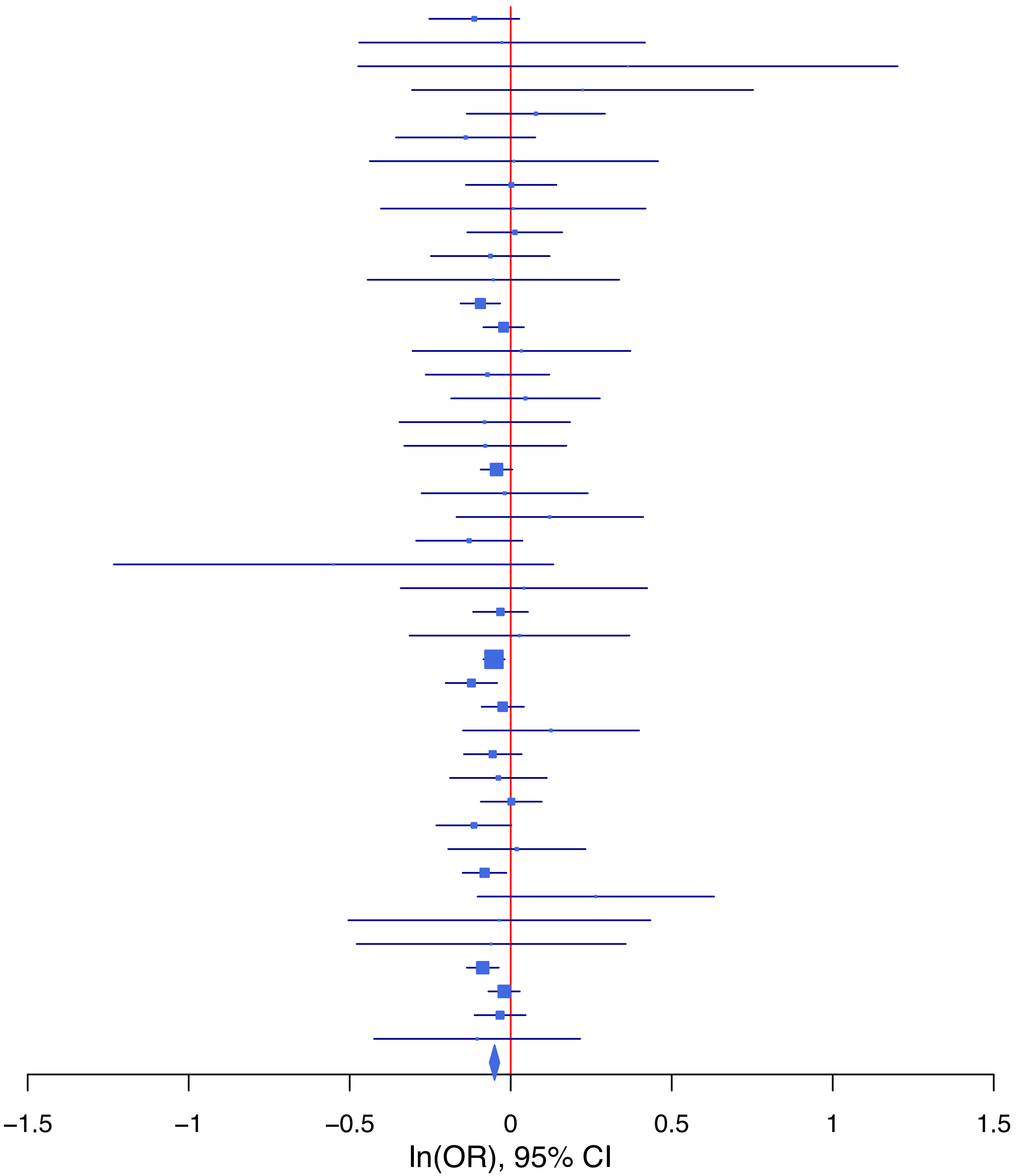

**rs62404522 C/T 6:19307114**

| Cohort | P | ln(OR) | SE |
|---|---|---|---|
| ADHEA_AFR | 0.1805 | −0.289 | 0.216 |
| ADHEA_EUR | 0.1413 | −0.154 | 0.105 |
| ALSPC_EUR | 0.2246 | 0.093 | 0.077 |
| BEPS7_EUR | 0.7783 | 0.07 | 0.25 |
| BHRCM_AFR | 0.9811 | 0.008 | 0.327 |
| BHRCM_EUR | 0.5224 | −0.166 | 0.26 |
| BHRCM_LAT | 0.9721 | −0.01 | 0.296 |
| BOR17_EUR | 0.9851 | −0.002 | 0.126 |
| BOR2C_EUR | 0.067 | 0.233 | 0.127 |
| BOR2E_EUR | 0.3309 | −0.281 | 0.29 |
| CNVRG_EAS | 0.7083 | −0.025 | 0.066 |
| COGA1_AFR | 0.0497 | 0.329 | 0.168 |
| COGA1_EUR | 0.1278 | 0.133 | 0.087 |
| CUINT_EUR | 0.9098 | −0.013 | 0.111 |
| CVEDA_CSA | 0.5702 | 0.101 | 0.177 |
| ESTB2_EUR | 0.3339 | 0.032 | 0.033 |
| FINNG_EUR | 0.0012 | 0.11 | 0.034 |
| GEDIS_EUR | 0.1439 | 0.257 | 0.176 |
| GISS1_EUR | 0.728 | 0.04 | 0.116 |
| GISS2_EUR | 0.0643 | 0.272 | 0.147 |
| GREAT_EAS | 0.6291 | −0.054 | 0.111 |
| GTPRJ_AFR | 0.0903 | 0.161 | 0.095 |
| IPSYC_EUR | 0.1929 | 0.034 | 0.026 |
| JANS3_EUR | 0.2348 | 0.171 | 0.144 |
| JANS4_EUR | 0.5364 | −0.116 | 0.187 |
| JAPAN_EAS | 0.1718 | 0.121 | 0.088 |
| MVPXQ_AFR | 0.0256 | 0.082 | 0.037 |
| MVPXQ_EAS | 0.5683 | −0.084 | 0.148 |
| MVPXQ_EUR | 3.92e−05 | 0.076 | 0.019 |
| MVPXQ_LAT | 0.0438 | 0.082 | 0.041 |
| PGCBD_EUR | 1e−04 | 0.146 | 0.037 |
| PGCPT_AFR | 0.3129 | −0.193 | 0.191 |
| PGCPT_EUR | 0.4313 | 0.063 | 0.08 |
| PGCSZ_EUR | 0.7464 | 0.018 | 0.054 |
| PRFCT_EUR | 0.0078 | 0.161 | 0.06 |
| PSYCR_EUR | 0.081 | −0.227 | 0.13 |
| QIMRB_EUR | 0.0417 | 0.081 | 0.04 |
| SNUBH−ASA_EAS | 0.9686 | −0.008 | 0.204 |
| SNUBH−KCHIP_EAS | 0.707 | 0.101 | 0.269 |
| STRR1_LAT | 0.9538 | −0.012 | 0.212 |
| UKBJC_EUR | 0.7566 | 0.009 | 0.029 |
| UTAH2_EUR | 0.2209 | 0.033 | 0.027 |
| UTAMR_LAT | 0.3477 | 0.037 | 0.04 |
| YPENN_EUR | 0.8099 | 0.045 | 0.188 |
| **meta** | **4.85e−13** | **0.06** | **0.008** |

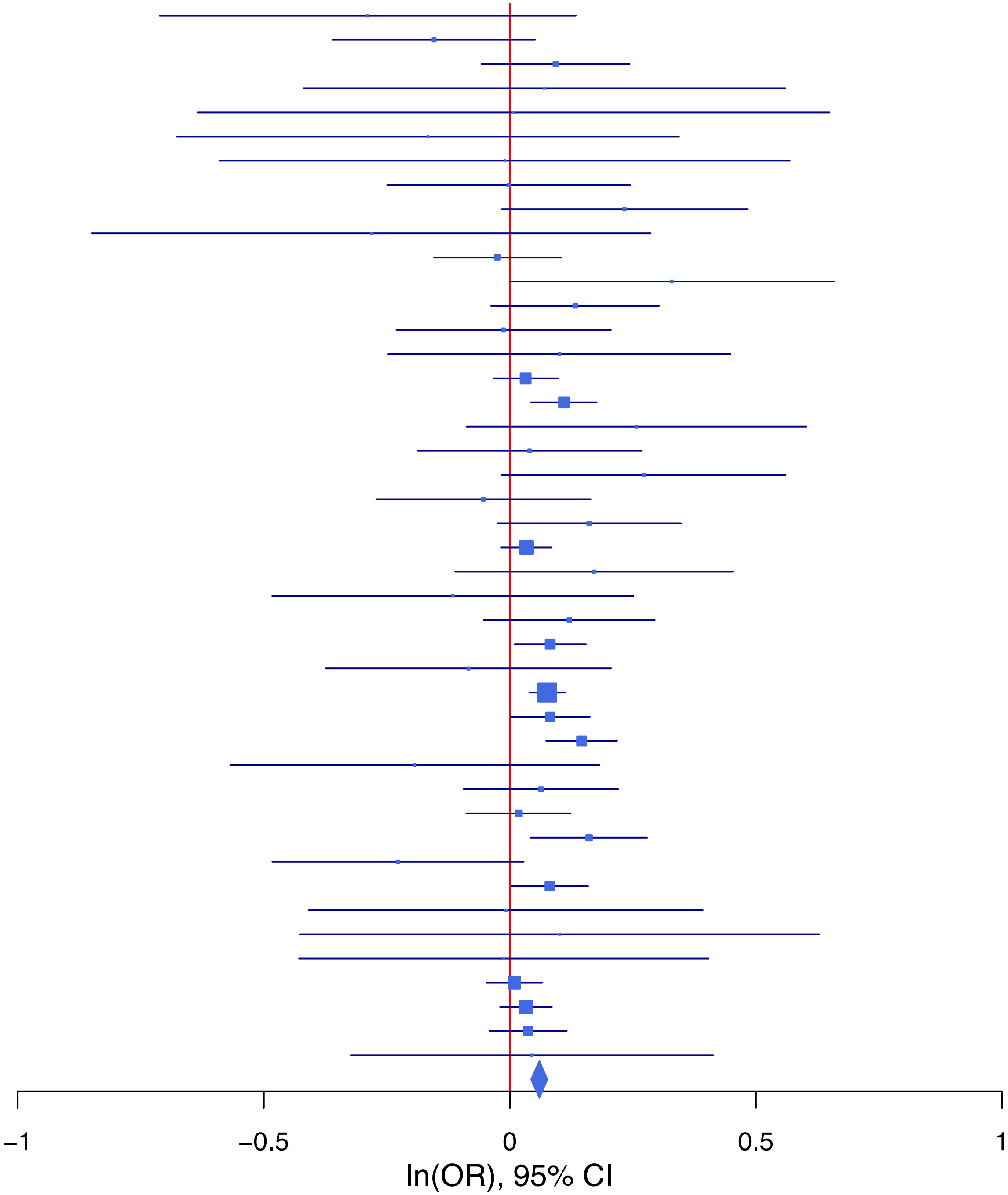

**rs62477308 T/C 7:114944408**

| Cohort | P | ln(OR) | SE |
|---|---|---|---|
| ADHEA_AFR | 0.4371 | −0.099 | 0.128 |
| ADHEA_EUR | 0.661 | −0.03 | 0.068 |
| ALSPC_EUR | 0.1506 | −0.078 | 0.054 |
| BEPS7_EUR | 0.3707 | −0.161 | 0.18 |
| BHRCM_AFR | 0.984 | 0.005 | 0.224 |
| BHRCM_EUR | 0.5132 | −0.105 | 0.16 |
| BHRCM_LAT | 0.7643 | −0.079 | 0.262 |
| BOR17_EUR | 0.5398 | 0.054 | 0.088 |
| BOR2C_EUR | 0.9154 | −0.009 | 0.087 |
| BOR2E_EUR | 0.545 | −0.115 | 0.19 |
| CNVRG_EAS | 0.2331 | 0.055 | 0.046 |
| COGA1_AFR | 0.5472 | 0.066 | 0.11 |
| COGA1_EUR | 0.5054 | −0.039 | 0.059 |
| CVEDA_CSA | 0.1905 | −0.211 | 0.161 |
| ESTB2_EUR | 2.01e−05 | −0.096 | 0.023 |
| FINNG_EUR | 0.1062 | −0.037 | 0.023 |
| GEDIS_EUR | 0.1601 | −0.183 | 0.13 |
| GISS1_EUR | 0.1989 | −0.1 | 0.078 |
| GISS2_EUR | 0.2092 | −0.115 | 0.092 |
| GREAT_EAS | 0.9595 | −0.004 | 0.086 |
| GTPRJ_AFR | 0.7026 | −0.026 | 0.069 |
| IPSYC_EUR | 1e−04 | −0.072 | 0.019 |
| JANS3_EUR | 0.4493 | −0.081 | 0.106 |
| JANS4_EUR | 0.1971 | −0.161 | 0.125 |
| JAPAN_EAS | 0.4649 | −0.041 | 0.056 |
| PGCBD_EUR | 0.0746 | −0.047 | 0.026 |
| PGCED_EUR | 0.3269 | 0.116 | 0.118 |
| PGCMD_EUR | 0.1787 | −0.047 | 0.035 |
| PGCPT_AFR | 0.8767 | −0.018 | 0.117 |
| PGCPT_EUR | 0.3554 | −0.054 | 0.059 |
| PGCSZ_EUR | 0.6802 | −0.015 | 0.038 |
| PRFCT_EUR | 0.44 | −0.034 | 0.044 |
| PSYCR_EUR | 0.9645 | −0.004 | 0.087 |
| QIMRB_EUR | 0.0144 | −0.068 | 0.028 |
| SNUBH−ASA_EAS | 0.2688 | −0.135 | 0.122 |
| STRR1_LAT | 0.0551 | 0.349 | 0.182 |
| UKBJC_EUR | 0.5582 | −0.012 | 0.02 |
| UTAH2_EUR | 0.0224 | −0.043 | 0.019 |
| UTAMR_LAT | 0.4062 | −0.03 | 0.036 |
| YPENN_EUR | 0.891 | 0.018 | 0.129 |
| **meta** | **2.09e−11** | **−0.046** | **0.007** |

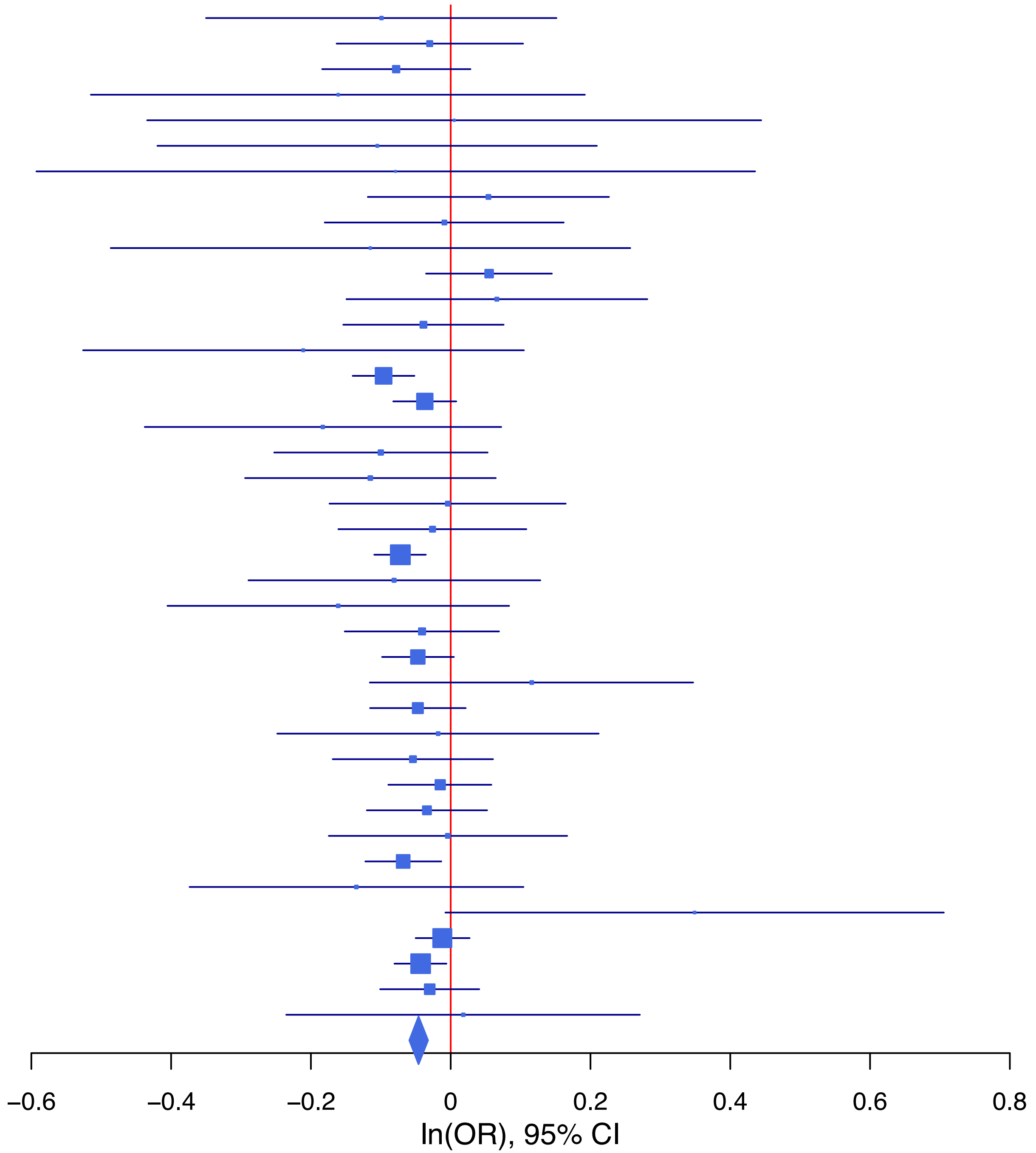

**rs631248 G/A 1:44071221**

| Cohort | P | ln(OR) | SE |
|---|---|---|---|
| ALSPC_EUR | 0.6215 | −0.033 | 0.067 |
| BEPS7_EUR | 0.263 | 0.244 | 0.218 |
| BHRCM_AFR | 0.0947 | 0.348 | 0.208 |
| BHRCM_EUR | 0.3554 | −0.186 | 0.201 |
| BHRCM_LAT | 0.0656 | 0.6 | 0.326 |
| BOR17_EUR | 0.531 | 0.065 | 0.103 |
| BOR2C_EUR | 0.9326 | −0.008 | 0.097 |
| BOR2E_EUR | 0.6073 | 0.115 | 0.225 |
| CNVRG_EAS | 0.1736 | 0.074 | 0.054 |
| COGA1_AFR | 0.5215 | 0.064 | 0.1 |
| COGA1_EUR | 0.3553 | −0.065 | 0.07 |
| CUINT_EUR | 0.3817 | −0.076 | 0.087 |
| CVEDA_CSA | 0.1391 | 0.24 | 0.162 |
| ESTB2_EUR | 0.5342 | 0.017 | 0.028 |
| FINNG_EUR | 0.3024 | 0.029 | 0.028 |
| GEDIS_EUR | 0.3597 | 0.137 | 0.15 |
| GISS1_EUR | 0.5194 | −0.059 | 0.092 |
| GISS2_EUR | 0.2986 | 0.114 | 0.11 |
| GREAT_EAS | 0.0348 | 0.211 | 0.1 |
| GTPRJ_AFR | 0.7552 | −0.018 | 0.059 |
| IPSYC_EUR | 0.0045 | 0.062 | 0.022 |
| JANS3_EUR | 0.3546 | 0.108 | 0.116 |
| JANS4_EUR | 0.6743 | −0.059 | 0.139 |
| JAPAN_EAS | 0.3973 | 0.05 | 0.059 |
| MVPXQ_EAS | 0.9062 | −0.013 | 0.112 |
| MVPXQ_EUR | 0.0172 | 0.035 | 0.015 |
| MVPXQ_LAT | 0.301 | 0.034 | 0.033 |
| PGCBD_EUR | 0.0571 | 0.058 | 0.03 |
| PGCED_EUR | 0.4375 | −0.106 | 0.136 |
| PGCMD_EUR | 0.051 | 0.079 | 0.04 |
| PGCPT_AFR | 0.4462 | 0.078 | 0.102 |
| PGCPT_EUR | 0.6231 | 0.033 | 0.068 |
| PGCSZ_EUR | 0.7772 | 0.012 | 0.044 |
| PRFCT_EUR | 0.0569 | 0.098 | 0.052 |
| PSYCR_EUR | 0.7628 | 0.031 | 0.104 |
| QIMRB_EUR | 0.0138 | 0.081 | 0.033 |
| SNUBH-ASA_EAS | 0.042 | −0.288 | 0.141 |
| SNUBH-KCHIP_EAS | 0.7981 | 0.05 | 0.196 |
| STRR1_LAT | 0.7137 | 0.065 | 0.177 |
| UKBJC_EUR | 0.5409 | 0.015 | 0.024 |
| UTAH2_EUR | 0.1291 | 0.034 | 0.022 |
| UTAMR_LAT | 0.3975 | 0.03 | 0.036 |
| YPENN_EUR | 0.5607 | 0.085 | 0.146 |
| **meta** | **3.20e−08** | **0.038** | **0.007** |

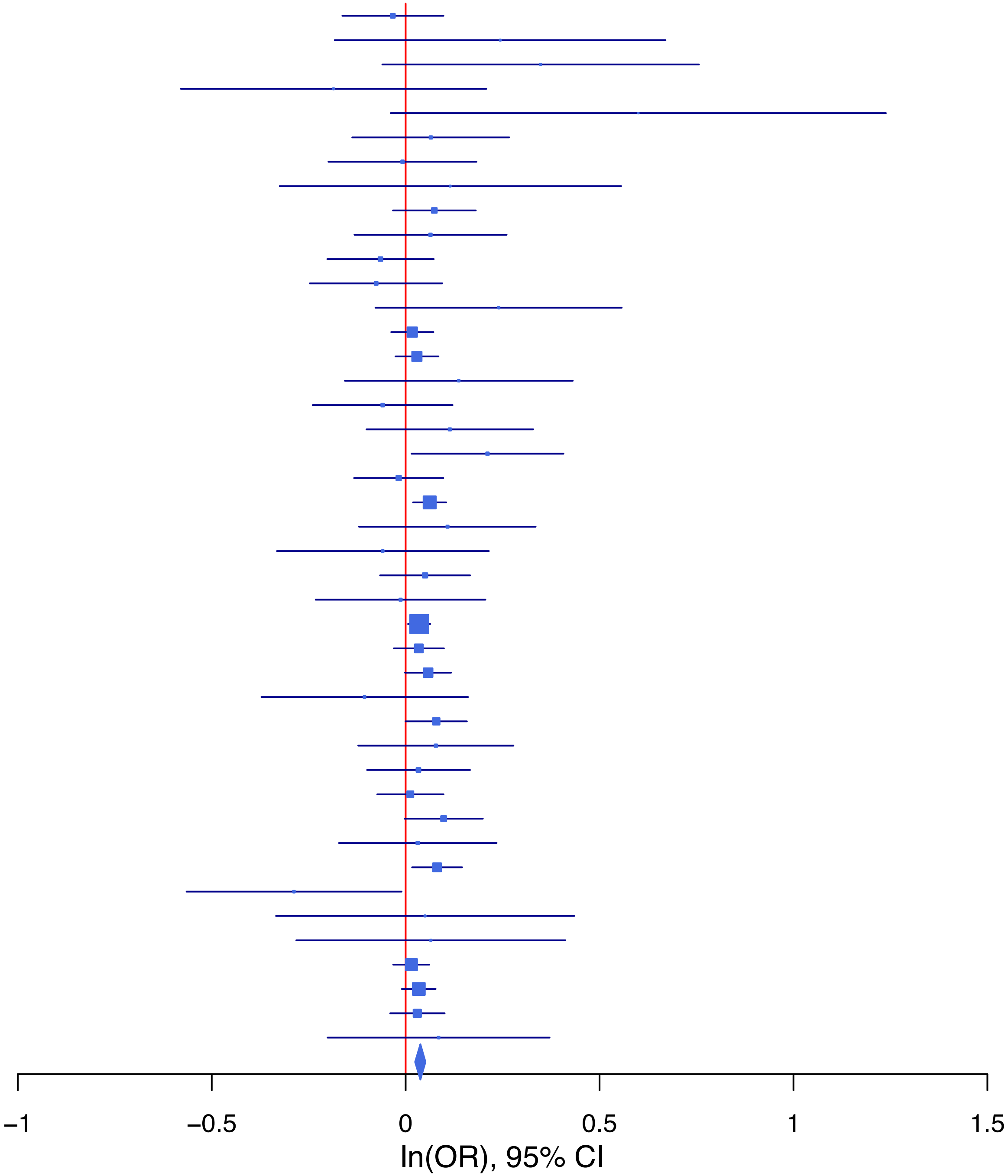

**rs653765 C/T 15:59042012**

| Cohort | P | ln(OR) | SE |
|---|---|---|---|
| ALSPC_EUR | 0.1641 | −0.084 | 0.061 |
| BEPS7_EUR | 0.8929 | 0.026 | 0.195 |
| BHRCM_AFR | 0.2395 | −0.254 | 0.216 |
| BHRCM_EUR | 0.9834 | −0.004 | 0.185 |
| BOR17_EUR | 0.5736 | −0.054 | 0.095 |
| BOR2C_EUR | 0.5038 | 0.064 | 0.096 |
| BOR2E_EUR | 0.8921 | −0.029 | 0.214 |
| CNVRG_EAS | 0.0896 | −0.101 | 0.06 |
| COGA1_AFR | 0.9355 | 0.009 | 0.111 |
| COGA1_EUR | 0.4536 | −0.049 | 0.065 |
| CUINT_EUR | 0.4552 | −0.063 | 0.085 |
| CVEDA_CSA | 0.6392 | −0.078 | 0.166 |
| ESTB2_EUR | 0.0065 | −0.067 | 0.025 |
| FINNG_EUR | 0.8225 | −0.005 | 0.023 |
| GEDIS_EUR | 0.6844 | 0.057 | 0.141 |
| GISS1_EUR | 0.8972 | 0.011 | 0.086 |
| GISS2_EUR | 0.2359 | −0.122 | 0.103 |
| IPSYC_EUR | 0.0028 | −0.06 | 0.02 |
| JANS3_EUR | 0.8897 | 0.016 | 0.117 |
| JANS4_EUR | 0.4711 | 0.1 | 0.139 |
| MVPXQ_EAS | 0.7452 | 0.04 | 0.125 |
| MVPXQ_EUR | 0.0433 | −0.028 | 0.014 |
| MVPXQ_LAT | 0.0452 | −0.071 | 0.036 |
| PGCBD_EUR | 0.0918 | −0.048 | 0.028 |
| PGCED_EUR | 0.439 | 0.097 | 0.126 |
| PGCMD_EUR | 0.9985 | 0 | 0.037 |
| PGCPT_AFR | 0.4766 | −0.082 | 0.116 |
| PGCPT_EUR | 0.6983 | 0.024 | 0.063 |
| PGCSZ_EUR | 0.0033 | −0.122 | 0.042 |
| PRFCT_EUR | 0.5067 | 0.031 | 0.047 |
| PSYCR_EUR | 0.8715 | 0.016 | 0.097 |
| QIMRB_EUR | 0.1317 | −0.045 | 0.03 |
| SNUBH−ASA_EAS | 0.5562 | −0.099 | 0.168 |
| SNUBH−KCHIP_EAS | 0.6106 | 0.13 | 0.256 |
| STRR1_LAT | 0.2728 | −0.201 | 0.183 |
| UKBJC_EUR | 0.038 | −0.045 | 0.022 |
| UTAMR_LAT | 0.0294 | −0.082 | 0.038 |
| YPENN_EUR | 0.8543 | 0.026 | 0.142 |
| **meta** | **3.21e−09** | **−0.04** | **0.007** |

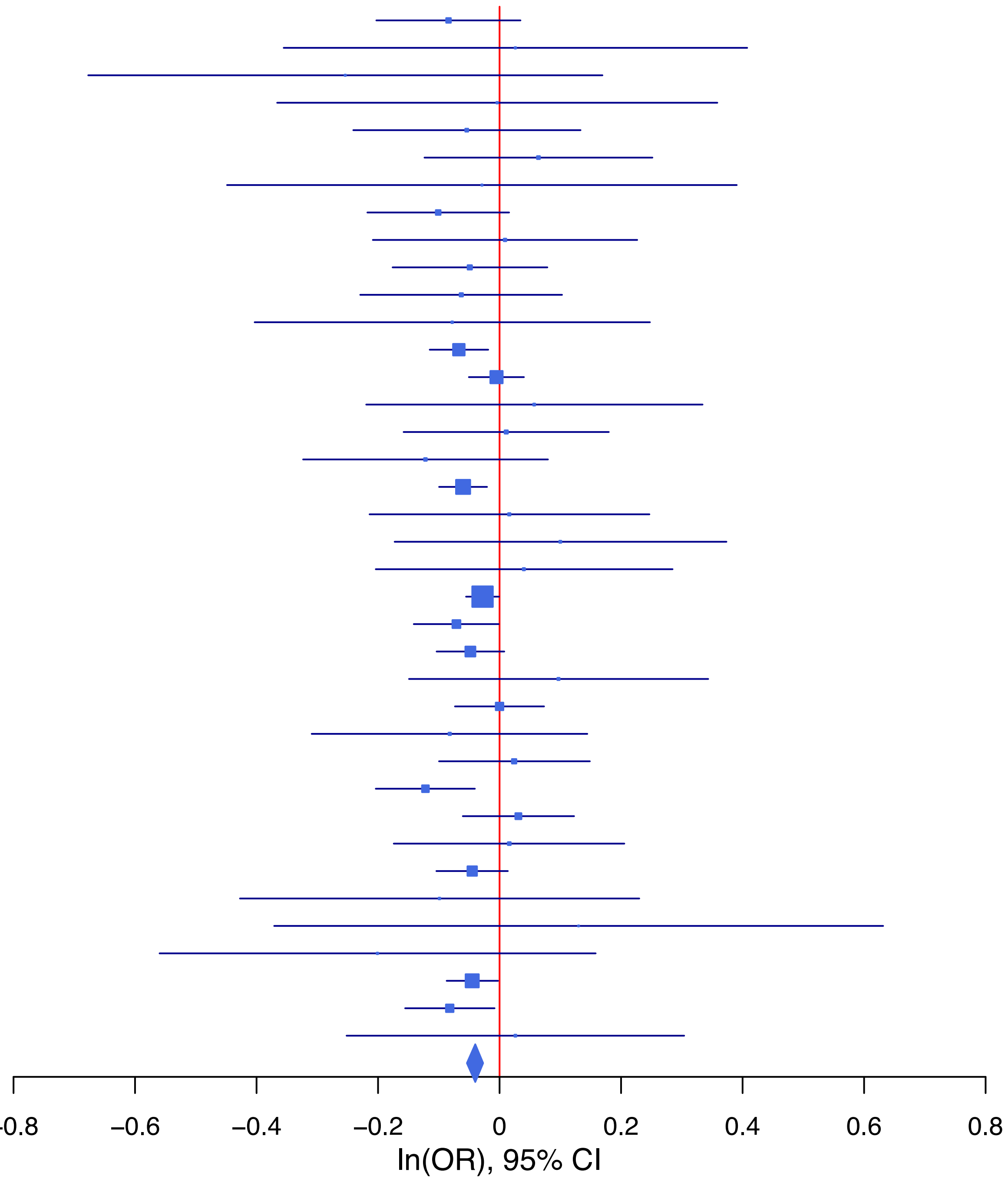

**rs66824958 T/C 4:166061559**

| Cohort | P | ln(OR) | SE |
|---|---|---|---|
| ADHEA_EUR | 0.6198 | −0.051 | 0.103 |
| ALSPC_EUR | 0.5444 | 0.047 | 0.078 |
| BEPS7_EUR | 0.7726 | 0.071 | 0.247 |
| BHRCM_EUR | 0.8864 | −0.041 | 0.284 |
| BHRCM_LAT | 0.4666 | 0.214 | 0.294 |
| BOR17_EUR | 0.6452 | −0.061 | 0.133 |
| BOR2C_EUR | 0.0168 | −0.308 | 0.129 |
| BOR2E_EUR | 0.6522 | −0.141 | 0.312 |
| CNVRG_EAS | 0.1551 | 0.173 | 0.122 |
| COGA1_AFR | 0.9161 | 0.032 | 0.304 |
| COGA1_EUR | 0.0248 | −0.203 | 0.091 |
| CUINT_EUR | 0.4481 | 0.087 | 0.114 |
| CVEDA_CSA | 0.8066 | 0.041 | 0.168 |
| ESTB2_EUR | 0.1504 | −0.047 | 0.032 |
| FINNG_EUR | 0.0212 | −0.068 | 0.03 |
| GEDIS_EUR | 0.8017 | −0.049 | 0.196 |
| GISS1_EUR | 0.128 | −0.179 | 0.117 |
| GISS2_EUR | 0.958 | 0.007 | 0.133 |
| GREAT_EAS | 0.5317 | −0.149 | 0.238 |
| GTPRJ_AFR | 0.661 | −0.092 | 0.21 |
| IPSYC_EUR | 0.0626 | −0.05 | 0.027 |
| JANS3_EUR | 0.1136 | −0.265 | 0.168 |
| JANS4_EUR | 0.6917 | 0.076 | 0.19 |
| JAPAN_EAS | 0.6335 | −0.065 | 0.136 |
| MVPXQ_AFR | 0.32 | −0.068 | 0.068 |
| MVPXQ_EAS | 0.4705 | −0.152 | 0.211 |
| MVPXQ_EUR | 0.0214 | −0.043 | 0.019 |
| MVPXQ_LAT | 0.8045 | −0.012 | 0.046 |
| PGCBD_EUR | 0.1373 | −0.059 | 0.04 |
| PGCED_EUR | 0.7918 | 0.05 | 0.189 |
| PGCMD_EUR | 0.761 | −0.016 | 0.053 |
| PGCPT_EUR | 0.6192 | −0.044 | 0.088 |
| PGCSZ_EUR | 0.5867 | −0.03 | 0.056 |
| PRFCT_EUR | 0.0145 | −0.146 | 0.06 |
| PSYCR_EUR | 0.0107 | −0.369 | 0.145 |
| QIMRB_EUR | 0.0907 | −0.068 | 0.04 |
| SNUBH−ASA_EAS | 0.0315 | 0.728 | 0.338 |
| STRR1_LAT | 0.2116 | −0.319 | 0.256 |
| UKBJC_EUR | 0.1811 | −0.038 | 0.028 |
| UTAMR_LAT | 0.649 | −0.021 | 0.046 |
| YPENN_EUR | 0.6152 | −0.095 | 0.189 |
| **meta** | **4.69e−08** | **−0.049** | **0.009** |

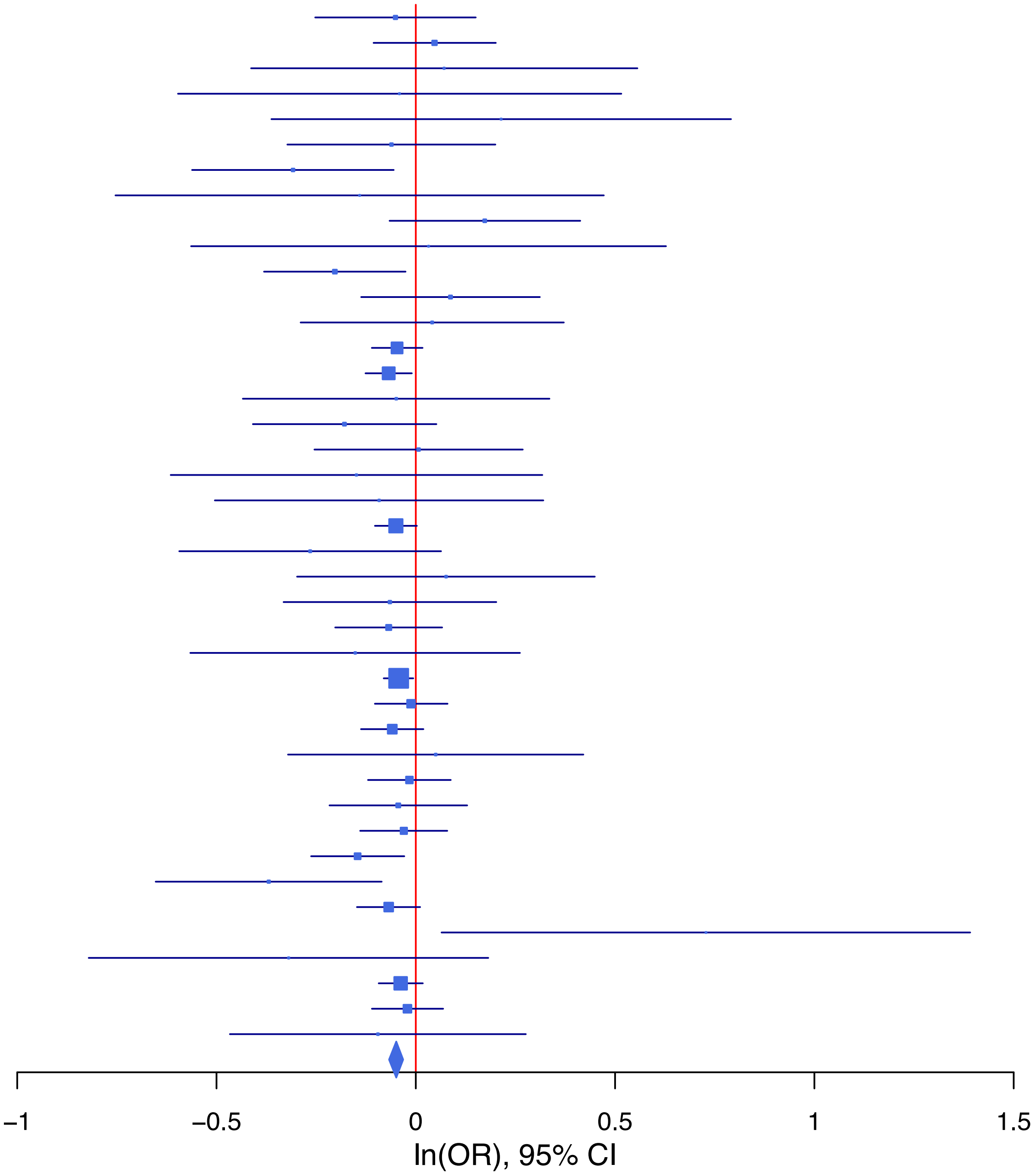

**rs7169340 C/T 15:47614645**

| Cohort | P | ln(OR) | SE |
|---|---|---|---|
| ADHEA_AFR | 0.3661 | −0.114 | 0.126 |
| ADHEA_EUR | 0.5505 | −0.052 | 0.087 |
| BEPS7_EUR | 0.6502 | 0.11 | 0.243 |
| BHRCM_AFR | 0.3234 | −0.206 | 0.208 |
| BHRCM_EUR | 0.9144 | 0.02 | 0.185 |
| BOR17_EUR | 0.3327 | 0.098 | 0.101 |
| BOR2C_EUR | 0.7579 | −0.034 | 0.11 |
| BOR2E_EUR | 0.4261 | 0.186 | 0.234 |
| COGA1_AFR | 0.2581 | 0.121 | 0.107 |
| COGA1_EUR | 0.803 | −0.018 | 0.072 |
| CUINT_EUR | 0.6831 | −0.039 | 0.096 |
| CVEDA_CSA | 0.7416 | −0.112 | 0.339 |
| ESTB2_EUR | 0.1632 | 0.043 | 0.031 |
| FINNG_EUR | 0.3617 | 0.028 | 0.031 |
| GEDIS_EUR | 0.1262 | −0.26 | 0.17 |
| GISS1_EUR | 0.4261 | 0.084 | 0.106 |
| GISS2_EUR | 0.0506 | 0.26 | 0.133 |
| GTPRJ_AFR | 0.1512 | 0.089 | 0.062 |
| IPSYC_EUR | 0.0022 | 0.068 | 0.022 |
| JANS3_EUR | 0.966 | 0.006 | 0.132 |
| JANS4_EUR | 0.9341 | −0.013 | 0.158 |
| MIREC_AFR | 0.571 | −0.083 | 0.147 |
| MIREC_EUR | 0.0703 | 0.324 | 0.179 |
| PGCBD_EUR | 0.0073 | 0.087 | 0.032 |
| PGCED_EUR | 0.3284 | 0.135 | 0.138 |
| PGCMD_EUR | 0.0078 | 0.112 | 0.042 |
| PGCPT_AFR | 0.675 | 0.045 | 0.107 |
| PGCPT_EUR | 0.1318 | −0.11 | 0.073 |
| PGCSZ_EUR | 9e−04 | 0.152 | 0.046 |
| PRFCT_EUR | 0.9847 | −0.001 | 0.055 |
| PSYCR_EUR | 0.418 | −0.094 | 0.116 |
| QIMRB_EUR | 7e−04 | 0.122 | 0.036 |
| STRR1_LAT | 0.6592 | 0.111 | 0.252 |
| UKBJC_EUR | 0.0116 | 0.062 | 0.024 |
| UTAH2_EUR | 0.0018 | 0.072 | 0.023 |
| UTAMR_LAT | 0.3449 | 0.055 | 0.058 |
| YPENN_EUR | 0.407 | −0.139 | 0.168 |
| **meta** | **2.70e−12** | **0.061** | **0.009** |

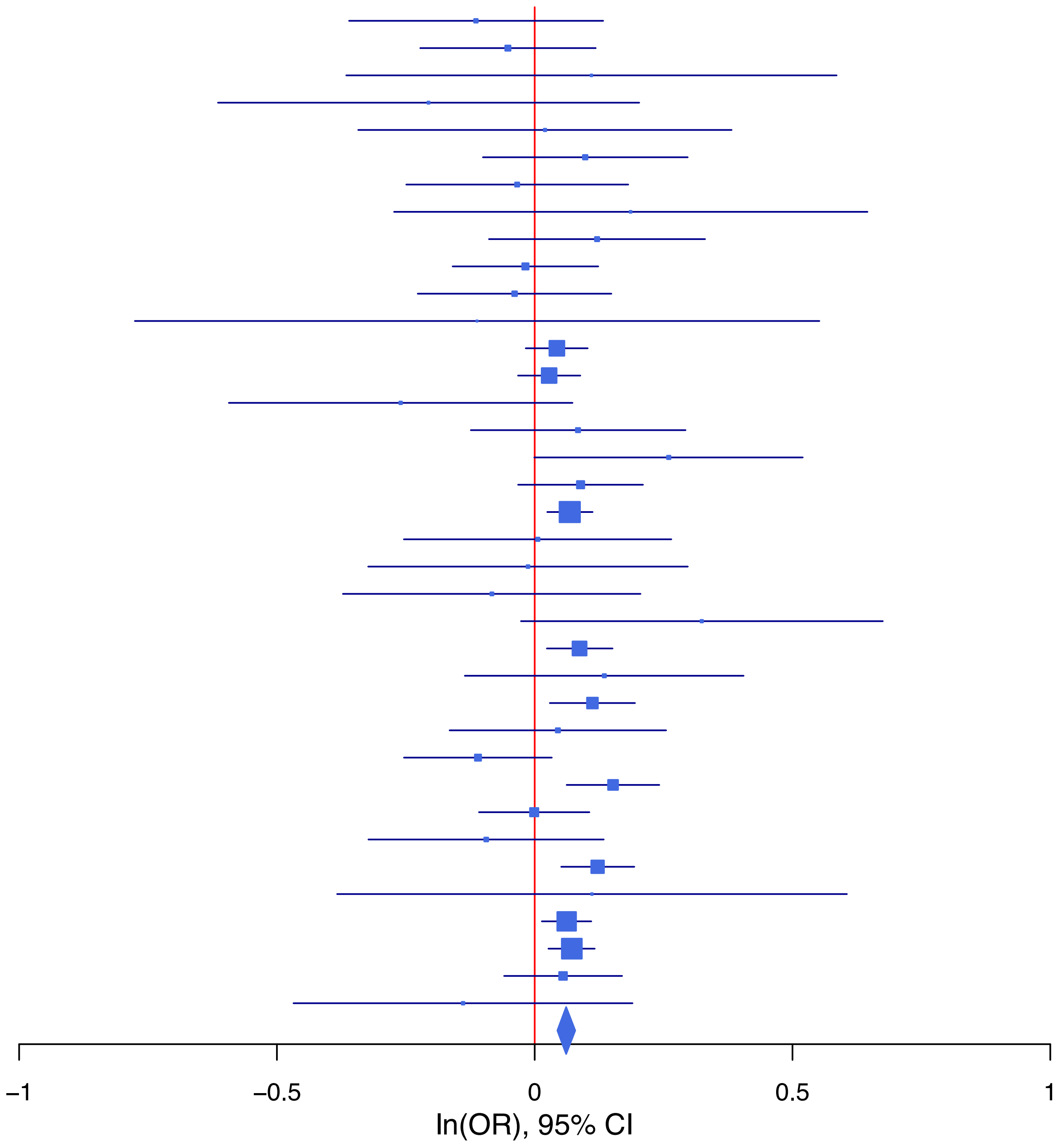

**rs72685056 A/G 1:66397344**

| Cohort | P | ln(OR) | SE |
|---|---|---|---|
| ALSPC_EUR | 0.5256 | −0.063 | 0.099 |
| BEPS7_EUR | 0.627 | 0.126 | 0.26 |
| BHRCM_EUR | 0.5638 | 0.148 | 0.256 |
| BOR17_EUR | 0.6235 | −0.059 | 0.121 |
| BOR2C_EUR | 0.2418 | 0.179 | 0.153 |
| BOR2E_EUR | 0.3041 | 0.318 | 0.309 |
| ESTB2_EUR | 0.0293 | 0.061 | 0.028 |
| FINNG_EUR | 0.0895 | 0.047 | 0.028 |
| GEDIS_EUR | 0.1911 | 0.236 | 0.18 |
| GISS2_EUR | 0.1644 | −0.188 | 0.135 |
| GTPRJ_AFR | 0.7527 | −0.081 | 0.257 |
| IPSYC_EUR | 8.25e−06 | 0.111 | 0.025 |
| JANS3_EUR | 0.9971 | −0.001 | 0.185 |
| JANS4_EUR | 0.9291 | −0.016 | 0.175 |
| MVPXQ_AFR | 0.546 | −0.041 | 0.068 |
| MVPXQ_EUR | 0.2447 | 0.021 | 0.018 |
| MVPXQ_LAT | 0.1569 | −0.101 | 0.071 |
| PGCBD_EUR | 0.1773 | 0.058 | 0.043 |
| PGCED_EUR | 0.249 | −0.309 | 0.268 |
| PGCMD_EUR | 0.0054 | 0.154 | 0.055 |
| PGCPT_EUR | 0.4778 | 0.064 | 0.091 |
| PGCSZ_EUR | 0.3355 | 0.058 | 0.06 |
| PRFCT_EUR | 0.1483 | 0.094 | 0.065 |
| PSYCR_EUR | 0.7831 | 0.039 | 0.14 |
| QIMRB_EUR | 0.1167 | 0.07 | 0.045 |
| STRR1_LAT | 0.1931 | −0.638 | 0.49 |
| UKBJC_EUR | 0.0314 | 0.063 | 0.029 |
| UTAH2_EUR | 0.0177 | 0.064 | 0.027 |
| UTAMR_LAT | 0.0474 | 0.181 | 0.091 |
| YPENN_EUR | 0.8174 | −0.044 | 0.193 |
| **meta** | **9.46e−10** | **0.054** | **0.009** |

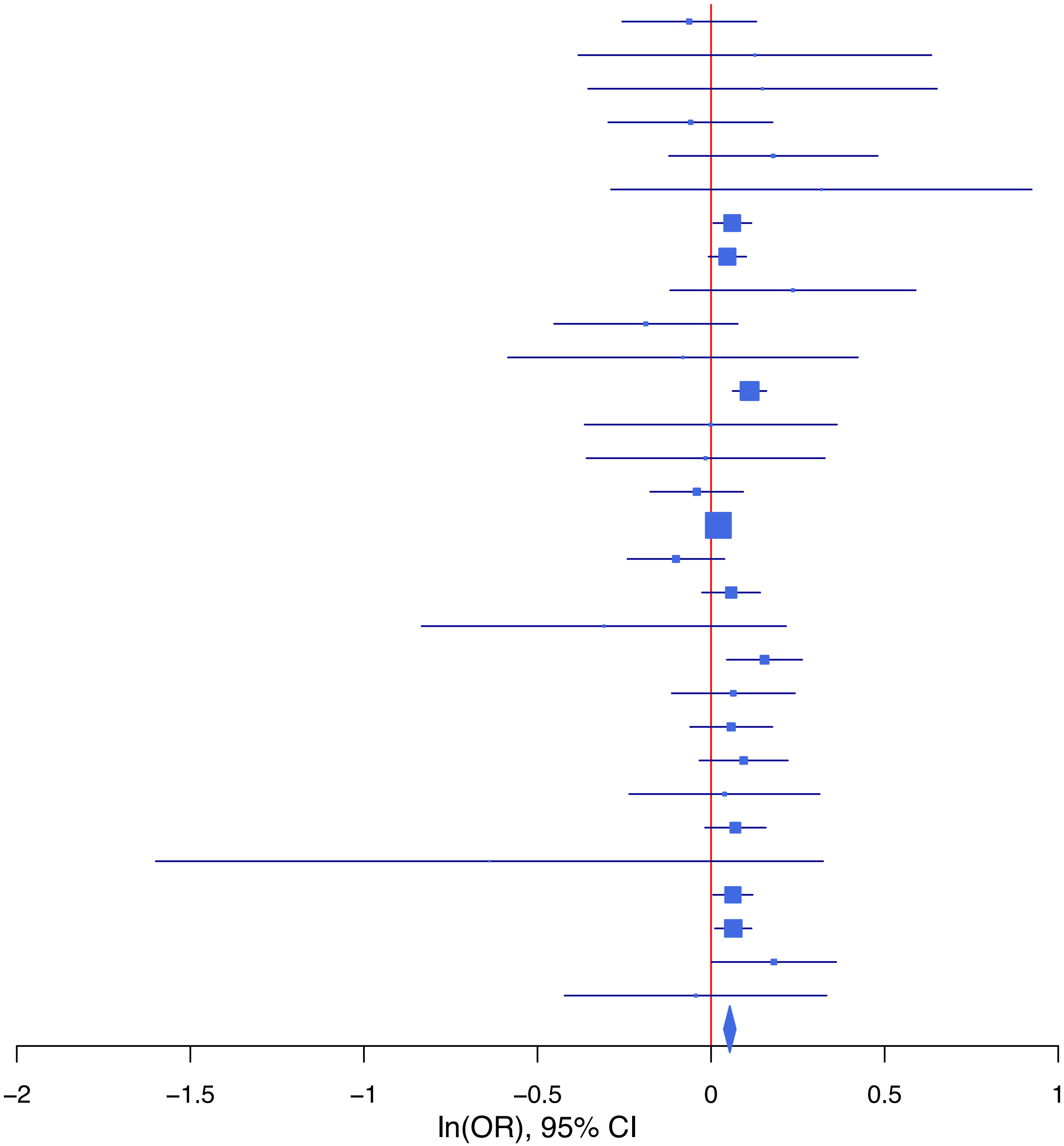

**rs72925321   A/G   18:50807202**

| Cohort | P | ln(OR) | SE |
|---|---|---|---|
| ADHEA_EUR | 0.9271 | −0.014 | 0.157 |
| ALSPC_EUR | 0.8505 | −0.021 | 0.114 |
| BHRCM_EUR | 0.0587 | 0.671 | 0.355 |
| BOR17_EUR | 0.281 | −0.197 | 0.183 |
| BOR2C_EUR | 0.9487 | 0.012 | 0.185 |
| BOR2E_EUR | 0.2693 | −0.45 | 0.407 |
| COGA1_EUR | 0.6371 | 0.06 | 0.128 |
| CUINT_EUR | 0.9387 | 0.014 | 0.184 |
| CVEDA_CSA | 0.8358 | −0.064 | 0.309 |
| ESTB2_EUR | 0.0526 | −0.084 | 0.043 |
| FINNG_EUR | 0.1095 | −0.066 | 0.041 |
| GEDIS_EUR | 0.8241 | −0.063 | 0.282 |
| GISS1_EUR | 0.4395 | −0.109 | 0.141 |
| GISS2_EUR | 0.4162 | 0.143 | 0.176 |
| IPSYC_EUR | 0.031 | −0.09 | 0.042 |
| JANS3_EUR | 0.8643 | −0.041 | 0.241 |
| JANS4_EUR | 0.5307 | −0.171 | 0.273 |
| MVPXQ_EUR | 9e−04 | −0.108 | 0.032 |
| PGCBD_EUR | 0.0437 | −0.134 | 0.067 |
| PGCMD_EUR | 0.073 | −0.164 | 0.091 |
| PGCPT_EUR | 0.6592 | −0.056 | 0.127 |
| PGCSZ_EUR | 0.0518 | −0.182 | 0.094 |
| PRFCT_EUR | 0.7534 | −0.031 | 0.098 |
| PSYCR_EUR | 0.4729 | −0.138 | 0.192 |
| QIMRB_EUR | 0.0468 | −0.116 | 0.058 |
| UKBJC_EUR | 0.243 | −0.049 | 0.042 |
| UTAH2_EUR | 0.2674 | −0.043 | 0.039 |
| UTAMR_LAT | 0.383 | 0.09 | 0.103 |
| YPENN_EUR | 0.7813 | −0.081 | 0.292 |
| **meta** | **2.43e−08** | **−0.076** | **0.014** |

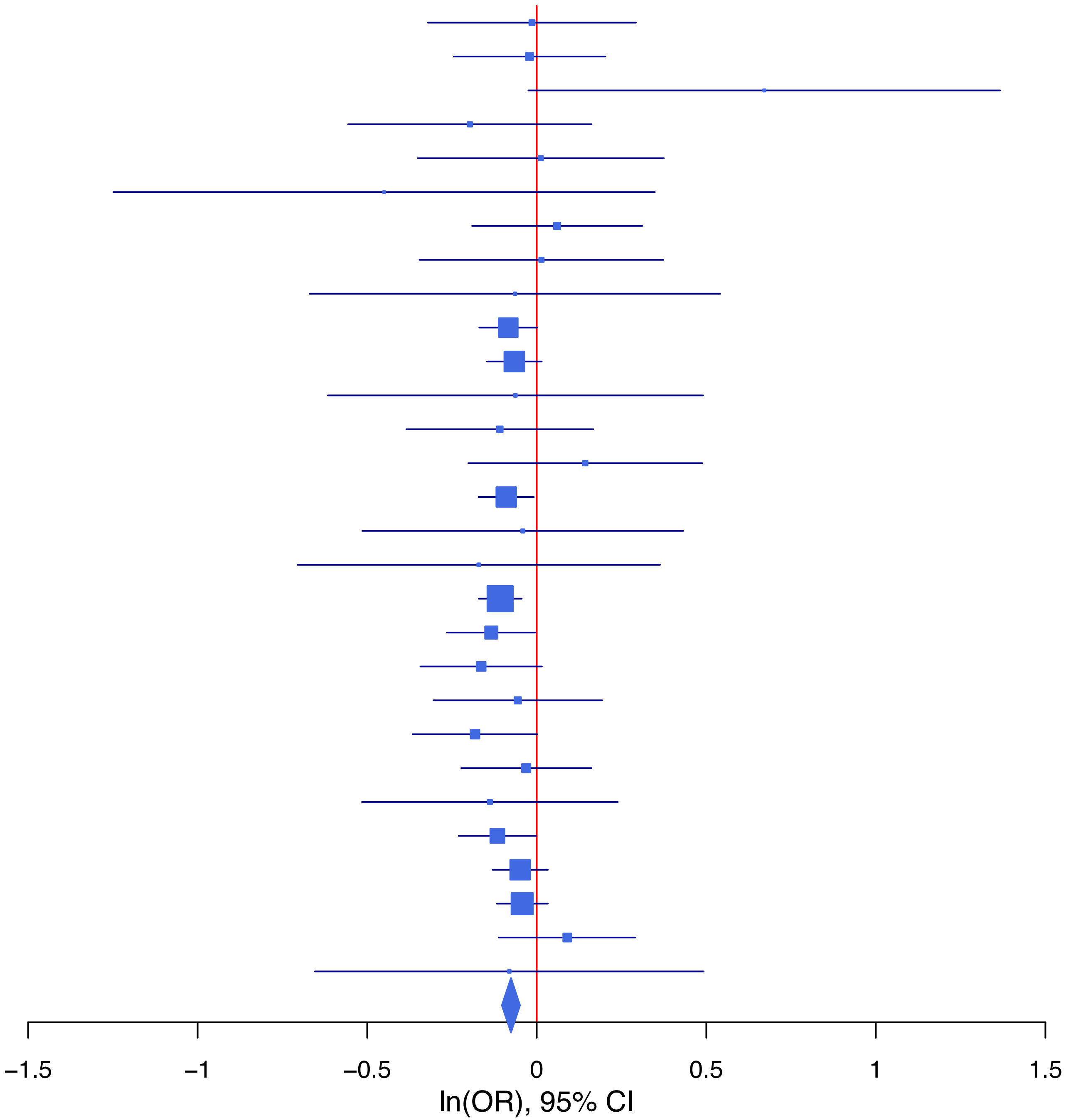

## rs7554968 T/C 1:73840843

| Cohort | P | ln(OR) | SE |
|---|---|---|---|
| ADHEA_AFR | 0.5818 | 0.063 | 0.114 |
| ADHEA_EUR | 0.5415 | 0.042 | 0.069 |
| BEPS7_EUR | 0.8846 | 0.026 | 0.179 |
| BHRCM_AFR | 0.8597 | 0.034 | 0.197 |
| BHRCM_EUR | 0.0197 | 0.372 | 0.16 |
| BHRCM_LAT | 0.6177 | 0.165 | 0.331 |
| BOR17_EUR | 0.8917 | −0.012 | 0.085 |
| BOR2C_EUR | 0.3385 | −0.081 | 0.085 |
| BOR2E_EUR | 0.6352 | 0.085 | 0.178 |
| CNVRG_EAS | 0.3113 | −0.057 | 0.056 |
| COGA1_AFR | 0.289 | 0.106 | 0.1 |
| COGA1_EUR | 0.1937 | 0.078 | 0.06 |
| CUINT_EUR | 0.2196 | 0.094 | 0.077 |
| CVEDA_CSA | 0.9627 | 0.008 | 0.163 |
| ESTB2_EUR | 0.8572 | 0.004 | 0.024 |
| FINNG_EUR | 0.2262 | 0.029 | 0.024 |
| GEDIS_EUR | 0.0055 | 0.366 | 0.132 |
| GISS1_EUR | 0.9363 | −0.006 | 0.08 |
| GISS2_EUR | 0.6179 | 0.047 | 0.093 |
| GREAT_EAS | 0.1538 | 0.145 | 0.102 |
| GTPRJ_AFR | 0.0421 | 0.12 | 0.059 |
| IPSYC_EUR | 0.2354 | 0.022 | 0.019 |
| JANS3_EUR | 0.0976 | −0.18 | 0.108 |
| JANS4_EUR | 0.0748 | −0.222 | 0.125 |
| JAPAN_EAS | 0.3333 | −0.065 | 0.067 |
| MIREC_AFR | 0.6177 | 0.07 | 0.139 |
| MIREC_EUR | 0.8457 | −0.03 | 0.152 |
| MVPXQ_AFR | 0.4747 | 0.015 | 0.021 |
| MVPXQ_EUR | 3.55e−05 | 0.051 | 0.012 |
| PGCBD_EUR | 0.9488 | 0.002 | 0.026 |
| PGCED_EUR | 0.331 | 0.112 | 0.115 |
| PGCMD_EUR | 0.1383 | 0.051 | 0.034 |
| PGCPT_EUR | 0.5008 | −0.04 | 0.059 |
| PGCSZ_EUR | 0.3234 | 0.037 | 0.038 |
| PRFCT_EUR | 0.0908 | 0.074 | 0.044 |
| PSYCR_EUR | 0.2049 | −0.116 | 0.091 |
| QIMRB_EUR | 0.1033 | 0.045 | 0.028 |
| SNUBH−ASA_EAS | 0.506 | 0.097 | 0.145 |
| SNUBH−KCHIP_EAS | 0.8625 | −0.035 | 0.205 |
| STRR1_LAT | 0.9114 | −0.02 | 0.176 |
| UKBJC_EUR | 0.0011 | 0.066 | 0.02 |
| UTAH2_EUR | 0.0578 | 0.037 | 0.02 |
| YPENN_EUR | 0.3662 | 0.118 | 0.13 |
| **meta** | **3.97e−09** | **0.035** | **0.006** |

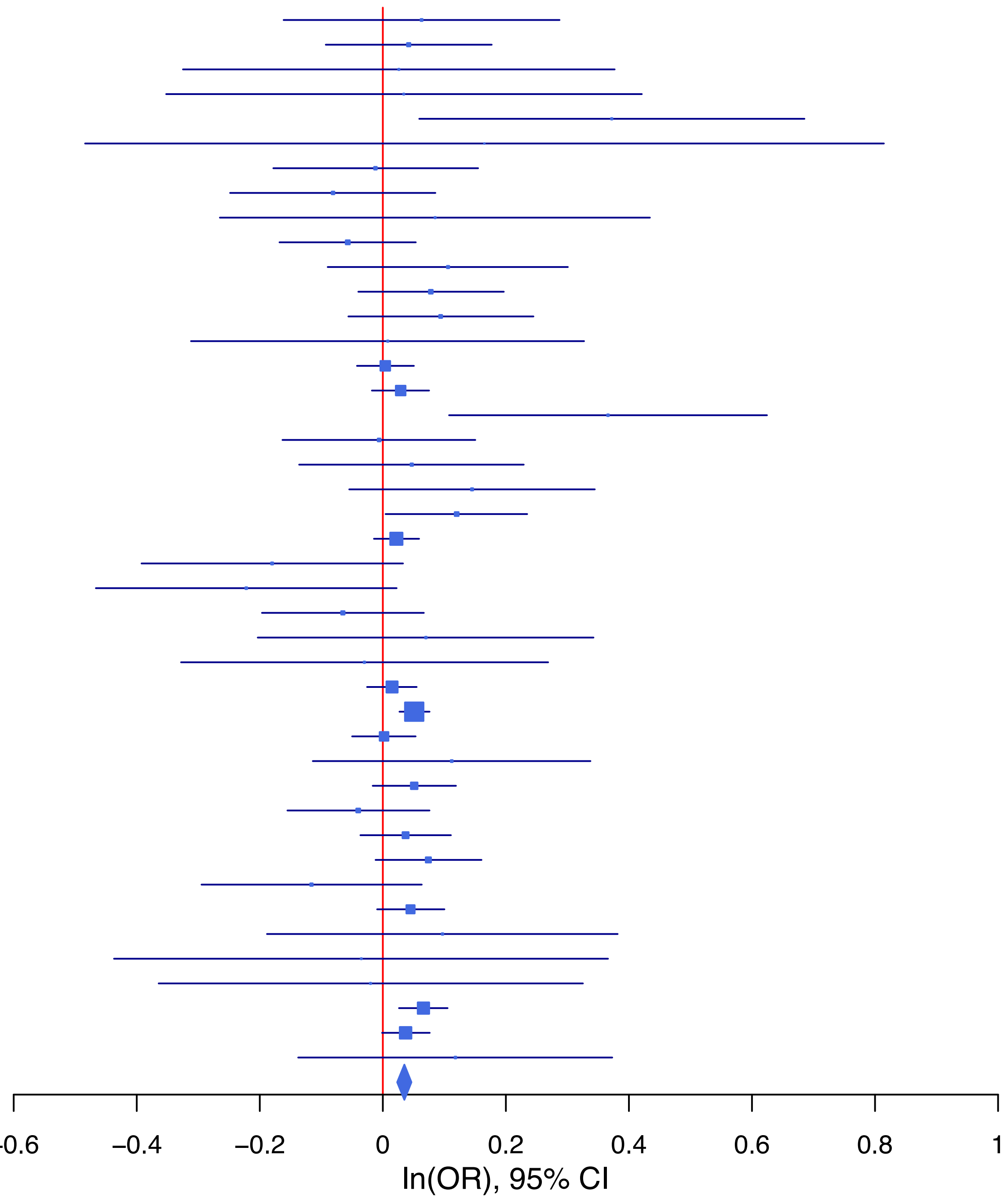

**rs7637711 G/A 3:49829653**

| Cohort | P | ln(OR) | SE |
|---|---|---|---|
| ADHEA_AFR | 0.9972 | −0.001 | 0.13 |
| ADHEA_EUR | 0.2641 | −0.099 | 0.089 |
| ALSPC_EUR | 0.7735 | 0.019 | 0.067 |
| BEPS7_EUR | 0.4612 | 0.166 | 0.225 |
| BHRCM_AFR | 0.2049 | 0.248 | 0.196 |
| BHRCM_EUR | 0.5998 | 0.102 | 0.194 |
| BOR17_EUR | 0.2726 | 0.121 | 0.111 |
| BOR2C_EUR | 0.0322 | 0.239 | 0.112 |
| BOR2E_EUR | 0.1133 | 0.363 | 0.229 |
| COGA1_AFR | 0.3425 | 0.108 | 0.114 |
| COGA1_EUR | 0.0569 | 0.143 | 0.075 |
| CUINT_EUR | 0.4163 | 0.073 | 0.09 |
| ESTB2_EUR | 0.5282 | 0.021 | 0.033 |
| FINNG_EUR | 0.3841 | 0.032 | 0.037 |
| GEDIS_EUR | 0.5904 | 0.087 | 0.161 |
| GISS1_EUR | 0.1382 | −0.163 | 0.11 |
| GISS2_EUR | 0.3661 | 0.114 | 0.126 |
| GTPRJ_AFR | 0.8322 | 0.014 | 0.065 |
| IPSYC_EUR | 0.0415 | 0.047 | 0.023 |
| JANS3_EUR | 0.7751 | −0.038 | 0.133 |
| JANS4_EUR | 0.8489 | −0.028 | 0.149 |
| MIREC_AFR | 0.1878 | 0.201 | 0.153 |
| MIREC_EUR | 0.6229 | −0.096 | 0.194 |
| MVPXQ_AFR | 0.4777 | 0.017 | 0.023 |
| MVPXQ_EUR | 6e−04 | 0.057 | 0.016 |
| MVPXQ_LAT | 0.6597 | 0.021 | 0.048 |
| PGCBD_EUR | 0.0057 | 0.091 | 0.033 |
| PGCED_EUR | 0.5704 | 0.082 | 0.144 |
| PGCPT_AFR | 0.2691 | 0.121 | 0.109 |
| PGCPT_EUR | 0.0049 | 0.196 | 0.07 |
| PGCSZ_EUR | 0.445 | 0.036 | 0.047 |
| PRFCT_EUR | 0.8413 | 0.011 | 0.056 |
| PSYCR_EUR | 0.3637 | −0.101 | 0.111 |
| QIMRB_EUR | 0.1003 | 0.058 | 0.035 |
| STRR1_LAT | 0.7848 | 0.07 | 0.257 |
| UKBJC_EUR | 0.0152 | 0.062 | 0.025 |
| UTAH2_EUR | 0.763 | 0.008 | 0.025 |
| UTAMR_LAT | 0.7128 | 0.024 | 0.066 |
| **meta** | **3.94e−09** | **0.045** | **0.008** |

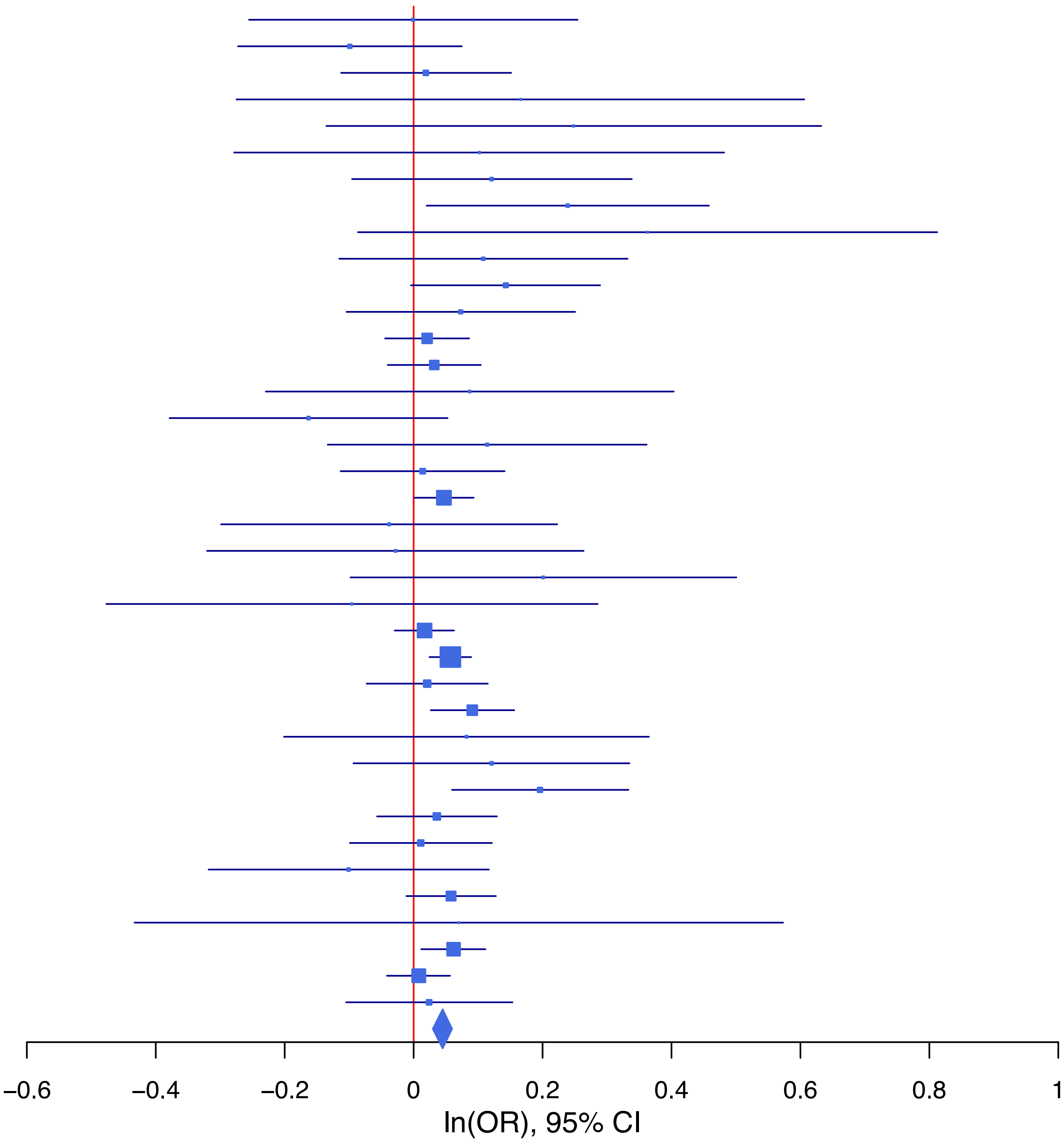

**rs7683962 A/G 4:143788874**

| Cohort | P | ln(OR) | SE |
|---|---|---|---|
| ADHEA_AFR | 0.1141 | −0.246 | 0.155 |
| ADHEA_EUR | 0.7213 | −0.029 | 0.08 |
| ALSPC_EUR | 0.8609 | −0.011 | 0.062 |
| BEPS7_EUR | 0.1933 | −0.298 | 0.229 |
| BHRCM_AFR | 0.7616 | −0.075 | 0.247 |
| BHRCM_EUR | 0.7984 | 0.048 | 0.187 |
| BHRCM_LAT | 0.8933 | −0.048 | 0.358 |
| BOR17_EUR | 0.7264 | 0.035 | 0.099 |
| BOR2C_EUR | 0.5118 | 0.067 | 0.101 |
| BOR2E_EUR | 0.0539 | −0.417 | 0.216 |
| CNVRG_EAS | 0.0232 | −0.152 | 0.067 |
| COGA1_AFR | 0.6774 | −0.053 | 0.128 |
| COGA1_EUR | 0.0039 | −0.198 | 0.069 |
| CUINT_EUR | 0.6931 | −0.033 | 0.084 |
| CVEDA_CSA | 0.8491 | 0.035 | 0.182 |
| ESTB2_EUR | 0.0168 | −0.065 | 0.027 |
| FINNG_EUR | 0.3699 | −0.026 | 0.029 |
| GEDIS_EUR | 0.9757 | −0.005 | 0.155 |
| GISS1_EUR | 0.2054 | −0.115 | 0.091 |
| GISS2_EUR | 0.1622 | −0.158 | 0.113 |
| GREAT_EAS | 0.8131 | −0.027 | 0.116 |
| GTPRJ_AFR | 0.5988 | −0.041 | 0.078 |
| IPSYC_EUR | 0.1319 | −0.033 | 0.022 |
| JANS3_EUR | 0.3229 | 0.119 | 0.121 |
| JANS4_EUR | 0.815 | −0.032 | 0.138 |
| JAPAN_EAS | 0.0887 | −0.13 | 0.077 |
| MVPXQ_AFR | 0.3393 | −0.026 | 0.027 |
| MVPXQ_EUR | 0.0416 | −0.03 | 0.015 |
| MVPXQ_LAT | 0.0441 | −0.082 | 0.041 |
| PGCBD_EUR | 0.4095 | −0.025 | 0.03 |
| PGCED_EUR | 0.5063 | 0.086 | 0.13 |
| PGCMD_EUR | 0.1664 | −0.057 | 0.041 |
| PGCPT_AFR | 0.3355 | −0.135 | 0.14 |
| PGCPT_EUR | 0.0714 | 0.118 | 0.065 |
| PGCSZ_EUR | 0.0264 | −0.099 | 0.045 |
| PRFCT_EUR | 0.514 | −0.035 | 0.054 |
| PSYCR_EUR | 0.2536 | −0.122 | 0.106 |
| QIMRB_EUR | 0.5826 | 0.017 | 0.032 |
| SNUBH−ASA_EAS | 0.5809 | −0.1 | 0.182 |
| SNUBH−KCHIP_EAS | 0.7086 | −0.089 | 0.238 |
| STRR1_LAT | 0.6082 | 0.104 | 0.203 |
| UKBJC_EUR | 0.4331 | −0.018 | 0.023 |
| UTAH2_EUR | 0.1517 | −0.032 | 0.022 |
| UTAMR_LAT | 0.013 | −0.12 | 0.048 |
| YPENN_AFR | 0.2026 | 0.235 | 0.184 |
| YPENN_EUR | 0.4463 | −0.116 | 0.153 |
| **meta** | **4.60e−08** | **−0.037** | **0.007** |

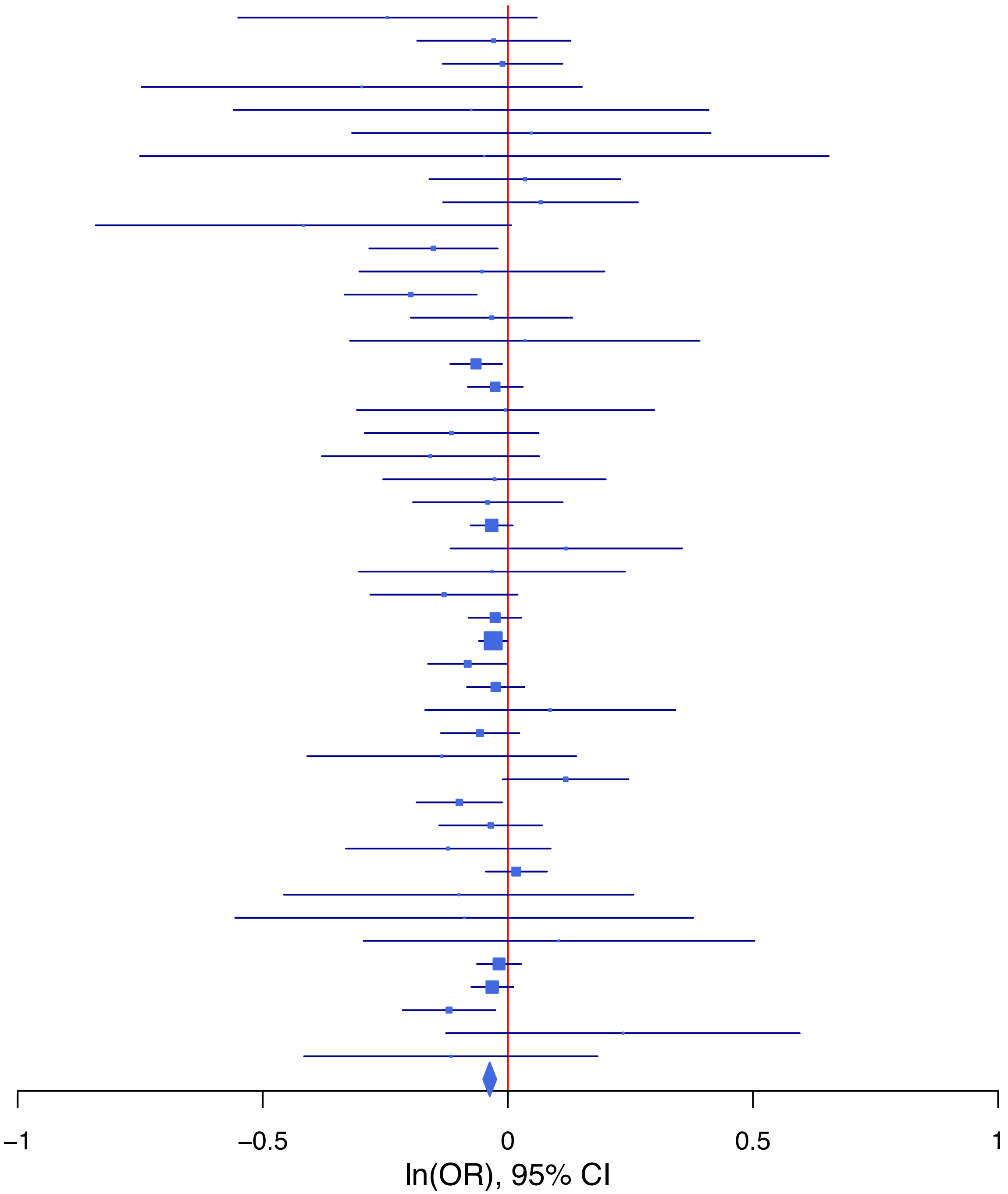

**rs77600213 G/A 6:65475897**

| Cohort | P | ln(OR) | SE |
|---|---|---|---|
| ADHEA_EUR | 0.9552 | 0.008 | 0.146 |
| ALSPC_EUR | 0.3262 | 0.107 | 0.109 |
| BHRCM_EUR | 0.4282 | −0.305 | 0.385 |
| BOR17_EUR | 0.5115 | 0.116 | 0.177 |
| BOR2C_EUR | 0.9636 | 0.009 | 0.187 |
| BOR2E_EUR | 0.9879 | −0.006 | 0.398 |
| CNVRG_EAS | 0.5422 | 0.033 | 0.055 |
| COGA1_EUR | 0.6399 | −0.059 | 0.126 |
| CUINT_EUR | 0.6384 | −0.079 | 0.168 |
| ESTB2_EUR | 0.0732 | 0.082 | 0.046 |
| FINNG_EUR | 0.3661 | 0.041 | 0.046 |
| GISS2_EUR | 0.2723 | 0.222 | 0.202 |
| GREAT_EAS | 0.077 | 0.16 | 0.09 |
| GTPRJ_AFR | 0.8767 | −0.045 | 0.291 |
| IPSYC_EUR | 0.1148 | 0.059 | 0.037 |
| JANS3_EUR | 0.8143 | 0.052 | 0.22 |
| JANS4_EUR | 0.3213 | 0.235 | 0.236 |
| JAPAN_EAS | 0.1042 | 0.117 | 0.072 |
| MIREC_EUR | 0.22 | −0.458 | 0.373 |
| MVPXQ_EUR | 0.0039 | 0.083 | 0.029 |
| MVPXQ_LAT | 0.0961 | 0.151 | 0.091 |
| PGCBD_EUR | 0.2276 | 0.074 | 0.061 |
| PGCMD_EUR | 0.5989 | −0.045 | 0.085 |
| PGCPT_EUR | 0.2725 | −0.144 | 0.132 |
| PGCSZ_EUR | 0.8504 | 0.018 | 0.095 |
| PRFCT_EUR | 0.9974 | 0 | 0.093 |
| PSYCR_EUR | 0.0897 | 0.338 | 0.199 |
| QIMRB_EUR | 0.2052 | 0.072 | 0.057 |
| SNUBH−ASA_EAS | 0.9034 | −0.019 | 0.153 |
| SNUBH−KCHIP_EAS | 0.0792 | 0.38 | 0.217 |
| UKBJC_EUR | 0.0152 | 0.097 | 0.04 |
| UTAH2_EUR | 0.0029 | 0.111 | 0.037 |
| UTAMR_LAT | 0.2908 | −0.103 | 0.097 |
| YPENN_EUR | 0.2829 | 0.273 | 0.254 |
| **meta** | **1.12e−08** | **0.07** | **0.012** |

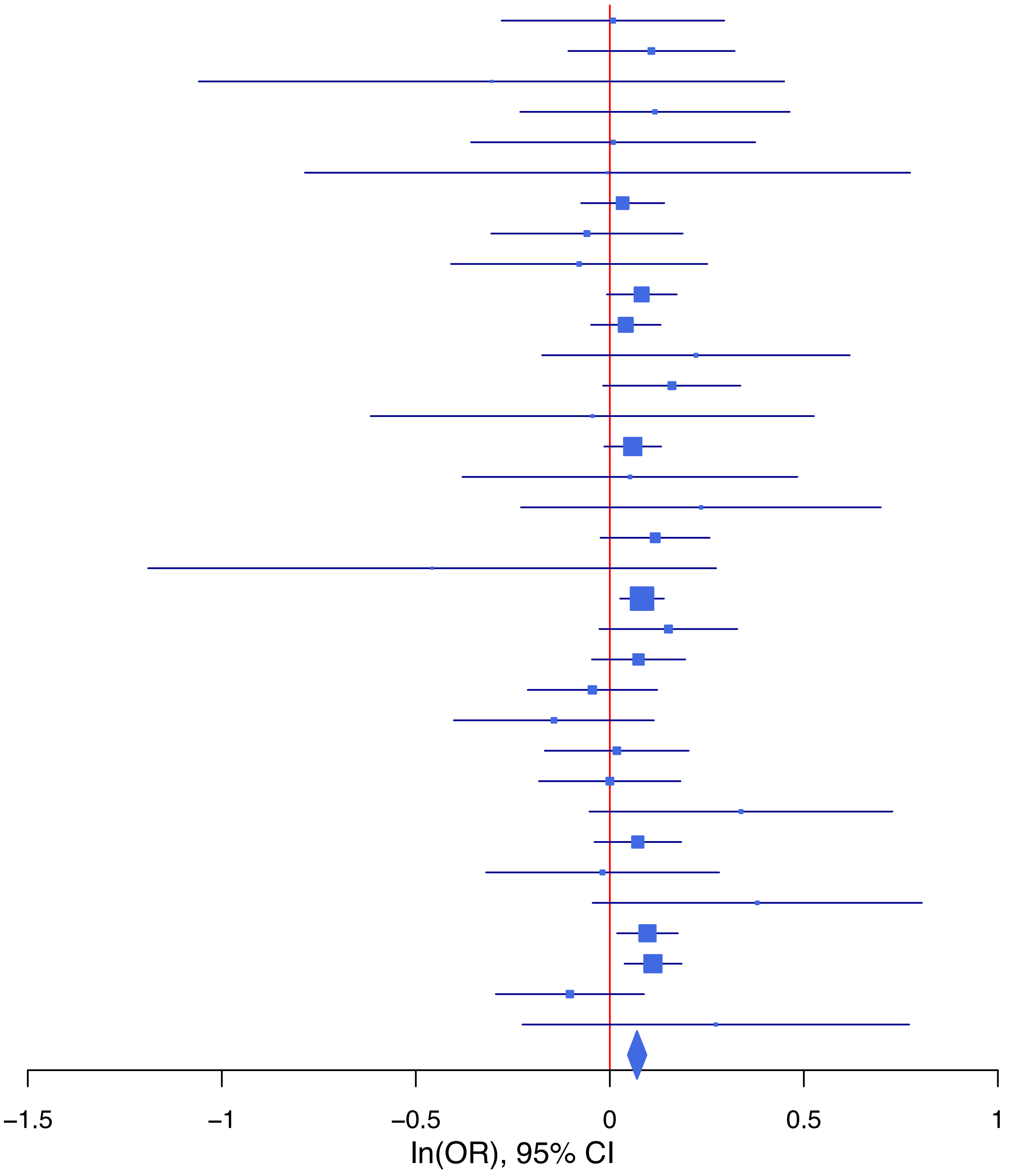

**rs7867749 C/T 9:127915210**

| Cohort | P | ln(OR) | SE |
|---|---|---|---|
| ALSPC_EUR | 0.0057 | −0.164 | 0.059 |
| BEPS7_EUR | 0.2604 | −0.225 | 0.2 |
| BHRCM_AFR | 0.0384 | 0.694 | 0.335 |
| BHRCM_EUR | 0.604 | 0.081 | 0.156 |
| BHRCM_LAT | 0.3276 | −0.585 | 0.598 |
| BOR17_EUR | 0.6142 | −0.046 | 0.092 |
| BOR2C_EUR | 0.1673 | −0.13 | 0.094 |
| BOR2E_EUR | 0.8812 | −0.028 | 0.19 |
| CNVRG_EAS | 0.3396 | −0.047 | 0.05 |
| COGA1_AFR | 0.1349 | −0.25 | 0.167 |
| COGA1_EUR | 0.0089 | −0.166 | 0.063 |
| CUINT_EUR | 0.8045 | −0.019 | 0.079 |
| CVEDA_CSA | 0.8045 | −0.05 | 0.204 |
| FINNG_EUR | 0.7801 | −0.008 | 0.03 |
| GEDIS_EUR | 0.8314 | −0.03 | 0.142 |
| GISS1_EUR | 0.3657 | −0.078 | 0.086 |
| GISS2_EUR | 0.6335 | −0.048 | 0.1 |
| GTPRJ_AFR | 0.3481 | −0.103 | 0.11 |
| IPSYC_EUR | 0.074 | −0.036 | 0.02 |
| JANS3_EUR | 0.0994 | −0.194 | 0.118 |
| JANS4_EUR | 0.6192 | −0.063 | 0.127 |
| JAPAN_EAS | 0.202 | −0.074 | 0.058 |
| MIREC_AFR | 0.4553 | 0.185 | 0.247 |
| MIREC_EUR | 0.0919 | 0.273 | 0.162 |
| MVPXQ_AFR | 0.7504 | −0.011 | 0.035 |
| MVPXQ_EAS | 0.1753 | 0.151 | 0.111 |
| MVPXQ_EUR | 1.83e−05 | −0.057 | 0.013 |
| MVPXQ_LAT | 0.2139 | −0.05 | 0.04 |
| PGCBD_EUR | 0.1611 | −0.039 | 0.028 |
| PGCED_EUR | 0.6409 | 0.059 | 0.128 |
| PGCMD_EUR | 0.2943 | −0.039 | 0.037 |
| PGCPT_EUR | 0.8862 | 0.009 | 0.062 |
| PGCSZ_EUR | 0.7938 | 0.011 | 0.04 |
| PRFCT_EUR | 0.862 | −0.008 | 0.048 |
| PSYCR_EUR | 0.3433 | −0.088 | 0.093 |
| QIMRB_EUR | 0.3257 | −0.029 | 0.029 |
| SNUBH−KCHIP_EAS | 0.3039 | 0.209 | 0.203 |
| STRR1_LAT | 0.8642 | −0.036 | 0.212 |
| UKBJC_EUR | 0.9287 | −0.002 | 0.021 |
| UTAH2_EUR | 0.0058 | −0.057 | 0.021 |
| UTAMR_LAT | 0.8161 | −0.012 | 0.052 |
| YPENN_AFR | 0.8428 | 0.046 | 0.23 |
| YPENN_EUR | 0.8089 | −0.033 | 0.137 |
| **meta** | **3.61e−09** | **−0.039** | **0.007** |

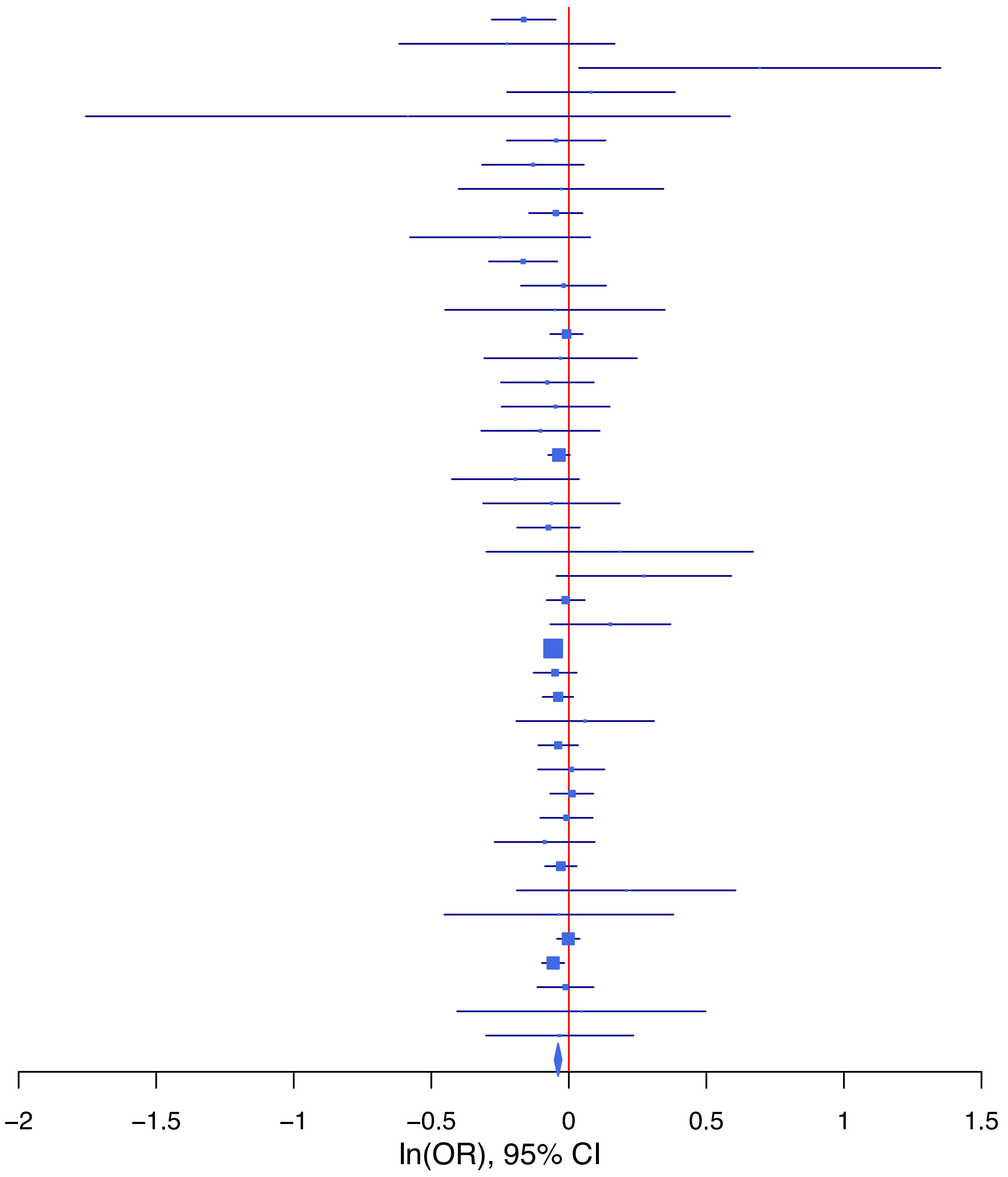

**rs78940908 G/C 2:58921049**

| Cohort | P | ln(OR) | SE |
|---|---|---|---|
| ADHEA_AFR | 0.3614 | −0.108 | 0.119 |
| ADHEA_EUR | 0.0089 | −0.183 | 0.07 |
| BEPS7_EUR | 0.9368 | −0.015 | 0.185 |
| BHRCM_AFR | 0.062 | −0.382 | 0.204 |
| BHRCM_LAT | 0.8783 | 0.037 | 0.241 |
| BOR17_EUR | 0.5683 | −0.05 | 0.087 |
| BOR2C_EUR | 0.565 | −0.05 | 0.087 |
| BOR2E_EUR | 0.3827 | −0.156 | 0.179 |
| CNVRG_EAS | 0.5883 | 0.026 | 0.048 |
| COGA1_AFR | 0.1021 | −0.167 | 0.102 |
| COGA1_EUR | 0.5856 | −0.032 | 0.059 |
| CUINT_EUR | 0.4746 | −0.055 | 0.077 |
| ESTB2_EUR | 0.2309 | −0.028 | 0.023 |
| FINNG_EUR | 0.7245 | 0.008 | 0.024 |
| GEDIS_EUR | 0.5801 | 0.071 | 0.129 |
| GISS1_EUR | 0.6598 | 0.035 | 0.08 |
| GISS2_EUR | 0.4138 | −0.076 | 0.093 |
| GREAT_EAS | 0.9599 | −0.004 | 0.076 |
| GTPRJ_AFR | 0.4787 | 0.042 | 0.059 |
| IPSYC_EUR | 0.0131 | −0.046 | 0.019 |
| JANS3_EUR | 0.7527 | 0.034 | 0.106 |
| JANS4_EUR | 0.9779 | −0.003 | 0.118 |
| MVPXQ_EUR | 0.0026 | −0.039 | 0.013 |
| PGCBD_EUR | 0.239 | −0.031 | 0.026 |
| PGCED_EUR | 0.4613 | 0.083 | 0.113 |
| PGCMD_EUR | 0.5107 | −0.023 | 0.035 |
| PGCPT_AFR | 0.0962 | −0.171 | 0.103 |
| PGCPT_EUR | 0.264 | −0.065 | 0.058 |
| PGCSZ_EUR | 0.4735 | −0.027 | 0.037 |
| PRFCT_EUR | 0.4512 | −0.033 | 0.044 |
| PSYCR_EUR | 0.7609 | −0.026 | 0.087 |
| QIMRB_EUR | 0.0193 | −0.064 | 0.028 |
| SNUBH−ASA_EAS | 0.0603 | −0.243 | 0.13 |
| SNUBH−KCHIP_EAS | 0.3454 | −0.16 | 0.17 |
| STRR1_LAT | 0.2594 | 0.197 | 0.174 |
| UKBJC_EUR | 0.0047 | −0.056 | 0.02 |
| UTAMR_LAT | 0.3649 | −0.034 | 0.037 |
| YPENN_AFR | 0.1816 | −0.197 | 0.148 |
| YPENN_EUR | 0.4275 | 0.103 | 0.13 |
| **meta** | **1.24e−08** | **−0.037** | **0.006** |

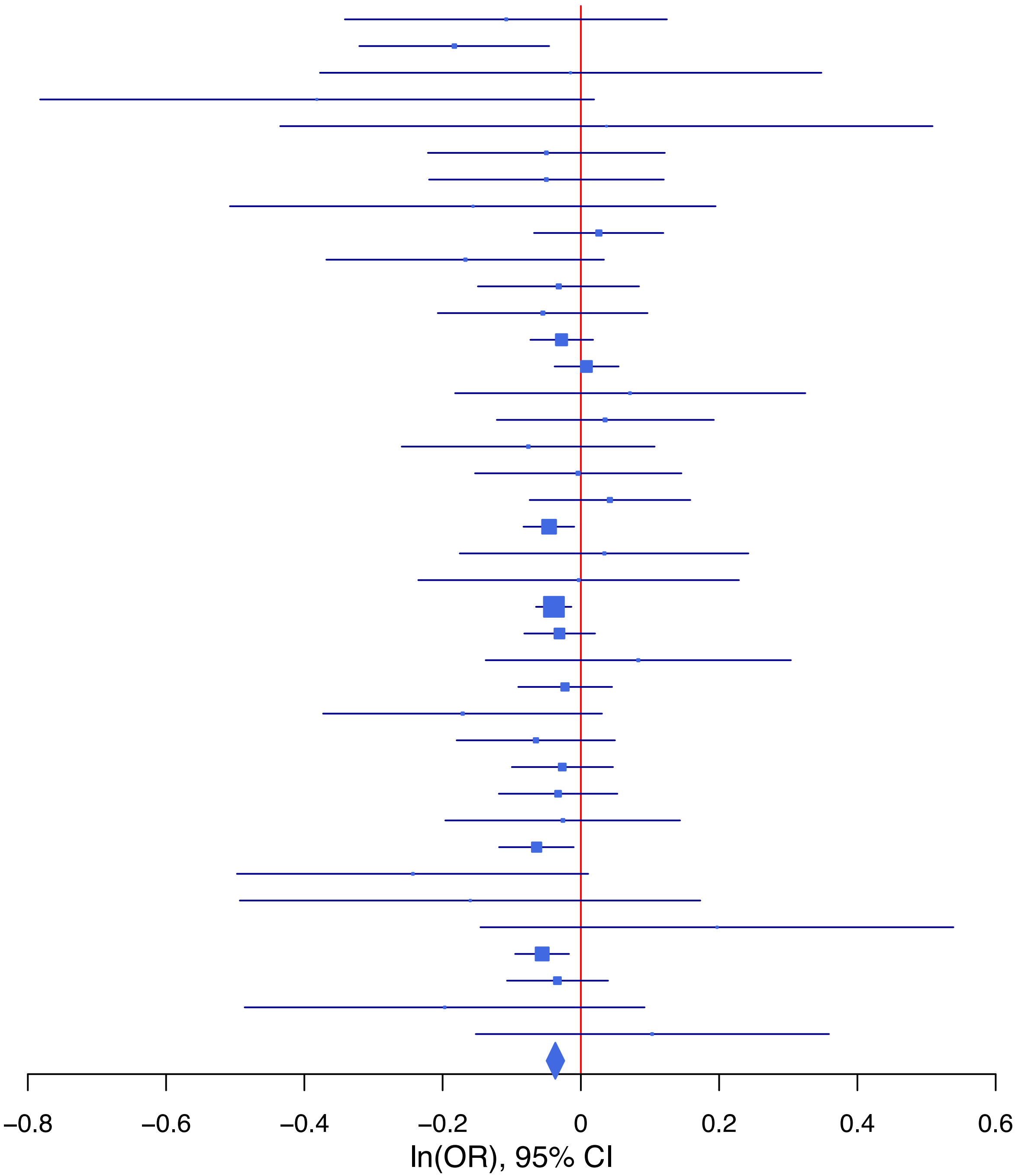

## rs8008844 G/T 14:98631075

| Cohort | P | ln(OR) | SE |
|---|---|---|---|
| ADHEA_AFR | 0.6282 | 0.056 | 0.116 |
| ADHEA_EUR | 0.9693 | 0.003 | 0.07 |
| ALSPC_EUR | 0.1294 | 0.085 | 0.056 |
| BEPS7_EUR | 0.6779 | 0.076 | 0.183 |
| BHRCM_AFR | 0.5397 | 0.11 | 0.179 |
| BHRCM_EUR | 0.0808 | 0.273 | 0.156 |
| BHRCM_LAT | 0.9156 | 0.046 | 0.437 |
| BOR17_EUR | 0.914 | 0.009 | 0.085 |
| BOR2C_EUR | 0.9711 | 0.003 | 0.089 |
| BOR2E_EUR | 0.0996 | 0.296 | 0.18 |
| CNVRG_EAS | 0.7539 | −0.02 | 0.062 |
| COGA1_AFR | 0.1602 | 0.143 | 0.102 |
| COGA1_EUR | 0.0452 | 0.123 | 0.062 |
| CUINT_EUR | 0.4444 | 0.058 | 0.076 |
| CVEDA_CSA | 0.3014 | 0.183 | 0.177 |
| ESTB2_EUR | 0.3671 | 0.022 | 0.024 |
| FINNG_EUR | 0.1171 | 0.041 | 0.026 |
| GEDIS_EUR | 0.813 | 0.031 | 0.129 |
| GISS1_EUR | 0.4611 | −0.06 | 0.082 |
| GISS2_EUR | 0.3881 | −0.082 | 0.095 |
| GREAT_EAS | 0.9098 | 0.011 | 0.095 |
| IPSYC_EUR | 0.1983 | 0.025 | 0.019 |
| JANS3_EUR | 0.4668 | 0.077 | 0.106 |
| JANS4_EUR | 0.4462 | 0.092 | 0.121 |
| JAPAN_EAS | 0.1925 | −0.137 | 0.105 |
| MIREC_AFR | 0.9526 | −0.008 | 0.141 |
| MIREC_EUR | 0.145 | 0.226 | 0.156 |
| MVPXQ_EAS | 0.0357 | −0.244 | 0.116 |
| MVPXQ_EUR | 0.071 | 0.024 | 0.013 |
| MVPXQ_LAT | 0.4476 | 0.028 | 0.037 |
| PGCBD_EUR | 0.0303 | 0.057 | 0.026 |
| PGCED_EUR | 0.0393 | 0.234 | 0.113 |
| PGCMD_EUR | 0.1417 | 0.052 | 0.035 |
| PGCPT_AFR | 0.8877 | 0.015 | 0.107 |
| PGCPT_EUR | 0.4645 | 0.043 | 0.059 |
| PGCSZ_EUR | 0.4022 | 0.032 | 0.038 |
| PRFCT_EUR | 0.1801 | 0.062 | 0.046 |
| PSYCR_EUR | 0.5201 | −0.058 | 0.09 |
| QIMRB_EUR | 0.0802 | 0.049 | 0.028 |
| SNUBH−ASA_EAS | 0.7628 | −0.059 | 0.195 |
| SNUBH−KCHIP_EAS | 0.9906 | −0.003 | 0.262 |
| STRR1_LAT | 0.7873 | −0.053 | 0.196 |
| UKBJC_EUR | 0.0039 | 0.059 | 0.02 |
| UTAH2_EUR | 0.0396 | 0.041 | 0.02 |
| UTAMR_LAT | 0.6669 | −0.019 | 0.045 |
| YPENN_AFR | 0.646 | 0.066 | 0.143 |
| YPENN_EUR | 0.6764 | −0.054 | 0.13 |
| **meta** | **1.44e−08** | **0.035** | **0.006** |

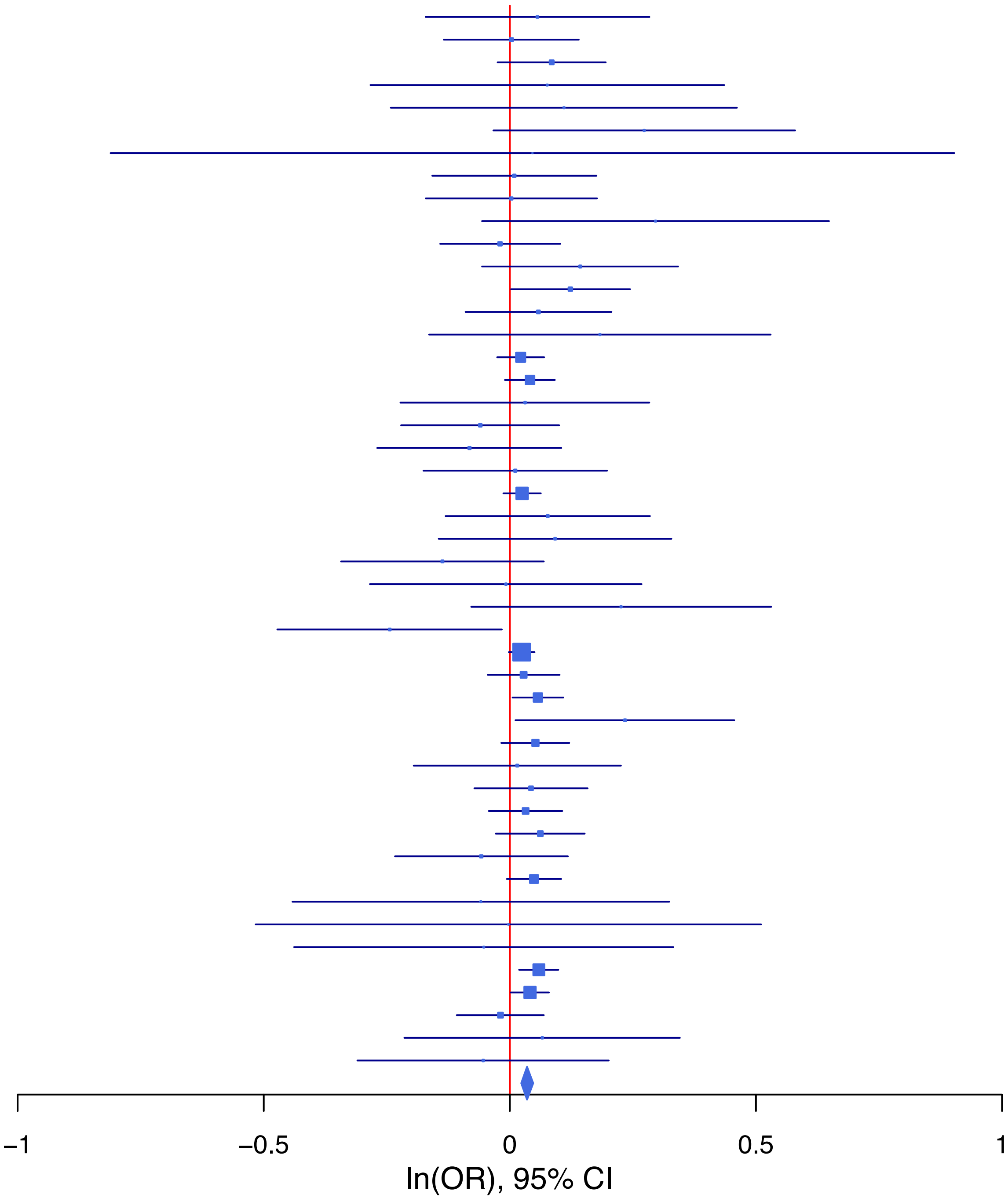

**rs8013266 C/T 14:103326780**

| Cohort | P | ln(OR) | SE |
|---|---|---|---|
| ALSPC_EUR | 0.009 | 0.169 | 0.065 |
| BEPS7_EUR | 0.8619 | 0.039 | 0.226 |
| BHRCM_AFR | 0.3574 | 0.241 | 0.261 |
| BHRCM_EUR | 0.8759 | 0.031 | 0.204 |
| BHRCM_LAT | 0.0641 | 0.434 | 0.234 |
| BOR17_EUR | 0.6035 | 0.056 | 0.109 |
| BOR2C_EUR | 0.3365 | −0.104 | 0.108 |
| BOR2E_EUR | 0.4177 | −0.168 | 0.208 |
| CNVRG_EAS | 0.0635 | 0.086 | 0.046 |
| COGA1_AFR | 0.5206 | −0.069 | 0.107 |
| CVEDA_CSA | 0.5384 | −0.157 | 0.255 |
| ESTB2_EUR | 0.1011 | 0.047 | 0.029 |
| FINNG_EUR | 0.0081 | 0.071 | 0.027 |
| GEDIS_EUR | 0.7571 | −0.049 | 0.16 |
| GISS1_EUR | 0.3859 | −0.089 | 0.102 |
| GISS2_EUR | 0.7295 | 0.04 | 0.116 |
| GREAT_EAS | 0.9233 | −0.008 | 0.078 |
| IPSYC_EUR | 0.1108 | 0.036 | 0.023 |
| JANS3_EUR | 0.9404 | −0.01 | 0.132 |
| JANS4_EUR | 0.1947 | −0.191 | 0.147 |
| MIREC_AFR | 0.346 | 0.138 | 0.146 |
| MIREC_EUR | 0.9825 | −0.004 | 0.19 |
| MVPXQ_EAS | 0.7098 | −0.039 | 0.105 |
| MVPXQ_EUR | 2.64e−06 | 0.074 | 0.016 |
| MVPXQ_LAT | 0.3619 | 0.032 | 0.035 |
| PGCBD_EUR | 0.5336 | −0.02 | 0.033 |
| PGCMD_EUR | 0.9094 | 0.005 | 0.044 |
| PGCPT_AFR | 0.8451 | 0.022 | 0.111 |
| PGCPT_EUR | 0.2249 | 0.086 | 0.07 |
| PGCSZ_EUR | 0.4226 | 0.037 | 0.046 |
| PRFCT_EUR | 0.0468 | 0.103 | 0.052 |
| PSYCR_EUR | 0.1717 | 0.154 | 0.113 |
| QIMRB_EUR | 0.6146 | 0.017 | 0.035 |
| SNUBH-ASA_EAS | 0.3091 | 0.127 | 0.125 |
| SNUBH-KCHIP_EAS | 0.6814 | −0.067 | 0.164 |
| STRR1_LAT | 0.2085 | −0.246 | 0.196 |
| UKBJC_EUR | 0.0014 | 0.078 | 0.024 |
| UTAH2_EUR | 0.2892 | 0.025 | 0.024 |
| UTAMR_LAT | 0.0621 | 0.069 | 0.037 |
| YPENN_AFR | 0.1448 | 0.232 | 0.159 |
| YPENN_EUR | 0.6367 | −0.075 | 0.159 |
| **meta** | **1.54e−11** | **0.049** | **0.007** |

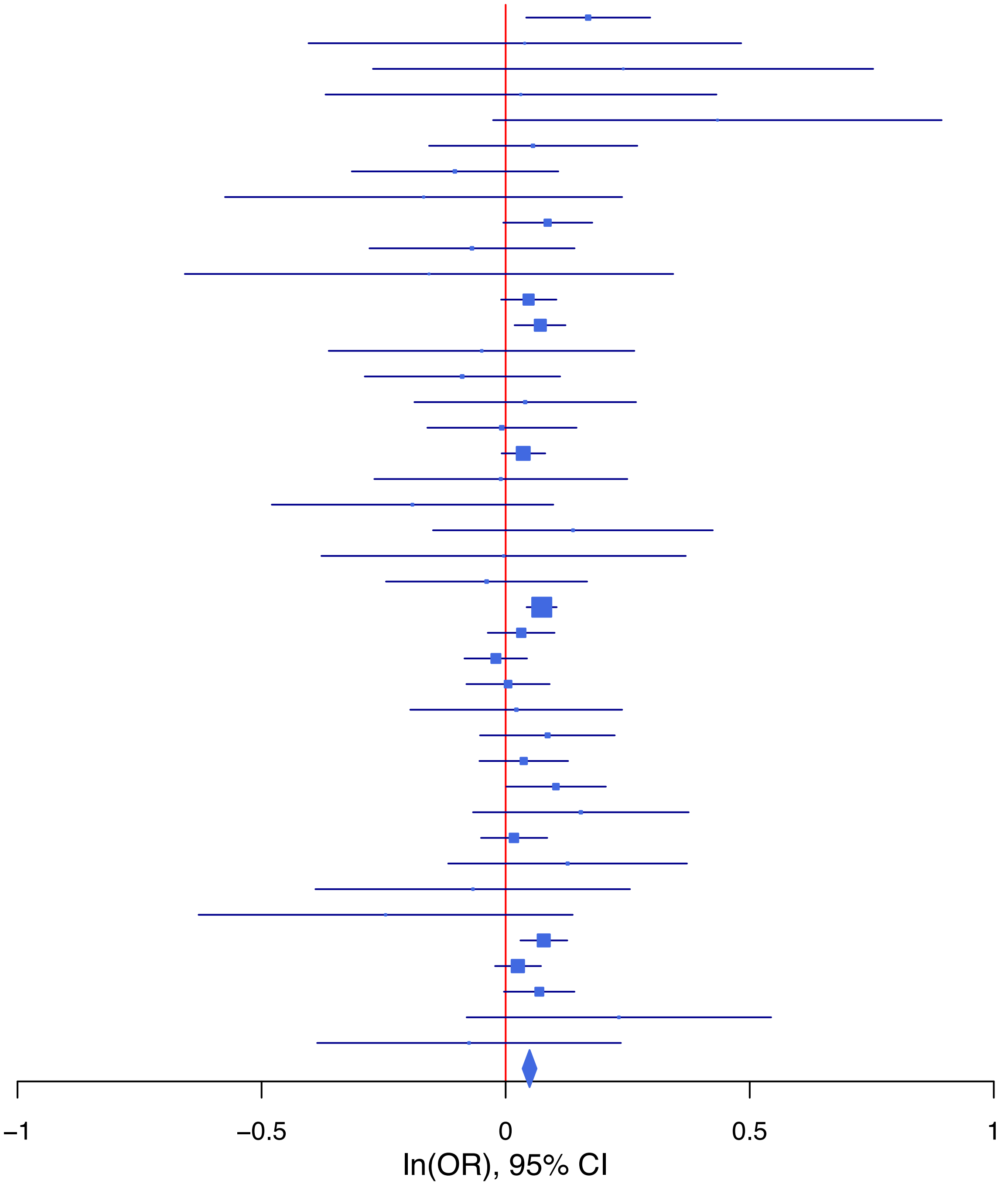

**rs9592599 A/T 13:69575338**

| Cohort | P | ln(OR) | SE |
|---|---|---|---|
| ALSPC_EUR | 0.7737 | −0.016 | 0.055 |
| BEPS7_EUR | 0.79 | −0.048 | 0.182 |
| BHRCM_AFR | 0.4093 | 0.166 | 0.202 |
| BHRCM_LAT | 0.1179 | 0.9 | 0.576 |
| BOR17_EUR | 0.1048 | −0.142 | 0.087 |
| BOR2C_EUR | 0.0125 | −0.219 | 0.088 |
| BOR2E_EUR | 0.8015 | 0.045 | 0.179 |
| CNVRG_EAS | 0.2296 | −0.058 | 0.048 |
| COGA1_AFR | 0.3208 | 0.106 | 0.107 |
| COGA1_EUR | 0.4793 | −0.042 | 0.059 |
| CVEDA_CSA | 0.6597 | −0.072 | 0.163 |
| ESTB2_EUR | 0.0028 | −0.069 | 0.023 |
| GEDIS_EUR | 0.6842 | 0.054 | 0.131 |
| GISS1_EUR | 0.0426 | −0.161 | 0.08 |
| GISS2_EUR | 0.3333 | 0.087 | 0.09 |
| GREAT_EAS | 0.9955 | 0 | 0.08 |
| GTPRJ_AFR | 0.7714 | −0.018 | 0.062 |
| IPSYC_EUR | 0.4804 | −0.013 | 0.019 |
| JANS3_EUR | 0.7732 | −0.03 | 0.104 |
| JANS4_EUR | 0.0904 | −0.206 | 0.122 |
| MIREC_EUR | 0.3195 | −0.153 | 0.154 |
| MVPXQ_EAS | 0.373 | −0.103 | 0.116 |
| PGCBD_EUR | 0.0152 | −0.063 | 0.026 |
| PGCED_EUR | 0.5551 | 0.069 | 0.118 |
| PGCMD_EUR | 0.175 | −0.047 | 0.035 |
| PGCPT_AFR | 0.2387 | −0.125 | 0.106 |
| PGCPT_EUR | 0.1014 | −0.095 | 0.058 |
| PGCSZ_EUR | 0.3029 | −0.039 | 0.037 |
| PRFCT_EUR | 0.1373 | −0.065 | 0.044 |
| PSYCR_EUR | 0.8664 | −0.015 | 0.088 |
| QIMRB_EUR | 0.2993 | −0.028 | 0.027 |
| SNUBH−ASA_EAS | 0.6974 | −0.055 | 0.141 |
| STRR1_LAT | 0.6281 | −0.083 | 0.172 |
| UKBJC_EUR | 0.2412 | −0.023 | 0.02 |
| UTAH2_EUR | 0.0074 | −0.052 | 0.02 |
| UTAMR_LAT | 0.0015 | −0.114 | 0.036 |
| YPENN_AFR | 0.0145 | −0.36 | 0.147 |
| YPENN_EUR | 0.4762 | −0.094 | 0.131 |
| **meta** | **2.36e−10** | **−0.046** | **0.007** |

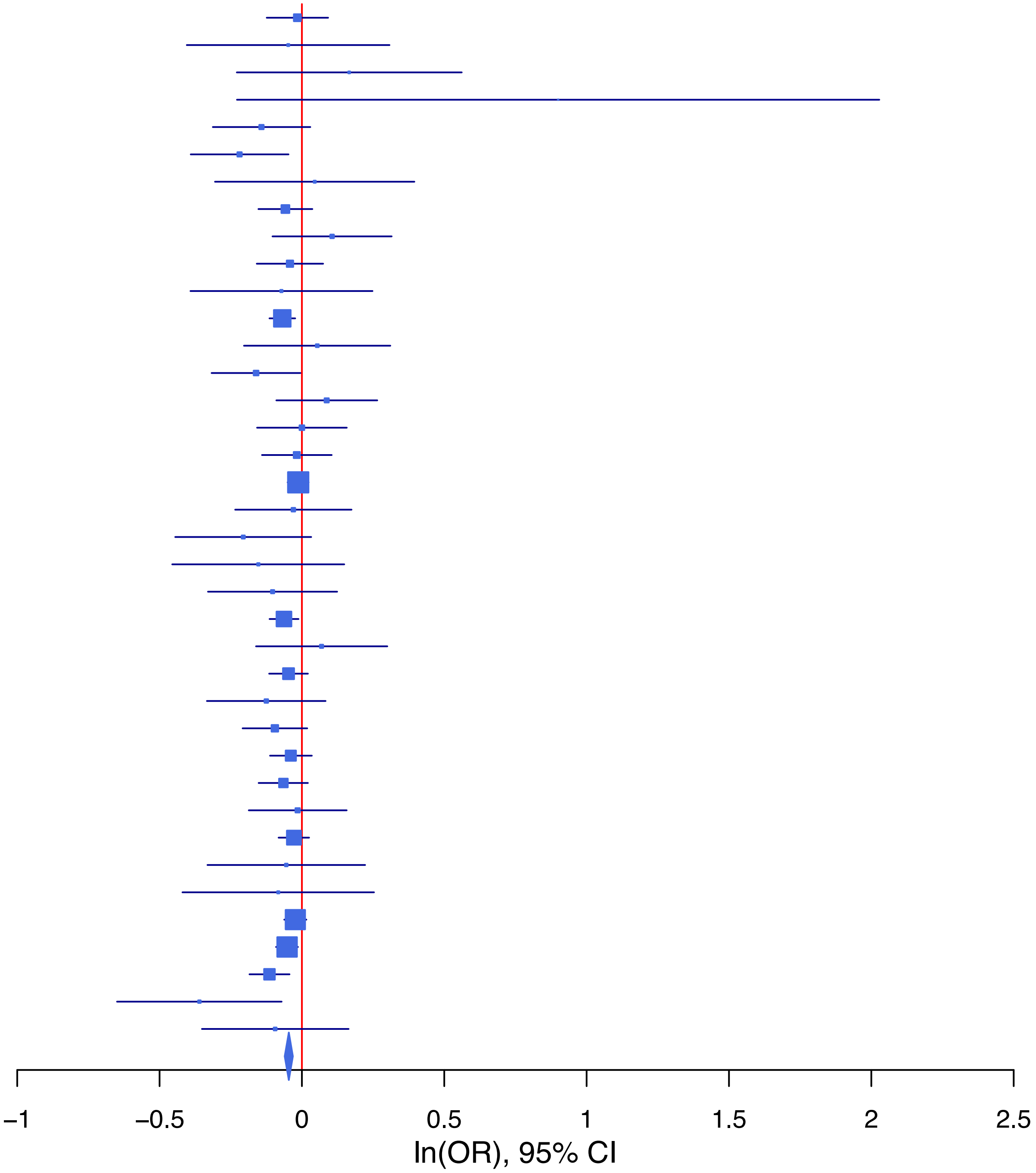

**Supplementary Data 1H: Forest plots of lead SNPs at the 46 genome-wide significant loci from the GWAS meta-analysis of suicidal behavior in European ancestry samples.**
Each box represents the log odds ratio (OR) from an individual contributing cohort, with horizontal lines indicating the 95% confidence interval (CI). The diamond represents the overall meta-analytic estimate across studies.

**rs1079595 C/A 11:113282669**

| Cohort | P | ln(OR) | SE |
|---|---|---|---|
| ADHEA_EUR | 0.0408 | 0.181 | 0.089 |
| ALSPC_EUR | 0.6574 | 0.033 | 0.074 |
| BEPS7_EUR | 0.8757 | −0.038 | 0.244 |
| BHRCM_EUR | 0.2036 | 0.29 | 0.228 |
| BOR17_EUR | 0.4863 | −0.082 | 0.118 |
| BOR2C_EUR | 0.3698 | 0.107 | 0.119 |
| BOR2E_EUR | 0.5201 | −0.184 | 0.286 |
| COGA1_EUR | 0.4003 | 0.069 | 0.082 |
| CUINT_EUR | 0.1645 | 0.145 | 0.104 |
| ESTB2_EUR | 0.2019 | 0.037 | 0.029 |
| FINNG_EUR | 0.226 | 0.035 | 0.029 |
| GEDIS_EUR | 0.4695 | −0.133 | 0.184 |
| GISS1_EUR | 0.2015 | 0.131 | 0.102 |
| GISS2_EUR | 0.4442 | −0.09 | 0.118 |
| IPSYC_EUR | 6e−04 | 0.086 | 0.025 |
| JANS3_EUR | 0.1307 | 0.202 | 0.134 |
| JANS4_EUR | 0.5566 | −0.096 | 0.163 |
| MIREC_EUR | 0.1844 | 0.259 | 0.196 |
| MVPXQ_EUR | 3e−04 | 0.06 | 0.017 |
| PGCBD_EUR | 0.3215 | 0.035 | 0.035 |
| PGCED_EUR | 0.7775 | 0.049 | 0.173 |
| PGCMD_EUR | 0.0563 | 0.092 | 0.048 |
| PGCPT_EUR | 0.244 | 0.088 | 0.076 |
| PGCSZ_EUR | 0.9622 | 0.002 | 0.051 |
| PRFCT_EUR | 0.0331 | 0.124 | 0.058 |
| PSYCR_EUR | 0.566 | 0.068 | 0.119 |
| QIMRB_EUR | 0.6018 | 0.019 | 0.037 |
| UKBJC_EUR | 0.1342 | 0.04 | 0.027 |
| UTAH2_EUR | 0.3006 | 0.026 | 0.025 |
| YPENN_EUR | 0.5229 | −0.114 | 0.179 |
| **meta** | **6.53e−10** | **0.051** | **0.008** |

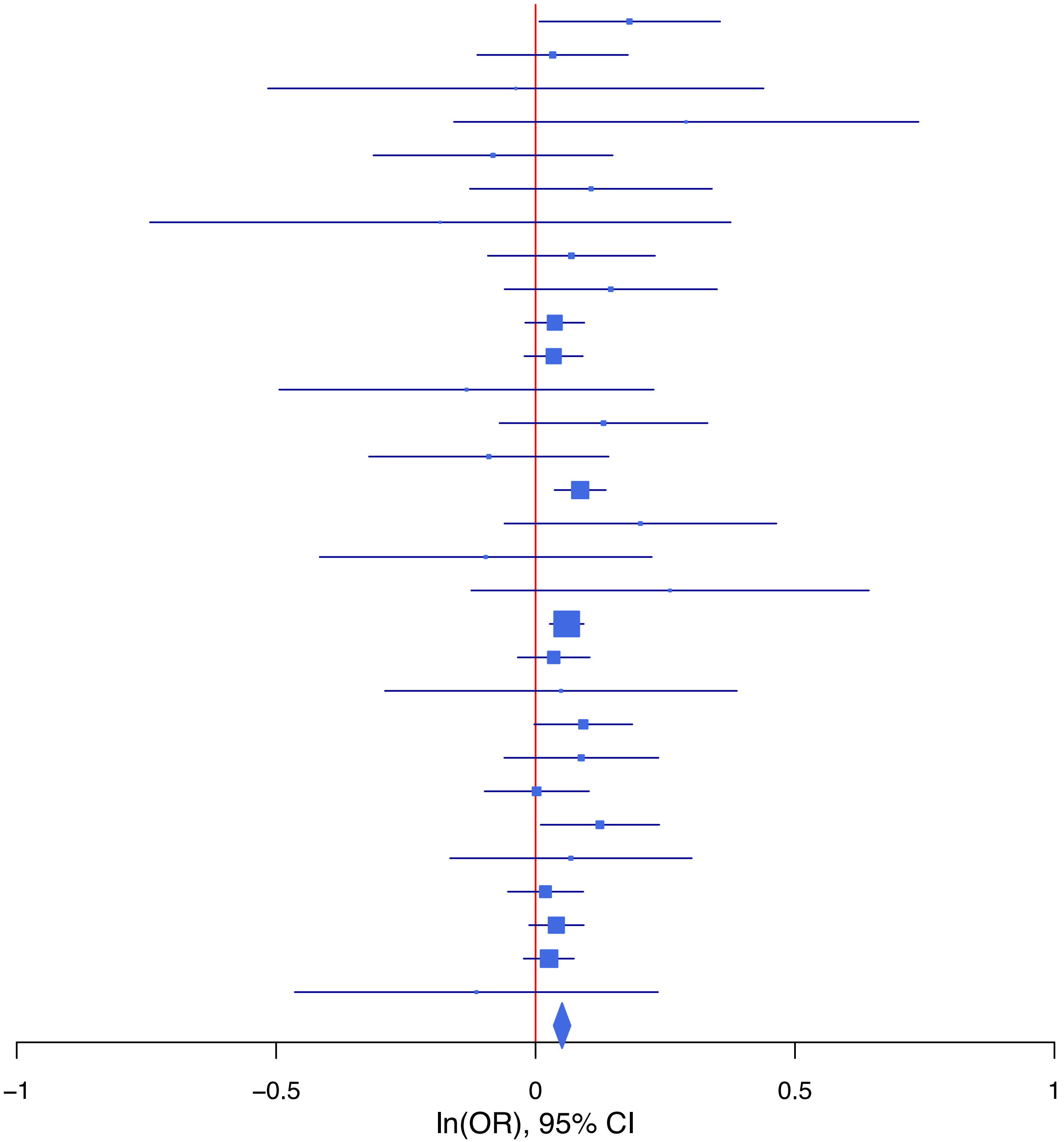

**rs10979816   G/A   9:112067488**

| Cohort | P | ln(OR) | SE |
|---|---|---|---|
| ADHEA_EUR | 0.0356 | 0.212 | 0.101 |
| ALSPC_EUR | 0.508 | −0.057 | 0.086 |
| BEPS7_EUR | 0.9336 | 0.025 | 0.296 |
| BOR17_EUR | 0.2007 | 0.194 | 0.152 |
| BOR2C_EUR | 0.4763 | 0.105 | 0.147 |
| BOR2E_EUR | 0.4387 | 0.234 | 0.303 |
| COGA1_EUR | 0.4048 | 0.077 | 0.093 |
| ESTB2_EUR | 0.0063 | 0.1 | 0.037 |
| GISS1_EUR | 0.7888 | 0.036 | 0.134 |
| GISS2_EUR | 0.0677 | 0.282 | 0.154 |
| IPSYC_EUR | 0.0776 | 0.052 | 0.03 |
| JANS3_EUR | 0.6483 | −0.077 | 0.17 |
| JANS4_EUR | 0.0913 | 0.322 | 0.19 |
| PGCBD_EUR | 0.0396 | 0.088 | 0.043 |
| PGCED_EUR | 0.9698 | 0.008 | 0.215 |
| PGCMD_EUR | 0.9144 | 0.006 | 0.059 |
| PGCPT_EUR | 0.3866 | 0.078 | 0.09 |
| PGCSZ_EUR | 0.0075 | 0.157 | 0.059 |
| PRFCT_EUR | 0.5306 | −0.045 | 0.072 |
| PSYCR_EUR | 0.9659 | 0.006 | 0.147 |
| QIMRB_EUR | 0.1555 | 0.06 | 0.042 |
| UKBJC_EUR | 0.0046 | 0.083 | 0.029 |
| YPENN_EUR | 0.9838 | 0.004 | 0.213 |
| **meta** | **3.21e−08** | **0.073** | **0.013** |

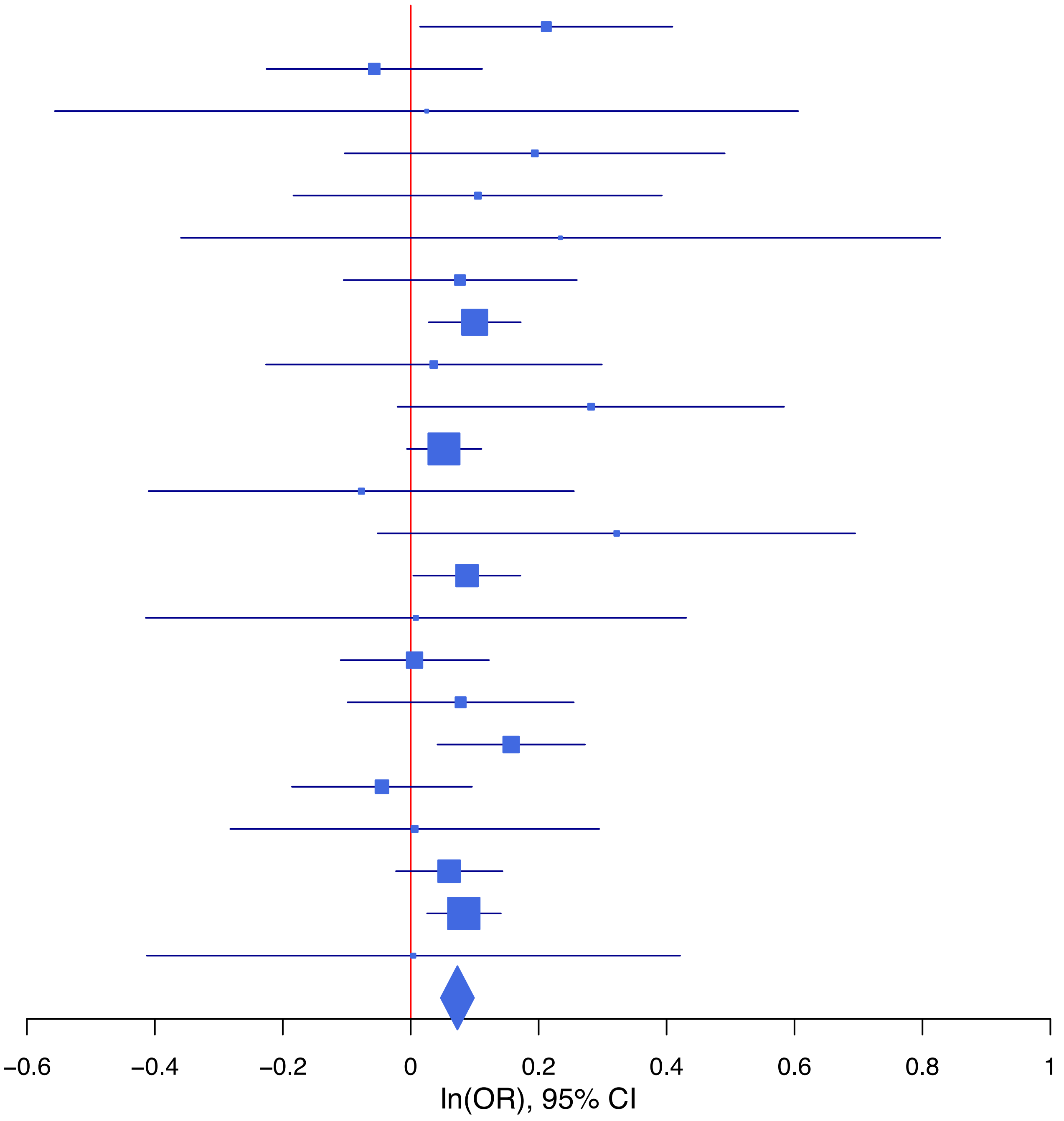

**rs11677638 G/A 2:212697656**

| Cohort | P | ln(OR) | SE |
|---|---|---|---|
| ALSPC_EUR | 0.2872 | −0.064 | 0.06 |
| BEPS7_EUR | 0.2876 | 0.221 | 0.208 |
| BHRCM_EUR | 0.1811 | −0.244 | 0.183 |
| BOR17_EUR | 0.5403 | −0.058 | 0.094 |
| BOR2C_EUR | 0.9008 | −0.012 | 0.097 |
| BOR2E_EUR | 0.6199 | 0.097 | 0.195 |
| COGA1_EUR | 0.1788 | −0.086 | 0.064 |
| CUINT_EUR | 0.061 | −0.148 | 0.079 |
| ESTB2_EUR | 0.0082 | −0.069 | 0.026 |
| FINNG_EUR | 0.9317 | 0.002 | 0.026 |
| GEDIS_EUR | 0.2858 | −0.156 | 0.146 |
| GISS1_EUR | 0.5876 | −0.045 | 0.083 |
| GISS2_EUR | 0.1884 | −0.13 | 0.099 |
| JANS3_EUR | 0.4297 | −0.088 | 0.112 |
| JANS4_EUR | 0.8082 | 0.033 | 0.135 |
| PGCBD_EUR | 0.6045 | −0.015 | 0.028 |
| PGCED_EUR | 0.3601 | −0.113 | 0.124 |
| PGCMD_EUR | 0.6738 | −0.016 | 0.038 |
| PGCPT_EUR | 0.0121 | 0.155 | 0.062 |
| PGCSZ_EUR | 0.0659 | −0.074 | 0.04 |
| PRFCT_EUR | 0.0148 | −0.126 | 0.052 |
| PSYCR_EUR | 0.2327 | 0.123 | 0.103 |
| QIMRB_EUR | 2e−04 | −0.123 | 0.033 |
| UKBJC_EUR | 0.0073 | −0.057 | 0.021 |
| UTAH2_EUR | 0.0022 | −0.064 | 0.021 |
| **meta** | **6.83e−09** | **−0.051** | **0.009** |

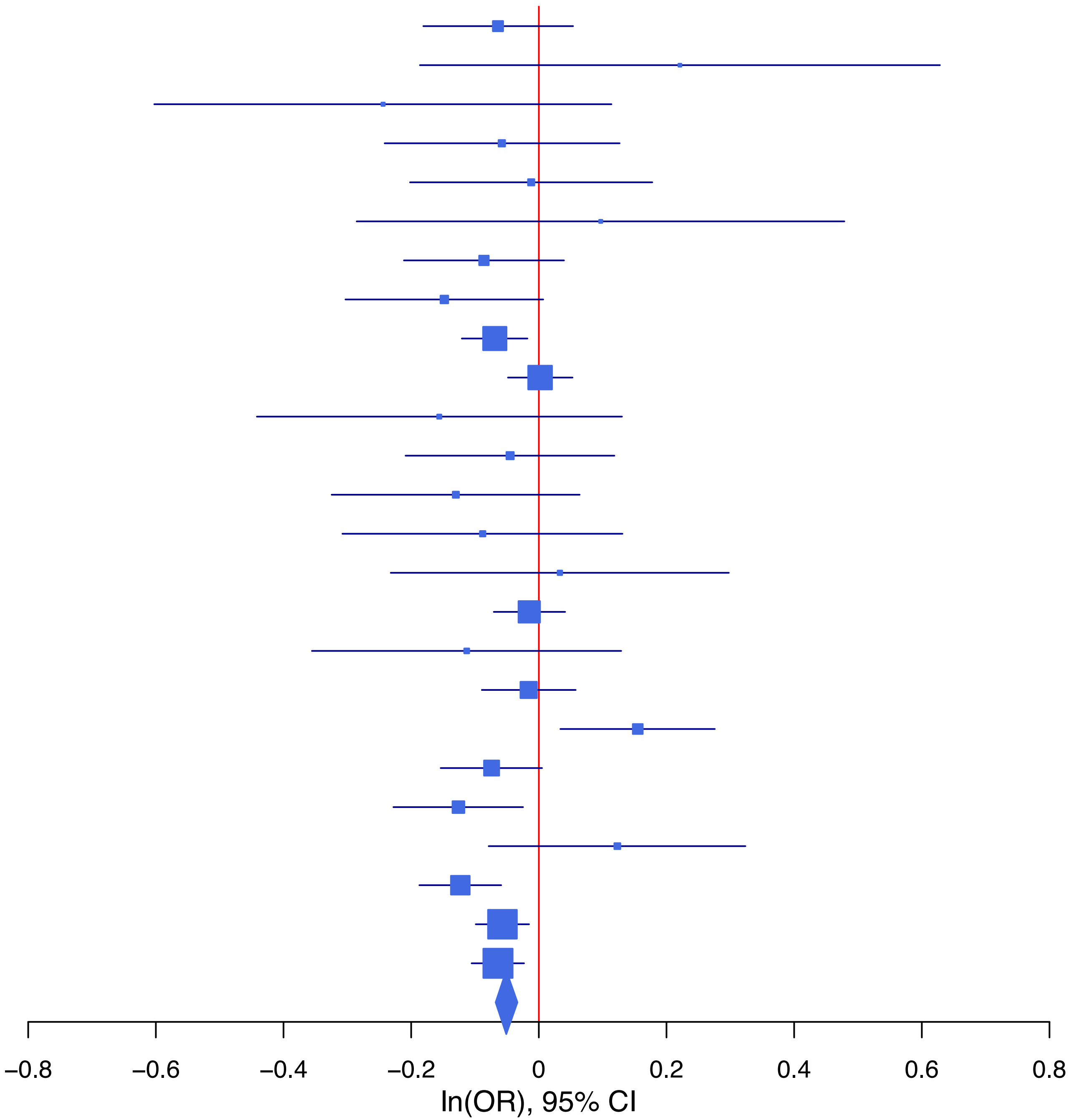

**rs12966785 A/G 18:22858941**

| Cohort | P | ln(OR) | SE |
|---|---|---|---|
| ALSPC_EUR | 0.8828 | −0.008 | 0.055 |
| BEPS7_EUR | 0.7455 | −0.059 | 0.183 |
| BHRCM_EUR | 0.3235 | 0.165 | 0.166 |
| BOR17_EUR | 0.5776 | −0.049 | 0.088 |
| BOR2C_EUR | 0.0777 | 0.157 | 0.089 |
| BOR2E_EUR | 0.7917 | −0.048 | 0.18 |
| COGA1_EUR | 0.5021 | −0.042 | 0.062 |
| ESTB2_EUR | 0.0845 | 0.041 | 0.023 |
| FINNG_EUR | 0.0582 | 0.046 | 0.024 |
| GEDIS_EUR | 0.9663 | 0.006 | 0.138 |
| GISS1_EUR | 0.6874 | −0.032 | 0.08 |
| GISS2_EUR | 0.1855 | −0.127 | 0.096 |
| IPSYC_EUR | 0.1381 | 0.028 | 0.019 |
| JANS3_EUR | 0.0618 | 0.193 | 0.104 |
| JANS4_EUR | 0.1685 | 0.165 | 0.12 |
| PGCBD_EUR | 0.4639 | 0.019 | 0.027 |
| PGCED_EUR | 0.1713 | 0.16 | 0.117 |
| PGCMD_EUR | 0.1829 | 0.047 | 0.035 |
| PGCPT_EUR | 0.8242 | 0.013 | 0.059 |
| PGCSZ_EUR | 0.2533 | 0.044 | 0.038 |
| PRFCT_EUR | 0.5933 | 0.024 | 0.045 |
| PSYCR_EUR | 0.7604 | 0.027 | 0.089 |
| QIMRB_EUR | 0.6809 | 0.012 | 0.029 |
| UKBJC_EUR | 1.45e−06 | 0.096 | 0.02 |
| UTAH2_EUR | 0.018 | 0.046 | 0.019 |
| YPENN_EUR | 0.9818 | −0.003 | 0.13 |
| **meta** | **2.51e−08** | **0.041** | **0.007** |

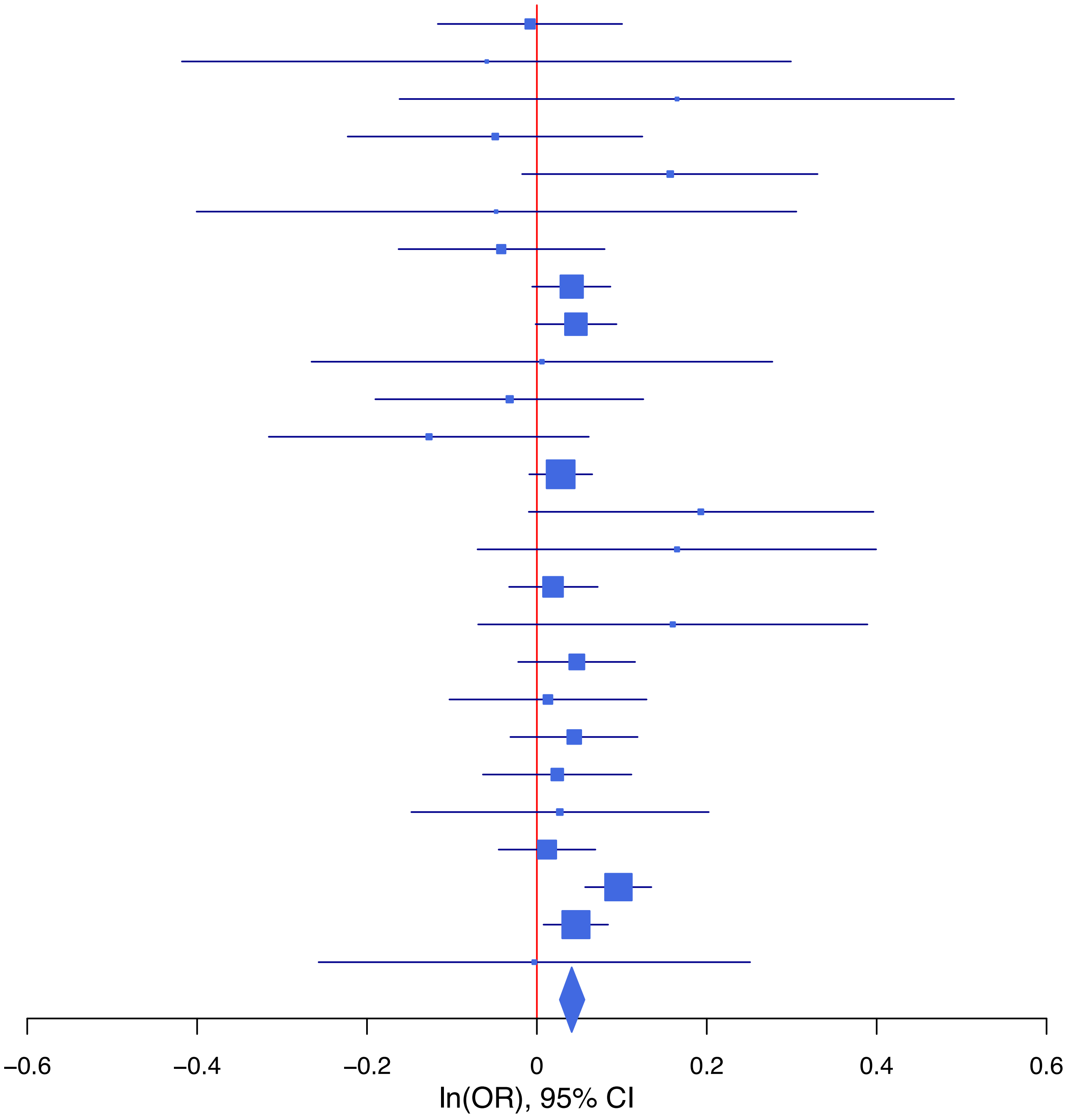

**rs13171784 A/C 5:153410363**

| Cohort | P | ln(OR) | SE |
|---|---|---|---|
| ADHEA_EUR | 0.0764 | 0.18 | 0.102 |
| ALSPC_EUR | 0.2743 | 0.089 | 0.081 |
| BEPS7_EUR | 0.2766 | 0.302 | 0.277 |
| BHRCM_EUR | 0.1086 | 0.33 | 0.206 |
| BOR17_EUR | 0.2464 | 0.155 | 0.134 |
| BOR2C_EUR | 0.0926 | 0.223 | 0.132 |
| BOR2E_EUR | 0.2614 | 0.297 | 0.264 |
| COGA1_EUR | 0.1464 | 0.134 | 0.092 |
| CUINT_EUR | 0.7665 | −0.034 | 0.116 |
| FINNG_EUR | 0.005 | 0.091 | 0.032 |
| GEDIS_EUR | 0.833 | −0.042 | 0.2 |
| GISS1_EUR | 0.3906 | −0.105 | 0.123 |
| GISS2_EUR | 0.9842 | 0.003 | 0.149 |
| IPSYC_EUR | 0.0058 | 0.077 | 0.028 |
| JANS3_EUR | 0.4694 | −0.116 | 0.161 |
| JANS4_EUR | 0.2138 | 0.217 | 0.175 |
| MIREC_EUR | 0.1941 | 0.274 | 0.211 |
| MVPXQ_EUR | 0.0033 | 0.058 | 0.02 |
| PGCBD_EUR | 0.7355 | 0.014 | 0.04 |
| PGCED_EUR | 0.3367 | 0.181 | 0.188 |
| PGCMD_EUR | 0.2659 | 0.063 | 0.057 |
| PGCPT_EUR | 0.1402 | 0.126 | 0.085 |
| PGCSZ_EUR | 0.7374 | 0.02 | 0.058 |
| PRFCT_EUR | 0.602 | 0.035 | 0.067 |
| PSYCR_EUR | 0.8822 | 0.021 | 0.142 |
| QIMRB_EUR | 0.526 | 0.026 | 0.041 |
| UKBJC_EUR | 0.2162 | 0.037 | 0.03 |
| UTAH2_EUR | 0.3356 | 0.028 | 0.029 |
| YPENN_EUR | 0.1408 | 0.261 | 0.178 |
| **meta** | **9.66e−09** | **0.057** | **0.01** |

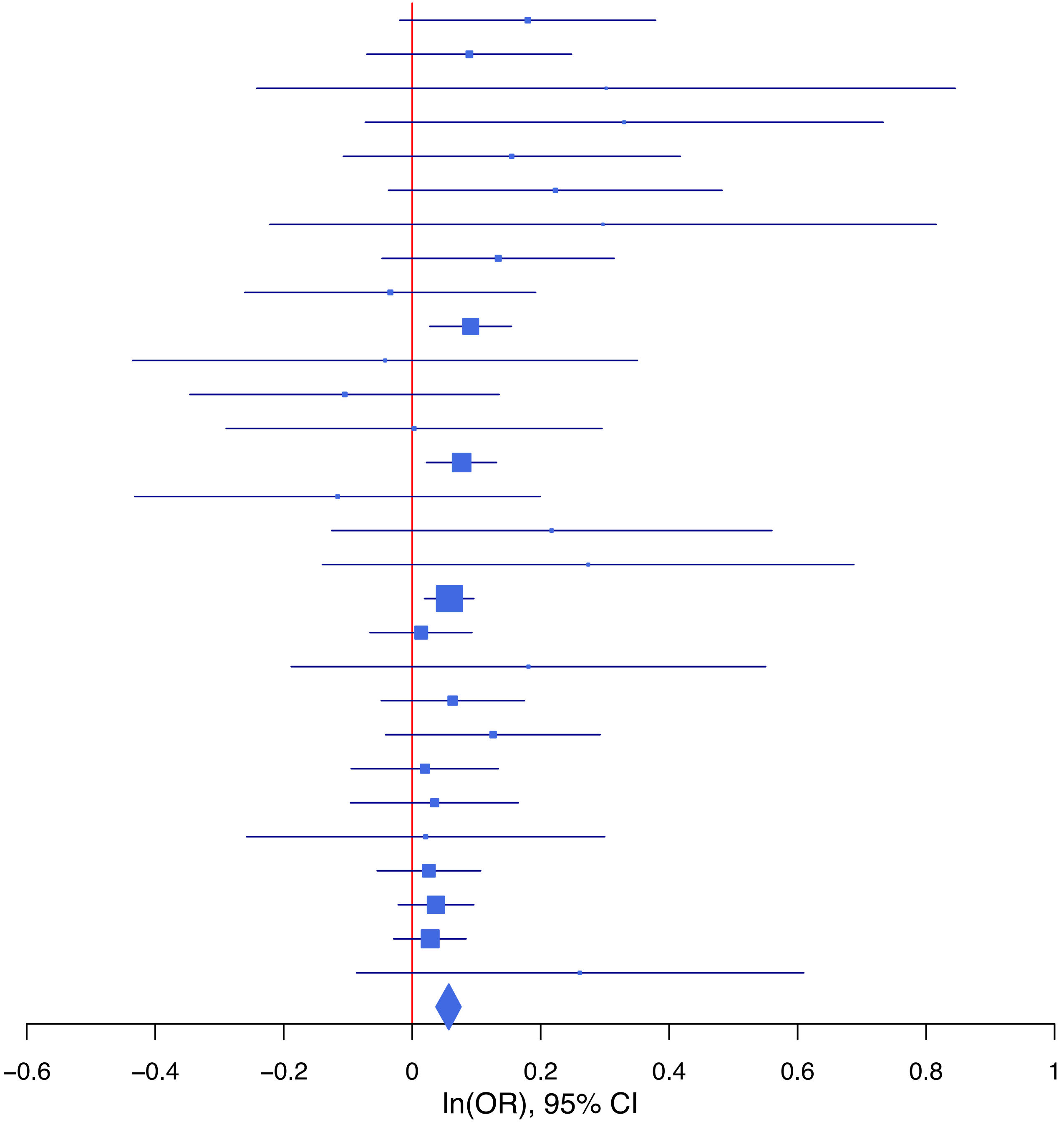

**rs13303 T/C 3:52558008**

| Cohort | P | ln(OR) | SE |
|---|---|---|---|
| ADHEA_EUR | 0.5936 | 0.037 | 0.069 |
| ALSPC_EUR | 0.3565 | 0.05 | 0.054 |
| BEPS7_EUR | 0.9011 | −0.022 | 0.174 |
| BHRCM_EUR | 0.1882 | 0.21 | 0.16 |
| BOR17_EUR | 0.1867 | 0.112 | 0.085 |
| BOR2C_EUR | 0.0811 | 0.149 | 0.085 |
| BOR2E_EUR | 0.7174 | −0.069 | 0.19 |
| COGA1_EUR | 0.017 | 0.143 | 0.06 |
| CUINT_EUR | 0.6975 | 0.029 | 0.074 |
| ESTB2_EUR | 0.0027 | 0.068 | 0.023 |
| FINNG_EUR | 0.5285 | 0.015 | 0.023 |
| GEDIS_EUR | 0.4708 | 0.095 | 0.132 |
| GISS2_EUR | 0.6999 | −0.036 | 0.093 |
| IPSYC_EUR | 0.347 | 0.018 | 0.019 |
| JANS3_EUR | 0.5878 | 0.056 | 0.102 |
| JANS4_EUR | 0.6711 | 0.05 | 0.117 |
| MIREC_EUR | 0.7553 | −0.046 | 0.149 |
| MVPXQ_EUR | 0.0039 | 0.053 | 0.019 |
| PGCBD_EUR | 0.0091 | 0.067 | 0.026 |
| PGCED_EUR | 0.581 | 0.064 | 0.116 |
| PGCMD_EUR | 0.6313 | −0.017 | 0.035 |
| PGCPT_EUR | 0.6222 | 0.028 | 0.057 |
| PGCSZ_EUR | 0.002 | 0.116 | 0.037 |
| PRFCT_EUR | 0.524 | 0.028 | 0.044 |
| PSYCR_EUR | 0.0307 | 0.189 | 0.087 |
| QIMRB_EUR | 0.0323 | 0.058 | 0.027 |
| UKBJC_EUR | 0.8357 | 0.004 | 0.02 |
| YPENN_EUR | 0.4728 | 0.093 | 0.129 |
| **meta** | **8.52e−09** | **0.041** | **0.007** |

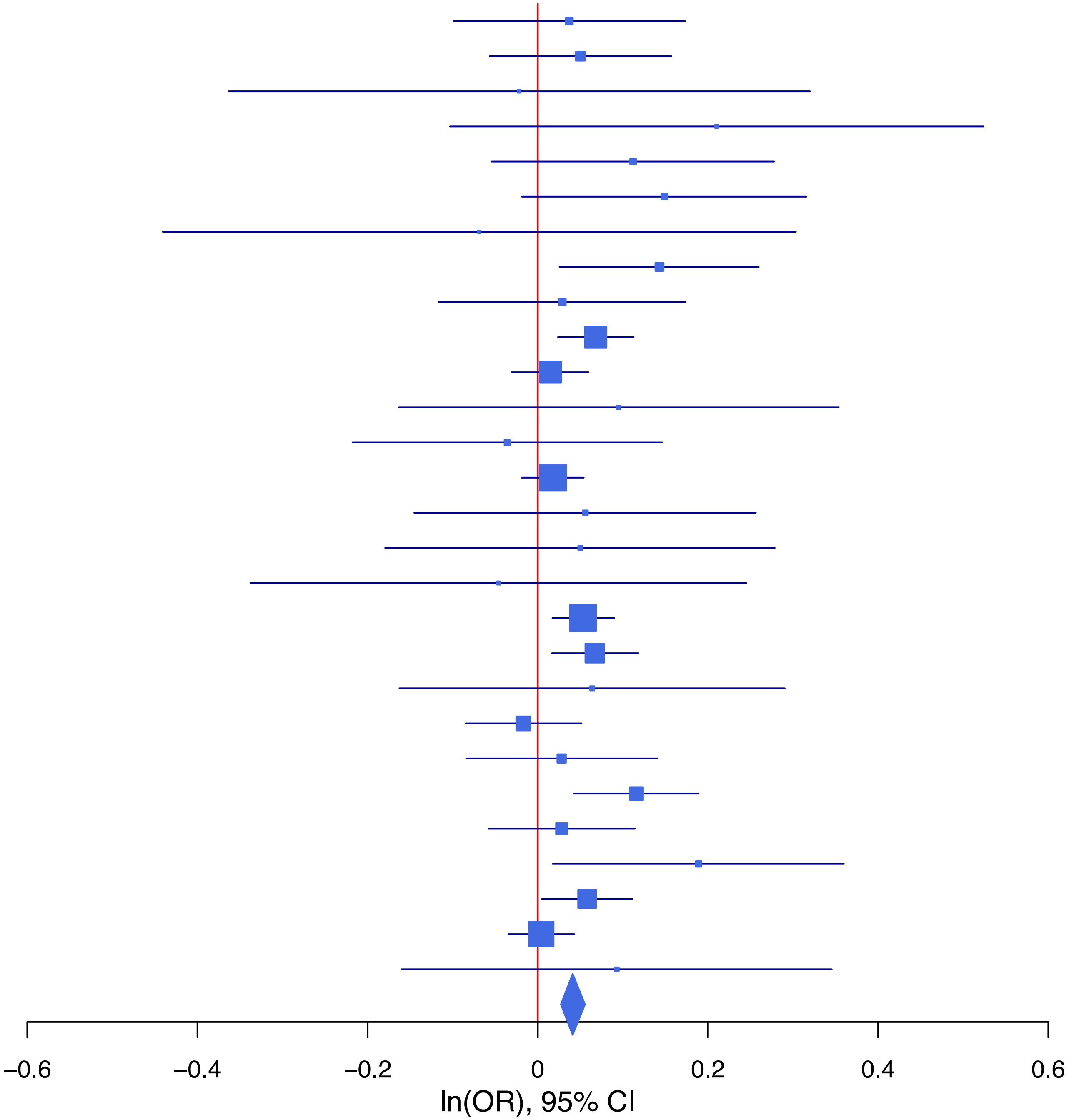

**rs13409451 G/A 2:144257639**

| Cohort | P | ln(OR) | SE |
|---|---|---|---|
| ALSPC_EUR | 0.5179 | 0.035 | 0.055 |
| BEPS7_EUR | 0.5857 | 0.102 | 0.186 |
| BHRCM_EUR | 0.799 | −0.042 | 0.164 |
| BOR17_EUR | 0.2649 | −0.098 | 0.088 |
| BOR2C_EUR | 0.3117 | −0.087 | 0.086 |
| BOR2E_EUR | 0.7528 | −0.058 | 0.184 |
| COGA1_EUR | 0.5976 | −0.032 | 0.061 |
| CUINT_EUR | 0.7733 | 0.022 | 0.076 |
| ESTB2_EUR | 0.0757 | −0.043 | 0.024 |
| FINNG_EUR | 0.0191 | −0.057 | 0.024 |
| GEDIS_EUR | 0.0027 | −0.421 | 0.141 |
| GISS1_EUR | 0.0631 | −0.157 | 0.085 |
| GISS2_EUR | 0.9151 | 0.011 | 0.1 |
| IPSYC_EUR | 0.013 | −0.047 | 0.019 |
| JANS3_EUR | 0.0638 | 0.198 | 0.107 |
| JANS4_EUR | 0.0708 | 0.223 | 0.123 |
| MVPXQ_EUR | 5.18e−06 | −0.061 | 0.013 |
| PGCED_EUR | 0.7979 | −0.03 | 0.116 |
| PGCMD_EUR | 0.17 | −0.049 | 0.035 |
| PGCPT_EUR | 0.5612 | 0.034 | 0.059 |
| PRFCT_EUR | 0.0167 | −0.109 | 0.045 |
| PSYCR_EUR | 0.809 | −0.022 | 0.092 |
| QIMRB_EUR | 0.0825 | −0.049 | 0.028 |
| UKBJC_EUR | 0.3595 | −0.018 | 0.02 |
| UTAH2_EUR | 0.0363 | −0.041 | 0.02 |
| YPENN_EUR | 0.8171 | −0.031 | 0.132 |
| **meta** | **3.47e−11** | **−0.045** | **0.007** |

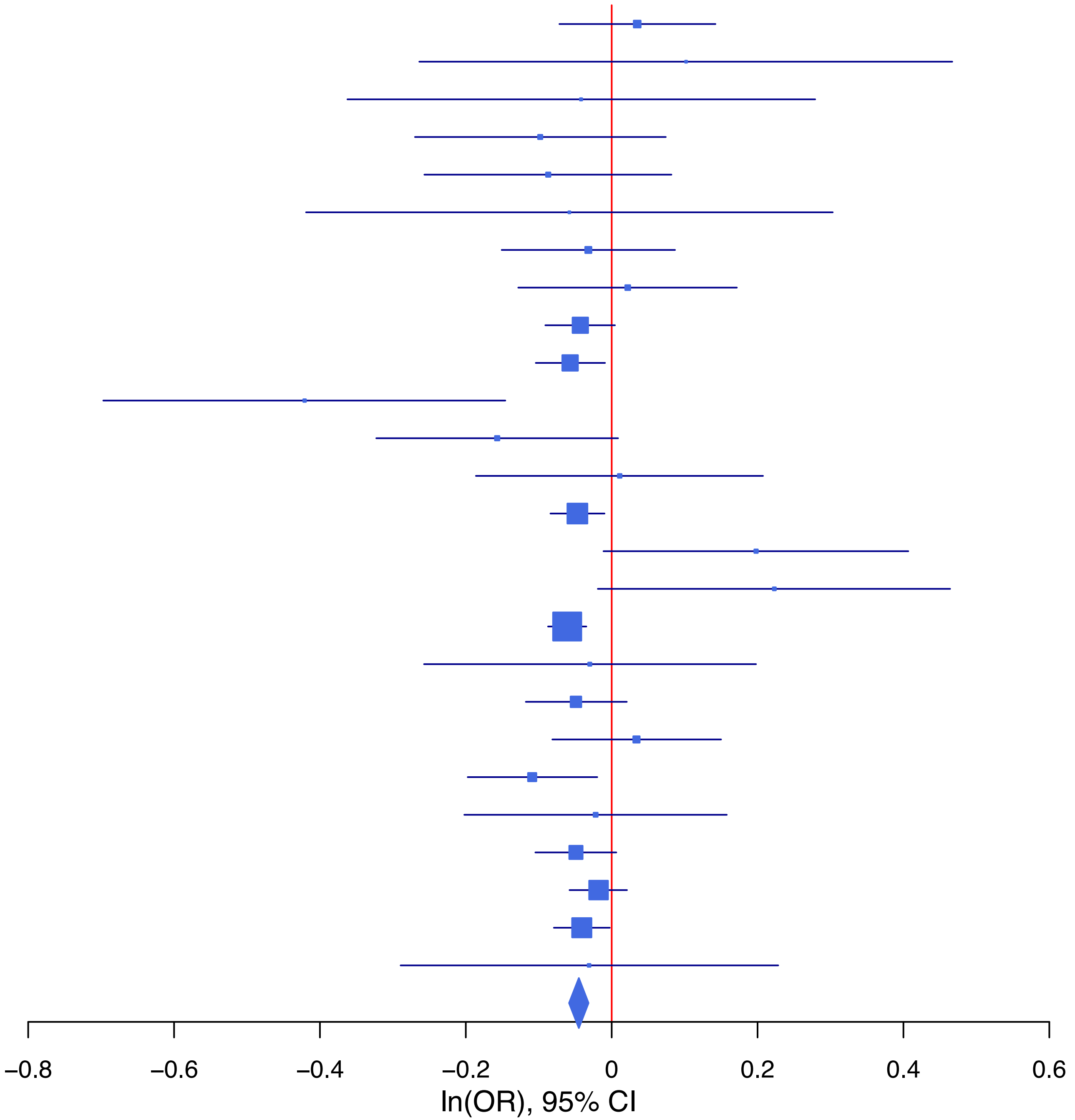

**rs1452787 G/A 18:53207207**

| Cohort | P | ln(OR) | SE |
|---|---|---|---|
| ADHEA_EUR | 0.3651 | 0.066 | 0.073 |
| ALSPC_EUR | 0.6703 | −0.026 | 0.061 |
| BEPS7_EUR | 0.3116 | 0.194 | 0.192 |
| BHRCM_EUR | 0.5528 | −0.103 | 0.173 |
| BOR17_EUR | 0.9278 | −0.008 | 0.094 |
| BOR2C_EUR | 0.1103 | −0.153 | 0.096 |
| BOR2E_EUR | 0.4452 | −0.152 | 0.199 |
| COGA1_EUR | 0.4333 | 0.05 | 0.064 |
| CUINT_EUR | 0.5911 | 0.044 | 0.082 |
| ESTB2_EUR | 0.1561 | 0.035 | 0.024 |
| FINNG_EUR | 0.1266 | 0.038 | 0.025 |
| GEDIS_EUR | 0.5074 | −0.097 | 0.146 |
| GISS1_EUR | 0.0353 | 0.174 | 0.083 |
| GISS2_EUR | 0.5508 | −0.057 | 0.096 |
| IPSYC_EUR | 0.0896 | 0.035 | 0.021 |
| JANS3_EUR | 0.0944 | −0.198 | 0.118 |
| JANS4_EUR | 0.6287 | −0.063 | 0.13 |
| MIREC_EUR | 0.5564 | −0.099 | 0.168 |
| MVPXQ_EUR | 1e−04 | 0.052 | 0.014 |
| PGCBD_EUR | 0.7559 | −0.009 | 0.029 |
| PGCED_EUR | 0.5108 | −0.083 | 0.127 |
| PGCMD_EUR | 0.8691 | −0.006 | 0.038 |
| PGCPT_EUR | 0.984 | −0.001 | 0.063 |
| PGCSZ_EUR | 0.0079 | 0.107 | 0.04 |
| PRFCT_EUR | 0.1278 | 0.074 | 0.048 |
| PSYCR_EUR | 0.5973 | 0.05 | 0.094 |
| QIMRB_EUR | 0.3472 | 0.028 | 0.03 |
| UKBJC_EUR | 0.0034 | 0.063 | 0.022 |
| UTAH2_EUR | 0.0198 | 0.048 | 0.021 |
| YPENN_EUR | 0.4574 | 0.099 | 0.133 |
| **meta** | **8.81e−09** | **0.039** | **0.007** |

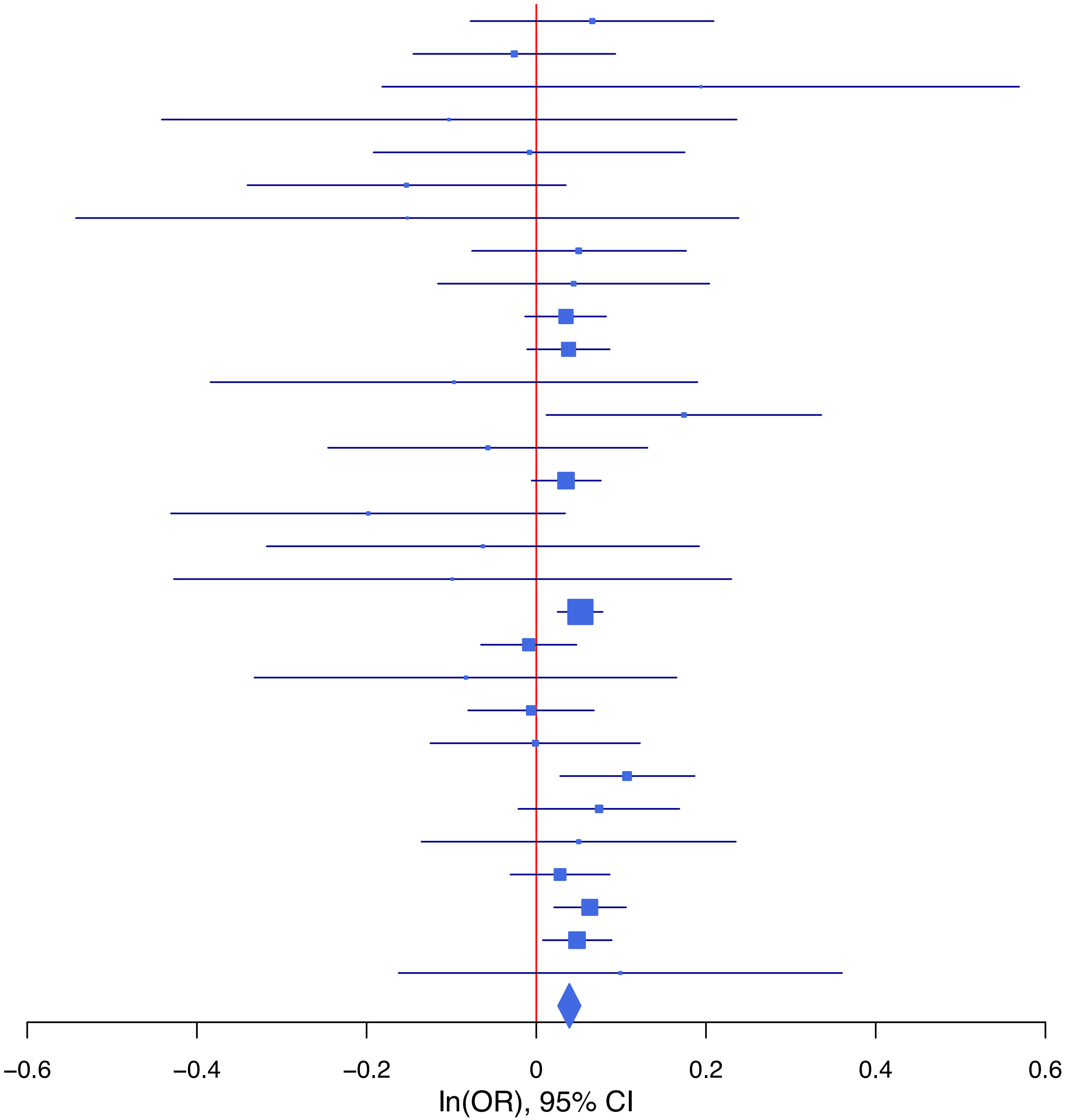

**rs1486900 T/G 15:47642886**

| Cohort | P | ln(OR) | SE |
|---|---|---|---|
| ADHEA_EUR | 0.9912 | −0.001 | 0.088 |
| BEPS7_EUR | 0.6461 | 0.112 | 0.245 |
| BHRCM_EUR | 0.7706 | −0.058 | 0.199 |
| BOR17_EUR | 0.3235 | 0.103 | 0.104 |
| BOR2C_EUR | 0.6565 | −0.049 | 0.11 |
| BOR2E_EUR | 0.6715 | 0.098 | 0.231 |
| COGA1_EUR | 0.6823 | 0.031 | 0.075 |
| CUINT_EUR | 0.7635 | −0.029 | 0.096 |
| ESTB2_EUR | 0.0486 | 0.063 | 0.032 |
| FINNG_EUR | 0.7385 | −0.011 | 0.033 |
| GEDIS_EUR | 0.2713 | −0.195 | 0.177 |
| GISS1_EUR | 0.5388 | 0.069 | 0.112 |
| GISS2_EUR | 0.0338 | 0.287 | 0.135 |
| IPSYC_EUR | 0.0024 | 0.07 | 0.023 |
| JANS3_EUR | 0.9739 | 0.004 | 0.136 |
| JANS4_EUR | 0.9512 | 0.01 | 0.161 |
| MIREC_EUR | 0.1238 | 0.281 | 0.182 |
| MVPXQ_EUR | 0.0151 | 0.039 | 0.016 |
| PGCBD_EUR | 0.0068 | 0.09 | 0.033 |
| PGCED_EUR | 0.2902 | 0.153 | 0.145 |
| PGCMD_EUR | 0.0021 | 0.134 | 0.044 |
| PGCPT_EUR | 0.1079 | −0.123 | 0.076 |
| PGCSZ_EUR | 0.007 | 0.127 | 0.047 |
| PRFCT_EUR | 0.2831 | −0.062 | 0.057 |
| PSYCR_EUR | 0.7897 | −0.031 | 0.117 |
| QIMRB_EUR | 1e−04 | 0.14 | 0.036 |
| UKBJC_EUR | 0.0015 | 0.081 | 0.026 |
| UTAH2_EUR | 0.0039 | 0.07 | 0.024 |
| **meta** | **3.58e−13** | **0.059** | **0.008** |

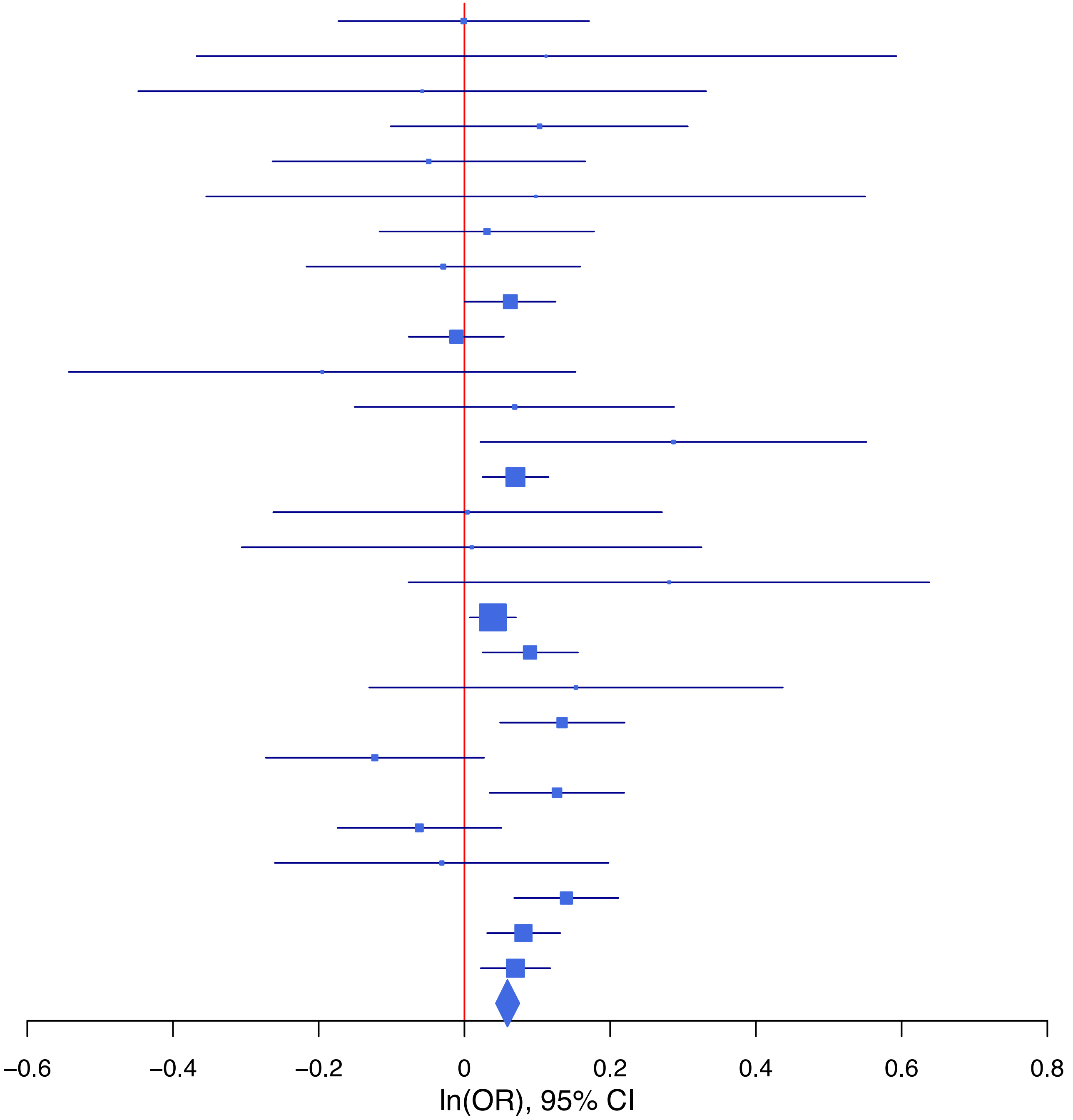

**rs1604350 G/C 1:73991651**

| Cohort | P | ln(OR) | SE |
|---|---|---|---|
| ADHEA_EUR | 0.8072 | 0.017 | 0.069 |
| ALSPC_EUR | 0.2468 | 0.064 | 0.055 |
| BEPS7_EUR | 0.4875 | −0.127 | 0.183 |
| BHRCM_EUR | 0.0239 | 0.355 | 0.157 |
| BOR17_EUR | 0.4482 | −0.065 | 0.085 |
| BOR2C_EUR | 0.355 | −0.079 | 0.086 |
| BOR2E_EUR | 0.9097 | 0.02 | 0.172 |
| COGA1_EUR | 0.492 | 0.041 | 0.06 |
| CUINT_EUR | 0.5018 | 0.05 | 0.074 |
| ESTB2_EUR | 0.9659 | 0.001 | 0.024 |
| FINNG_EUR | 0.4727 | 0.017 | 0.024 |
| GEDIS_EUR | 0.0183 | 0.31 | 0.132 |
| GISS1_EUR | 0.9363 | −0.006 | 0.08 |
| GISS2_EUR | 0.4527 | 0.07 | 0.093 |
| IPSYC_EUR | 0.0979 | 0.031 | 0.019 |
| JANS3_EUR | 0.1228 | −0.168 | 0.109 |
| JANS4_EUR | 0.1589 | −0.176 | 0.125 |
| MVPXQ_EUR | 1.09e−05 | 0.057 | 0.013 |
| PGCBD_EUR | 0.7202 | 0.009 | 0.026 |
| PGCED_EUR | 0.3539 | 0.107 | 0.115 |
| PGCMD_EUR | 0.1227 | 0.054 | 0.035 |
| PGCPT_EUR | 0.7587 | 0.018 | 0.058 |
| PGCSZ_EUR | 0.2662 | 0.042 | 0.038 |
| PRFCT_EUR | 0.1675 | 0.061 | 0.044 |
| PSYCR_EUR | 0.2929 | −0.095 | 0.091 |
| QIMRB_EUR | 0.0485 | 0.055 | 0.028 |
| UKBJC_EUR | 0.0037 | 0.058 | 0.02 |
| UTAH2_EUR | 0.0295 | 0.043 | 0.02 |
| YPENN_EUR | 0.2375 | 0.153 | 0.13 |
| **meta** | **1.33e−09** | **0.039** | **0.006** |

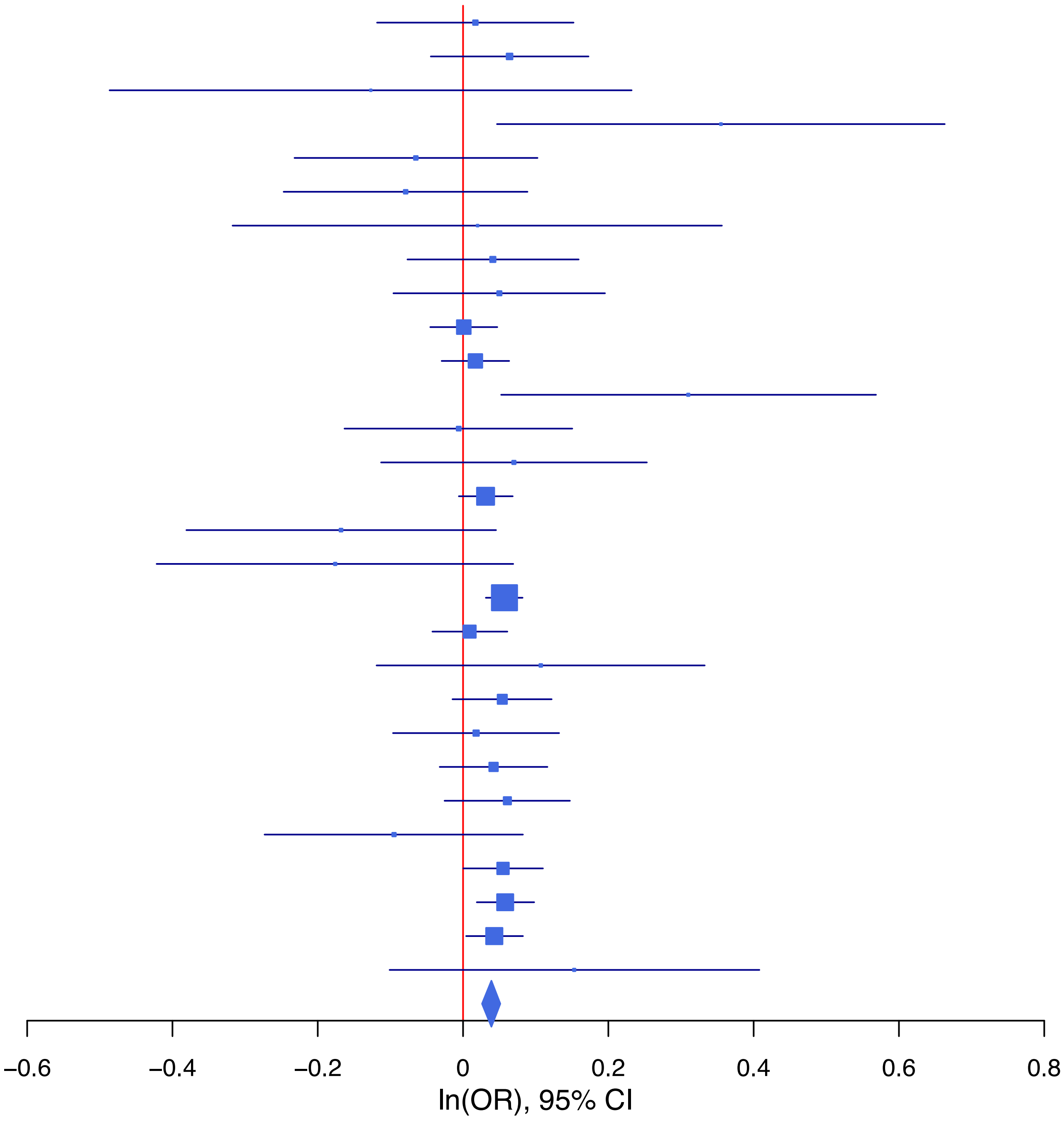

**rs17137753 G/A 7:115030592**

| Cohort | P | ln(OR) | SE |
|---|---|---|---|
| ALSPC_EUR | 0.4013 | 0.045 | 0.054 |
| BEPS7_EUR | 0.2575 | 0.194 | 0.171 |
| BHRCM_EUR | 0.2787 | 0.171 | 0.158 |
| BOR17_EUR | 0.8084 | −0.021 | 0.085 |
| BOR2C_EUR | 0.5652 | −0.049 | 0.085 |
| BOR2E_EUR | 0.655 | 0.079 | 0.176 |
| COGA1_EUR | 0.829 | −0.013 | 0.059 |
| CUINT_EUR | 0.0095 | 0.193 | 0.075 |
| ESTB2_EUR | 8e−04 | 0.075 | 0.022 |
| FINNG_EUR | 0.1565 | 0.033 | 0.023 |
| GEDIS_EUR | 0.0059 | 0.36 | 0.131 |
| GISS1_EUR | 0.6683 | 0.033 | 0.078 |
| GISS2_EUR | 0.9185 | −0.009 | 0.092 |
| IPSYC_EUR | 8e−04 | 0.062 | 0.018 |
| JANS3_EUR | 0.714 | −0.038 | 0.105 |
| JANS4_EUR | 0.1349 | 0.181 | 0.121 |
| PGCBD_EUR | 0.0071 | 0.069 | 0.026 |
| PGCED_EUR | 0.565 | −0.066 | 0.115 |
| PGCMD_EUR | 0.0486 | 0.067 | 0.034 |
| PGCPT_EUR | 0.4042 | 0.048 | 0.057 |
| PGCSZ_EUR | 0.4144 | 0.03 | 0.036 |
| PRFCT_EUR | 0.3629 | 0.039 | 0.043 |
| PSYCR_EUR | 0.1831 | −0.115 | 0.086 |
| QIMRB_EUR | 2e−04 | 0.102 | 0.027 |
| UKBJC_EUR | 0.0761 | 0.035 | 0.02 |
| UTAH2_EUR | 0.3102 | 0.019 | 0.019 |
| YPENN_EUR | 0.8597 | 0.022 | 0.127 |
| **meta** | **4.87e−12** | **0.05** | **0.007** |

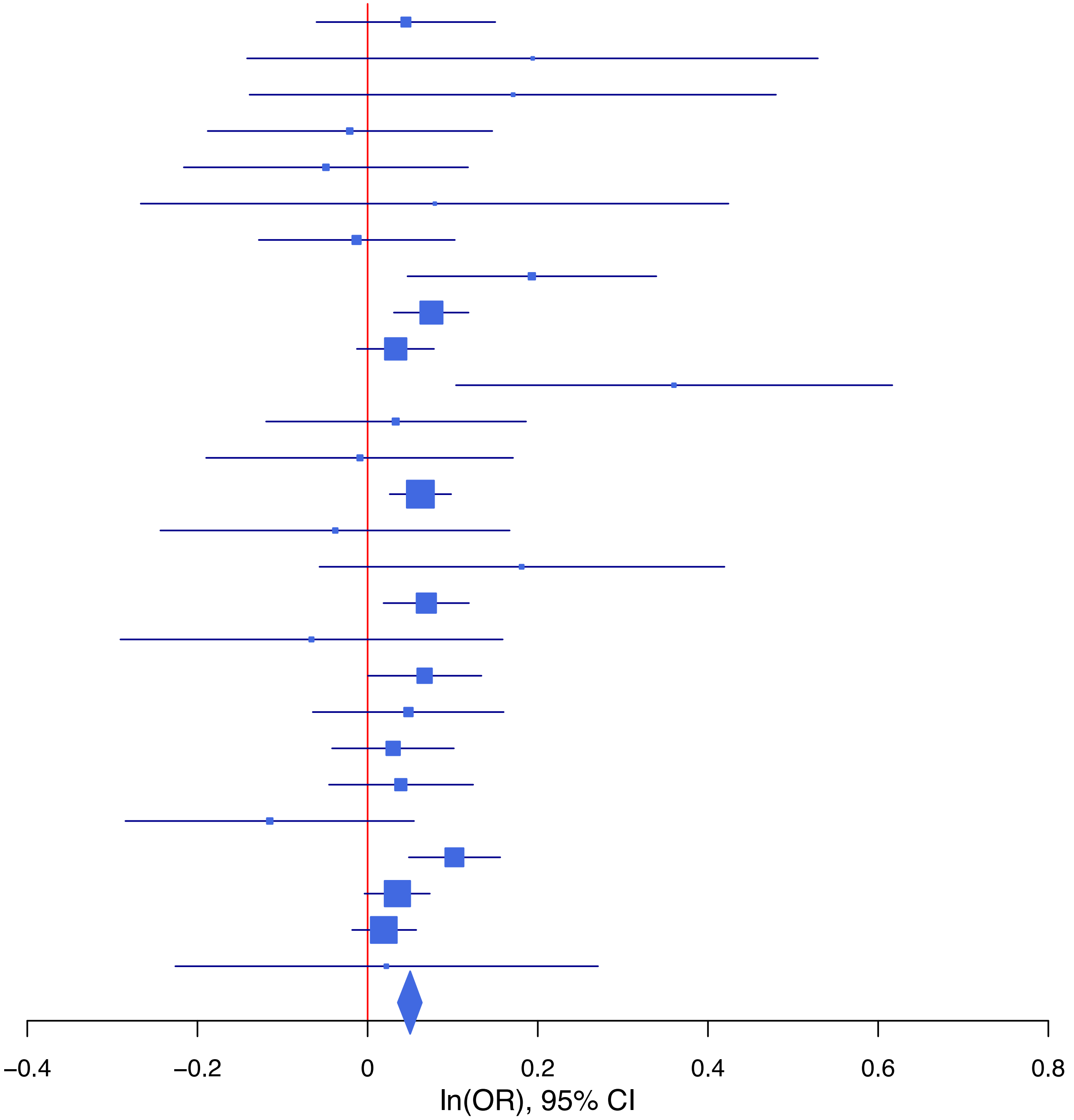

**rs185782836 A/G 14:64520663**

| Cohort | P | ln(OR) | SE |
|---|---|---|---|
| ALSPC_EUR | 0.1242 | 0.272 | 0.177 |
| BHRCM_EUR | 0.1301 | 0.751 | 0.496 |
| BOR17_EUR | 0.2941 | 0.309 | 0.294 |
| BOR2C_EUR | 0.6106 | 0.15 | 0.294 |
| CUINT_EUR | 0.8638 | 0.054 | 0.314 |
| ESTB2_EUR | 0.0162 | 0.124 | 0.052 |
| FINNG_EUR | 0.0281 | 0.235 | 0.107 |
| GISS2_EUR | 0.8451 | 0.051 | 0.259 |
| IPSYC_EUR | 2e−04 | 0.261 | 0.071 |
| MVPXQ_EUR | 0.0027 | 0.169 | 0.056 |
| PGCPT_EUR | 0.2954 | −0.287 | 0.274 |
| PGCSZ_EUR | 0.7751 | −0.053 | 0.187 |
| PRFCT_EUR | 0.6617 | 0.068 | 0.156 |
| PSYCR_EUR | 0.4005 | 0.245 | 0.292 |
| QIMRB_EUR | 0.4612 | 0.077 | 0.104 |
| UKBJC_EUR | 0.1218 | 0.114 | 0.073 |
| UTAH2_EUR | 0.2838 | 0.071 | 0.066 |
| **meta** | **7.67e−09** | **0.142** | **0.024** |

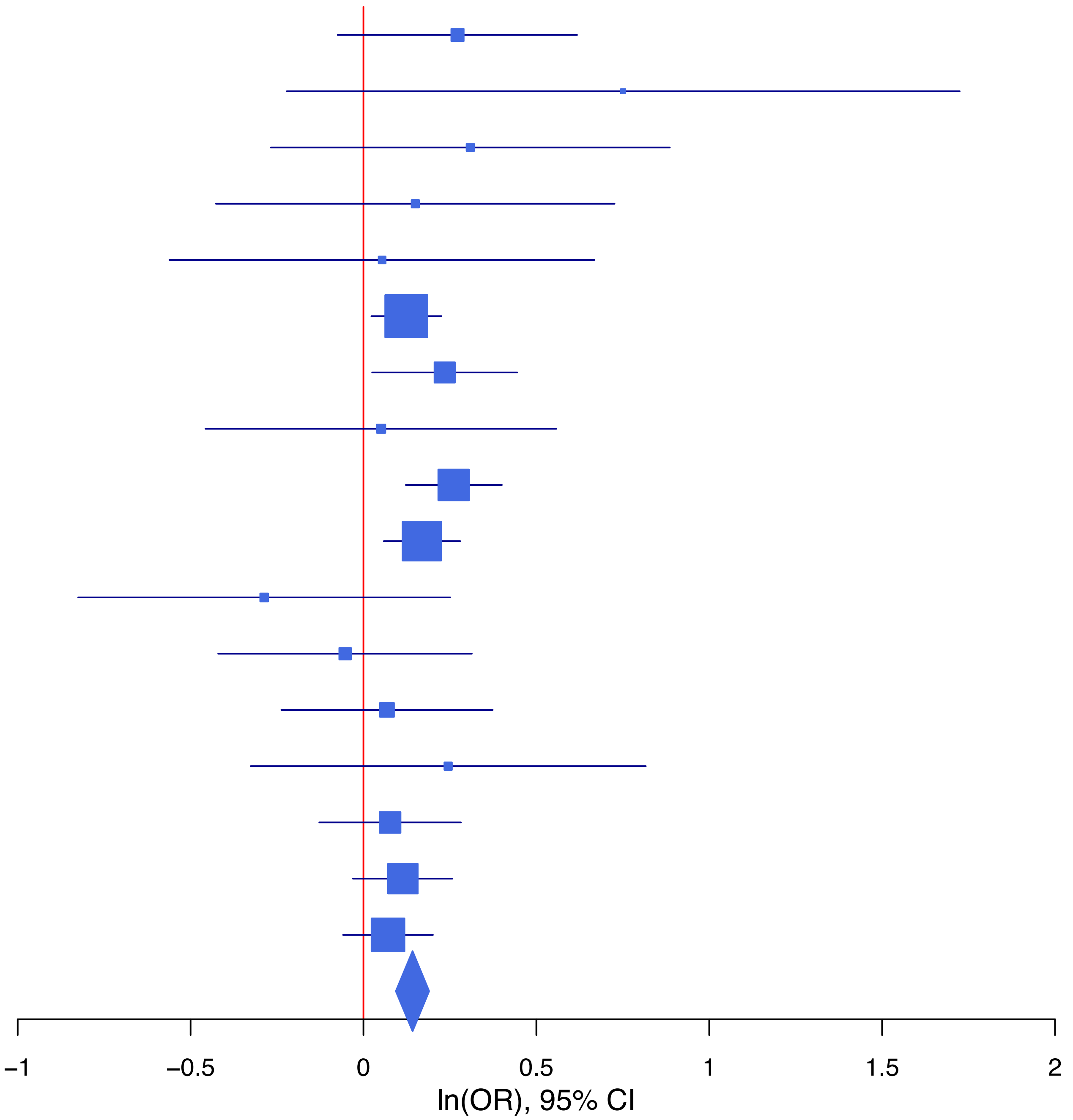

**rs1883987 C/T 22:37053469**

| Cohort | P | ln(OR) | SE |
|---|---|---|---|
| ALSPC_EUR | 0.7298 | 0.019 | 0.056 |
| BEPS7_EUR | 0.3171 | 0.215 | 0.215 |
| BOR17_EUR | 0.8451 | 0.019 | 0.097 |
| BOR2C_EUR | 0.5889 | −0.053 | 0.098 |
| BOR2E_EUR | 0.5946 | −0.113 | 0.211 |
| COGA1_EUR | 0.4449 | −0.047 | 0.062 |
| ESTB2_EUR | 0.1022 | −0.043 | 0.026 |
| FINNG_EUR | 0.5202 | 0.016 | 0.024 |
| GEDIS_EUR | 0.6507 | −0.061 | 0.135 |
| GISS1_EUR | 0.7353 | 0.029 | 0.085 |
| GISS2_EUR | 0.1038 | −0.197 | 0.121 |
| IPSYC_EUR | 0.1073 | −0.033 | 0.02 |
| JANS3_EUR | 0.3567 | −0.11 | 0.119 |
| JANS4_EUR | 0.6615 | 0.058 | 0.132 |
| MVPXQ_EUR | 0.0089 | −0.035 | 0.013 |
| PGCBD_EUR | 0.0014 | −0.092 | 0.029 |
| PGCMD_EUR | 0.3805 | −0.033 | 0.037 |
| PGCPT_EUR | 0.3099 | −0.064 | 0.063 |
| PGCSZ_EUR | 0.3522 | −0.038 | 0.041 |
| PRFCT_EUR | 0.0342 | −0.11 | 0.052 |
| PSYCR_EUR | 0.2351 | 0.125 | 0.106 |
| QIMRB_EUR | 0.1993 | −0.042 | 0.033 |
| UKBJC_EUR | 0.0026 | −0.062 | 0.021 |
| UTAH2_EUR | 0.0673 | −0.036 | 0.019 |
| **meta** | **2.49e−08** | **−0.038** | **0.007** |

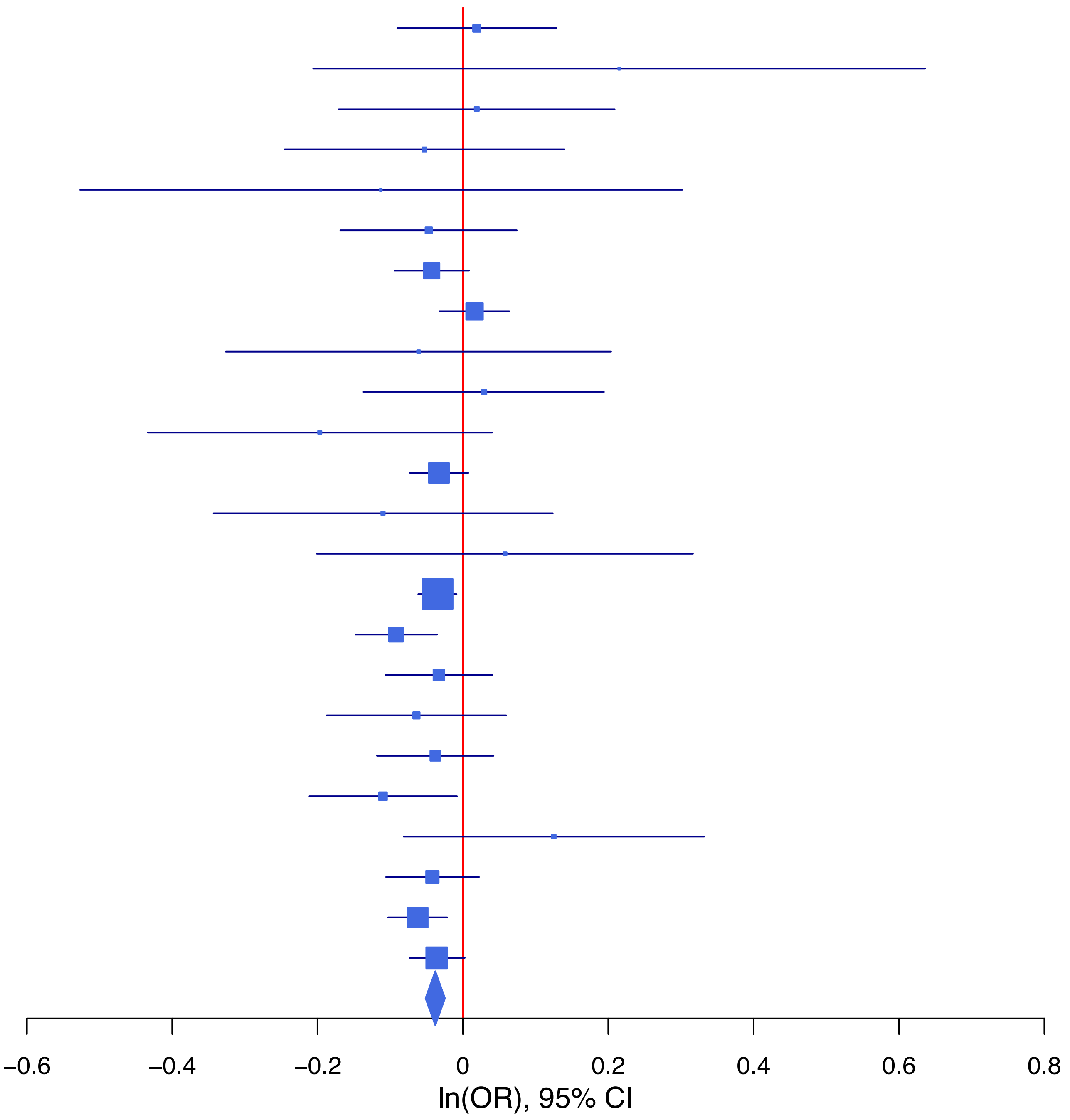

**rs1894401 G/A 15:91429042**

| Cohort | P | ln(OR) | SE |
|---|---|---|---|
| ALSPC_EUR | 0.218 | −0.068 | 0.055 |
| BEPS7_EUR | 0.0995 | −0.278 | 0.169 |
| BHRCM_EUR | 0.917 | 0.016 | 0.153 |
| BOR17_EUR | 0.2113 | −0.106 | 0.085 |
| BOR2C_EUR | 0.8583 | 0.015 | 0.086 |
| BOR2E_EUR | 0.187 | 0.232 | 0.176 |
| COGA1_EUR | 0.5051 | −0.04 | 0.059 |
| CUINT_EUR | 0.4018 | −0.061 | 0.073 |
| ESTB2_EUR | 0.8601 | 0.004 | 0.022 |
| FINNG_EUR | 0.1938 | −0.03 | 0.023 |
| GEDIS_EUR | 0.4488 | 0.098 | 0.129 |
| GISS1_EUR | 0.1021 | −0.127 | 0.078 |
| GISS2_EUR | 0.4938 | −0.063 | 0.092 |
| IPSYC_EUR | 0.0835 | −0.032 | 0.018 |
| JANS3_EUR | 0.3362 | 0.098 | 0.102 |
| JANS4_EUR | 0.2652 | −0.135 | 0.121 |
| MVPXQ_EUR | 0.0046 | −0.036 | 0.013 |
| PGCBD_EUR | 0.2614 | −0.03 | 0.026 |
| PGCMD_EUR | 0.0367 | −0.074 | 0.035 |
| PGCPT_EUR | 0.4035 | −0.048 | 0.058 |
| PGCSZ_EUR | 0.3317 | −0.037 | 0.038 |
| PRFCT_EUR | 0.3773 | −0.039 | 0.044 |
| PSYCR_EUR | 0.8819 | 0.013 | 0.087 |
| QIMRB_EUR | 0.0081 | −0.071 | 0.027 |
| UKBJC_EUR | 0.0018 | −0.061 | 0.02 |
| UTAH2_EUR | 0.1302 | −0.028 | 0.019 |
| YPENN_EUR | 0.7638 | −0.038 | 0.128 |
| **meta** | **2.75e−09** | **−0.037** | **0.006** |

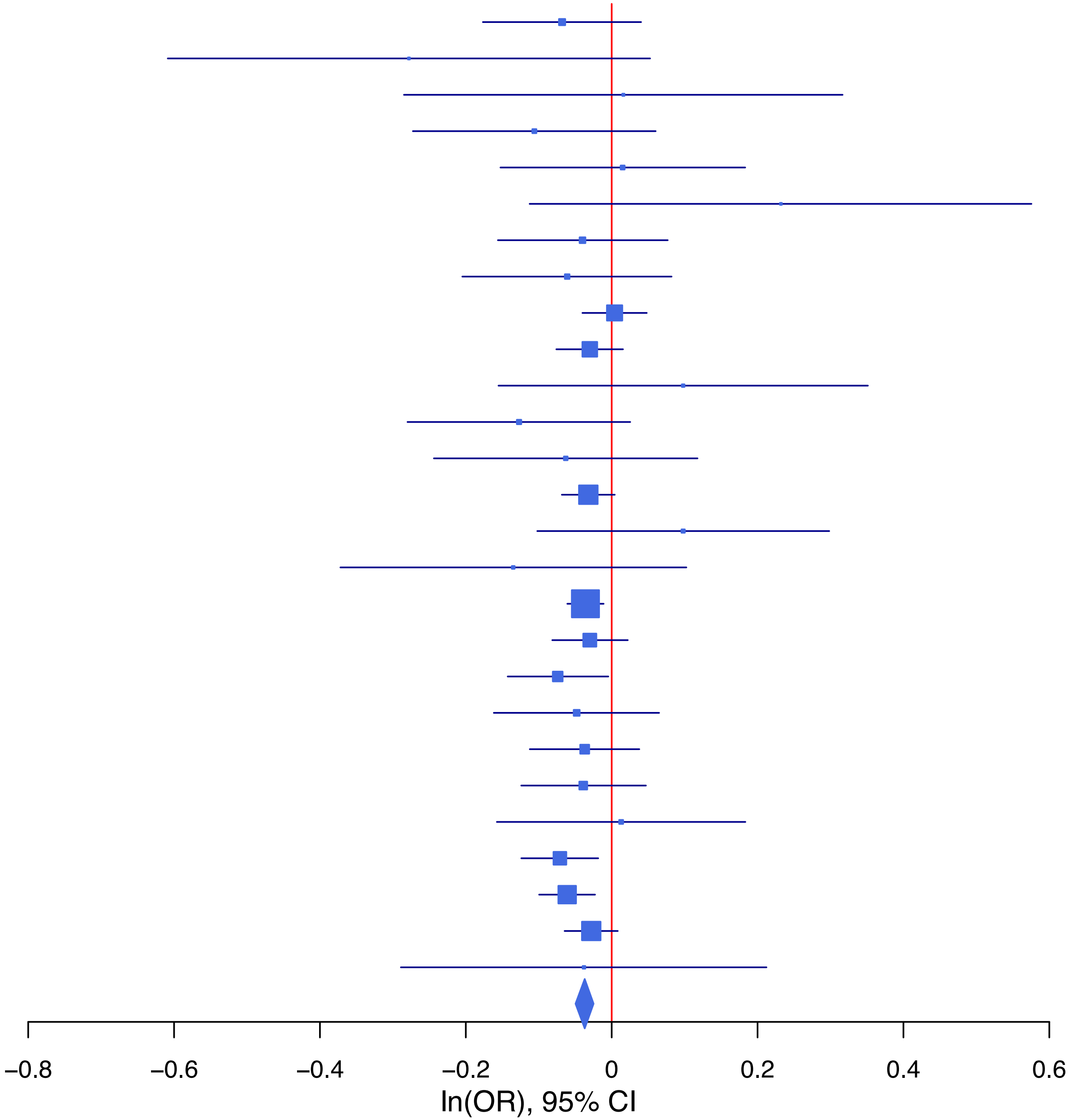

**rs2155281 A/G 11:112838338**

| Cohort | P | ln(OR) | SE |
|---|---|---|---|
| ADHEA_EUR | 0.4464 | 0.053 | 0.069 |
| ALSPC_EUR | 0.6289 | −0.027 | 0.055 |
| BEPS7_EUR | 0.5775 | 0.108 | 0.194 |
| BHRCM_EUR | 0.9287 | 0.015 | 0.17 |
| BOR17_EUR | 0.7363 | 0.029 | 0.087 |
| BOR2C_EUR | 0.0787 | 0.158 | 0.09 |
| BOR2E_EUR | 0.3841 | −0.177 | 0.204 |
| COGA1_EUR | 0.4002 | 0.05 | 0.059 |
| CUINT_EUR | 0.4539 | 0.056 | 0.075 |
| ESTB2_EUR | 0.0862 | 0.039 | 0.023 |
| FINNG_EUR | 0.1467 | 0.034 | 0.024 |
| GEDIS_EUR | 0.8247 | −0.03 | 0.136 |
| GISS1_EUR | 0.4595 | −0.058 | 0.078 |
| GISS2_EUR | 0.0712 | 0.174 | 0.096 |
| IPSYC_EUR | 0.1056 | 0.031 | 0.019 |
| JANS3_EUR | 0.0781 | 0.187 | 0.106 |
| JANS4_EUR | 0.4831 | 0.086 | 0.123 |
| MVPXQ_EUR | 1.72e−05 | 0.056 | 0.013 |
| PGCBD_EUR | 0.0797 | 0.046 | 0.026 |
| PGCED_EUR | 0.2986 | 0.12 | 0.115 |
| PGCMD_EUR | 0.5444 | 0.021 | 0.035 |
| PGCPT_EUR | 0.3805 | −0.052 | 0.059 |
| PGCSZ_EUR | 0.1023 | 0.061 | 0.037 |
| PRFCT_EUR | 0.5988 | −0.023 | 0.044 |
| PSYCR_EUR | 0.387 | 0.078 | 0.09 |
| QIMRB_EUR | 0.1797 | 0.038 | 0.029 |
| UKBJC_EUR | 0.0513 | 0.039 | 0.02 |
| UTAH2_EUR | 0.9756 | 0.001 | 0.019 |
| YPENN_EUR | 0.8922 | −0.017 | 0.128 |
| **meta** | **1.23e−08** | **0.036** | **0.006** |

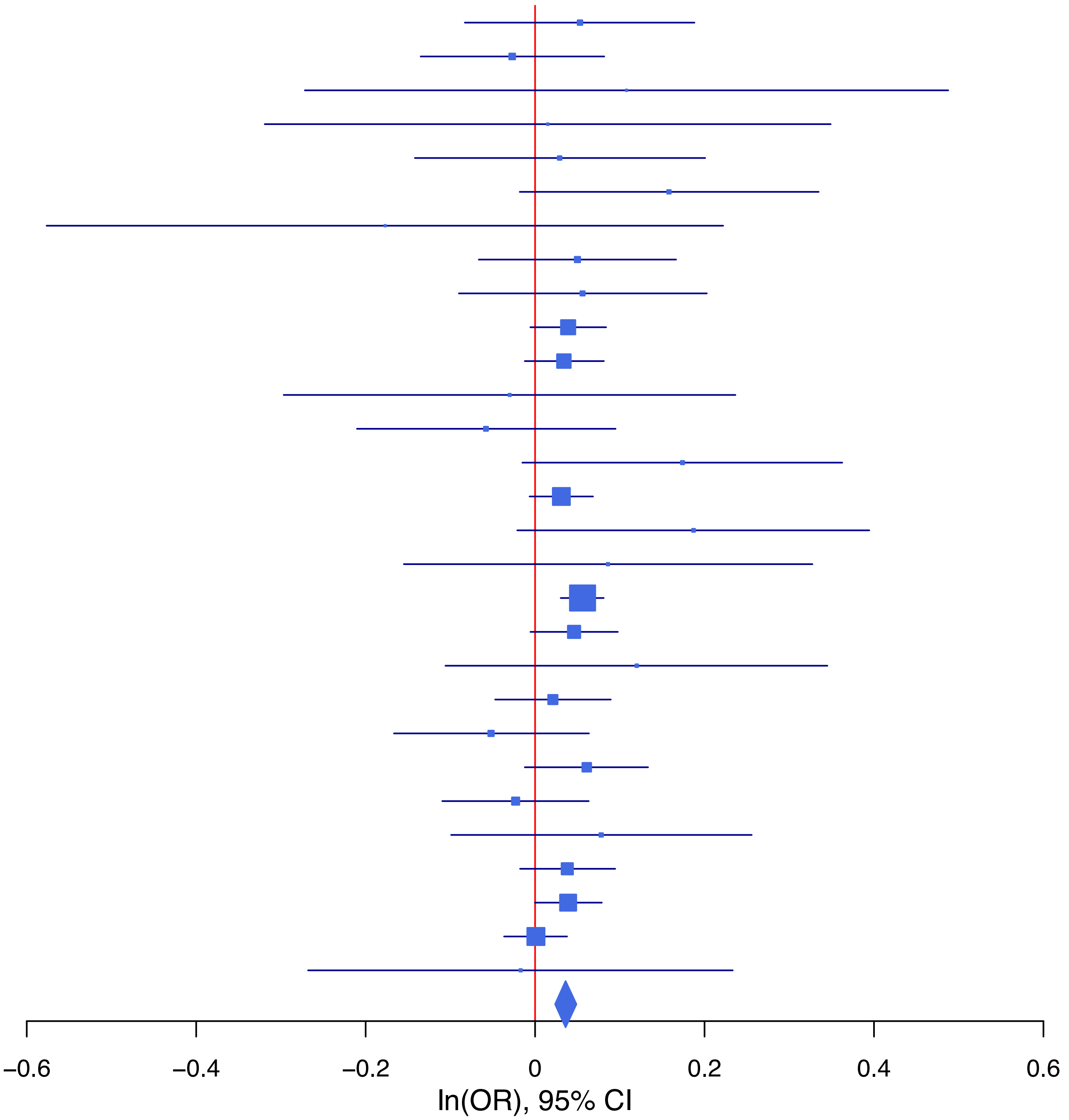

**rs2295402 A/T 14:103338324**

| Cohort | P | ln(OR) | SE |
|---|---|---|---|
| ALSPC_EUR | 0.0088 | 0.17 | 0.065 |
| BEPS7_EUR | 0.8371 | 0.046 | 0.226 |
| BHRCM_EUR | 0.9071 | 0.024 | 0.204 |
| BOR17_EUR | 0.6678 | 0.047 | 0.109 |
| BOR2C_EUR | 0.2711 | −0.119 | 0.108 |
| BOR2E_EUR | 0.4328 | −0.163 | 0.208 |
| COGA1_EUR | 0.7583 | −0.022 | 0.073 |
| ESTB2_EUR | 0.1044 | 0.047 | 0.029 |
| FINNG_EUR | 0.006 | 0.073 | 0.027 |
| GEDIS_EUR | 0.7434 | −0.052 | 0.16 |
| GISS1_EUR | 0.4141 | −0.083 | 0.102 |
| GISS2_EUR | 0.7618 | 0.035 | 0.116 |
| IPSYC_EUR | 0.0979 | 0.038 | 0.023 |
| JANS3_EUR | 0.8992 | −0.017 | 0.132 |
| JANS4_EUR | 0.1983 | −0.189 | 0.147 |
| MIREC_EUR | 0.9735 | −0.006 | 0.19 |
| MVPXQ_EUR | 1.46e−06 | 0.074 | 0.015 |
| PGCBD_EUR | 0.5228 | −0.021 | 0.033 |
| PGCPT_EUR | 0.2403 | 0.083 | 0.07 |
| PGCSZ_EUR | 0.4012 | 0.039 | 0.046 |
| PRFCT_EUR | 0.0462 | 0.104 | 0.052 |
| PSYCR_EUR | 0.1992 | 0.145 | 0.113 |
| QIMRB_EUR | 0.6521 | 0.016 | 0.035 |
| UKBJC_EUR | 0.002 | 0.076 | 0.025 |
| UTAH2_EUR | 0.2729 | 0.026 | 0.024 |
| YPENN_EUR | 0.622 | −0.078 | 0.159 |
| **meta** | **2.57e−10** | **0.05** | **0.008** |

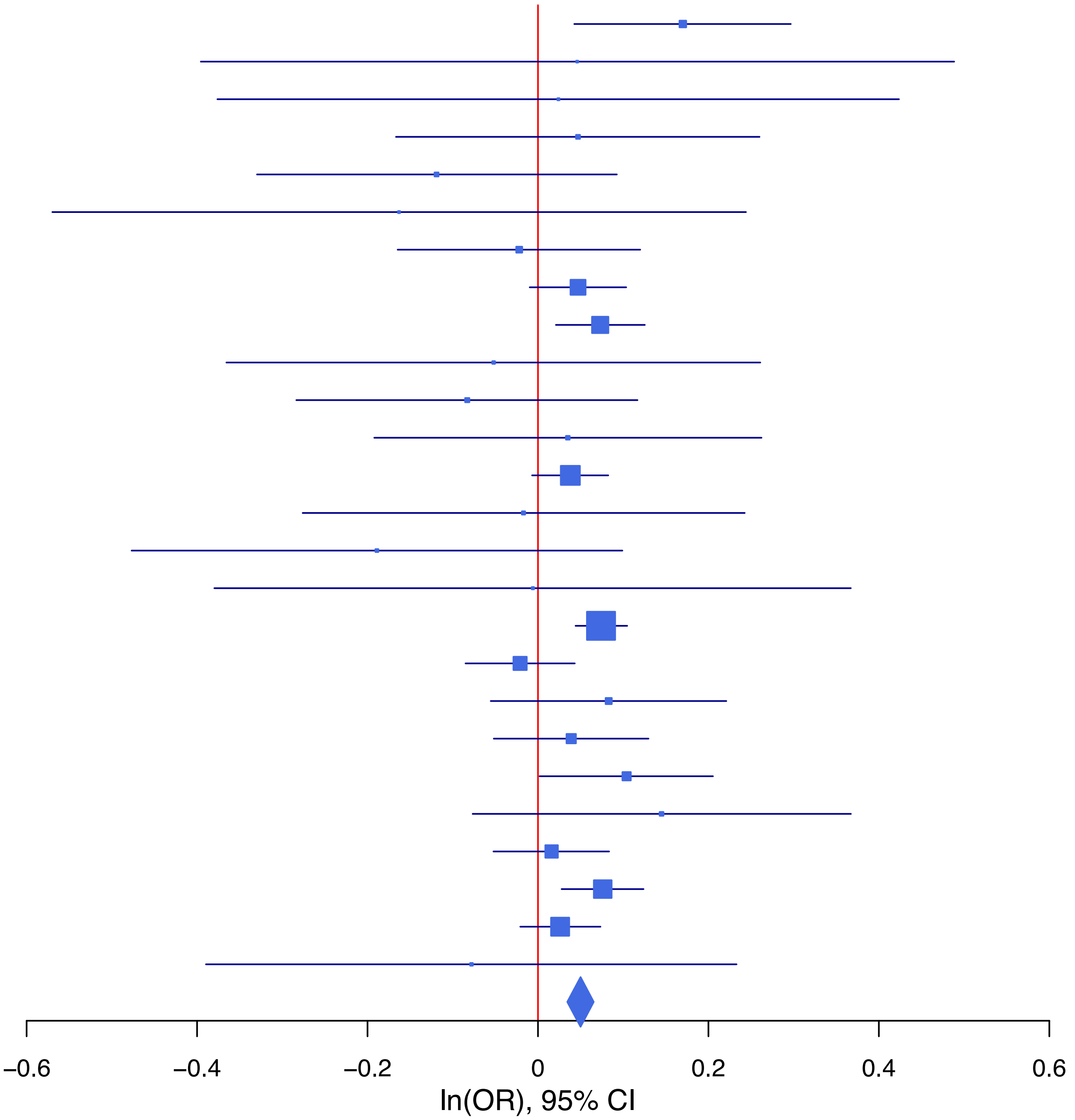

**rs2503185 G/A 1:66461401**

| Cohort | P | ln(OR) | SE |
|---|---|---|---|
| ALSPC_EUR | 0.525 | −0.034 | 0.054 |
| BEPS7_EUR | 0.7202 | −0.063 | 0.176 |
| BHRCM_EUR | 0.3888 | −0.136 | 0.158 |
| BOR17_EUR | 0.181 | 0.113 | 0.084 |
| BOR2C_EUR | 0.0609 | −0.162 | 0.086 |
| BOR2E_EUR | 0.5384 | 0.108 | 0.176 |
| COGA1_EUR | 0.9308 | −0.005 | 0.059 |
| CUINT_EUR | 0.6235 | 0.036 | 0.074 |
| ESTB2_EUR | 0.1457 | −0.033 | 0.023 |
| FINNG_EUR | 0.0782 | −0.04 | 0.023 |
| GEDIS_EUR | 0.5361 | −0.079 | 0.128 |
| GISS1_EUR | 0.258 | 0.088 | 0.078 |
| GISS2_EUR | 0.1551 | 0.13 | 0.091 |
| IPSYC_EUR | 1.25e−05 | −0.081 | 0.018 |
| JANS3_EUR | 0.8184 | −0.023 | 0.102 |
| JANS4_EUR | 0.9833 | −0.002 | 0.114 |
| MIREC_EUR | 0.525 | 0.096 | 0.151 |
| MVPXQ_EUR | 0.0013 | −0.041 | 0.013 |
| PGCBD_EUR | 0.0503 | −0.05 | 0.026 |
| PGCED_EUR | 0.4949 | −0.078 | 0.114 |
| PGCMD_EUR | 0.0648 | −0.063 | 0.034 |
| PGCPT_EUR | 0.642 | −0.027 | 0.058 |
| PGCSZ_EUR | 0.2226 | −0.045 | 0.037 |
| PRFCT_EUR | 0.0068 | −0.118 | 0.043 |
| PSYCR_EUR | 0.7537 | −0.027 | 0.087 |
| QIMRB_EUR | 0.8134 | −0.006 | 0.027 |
| UKBJC_EUR | 0.0249 | −0.044 | 0.02 |
| UTAH2_EUR | 0.0064 | −0.052 | 0.019 |
| YPENN_EUR | 0.0715 | 0.229 | 0.127 |
| **meta** | **3.12e−12** | **−0.044** | **0.006** |

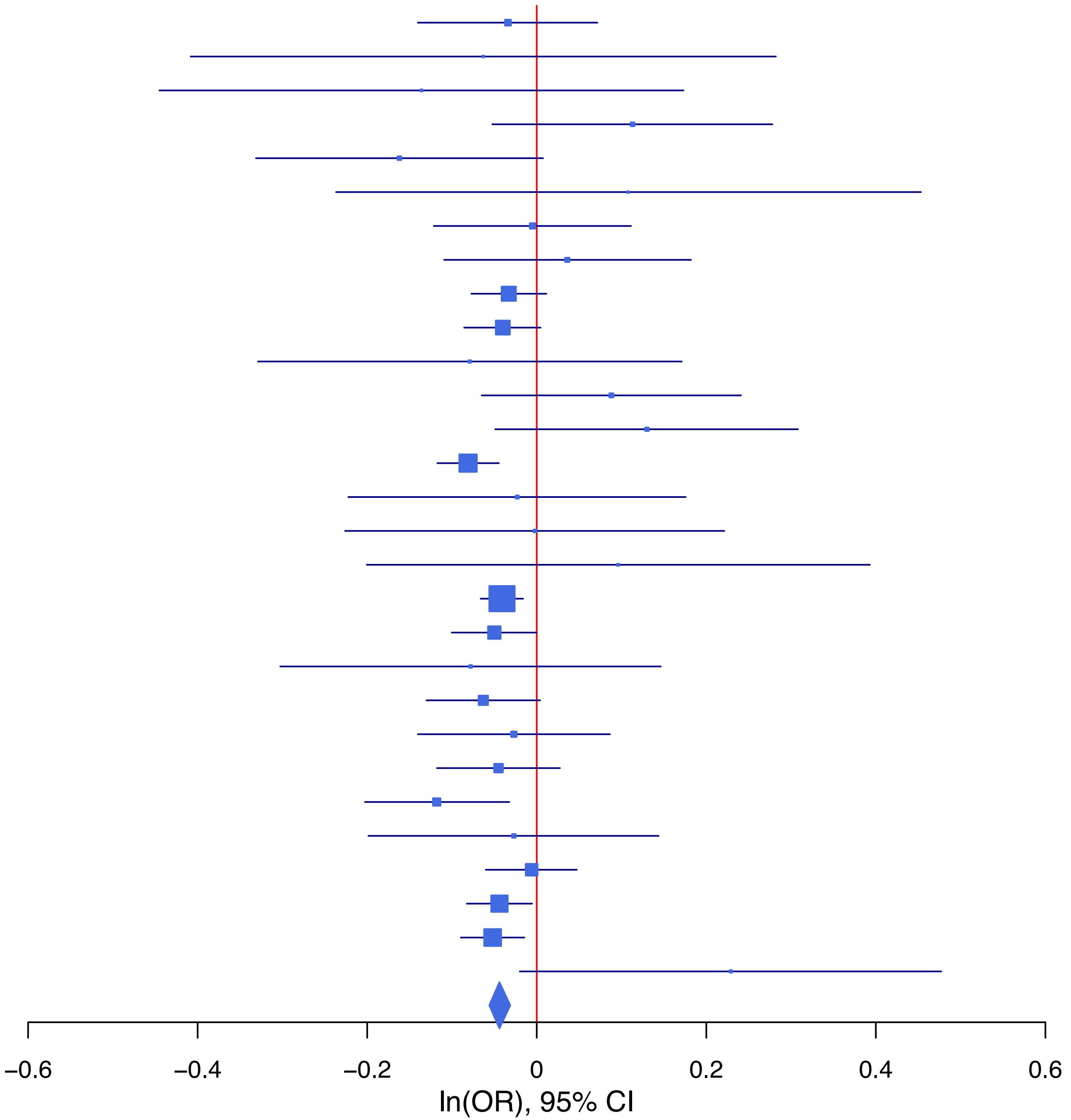

**rs4305732 A/G 6:152240448**

| Cohort | P | ln(OR) | SE |
|---|---|---|---|
| ALSPC_EUR | 0.6778 | −0.023 | 0.056 |
| BEPS7_EUR | 0.6492 | 0.085 | 0.187 |
| BHRCM_EUR | 0.5377 | −0.111 | 0.18 |
| BOR17_EUR | 0.1939 | 0.118 | 0.091 |
| BOR2C_EUR | 0.3861 | 0.078 | 0.09 |
| BOR2E_EUR | 0.1923 | 0.242 | 0.186 |
| COGA1_EUR | 0.1446 | 0.089 | 0.061 |
| CUINT_EUR | 0.1481 | −0.114 | 0.079 |
| ESTB2_EUR | 0.001 | 0.08 | 0.024 |
| FINNG_EUR | 0.5498 | 0.014 | 0.024 |
| GEDIS_EUR | 0.8972 | 0.018 | 0.136 |
| GISS1_EUR | 0.4307 | −0.069 | 0.088 |
| GISS2_EUR | 0.8851 | 0.014 | 0.099 |
| IPSYC_EUR | 0.6937 | 0.008 | 0.019 |
| JANS3_EUR | 0.4963 | −0.075 | 0.11 |
| JANS4_EUR | 0.7285 | 0.044 | 0.128 |
| MVPXQ_EUR | 7.96e−07 | 0.066 | 0.013 |
| PGCBD_EUR | 0.3649 | 0.025 | 0.027 |
| PGCED_EUR | 0.0702 | −0.227 | 0.125 |
| PGCMD_EUR | 0.233 | 0.043 | 0.036 |
| PGCPT_EUR | 0.1496 | 0.086 | 0.06 |
| PGCSZ_EUR | 0.419 | 0.031 | 0.039 |
| PRFCT_EUR | 0.0011 | 0.149 | 0.046 |
| PSYCR_EUR | 0.1498 | 0.132 | 0.092 |
| QIMRB_EUR | 0.2196 | 0.035 | 0.028 |
| UKBJC_EUR | 7.44e−06 | 0.09 | 0.02 |
| UTAH2_EUR | 0.6414 | 0.009 | 0.02 |
| YPENN_EUR | 0.6556 | 0.058 | 0.131 |
| **meta** | **7.71e−12** | **0.045** | **0.006** |

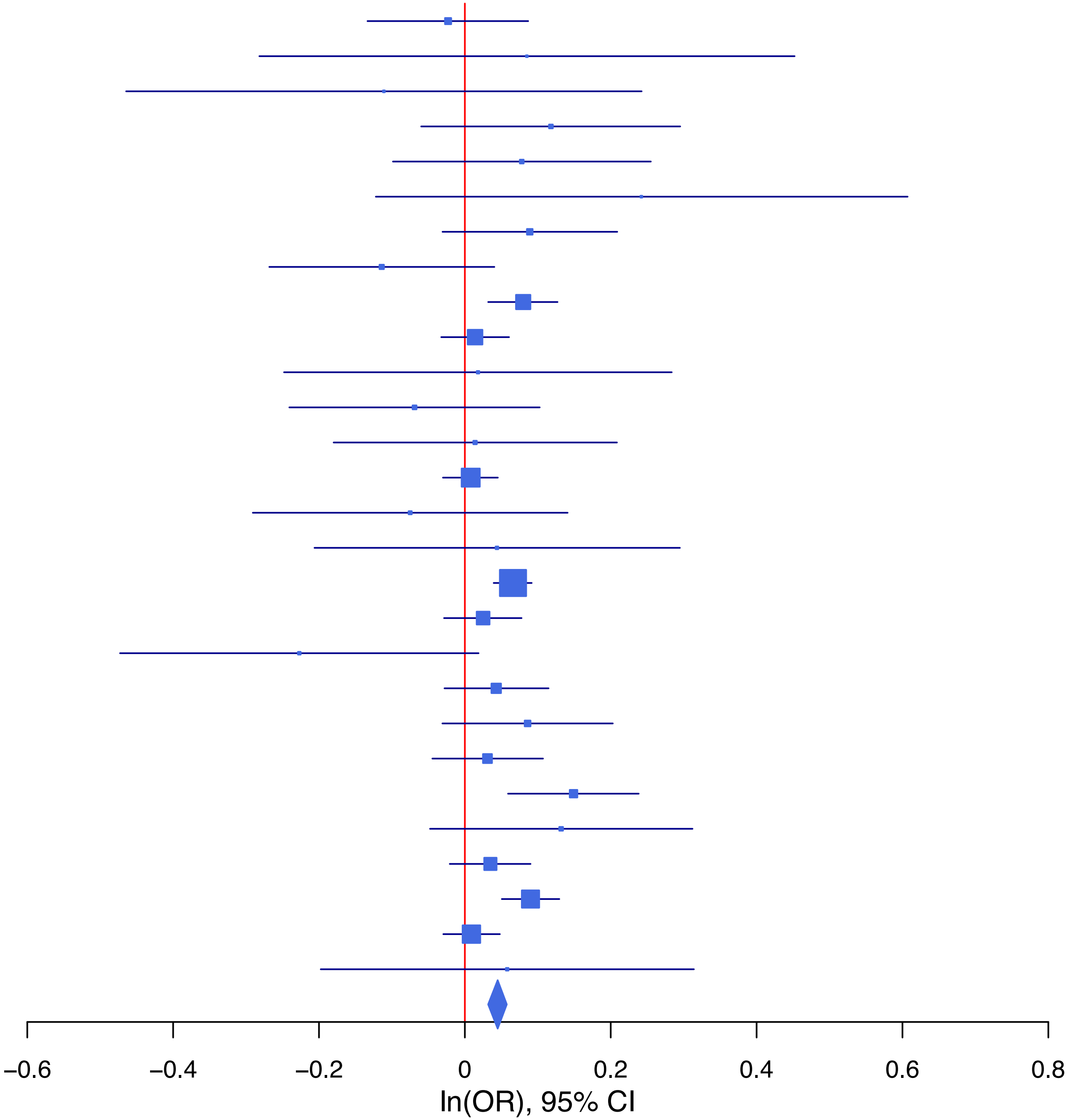

**rs4632195 C/T 18:50746748**

| Cohort | P | ln(OR) | SE |
|---|---|---|---|
| ADHEA_EUR | 0.5467 | 0.041 | 0.068 |
| ALSPC_EUR | 0.4583 | −0.04 | 0.054 |
| BEPS7_EUR | 0.3121 | 0.179 | 0.177 |
| BHRCM_EUR | 0.5189 | −0.102 | 0.158 |
| BOR17_EUR | 0.0012 | −0.273 | 0.084 |
| BOR2C_EUR | 0.0104 | −0.221 | 0.086 |
| BOR2E_EUR | 0.9627 | −0.008 | 0.179 |
| COGA1_EUR | 0.7583 | −0.018 | 0.059 |
| CUINT_EUR | 0.2731 | −0.08 | 0.073 |
| ESTB2_EUR | 0.0657 | −0.042 | 0.023 |
| FINNG_EUR | 0.8201 | 0.005 | 0.023 |
| GEDIS_EUR | 0.1004 | 0.218 | 0.132 |
| GISS1_EUR | 0.4107 | −0.064 | 0.078 |
| GISS2_EUR | 0.6135 | 0.047 | 0.092 |
| IPSYC_EUR | 0.141 | −0.027 | 0.018 |
| JANS3_EUR | 0.9745 | −0.003 | 0.103 |
| JANS4_EUR | 0.0554 | 0.223 | 0.116 |
| MIREC_EUR | 0.9089 | 0.018 | 0.154 |
| MVPXQ_EUR | 0.0044 | −0.035 | 0.012 |
| PGCBD_EUR | 0.539 | −0.016 | 0.026 |
| PGCED_EUR | 0.0638 | −0.213 | 0.115 |
| PGCMD_EUR | 0.1361 | −0.051 | 0.034 |
| PGCPT_EUR | 0.6744 | 0.024 | 0.057 |
| PGCSZ_EUR | 0.3181 | −0.037 | 0.037 |
| PRFCT_EUR | 0.9267 | −0.004 | 0.044 |
| PSYCR_EUR | 0.2269 | −0.102 | 0.084 |
| QIMRB_EUR | 0.2559 | −0.031 | 0.027 |
| UKBJC_EUR | 4e−04 | −0.069 | 0.02 |
| UTAH2_EUR | 0.002 | −0.06 | 0.019 |
| YPENN_EUR | 0.0501 | −0.249 | 0.127 |
| **meta** | **2.15e−09** | **−0.037** | **0.006** |

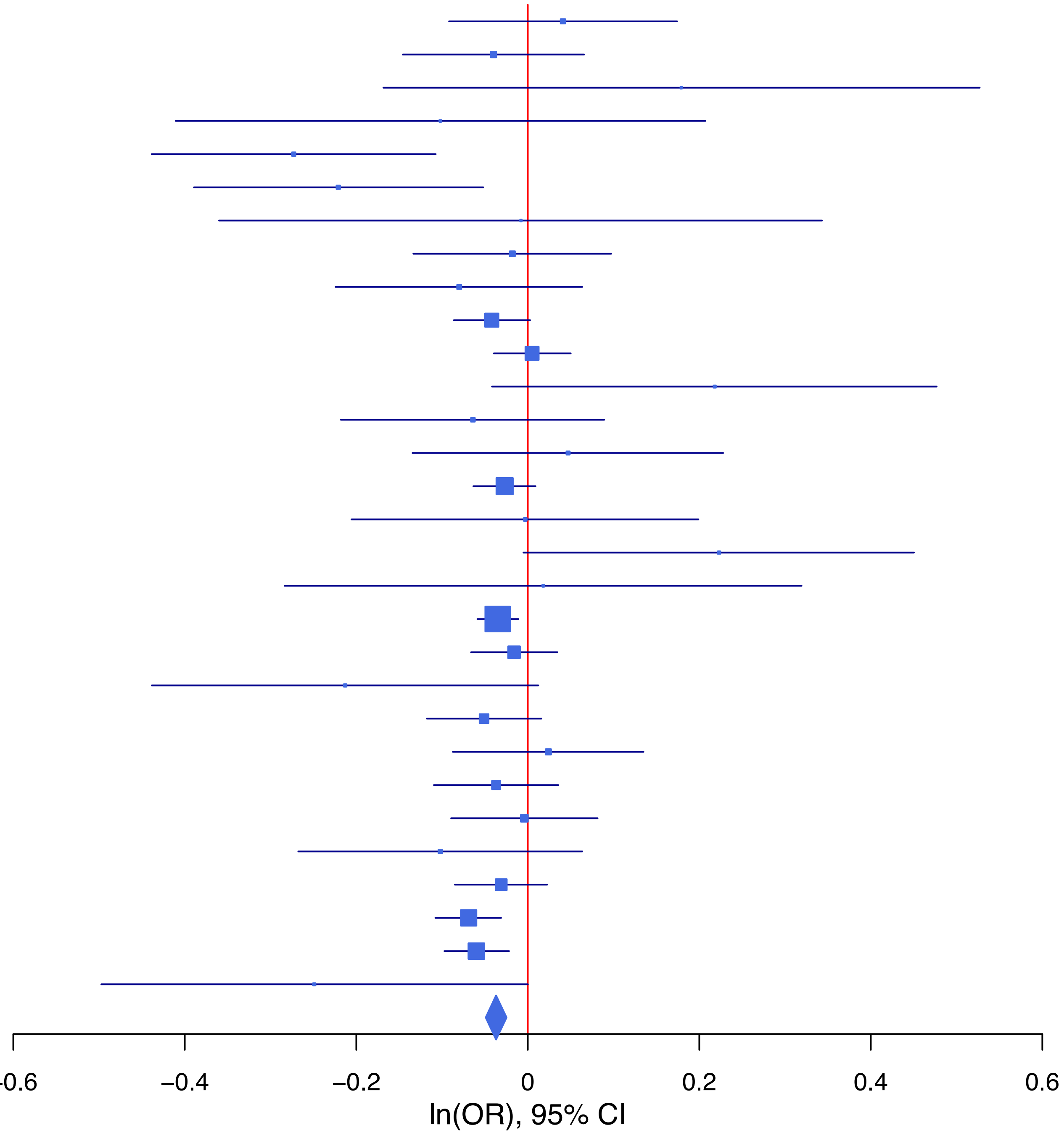

**rs4771932 A/G 13:96950542**

| Cohort | P | ln(OR) | SE |
|---|---|---|---|
| ADHEA_EUR | 0.1195 | 0.111 | 0.071 |
| ALSPC_EUR | 0.4649 | −0.043 | 0.059 |
| BEPS7_EUR | 0.3173 | −0.185 | 0.185 |
| BOR17_EUR | 0.0054 | 0.247 | 0.089 |
| BOR2C_EUR | 0.6638 | 0.04 | 0.092 |
| BOR2E_EUR | 0.1954 | −0.245 | 0.189 |
| COGA1_EUR | 0.7169 | 0.022 | 0.062 |
| CUINT_EUR | 0.1119 | −0.127 | 0.08 |
| ESTB2_EUR | 0.0023 | 0.071 | 0.023 |
| FINNG_EUR | 0.2983 | 0.024 | 0.023 |
| GISS2_EUR | 0.8402 | −0.019 | 0.096 |
| IPSYC_EUR | 0.0837 | 0.034 | 0.02 |
| JANS3_EUR | 0.9363 | 0.009 | 0.109 |
| JANS4_EUR | 0.3807 | −0.111 | 0.127 |
| MIREC_EUR | 0.9268 | 0.014 | 0.157 |
| MVPXQ_EUR | 0.0246 | 0.03 | 0.013 |
| PGCBD_EUR | 0.2496 | 0.031 | 0.027 |
| PGCED_EUR | 0.1783 | −0.166 | 0.124 |
| PGCMD_EUR | 0.5736 | 0.02 | 0.036 |
| PGCPT_EUR | 0.072 | 0.108 | 0.06 |
| PGCSZ_EUR | 0.6293 | 0.019 | 0.04 |
| PRFCT_EUR | 0.0756 | 0.083 | 0.046 |
| PSYCR_EUR | 0.9156 | −0.01 | 0.093 |
| QIMRB_EUR | 0.0508 | 0.057 | 0.029 |
| UKBJC_EUR | 0.0036 | 0.061 | 0.021 |
| UTAH2_EUR | 0.0093 | 0.052 | 0.02 |
| YPENN_EUR | 0.9698 | 0.005 | 0.135 |
| **meta** | **2.39e−09** | **0.039** | **0.007** |

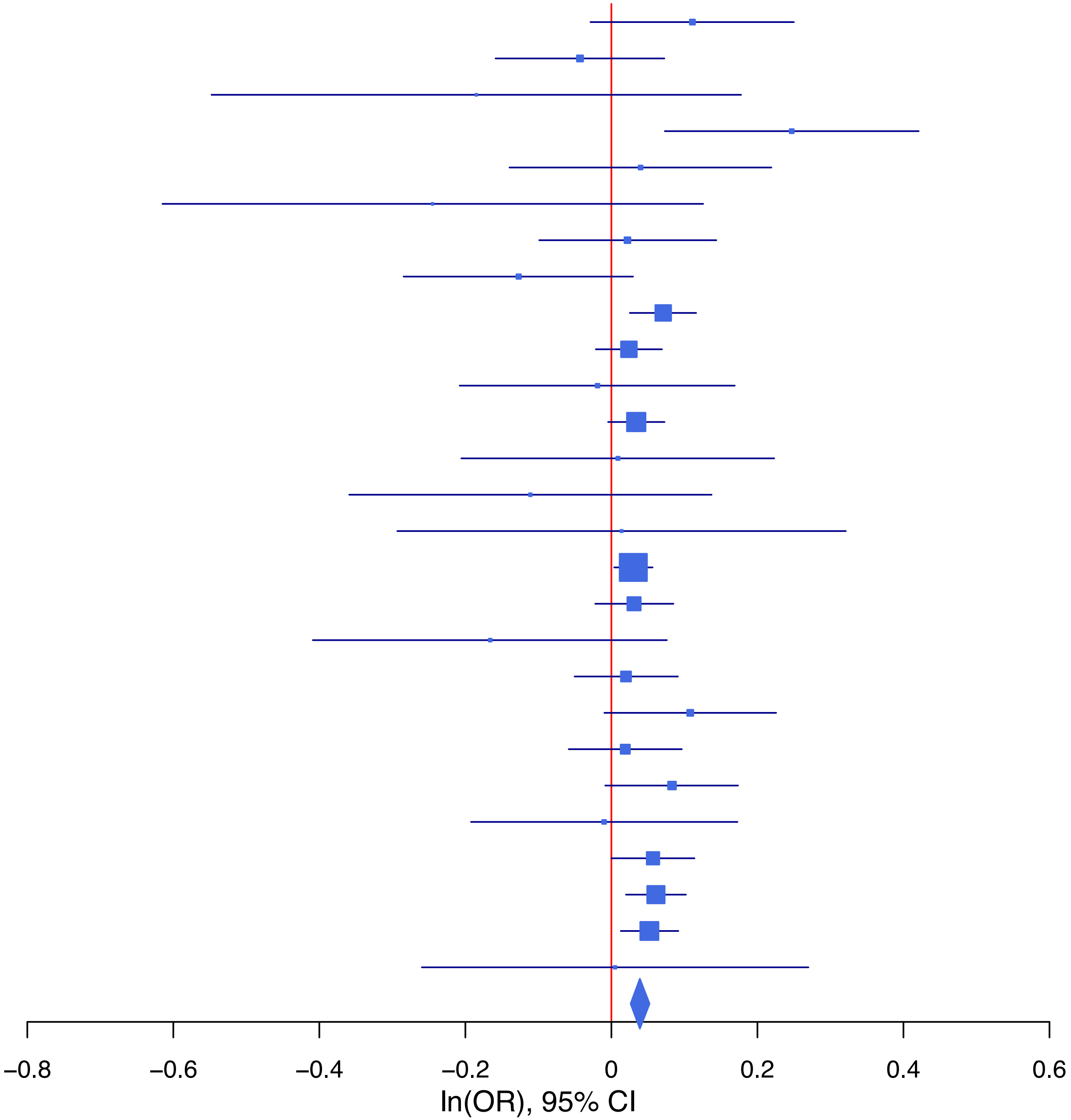

**rs4974203 T/C 3:56390992**

| Cohort | P | ln(OR) | SE |
|---|---|---|---|
| ADHEA_EUR | 0.97 | 0.003 | 0.075 |
| ALSPC_EUR | 0.1236 | 0.091 | 0.059 |
| BEPS7_EUR | 0.8633 | −0.036 | 0.207 |
| BHRCM_EUR | 0.3389 | −0.193 | 0.202 |
| BOR17_EUR | 0.0658 | 0.181 | 0.098 |
| BOR2C_EUR | 0.6214 | −0.049 | 0.099 |
| BOR2E_EUR | 0.1239 | 0.304 | 0.198 |
| COGA1_EUR | 0.8579 | 0.012 | 0.064 |
| CUINT_EUR | 0.8762 | −0.013 | 0.086 |
| ESTB2_EUR | 0.0914 | 0.044 | 0.026 |
| FINNG_EUR | 0.4073 | 0.021 | 0.025 |
| GEDIS_EUR | 0.2394 | 0.166 | 0.141 |
| GISS1_EUR | 0.5787 | 0.052 | 0.093 |
| GISS2_EUR | 0.9132 | −0.012 | 0.109 |
| IPSYC_EUR | 0.0069 | 0.054 | 0.02 |
| JANS3_EUR | 0.5429 | −0.071 | 0.117 |
| JANS4_EUR | 0.661 | 0.058 | 0.132 |
| MIREC_EUR | 0.2058 | −0.223 | 0.176 |
| MVPXQ_EUR | 0.0236 | 0.031 | 0.014 |
| PGCBD_EUR | 0.2811 | 0.031 | 0.029 |
| PGCED_EUR | 0.7211 | −0.047 | 0.131 |
| PGCMD_EUR | 0.1431 | 0.056 | 0.038 |
| PGCPT_EUR | 0.5048 | −0.043 | 0.064 |
| PGCSZ_EUR | 0.4098 | 0.034 | 0.041 |
| PRFCT_EUR | 0.117 | 0.075 | 0.048 |
| PSYCR_EUR | 0.9863 | −0.002 | 0.099 |
| QIMRB_EUR | 0.0315 | 0.066 | 0.031 |
| UKBJC_EUR | 0.0091 | 0.056 | 0.021 |
| UTAH2_EUR | 0.0097 | 0.054 | 0.021 |
| YPENN_EUR | 0.7014 | −0.056 | 0.147 |
| **meta** | **2.15e−09** | **0.041** | **0.007** |

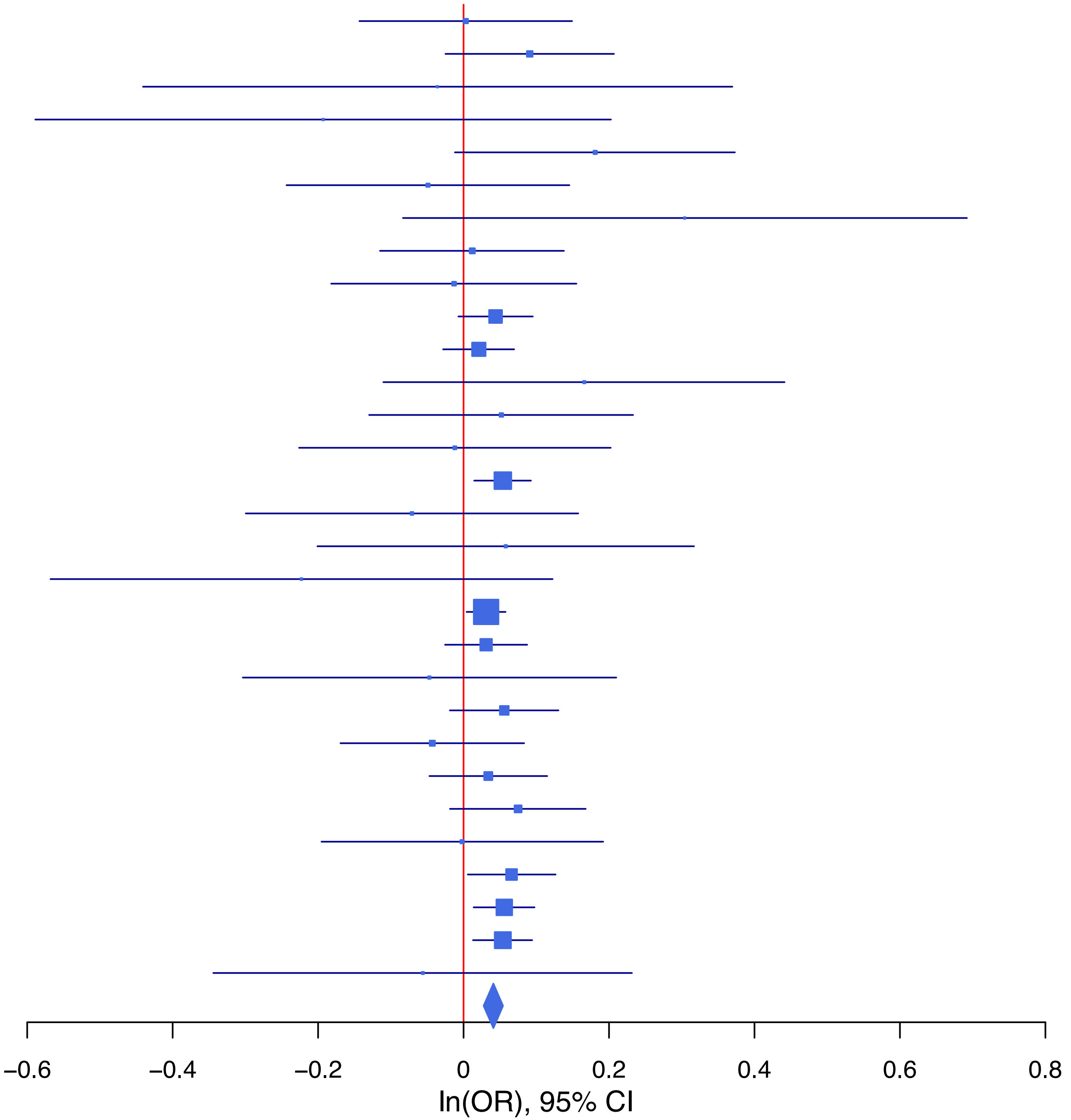

**rs499168 A/T 1:38228797**

| Cohort | P | ln(OR) | SE |
|---|---|---|---|
| ADHEA_EUR | 0.37 | −0.061 | 0.068 |
| ALSPC_EUR | 0.4812 | 0.038 | 0.054 |
| BEPS7_EUR | 0.257 | −0.202 | 0.178 |
| BHRCM_EUR | 0.6076 | −0.08 | 0.156 |
| BOR17_EUR | 0.4035 | −0.073 | 0.087 |
| BOR2C_EUR | 0.3764 | 0.077 | 0.088 |
| BOR2E_EUR | 0.7104 | 0.069 | 0.185 |
| COGA1_EUR | 0.4647 | 0.043 | 0.059 |
| CUINT_EUR | 0.4989 | 0.051 | 0.074 |
| ESTB2_EUR | 0.0465 | −0.046 | 0.023 |
| FINNG_EUR | 0.0425 | −0.046 | 0.023 |
| GEDIS_EUR | 0.6274 | −0.063 | 0.13 |
| GISS1_EUR | 0.1036 | −0.129 | 0.079 |
| GISS2_EUR | 0.4017 | 0.08 | 0.095 |
| IPSYC_EUR | 0.0347 | −0.039 | 0.018 |
| JANS3_EUR | 0.8317 | −0.022 | 0.105 |
| JANS4_EUR | 0.6562 | 0.054 | 0.12 |
| MVPXQ_EUR | 2e−04 | −0.046 | 0.012 |
| PGCBD_EUR | 0.1098 | −0.042 | 0.026 |
| PGCED_EUR | 0.3287 | −0.116 | 0.119 |
| PGCMD_EUR | 0.1233 | −0.055 | 0.035 |
| PGCPT_EUR | 0.8999 | −0.007 | 0.058 |
| PGCSZ_EUR | 0.7789 | 0.011 | 0.038 |
| PRFCT_EUR | 8e−04 | −0.147 | 0.044 |
| PSYCR_EUR | 0.1234 | −0.136 | 0.088 |
| QIMRB_EUR | 0.7763 | 0.008 | 0.027 |
| UKBJC_EUR | 0.0132 | −0.049 | 0.02 |
| UTAH2_EUR | 0.0019 | −0.059 | 0.019 |
| YPENN_EUR | 0.0988 | 0.211 | 0.128 |
| **meta** | **5.24e−11** | **−0.041** | **0.006** |

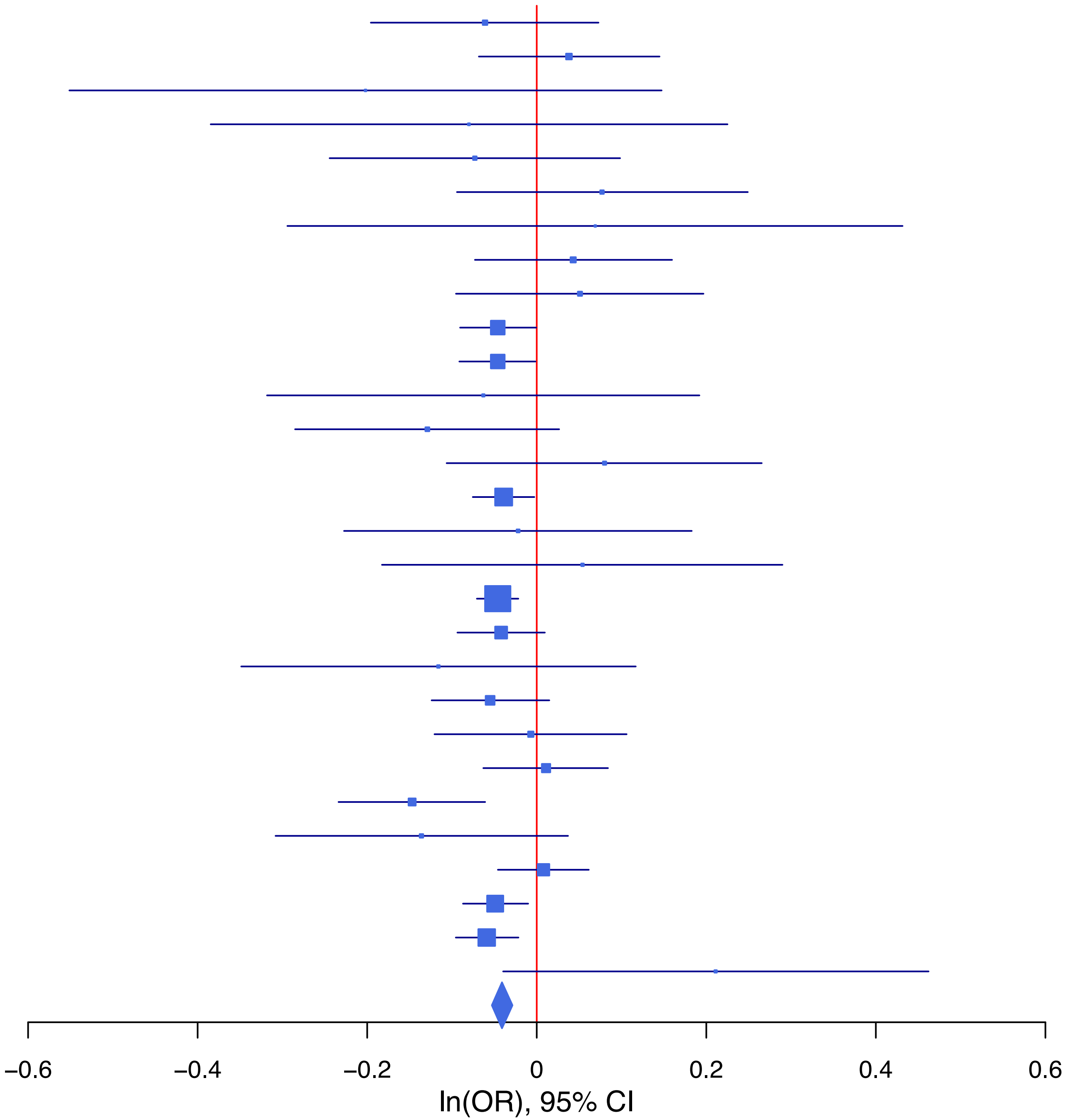

**rs55995895   T/C   7:1862183**

| Cohort | P | ln(OR) | SE |
|---|---|---|---|
| ADHEA_EUR | 0.2072 | −0.121 | 0.096 |
| ALSPC_EUR | 0.7761 | 0.022 | 0.077 |
| BEPS7_EUR | 0.2763 | −0.254 | 0.233 |
| BOR17_EUR | 0.8961 | 0.014 | 0.11 |
| BOR2C_EUR | 0.4235 | −0.088 | 0.11 |
| BOR2E_EUR | 0.4332 | −0.183 | 0.234 |
| COGA1_EUR | 0.1027 | −0.132 | 0.081 |
| CUINT_EUR | 0.626 | 0.048 | 0.098 |
| ESTB2_EUR | 0.0091 | −0.07 | 0.027 |
| FINNG_EUR | 8e−04 | −0.09 | 0.027 |
| GISS1_EUR | 0.7848 | 0.025 | 0.091 |
| GISS2_EUR | 0.7959 | −0.027 | 0.103 |
| IPSYC_EUR | 0.0025 | −0.077 | 0.025 |
| JANS3_EUR | 0.277 | −0.157 | 0.144 |
| JANS4_EUR | 0.0214 | −0.376 | 0.163 |
| MIREC_EUR | 0.7999 | −0.053 | 0.208 |
| MVPXQ_EUR | 0.0014 | −0.056 | 0.017 |
| PGCBD_EUR | 0.1966 | 0.044 | 0.034 |
| PGCED_EUR | 0.5168 | −0.108 | 0.167 |
| PGCMD_EUR | 0.2842 | 0.049 | 0.046 |
| PGCPT_EUR | 0.2804 | −0.086 | 0.079 |
| PGCSZ_EUR | 0.3063 | 0.05 | 0.049 |
| PRFCT_EUR | 0.57 | −0.032 | 0.056 |
| PSYCR_EUR | 0.0621 | −0.204 | 0.109 |
| QIMRB_EUR | 0.0018 | −0.115 | 0.037 |
| UKBJC_EUR | 0.0678 | −0.049 | 0.027 |
| YPENN_EUR | 0.4864 | −0.123 | 0.177 |
| **meta** | **7.84e−10** | **−0.053** | **0.009** |

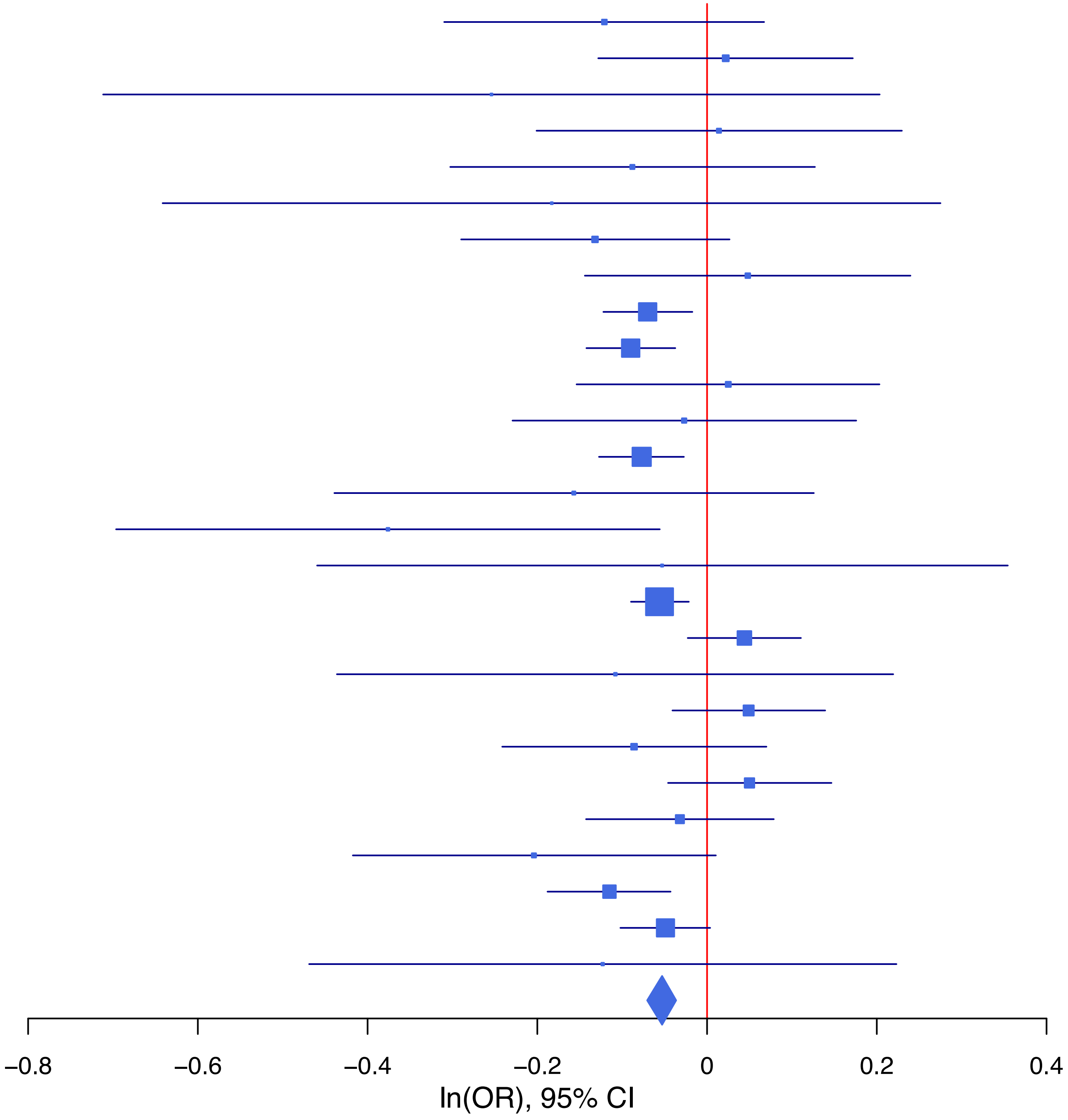

**rs58907748 G/T 6:156468874**

| Cohort | P | ln(OR) | SE |
|---|---|---|---|
| ADHEA_EUR | 0.3721 | 0.111 | 0.124 |
| ALSPC_EUR | 0.6451 | −0.05 | 0.108 |
| BEPS7_EUR | 0.6622 | −0.16 | 0.366 |
| BHRCM_EUR | 0.8506 | −0.057 | 0.302 |
| BOR17_EUR | 0.1115 | −0.273 | 0.172 |
| BOR2C_EUR | 0.0867 | 0.266 | 0.155 |
| BOR2E_EUR | 0.4477 | 0.247 | 0.326 |
| COGA1_EUR | 0.0856 | 0.192 | 0.112 |
| CUINT_EUR | 0.1023 | 0.219 | 0.134 |
| ESTB2_EUR | 0.0929 | 0.075 | 0.045 |
| FINNG_EUR | 0.0145 | 0.14 | 0.058 |
| GEDIS_EUR | 0.8416 | 0.05 | 0.247 |
| GISS1_EUR | 0.5684 | 0.082 | 0.143 |
| GISS2_EUR | 0.5235 | 0.108 | 0.169 |
| IPSYC_EUR | 0.0726 | 0.064 | 0.035 |
| JANS3_EUR | 0.1003 | 0.291 | 0.177 |
| JANS4_EUR | 0.7259 | −0.08 | 0.229 |
| MVPXQ_EUR | 0.0035 | 0.075 | 0.026 |
| PGCBD_EUR | 0.0069 | 0.148 | 0.055 |
| PGCMD_EUR | 0.3937 | 0.062 | 0.072 |
| PGCPT_EUR | 0.7354 | 0.037 | 0.11 |
| PGCSZ_EUR | 0.0683 | 0.134 | 0.074 |
| PRFCT_EUR | 0.0272 | 0.186 | 0.084 |
| PSYCR_EUR | 0.7352 | 0.054 | 0.161 |
| QIMRB_EUR | 0.521 | 0.034 | 0.053 |
| UKBJC_EUR | 0.0017 | 0.12 | 0.038 |
| UTAH2_EUR | 0.808 | 0.009 | 0.038 |
| YPENN_EUR | 0.5045 | 0.154 | 0.231 |
| **meta** | **1.24e−10** | **0.08** | **0.013** |

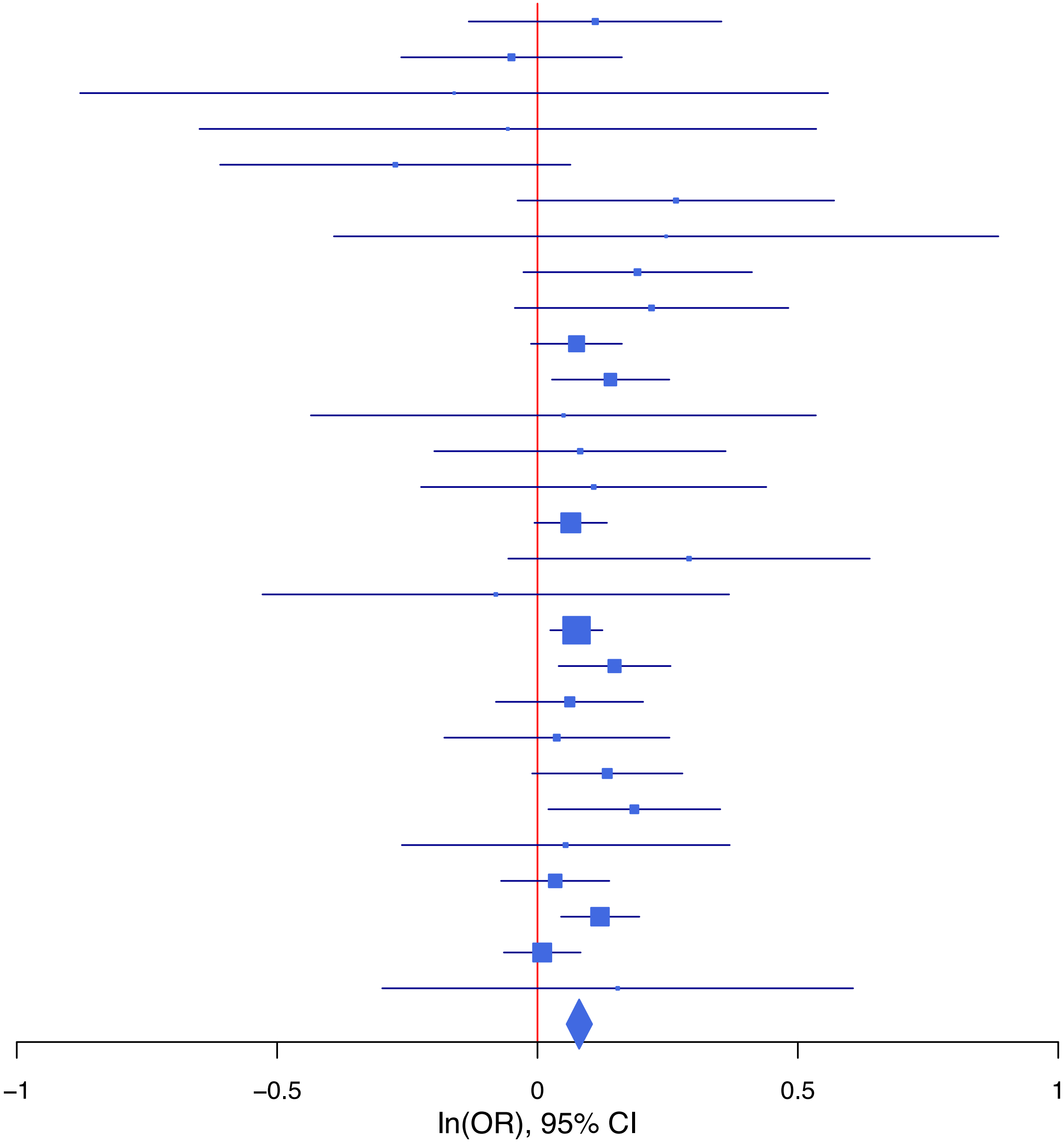

**rs61513449 G/A 5:92570831**

| Cohort | P | ln(OR) | SE |
|---|---|---|---|
| ADHEA_EUR | 0.5936 | 0.05 | 0.093 |
| ALSPC_EUR | 0.2387 | −0.086 | 0.073 |
| BEPS7_EUR | 0.133 | −0.412 | 0.274 |
| BHRCM_EUR | 0.791 | 0.054 | 0.206 |
| BOR17_EUR | 0.5443 | −0.071 | 0.118 |
| BOR2C_EUR | 0.3715 | −0.112 | 0.125 |
| BOR2E_EUR | 0.9893 | −0.003 | 0.242 |
| COGA1_EUR | 0.0015 | −0.256 | 0.081 |
| CUINT_EUR | 0.9674 | 0.004 | 0.107 |
| ESTB2_EUR | 0.9869 | 0.001 | 0.032 |
| FINNG_EUR | 0.4188 | −0.024 | 0.029 |
| GEDIS_EUR | 0.7186 | 0.062 | 0.172 |
| GISS1_EUR | 0.5697 | −0.065 | 0.114 |
| GISS2_EUR | 0.8296 | 0.029 | 0.137 |
| IPSYC_EUR | 0.0051 | −0.071 | 0.025 |
| JANS3_EUR | 0.7721 | −0.041 | 0.142 |
| JANS4_EUR | 0.6652 | −0.073 | 0.17 |
| MIREC_EUR | 0.8713 | 0.032 | 0.202 |
| MVPXQ_EUR | 0.0426 | −0.035 | 0.017 |
| PGCBD_EUR | 1e−04 | −0.14 | 0.037 |
| PGCED_EUR | 0.1356 | −0.254 | 0.17 |
| PGCMD_EUR | 0.385 | −0.042 | 0.049 |
| PGCPT_EUR | 0.0647 | −0.15 | 0.081 |
| PGCSZ_EUR | 0.4413 | −0.039 | 0.051 |
| PRFCT_EUR | 0.9755 | 0.002 | 0.062 |
| PSYCR_EUR | 0.5214 | 0.079 | 0.124 |
| QIMRB_EUR | 0.6802 | −0.016 | 0.038 |
| UKBJC_EUR | 0.0047 | −0.075 | 0.027 |
| UTAH2_EUR | 0.338 | −0.024 | 0.025 |
| YPENN_EUR | 0.6626 | −0.079 | 0.181 |
| **meta** | **3.97e−08** | **−0.047** | **0.009** |

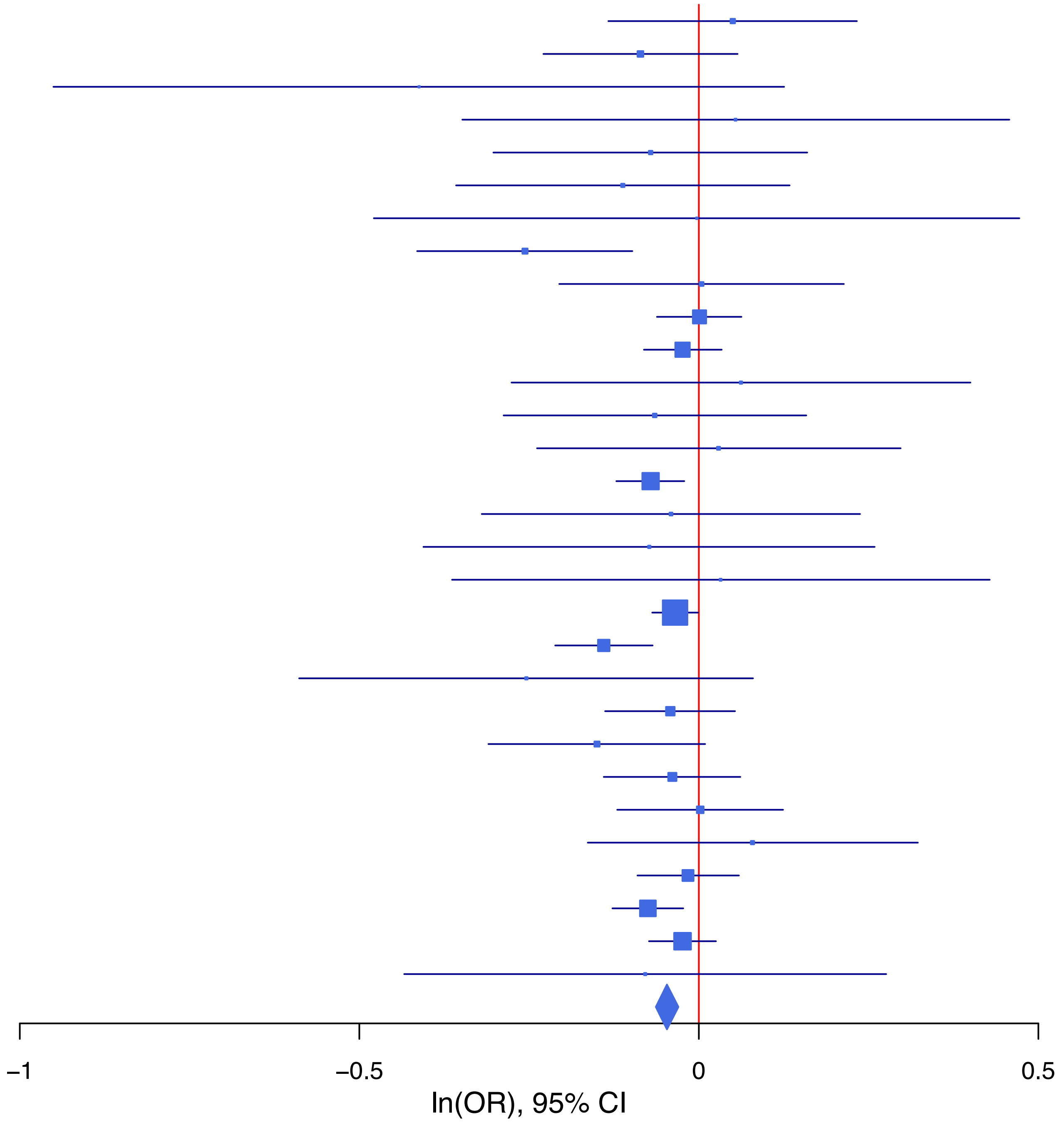

**rs62084682 C/G 17:66031521**

| Cohort | P | ln(OR) | SE |
|---|---|---|---|
| ALSPC_EUR | 0.991 | −0.001 | 0.074 |
| BEPS7_EUR | 0.6129 | 0.111 | 0.219 |
| BHRCM_EUR | 0.4395 | −0.161 | 0.208 |
| BOR17_EUR | 0.9128 | −0.012 | 0.108 |
| BOR2C_EUR | 0.0621 | 0.199 | 0.106 |
| BOR2E_EUR | 0.9438 | −0.017 | 0.242 |
| ESTB2_EUR | 0.0784 | 0.047 | 0.027 |
| GEDIS_EUR | 0.0735 | 0.273 | 0.152 |
| GISS1_EUR | 0.6551 | −0.044 | 0.099 |
| GISS2_EUR | 0.0269 | 0.257 | 0.116 |
| IPSYC_EUR | 0.5016 | 0.015 | 0.022 |
| JANS3_EUR | 0.3448 | −0.122 | 0.129 |
| JANS4_EUR | 0.4741 | 0.103 | 0.144 |
| MVPXQ_EUR | 5e−04 | 0.051 | 0.015 |
| PGCBD_EUR | 0.5011 | 0.023 | 0.033 |
| PGCED_EUR | 0.2984 | 0.19 | 0.183 |
| PGCMD_EUR | 0.4501 | 0.033 | 0.044 |
| PGCPT_EUR | 0.4165 | 0.056 | 0.069 |
| PGCSZ_EUR | 0.9496 | −0.003 | 0.048 |
| PRFCT_EUR | 0.0765 | 0.092 | 0.052 |
| PSYCR_EUR | 0.4048 | 0.084 | 0.101 |
| QIMRB_EUR | 0.819 | 0.008 | 0.034 |
| UKBJC_EUR | 0.0226 | 0.054 | 0.024 |
| UTAH2_EUR | 0.0018 | 0.069 | 0.022 |
| YPENN_EUR | 0.592 | −0.085 | 0.159 |
| **meta** | **2.50e−08** | **0.044** | **0.008** |

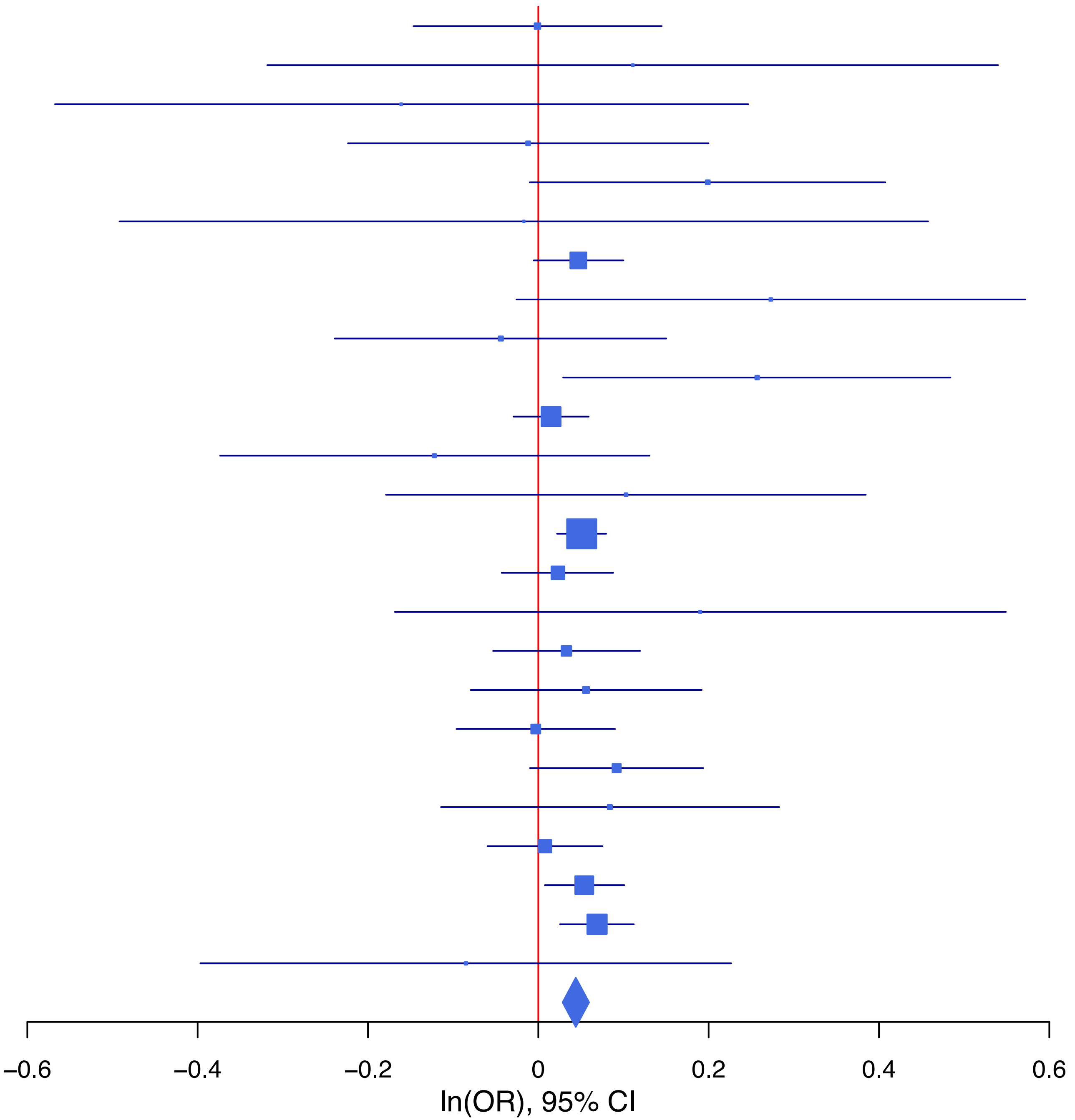

**rs62262722 C/G 3:49769419**

| Cohort | P | ln(OR) | SE |
|---|---|---|---|
| ADHEA_EUR | 0.3354 | −0.086 | 0.089 |
| ALSPC_EUR | 0.7337 | 0.023 | 0.067 |
| BEPS7_EUR | 0.4359 | 0.176 | 0.226 |
| BHRCM_EUR | 0.5168 | 0.126 | 0.194 |
| BOR17_EUR | 0.2607 | 0.125 | 0.111 |
| BOR2C_EUR | 0.0241 | 0.256 | 0.114 |
| BOR2E_EUR | 0.061 | 0.454 | 0.242 |
| COGA1_EUR | 0.0975 | 0.125 | 0.076 |
| CUINT_EUR | 0.3498 | 0.088 | 0.094 |
| ESTB2_EUR | 0.4985 | 0.023 | 0.033 |
| FINNG_EUR | 0.3841 | 0.032 | 0.037 |
| GEDIS_EUR | 0.5506 | 0.097 | 0.163 |
| GISS1_EUR | 0.1556 | −0.155 | 0.109 |
| GISS2_EUR | 0.327 | 0.125 | 0.127 |
| IPSYC_EUR | 0.0386 | 0.048 | 0.023 |
| JANS3_EUR | 0.9224 | −0.013 | 0.135 |
| JANS4_EUR | 0.8354 | 0.032 | 0.153 |
| MVPXQ_EUR | 0.0011 | 0.055 | 0.017 |
| PGCBD_EUR | 0.0039 | 0.096 | 0.033 |
| PGCED_EUR | 0.4369 | 0.126 | 0.162 |
| PGCMD_EUR | 0.5221 | 0.029 | 0.045 |
| PGCPT_EUR | 0.0024 | 0.212 | 0.07 |
| PGCSZ_EUR | 0.5332 | 0.03 | 0.048 |
| PRFCT_EUR | 0.891 | 0.008 | 0.056 |
| PSYCR_EUR | 0.3168 | −0.112 | 0.112 |
| QIMRB_EUR | 0.1166 | 0.056 | 0.036 |
| UKBJC_EUR | 0.0166 | 0.061 | 0.026 |
| YPENN_EUR | 0.1619 | −0.247 | 0.177 |
| **meta** | **2.25e−09** | **0.052** | **0.009** |

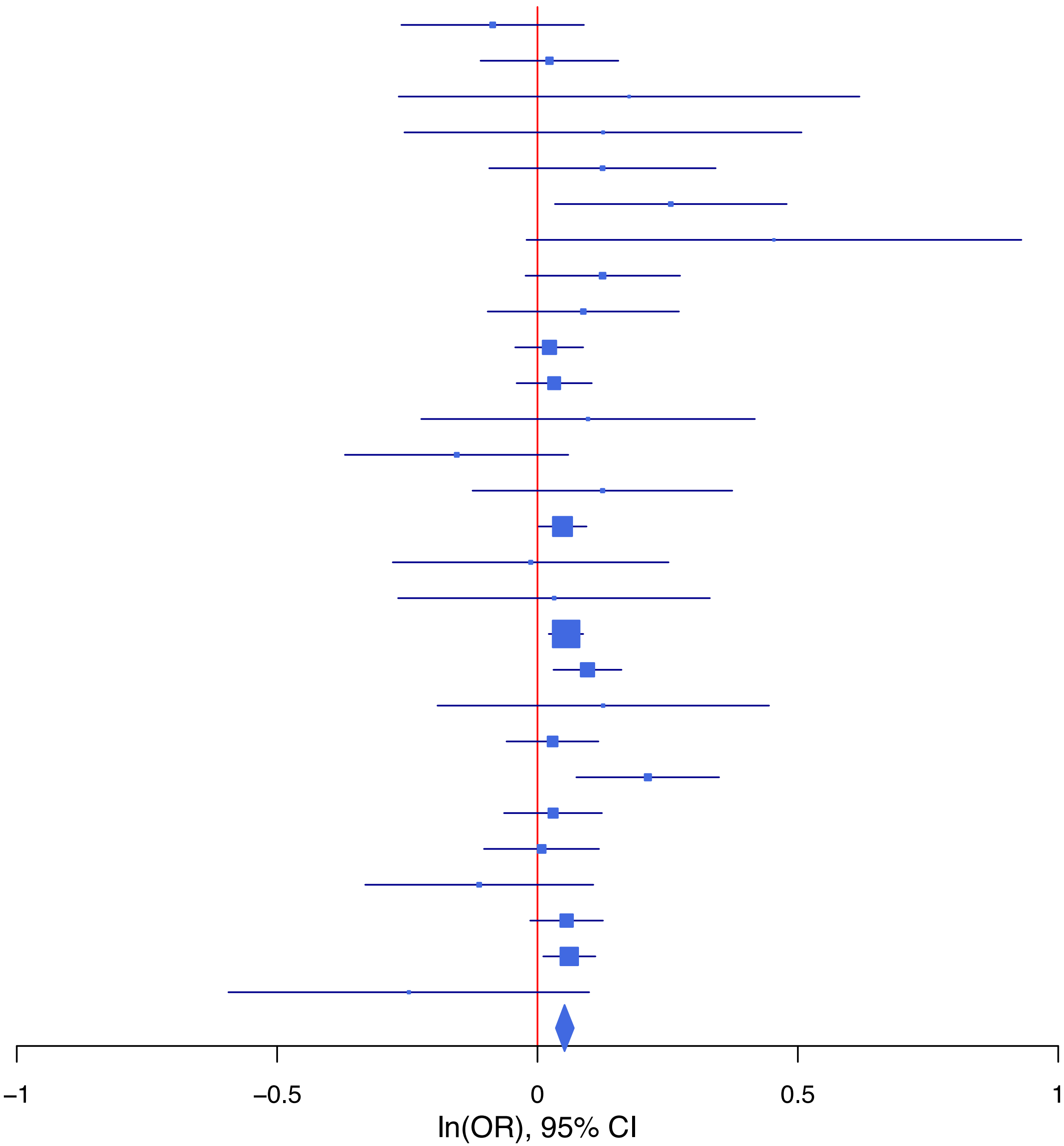

**rs62367522 C/A 5:45280212**

| Cohort | P | ln(OR) | SE |
|---|---|---|---|
| ALSPC_EUR | 0.1126 | −0.113 | 0.071 |
| BEPS7_EUR | 0.9049 | −0.027 | 0.226 |
| BOR17_EUR | 0.4751 | 0.078 | 0.11 |
| BOR2C_EUR | 0.2053 | −0.14 | 0.11 |
| BOR2E_EUR | 0.9648 | 0.01 | 0.228 |
| COGA1_EUR | 0.8662 | 0.013 | 0.075 |
| CUINT_EUR | 0.4997 | −0.063 | 0.094 |
| ESTB2_EUR | 0.0027 | −0.094 | 0.031 |
| FINNG_EUR | 0.4942 | −0.022 | 0.032 |
| GEDIS_EUR | 0.8482 | 0.033 | 0.173 |
| GISS1_EUR | 0.4336 | −0.077 | 0.098 |
| GISS2_EUR | 0.691 | 0.047 | 0.118 |
| IPSYC_EUR | 0.0788 | −0.044 | 0.025 |
| JANS3_EUR | 0.8927 | −0.018 | 0.132 |
| JANS4_EUR | 0.4105 | 0.122 | 0.148 |
| MIREC_EUR | 0.8345 | 0.041 | 0.195 |
| MVPXQ_EUR | 0.002 | −0.052 | 0.017 |
| PGCBD_EUR | 0.4479 | −0.025 | 0.034 |
| PGCED_EUR | 0.369 | 0.125 | 0.14 |
| PGCMD_EUR | 0.2249 | −0.056 | 0.046 |
| PGCPT_EUR | 0.6207 | −0.038 | 0.077 |
| PGCSZ_EUR | 0.9696 | 0.002 | 0.048 |
| PRFCT_EUR | 0.0488 | −0.117 | 0.06 |
| PSYCR_EUR | 0.8606 | 0.019 | 0.109 |
| QIMRB_EUR | 0.0191 | −0.081 | 0.035 |
| UKBJC_EUR | 7e−04 | −0.086 | 0.025 |
| UTAH2_EUR | 0.4114 | −0.02 | 0.025 |
| YPENN_EUR | 0.5228 | −0.104 | 0.163 |
| **meta** | **2.45e−09** | **−0.049** | **0.008** |

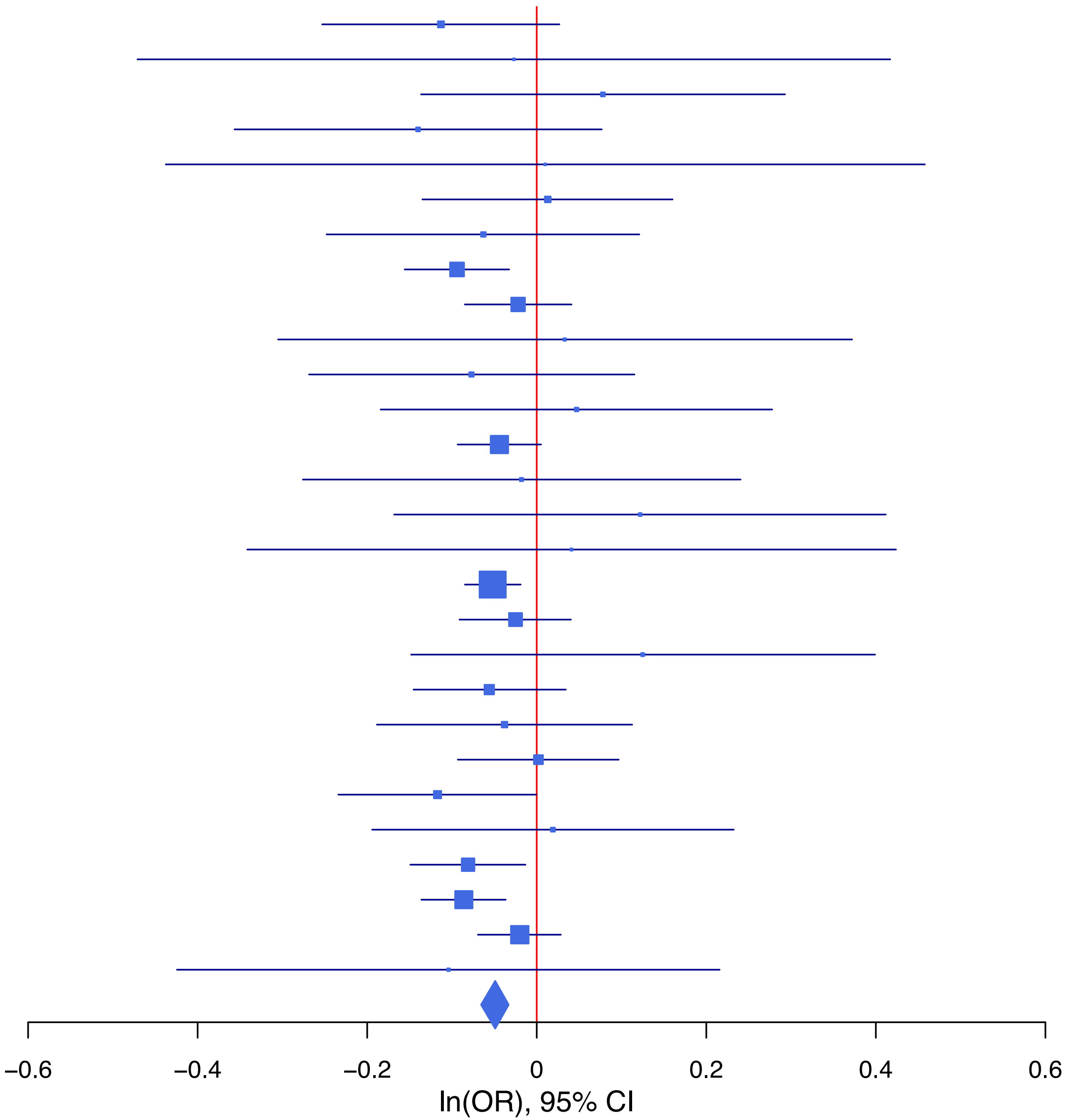

**rs62404522   C/T   6:19307114**

| Cohort | P | ln(OR) | SE |
|---|---|---|---|
| ADHEA_EUR | 0.1413 | −0.154 | 0.105 |
| ALSPC_EUR | 0.2246 | 0.093 | 0.077 |
| BEPS7_EUR | 0.7783 | 0.07 | 0.25 |
| BHRCM_EUR | 0.5224 | −0.166 | 0.26 |
| BOR17_EUR | 0.9851 | −0.002 | 0.126 |
| BOR2C_EUR | 0.067 | 0.233 | 0.127 |
| BOR2E_EUR | 0.3309 | −0.281 | 0.29 |
| COGA1_EUR | 0.1278 | 0.133 | 0.087 |
| CUINT_EUR | 0.9098 | −0.013 | 0.111 |
| ESTB2_EUR | 0.3339 | 0.032 | 0.033 |
| FINNG_EUR | 0.0012 | 0.11 | 0.034 |
| GEDIS_EUR | 0.1439 | 0.257 | 0.176 |
| GISS1_EUR | 0.728 | 0.04 | 0.116 |
| GISS2_EUR | 0.0643 | 0.272 | 0.147 |
| IPSYC_EUR | 0.1929 | 0.034 | 0.026 |
| JANS3_EUR | 0.2348 | 0.171 | 0.144 |
| JANS4_EUR | 0.5364 | −0.116 | 0.187 |
| MVPXQ_EUR | 3.92e−05 | 0.076 | 0.019 |
| PGCBD_EUR | 1e−04 | 0.146 | 0.037 |
| PGCPT_EUR | 0.4313 | 0.063 | 0.08 |
| PGCSZ_EUR | 0.7464 | 0.018 | 0.054 |
| PRFCT_EUR | 0.0078 | 0.161 | 0.06 |
| PSYCR_EUR | 0.081 | −0.227 | 0.13 |
| QIMRB_EUR | 0.0417 | 0.081 | 0.04 |
| UKBJC_EUR | 0.7566 | 0.009 | 0.029 |
| UTAH2_EUR | 0.2209 | 0.033 | 0.027 |
| YPENN_EUR | 0.8099 | 0.045 | 0.188 |
| **meta** | **4.62e−11** | **0.06** | **0.009** |

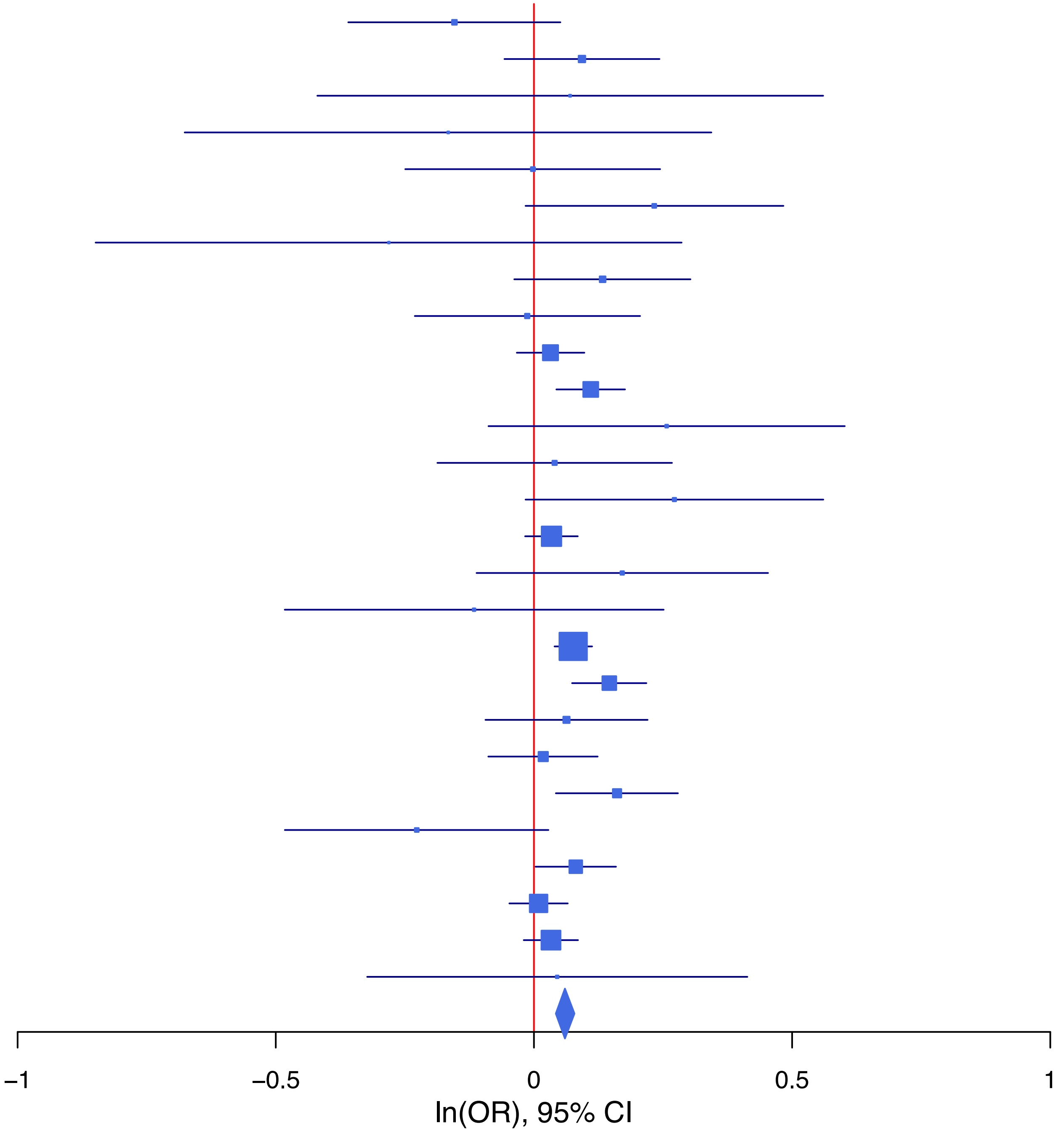

**rs631791 A/G 11:57675410**

| Cohort | P | ln(OR) | SE |
|---|---|---|---|
| ADHEA_EUR | 0.3596 | 0.066 | 0.072 |
| ALSPC_EUR | 0.5726 | −0.033 | 0.058 |
| BEPS7_EUR | 0.8238 | −0.042 | 0.189 |
| BHRCM_EUR | 0.0387 | 0.33 | 0.16 |
| BOR17_EUR | 0.3826 | 0.081 | 0.093 |
| BOR2C_EUR | 0.4181 | 0.075 | 0.093 |
| BOR2E_EUR | 0.9773 | −0.005 | 0.19 |
| COGA1_EUR | 0.6861 | 0.025 | 0.063 |
| CUINT_EUR | 0.6615 | −0.035 | 0.08 |
| ESTB2_EUR | 5e−04 | 0.083 | 0.024 |
| FINNG_EUR | 0.7248 | 0.008 | 0.024 |
| GEDIS_EUR | 0.7787 | 0.039 | 0.14 |
| GISS1_EUR | 0.1541 | 0.12 | 0.084 |
| GISS2_EUR | 0.4854 | 0.07 | 0.1 |
| IPSYC_EUR | 0.0028 | 0.058 | 0.019 |
| JANS3_EUR | 0.5756 | 0.062 | 0.111 |
| JANS4_EUR | 0.181 | 0.166 | 0.124 |
| MVPXQ_EUR | 0.0482 | 0.026 | 0.013 |
| PGCBD_EUR | 0.2287 | 0.033 | 0.027 |
| PGCED_EUR | 0.4579 | −0.097 | 0.13 |
| PGCMD_EUR | 0.3265 | 0.036 | 0.037 |
| PGCPT_EUR | 0.2409 | 0.072 | 0.061 |
| PGCSZ_EUR | 0.8621 | 0.007 | 0.04 |
| PRFCT_EUR | 0.1902 | −0.062 | 0.048 |
| PSYCR_EUR | 0.6578 | 0.043 | 0.097 |
| QIMRB_EUR | 0.0041 | 0.085 | 0.03 |
| UKBJC_EUR | 0.3828 | 0.018 | 0.021 |
| UTAH2_EUR | 0.084 | 0.034 | 0.02 |
| YPENN_EUR | 0.1667 | 0.189 | 0.137 |
| **meta** | **2.96e−08** | **0.036** | **0.007** |

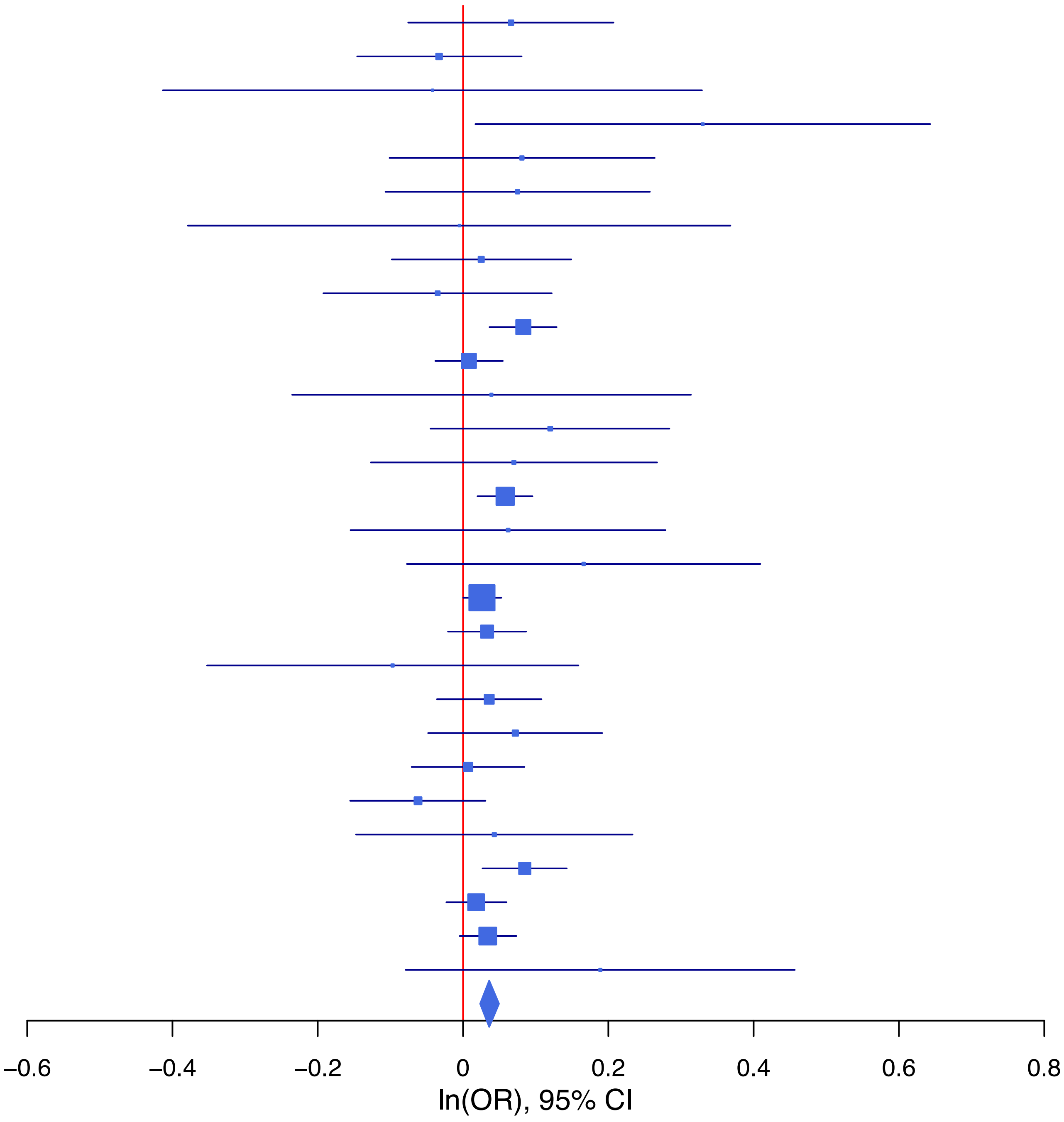

**rs6539788 T/G 12:84226327**

| Cohort | P | ln(OR) | SE |
|---|---|---|---|
| ADHEA_EUR | 0.2718 | −0.075 | 0.068 |
| ALSPC_EUR | 0.0531 | −0.106 | 0.055 |
| BEPS7_EUR | 0.3793 | −0.155 | 0.176 |
| BHRCM_EUR | 0.3214 | 0.15 | 0.152 |
| BOR17_EUR | 0.0508 | −0.17 | 0.087 |
| BOR2C_EUR | 0.4333 | −0.067 | 0.086 |
| BOR2E_EUR | 0.9028 | −0.022 | 0.184 |
| COGA1_EUR | 0.7323 | −0.02 | 0.059 |
| CUINT_EUR | 0.3245 | −0.073 | 0.074 |
| ESTB2_EUR | 0.4791 | −0.016 | 0.023 |
| FINNG_EUR | 0.0239 | −0.051 | 0.023 |
| GEDIS_EUR | 0.9992 | 0 | 0.131 |
| GISS1_EUR | 0.7554 | 0.024 | 0.078 |
| GISS2_EUR | 0.049 | −0.176 | 0.09 |
| IPSYC_EUR | 0.5213 | −0.012 | 0.018 |
| JANS3_EUR | 0.1733 | 0.144 | 0.106 |
| JANS4_EUR | 0.9686 | −0.005 | 0.12 |
| MVPXQ_EUR | 0.0028 | −0.037 | 0.012 |
| PGCBD_EUR | 0.0035 | −0.075 | 0.026 |
| PGCED_EUR | 0.6692 | 0.05 | 0.117 |
| PGCMD_EUR | 0.237 | −0.041 | 0.034 |
| PGCPT_EUR | 0.9824 | 0.001 | 0.058 |
| PGCSZ_EUR | 0.9065 | 0.004 | 0.037 |
| PRFCT_EUR | 0.2789 | −0.047 | 0.043 |
| PSYCR_EUR | 0.5712 | −0.049 | 0.087 |
| QIMRB_EUR | 0.2897 | −0.029 | 0.027 |
| UKBJC_EUR | 0.0078 | −0.052 | 0.02 |
| UTAH2_EUR | 0.1586 | −0.028 | 0.02 |
| YPENN_EUR | 0.088 | −0.225 | 0.132 |
| **meta** | **4.61e−09** | **−0.036** | **0.006** |

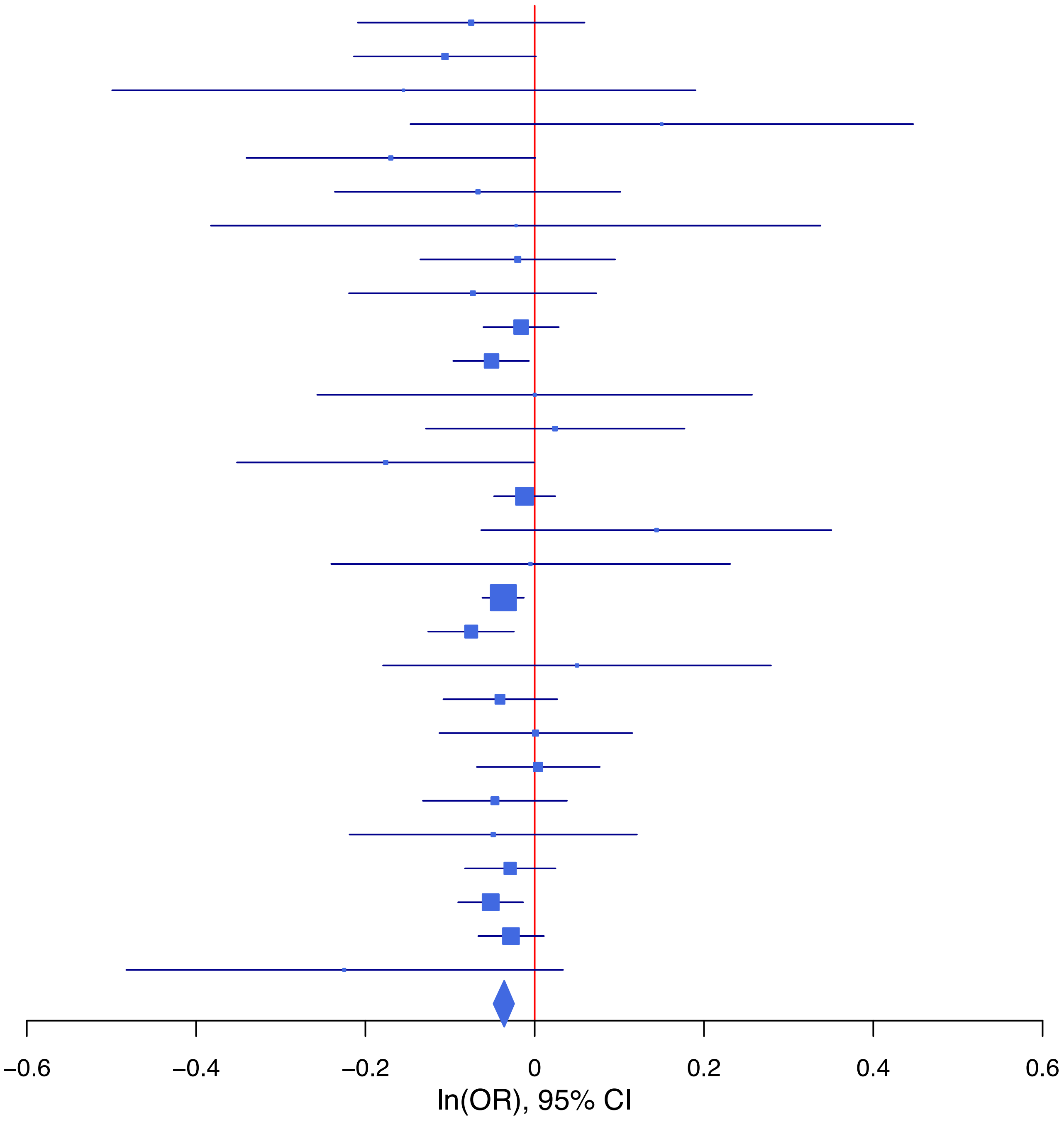

**rs6589377 G/A 11:113355736**

| Cohort | P | ln(OR) | SE |
|---|---|---|---|
| ADHEA_EUR | 0.2158 | −0.088 | 0.071 |
| ALSPC_EUR | 0.6826 | −0.023 | 0.055 |
| BEPS7_EUR | 0.8831 | 0.028 | 0.193 |
| BHRCM_EUR | 0.8435 | −0.032 | 0.163 |
| BOR17_EUR | 0.6963 | 0.034 | 0.086 |
| BOR2C_EUR | 0.3381 | −0.085 | 0.089 |
| BOR2E_EUR | 0.4234 | 0.154 | 0.193 |
| COGA1_EUR | 0.4617 | −0.045 | 0.061 |
| CUINT_EUR | 0.9528 | 0.005 | 0.076 |
| ESTB2_EUR | 0.0072 | −0.069 | 0.026 |
| FINNG_EUR | 0.0399 | −0.058 | 0.028 |
| GEDIS_EUR | 0.13 | −0.21 | 0.139 |
| GISS1_EUR | 0.8354 | −0.017 | 0.083 |
| GISS2_EUR | 0.9638 | 0.004 | 0.094 |
| IPSYC_EUR | 0.0095 | −0.05 | 0.019 |
| JANS3_EUR | 0.0964 | −0.18 | 0.108 |
| JANS4_EUR | 0.1245 | 0.183 | 0.119 |
| MIREC_EUR | 0.0158 | −0.406 | 0.168 |
| MVPXQ_EUR | 2.03e−06 | −0.062 | 0.013 |
| PGCBD_EUR | 0.1923 | −0.035 | 0.027 |
| PGCED_EUR | 0.3179 | −0.116 | 0.116 |
| PGCPT_EUR | 0.9283 | −0.005 | 0.059 |
| PGCSZ_EUR | 0.0784 | −0.068 | 0.038 |
| PRFCT_EUR | 0.6107 | −0.024 | 0.047 |
| PSYCR_EUR | 0.0211 | 0.204 | 0.088 |
| QIMRB_EUR | 0.1029 | −0.046 | 0.028 |
| UKBJC_EUR | 0.2584 | −0.023 | 0.02 |
| UTAH2_EUR | 0.6352 | −0.009 | 0.02 |
| YPENN_EUR | 0.9107 | −0.015 | 0.132 |
| **meta** | **8.69e−11** | **−0.043** | **0.007** |

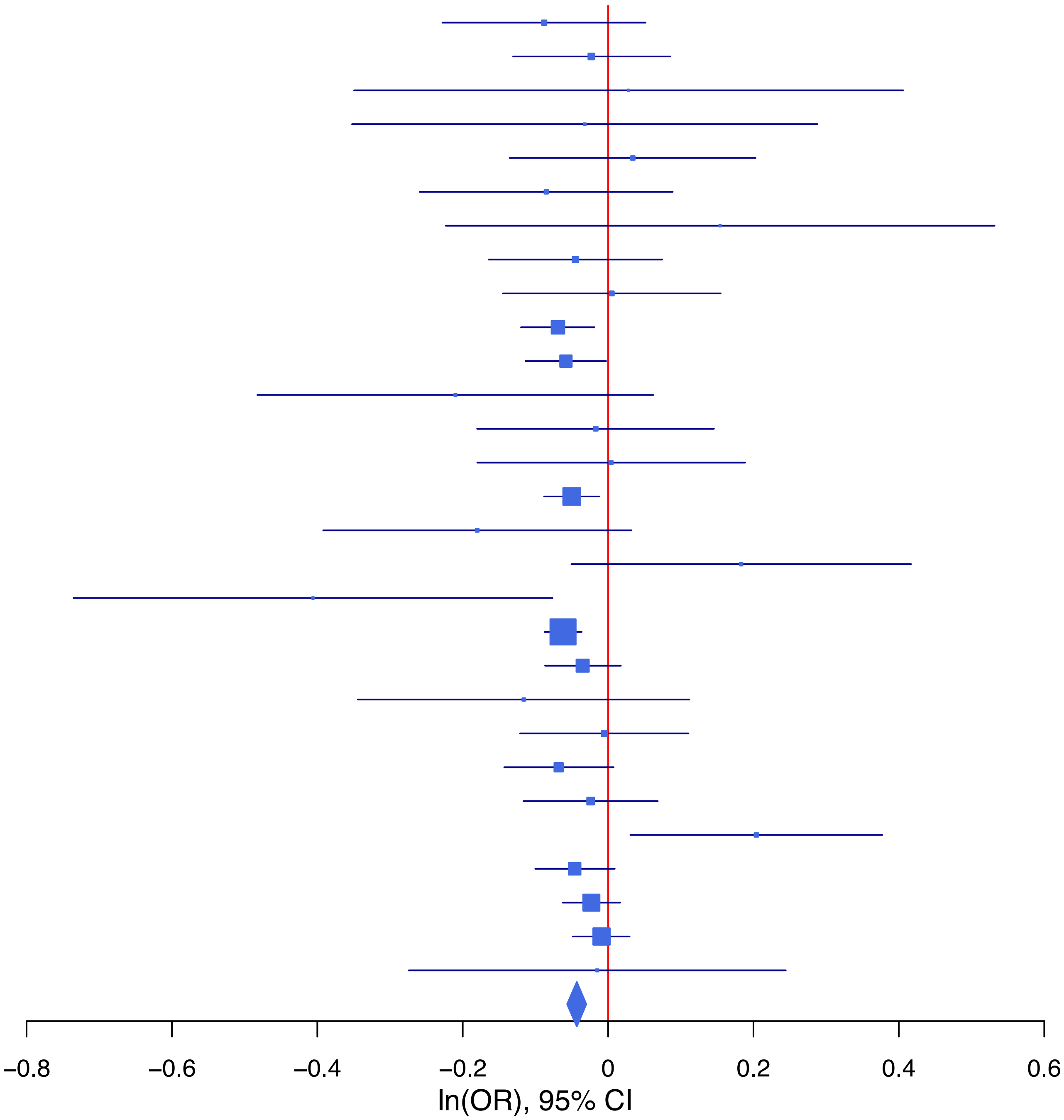

**rs66824958 T/C 4:166061559**

| Cohort | P | ln(OR) | SE |
|---|---|---|---|
| ADHEA_EUR | 0.6198 | −0.051 | 0.103 |
| ALSPC_EUR | 0.5444 | 0.047 | 0.078 |
| BEPS7_EUR | 0.7726 | 0.071 | 0.247 |
| BHRCM_EUR | 0.8864 | −0.041 | 0.284 |
| BOR17_EUR | 0.6452 | −0.061 | 0.133 |
| BOR2C_EUR | 0.0168 | −0.308 | 0.129 |
| BOR2E_EUR | 0.6522 | −0.141 | 0.312 |
| COGA1_EUR | 0.0248 | −0.203 | 0.091 |
| CUINT_EUR | 0.4481 | 0.087 | 0.114 |
| ESTB2_EUR | 0.1504 | −0.047 | 0.032 |
| FINNG_EUR | 0.0212 | −0.068 | 0.03 |
| GEDIS_EUR | 0.8017 | −0.049 | 0.196 |
| GISS1_EUR | 0.128 | −0.179 | 0.117 |
| GISS2_EUR | 0.958 | 0.007 | 0.133 |
| IPSYC_EUR | 0.0626 | −0.05 | 0.027 |
| JANS3_EUR | 0.1136 | −0.265 | 0.168 |
| JANS4_EUR | 0.6917 | 0.076 | 0.19 |
| MVPXQ_EUR | 0.0214 | −0.043 | 0.019 |
| PGCBD_EUR | 0.1373 | −0.059 | 0.04 |
| PGCED_EUR | 0.7918 | 0.05 | 0.189 |
| PGCMD_EUR | 0.761 | −0.016 | 0.053 |
| PGCPT_EUR | 0.6192 | −0.044 | 0.088 |
| PGCSZ_EUR | 0.5867 | −0.03 | 0.056 |
| PRFCT_EUR | 0.0145 | −0.146 | 0.06 |
| PSYCR_EUR | 0.0107 | −0.369 | 0.145 |
| QIMRB_EUR | 0.0907 | −0.068 | 0.04 |
| UKBJC_EUR | 0.1811 | −0.038 | 0.028 |
| YPENN_EUR | 0.6152 | −0.095 | 0.189 |
| **meta** | **2.41e−08** | **−0.054** | **0.01** |

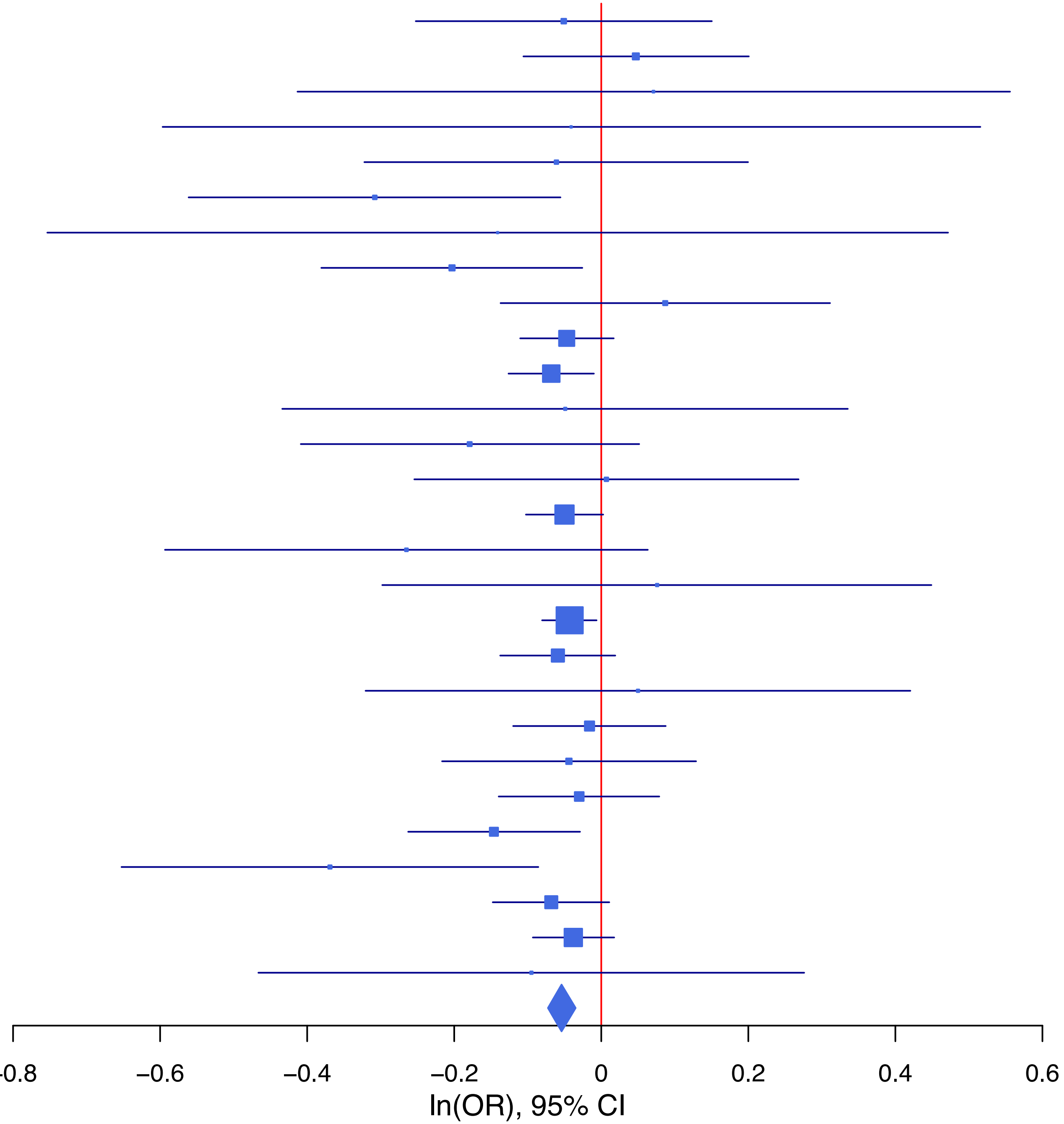

**rs687654 T/C 9:127833905**

| Cohort | P | ln(OR) | SE |
|---|---|---|---|
| ADHEA_EUR | 0.7646 | −0.022 | 0.072 |
| ALSPC_EUR | 0.0063 | −0.161 | 0.059 |
| BEPS7_EUR | 0.1771 | −0.267 | 0.198 |
| BHRCM_EUR | 0.5874 | 0.084 | 0.155 |
| BOR17_EUR | 0.5953 | −0.049 | 0.091 |
| BOR2C_EUR | 0.2374 | −0.11 | 0.094 |
| BOR2E_EUR | 0.9066 | −0.022 | 0.188 |
| COGA1_EUR | 0.023 | −0.144 | 0.063 |
| CUINT_EUR | 0.7867 | −0.021 | 0.079 |
| ESTB2_EUR | 0.2202 | −0.032 | 0.026 |
| FINNG_EUR | 0.4888 | −0.02 | 0.029 |
| GEDIS_EUR | 0.8042 | −0.035 | 0.141 |
| GISS1_EUR | 0.581 | −0.047 | 0.085 |
| GISS2_EUR | 0.963 | −0.005 | 0.099 |
| IPSYC_EUR | 0.085 | −0.034 | 0.02 |
| JANS3_EUR | 0.1476 | −0.169 | 0.117 |
| JANS4_EUR | 0.7399 | −0.041 | 0.122 |
| MIREC_EUR | 0.0221 | 0.369 | 0.161 |
| MVPXQ_EUR | 4.20e−06 | −0.061 | 0.013 |
| PGCBD_EUR | 0.1417 | −0.041 | 0.028 |
| PGCED_EUR | 0.793 | 0.032 | 0.123 |
| PGCMD_EUR | 0.2178 | −0.045 | 0.037 |
| PGCPT_EUR | 0.9065 | 0.007 | 0.061 |
| PGCSZ_EUR | 0.9877 | 0.001 | 0.04 |
| PRFCT_EUR | 0.9237 | 0.005 | 0.048 |
| PSYCR_EUR | 0.3911 | −0.08 | 0.093 |
| QIMRB_EUR | 0.3146 | −0.029 | 0.029 |
| UKBJC_EUR | 0.8297 | −0.005 | 0.021 |
| UTAH2_EUR | 0.0084 | −0.054 | 0.02 |
| YPENN_EUR | 0.8586 | −0.024 | 0.136 |
| **meta** | **1.90e−09** | **−0.041** | **0.007** |

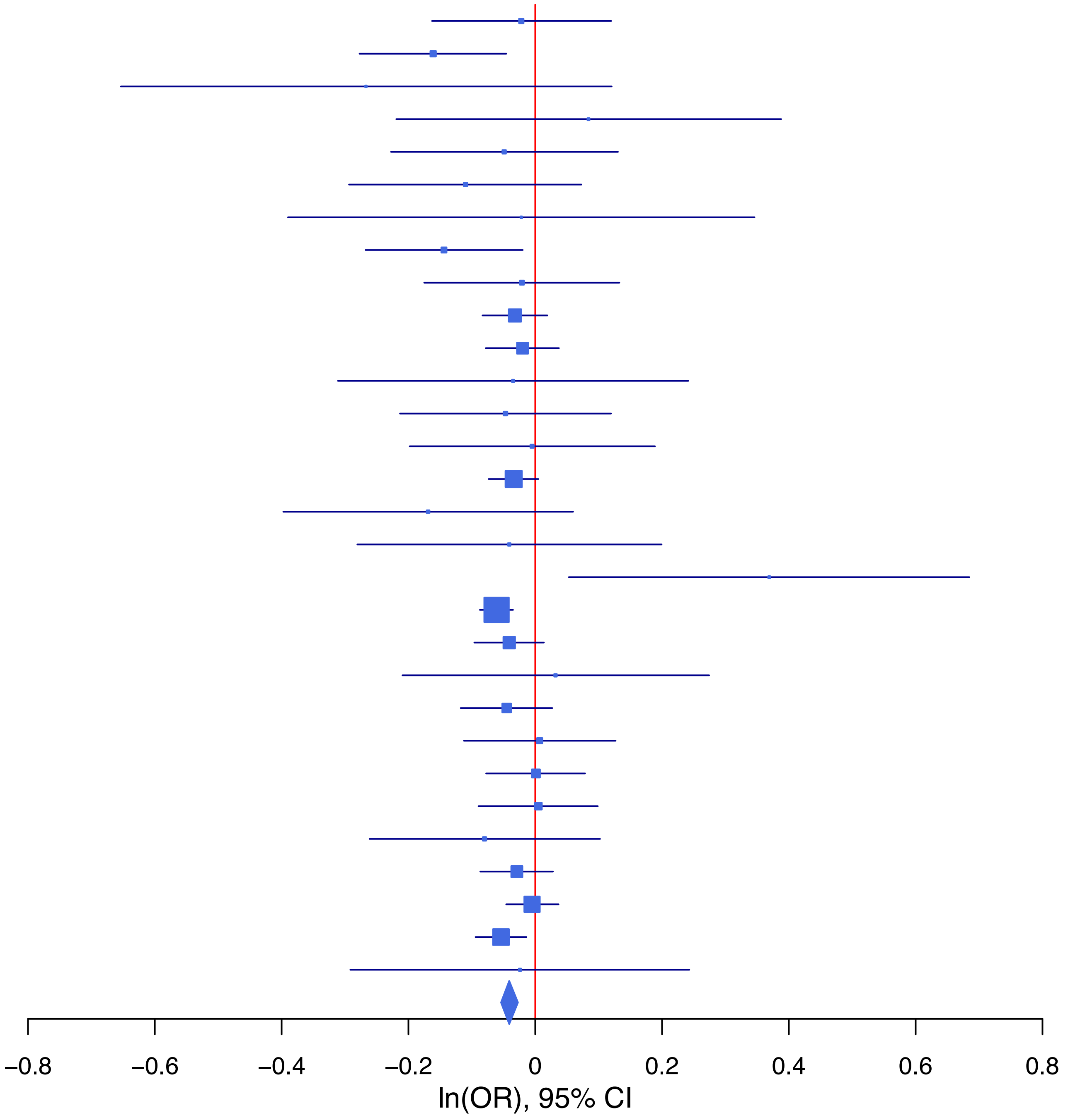

**rs6959688   G/A   7:1966831**

| Cohort | P | ln(OR) | SE |
|---|---|---|---|
| ADHEA_EUR | 0.3461 | 0.065 | 0.069 |
| ALSPC_EUR | 0.3065 | 0.057 | 0.056 |
| BEPS7_EUR | 0.0642 | 0.339 | 0.183 |
| BHRCM_EUR | 0.9133 | −0.018 | 0.166 |
| BOR17_EUR | 0.5783 | −0.048 | 0.087 |
| BOR2C_EUR | 0.0097 | 0.222 | 0.086 |
| BOR2E_EUR | 0.1128 | −0.298 | 0.188 |
| COGA1_EUR | 0.1911 | 0.079 | 0.06 |
| ESTB2_EUR | 0.9879 | 0 | 0.023 |
| FINNG_EUR | 0.0575 | 0.045 | 0.023 |
| GEDIS_EUR | 0.0838 | 0.221 | 0.128 |
| GISS1_EUR | 0.9665 | 0.004 | 0.084 |
| GISS2_EUR | 0.8088 | 0.023 | 0.095 |
| IPSYC_EUR | 0.0154 | 0.045 | 0.019 |
| JANS3_EUR | 0.2813 | 0.114 | 0.106 |
| JANS4_EUR | 0.1698 | 0.162 | 0.118 |
| MVPXQ_EUR | 0.0079 | 0.035 | 0.013 |
| PGCBD_EUR | 0.2113 | 0.033 | 0.026 |
| PGCED_EUR | 0.4801 | −0.085 | 0.12 |
| PGCMD_EUR | 0.1072 | −0.057 | 0.035 |
| PGCPT_EUR | 0.332 | 0.056 | 0.058 |
| PGCSZ_EUR | 0.6581 | 0.017 | 0.038 |
| PRFCT_EUR | 0.82 | −0.01 | 0.045 |
| PSYCR_EUR | 0.3533 | 0.079 | 0.085 |
| QIMRB_EUR | 0.0542 | 0.053 | 0.027 |
| UKBJC_EUR | 0.0022 | 0.061 | 0.02 |
| UTAH2_EUR | 0.1474 | 0.028 | 0.019 |
| YPENN_EUR | 0.4516 | 0.096 | 0.128 |
| **meta** | **3.33e−08** | **0.035** | **0.006** |

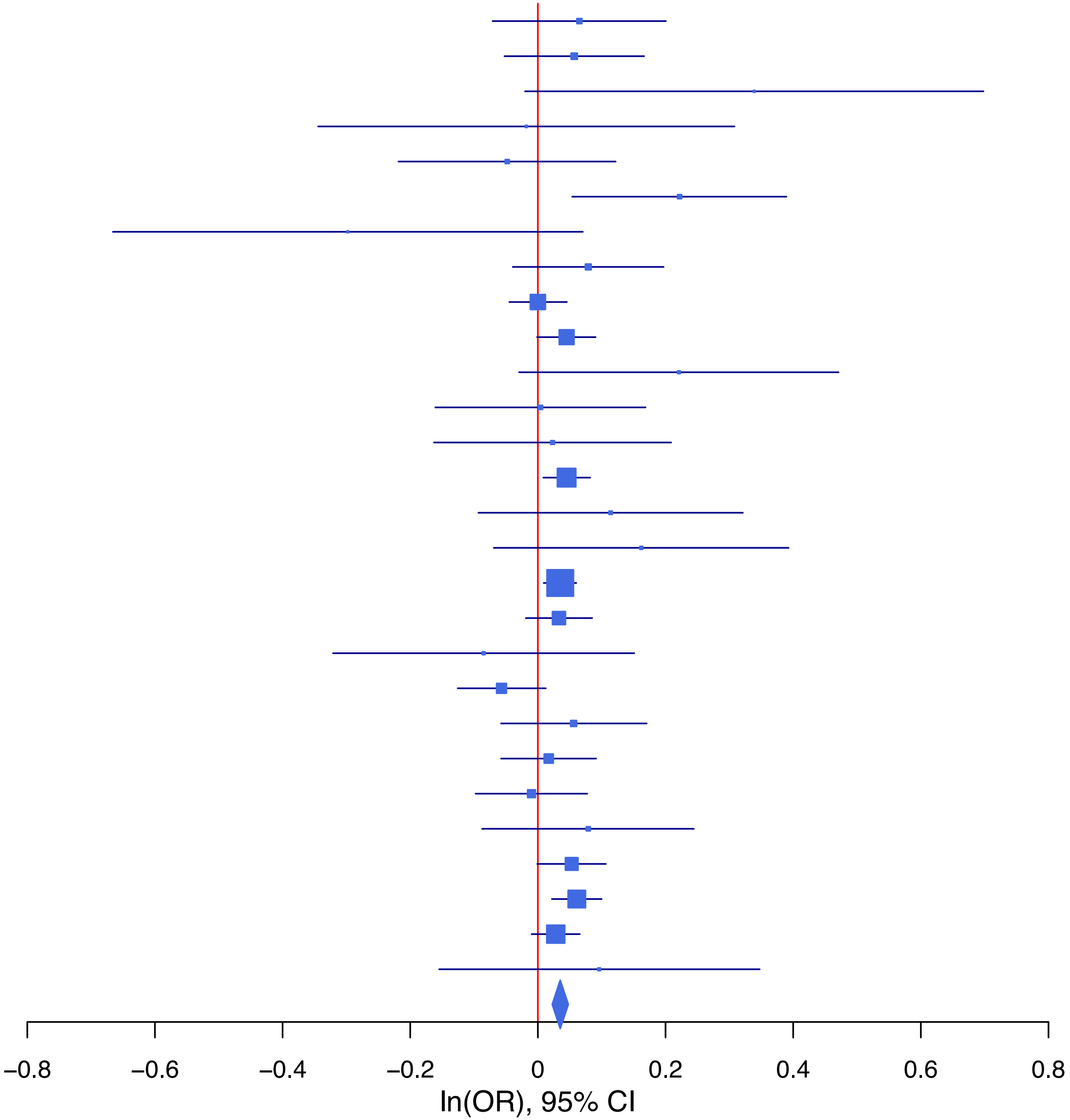

**rs7152530 A/G 14:98641215**

| Cohort | P | ln(OR) | SE |
|---|---|---|---|
| ADHEA_EUR | 0.7157 | 0.026 | 0.07 |
| ALSPC_EUR | 0.2087 | 0.071 | 0.056 |
| BEPS7_EUR | 0.8337 | 0.039 | 0.185 |
| BHRCM_EUR | 0.0501 | 0.305 | 0.156 |
| BOR17_EUR | 0.9241 | −0.008 | 0.086 |
| BOR2C_EUR | 0.9216 | 0.009 | 0.089 |
| BOR2E_EUR | 0.2956 | 0.185 | 0.177 |
| COGA1_EUR | 0.0639 | 0.115 | 0.062 |
| CUINT_EUR | 0.3419 | 0.072 | 0.076 |
| ESTB2_EUR | 0.5984 | 0.013 | 0.025 |
| FINNG_EUR | 0.0244 | 0.06 | 0.027 |
| GEDIS_EUR | 0.8335 | 0.028 | 0.131 |
| GISS1_EUR | 0.5347 | −0.051 | 0.083 |
| GISS2_EUR | 0.5199 | −0.062 | 0.097 |
| IPSYC_EUR | 0.268 | 0.022 | 0.02 |
| JANS3_EUR | 0.6918 | 0.042 | 0.106 |
| JANS4_EUR | 0.1798 | 0.162 | 0.121 |
| MIREC_EUR | 0.1537 | 0.222 | 0.156 |
| MVPXQ_EUR | 0.0822 | 0.023 | 0.013 |
| PGCBD_EUR | 0.0148 | 0.065 | 0.027 |
| PGCED_EUR | 0.0928 | 0.192 | 0.114 |
| PGCMD_EUR | 0.1864 | 0.047 | 0.035 |
| PGCPT_EUR | 0.4516 | 0.045 | 0.059 |
| PGCSZ_EUR | 0.5578 | 0.023 | 0.038 |
| PRFCT_EUR | 0.0905 | 0.079 | 0.046 |
| PSYCR_EUR | 0.404 | −0.076 | 0.091 |
| QIMRB_EUR | 0.031 | 0.061 | 0.028 |
| UKBJC_EUR | 0.0035 | 0.06 | 0.02 |
| UTAH2_EUR | 0.0195 | 0.047 | 0.02 |
| YPENN_EUR | 0.5294 | −0.083 | 0.132 |
| **meta** | **1.24e−09** | **0.04** | **0.007** |

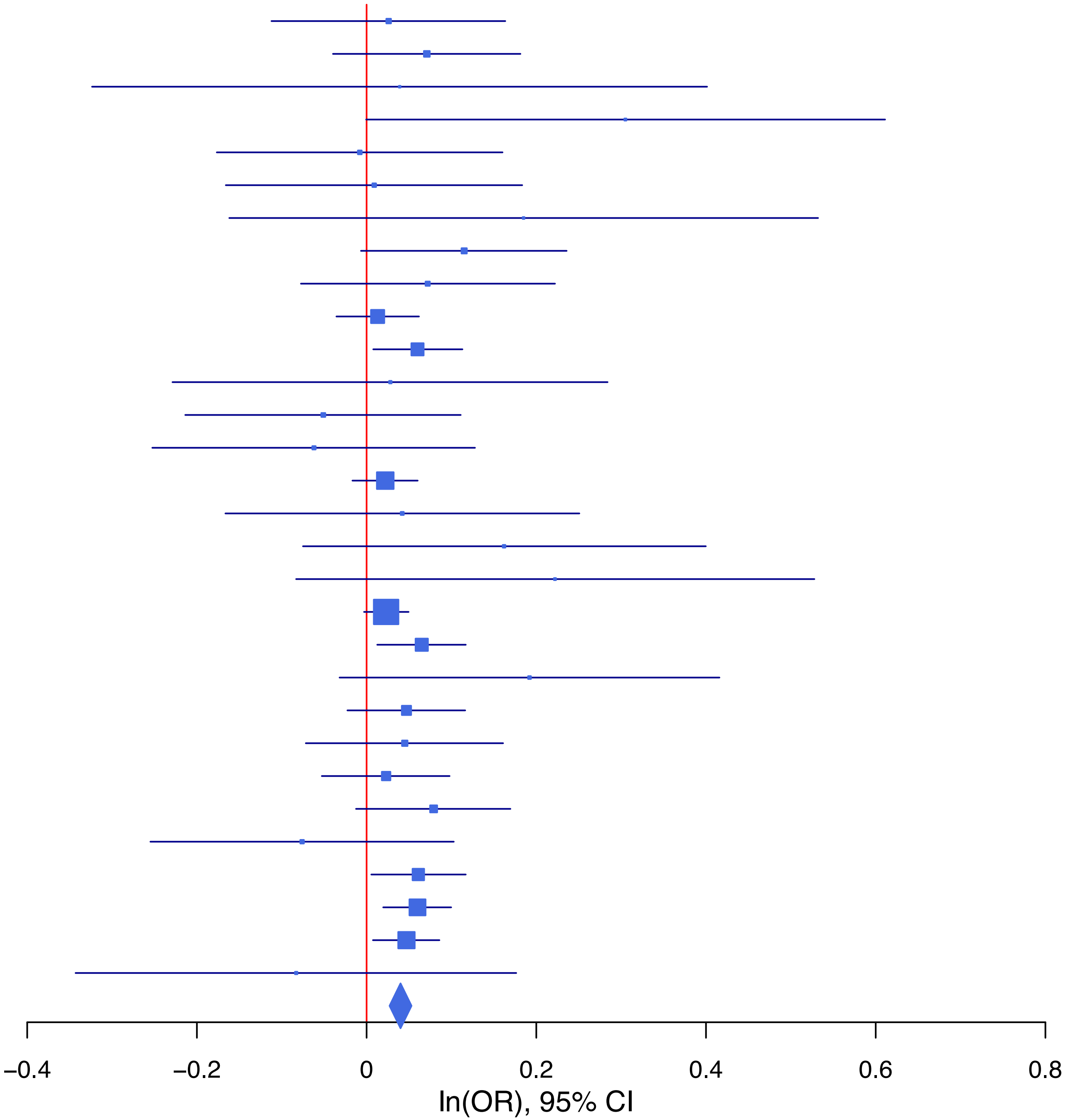

**rs72925321 A/G 18:50807202**

| Cohort | P | ln(OR) | SE |
|---|---|---|---|
| ADHEA_EUR | 0.9271 | −0.014 | 0.157 |
| ALSPC_EUR | 0.8505 | −0.021 | 0.114 |
| BHRCM_EUR | 0.0587 | 0.671 | 0.355 |
| BOR17_EUR | 0.281 | −0.197 | 0.183 |
| BOR2C_EUR | 0.9487 | 0.012 | 0.185 |
| BOR2E_EUR | 0.2693 | −0.45 | 0.407 |
| COGA1_EUR | 0.6371 | 0.06 | 0.128 |
| CUINT_EUR | 0.9387 | 0.014 | 0.184 |
| ESTB2_EUR | 0.0526 | −0.084 | 0.043 |
| FINNG_EUR | 0.1095 | −0.066 | 0.041 |
| GEDIS_EUR | 0.8241 | −0.063 | 0.282 |
| GISS1_EUR | 0.4395 | −0.109 | 0.141 |
| GISS2_EUR | 0.4162 | 0.143 | 0.176 |
| IPSYC_EUR | 0.031 | −0.09 | 0.042 |
| JANS3_EUR | 0.8643 | −0.041 | 0.241 |
| JANS4_EUR | 0.5307 | −0.171 | 0.273 |
| MVPXQ_EUR | 9e−04 | −0.108 | 0.032 |
| PGCBD_EUR | 0.0437 | −0.134 | 0.067 |
| PGCMD_EUR | 0.073 | −0.164 | 0.091 |
| PGCPT_EUR | 0.6592 | −0.056 | 0.127 |
| PGCSZ_EUR | 0.0518 | −0.182 | 0.094 |
| PRFCT_EUR | 0.7534 | −0.031 | 0.098 |
| PSYCR_EUR | 0.4729 | −0.138 | 0.192 |
| QIMRB_EUR | 0.0468 | −0.116 | 0.058 |
| UKBJC_EUR | 0.243 | −0.049 | 0.042 |
| UTAH2_EUR | 0.2674 | −0.043 | 0.039 |
| YPENN_EUR | 0.7813 | −0.081 | 0.292 |
| **meta** | **9.39e−09** | **−0.079** | **0.014** |

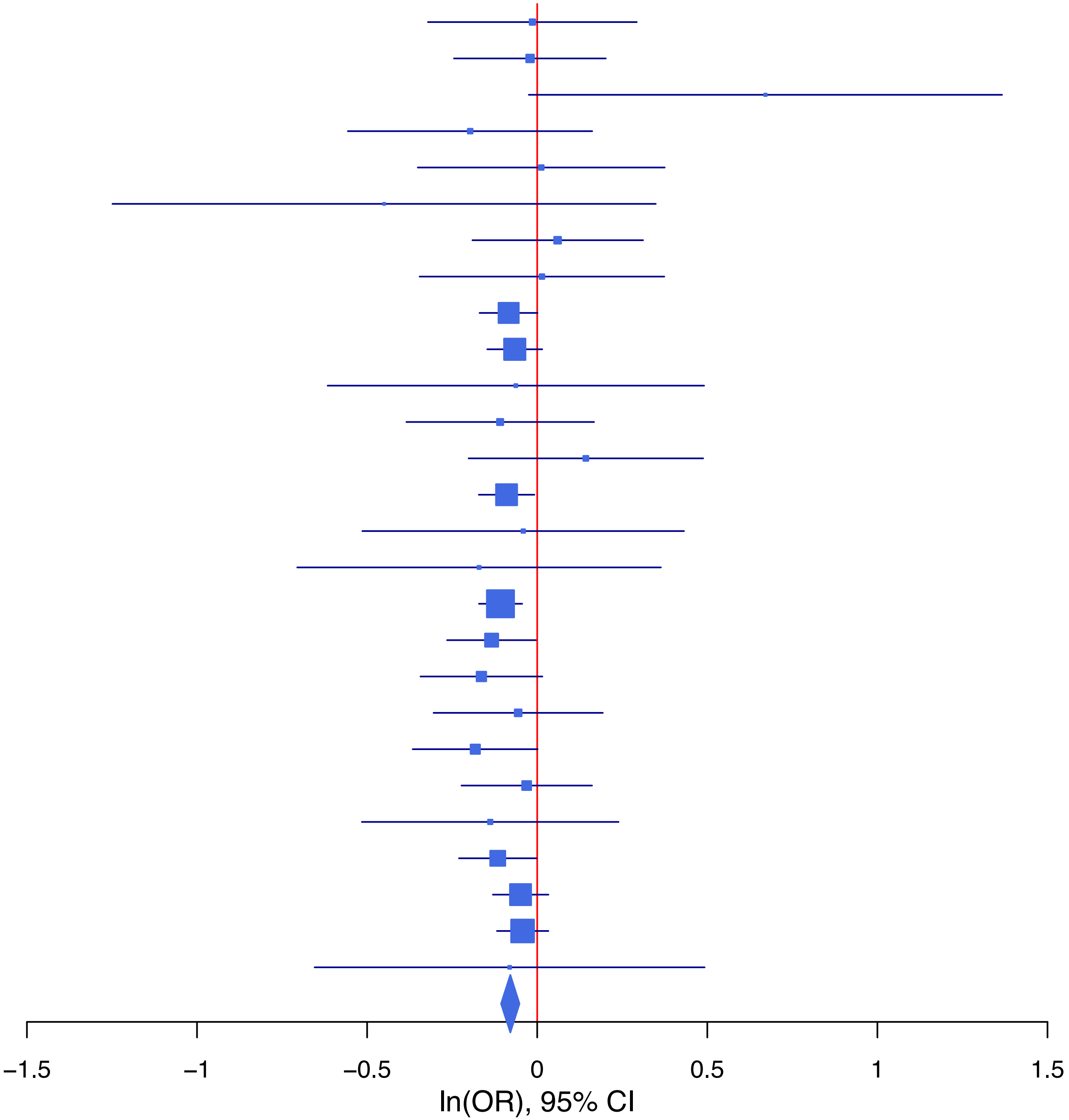

**rs75633418   G/C   9:122663012**

| Cohort | P | ln(OR) | SE |
|---|---|---|---|
| ADHEA_EUR | 0.2717 | −0.078 | 0.071 |
| ALSPC_EUR | 0.646 | −0.025 | 0.055 |
| BEPS7_EUR | 0.0244 | −0.434 | 0.193 |
| BOR17_EUR | 0.4426 | 0.069 | 0.089 |
| BOR2C_EUR | 0.7596 | 0.027 | 0.088 |
| BOR2E_EUR | 0.2531 | 0.205 | 0.179 |
| COGA1_EUR | 0.2261 | −0.072 | 0.059 |
| CUINT_EUR | 0.547 | −0.045 | 0.074 |
| ESTB2_EUR | 0.0697 | −0.043 | 0.023 |
| FINNG_EUR | 0.0237 | −0.053 | 0.024 |
| GEDIS_EUR | 0.5318 | 0.083 | 0.134 |
| GISS1_EUR | 0.4948 | −0.055 | 0.08 |
| GISS2_EUR | 0.7291 | 0.034 | 0.099 |
| IPSYC_EUR | 2e−04 | −0.071 | 0.019 |
| JANS3_EUR | 0.1583 | −0.156 | 0.11 |
| JANS4_EUR | 0.9212 | −0.012 | 0.123 |
| MVPXQ_EUR | 0.0127 | −0.032 | 0.013 |
| PGCBD_EUR | 0.0495 | −0.052 | 0.027 |
| PGCED_EUR | 0.8448 | −0.023 | 0.119 |
| PGCMD_EUR | 0.0569 | −0.067 | 0.035 |
| PGCPT_EUR | 0.3737 | −0.053 | 0.059 |
| PGCSZ_EUR | 0.9885 | 0 | 0.038 |
| PRFCT_EUR | 0.4157 | −0.037 | 0.045 |
| PSYCR_EUR | 0.9139 | 0.01 | 0.09 |
| QIMRB_EUR | 0.5218 | −0.018 | 0.028 |
| UKBJC_EUR | 0.0767 | −0.035 | 0.02 |
| UTAH2_EUR | 0.9221 | −0.002 | 0.02 |
| YPENN_EUR | 0.7885 | −0.035 | 0.131 |
| **meta** | **8.73e−09** | **−0.037** | **0.006** |

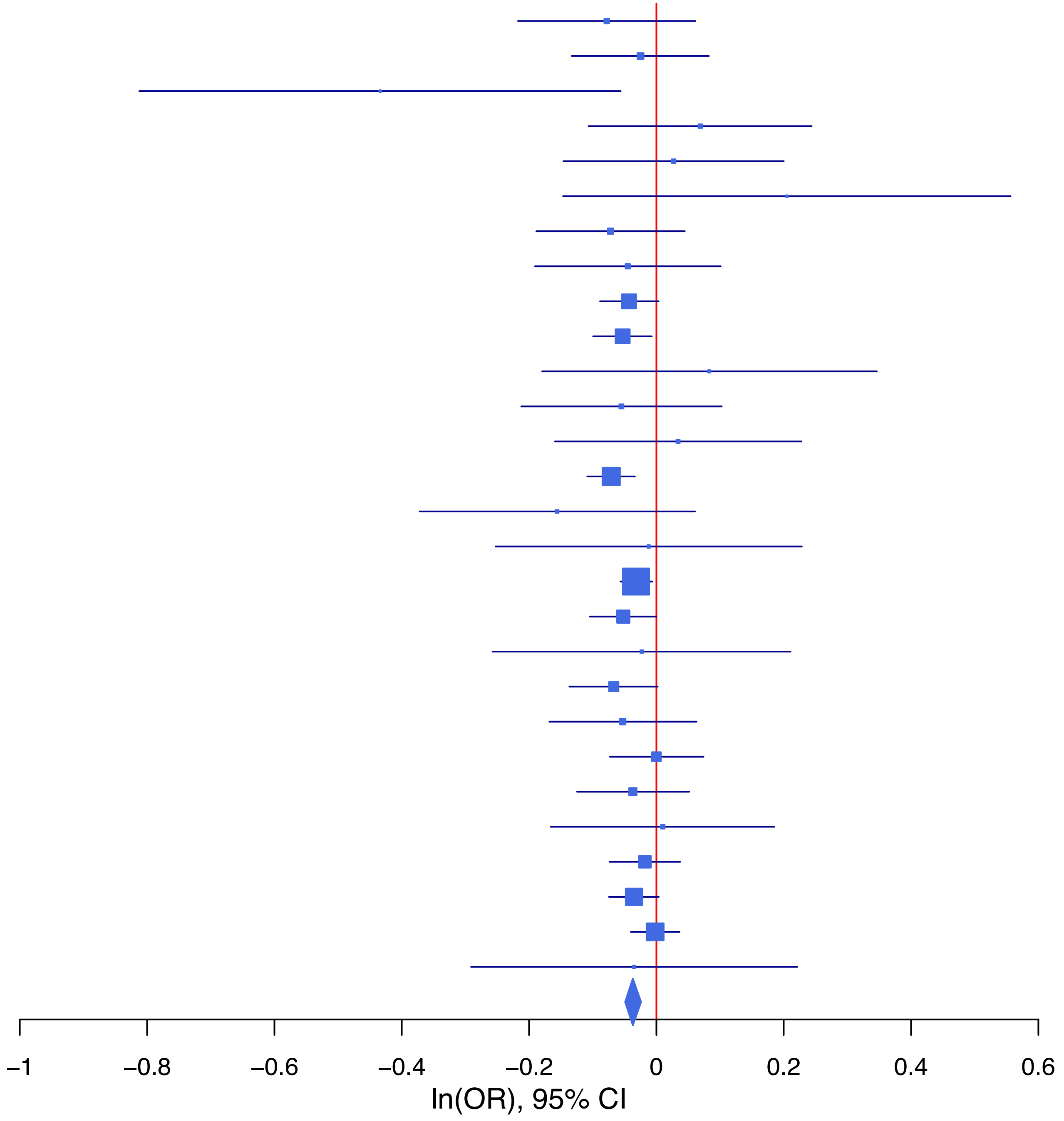

## rs78470849 T/C 6:31382675

| Cohort | P | ln(OR) | SE |
|---|---|---|---|
| ADHEA_EUR | 0.2204 | 0.107 | 0.087 |
| ALSPC_EUR | 0.7815 | 0.021 | 0.075 |
| BEPS7_EUR | 0.2714 | −0.252 | 0.229 |
| BOR17_EUR | 0.3991 | −0.09 | 0.107 |
| BOR2C_EUR | 0.926 | 0.01 | 0.111 |
| BOR2E_EUR | 0.3763 | 0.195 | 0.221 |
| COGA1_EUR | 0.5488 | 0.047 | 0.078 |
| CUINT_EUR | 0.9356 | −0.008 | 0.095 |
| ESTB2_EUR | 0.0692 | 0.051 | 0.028 |
| FINNG_EUR | 0.0755 | 0.047 | 0.027 |
| GISS2_EUR | 0.7858 | 0.032 | 0.119 |
| IPSYC_EUR | 1e−04 | 0.096 | 0.024 |
| JANS3_EUR | 0.353 | −0.131 | 0.142 |
| PGCBD_EUR | 0.0754 | 0.06 | 0.034 |
| PGCED_EUR | 0.9866 | −0.003 | 0.176 |
| PGCMD_EUR | 0.9022 | 0.006 | 0.047 |
| PGCPT_EUR | 0.1022 | 0.121 | 0.074 |
| PGCSZ_EUR | 0.2476 | 0.057 | 0.049 |
| PRFCT_EUR | 0.7123 | 0.022 | 0.059 |
| PSYCR_EUR | 0.7677 | 0.032 | 0.109 |
| QIMRB_EUR | 0.01 | 0.095 | 0.037 |
| UKBJC_EUR | 0.0898 | 0.046 | 0.027 |
| YPENN_EUR | 0.6328 | −0.08 | 0.167 |
| **meta** | **3.03e−08** | **0.056** | **0.01** |

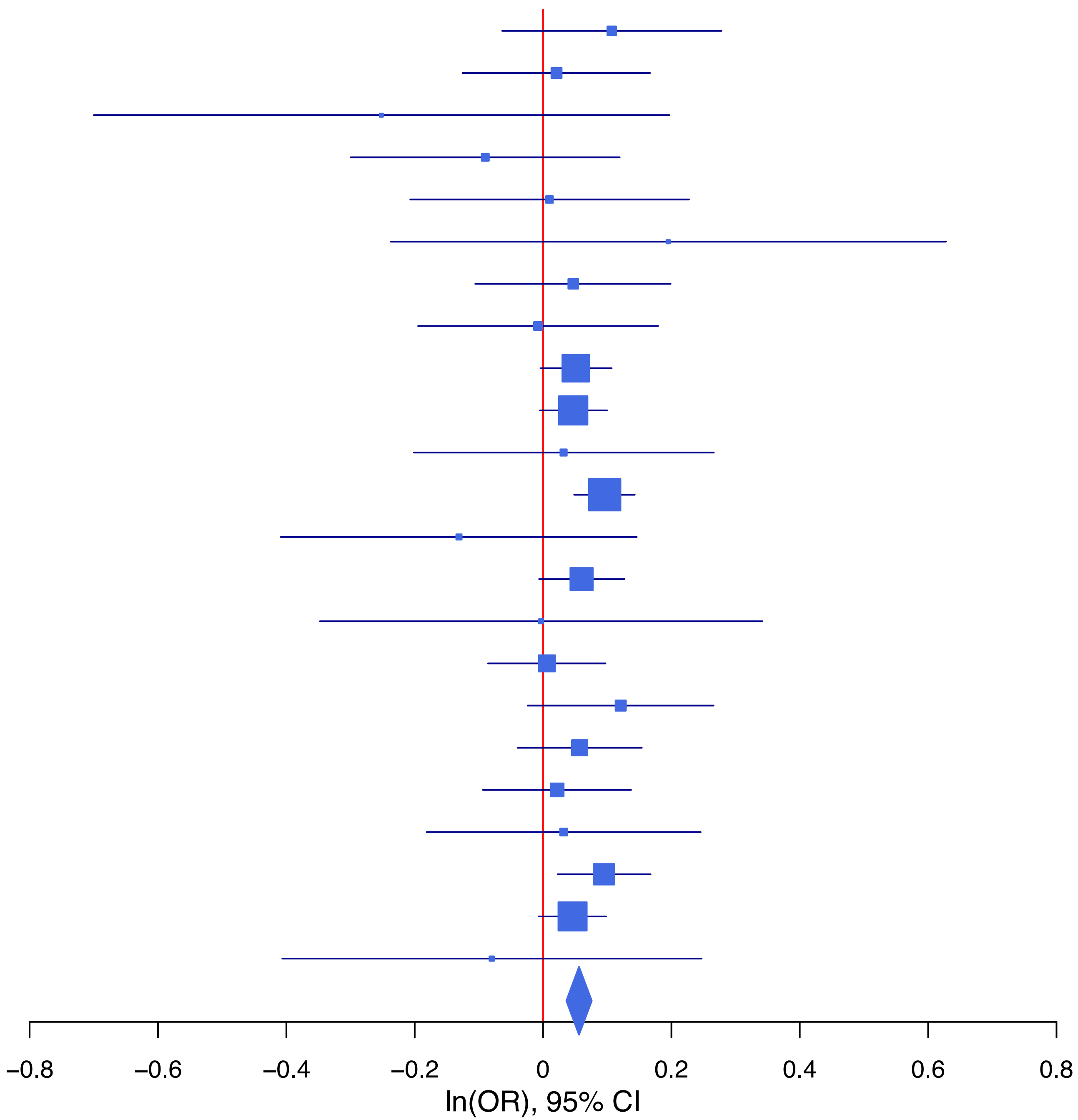

**rs78940908 G/C 2:58921049**

| Cohort | P | ln(OR) | SE |
|---|---|---|---|
| ADHEA_EUR | 0.0089 | −0.183 | 0.07 |
| BEPS7_EUR | 0.9368 | −0.015 | 0.185 |
| BOR17_EUR | 0.5683 | −0.05 | 0.087 |
| BOR2C_EUR | 0.565 | −0.05 | 0.087 |
| BOR2E_EUR | 0.3827 | −0.156 | 0.179 |
| COGA1_EUR | 0.5856 | −0.032 | 0.059 |
| CUINT_EUR | 0.4746 | −0.055 | 0.077 |
| ESTB2_EUR | 0.2309 | −0.028 | 0.023 |
| FINNG_EUR | 0.7245 | 0.008 | 0.024 |
| GEDIS_EUR | 0.5801 | 0.071 | 0.129 |
| GISS1_EUR | 0.6598 | 0.035 | 0.08 |
| GISS2_EUR | 0.4138 | −0.076 | 0.093 |
| IPSYC_EUR | 0.0131 | −0.046 | 0.019 |
| JANS3_EUR | 0.7527 | 0.034 | 0.106 |
| JANS4_EUR | 0.9779 | −0.003 | 0.118 |
| MVPXQ_EUR | 0.0026 | −0.039 | 0.013 |
| PGCBD_EUR | 0.239 | −0.031 | 0.026 |
| PGCED_EUR | 0.4613 | 0.083 | 0.113 |
| PGCMD_EUR | 0.5107 | −0.023 | 0.035 |
| PGCPT_EUR | 0.264 | −0.065 | 0.058 |
| PGCSZ_EUR | 0.4735 | −0.027 | 0.037 |
| PRFCT_EUR | 0.4512 | −0.033 | 0.044 |
| PSYCR_EUR | 0.7609 | −0.026 | 0.087 |
| QIMRB_EUR | 0.0193 | −0.064 | 0.028 |
| UKBJC_EUR | 0.0047 | −0.056 | 0.02 |
| YPENN_EUR | 0.4275 | 0.103 | 0.13 |
| **meta** | **4.54e−08** | **−0.037** | **0.007** |

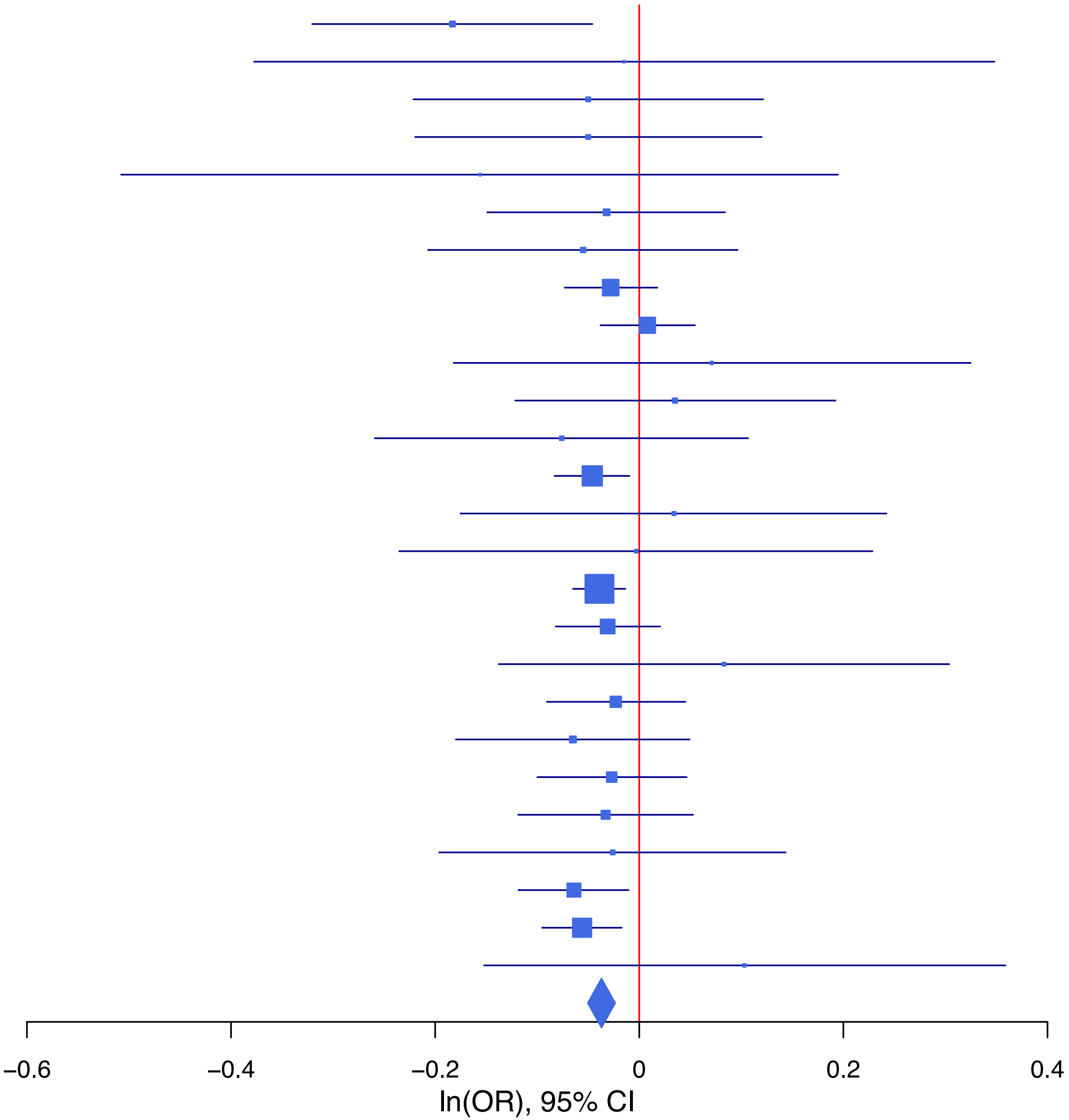

**rs7931884 C/G 11:28648378**

| Cohort | P | ln(OR) | SE |
|---|---|---|---|
| ADHEA_EUR | 0.1175 | −0.113 | 0.072 |
| ALSPC_EUR | 0.5242 | −0.036 | 0.057 |
| BEPS7_EUR | 0.3974 | 0.151 | 0.178 |
| BHRCM_EUR | 0.0714 | −0.331 | 0.183 |
| BOR17_EUR | 0.9304 | 0.008 | 0.09 |
| BOR2C_EUR | 0.3218 | −0.088 | 0.089 |
| BOR2E_EUR | 0.0125 | −0.5 | 0.2 |
| COGA1_EUR | 0.646 | −0.028 | 0.062 |
| CUINT_EUR | 0.1789 | −0.11 | 0.082 |
| ESTB2_EUR | 0.2309 | −0.028 | 0.023 |
| FINNG_EUR | 0.4487 | 0.018 | 0.024 |
| GEDIS_EUR | 0.8074 | −0.034 | 0.14 |
| GISS2_EUR | 0.7167 | −0.034 | 0.094 |
| IPSYC_EUR | 0.4939 | −0.013 | 0.02 |
| JANS3_EUR | 0.5291 | −0.069 | 0.11 |
| JANS4_EUR | 0.2418 | −0.145 | 0.124 |
| MVPXQ_EUR | 4.76e−05 | −0.056 | 0.014 |
| PGCBD_EUR | 0.1114 | −0.043 | 0.027 |
| PGCED_EUR | 0.3102 | −0.124 | 0.123 |
| PGCMD_EUR | 0.0544 | −0.069 | 0.036 |
| PGCPT_EUR | 0.4159 | 0.049 | 0.06 |
| PGCSZ_EUR | 0.1059 | −0.063 | 0.039 |
| PRFCT_EUR | 0.2797 | −0.05 | 0.047 |
| PSYCR_EUR | 0.0641 | −0.172 | 0.093 |
| QIMRB_EUR | 0.0358 | −0.06 | 0.028 |
| UKBJC_EUR | 0.0319 | −0.044 | 0.021 |
| UTAH2_EUR | 0.0698 | −0.036 | 0.02 |
| YPENN_EUR | 0.7575 | −0.041 | 0.133 |
| **meta** | **1.26e−09** | **−0.04** | **0.007** |

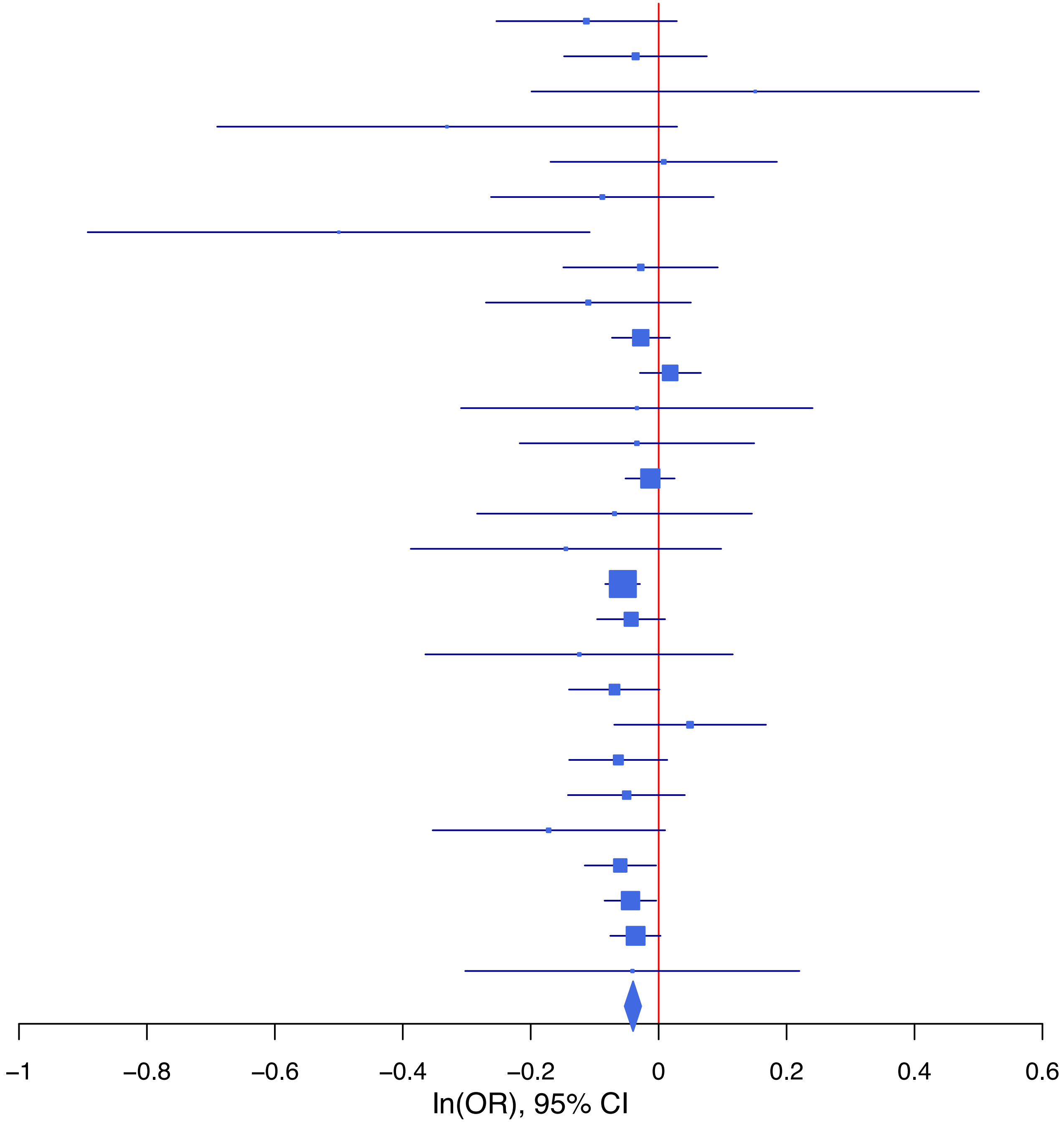

**rs8061310 C/A 16:79319475**

| Cohort | P | ln(OR) | SE |
|---|---|---|---|
| ALSPC_EUR | 0.2204 | 0.083 | 0.068 |
| BEPS7_EUR | 0.9056 | 0.026 | 0.222 |
| BHRCM_EUR | 0.3785 | 0.16 | 0.181 |
| BOR17_EUR | 0.0151 | 0.257 | 0.106 |
| BOR2C_EUR | 0.4975 | 0.072 | 0.106 |
| BOR2E_EUR | 0.9702 | −0.009 | 0.231 |
| COGA1_EUR | 0.5882 | 0.04 | 0.074 |
| CUINT_EUR | 0.0163 | 0.211 | 0.088 |
| ESTB2_EUR | 0.0199 | 0.061 | 0.026 |
| FINNG_EUR | 0.2342 | 0.035 | 0.029 |
| GEDIS_EUR | 0.828 | −0.036 | 0.166 |
| GISS1_EUR | 0.4868 | 0.065 | 0.093 |
| GISS2_EUR | 0.8133 | −0.027 | 0.114 |
| IPSYC_EUR | 0.1426 | 0.034 | 0.023 |
| JANS3_EUR | 0.6239 | 0.062 | 0.127 |
| JANS4_EUR | 0.657 | 0.065 | 0.146 |
| MVPXQ_EUR | 0.0132 | 0.04 | 0.016 |
| PGCBD_EUR | 0.5293 | 0.02 | 0.032 |
| PGCED_EUR | 0.1984 | −0.197 | 0.154 |
| PGCMD_EUR | 0.016 | 0.102 | 0.042 |
| PGCPT_EUR | 0.6894 | 0.028 | 0.071 |
| PGCSZ_EUR | 0.257 | 0.053 | 0.046 |
| PRFCT_EUR | 0.0714 | 0.1 | 0.056 |
| PSYCR_EUR | 0.7059 | 0.039 | 0.103 |
| QIMRB_EUR | 0.874 | 0.005 | 0.034 |
| UKBJC_EUR | 0.17 | 0.034 | 0.025 |
| UTAH2_EUR | 0.0659 | 0.043 | 0.023 |
| YPENN_EUR | 0.6427 | 0.074 | 0.159 |
| **meta** | **1.96e−08** | **0.044** | **0.008** |

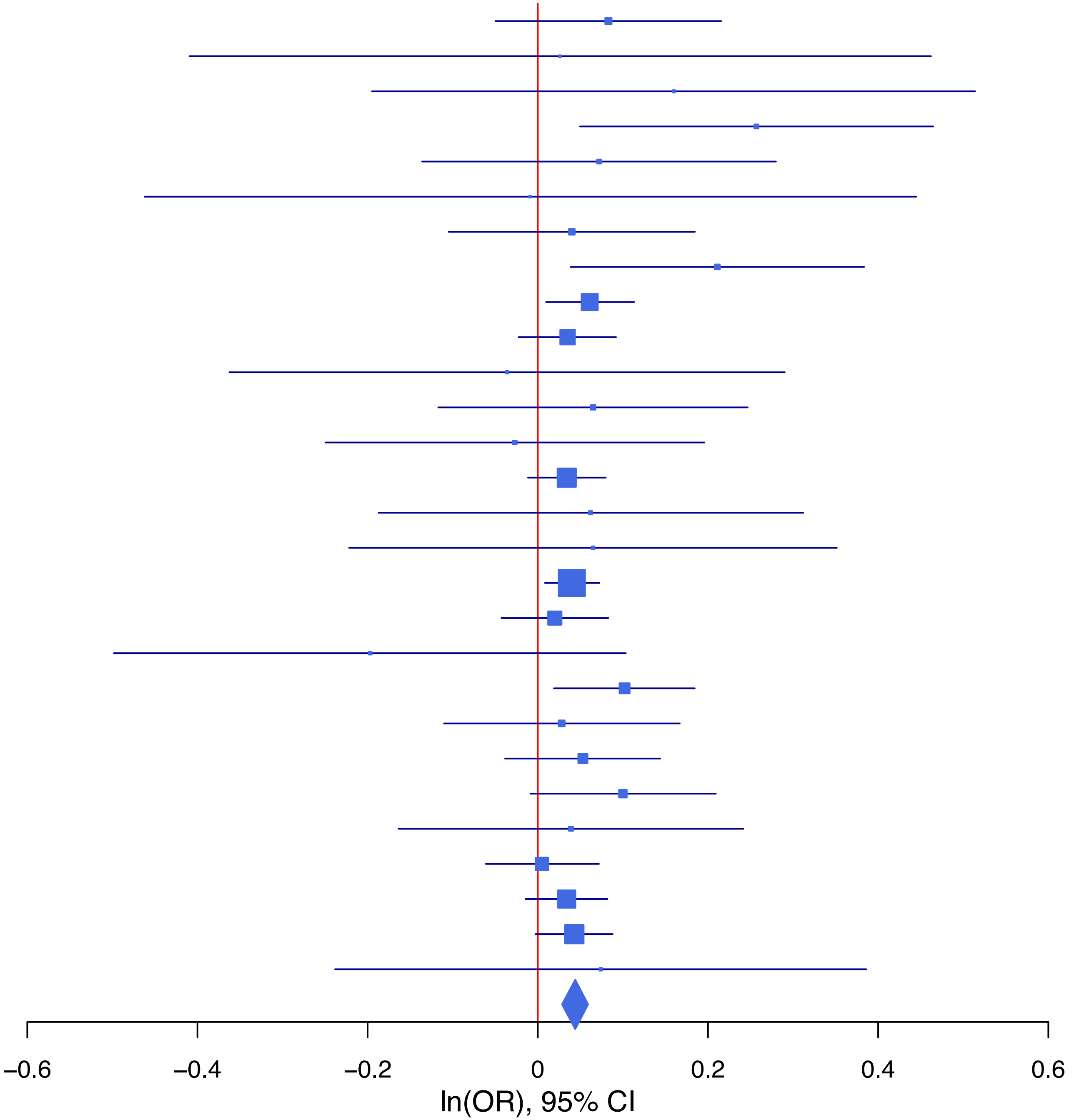

**rs8072008 C/G 17:27407775**

| Cohort | P | ln(OR) | SE |
|---|---|---|---|
| ALSPC_EUR | 0.66 | 0.025 | 0.056 |
| BEPS7_EUR | 0.9013 | 0.024 | 0.194 |
| BHRCM_EUR | 0.6112 | 0.081 | 0.158 |
| BOR17_EUR | 0.4493 | 0.069 | 0.091 |
| BOR2C_EUR | 0.4562 | 0.069 | 0.092 |
| BOR2E_EUR | 0.6289 | −0.091 | 0.189 |
| COGA1_EUR | 0.2812 | 0.065 | 0.061 |
| CUINT_EUR | 0.0798 | 0.138 | 0.079 |
| ESTB2_EUR | 0.0249 | 0.055 | 0.024 |
| FINNG_EUR | 0.029 | 0.053 | 0.024 |
| GEDIS_EUR | 0.2025 | 0.172 | 0.135 |
| GISS1_EUR | 0.7997 | 0.022 | 0.085 |
| GISS2_EUR | 0.2974 | 0.104 | 0.1 |
| IPSYC_EUR | 0.003 | 0.058 | 0.02 |
| JANS3_EUR | 0.3508 | 0.101 | 0.109 |
| JANS4_EUR | 0.4765 | −0.09 | 0.126 |
| MIREC_EUR | 0.4044 | 0.128 | 0.154 |
| MVPXQ_EUR | 0.0383 | 0.028 | 0.013 |
| PGCBD_EUR | 0.0019 | 0.085 | 0.027 |
| PGCED_EUR | 0.8654 | 0.02 | 0.119 |
| PGCMD_EUR | 0.176 | 0.049 | 0.036 |
| PGCPT_EUR | 0.8065 | −0.015 | 0.061 |
| PGCSZ_EUR | 0.2416 | 0.045 | 0.039 |
| PRFCT_EUR | 0.8308 | −0.01 | 0.046 |
| PSYCR_EUR | 0.0348 | 0.193 | 0.091 |
| QIMRB_EUR | 0.082 | 0.05 | 0.029 |
| UKBJC_EUR | 0.7395 | −0.007 | 0.021 |
| UTAH2_EUR | 0.1223 | 0.031 | 0.02 |
| YPENN_EUR | 0.1191 | −0.227 | 0.145 |
| **meta** | **3.61e−09** | **0.039** | **0.007** |

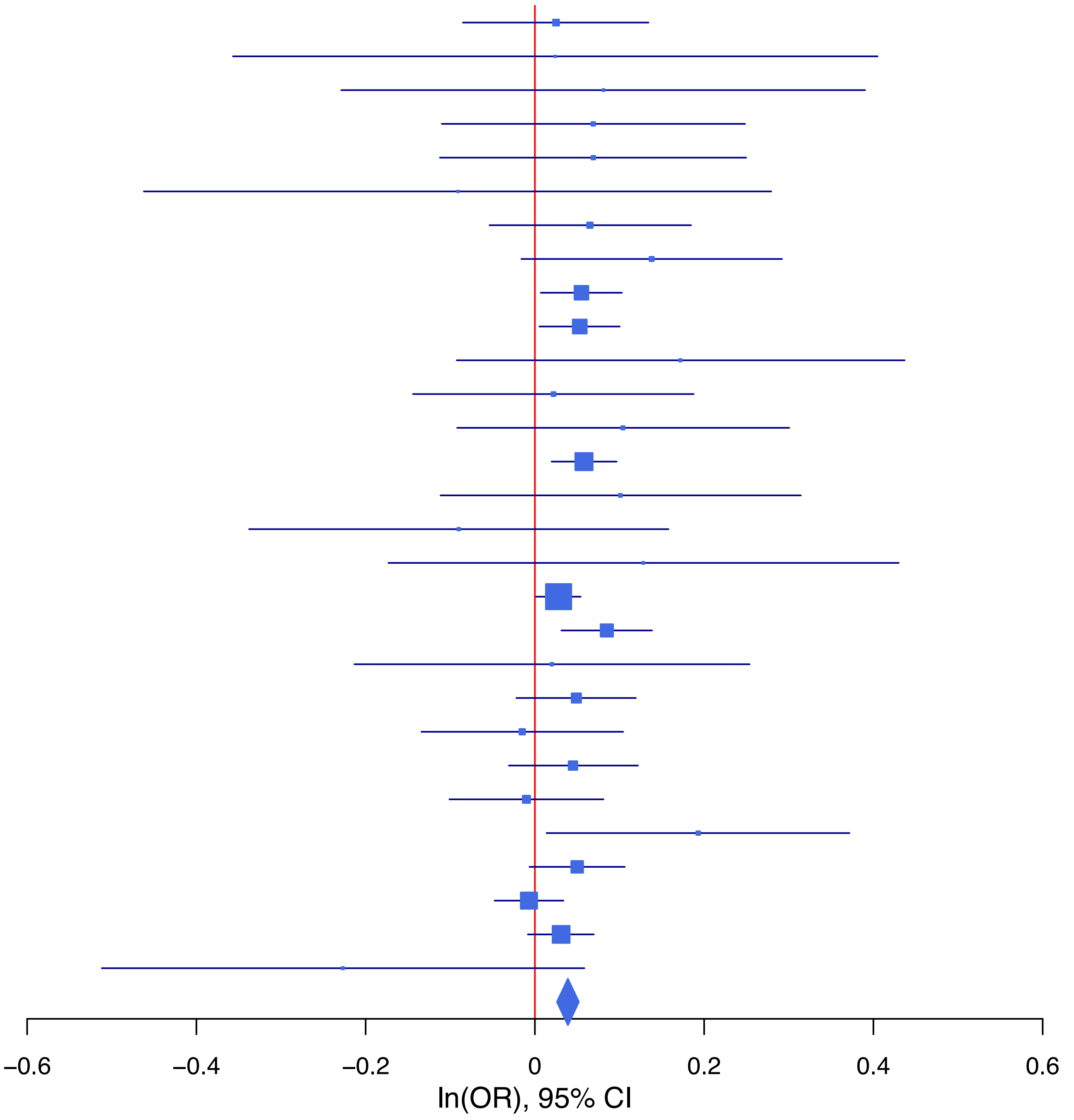

**rs9399221 T/C 6:137914871**

| Cohort | P | ln(OR) | SE |
|---|---|---|---|
| ALSPC_EUR | 0.8003 | −0.018 | 0.07 |
| BEPS7_EUR | 0.215 | 0.266 | 0.215 |
| BHRCM_EUR | 0.0864 | −0.361 | 0.21 |
| BOR17_EUR | 0.629 | −0.054 | 0.111 |
| BOR2C_EUR | 0.7739 | −0.032 | 0.11 |
| BOR2E_EUR | 0.2814 | −0.229 | 0.212 |
| COGA1_EUR | 0.2402 | −0.09 | 0.076 |
| CUINT_EUR | 0.2844 | −0.105 | 0.098 |
| ESTB2_EUR | 0.0512 | −0.054 | 0.028 |
| FINNG_EUR | 0.0018 | −0.078 | 0.025 |
| GEDIS_EUR | 0.7859 | −0.047 | 0.172 |
| GISS1_EUR | 0.5168 | −0.065 | 0.1 |
| GISS2_EUR | 0.3854 | 0.102 | 0.118 |
| IPSYC_EUR | 0.0669 | −0.042 | 0.023 |
| JANS3_EUR | 0.8219 | −0.031 | 0.138 |
| JANS4_EUR | 0.0499 | 0.282 | 0.144 |
| MVPXQ_EUR | 0.0039 | −0.049 | 0.017 |
| PGCBD_EUR | 0.3144 | −0.034 | 0.033 |
| PGCED_EUR | 0.7945 | −0.039 | 0.149 |
| PGCMD_EUR | 0.0904 | −0.077 | 0.045 |
| PGCPT_EUR | 0.6416 | 0.034 | 0.073 |
| PGCSZ_EUR | 0.2935 | −0.051 | 0.048 |
| PRFCT_EUR | 0.6752 | −0.023 | 0.055 |
| PSYCR_EUR | 0.6377 | −0.053 | 0.113 |
| QIMRB_EUR | 0.2839 | −0.037 | 0.035 |
| UKBJC_EUR | 0.1671 | −0.035 | 0.025 |
| UTAH2_EUR | 0.0023 | −0.072 | 0.024 |
| YPENN_EUR | 0.5532 | 0.094 | 0.159 |
| **meta** | **5.34e−10** | **−0.049** | **0.008** |

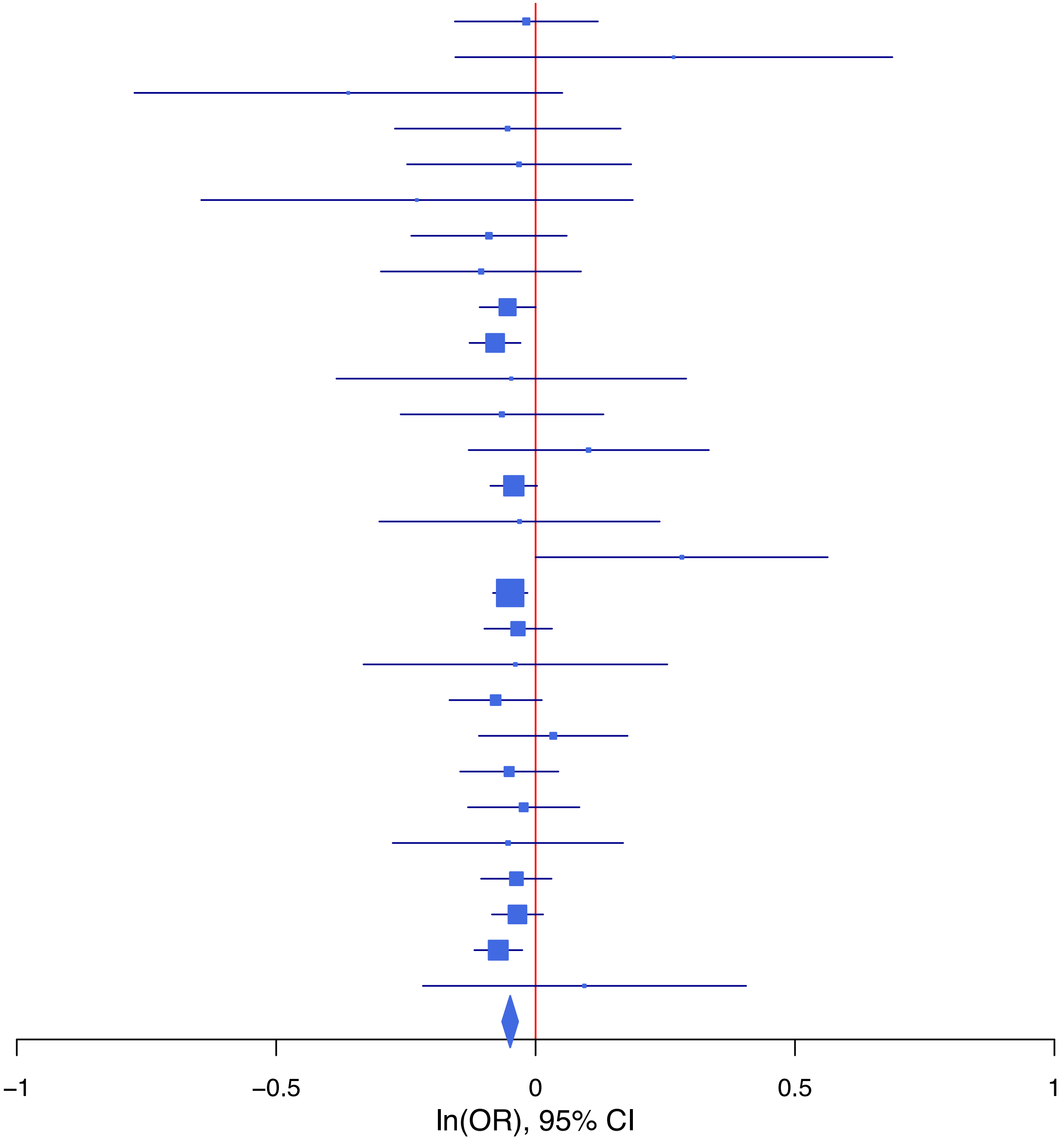

**rs9592599 A/T 13:69575338**

| Cohort | P | ln(OR) | SE |
|---|---|---|---|
| ALSPC_EUR | 0.7737 | −0.016 | 0.055 |
| BEPS7_EUR | 0.79 | −0.048 | 0.182 |
| BOR17_EUR | 0.1048 | −0.142 | 0.087 |
| BOR2C_EUR | 0.0125 | −0.219 | 0.088 |
| BOR2E_EUR | 0.8015 | 0.045 | 0.179 |
| COGA1_EUR | 0.4793 | −0.042 | 0.059 |
| ESTB2_EUR | 0.0028 | −0.069 | 0.023 |
| GEDIS_EUR | 0.6842 | 0.054 | 0.131 |
| GISS1_EUR | 0.0426 | −0.161 | 0.08 |
| GISS2_EUR | 0.3333 | 0.087 | 0.09 |
| IPSYC_EUR | 0.4804 | −0.013 | 0.019 |
| JANS3_EUR | 0.7732 | −0.03 | 0.104 |
| JANS4_EUR | 0.0904 | −0.206 | 0.122 |
| MIREC_EUR | 0.3195 | −0.153 | 0.154 |
| PGCBD_EUR | 0.0152 | −0.063 | 0.026 |
| PGCED_EUR | 0.5551 | 0.069 | 0.118 |
| PGCMD_EUR | 0.175 | −0.047 | 0.035 |
| PGCPT_EUR | 0.1014 | −0.095 | 0.058 |
| PGCSZ_EUR | 0.3029 | −0.039 | 0.037 |
| PRFCT_EUR | 0.1373 | −0.065 | 0.044 |
| PSYCR_EUR | 0.8664 | −0.015 | 0.088 |
| QIMRB_EUR | 0.2993 | −0.028 | 0.027 |
| UKBJC_EUR | 0.2412 | −0.023 | 0.02 |
| UTAH2_EUR | 0.0074 | −0.052 | 0.02 |
| YPENN_EUR | 0.4762 | −0.094 | 0.131 |
| **meta** | **2.25e−08** | **−0.043** | **0.008** |

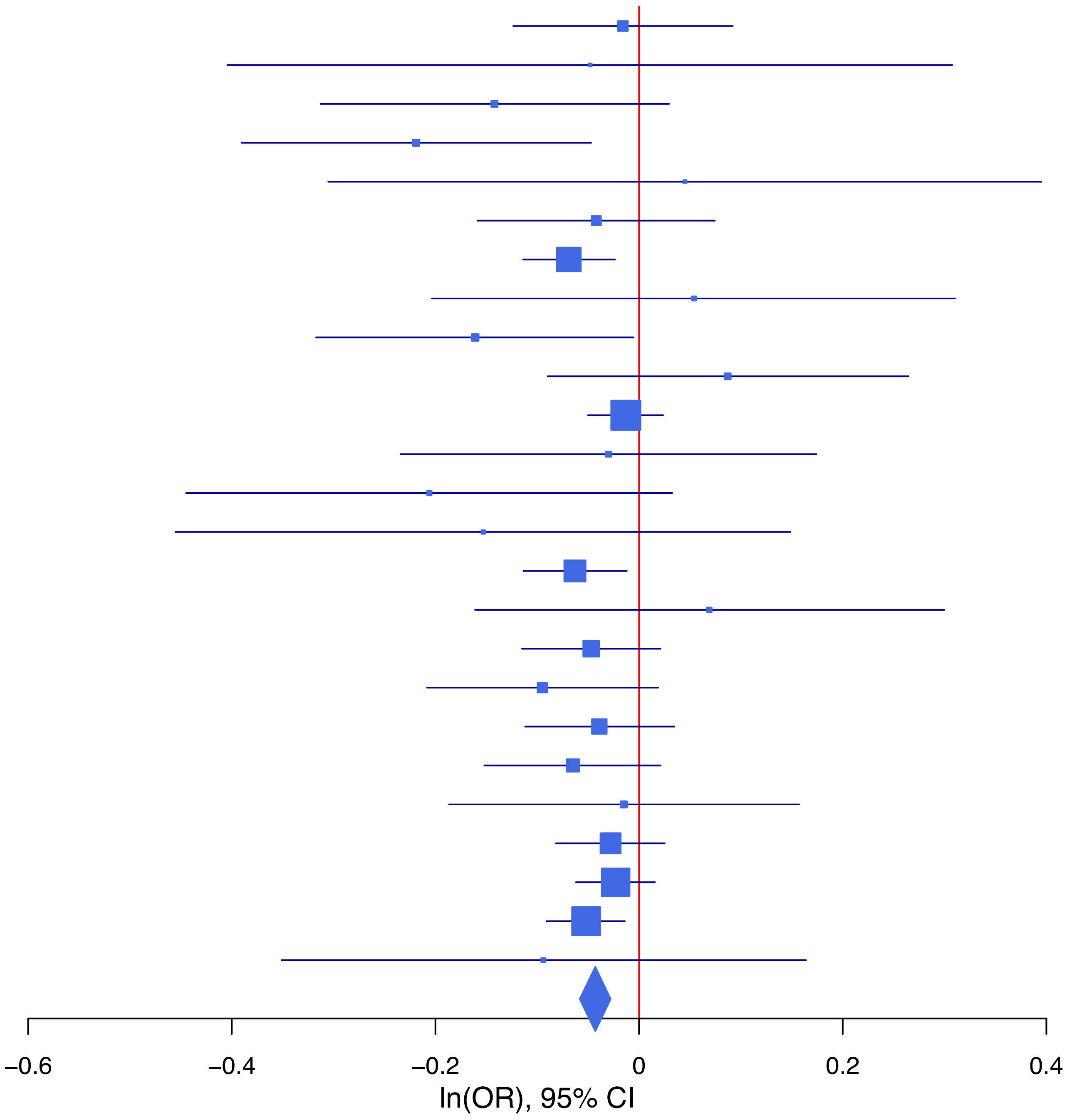

**rs989652 A/G 15:47823734**

| Cohort | P | ln(OR) | SE |
|---|---|---|---|
| ALSPC_EUR | 0.8165 | 0.013 | 0.055 |
| BEPS7_EUR | 0.0195 | 0.433 | 0.185 |
| BHRCM_EUR | 0.954 | 0.009 | 0.155 |
| BOR17_EUR | 0.7545 | 0.027 | 0.086 |
| BOR2C_EUR | 0.5761 | −0.05 | 0.089 |
| BOR2E_EUR | 0.464 | −0.138 | 0.189 |
| COGA1_EUR | 0.2873 | 0.063 | 0.059 |
| CUINT_EUR | 0.0241 | −0.169 | 0.075 |
| ESTB2_EUR | 0.0942 | 0.039 | 0.023 |
| FINNG_EUR | 0.1368 | 0.035 | 0.024 |
| GEDIS_EUR | 0.8775 | 0.021 | 0.132 |
| GISS1_EUR | 0.7197 | −0.029 | 0.08 |
| GISS2_EUR | 0.0232 | 0.218 | 0.096 |
| IPSYC_EUR | 0.1818 | 0.025 | 0.019 |
| JANS3_EUR | 0.174 | 0.141 | 0.104 |
| JANS4_EUR | 0.5348 | −0.074 | 0.118 |
| MIREC_EUR | 0.6493 | 0.069 | 0.151 |
| MVPXQ_EUR | 0.0338 | 0.027 | 0.013 |
| PGCBD_EUR | 0.694 | 0.01 | 0.026 |
| PGCED_EUR | 0.5807 | 0.063 | 0.114 |
| PGCMD_EUR | 0.0449 | 0.069 | 0.035 |
| PGCPT_EUR | 0.6432 | −0.027 | 0.058 |
| PGCSZ_EUR | 1e−04 | 0.15 | 0.037 |
| PRFCT_EUR | 0.016 | −0.109 | 0.045 |
| PSYCR_EUR | 0.1544 | 0.126 | 0.089 |
| QIMRB_EUR | 0.2791 | 0.03 | 0.028 |
| UKBJC_EUR | 2e−04 | 0.074 | 0.02 |
| UTAH2_EUR | 5e−04 | 0.067 | 0.019 |
| YPENN_EUR | 0.2398 | 0.148 | 0.126 |
| **meta** | **1.46e−09** | **0.038** | **0.006** |

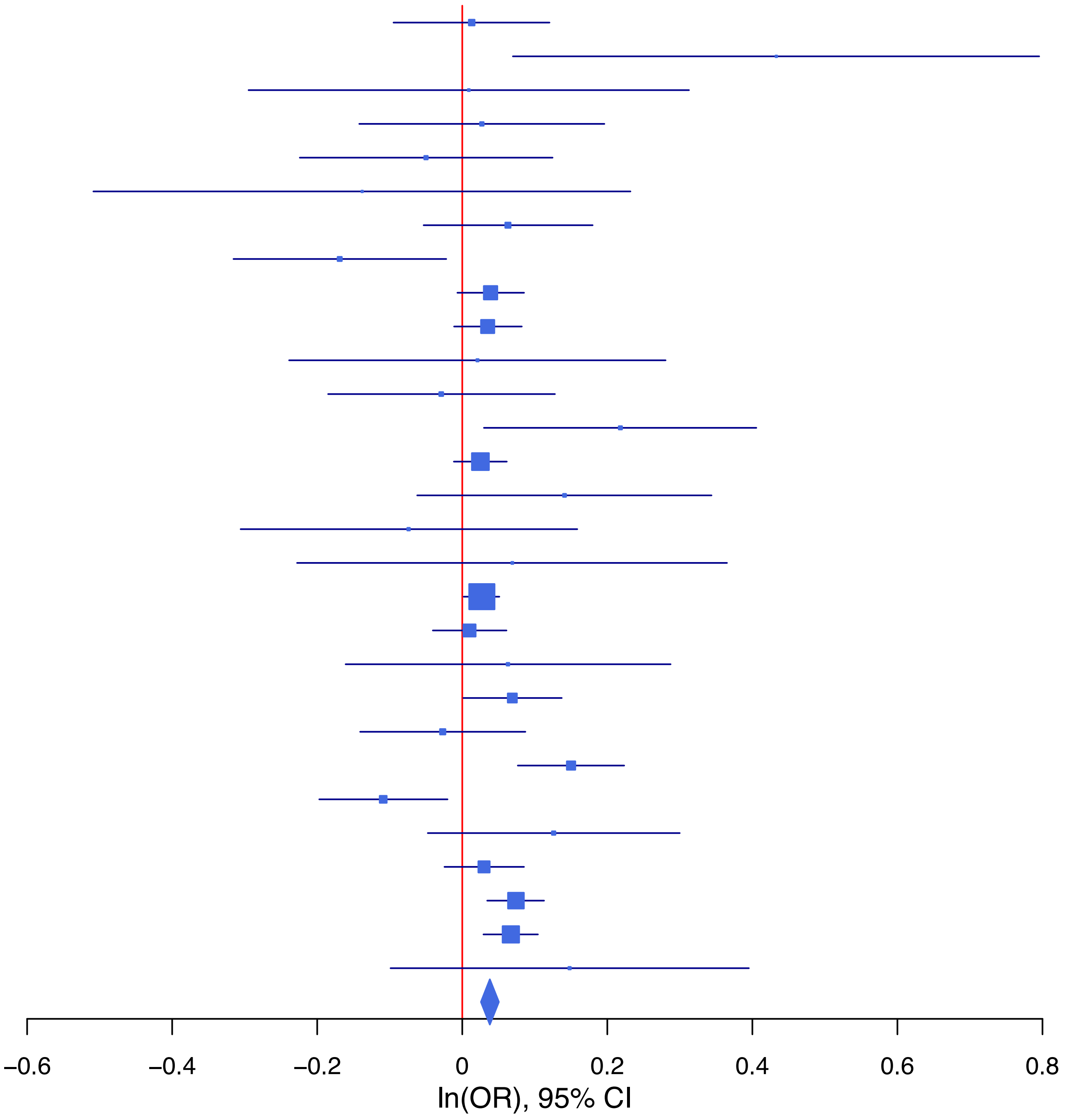
