## Supplementary Data 2 for "Genome-wide association studies identify 77 loci for suicidality and provide novel biological insights"

**Supplementary Data 2A: Area plots of lead SNPs at the 13 genome-wide significant loci from the multi-ancestry GWAS meta-analysis of suicidal ideation.**
If >1 independent SNPs are in the same plot, then each is given a different color, and their LD partners are shaded with the same color. Detailed info about each independent index SNP (a., b., c. ...) is provided in the right upper corner. If SNPs share independent index SNPs as an LD partner, the SNP is assigned to the more significant one. Information regarding other GWAS associations at the locus is provided in the left upper corner.

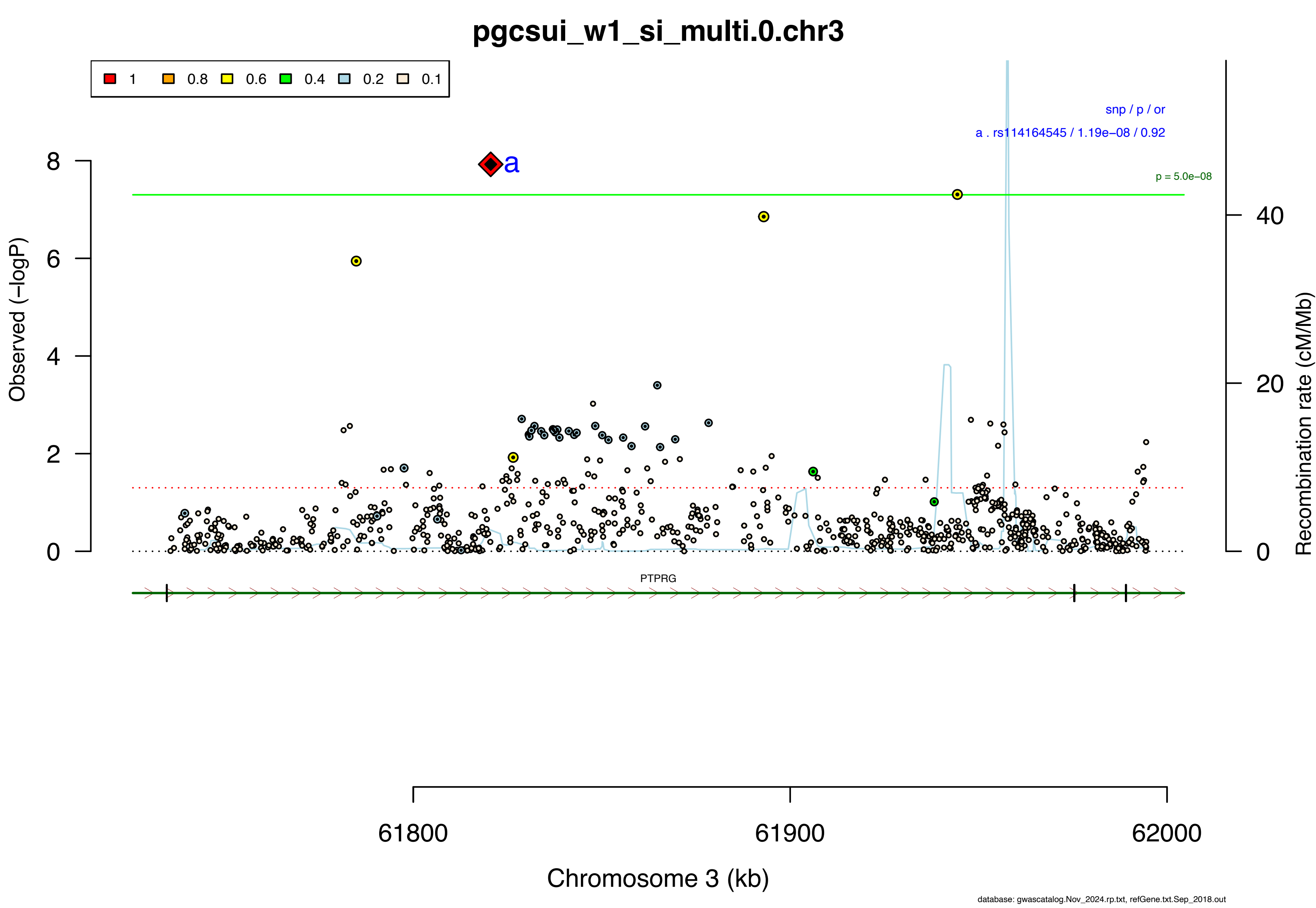
pgcsui_w1_si_multi.0.chr3
1
0.8
0.6
0.4
0.2
0.1
snp / p / or
a . rs114164545 / 1.19e−08 / 0.92
p = 5.0e−08
a
Observed (−logP)
Recombination rate (cM/Mb)
PTPRG
61800
61900
62000
Chromosome 3 (kb)
database: gwascatalog.Nov_2024.rp.txt, refGene.txt.Sep_2018.out

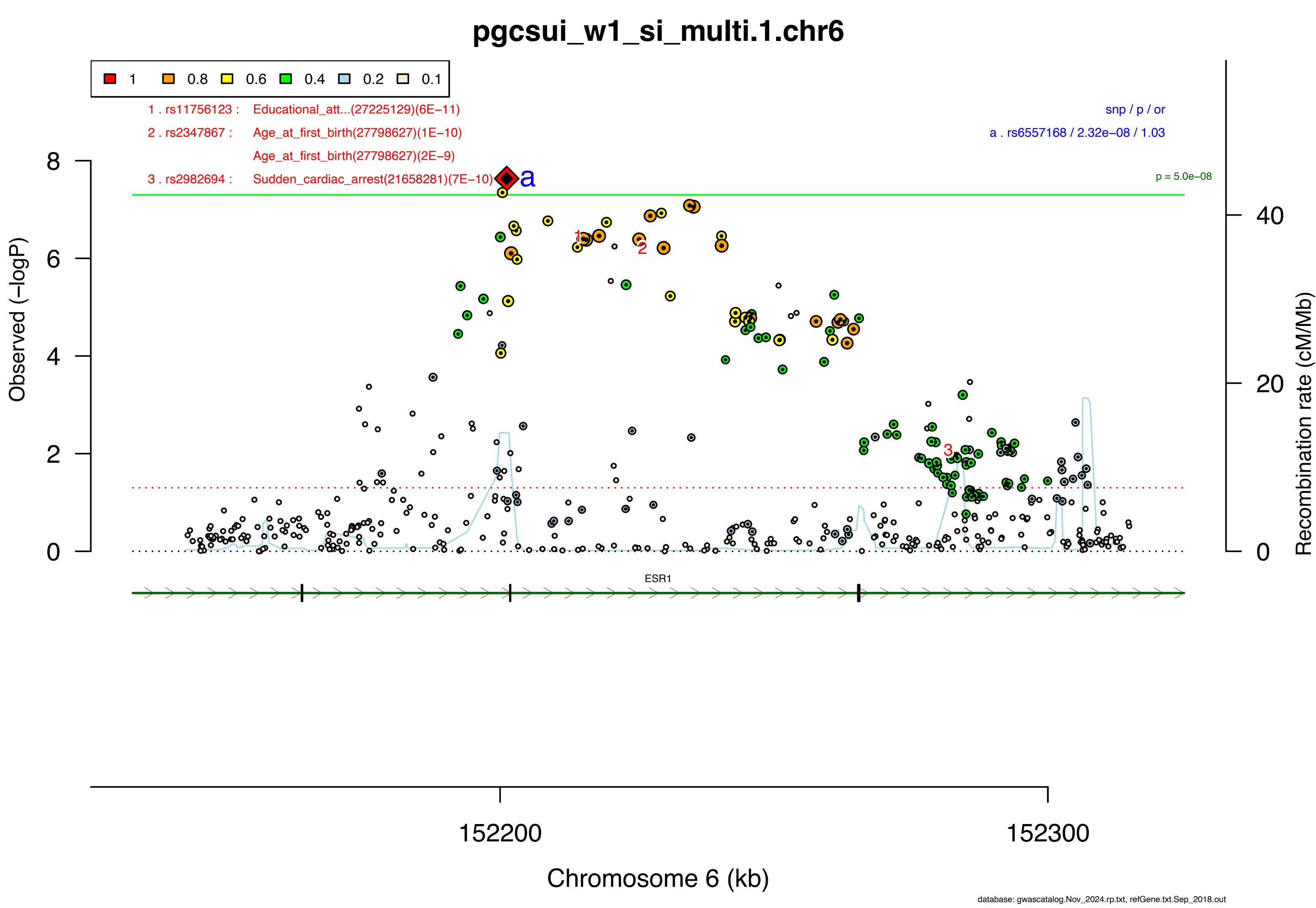

pgcsui_w1_si_multi.1.chr6
1
0.8
0.6
0.4
0.2
0.1
1 . rs11756123 : Educational_att...(27225129)(6E−11)
2 . rs2347867 : Age_at_first_birth(27798627)(1E−10)
Age_at_first_birth(27798627)(2E−9)
3 . rs2982694 : Sudden_cardiac_arrest(21658281)(7E−10)
a
snp / p / or
a . rs6557168 / 2.32e−08 / 1.03
p = 5.0e−08
Observed (−logP)
Recombination rate (cM/Mb)
ESR1
152200
152300
Chromosome 6 (kb)
database: gwascatalog.Nov_2024.rp.txt, refGene.txt.Sep_2018.out

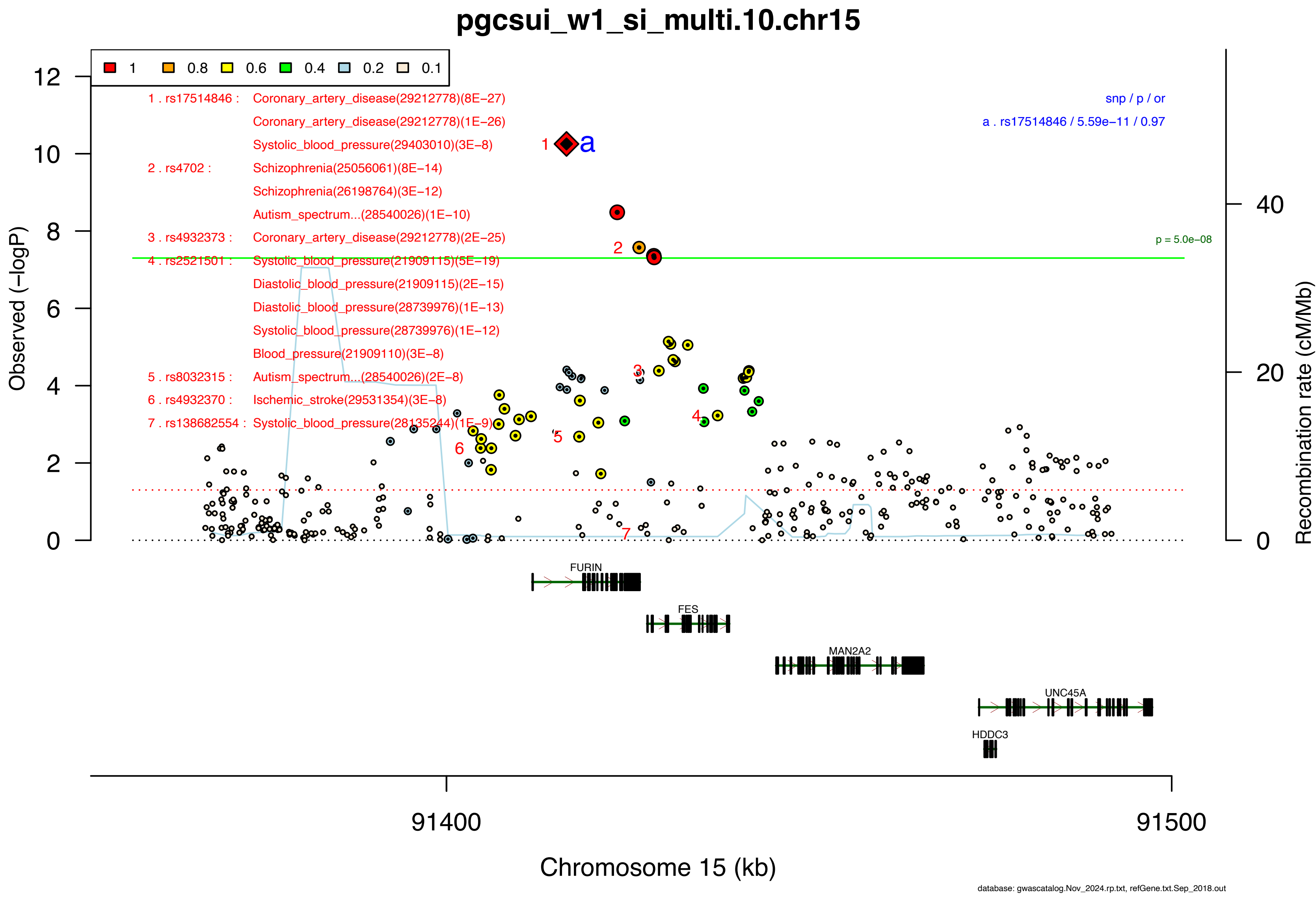
pgcsui_w1_si_multi.10.chr15
1
0.8
0.6
0.4
0.2
0.1
1 . rs17514846 : Coronary_artery_disease(29212778)(8E−27)
Coronary_artery_disease(29212778)(1E−26)
Systolic_blood_pressure(29403010)(3E−8)
2 . rs4702 : Schizophrenia(25056061)(8E−14)
Schizophrenia(26198764)(3E−12)
Autism_spectrum...(28540026)(1E−10)
3 . rs4932373 : Coronary_artery_disease(29212778)(2E−25)
4 . rs2521501 : Systolic_blood_pressure(21909115)(5E−19)
Diastolic_blood_pressure(21909115)(2E−15)
Diastolic_blood_pressure(28739976)(1E−13)
Systolic_blood_pressure(28739976)(1E−12)
Blood_pressure(21909110)(3E−8)
5 . rs8032315 : Autism_spectrum...(28540026)(2E−8)
6 . rs4932370 : Ischemic_stroke(29531354)(3E−8)
7 . rs138682554 : Systolic_blood_pressure(28135244)(1E−9)
snp / p / or
a . rs17514846 / 5.59e−11 / 0.97
p = 5.0e−08
Observed (−logP)
Recombination rate (cM/Mb)
FURIN
FES
MAN2A2
UNC45A
HDDC3
91400
91500
Chromosome 15 (kb)
database: gwascatalog.Nov_2024.rp.txt, refGene.txt.Sep_2018.out

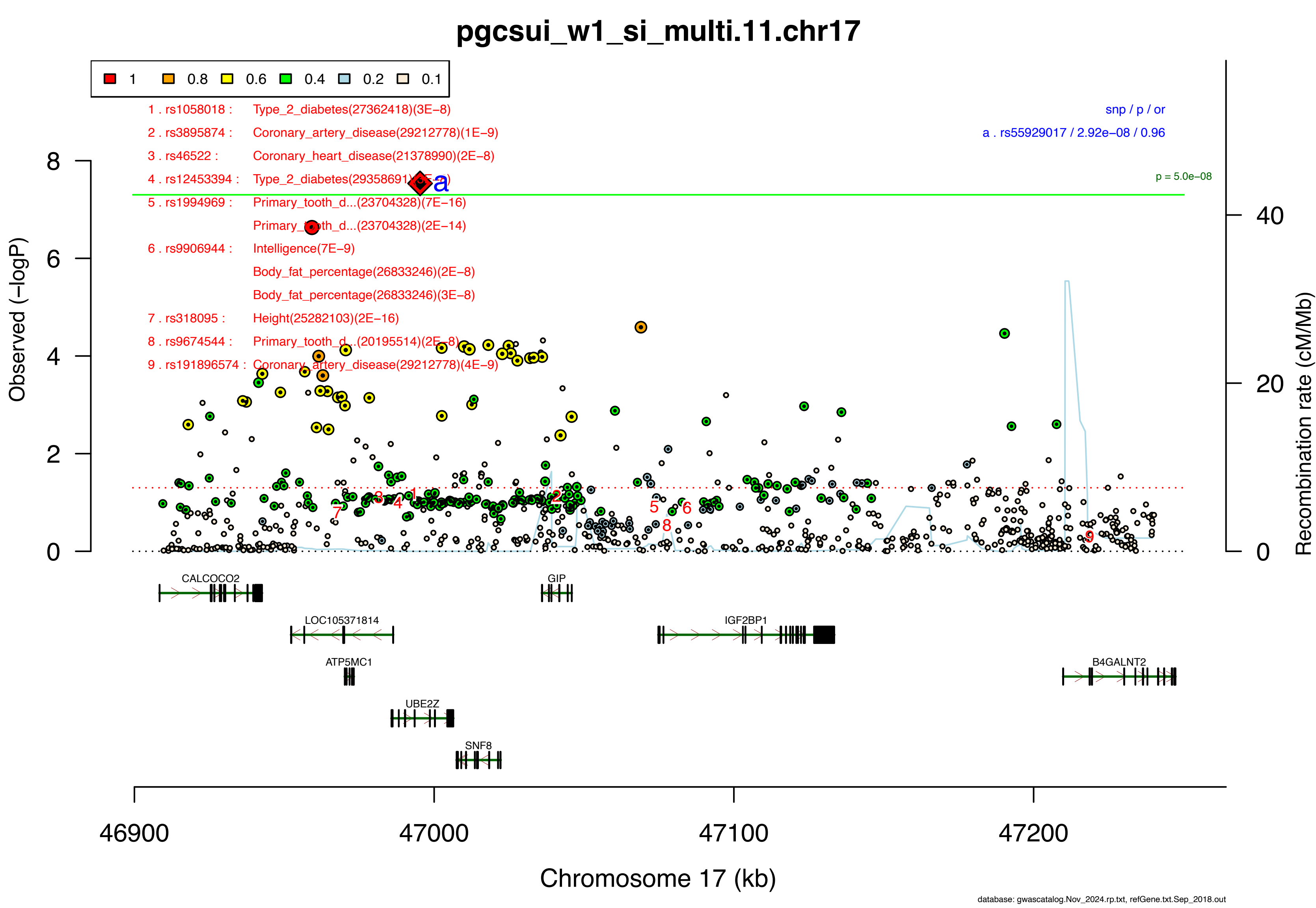

pgcsui_w1_si_multi.11.chr17
1
0.8
0.6
0.4
0.2
0.1
1 . rs1058018 : Type_2_diabetes(27362418)(3E−8)
2 . rs3895874 : Coronary_artery_disease(29212778)(1E−9)
3 . rs46522 : Coronary_heart_disease(21378990)(2E−8)
4 . rs12453394 : Type_2_diabetes(29358691)(5E−8)
5 . rs1994969 : Primary_tooth_d...(23704328)(7E−16)
Primary_tooth_d...(23704328)(2E−14)
6 . rs9906944 : Intelligence(7E−9)
Body_fat_percentage(26833246)(2E−8)
Body_fat_percentage(26833246)(3E−8)
7 . rs318095 : Height(25282103)(2E−16)
8 . rs9674544 : Primary_tooth_d...(20195514)(2E−8)
9 . rs191896574 : Coronary_artery_disease(29212778)(4E−9)
snp / p / or
a . rs55929017 / 2.92e−08 / 0.96
p = 5.0e−08
Observed (−logP)
Recombination rate (cM/Mb)
CALCOCO2
GIP
LOC105371814
IGF2BP1
ATP5MC1
B4GALNT2
UBE2Z
SNF8
46900
47000
47100
47200
Chromosome 17 (kb)
database: gwascatalog.Nov_2024.rp.txt, refGene.txt.Sep_2018.out

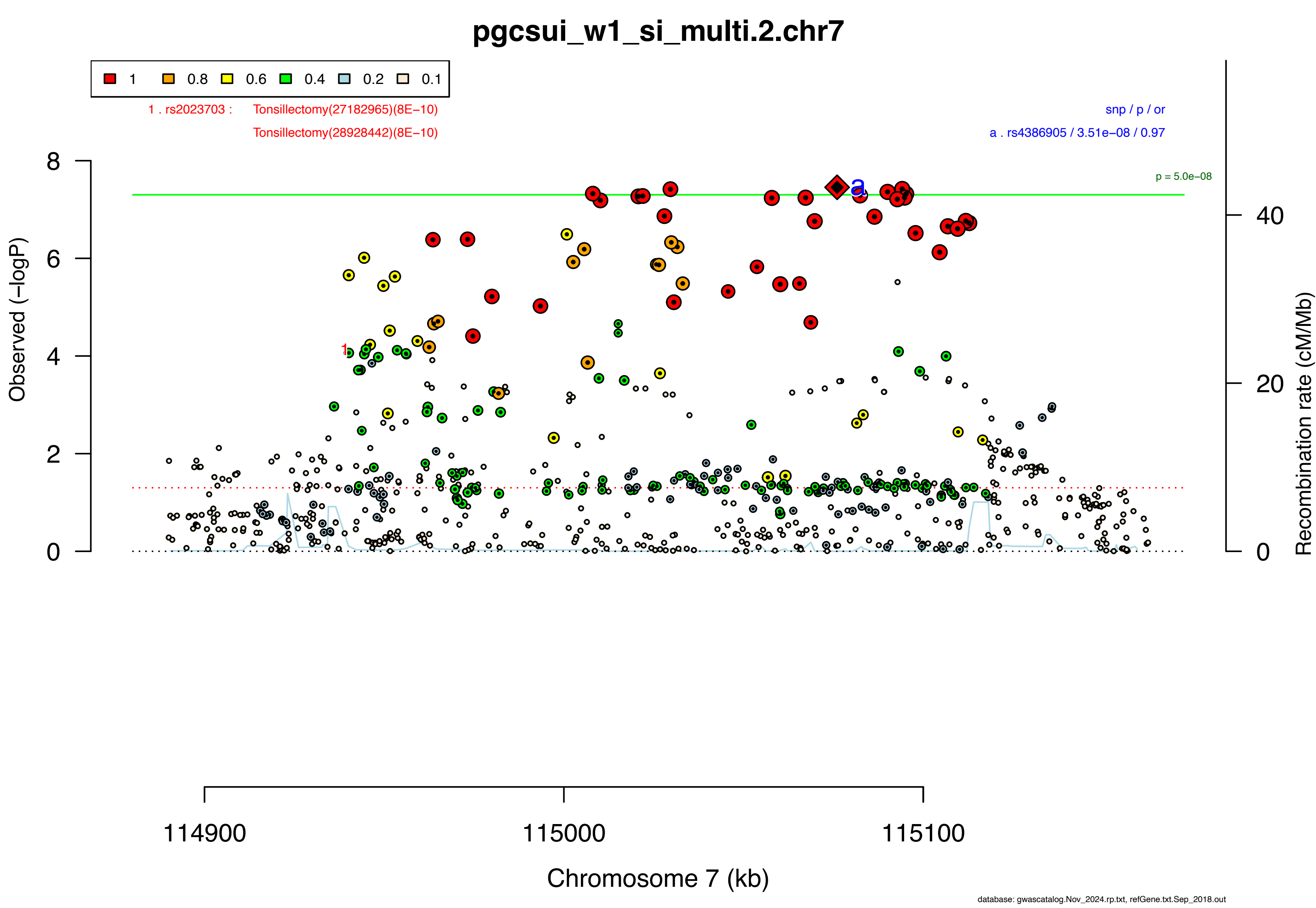
pgcsui_w1_si_multi.2.chr7
1
0.8
0.6
0.4
0.2
0.1
1 . rs2023703 : Tonsillectomy(27182965)(8E−10)
Tonsillectomy(28928442)(8E−10)
snp / p / or
a . rs4386905 / 3.51e−08 / 0.97
p = 5.0e−08
Observed (−logP)
Recombination rate (cM/Mb)
0
2
4
6
8
0
20
40
114900
115000
115100
Chromosome 7 (kb)
database: gwascatalog.Nov_2024.rp.txt, refGene.txt.Sep_2018.out

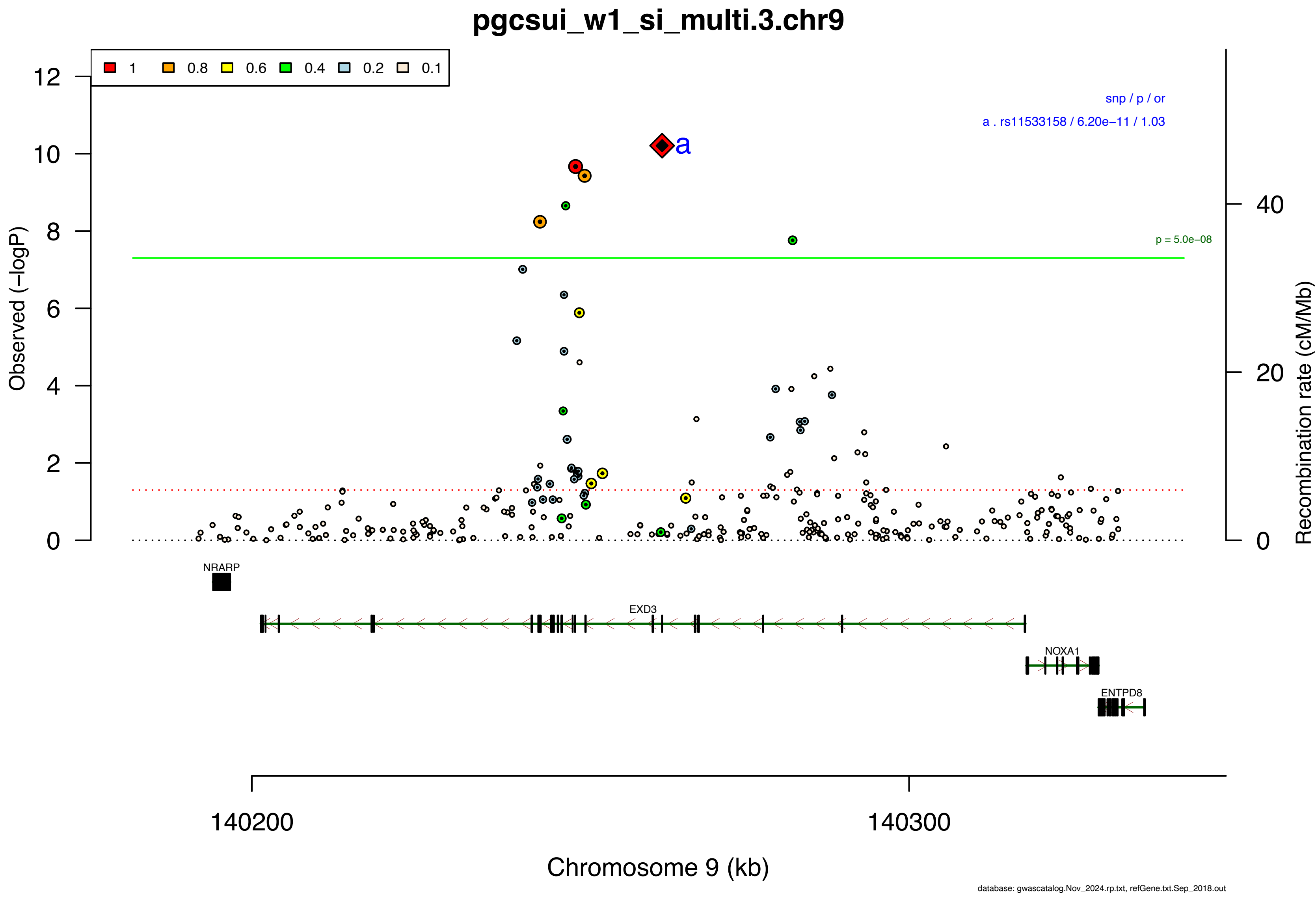
pgcsui_w1_si_multi.3.chr9
1
0.8
0.6
0.4
0.2
0.1
snp / p / or
a . rs11533158 / 6.20e−11 / 1.03
a
p = 5.0e−08
Observed (−logP)
Recombination rate (cM/Mb)
NRARP
EXD3
NOXA1
ENTPD8
140200
140300
Chromosome 9 (kb)
database: gwascatalog.Nov_2024.rp.txt, refGene.txt.Sep_2018.out

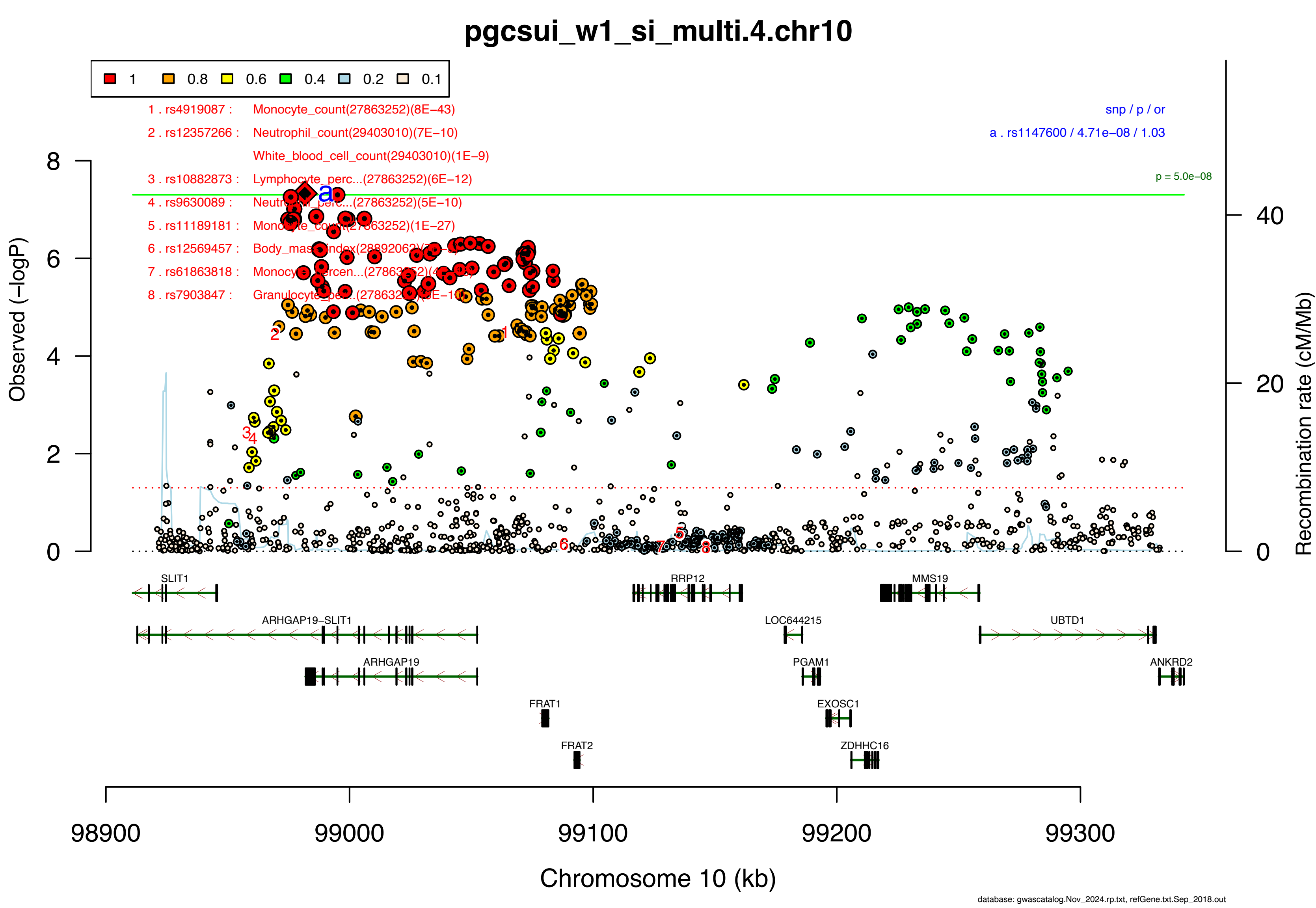
pgcsui_w1_si_multi.4.chr10
1
0.8
0.6
0.4
0.2
0.1
1 . rs4919087 : Monocyte_count(27863252)(8E−43)
2 . rs12357266 : Neutrophil_count(29403010)(7E−10)
White_blood_cell_count(29403010)(1E−9)
3 . rs10882873 : Lymphocyte_perc...(27863252)(6E−12)
4 . rs9630089 :
5 . rs11189181 :
6 . rs12569457 : Body_mass_index
7 . rs61863818 :
8 . rs7903847 : Granulocyte_perc...
snp / p / or
a . rs1147600 / 4.71e−08 / 1.03
p = 5.0e−08
Observed (−logP)
Recombination rate (cM/Mb)
SLIT1
RRP12
MMS19
ARHGAP19−SLIT1
LOC644215
UBTD1
ARHGAP19
PGAM1
ANKRD2
FRAT1
EXOSC1
FRAT2
ZDHHC16
98900
99000
99100
99200
99300
Chromosome 10 (kb)
database: gwascatalog.Nov_2024.rp.txt, refGene.txt.Sep_2018.out

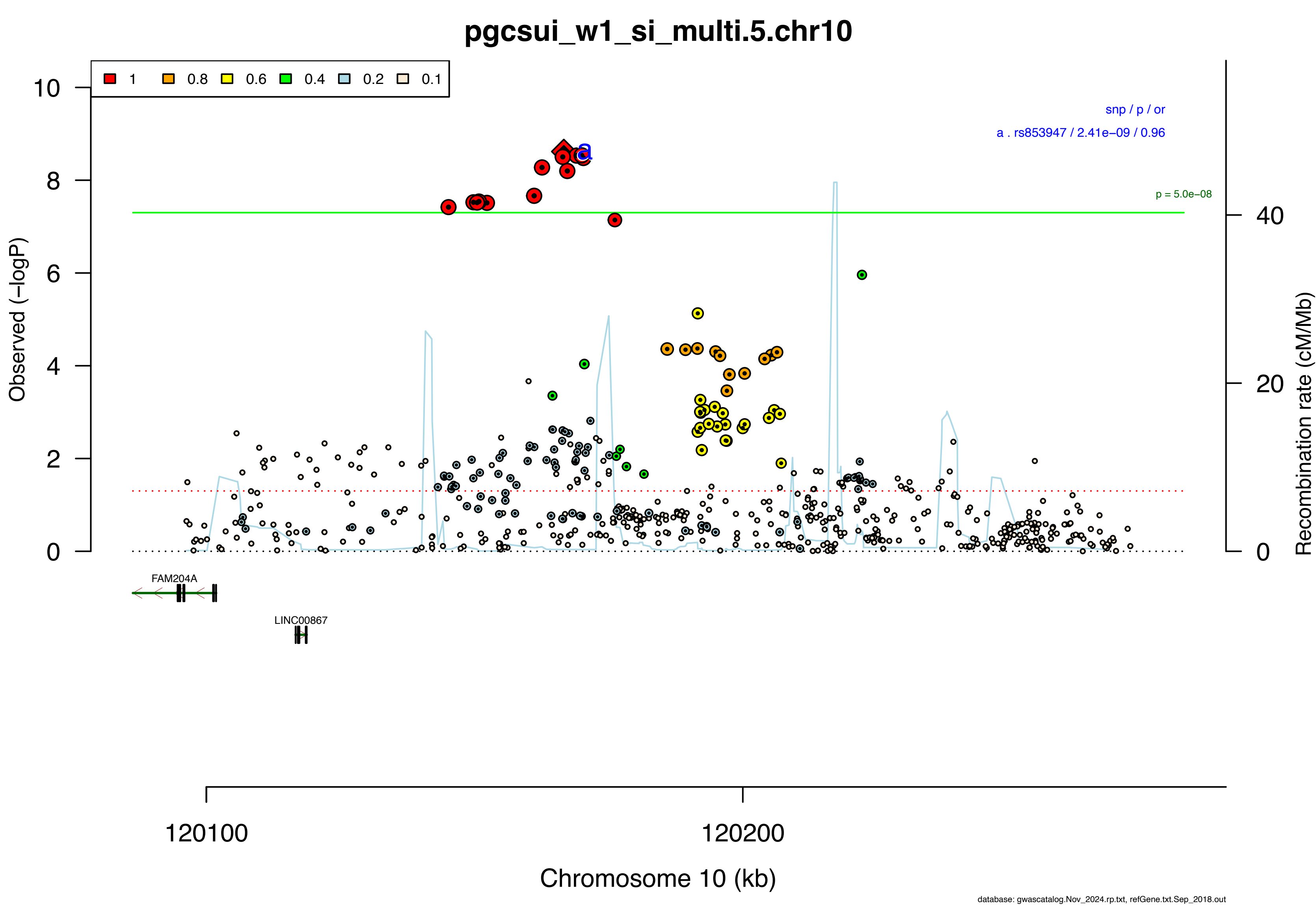

pgcsui_w1_si_multi.5.chr10
1
0.8
0.6
0.4
0.2
0.1
snp / p / or
a . rs853947 / 2.41e−09 / 0.96
p = 5.0e−08
a
Observed (−logP)
Recombination rate (cM/Mb)
10
8
6
4
2
0
40
20
0
FAM204A
LINC00867
120100
120200
Chromosome 10 (kb)
database: gwascatalog.Nov_2024.rp.txt, refGene.txt.Sep_2018.out

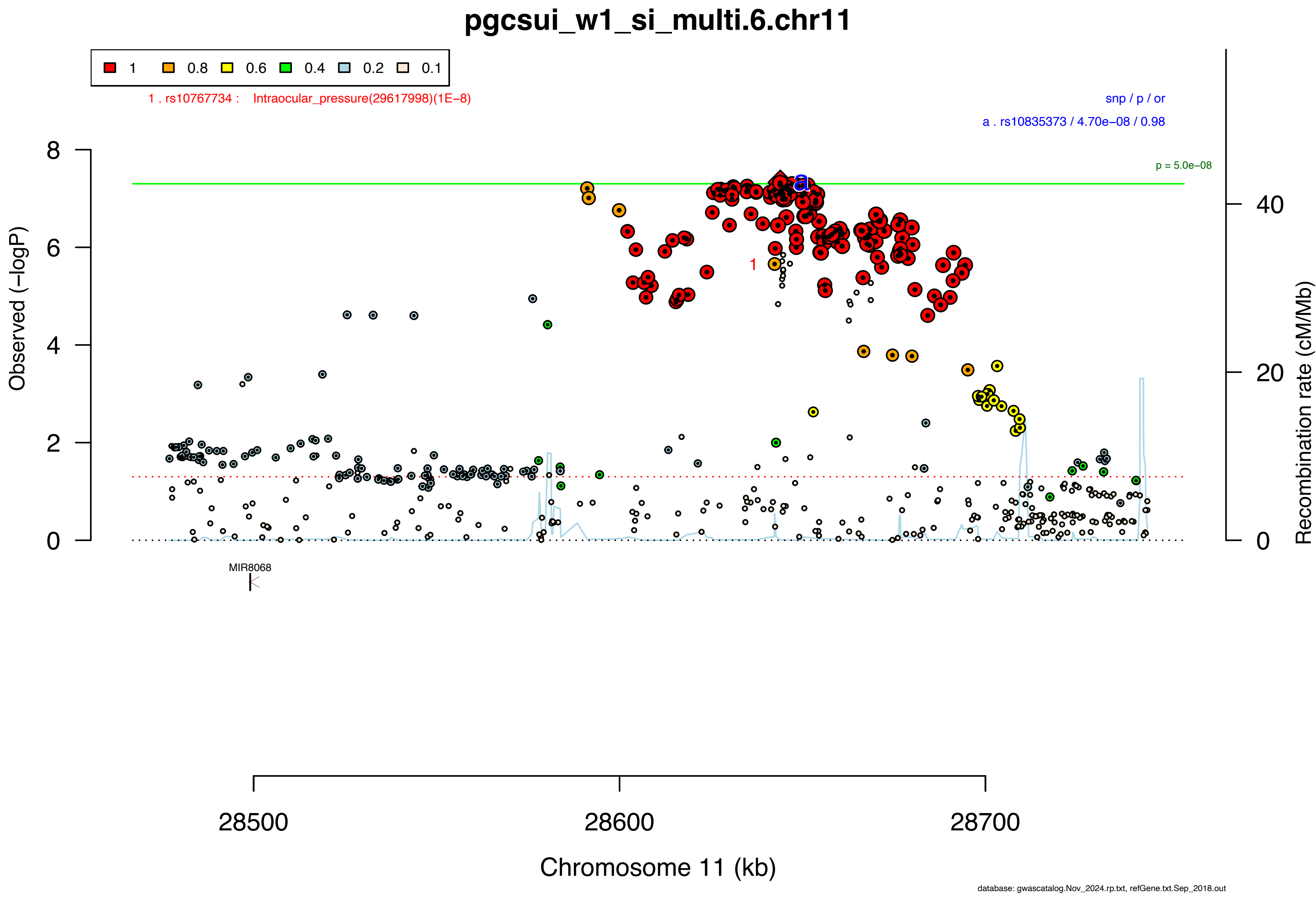

pgcsui_w1_si_multi.6.chr11
1
0.8
0.6
0.4
0.2
0.1
1 . rs10767734 : Intraocular_pressure(29617998)(1E−8)
snp / p / or
a . rs10835373 / 4.70e−08 / 0.98
p = 5.0e−08
Observed (−logP)
Recombination rate (cM/Mb)
MIR8068
28500
28600
28700
Chromosome 11 (kb)
database: gwascatalog.Nov_2024.rp.txt, refGene.txt.Sep_2018.out

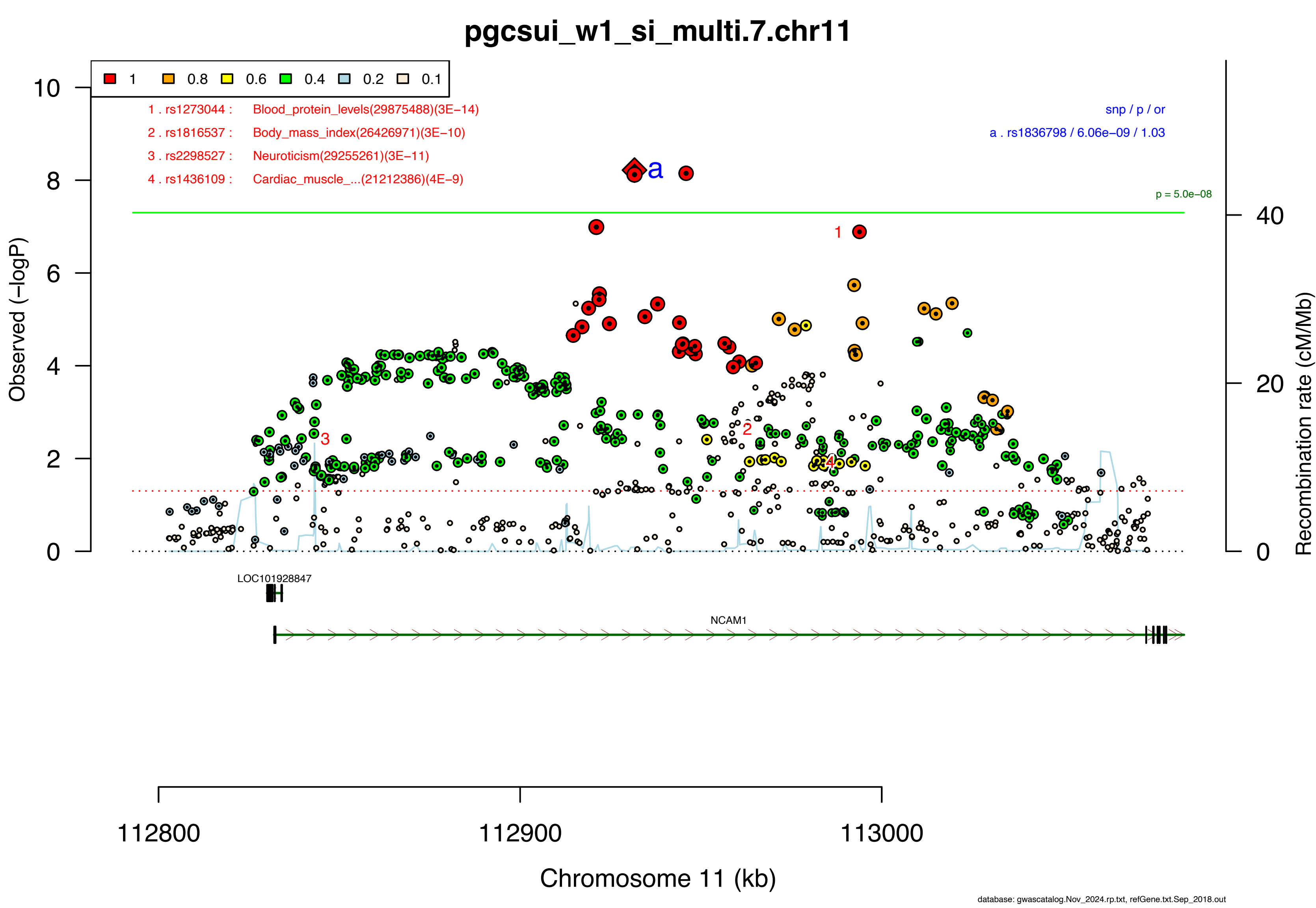
pgcsui_w1_si_multi.7.chr11
1
0.8
0.6
0.4
0.2
0.1
1 . rs1273044 : Blood_protein_levels(29875488)(3E−14)
2 . rs1816537 : Body_mass_index(26426971)(3E−10)
3 . rs2298527 : Neuroticism(29255261)(3E−11)
4 . rs1436109 : Cardiac_muscle_...(21212386)(4E−9)
snp / p / or
a . rs1836798 / 6.06e−09 / 1.03
p = 5.0e−08
Observed (−logP)
Recombination rate (cM/Mb)
LOC101928847
NCAM1
112800
112900
113000
Chromosome 11 (kb)
database: gwascatalog.Nov_2024.rp.txt, refGene.txt.Sep_2018.out

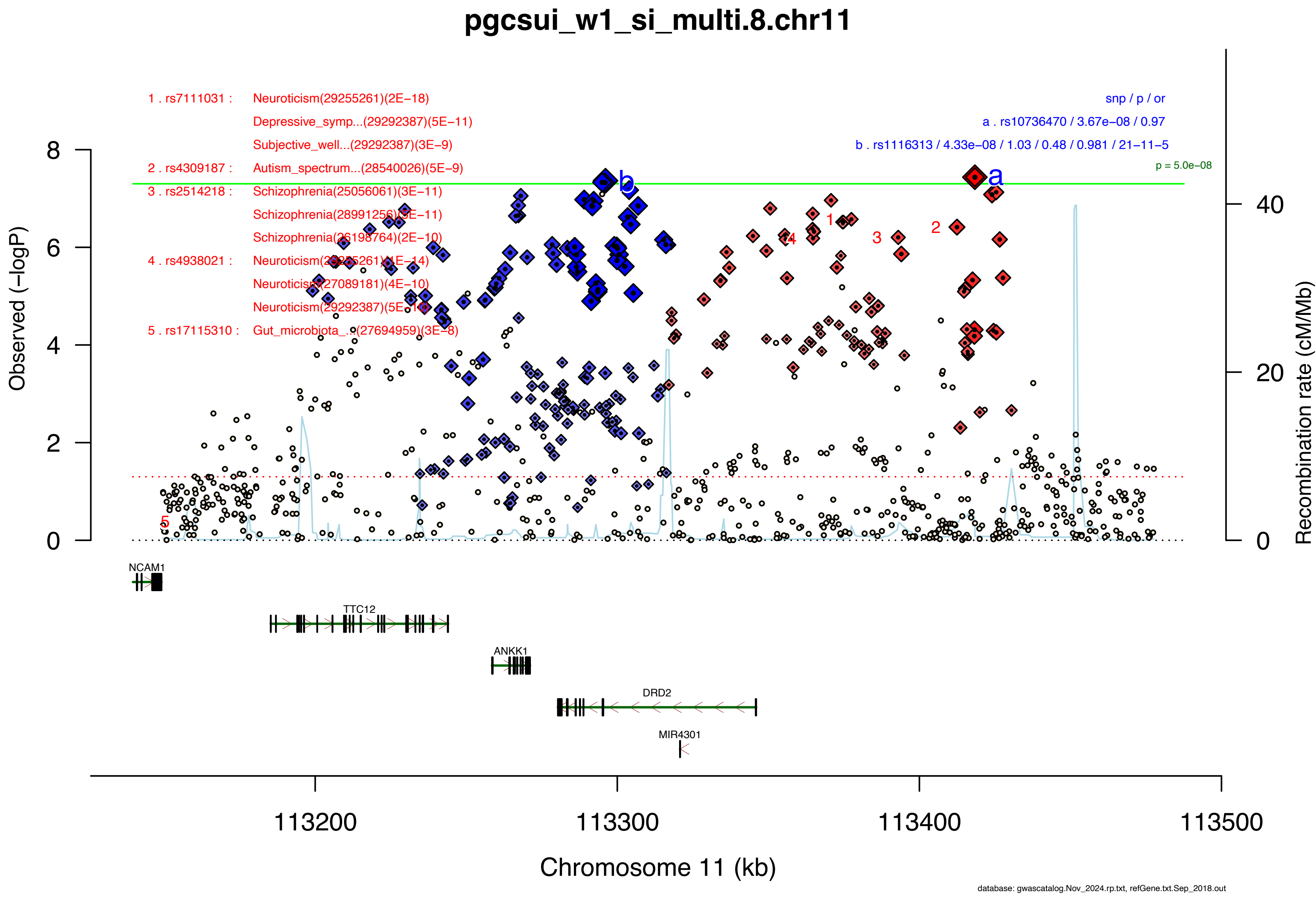

pgcsui_w1_si_multi.8.chr11
1 . rs7111031 : Neuroticism(29255261)(2E−18)
Depressive_symp...(29292387)(5E−11)
Subjective_well...(29292387)(3E−9)
2 . rs4309187 : Autism_spectrum...(28540026)(5E−9)
3 . rs2514218 : Schizophrenia(25056061)(3E−11)
Schizophrenia(28991256)(3E−11)
Schizophrenia(26198764)(2E−10)
4 . rs4938021 : Neuroticism(29255261)(1E−14)
Neuroticism(27089181)(4E−10)
Neuroticism(29292387)(5E−10)
5 . rs17115310 : Gut_microbiota_...(27694959)(3E−8)
snp / p / or
a . rs10736470 / 3.67e−08 / 0.97
b . rs1116313 / 4.33e−08 / 1.03 / 0.48 / 0.981 / 21−11−5
p = 5.0e−08
Observed (−logP)
Recombination rate (cM/Mb)
NCAM1
TTC12
ANKK1
DRD2
MIR4301
113200
113300
113400
113500
Chromosome 11 (kb)
database: gwascatalog.Nov_2024.rp.txt, refGene.txt.Sep_2018.out

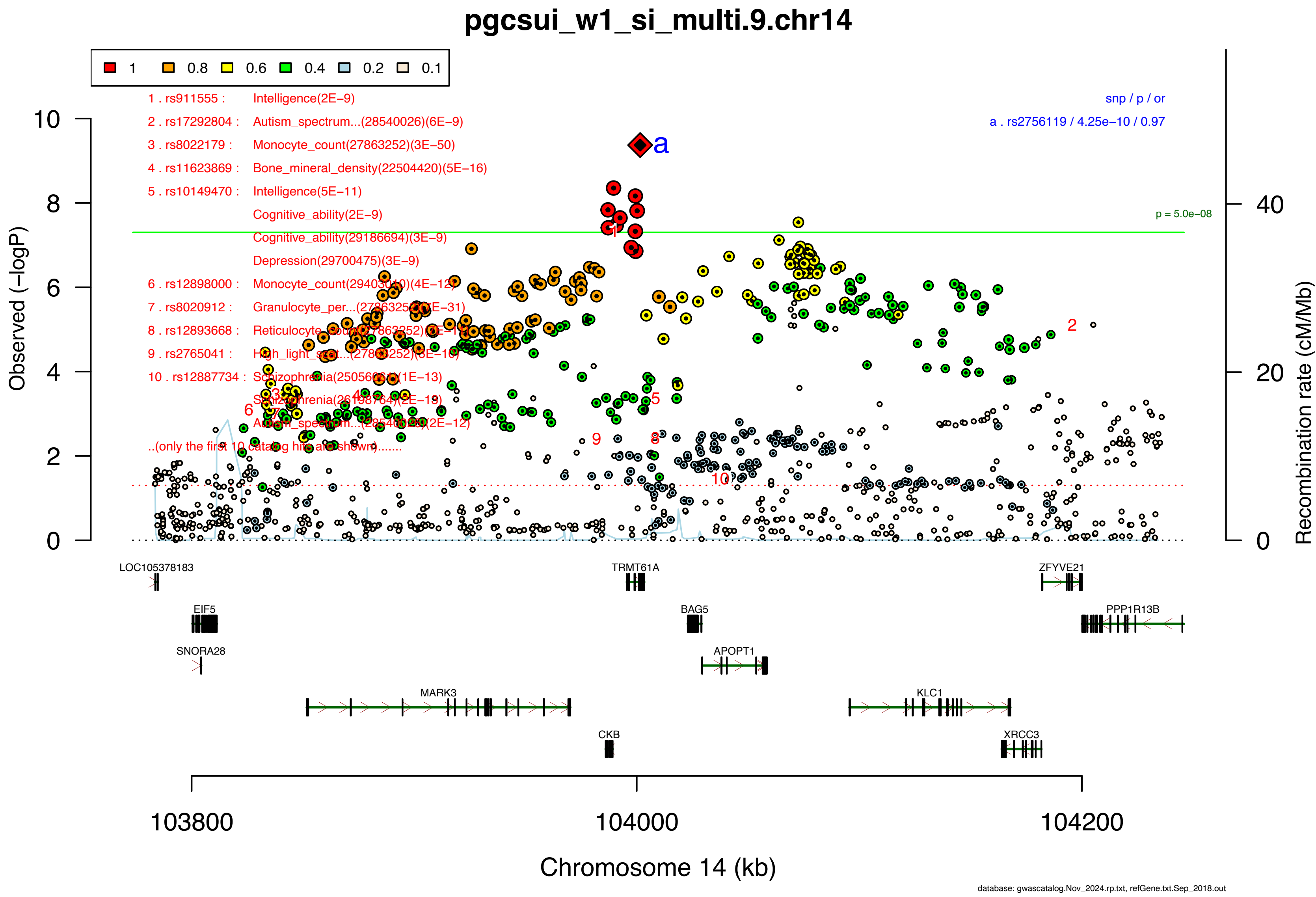
pgcsui_w1_si_multi.9.chr14
1
0.8
0.6
0.4
0.2
0.1
1 . rs911555 : Intelligence(2E−9)
2 . rs17292804 : Autism_spectrum...(28540026)(6E−9)
3 . rs8022179 : Monocyte_count(27863252)(3E−50)
4 . rs11623869 : Bone_mineral_density(22504420)(5E−16)
5 . rs10149470 : Intelligence(5E−11)
Cognitive_ability(2E−9)
Cognitive_ability(29186694)(3E−9)
Depression(29700475)(3E−9)
6 . rs12898000 : Monocyte_count(29403010)(4E−12)
7 . rs8020912 : Granulocyte_per...(27863252)(2E−31)
8 . rs12893668 : Reticulocyte_count(27863252)(2E−14)
9 . rs2765041 : High_light_scat...(27863252)(3E−10)
10 . rs12887734 : Schizophrenia(25056061)(1E−13)
Schizophrenia(26198764)(2E−13)
Autism_spectrum...(28540026)(2E−12)
..(only the first 10 catalog hits are shown).......
snp / p / or
a . rs2756119 / 4.25e−10 / 0.97
p = 5.0e−08
Observed (−logP)
Recombination rate (cM/Mb)
LOC105378183
TRMT61A
ZFYVE21
EIF5
BAG5
PPP1R13B
SNORA28
APOPT1
MARK3
KLC1
CKB
XRCC3
103800
104000
104200
Chromosome 14 (kb)
database: gwascatalog.Nov_2024.rp.txt, refGene.txt.Sep_2018.out

**Supplementary Data 2B: Area plots of lead SNPs at the 10 genome-wide significant loci from the GWAS meta-analysis of suicidal ideation in European ancestry samples.**

If >1 independent SNPs are in the same plot, then each is given a different color, and their LD partners are shaded with the same color. Detailed info about each independent index SNP (a., b., c. ...) is provided in the right upper corner. If SNPs share independent index SNPs as an LD partner, the SNP is assigned to the more significant one. Information regarding other GWAS associations at the locus is provided in the left upper corner.

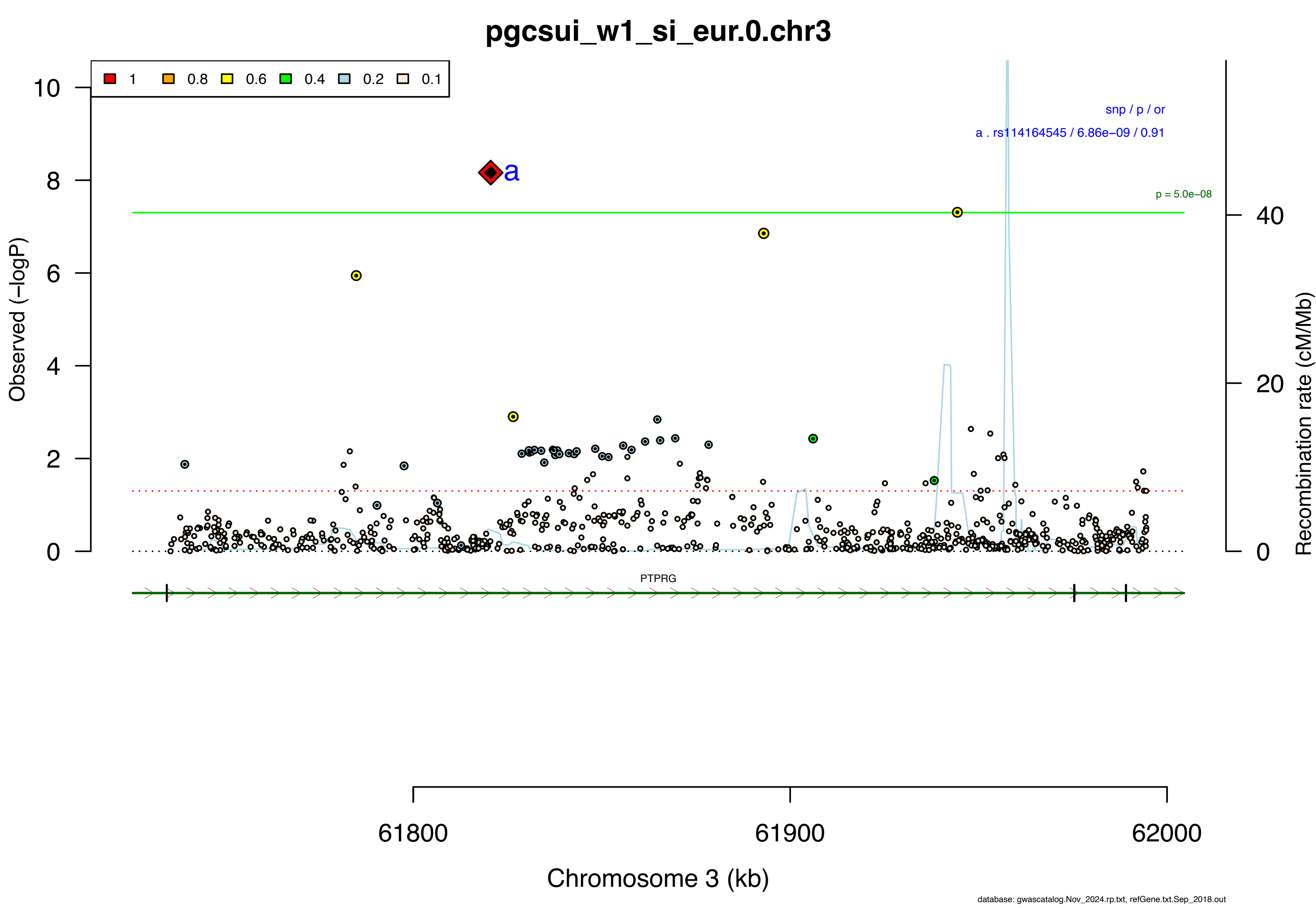

pgcsui_w1_si_eur.0.chr3
1
0.8
0.6
0.4
0.2
0.1
snp / p / or
a . rs114164545 / 6.86e−09 / 0.91
p = 5.0e−08
a
Observed (−logP)
Recombination rate (cM/Mb)
PTPRG
61800
61900
62000
Chromosome 3 (kb)
database: gwascatalog.Nov_2024.rp.txt, refGene.txt.Sep_2018.out

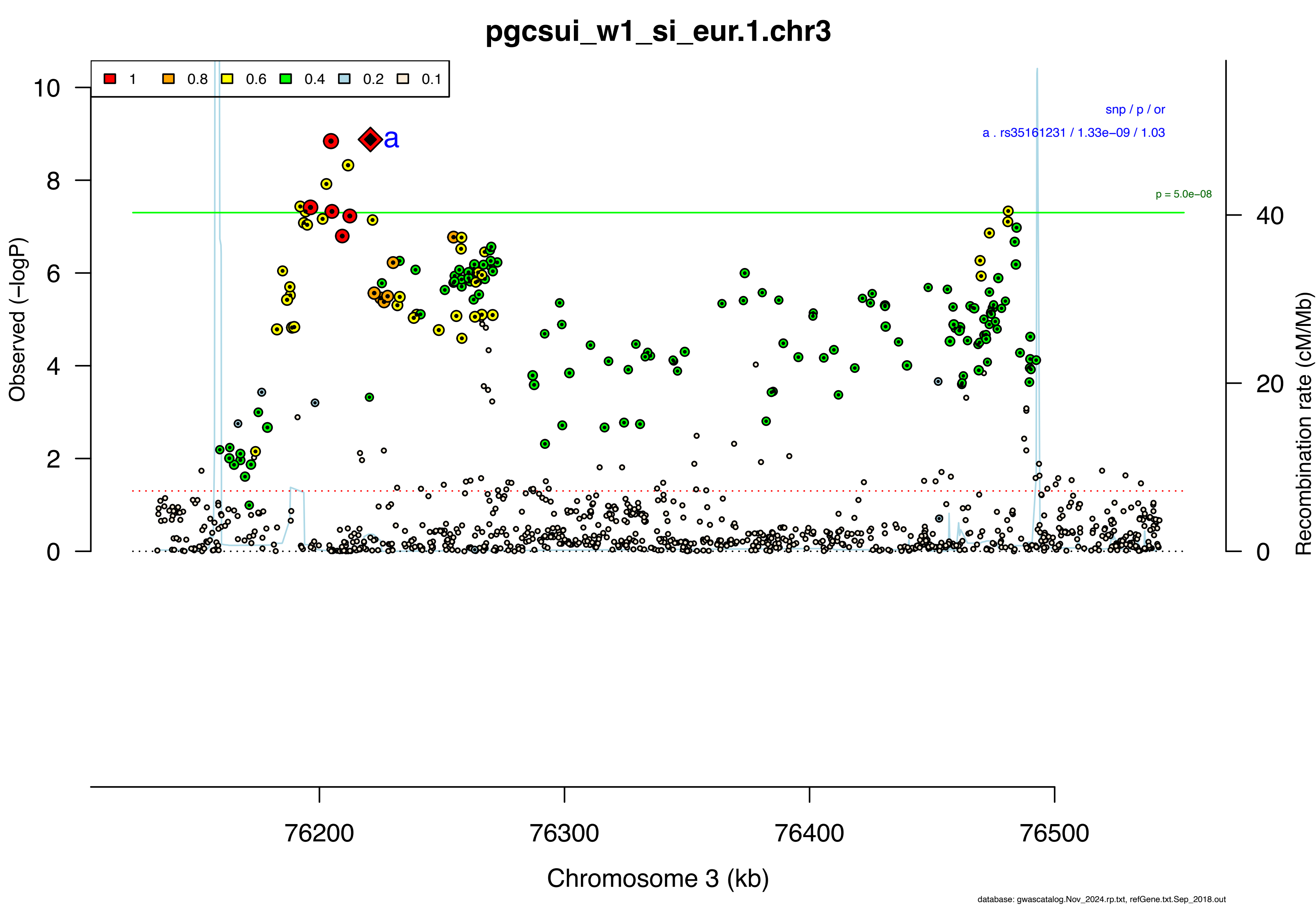

pgcsui_w1_si_eur.1.chr3
1
0.8
0.6
0.4
0.2
0.1
snp / p / or
a . rs35161231 / 1.33e−09 / 1.03
p = 5.0e−08
a
Observed (−logP)
Recombination rate (cM/Mb)
Chromosome 3 (kb)
76200
76300
76400
76500
database: gwascatalog.Nov_2024.rp.txt, refGene.txt.Sep_2018.out

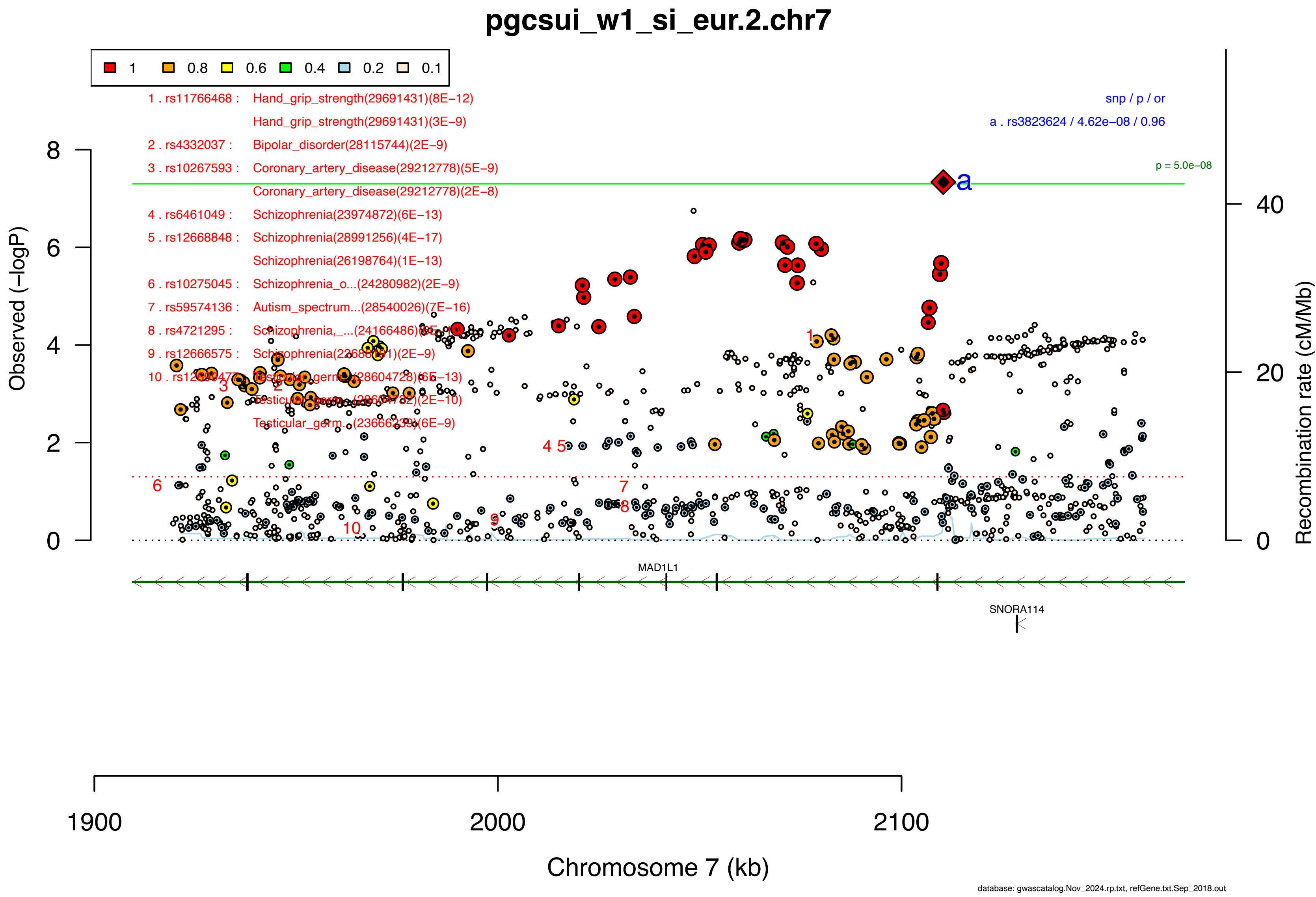
pgcsui_w1_si_eur.2.chr7
1
0.8
0.6
0.4
0.2
0.1
1 . rs11766468 : Hand_grip_strength(29691431)(8E−12)
Hand_grip_strength(29691431)(3E−9)
2 . rs4332037 : Bipolar_disorder(28115744)(2E−9)
3 . rs10267593 : Coronary_artery_disease(29212778)(5E−9)
Coronary_artery_disease(29212778)(2E−8)
4 . rs6461049 : Schizophrenia(23974872)(6E−13)
5 . rs12668848 : Schizophrenia(28991256)(4E−17)
Schizophrenia(26198764)(1E−13)
6 . rs10275045 : Schizophrenia_o...(24280982)(2E−9)
7 . rs59574136 : Autism_spectrum...(28540026)(7E−16)
8 . rs4721295 : Schizophrenia,_...(24166486)(8E−10)
9 . rs12666575 : Schizophrenia(22688191)(2E−9)
10 . rs12699477 : Testicular_germ...(28604728)(6E−13)
Testicular_germ...(28604732)(2E−10)
Testicular_germ...(23666239)(6E−9)
snp / p / or
a . rs3823624 / 4.62e−08 / 0.96
p = 5.0e−08
a
Observed (−logP)
Recombination rate (cM/Mb)
MAD1L1
SNORA114
1900
2000
2100
Chromosome 7 (kb)
database: gwascatalog.Nov_2024.rp.txt, refGene.txt.Sep_2018.out

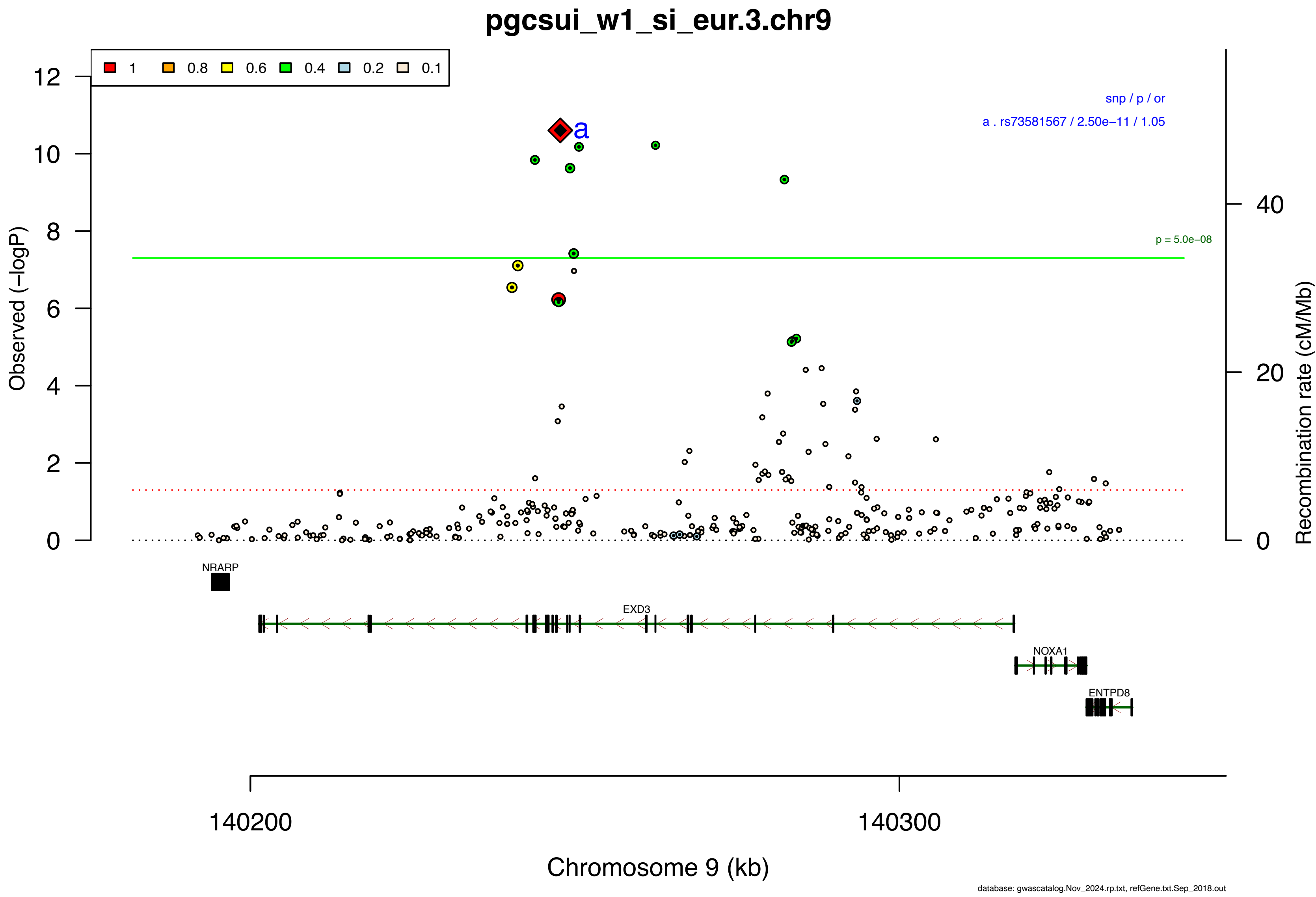

pgcsui_w1_si_eur.3.chr9
1
0.8
0.6
0.4
0.2
0.1
snp / p / or
a . rs73581567 / 2.50e−11 / 1.05
a
p = 5.0e−08
Observed (−logP)
Recombination rate (cM/Mb)
0
2
4
6
8
10
12
20
40
NRARP
EXD3
NOXA1
ENTPD8
140200
140300
Chromosome 9 (kb)
database: gwascatalog.Nov_2024.rp.txt, refGene.txt.Sep_2018.out

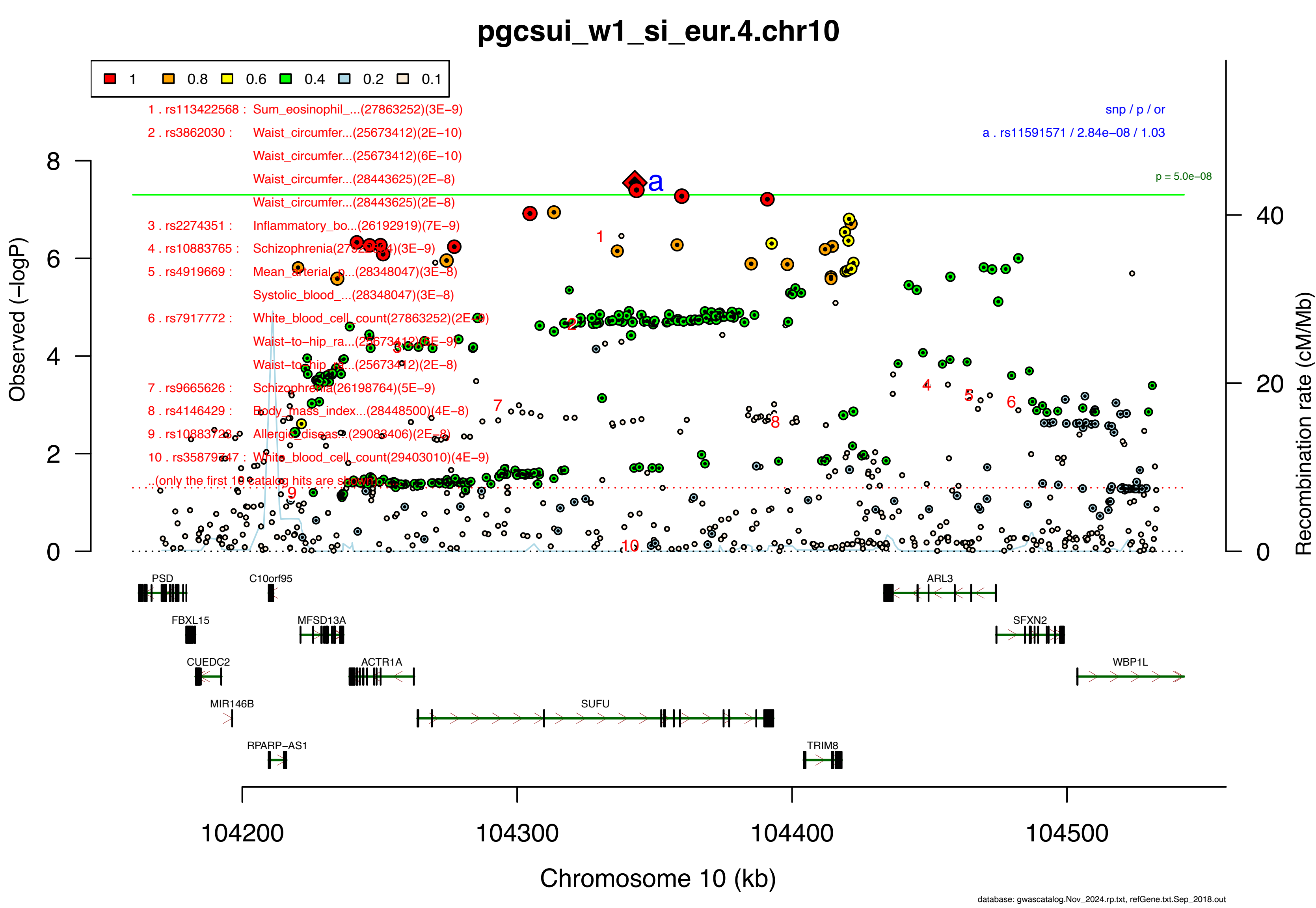

pgcsui_w1_si_eur.4.chr10
1
0.8
0.6
0.4
0.2
0.1
1 . rs113422568 : Sum_eosinophil_...(27863252)(3E−9)
2 . rs3862030 : Waist_circumfer...(25673412)(2E−10)
Waist_circumfer...(25673412)(6E−10)
Waist_circumfer...(28443625)(2E−8)
Waist_circumfer...(28443625)(2E−8)
3 . rs2274351 : Inflammatory_bo...(26192919)(7E−9)
4 . rs10883765 : Schizophrenia(27922834)(3E−9)
5 . rs4919669 : Mean_arterial_p...(28348047)(3E−8)
Systolic_blood_...(28348047)(3E−8)
6 . rs7917772 : White_blood_cell_count(27863252)(2E−9)
Waist−to−hip_ra...(25673412)(6E−9)
Waist−to−hip_ra...(25673412)(2E−8)
7 . rs9665626 : Schizophrenia(26198764)(5E−9)
8 . rs4146429 : Body_mass_index...(28448500)(4E−8)
9 . rs10883723 : Allergic_diseas...(29083406)(2E−8)
10 . rs35879747 : White_blood_cell_count(29403010)(4E−9)
..(only the first 10 catalog hits are shown)
snp / p / or
a . rs11591571 / 2.84e−08 / 1.03
p = 5.0e−08
Observed (−logP)
Recombination rate (cM/Mb)
PSD
C10orf95
ARL3
FBXL15
MFSD13A
SFXN2
CUEDC2
ACTR1A
WBP1L
MIR146B
SUFU
RPARP−AS1
TRIM8
104200
104300
104400
104500
Chromosome 10 (kb)
database: gwascatalog.Nov_2024.rp.txt, refGene.txt.Sep_2018.out

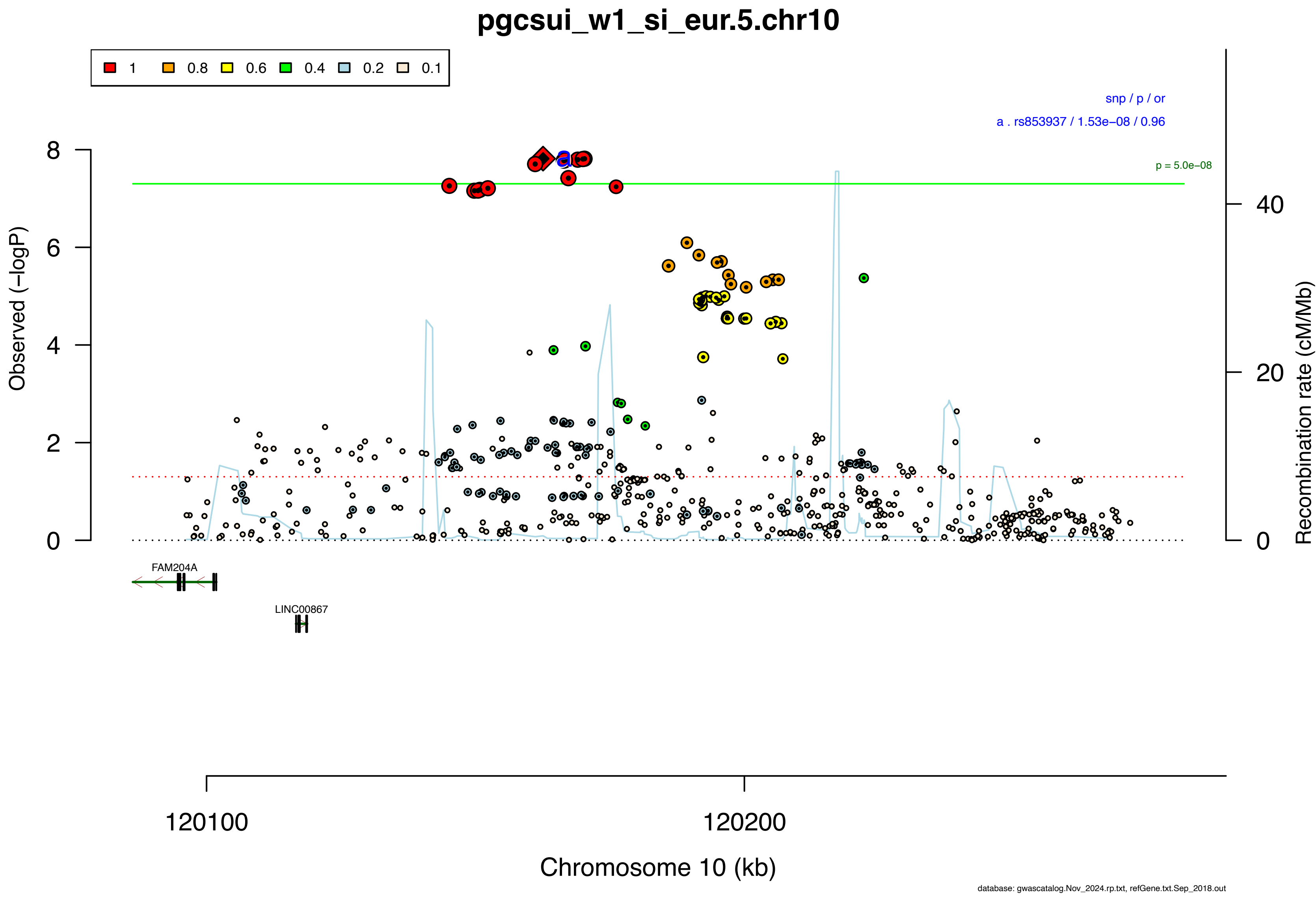

pgcsui_w1_si_eur.5.chr10
1
0.8
0.6
0.4
0.2
0.1
snp / p / or
a . rs853937 / 1.53e−08 / 0.96
p = 5.0e−08
Observed (−logP)
Recombination rate (cM/Mb)
FAM204A
LINC00867
120100
120200
Chromosome 10 (kb)
database: gwascatalog.Nov_2024.rp.txt, refGene.txt.Sep_2018.out

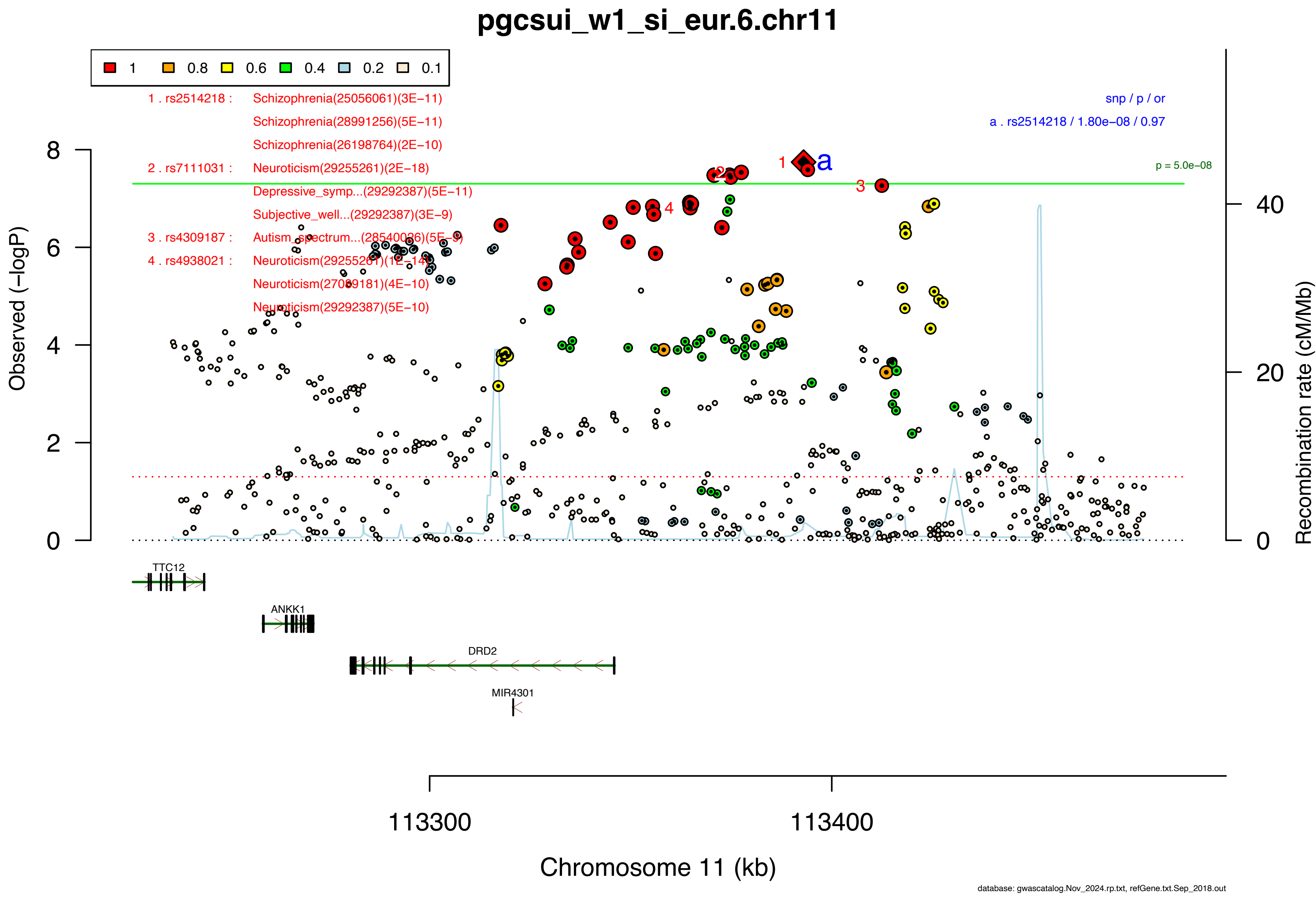
pgcsui_w1_si_eur.6.chr11
1 0.8 0.6 0.4 0.2 0.1
1 . rs2514218 : Schizophrenia(25056061)(3E−11)
Schizophrenia(28991256)(5E−11)
Schizophrenia(26198764)(2E−10)
2 . rs7111031 : Neuroticism(29255261)(2E−18)
Depressive_symp...(29292387)(5E−11)
Subjective_well...(29292387)(3E−9)
3 . rs4309187 : Autism_spectrum...(28540026)(5E−9)
4 . rs4938021 : Neuroticism(29255261)(1E−14)
Neuroticism(27089181)(4E−10)
Neuroticism(29292387)(5E−10)
snp / p / or
a . rs2514218 / 1.80e−08 / 0.97
p = 5.0e−08
Observed (−logP)
Recombination rate (cM/Mb)
TTC12
ANKK1
DRD2
MIR4301
113300
113400
Chromosome 11 (kb)
database: gwascatalog.Nov_2024.rp.txt, refGene.txt.Sep_2018.out

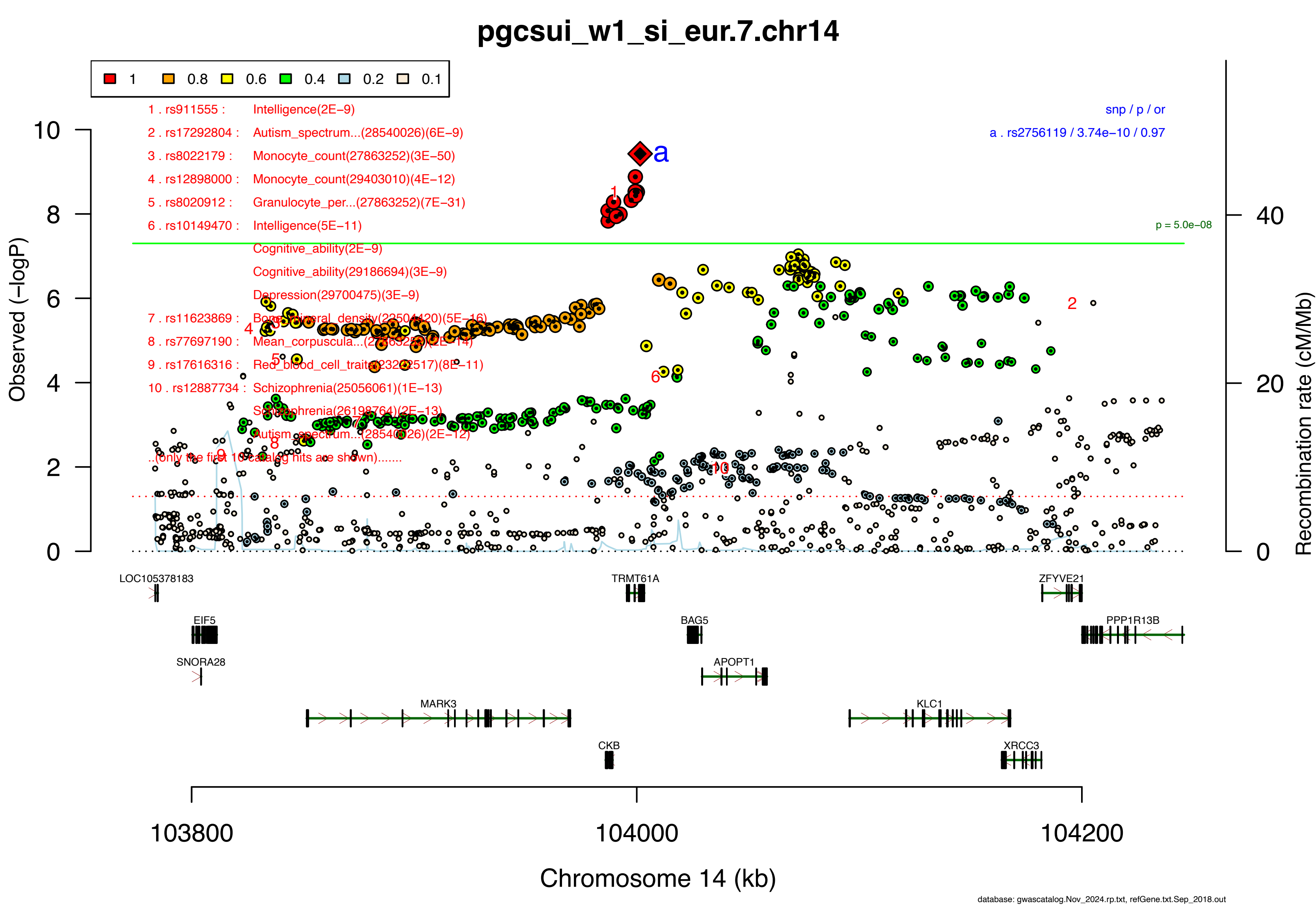
pgcsui_w1_si_eur.7.chr14
1
0.8
0.6
0.4
0.2
0.1
1 . rs911555 : Intelligence(2E−9)
2 . rs17292804 : Autism_spectrum...(28540026)(6E−9)
3 . rs8022179 : Monocyte_count(27863252)(3E−50)
4 . rs12898000 : Monocyte_count(29403010)(4E−12)
5 . rs8020912 : Granulocyte_per...(27863252)(7E−31)
6 . rs10149470 : Intelligence(5E−11)
Cognitive_ability(2E−9)
Cognitive_ability(29186694)(3E−9)
Depression(29700475)(3E−9)
7 . rs11623869 : Bone_mineral_density(22504420)(5E−16)
8 . rs77697190 : Mean_corpuscula...(27863252)(2E−14)
9 . rs17616316 : Red_blood_cell_traits(23222517)(8E−11)
10 . rs12887734 : Schizophrenia(25056061)(1E−13)
Schizophrenia(26198764)(2E−13)
Autism_spectrum...(28540026)(2E−12)
..(only the first 10 catalog hits are shown).......
snp / p / or
a . rs2756119 / 3.74e−10 / 0.97
p = 5.0e−08
Observed (−logP)
Recombination rate (cM/Mb)
LOC105378183
TRMT61A
ZFYVE21
EIF5
BAG5
PPP1R13B
SNORA28
APOPT1
MARK3
KLC1
CKB
XRCC3
103800
104000
104200
Chromosome 14 (kb)
database: gwascatalog.Nov_2024.rp.txt, refGene.txt.Sep_2018.out

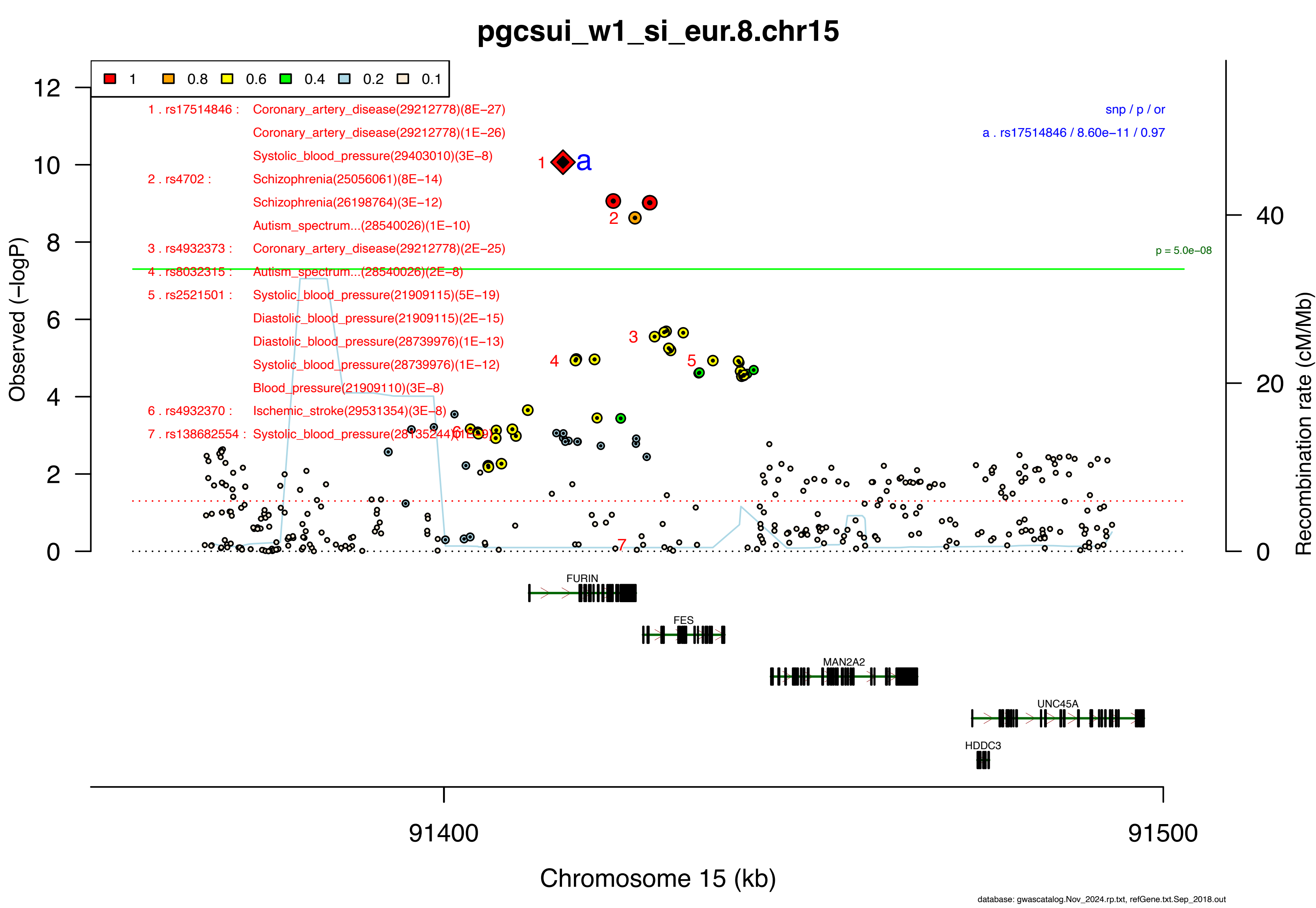

pgcsui_w1_si_eur.8.chr15
1
0.8
0.6
0.4
0.2
0.1
1 . rs17514846 : Coronary_artery_disease(29212778)(8E−27)
Coronary_artery_disease(29212778)(1E−26)
Systolic_blood_pressure(29403010)(3E−8)
2 . rs4702 : Schizophrenia(25056061)(8E−14)
Schizophrenia(26198764)(3E−12)
Autism_spectrum...(28540026)(1E−10)
3 . rs4932373 : Coronary_artery_disease(29212778)(2E−25)
4 . rs8032315 : Autism_spectrum...(28540026)(2E−8)
5 . rs2521501 : Systolic_blood_pressure(21909115)(5E−19)
Diastolic_blood_pressure(21909115)(2E−15)
Diastolic_blood_pressure(28739976)(1E−13)
Systolic_blood_pressure(28739976)(1E−12)
Blood_pressure(21909110)(3E−8)
6 . rs4932370 : Ischemic_stroke(29531354)(3E−8)
7 . rs138682554 : Systolic_blood_pressure(28135244)(1E−9)
snp / p / or
a . rs17514846 / 8.60e−11 / 0.97
p = 5.0e−08
Observed (−logP)
Recombination rate (cM/Mb)
FURIN
FES
MAN2A2
UNC45A
HDDC3
91400
91500
Chromosome 15 (kb)
database: gwascatalog.Nov_2024.rp.txt, refGene.txt.Sep_2018.out

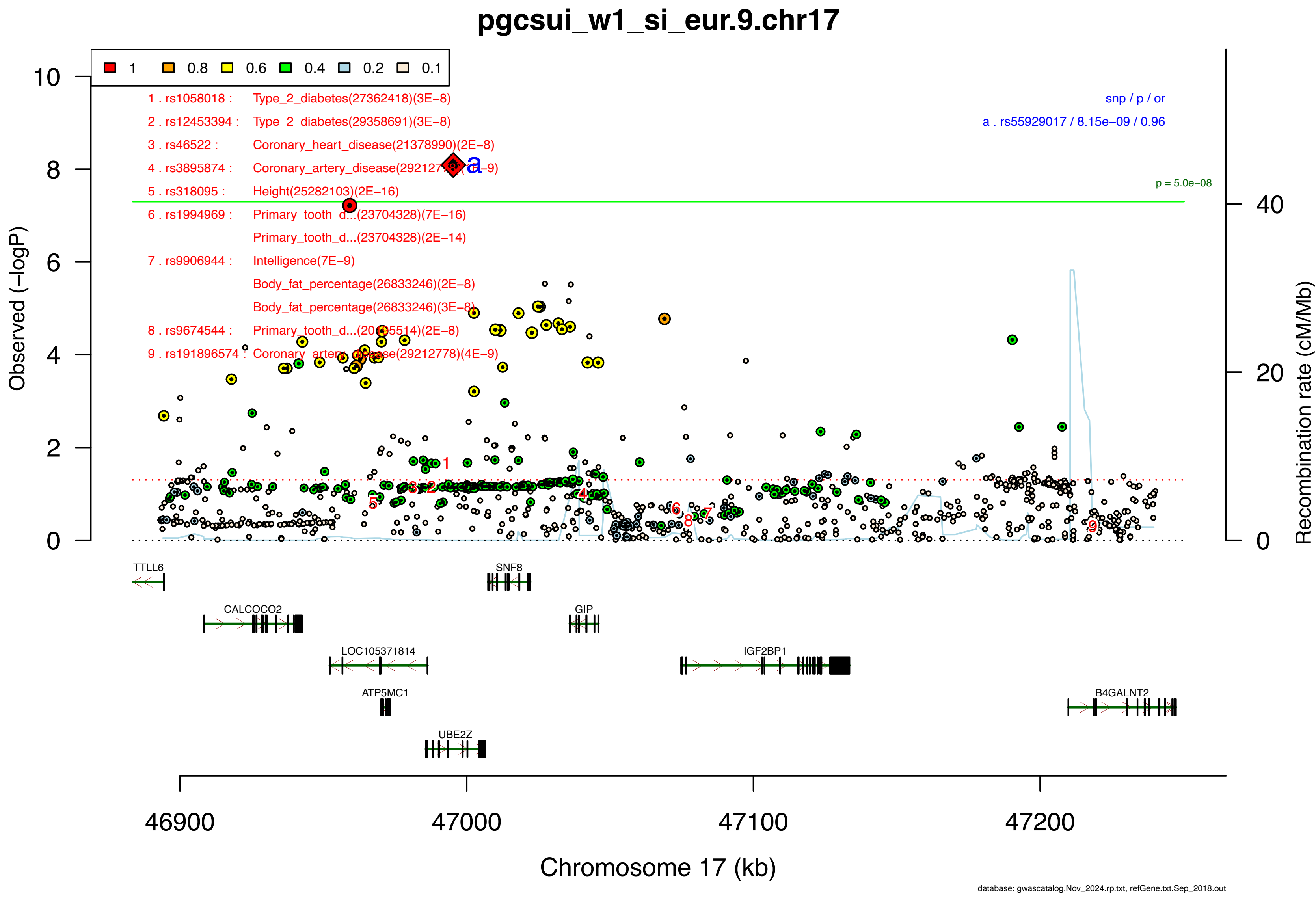

pgcsui_w1_si_eur.9.chr17
1
0.8
0.6
0.4
0.2
0.1
1 . rs1058018 : Type_2_diabetes(27362418)(3E−8)
2 . rs12453394 : Type_2_diabetes(29358691)(3E−8)
3 . rs46522 : Coronary_heart_disease(21378990)(2E−8)
4 . rs3895874 : Coronary_artery_disease(29212778)(1E−9)
5 . rs318095 : Height(25282103)(2E−16)
6 . rs1994969 : Primary_tooth_d...(23704328)(7E−16)
Primary_tooth_d...(23704328)(2E−14)
7 . rs9906944 : Intelligence(7E−9)
Body_fat_percentage(26833246)(2E−8)
Body_fat_percentage(26833246)(3E−8)
8 . rs9674544 : Primary_tooth_d...(20195514)(2E−8)
9 . rs191896574 : Coronary_artery_disease(29212778)(4E−9)
snp / p / or
a . rs55929017 / 8.15e−09 / 0.96
p = 5.0e−08
Observed (−logP)
Recombination rate (cM/Mb)
TTLL6
SNF8
CALCOCO2
GIP
LOC105371814
IGF2BP1
ATP5MC1
B4GALNT2
UBE2Z
46900
47000
47100
47200
Chromosome 17 (kb)
database: gwascatalog.Nov_2024.rp.txt, refGene.txt.Sep_2018.out

**Supplementary Data 2C: Area plot of the lead SNP at the genome-wide significant locus from the GWAS meta-analysis of suicidal ideation in East Asian ancestry samples.** Detailed info about each independent index SNP (a., b., c. ...) is provided in the right upper corner. If SNPs share independent index SNPs as an LD partner, the SNP is assigned to the more significant one. Information regarding other GWAS associations at the locus is provided in the left upper corner.

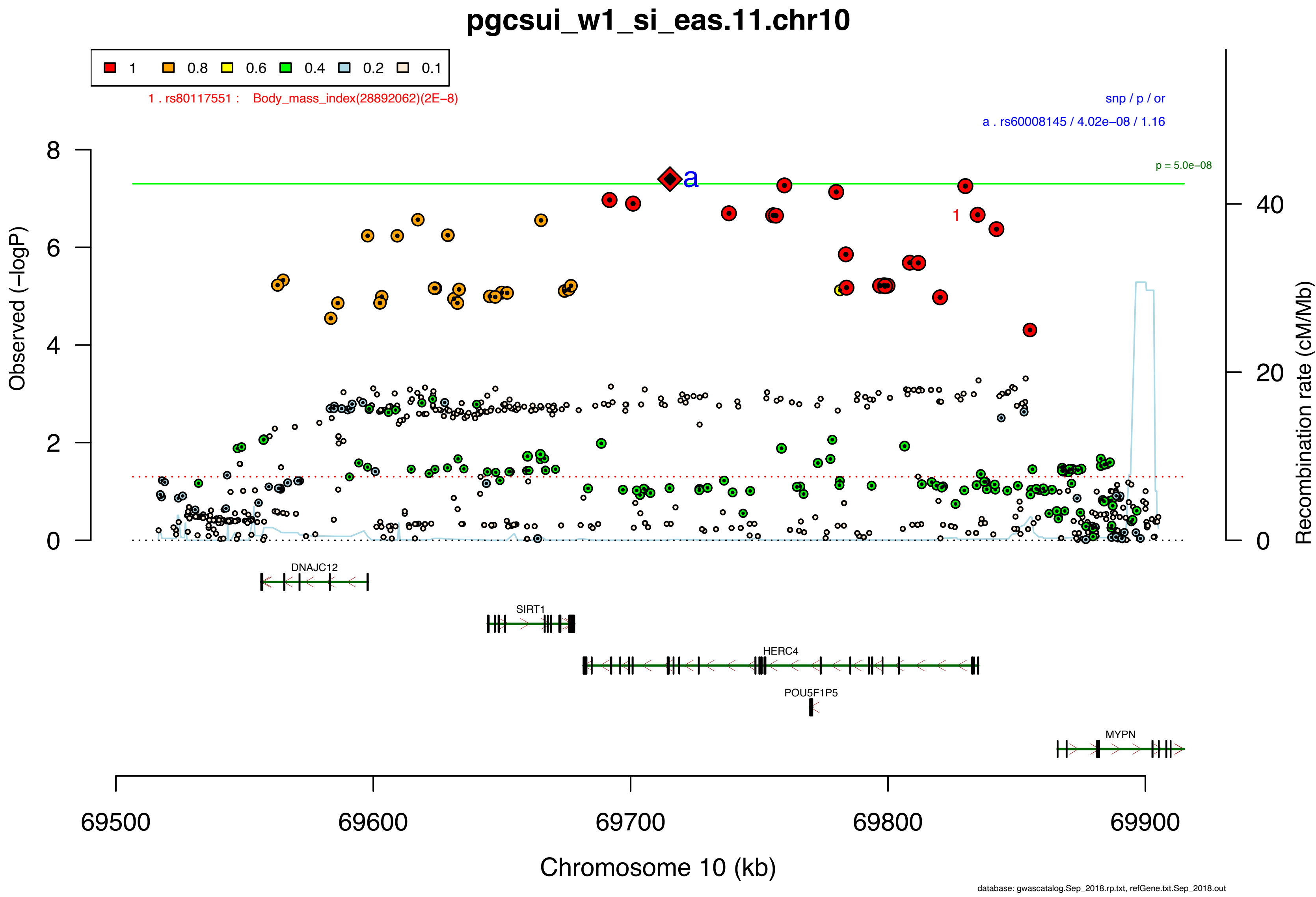
pgcsui_w1_si_eas.11.chr10
1
0.8
0.6
0.4
0.2
0.1
1 . rs80117551 : Body_mass_index(28892062)(2E−8)
snp / p / or
a . rs60008145 / 4.02e−08 / 1.16
p = 5.0e−08
a
1
Observed (−logP)
Recombination rate (cM/Mb)
0
2
4
6
8
0
20
40
DNAJC12
SIRT1
HERC4
POU5F1P5
MYPN
69500
69600
69700
69800
69900
Chromosome 10 (kb)
database: gwascatalog.Sep_2018.rp.txt, refGene.txt.Sep_2018.out

**Supplementary Data 2D: Area plots of lead SNPs at the 37 genome-wide significant loci from the multi-ancestry GWAS meta-analysis of suicide attempt.**

If >1 independent SNPs are in the same plot, then each is given a different color, and their LD partners are shaded with the same color. Detailed info about each independent index SNP (a., b., c. ...) is provided in the right upper corner. If SNPs share independent index SNPs as an LD partner, the SNP is assigned to the more significant one. Information regarding other GWAS associations at the locus is provided in the left upper corner.

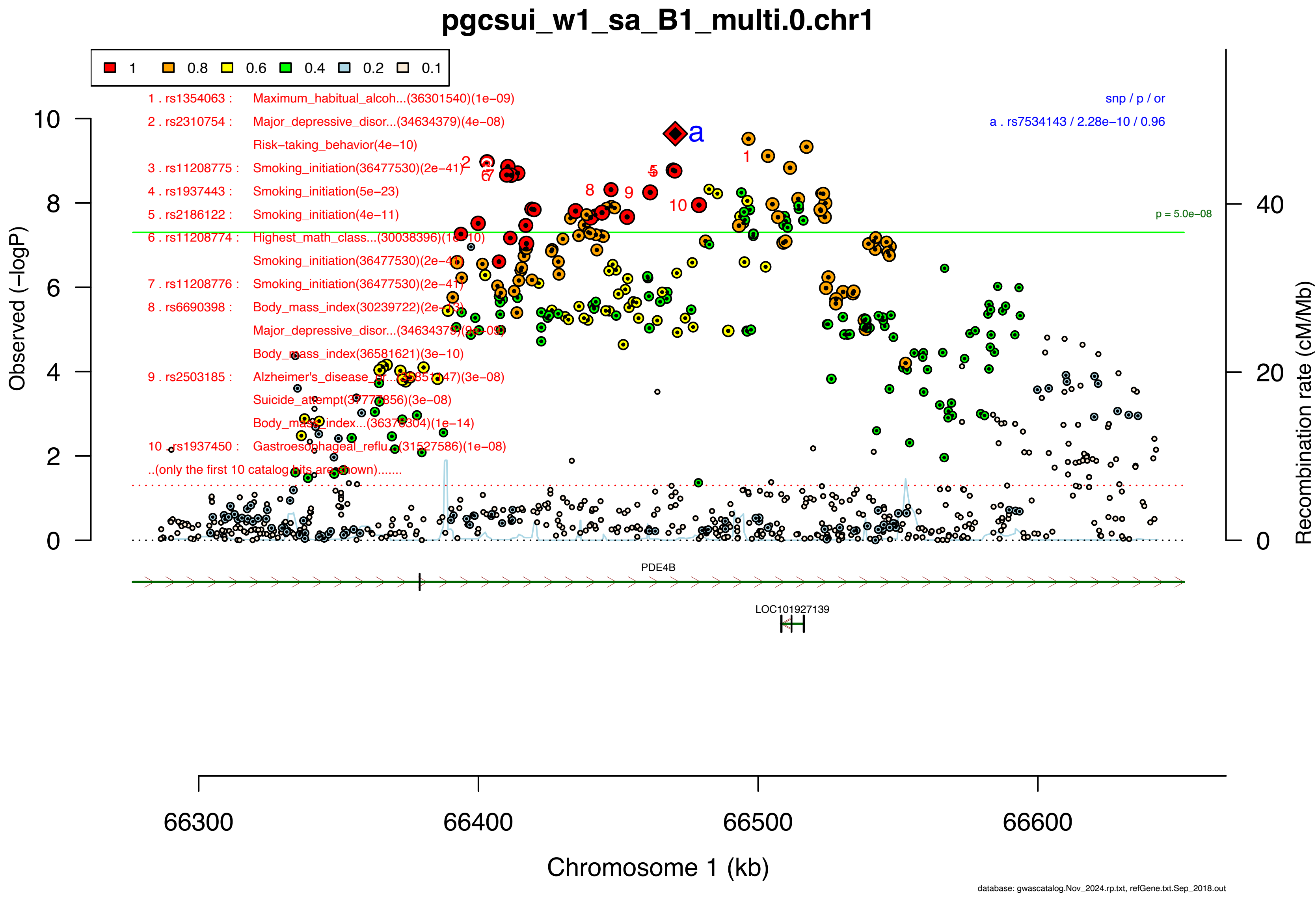
pgcsui_w1_sa_B1_multi.0.chr1
1
0.8
0.6
0.4
0.2
0.1
1 . rs1354063 : Maximum_habitual_alcoh...(36301540)(1e−09)
2 . rs2310754 : Major_depressive_disor...(34634379)(4e−08)
Risk−taking_behavior(4e−10)
3 . rs11208775 : Smoking_initiation(36477530)(2e−41)
4 . rs1937443 : Smoking_initiation(5e−23)
5 . rs2186122 : Smoking_initiation(4e−11)
6 . rs11208774 : Highest_math_class...(30038396)(1e−10)
Smoking_initiation(36477530)(2e−41)
7 . rs11208776 : Smoking_initiation(36477530)(2e−41)
8 . rs6690398 : Body_mass_index(30239722)(2e−13)
Major_depressive_disor...(34634379)(9e−09)
Body_mass_index(36581621)(3e−10)
9 . rs2503185 : Alzheimer's_disease_or...(35851147)(3e−08)
Suicide_attempt(37777856)(3e−08)
Body_mass_index...(36376304)(1e−14)
10 . rs1937450 : Gastroesophageal_reflu...(31527586)(1e−08)
..(only the first 10 catalog hits are shown).......
snp / p / or
a . rs7534143 / 2.28e−10 / 0.96
p = 5.0e−08
Observed (−logP)
Recombination rate (cM/Mb)
PDE4B
LOC101927139
66300
66400
66500
66600
Chromosome 1 (kb)
database: gwascatalog.Nov_2024.rp.txt, refGene.txt.Sep_2018.out

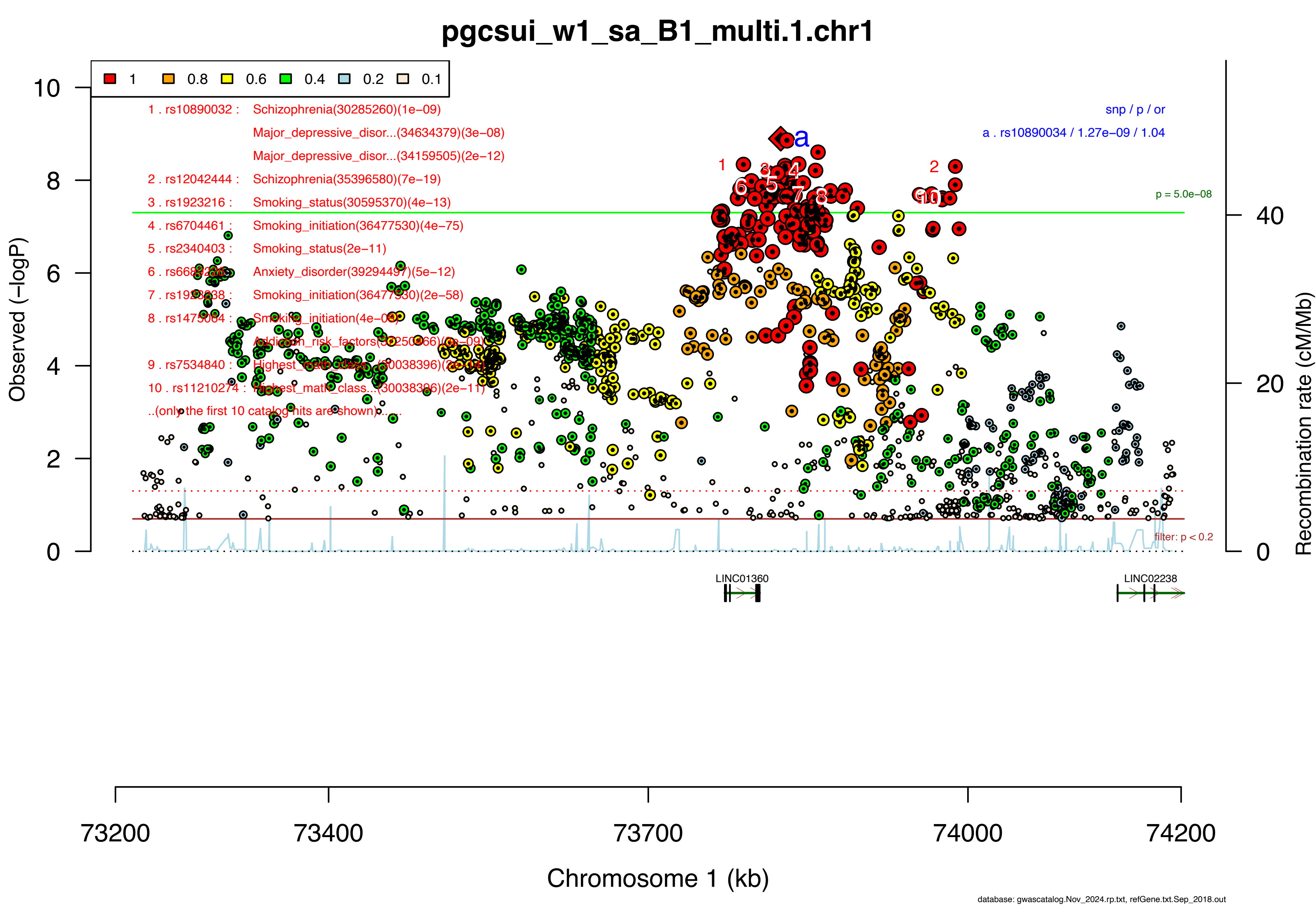

pgcsui_w1_sa_B1_multi.1.chr1
1 0.8 0.6 0.4 0.2 0.1
1 . rs10890032 : Schizophrenia(30285260)(1e−09)
Major_depressive_disor...(34634379)(3e−08)
Major_depressive_disor...(34159505)(2e−12)
2 . rs12042444 : Schizophrenia(35396580)(7e−19)
3 . rs1923216 : Smoking_status(30595370)(4e−13)
4 . rs6704461 : Smoking_initiation(36477530)(4e−75)
5 . rs2340403 : Smoking_status(2e−11)
6 . rs66[illegible] : Anxiety_disorder(39294497)(5e−12)
7 . rs19[illegible] : Smoking_initiation(36477530)(2e−58)
8 . rs14[illegible] : Smoking_initiation(4e−08)
Addiction_risk_factors(3[illegible])(6e−09)
9 . rs7534840 : Highest_math_class...(30038396)([illegible])
10 . rs11210274 : Highest_math_class...(30038396)(2e−11)
..(only the first 10 catalog hits are shown)......
snp / p / or
a . rs10890034 / 1.27e−09 / 1.04
p = 5.0e−08
filter: p < 0.2
Observed (−logP)
Recombination rate (cM/Mb)
LINC01360
LINC02238
73200 73400 73700 74000 74200
Chromosome 1 (kb)
database: gwascatalog.Nov_2024.rp.txt, refGene.txt.Sep_2018.out

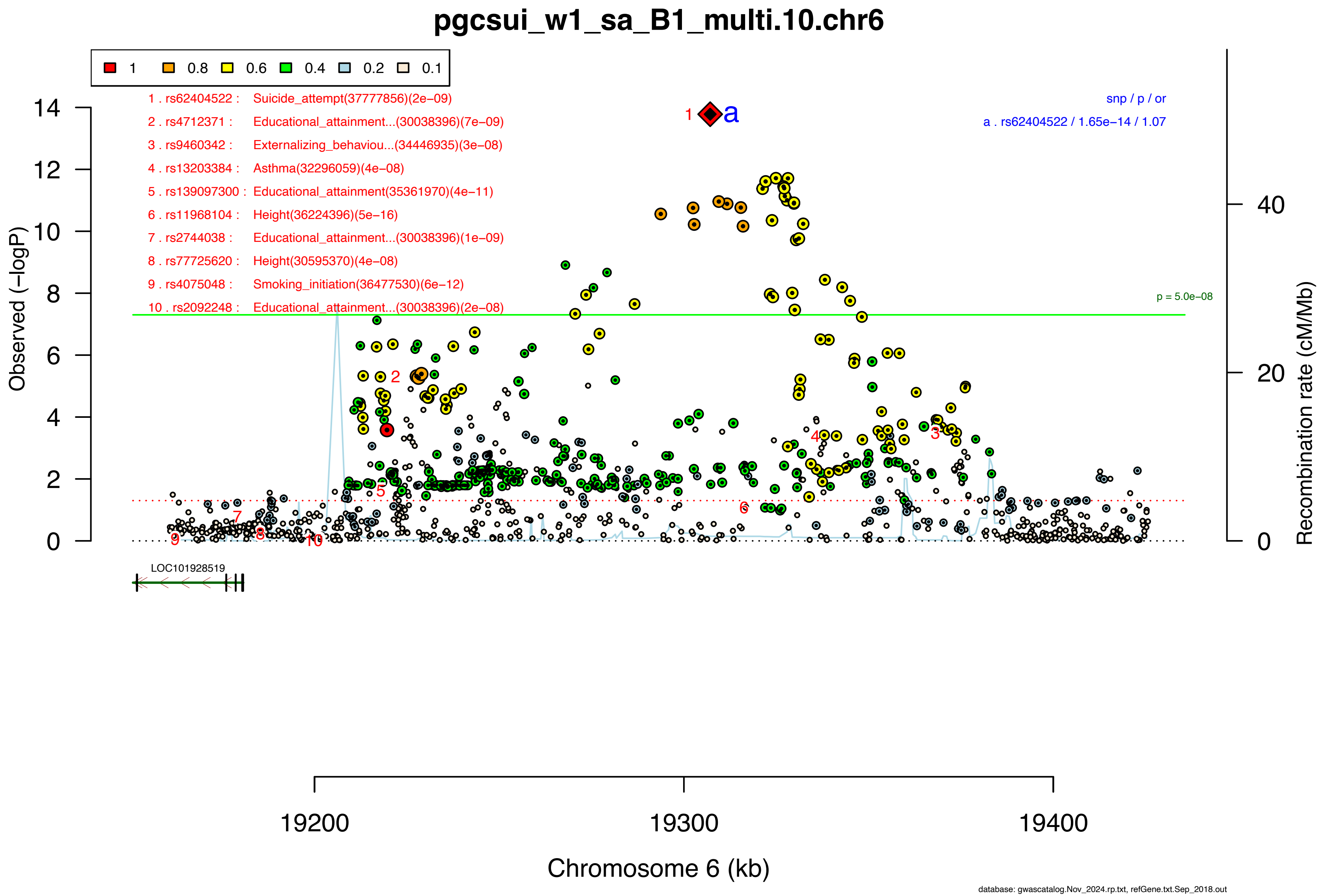

pgcsui_w1_sa_B1_multi.10.chr6
1
0.8
0.6
0.4
0.2
0.1
1 . rs62404522 : Suicide_attempt(37777856)(2e−09)
2 . rs4712371 : Educational_attainment...(30038396)(7e−09)
3 . rs9460342 : Externalizing_behaviou...(34446935)(3e−08)
4 . rs13203384 : Asthma(32296059)(4e−08)
5 . rs139097300 : Educational_attainment(35361970)(4e−11)
6 . rs11968104 : Height(36224396)(5e−16)
7 . rs2744038 : Educational_attainment...(30038396)(1e−09)
8 . rs77725620 : Height(30595370)(4e−08)
9 . rs4075048 : Smoking_initiation(36477530)(6e−12)
10 . rs2092248 : Educational_attainment...(30038396)(2e−08)
snp / p / or
a . rs62404522 / 1.65e−14 / 1.07
p = 5.0e−08
Observed (−logP)
Recombination rate (cM/Mb)
LOC101928519
19200
19300
19400
Chromosome 6 (kb)
database: gwascatalog.Nov_2024.rp.txt, refGene.txt.Sep_2018.out

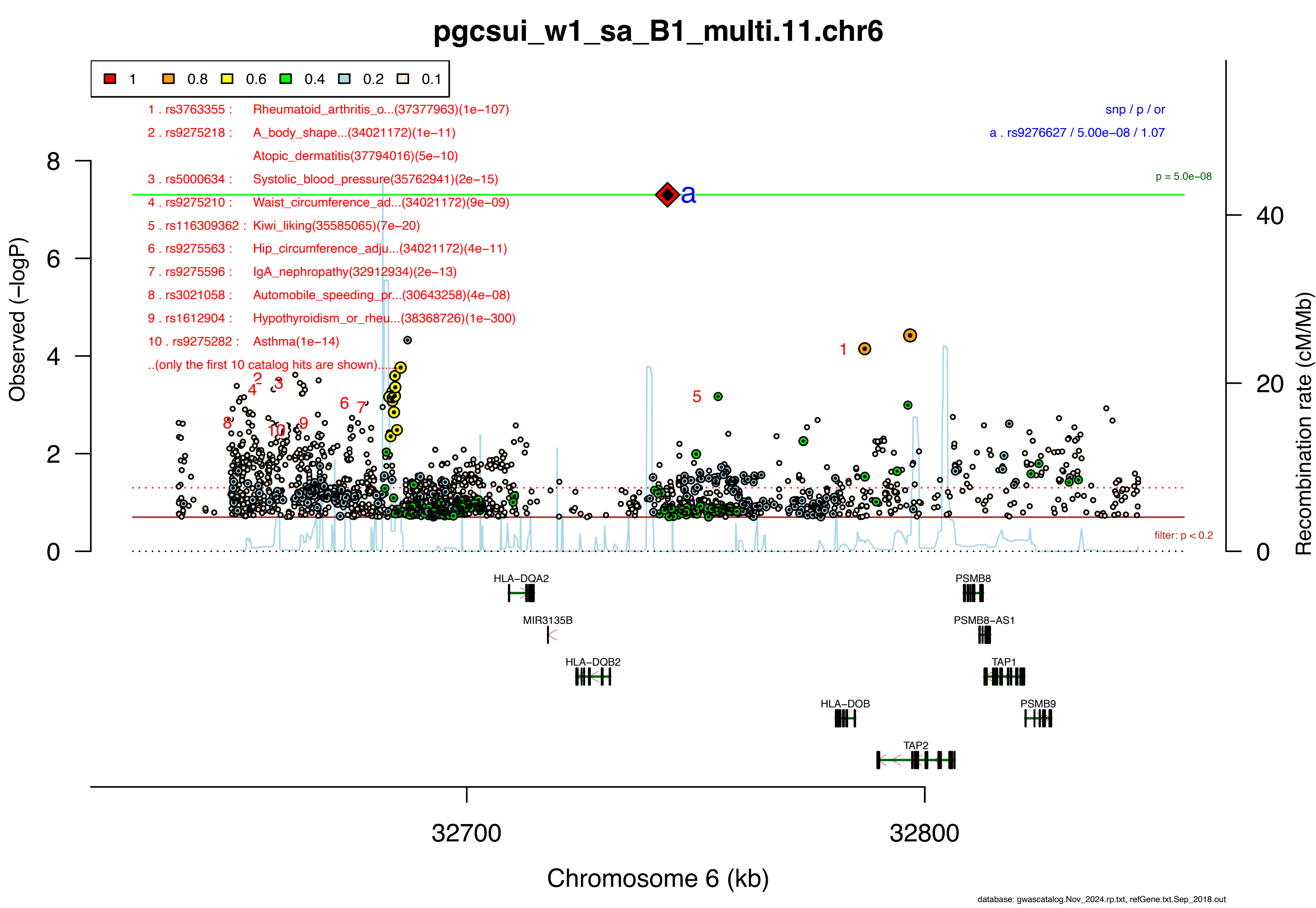

pgcsui_w1_sa_B1_multi.11.chr6
1
0.8
0.6
0.4
0.2
0.1
1 . rs3763355 : Rheumatoid_arthritis_o...(37377963)(1e−107)
2 . rs9275218 : A_body_shape...(34021172)(1e−11)
Atopic_dermatitis(37794016)(5e−10)
3 . rs5000634 : Systolic_blood_pressure(35762941)(2e−15)
4 . rs9275210 : Waist_circumference_ad...(34021172)(9e−09)
5 . rs116309362 : Kiwi_liking(35585065)(7e−20)
6 . rs9275563 : Hip_circumference_adju...(34021172)(4e−11)
7 . rs9275596 : IgA_nephropathy(32912934)(2e−13)
8 . rs3021058 : Automobile_speeding_pr...(30643258)(4e−08)
9 . rs1612904 : Hypothyroidism_or_rheu...(38368726)(1e−300)
10 . rs9275282 : Asthma(1e−14)
..(only the first 10 catalog hits are shown)...
snp / p / or
a . rs9276627 / 5.00e−08 / 1.07
p = 5.0e−08
filter: p < 0.2
Observed (−logP)
Recombination rate (cM/Mb)
HLA−DQA2
MIR3135B
HLA−DQB2
HLA−DOB
TAP2
PSMB8
PSMB8−AS1
TAP1
PSMB9
32700
32800
Chromosome 6 (kb)
database: gwascatalog.Nov_2024.rp.txt, refGene.txt.Sep_2018.out

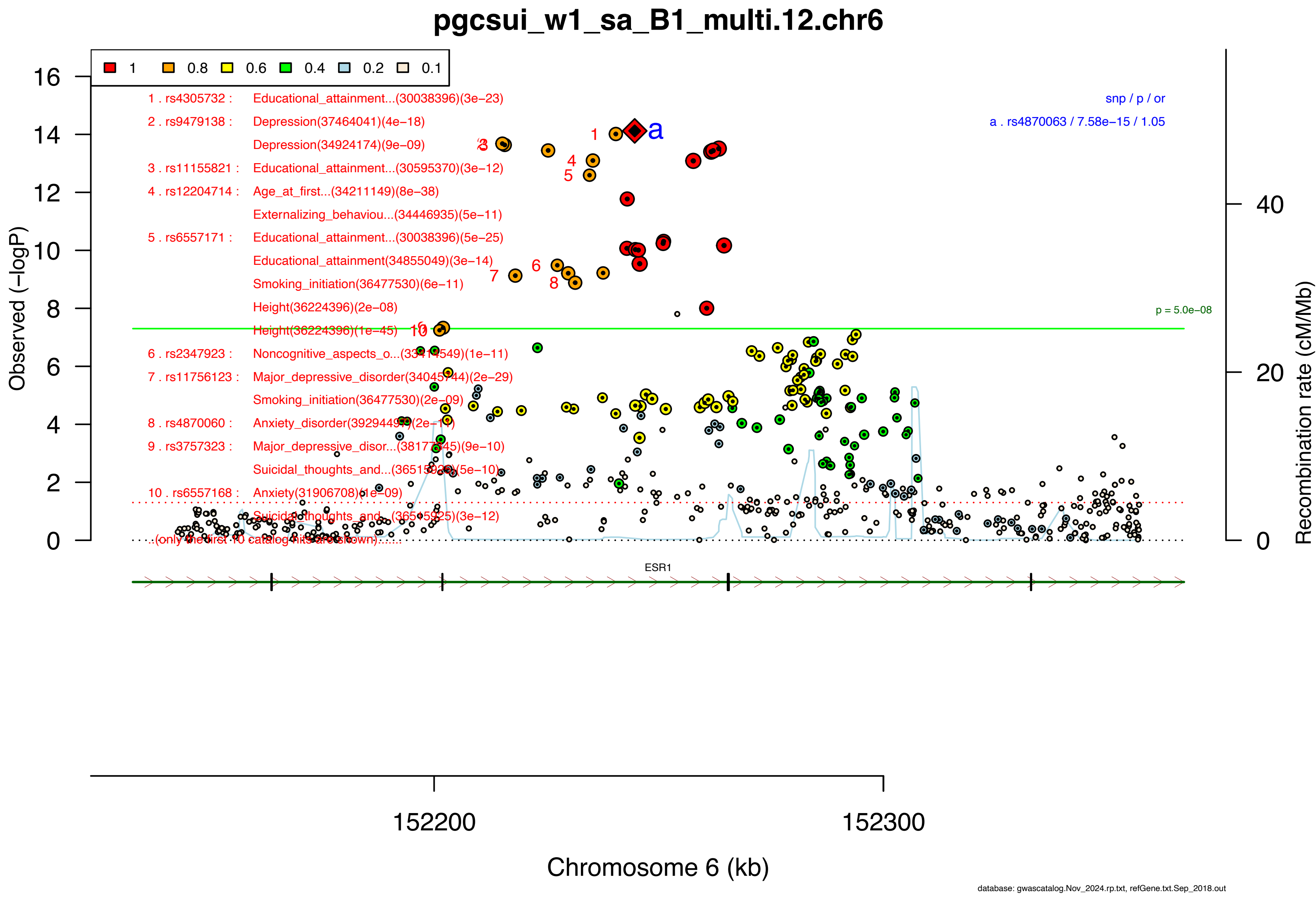
pgcsui_w1_sa_B1_multi.12.chr6
1 0.8 0.6 0.4 0.2 0.1
1 . rs4305732 : Educational_attainment...(30038396)(3e−23)
2 . rs9479138 : Depression(37464041)(4e−18)
Depression(34924174)(9e−09)
3 . rs11155821 : Educational_attainment...(30595370)(3e−12)
4 . rs12204714 : Age_at_first...(34211149)(8e−38)
Externalizing_behaviou...(34446935)(5e−11)
5 . rs6557171 : Educational_attainment...(30038396)(5e−25)
Educational_attainment(34855049)(3e−14)
Smoking_initiation(36477530)(6e−11)
Height(36224396)(2e−08)
Height(36224396)(1e−45)
6 . rs2347923 : Noncognitive_aspects_o...(33414549)(1e−11)
7 . rs11756123 : Major_depressive_disorder(34045744)(2e−29)
Smoking_initiation(36477530)(2e−09)
8 . rs4870060 : Anxiety_disorder(39294497)(2e−11)
9 . rs3757323 : Major_depressive_disor...(38177345)(9e−10)
Suicidal_thoughts_and...(36515925)(5e−10)
10 . rs6557168 : Anxiety(31906708)(1e−09)
Suicidal_thoughts_and...(36515925)(3e−12)
..(only the first 10 catalog hits are shown)......
snp / p / or
a . rs4870063 / 7.58e−15 / 1.05
p = 5.0e−08
Observed (−logP)
Recombination rate (cM/Mb)
ESR1
152200
152300
Chromosome 6 (kb)
database: gwascatalog.Nov_2024.rp.txt, refGene.txt.Sep_2018.out

### pgcsui_w1_sa_B1_multi.13.chr6

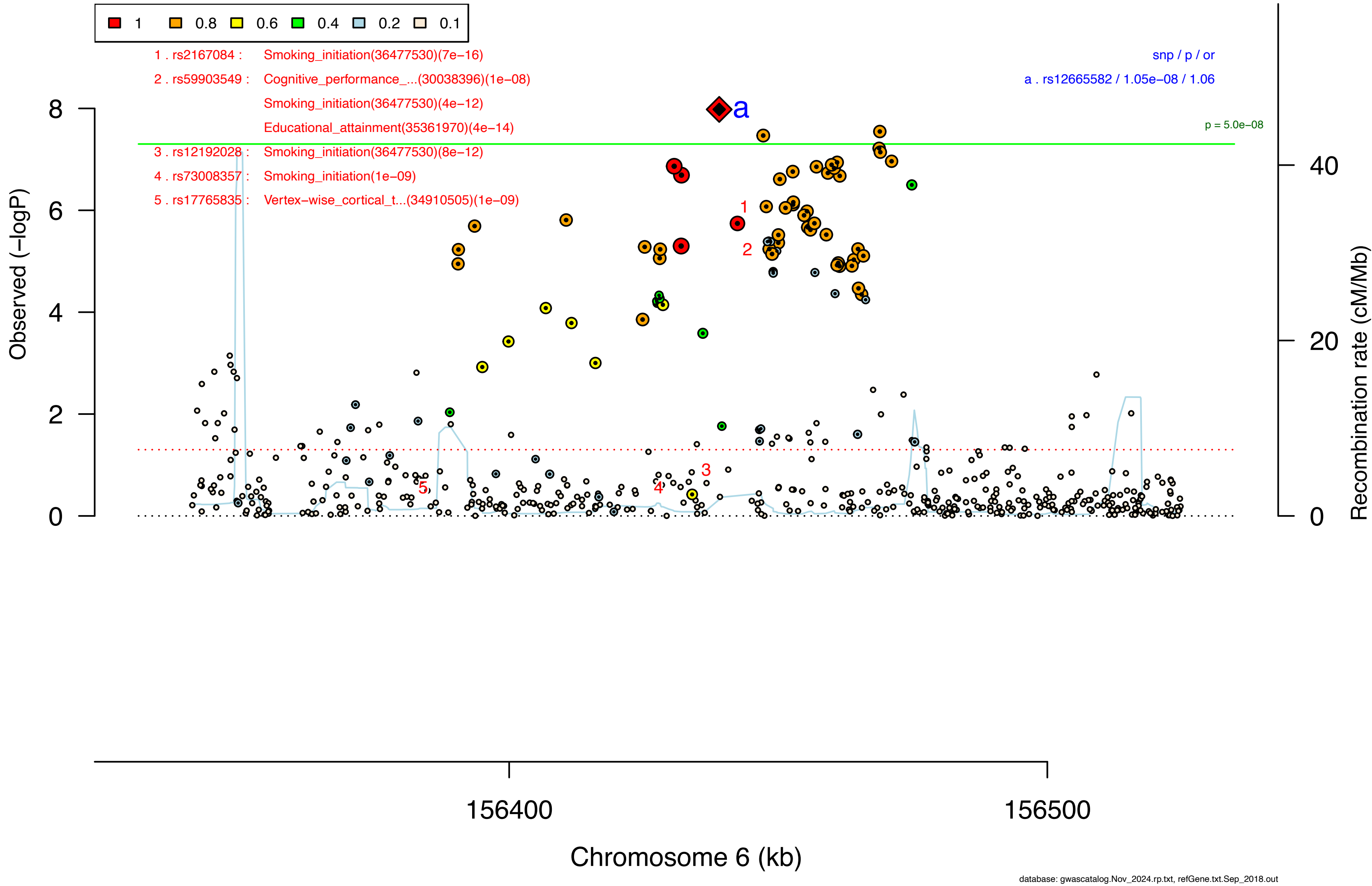


database: gwascatalog.Nov_2024.rp.txt, refGene.txt.Sep_2018.out

### pgcsui_w1_sa_B1_multi.14.chr7

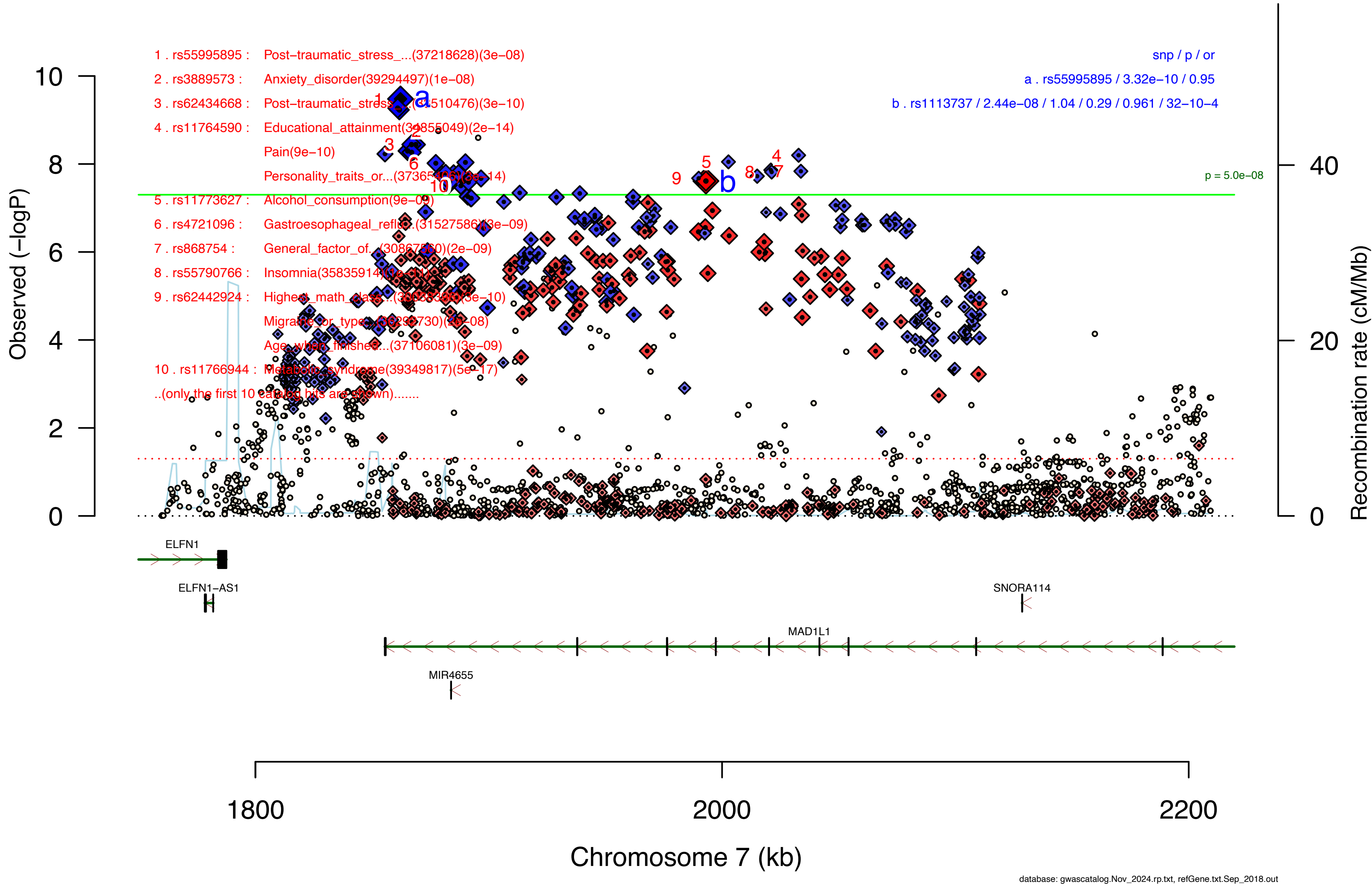


database: gwascatalog.Nov_2024.rp.txt, refGene.txt.Sep_2018.out

#### pgcsui_w1_sa_B1_multi.15.chr7

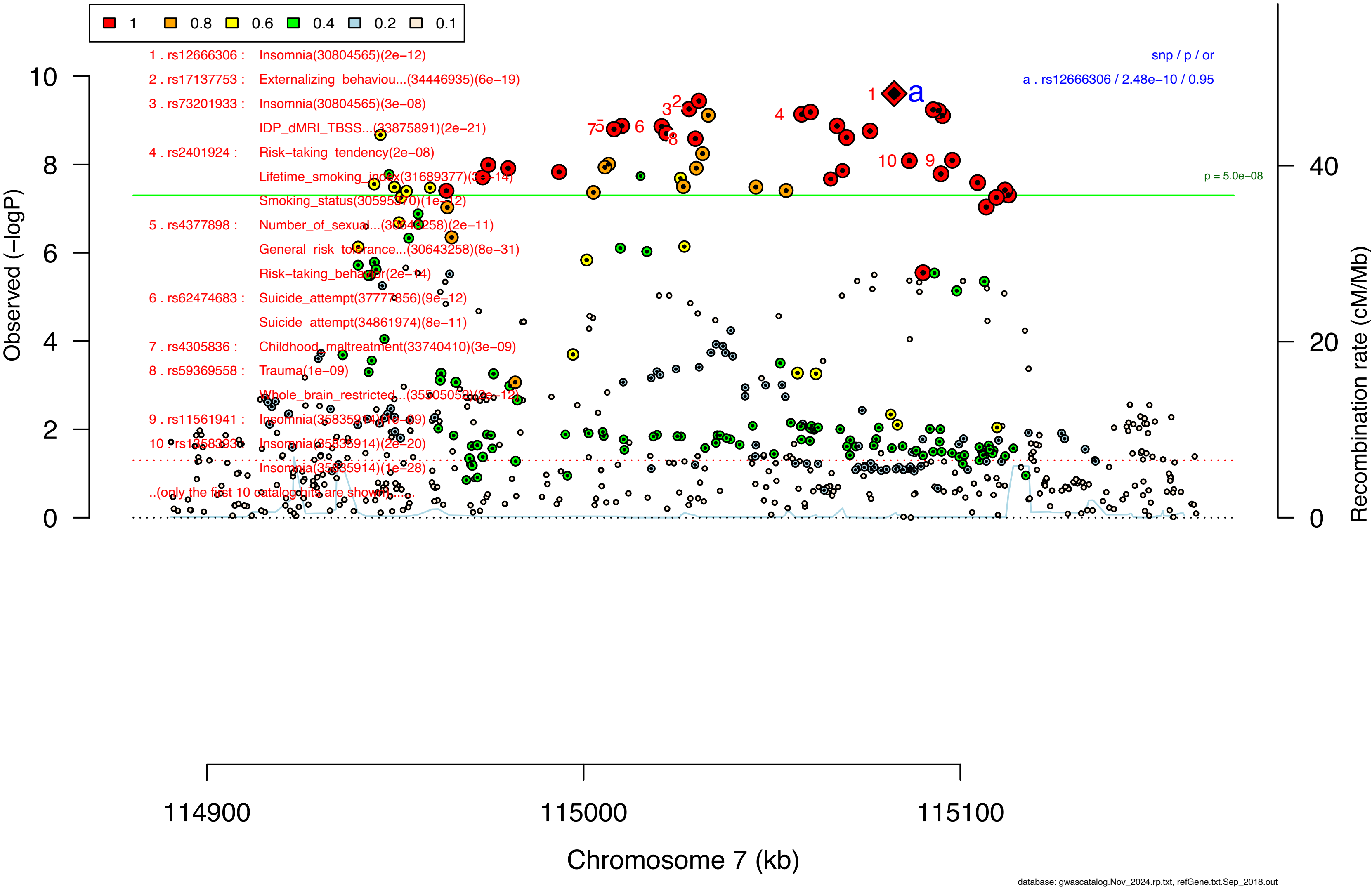

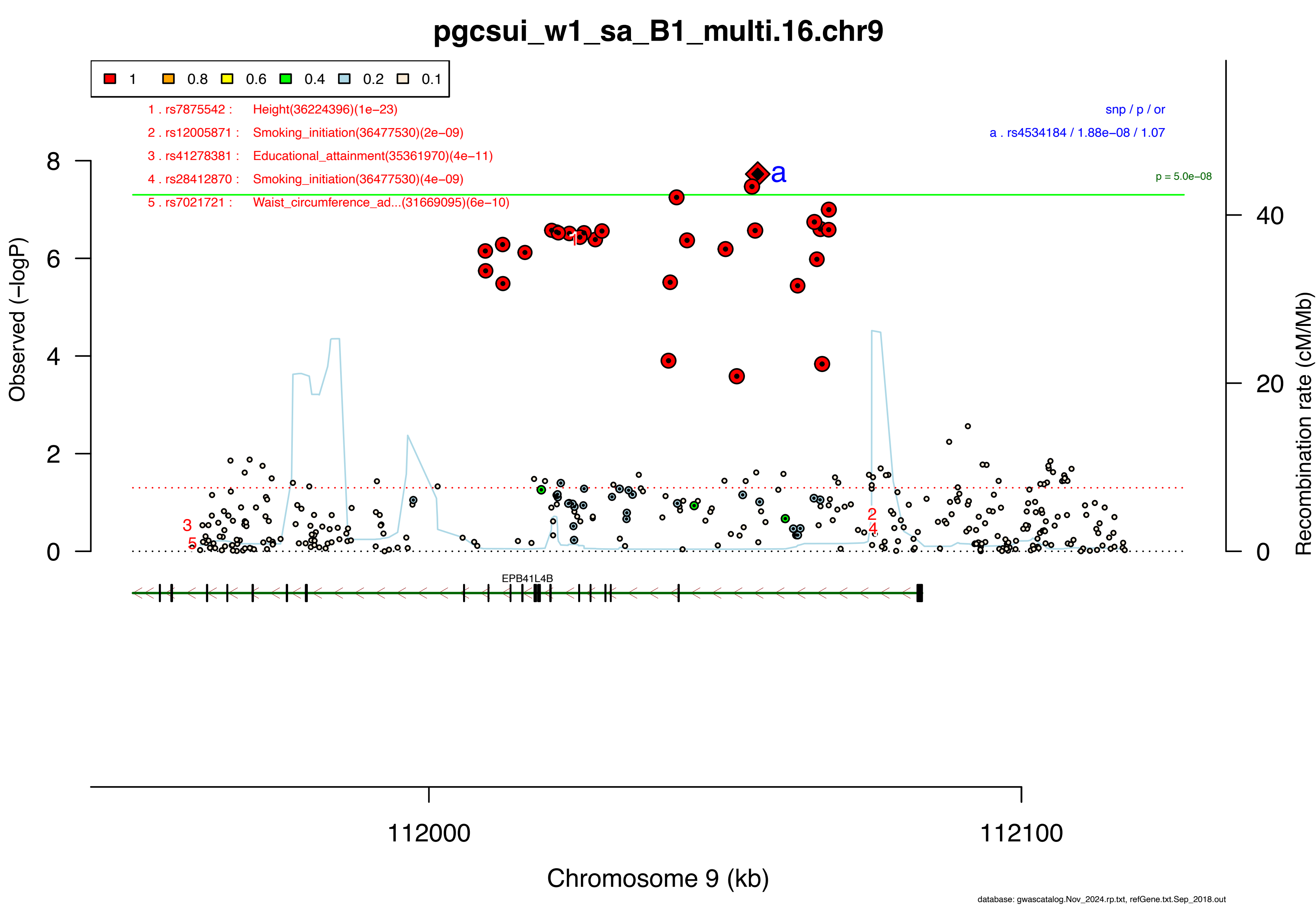
pgcsui_w1_sa_B1_multi.16.chr9
1
0.8
0.6
0.4
0.2
0.1
1 . rs7875542 : Height(36224396)(1e−23)
2 . rs12005871 : Smoking_initiation(36477530)(2e−09)
3 . rs41278381 : Educational_attainment(35361970)(4e−11)
4 . rs28412870 : Smoking_initiation(36477530)(4e−09)
5 . rs7021721 : Waist_circumference_ad...(31669095)(6e−10)
snp / p / or
a . rs4534184 / 1.88e−08 / 1.07
p = 5.0e−08
Observed (−logP)
Recombination rate (cM/Mb)
EPB41L4B
112000
112100
Chromosome 9 (kb)
database: gwascatalog.Nov_2024.rp.txt, refGene.txt.Sep_2018.out

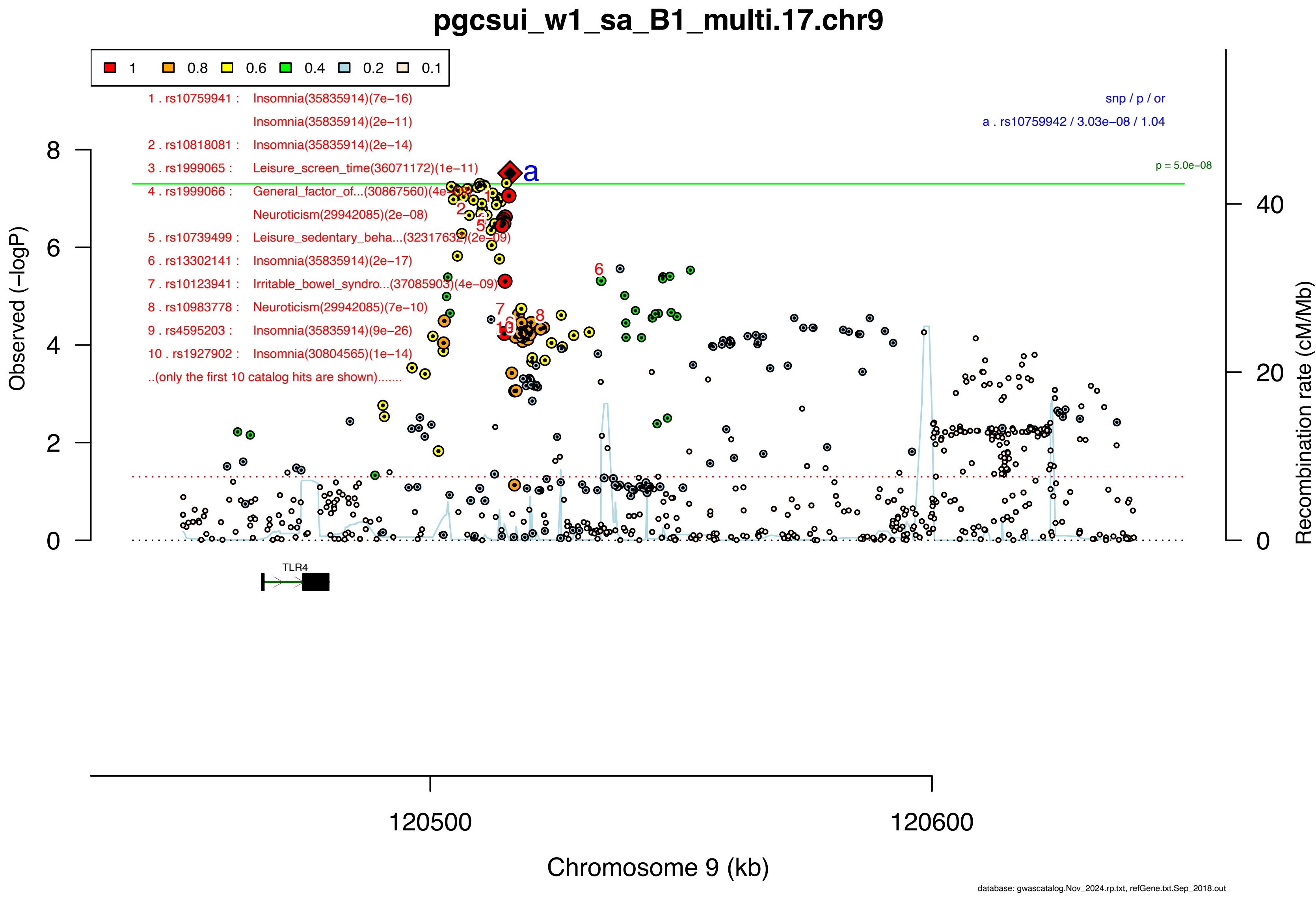

pgcsui_w1_sa_B1_multi.17.chr9
1
0.8
0.6
0.4
0.2
0.1
1 . rs10759941 : Insomnia(35835914)(7e−16)
Insomnia(35835914)(2e−11)
2 . rs10818081 : Insomnia(35835914)(2e−14)
3 . rs1999065 : Leisure_screen_time(36071172)(1e−11)
4 . rs1999066 : General_factor_of...(30867560)(4e−08)
Neuroticism(29942085)(2e−08)
5 . rs10739499 : Leisure_sedentary_beha...(32317632)(2e−09)
6 . rs13302141 : Insomnia(35835914)(2e−17)
7 . rs10123941 : Irritable_bowel_syndro...(37085903)(4e−09)
8 . rs10983778 : Neuroticism(29942085)(7e−10)
9 . rs4595203 : Insomnia(35835914)(9e−26)
10 . rs1927902 : Insomnia(30804565)(1e−14)
..(only the first 10 catalog hits are shown).......
snp / p / or
a . rs10759942 / 3.03e−08 / 1.04
p = 5.0e−08
Observed (−logP)
Recombination rate (cM/Mb)
TLR4
120500
120600
Chromosome 9 (kb)
database: gwascatalog.Nov_2024.rp.txt, refGene.txt.Sep_2018.out

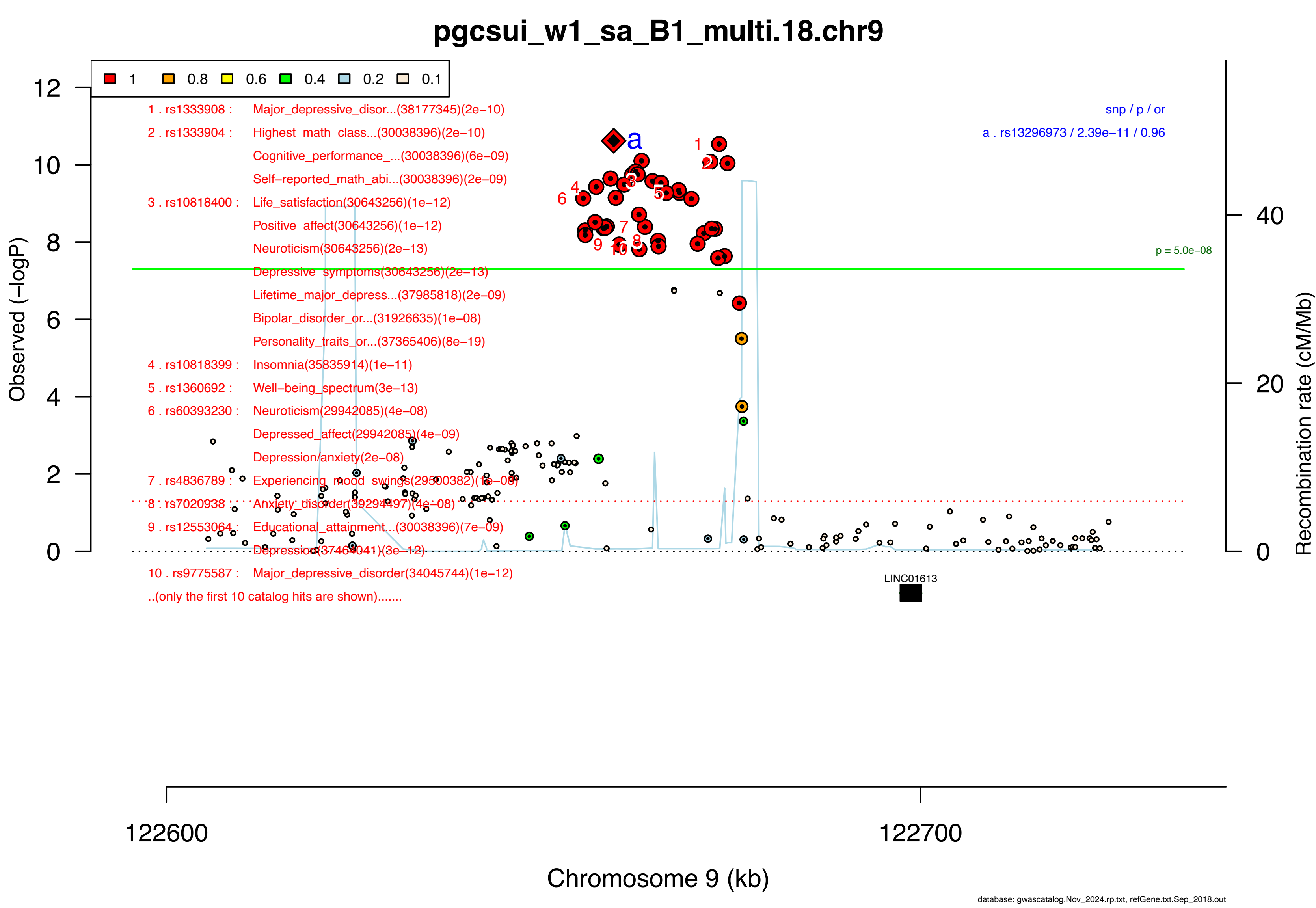
pgcsui_w1_sa_B1_multi.18.chr9
1
0.8
0.6
0.4
0.2
0.1
1 . rs1333908 : Major_depressive_disor...(38177345)(2e−10)
2 . rs1333904 : Highest_math_class...(30038396)(2e−10)
Cognitive_performance_...(30038396)(6e−09)
Self−reported_math_abi...(30038396)(2e−09)
3 . rs10818400 : Life_satisfaction(30643256)(1e−12)
Positive_affect(30643256)(1e−12)
Neuroticism(30643256)(2e−13)
Depressive_symptoms(30643256)(2e−13)
Lifetime_major_depress...(37985818)(2e−09)
Bipolar_disorder_or...(31926635)(1e−08)
Personality_traits_or...(37365406)(8e−19)
4 . rs10818399 : Insomnia(35835914)(1e−11)
5 . rs1360692 : Well−being_spectrum(3e−13)
6 . rs60393230 : Neuroticism(29942085)(4e−08)
Depressed_affect(29942085)(4e−09)
Depression/anxiety(2e−08)
7 . rs4836789 : Experiencing_mood_swings(29500382)(1e−08)
8 . rs7020938 : Anxiety_disorder(39294497)(4e−08)
9 . rs12553064 : Educational_attainment...(30038396)(7e−09)
Depression(37464041)(3e−12)
10 . rs9775587 : Major_depressive_disorder(34045744)(1e−12)
..(only the first 10 catalog hits are shown).......
snp / p / or
a . rs13296973 / 2.39e−11 / 0.96
p = 5.0e−08
Observed (−logP)
Recombination rate (cM/Mb)
LINC01613
122600
122700
Chromosome 9 (kb)
database: gwascatalog.Nov_2024.rp.txt, refGene.txt.Sep_2018.out

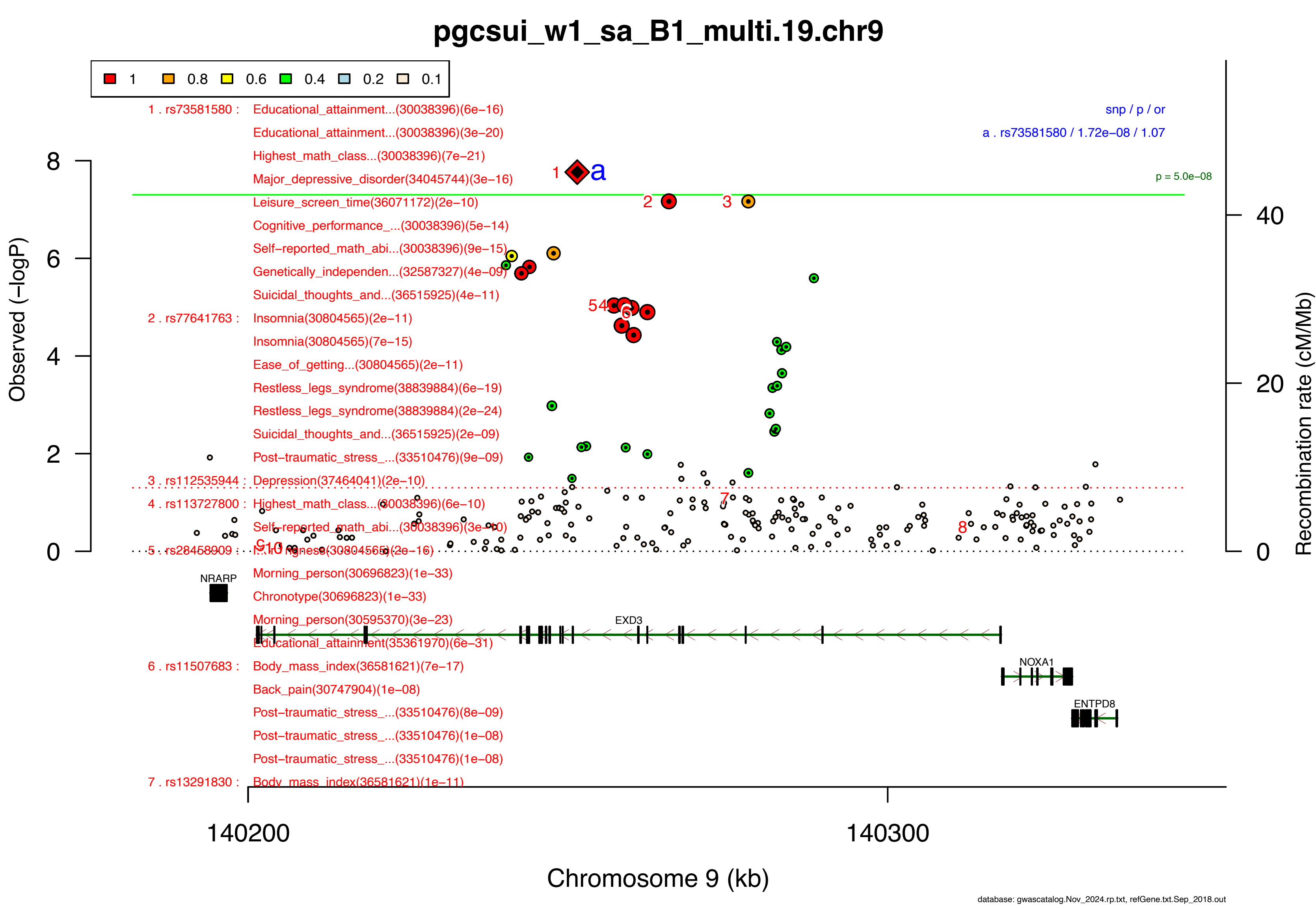

pgcsui_w1_sa_B1_multi.19.chr9
1
0.8
0.6
0.4
0.2
0.1
1 . rs73581580 : Educational_attainment...(30038396)(6e−16)
Educational_attainment...(30038396)(3e−20)
Highest_math_class...(30038396)(7e−21)
Major_depressive_disorder(34045744)(3e−16)
Leisure_screen_time(36071172)(2e−10)
Cognitive_performance_...(30038396)(5e−14)
Self−reported_math_abi...(30038396)(9e−15)
Genetically_independen...(32587327)(4e−09)
Suicidal_thoughts_and...(36515925)(4e−11)
2 . rs77641763 : Insomnia(30804565)(2e−11)
Insomnia(30804565)(7e−15)
Ease_of_getting...(30804565)(2e−11)
Restless_legs_syndrome(38839884)(6e−19)
Restless_legs_syndrome(38839884)(2e−24)
Suicidal_thoughts_and...(36515925)(2e−09)
Post−traumatic_stress_...(33510476)(9e−09)
3 . rs112535944 : Depression(37464041)(2e−10)
4 . rs113727800 : Highest_math_class...(30038396)(6e−10)
Self−reported_math_abi...(30038396)(3e−10)
5 . rs28458909 : Morningness(30804565)(2e−16)
Morning_person(30696823)(1e−33)
Chronotype(30696823)(1e−33)
Morning_person(30595370)(3e−23)
Educational_attainment(35361970)(6e−31)
6 . rs11507683 : Body_mass_index(36581621)(7e−17)
Back_pain(30747904)(1e−08)
Post−traumatic_stress_...(33510476)(8e−09)
Post−traumatic_stress_...(33510476)(1e−08)
Post−traumatic_stress_...(33510476)(1e−08)
7 . rs13291830 : Body_mass_index(36581621)(1e−11)
snp / p / or
a . rs73581580 / 1.72e−08 / 1.07
p = 5.0e−08
Observed (−logP)
Recombination rate (cM/Mb)
NRARP
EXD3
NOXA1
ENTPD8
140200
140300
Chromosome 9 (kb)
database: gwascatalog.Nov_2024.rp.txt, refGene.txt.Sep_2018.out

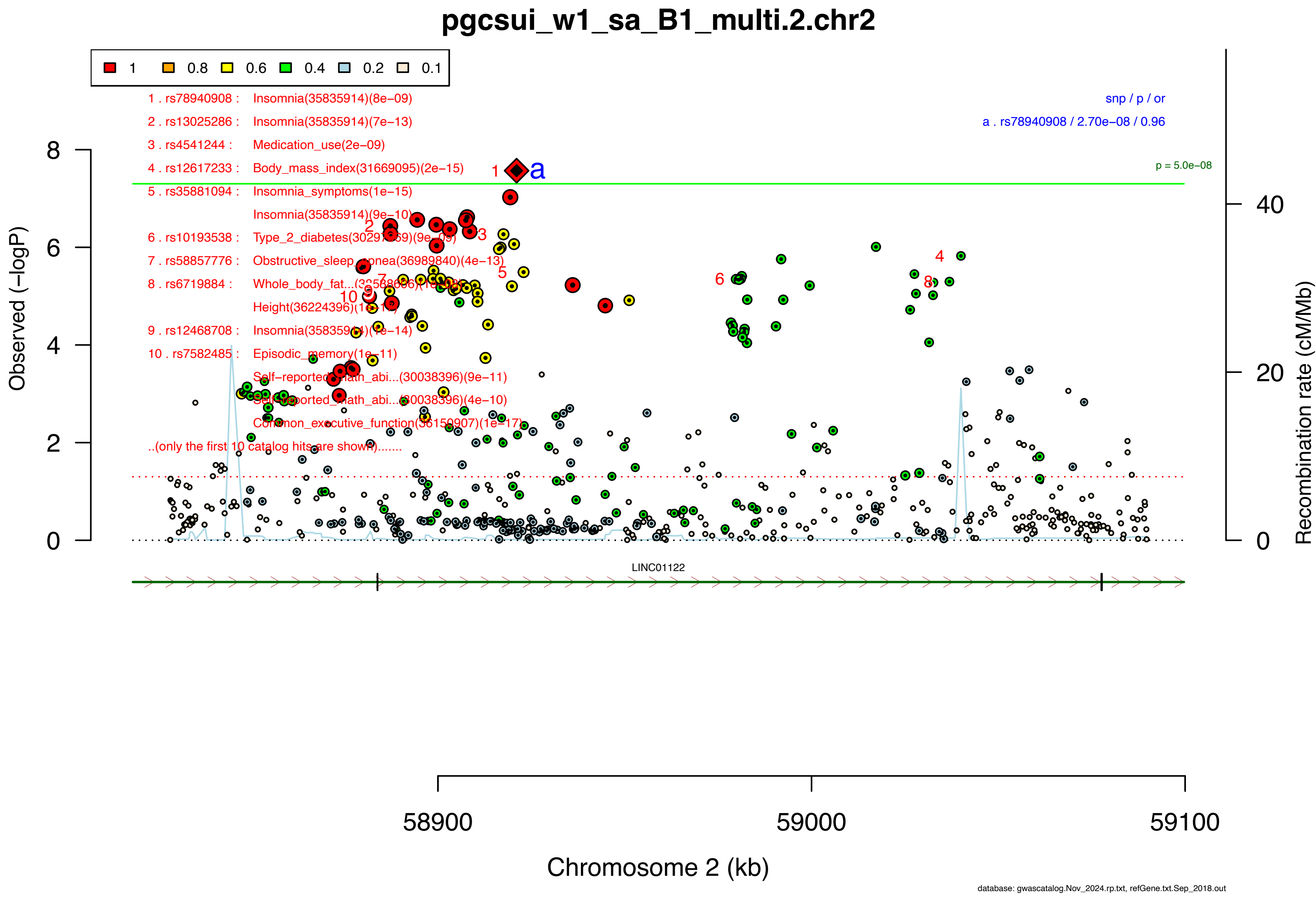
pgcsui_w1_sa_B1_multi.2.chr2
1
0.8
0.6
0.4
0.2
0.1
1 . rs78940908 : Insomnia(35835914)(8e−09)
2 . rs13025286 : Insomnia(35835914)(7e−13)
3 . rs4541244 : Medication_use(2e−09)
4 . rs12617233 : Body_mass_index(31669095)(2e−15)
5 . rs35881094 : Insomnia_symptoms(1e−15)
Insomnia(35835914)(9e−10)
6 . rs10193538 : Type_2_diabetes(30297969)(9e−09)
7 . rs58857776 : Obstructive_sleep_apnea(36989840)(4e−13)
8 . rs6719884 : Whole_body_fat...(32568606)(1e−08)
Height(36224396)(1e−11)
9 . rs12468708 : Insomnia(35835914)(1e−14)
10 . rs7582485 : Episodic_memory(1e−11)
Self−reported_math_abi...(30038396)(9e−11)
Self−reported_math_abi...(30038396)(4e−10)
Common_executive_function(36150907)(1e−17)
..(only the first 10 catalog hits are shown).......
snp / p / or
a . rs78940908 / 2.70e−08 / 0.96
p = 5.0e−08
Observed (−logP)
Recombination rate (cM/Mb)
LINC01122
58900
59000
59100
Chromosome 2 (kb)
database: gwascatalog.Nov_2024.rp.txt, refGene.txt.Sep_2018.out

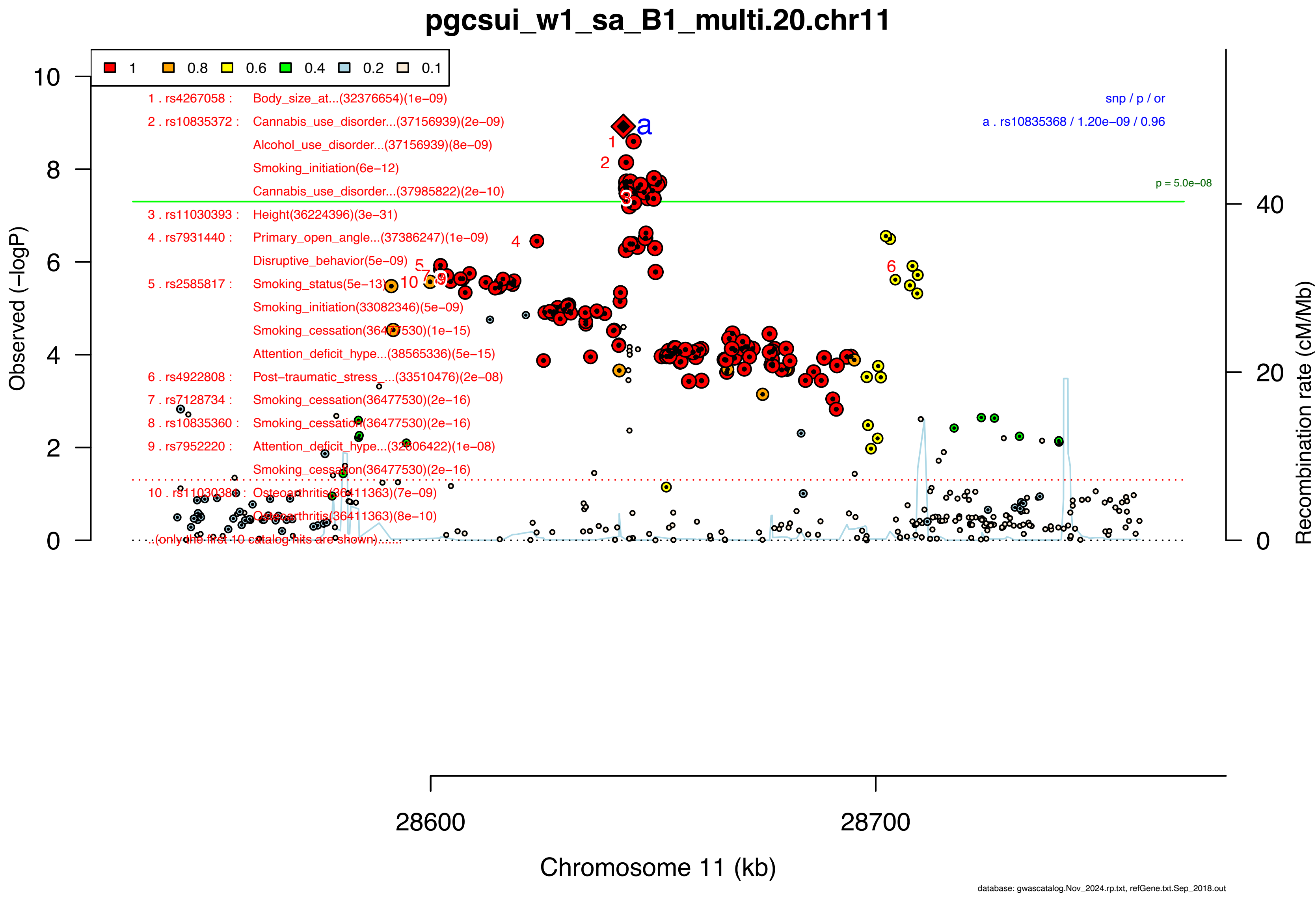

pgcsui_w1_sa_B1_multi.20.chr11
1
0.8
0.6
0.4
0.2
0.1
1 . rs4267058 : Body_size_at...(32376654)(1e−09)
2 . rs10835372 : Cannabis_use_disorder...(37156939)(2e−09)
Alcohol_use_disorder...(37156939)(8e−09)
Smoking_initiation(6e−12)
Cannabis_use_disorder...(37985822)(2e−10)
3 . rs11030393 : Height(36224396)(3e−31)
4 . rs7931440 : Primary_open_angle...(37386247)(1e−09)
Disruptive_behavior(5e−09)
5 . rs2585817 : Smoking_status(5e−13)
Smoking_initiation(33082346)(5e−09)
Smoking_cessation(36477530)(1e−15)
Attention_deficit_hype...(38565336)(5e−15)
6 . rs4922808 : Post−traumatic_stress_...(33510476)(2e−08)
7 . rs7128734 : Smoking_cessation(36477530)(2e−16)
8 . rs10835360 : Smoking_cessation(36477530)(2e−16)
9 . rs7952220 : Attention_deficit_hype...(32606422)(1e−08)
Smoking_cessation(36477530)(2e−16)
10 . rs11030381 : Osteoarthritis(36411363)(7e−09)
Osteoarthritis(36411363)(8e−10)
..(only the first 10 catalog hits are shown)......
snp / p / or
a . rs10835368 / 1.20e−09 / 0.96
p = 5.0e−08
Observed (−logP)
Recombination rate (cM/Mb)
28600
28700
Chromosome 11 (kb)
database: gwascatalog.Nov_2024.rp.txt, refGene.txt.Sep_2018.out

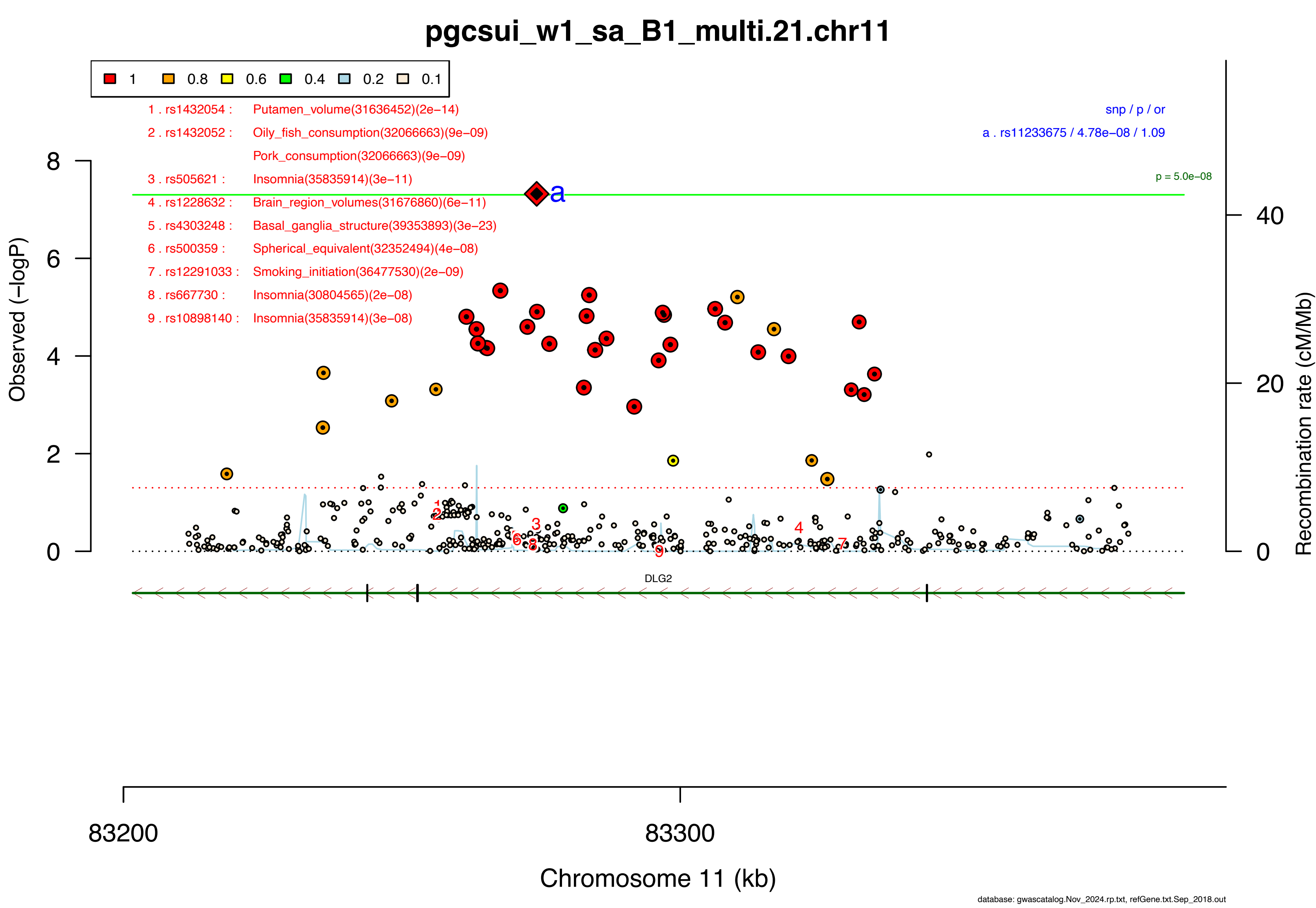

pgcsui_w1_sa_B1_multi.21.chr11
1
0.8
0.6
0.4
0.2
0.1
1 . rs1432054 : Putamen_volume(31636452)(2e−14)
2 . rs1432052 : Oily_fish_consumption(32066663)(9e−09)
Pork_consumption(32066663)(9e−09)
3 . rs505621 : Insomnia(35835914)(3e−11)
4 . rs1228632 : Brain_region_volumes(31676860)(6e−11)
5 . rs4303248 : Basal_ganglia_structure(39353893)(3e−23)
6 . rs500359 : Spherical_equivalent(32352494)(4e−08)
7 . rs12291033 : Smoking_initiation(36477530)(2e−09)
8 . rs667730 : Insomnia(30804565)(2e−08)
9 . rs10898140 : Insomnia(35835914)(3e−08)
snp / p / or
a . rs11233675 / 4.78e−08 / 1.09
p = 5.0e−08
a
Observed (−logP)
8
6
4
2
0
Recombination rate (cM/Mb)
40
20
0
DLG2
83200
83300
Chromosome 11 (kb)
database: gwascatalog.Nov_2024.rp.txt, refGene.txt.Sep_2018.out

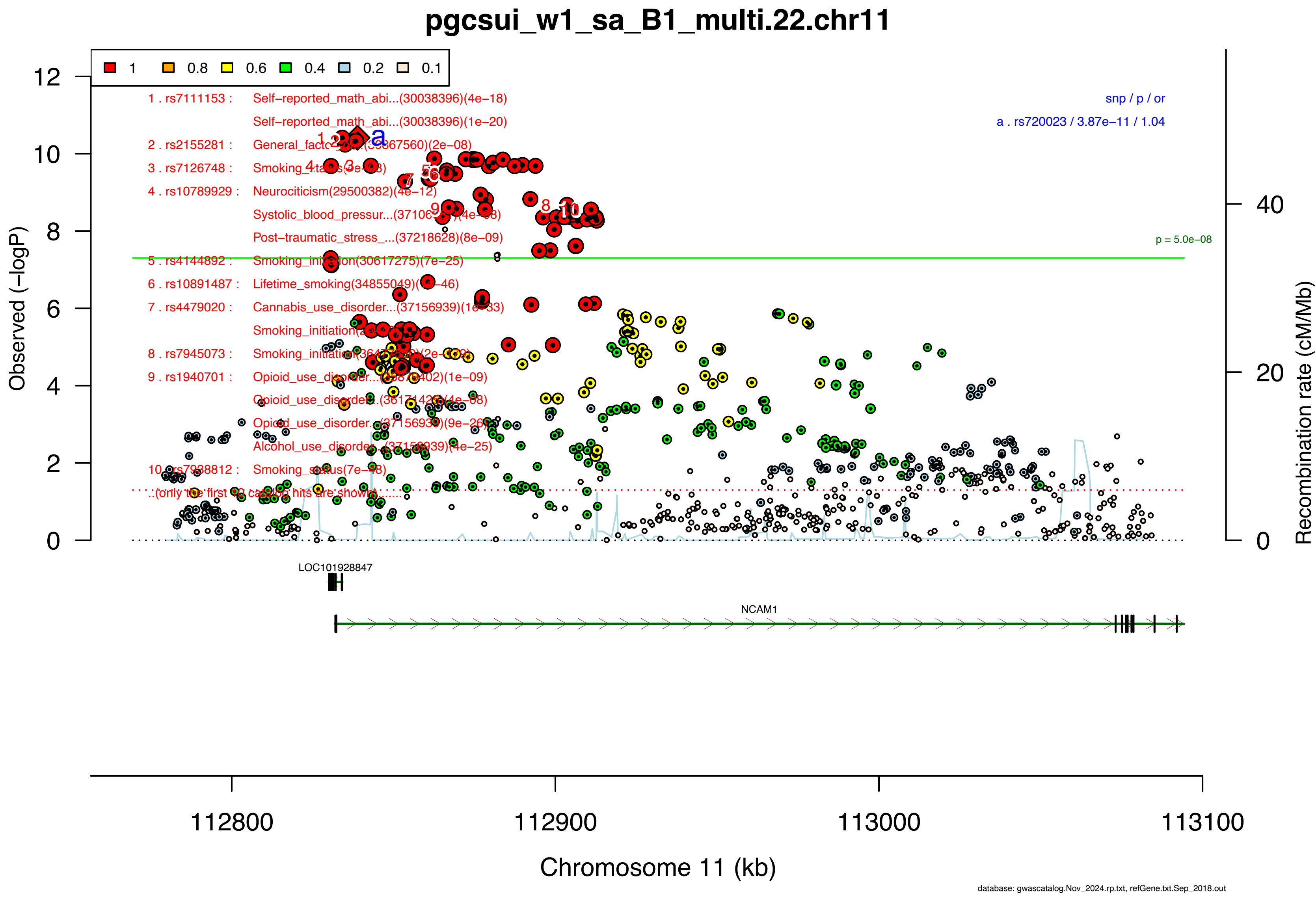

pgcsui_w1_sa_B1_multi.22.chr11
1
0.8
0.6
0.4
0.2
0.1
1 . rs7111153 : Self−reported_math_abi...(30038396)(4e−18)
Self−reported_math_abi...(30038396)(1e−20)
2 . rs2155281 : General_facto (2e−08)
3 . rs7126748 : Smoking
4 . rs10789929 : Neurociticism(29500382)(4e−12)
Systolic_blood_pressur...(3710 (4e−08)
Post−traumatic_stress_...(37218628)(8e−09)
5 . rs4144892 : Smoking_ini on(30617275)(7e−25)
6 . rs10891487 : Lifetime_smoking(34855049)( −46)
7 . rs4479020 : Cannabis_use_disorder...(37156939)(1e−33)
Smoking_initiation
8 . rs7945073 : Smoking_initiation
9 . rs1940701 : Opioid_use_disorder...
Opioid_use_disorder...
Opioid_use_disorder...
Alcohol_use_disorder...
10 . rs7938812 : Smoking_status(7e−48)
..(only the first 10 catalog hits are shown)......
snp / p / or
a . rs720023 / 3.87e−11 / 1.04
a
p = 5.0e−08
Observed (−logP)
Recombination rate (cM/Mb)
LOC101928847
NCAM1
112800
112900
113000
113100
Chromosome 11 (kb)
database: gwascatalog.Nov_2024.rp.txt, refGene.txt.Sep_2018.out

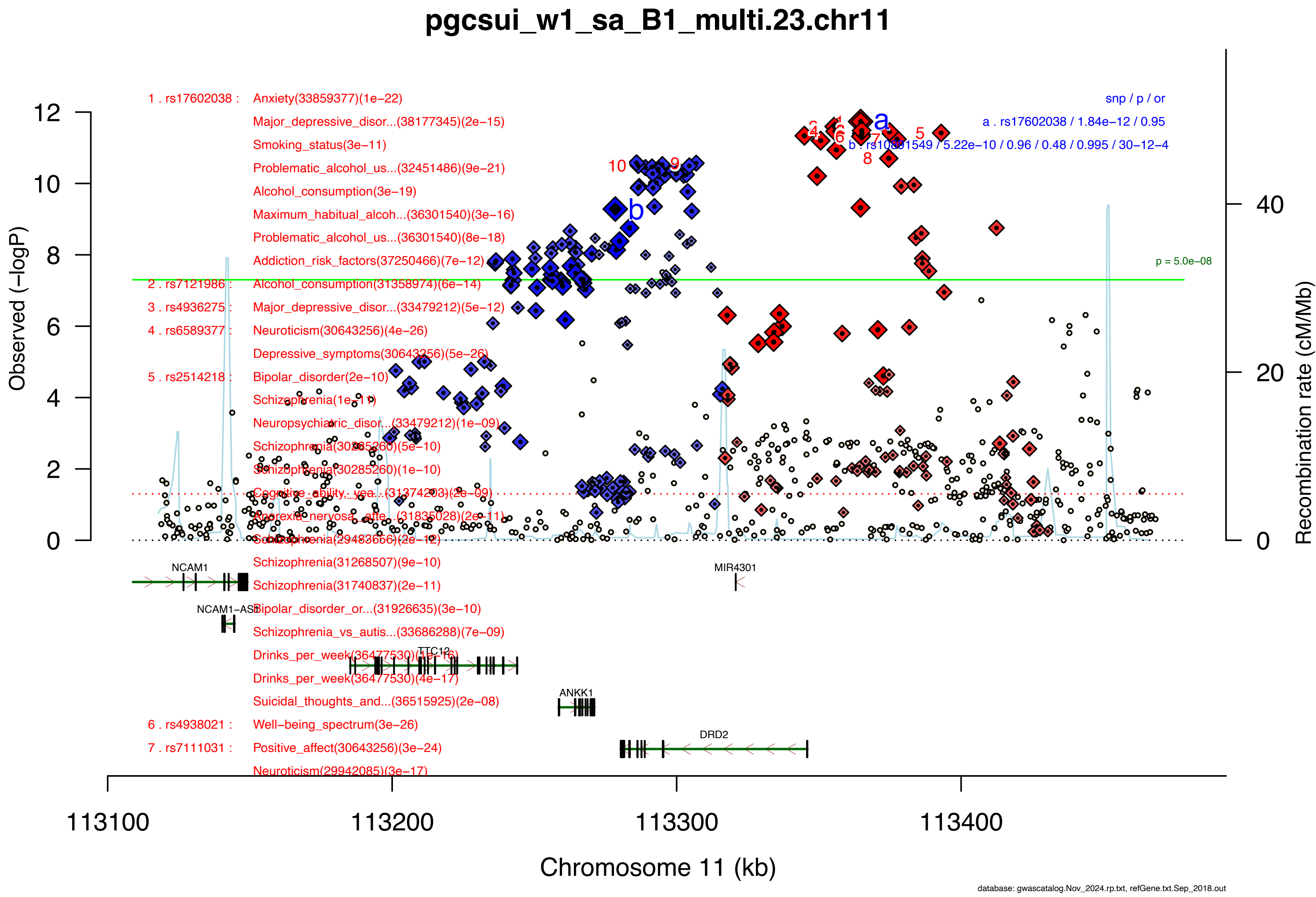

pgcsui_w1_sa_B1_multi.23.chr11
1 . rs17602038 :
Anxiety(33859377)(1e−22)
Major_depressive_disor...(38177345)(2e−15)
Smoking_status(3e−11)
Problematic_alcohol_us...(32451486)(9e−21)
Alcohol_consumption(3e−19)
Maximum_habitual_alcoh...(36301540)(3e−16)
Problematic_alcohol_us...(36301540)(8e−18)
Addiction_risk_factors(37250466)(7e−12)
2 . rs7121986 :
Alcohol_consumption(31358974)(6e−14)
3 . rs4936275 :
Major_depressive_disor...(33479212)(5e−12)
4 . rs6589377 :
Neuroticism(30643256)(4e−26)
Depressive_symptoms(30643256)(5e−26)
5 . rs2514218 :
Bipolar_disorder(2e−10)
Schizophrenia(1e−11)
Neuropsychiatric_disor...(33479212)(1e−09)
Schizophrenia(30285260)(5e−10)
Schizophrenia(30285260)(1e−10)
Cognitive_ability,_yea...(31374203)(2e−09)
Anorexia_nervosa_atte...(31835028)(2e−11)
Schizophrenia(29483656)(2e−12)
Schizophrenia(31268507)(9e−10)
Schizophrenia(31740837)(2e−11)
Bipolar_disorder_or...(31926635)(3e−10)
Schizophrenia_vs_autis...(33686288)(7e−09)
Drinks_per_week(36477530)(1e−16)
Drinks_per_week(36477530)(4e−17)
Suicidal_thoughts_and...(36515925)(2e−08)
6 . rs4938021 :
Well−being_spectrum(3e−26)
7 . rs7111031 :
Positive_affect(30643256)(3e−24)
Neuroticism(29942085)(3e−17)
snp / p / or
a . rs17602038 / 1.84e−12 / 0.95
b . rs10891549 / 5.22e−10 / 0.96 / 0.48 / 0.995 / 30−12−4
p = 5.0e−08
a
b
Observed (−logP)
Recombination rate (cM/Mb)
NCAM1
NCAM1−AS1
TTC12
ANKK1
DRD2
MIR4301
113100
113200
113300
113400
Chromosome 11 (kb)
database: gwascatalog.Nov_2024.rp.txt, refGene.txt.Sep_2018.out

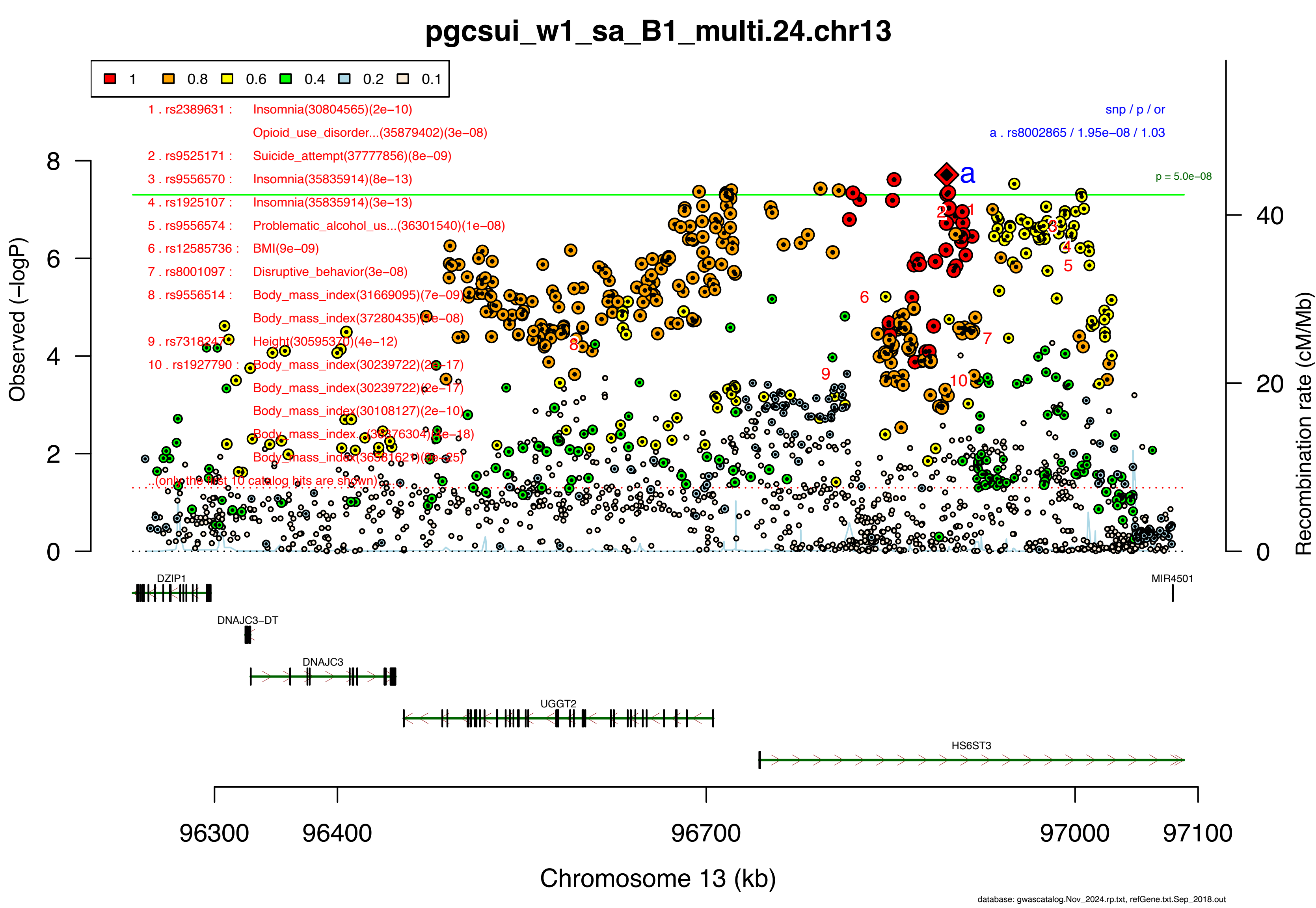

pgcsui_w1_sa_B1_multi.24.chr13
1 0.8 0.6 0.4 0.2 0.1
1 . rs2389631 : Insomnia(30804565)(2e−10)
Opioid_use_disorder...(35879402)(3e−08)
2 . rs9525171 : Suicide_attempt(37777856)(8e−09)
3 . rs9556570 : Insomnia(35835914)(8e−13)
4 . rs1925107 : Insomnia(35835914)(3e−13)
5 . rs9556574 : Problematic_alcohol_us...(36301540)(1e−08)
6 . rs12585736 : BMI(9e−09)
7 . rs8001097 : Disruptive_behavior(3e−08)
8 . rs9556514 : Body_mass_index(31669095)(7e−09)
Body_mass_index(37280435)(7e−08)
9 . rs7318247 : Height(30595370)(4e−12)
10 . rs1927790 : Body_mass_index(30239722)(2e−17)
Body_mass_index(30239722)(2e−17)
Body_mass_index(30108127)(2e−10)
Body_mass_index...(36376304)(4e−18)
Body_mass_index(36581621)(8e−25)
..(only the first 10 catalog hits are shown)...
snp / p / or
a . rs8002865 / 1.95e−08 / 1.03
p = 5.0e−08
Observed (−logP)
Recombination rate (cM/Mb)
DZIP1
DNAJC3−DT
DNAJC3
UGGT2
HS6ST3
MIR4501
96300 96400 96700 97000 97100
Chromosome 13 (kb)
database: gwascatalog.Nov_2024.rp.txt, refGene.txt.Sep_2018.out

### pgcsui_w1_sa_B1_multi.25.chr14

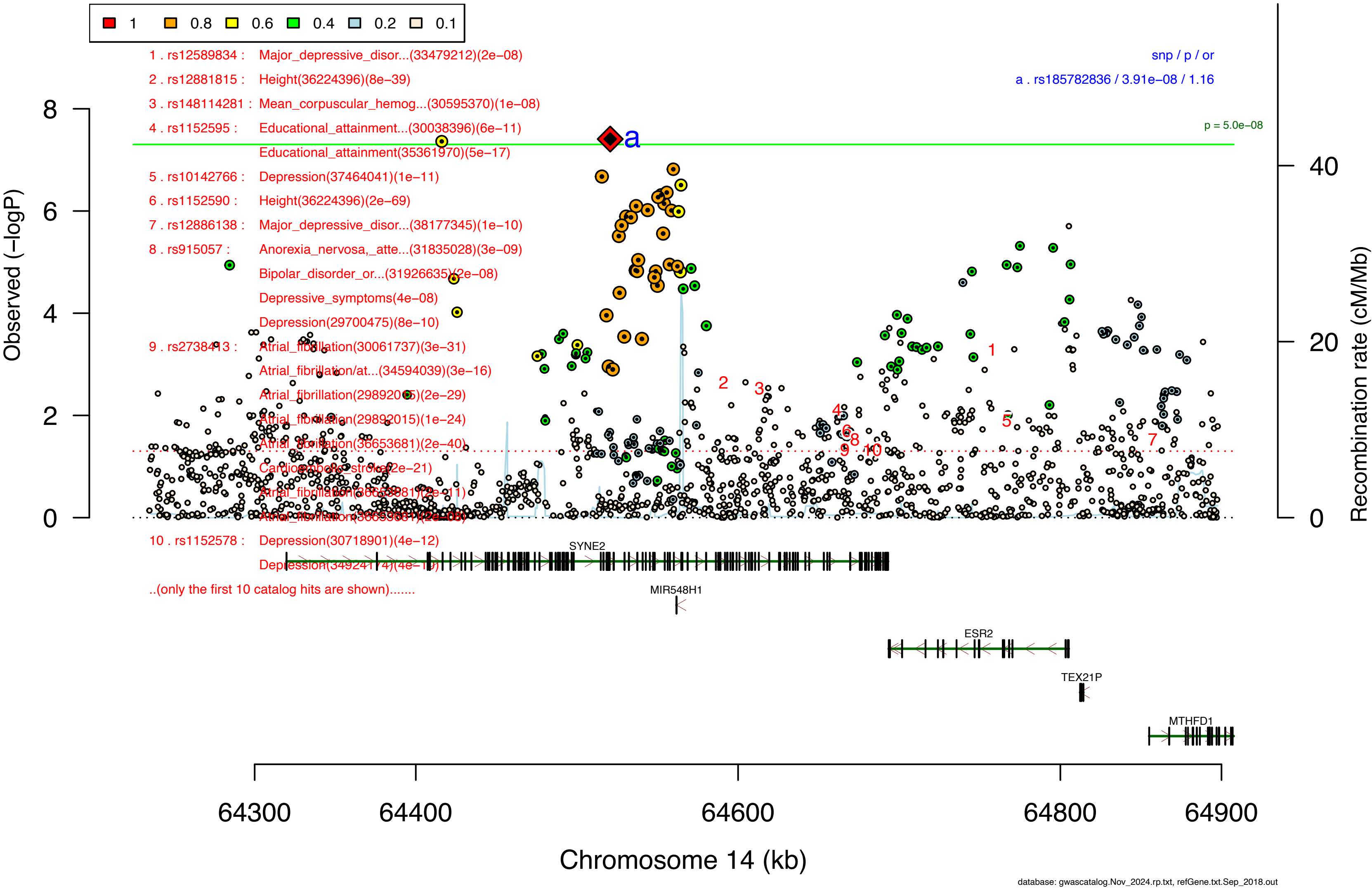

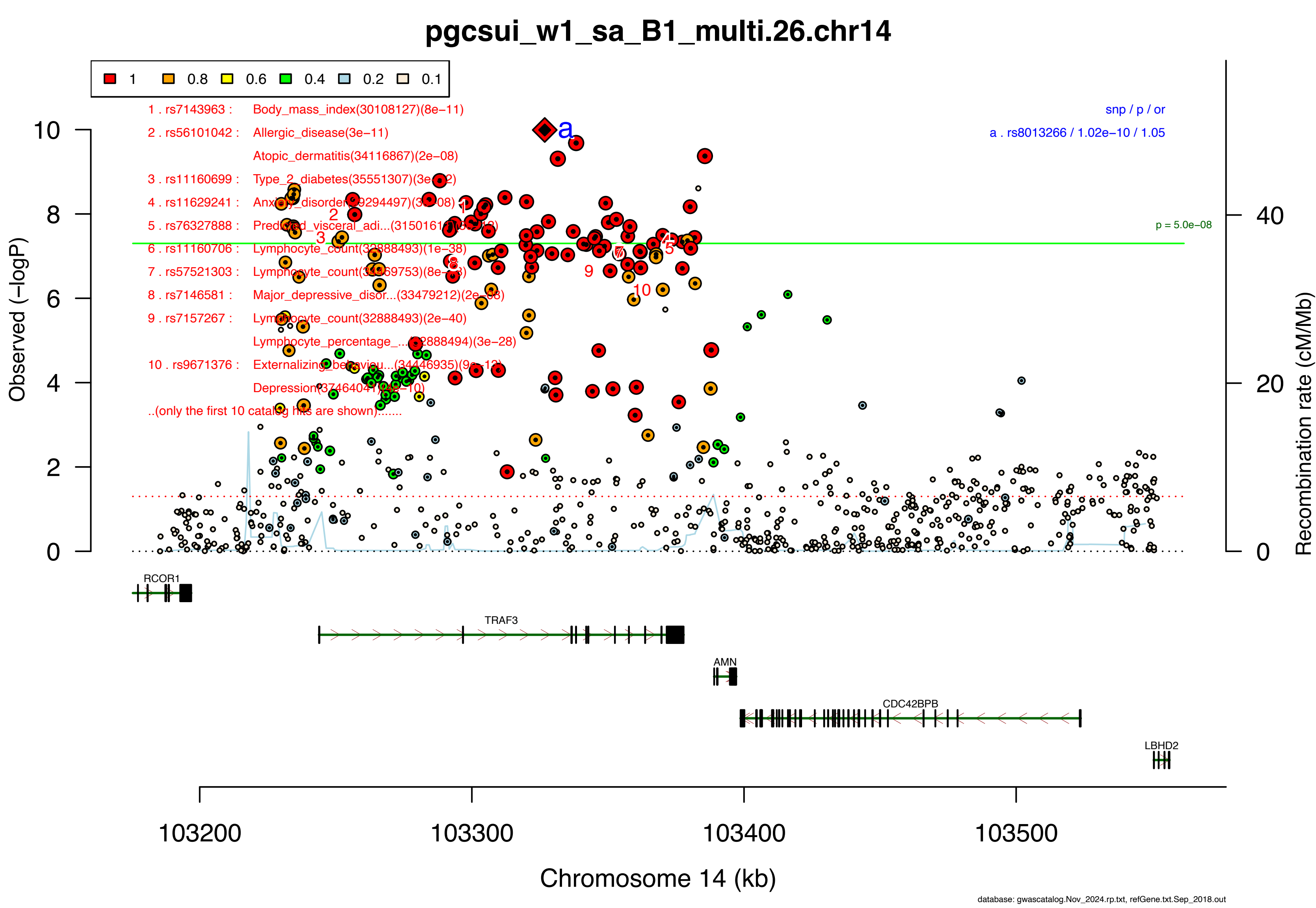

pgcsui_w1_sa_B1_multi.26.chr14
1
0.8
0.6
0.4
0.2
0.1
1 . rs7143963 : Body_mass_index(30108127)(8e−11)
2 . rs56101042 : Allergic_disease(3e−11)
Atopic_dermatitis(34116867)(2e−08)
3 . rs11160699 : Type_2_diabetes(35551307)(3e−12)
4 . rs11629241 : Anxiety_disorder(29294497)(3e−08)
5 . rs76327888 : Predicted_visceral_adi...(31501611)(8e−12)
6 . rs11160706 : Lymphocyte_count(32888493)(1e−38)
7 . rs57521303 : Lymphocyte_count(34469753)(8e−43)
8 . rs7146581 : Major_depressive_disor...(33479212)(2e−08)
9 . rs7157267 : Lymphocyte_count(32888493)(2e−40)
Lymphocyte_percentage_...(32888494)(3e−28)
10 . rs9671376 : Externalizing_behaviou...(34446935)(9e−12)
Depression(37464041)(9e−10)
..(only the first 10 catalog hits are shown).......
snp / p / or
a . rs8013266 / 1.02e−10 / 1.05
p = 5.0e−08
Observed (−logP)
Recombination rate (cM/Mb)
RCOR1
TRAF3
AMN
CDC42BPB
LBHD2
103200
103300
103400
103500
Chromosome 14 (kb)
database: gwascatalog.Nov_2024.rp.txt, refGene.txt.Sep_2018.out

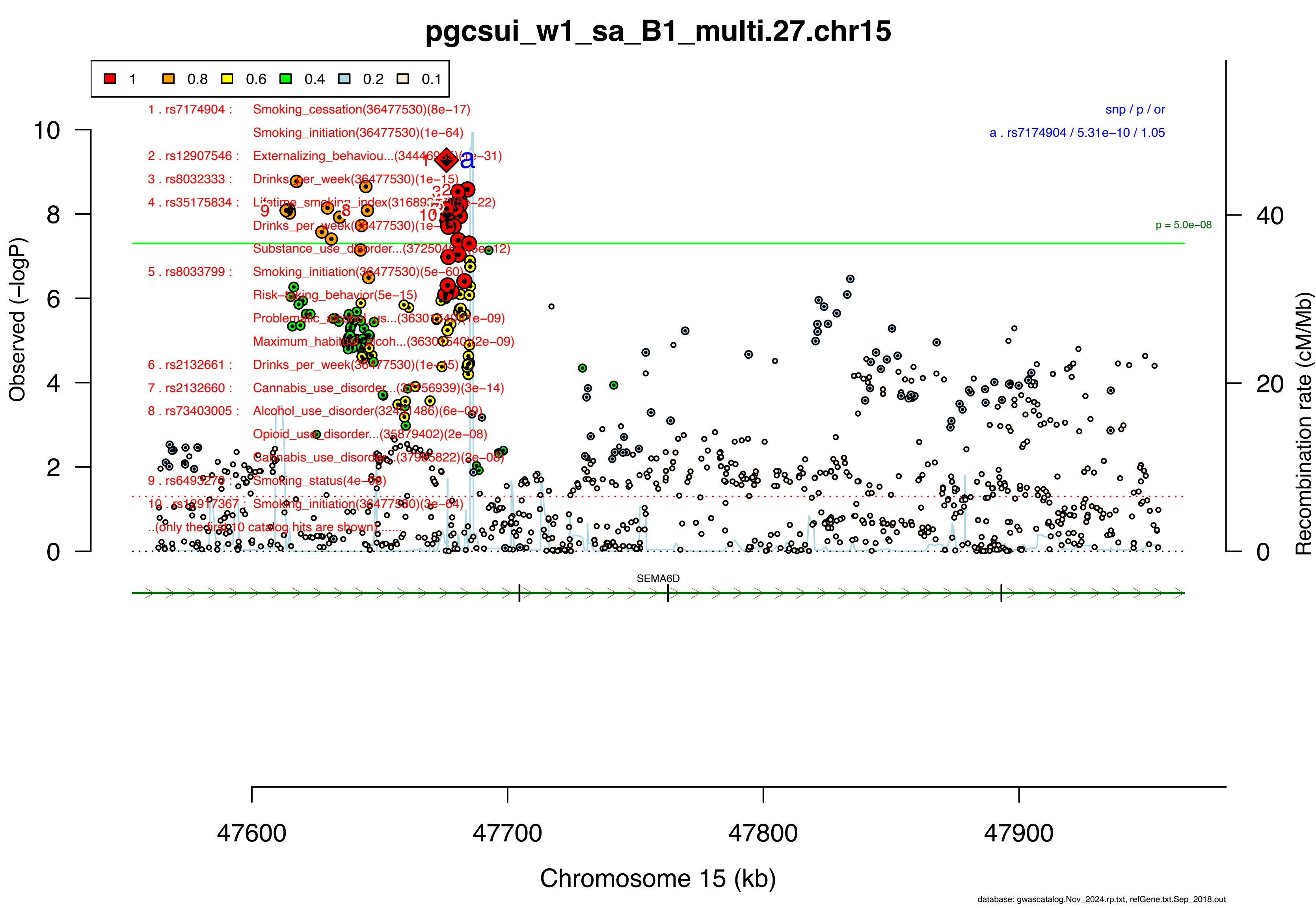

pgcsui_w1_sa_B1_multi.27.chr15
1
0.8
0.6
0.4
0.2
0.1
1 . rs7174904 : Smoking_cessation(36477530)(8e−17)
Smoking_initiation(36477530)(1e−64)
2 . rs12907546 : Externalizing_behaviou...(3444[illegible])(2e−31)
3 . rs8032333 : Drinks_per_week(36477530)(1e−15)
4 . rs35175834 : Lifetime_smoking_index(3168[illegible])(5e−22)
Drinks_per_week(36477530)(1e−15)
Substance_use_disorder...(3725[illegible])(8e−12)
5 . rs8033799 : Smoking_initiation(36477530)(5e−60)
Risk−taking_behavior(5e−15)
Problematic_alcohol_us...(36303540)(1e−09)
Maximum_habitual_alcoh...(36303540)(2e−09)
6 . rs2132661 : Drinks_per_week(36477530)(1e−15)
7 . rs2132660 : Cannabis_use_disorder...(37956939)(3e−14)
8 . rs73403005 : Alcohol_use_disorder(32451486)(6e−09)
Opioid_use_disorder...(35879402)(2e−08)
Cannabis_use_disorder...(37985822)(3e−08)
9 . rs6493276 : Smoking_status(4e−08)
10 . rs12917367 : Smoking_initiation(36477530)(3e−64)
..(only the first 10 catalog hits are shown)......
snp / p / or
a . rs7174904 / 5.31e−10 / 1.05
p = 5.0e−08
Observed (−logP)
Recombination rate (cM/Mb)
SEMA6D
Chromosome 15 (kb)
47600
47700
47800
47900
database: gwascatalog.Nov_2024.rp.txt, refGene.txt.Sep_2018.out

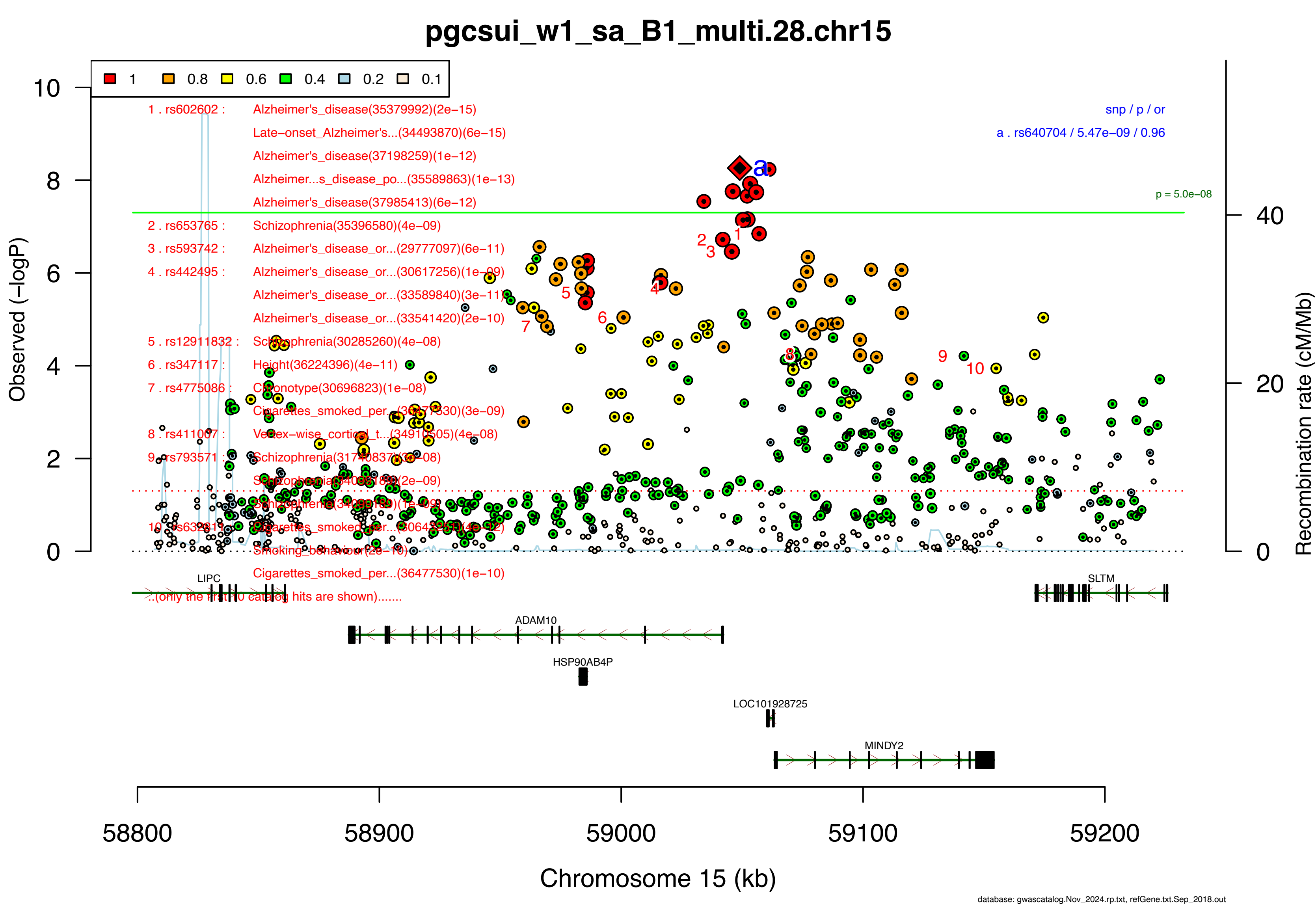
pgcsui_w1_sa_B1_multi.28.chr15
1 0.8 0.6 0.4 0.2 0.1
1 . rs602602 : Alzheimer's_disease(35379992)(2e−15)
Late−onset_Alzheimer's...(34493870)(6e−15)
Alzheimer's_disease(37198259)(1e−12)
Alzheimer...s_disease_po...(35589863)(1e−13)
Alzheimer's_disease(37985413)(6e−12)
2 . rs653765 : Schizophrenia(35396580)(4e−09)
3 . rs593742 : Alzheimer's_disease_or...(29777097)(6e−11)
4 . rs442495 : Alzheimer's_disease_or...(30617256)(1e−09)
Alzheimer's_disease_or...(33589840)(3e−11)
Alzheimer's_disease_or...(33541420)(2e−10)
5 . rs12911832 : Schizophrenia(30285260)(4e−08)
6 . rs347117 : Height(36224396)(4e−11)
7 . rs4775086 : Chronotype(30696823)(1e−08)
Cigarettes_smoked_per...(36477530)(3e−09)
8 . rs411007 : Vertex−wise_cortical_t...(34910505)(4e−08)
9 . rs793571 : Schizophrenia(31740837)(3e−08)
Schizophrenia(34099189)(2e−09)
Schizophrenia(34099189)(1e−09)
10 . rs632811 : Cigarettes_smoked_per...(30643251)(4e−12)
Smoking_behaviour(2e−10)
Cigarettes_smoked_per...(36477530)(1e−10)
...(only the first 10 catalog hits are shown).......
snp / p / or
a . rs640704 / 5.47e−09 / 0.96
p = 5.0e−08
Observed (−logP)
Recombination rate (cM/Mb)
LIPC
SLTM
ADAM10
HSP90AB4P
LOC101928725
MINDY2
58800 58900 59000 59100 59200
Chromosome 15 (kb)
database: gwascatalog.Nov_2024.rp.txt, refGene.txt.Sep_2018.out

### pgcsui_w1_sa_B1_multi.29.chr15

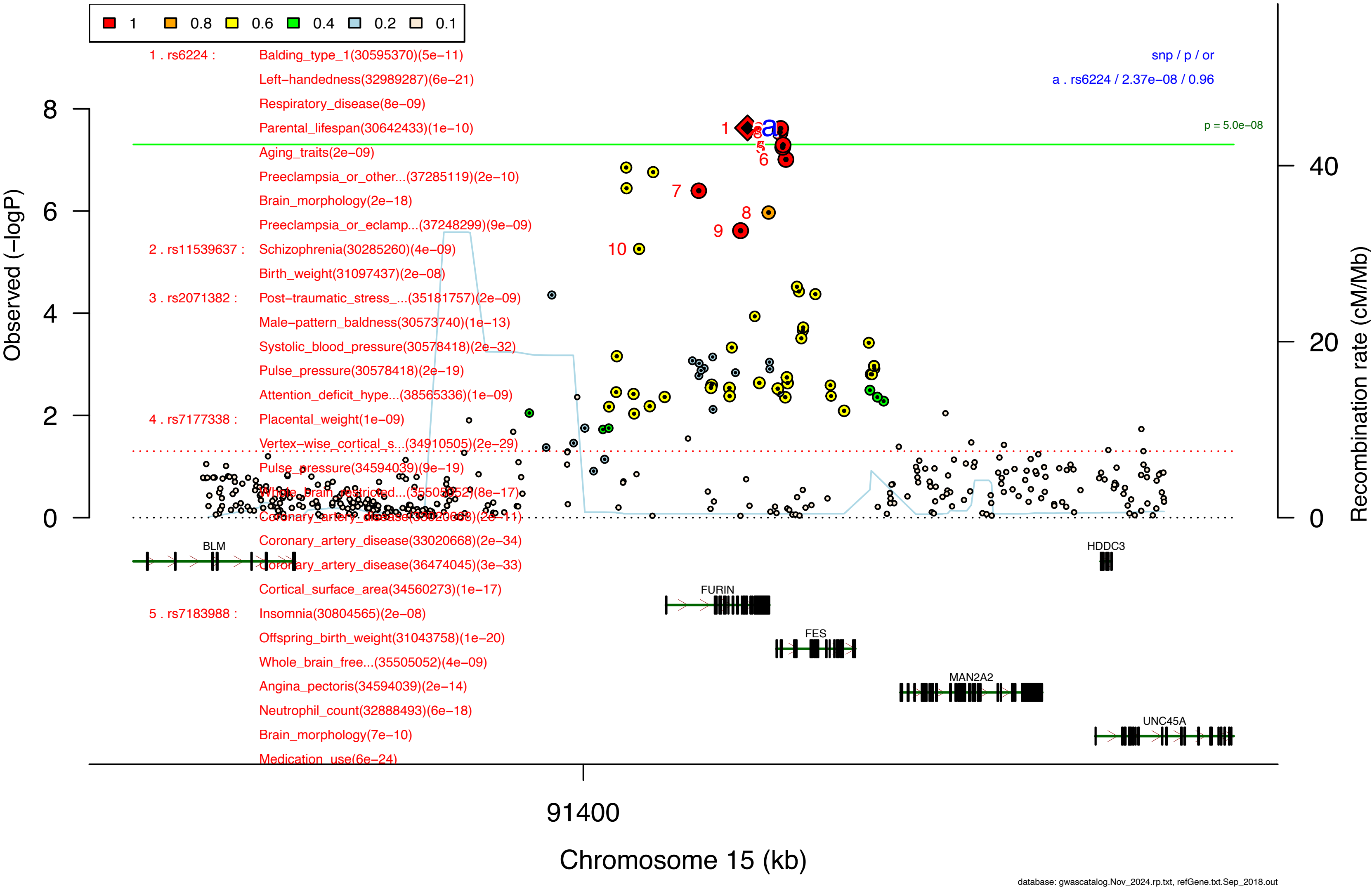

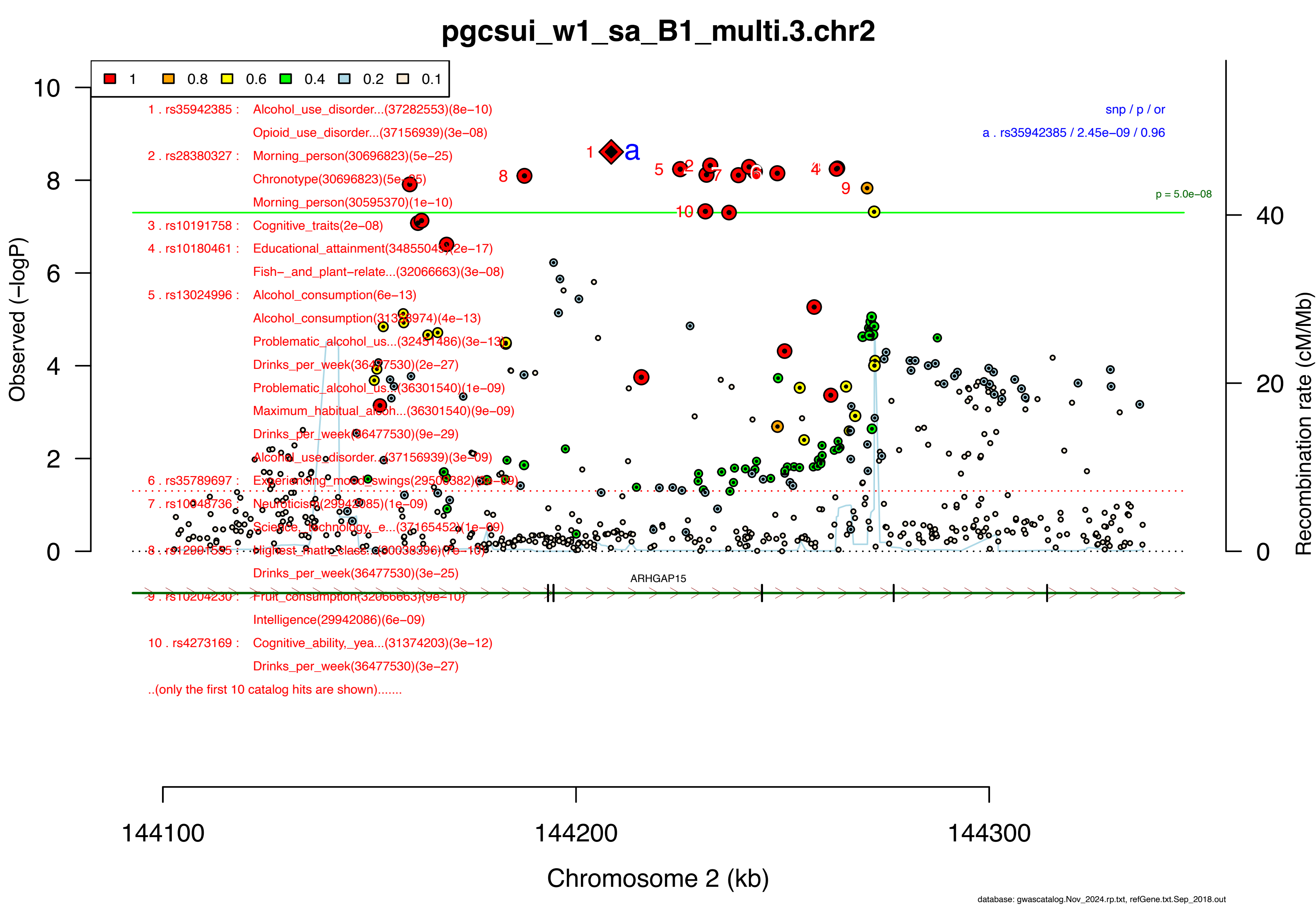

pgcsui_w1_sa_B1_multi.3.chr2
1
0.8
0.6
0.4
0.2
0.1
1 . rs35942385 : Alcohol_use_disorder...(37282553)(8e−10)
Opioid_use_disorder...(37156939)(3e−08)
2 . rs28380327 : Morning_person(30696823)(5e−25)
Chronotype(30696823)(5e−25)
Morning_person(30595370)(1e−10)
3 . rs10191758 : Cognitive_traits(2e−08)
4 . rs10180461 : Educational_attainment(34855049)(2e−17)
Fish−_and_plant−relate...(32066663)(3e−08)
5 . rs13024996 : Alcohol_consumption(6e−13)
Alcohol_consumption(31358974)(4e−13)
Problematic_alcohol_us...(32451486)(3e−13)
Drinks_per_week(36477530)(2e−27)
Problematic_alcohol_us...(36301540)(1e−09)
Maximum_habitual_alcoh...(36301540)(9e−09)
Drinks_per_week(36477530)(9e−29)
Alcohol_use_disorder...(37156939)(3e−09)
6 . rs35789697 : Experiencing_mood_swings(29500382)(3e−09)
7 . rs10048736 : Neuroticism(29942085)(1e−09)
Science,_technology,_e...(37165452)(1e−09)
8 . rs12991595 : Highest_math_class...(30038396)(7e−10)
Drinks_per_week(36477530)(3e−25)
9 . rs10204230 : Fruit_consumption(32066663)(9e−10)
Intelligence(29942086)(6e−09)
10 . rs4273169 : Cognitive_ability,_yea...(31374203)(3e−12)
Drinks_per_week(36477530)(3e−27)
..(only the first 10 catalog hits are shown).......
snp / p / or
a . rs35942385 / 2.45e−09 / 0.96
p = 5.0e−08
ARHGAP15
Observed (−logP)
Recombination rate (cM/Mb)
Chromosome 2 (kb)
144100
144200
144300
database: gwascatalog.Nov_2024.rp.txt, refGene.txt.Sep_2018.out

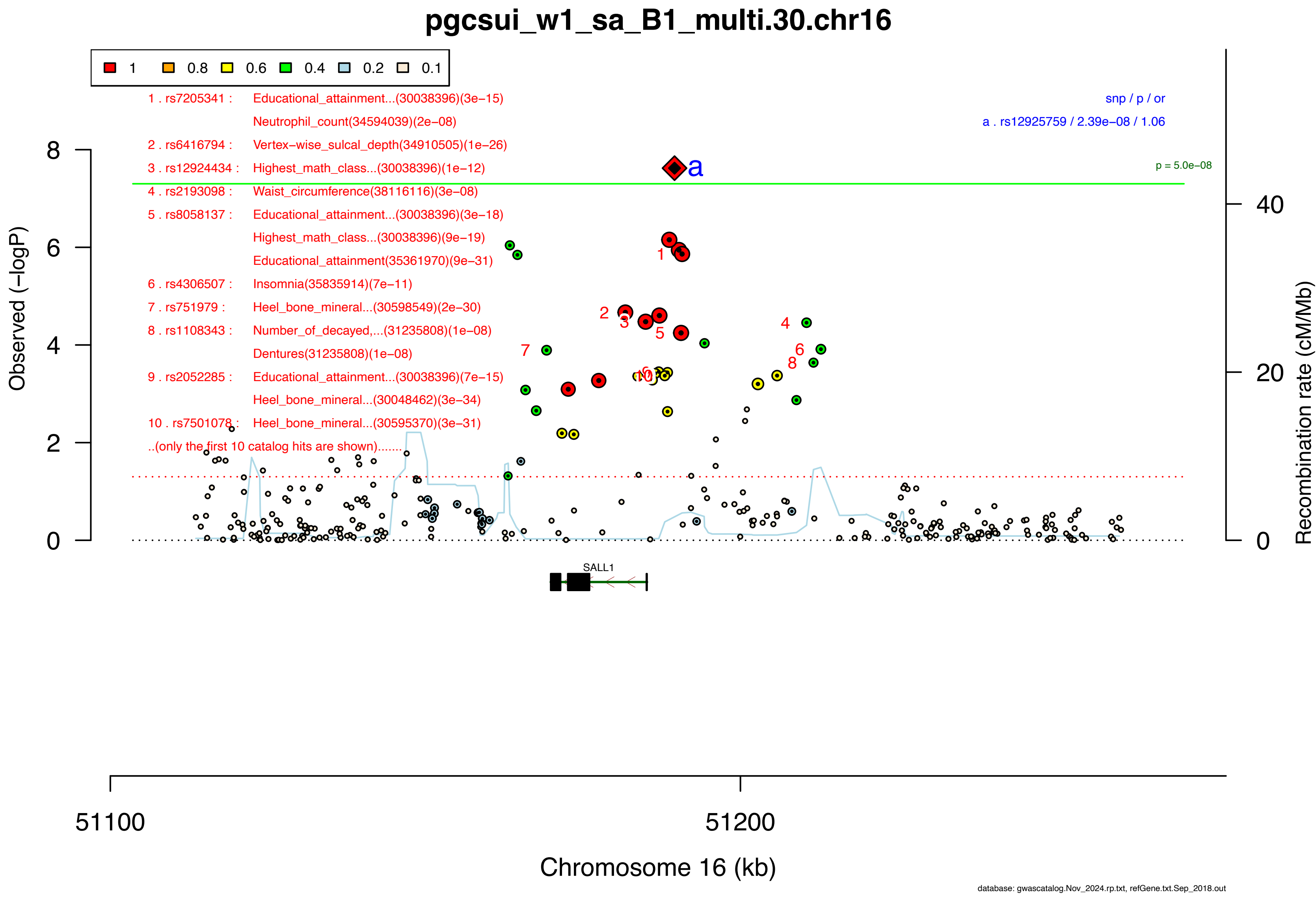
pgcsui_w1_sa_B1_multi.30.chr16
1
0.8
0.6
0.4
0.2
0.1
1 . rs7205341 : Educational_attainment...(30038396)(3e−15)
Neutrophil_count(34594039)(2e−08)
2 . rs6416794 : Vertex−wise_sulcal_depth(34910505)(1e−26)
3 . rs12924434 : Highest_math_class...(30038396)(1e−12)
4 . rs2193098 : Waist_circumference(38116116)(3e−08)
5 . rs8058137 : Educational_attainment...(30038396)(3e−18)
Highest_math_class...(30038396)(9e−19)
Educational_attainment(35361970)(9e−31)
6 . rs4306507 : Insomnia(35835914)(7e−11)
7 . rs751979 : Heel_bone_mineral...(30598549)(2e−30)
8 . rs1108343 : Number_of_decayed,...(31235808)(1e−08)
Dentures(31235808)(1e−08)
9 . rs2052285 : Educational_attainment...(30038396)(7e−15)
Heel_bone_mineral...(30048462)(3e−34)
10 . rs7501078 : Heel_bone_mineral...(30595370)(3e−31)
..(only the first 10 catalog hits are shown).......
snp / p / or
a . rs12925759 / 2.39e−08 / 1.06
p = 5.0e−08
Observed (−logP)
Recombination rate (cM/Mb)
SALL1
51100
51200
Chromosome 16 (kb)
database: gwascatalog.Nov_2024.rp.txt, refGene.txt.Sep_2018.out

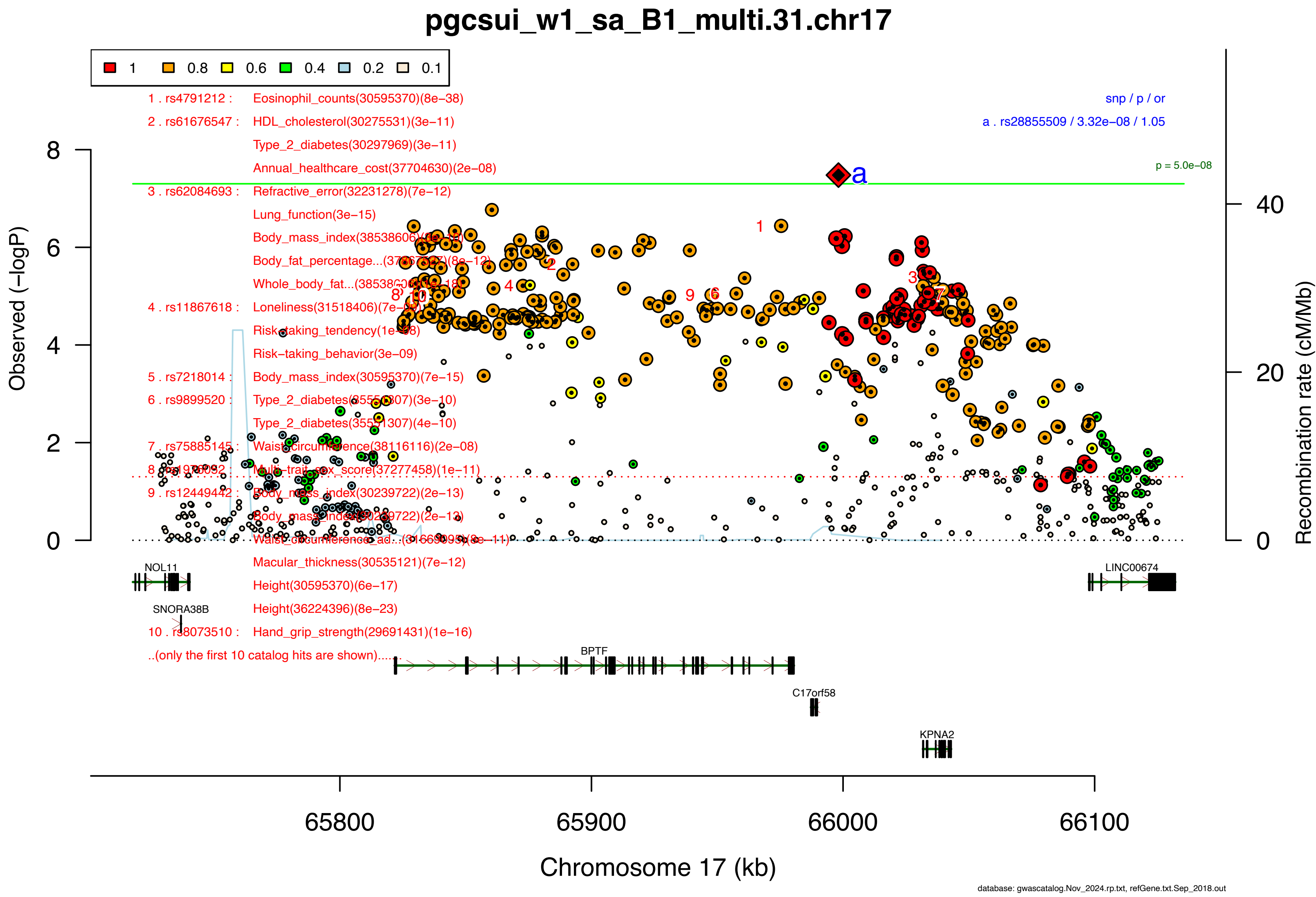
pgcsui_w1_sa_B1_multi.31.chr17
1 0.8 0.6 0.4 0.2 0.1
1 . rs4791212 : Eosinophil_counts(30595370)(8e−38)
2 . rs61676547 : HDL_cholesterol(30275531)(3e−11)
Type_2_diabetes(30297969)(3e−11)
Annual_healthcare_cost(37704630)(2e−08)
3 . rs62084693 : Refractive_error(32231278)(7e−12)
Lung_function(3e−15)
4 . rs11867618 : Loneliness(31518406)(7e−[illegible])
Risk-taking_tendency(1e−[illegible])
Risk−taking_behavior(3e−09)
5 . rs7218014 : Body_mass_index(30595370)(7e−15)
6 . rs9899520 : Type_2_diabetes(35551307)(3e−10)
Type_2_diabetes(35551307)(4e−10)
7 . rs75885145 : Waist_circumference(38116116)(2e−08)
Multi−trait_sex_score(37277458)(1e−11)
9 . rs12449442 : Body_mass_index(30239722)(2e−13)
Body_mass_index(30239722)(2e−13)
Macular_thickness(30535121)(7e−12)
Height(30595370)(6e−17)
Height(36224396)(8e−23)
10 . rs8073510 : Hand_grip_strength(29691431)(1e−16)
..(only the first 10 catalog hits are shown).....
snp / p / or
a . rs28855509 / 3.32e−08 / 1.05
p = 5.0e−08
Observed (−logP)
Recombination rate (cM/Mb)
NOL11
SNORA38B
BPTF
C17orf58
KPNA2
LINC00674
65800 65900 66000 66100
Chromosome 17 (kb)
database: gwascatalog.Nov_2024.rp.txt, refGene.txt.Sep_2018.out

### pgcsui_w1_sa_B1_multi.32.chr18

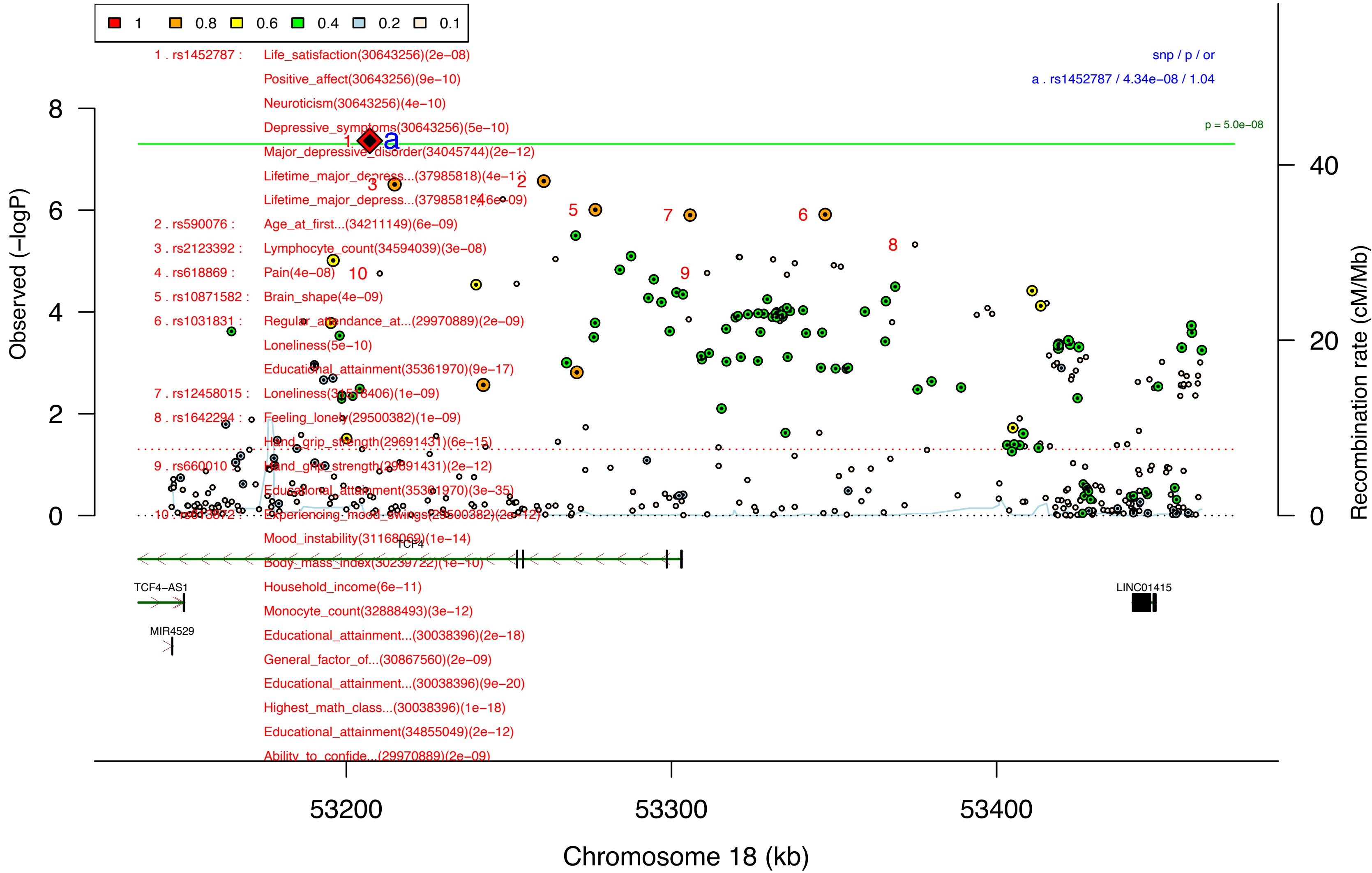


database: gwascatalog.Nov_2024.rp.txt, refGene.txt.Sep_2018.out

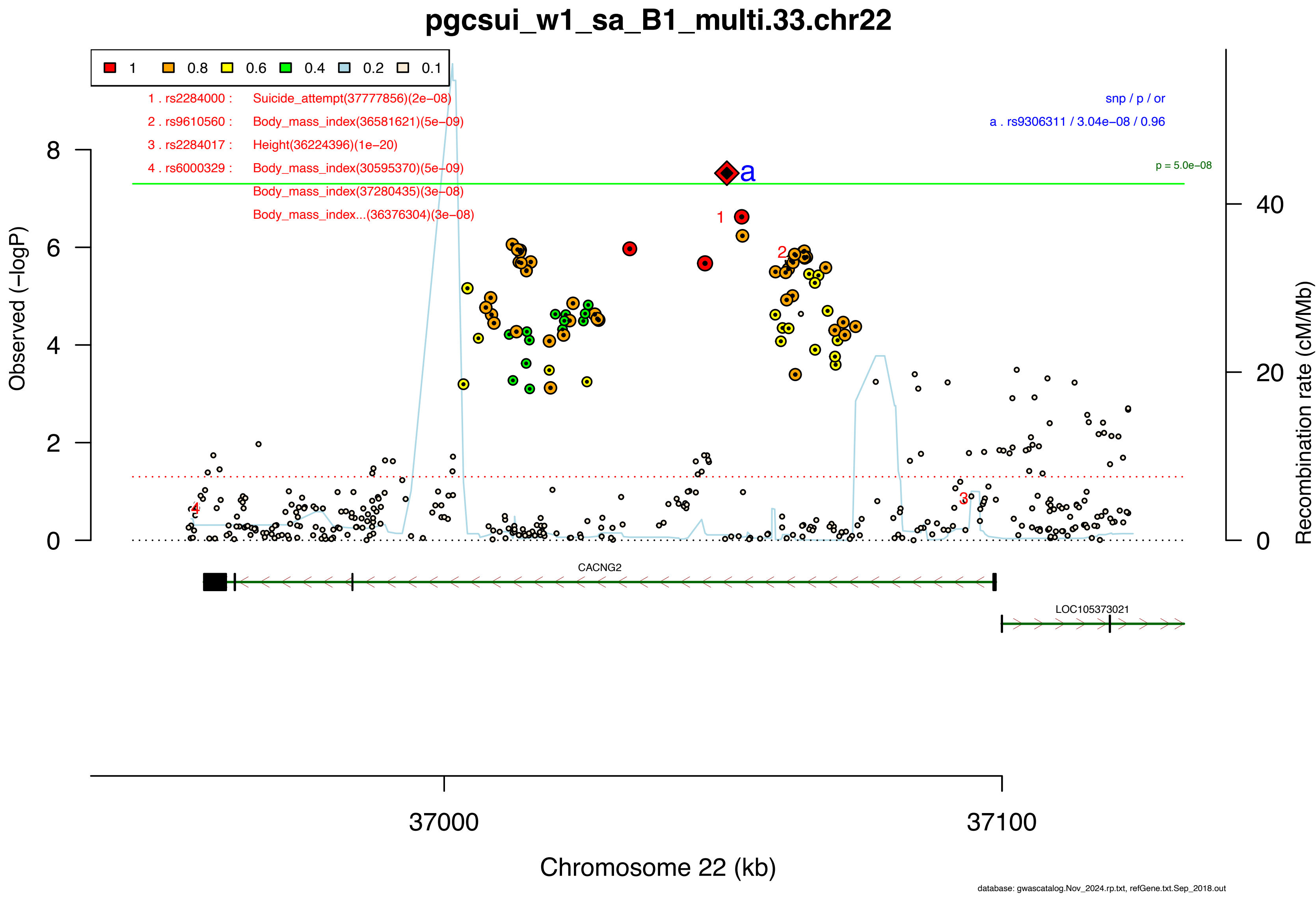

pgcsui_w1_sa_B1_multi.33.chr22
1
0.8
0.6
0.4
0.2
0.1
1 . rs2284000 : Suicide_attempt(37777856)(2e−08)
2 . rs9610560 : Body_mass_index(36581621)(5e−09)
3 . rs2284017 : Height(36224396)(1e−20)
4 . rs6000329 : Body_mass_index(30595370)(5e−09)
Body_mass_index(37280435)(3e−08)
Body_mass_index...(36376304)(3e−08)
snp / p / or
a . rs9306311 / 3.04e−08 / 0.96
p = 5.0e−08
Observed (−logP)
Recombination rate (cM/Mb)
CACNG2
LOC105373021
37000
37100
Chromosome 22 (kb)
database: gwascatalog.Nov_2024.rp.txt, refGene.txt.Sep_2018.out

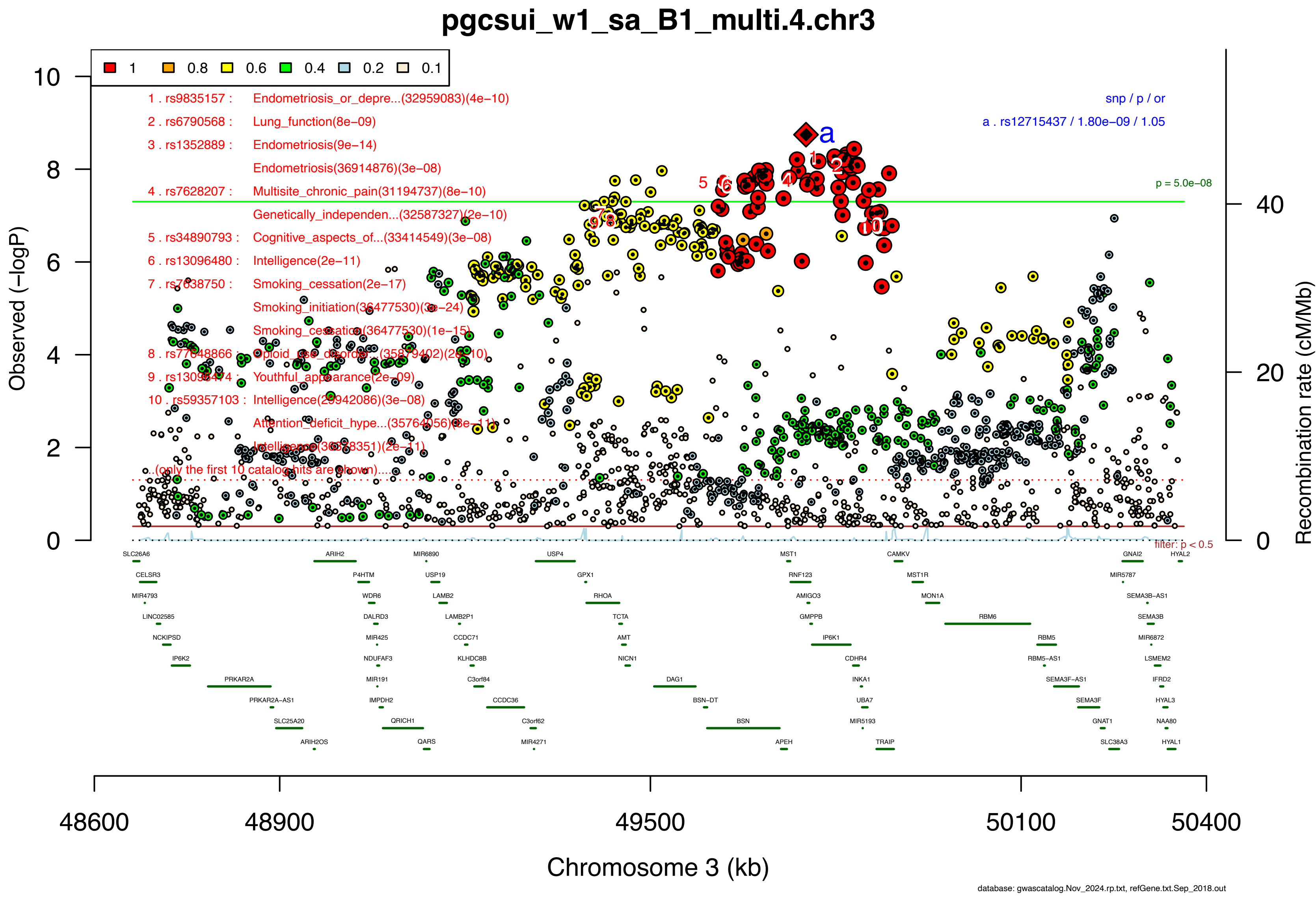

pgcsui_w1_sa_B1_multi.4.chr3
1 0.8 0.6 0.4 0.2 0.1
1 . rs9835157 : Endometriosis_or_depre...(32959083)(4e−10)
2 . rs6790568 : Lung_function(8e−09)
3 . rs1352889 : Endometriosis(9e−14)
Endometriosis(36914876)(3e−08)
4 . rs7628207 : Multisite_chronic_pain(31194737)(8e−10)
Genetically_independen...(32587327)(2e−10)
5 . rs34890793 : Cognitive_aspects_of...(33414549)(3e−08)
6 . rs13096480 : Intelligence(2e−11)
7 . rs7638750 : Smoking_cessation(2e−17)
Smoking_initiation(36477530)(3e−24)
Smoking_cessation(36477530)(1e−15)
8 . rs77648866 : Opioid_use_disorde...(35879402)(2e−10)
9 . rs13096474 : Youthful_appearance(2e−09)
10 . rs59357103 : Intelligence(29942086)(3e−08)
Attention_deficit_hype...(35764056)(8e−11)
Intelligence(30278351)(2e−11)
...(only the first 10 catalog hits are shown)...
snp / p / or
a . rs12715437 / 1.80e−09 / 1.05
p = 5.0e−08
filter: p < 0.5
Observed (−logP)
Recombination rate (cM/Mb)
Chromosome 3 (kb)
48600 48900 49500 50100 50400
database: gwascatalog.Nov_2024.rp.txt, refGene.txt.Sep_2018.out

### pgcsui_w1_sa_B1_multi.5.chr3

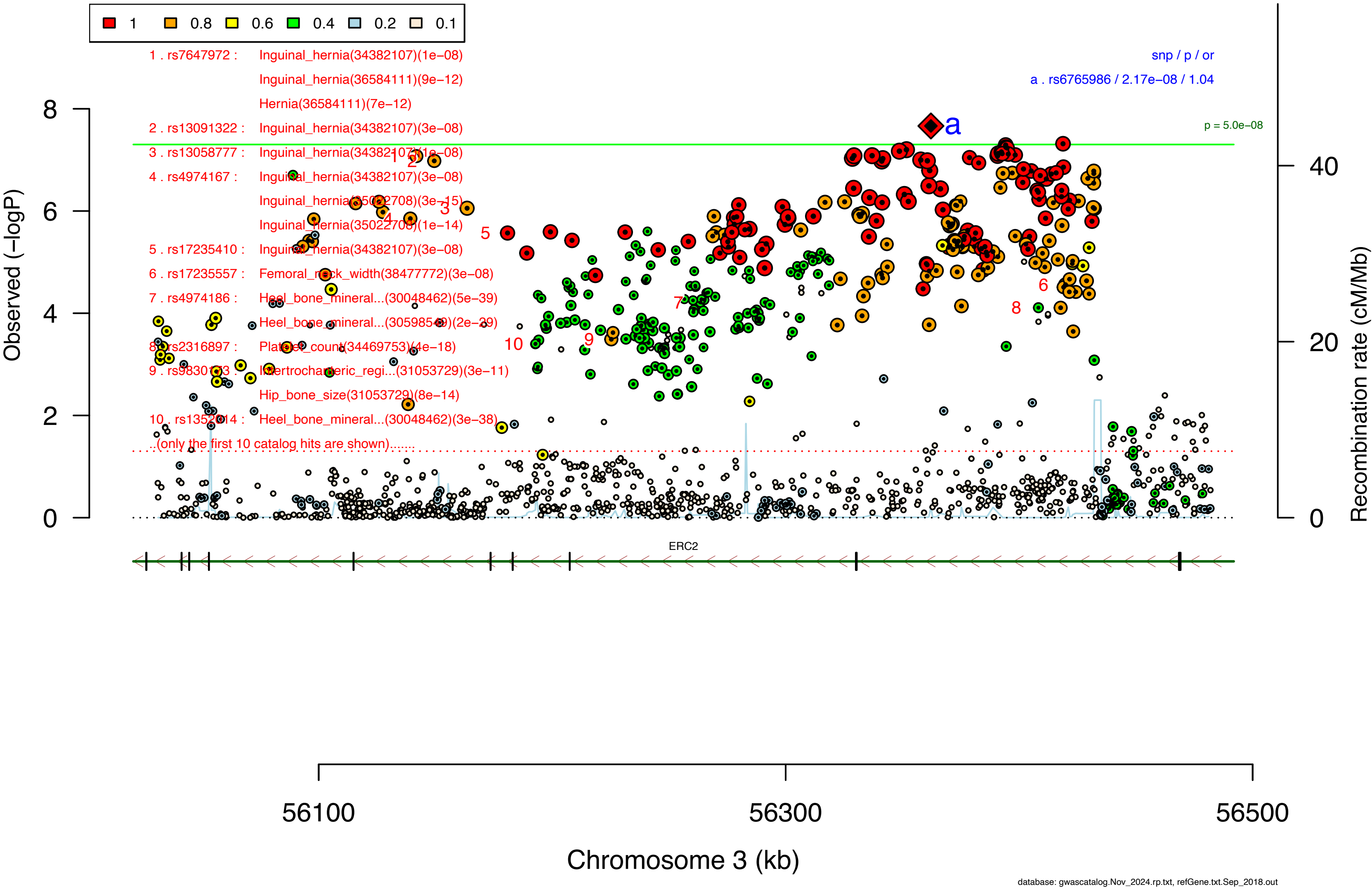

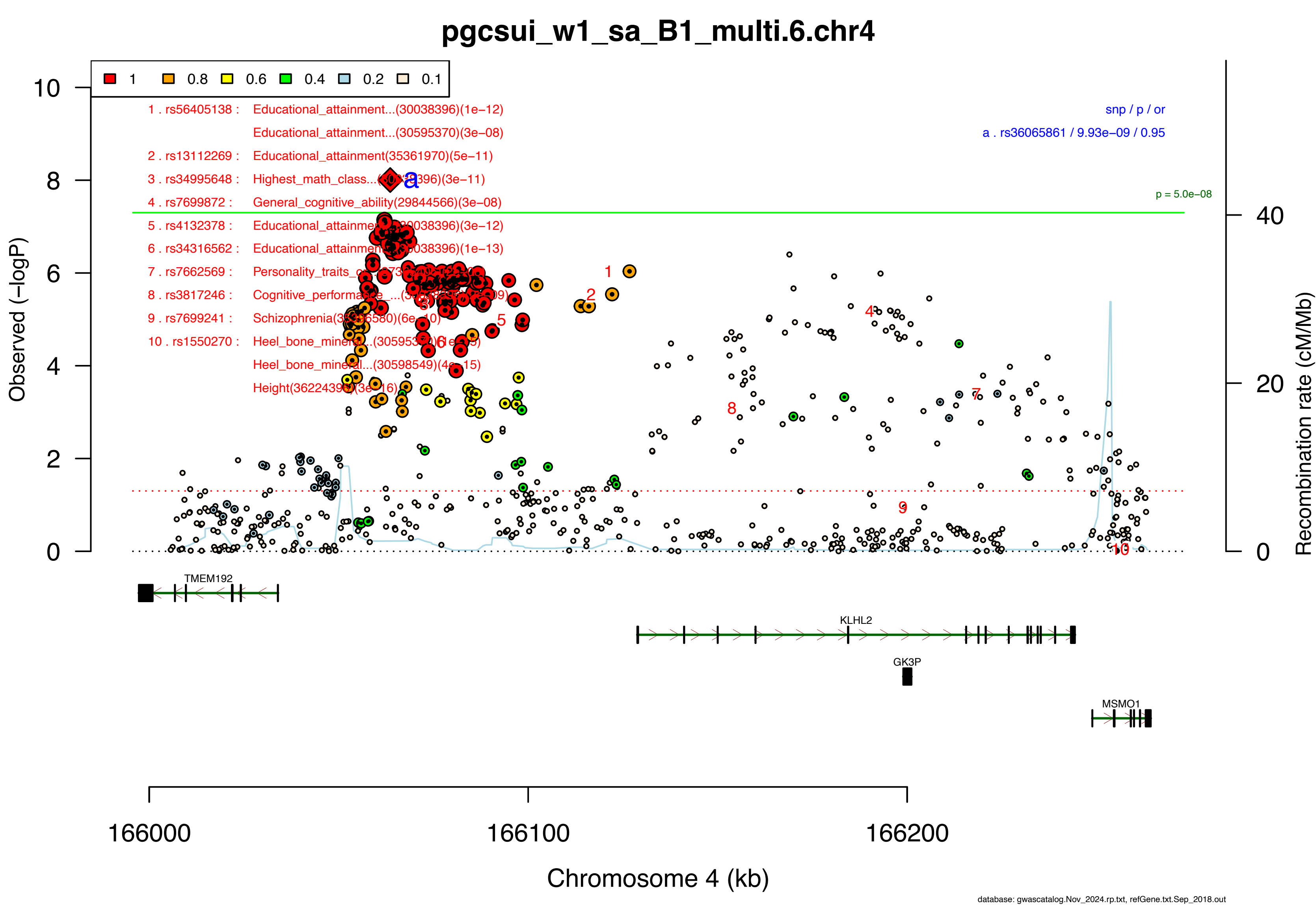
pgcsui_w1_sa_B1_multi.6.chr4
1
0.8
0.6
0.4
0.2
0.1
1 . rs56405138 : Educational_attainment...(30038396)(1e−12)
Educational_attainment...(30595370)(3e−08)
2 . rs13112269 : Educational_attainment(35361970)(5e−11)
3 . rs34995648 : Highest_math_class...
4 . rs7699872 : General_cognitive_ability(29844566)(3e−08)
5 . rs4132378 : Educational_attainme
6 . rs34316562 : Educational_attainmen
7 . rs7662569 : Personality_traits_
8 . rs3817246 : Cognitive_performance
9 . rs7699241 : Schizophrenia
10 . rs1550270 : Heel_bone_mine
Heel_bone_mine
Height
snp / p / or
a . rs36065861 / 9.93e−09 / 0.95
p = 5.0e−08
Observed (−logP)
Recombination rate (cM/Mb)
TMEM192
KLHL2
GK3P
MSMO1
166000
166100
166200
Chromosome 4 (kb)
database: gwascatalog.Nov_2024.rp.txt, refGene.txt.Sep_2018.out

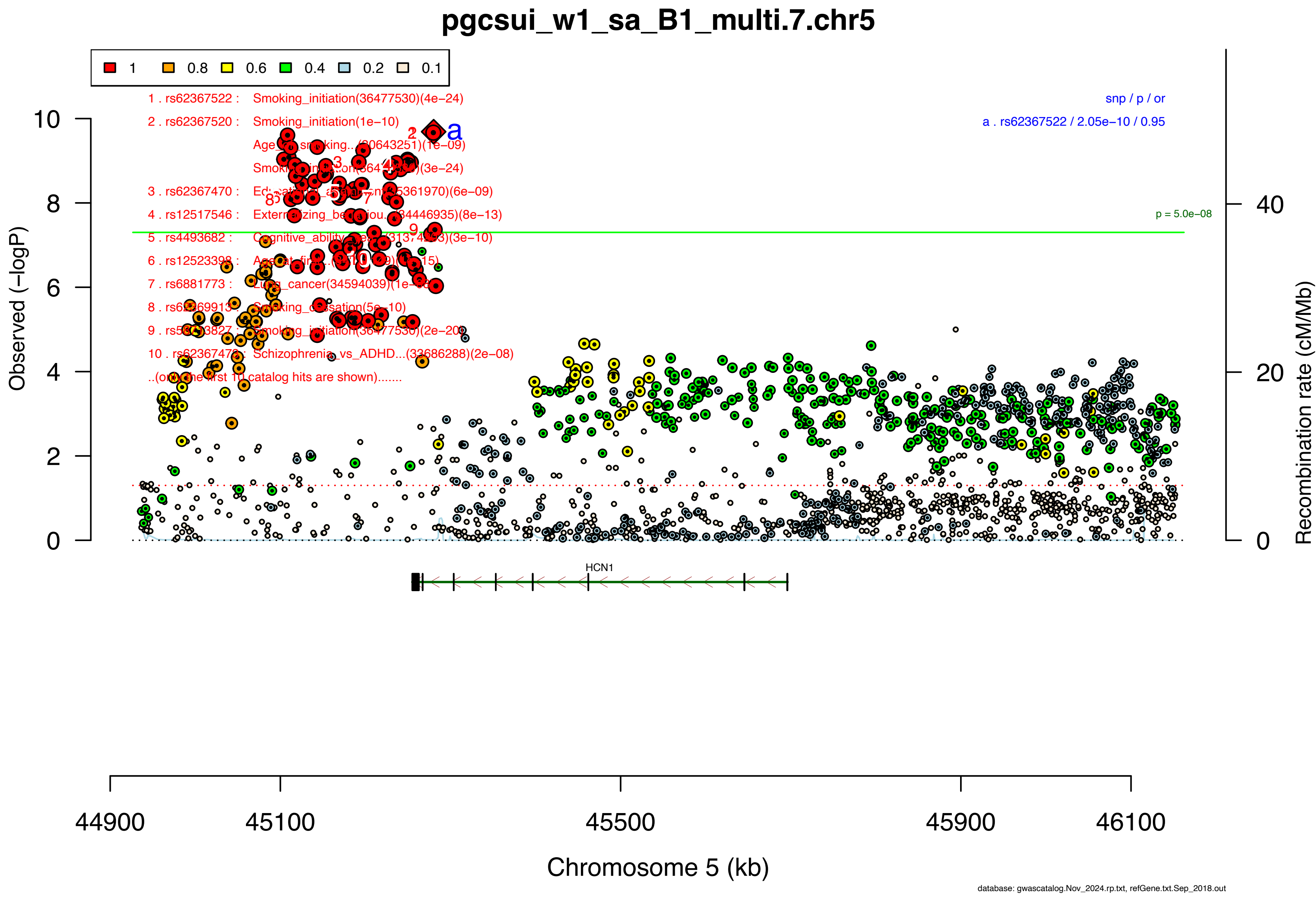

pgcsui_w1_sa_B1_multi.7.chr5
1
0.8
0.6
0.4
0.2
0.1
1 . rs62367522 : Smoking_initiation(36477530)(4e−24)
2 . rs62367520 : Smoking_initiation(1e−10)
Smoking_initiation(36477530)(3e−24)
3 . rs62367470 :
4 . rs12517546 :
5 . rs4493682 :
6 . rs12523398 :
7 . rs6881773 :
8 . rs62369913 : Smoking_cessation(5e−10)
9 . rs5
10 . rs62367473 : Schizophrenia_vs_ADHD...(33686288)(2e−08)
..(only the first 10 catalog hits are shown).......
snp / p / or
a . rs62367522 / 2.05e−10 / 0.95
p = 5.0e−08
HCN1
Observed (−logP)
Recombination rate (cM/Mb)
Chromosome 5 (kb)
44900
45100
45500
45900
46100
database: gwascatalog.Nov_2024.rp.txt, refGene.txt.Sep_2018.out

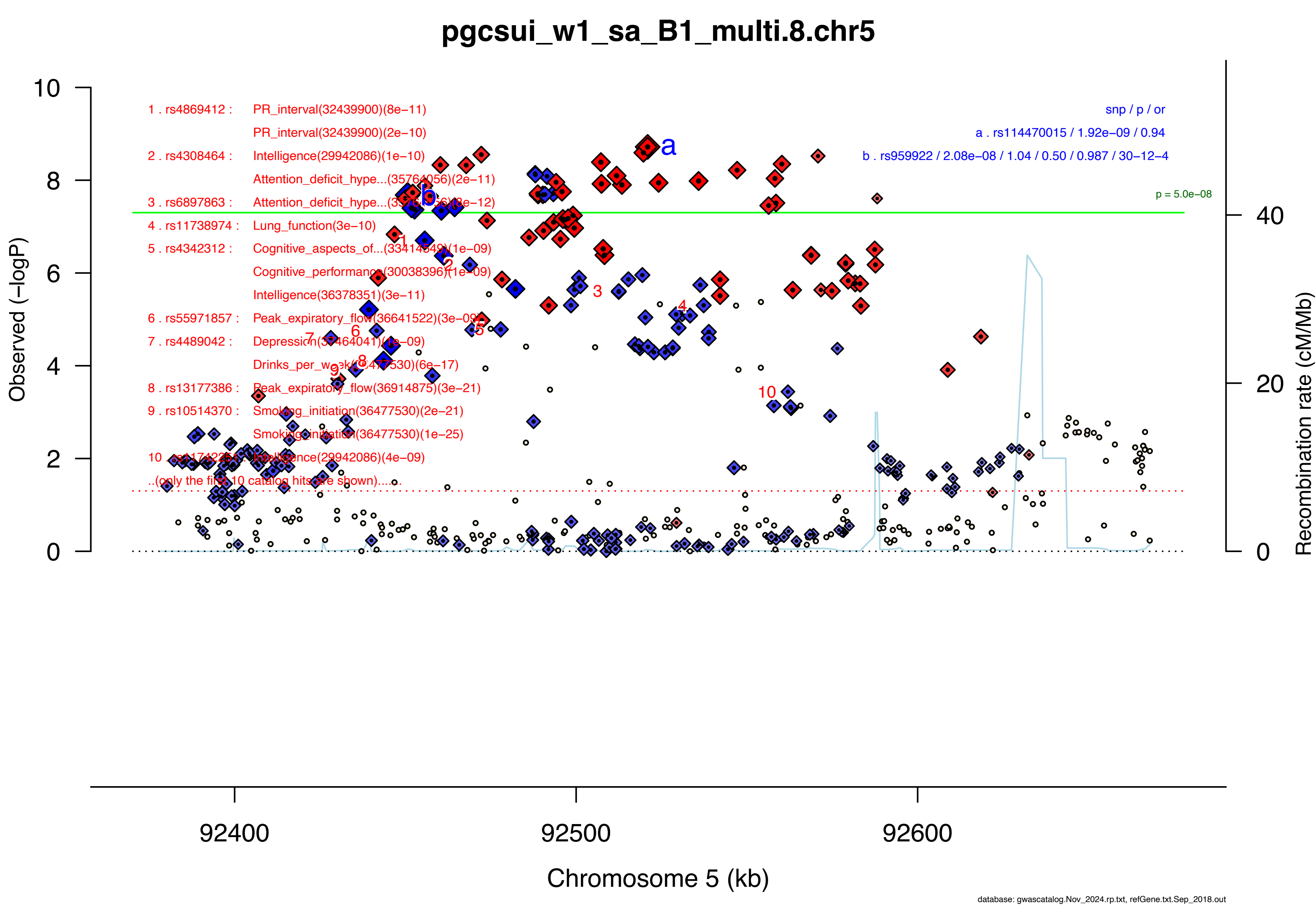
pgcsui_w1_sa_B1_multi.8.chr5
1 . rs4869412 : PR_interval(32439900)(8e−11)
PR_interval(32439900)(2e−10)
2 . rs4308464 : Intelligence(29942086)(1e−10)
Attention_deficit_hype...(35764056)(2e−11)
3 . rs6897863 : Attention_deficit_hype...(35764056)(8e−12)
4 . rs11738974 : Lung_function(3e−10)
5 . rs4342312 : Cognitive_aspects_of...(33414549)(1e−09)
Cognitive_performance(30038396)(1e−09)
Intelligence(36378351)(3e−11)
6 . rs55971857 : Peak_expiratory_flow(36641522)(3e−09)
7 . rs4489042 : Depression(37464041)(1e−09)
Drinks_per_week(36477530)(6e−17)
8 . rs13177386 : Peak_expiratory_flow(36914875)(3e−21)
9 . rs10514370 : Smoking_initiation(36477530)(2e−21)
Smoking_initiation(36477530)(1e−25)
10 . rs11742251 : Intelligence(29942086)(4e−09)
..(only the first 10 catalog hits are shown).....
snp / p / or
a . rs114470015 / 1.92e−09 / 0.94
b . rs959922 / 2.08e−08 / 1.04 / 0.50 / 0.987 / 30−12−4
p = 5.0e−08
Observed (−logP)
Recombination rate (cM/Mb)
Chromosome 5 (kb)
92400
92500
92600
database: gwascatalog.Nov_2024.rp.txt, refGene.txt.Sep_2018.out

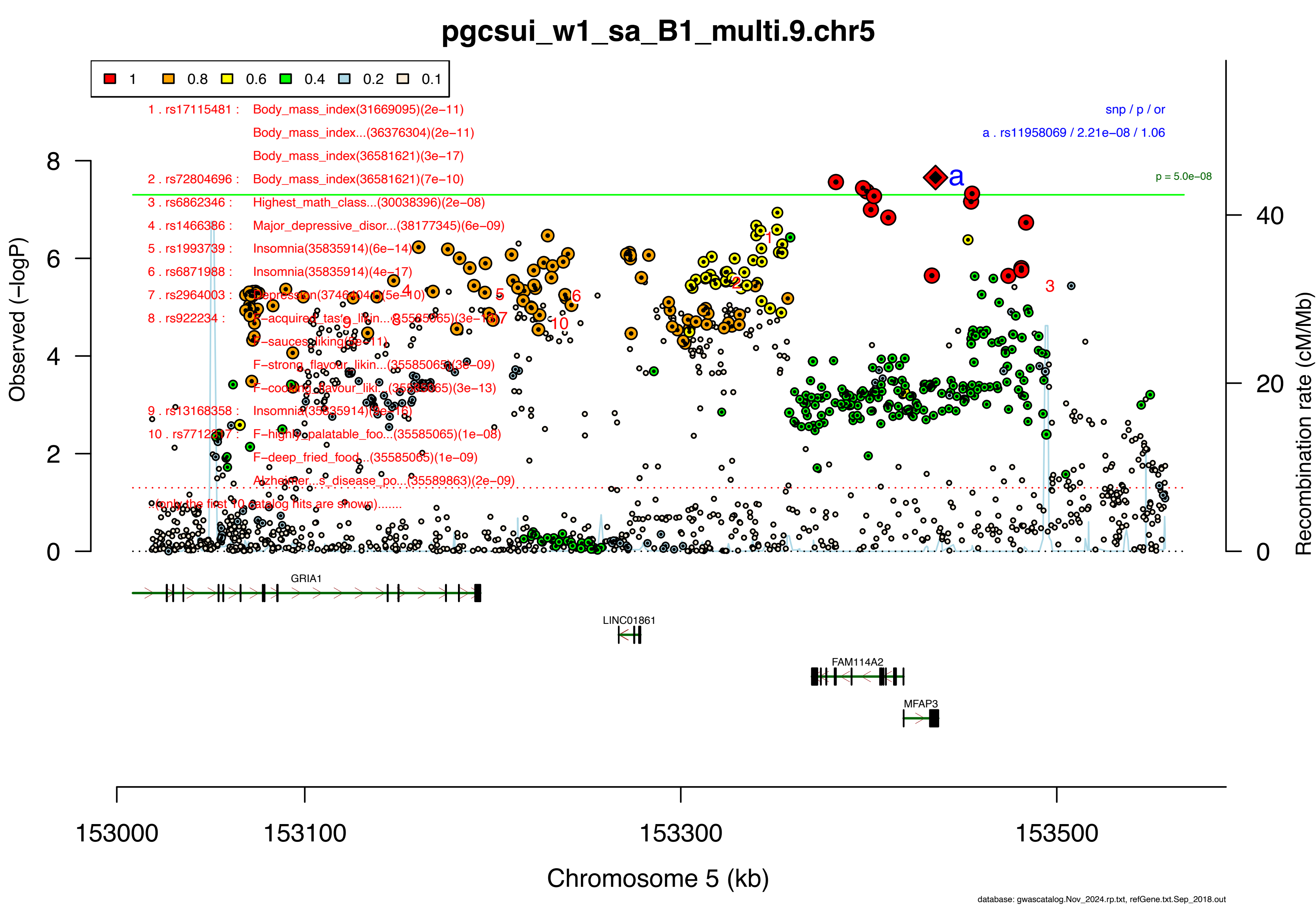

pgcsui_w1_sa_B1_multi.9.chr5
1 0.8 0.6 0.4 0.2 0.1
1 . rs17115481 : Body_mass_index(31669095)(2e−11)
Body_mass_index...(36376304)(2e−11)
Body_mass_index(36581621)(3e−17)
2 . rs72804696 : Body_mass_index(36581621)(7e−10)
3 . rs6862346 : Highest_math_class...(30038396)(2e−08)
4 . rs1466386 : Major_depressive_disor...(38177345)(6e−09)
5 . rs1993739 : Insomnia(35835914)(6e−14)
6 . rs6871988 : Insomnia(35835914)(4e−17)
7 . rs2964003 : Depression(37464041)(5e−10)
8 . rs922234 : F−acquired_taste_likin...(35585065)(3e−18)
F−sauces_liking(9e−11)
F−strong_flavour_likin...(35585065)(3e−09)
F−cooking_flavour_liki...(35585065)(3e−13)
9 . rs13168358 : Insomnia(35835914)(3e−16)
10 . rs7712817 : F−highly_palatable_foo...(35585065)(1e−08)
F−deep_fried_food...(35585065)(1e−09)
Alzheimer...s_disease_po...(35589863)(2e−09)
(only the first 10 catalog hits are shown).......
snp / p / or
a . rs11958069 / 2.21e−08 / 1.06
p = 5.0e−08
Observed (−logP)
Recombination rate (cM/Mb)
GRIA1
LINC01861
FAM114A2
MFAP3
153000 153100 153300 153500
Chromosome 5 (kb)
database: gwascatalog.Nov_2024.rp.txt, refGene.txt.Sep_2018.out

**Supplementary Data 2E: Area plots of lead SNPs at the 35 genome-wide significant loci from the GWAS meta-analysis of suicide attempt in European ancestry samples.**

If >1 independent SNPs are in the same plot, then each is given a different color, and their LD partners are shaded with the same color. Detailed info about each independent index SNP (a., b., c. ...) is provided in the right upper corner. If SNPs share independent index SNPs as an LD partner, the SNP is assigned to the more significant one. Information regarding other GWAS associations at the locus is provided in the left upper corner.

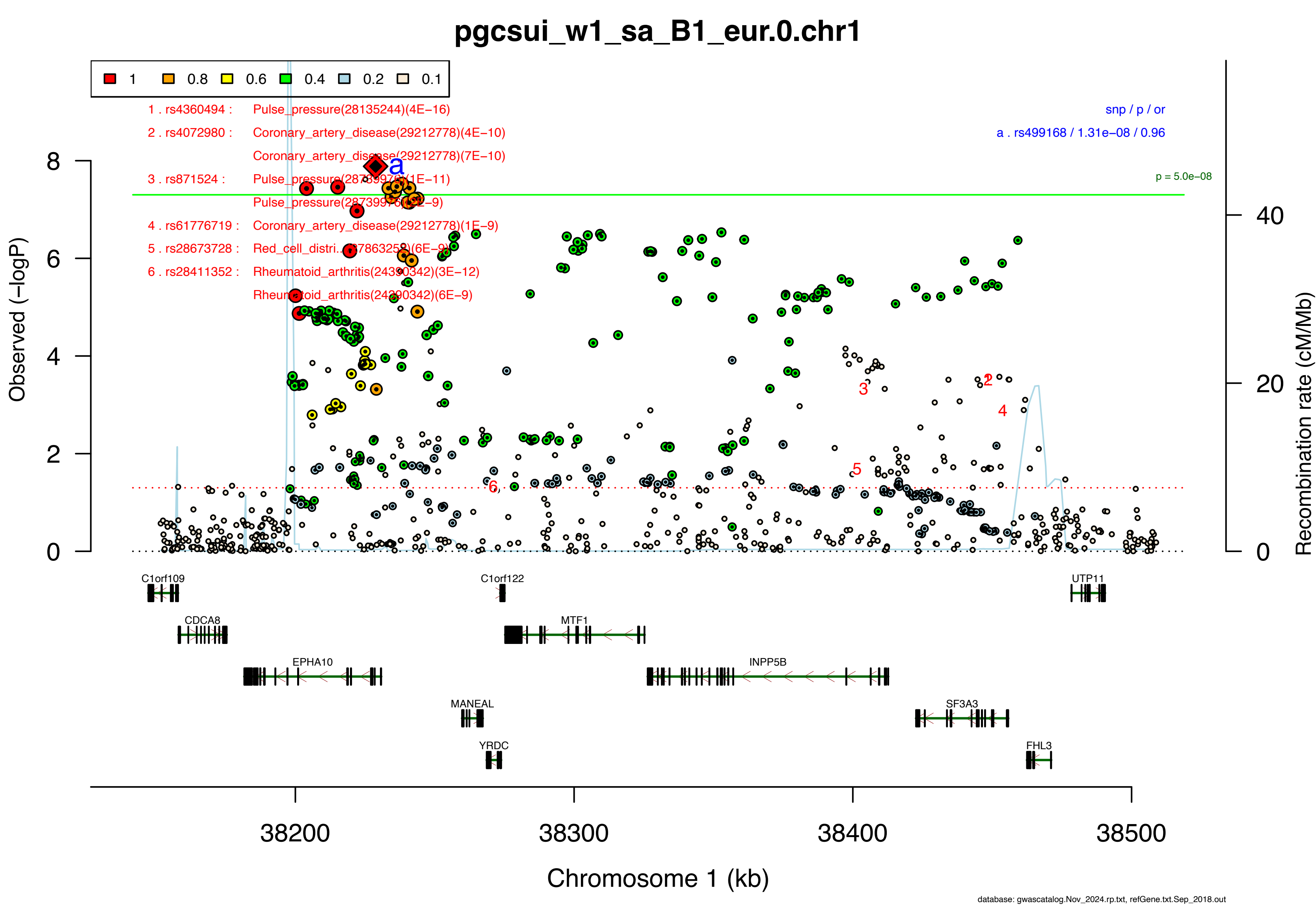

pgcsui_w1_sa_B1_eur.0.chr1
1
0.8
0.6
0.4
0.2
0.1
1 . rs4360494 : Pulse_pressure(28135244)(4E−16)
2 . rs4072980 : Coronary_artery_disease(29212778)(4E−10)
Coronary_artery_disease(29212778)(7E−10)
3 . rs871524 : Pulse_pressure(28739976)(1E−11)
Pulse_pressure(28739976)(2E−9)
4 . rs61776719 : Coronary_artery_disease(29212778)(1E−9)
5 . rs28673728 : Red_cell_distri...(27863252)(6E−9)
6 . rs28411352 : Rheumatoid_arthritis(24390342)(3E−12)
Rheumatoid_arthritis(24390342)(6E−9)
snp / p / or
a . rs499168 / 1.31e−08 / 0.96
p = 5.0e−08
Observed (−logP)
Recombination rate (cM/Mb)
C1orf109
C1orf122
UTP11
CDCA8
MTF1
EPHA10
INPP5B
MANEAL
SF3A3
YRDC
FHL3
38200
38300
38400
38500
Chromosome 1 (kb)
database: gwascatalog.Nov_2024.rp.txt, refGene.txt.Sep_2018.out

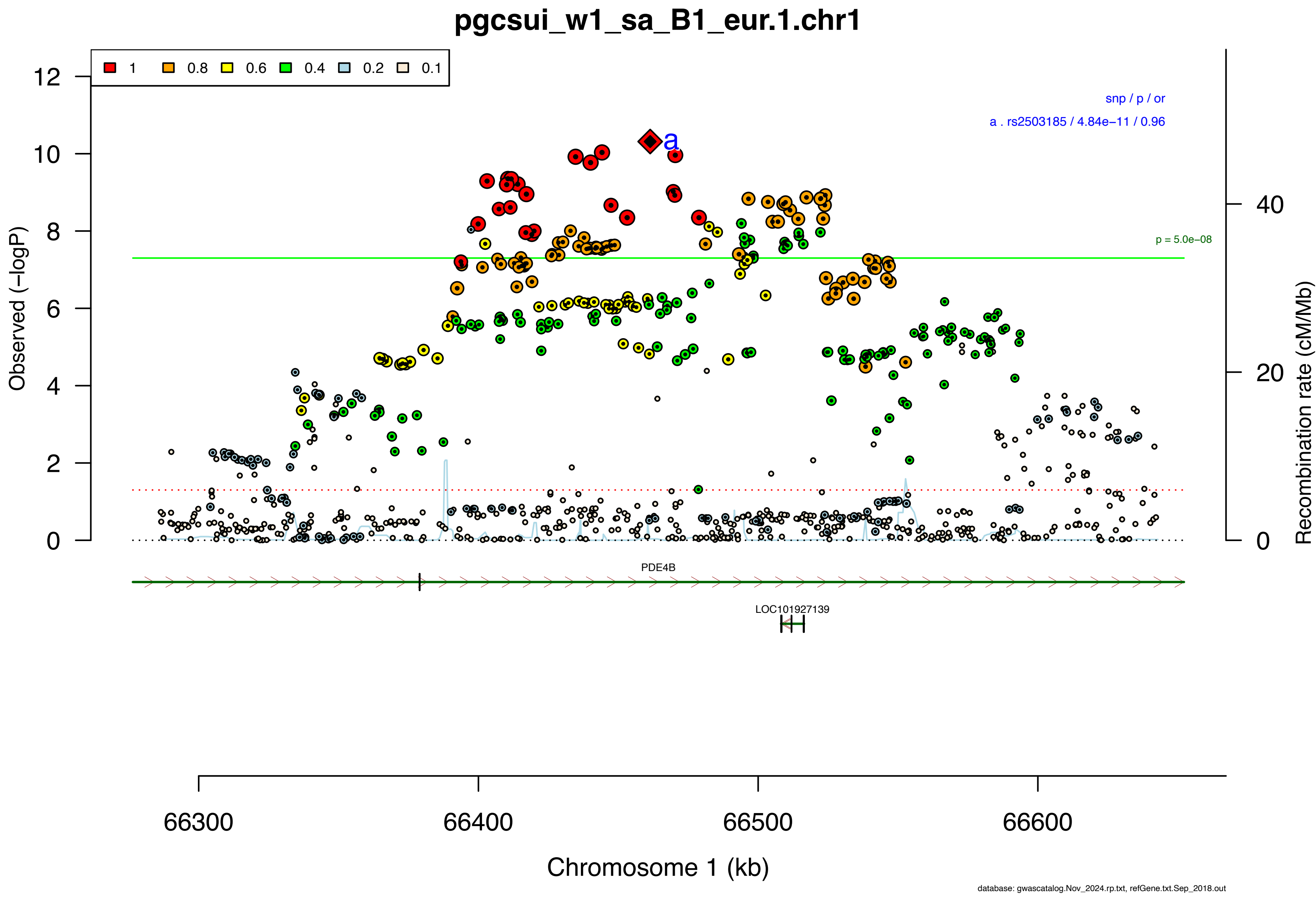

pgcsui_w1_sa_B1_eur.1.chr1
1
0.8
0.6
0.4
0.2
0.1
snp / p / or
a . rs2503185 / 4.84e−11 / 0.96
a
p = 5.0e−08
Observed (−logP)
Recombination rate (cM/Mb)
0
2
4
6
8
10
12
20
40
PDE4B
LOC101927139
66300
66400
66500
66600
Chromosome 1 (kb)
database: gwascatalog.Nov_2024.rp.txt, refGene.txt.Sep_2018.out

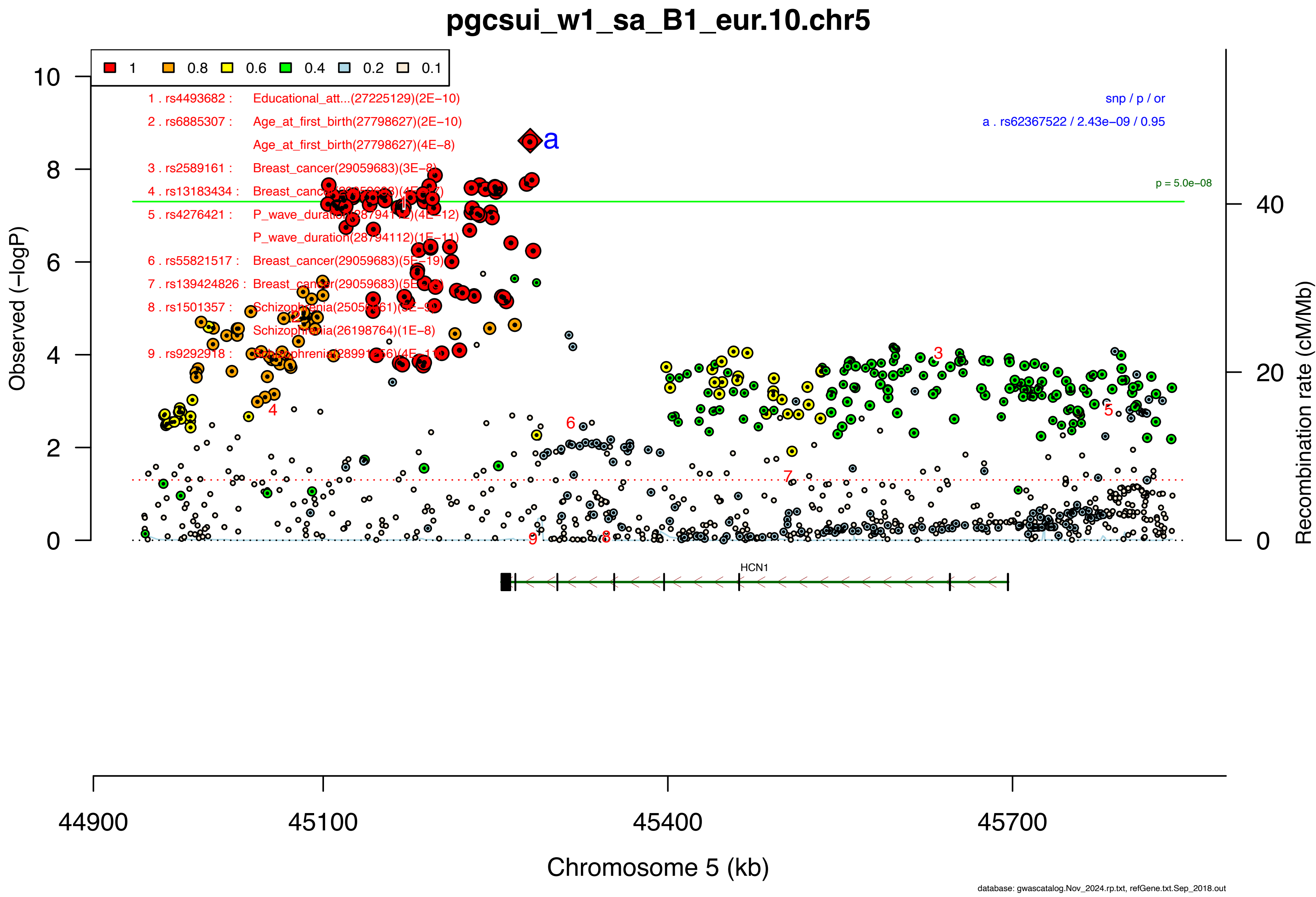

pgcsui_w1_sa_B1_eur.10.chr5
1
0.8
0.6
0.4
0.2
0.1
1 . rs4493682 : Educational_att...(27225129)(2E−10)
2 . rs6885307 : Age_at_first_birth(27798627)(2E−10)
Age_at_first_birth(27798627)(4E−8)
3 . rs2589161 : Breast_cancer(29059683)(3E−8)
4 . rs13183434 : Breast_cancer(29059683)(4E−7)
5 . rs4276421 : P_wave_duration(28794112)(4E−12)
P_wave_duration(28794112)(1E−11)
6 . rs55821517 : Breast_cancer(29059683)(5E−19)
7 . rs139424826 : Breast_cancer(29059683)(5E−8)
8 . rs1501357 : Schizophrenia(25056061)(3E−9)
Schizophrenia(26198764)(1E−8)
9 . rs9292918 : Schizophrenia(28991256)(4E−11)
snp / p / or
a . rs62367522 / 2.43e−09 / 0.95
a
p = 5.0e−08
Observed (−logP)
Recombination rate (cM/Mb)
HCN1
44900
45100
45400
45700
Chromosome 5 (kb)
database: gwascatalog.Nov_2024.rp.txt, refGene.txt.Sep_2018.out

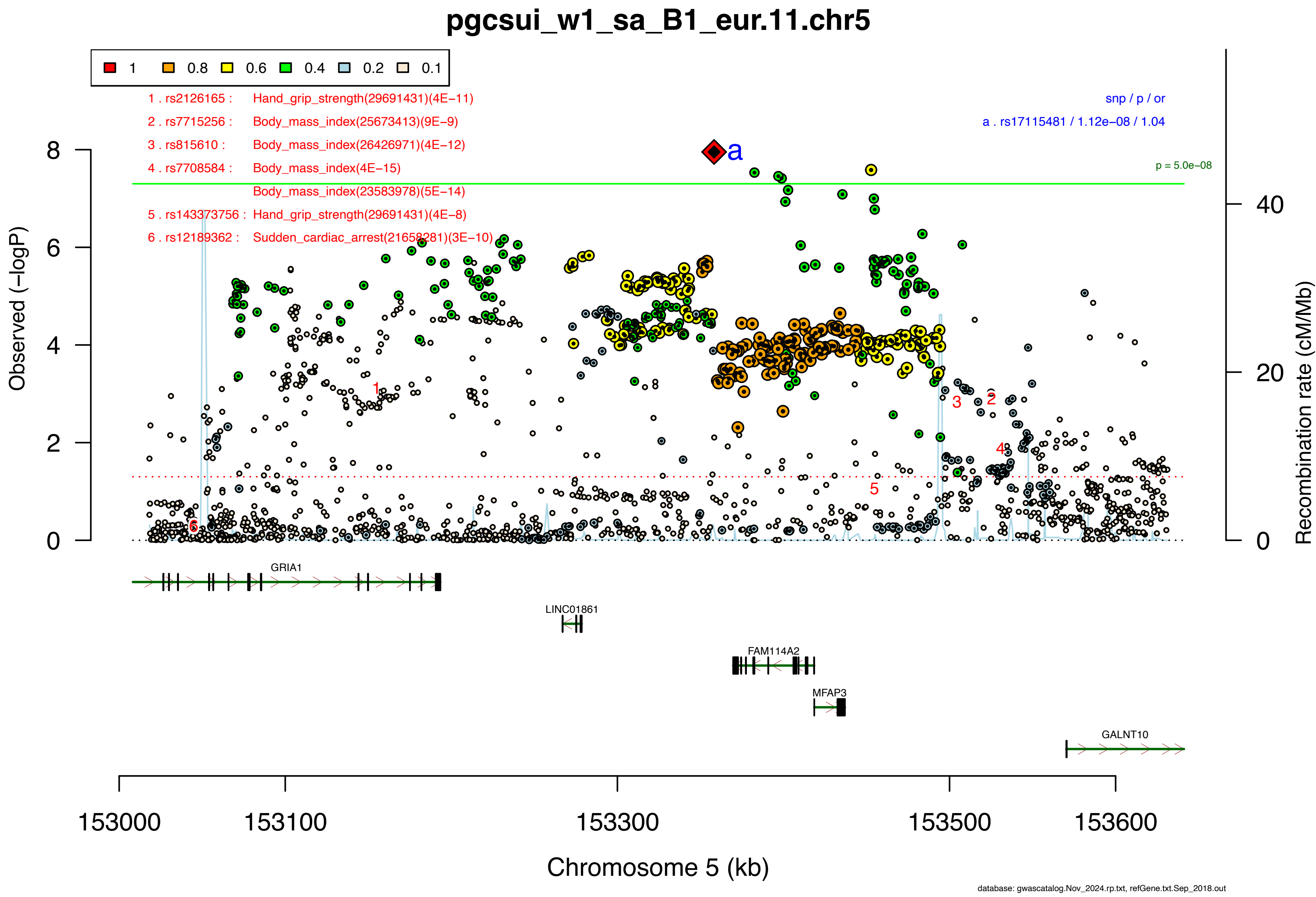

pgcsui_w1_sa_B1_eur.11.chr5
1 0.8 0.6 0.4 0.2 0.1
1 . rs2126165 : Hand_grip_strength(29691431)(4E−11)
2 . rs7715256 : Body_mass_index(25673413)(9E−9)
3 . rs815610 : Body_mass_index(26426971)(4E−12)
4 . rs7708584 : Body_mass_index(4E−15)
Body_mass_index(23583978)(5E−14)
5 . rs143373756 : Hand_grip_strength(29691431)(4E−8)
6 . rs12189362 : Sudden_cardiac_arrest(21658281)(3E−10)
snp / p / or
a . rs17115481 / 1.12e−08 / 1.04
p = 5.0e−08
Observed (−logP)
Recombination rate (cM/Mb)
GRIA1
LINC01861
FAM114A2
MFAP3
GALNT10
153000 153100 153300 153500 153600
Chromosome 5 (kb)
database: gwascatalog.Nov_2024.rp.txt, refGene.txt.Sep_2018.out

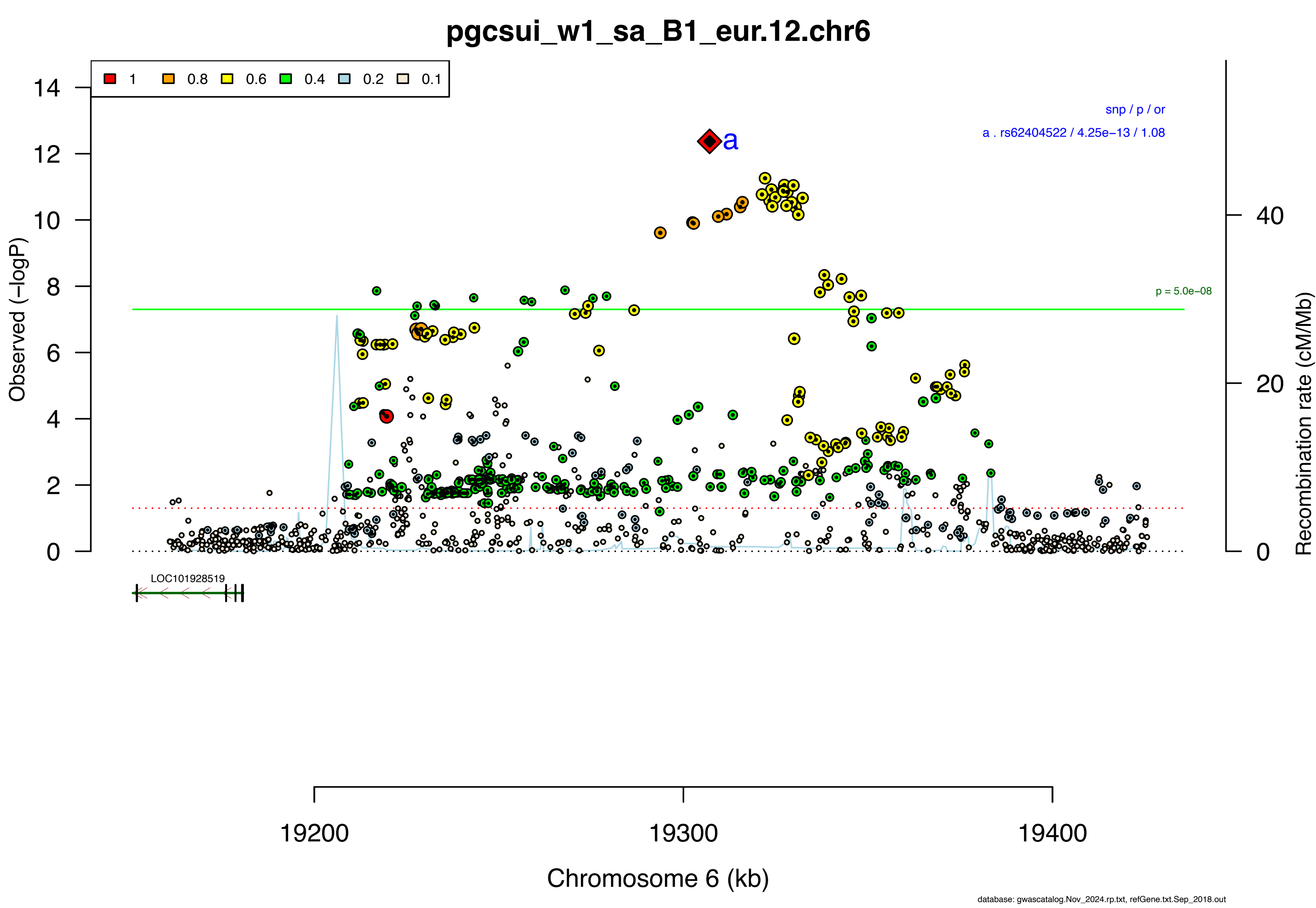

pgcsui_w1_sa_B1_eur.12.chr6
1
0.8
0.6
0.4
0.2
0.1
snp / p / or
a . rs62404522 / 4.25e−13 / 1.08
a
p = 5.0e−08
Observed (−logP)
Recombination rate (cM/Mb)
LOC101928519
19200
19300
19400
Chromosome 6 (kb)
database: gwascatalog.Nov_2024.rp.txt, refGene.txt.Sep_2018.out

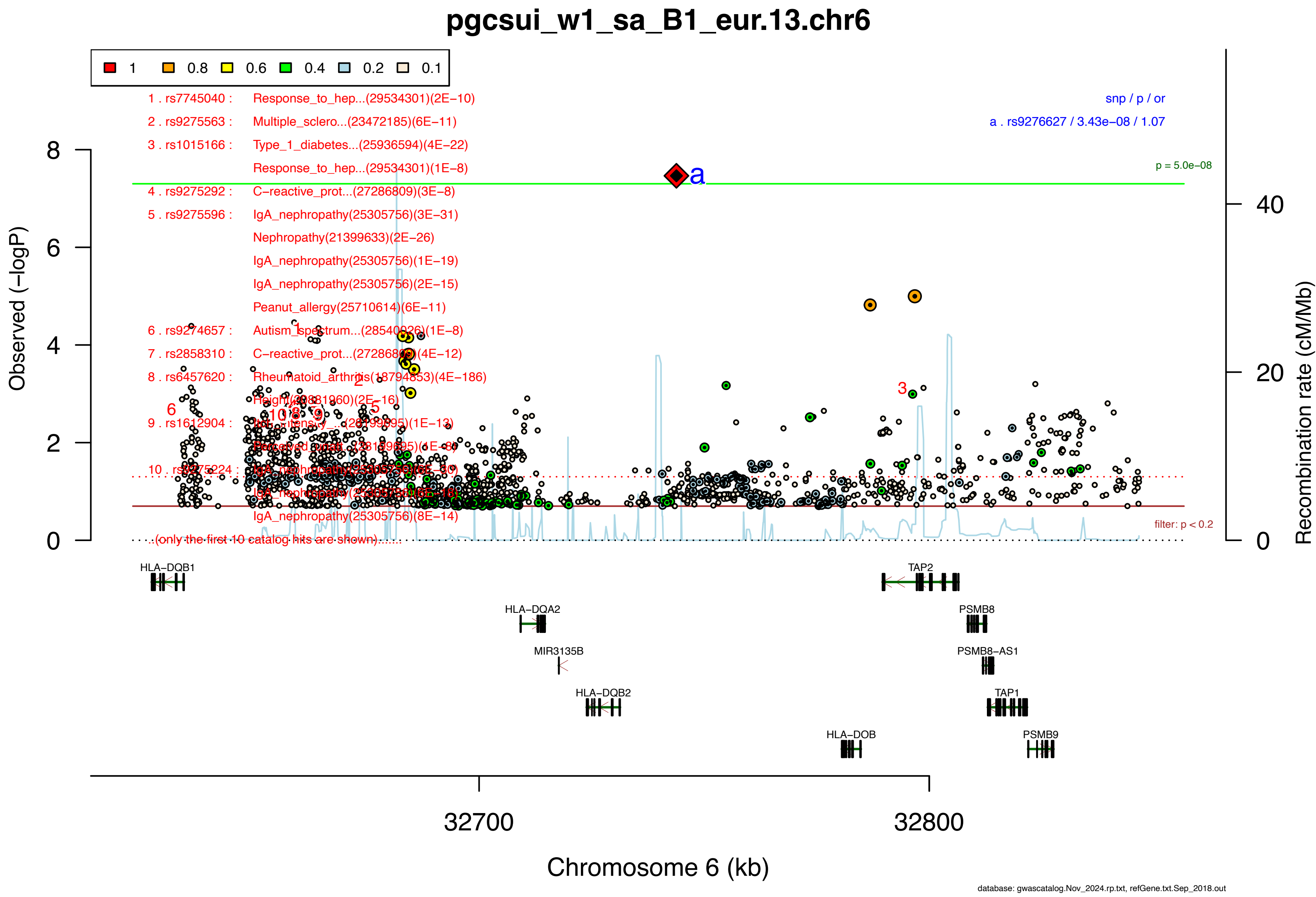

pgcsui_w1_sa_B1_eur.13.chr6
1
0.8
0.6
0.4
0.2
0.1
1 . rs7745040 : Response_to_hep...(29534301)(2E−10)
2 . rs9275563 : Multiple_sclero...(23472185)(6E−11)
3 . rs1015166 : Type_1_diabetes...(25936594)(4E−22)
Response_to_hep...(29534301)(1E−8)
4 . rs9275292 : C−reactive_prot...(27286809)(3E−8)
5 . rs9275596 : IgA_nephropathy(25305756)(3E−31)
Nephropathy(21399633)(2E−26)
IgA_nephropathy(25305756)(1E−19)
IgA_nephropathy(25305756)(2E−15)
Peanut_allergy(25710614)(6E−11)
6 . rs9274657 : Autism_spectrum...(28540026)(1E−8)
7 . rs2858310 : C−reactive_prot...(27286809)(4E−12)
8 . rs6457620 : Rheumatoid_arthritis(18794853)(4E−186)
Height(20881960)(2E−16)
9 . rs1612904 : Pain_intensity...(28199695)(1E−13)
Perceived_unpl...(28199695)(1E−8)
10 . rs9275224 : IgA_nephropathy(25305756)(6E−30)
IgA_nephropathy(25305756)(6E−16)
IgA_nephropathy(25305756)(8E−14)
...(only the first 10 catalog hits are shown)...
snp / p / or
a . rs9276627 / 3.43e−08 / 1.07
p = 5.0e−08
filter: p < 0.2
Observed (−logP)
Recombination rate (cM/Mb)
HLA−DQB1
HLA−DQA2
MIR3135B
HLA−DQB2
HLA−DOB
TAP2
PSMB8
PSMB8−AS1
TAP1
PSMB9
32700
32800
Chromosome 6 (kb)
database: gwascatalog.Nov_2024.rp.txt, refGene.txt.Sep_2018.out

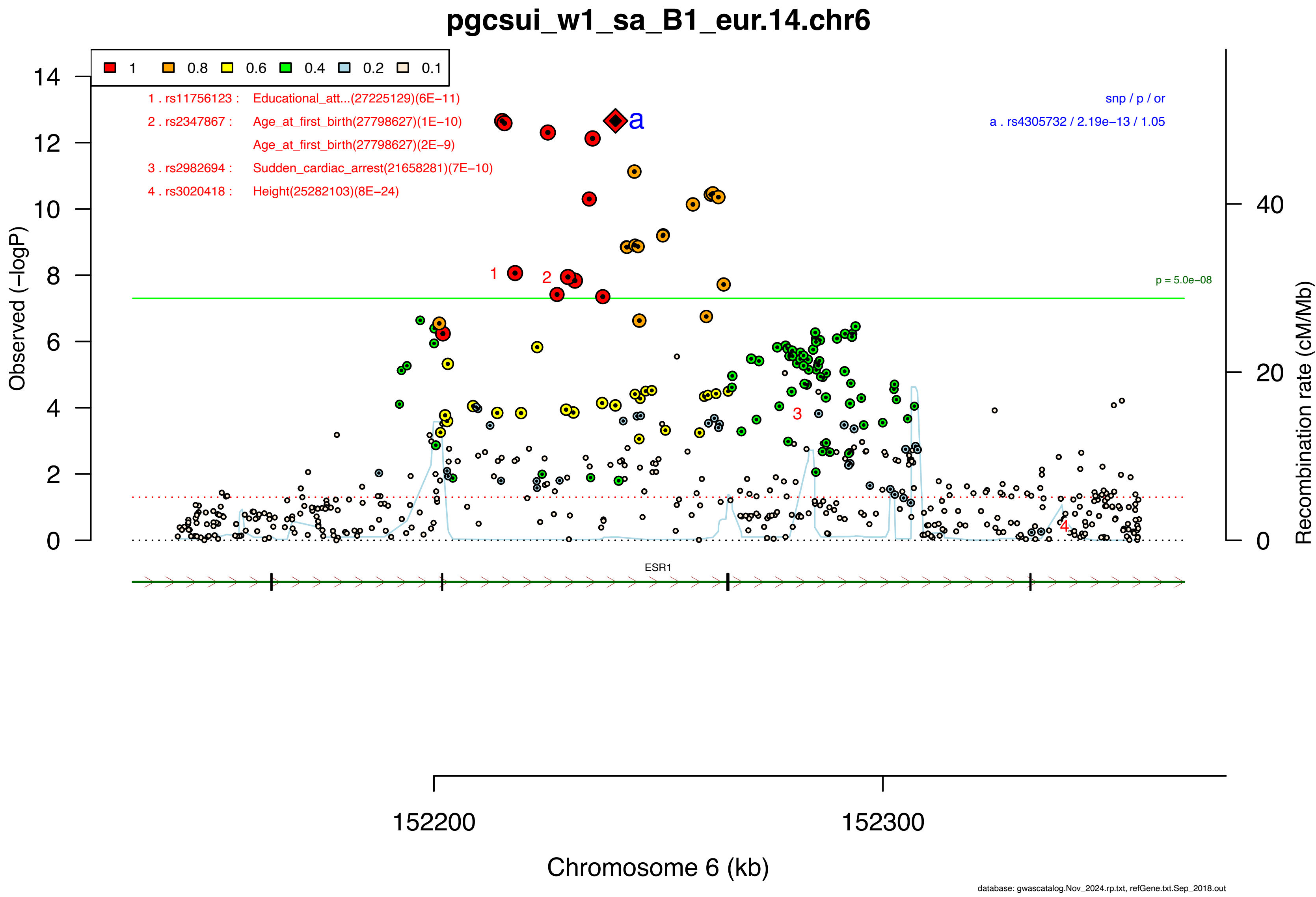

pgcsui_w1_sa_B1_eur.14.chr6
1
0.8
0.6
0.4
0.2
0.1
1 . rs11756123 : Educational_att...(27225129)(6E−11)
2 . rs2347867 : Age_at_first_birth(27798627)(1E−10)
Age_at_first_birth(27798627)(2E−9)
3 . rs2982694 : Sudden_cardiac_arrest(21658281)(7E−10)
4 . rs3020418 : Height(25282103)(8E−24)
snp / p / or
a . rs4305732 / 2.19e−13 / 1.05
p = 5.0e−08
Observed (−logP)
Recombination rate (cM/Mb)
ESR1
152200
152300
Chromosome 6 (kb)
database: gwascatalog.Nov_2024.rp.txt, refGene.txt.Sep_2018.out

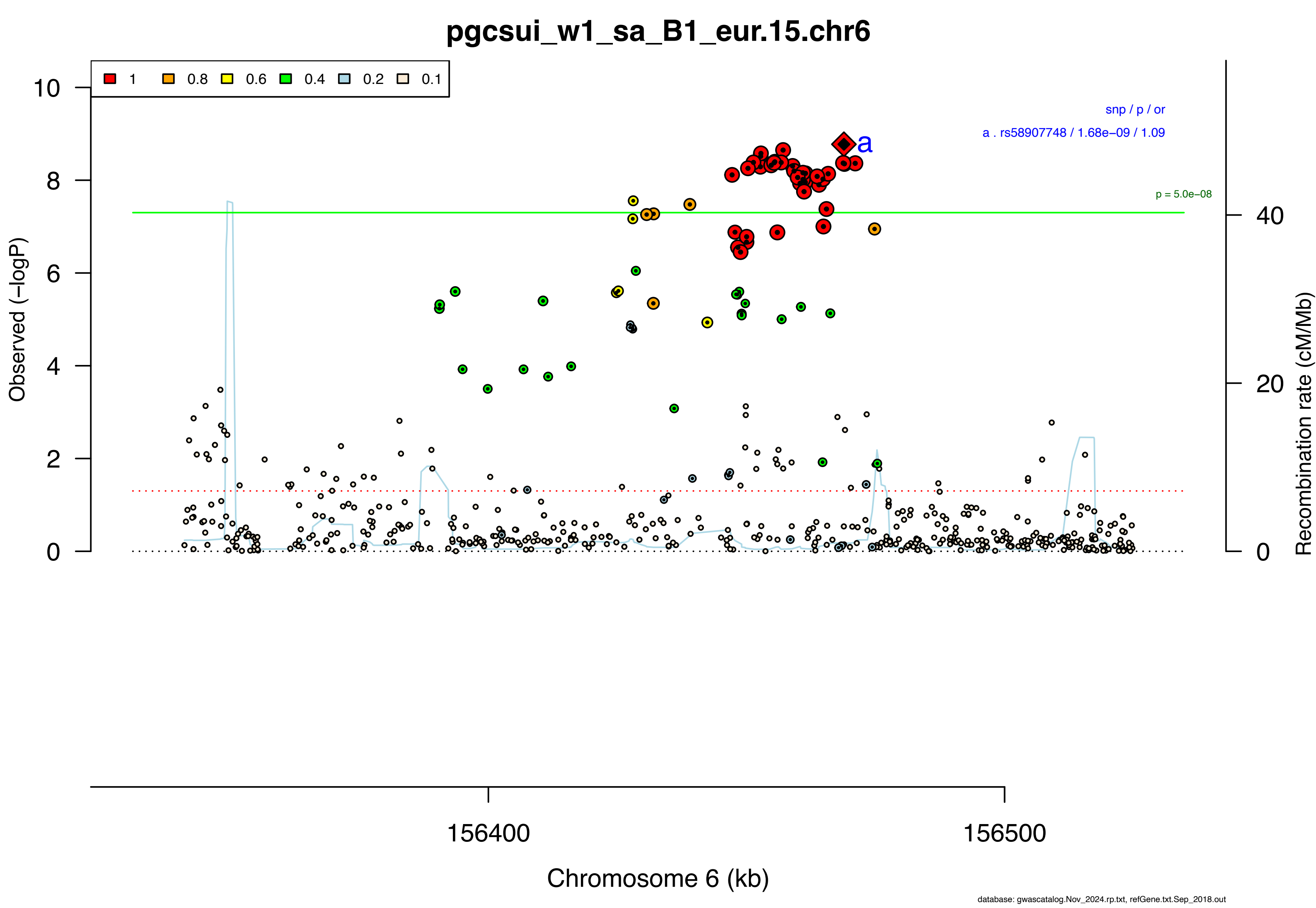

pgcsui_w1_sa_B1_eur.15.chr6
1
0.8
0.6
0.4
0.2
0.1
snp / p / or
a . rs58907748 / 1.68e−09 / 1.09
a
p = 5.0e−08
Observed (−logP)
Recombination rate (cM/Mb)
0
2
4
6
8
10
0
20
40
156400
156500
Chromosome 6 (kb)
database: gwascatalog.Nov_2024.rp.txt, refGene.txt.Sep_2018.out

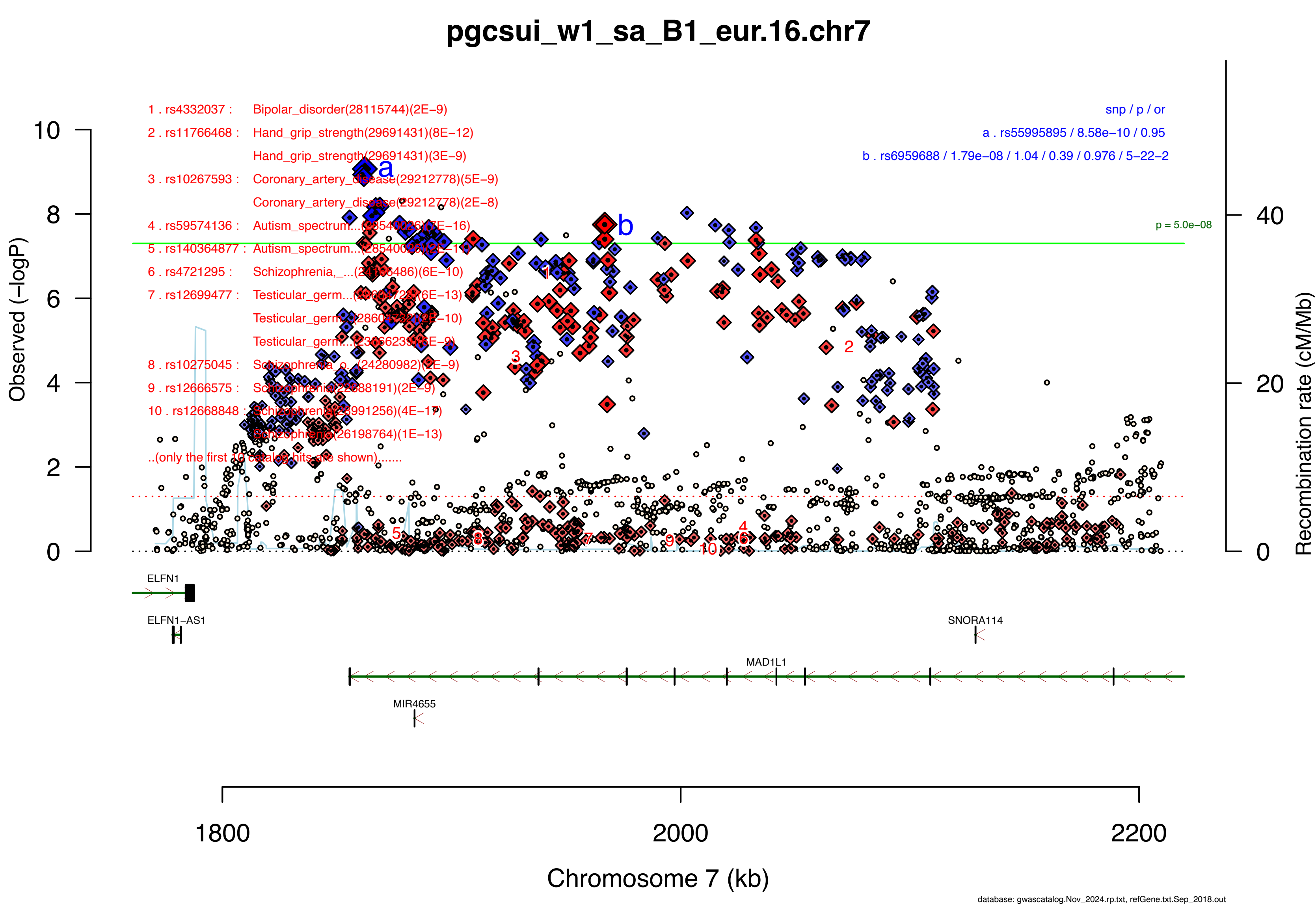

pgcsui_w1_sa_B1_eur.16.chr7
1 . rs4332037 : Bipolar_disorder(28115744)(2E−9)
2 . rs11766468 : Hand_grip_strength(29691431)(8E−12)
Hand_grip_strength(29691431)(3E−9)
3 . rs10267593 : Coronary_artery_disease(29212778)(5E−9)
Coronary_artery_disease(29212778)(2E−8)
4 . rs59574136 : Autism_spectrum...(28540026)(7E−16)
5 . rs140364877 : Autism_spectrum...(28540026)(2E−11)
6 . rs4721295 : Schizophrenia,_...(24166486)(6E−10)
7 . rs12699477 : Testicular_germ...(28604728)(6E−13)
Testicular_germ...(28604728)(2E−10)
Testicular_germ...(23666239)(3E−9)
8 . rs10275045 : Schizophrenia_o...(24280982)(2E−9)
9 . rs12666575 : Schizophrenia(22688191)(2E−9)
10 . rs12668848 : Schizophrenia(28991256)(4E−17)
Schizophrenia(26198764)(1E−13)
..(only the first 10 catalog hits are shown).......
snp / p / or
a . rs55995895 / 8.58e−10 / 0.95
b . rs6959688 / 1.79e−08 / 1.04 / 0.39 / 0.976 / 5−22−2
p = 5.0e−08
Observed (−logP)
Recombination rate (cM/Mb)
ELFN1
ELFN1−AS1
SNORA114
MAD1L1
MIR4655
1800
2000
2200
Chromosome 7 (kb)
database: gwascatalog.Nov_2024.rp.txt, refGene.txt.Sep_2018.out

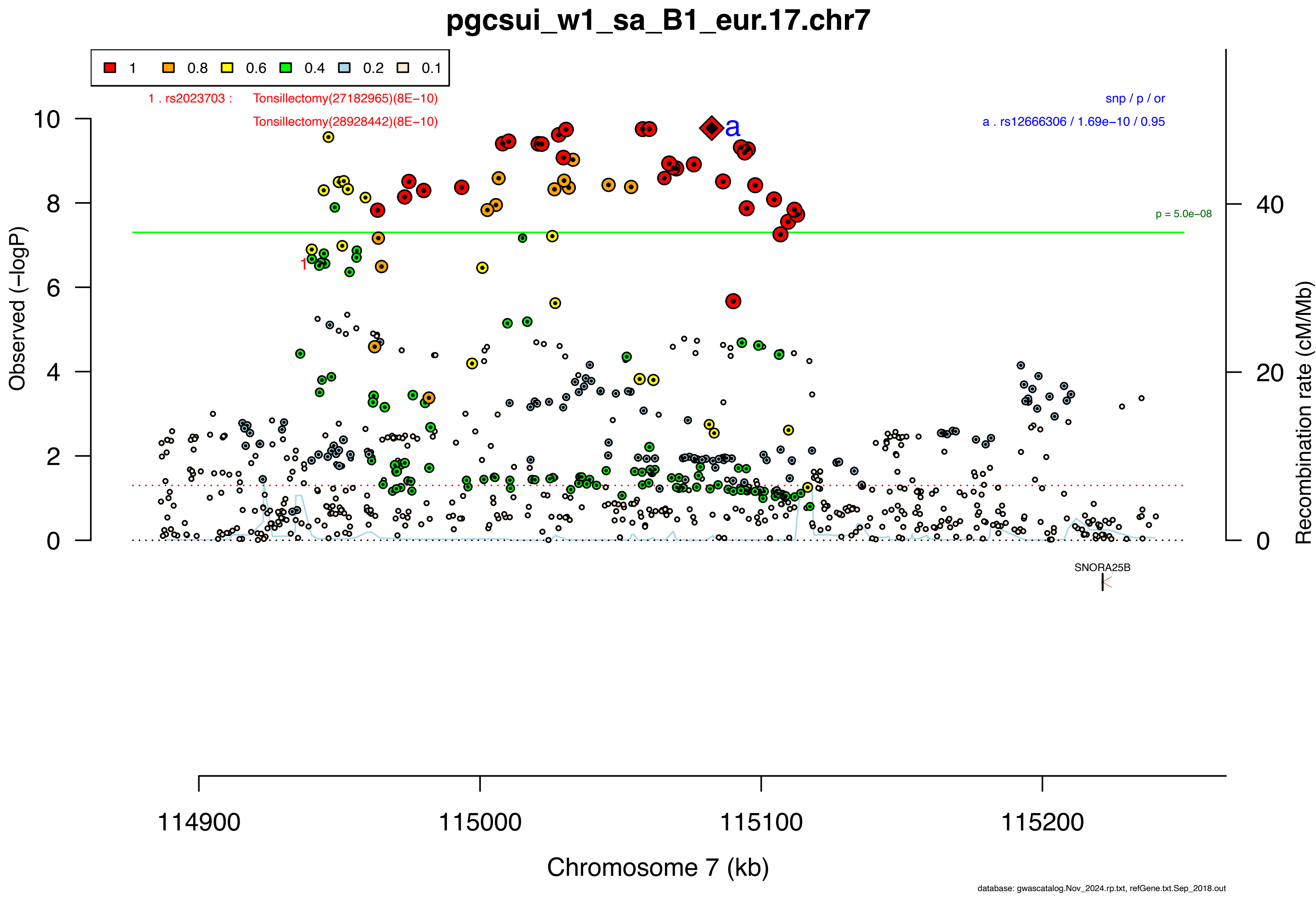

pgcsui_w1_sa_B1_eur.17.chr7
1
0.8
0.6
0.4
0.2
0.1
1 . rs2023703 : Tonsillectomy(27182965)(8E−10)
Tonsillectomy(28928442)(8E−10)
snp / p / or
a . rs12666306 / 1.69e−10 / 0.95
a
1
p = 5.0e−08
Observed (−logP)
Recombination rate (cM/Mb)
0
2
4
6
8
10
0
20
40
SNORA25B
114900
115000
115100
115200
Chromosome 7 (kb)
database: gwascatalog.Nov_2024.rp.txt, refGene.txt.Sep_2018.out

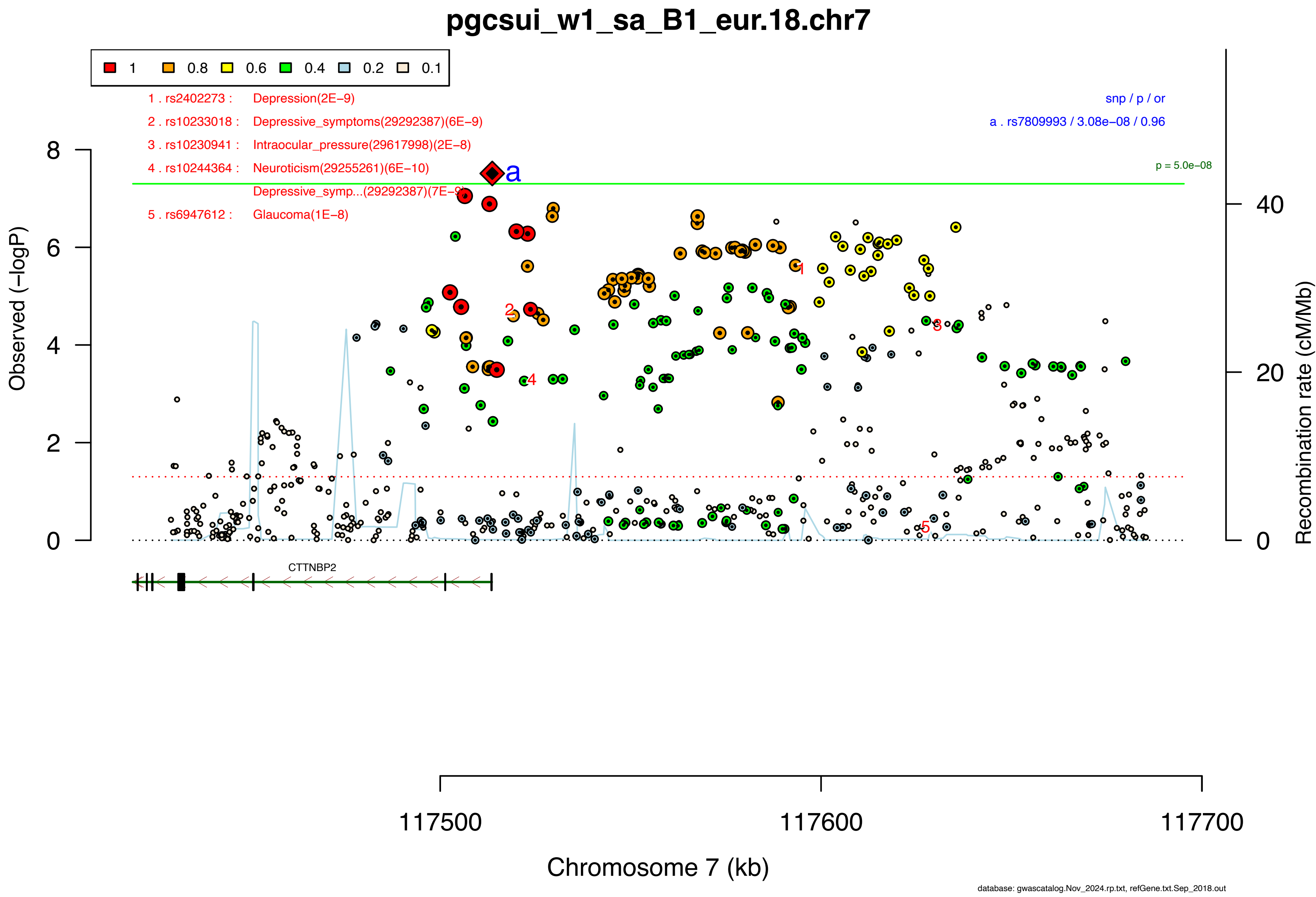
pgcsui_w1_sa_B1_eur.18.chr7
1 0.8 0.6 0.4 0.2 0.1
1 . rs2402273 : Depression(2E−9)
2 . rs10233018 : Depressive_symptoms(29292387)(6E−9)
3 . rs10230941 : Intraocular_pressure(29617998)(2E−8)
4 . rs10244364 : Neuroticism(29255261)(6E−10)
Depressive_symp...(29292387)(7E−9)
5 . rs6947612 : Glaucoma(1E−8)
snp / p / or
a . rs7809993 / 3.08e−08 / 0.96
p = 5.0e−08
Observed (−logP)
Recombination rate (cM/Mb)
CTTNBP2
117500
117600
117700
Chromosome 7 (kb)
database: gwascatalog.Nov_2024.rp.txt, refGene.txt.Sep_2018.out

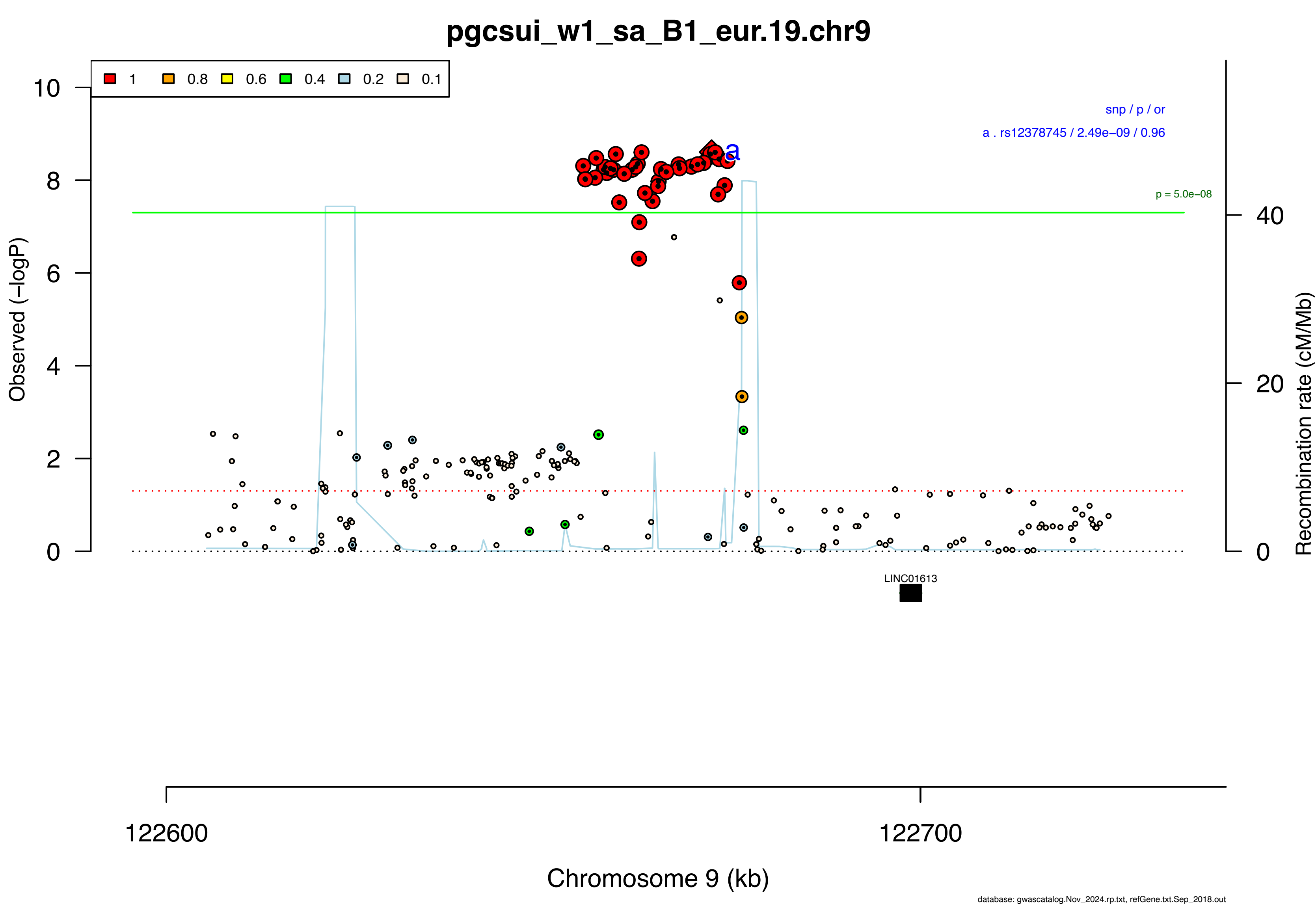
pgcsui_w1_sa_B1_eur.19.chr9
1
0.8
0.6
0.4
0.2
0.1
snp / p / or
a . rs12378745 / 2.49e−09 / 0.96
p = 5.0e−08
a
Observed (−logP)
Recombination rate (cM/Mb)
LINC01613
122600
122700
Chromosome 9 (kb)
database: gwascatalog.Nov_2024.rp.txt, refGene.txt.Sep_2018.out

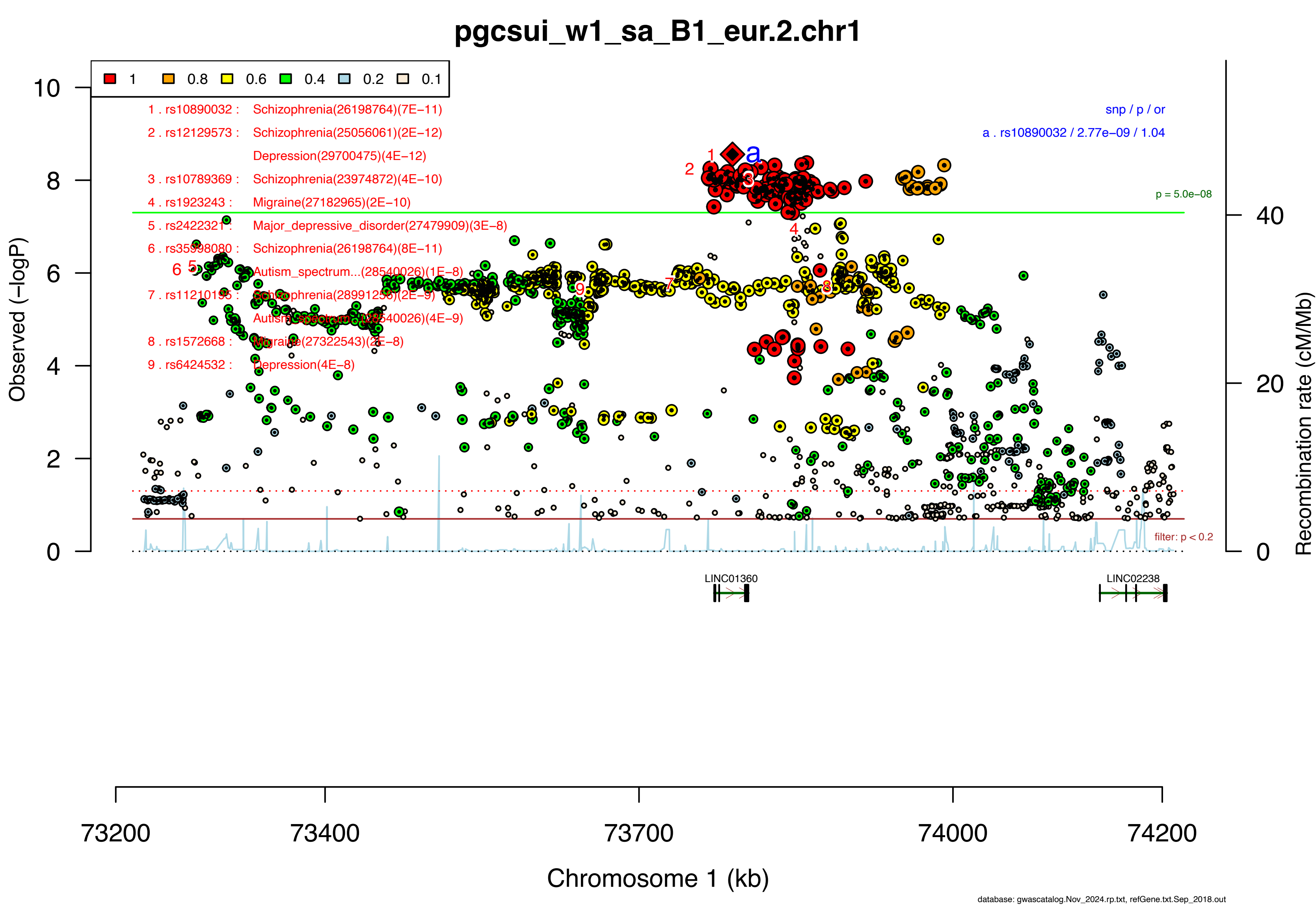
pgcsui_w1_sa_B1_eur.2.chr1
1 0.8 0.6 0.4 0.2 0.1
1 . rs10890032 : Schizophrenia(26198764)(7E−11)
2 . rs12129573 : Schizophrenia(25056061)(2E−12)
Depression(29700475)(4E−12)
3 . rs10789369 : Schizophrenia(23974872)(4E−10)
4 . rs1923243 : Migraine(27182965)(2E−10)
5 . rs2422321 : Major_depressive_disorder(27479909)(3E−8)
6 . rs35998080 : Schizophrenia(26198764)(8E−11)
Autism_spectrum...(28540026)(1E−8)
7 . rs11210195 : Schizophrenia(28991256)(2E−9)
Autism_spectrum...(28540026)(4E−9)
8 . rs1572668 : Migraine(27322543)(2E−8)
9 . rs6424532 : Depression(4E−8)
snp / p / or
a . rs10890032 / 2.77e−09 / 1.04
p = 5.0e−08
filter: p < 0.2
Observed (−logP)
Recombination rate (cM/Mb)
LINC01360
LINC02238
73200
73400
73700
74000
74200
Chromosome 1 (kb)
database: gwascatalog.Nov_2024.rp.txt, refGene.txt.Sep_2018.out

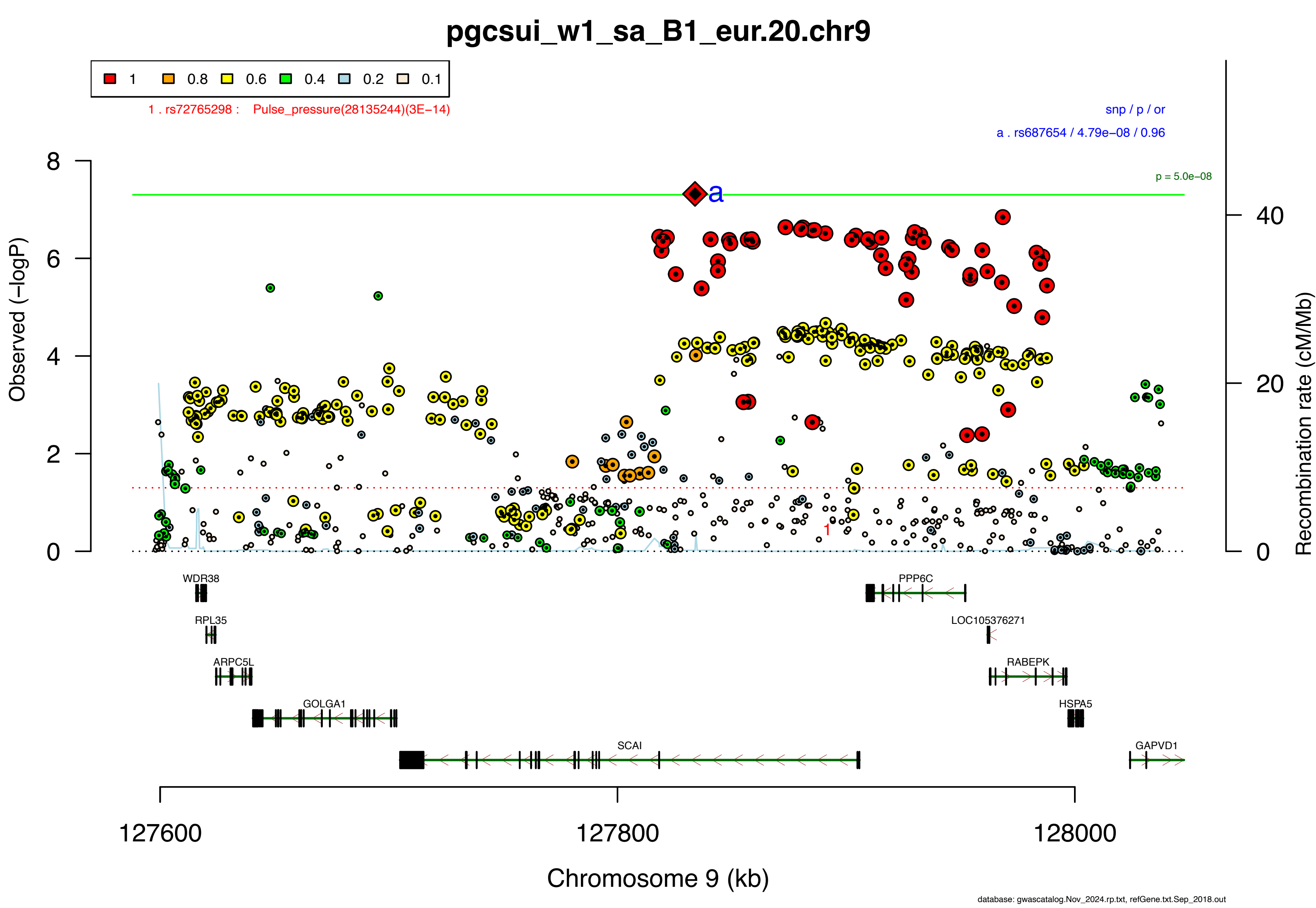

pgcsui_w1_sa_B1_eur.20.chr9
1
0.8
0.6
0.4
0.2
0.1
1 . rs72765298 : Pulse_pressure(28135244)(3E−14)
snp / p / or
a . rs687654 / 4.79e−08 / 0.96
p = 5.0e−08
a
Observed (−logP)
Recombination rate (cM/Mb)
WDR38
RPL35
ARPC5L
GOLGA1
SCAI
PPP6C
LOC105376271
RABEPK
HSPA5
GAPVD1
127600
127800
128000
Chromosome 9 (kb)
database: gwascatalog.Nov_2024.rp.txt, refGene.txt.Sep_2018.out

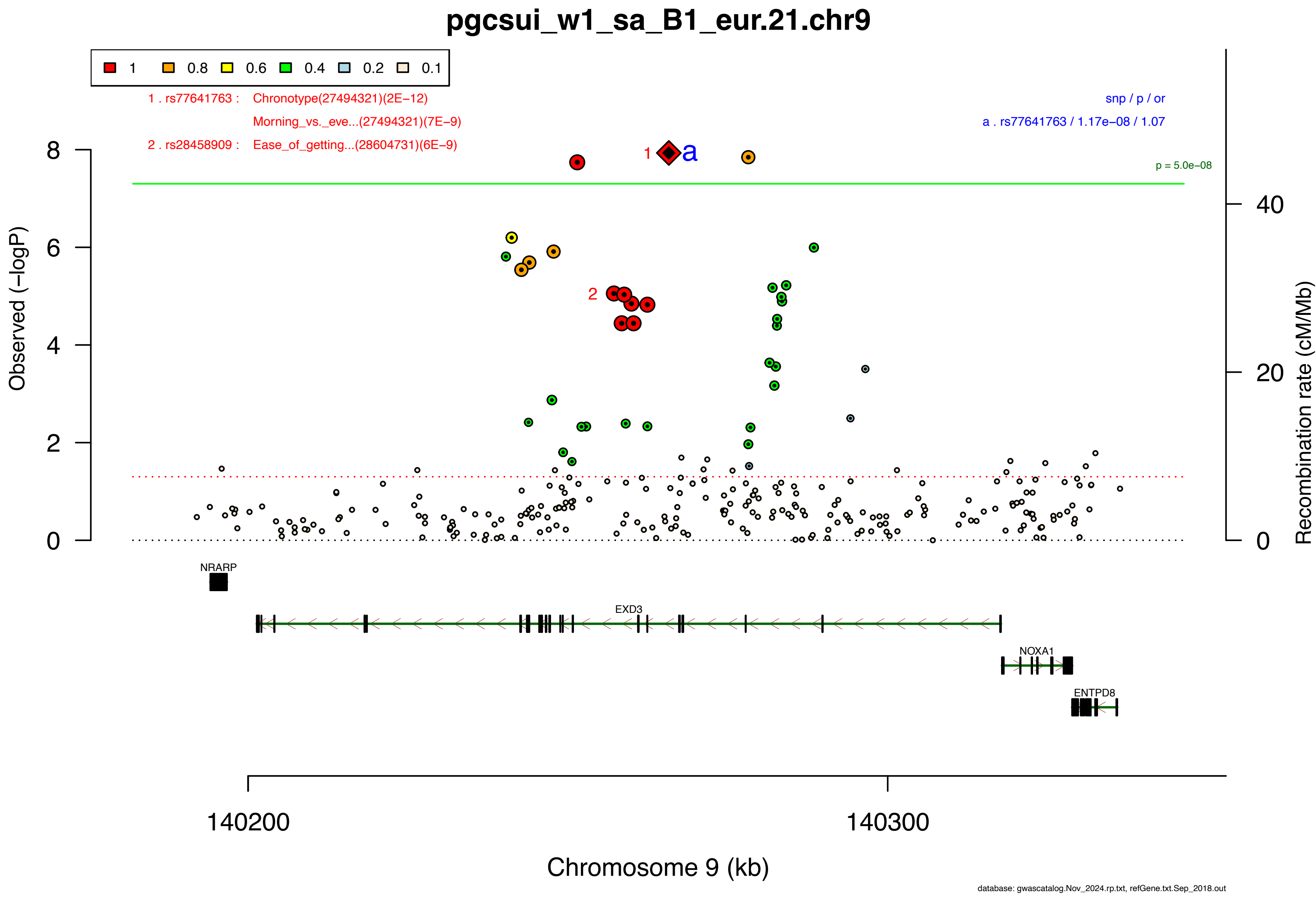

pgcsui_w1_sa_B1_eur.21.chr9
1
0.8
0.6
0.4
0.2
0.1
1 . rs77641763 : Chronotype(27494321)(2E−12)
Morning_vs._eve...(27494321)(7E−9)
2 . rs28458909 : Ease_of_getting...(28604731)(6E−9)
snp / p / or
a . rs77641763 / 1.17e−08 / 1.07
p = 5.0e−08
Observed (−logP)
Recombination rate (cM/Mb)
NRARP
EXD3
NOXA1
ENTPD8
140200
140300
Chromosome 9 (kb)
database: gwascatalog.Nov_2024.rp.txt, refGene.txt.Sep_2018.out

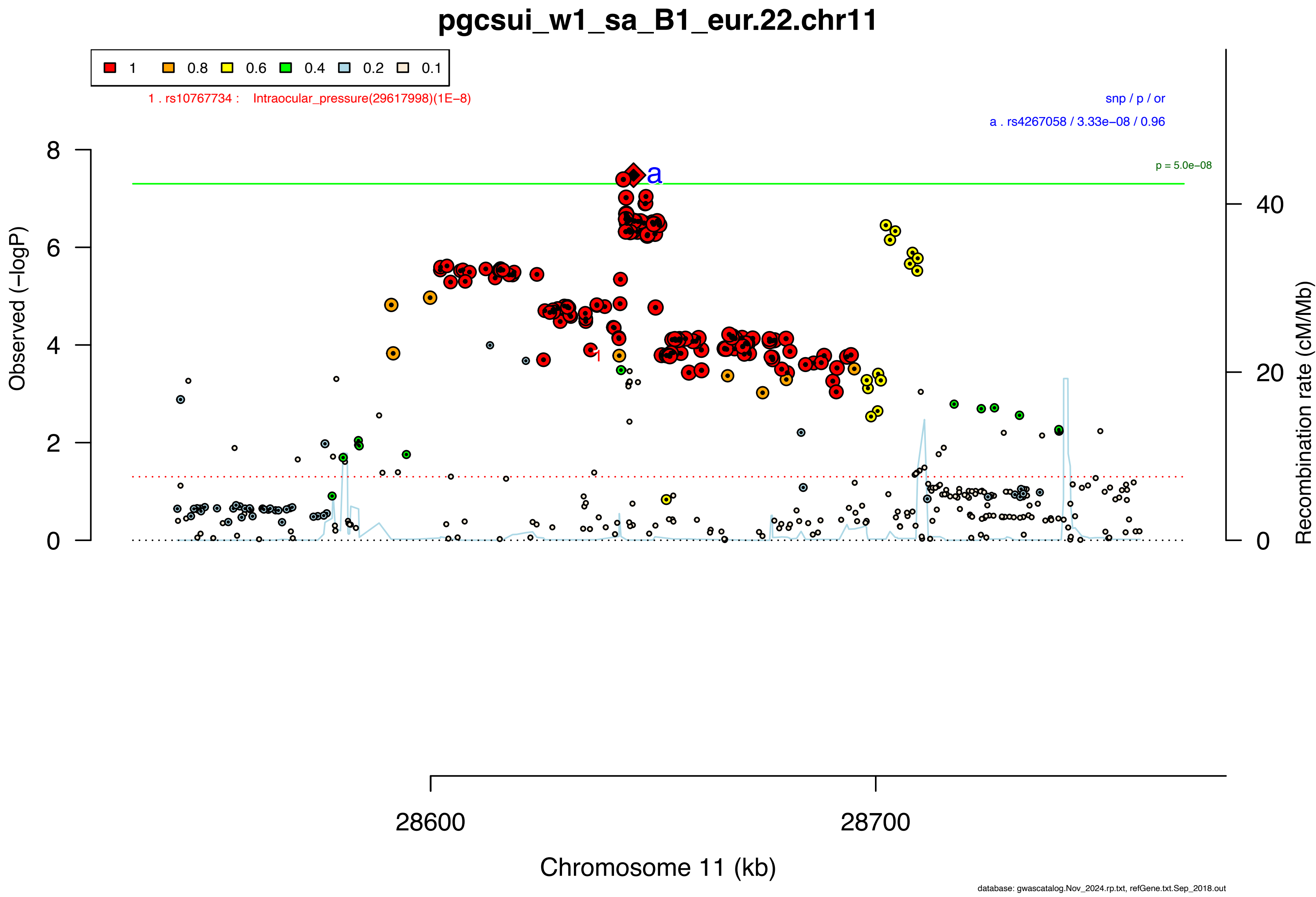
pgcsui_w1_sa_B1_eur.22.chr11
1 0.8 0.6 0.4 0.2 0.1
1 . rs10767734 : Intraocular_pressure(29617998)(1E−8)
snp / p / or
a . rs4267058 / 3.33e−08 / 0.96
p = 5.0e−08
Observed (−logP)
Recombination rate (cM/Mb)
28600
28700
Chromosome 11 (kb)
database: gwascatalog.Nov_2024.rp.txt, refGene.txt.Sep_2018.out

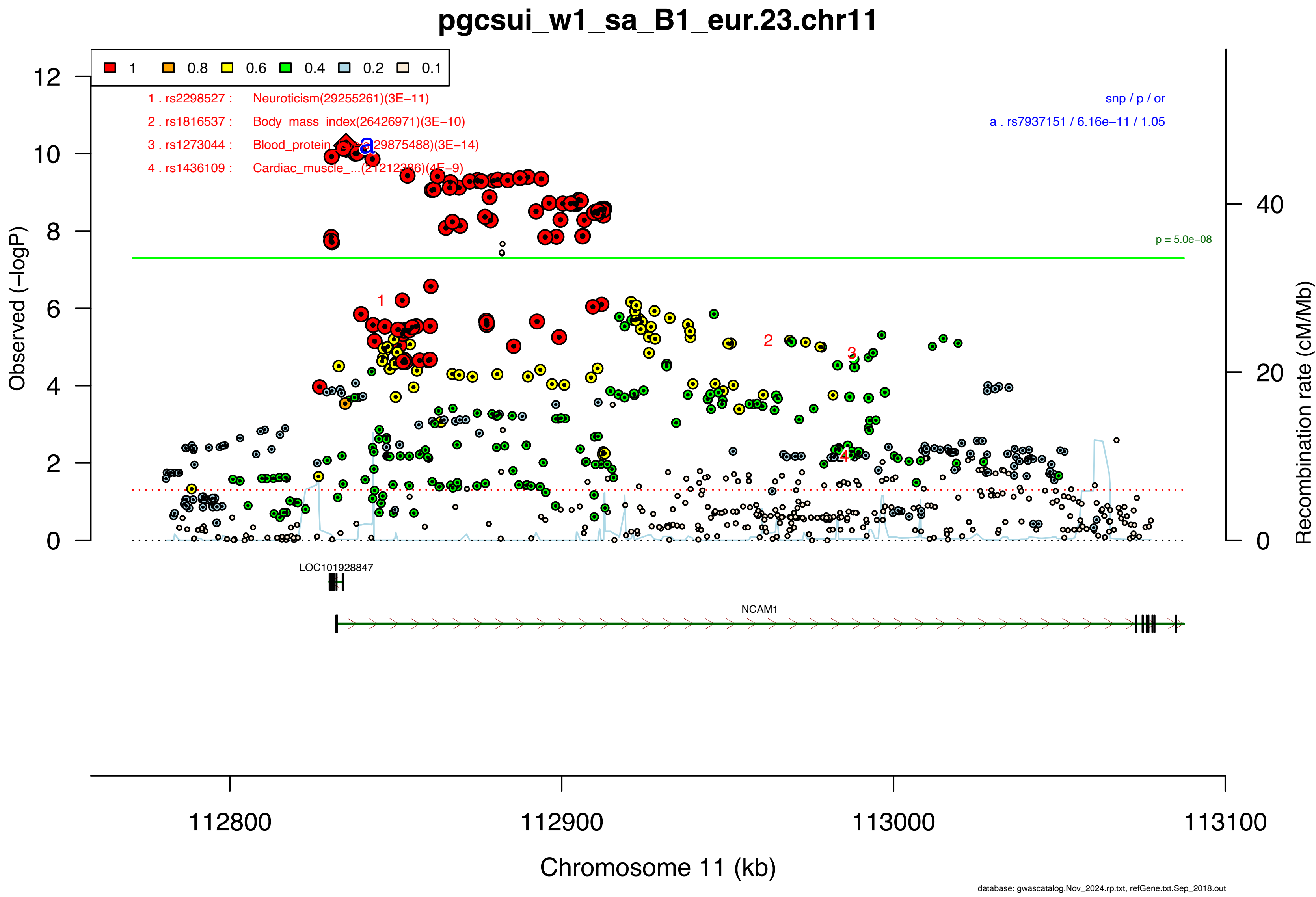
pgcsui_w1_sa_B1_eur.23.chr11
1
0.8
0.6
0.4
0.2
0.1
1 . rs2298527 : Neuroticism(29255261)(3E−11)
2 . rs1816537 : Body_mass_index(26426971)(3E−10)
3 . rs1273044 : Blood_protein_...(29875488)(3E−14)
4 . rs1436109 : Cardiac_muscle_...(21212386)(4E−9)
snp / p / or
a . rs7937151 / 6.16e−11 / 1.05
p = 5.0e−08
Observed (−logP)
Recombination rate (cM/Mb)
LOC101928847
NCAM1
112800
112900
113000
113100
Chromosome 11 (kb)
database: gwascatalog.Nov_2024.rp.txt, refGene.txt.Sep_2018.out

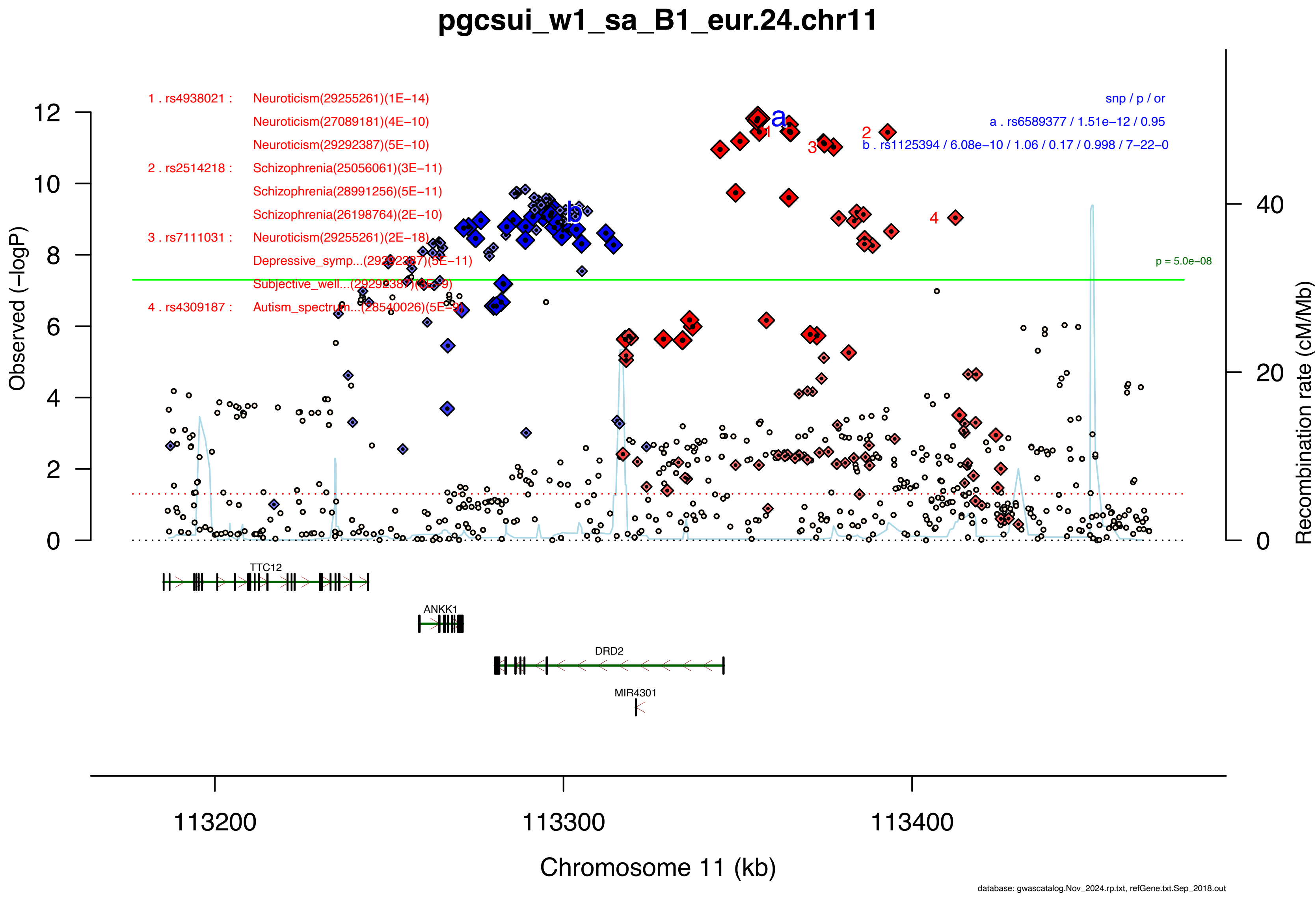

pgcsui_w1_sa_B1_eur.24.chr11
1 . rs4938021 : Neuroticism(29255261)(1E−14)
Neuroticism(27089181)(4E−10)
Neuroticism(29292387)(5E−10)
2 . rs2514218 : Schizophrenia(25056061)(3E−11)
Schizophrenia(28991256)(5E−11)
Schizophrenia(26198764)(2E−10)
3 . rs7111031 : Neuroticism(29255261)(2E−18)
Depressive_symp...(29292387)(5E−11)
Subjective_well...(29292387)(4E−9)
4 . rs4309187 : Autism_spectrum...(28540026)(5E−9)
snp / p / or
a . rs6589377 / 1.51e−12 / 0.95
b . rs1125394 / 6.08e−10 / 1.06 / 0.17 / 0.998 / 7−22−0
p = 5.0e−08
Observed (−logP)
Recombination rate (cM/Mb)
TTC12
ANKK1
DRD2
MIR4301
113200
113300
113400
Chromosome 11 (kb)
database: gwascatalog.Nov_2024.rp.txt, refGene.txt.Sep_2018.out

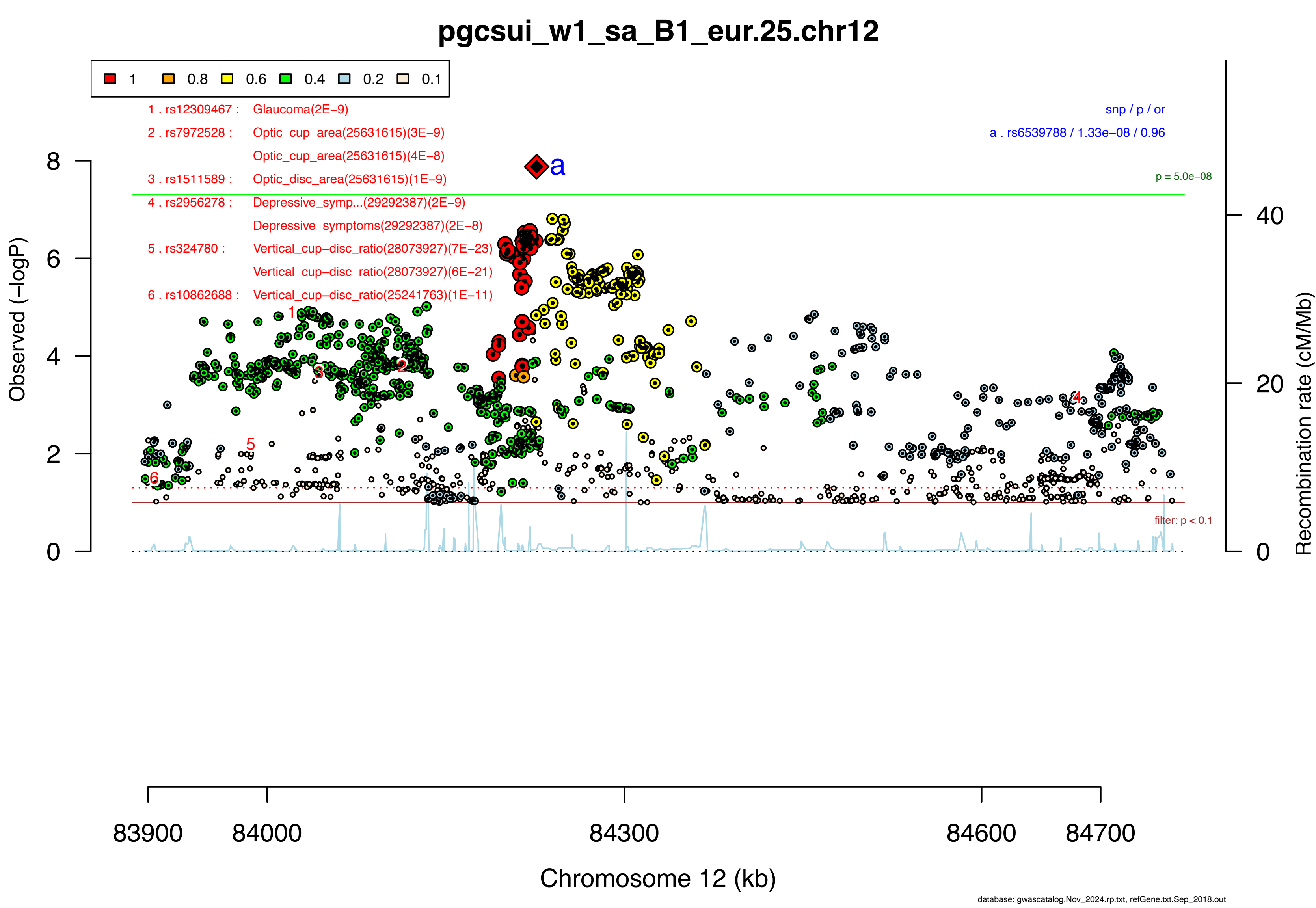
pgcsui_w1_sa_B1_eur.25.chr12
1
0.8
0.6
0.4
0.2
0.1
1 . rs12309467 : Glaucoma(2E−9)
2 . rs7972528 : Optic_cup_area(25631615)(3E−9)
Optic_cup_area(25631615)(4E−8)
3 . rs1511589 : Optic_disc_area(25631615)(1E−9)
4 . rs2956278 : Depressive_symp...(29292387)(2E−9)
Depressive_symptoms(29292387)(2E−8)
5 . rs324780 : Vertical_cup−disc_ratio(28073927)(7E−23)
Vertical_cup−disc_ratio(28073927)(6E−21)
6 . rs10862688 : Vertical_cup−disc_ratio(25241763)(1E−11)
snp / p / or
a . rs6539788 / 1.33e−08 / 0.96
p = 5.0e−08
filter: p < 0.1
Observed (−logP)
Recombination rate (cM/Mb)
Chromosome 12 (kb)
83900
84000
84300
84600
84700
database: gwascatalog.Nov_2024.rp.txt, refGene.txt.Sep_2018.out

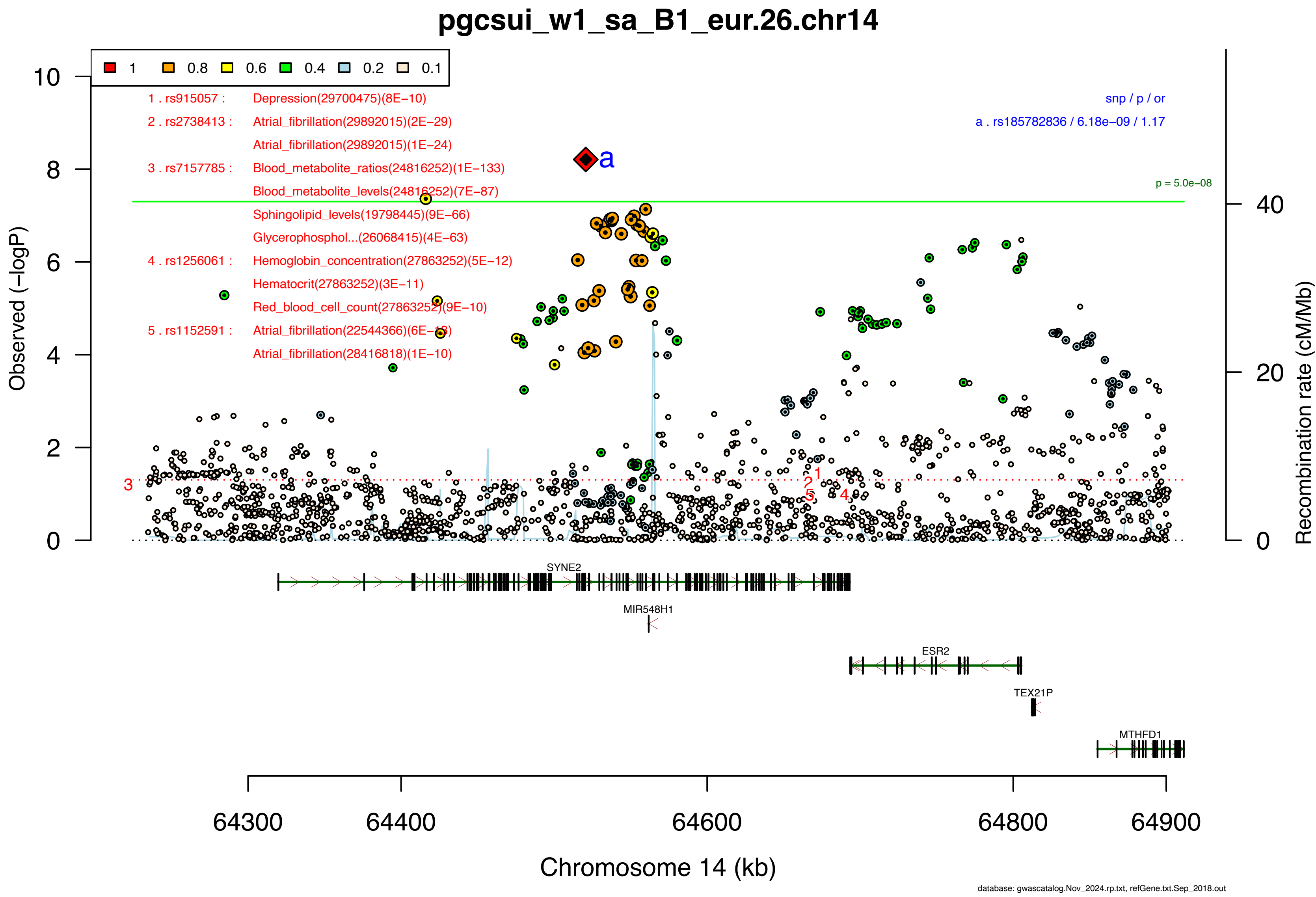

pgcsui_w1_sa_B1_eur.26.chr14
1 0.8 0.6 0.4 0.2 0.1
1 . rs915057 : Depression(29700475)(8E−10)
2 . rs2738413 : Atrial_fibrillation(29892015)(2E−29)
Atrial_fibrillation(29892015)(1E−24)
3 . rs7157785 : Blood_metabolite_ratios(24816252)(1E−133)
Blood_metabolite_levels(24816252)(7E−87)
Sphingolipid_levels(19798445)(9E−66)
Glycerophosphol...(26068415)(4E−63)
4 . rs1256061 : Hemoglobin_concentration(27863252)(5E−12)
Hematocrit(27863252)(3E−11)
Red_blood_cell_count(27863252)(9E−10)
5 . rs1152591 : Atrial_fibrillation(22544366)(6E−18)
Atrial_fibrillation(28416818)(1E−10)
snp / p / or
a . rs185782836 / 6.18e−09 / 1.17
p = 5.0e−08
Observed (−logP)
Recombination rate (cM/Mb)
SYNE2
MIR548H1
ESR2
TEX21P
MTHFD1
64300 64400 64600 64800 64900
Chromosome 14 (kb)
database: gwascatalog.Nov_2024.rp.txt, refGene.txt.Sep_2018.out

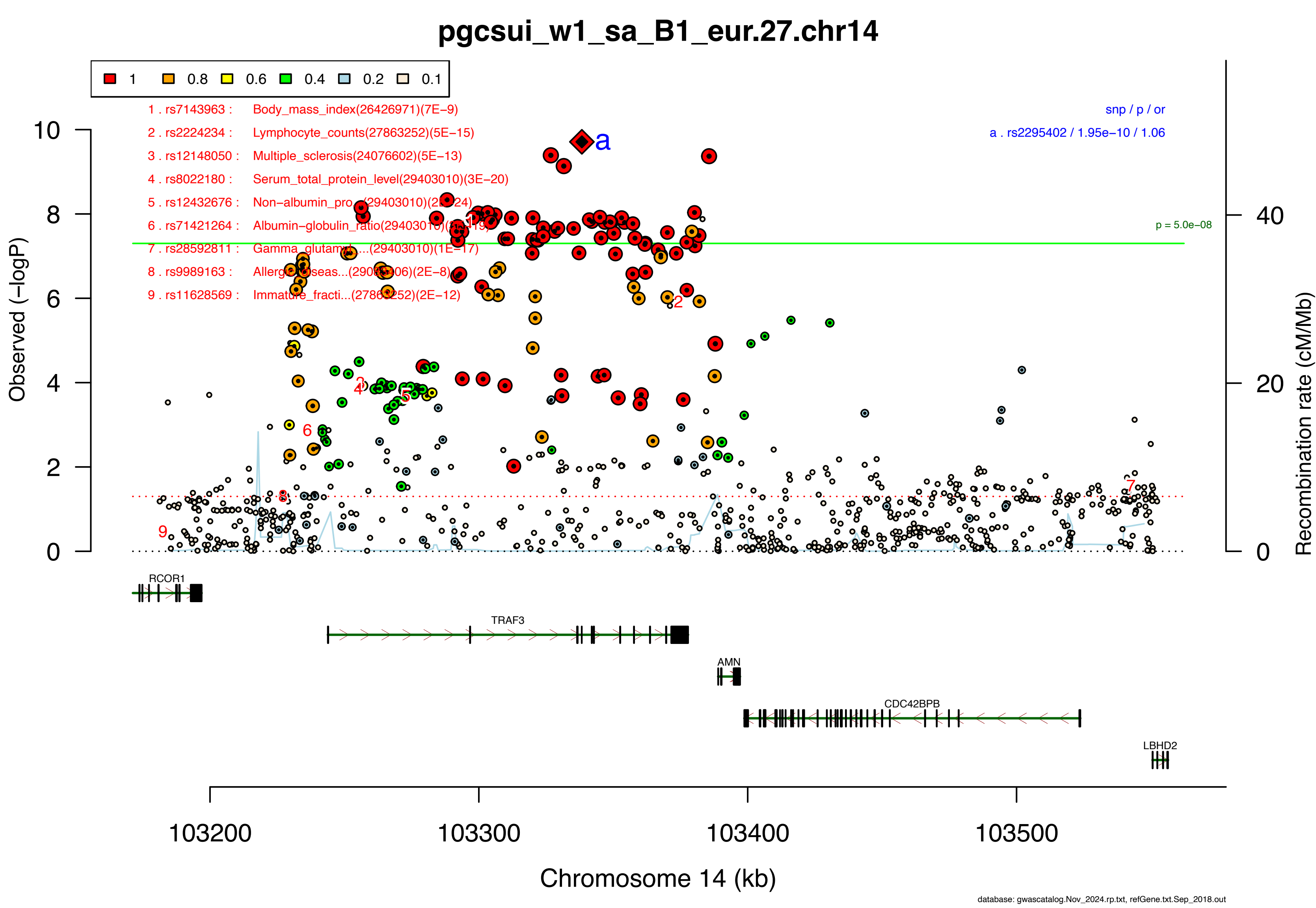

pgcsui_w1_sa_B1_eur.27.chr14
1 0.8 0.6 0.4 0.2 0.1
1 . rs7143963 : Body_mass_index(26426971)(7E−9)
2 . rs2224234 : Lymphocyte_counts(27863252)(5E−15)
3 . rs12148050 : Multiple_sclerosis(24076602)(5E−13)
4 . rs8022180 : Serum_total_protein_level(29403010)(3E−20)
5 . rs12432676 : Non−albumin_pro...(29403010)(2E−24)
6 . rs71421264 : Albumin−globulin_ratio(29403010)(3E−19)
7 . rs28592811 : Gamma_glutamyl...(29403010)(1E−17)
8 . rs9989163 : Allergic_diseas...(29083406)(2E−8)
9 . rs11628569 : Immature_fracti...(27863252)(2E−12)
snp / p / or
a . rs2295402 / 1.95e−10 / 1.06
p = 5.0e−08
Observed (−logP)
Recombination rate (cM/Mb)
RCOR1
TRAF3
AMN
CDC42BPB
LBHD2
103200
103300
103400
103500
Chromosome 14 (kb)
database: gwascatalog.Nov_2024.rp.txt, refGene.txt.Sep_2018.out

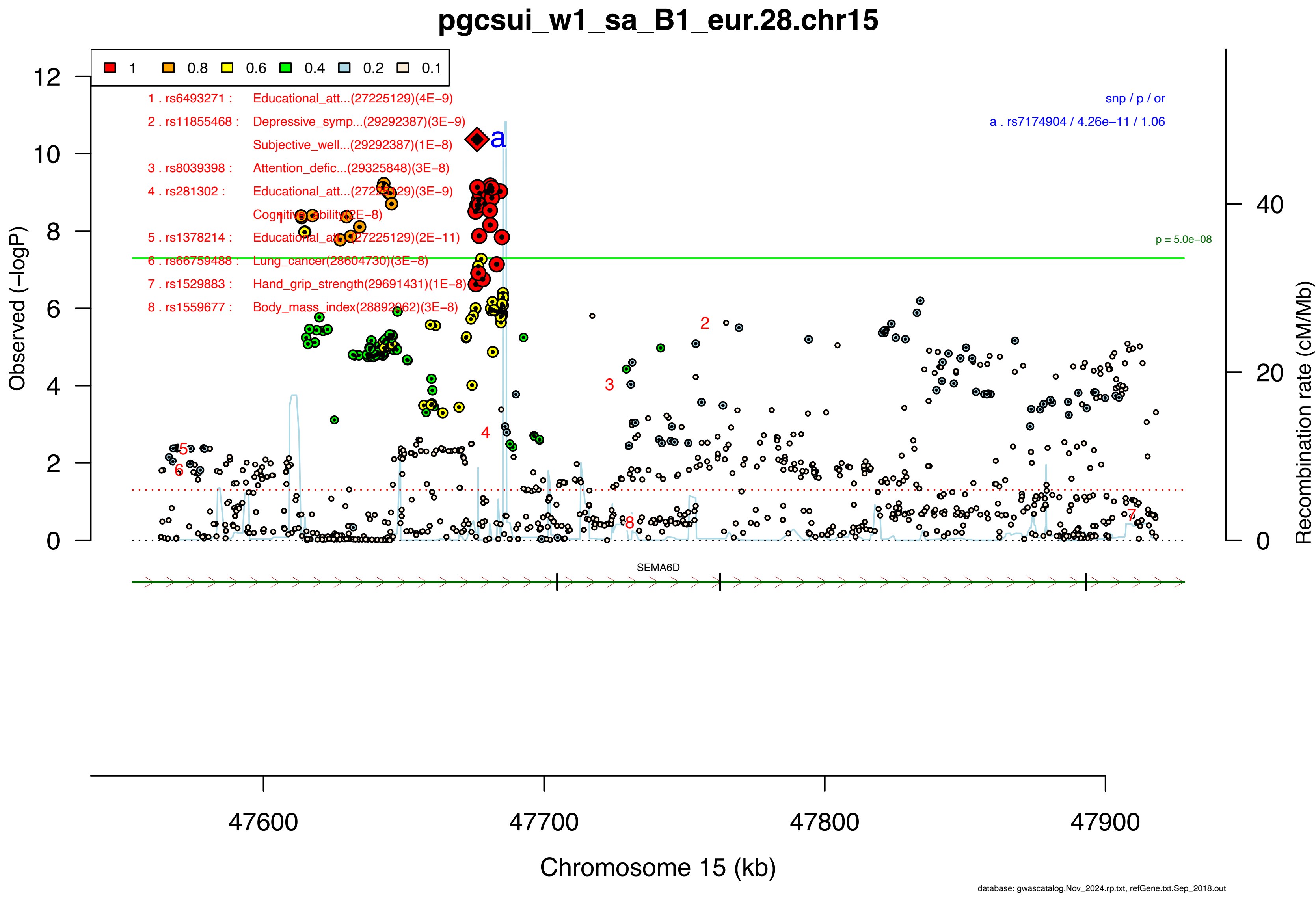

pgcsui_w1_sa_B1_eur.28.chr15
1
0.8
0.6
0.4
0.2
0.1
1 . rs6493271 : Educational_att...(27225129)(4E−9)
2 . rs11855468 : Depressive_symp...(29292387)(3E−9)
Subjective_well...(29292387)(1E−8)
3 . rs8039398 : Attention_defic...(29325848)(3E−8)
4 . rs281302 : Educational_att...(27225129)(3E−9)
Cognitive_ability(2E−8)
5 . rs1378214 : Educational_att...(27225129)(2E−11)
6 . rs66759488 : Lung_cancer(28604730)(3E−8)
7 . rs1529883 : Hand_grip_strength(29691431)(1E−8)
8 . rs1559677 : Body_mass_index(28892062)(3E−8)
snp / p / or
a . rs7174904 / 4.26e−11 / 1.06
p = 5.0e−08
Observed (−logP)
Recombination rate (cM/Mb)
SEMA6D
47600
47700
47800
47900
Chromosome 15 (kb)
database: gwascatalog.Nov_2024.rp.txt, refGene.txt.Sep_2018.out

#### pgcsui_w1_sa_B1_eur.29.chr15

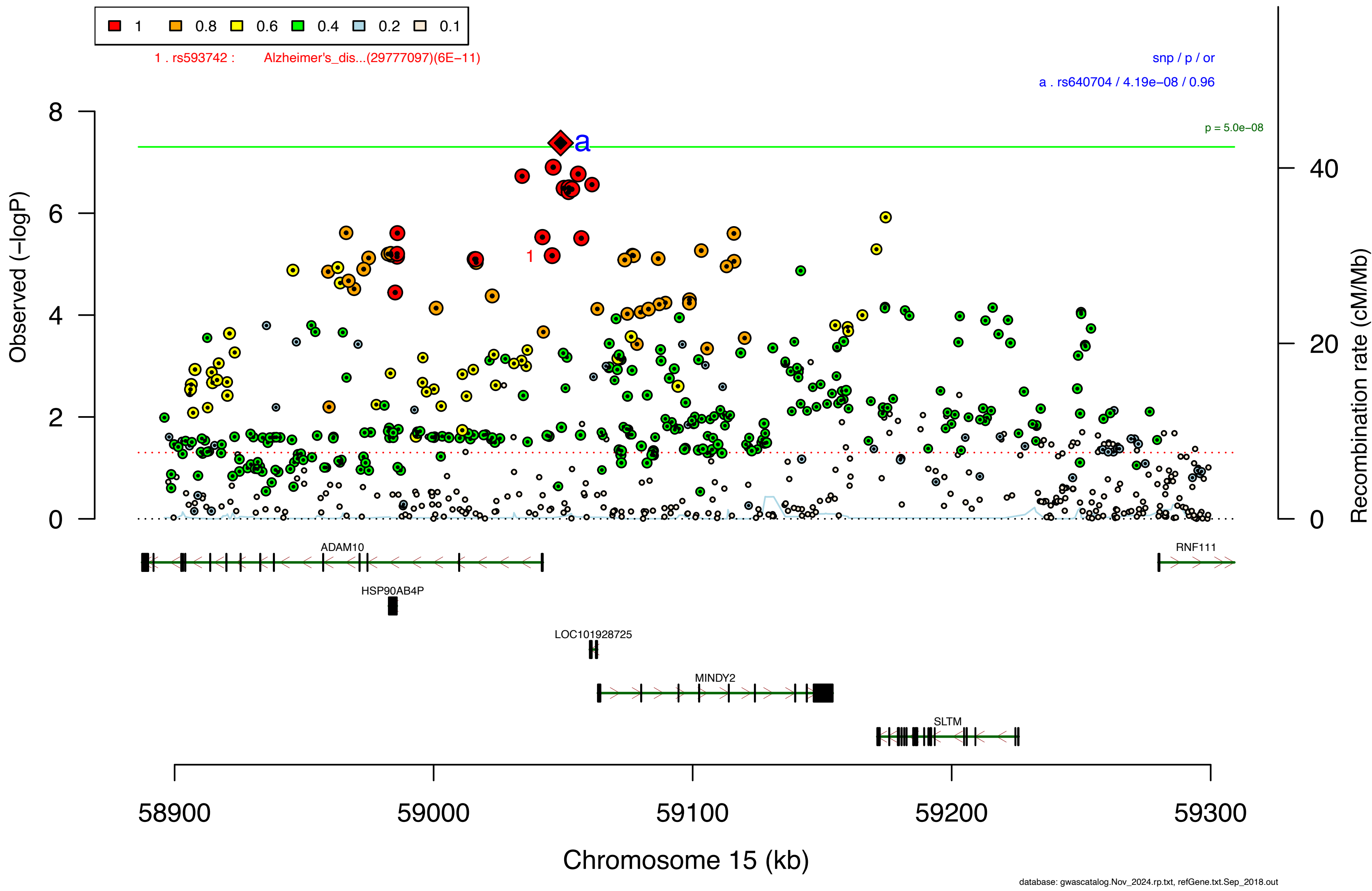

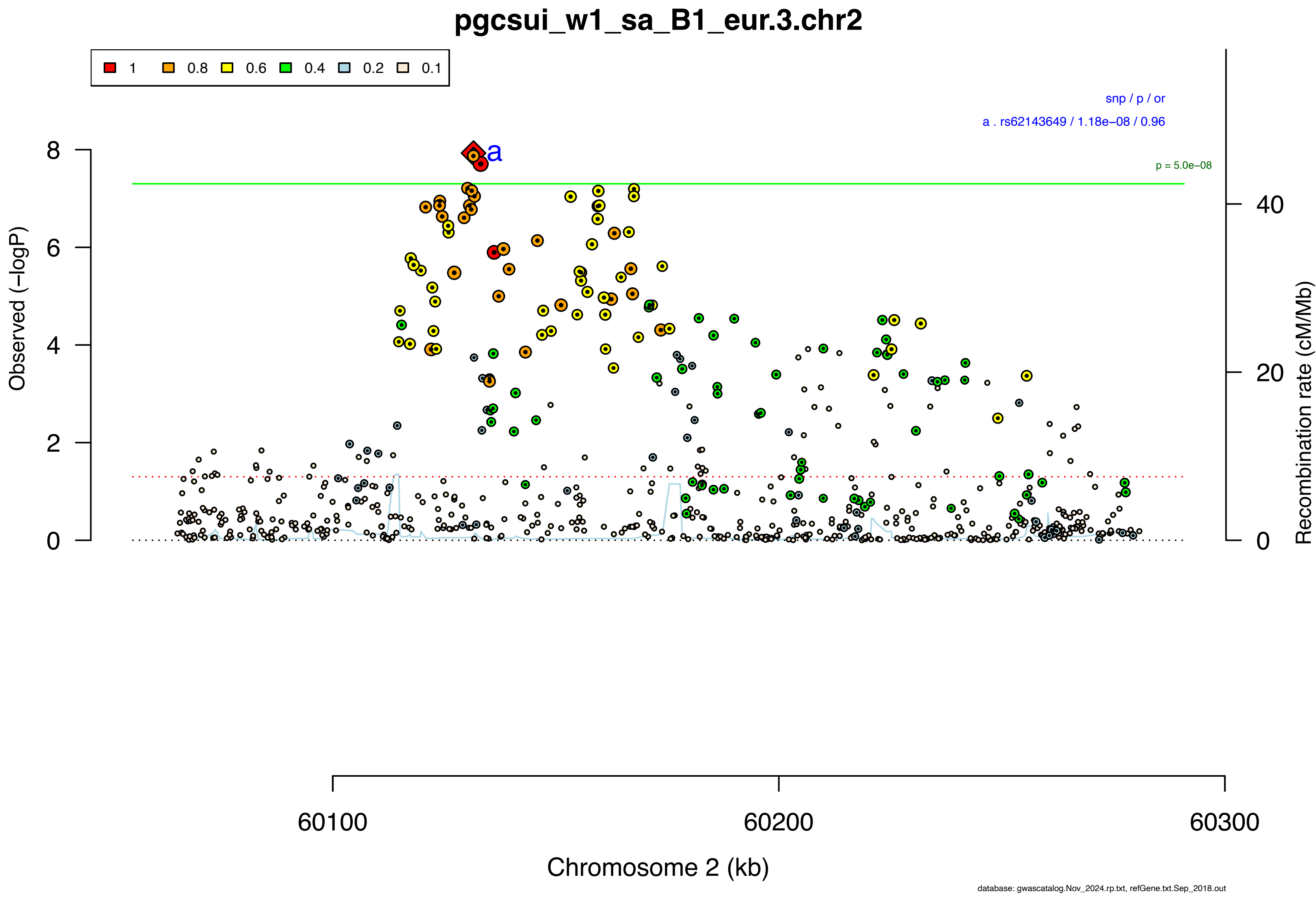
pgcsui_w1_sa_B1_eur.3.chr2
1
0.8
0.6
0.4
0.2
0.1
snp / p / or
a . rs62143649 / 1.18e−08 / 0.96
p = 5.0e−08
a
Observed (−logP)
Recombination rate (cM/Mb)
Chromosome 2 (kb)
60100
60200
60300
database: gwascatalog.Nov_2024.rp.txt, refGene.txt.Sep_2018.out

### pgcsui_w1_sa_B1_eur.30.chr15

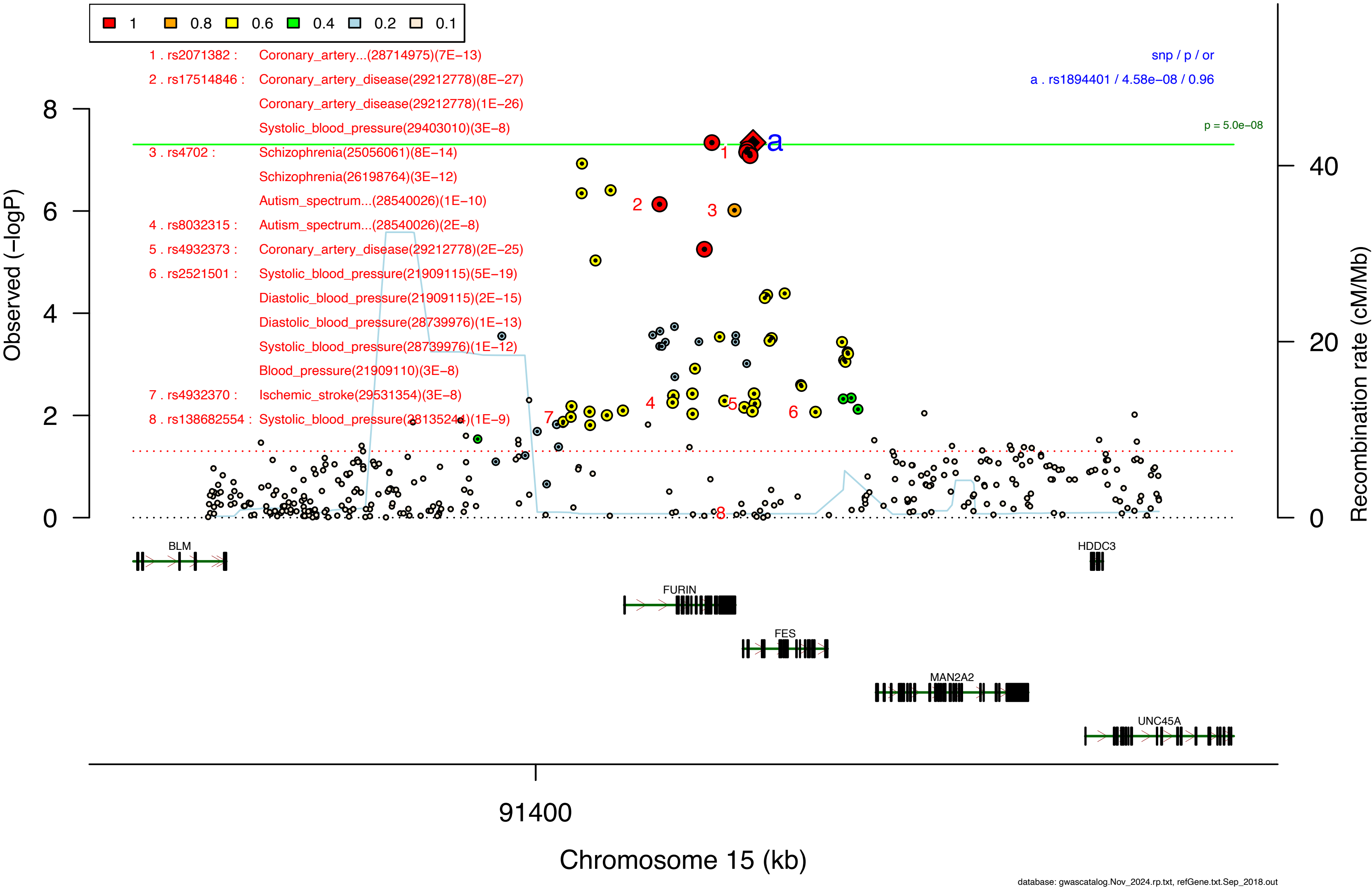

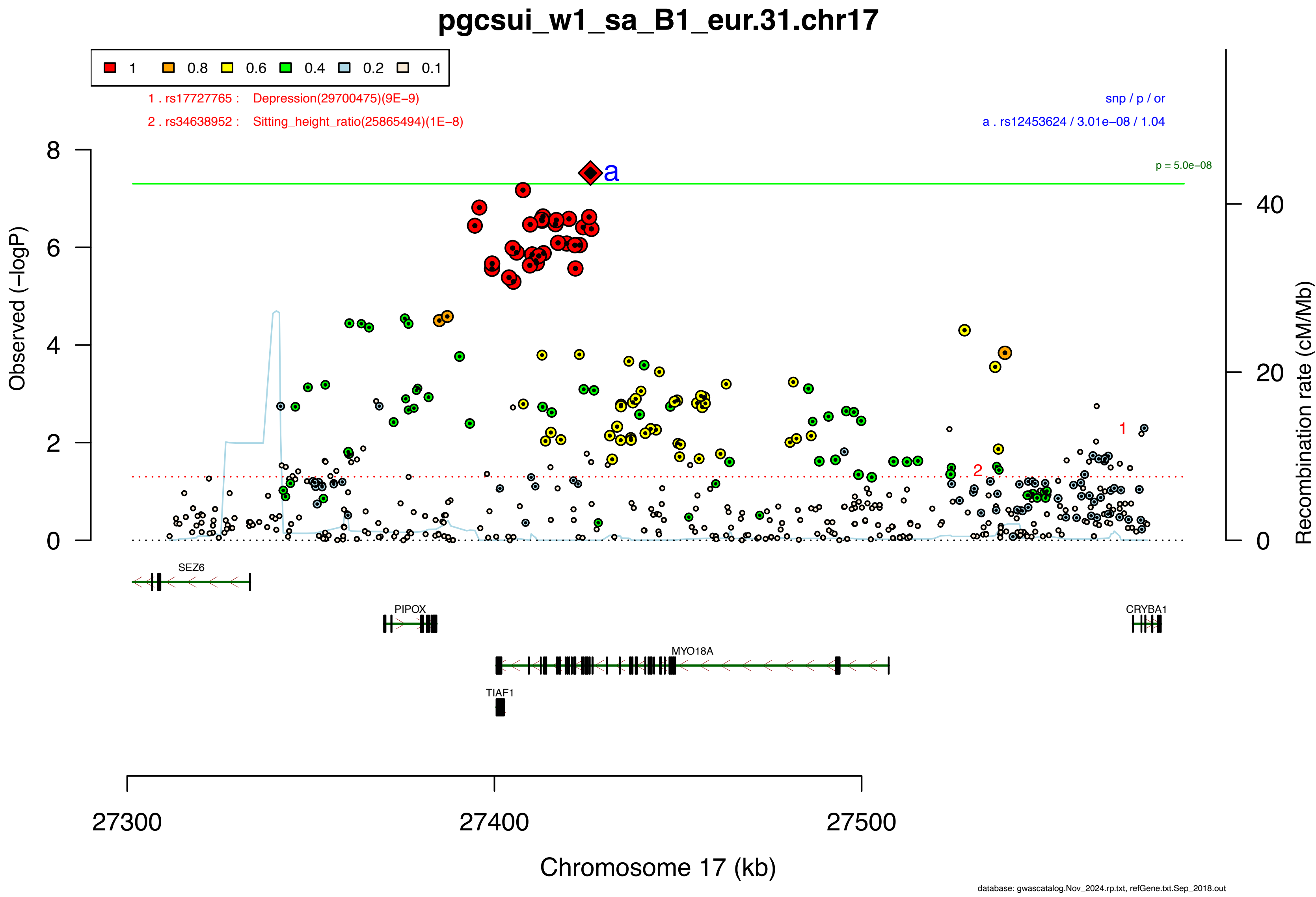

pgcsui_w1_sa_B1_eur.31.chr17
1 0.8 0.6 0.4 0.2 0.1
1 . rs17727765 : Depression(29700475)(9E−9)
2 . rs34638952 : Sitting_height_ratio(25865494)(1E−8)
snp / p / or
a . rs12453624 / 3.01e−08 / 1.04
p = 5.0e−08
a
1
2
Observed (−logP)
Recombination rate (cM/Mb)
0
2
4
6
8
0
20
40
SEZ6
PIPOX
MYO18A
TIAF1
CRYBA1
27300
27400
27500
Chromosome 17 (kb)
database: gwascatalog.Nov_2024.rp.txt, refGene.txt.Sep_2018.out

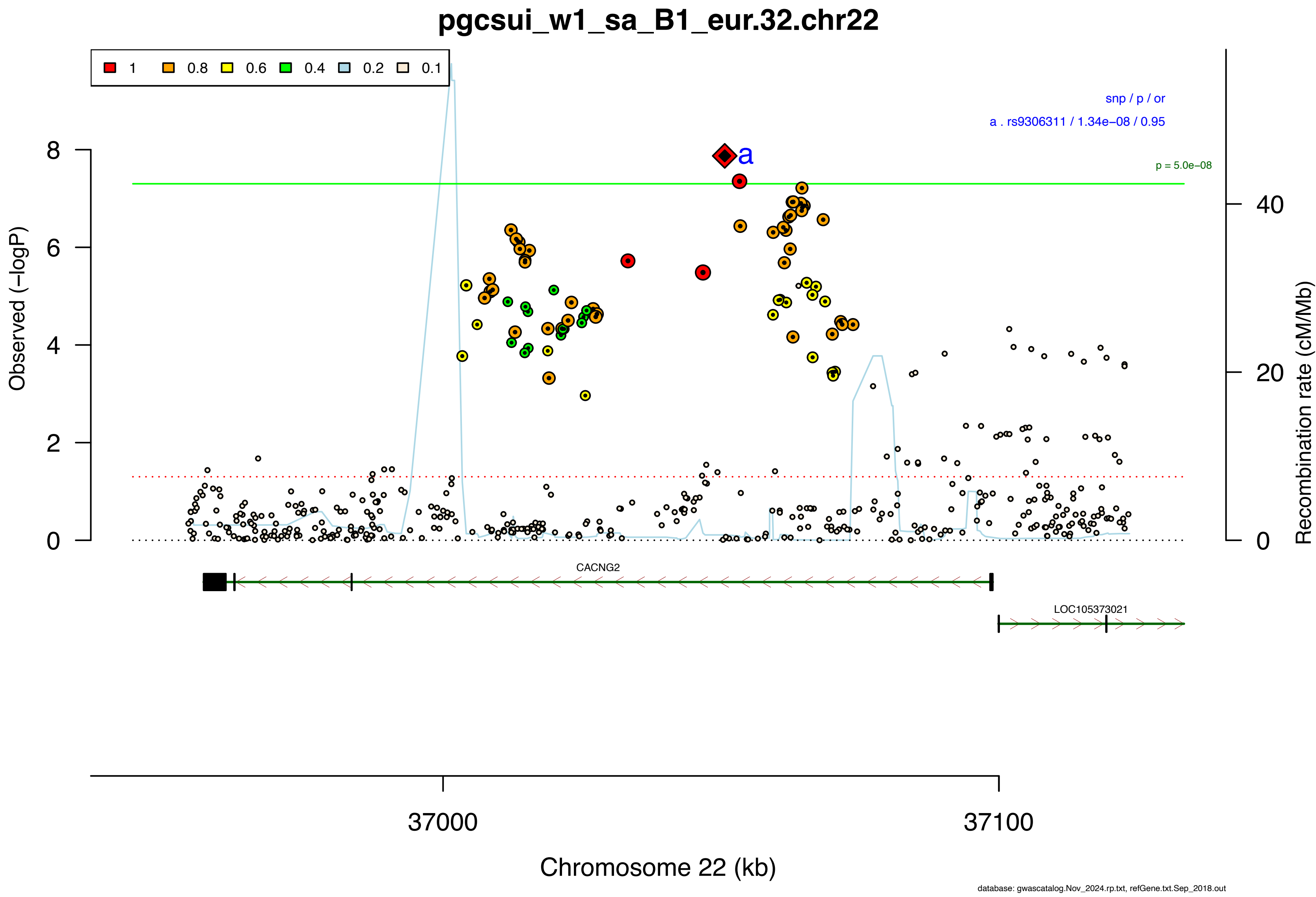
pgcsui_w1_sa_B1_eur.32.chr22
1
0.8
0.6
0.4
0.2
0.1
snp / p / or
a . rs9306311 / 1.34e−08 / 0.95
a
p = 5.0e−08
Observed (−logP)
Recombination rate (cM/Mb)
0
2
4
6
8
0
20
40
CACNG2
LOC105373021
37000
37100
Chromosome 22 (kb)
database: gwascatalog.Nov_2024.rp.txt, refGene.txt.Sep_2018.out

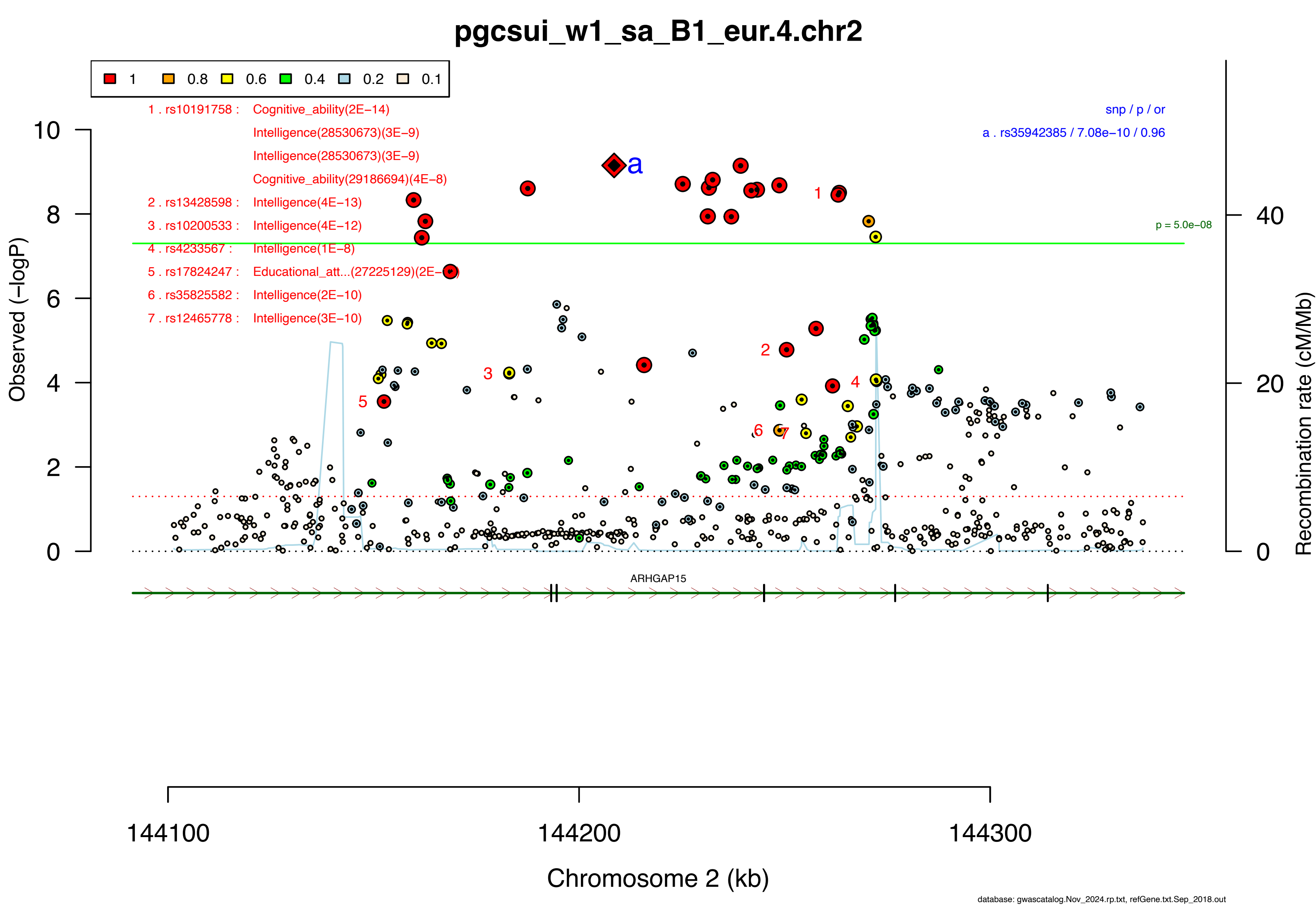

pgcsui_w1_sa_B1_eur.4.chr2
1
0.8
0.6
0.4
0.2
0.1
1 . rs10191758 : Cognitive_ability(2E−14)
Intelligence(28530673)(3E−9)
Intelligence(28530673)(3E−9)
Cognitive_ability(29186694)(4E−8)
2 . rs13428598 : Intelligence(4E−13)
3 . rs10200533 : Intelligence(4E−12)
4 . rs4233567 : Intelligence(1E−8)
5 . rs17824247 : Educational_att...(27225129)(2E−
6 . rs35825582 : Intelligence(2E−10)
7 . rs12465778 : Intelligence(3E−10)
snp / p / or
a . rs35942385 / 7.08e−10 / 0.96
p = 5.0e−08
Observed (−logP)
Recombination rate (cM/Mb)
ARHGAP15
144100
144200
144300
Chromosome 2 (kb)
database: gwascatalog.Nov_2024.rp.txt, refGene.txt.Sep_2018.out

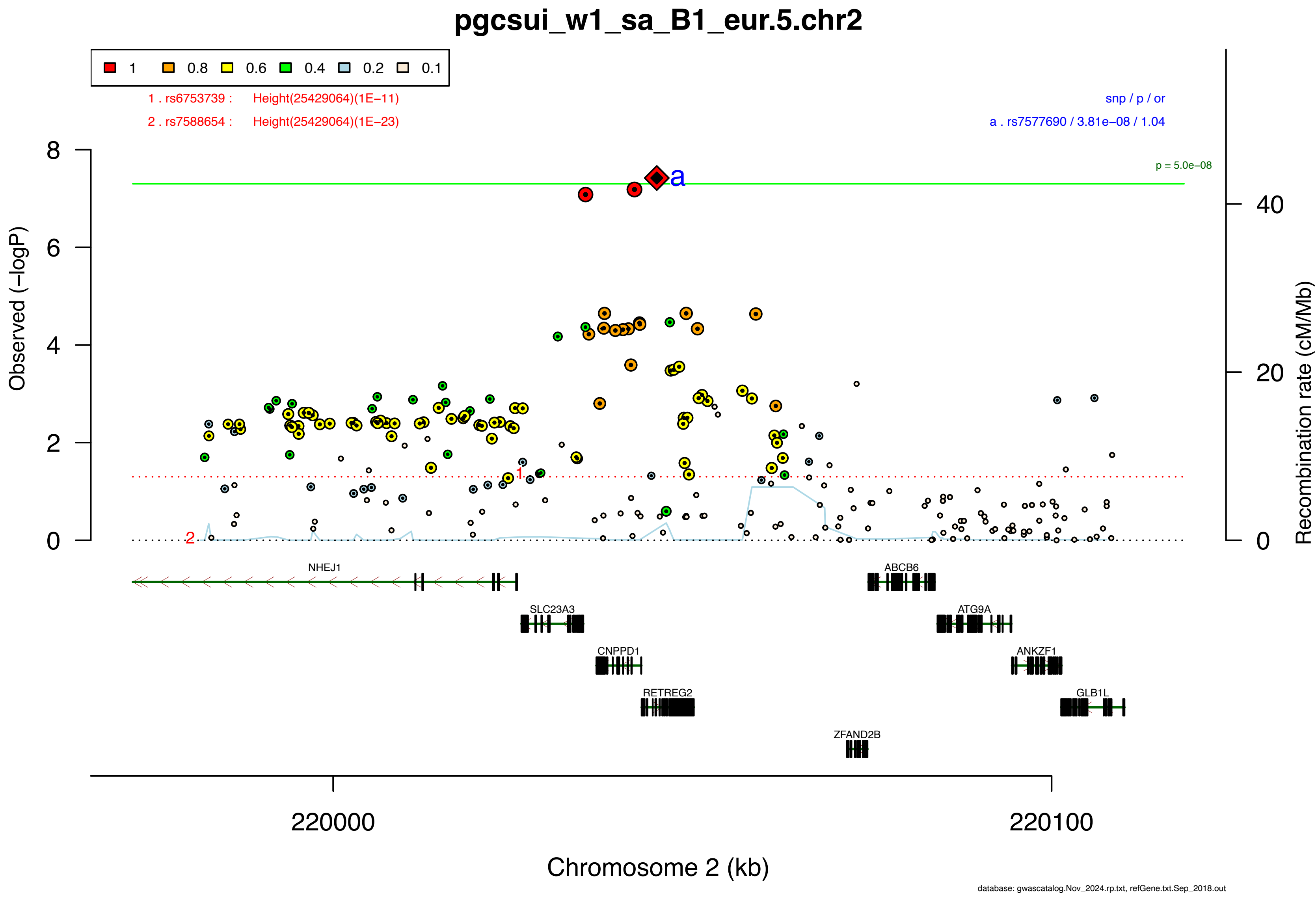

pgcsui_w1_sa_B1_eur.5.chr2
1
0.8
0.6
0.4
0.2
0.1
1 . rs6753739 : Height(25429064)(1E−11)
2 . rs7588654 : Height(25429064)(1E−23)
snp / p / or
a . rs7577690 / 3.81e−08 / 1.04
p = 5.0e−08
Observed (−logP)
Recombination rate (cM/Mb)
NHEJ1
SLC23A3
CNPPD1
RETREG2
ZFAND2B
ABCB6
ATG9A
ANKZF1
GLB1L
220000
220100
Chromosome 2 (kb)
database: gwascatalog.Nov_2024.rp.txt, refGene.txt.Sep_2018.out

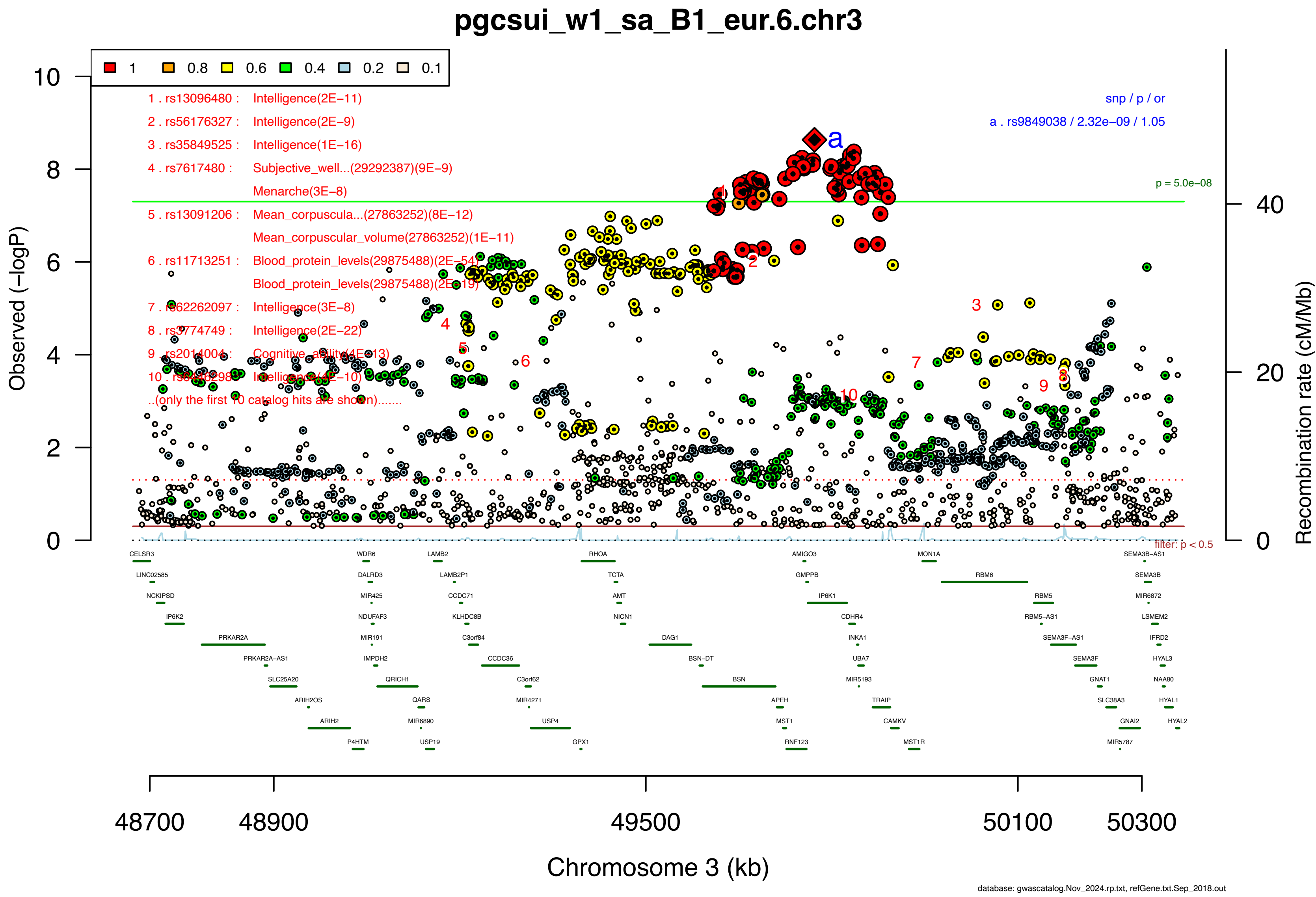
pgcsui_w1_sa_B1_eur.6.chr3
1
0.8
0.6
0.4
0.2
0.1
1 . rs13096480 : Intelligence(2E−11)
2 . rs56176327 : Intelligence(2E−9)
3 . rs35849525 : Intelligence(1E−16)
4 . rs7617480 : Subjective_well...(29292387)(9E−9)
Menarche(3E−8)
5 . rs13091206 : Mean_corpuscula...(27863252)(8E−12)
Mean_corpuscular_volume(27863252)(1E−11)
6 . rs11713251 : Blood_protein_levels(29875488)(2E−54)
Blood_protein_levels(29875488)(2E−19)
7 . rs62262097 : Intelligence(3E−8)
8 . rs3774749 : Intelligence(2E−22)
9 . rs2014004 : Cognitive_ability(4E−13)
10 . rs62262982 : Intelligence(4E−10)
..(only the first 10 catalog hits are shown).......
snp / p / or
a . rs9849038 / 2.32e−09 / 1.05
p = 5.0e−08
filter: p < 0.5
Observed (−logP)
Recombination rate (cM/Mb)
Chromosome 3 (kb)
48700
48900
49500
50100
50300
CELSR3
LINC02585
NCKIPSD
IP6K2
PRKAR2A
PRKAR2A-AS1
SLC25A20
ARIH2OS
ARIH2
P4HTM
WDR6
DALRD3
MIR425
NDUFAF3
MIR191
IMPDH2
QRICH1
QARS
MIR6890
USP19
LAMB2
LAMB2P1
CCDC71
KLHDC8B
C3orf84
CCDC36
C3orf62
MIR4271
USP4
GPX1
RHOA
TCTA
AMT
NICN1
DAG1
BSN-DT
BSN
APEH
MST1
RNF123
AMIGO3
GMPPB
IP6K1
CDHR4
INKA1
UBA7
MIR5193
TRAIP
CAMKV
MST1R
MON1A
RBM6
RBM5
RBM5-AS1
SEMA3F-AS1
SEMA3F
GNAT1
SLC38A3
GNAI2
MIR5787
SEMA3B-AS1
SEMA3B
MIR6872
LSMEM2
IFRD2
HYAL3
NAA80
HYAL1
HYAL2
database: gwascatalog.Nov_2024.rp.txt, refGene.txt.Sep_2018.out

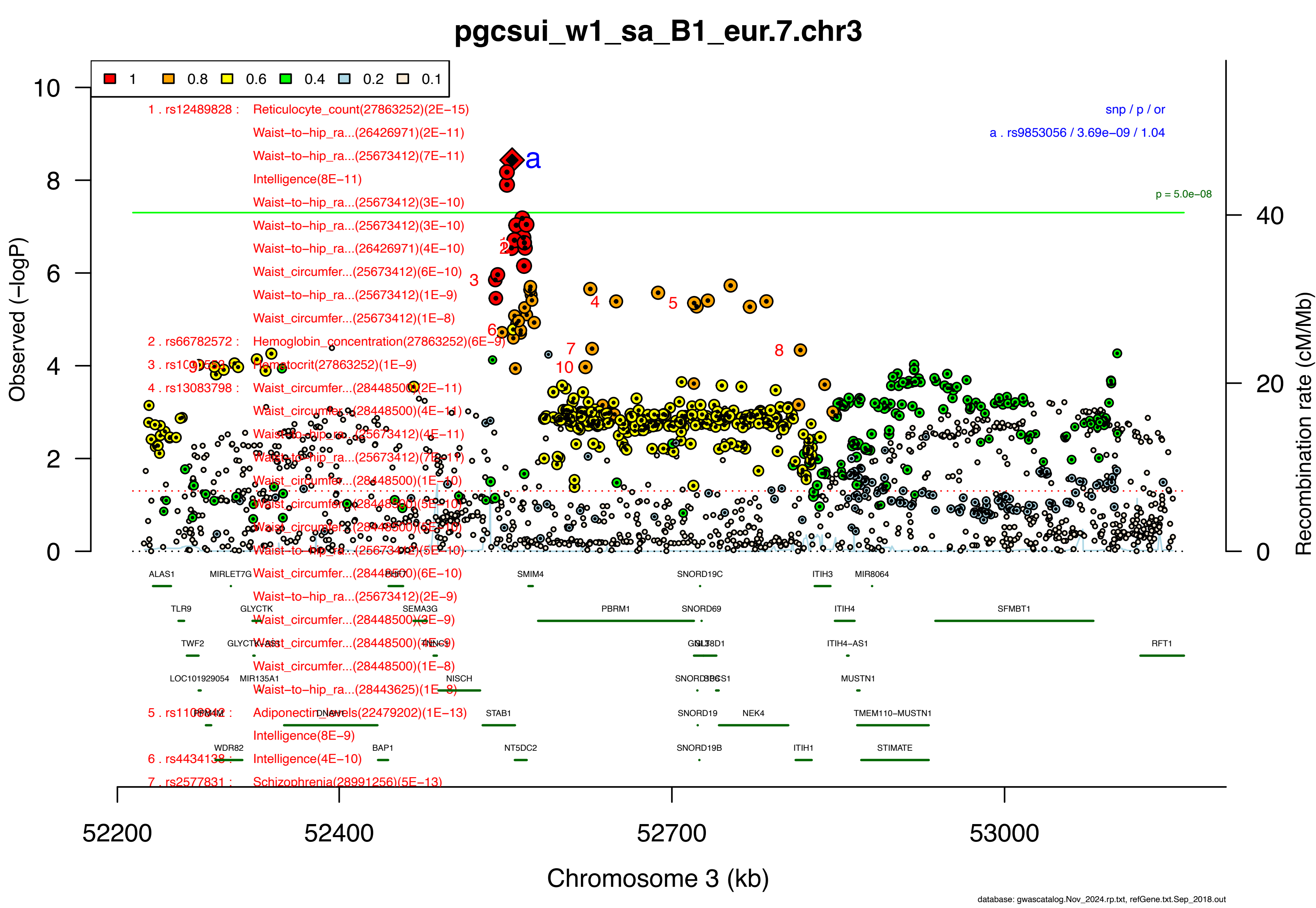
pgcsui_w1_sa_B1_eur.7.chr3
1 0.8 0.6 0.4 0.2 0.1
1 . rs12489828 : Reticulocyte_count(27863252)(2E−15)
Waist−to−hip_ra...(26426971)(2E−11)
Waist−to−hip_ra...(25673412)(7E−11)
Intelligence(8E−11)
Waist−to−hip_ra...(25673412)(3E−10)
Waist−to−hip_ra...(25673412)(3E−10)
Waist−to−hip_ra...(26426971)(4E−10)
Waist_circumfer...(25673412)(6E−10)
Waist−to−hip_ra...(25673412)(1E−9)
Waist_circumfer...(25673412)(1E−8)
2 . rs66782572 : Hemoglobin_concentration(27863252)(6E−9)
Hematocrit(27863252)(1E−9)
4 . rs13083798 : Waist_circumfer...(28448500)(2E−11)
Waist_circumfer...(28448500)(6E−10)
Waist−to−hip_ra...(25673412)(2E−9)
Intelligence(8E−9)
6 . rs4434138 : Intelligence(4E−10)
7 . rs2577831 : Schizophrenia(28991256)(5E−13)
snp / p / or
a . rs9853056 / 3.69e−09 / 1.04
p = 5.0e−08
Observed (−logP)
Recombination rate (cM/Mb)
Chromosome 3 (kb)
52200
52400
52700
53000
ALAS1
MIRLET7G
SMIM4
SNORD19C
ITIH3
MIR8064
TLR9
GLYCTK
SEMA3G
PBRM1
SNORD69
ITIH4
SFMBT1
TWF2
ITIH4−AS1
RFT1
LOC101929054
MIR135A1
NISCH
MUSTN1
STAB1
SNORD19
NEK4
TMEM110−MUSTN1
WDR82
BAP1
NT5DC2
SNORD19B
ITIH1
STIMATE
database: gwascatalog.Nov_2024.rp.txt, refGene.txt.Sep_2018.out

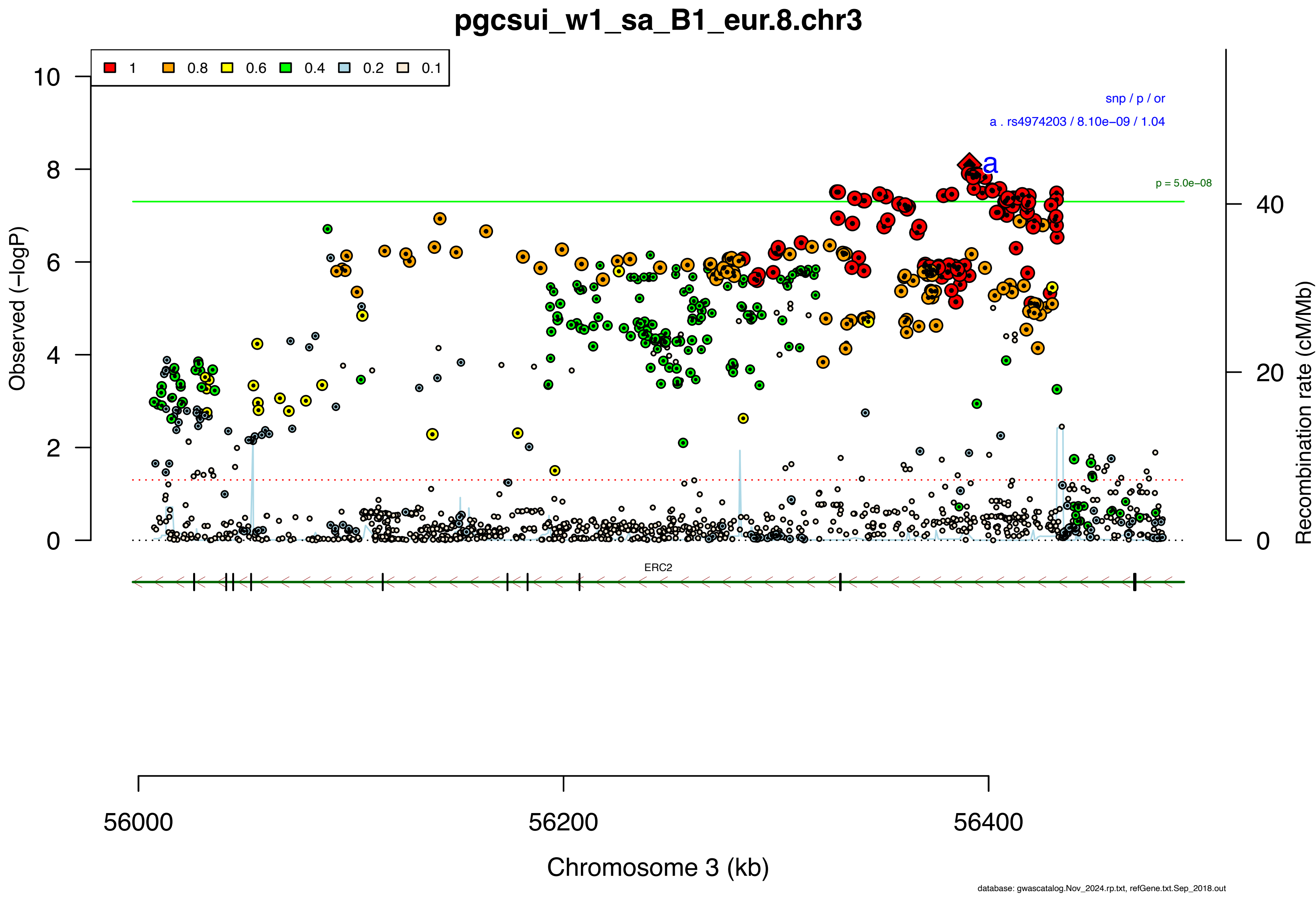

pgcsui_w1_sa_B1_eur.8.chr3
1 0.8 0.6 0.4 0.2 0.1
snp / p / or
a . rs4974203 / 8.10e−09 / 1.04
a
p = 5.0e−08
Observed (−logP)
Recombination rate (cM/Mb)
ERC2
56000
56200
56400
Chromosome 3 (kb)
database: gwascatalog.Nov_2024.rp.txt, refGene.txt.Sep_2018.out

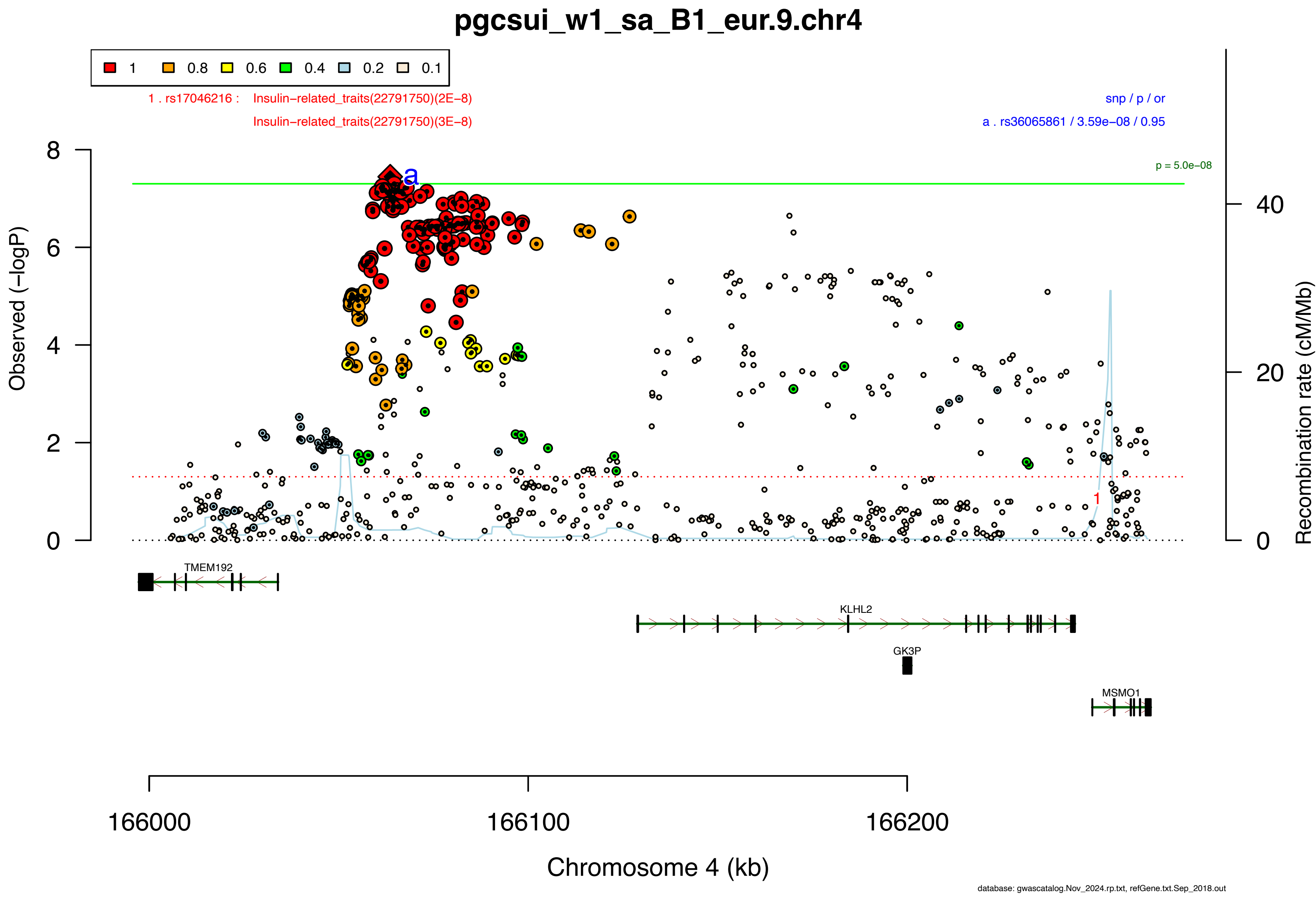
pgcsui_w1_sa_B1_eur.9.chr4
1
0.8
0.6
0.4
0.2
0.1
1 . rs17046216 : Insulin−related_traits(22791750)(2E−8)
Insulin−related_traits(22791750)(3E−8)
snp / p / or
a . rs36065861 / 3.59e−08 / 0.95
p = 5.0e−08
Observed (−logP)
Recombination rate (cM/Mb)
TMEM192
KLHL2
GK3P
MSMO1
166000
166100
166200
Chromosome 4 (kb)
database: gwascatalog.Nov_2024.rp.txt, refGene.txt.Sep_2018.out

**Supplementary Data 2F: Area plots of lead SNPs at the 2 genome-wide significant loci from the GWAS meta-analysis of suicide death in European ancestry samples.**

If >1 independent SNPs are in the same plot, then each is given a different color, and their LD partners are shaded with the same color. Detailed info about each independent index SNP (a., b., c. ...) is provided in the right upper corner. If SNPs share independent index SNPs as an LD partner, the SNP is assigned to the more significant one. Information regarding other GWAS associations at the locus is provided in the left upper corner.

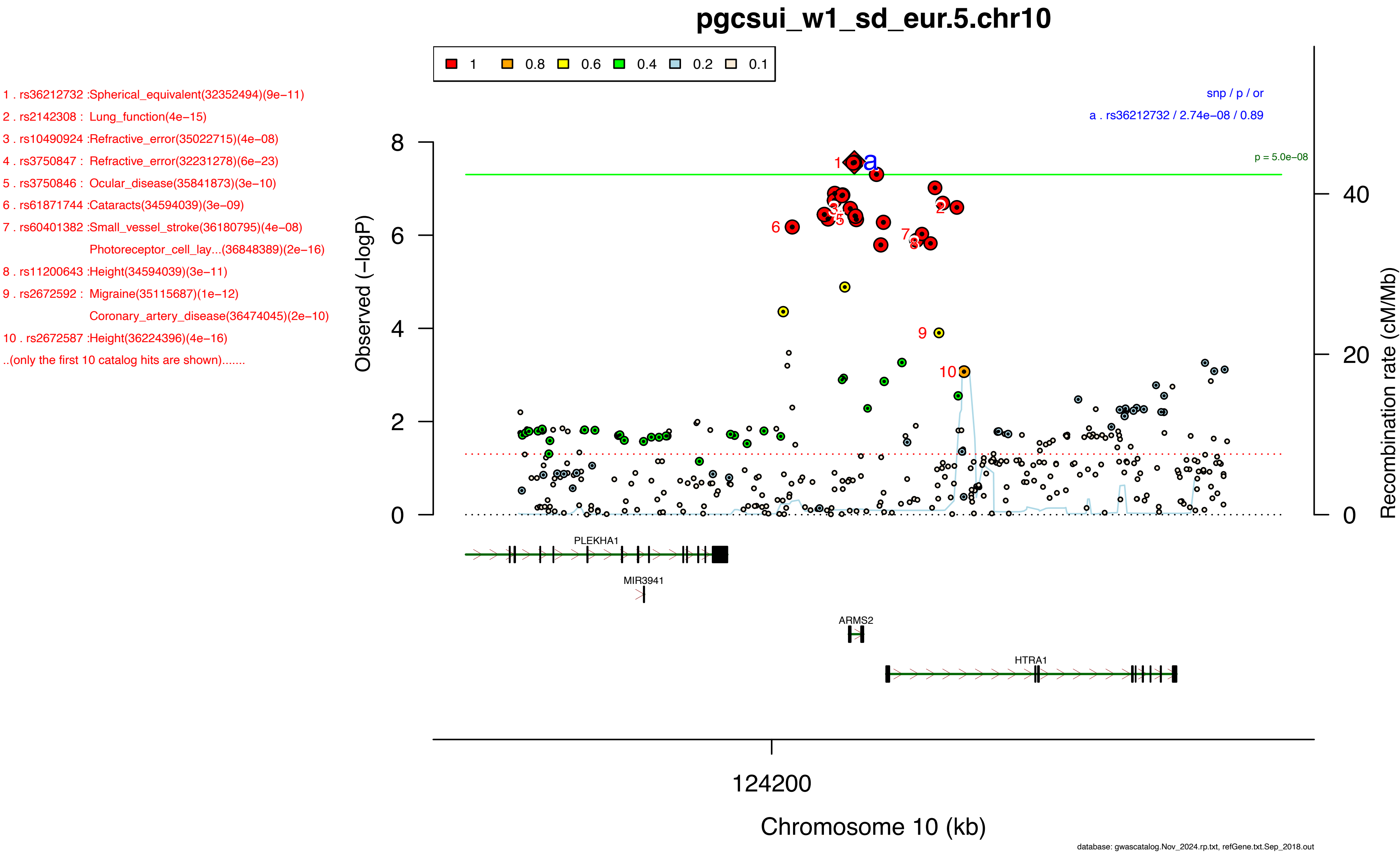
pgcsui_w1_sd_eur.5.chr10
1 0.8 0.6 0.4 0.2 0.1
1 . rs36212732 :Spherical_equivalent(32352494)(9e−11)
2 . rs2142308 : Lung_function(4e−15)
3 . rs10490924 :Refractive_error(35022715)(4e−08)
4 . rs3750847 : Refractive_error(32231278)(6e−23)
5 . rs3750846 : Ocular_disease(35841873)(3e−10)
6 . rs61871744 :Cataracts(34594039)(3e−09)
7 . rs60401382 :Small_vessel_stroke(36180795)(4e−08)
Photoreceptor_cell_lay...(36848389)(2e−16)
8 . rs11200643 :Height(34594039)(3e−11)
9 . rs2672592 : Migraine(35115687)(1e−12)
Coronary_artery_disease(36474045)(2e−10)
10 . rs2672587 :Height(36224396)(4e−16)
..(only the first 10 catalog hits are shown).......
snp / p / or
a . rs36212732 / 2.74e−08 / 0.89
p = 5.0e−08
Observed (−logP)
Recombination rate (cM/Mb)
PLEKHA1
MIR3941
ARMS2
HTRA1
124200
Chromosome 10 (kb)
database: gwascatalog.Nov_2024.rp.txt, refGene.txt.Sep_2018.out

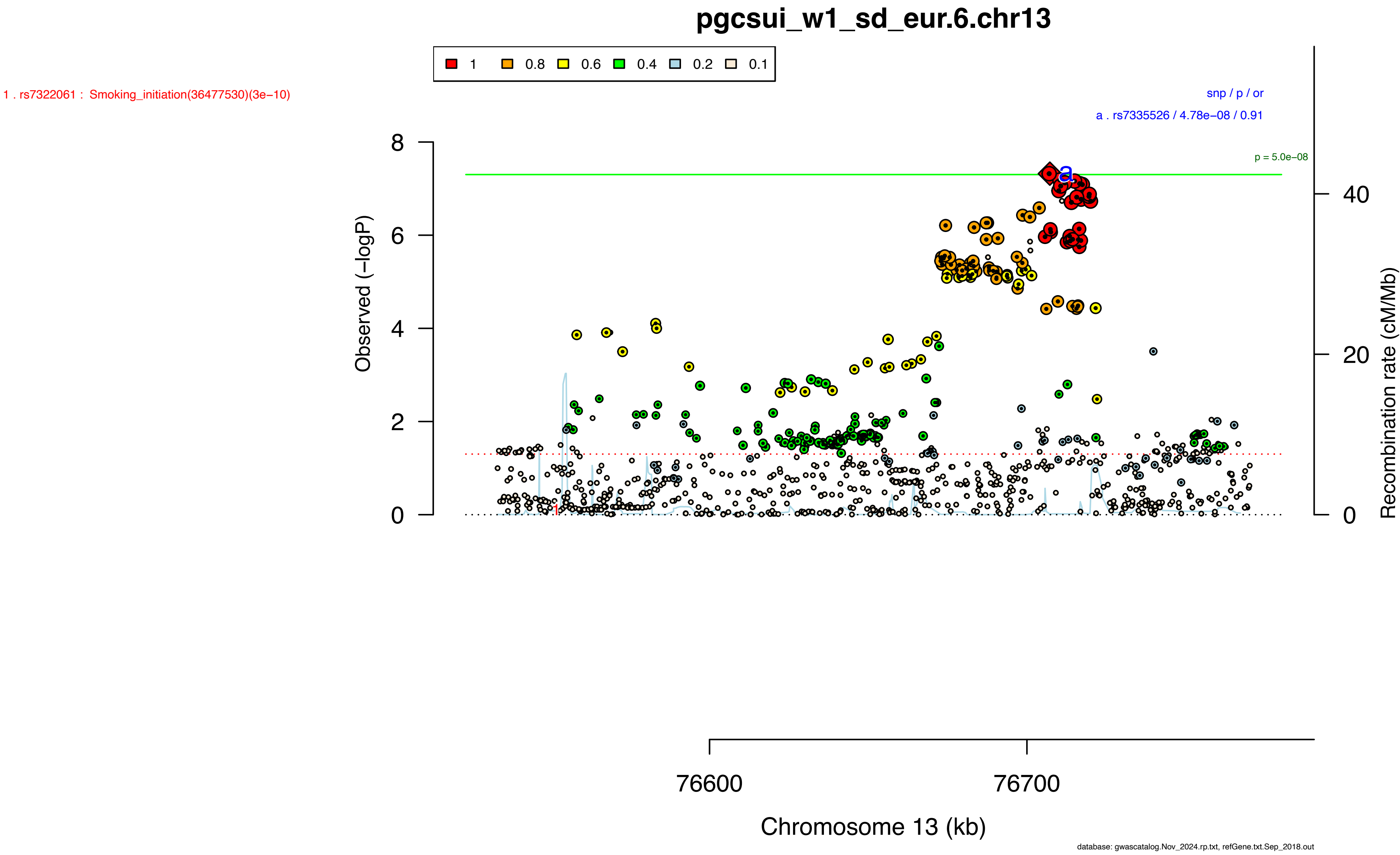

pgcsui_w1_sd_eur.6.chr13
1
0.8
0.6
0.4
0.2
0.1
1 . rs7322061 : Smoking_initiation(36477530)(3e−10)
snp / p / or
a . rs7335526 / 4.78e−08 / 0.91
p = 5.0e−08
Observed (−logP)
Recombination rate (cM/Mb)
Chromosome 13 (kb)
76600
76700
database: gwascatalog.Nov_2024.rp.txt, refGene.txt.Sep_2018.out

**Supplementary Data 2G: Area plots of lead SNPs at the 53 genome-wide significant loci from the multi-ancestry GWAS meta-analysis of suicidal behavior.**

If >1 independent SNPs are in the same plot, then each is given a different color, and their LD partners are shaded with the same color. Detailed info about each independent index SNP (a., b., c. ...) is provided in the right upper corner. If SNPs share independent index SNPs as an LD partner, the SNP is assigned to the more significant one. Information regarding other GWAS associations at the locus is provided in the left upper corner.

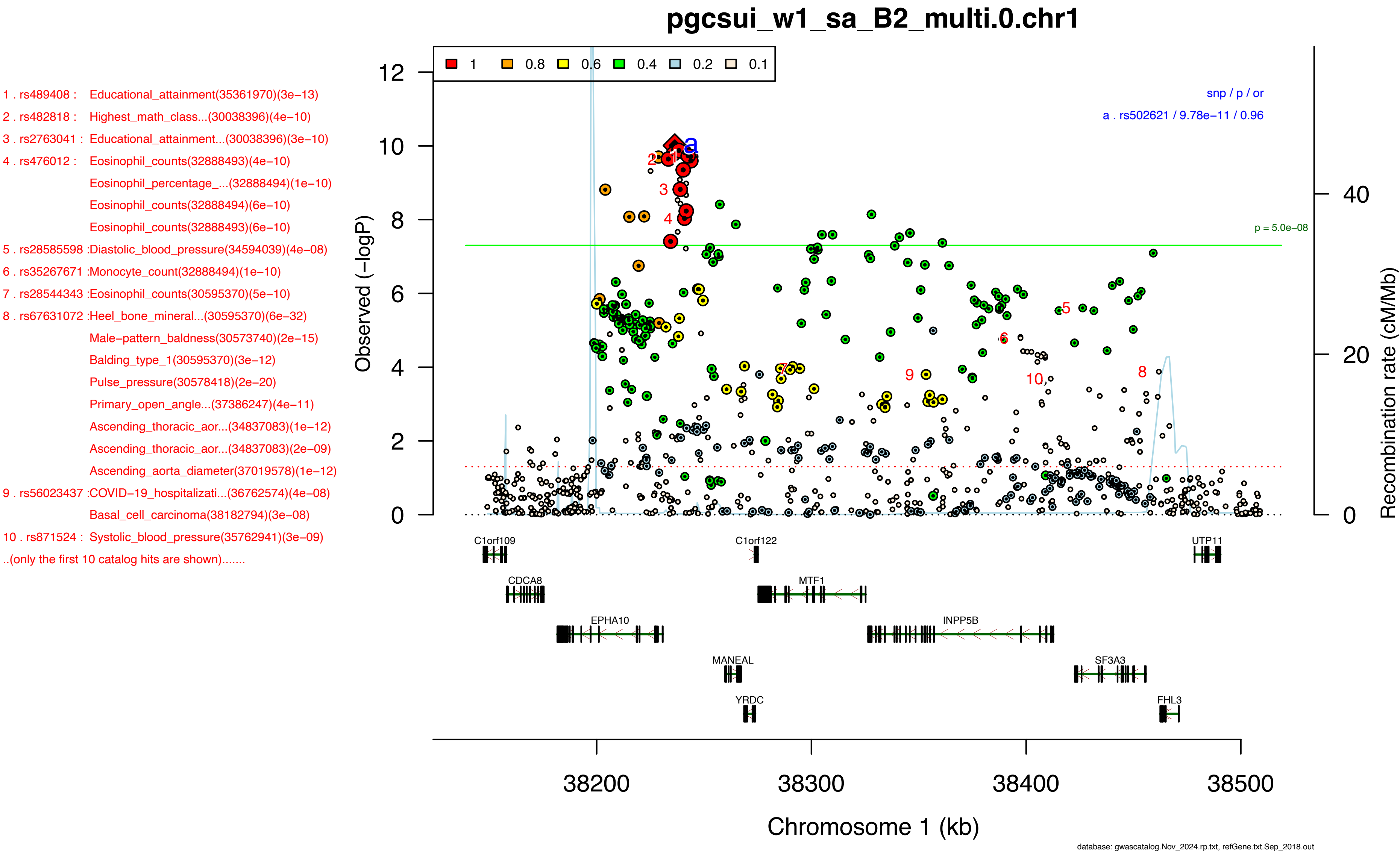
pgcsui_w1_sa_B2_multi.0.chr1
1 . rs489408 : Educational_attainment(35361970)(3e−13)
2 . rs482818 : Highest_math_class...(30038396)(4e−10)
3 . rs2763041 : Educational_attainment...(30038396)(3e−10)
4 . rs476012 : Eosinophil_counts(32888493)(4e−10)
Eosinophil_percentage_...(32888494)(1e−10)
Eosinophil_counts(32888494)(6e−10)
Eosinophil_counts(32888493)(6e−10)
5 . rs28585598 :Diastolic_blood_pressure(34594039)(4e−08)
6 . rs35267671 :Monocyte_count(32888494)(1e−10)
7 . rs28544343 :Eosinophil_counts(30595370)(5e−10)
8 . rs67631072 :Heel_bone_mineral...(30595370)(6e−32)
Male−pattern_baldness(30573740)(2e−15)
Balding_type_1(30595370)(3e−12)
Pulse_pressure(30578418)(2e−20)
Primary_open_angle...(37386247)(4e−11)
Ascending_thoracic_aor...(34837083)(1e−12)
Ascending_thoracic_aor...(34837083)(2e−09)
Ascending_aorta_diameter(37019578)(1e−12)
9 . rs56023437 :COVID−19_hospitalizati...(36762574)(4e−08)
Basal_cell_carcinoma(38182794)(3e−08)
10 . rs871524 : Systolic_blood_pressure(35762941)(3e−09)
..(only the first 10 catalog hits are shown).......
1
0.8
0.6
0.4
0.2
0.1
snp / p / or
a . rs502621 / 9.78e−11 / 0.96
p = 5.0e−08
Observed (−logP)
Recombination rate (cM/Mb)
C1orf109
C1orf122
UTP11
CDCA8
MTF1
EPHA10
INPP5B
MANEAL
SF3A3
YRDC
FHL3
38200
38300
38400
38500
Chromosome 1 (kb)
database: gwascatalog.Nov_2024.rp.txt, refGene.txt.Sep_2018.out

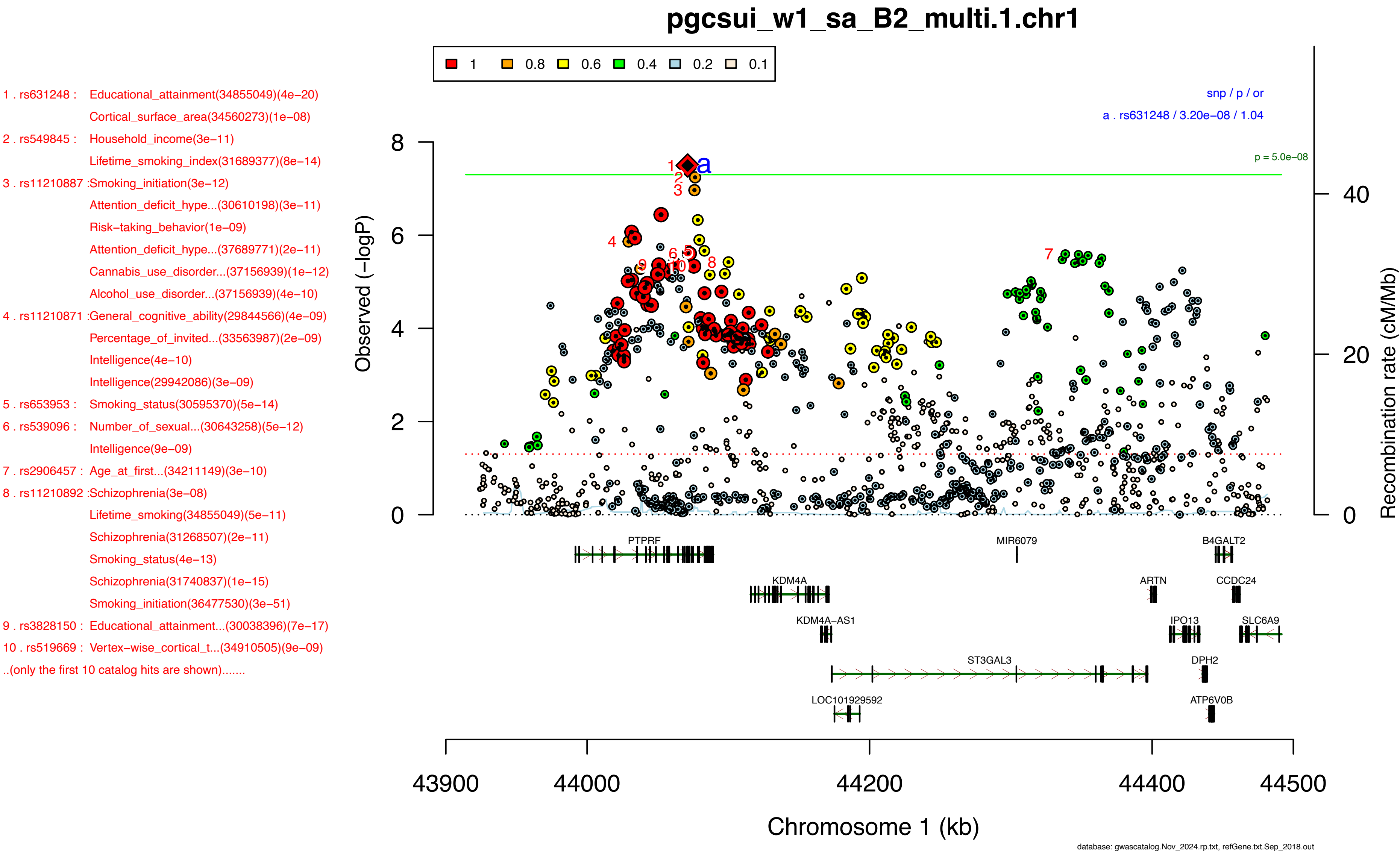
pgcsui_w1_sa_B2_multi.1.chr1
1 0.8 0.6 0.4 0.2 0.1
1 . rs631248 : Educational_attainment(34855049)(4e−20)
Cortical_surface_area(34560273)(1e−08)
2 . rs549845 : Household_income(3e−11)
Lifetime_smoking_index(31689377)(8e−14)
3 . rs11210887 :Smoking_initiation(3e−12)
Attention_deficit_hype...(30610198)(3e−11)
Risk−taking_behavior(1e−09)
Attention_deficit_hype...(37689771)(2e−11)
Cannabis_use_disorder...(37156939)(1e−12)
Alcohol_use_disorder...(37156939)(4e−10)
4 . rs11210871 :General_cognitive_ability(29844566)(4e−09)
Percentage_of_invited...(33563987)(2e−09)
Intelligence(4e−10)
Intelligence(29942086)(3e−09)
5 . rs653953 : Smoking_status(30595370)(5e−14)
6 . rs539096 : Number_of_sexual...(30643258)(5e−12)
Intelligence(9e−09)
7 . rs2906457 : Age_at_first...(34211149)(3e−10)
8 . rs11210892 :Schizophrenia(3e−08)
Lifetime_smoking(34855049)(5e−11)
Schizophrenia(31268507)(2e−11)
Smoking_status(4e−13)
Schizophrenia(31740837)(1e−15)
Smoking_initiation(36477530)(3e−51)
9 . rs3828150 : Educational_attainment...(30038396)(7e−17)
10 . rs519669 : Vertex−wise_cortical_t...(34910505)(9e−09)
..(only the first 10 catalog hits are shown).......
snp / p / or
a . rs631248 / 3.20e−08 / 1.04
p = 5.0e−08
Observed (−logP)
Recombination rate (cM/Mb)
PTPRF
KDM4A
KDM4A-AS1
ST3GAL3
LOC101929592
MIR6079
ARTN
IPO13
DPH2
ATP6V0B
B4GALT2
CCDC24
SLC6A9
43900 44000 44200 44400 44500
Chromosome 1 (kb)
database: gwascatalog.Nov_2024.rp.txt, refGene.txt.Sep_2018.out

1 . rs7681616 : Schizophrenia(35396580)(4e−08)
2 . rs6831966 : Externalizing_behaviou...(34446935)(3e−14)
3 . rs966457 : Body_mass_index(39375568)(1e−10)
Body_mass_index(36581621)(1e−12)
4 . rs331949 : Adventurousness(30643258)(2e−09)
5 . rs1443186 : Metabolic_syndrome(39349817)(1e−16)
6 . rs34959108 :Educational_attainment(35361970)(9e−09)
7 . rs11933763 :Hypertension(37947095)(4e−09)
8 . rs1116690 : Smoking_initiation(2e−08)
Decaffeinated_coffee_c...(35898629)(1e−08)
Smoking_initiation(36477530)(1e−24)
9 . rs1425523 : Smoking_initiation(36477530)(5e−24)
10 . rs2081474 :Smoking_initiation(36477530)(6e−24)
..(only the first 10 catalog hits are shown).......

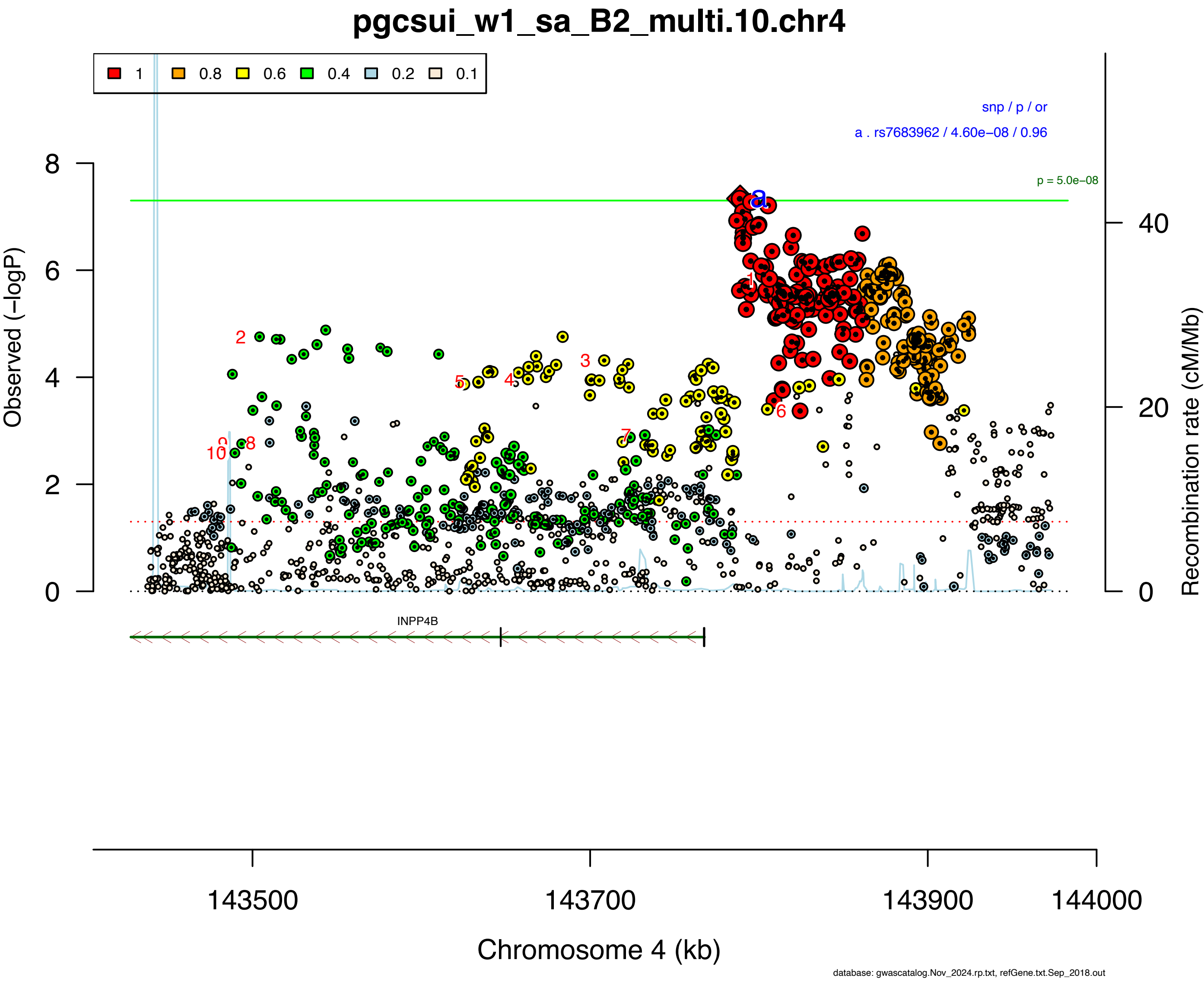

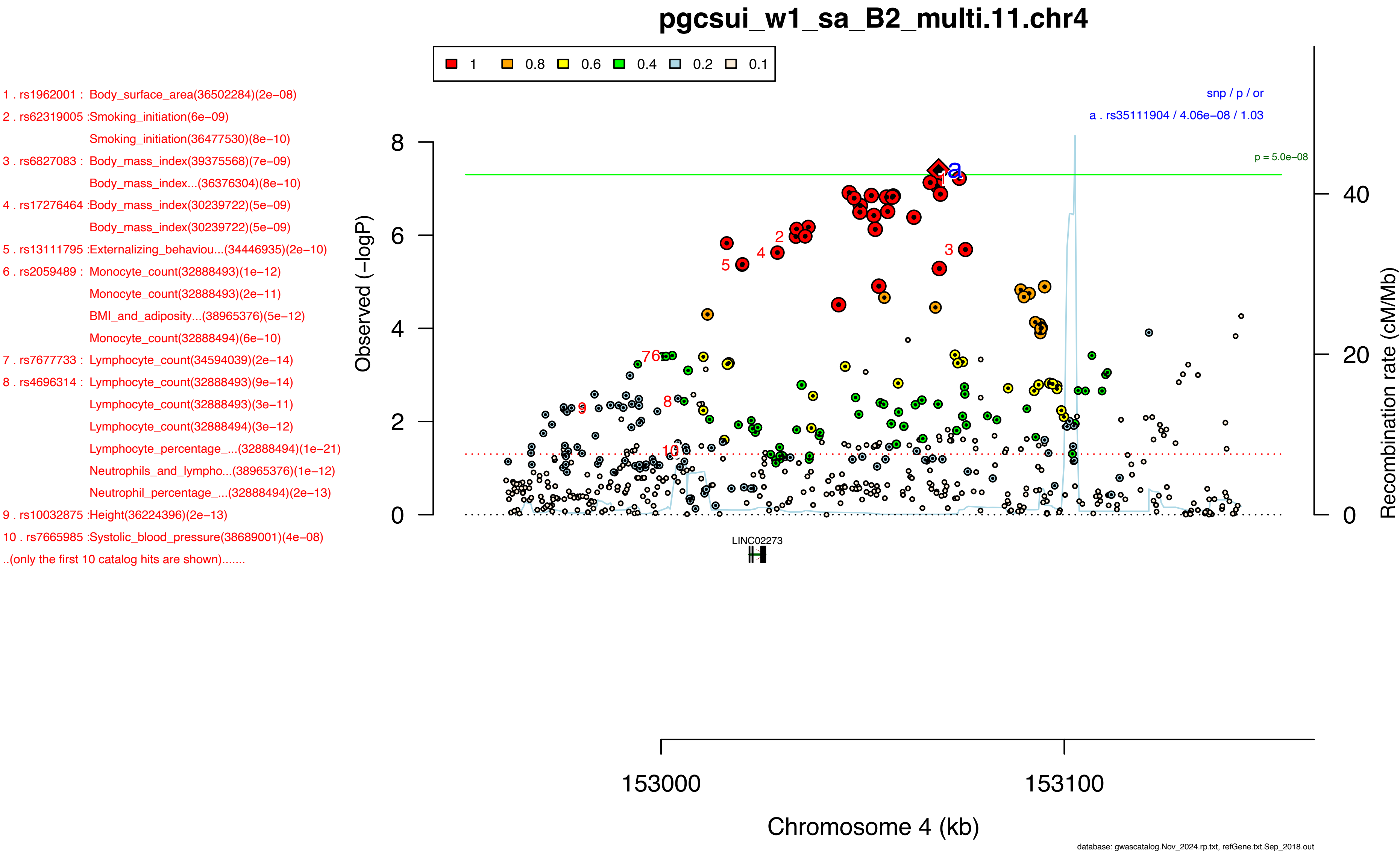

pgcsui_w1_sa_B2_multi.11.chr4
1
0.8
0.6
0.4
0.2
0.1
1 . rs1962001 : Body_surface_area(36502284)(2e−08)
2 . rs62319005 :Smoking_initiation(6e−09)
Smoking_initiation(36477530)(8e−10)
3 . rs6827083 : Body_mass_index(39375568)(7e−09)
Body_mass_index...(36376304)(8e−10)
4 . rs17276464 :Body_mass_index(30239722)(5e−09)
Body_mass_index(30239722)(5e−09)
5 . rs13111795 :Externalizing_behaviou...(34446935)(2e−10)
6 . rs2059489 : Monocyte_count(32888493)(1e−12)
Monocyte_count(32888493)(2e−11)
BMI_and_adiposity...(38965376)(5e−12)
Monocyte_count(32888494)(6e−10)
7 . rs7677733 : Lymphocyte_count(34594039)(2e−14)
8 . rs4696314 : Lymphocyte_count(32888493)(9e−14)
Lymphocyte_count(32888493)(3e−11)
Lymphocyte_count(32888494)(3e−12)
Lymphocyte_percentage_...(32888494)(1e−21)
Neutrophils_and_lympho...(38965376)(1e−12)
Neutrophil_percentage_...(32888494)(2e−13)
9 . rs10032875 :Height(36224396)(2e−13)
10 . rs7665985 :Systolic_blood_pressure(38689001)(4e−08)
..(only the first 10 catalog hits are shown).......
snp / p / or
a . rs35111904 / 4.06e−08 / 1.03
p = 5.0e−08
Observed (−logP)
Recombination rate (cM/Mb)
LINC02273
153000
153100
Chromosome 4 (kb)
database: gwascatalog.Nov_2024.rp.txt, refGene.txt.Sep_2018.out

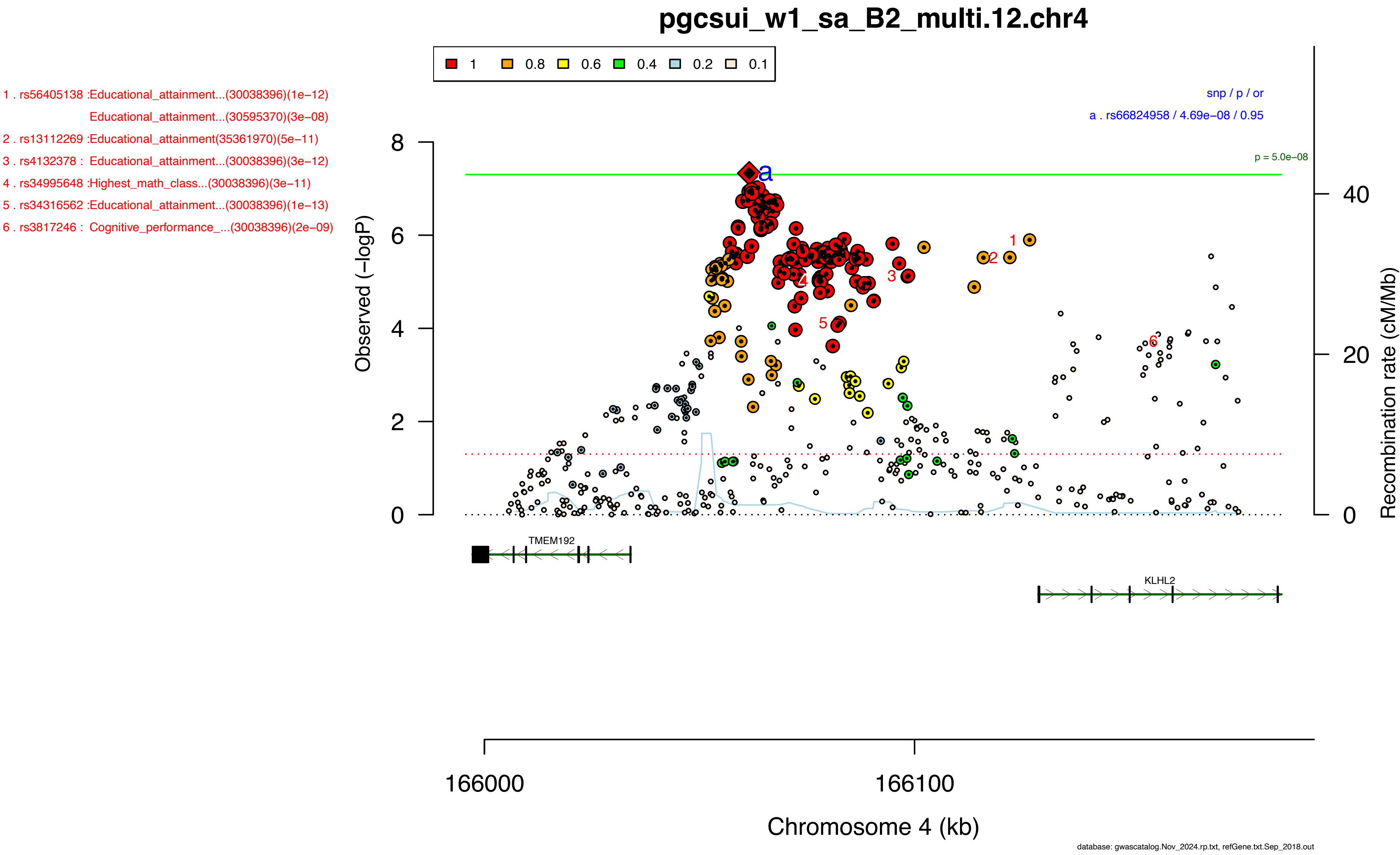
pgcsui_w1_sa_B2_multi.12.chr4
1 0.8 0.6 0.4 0.2 0.1
1 . rs56405138 :Educational_attainment...(30038396)(1e−12)
Educational_attainment...(30595370)(3e−08)
2 . rs13112269 :Educational_attainment(35361970)(5e−11)
3 . rs4132378 : Educational_attainment...(30038396)(3e−12)
4 . rs34995648 :Highest_math_class...(30038396)(3e−11)
5 . rs34316562 :Educational_attainment...(30038396)(1e−13)
6 . rs3817246 : Cognitive_performance_...(30038396)(2e−09)
snp / p / or
a . rs66824958 / 4.69e−08 / 0.95
p = 5.0e−08
Observed (−logP)
Recombination rate (cM/Mb)
TMEM192
KLHL2
166000
166100
Chromosome 4 (kb)
database: gwascatalog.Nov_2024.rp.txt, refGene.txt.Sep_2018.out

### pgcsui_w1_sa_B2_multi.13.chr5

1 . rs62367520 :Smoking_initiation(1e−10)
Age_of_smoking...(30643251)(1e−09)
Smoking_initiation(36477530)(3e−24)
2 . rs62367522 :Smoking_initiation(36477530)(4e−24)
3 . rs62367470 :Educational_attainment(35361970)(6e−09)
4 . rs12517546 :Externalizing_behaviou...(34446935)(8e−13)
5 . rs12523398 :Age_at_first...(34211149)(1e−15)
6 . rs4493682 : Cognitive_ability,_yea...(31374203)(3e−10)
7 . rs62369913 :Smoking_cessation(5e−10)
8 . rs58713827 :Smoking_initiation(36477530)(2e−20)
9 . rs62367473 :Schizophrenia_vs_ADHD...(33686288)(2e−08)
10 . rs6881773 :Lung_cancer(34594039)(1e−08)
..(only the first 10 catalog hits are shown).......

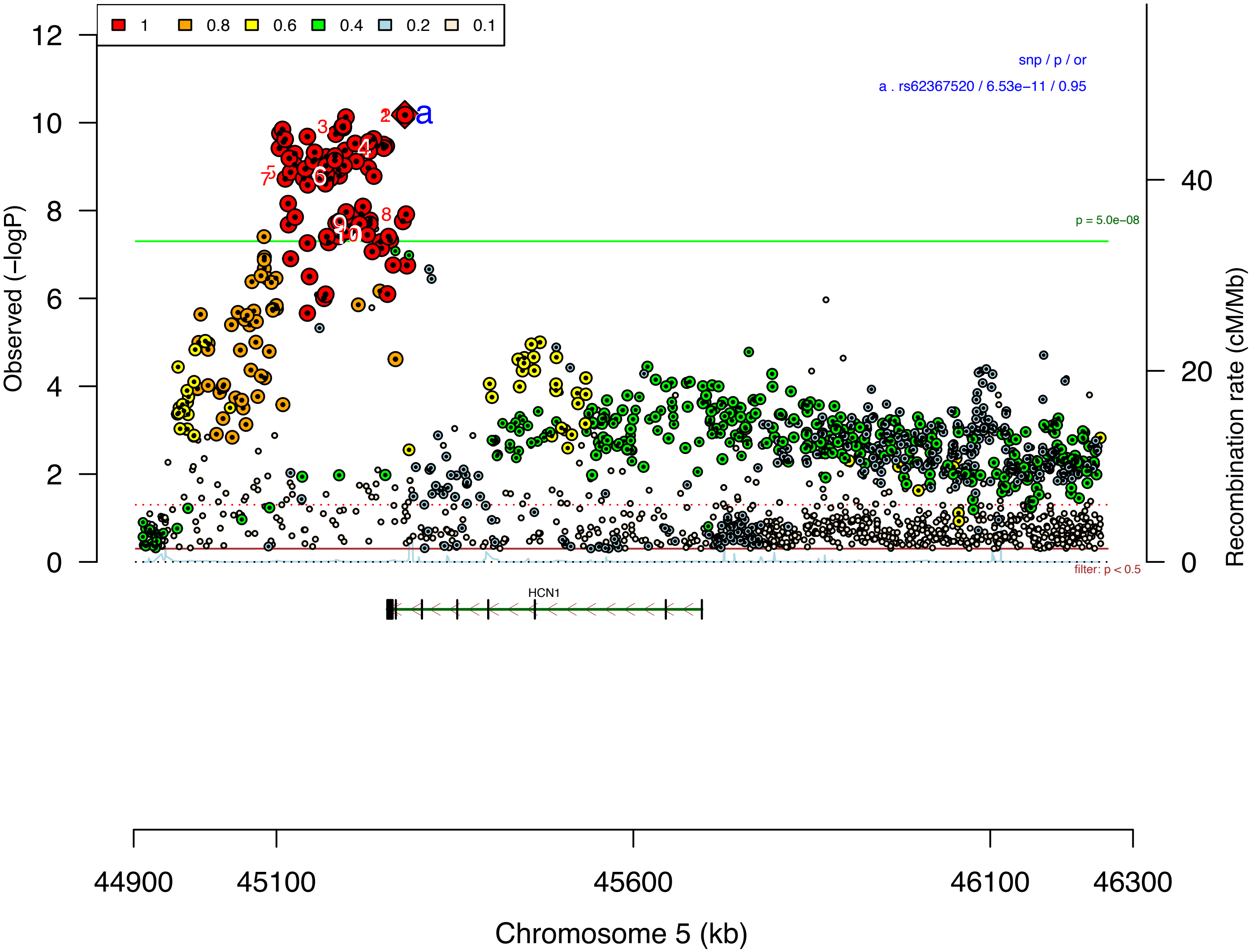


database: gwascatalog.Nov_2024.rp.txt, refGene.txt.Sep_2018.out

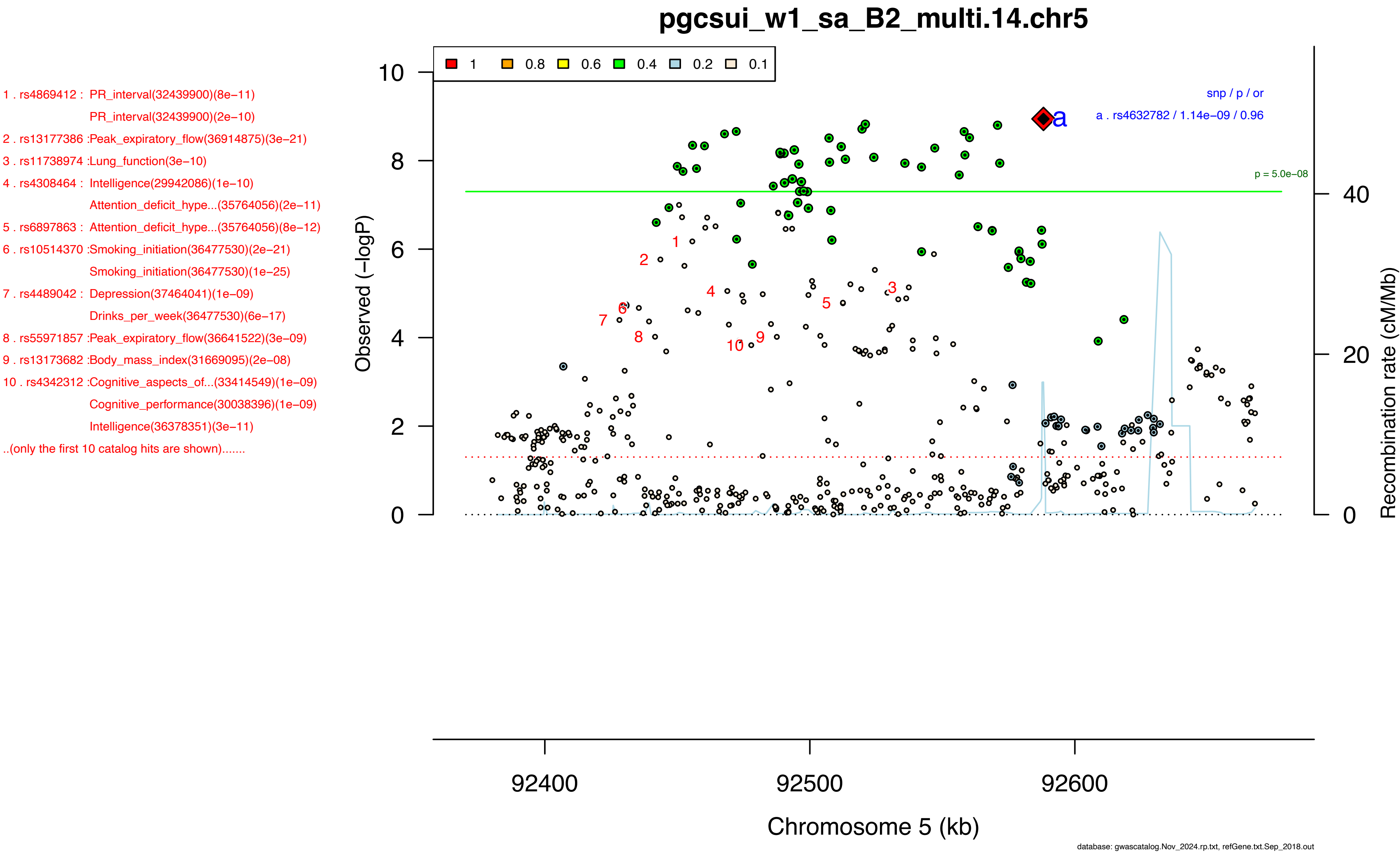
pgcsui_w1_sa_B2_multi.14.chr5
1 . rs4869412 : PR_interval(32439900)(8e−11)
PR_interval(32439900)(2e−10)
2 . rs13177386 :Peak_expiratory_flow(36914875)(3e−21)
3 . rs11738974 :Lung_function(3e−10)
4 . rs4308464 : Intelligence(29942086)(1e−10)
Attention_deficit_hype...(35764056)(2e−11)
5 . rs6897863 : Attention_deficit_hype...(35764056)(8e−12)
6 . rs10514370 :Smoking_initiation(36477530)(2e−21)
Smoking_initiation(36477530)(1e−25)
7 . rs4489042 : Depression(37464041)(1e−09)
Drinks_per_week(36477530)(6e−17)
8 . rs55971857 :Peak_expiratory_flow(36641522)(3e−09)
9 . rs13173682 :Body_mass_index(31669095)(2e−08)
10 . rs4342312 :Cognitive_aspects_of...(33414549)(1e−09)
Cognitive_performance(30038396)(1e−09)
Intelligence(36378351)(3e−11)
..(only the first 10 catalog hits are shown).......
1
0.8
0.6
0.4
0.2
0.1
snp / p / or
a . rs4632782 / 1.14e−09 / 0.96
p = 5.0e−08
Observed (−logP)
Recombination rate (cM/Mb)
Chromosome 5 (kb)
92400
92500
92600
database: gwascatalog.Nov_2024.rp.txt, refGene.txt.Sep_2018.out

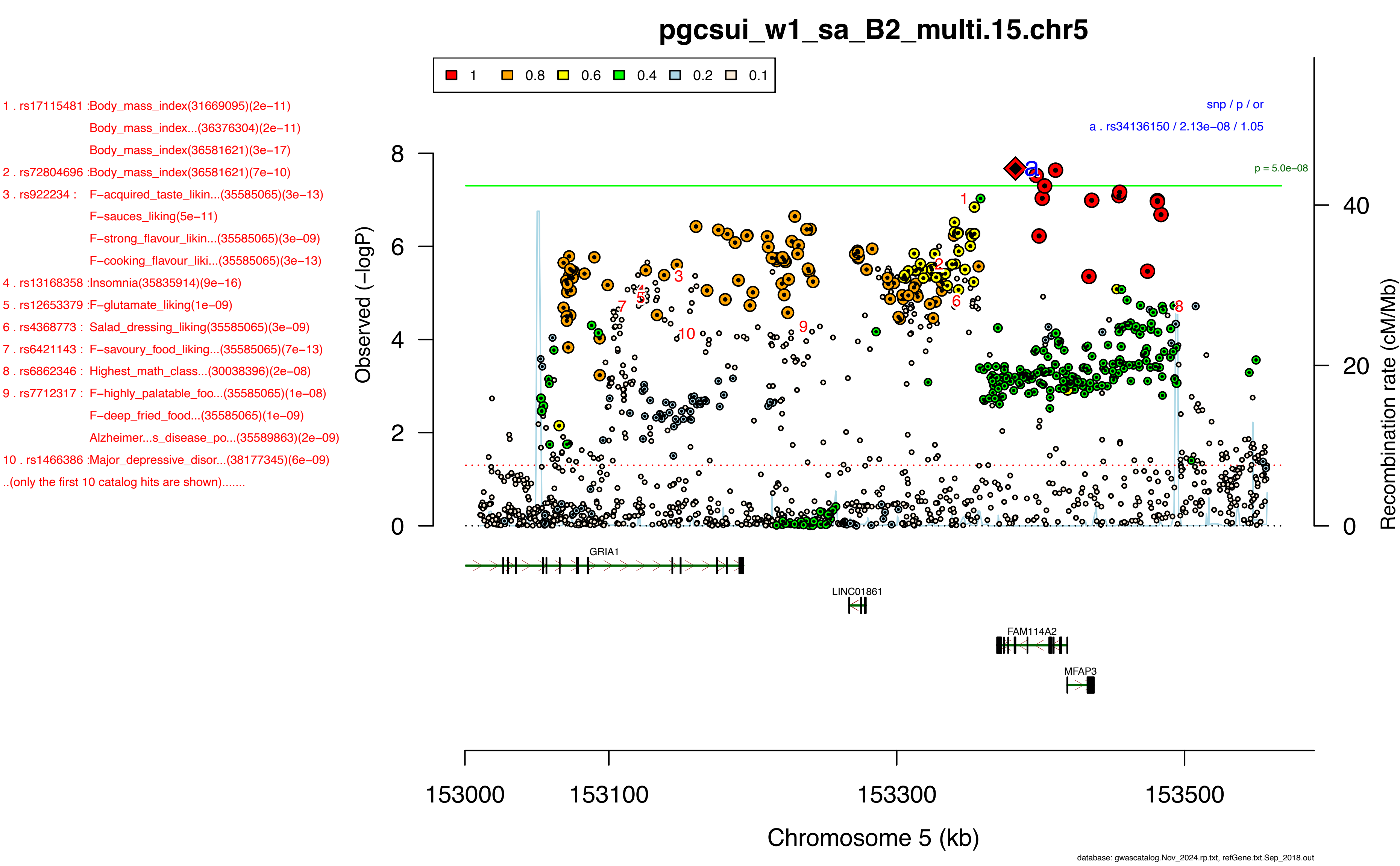

pgcsui_w1_sa_B2_multi.15.chr5
1 0.8 0.6 0.4 0.2 0.1
1 . rs17115481 :Body_mass_index(31669095)(2e−11)
Body_mass_index...(36376304)(2e−11)
Body_mass_index(36581621)(3e−17)
2 . rs72804696 :Body_mass_index(36581621)(7e−10)
3 . rs922234 : F−acquired_taste_likin...(35585065)(3e−13)
F−sauces_liking(5e−11)
F−strong_flavour_likin...(35585065)(3e−09)
F−cooking_flavour_liki...(35585065)(3e−13)
4 . rs13168358 :Insomnia(35835914)(9e−16)
5 . rs12653379 :F−glutamate_liking(1e−09)
6 . rs4368773 : Salad_dressing_liking(35585065)(3e−09)
7 . rs6421143 : F−savoury_food_liking...(35585065)(7e−13)
8 . rs6862346 : Highest_math_class...(30038396)(2e−08)
9 . rs7712317 : F−highly_palatable_foo...(35585065)(1e−08)
F−deep_fried_food...(35585065)(1e−09)
Alzheimer...s_disease_po...(35589863)(2e−09)
10 . rs1466386 :Major_depressive_disor...(38177345)(6e−09)
..(only the first 10 catalog hits are shown).......
snp / p / or
a . rs34136150 / 2.13e−08 / 1.05
p = 5.0e−08
Observed (−logP)
Recombination rate (cM/Mb)
GRIA1
LINC01861
FAM114A2
MFAP3
153000 153100 153300 153500
Chromosome 5 (kb)
database: gwascatalog.Nov_2024.rp.txt, refGene.txt.Sep_2018.out

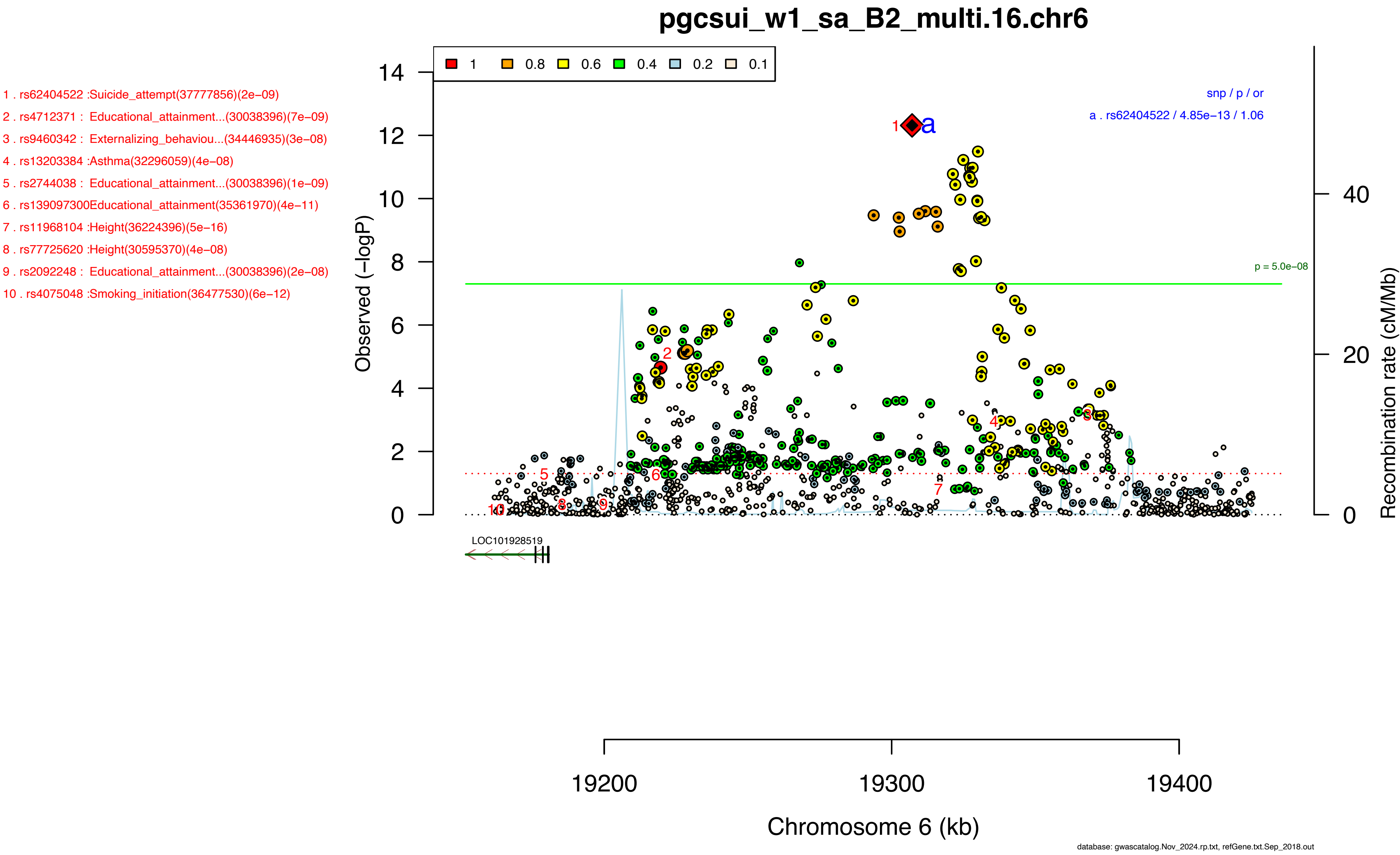

pgcsui_w1_sa_B2_multi.16.chr6
1 . rs62404522 :Suicide_attempt(37777856)(2e−09)
2 . rs4712371 : Educational_attainment...(30038396)(7e−09)
3 . rs9460342 : Externalizing_behaviou...(34446935)(3e−08)
4 . rs13203384 :Asthma(32296059)(4e−08)
5 . rs2744038 : Educational_attainment...(30038396)(1e−09)
6 . rs139097300Educational_attainment(35361970)(4e−11)
7 . rs11968104 :Height(36224396)(5e−16)
8 . rs77725620 :Height(30595370)(4e−08)
9 . rs2092248 : Educational_attainment...(30038396)(2e−08)
10 . rs4075048 :Smoking_initiation(36477530)(6e−12)
1 0.8 0.6 0.4 0.2 0.1
snp / p / or
a . rs62404522 / 4.85e−13 / 1.06
p = 5.0e−08
Observed (−logP)
Recombination rate (cM/Mb)
LOC101928519
19200 19300 19400
Chromosome 6 (kb)
database: gwascatalog.Nov_2024.rp.txt, refGene.txt.Sep_2018.out

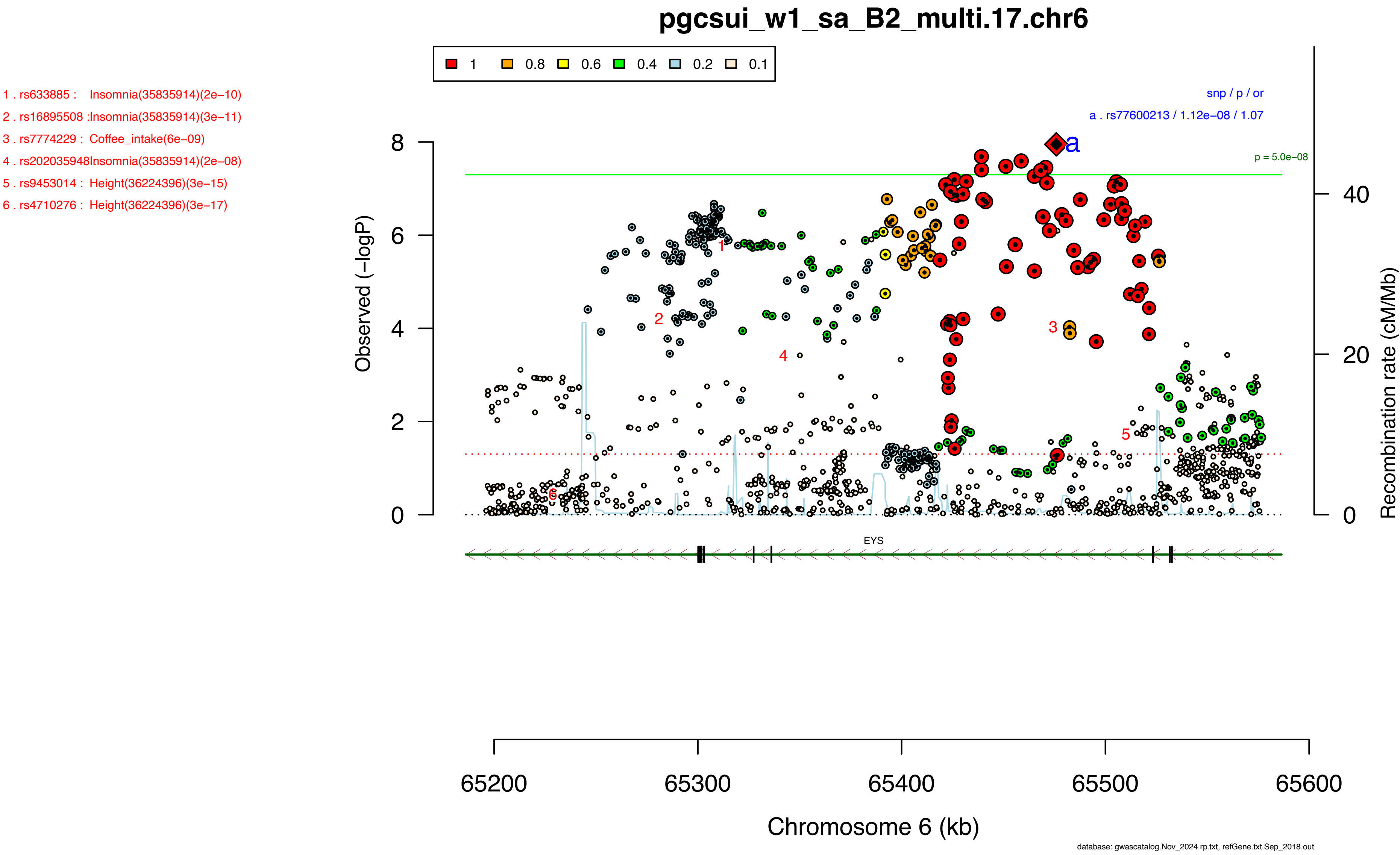

pgcsui_w1_sa_B2_multi.17.chr6
1 0.8 0.6 0.4 0.2 0.1
1 . rs633885 : Insomnia(35835914)(2e−10)
2 . rs16895508 :Insomnia(35835914)(3e−11)
3 . rs7774229 : Coffee_intake(6e−09)
4 . rs202035948Insomnia(35835914)(2e−08)
5 . rs9453014 : Height(36224396)(3e−15)
6 . rs4710276 : Height(36224396)(3e−17)
snp / p / or
a . rs77600213 / 1.12e−08 / 1.07
p = 5.0e−08
Observed (−logP)
Recombination rate (cM/Mb)
EYS
Chromosome 6 (kb)
65200
65300
65400
65500
65600
database: gwascatalog.Nov_2024.rp.txt, refGene.txt.Sep_2018.out

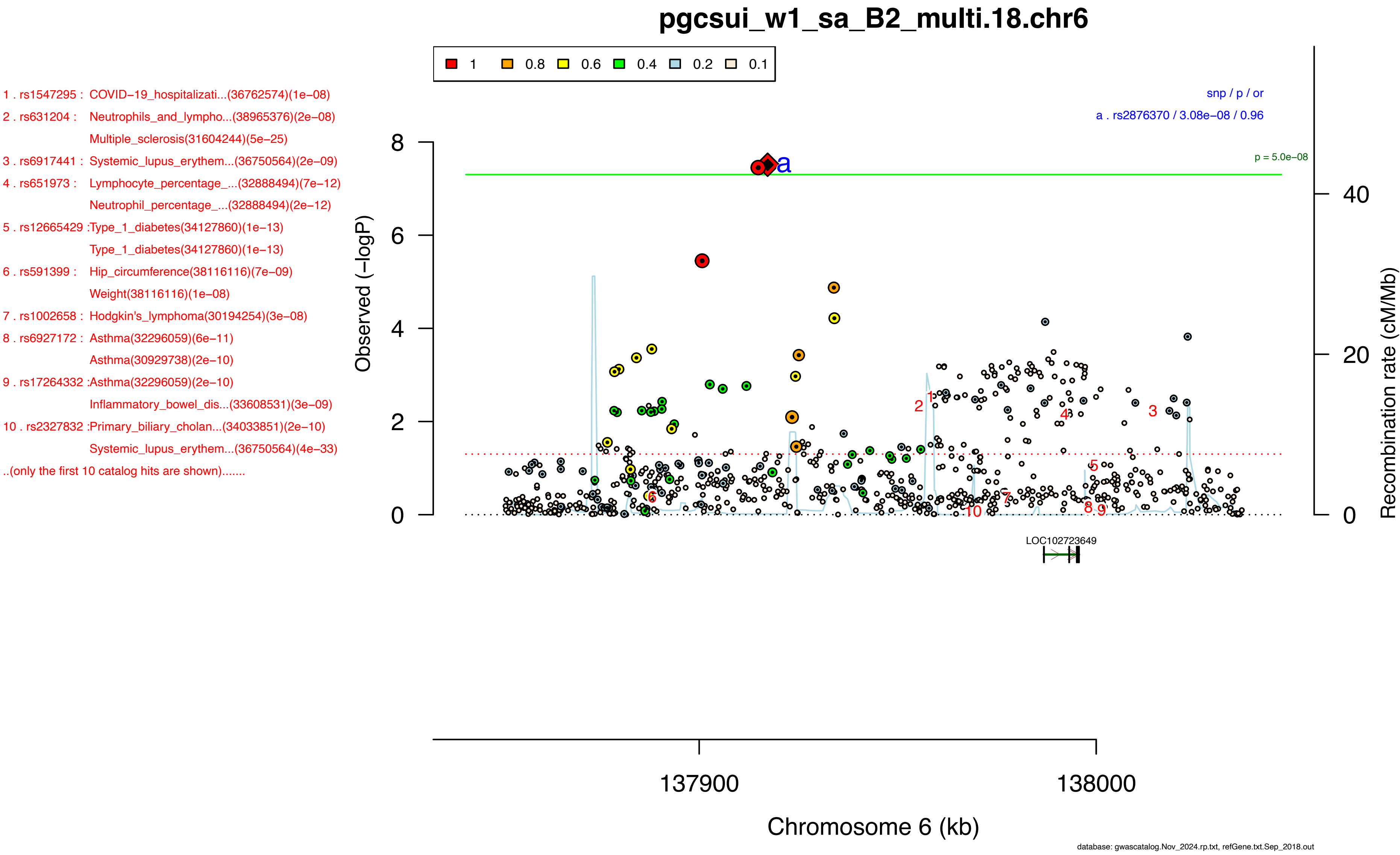
pgcsui_w1_sa_B2_multi.18.chr6
1 0.8 0.6 0.4 0.2 0.1
1 . rs1547295 : COVID−19_hospitalizati...(36762574)(1e−08)
2 . rs631204 : Neutrophils_and_lympho...(38965376)(2e−08)
Multiple_sclerosis(31604244)(5e−25)
3 . rs6917441 : Systemic_lupus_erythem...(36750564)(2e−09)
4 . rs651973 : Lymphocyte_percentage_...(32888494)(7e−12)
Neutrophil_percentage_...(32888494)(2e−12)
5 . rs12665429 :Type_1_diabetes(34127860)(1e−13)
Type_1_diabetes(34127860)(1e−13)
6 . rs591399 : Hip_circumference(38116116)(7e−09)
Weight(38116116)(1e−08)
7 . rs1002658 : Hodgkin's_lymphoma(30194254)(3e−08)
8 . rs6927172 : Asthma(32296059)(6e−11)
Asthma(30929738)(2e−10)
9 . rs17264332 :Asthma(32296059)(2e−10)
Inflammatory_bowel_dis...(33608531)(3e−09)
10 . rs2327832 :Primary_biliary_cholan...(34033851)(2e−10)
Systemic_lupus_erythem...(36750564)(4e−33)
..(only the first 10 catalog hits are shown).......
snp / p / or
a . rs2876370 / 3.08e−08 / 0.96
p = 5.0e−08
Observed (−logP)
8
6
4
2
0
Recombination rate (cM/Mb)
40
20
0
LOC102723649
137900
138000
Chromosome 6 (kb)
database: gwascatalog.Nov_2024.rp.txt, refGene.txt.Sep_2018.out

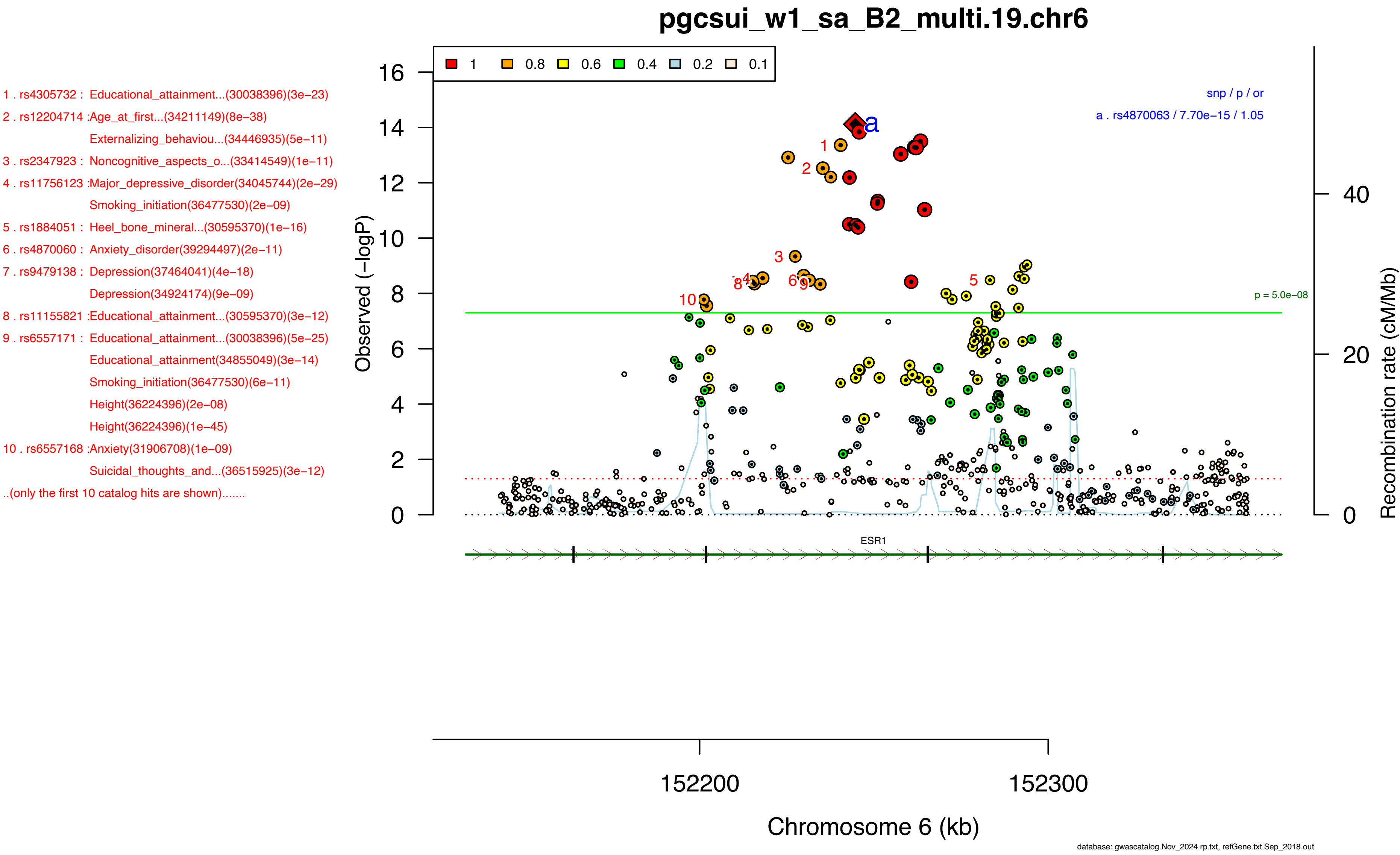

pgcsui_w1_sa_B2_multi.19.chr6
1 . rs4305732 : Educational_attainment...(30038396)(3e−23)
2 . rs12204714 :Age_at_first...(34211149)(8e−38)
Externalizing_behaviou...(34446935)(5e−11)
3 . rs2347923 : Noncognitive_aspects_o...(33414549)(1e−11)
4 . rs11756123 :Major_depressive_disorder(34045744)(2e−29)
Smoking_initiation(36477530)(2e−09)
5 . rs1884051 : Heel_bone_mineral...(30595370)(1e−16)
6 . rs4870060 : Anxiety_disorder(39294497)(2e−11)
7 . rs9479138 : Depression(37464041)(4e−18)
Depression(34924174)(9e−09)
8 . rs11155821 :Educational_attainment...(30595370)(3e−12)
9 . rs6557171 : Educational_attainment...(30038396)(5e−25)
Educational_attainment(34855049)(3e−14)
Smoking_initiation(36477530)(6e−11)
Height(36224396)(2e−08)
Height(36224396)(1e−45)
10 . rs6557168 :Anxiety(31906708)(1e−09)
Suicidal_thoughts_and...(36515925)(3e−12)
..(only the first 10 catalog hits are shown).......
1 0.8 0.6 0.4 0.2 0.1
snp / p / or
a . rs4870063 / 7.70e−15 / 1.05
p = 5.0e−08
Observed (−logP)
Recombination rate (cM/Mb)
ESR1
152200
152300
Chromosome 6 (kb)
database: gwascatalog.Nov_2024.rp.txt, refGene.txt.Sep_2018.out

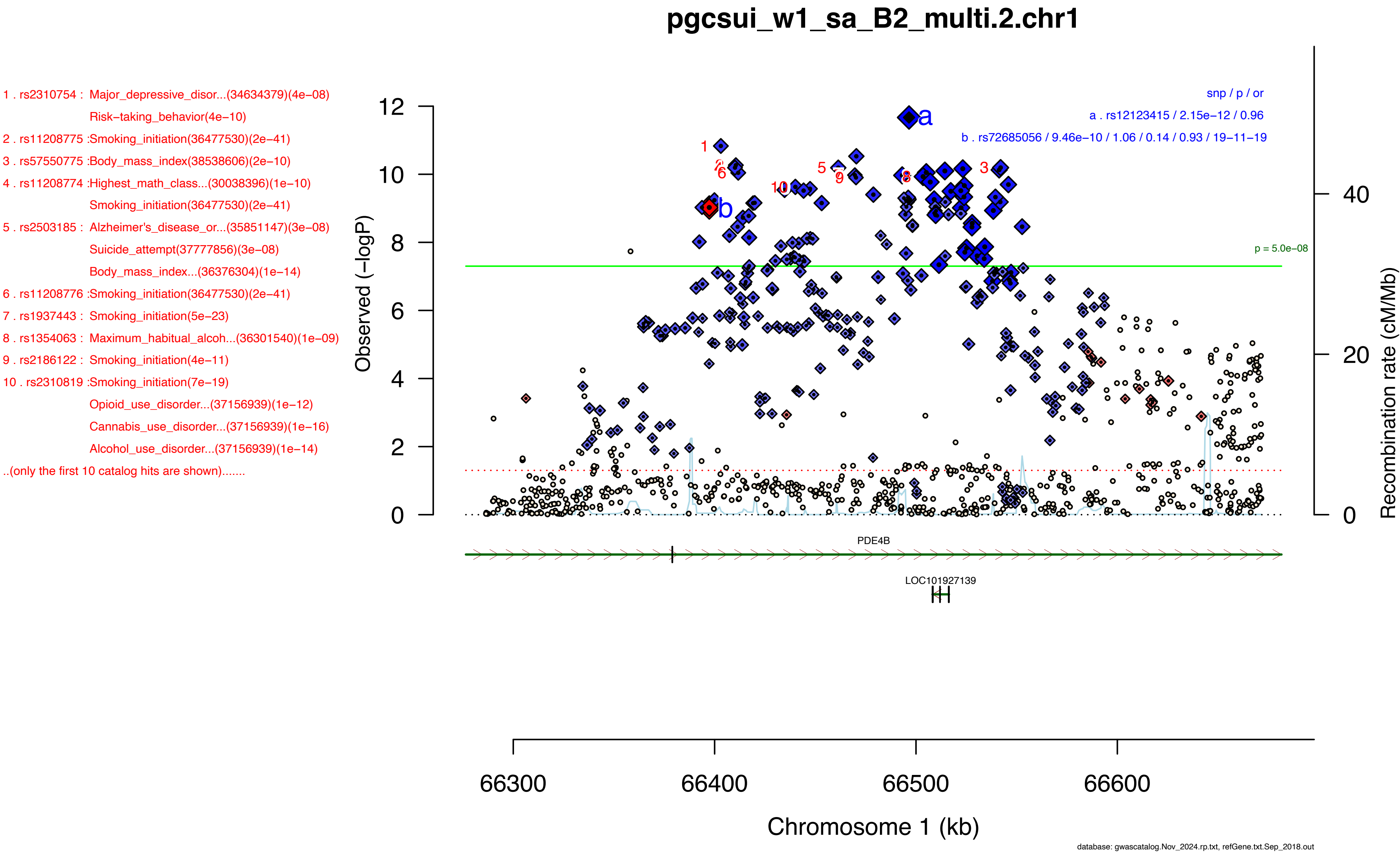
pgcsui_w1_sa_B2_multi.2.chr1
1 . rs2310754 : Major_depressive_disor...(34634379)(4e–08)
Risk–taking_behavior(4e–10)
2 . rs11208775 :Smoking_initiation(36477530)(2e–41)
3 . rs57550775 :Body_mass_index(38538606)(2e–10)
4 . rs11208774 :Highest_math_class...(30038396)(1e–10)
Smoking_initiation(36477530)(2e–41)
5 . rs2503185 : Alzheimer's_disease_or...(35851147)(3e–08)
Suicide_attempt(37777856)(3e–08)
Body_mass_index...(36376304)(1e–14)
6 . rs11208776 :Smoking_initiation(36477530)(2e–41)
7 . rs1937443 : Smoking_initiation(5e–23)
8 . rs1354063 : Maximum_habitual_alcoh...(36301540)(1e–09)
9 . rs2186122 : Smoking_initiation(4e–11)
10 . rs2310819 :Smoking_initiation(7e–19)
Opioid_use_disorder...(37156939)(1e–12)
Cannabis_use_disorder...(37156939)(1e–16)
Alcohol_use_disorder...(37156939)(1e–14)
..(only the first 10 catalog hits are shown).......
snp / p / or
a . rs12123415 / 2.15e–12 / 0.96
b . rs72685056 / 9.46e–10 / 1.06 / 0.14 / 0.93 / 19–11–19
p = 5.0e–08
Observed (–logP)
Recombination rate (cM/Mb)
PDE4B
LOC101927139
66300
66400
66500
66600
Chromosome 1 (kb)
database: gwascatalog.Nov_2024.rp.txt, refGene.txt.Sep_2018.out

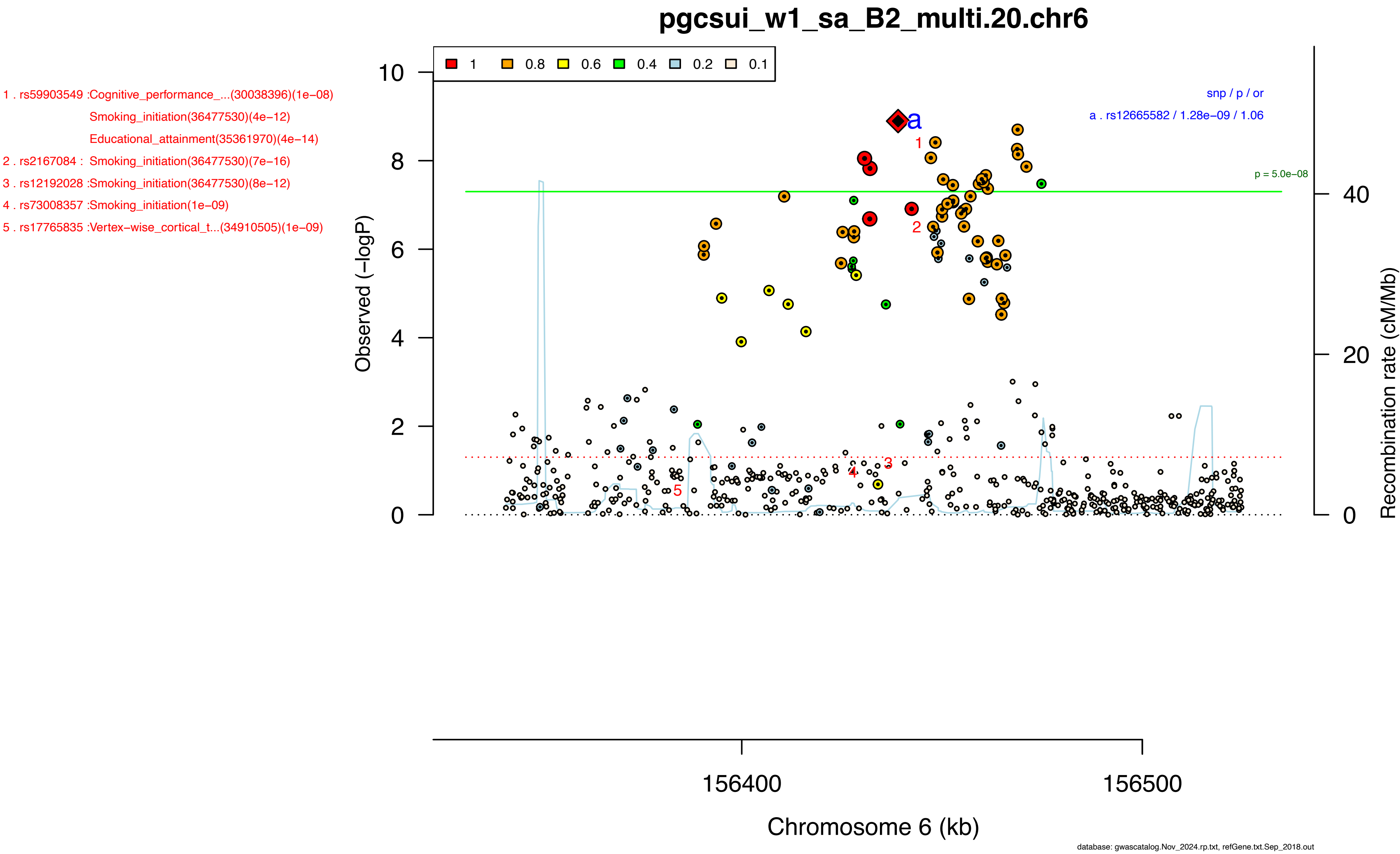

pgcsui_w1_sa_B2_multi.20.chr6
1 . rs59903549 :Cognitive_performance_...(30038396)(1e−08)
Smoking_initiation(36477530)(4e−12)
Educational_attainment(35361970)(4e−14)
2 . rs2167084 : Smoking_initiation(36477530)(7e−16)
3 . rs12192028 :Smoking_initiation(36477530)(8e−12)
4 . rs73008357 :Smoking_initiation(1e−09)
5 . rs17765835 :Vertex−wise_cortical_t...(34910505)(1e−09)
1 0.8 0.6 0.4 0.2 0.1
snp / p / or
a . rs12665582 / 1.28e−09 / 1.06
p = 5.0e−08
Observed (−logP)
Recombination rate (cM/Mb)
156400
156500
Chromosome 6 (kb)
database: gwascatalog.Nov_2024.rp.txt, refGene.txt.Sep_2018.out

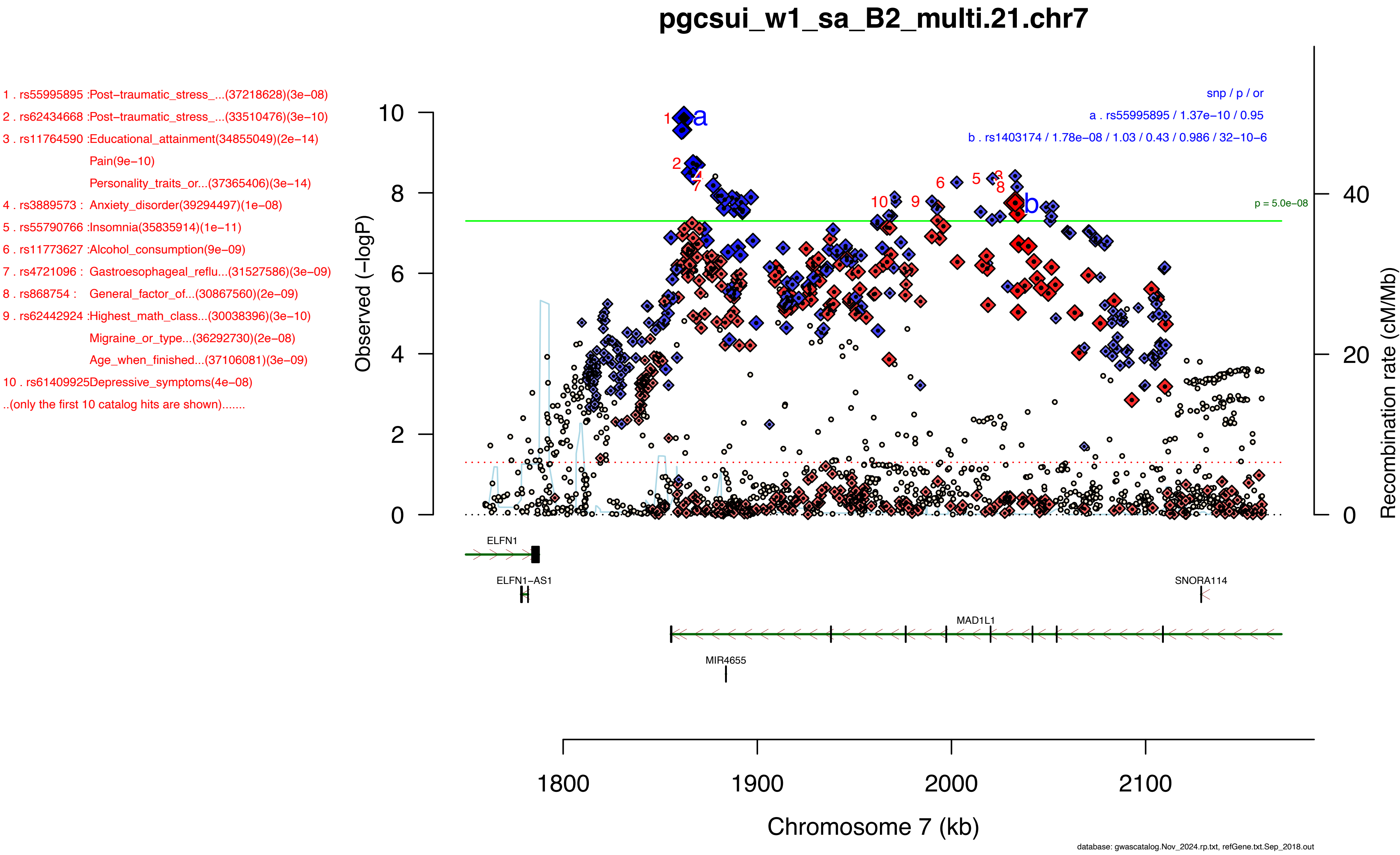
pgcsui_w1_sa_B2_multi.21.chr7
1 . rs55995895 :Post−traumatic_stress_...(37218628)(3e−08)
2 . rs62434668 :Post−traumatic_stress_...(33510476)(3e−10)
3 . rs11764590 :Educational_attainment(34855049)(2e−14)
Pain(9e−10)
Personality_traits_or...(37365406)(3e−14)
4 . rs3889573 : Anxiety_disorder(39294497)(1e−08)
5 . rs55790766 :Insomnia(35835914)(1e−11)
6 . rs11773627 :Alcohol_consumption(9e−09)
7 . rs4721096 : Gastroesophageal_reflu...(31527586)(3e−09)
8 . rs868754 : General_factor_of...(30867560)(2e−09)
9 . rs62442924 :Highest_math_class...(30038396)(3e−10)
Migraine_or_type...(36292730)(2e−08)
Age_when_finished...(37106081)(3e−09)
10 . rs61409925Depressive_symptoms(4e−08)
..(only the first 10 catalog hits are shown).......
snp / p / or
a . rs55995895 / 1.37e−10 / 0.95
b . rs1403174 / 1.78e−08 / 1.03 / 0.43 / 0.986 / 32−10−6
p = 5.0e−08
Observed (−logP)
Recombination rate (cM/Mb)
ELFN1
ELFN1−AS1
SNORA114
MAD1L1
MIR4655
Chromosome 7 (kb)
database: gwascatalog.Nov_2024.rp.txt, refGene.txt.Sep_2018.out

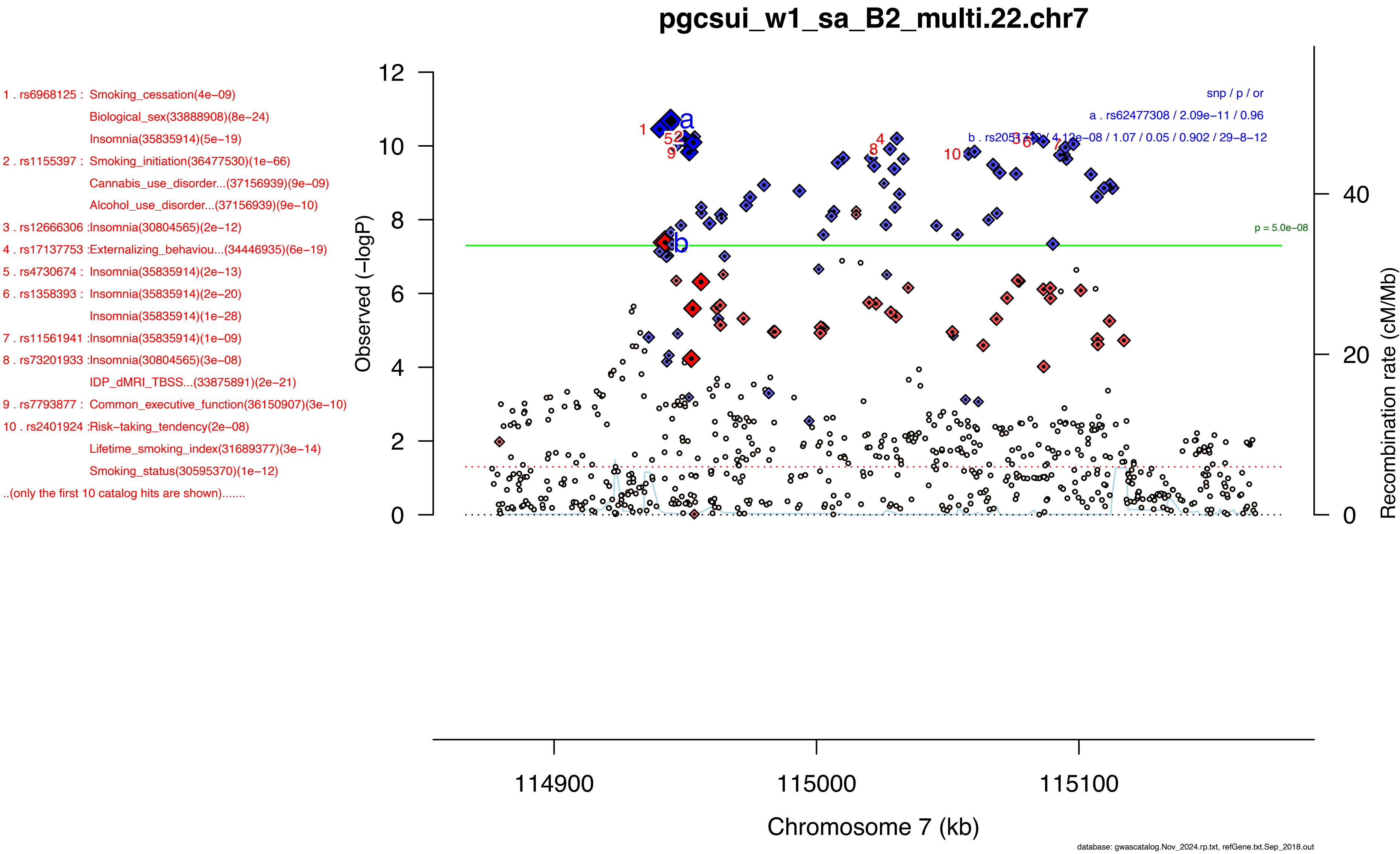

pgcsui_w1_sa_B2_multi.22.chr7
1 . rs6968125 : Smoking_cessation(4e−09)
Biological_sex(33888908)(8e−24)
Insomnia(35835914)(5e−19)
2 . rs1155397 : Smoking_initiation(36477530)(1e−66)
Cannabis_use_disorder...(37156939)(9e−09)
Alcohol_use_disorder...(37156939)(9e−10)
3 . rs12666306 :Insomnia(30804565)(2e−12)
4 . rs17137753 :Externalizing_behaviou...(34446935)(6e−19)
5 . rs4730674 : Insomnia(35835914)(2e−13)
6 . rs1358393 : Insomnia(35835914)(2e−20)
Insomnia(35835914)(1e−28)
7 . rs11561941 :Insomnia(35835914)(1e−09)
8 . rs73201933 :Insomnia(30804565)(3e−08)
IDP_dMRI_TBSS...(33875891)(2e−21)
9 . rs7793877 : Common_executive_function(36150907)(3e−10)
10 . rs2401924 :Risk−taking_tendency(2e−08)
Lifetime_smoking_index(31689377)(3e−14)
Smoking_status(30595370)(1e−12)
..(only the first 10 catalog hits are shown).......
snp / p / or
a . rs62477308 / 2.09e−11 / 0.96
b . rs2051710 / 4.12e−08 / 1.07 / 0.05 / 0.902 / 29−8−12
p = 5.0e−08
Observed (−logP)
Recombination rate (cM/Mb)
Chromosome 7 (kb)
114900
115000
115100
database: gwascatalog.Nov_2024.rp.txt, refGene.txt.Sep_2018.out

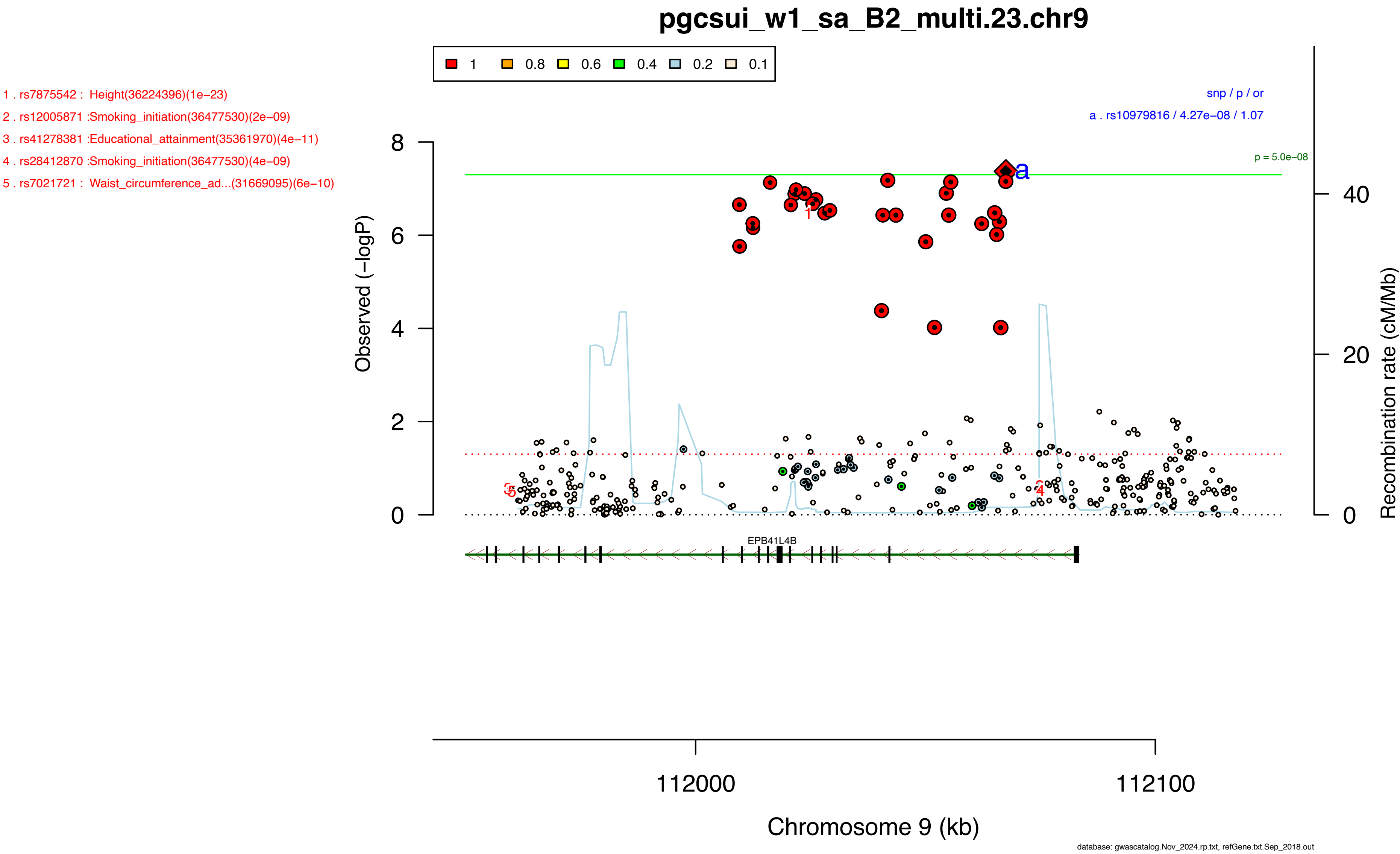

pgcsui_w1_sa_B2_multi.23.chr9
1 0.8 0.6 0.4 0.2 0.1
1 . rs7875542 : Height(36224396)(1e−23)
2 . rs12005871 :Smoking_initiation(36477530)(2e−09)
3 . rs41278381 :Educational_attainment(35361970)(4e−11)
4 . rs28412870 :Smoking_initiation(36477530)(4e−09)
5 . rs7021721 : Waist_circumference_ad...(31669095)(6e−10)
snp / p / or
a . rs10979816 / 4.27e−08 / 1.07
p = 5.0e−08
Observed (−logP)
Recombination rate (cM/Mb)
EPB41L4B
112000
112100
Chromosome 9 (kb)
database: gwascatalog.Nov_2024.rp.txt, refGene.txt.Sep_2018.out

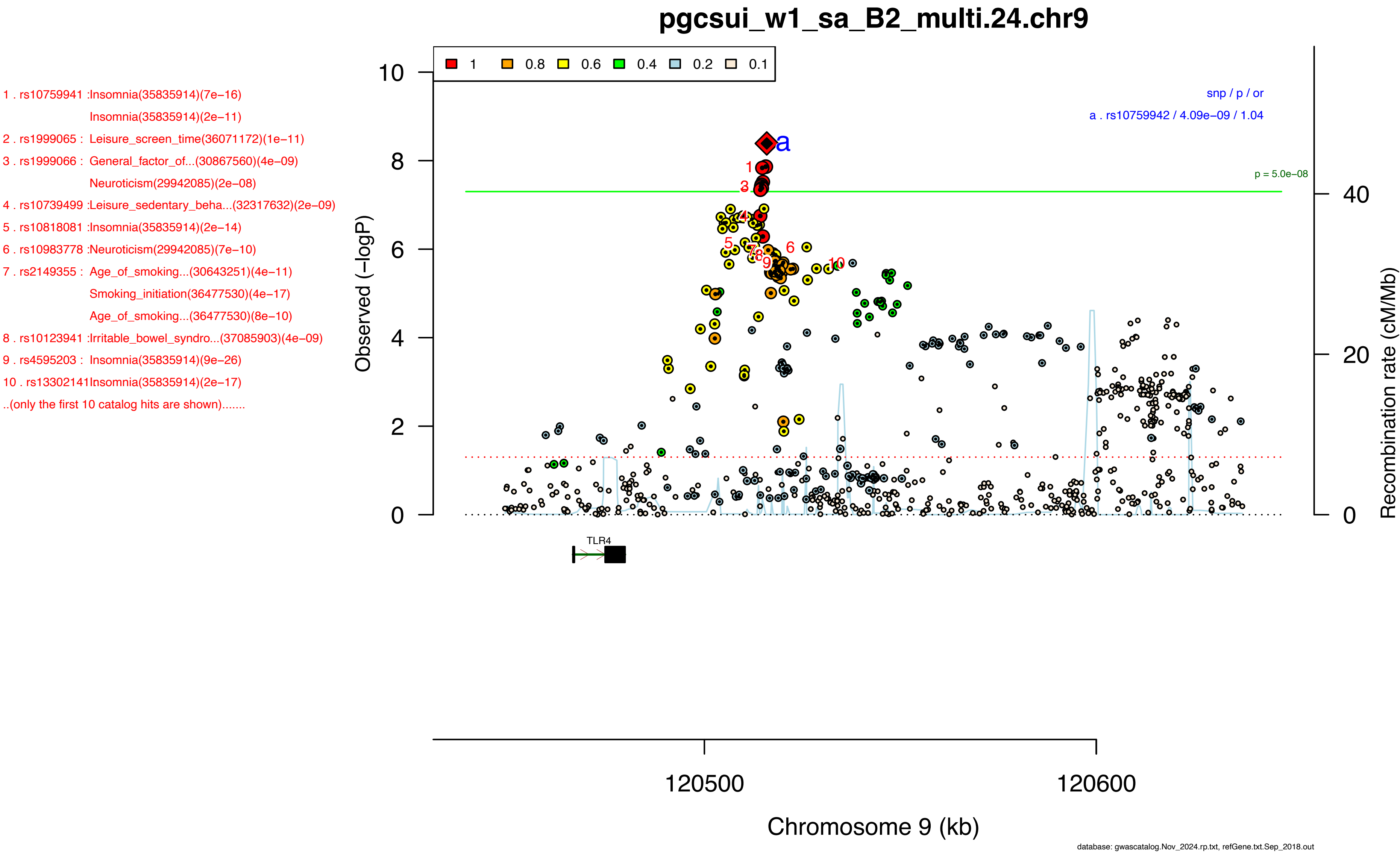
pgcsui_w1_sa_B2_multi.24.chr9
1 . rs10759941 :Insomnia(35835914)(7e−16)
Insomnia(35835914)(2e−11)
2 . rs1999065 : Leisure_screen_time(36071172)(1e−11)
3 . rs1999066 : General_factor_of...(30867560)(4e−09)
Neuroticism(29942085)(2e−08)
4 . rs10739499 :Leisure_sedentary_beha...(32317632)(2e−09)
5 . rs10818081 :Insomnia(35835914)(2e−14)
6 . rs10983778 :Neuroticism(29942085)(7e−10)
7 . rs2149355 : Age_of_smoking...(30643251)(4e−11)
Smoking_initiation(36477530)(4e−17)
Age_of_smoking...(36477530)(8e−10)
8 . rs10123941 :Irritable_bowel_syndro...(37085903)(4e−09)
9 . rs4595203 : Insomnia(35835914)(9e−26)
10 . rs13302141Insomnia(35835914)(2e−17)
..(only the first 10 catalog hits are shown).......
1 0.8 0.6 0.4 0.2 0.1
snp / p / or
a . rs10759942 / 4.09e−09 / 1.04
p = 5.0e−08
Observed (−logP)
Recombination rate (cM/Mb)
TLR4
120500
120600
Chromosome 9 (kb)
database: gwascatalog.Nov_2024.rp.txt, refGene.txt.Sep_2018.out

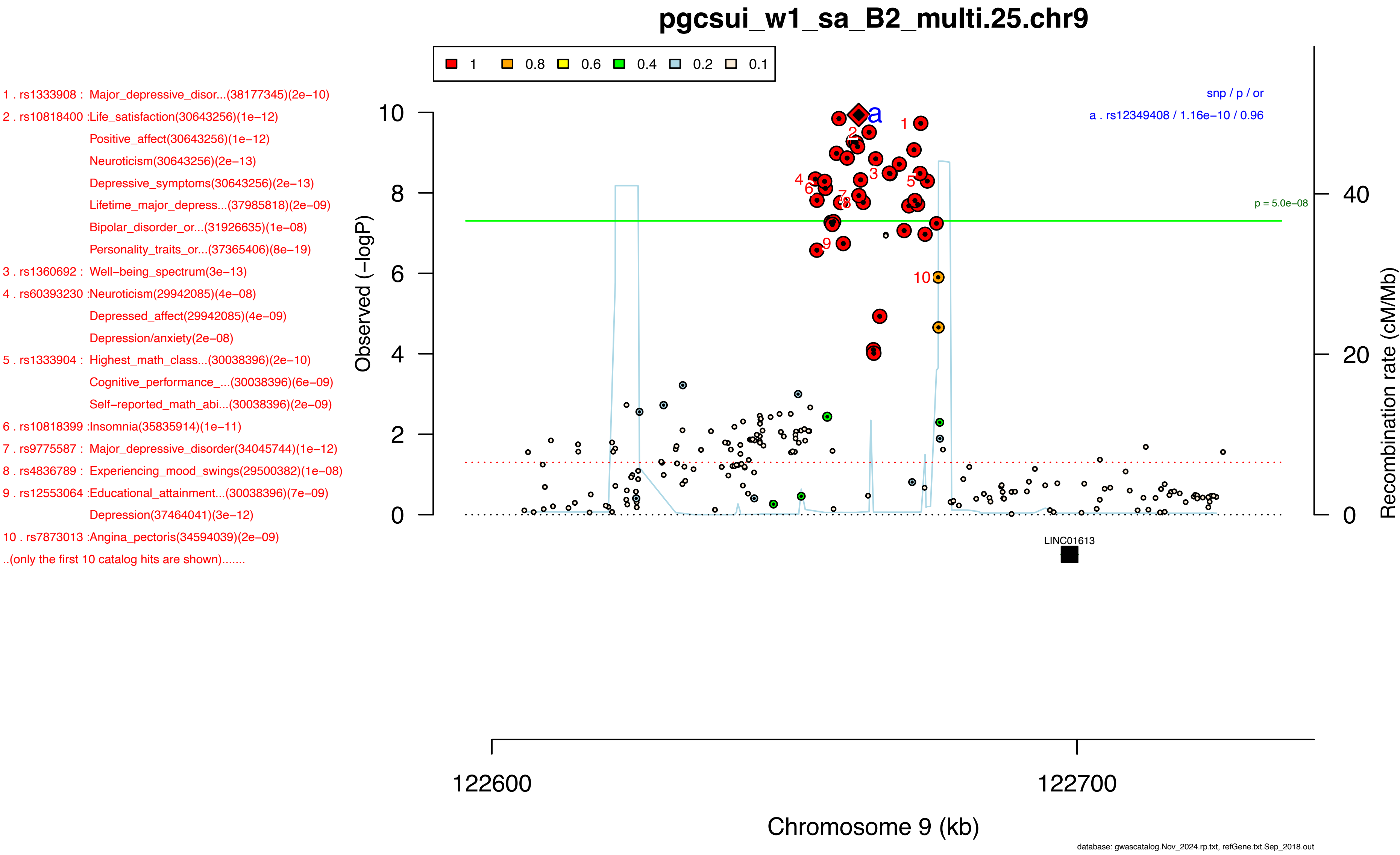

pgcsui_w1_sa_B2_multi.25.chr9
1 0.8 0.6 0.4 0.2 0.1
1 . rs1333908 : Major_depressive_disor...(38177345)(2e−10)
2 . rs10818400 :Life_satisfaction(30643256)(1e−12)
Positive_affect(30643256)(1e−12)
Neuroticism(30643256)(2e−13)
Depressive_symptoms(30643256)(2e−13)
Lifetime_major_depress...(37985818)(2e−09)
Bipolar_disorder_or...(31926635)(1e−08)
Personality_traits_or...(37365406)(8e−19)
3 . rs1360692 : Well−being_spectrum(3e−13)
4 . rs60393230 :Neuroticism(29942085)(4e−08)
Depressed_affect(29942085)(4e−09)
Depression/anxiety(2e−08)
5 . rs1333904 : Highest_math_class...(30038396)(2e−10)
Cognitive_performance_...(30038396)(6e−09)
Self−reported_math_abi...(30038396)(2e−09)
6 . rs10818399 :Insomnia(35835914)(1e−11)
7 . rs9775587 : Major_depressive_disorder(34045744)(1e−12)
8 . rs4836789 : Experiencing_mood_swings(29500382)(1e−08)
9 . rs12553064 :Educational_attainment...(30038396)(7e−09)
Depression(37464041)(3e−12)
10 . rs7873013 :Angina_pectoris(34594039)(2e−09)
..(only the first 10 catalog hits are shown).......
snp / p / or
a . rs12349408 / 1.16e−10 / 0.96
p = 5.0e−08
Observed (−logP)
Recombination rate (cM/Mb)
LINC01613
122600
122700
Chromosome 9 (kb)
database: gwascatalog.Nov_2024.rp.txt, refGene.txt.Sep_2018.out

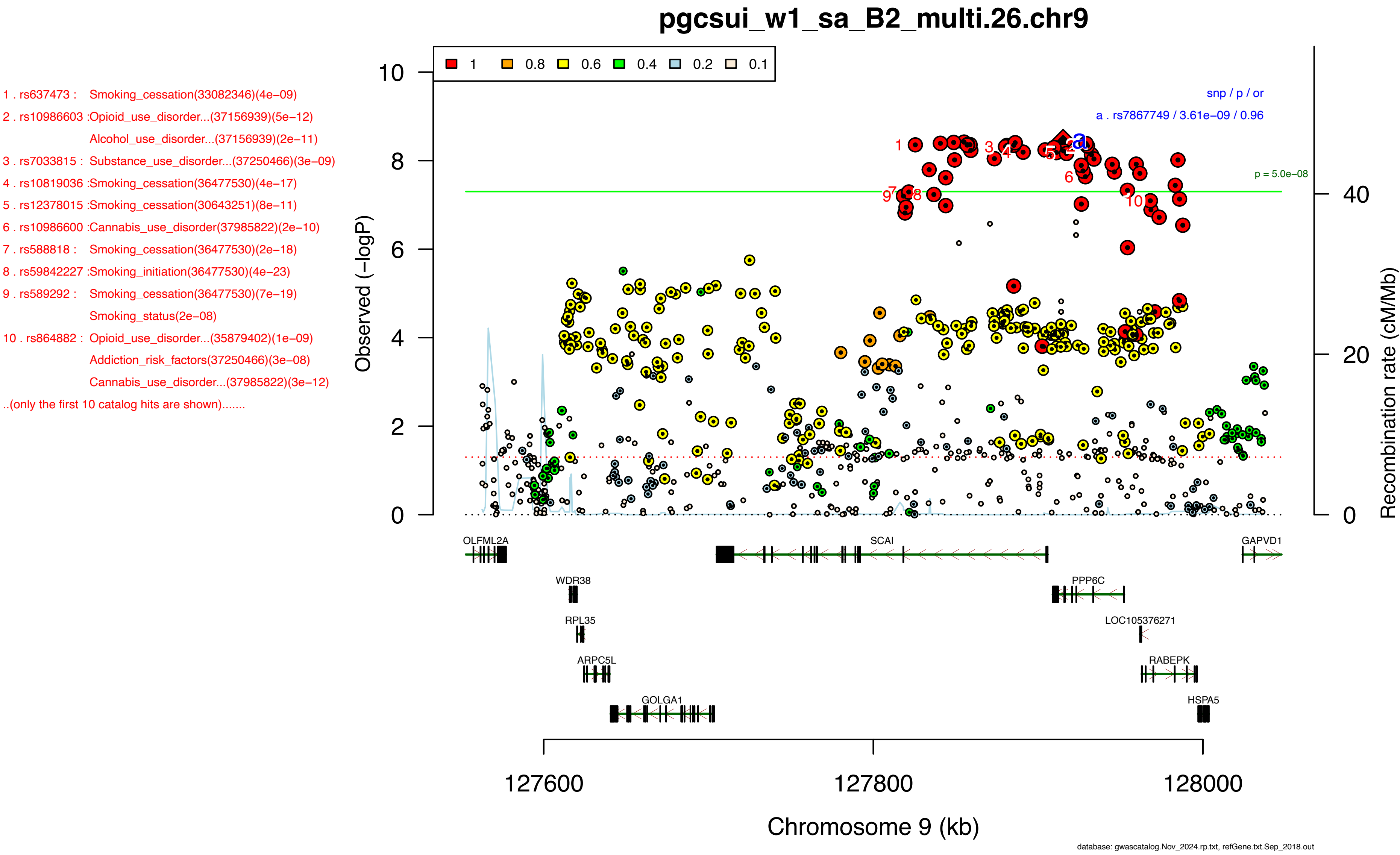

pgcsui_w1_sa_B2_multi.26.chr9
1 . rs637473 : Smoking_cessation(33082346)(4e−09)
2 . rs10986603 :Opioid_use_disorder...(37156939)(5e−12)
Alcohol_use_disorder...(37156939)(2e−11)
3 . rs7033815 : Substance_use_disorder...(37250466)(3e−09)
4 . rs10819036 :Smoking_cessation(36477530)(4e−17)
5 . rs12378015 :Smoking_cessation(30643251)(8e−11)
6 . rs10986600 :Cannabis_use_disorder(37985822)(2e−10)
7 . rs588818 : Smoking_cessation(36477530)(2e−18)
8 . rs59842227 :Smoking_initiation(36477530)(4e−23)
9 . rs589292 : Smoking_cessation(36477530)(7e−19)
Smoking_status(2e−08)
10 . rs864882 : Opioid_use_disorder...(35879402)(1e−09)
Addiction_risk_factors(37250466)(3e−08)
Cannabis_use_disorder...(37985822)(3e−12)
..(only the first 10 catalog hits are shown).......
1 0.8 0.6 0.4 0.2 0.1
snp / p / or
a . rs7867749 / 3.61e−09 / 0.96
p = 5.0e−08
Observed (−logP)
Recombination rate (cM/Mb)
OLFML2A
SCAI
GAPVD1
WDR38
PPP6C
RPL35
LOC105376271
ARPC5L
RABEPK
GOLGA1
HSPA5
127600
127800
128000
Chromosome 9 (kb)
database: gwascatalog.Nov_2024.rp.txt, refGene.txt.Sep_2018.out

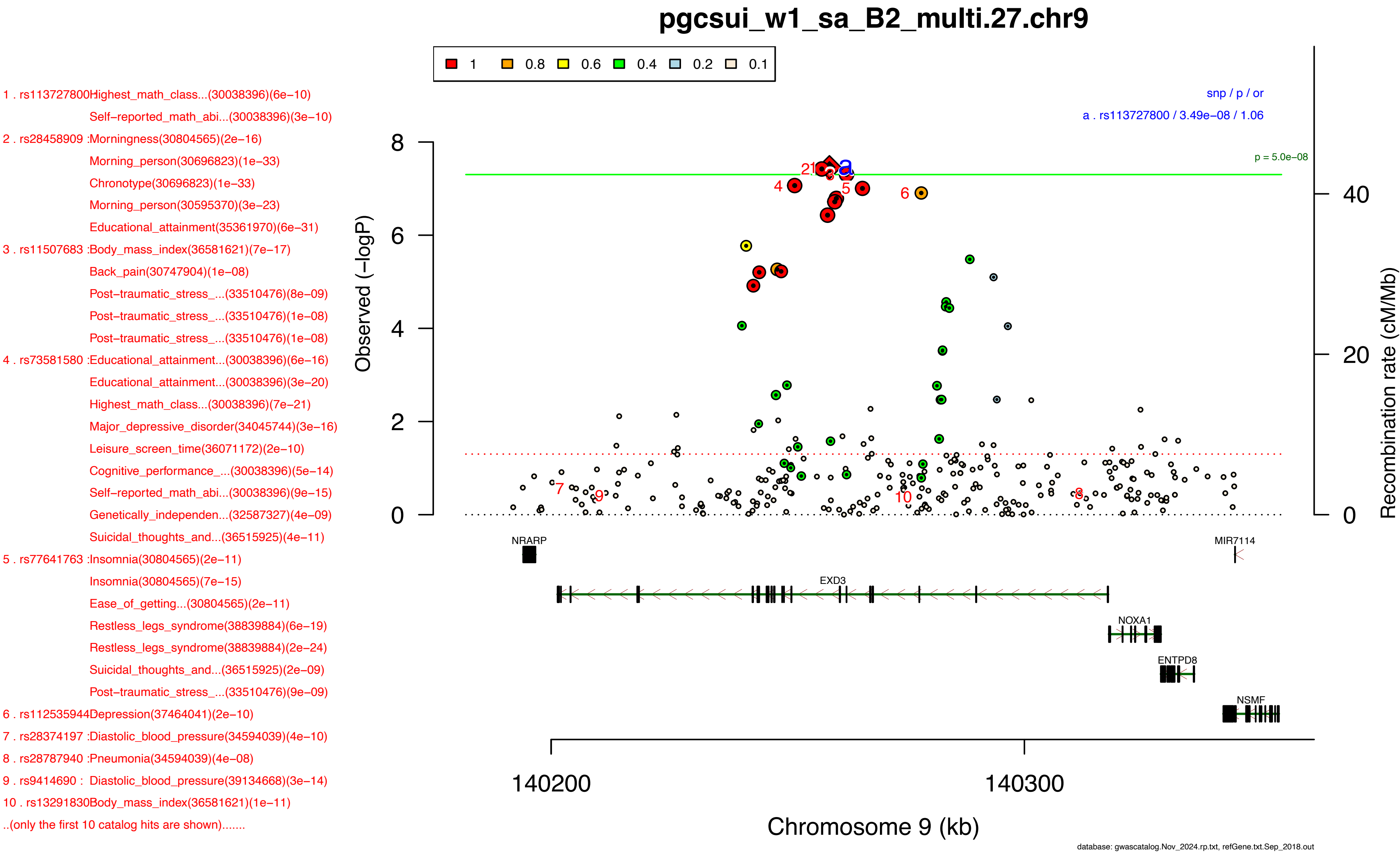
pgcsui_w1_sa_B2_multi.27.chr9
1 . rs113727800Highest_math_class...(30038396)(6e−10)
Self−reported_math_abi...(30038396)(3e−10)
2 . rs28458909 :Morningness(30804565)(2e−16)
Morning_person(30696823)(1e−33)
Chronotype(30696823)(1e−33)
Morning_person(30595370)(3e−23)
Educational_attainment(35361970)(6e−31)
3 . rs11507683 :Body_mass_index(36581621)(7e−17)
Back_pain(30747904)(1e−08)
Post−traumatic_stress_...(33510476)(8e−09)
Post−traumatic_stress_...(33510476)(1e−08)
Post−traumatic_stress_...(33510476)(1e−08)
4 . rs73581580 :Educational_attainment...(30038396)(6e−16)
Educational_attainment...(30038396)(3e−20)
Highest_math_class...(30038396)(7e−21)
Major_depressive_disorder(34045744)(3e−16)
Leisure_screen_time(36071172)(2e−10)
Cognitive_performance_...(30038396)(5e−14)
Self−reported_math_abi...(30038396)(9e−15)
Genetically_independen...(32587327)(4e−09)
Suicidal_thoughts_and...(36515925)(4e−11)
5 . rs77641763 :Insomnia(30804565)(2e−11)
Insomnia(30804565)(7e−15)
Ease_of_getting...(30804565)(2e−11)
Restless_legs_syndrome(38839884)(6e−19)
Restless_legs_syndrome(38839884)(2e−24)
Suicidal_thoughts_and...(36515925)(2e−09)
Post−traumatic_stress_...(33510476)(9e−09)
6 . rs112535944Depression(37464041)(2e−10)
7 . rs28374197 :Diastolic_blood_pressure(34594039)(4e−10)
8 . rs28787940 :Pneumonia(34594039)(4e−08)
9 . rs9414690 : Diastolic_blood_pressure(39134668)(3e−14)
10 . rs13291830Body_mass_index(36581621)(1e−11)
..(only the first 10 catalog hits are shown).......
1 0.8 0.6 0.4 0.2 0.1
snp / p / or
a . rs113727800 / 3.49e−08 / 1.06
p = 5.0e−08
Observed (−logP)
Recombination rate (cM/Mb)
NRARP
MIR7114
EXD3
NOXA1
ENTPD8
NSMF
140200
140300
Chromosome 9 (kb)
database: gwascatalog.Nov_2024.rp.txt, refGene.txt.Sep_2018.out

1 . rs10835372 :Cannabis_use_disorder...(37156939)(2e−09)
Alcohol_use_disorder...(37156939)(8e−09)
Smoking_initiation(6e−12)
Cannabis_use_disorder...(37985822)(2e−10)
2 . rs7931440 : Primary_open_angle...(37386247)(1e−09)
Disruptive_behavior(5e−09)
3 . rs11030393 :Height(36224396)(3e−31)
4 . rs4267058 : Body_size_at...(32376654)(1e−09)
5 . rs11030381 :Osteoarthritis(36411363)(7e−09)
Osteoarthritis(36411363)(8e−10)
6 . rs11030380 :Depression(37464041)(1e−14)
7 . rs7127383 : Cigarettes_smoked_per...(30643251)(2e−08)
Smoking_initiation(36477530)(2e−34)
Suicidal_thoughts_and...(36515925)(1e−08)
8 . rs2585817 : Smoking_status(5e−13)
Smoking_initiation(33082346)(5e−09)
Smoking_cessation(36477530)(1e−15)
Attention_deficit_hype...(38565336)(5e−15)
9 . rs7128734 : Smoking_cessation(36477530)(2e−16)
10 . rs7952220 :Attention_deficit_hype...(32606422)(1e−08)
Smoking_cessation(36477530)(2e−16)
..(only the first 10 catalog hits are shown).......

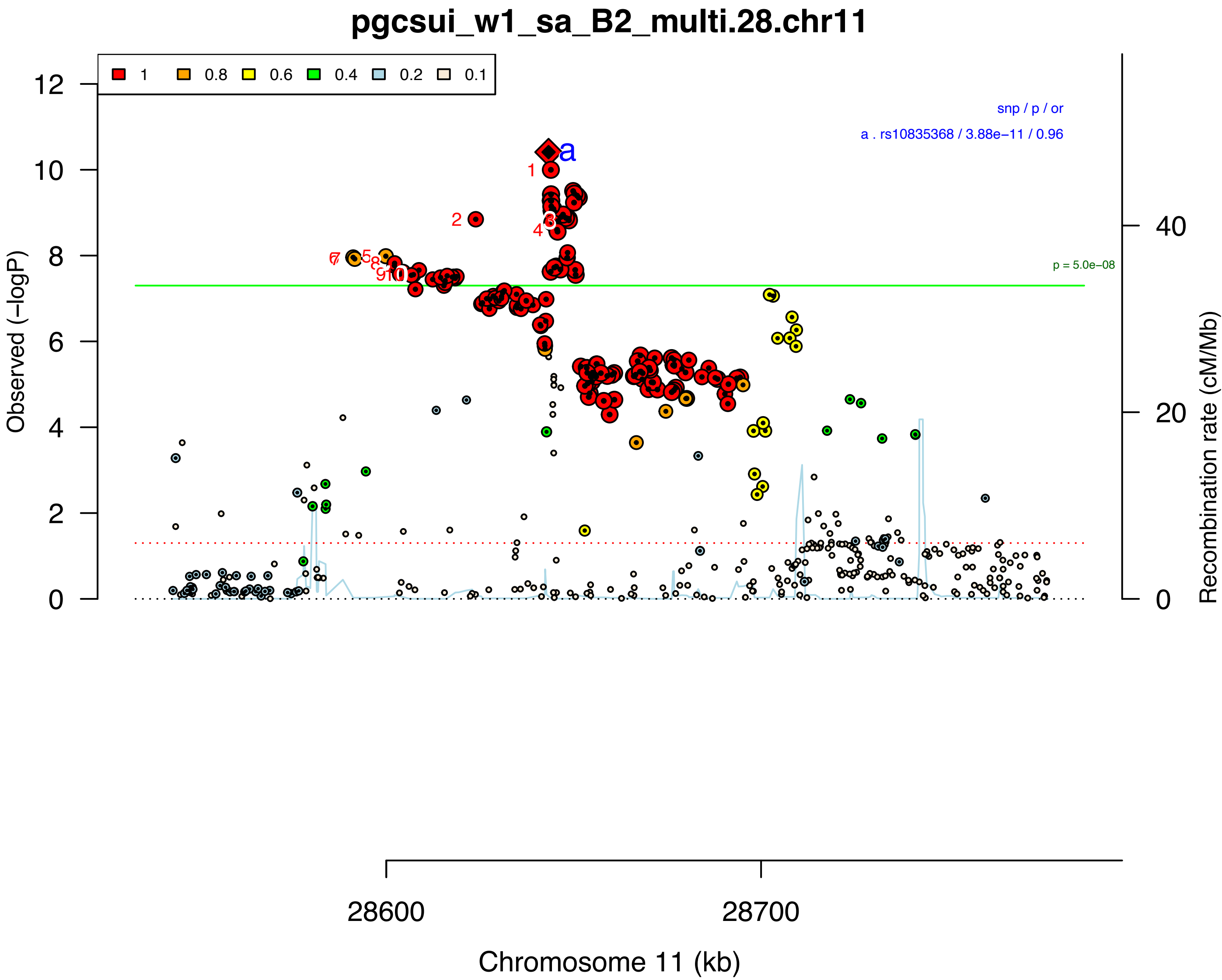


database: gwascatalog.Nov_2024.rp.txt, refGene.txt.Sep_2018.out

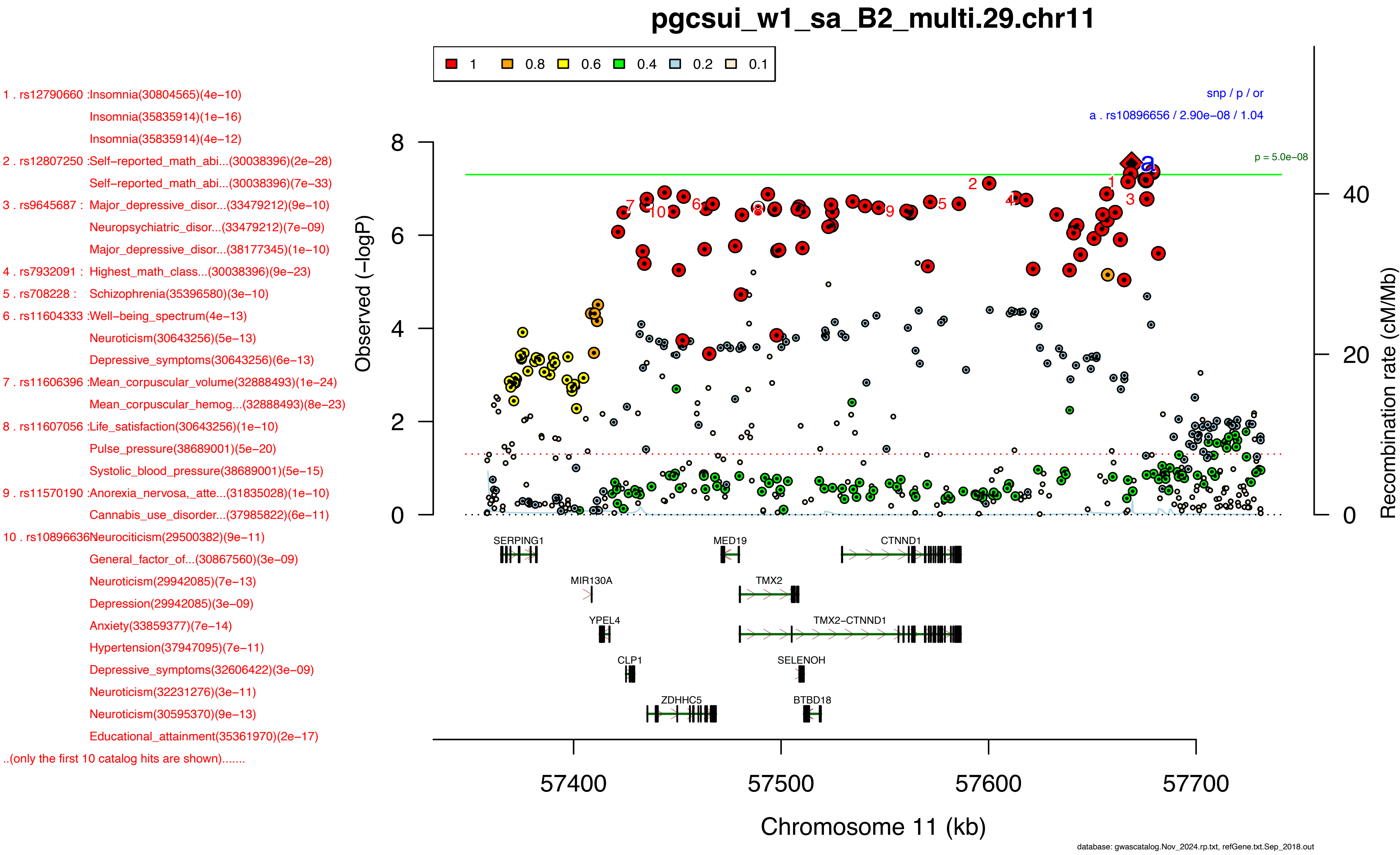
pgcsui_w1_sa_B2_multi.29.chr11
1
0.8
0.6
0.4
0.2
0.1
1 . rs12790660 :Insomnia(30804565)(4e−10)
Insomnia(35835914)(1e−16)
Insomnia(35835914)(4e−12)
2 . rs12807250 :Self−reported_math_abi...(30038396)(2e−28)
Self−reported_math_abi...(30038396)(7e−33)
3 . rs9645687 : Major_depressive_disor...(33479212)(9e−10)
Neuropsychiatric_disor...(33479212)(7e−09)
Major_depressive_disor...(38177345)(1e−10)
4 . rs7932091 : Highest_math_class...(30038396)(9e−23)
5 . rs708228 : Schizophrenia(35396580)(3e−10)
6 . rs11604333 :Well−being_spectrum(4e−13)
Neuroticism(30643256)(5e−13)
Depressive_symptoms(30643256)(6e−13)
7 . rs11606396 :Mean_corpuscular_volume(32888493)(1e−24)
Mean_corpuscular_hemog...(32888493)(8e−23)
8 . rs11607056 :Life_satisfaction(30643256)(1e−10)
Pulse_pressure(38689001)(5e−20)
Systolic_blood_pressure(38689001)(5e−15)
9 . rs11570190 :Anorexia_nervosa,_atte...(31835028)(1e−10)
Cannabis_use_disorder...(37985822)(6e−11)
10 . rs10896636Neurociticism(29500382)(9e−11)
General_factor_of...(30867560)(3e−09)
Neuroticism(29942085)(7e−13)
Depression(29942085)(3e−09)
Anxiety(33859377)(7e−14)
Hypertension(37947095)(7e−11)
Depressive_symptoms(32606422)(3e−09)
Neuroticism(32231276)(3e−11)
Neuroticism(30595370)(9e−13)
Educational_attainment(35361970)(2e−17)
..(only the first 10 catalog hits are shown).......
snp / p / or
a . rs10896656 / 2.90e−08 / 1.04
p = 5.0e−08
Observed (−logP)
Recombination rate (cM/Mb)
SERPING1
MED19
CTNND1
MIR130A
TMX2
YPEL4
TMX2−CTNND1
CLP1
SELENOH
ZDHHC5
BTBD18
57400
57500
57600
57700
Chromosome 11 (kb)
database: gwascatalog.Nov_2024.rp.txt, refGene.txt.Sep_2018.out

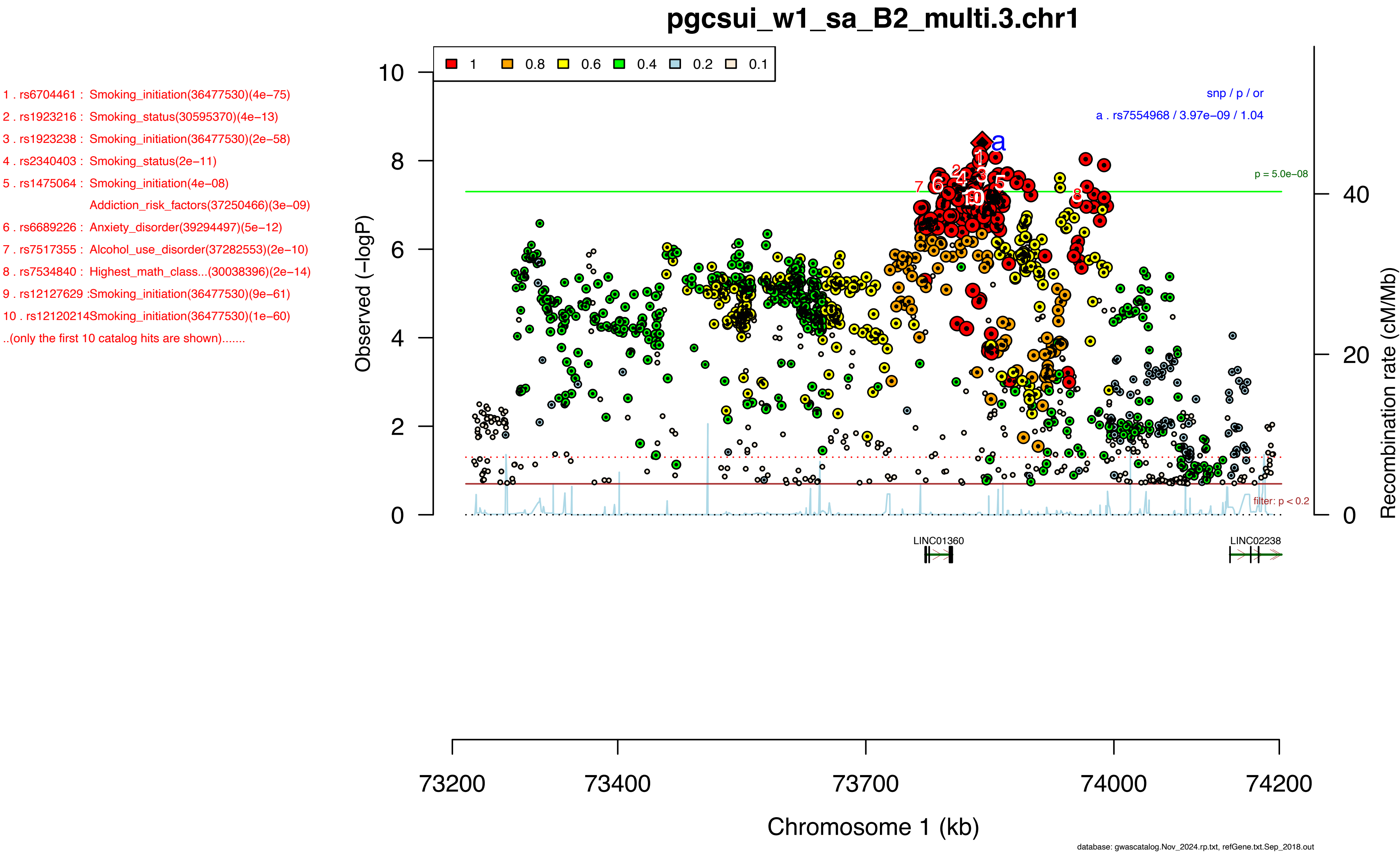
pgcsui_w1_sa_B2_multi.3.chr1
1 . rs6704461 : Smoking_initiation(36477530)(4e−75)
2 . rs1923216 : Smoking_status(30595370)(4e−13)
3 . rs1923238 : Smoking_initiation(36477530)(2e−58)
4 . rs2340403 : Smoking_status(2e−11)
5 . rs1475064 : Smoking_initiation(4e−08)
Addiction_risk_factors(37250466)(3e−09)
6 . rs6689226 : Anxiety_disorder(39294497)(5e−12)
7 . rs7517355 : Alcohol_use_disorder(37282553)(2e−10)
8 . rs7534840 : Highest_math_class...(30038396)(2e−14)
9 . rs12127629 :Smoking_initiation(36477530)(9e−61)
10 . rs12120214Smoking_initiation(36477530)(1e−60)
..(only the first 10 catalog hits are shown).......
1
0.8
0.6
0.4
0.2
0.1
snp / p / or
a . rs7554968 / 3.97e−09 / 1.04
p = 5.0e−08
filter: p < 0.2
Observed (−logP)
Recombination rate (cM/Mb)
LINC01360
LINC02238
73200
73400
73700
74000
74200
Chromosome 1 (kb)
database: gwascatalog.Nov_2024.rp.txt, refGene.txt.Sep_2018.out

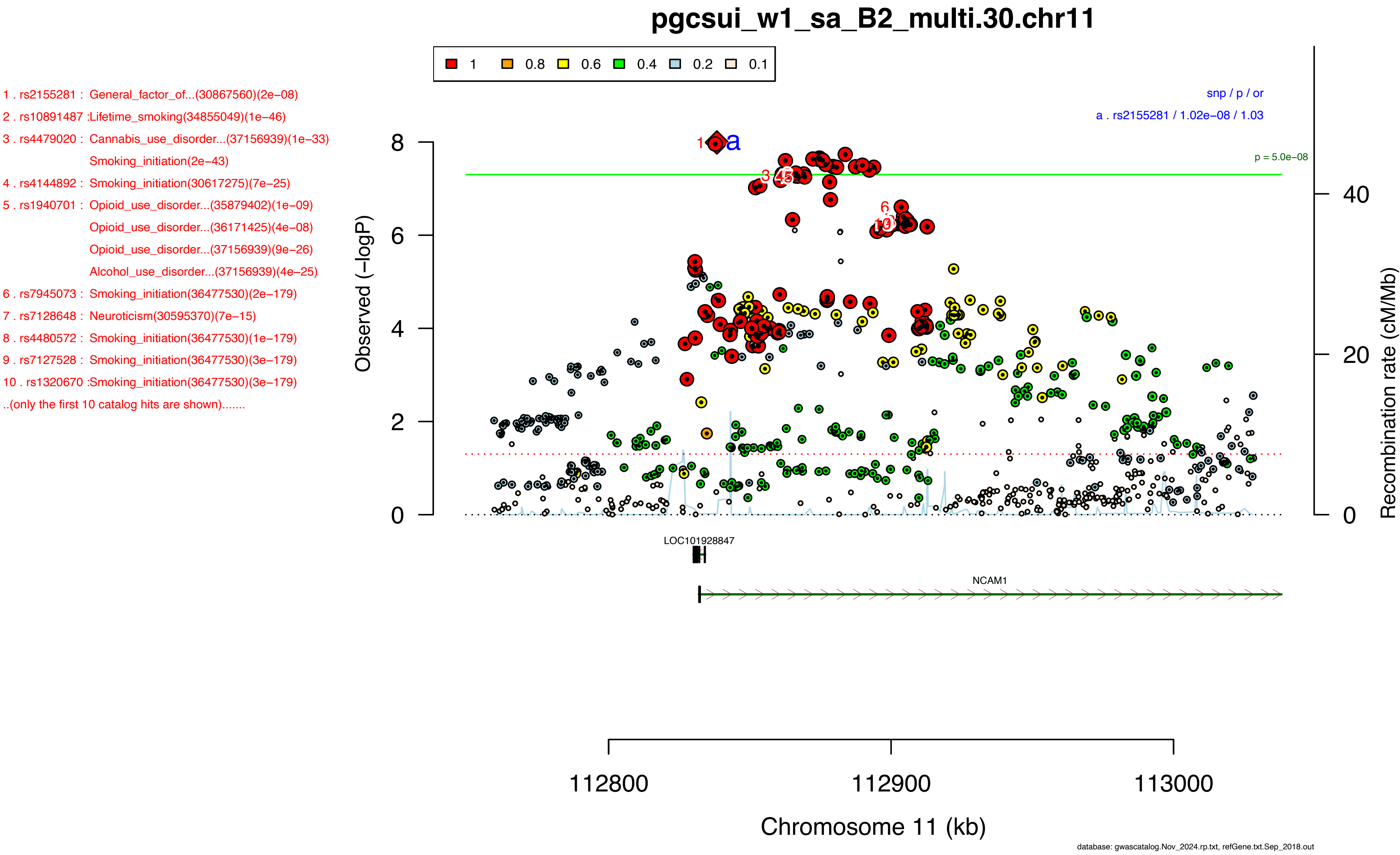

pgcsui_w1_sa_B2_multi.30.chr11
1 0.8 0.6 0.4 0.2 0.1
1 . rs2155281 : General_factor_of...(30867560)(2e−08)
2 . rs10891487 :Lifetime_smoking(34855049)(1e−46)
3 . rs4479020 : Cannabis_use_disorder...(37156939)(1e−33)
Smoking_initiation(2e−43)
4 . rs4144892 : Smoking_initiation(30617275)(7e−25)
5 . rs1940701 : Opioid_use_disorder...(35879402)(1e−09)
Opioid_use_disorder...(36171425)(4e−08)
Opioid_use_disorder...(37156939)(9e−26)
Alcohol_use_disorder...(37156939)(4e−25)
6 . rs7945073 : Smoking_initiation(36477530)(2e−179)
7 . rs7128648 : Neuroticism(30595370)(7e−15)
8 . rs4480572 : Smoking_initiation(36477530)(1e−179)
9 . rs7127528 : Smoking_initiation(36477530)(3e−179)
10 . rs1320670 :Smoking_initiation(36477530)(3e−179)
..(only the first 10 catalog hits are shown).......
snp / p / or
a . rs2155281 / 1.02e−08 / 1.03
p = 5.0e−08
Observed (−logP)
Recombination rate (cM/Mb)
LOC101928847
NCAM1
112800
112900
113000
Chromosome 11 (kb)
database: gwascatalog.Nov_2024.rp.txt, refGene.txt.Sep_2018.out

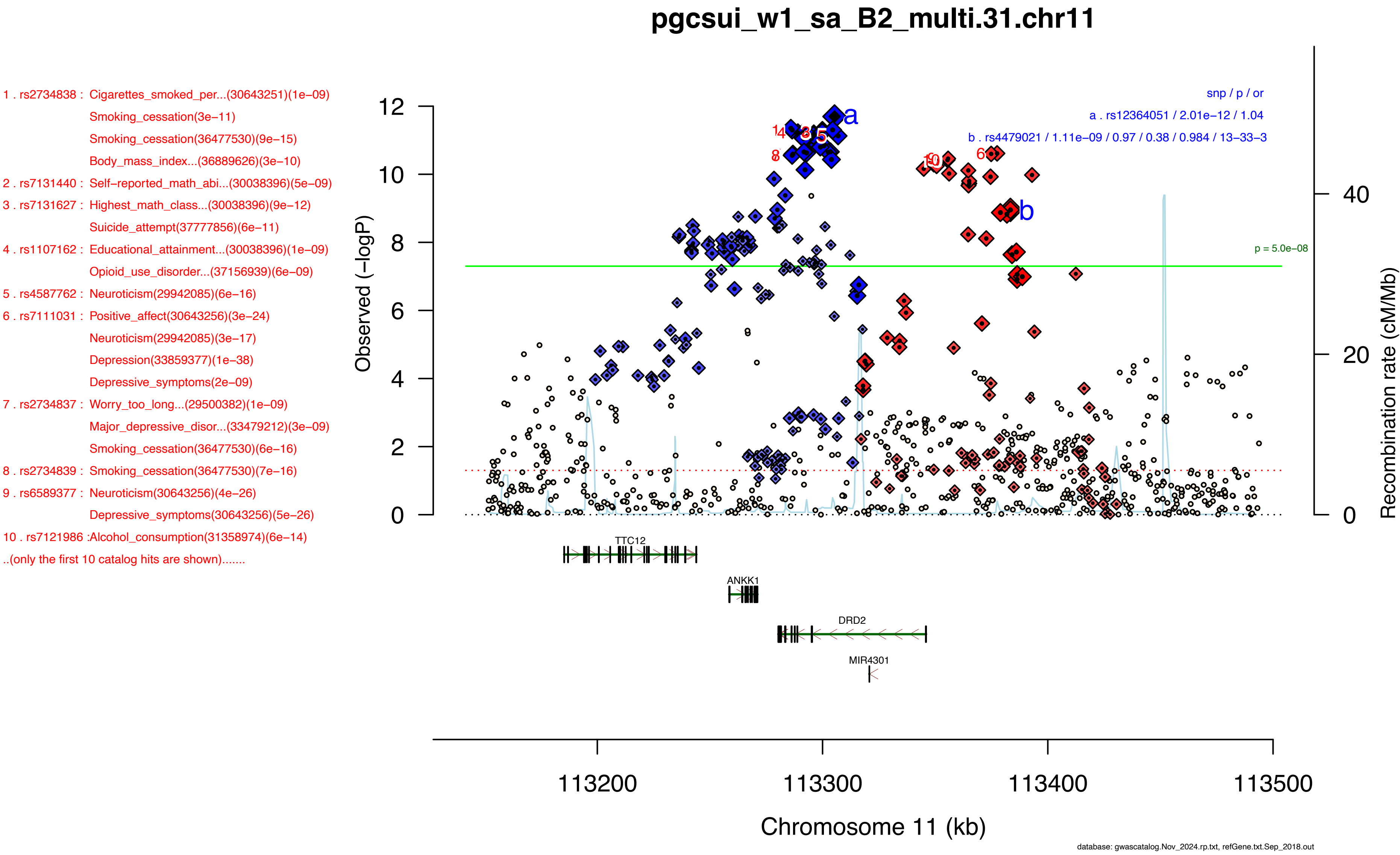
pgcsui_w1_sa_B2_multi.31.chr11
1 . rs2734838 : Cigarettes_smoked_per...(30643251)(1e−09)
Smoking_cessation(3e−11)
Smoking_cessation(36477530)(9e−15)
Body_mass_index...(36889626)(3e−10)
2 . rs7131440 : Self−reported_math_abi...(30038396)(5e−09)
3 . rs7131627 : Highest_math_class...(30038396)(9e−12)
Suicide_attempt(37777856)(6e−11)
4 . rs1107162 : Educational_attainment...(30038396)(1e−09)
Opioid_use_disorder...(37156939)(6e−09)
5 . rs4587762 : Neuroticism(29942085)(6e−16)
6 . rs7111031 : Positive_affect(30643256)(3e−24)
Neuroticism(29942085)(3e−17)
Depression(33859377)(1e−38)
Depressive_symptoms(2e−09)
7 . rs2734837 : Worry_too_long...(29500382)(1e−09)
Major_depressive_disor...(33479212)(3e−09)
Smoking_cessation(36477530)(6e−16)
8 . rs2734839 : Smoking_cessation(36477530)(7e−16)
9 . rs6589377 : Neuroticism(30643256)(4e−26)
Depressive_symptoms(30643256)(5e−26)
10 . rs7121986 :Alcohol_consumption(31358974)(6e−14)
..(only the first 10 catalog hits are shown).......
snp / p / or
a . rs12364051 / 2.01e−12 / 1.04
b . rs4479021 / 1.11e−09 / 0.97 / 0.38 / 0.984 / 13−33−3
p = 5.0e−08
Observed (−logP)
Recombination rate (cM/Mb)
TTC12
ANKK1
DRD2
MIR4301
113200
113300
113400
113500
Chromosome 11 (kb)
database: gwascatalog.Nov_2024.rp.txt, refGene.txt.Sep_2018.out

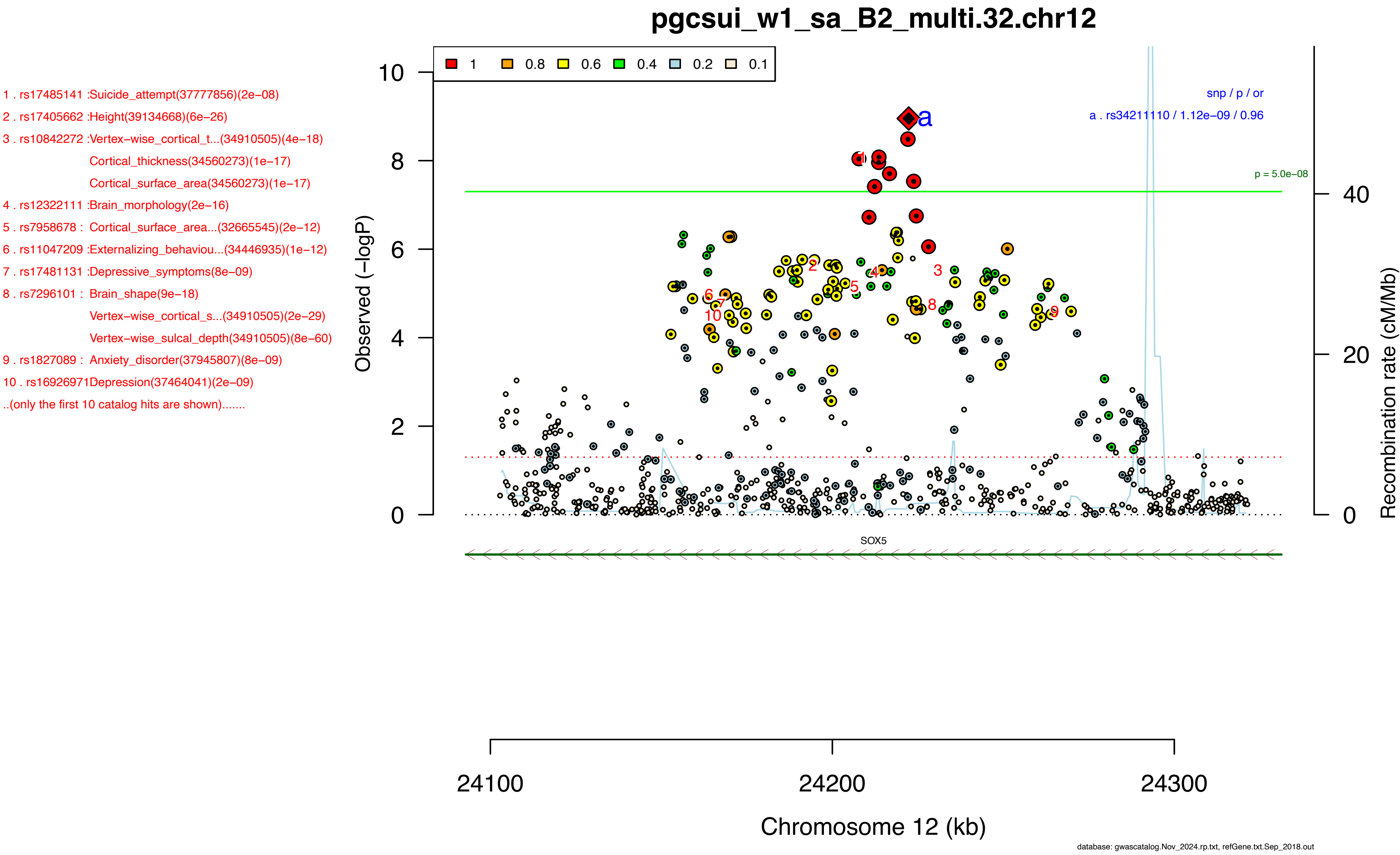
pgcsui_w1_sa_B2_multi.32.chr12
1 . rs17485141 :Suicide_attempt(37777856)(2e−08)
2 . rs17405662 :Height(39134668)(6e−26)
3 . rs10842272 :Vertex−wise_cortical_t...(34910505)(4e−18)
Cortical_thickness(34560273)(1e−17)
Cortical_surface_area(34560273)(1e−17)
4 . rs12322111 :Brain_morphology(2e−16)
5 . rs7958678 : Cortical_surface_area...(32665545)(2e−12)
6 . rs11047209 :Externalizing_behaviou...(34446935)(1e−12)
7 . rs17481131 :Depressive_symptoms(8e−09)
8 . rs7296101 : Brain_shape(9e−18)
Vertex−wise_cortical_s...(34910505)(2e−29)
Vertex−wise_sulcal_depth(34910505)(8e−60)
9 . rs1827089 : Anxiety_disorder(37945807)(8e−09)
10 . rs16926971Depression(37464041)(2e−09)
..(only the first 10 catalog hits are shown).......
1 0.8 0.6 0.4 0.2 0.1
snp / p / or
a . rs34211110 / 1.12e−09 / 0.96
p = 5.0e−08
Observed (−logP)
Recombination rate (cM/Mb)
SOX5
24100 24200 24300
Chromosome 12 (kb)
database: gwascatalog.Nov_2024.rp.txt, refGene.txt.Sep_2018.out

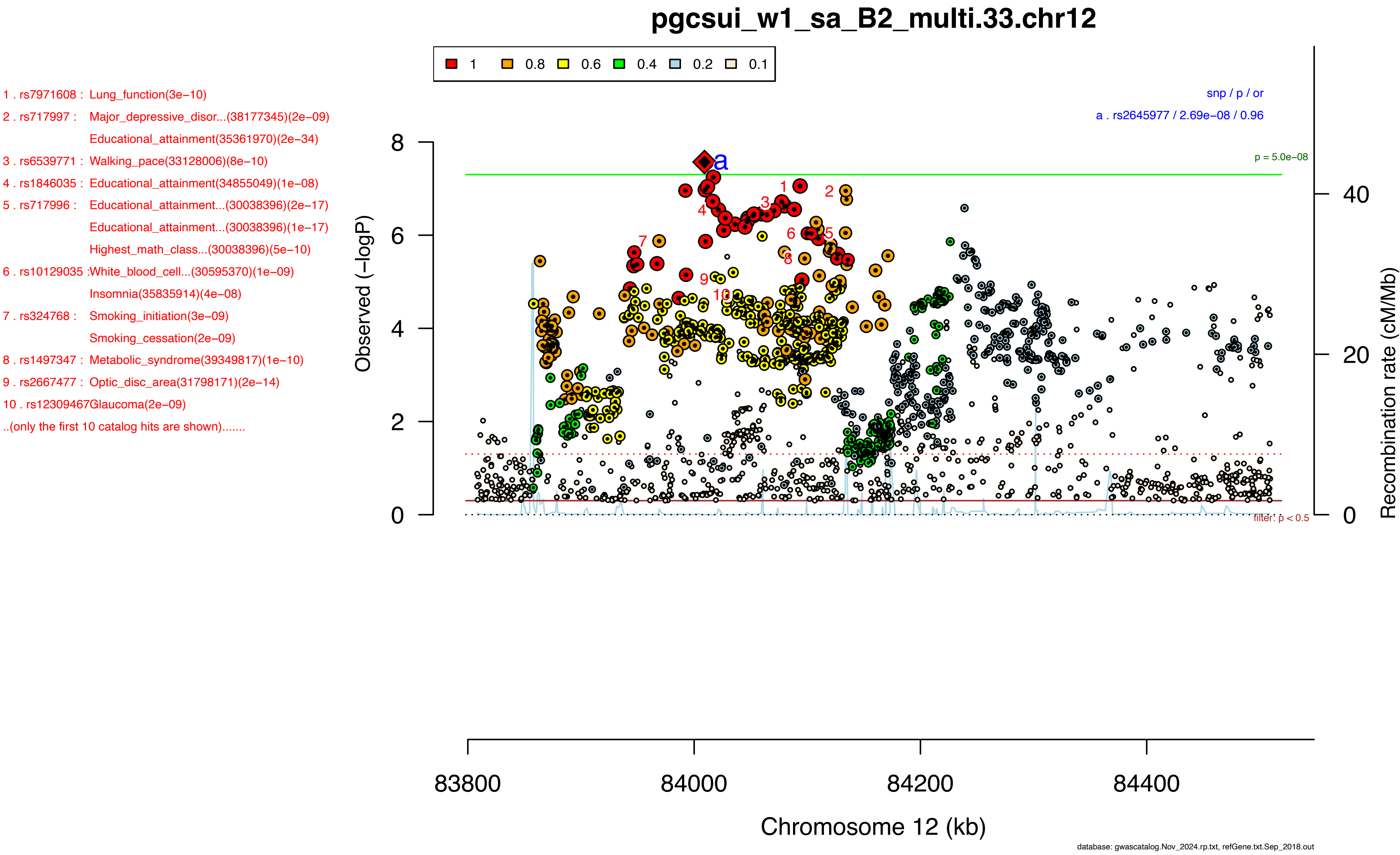

pgcsui_w1_sa_B2_multi.33.chr12
1
0.8
0.6
0.4
0.2
0.1
1 . rs7971608 : Lung_function(3e−10)
2 . rs717997 : Major_depressive_disor...(38177345)(2e−09)
Educational_attainment(35361970)(2e−34)
3 . rs6539771 : Walking_pace(33128006)(8e−10)
4 . rs1846035 : Educational_attainment(34855049)(1e−08)
5 . rs717996 : Educational_attainment...(30038396)(2e−17)
Educational_attainment...(30038396)(1e−17)
Highest_math_class...(30038396)(5e−10)
6 . rs10129035 :White_blood_cell...(30595370)(1e−09)
Insomnia(35835914)(4e−08)
7 . rs324768 : Smoking_initiation(3e−09)
Smoking_cessation(2e−09)
8 . rs1497347 : Metabolic_syndrome(39349817)(1e−10)
9 . rs2667477 : Optic_disc_area(31798171)(2e−14)
10 . rs12309467Glaucoma(2e−09)
..(only the first 10 catalog hits are shown).......
snp / p / or
a . rs2645977 / 2.69e−08 / 0.96
p = 5.0e−08
Observed (−logP)
Recombination rate (cM/Mb)
filter: p < 0.5
83800
84000
84200
84400
Chromosome 12 (kb)
database: gwascatalog.Nov_2024.rp.txt, refGene.txt.Sep_2018.out

1 . rs9317758 : Neuroticism(29942085)(4e−09)
Depression(33859377)(2e−15)
2 . rs7984597 : Neuroticism(36244801)(9e−10)
3 . rs11841876 :Neuroticism(30595370)(3e−09)
4 . rs2210903 : General_factor_of...(30867560)(2e−09)
Neuroticism(32231276)(1e−09)
5 . rs8002479 : Neuroticism(29942085)(1e−09)
6 . rs9541703 : Neurociticism(29500382)(7e−09)
7 . rs9541650 : General_factor_of...(30867560)(7e−09)
8 . rs8002150 : General_factor_of...(30867560)(2e−09)
9 . rs9541694 : Anxiety(33859377)(3e−12)
10 . rs9529502 :Neuroticism(29942085)(4e−08)
..(only the first 10 catalog hits are shown).......

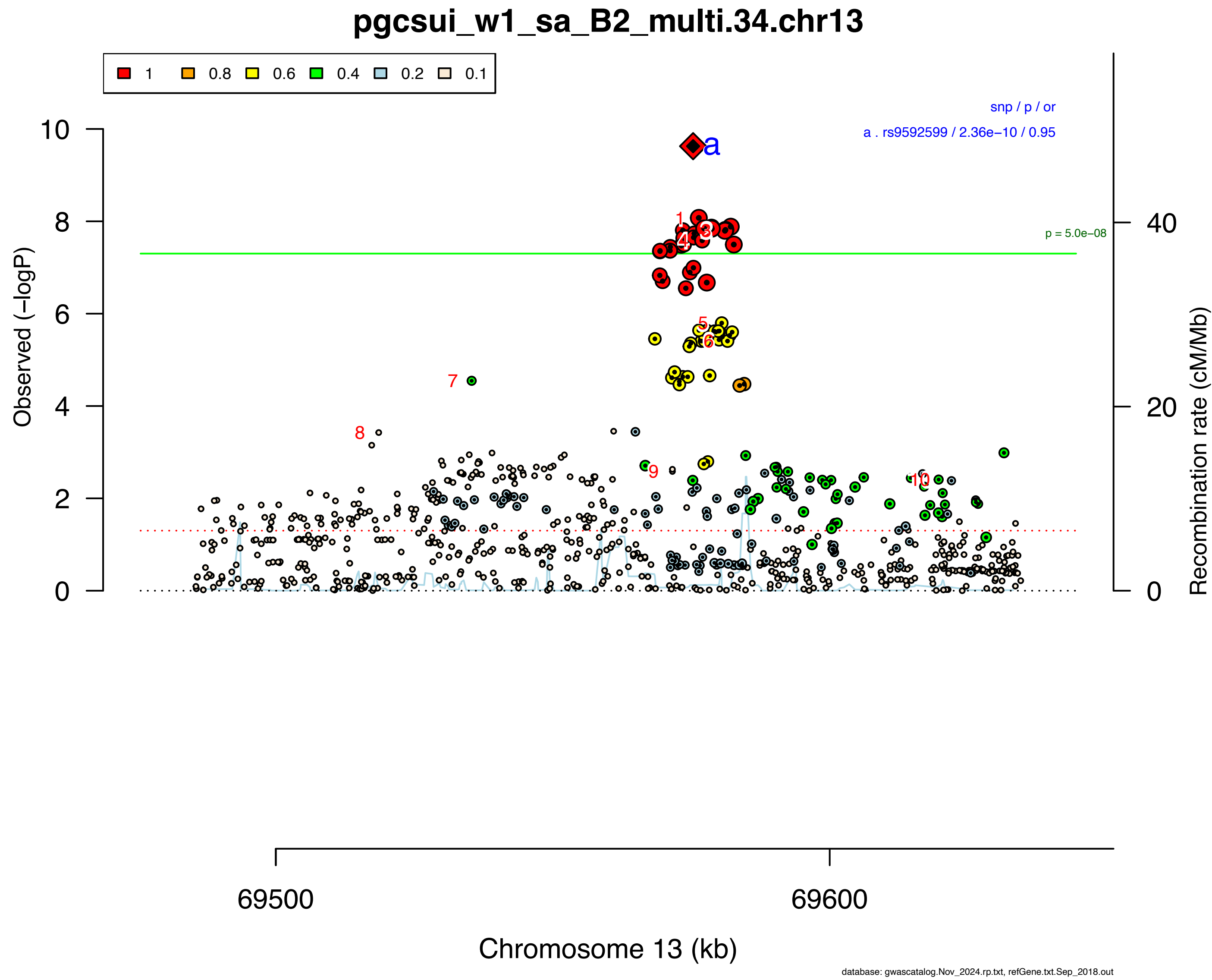

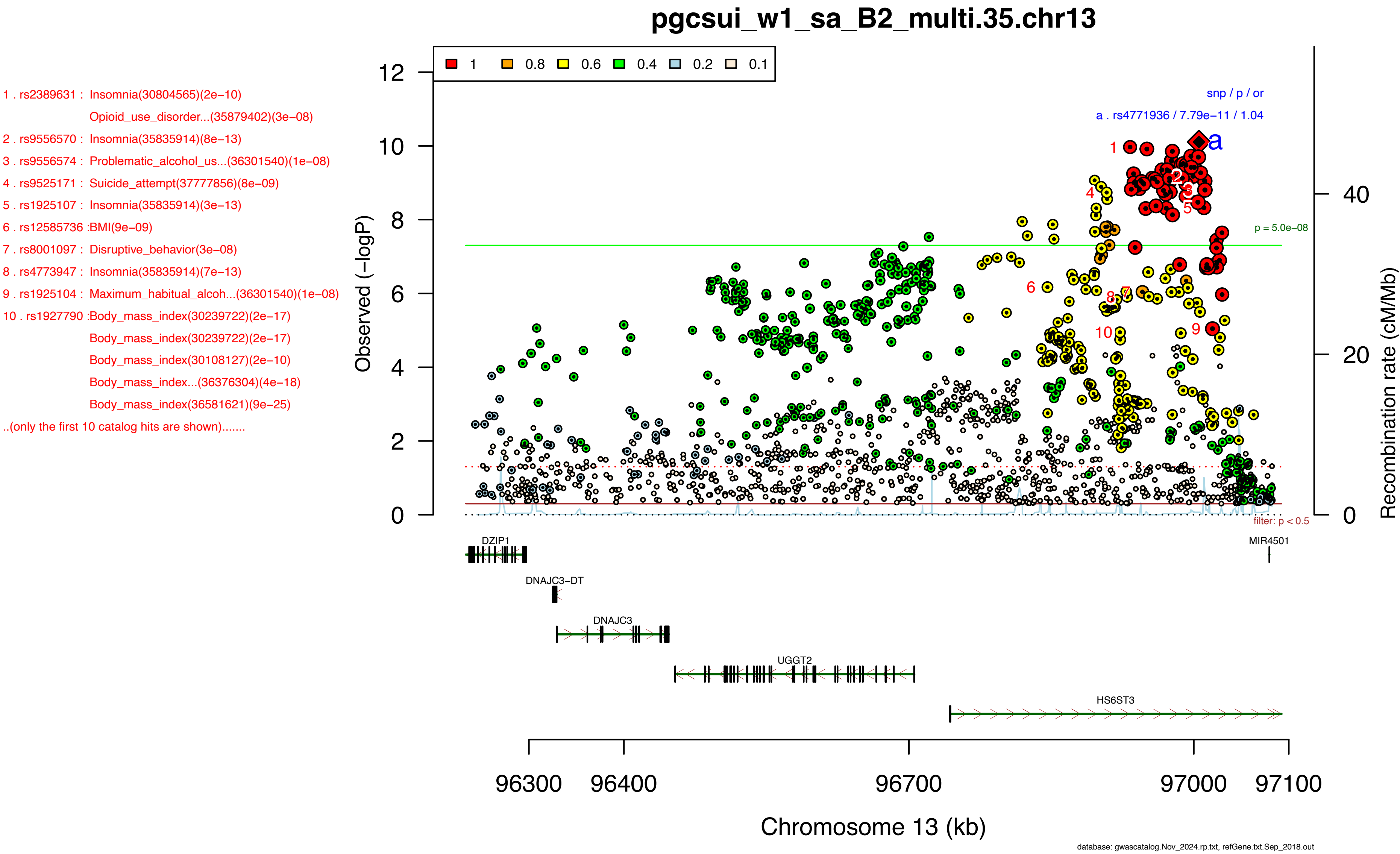
pgcsui_w1_sa_B2_multi.35.chr13
1 . rs2389631 : Insomnia(30804565)(2e−10)
Opioid_use_disorder...(35879402)(3e−08)
2 . rs9556570 : Insomnia(35835914)(8e−13)
3 . rs9556574 : Problematic_alcohol_us...(36301540)(1e−08)
4 . rs9525171 : Suicide_attempt(37777856)(8e−09)
5 . rs1925107 : Insomnia(35835914)(3e−13)
6 . rs12585736 :BMI(9e−09)
7 . rs8001097 : Disruptive_behavior(3e−08)
8 . rs4773947 : Insomnia(35835914)(7e−13)
9 . rs1925104 : Maximum_habitual_alcoh...(36301540)(1e−08)
10 . rs1927790 :Body_mass_index(30239722)(2e−17)
Body_mass_index(30239722)(2e−17)
Body_mass_index(30108127)(2e−10)
Body_mass_index...(36376304)(4e−18)
Body_mass_index(36581621)(9e−25)
..(only the first 10 catalog hits are shown).......
1
0.8
0.6
0.4
0.2
0.1
snp / p / or
a . rs4771936 / 7.79e−11 / 1.04
p = 5.0e−08
filter: p < 0.5
Observed (−logP)
Recombination rate (cM/Mb)
DZIP1
DNAJC3−DT
DNAJC3
UGGT2
HS6ST3
MIR4501
96300
96400
96700
97000
97100
Chromosome 13 (kb)
database: gwascatalog.Nov_2024.rp.txt, refGene.txt.Sep_2018.out

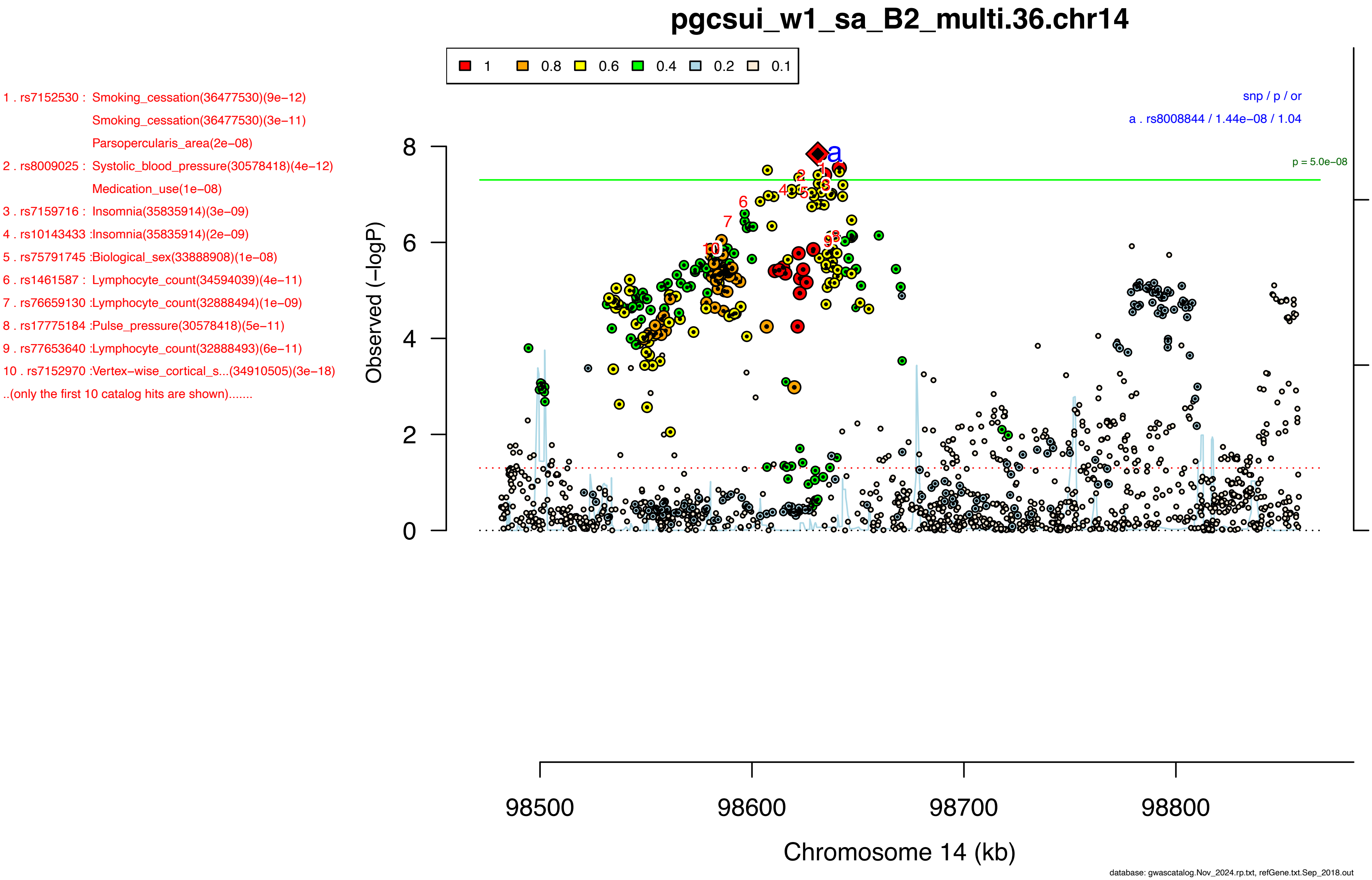
pgcsui_w1_sa_B2_multi.36.chr14
1 0.8 0.6 0.4 0.2 0.1
1 . rs7152530 : Smoking_cessation(36477530)(9e−12)
Smoking_cessation(36477530)(3e−11)
Parsopercularis_area(2e−08)
2 . rs8009025 : Systolic_blood_pressure(30578418)(4e−12)
Medication_use(1e−08)
3 . rs7159716 : Insomnia(35835914)(3e−09)
4 . rs10143433 :Insomnia(35835914)(2e−09)
5 . rs75791745 :Biological_sex(33888908)(1e−08)
6 . rs1461587 : Lymphocyte_count(34594039)(4e−11)
7 . rs76659130 :Lymphocyte_count(32888494)(1e−09)
8 . rs17775184 :Pulse_pressure(30578418)(5e−11)
9 . rs77653640 :Lymphocyte_count(32888493)(6e−11)
10 . rs7152970 :Vertex−wise_cortical_s...(34910505)(3e−18)
..(only the first 10 catalog hits are shown).......
snp / p / or
a . rs8008844 / 1.44e−08 / 1.04
p = 5.0e−08
Observed (−logP)
Recombination rate (cM/Mb)
Chromosome 14 (kb)
98500
98600
98700
98800
database: gwascatalog.Nov_2024.rp.txt, refGene.txt.Sep_2018.out

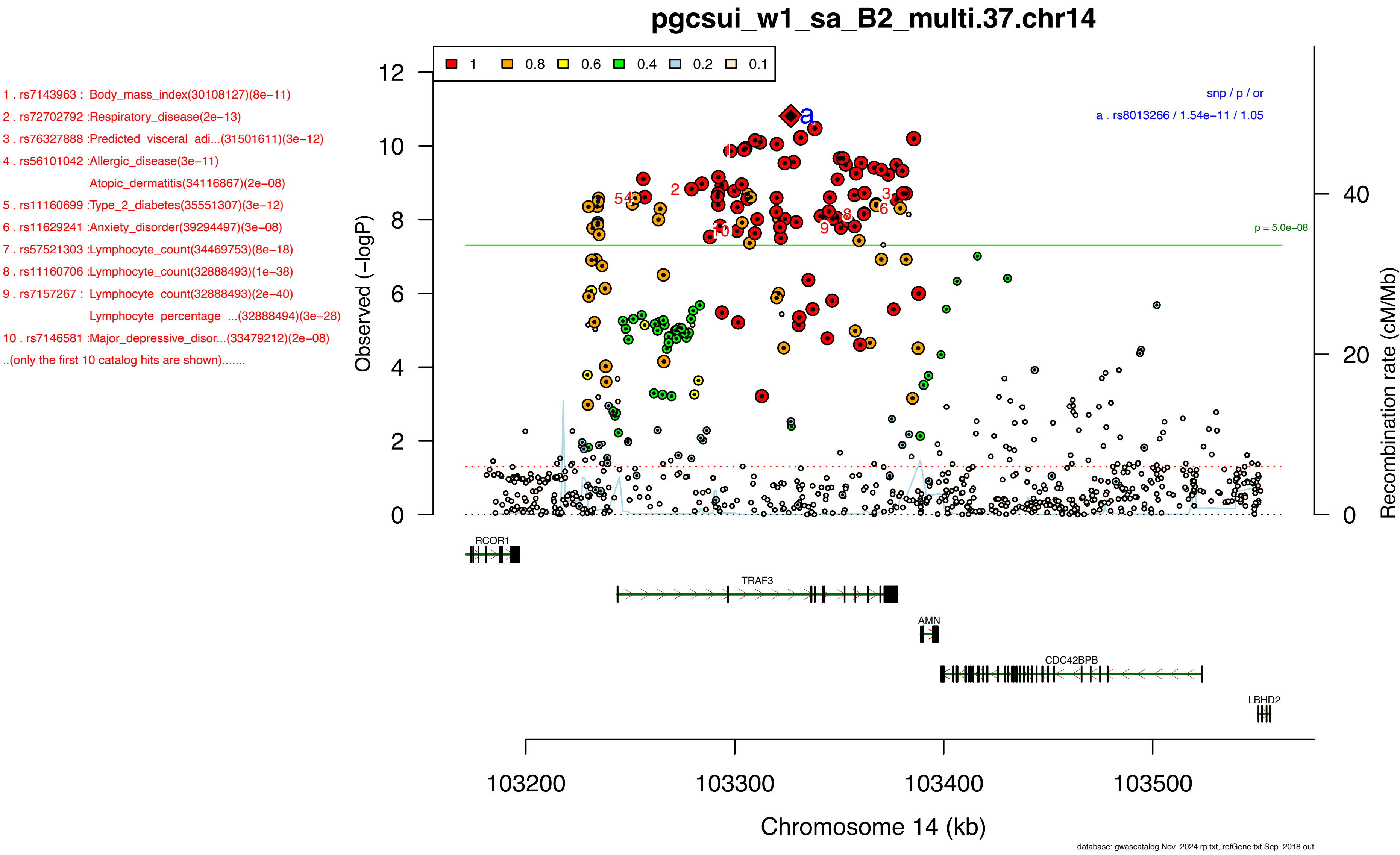
pgcsui_w1_sa_B2_multi.37.chr14
1 . rs7143963 : Body_mass_index(30108127)(8e−11)
2 . rs72702792 :Respiratory_disease(2e−13)
3 . rs76327888 :Predicted_visceral_adi...(31501611)(3e−12)
4 . rs56101042 :Allergic_disease(3e−11)
Atopic_dermatitis(34116867)(2e−08)
5 . rs11160699 :Type_2_diabetes(35551307)(3e−12)
6 . rs11629241 :Anxiety_disorder(39294497)(3e−08)
7 . rs57521303 :Lymphocyte_count(34469753)(8e−18)
8 . rs11160706 :Lymphocyte_count(32888493)(1e−38)
9 . rs7157267 : Lymphocyte_count(32888493)(2e−40)
Lymphocyte_percentage_...(32888494)(3e−28)
10 . rs7146581 :Major_depressive_disor...(33479212)(2e−08)
..(only the first 10 catalog hits are shown).......
1 0.8 0.6 0.4 0.2 0.1
snp / p / or
a . rs8013266 / 1.54e−11 / 1.05
p = 5.0e−08
Observed (−logP)
Recombination rate (cM/Mb)
RCOR1
TRAF3
AMN
CDC42BPB
LBHD2
103200 103300 103400 103500
Chromosome 14 (kb)
database: gwascatalog.Nov_2024.rp.txt, refGene.txt.Sep_2018.out

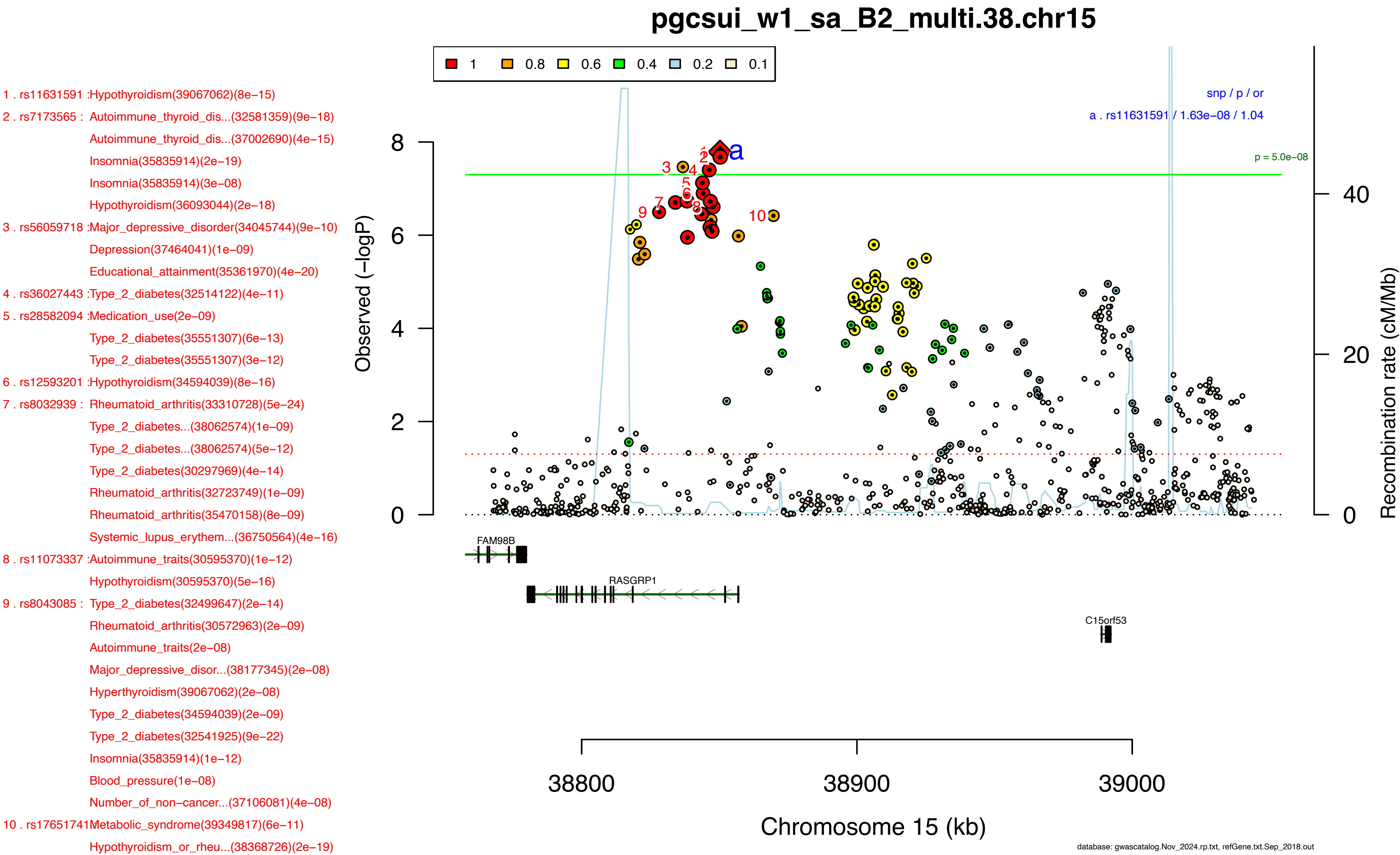

pgcsui_w1_sa_B2_multi.38.chr15
1 0.8 0.6 0.4 0.2 0.1
snp / p / or
a . rs11631591 / 1.63e−08 / 1.04
p = 5.0e−08
Observed (−logP)
Recombination rate (cM/Mb)
Chromosome 15 (kb)
38800
38900
39000
FAM98B
RASGRP1
C15orf53
1 . rs11631591 :Hypothyroidism(39067062)(8e−15)
2 . rs7173565 : Autoimmune_thyroid_dis...(32581359)(9e−18)
Autoimmune_thyroid_dis...(37002690)(4e−15)
Insomnia(35835914)(2e−19)
Insomnia(35835914)(3e−08)
Hypothyroidism(36093044)(2e−18)
3 . rs56059718 :Major_depressive_disorder(34045744)(9e−10)
Depression(37464041)(1e−09)
Educational_attainment(35361970)(4e−20)
4 . rs36027443 :Type_2_diabetes(32514122)(4e−11)
5 . rs28582094 :Medication_use(2e−09)
Type_2_diabetes(35551307)(6e−13)
Type_2_diabetes(35551307)(3e−12)
6 . rs12593201 :Hypothyroidism(34594039)(8e−16)
7 . rs8032939 : Rheumatoid_arthritis(33310728)(5e−24)
Type_2_diabetes...(38062574)(1e−09)
Type_2_diabetes...(38062574)(5e−12)
Type_2_diabetes(30297969)(4e−14)
Rheumatoid_arthritis(32723749)(1e−09)
Rheumatoid_arthritis(35470158)(8e−09)
Systemic_lupus_erythem...(36750564)(4e−16)
8 . rs11073337 :Autoimmune_traits(30595370)(1e−12)
Hypothyroidism(30595370)(5e−16)
9 . rs8043085 : Type_2_diabetes(32499647)(2e−14)
Rheumatoid_arthritis(30572963)(2e−09)
Autoimmune_traits(2e−08)
Major_depressive_disor...(38177345)(2e−08)
Hyperthyroidism(39067062)(2e−08)
Type_2_diabetes(34594039)(2e−09)
Type_2_diabetes(32541925)(9e−22)
Insomnia(35835914)(1e−12)
Blood_pressure(1e−08)
Number_of_non−cancer...(37106081)(4e−08)
10 . rs17651741Metabolic_syndrome(39349817)(6e−11)
Hypothyroidism_or_rheu...(38368726)(2e−19)
database: gwascatalog.Nov_2024.rp.txt, refGene.txt.Sep_2018.out

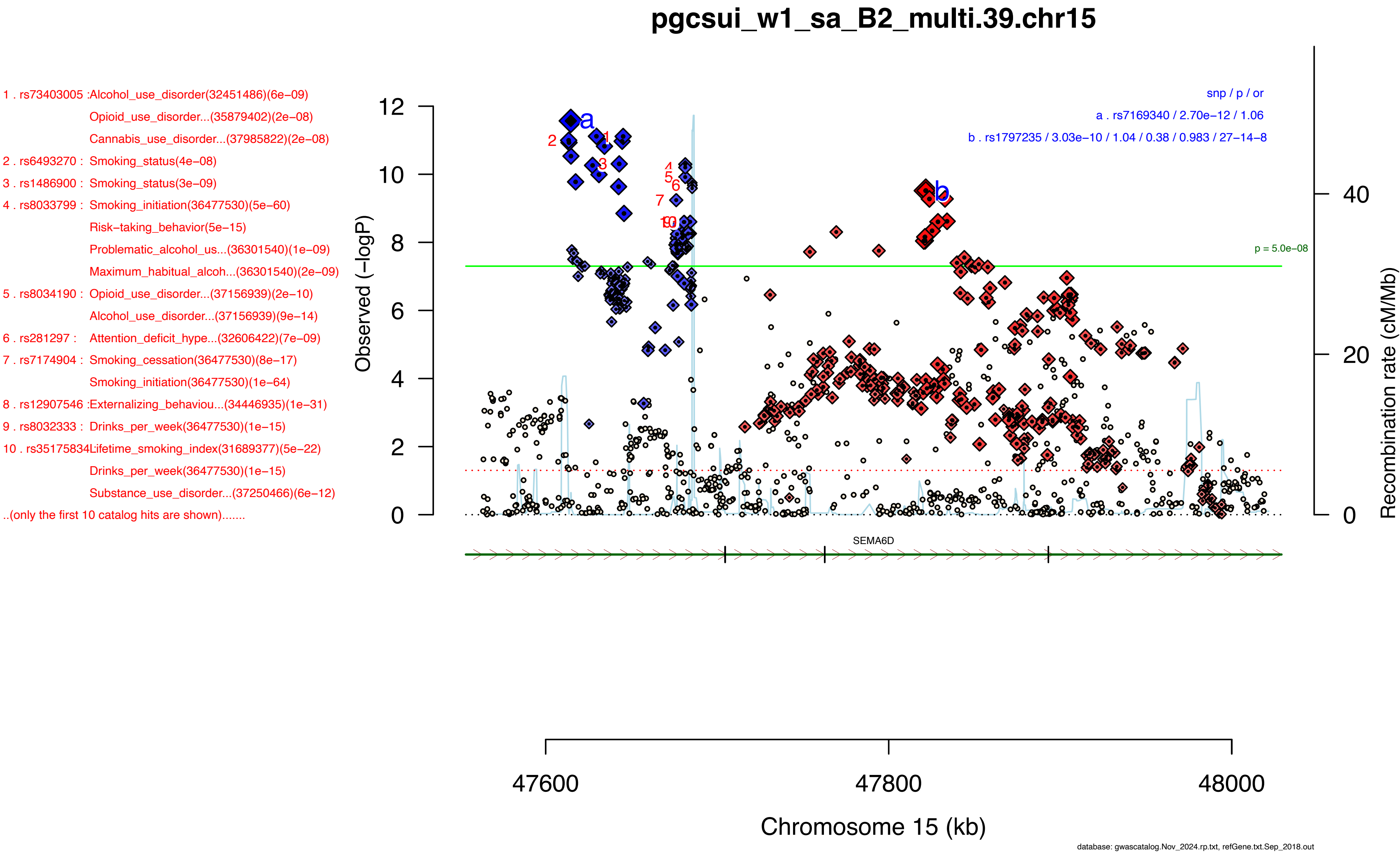

pgcsui_w1_sa_B2_multi.39.chr15
1 . rs73403005 :Alcohol_use_disorder(32451486)(6e−09)
Opioid_use_disorder...(35879402)(2e−08)
Cannabis_use_disorder...(37985822)(2e−08)
2 . rs6493270 : Smoking_status(4e−08)
3 . rs1486900 : Smoking_status(3e−09)
4 . rs8033799 : Smoking_initiation(36477530)(5e−60)
Risk−taking_behavior(5e−15)
Problematic_alcohol_us...(36301540)(1e−09)
Maximum_habitual_alcoh...(36301540)(2e−09)
5 . rs8034190 : Opioid_use_disorder...(37156939)(2e−10)
Alcohol_use_disorder...(37156939)(9e−14)
6 . rs281297 : Attention_deficit_hype...(32606422)(7e−09)
7 . rs7174904 : Smoking_cessation(36477530)(8e−17)
Smoking_initiation(36477530)(1e−64)
8 . rs12907546 :Externalizing_behaviou...(34446935)(1e−31)
9 . rs8032333 : Drinks_per_week(36477530)(1e−15)
10 . rs35175834Lifetime_smoking_index(31689377)(5e−22)
Drinks_per_week(36477530)(1e−15)
Substance_use_disorder...(37250466)(6e−12)
..(only the first 10 catalog hits are shown).......
snp / p / or
a . rs7169340 / 2.70e−12 / 1.06
b . rs1797235 / 3.03e−10 / 1.04 / 0.38 / 0.983 / 27−14−8
p = 5.0e−08
Observed (−logP)
Recombination rate (cM/Mb)
SEMA6D
47600
47800
48000
Chromosome 15 (kb)
database: gwascatalog.Nov_2024.rp.txt, refGene.txt.Sep_2018.out

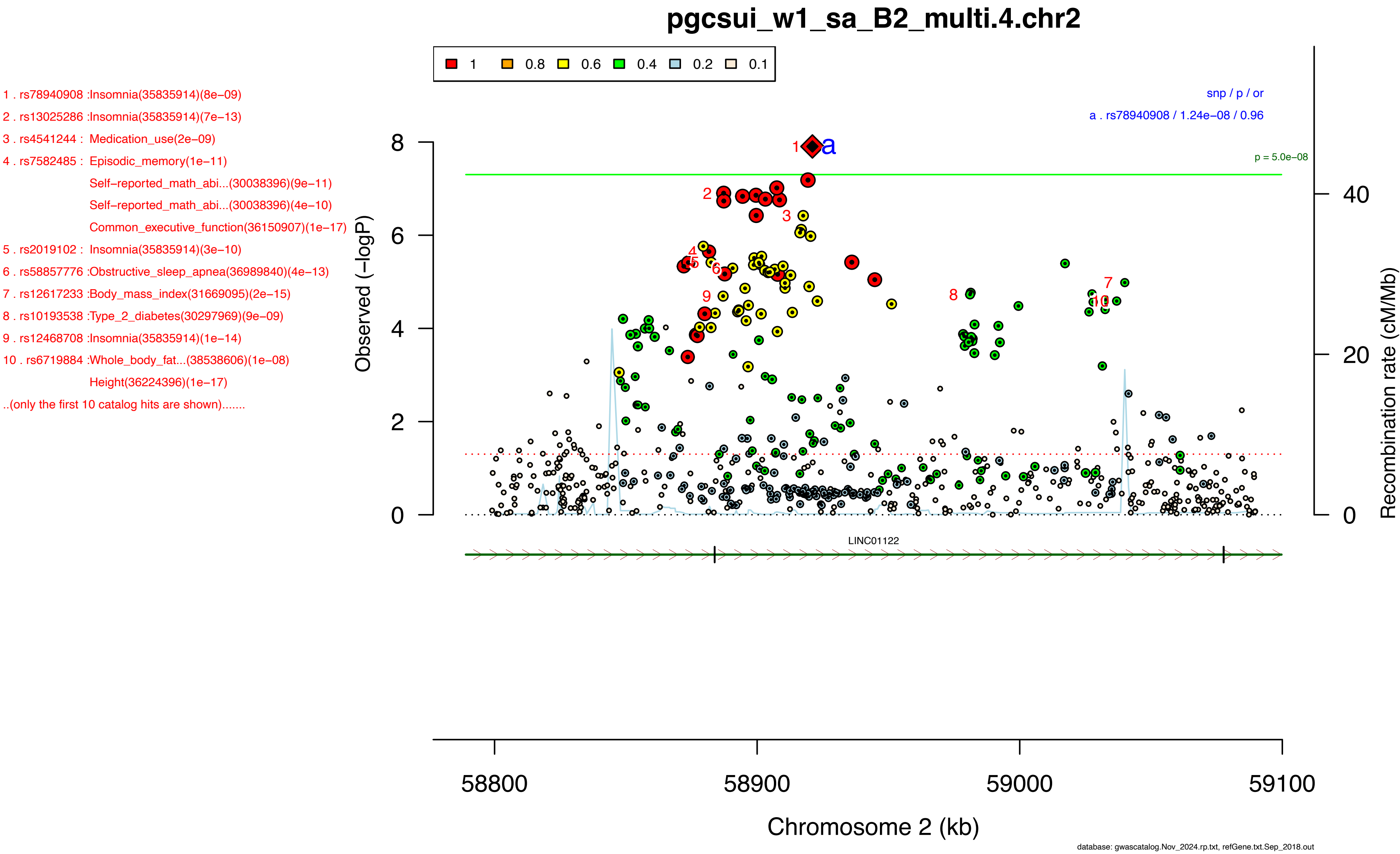
pgcsui_w1_sa_B2_multi.4.chr2
1 . rs78940908 :Insomnia(35835914)(8e−09)
2 . rs13025286 :Insomnia(35835914)(7e−13)
3 . rs4541244 : Medication_use(2e−09)
4 . rs7582485 : Episodic_memory(1e−11)
Self−reported_math_abi...(30038396)(9e−11)
Self−reported_math_abi...(30038396)(4e−10)
Common_executive_function(36150907)(1e−17)
5 . rs2019102 : Insomnia(35835914)(3e−10)
6 . rs58857776 :Obstructive_sleep_apnea(36989840)(4e−13)
7 . rs12617233 :Body_mass_index(31669095)(2e−15)
8 . rs10193538 :Type_2_diabetes(30297969)(9e−09)
9 . rs12468708 :Insomnia(35835914)(1e−14)
10 . rs6719884 :Whole_body_fat...(38538606)(1e−08)
Height(36224396)(1e−17)
..(only the first 10 catalog hits are shown).......
1 0.8 0.6 0.4 0.2 0.1
snp / p / or
a . rs78940908 / 1.24e−08 / 0.96
p = 5.0e−08
Observed (−logP)
Recombination rate (cM/Mb)
LINC01122
58800
58900
59000
59100
Chromosome 2 (kb)
database: gwascatalog.Nov_2024.rp.txt, refGene.txt.Sep_2018.out

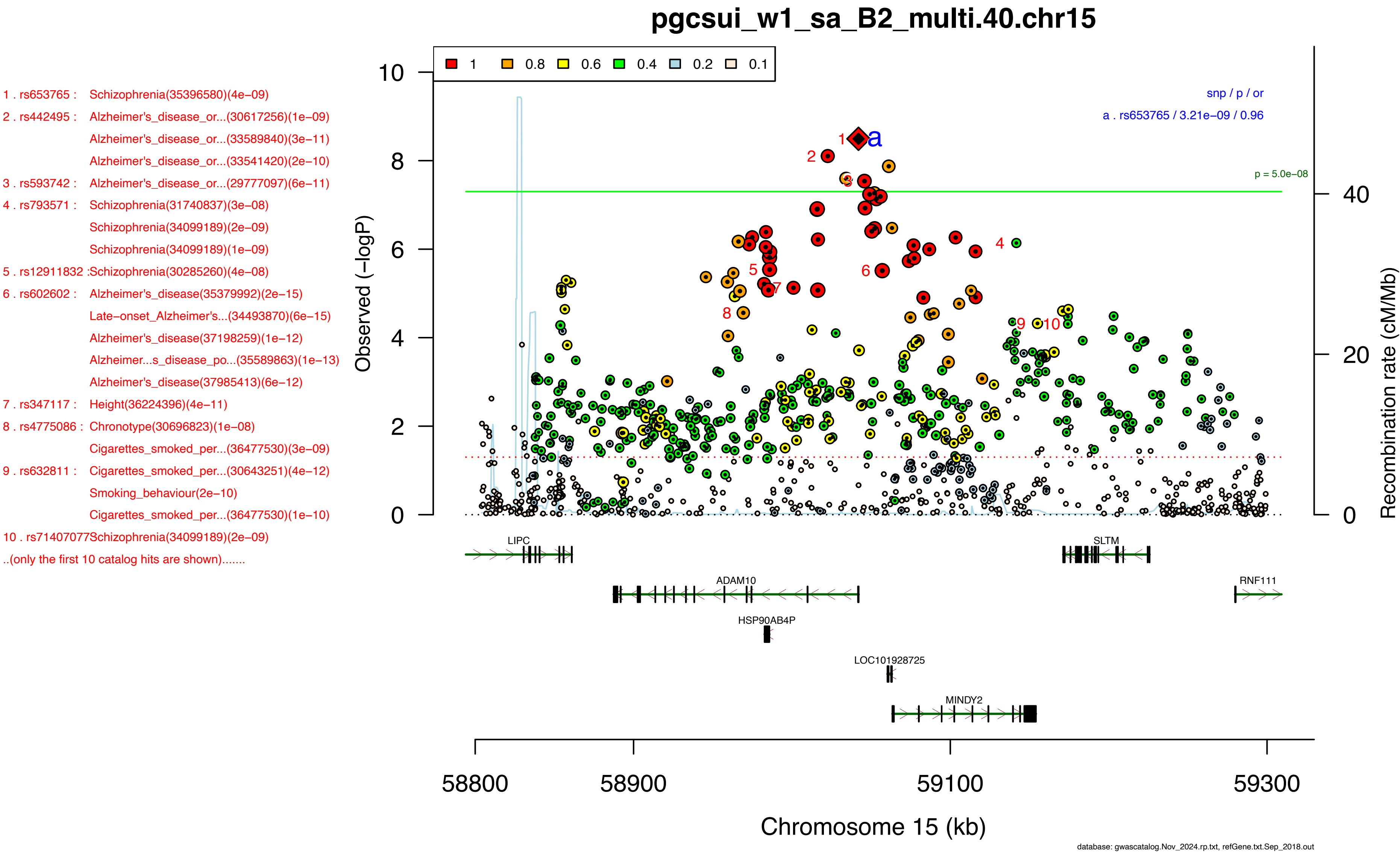
pgcsui_w1_sa_B2_multi.40.chr15
1 . rs653765 : Schizophrenia(35396580)(4e−09)
2 . rs442495 : Alzheimer's_disease_or...(30617256)(1e−09)
Alzheimer's_disease_or...(33589840)(3e−11)
Alzheimer's_disease_or...(33541420)(2e−10)
3 . rs593742 : Alzheimer's_disease_or...(29777097)(6e−11)
4 . rs793571 : Schizophrenia(31740837)(3e−08)
Schizophrenia(34099189)(2e−09)
Schizophrenia(34099189)(1e−09)
5 . rs12911832 :Schizophrenia(30285260)(4e−08)
6 . rs602602 : Alzheimer's_disease(35379992)(2e−15)
Late−onset_Alzheimer's...(34493870)(6e−15)
Alzheimer's_disease(37198259)(1e−12)
Alzheimer...s_disease_po...(35589863)(1e−13)
Alzheimer's_disease(37985413)(6e−12)
7 . rs347117 : Height(36224396)(4e−11)
8 . rs4775086 : Chronotype(30696823)(1e−08)
Cigarettes_smoked_per...(36477530)(3e−09)
9 . rs632811 : Cigarettes_smoked_per...(30643251)(4e−12)
Smoking_behaviour(2e−10)
Cigarettes_smoked_per...(36477530)(1e−10)
10 . rs71407077Schizophrenia(34099189)(2e−09)
..(only the first 10 catalog hits are shown).......
1 0.8 0.6 0.4 0.2 0.1
snp / p / or
a . rs653765 / 3.21e−09 / 0.96
p = 5.0e−08
Observed (−logP)
Recombination rate (cM/Mb)
LIPC
SLTM
ADAM10
RNF111
HSP90AB4P
LOC101928725
MINDY2
58800
58900
59100
59300
Chromosome 15 (kb)
database: gwascatalog.Nov_2024.rp.txt, refGene.txt.Sep_2018.out

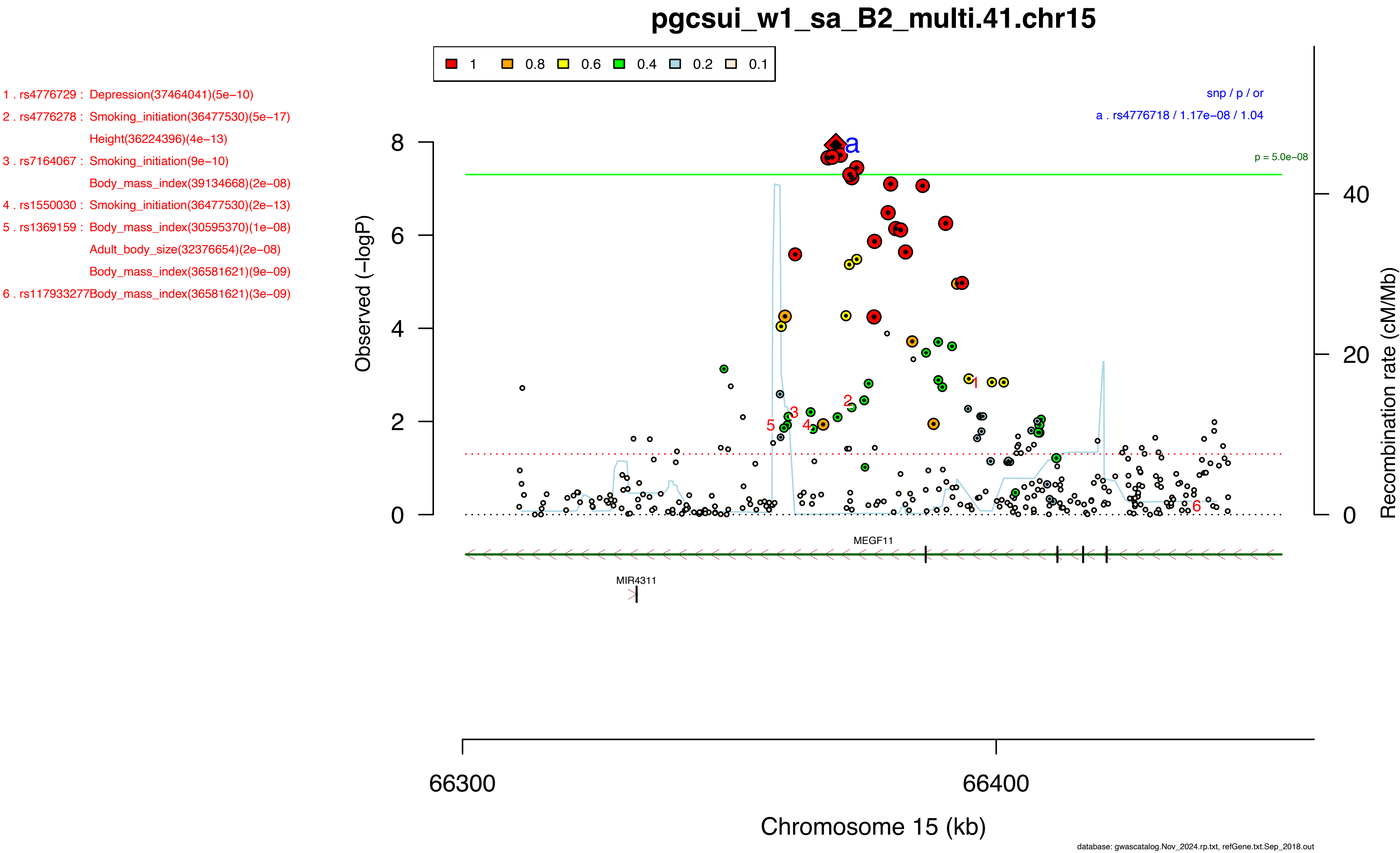
pgcsui_w1_sa_B2_multi.41.chr15
1 0.8 0.6 0.4 0.2 0.1
1 . rs4776729 : Depression(37464041)(5e−10)
2 . rs4776278 : Smoking_initiation(36477530)(5e−17)
Height(36224396)(4e−13)
3 . rs7164067 : Smoking_initiation(9e−10)
Body_mass_index(39134668)(2e−08)
4 . rs1550030 : Smoking_initiation(36477530)(2e−13)
5 . rs1369159 : Body_mass_index(30595370)(1e−08)
Adult_body_size(32376654)(2e−08)
Body_mass_index(36581621)(9e−09)
6 . rs117933277Body_mass_index(36581621)(3e−09)
snp / p / or
a . rs4776718 / 1.17e−08 / 1.04
p = 5.0e−08
Observed (−logP)
Recombination rate (cM/Mb)
MEGF11
MIR4311
66300
66400
Chromosome 15 (kb)
database: gwascatalog.Nov_2024.rp.txt, refGene.txt.Sep_2018.out

1 . rs2071382 : Post−traumatic_stress_...(35181757)(2e−09)
Male−pattern_baldness(30573740)(1e−13)
Systolic_blood_pressure(30578418)(2e−32)
Pulse_pressure(30578418)(2e−19)
Attention_deficit_hype...(38565336)(1e−09)
2 . rs11539637 :Schizophrenia(30285260)(4e−09)
Birth_weight(31097437)(2e−08)
3 . rs8039305 : Hypertension(30487518)(7e−09)
Neutrophil_count(32888494)(6e−15)
Neutrophil_count(34469753)(9e−09)
Major_depressive_disor...(34159505)(2e−08)
Bipolar_disorder(3e−09)
Schizophrenia(5e−11)
Medication_use(4e−09)
4 . rs7177338 : Placental_weight(1e−09)
Vertex−wise_cortical_s...(34910505)(2e−29)
Pulse_pressure(34594039)(9e−19)
Whole_brain_restricted...(35505052)(8e−17)
Coronary_artery_disease(33020668)(2e−11)
Coronary_artery_disease(33020668)(2e−34)
Coronary_artery_disease(36474045)(3e−33)
Cortical_surface_area(34560273)(1e−17)
5 . rs7183988 : Insomnia(30804565)(2e−08)
Offspring_birth_weight(31043758)(1e−20)
Whole_brain_free...(35505052)(4e−09)
Angina_pectoris(34594039)(2e−14)
Neutrophil_count(32888493)(6e−18)
Brain_morphology(7e−10)
Medication_use(6e−24)
Coronary_artery_disease(36474045)(3e−33)
6 . rs6224 : Balding_type_1(30595370)(5e−11)
Left−handedness(32989287)(6e−21)
Respiratory_disease(8e−09)
Parental_lifespan(30642433)(1e−10)
Aging_traits(2e−09)

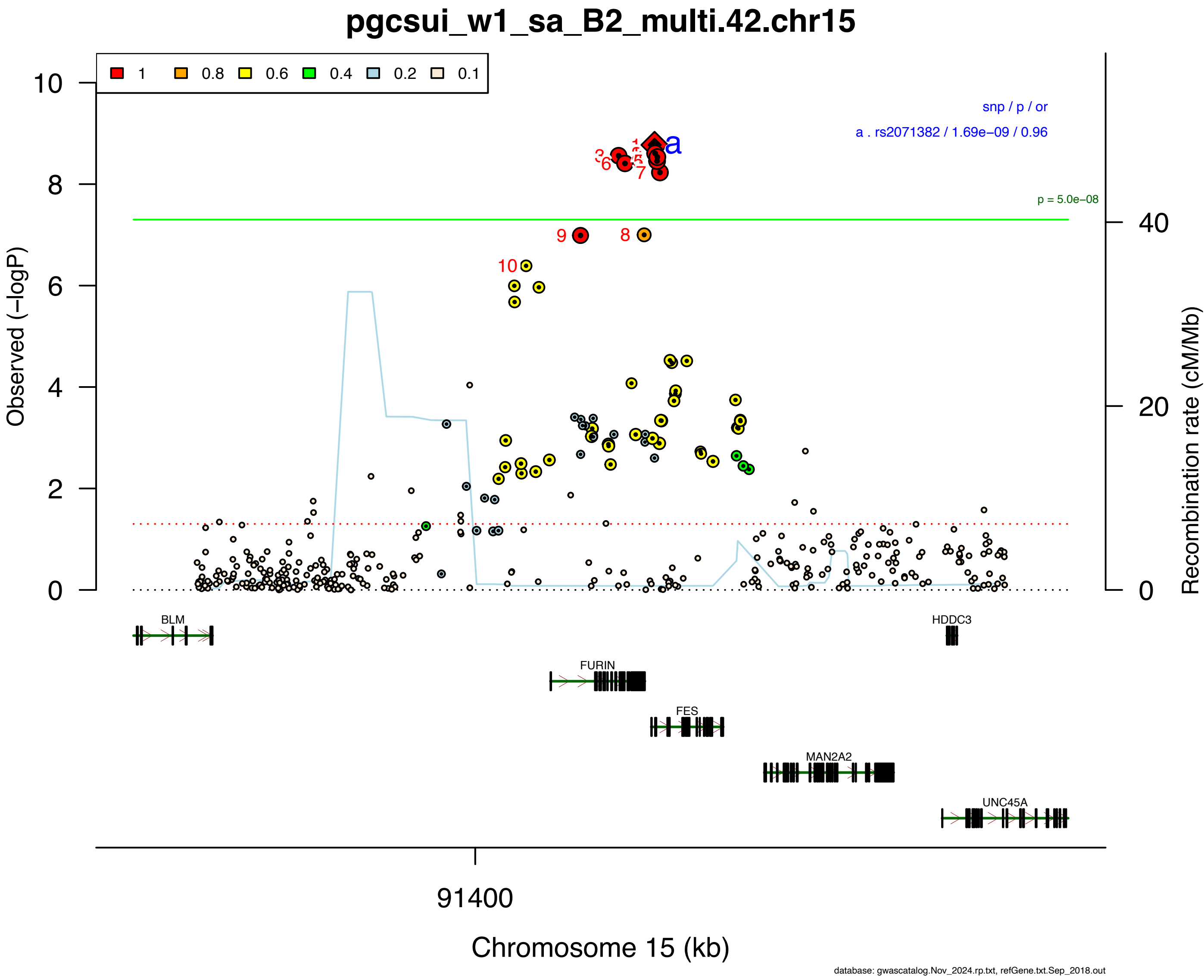

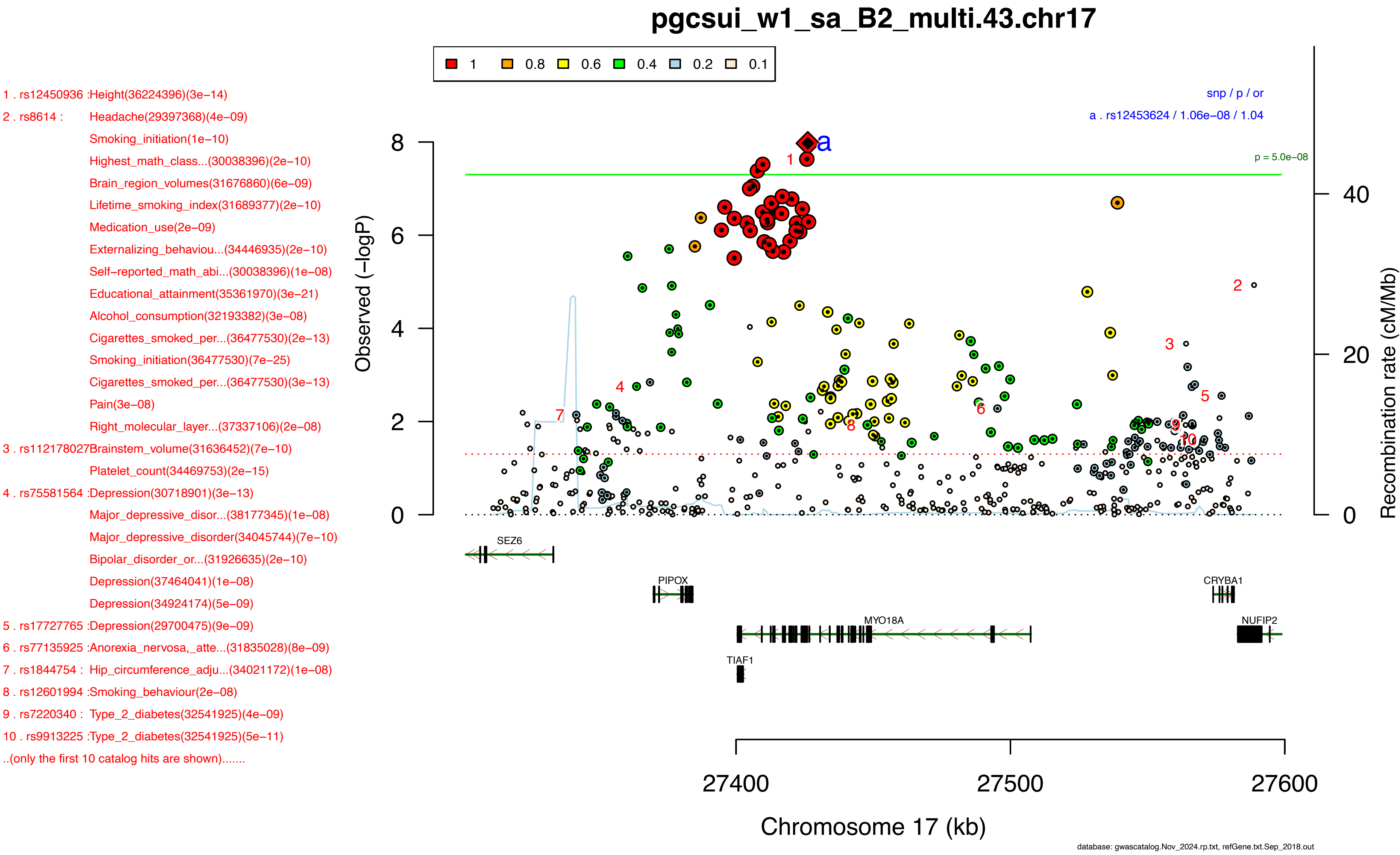
pgcsui_w1_sa_B2_multi.43.chr17
1
0.8
0.6
0.4
0.2
0.1
snp / p / or
a . rs12453624 / 1.06e−08 / 1.04
p = 5.0e−08
1 . rs12450936 :Height(36224396)(3e−14)
2 . rs8614 : Headache(29397368)(4e−09)
Smoking_initiation(1e−10)
Highest_math_class...(30038396)(2e−10)
Brain_region_volumes(31676860)(6e−09)
Lifetime_smoking_index(31689377)(2e−10)
Medication_use(2e−09)
Externalizing_behaviou...(34446935)(2e−10)
Self−reported_math_abi...(30038396)(1e−08)
Educational_attainment(35361970)(3e−21)
Alcohol_consumption(32193382)(3e−08)
Cigarettes_smoked_per...(36477530)(2e−13)
Smoking_initiation(36477530)(7e−25)
Cigarettes_smoked_per...(36477530)(3e−13)
Pain(3e−08)
Right_molecular_layer...(37337106)(2e−08)
3 . rs112178027Brainstem_volume(31636452)(7e−10)
Platelet_count(34469753)(2e−15)
4 . rs75581564 :Depression(30718901)(3e−13)
Major_depressive_disor...(38177345)(1e−08)
Major_depressive_disorder(34045744)(7e−10)
Bipolar_disorder_or...(31926635)(2e−10)
Depression(37464041)(1e−08)
Depression(34924174)(5e−09)
5 . rs17727765 :Depression(29700475)(9e−09)
6 . rs77135925 :Anorexia_nervosa,_atte...(31835028)(8e−09)
7 . rs1844754 : Hip_circumference_adju...(34021172)(1e−08)
8 . rs12601994 :Smoking_behaviour(2e−08)
9 . rs7220340 : Type_2_diabetes(32541925)(4e−09)
10 . rs9913225 :Type_2_diabetes(32541925)(5e−11)
..(only the first 10 catalog hits are shown).......
Observed (−logP)
Recombination rate (cM/Mb)
SEZ6
PIPOX
CRYBA1
MYO18A
NUFIP2
TIAF1
27400
27500
27600
Chromosome 17 (kb)
database: gwascatalog.Nov_2024.rp.txt, refGene.txt.Sep_2018.out

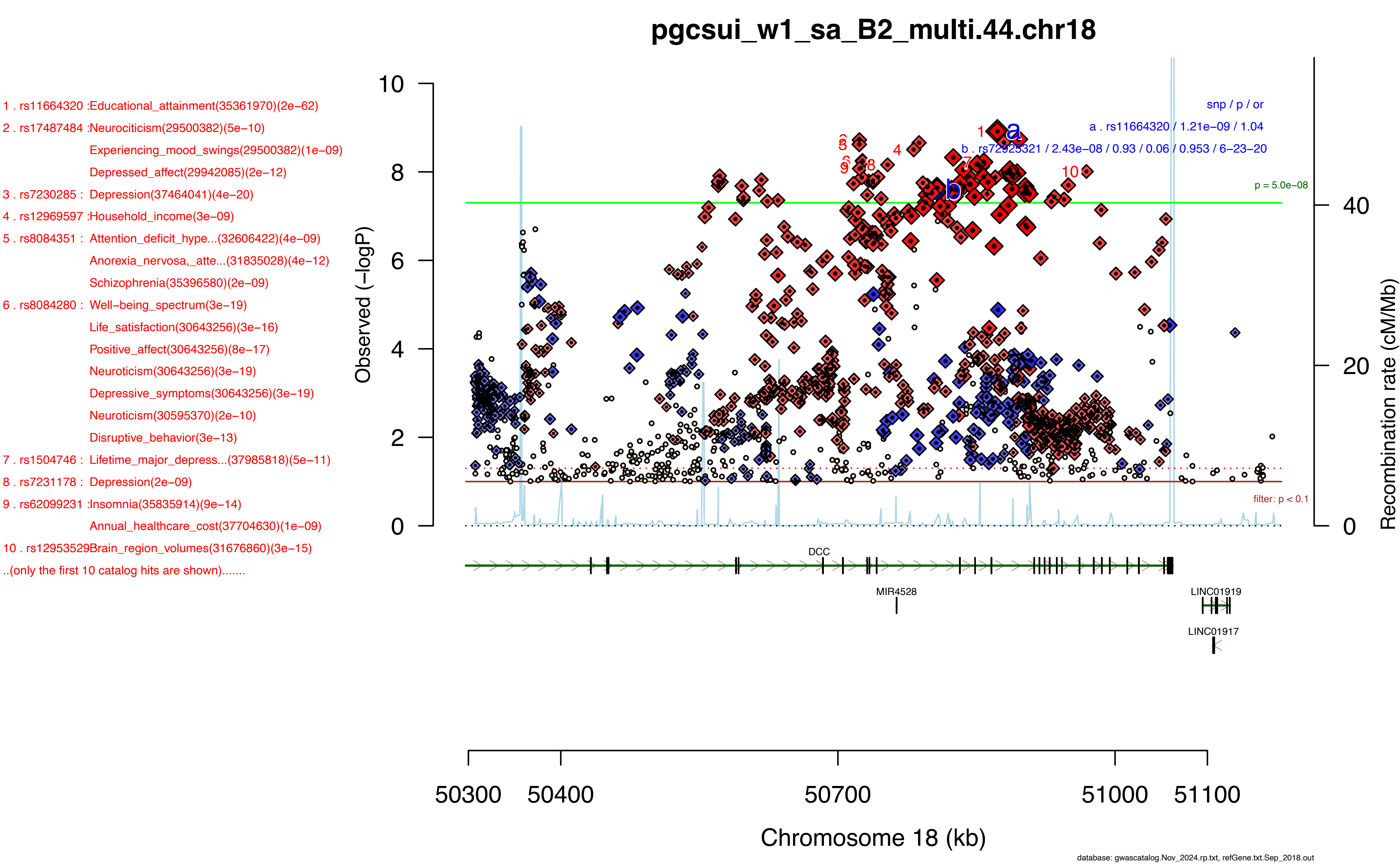
pgcsui_w1_sa_B2_multi.44.chr18
1 . rs11664320 :Educational_attainment(35361970)(2e−62)
2 . rs17487484 :Neurociticism(29500382)(5e−10)
Experiencing_mood_swings(29500382)(1e−09)
Depressed_affect(29942085)(2e−12)
3 . rs7230285 : Depression(37464041)(4e−20)
4 . rs12969597 :Household_income(3e−09)
5 . rs8084351 : Attention_deficit_hype...(32606422)(4e−09)
Anorexia_nervosa,_atte...(31835028)(4e−12)
Schizophrenia(35396580)(2e−09)
6 . rs8084280 : Well−being_spectrum(3e−19)
Life_satisfaction(30643256)(3e−16)
Positive_affect(30643256)(8e−17)
Neuroticism(30643256)(3e−19)
Depressive_symptoms(30643256)(3e−19)
Neuroticism(30595370)(2e−10)
Disruptive_behavior(3e−13)
7 . rs1504746 : Lifetime_major_depress...(37985818)(5e−11)
8 . rs7231178 : Depression(2e−09)
9 . rs62099231 :Insomnia(35835914)(9e−14)
Annual_healthcare_cost(37704630)(1e−09)
10 . rs12953529Brain_region_volumes(31676860)(3e−15)
..(only the first 10 catalog hits are shown).......
snp / p / or
a . rs11664320 / 1.21e−09 / 1.04
b . rs72925321 / 2.43e−08 / 0.93 / 0.06 / 0.953 / 6−23−20
p = 5.0e−08
filter: p < 0.1
Observed (−logP)
Recombination rate (cM/Mb)
DCC
MIR4528
LINC01919
LINC01917
50300
50400
50700
51000
51100
Chromosome 18 (kb)
database: gwascatalog.Nov_2024.rp.txt, refGene.txt.Sep_2018.out

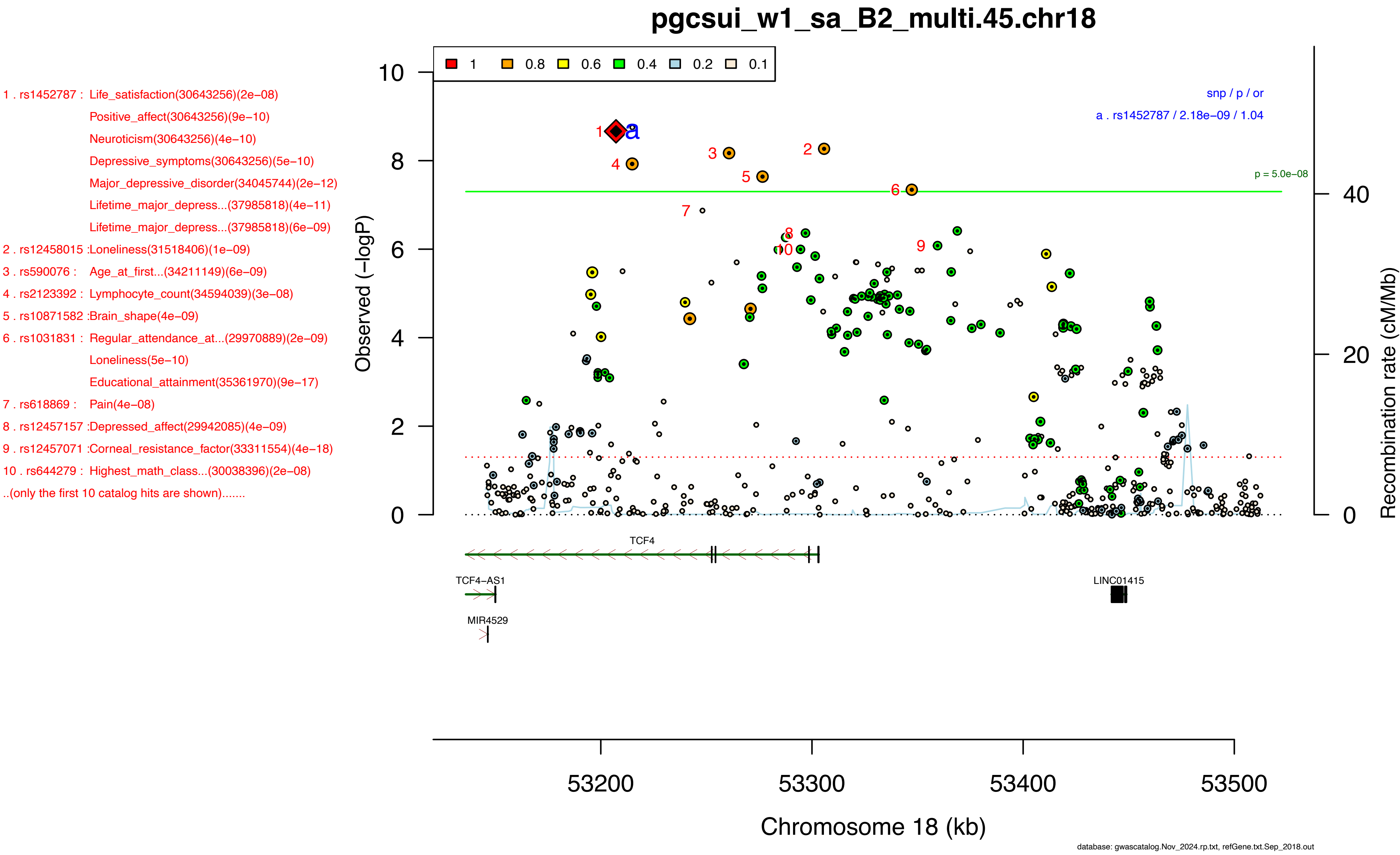

pgcsui_w1_sa_B2_multi.45.chr18
1 . rs1452787 : Life_satisfaction(30643256)(2e−08)
Positive_affect(30643256)(9e−10)
Neuroticism(30643256)(4e−10)
Depressive_symptoms(30643256)(5e−10)
Major_depressive_disorder(34045744)(2e−12)
Lifetime_major_depress...(37985818)(4e−11)
Lifetime_major_depress...(37985818)(6e−09)
2 . rs12458015 :Loneliness(31518406)(1e−09)
3 . rs590076 : Age_at_first...(34211149)(6e−09)
4 . rs2123392 : Lymphocyte_count(34594039)(3e−08)
5 . rs10871582 :Brain_shape(4e−09)
6 . rs1031831 : Regular_attendance_at...(29970889)(2e−09)
Loneliness(5e−10)
Educational_attainment(35361970)(9e−17)
7 . rs618869 : Pain(4e−08)
8 . rs12457157 :Depressed_affect(29942085)(4e−09)
9 . rs12457071 :Corneal_resistance_factor(33311554)(4e−18)
10 . rs644279 : Highest_math_class...(30038396)(2e−08)
..(only the first 10 catalog hits are shown).......
1 0.8 0.6 0.4 0.2 0.1
snp / p / or
a . rs1452787 / 2.18e−09 / 1.04
p = 5.0e−08
Observed (−logP)
Recombination rate (cM/Mb)
TCF4
TCF4−AS1
MIR4529
LINC01415
53200 53300 53400 53500
Chromosome 18 (kb)
database: gwascatalog.Nov_2024.rp.txt, refGene.txt.Sep_2018.out

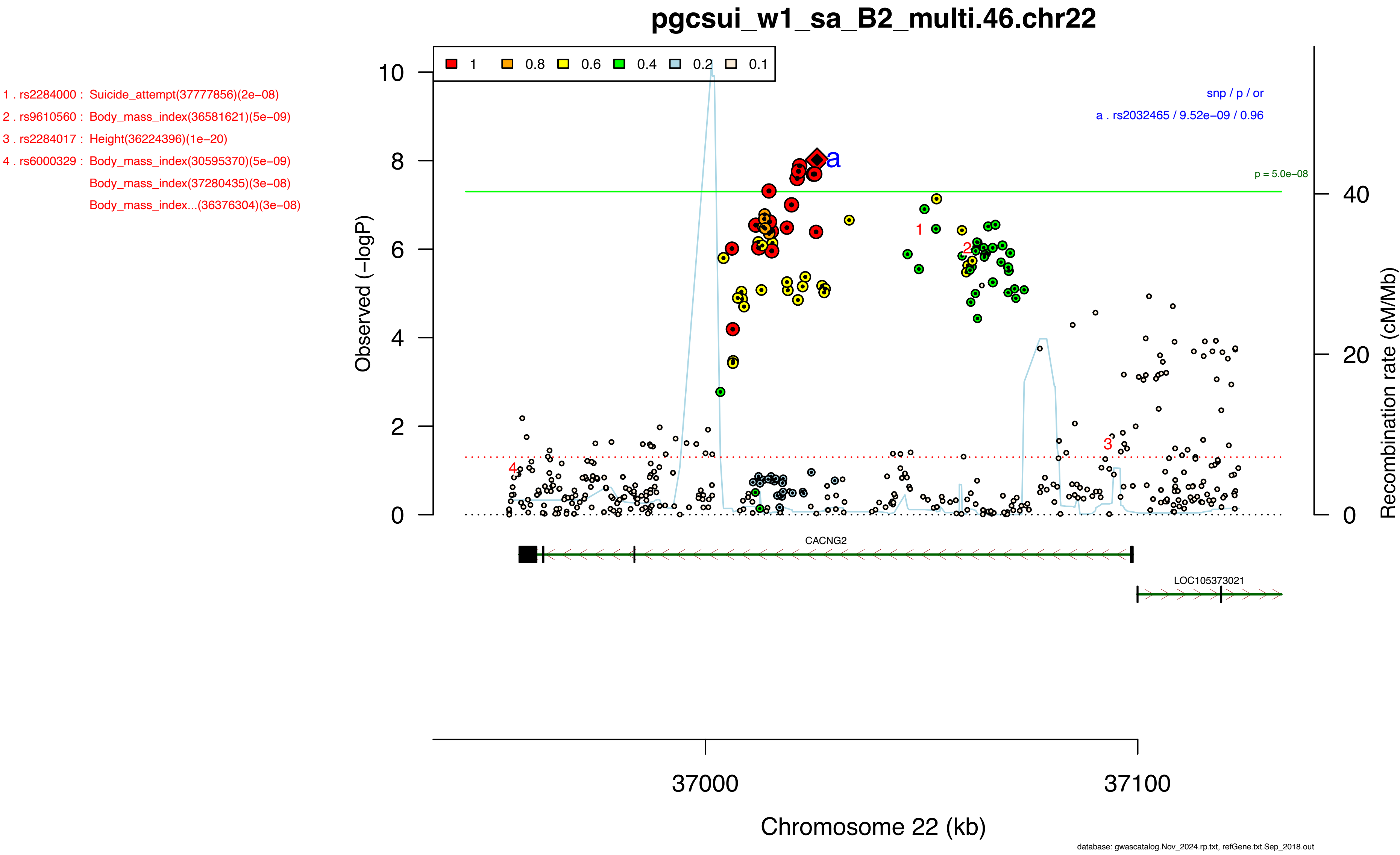
pgcsui_w1_sa_B2_multi.46.chr22
1 . rs2284000 : Suicide_attempt(37777856)(2e−08)
2 . rs9610560 : Body_mass_index(36581621)(5e−09)
3 . rs2284017 : Height(36224396)(1e−20)
4 . rs6000329 : Body_mass_index(30595370)(5e−09)
Body_mass_index(37280435)(3e−08)
Body_mass_index...(36376304)(3e−08)
1 0.8 0.6 0.4 0.2 0.1
snp / p / or
a . rs2032465 / 9.52e−09 / 0.96
p = 5.0e−08
Observed (−logP)
Recombination rate (cM/Mb)
CACNG2
LOC105373021
37000
37100
Chromosome 22 (kb)
database: gwascatalog.Nov_2024.rp.txt, refGene.txt.Sep_2018.out

1 . rs13409451 :Anxiety_disorder(39294497)(2e−08)
2 . rs35942385 :Alcohol_use_disorder...(37282553)(8e−10)
Opioid_use_disorder...(37156939)(3e−08)
3 . rs10180461 :Educational_attainment(34855049)(2e−17)
Fish−_and_plant−relate...(32066663)(3e−08)
4 . rs10191758 :Cognitive_traits(2e−08)
5 . rs28380327 :Morning_person(30696823)(5e−25)
Chronotype(30696823)(5e−25)
Morning_person(30595370)(1e−10)
6 . rs13024996 :Alcohol_consumption(6e−13)
Alcohol_consumption(31358974)(4e−13)
Problematic_alcohol_us...(32451486)(3e−13)
Drinks_per_week(36477530)(2e−27)
Problematic_alcohol_us...(36301540)(1e−09)
Maximum_habitual_alcoh...(36301540)(9e−09)
Drinks_per_week(36477530)(9e−29)
Alcohol_use_disorder...(37156939)(3e−09)
7 . rs35789697 :Experiencing_mood_swings(29500382)(3e−09)
8 . rs10204230 :Fruit_consumption(32066663)(9e−10)
Intelligence(29942086)(6e−09)
9 . rs10048736 :Neuroticism(29942085)(1e−09)
Science,_technology,_e...(37165452)(1e−09)
10 . rs12991555Highest_math_class...(30038396)(7e−10)
Drinks_per_week(36477530)(3e−25)
..(only the first 10 catalog hits are shown).......

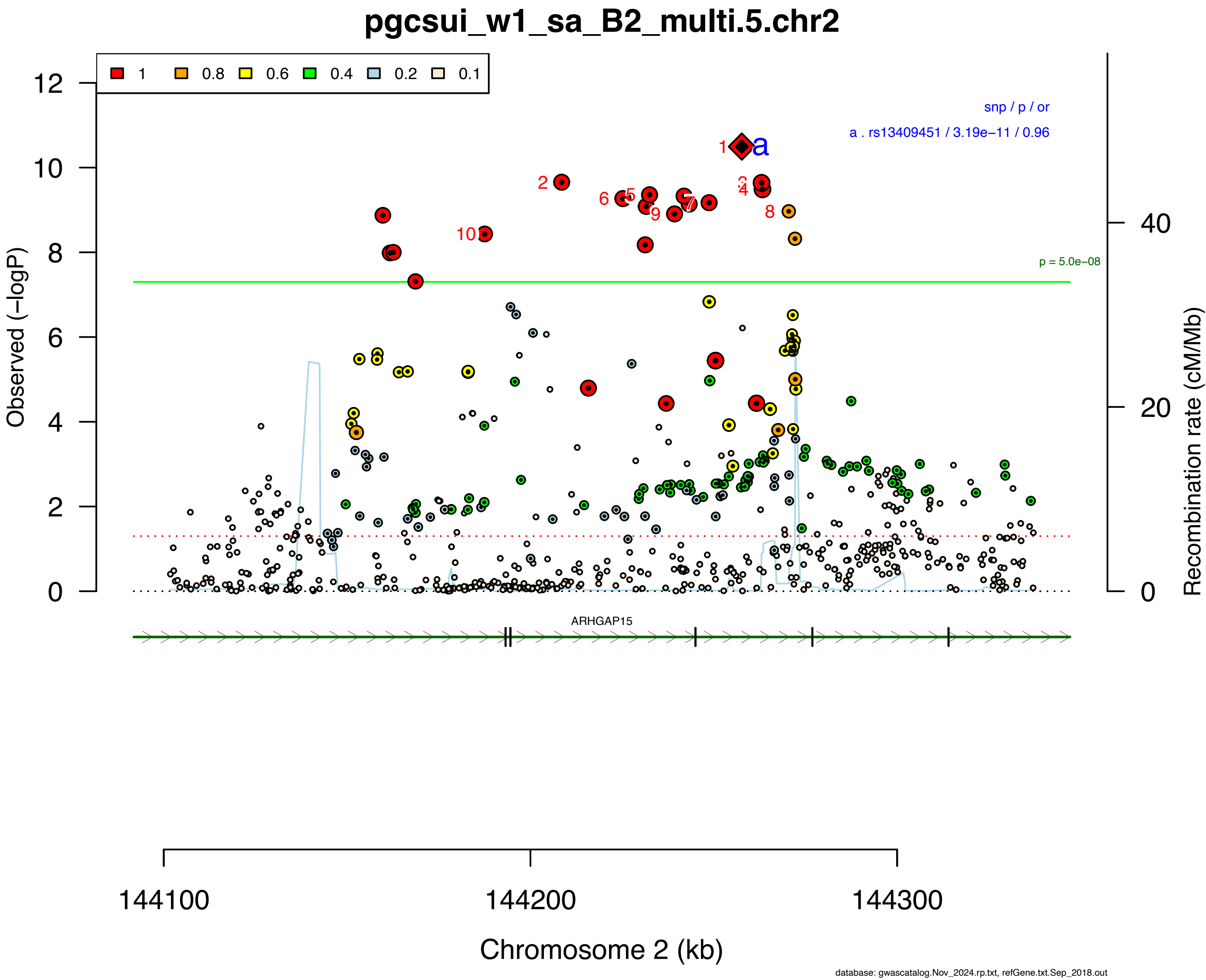

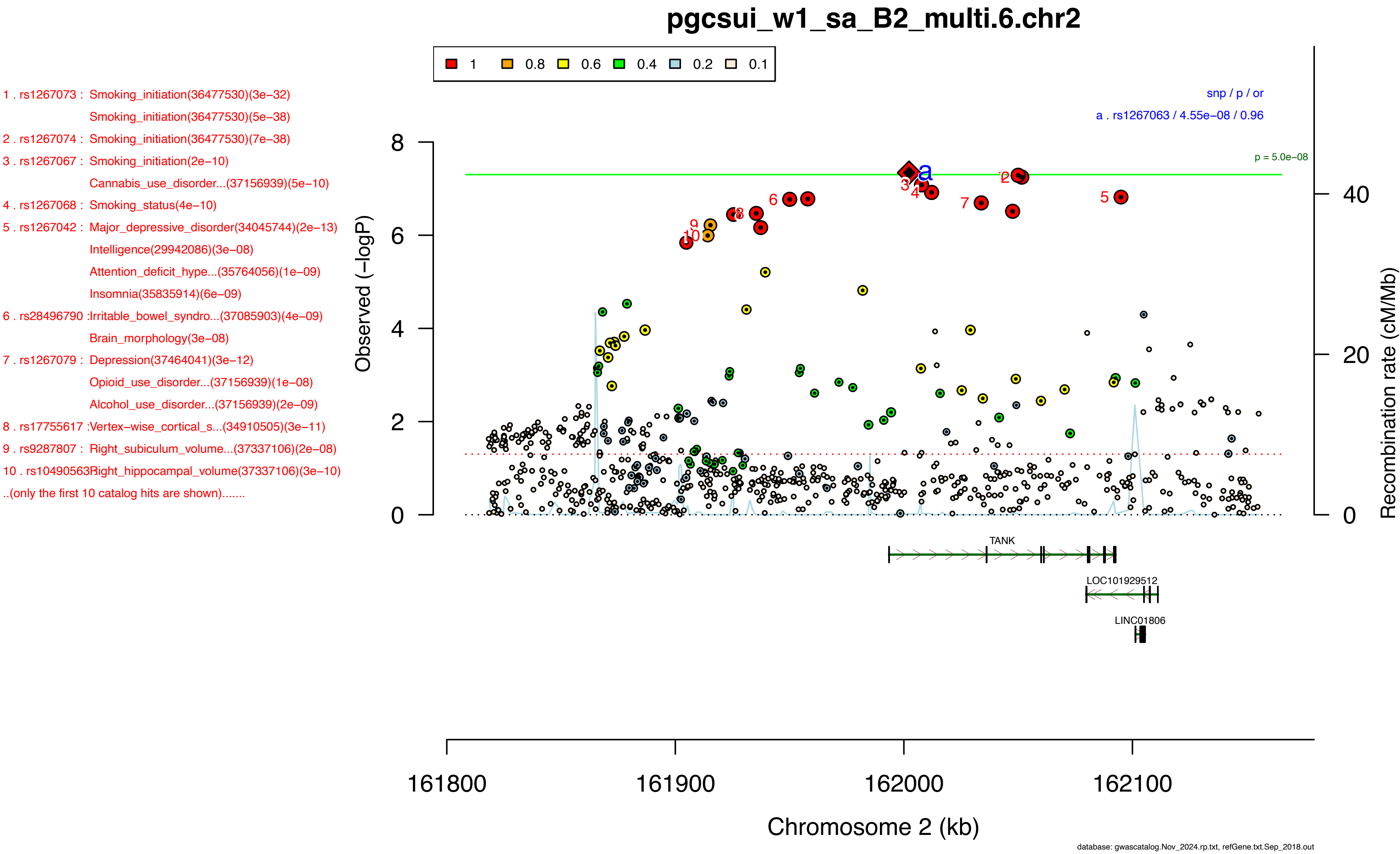
pgcsui_w1_sa_B2_multi.6.chr2
1 0.8 0.6 0.4 0.2 0.1
1 . rs1267073 : Smoking_initiation(36477530)(3e−32)
Smoking_initiation(36477530)(5e−38)
2 . rs1267074 : Smoking_initiation(36477530)(7e−38)
3 . rs1267067 : Smoking_initiation(2e−10)
Cannabis_use_disorder...(37156939)(5e−10)
4 . rs1267068 : Smoking_status(4e−10)
5 . rs1267042 : Major_depressive_disorder(34045744)(2e−13)
Intelligence(29942086)(3e−08)
Attention_deficit_hype...(35764056)(1e−09)
Insomnia(35835914)(6e−09)
6 . rs28496790 :Irritable_bowel_syndro...(37085903)(4e−09)
Brain_morphology(3e−08)
7 . rs1267079 : Depression(37464041)(3e−12)
Opioid_use_disorder...(37156939)(1e−08)
Alcohol_use_disorder...(37156939)(2e−09)
8 . rs17755617 :Vertex-wise_cortical_s...(34910505)(3e−11)
9 . rs9287807 : Right_subiculum_volume...(37337106)(2e−08)
10 . rs10490563Right_hippocampal_volume(37337106)(3e−10)
..(only the first 10 catalog hits are shown).......
snp / p / or
a . rs1267063 / 4.55e−08 / 0.96
p = 5.0e−08
Observed (−logP)
Recombination rate (cM/Mb)
TANK
LOC101929512
LINC01806
161800
161900
162000
162100
Chromosome 2 (kb)
database: gwascatalog.Nov_2024.rp.txt, refGene.txt.Sep_2018.out

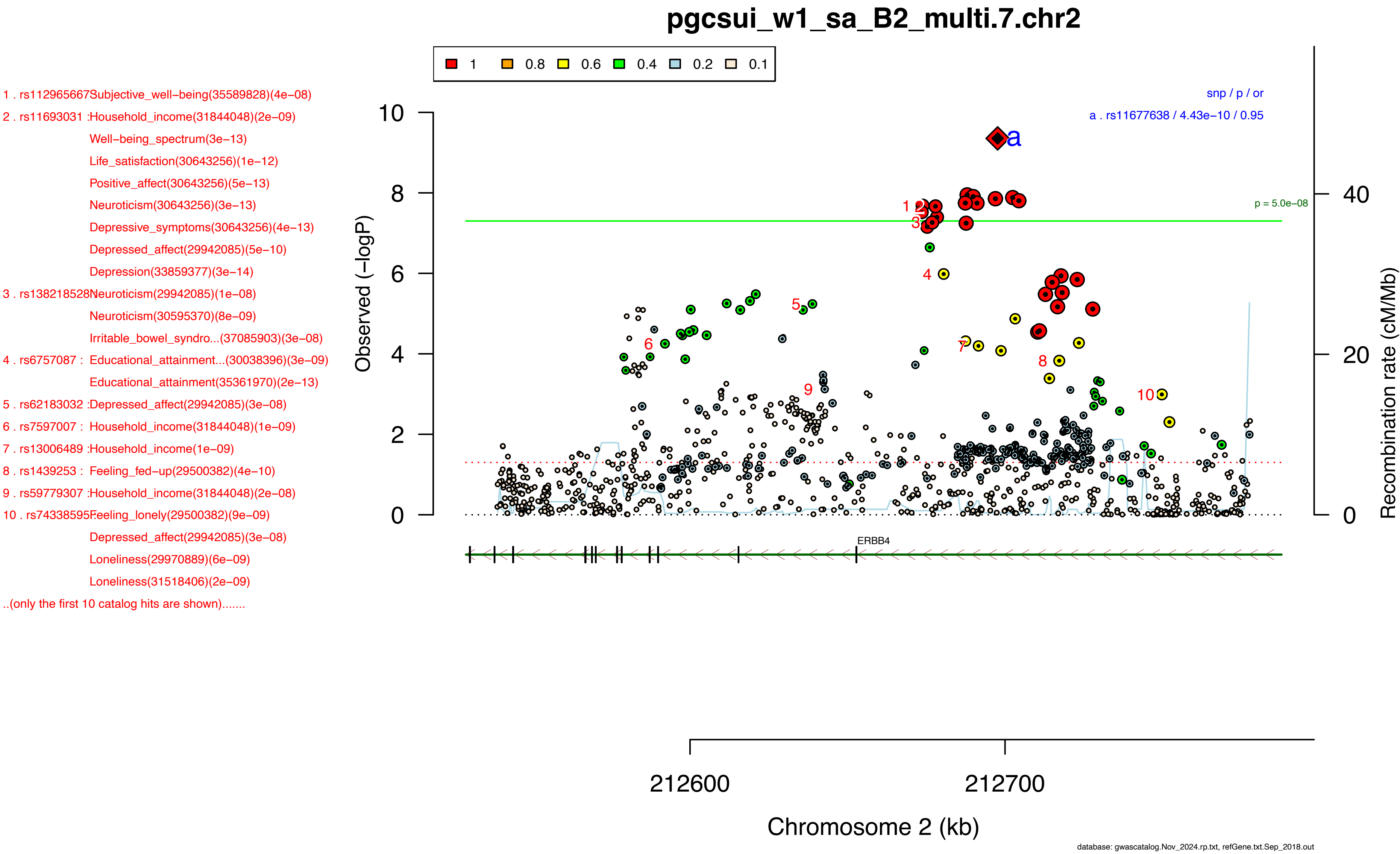
pgcsui_w1_sa_B2_multi.7.chr2
1 0.8 0.6 0.4 0.2 0.1
1 . rs112965667Subjective_well-being(35589828)(4e−08)
2 . rs11693031 :Household_income(31844048)(2e−09)
Well-being_spectrum(3e−13)
Life_satisfaction(30643256)(1e−12)
Positive_affect(30643256)(5e−13)
Neuroticism(30643256)(3e−13)
Depressive_symptoms(30643256)(4e−13)
Depressed_affect(29942085)(5e−10)
Depression(33859377)(3e−14)
3 . rs138218528Neuroticism(29942085)(1e−08)
Neuroticism(30595370)(8e−09)
Irritable_bowel_syndro...(37085903)(3e−08)
4 . rs6757087 : Educational_attainment...(30038396)(3e−09)
Educational_attainment(35361970)(2e−13)
5 . rs62183032 :Depressed_affect(29942085)(3e−08)
6 . rs7597007 : Household_income(31844048)(1e−09)
7 . rs13006489 :Household_income(1e−09)
8 . rs1439253 : Feeling_fed−up(29500382)(4e−10)
9 . rs59779307 :Household_income(31844048)(2e−08)
10 . rs74338595Feeling_lonely(29500382)(9e−09)
Depressed_affect(29942085)(3e−08)
Loneliness(29970889)(6e−09)
Loneliness(31518406)(2e−09)
..(only the first 10 catalog hits are shown).......
snp / p / or
a . rs11677638 / 4.43e−10 / 0.95
p = 5.0e−08
Observed (−logP)
Recombination rate (cM/Mb)
ERBB4
212600
212700
Chromosome 2 (kb)
database: gwascatalog.Nov_2024.rp.txt, refGene.txt.Sep_2018.out

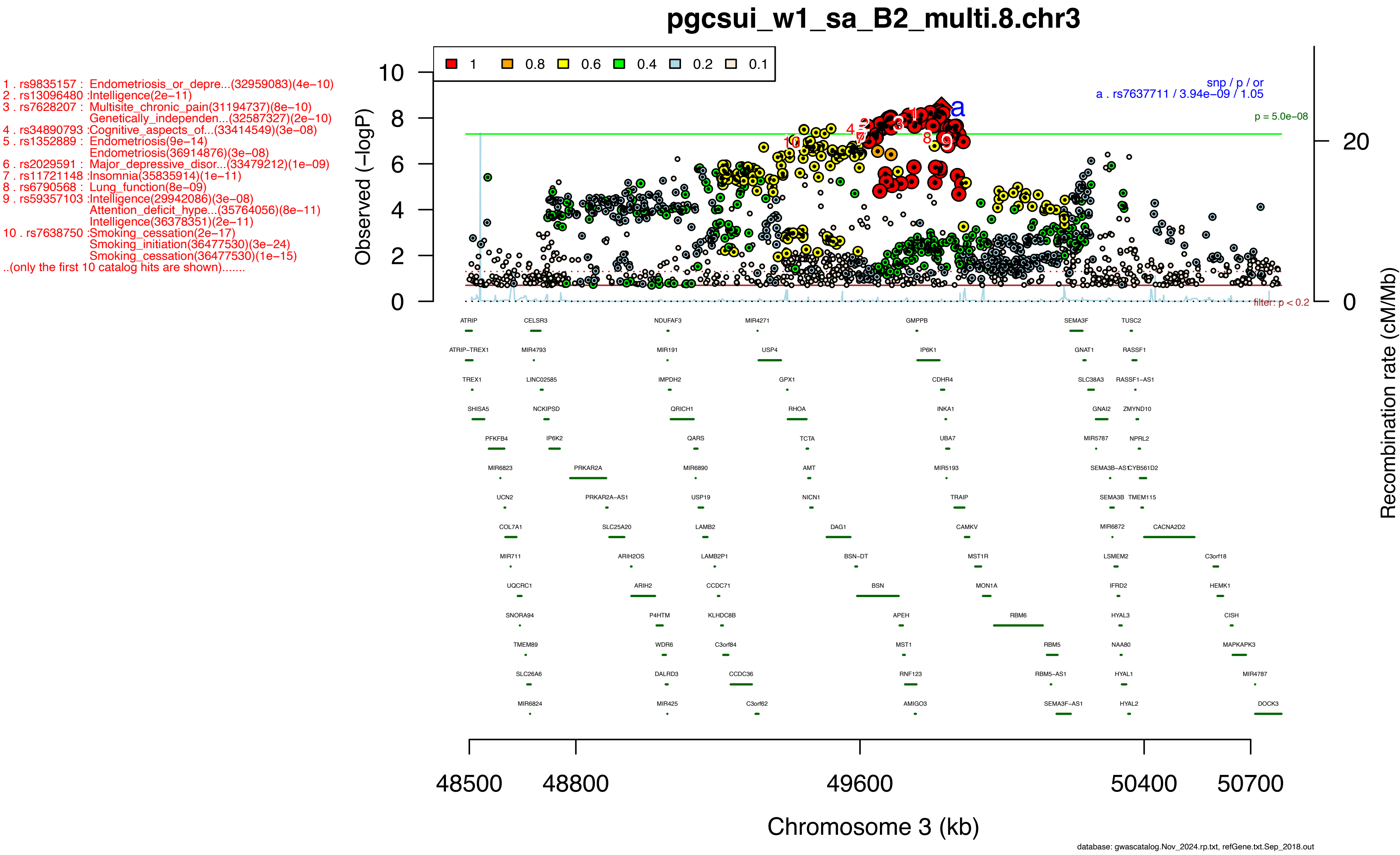
pgcsui_w1_sa_B2_multi.8.chr3
1
0.8
0.6
0.4
0.2
0.1
1 . rs9835157 : Endometriosis_or_depre...(32959083)(4e−10)
2 . rs13096480 :Intelligence(2e−11)
3 . rs7628207 : Multisite_chronic_pain(31194737)(8e−10)
Genetically_independen...(32587327)(2e−10)
4 . rs34890793 :Cognitive_aspects_of...(33414549)(3e−08)
5 . rs1352889 : Endometriosis(9e−14)
Endometriosis(36914876)(3e−08)
6 . rs2029591 : Major_depressive_disor...(33479212)(1e−09)
7 . rs11721148 :Insomnia(35835914)(1e−11)
8 . rs6790568 : Lung_function(8e−09)
9 . rs59357103 :Intelligence(29942086)(3e−08)
Attention_deficit_hype...(35764056)(8e−11)
Intelligence(36378351)(2e−11)
10 . rs7638750 :Smoking_cessation(2e−17)
Smoking_initiation(36477530)(3e−24)
Smoking_cessation(36477530)(1e−15)
..(only the first 10 catalog hits are shown).......
snp / p / or
a . rs7637711 / 3.94e−09 / 1.05
p = 5.0e−08
filter: p < 0.2
Observed (−logP)
Recombination rate (cM/Mb)
ATRIP
ATRIP−TREX1
TREX1
SHISA5
PFKFB4
MIR6823
UCN2
COL7A1
MIR711
UQCRC1
SNORA94
TMEM89
SLC26A6
MIR6824
CELSR3
MIR4793
LINC02585
NCKIPSD
IP6K2
PRKAR2A
PRKAR2A−AS1
SLC25A20
ARIH2OS
ARIH2
P4HTM
WDR6
DALRD3
MIR425
NDUFAF3
MIR191
IMPDH2
QRICH1
QARS
MIR6890
USP19
LAMB2
LAMB2P1
CCDC71
KLHDC8B
C3orf84
CCDC36
C3orf62
MIR4271
USP4
GPX1
RHOA
TCTA
AMT
NICN1
DAG1
BSN−DT
BSN
APEH
MST1
RNF123
AMIGO3
GMPPB
IP6K1
CDHR4
INKA1
UBA7
MIR5193
TRAIP
CAMKV
MST1R
MON1A
RBM6
RBM5
RBM5−AS1
SEMA3F−AS1
SEMA3F
GNAT1
SLC38A3
GNAI2
MIR5787
SEMA3B−AS1
SEMA3B
MIR6872
LSMEM2
IFRD2
HYAL3
NAA80
HYAL1
HYAL2
TUSC2
RASSF1
RASSF1−AS1
ZMYND10
NPRL2
CYB561D2
TMEM115
CACNA2D2
C3orf18
HEMK1
CISH
MAPKAPK3
MIR4787
DOCK3
48500
48800
49600
50400
50700
Chromosome 3 (kb)
database: gwascatalog.Nov_2024.rp.txt, refGene.txt.Sep_2018.out

### pgcsui_w1_sa_B2_multi.9.chr3

1 . rs7647972 : Inguinal_hernia(34382107)(1e−08)
Inguinal_hernia(36584111)(9e−12)
Hernia(36584111)(7e−12)
2 . rs13091322 :Inguinal_hernia(34382107)(3e−08)
3 . rs13058777 :Inguinal_hernia(34382107)(1e−08)
4 . rs4974167 : Inguinal_hernia(34382107)(3e−08)
Inguinal_hernia(35022708)(3e−15)
Inguinal_hernia(35022708)(1e−14)
5 . rs17235410 :Inguinal_hernia(34382107)(3e−08)
6 . rs17235557 :Femoral_neck_width(38477772)(3e−08)
7 . rs4974186 : Heel_bone_mineral...(30048462)(5e−39)
Heel_bone_mineral...(30598549)(2e−29)
8 . rs1352014 : Heel_bone_mineral...(30048462)(3e−38)
9 . rs11130496 :Handedness(3e−08)
10 . rs2316897 :Platelet_count(34469753)(4e−18)
..(only the first 10 catalog hits are shown).......

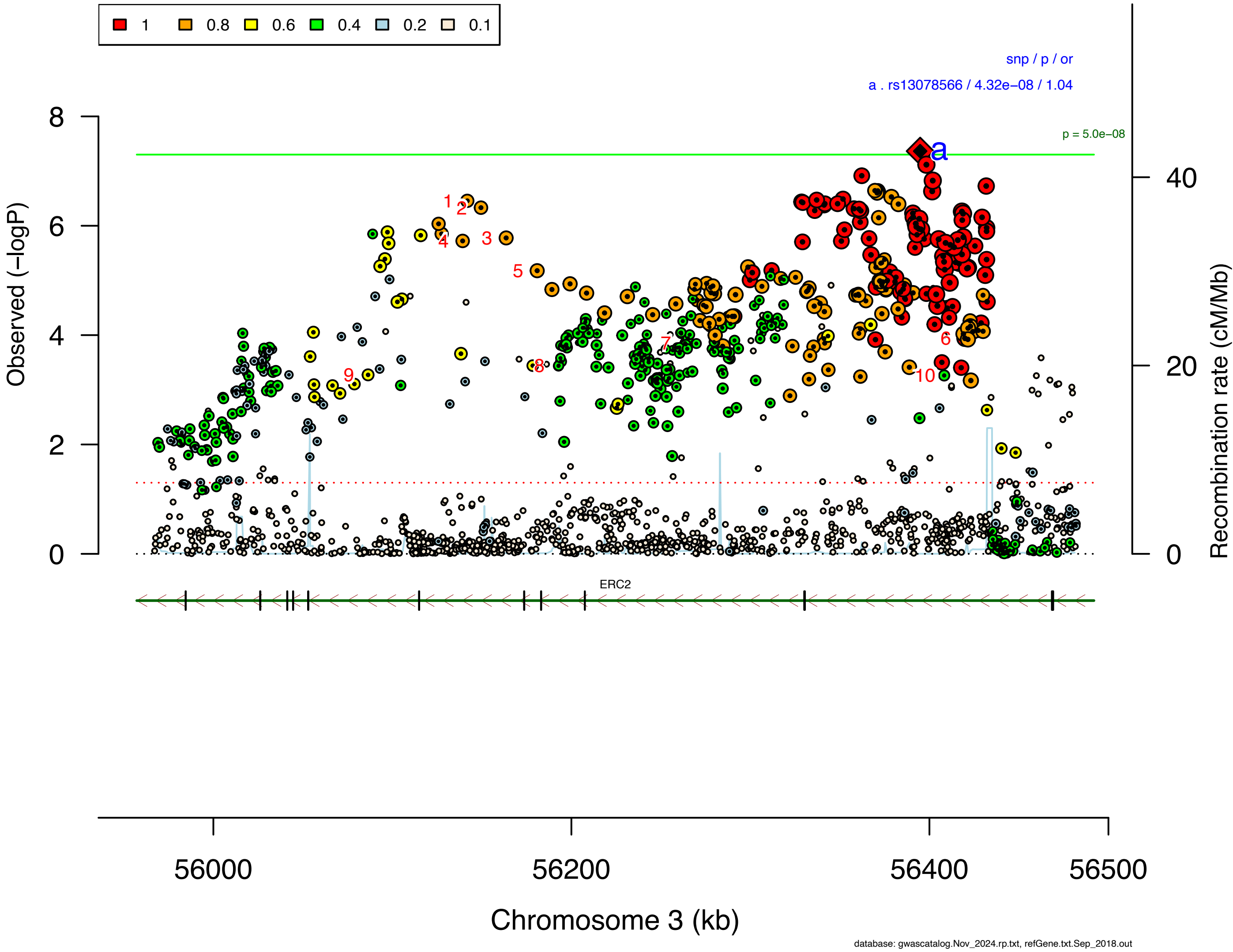

**Supplementary Data 2H: Area plots of lead SNPs at the 46 genome-wide significant loci from the GWAS meta-analysis of suicidal behavior in European ancestry samples.**

If >1 independent SNPs are in the same plot, then each is given a different color, and their LD partners are shaded with the same color. Detailed info about each independent index SNP (a., b., c. ...) is provided in the right upper corner. If SNPs share independent index SNPs as an LD partner, the SNP is assigned to the more significant one. Information regarding other GWAS associations at the locus is provided in the left upper corner.

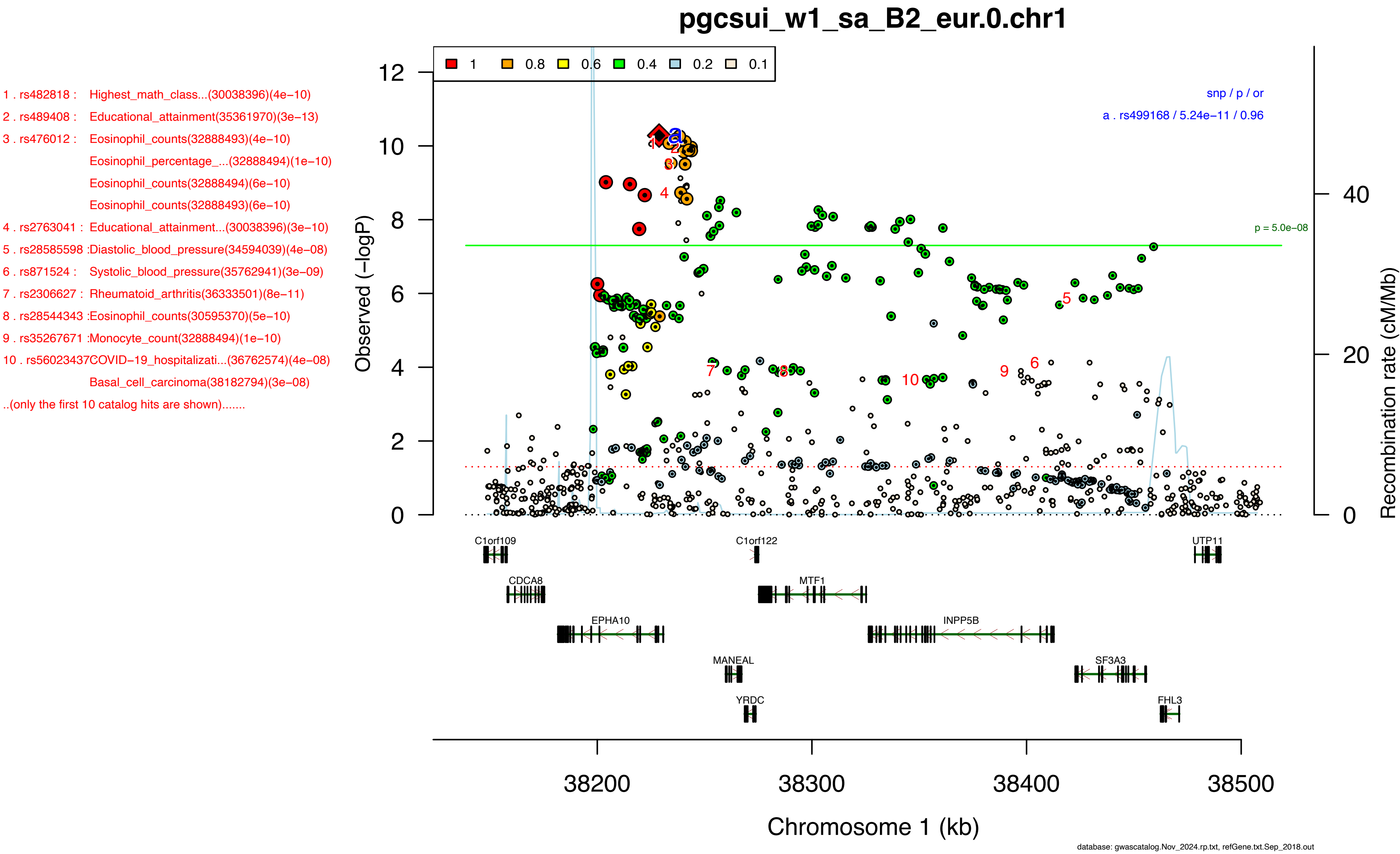

pgcsui_w1_sa_B2_eur.0.chr1
1 . rs482818 : Highest_math_class...(30038396)(4e−10)
2 . rs489408 : Educational_attainment(35361970)(3e−13)
3 . rs476012 : Eosinophil_counts(32888493)(4e−10)
Eosinophil_percentage_...(32888494)(1e−10)
Eosinophil_counts(32888494)(6e−10)
Eosinophil_counts(32888493)(6e−10)
4 . rs2763041 : Educational_attainment...(30038396)(3e−10)
5 . rs28585598 :Diastolic_blood_pressure(34594039)(4e−08)
6 . rs871524 : Systolic_blood_pressure(35762941)(3e−09)
7 . rs2306627 : Rheumatoid_arthritis(36333501)(8e−11)
8 . rs28544343 :Eosinophil_counts(30595370)(5e−10)
9 . rs35267671 :Monocyte_count(32888494)(1e−10)
10 . rs56023437COVID−19_hospitalizati...(36762574)(4e−08)
Basal_cell_carcinoma(38182794)(3e−08)
..(only the first 10 catalog hits are shown).......
1 0.8 0.6 0.4 0.2 0.1
snp / p / or
a . rs499168 / 5.24e−11 / 0.96
p = 5.0e−08
Observed (−logP)
Recombination rate (cM/Mb)
C1orf109
C1orf122
UTP11
CDCA8
MTF1
EPHA10
INPP5B
MANEAL
SF3A3
YRDC
FHL3
38200 38300 38400 38500
Chromosome 1 (kb)
database: gwascatalog.Nov_2024.rp.txt, refGene.txt.Sep_2018.out

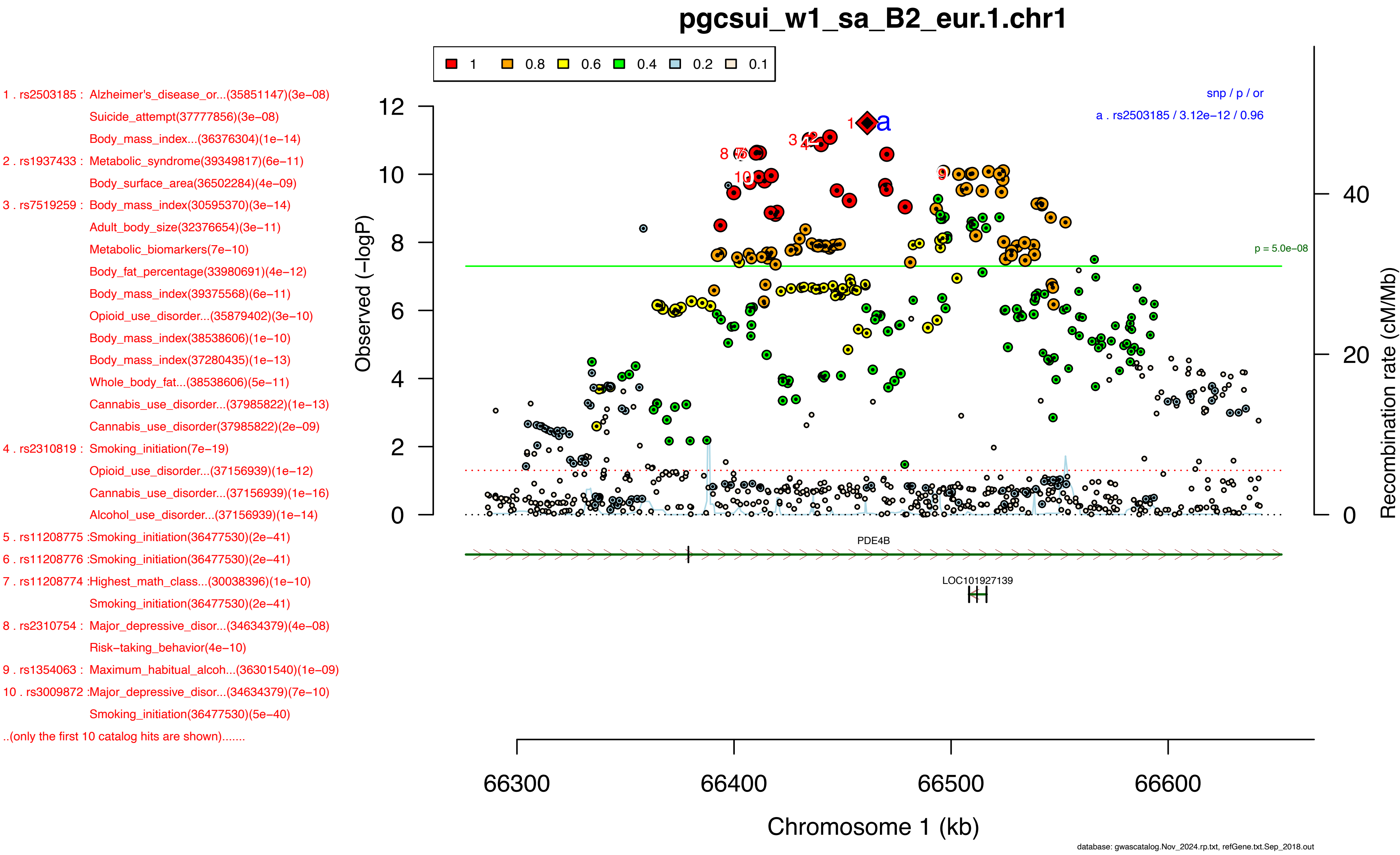
pgcsui_w1_sa_B2_eur.1.chr1
1 0.8 0.6 0.4 0.2 0.1
snp / p / or
a . rs2503185 / 3.12e−12 / 0.96
p = 5.0e−08
Observed (−logP)
Recombination rate (cM/Mb)
Chromosome 1 (kb)
66300 66400 66500 66600
PDE4B
LOC101927139
1 . rs2503185 : Alzheimer's_disease_or...(35851147)(3e−08)
Suicide_attempt(37777856)(3e−08)
Body_mass_index...(36376304)(1e−14)
2 . rs1937433 : Metabolic_syndrome(39349817)(6e−11)
Body_surface_area(36502284)(4e−09)
3 . rs7519259 : Body_mass_index(30595370)(3e−14)
Adult_body_size(32376654)(3e−11)
Metabolic_biomarkers(7e−10)
Body_fat_percentage(33980691)(4e−12)
Body_mass_index(39375568)(6e−11)
Opioid_use_disorder...(35879402)(3e−10)
Body_mass_index(38538606)(1e−10)
Body_mass_index(37280435)(1e−13)
Whole_body_fat...(38538606)(5e−11)
Cannabis_use_disorder...(37985822)(1e−13)
Cannabis_use_disorder(37985822)(2e−09)
4 . rs2310819 : Smoking_initiation(7e−19)
Opioid_use_disorder...(37156939)(1e−12)
Cannabis_use_disorder...(37156939)(1e−16)
Alcohol_use_disorder...(37156939)(1e−14)
5 . rs11208775 :Smoking_initiation(36477530)(2e−41)
6 . rs11208776 :Smoking_initiation(36477530)(2e−41)
7 . rs11208774 :Highest_math_class...(30038396)(1e−10)
Smoking_initiation(36477530)(2e−41)
8 . rs2310754 : Major_depressive_disor...(34634379)(4e−08)
Risk−taking_behavior(4e−10)
9 . rs1354063 : Maximum_habitual_alcoh...(36301540)(1e−09)
10 . rs3009872 :Major_depressive_disor...(34634379)(7e−10)
Smoking_initiation(36477530)(5e−40)
..(only the first 10 catalog hits are shown).......
database: gwascatalog.Nov_2024.rp.txt, refGene.txt.Sep_2018.out

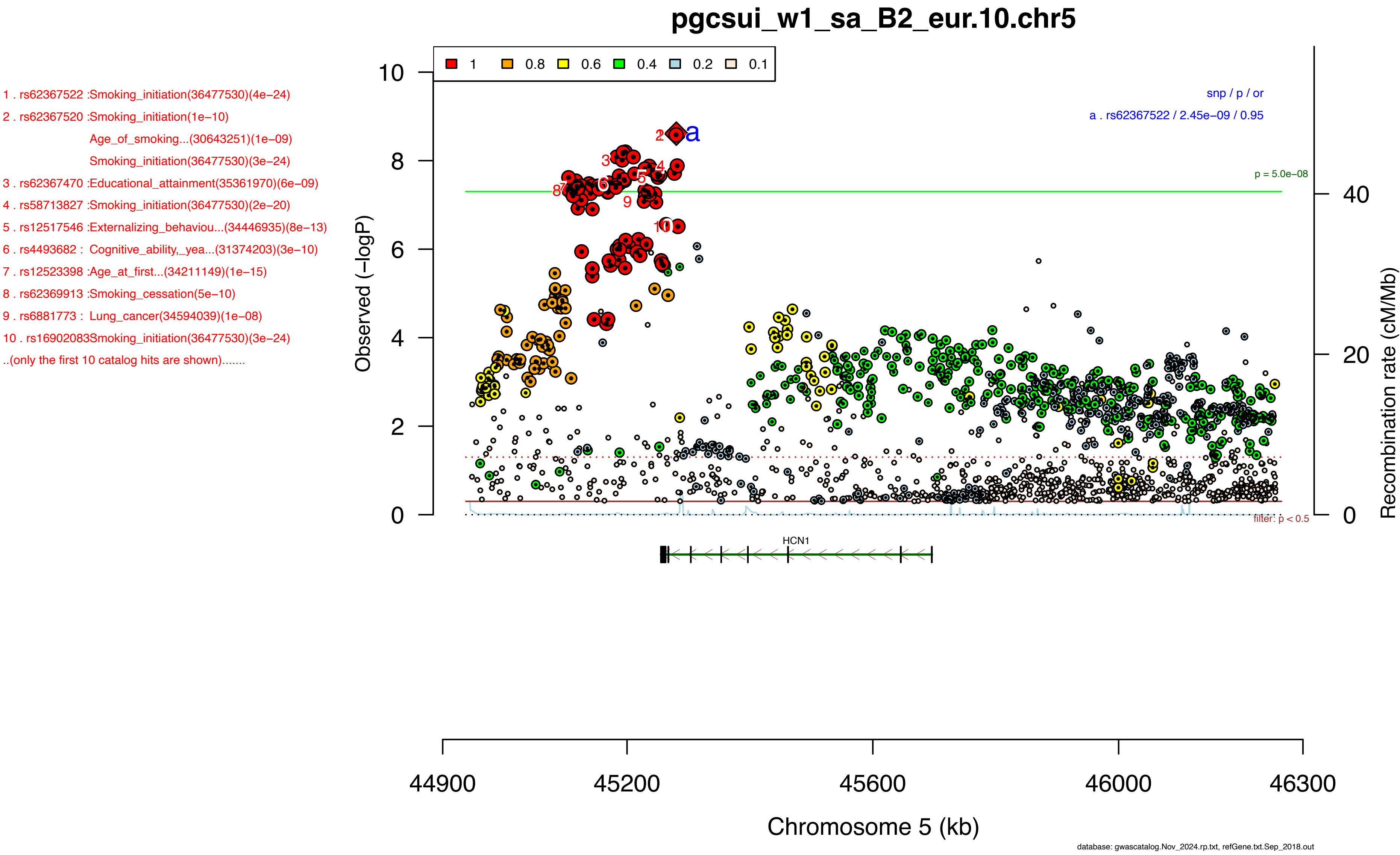
pgcsui_w1_sa_B2_eur.10.chr5
1 . rs62367522 :Smoking_initiation(36477530)(4e−24)
2 . rs62367520 :Smoking_initiation(1e−10)
Age_of_smoking...(30643251)(1e−09)
Smoking_initiation(36477530)(3e−24)
3 . rs62367470 :Educational_attainment(35361970)(6e−09)
4 . rs58713827 :Smoking_initiation(36477530)(2e−20)
5 . rs12517546 :Externalizing_behaviou...(34446935)(8e−13)
6 . rs4493682 : Cognitive_ability,_yea...(31374203)(3e−10)
7 . rs12523398 :Age_at_first...(34211149)(1e−15)
8 . rs62369913 :Smoking_cessation(5e−10)
9 . rs6881773 : Lung_cancer(34594039)(1e−08)
10 . rs16902083Smoking_initiation(36477530)(3e−24)
..(only the first 10 catalog hits are shown).......
1 0.8 0.6 0.4 0.2 0.1
snp / p / or
a . rs62367522 / 2.45e−09 / 0.95
p = 5.0e−08
filter: p < 0.5
Observed (−logP)
Recombination rate (cM/Mb)
HCN1
44900 45200 45600 46000 46300
Chromosome 5 (kb)
database: gwascatalog.Nov_2024.rp.txt, refGene.txt.Sep_2018.out

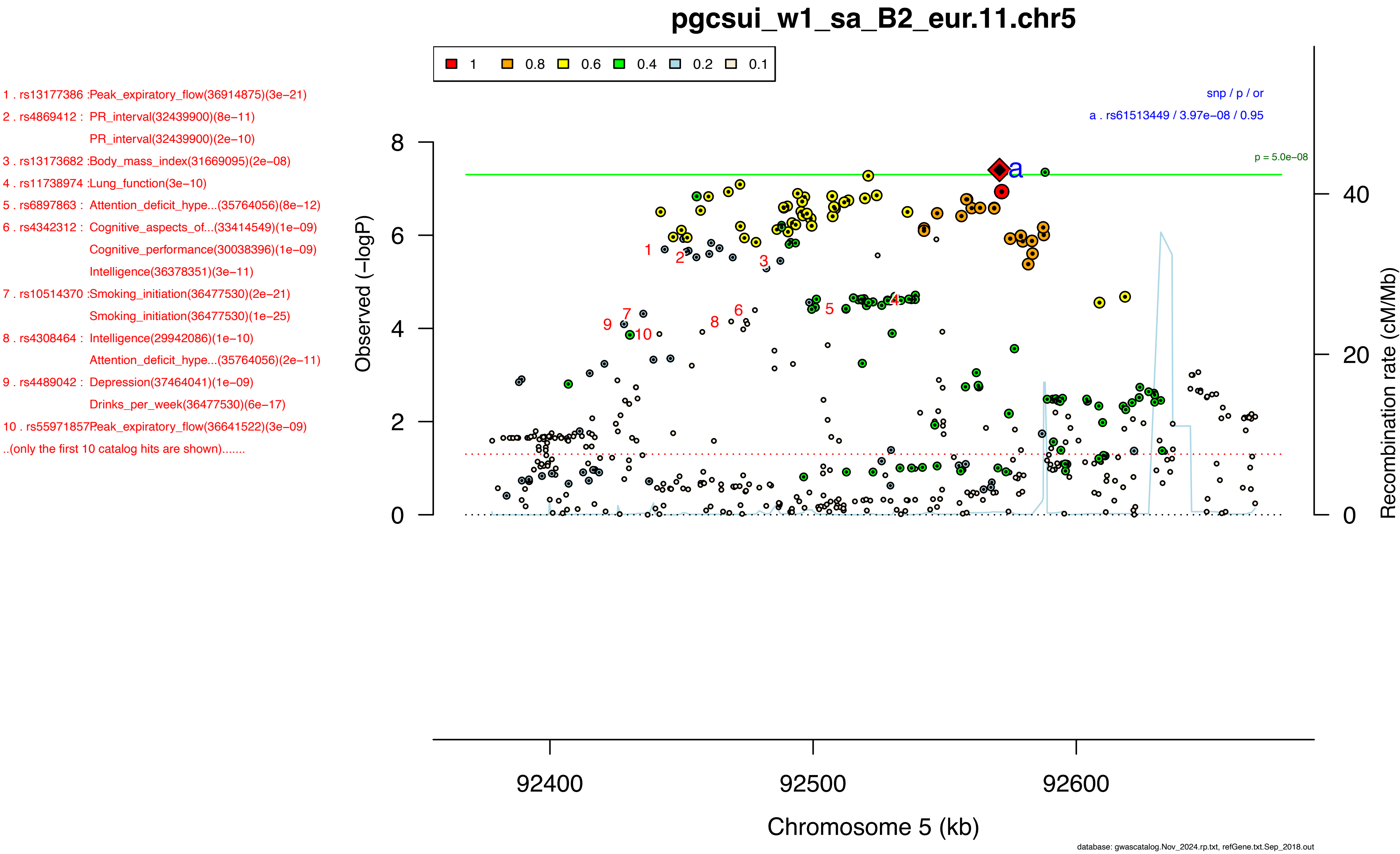

pgcsui_w1_sa_B2_eur.11.chr5
1 0.8 0.6 0.4 0.2 0.1
1 . rs13177386 :Peak_expiratory_flow(36914875)(3e−21)
2 . rs4869412 : PR_interval(32439900)(8e−11)
PR_interval(32439900)(2e−10)
3 . rs13173682 :Body_mass_index(31669095)(2e−08)
4 . rs11738974 :Lung_function(3e−10)
5 . rs6897863 : Attention_deficit_hype...(35764056)(8e−12)
6 . rs4342312 : Cognitive_aspects_of...(33414549)(1e−09)
Cognitive_performance(30038396)(1e−09)
Intelligence(36378351)(3e−11)
7 . rs10514370 :Smoking_initiation(36477530)(2e−21)
Smoking_initiation(36477530)(1e−25)
8 . rs4308464 : Intelligence(29942086)(1e−10)
Attention_deficit_hype...(35764056)(2e−11)
9 . rs4489042 : Depression(37464041)(1e−09)
Drinks_per_week(36477530)(6e−17)
10 . rs55971857Peak_expiratory_flow(36641522)(3e−09)
..(only the first 10 catalog hits are shown).......
snp / p / or
a . rs61513449 / 3.97e−08 / 0.95
p = 5.0e−08
Observed (−logP)
Recombination rate (cM/Mb)
Chromosome 5 (kb)
92400
92500
92600
database: gwascatalog.Nov_2024.rp.txt, refGene.txt.Sep_2018.out

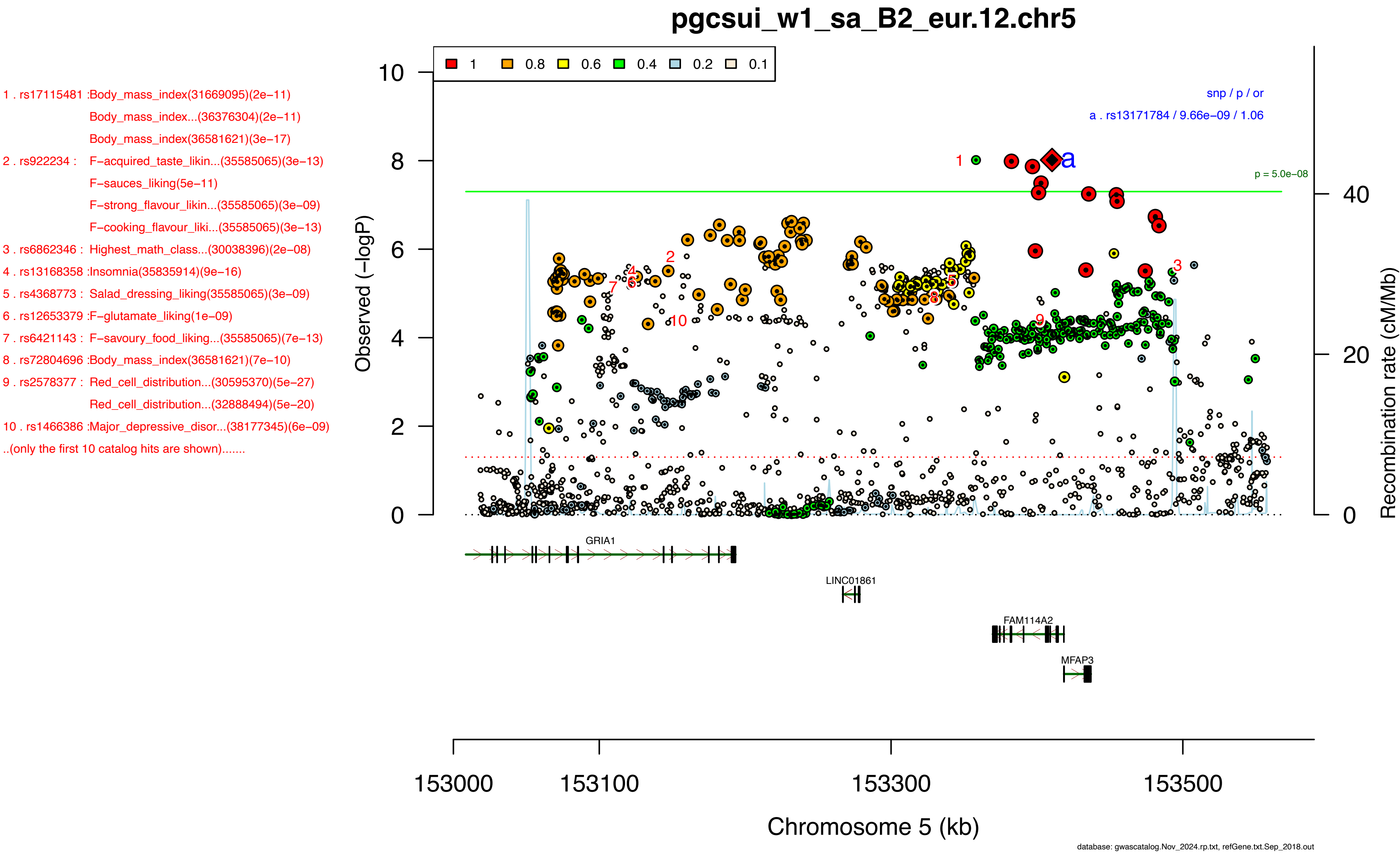
pgcsui_w1_sa_B2_eur.12.chr5
1 . rs17115481 :Body_mass_index(31669095)(2e−11)
Body_mass_index...(36376304)(2e−11)
Body_mass_index(36581621)(3e−17)
2 . rs922234 : F−acquired_taste_likin...(35585065)(3e−13)
F−sauces_liking(5e−11)
F−strong_flavour_likin...(35585065)(3e−09)
F−cooking_flavour_liki...(35585065)(3e−13)
3 . rs6862346 : Highest_math_class...(30038396)(2e−08)
4 . rs13168358 :Insomnia(35835914)(9e−16)
5 . rs4368773 : Salad_dressing_liking(35585065)(3e−09)
6 . rs12653379 :F−glutamate_liking(1e−09)
7 . rs6421143 : F−savoury_food_liking...(35585065)(7e−13)
8 . rs72804696 :Body_mass_index(36581621)(7e−10)
9 . rs2578377 : Red_cell_distribution...(30595370)(5e−27)
Red_cell_distribution...(32888494)(5e−20)
10 . rs1466386 :Major_depressive_disor...(38177345)(6e−09)
..(only the first 10 catalog hits are shown).......
1
0.8
0.6
0.4
0.2
0.1
snp / p / or
a . rs13171784 / 9.66e−09 / 1.06
p = 5.0e−08
Observed (−logP)
Recombination rate (cM/Mb)
GRIA1
LINC01861
FAM114A2
MFAP3
153000
153100
153300
153500
Chromosome 5 (kb)
database: gwascatalog.Nov_2024.rp.txt, refGene.txt.Sep_2018.out

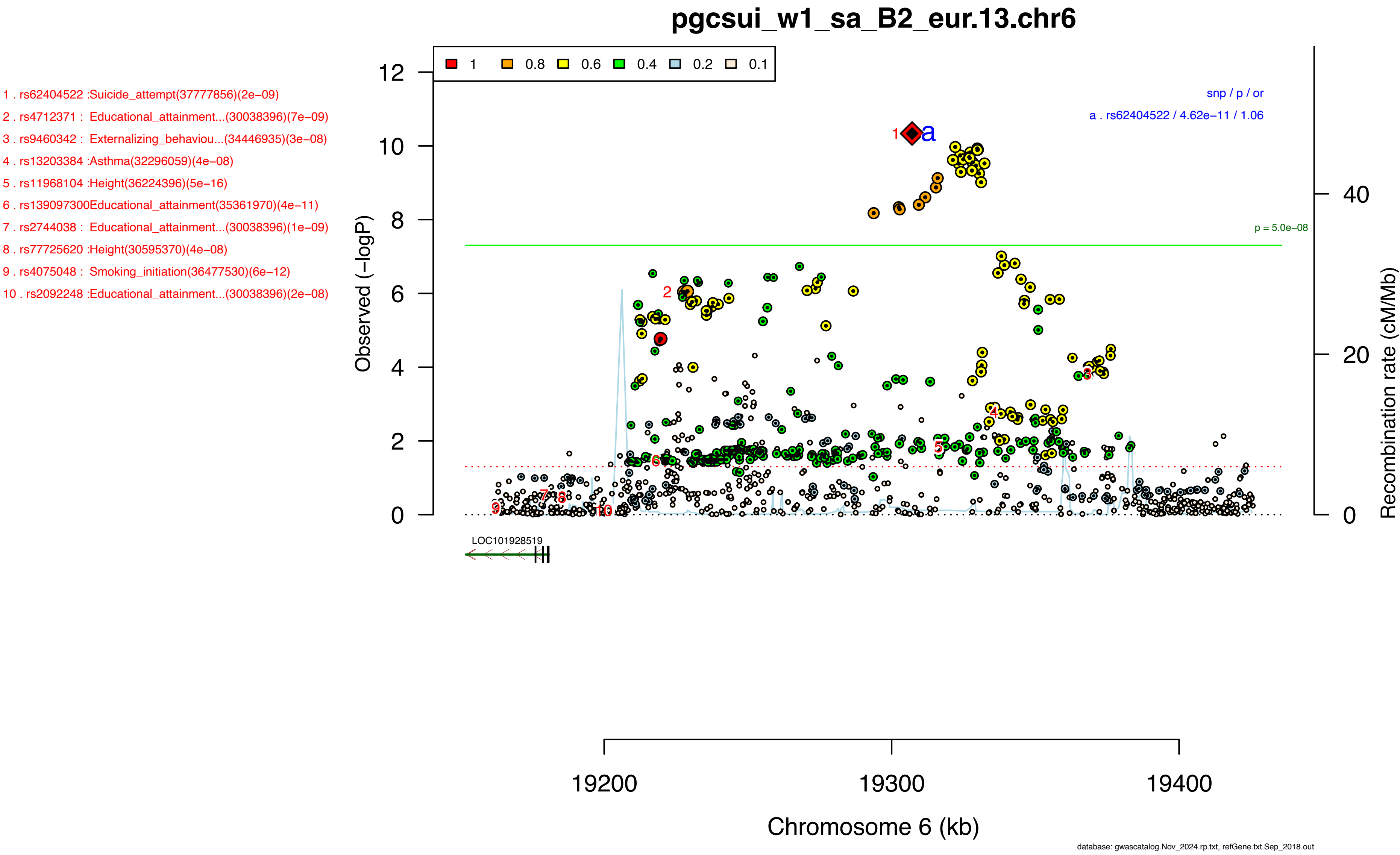

pgcsui_w1_sa_B2_eur.13.chr6
1 . rs62404522 :Suicide_attempt(37777856)(2e−09)
2 . rs4712371 : Educational_attainment...(30038396)(7e−09)
3 . rs9460342 : Externalizing_behaviou...(34446935)(3e−08)
4 . rs13203384 :Asthma(32296059)(4e−08)
5 . rs11968104 :Height(36224396)(5e−16)
6 . rs139097300Educational_attainment(35361970)(4e−11)
7 . rs2744038 : Educational_attainment...(30038396)(1e−09)
8 . rs77725620 :Height(30595370)(4e−08)
9 . rs4075048 : Smoking_initiation(36477530)(6e−12)
10 . rs2092248 :Educational_attainment...(30038396)(2e−08)
1
0.8
0.6
0.4
0.2
0.1
snp / p / or
a . rs62404522 / 4.62e−11 / 1.06
p = 5.0e−08
Observed (−logP)
Recombination rate (cM/Mb)
LOC101928519
19200
19300
19400
Chromosome 6 (kb)
database: gwascatalog.Nov_2024.rp.txt, refGene.txt.Sep_2018.out

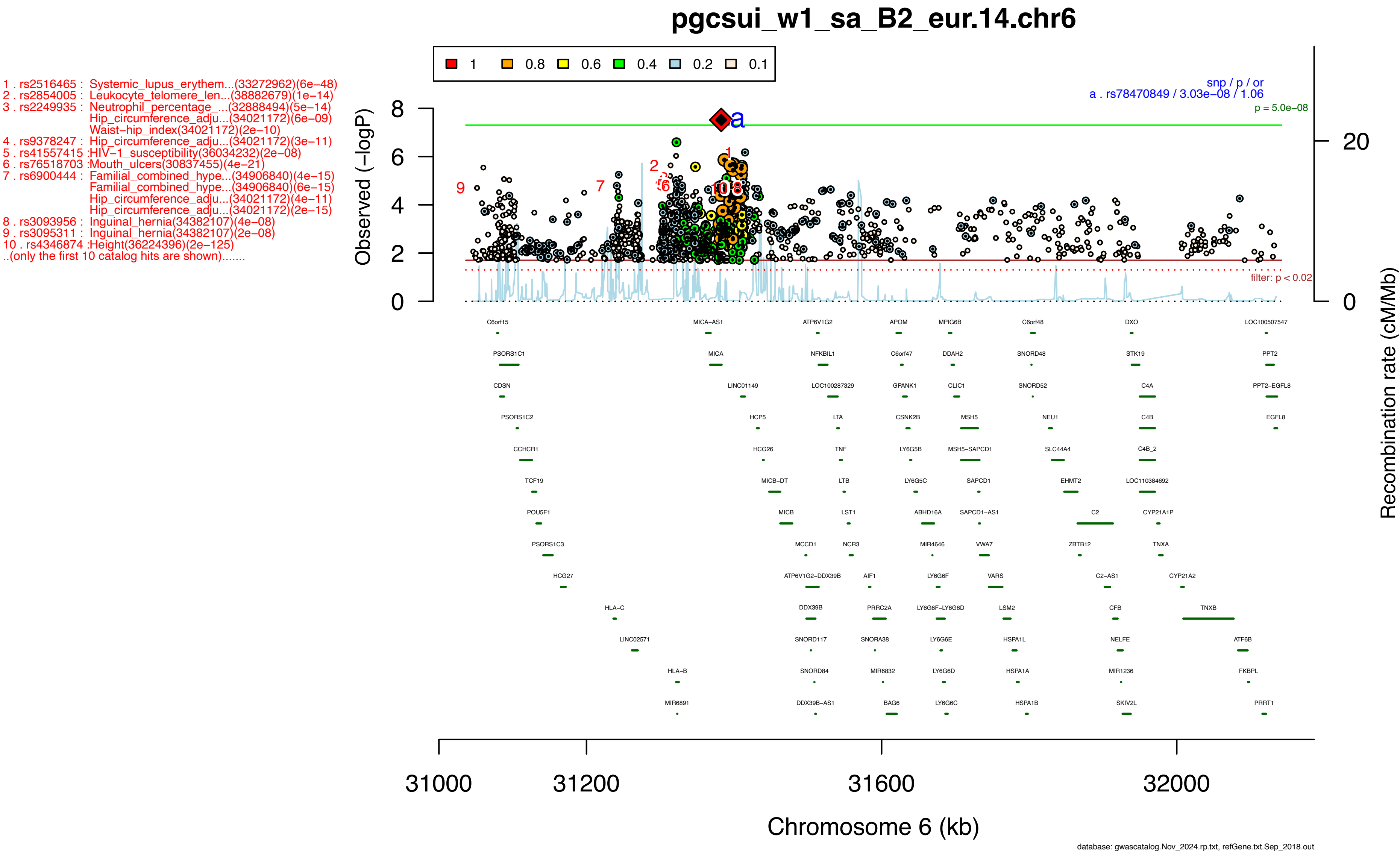

pgcsui_w1_sa_B2_eur.14.chr6
1 . rs2516465 : Systemic_lupus_erythem...(33272962)(6e−48)
2 . rs2854005 : Leukocyte_telomere_len...(38882679)(1e−14)
3 . rs2249935 : Neutrophil_percentage_...(32888494)(5e−14)
Hip_circumference_adju...(34021172)(6e−09)
Waist−hip_index(34021172)(2e−10)
4 . rs9378247 : Hip_circumference_adju...(34021172)(3e−11)
5 . rs41557415 :HIV−1_susceptibility(36034232)(2e−08)
6 . rs76518703 :Mouth_ulcers(30837455)(4e−21)
7 . rs6900444 : Familial_combined_hype...(34906840)(4e−15)
Familial_combined_hype...(34906840)(6e−15)
Hip_circumference_adju...(34021172)(4e−11)
Hip_circumference_adju...(34021172)(2e−15)
8 . rs3093956 : Inguinal_hernia(34382107)(4e−08)
9 . rs3095311 : Inguinal_hernia(34382107)(2e−08)
10 . rs4346874 :Height(36224396)(2e−125)
..(only the first 10 catalog hits are shown).......
1 0.8 0.6 0.4 0.2 0.1
snp / p / or
a . rs78470849 / 3.03e−08 / 1.06
p = 5.0e−08
filter: p < 0.02
Observed (−logP)
Recombination rate (cM/Mb)
C6orf15 MICA−AS1 ATP6V1G2 APOM MPIG6B C6orf48 DXO LOC100507547
PSORS1C1 MICA NFKBIL1 C6orf47 DDAH2 SNORD48 STK19 PPT2
CDSN LINC01149 LOC100287329 GPANK1 CLIC1 SNORD52 C4A PPT2−EGFL8
PSORS1C2 HCP5 LTA CSNK2B MSH5 NEU1 C4B EGFL8
CCHCR1 HCG26 TNF LY6G5B MSH5−SAPCD1 SLC44A4 C4B_2
TCF19 MICB−DT LTB LY6G5C SAPCD1 EHMT2 LOC110384692
POU5F1 MICB LST1 ABHD16A SAPCD1−AS1 C2 CYP21A1P
PSORS1C3 MCCD1 NCR3 MIR4646 VWA7 ZBTB12 TNXA
HCG27 ATP6V1G2−DDX39B AIF1 LY6G6F VARS C2−AS1 CYP21A2
HLA−C DDX39B PRRC2A LY6G6F−LY6G6D LSM2 CFB TNXB
LINC02571 SNORD117 SNORA38 LY6G6E HSPA1L NELFE ATF6B
HLA−B SNORD84 MIR6832 LY6G6D HSPA1A MIR1236 FKBPL
MIR6891 DDX39B−AS1 BAG6 LY6G6C HSPA1B SKIV2L PRRT1
31000 31200 31600 32000
Chromosome 6 (kb)
database: gwascatalog.Nov_2024.rp.txt, refGene.txt.Sep_2018.out

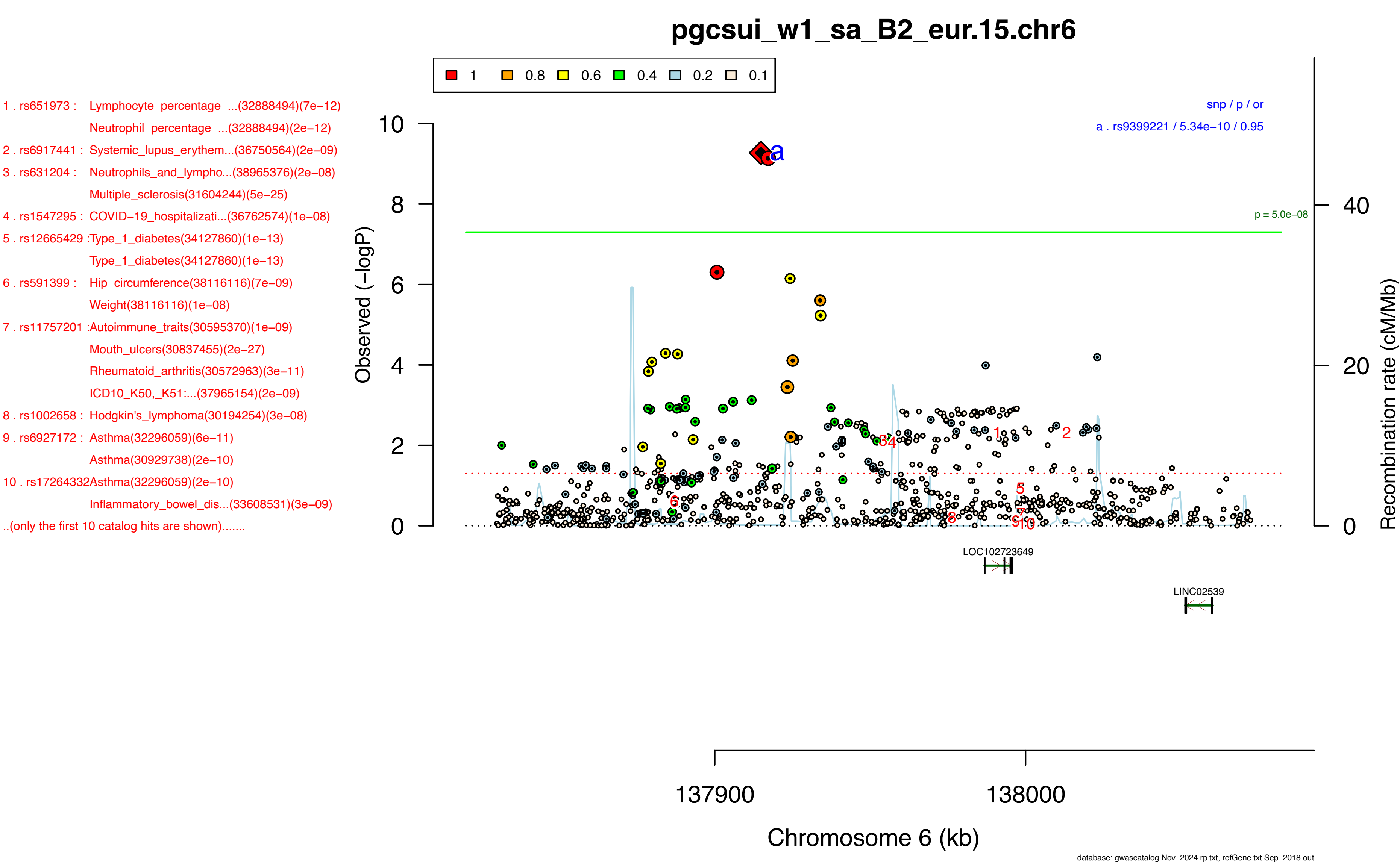
pgcsui_w1_sa_B2_eur.15.chr6
1 0.8 0.6 0.4 0.2 0.1
1 . rs651973 : Lymphocyte_percentage_...(32888494)(7e−12)
Neutrophil_percentage_...(32888494)(2e−12)
2 . rs6917441 : Systemic_lupus_erythem...(36750564)(2e−09)
3 . rs631204 : Neutrophils_and_lympho...(38965376)(2e−08)
Multiple_sclerosis(31604244)(5e−25)
4 . rs1547295 : COVID−19_hospitalizati...(36762574)(1e−08)
5 . rs12665429 :Type_1_diabetes(34127860)(1e−13)
Type_1_diabetes(34127860)(1e−13)
6 . rs591399 : Hip_circumference(38116116)(7e−09)
Weight(38116116)(1e−08)
7 . rs11757201 :Autoimmune_traits(30595370)(1e−09)
Mouth_ulcers(30837455)(2e−27)
Rheumatoid_arthritis(30572963)(3e−11)
ICD10_K50,_K51:...(37965154)(2e−09)
8 . rs1002658 : Hodgkin's_lymphoma(30194254)(3e−08)
9 . rs6927172 : Asthma(32296059)(6e−11)
Asthma(30929738)(2e−10)
10 . rs17264332Asthma(32296059)(2e−10)
Inflammatory_bowel_dis...(33608531)(3e−09)
..(only the first 10 catalog hits are shown).......
snp / p / or
a . rs9399221 / 5.34e−10 / 0.95
p = 5.0e−08
Observed (−logP)
Recombination rate (cM/Mb)
LOC102723649
LINC02539
137900
138000
Chromosome 6 (kb)
database: gwascatalog.Nov_2024.rp.txt, refGene.txt.Sep_2018.out

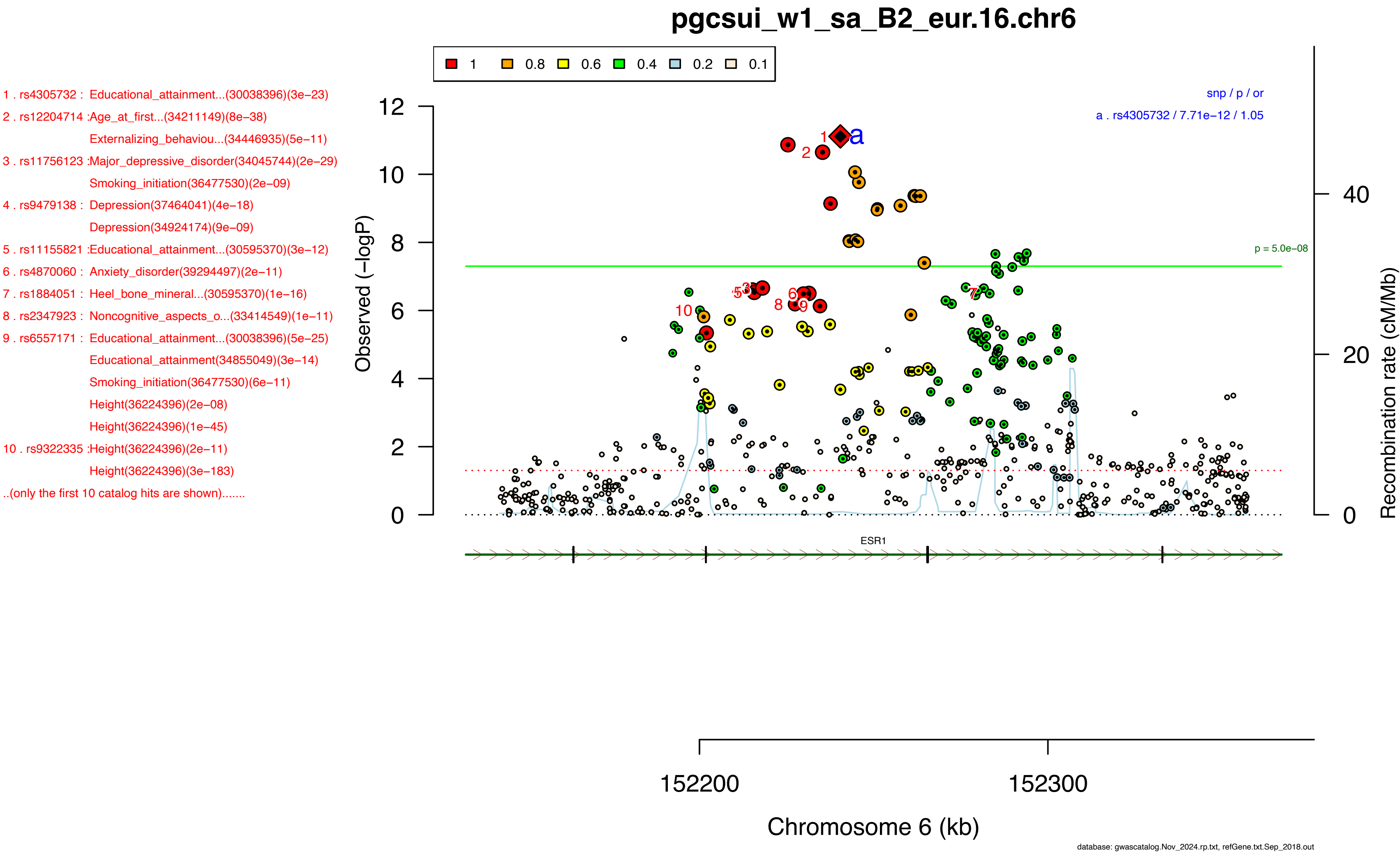
pgcsui_w1_sa_B2_eur.16.chr6
1 0.8 0.6 0.4 0.2 0.1
1 . rs4305732 : Educational_attainment...(30038396)(3e−23)
2 . rs12204714 :Age_at_first...(34211149)(8e−38)
Externalizing_behaviou...(34446935)(5e−11)
3 . rs11756123 :Major_depressive_disorder(34045744)(2e−29)
Smoking_initiation(36477530)(2e−09)
4 . rs9479138 : Depression(37464041)(4e−18)
Depression(34924174)(9e−09)
5 . rs11155821 :Educational_attainment...(30595370)(3e−12)
6 . rs4870060 : Anxiety_disorder(39294497)(2e−11)
7 . rs1884051 : Heel_bone_mineral...(30595370)(1e−16)
8 . rs2347923 : Noncognitive_aspects_o...(33414549)(1e−11)
9 . rs6557171 : Educational_attainment...(30038396)(5e−25)
Educational_attainment(34855049)(3e−14)
Smoking_initiation(36477530)(6e−11)
Height(36224396)(2e−08)
Height(36224396)(1e−45)
10 . rs9322335 :Height(36224396)(2e−11)
Height(36224396)(3e−183)
..(only the first 10 catalog hits are shown).......
snp / p / or
a . rs4305732 / 7.71e−12 / 1.05
p = 5.0e−08
Observed (−logP)
Recombination rate (cM/Mb)
ESR1
152200
152300
Chromosome 6 (kb)
database: gwascatalog.Nov_2024.rp.txt, refGene.txt.Sep_2018.out

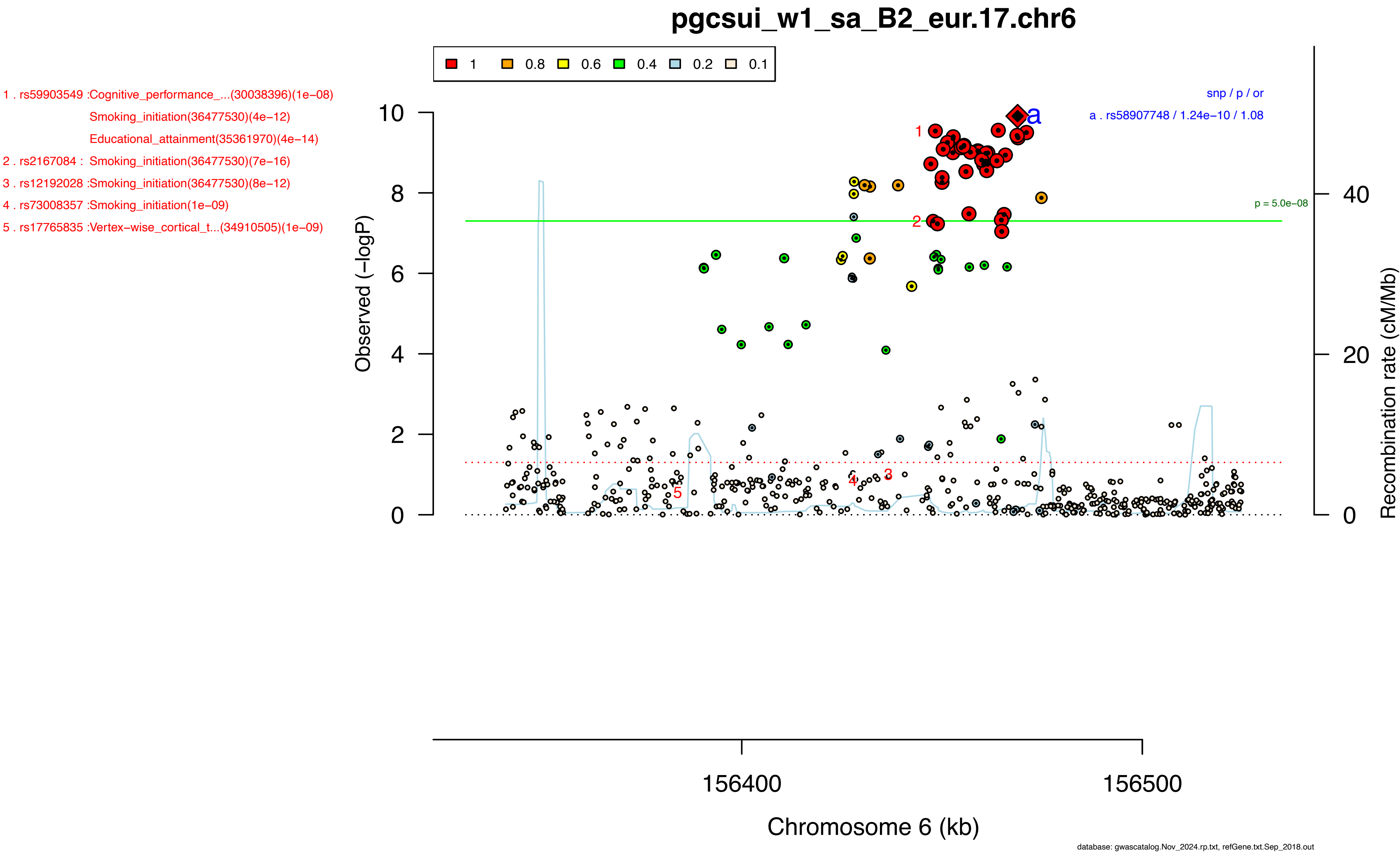
pgcsui_w1_sa_B2_eur.17.chr6
1 0.8 0.6 0.4 0.2 0.1
1 . rs59903549 :Cognitive_performance_...(30038396)(1e−08)
Smoking_initiation(36477530)(4e−12)
Educational_attainment(35361970)(4e−14)
2 . rs2167084 : Smoking_initiation(36477530)(7e−16)
3 . rs12192028 :Smoking_initiation(36477530)(8e−12)
4 . rs73008357 :Smoking_initiation(1e−09)
5 . rs17765835 :Vertex−wise_cortical_t...(34910505)(1e−09)
snp / p / or
a . rs58907748 / 1.24e−10 / 1.08
p = 5.0e−08
Observed (−logP)
Recombination rate (cM/Mb)
156400
156500
Chromosome 6 (kb)
database: gwascatalog.Nov_2024.rp.txt, refGene.txt.Sep_2018.out

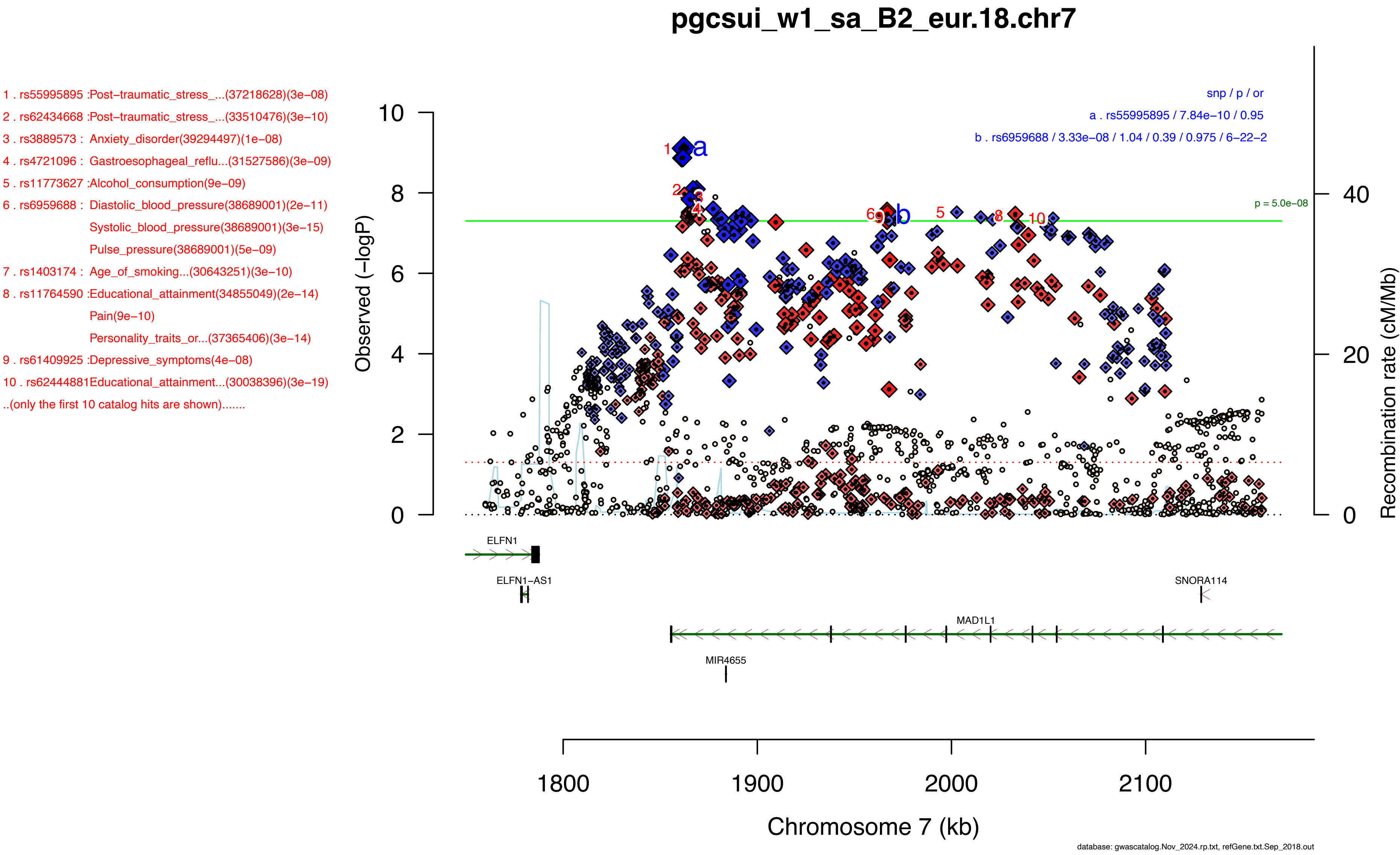
pgcsui_w1_sa_B2_eur.18.chr7
1 . rs55995895 :Post−traumatic_stress_...(37218628)(3e−08)
2 . rs62434668 :Post−traumatic_stress_...(33510476)(3e−10)
3 . rs3889573 : Anxiety_disorder(39294497)(1e−08)
4 . rs4721096 : Gastroesophageal_reflu...(31527586)(3e−09)
5 . rs11773627 :Alcohol_consumption(9e−09)
6 . rs6959688 : Diastolic_blood_pressure(38689001)(2e−11)
Systolic_blood_pressure(38689001)(3e−15)
Pulse_pressure(38689001)(5e−09)
7 . rs1403174 : Age_of_smoking...(30643251)(3e−10)
8 . rs11764590 :Educational_attainment(34855049)(2e−14)
Pain(9e−10)
Personality_traits_or...(37365406)(3e−14)
9 . rs61409925 :Depressive_symptoms(4e−08)
10 . rs62444881Educational_attainment...(30038396)(3e−19)
..(only the first 10 catalog hits are shown).......
snp / p / or
a . rs55995895 / 7.84e−10 / 0.95
b . rs6959688 / 3.33e−08 / 1.04 / 0.39 / 0.975 / 6−22−2
p = 5.0e−08
Observed (−logP)
Recombination rate (cM/Mb)
ELFN1
ELFN1−AS1
SNORA114
MAD1L1
MIR4655
1800
1900
2000
2100
Chromosome 7 (kb)
database: gwascatalog.Nov_2024.rp.txt, refGene.txt.Sep_2018.out

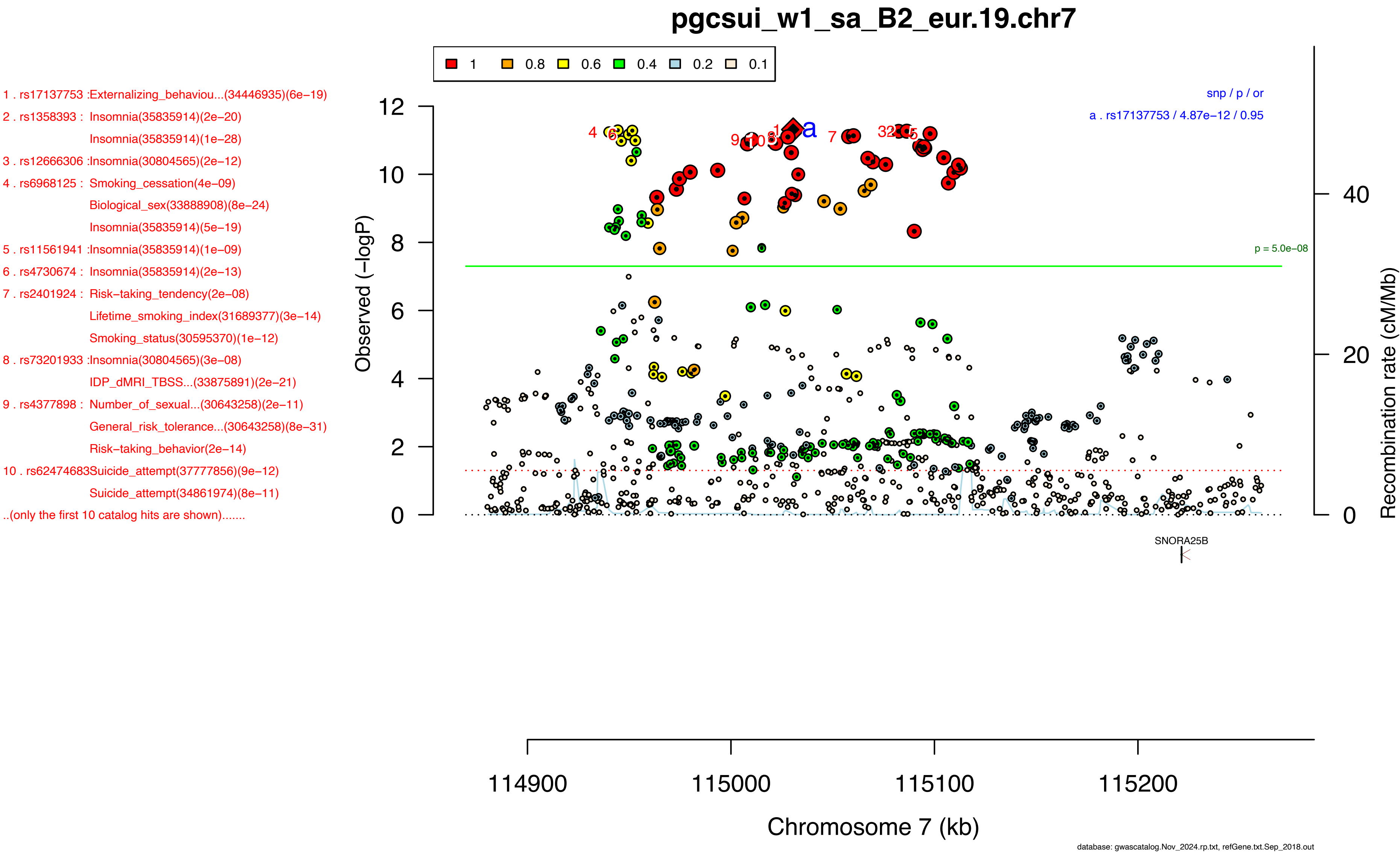

pgcsui_w1_sa_B2_eur.19.chr7
1 . rs17137753 :Externalizing_behaviou...(34446935)(6e−19)
2 . rs1358393 : Insomnia(35835914)(2e−20)
Insomnia(35835914)(1e−28)
3 . rs12666306 :Insomnia(30804565)(2e−12)
4 . rs6968125 : Smoking_cessation(4e−09)
Biological_sex(33888908)(8e−24)
Insomnia(35835914)(5e−19)
5 . rs11561941 :Insomnia(35835914)(1e−09)
6 . rs4730674 : Insomnia(35835914)(2e−13)
7 . rs2401924 : Risk−taking_tendency(2e−08)
Lifetime_smoking_index(31689377)(3e−14)
Smoking_status(30595370)(1e−12)
8 . rs73201933 :Insomnia(30804565)(3e−08)
IDP_dMRI_TBSS...(33875891)(2e−21)
9 . rs4377898 : Number_of_sexual...(30643258)(2e−11)
General_risk_tolerance...(30643258)(8e−31)
Risk−taking_behavior(2e−14)
10 . rs62474683Suicide_attempt(37777856)(9e−12)
Suicide_attempt(34861974)(8e−11)
..(only the first 10 catalog hits are shown).......
1 0.8 0.6 0.4 0.2 0.1
snp / p / or
a . rs17137753 / 4.87e−12 / 0.95
p = 5.0e−08
Observed (−logP)
Recombination rate (cM/Mb)
SNORA25B
114900 115000 115100 115200
Chromosome 7 (kb)
database: gwascatalog.Nov_2024.rp.txt, refGene.txt.Sep_2018.out

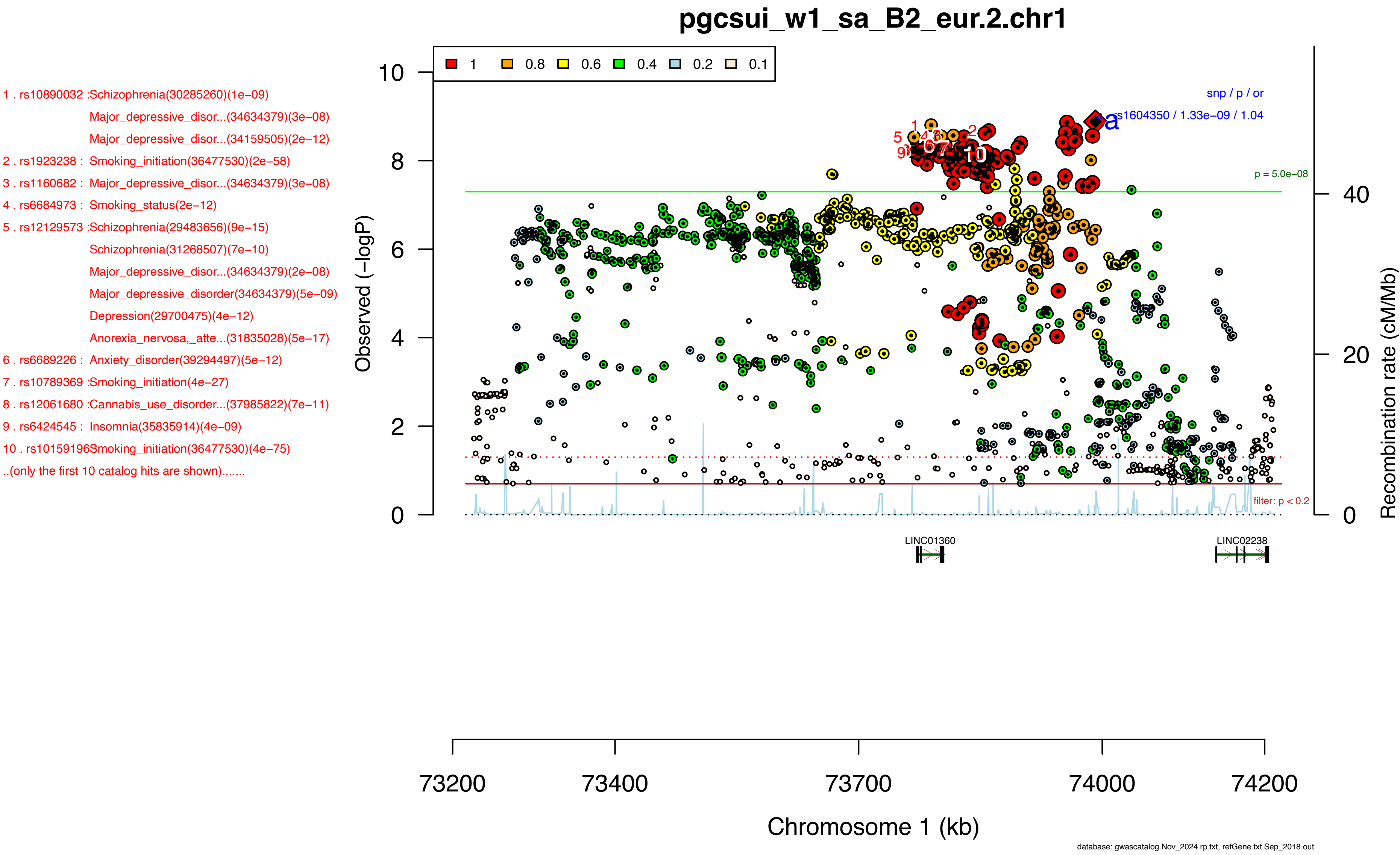
pgcsui_w1_sa_B2_eur.2.chr1
1 . rs10890032 :Schizophrenia(30285260)(1e–09)
Major_depressive_disor...(34634379)(3e–08)
Major_depressive_disor...(34159505)(2e–12)
2 . rs1923238 : Smoking_initiation(36477530)(2e–58)
3 . rs1160682 : Major_depressive_disor...(34634379)(3e–08)
4 . rs6684973 : Smoking_status(2e–12)
5 . rs12129573 :Schizophrenia(29483656)(9e–15)
Schizophrenia(31268507)(7e–10)
Major_depressive_disor...(34634379)(2e–08)
Major_depressive_disorder(34634379)(5e–09)
Depression(29700475)(4e–12)
Anorexia_nervosa,_atte...(31835028)(5e–17)
6 . rs6689226 : Anxiety_disorder(39294497)(5e–12)
7 . rs10789369 :Smoking_initiation(4e–27)
8 . rs12061680 :Cannabis_use_disorder...(37985822)(7e–11)
9 . rs6424545 : Insomnia(35835914)(4e–09)
10 . rs10159196Smoking_initiation(36477530)(4e–75)
..(only the first 10 catalog hits are shown).......
1
0.8
0.6
0.4
0.2
0.1
snp / p / or
rs1604350 / 1.33e–09 / 1.04
a
p = 5.0e–08
filter: p < 0.2
Observed (–logP)
Recombination rate (cM/Mb)
LINC01360
LINC02238
Chromosome 1 (kb)
73200
73400
73700
74000
74200
database: gwascatalog.Nov_2024.rp.txt, refGene.txt.Sep_2018.out

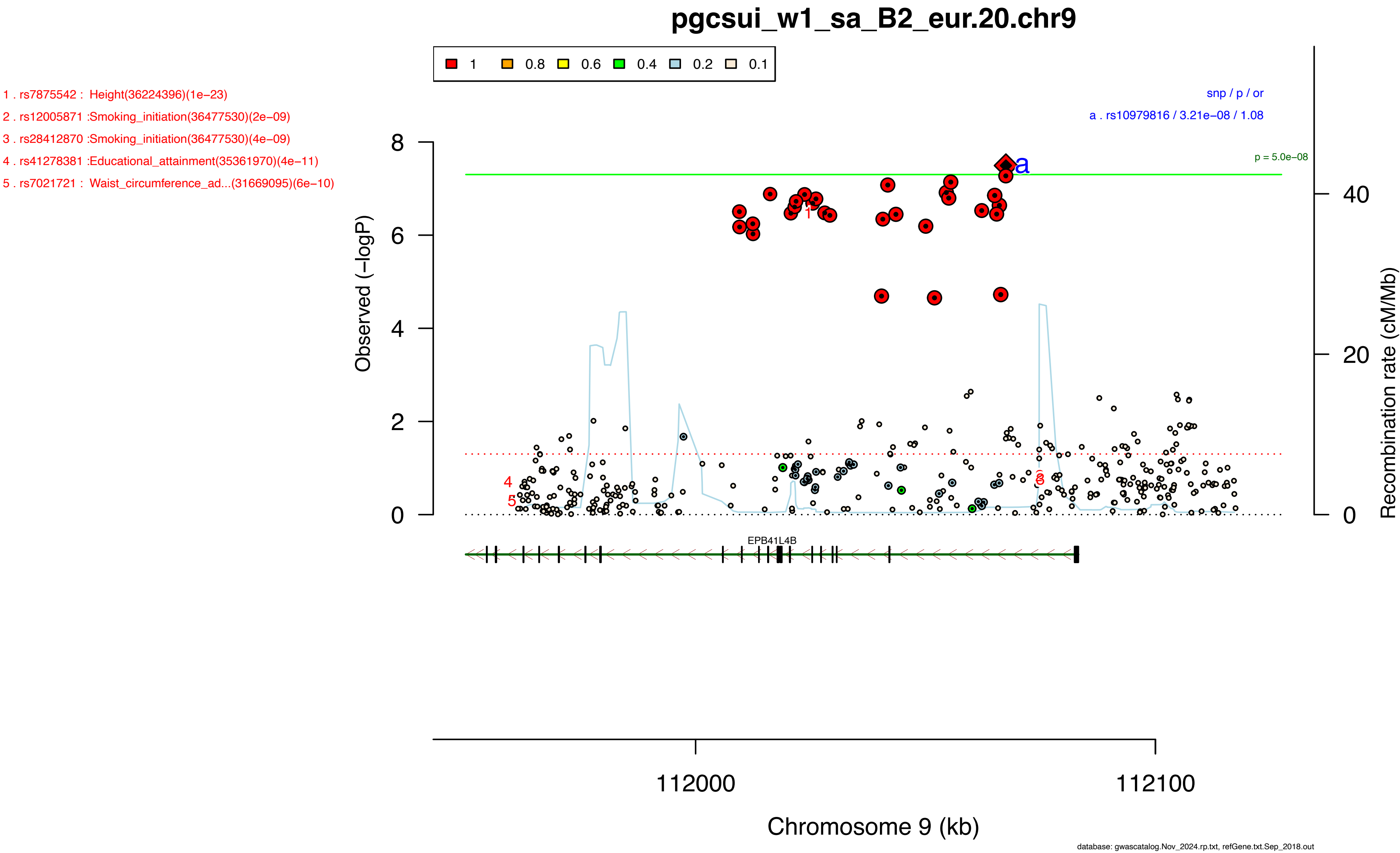
pgcsui_w1_sa_B2_eur.20.chr9
1 . rs7875542 : Height(36224396)(1e−23)
2 . rs12005871 :Smoking_initiation(36477530)(2e−09)
3 . rs28412870 :Smoking_initiation(36477530)(4e−09)
4 . rs41278381 :Educational_attainment(35361970)(4e−11)
5 . rs7021721 : Waist_circumference_ad...(31669095)(6e−10)
1 0.8 0.6 0.4 0.2 0.1
snp / p / or
a . rs10979816 / 3.21e−08 / 1.08
p = 5.0e−08
Observed (−logP)
Recombination rate (cM/Mb)
EPB41L4B
112000
112100
Chromosome 9 (kb)
database: gwascatalog.Nov_2024.rp.txt, refGene.txt.Sep_2018.out

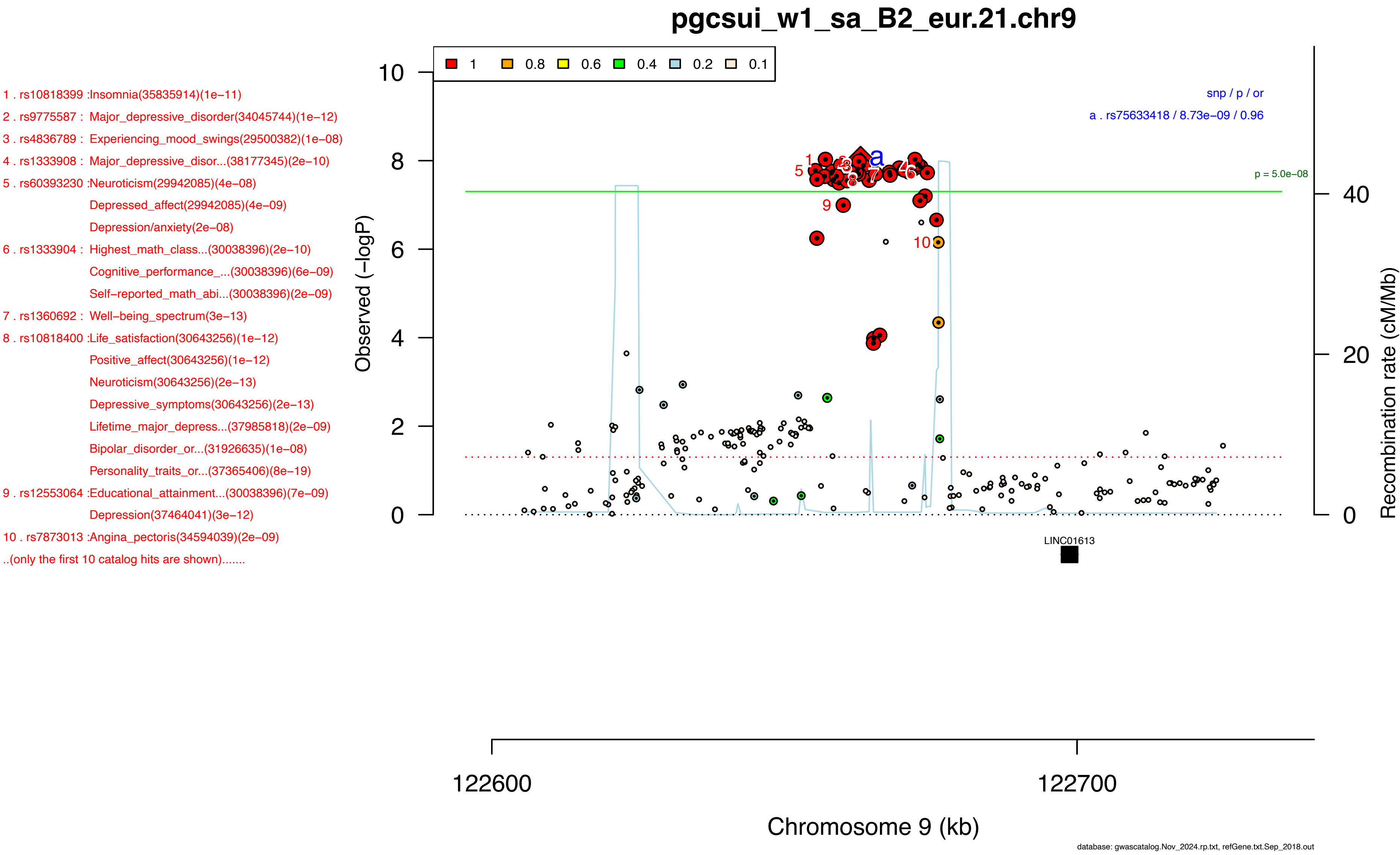
pgcsui_w1_sa_B2_eur.21.chr9
1 . rs10818399 :Insomnia(35835914)(1e−11)
2 . rs9775587 : Major_depressive_disorder(34045744)(1e−12)
3 . rs4836789 : Experiencing_mood_swings(29500382)(1e−08)
4 . rs1333908 : Major_depressive_disor...(38177345)(2e−10)
5 . rs60393230 :Neuroticism(29942085)(4e−08)
Depressed_affect(29942085)(4e−09)
Depression/anxiety(2e−08)
6 . rs1333904 : Highest_math_class...(30038396)(2e−10)
Cognitive_performance_...(30038396)(6e−09)
Self−reported_math_abi...(30038396)(2e−09)
7 . rs1360692 : Well−being_spectrum(3e−13)
8 . rs10818400 :Life_satisfaction(30643256)(1e−12)
Positive_affect(30643256)(1e−12)
Neuroticism(30643256)(2e−13)
Depressive_symptoms(30643256)(2e−13)
Lifetime_major_depress...(37985818)(2e−09)
Bipolar_disorder_or...(31926635)(1e−08)
Personality_traits_or...(37365406)(8e−19)
9 . rs12553064 :Educational_attainment...(30038396)(7e−09)
Depression(37464041)(3e−12)
10 . rs7873013 :Angina_pectoris(34594039)(2e−09)
..(only the first 10 catalog hits are shown).......
1
0.8
0.6
0.4
0.2
0.1
snp / p / or
a . rs75633418 / 8.73e−09 / 0.96
p = 5.0e−08
Observed (−logP)
Recombination rate (cM/Mb)
LINC01613
122600
122700
Chromosome 9 (kb)
database: gwascatalog.Nov_2024.rp.txt, refGene.txt.Sep_2018.out

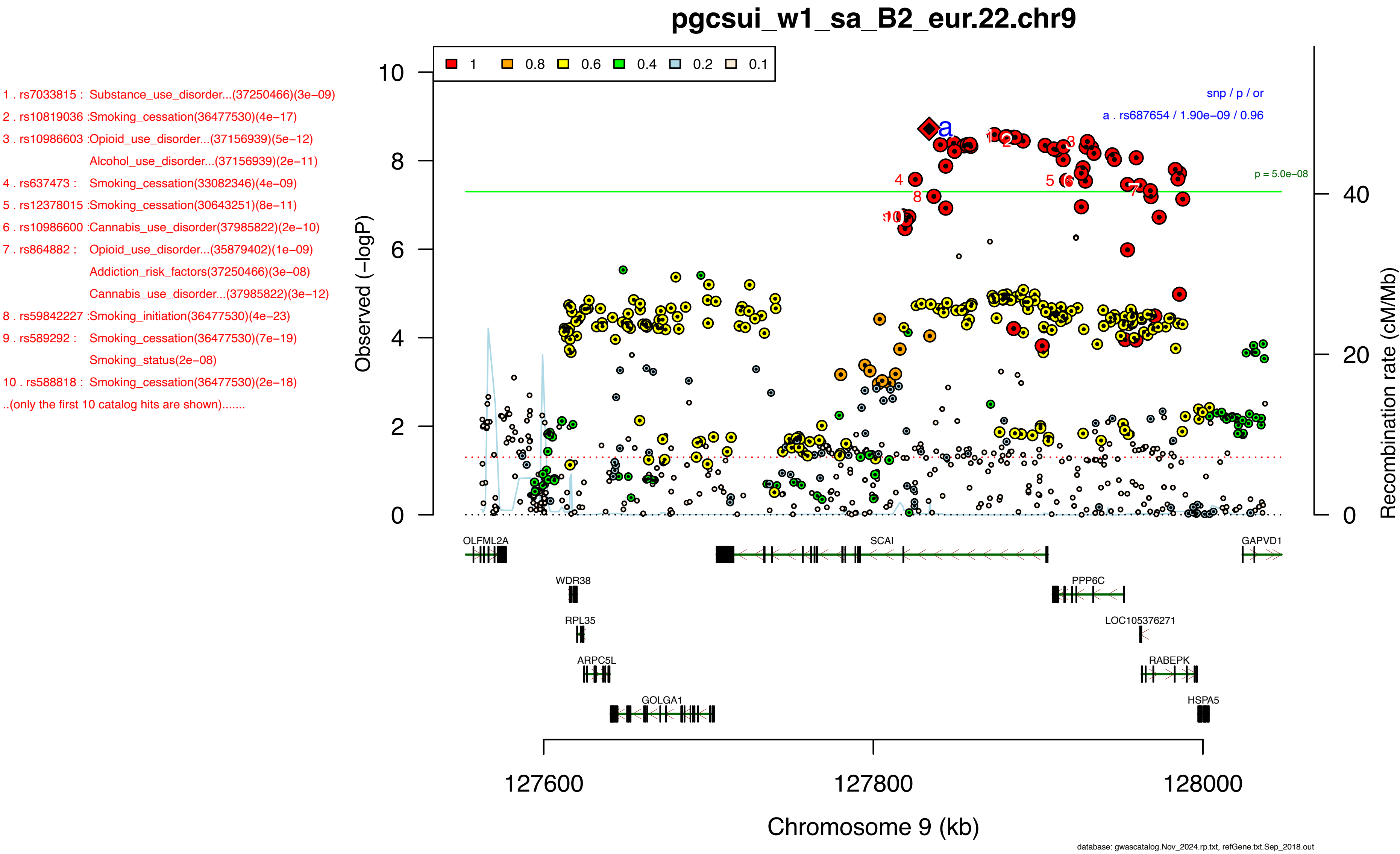

pgcsui_w1_sa_B2_eur.22.chr9
1 . rs7033815 : Substance_use_disorder...(37250466)(3e−09)
2 . rs10819036 :Smoking_cessation(36477530)(4e−17)
3 . rs10986603 :Opioid_use_disorder...(37156939)(5e−12)
Alcohol_use_disorder...(37156939)(2e−11)
4 . rs637473 : Smoking_cessation(33082346)(4e−09)
5 . rs12378015 :Smoking_cessation(30643251)(8e−11)
6 . rs10986600 :Cannabis_use_disorder(37985822)(2e−10)
7 . rs864882 : Opioid_use_disorder...(35879402)(1e−09)
Addiction_risk_factors(37250466)(3e−08)
Cannabis_use_disorder...(37985822)(3e−12)
8 . rs59842227 :Smoking_initiation(36477530)(4e−23)
9 . rs589292 : Smoking_cessation(36477530)(7e−19)
Smoking_status(2e−08)
10 . rs588818 : Smoking_cessation(36477530)(2e−18)
..(only the first 10 catalog hits are shown).......
1 0.8 0.6 0.4 0.2 0.1
snp / p / or
a . rs687654 / 1.90e−09 / 0.96
p = 5.0e−08
Observed (−logP)
Recombination rate (cM/Mb)
OLFML2A
SCAI
GAPVD1
WDR38
PPP6C
RPL35
LOC105376271
ARPC5L
RABEPK
GOLGA1
HSPA5
127600
127800
128000
Chromosome 9 (kb)
database: gwascatalog.Nov_2024.rp.txt, refGene.txt.Sep_2018.out

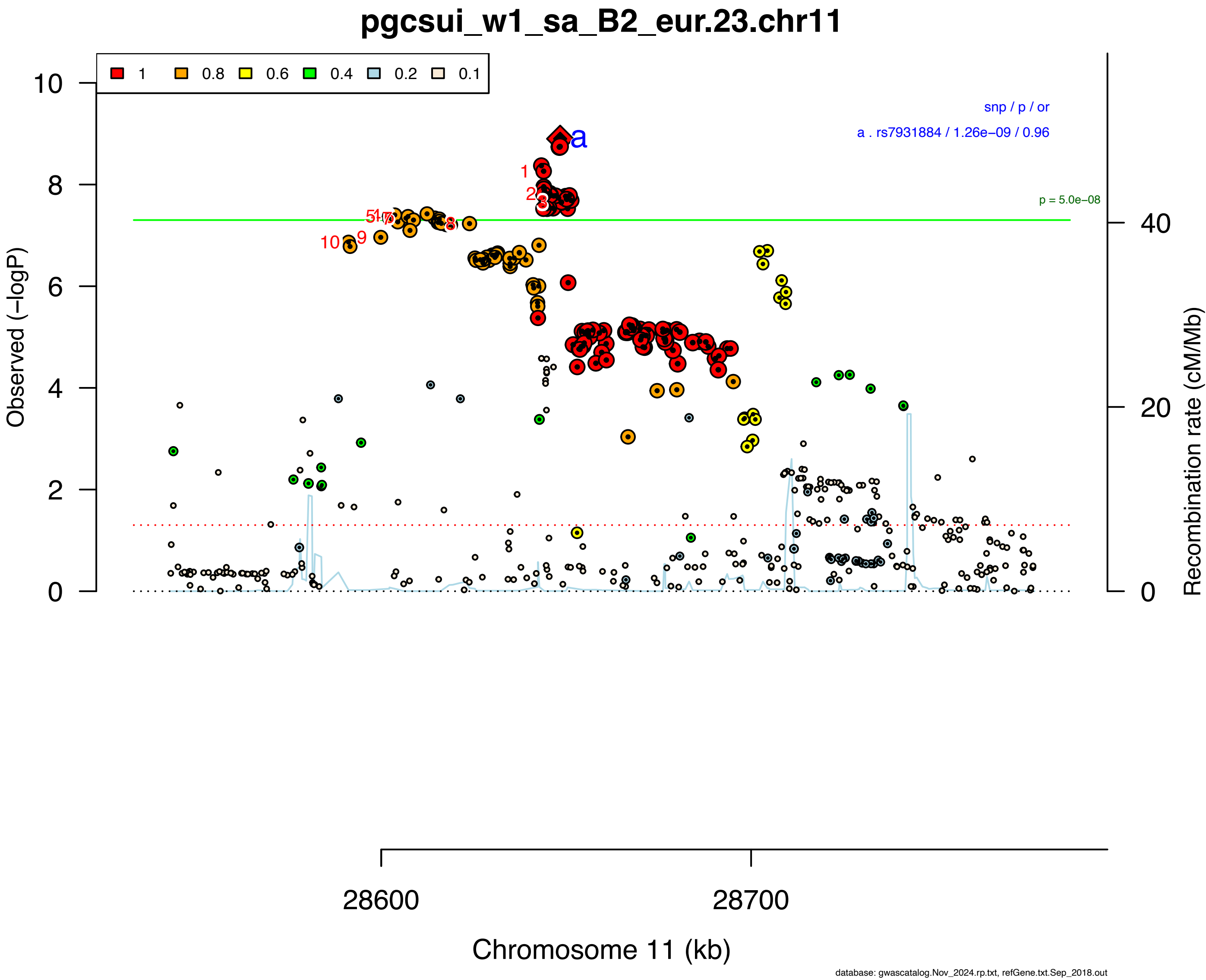


1 . rs10835372 :Cannabis_use_disorder...(37156939)(2e−09)
Alcohol_use_disorder...(37156939)(8e−09)
Smoking_initiation(6e−12)
Cannabis_use_disorder...(37985822)(2e−10)
2 . rs4267058 : Body_size_at...(32376654)(1e−09)
3 . rs11030393 :Height(36224396)(3e−31)
4 . rs7128734 : Smoking_cessation(36477530)(2e−16)
5 . rs2585817 : Smoking_status(5e−13)
Smoking_initiation(33082346)(5e−09)
Smoking_cessation(36477530)(1e−15)
Attention_deficit_hype...(38565336)(5e−15)
6 . rs7952220 : Attention_deficit_hype...(32606422)(1e−08)
Smoking_cessation(36477530)(2e−16)
7 . rs10835360 :Smoking_cessation(36477530)(2e−16)
8 . rs7931440 : Primary_open_angle...(37386247)(1e−09)
Disruptive_behavior(5e−09)
9 . rs11030381 :Osteoarthritis(36411363)(7e−09)
Osteoarthritis(36411363)(8e−10)
10 . rs11030380Depression(37464041)(1e−14)
..(only the first 10 catalog hits are shown).......

1 . rs12790660 :Insomnia(30804565)(4e−10)
Insomnia(35835914)(1e−16)
Insomnia(35835914)(4e−12)
2 . rs12807250 :Self−reported_math_abi...(30038396)(2e−28)
Self−reported_math_abi...(30038396)(7e−33)
3 . rs10896636 :Neurociticism(29500382)(9e−11)
General_factor_of...(30867560)(3e−09)
Neuroticism(29942085)(7e−13)
Depression(29942085)(3e−09)
Anxiety(33859377)(7e−14)
Hypertension(37947095)(7e−11)
Depressive_symptoms(32606422)(3e−09)
Neuroticism(32231276)(3e−11)
Neuroticism(30595370)(9e−13)
Educational_attainment(35361970)(2e−17)
4 . rs9645687 : Major_depressive_disor...(33479212)(9e−10)
Neuropsychiatric_disor...(33479212)(7e−09)
Major_depressive_disor...(38177345)(1e−10)
5 . rs9420 : Schizophrenia(30285260)(2e−08)
Schizophrenia(31740837)(7e−09)
6 . rs7117205 : Neurociticism(29500382)(6e−10)
Educational_attainment(34855049)(2e−10)
7 . rs78587207 :Mean_corpuscular_volume(34104963)(8e−10)
8 . rs527528 : Sensitivity_to_environ...(31972866)(7e−14)
Depressive_symptoms(3e−12)
Sociability_score(34054130)(4e−10)
Worry_too_long...(34054130)(4e−11)
9 . rs488769 : Mean_corpuscular_volume(32888493)(5e−23)
Mean_corpuscular_hemog...(32888493)(3e−21)
10 . rs7932091 :Highest_math_class...(30038396)(9e−23)
..(only the first 10 catalog hits are shown).......

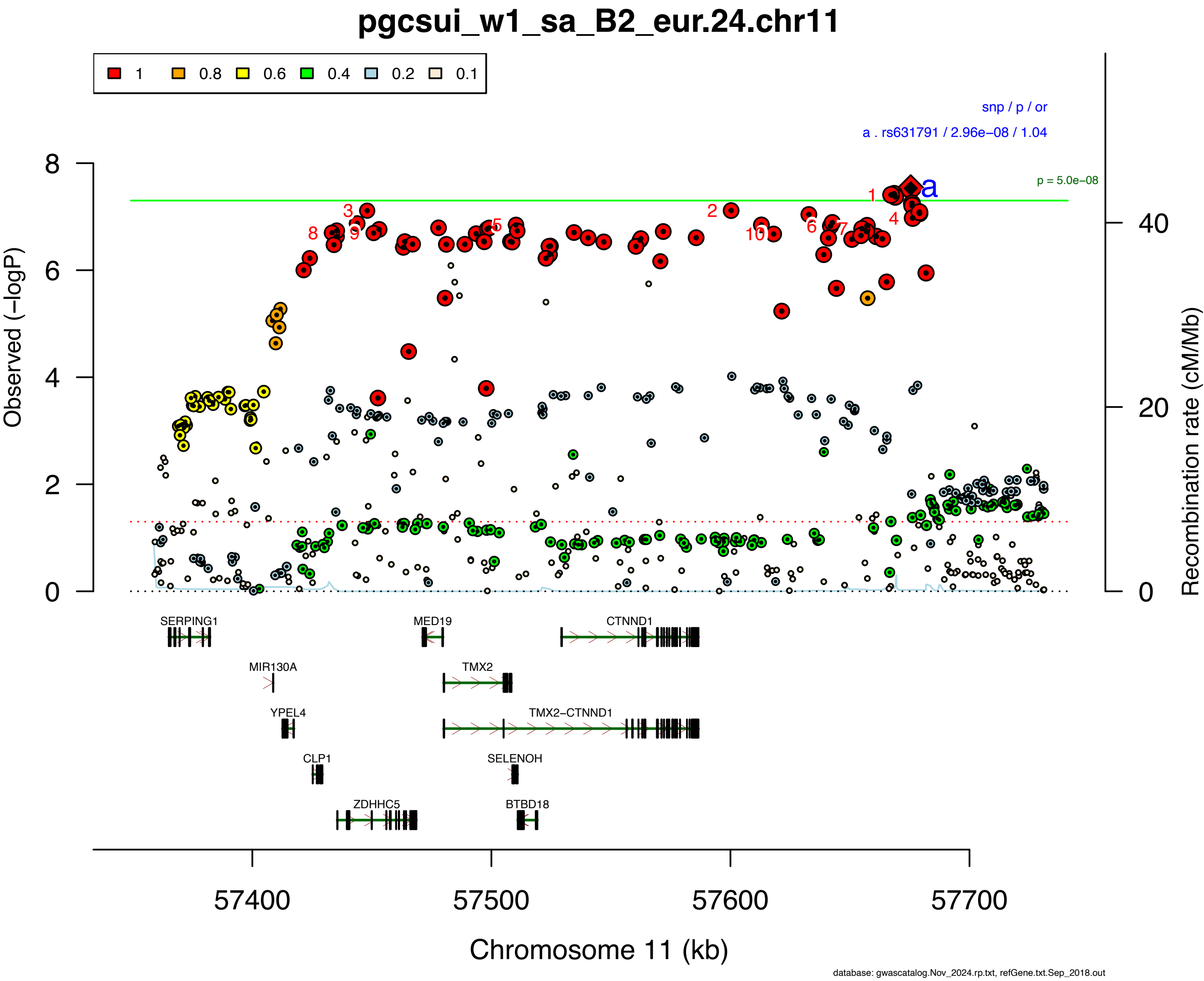

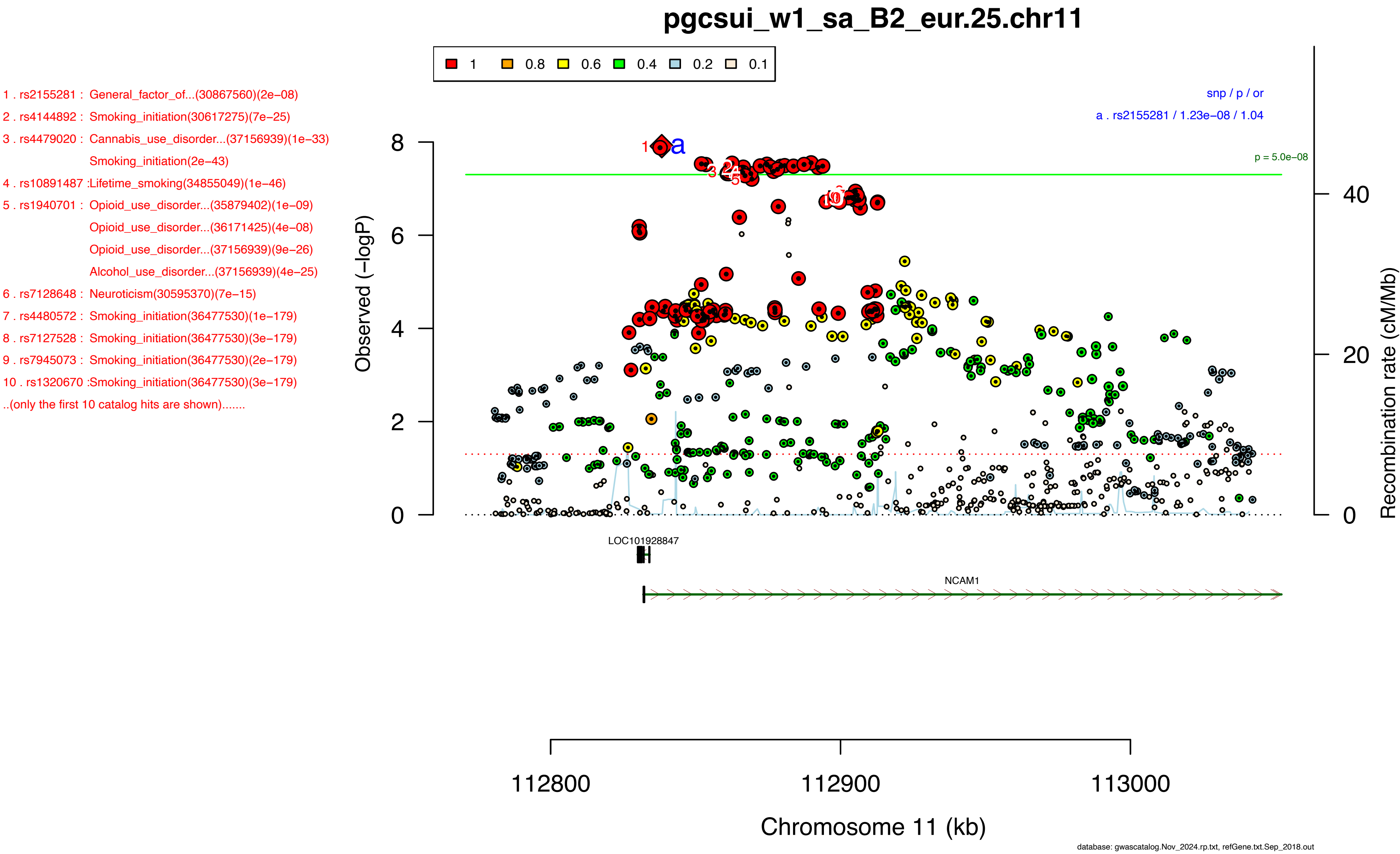
pgcsui_w1_sa_B2_eur.25.chr11
1 0.8 0.6 0.4 0.2 0.1
1 . rs2155281 : General_factor_of...(30867560)(2e−08)
2 . rs4144892 : Smoking_initiation(30617275)(7e−25)
3 . rs4479020 : Cannabis_use_disorder...(37156939)(1e−33)
Smoking_initiation(2e−43)
4 . rs10891487 :Lifetime_smoking(34855049)(1e−46)
5 . rs1940701 : Opioid_use_disorder...(35879402)(1e−09)
Opioid_use_disorder...(36171425)(4e−08)
Opioid_use_disorder...(37156939)(9e−26)
Alcohol_use_disorder...(37156939)(4e−25)
6 . rs7128648 : Neuroticism(30595370)(7e−15)
7 . rs4480572 : Smoking_initiation(36477530)(1e−179)
8 . rs7127528 : Smoking_initiation(36477530)(3e−179)
9 . rs7945073 : Smoking_initiation(36477530)(2e−179)
10 . rs1320670 :Smoking_initiation(36477530)(3e−179)
..(only the first 10 catalog hits are shown).......
snp / p / or
a . rs2155281 / 1.23e−08 / 1.04
p = 5.0e−08
Observed (−logP)
Recombination rate (cM/Mb)
LOC101928847
NCAM1
112800
112900
113000
Chromosome 11 (kb)
database: gwascatalog.Nov_2024.rp.txt, refGene.txt.Sep_2018.out

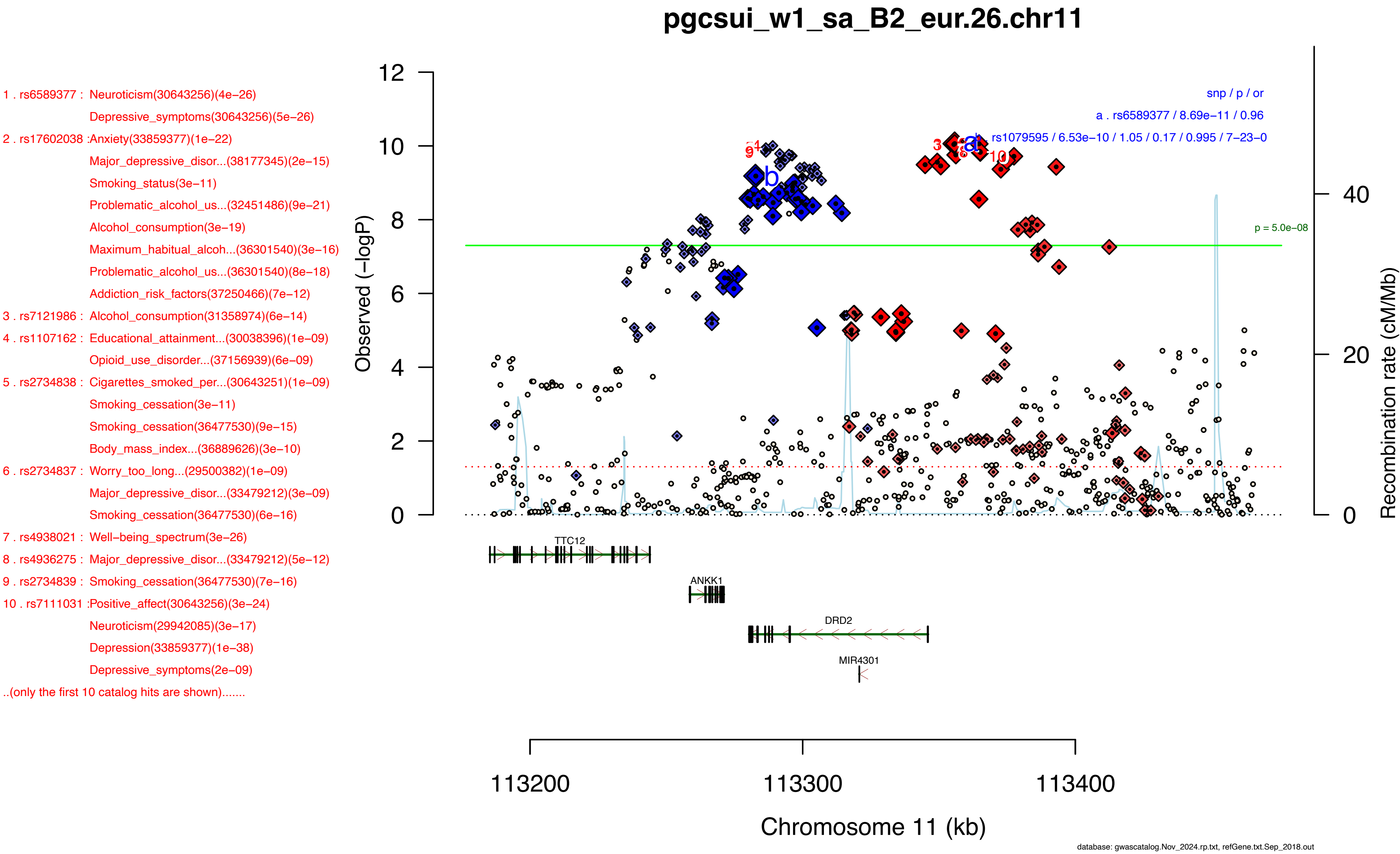

pgcsui_w1_sa_B2_eur.26.chr11
1 . rs6589377 : Neuroticism(30643256)(4e−26)
Depressive_symptoms(30643256)(5e−26)
2 . rs17602038 :Anxiety(33859377)(1e−22)
Major_depressive_disor...(38177345)(2e−15)
Smoking_status(3e−11)
Problematic_alcohol_us...(32451486)(9e−21)
Alcohol_consumption(3e−19)
Maximum_habitual_alcoh...(36301540)(3e−16)
Problematic_alcohol_us...(36301540)(8e−18)
Addiction_risk_factors(37250466)(7e−12)
3 . rs7121986 : Alcohol_consumption(31358974)(6e−14)
4 . rs1107162 : Educational_attainment...(30038396)(1e−09)
Opioid_use_disorder...(37156939)(6e−09)
5 . rs2734838 : Cigarettes_smoked_per...(30643251)(1e−09)
Smoking_cessation(3e−11)
Smoking_cessation(36477530)(9e−15)
Body_mass_index...(36889626)(3e−10)
6 . rs2734837 : Worry_too_long...(29500382)(1e−09)
Major_depressive_disor...(33479212)(3e−09)
Smoking_cessation(36477530)(6e−16)
7 . rs4938021 : Well−being_spectrum(3e−26)
8 . rs4936275 : Major_depressive_disor...(33479212)(5e−12)
9 . rs2734839 : Smoking_cessation(36477530)(7e−16)
10 . rs7111031 :Positive_affect(30643256)(3e−24)
Neuroticism(29942085)(3e−17)
Depression(33859377)(1e−38)
Depressive_symptoms(2e−09)
..(only the first 10 catalog hits are shown).......
snp / p / or
a . rs6589377 / 8.69e−11 / 0.96
b . rs1079595 / 6.53e−10 / 1.05 / 0.17 / 0.995 / 7−23−0
p = 5.0e−08
Observed (−logP)
Recombination rate (cM/Mb)
TTC12
ANKK1
DRD2
MIR4301
113200
113300
113400
Chromosome 11 (kb)
database: gwascatalog.Nov_2024.rp.txt, refGene.txt.Sep_2018.out

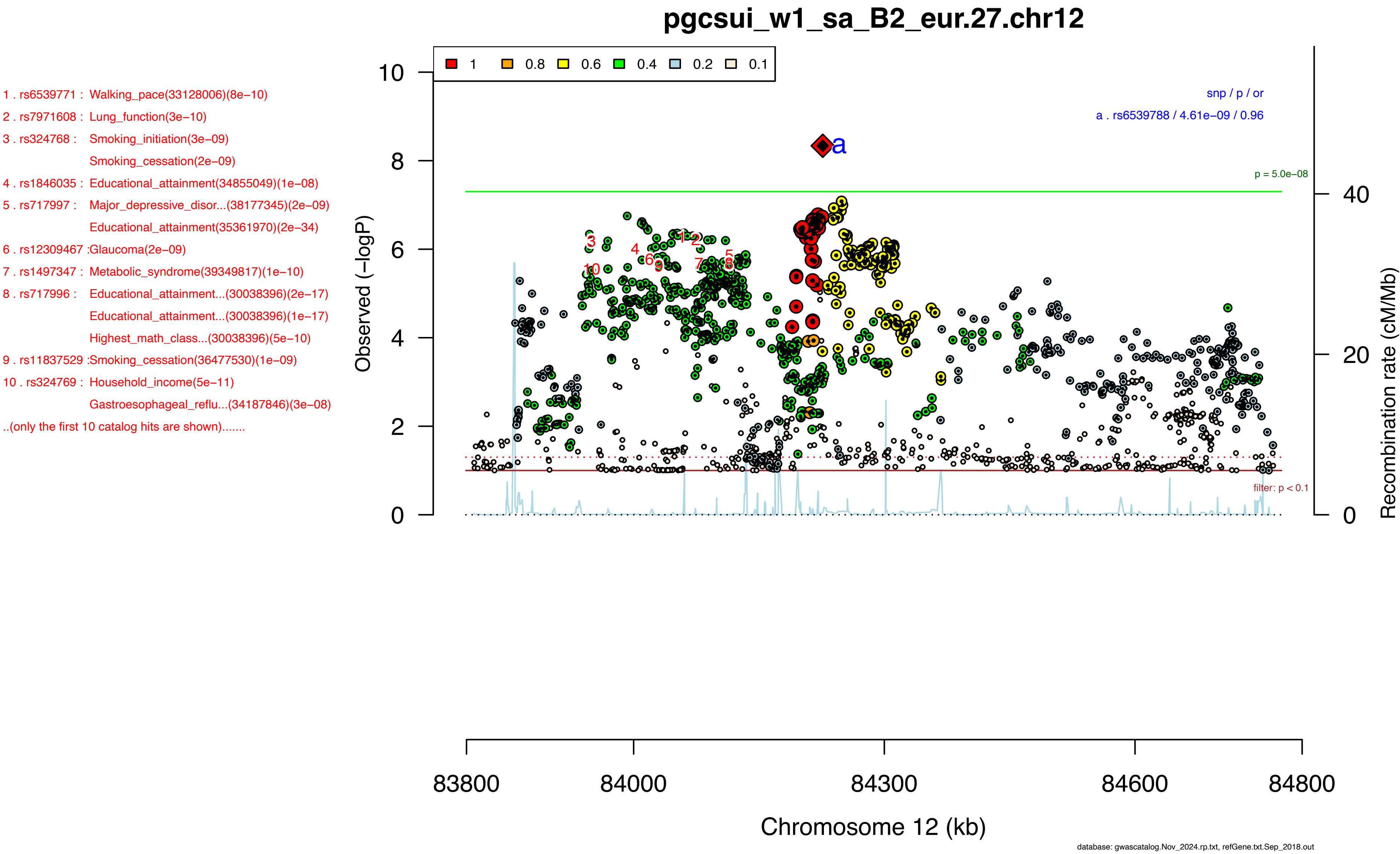

pgcsui_w1_sa_B2_eur.27.chr12
1 . rs6539771 : Walking_pace(33128006)(8e−10)
2 . rs7971608 : Lung_function(3e−10)
3 . rs324768 : Smoking_initiation(3e−09)
Smoking_cessation(2e−09)
4 . rs1846035 : Educational_attainment(34855049)(1e−08)
5 . rs717997 : Major_depressive_disor...(38177345)(2e−09)
Educational_attainment(35361970)(2e−34)
6 . rs12309467 :Glaucoma(2e−09)
7 . rs1497347 : Metabolic_syndrome(39349817)(1e−10)
8 . rs717996 : Educational_attainment...(30038396)(2e−17)
Educational_attainment...(30038396)(1e−17)
Highest_math_class...(30038396)(5e−10)
9 . rs11837529 :Smoking_cessation(36477530)(1e−09)
10 . rs324769 : Household_income(5e−11)
Gastroesophageal_reflu...(34187846)(3e−08)
..(only the first 10 catalog hits are shown).......
1 0.8 0.6 0.4 0.2 0.1
snp / p / or
a . rs6539788 / 4.61e−09 / 0.96
p = 5.0e−08
filter: p < 0.1
Observed (−logP)
Recombination rate (cM/Mb)
Chromosome 12 (kb)
83800 84000 84300 84600 84800
database: gwascatalog.Nov_2024.rp.txt, refGene.txt.Sep_2018.out

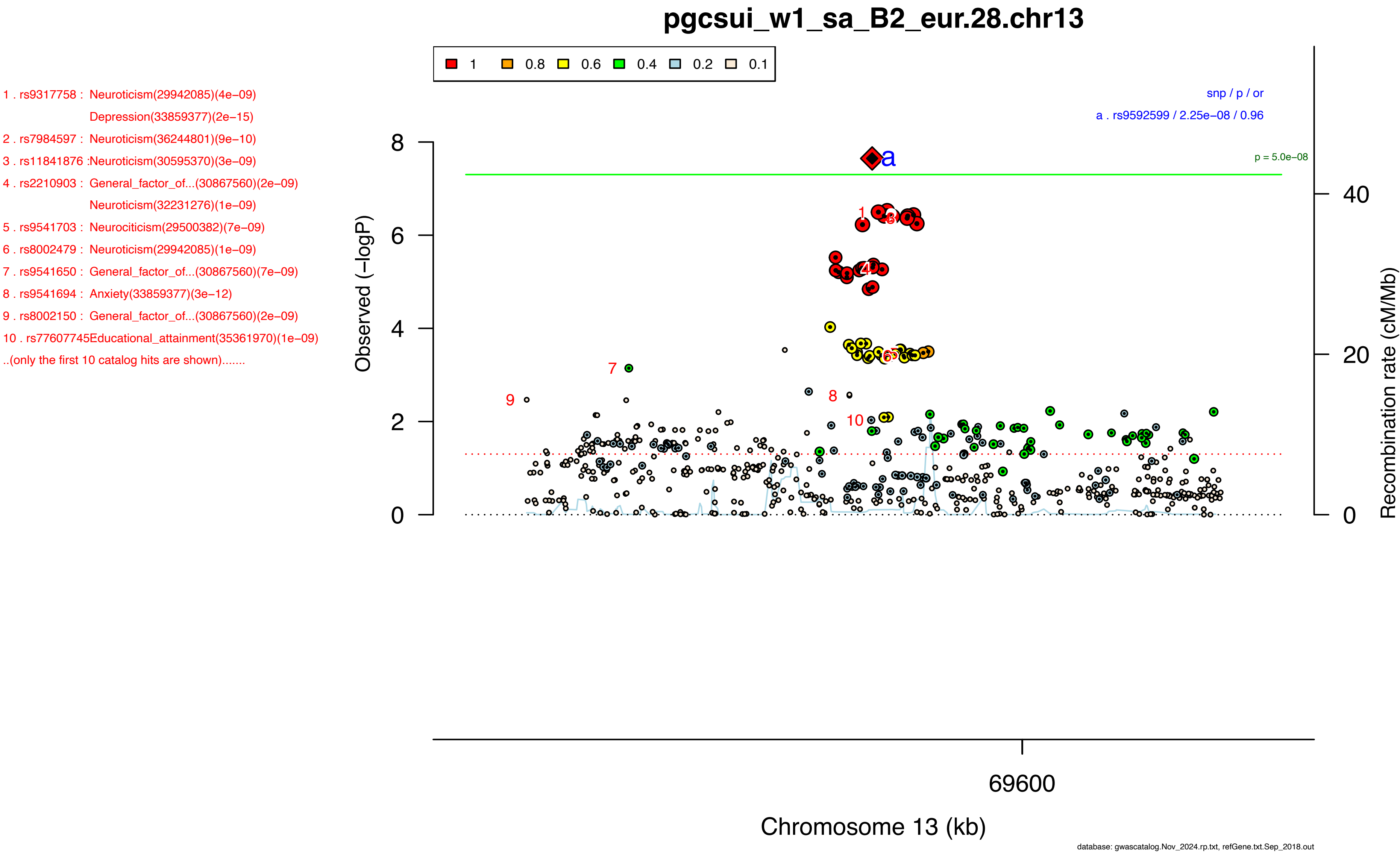
pgcsui_w1_sa_B2_eur.28.chr13
1
0.8
0.6
0.4
0.2
0.1
1 . rs9317758 : Neuroticism(29942085)(4e−09)
Depression(33859377)(2e−15)
2 . rs7984597 : Neuroticism(36244801)(9e−10)
3 . rs11841876 :Neuroticism(30595370)(3e−09)
4 . rs2210903 : General_factor_of...(30867560)(2e−09)
Neuroticism(32231276)(1e−09)
5 . rs9541703 : Neurociticism(29500382)(7e−09)
6 . rs8002479 : Neuroticism(29942085)(1e−09)
7 . rs9541650 : General_factor_of...(30867560)(7e−09)
8 . rs9541694 : Anxiety(33859377)(3e−12)
9 . rs8002150 : General_factor_of...(30867560)(2e−09)
10 . rs77607745Educational_attainment(35361970)(1e−09)
..(only the first 10 catalog hits are shown).......
snp / p / or
a . rs9592599 / 2.25e−08 / 0.96
p = 5.0e−08
Observed (−logP)
Recombination rate (cM/Mb)
69600
Chromosome 13 (kb)
database: gwascatalog.Nov_2024.rp.txt, refGene.txt.Sep_2018.out

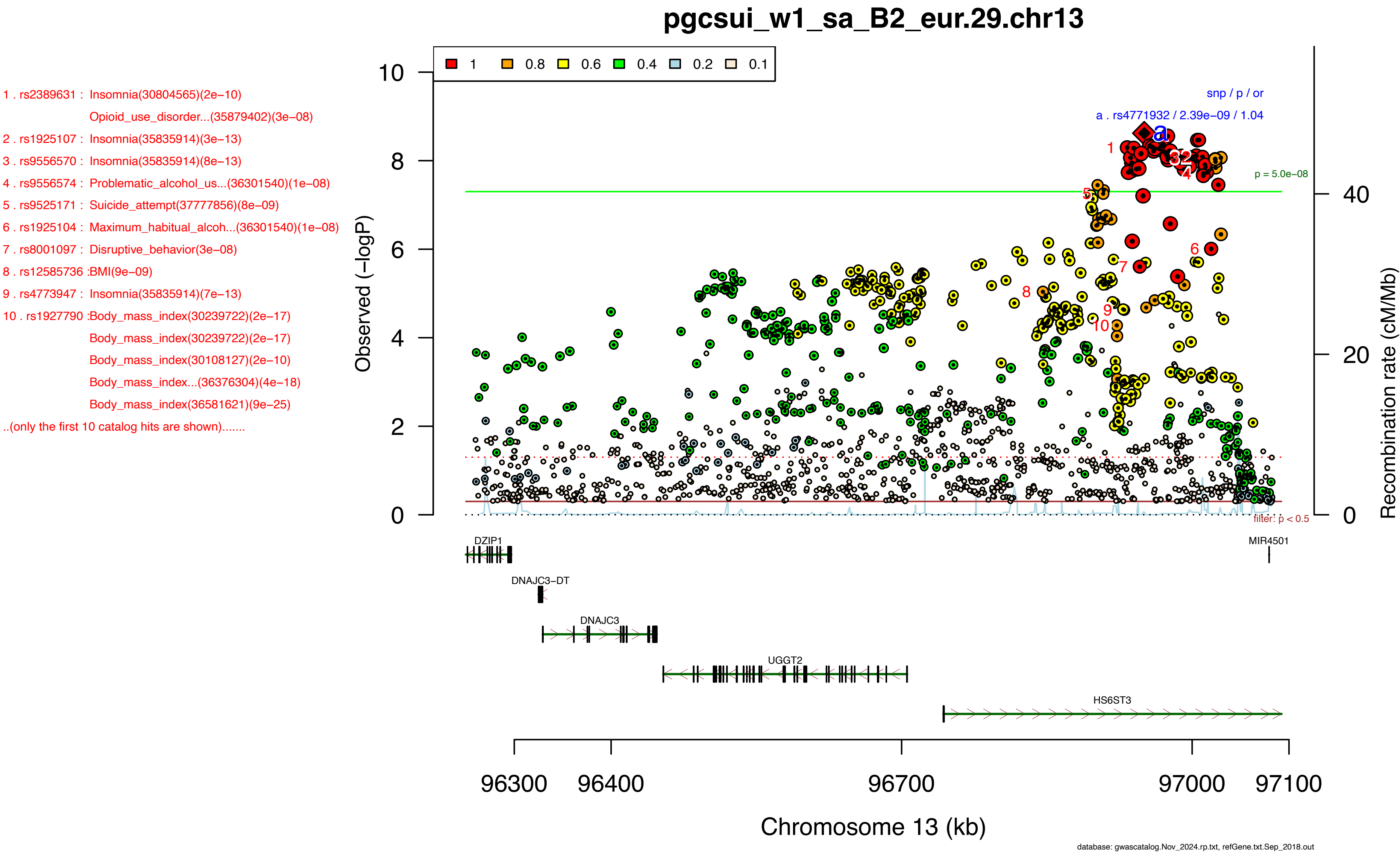
pgcsui_w1_sa_B2_eur.29.chr13
1 . rs2389631 : Insomnia(30804565)(2e−10)
Opioid_use_disorder...(35879402)(3e−08)
2 . rs1925107 : Insomnia(35835914)(3e−13)
3 . rs9556570 : Insomnia(35835914)(8e−13)
4 . rs9556574 : Problematic_alcohol_us...(36301540)(1e−08)
5 . rs9525171 : Suicide_attempt(37777856)(8e−09)
6 . rs1925104 : Maximum_habitual_alcoh...(36301540)(1e−08)
7 . rs8001097 : Disruptive_behavior(3e−08)
8 . rs12585736 :BMI(9e−09)
9 . rs4773947 : Insomnia(35835914)(7e−13)
10 . rs1927790 :Body_mass_index(30239722)(2e−17)
Body_mass_index(30239722)(2e−17)
Body_mass_index(30108127)(2e−10)
Body_mass_index...(36376304)(4e−18)
Body_mass_index(36581621)(9e−25)
..(only the first 10 catalog hits are shown).......
1 0.8 0.6 0.4 0.2 0.1
snp / p / or
a . rs4771932 / 2.39e−09 / 1.04
p = 5.0e−08
filter: p < 0.5
Observed (−logP)
Recombination rate (cM/Mb)
DZIP1
MIR4501
DNAJC3−DT
DNAJC3
UGGT2
HS6ST3
96300 96400 96700 97000 97100
Chromosome 13 (kb)
database: gwascatalog.Nov_2024.rp.txt, refGene.txt.Sep_2018.out

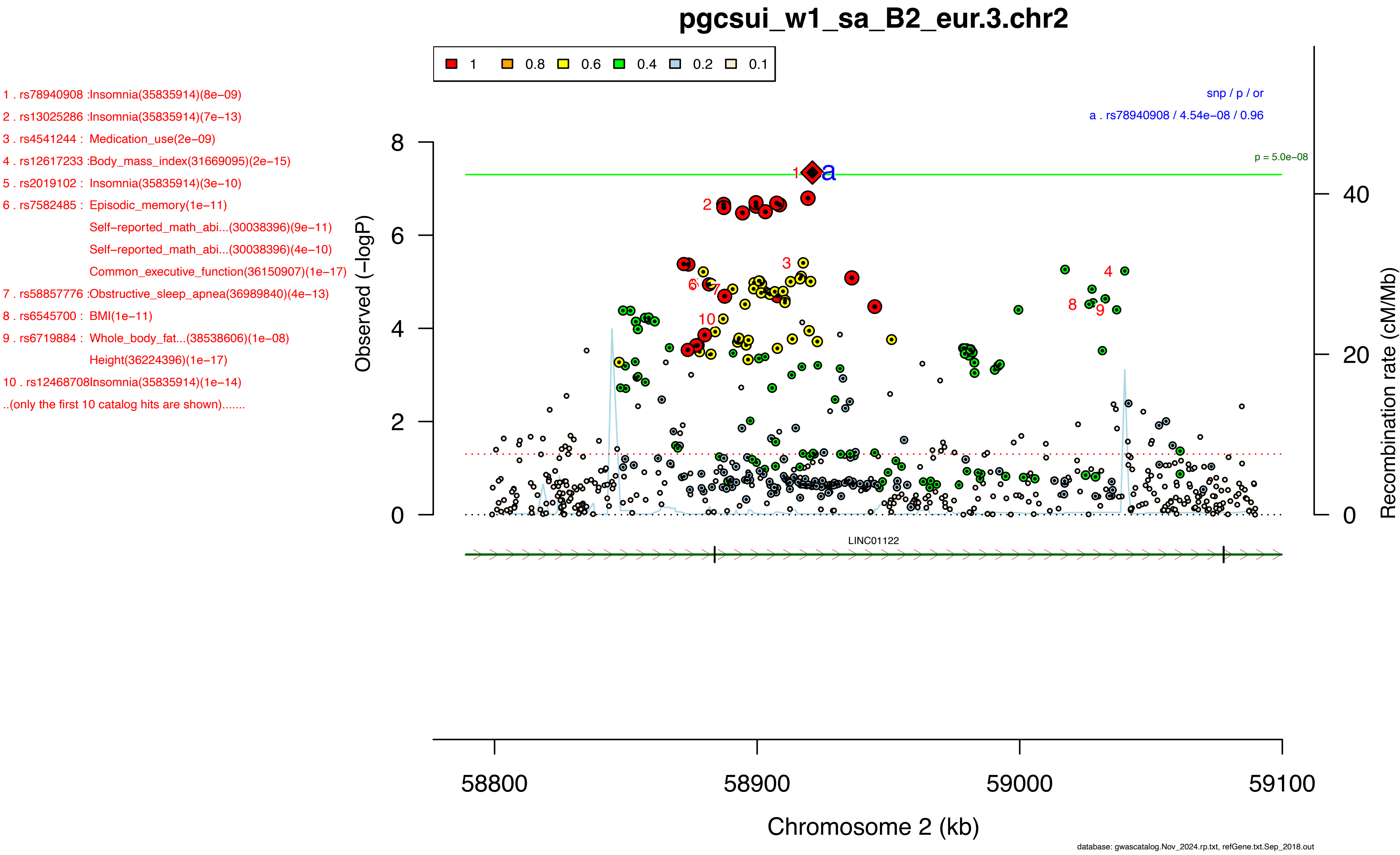
pgcsui_w1_sa_B2_eur.3.chr2
1 0.8 0.6 0.4 0.2 0.1
1 . rs78940908 :Insomnia(35835914)(8e−09)
2 . rs13025286 :Insomnia(35835914)(7e−13)
3 . rs4541244 : Medication_use(2e−09)
4 . rs12617233 :Body_mass_index(31669095)(2e−15)
5 . rs2019102 : Insomnia(35835914)(3e−10)
6 . rs7582485 : Episodic_memory(1e−11)
Self−reported_math_abi...(30038396)(9e−11)
Self−reported_math_abi...(30038396)(4e−10)
Common_executive_function(36150907)(1e−17)
7 . rs58857776 :Obstructive_sleep_apnea(36989840)(4e−13)
8 . rs6545700 : BMI(1e−11)
9 . rs6719884 : Whole_body_fat...(38538606)(1e−08)
Height(36224396)(1e−17)
10 . rs12468708Insomnia(35835914)(1e−14)
..(only the first 10 catalog hits are shown).......
snp / p / or
a . rs78940908 / 4.54e−08 / 0.96
p = 5.0e−08
Observed (−logP)
Recombination rate (cM/Mb)
LINC01122
58800 58900 59000 59100
Chromosome 2 (kb)
database: gwascatalog.Nov_2024.rp.txt, refGene.txt.Sep_2018.out

1 . rs12589834 :Major_depressive_disor...(33479212)(2e−08)
2 . rs1152595 : Educational_attainment...(30038396)(6e−11)
Educational_attainment(35361970)(5e−17)
3 . rs1152590 : Height(36224396)(2e−69)
4 . rs148114281Mean_corpuscular_hemog...(30595370)(1e−08)
5 . rs10142766 :Depression(37464041)(1e−11)
6 . rs1152578 : Depression(30718901)(4e−12)
Depression(34924174)(4e−10)
7 . rs3783727 : Heel_bone_mineral...(30595370)(5e−13)
8 . rs12881815 :Height(36224396)(8e−39)
9 . rs915057 : Anorexia_nervosa,_atte...(31835028)(3e−09)
Bipolar_disorder_or...(31926635)(2e−08)
Depressive_symptoms(4e−08)
Depression(29700475)(8e−10)
10 . rs8016556 :Cigarettes_smoked_per...(36477530)(2e−10)
..(only the first 10 catalog hits are shown).......

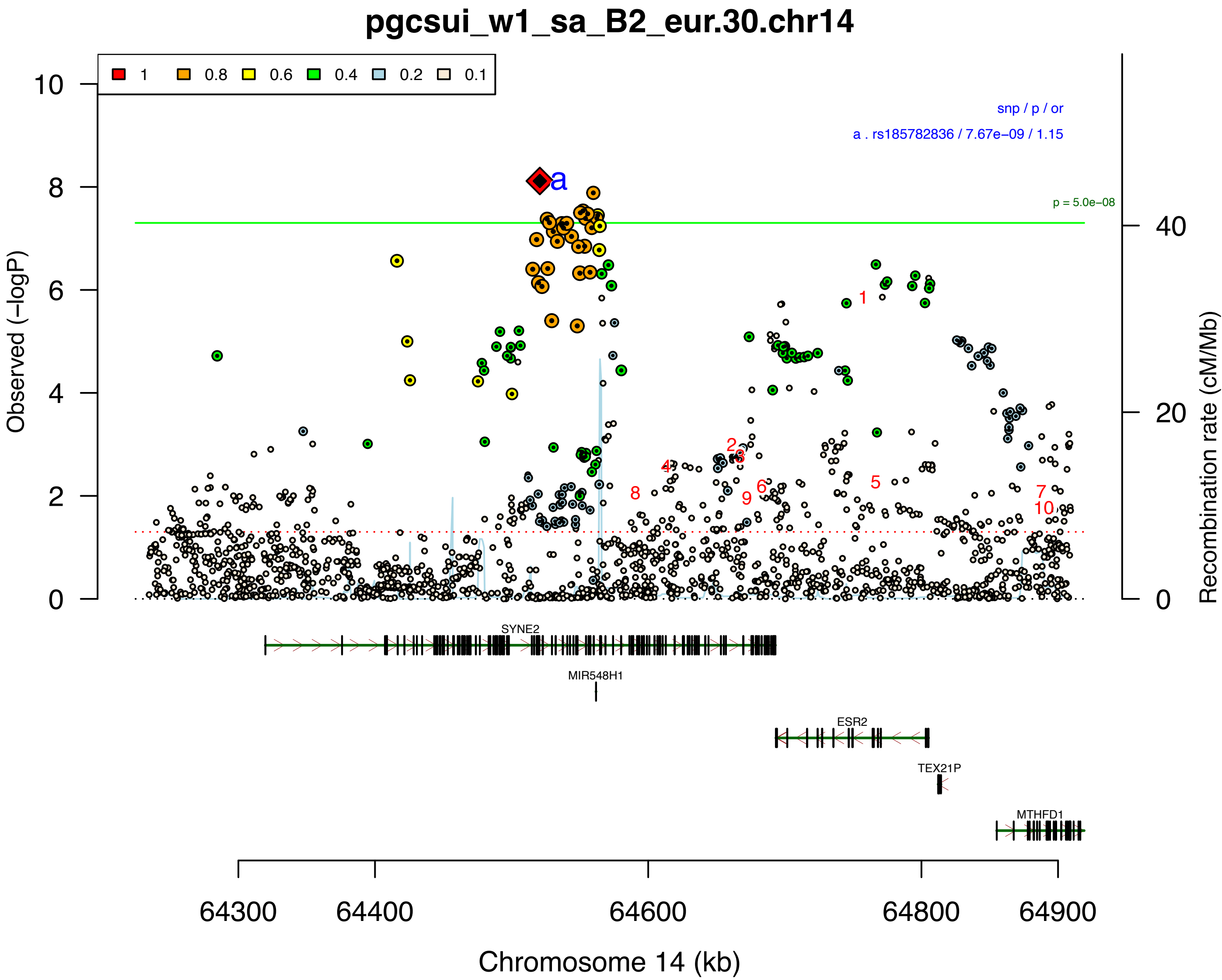


database: gwascatalog.Nov_2024.rp.txt, refGene.txt.Sep_2018.out

1 . rs7152530 : Smoking_cessation(36477530)(9e−12)
Smoking_cessation(36477530)(3e−11)
Parsopercularis_area(2e−08)
2 . rs8009025 : Systolic_blood_pressure(30578418)(4e−12)
Medication_use(1e−08)
3 . rs10143433 :Insomnia(35835914)(2e−09)
4 . rs75791745 :Biological_sex(33888908)(1e−08)
5 . rs7159716 : Insomnia(35835914)(3e−09)
6 . rs1461583 : Self−reported_math_abi...(30038396)(4e−08)
7 . rs1461584 : Lymphocyte_percentage_...(32888494)(6e−09)
8 . rs1461587 : Lymphocyte_count(34594039)(4e−11)
9 . rs1381274 : Age_of_smoking...(30643251)(1e−10)
Eosinophil_counts(34594039)(2e−10)
Risk−taking_tendency(6e−13)
Eosinophil_counts(32888494)(2e−09)
Eosinophil_percentage_...(32888494)(5e−10)
Smoking_status(30595370)(5e−09)
Water_consumption(6e−16)
Number_of_sexual...(34426668)(7e−12)
Smoking_initiation(2e−08)
10 . rs7160389 :Eosinophil_counts(30595370)(3e−08)
Smoking_status(4e−09)
..(only the first 10 catalog hits are shown).......

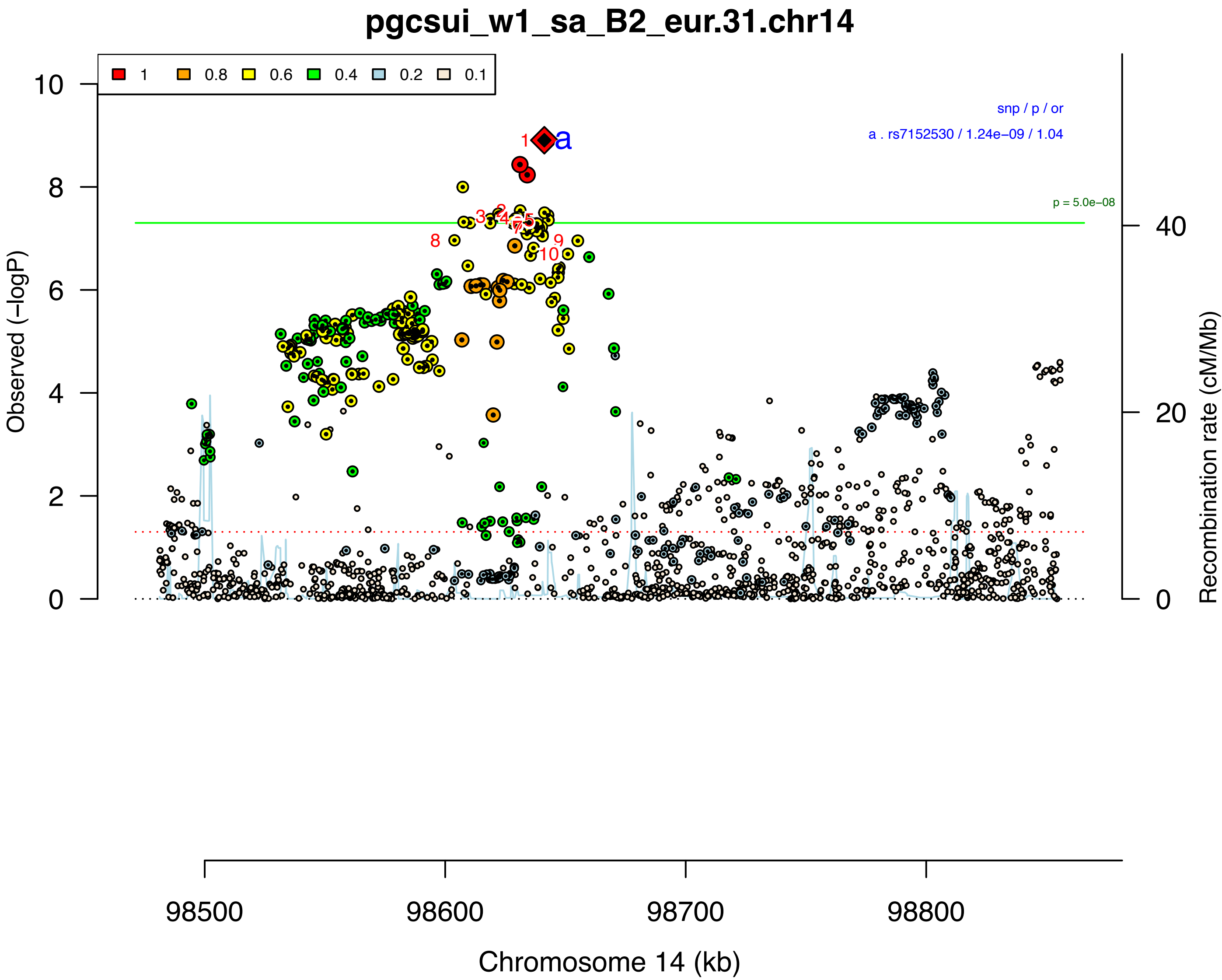


database: gwascatalog.Nov_2024.rp.txt, refGene.txt.Sep_2018.out

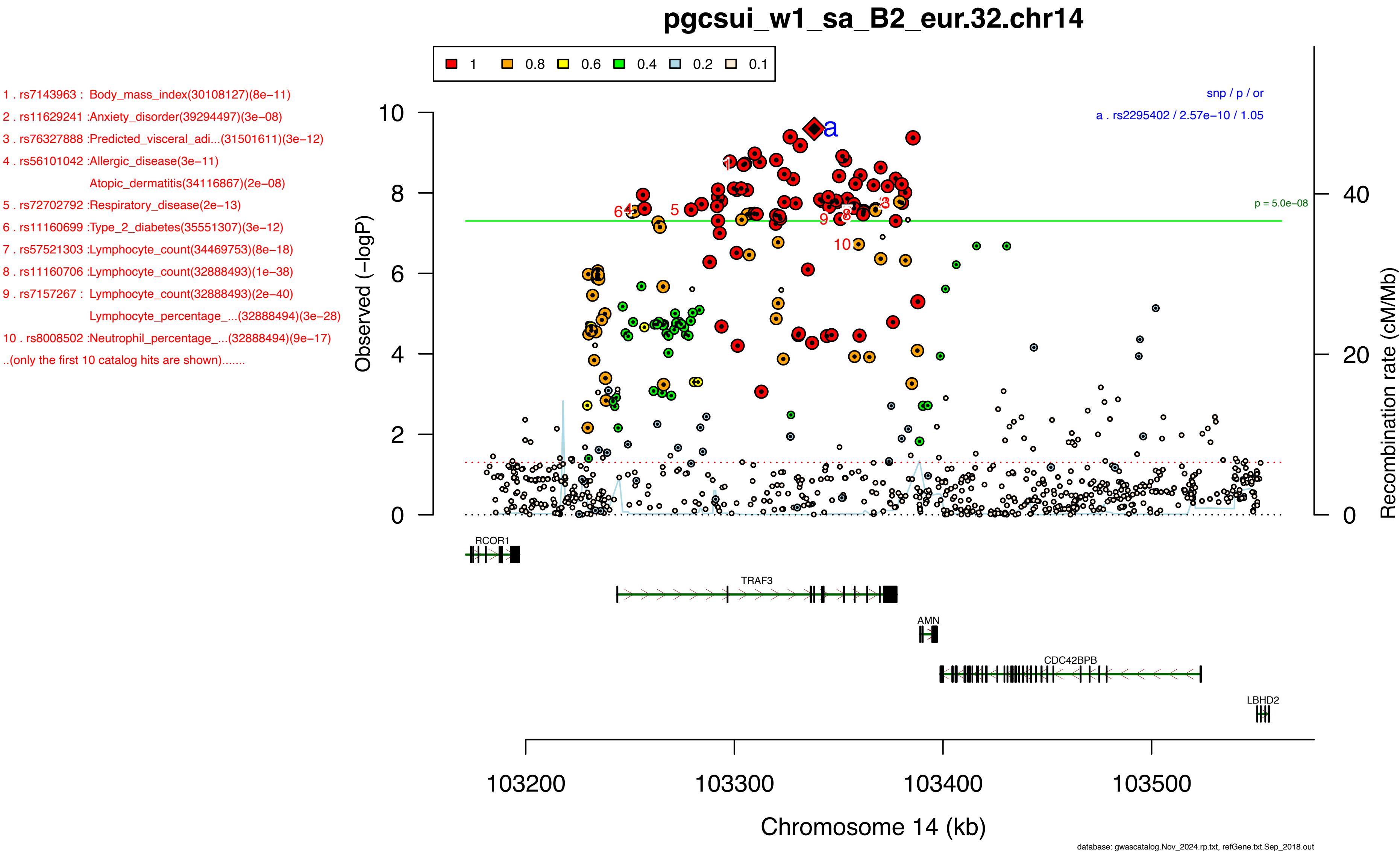

pgcsui_w1_sa_B2_eur.32.chr14
1 0.8 0.6 0.4 0.2 0.1
1 . rs7143963 : Body_mass_index(30108127)(8e−11)
2 . rs11629241 :Anxiety_disorder(39294497)(3e−08)
3 . rs76327888 :Predicted_visceral_adi...(31501611)(3e−12)
4 . rs56101042 :Allergic_disease(3e−11)
Atopic_dermatitis(34116867)(2e−08)
5 . rs72702792 :Respiratory_disease(2e−13)
6 . rs11160699 :Type_2_diabetes(35551307)(3e−12)
7 . rs57521303 :Lymphocyte_count(34469753)(8e−18)
8 . rs11160706 :Lymphocyte_count(32888493)(1e−38)
9 . rs7157267 : Lymphocyte_count(32888493)(2e−40)
Lymphocyte_percentage_...(32888494)(3e−28)
10 . rs8008502 :Neutrophil_percentage_...(32888494)(9e−17)
..(only the first 10 catalog hits are shown).......
snp / p / or
a . rs2295402 / 2.57e−10 / 1.05
p = 5.0e−08
Observed (−logP)
Recombination rate (cM/Mb)
RCOR1
TRAF3
AMN
CDC42BPB
LBHD2
103200 103300 103400 103500
Chromosome 14 (kb)
database: gwascatalog.Nov_2024.rp.txt, refGene.txt.Sep_2018.out

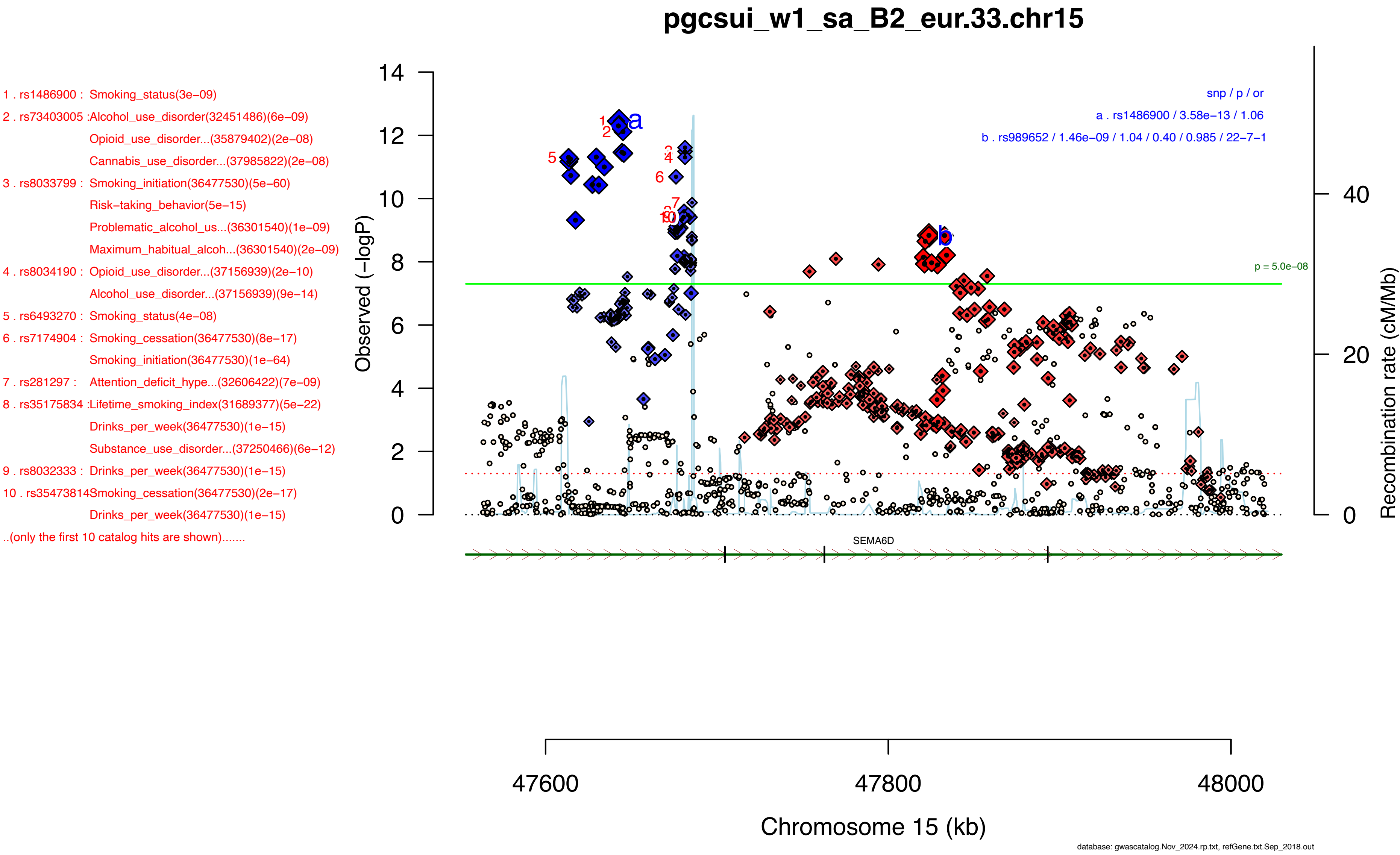

pgcsui_w1_sa_B2_eur.33.chr15
1 . rs1486900 : Smoking_status(3e−09)
2 . rs73403005 :Alcohol_use_disorder(32451486)(6e−09)
Opioid_use_disorder...(35879402)(2e−08)
Cannabis_use_disorder...(37985822)(2e−08)
3 . rs8033799 : Smoking_initiation(36477530)(5e−60)
Risk−taking_behavior(5e−15)
Problematic_alcohol_us...(36301540)(1e−09)
Maximum_habitual_alcoh...(36301540)(2e−09)
4 . rs8034190 : Opioid_use_disorder...(37156939)(2e−10)
Alcohol_use_disorder...(37156939)(9e−14)
5 . rs6493270 : Smoking_status(4e−08)
6 . rs7174904 : Smoking_cessation(36477530)(8e−17)
Smoking_initiation(36477530)(1e−64)
7 . rs281297 : Attention_deficit_hype...(32606422)(7e−09)
8 . rs35175834 :Lifetime_smoking_index(31689377)(5e−22)
Drinks_per_week(36477530)(1e−15)
Substance_use_disorder...(37250466)(6e−12)
9 . rs8032333 : Drinks_per_week(36477530)(1e−15)
10 . rs35473814Smoking_cessation(36477530)(2e−17)
Drinks_per_week(36477530)(1e−15)
..(only the first 10 catalog hits are shown).......
snp / p / or
a . rs1486900 / 3.58e−13 / 1.06
b . rs989652 / 1.46e−09 / 1.04 / 0.40 / 0.985 / 22−7−1
p = 5.0e−08
Observed (−logP)
Recombination rate (cM/Mb)
SEMA6D
47600
47800
48000
Chromosome 15 (kb)
database: gwascatalog.Nov_2024.rp.txt, refGene.txt.Sep_2018.out

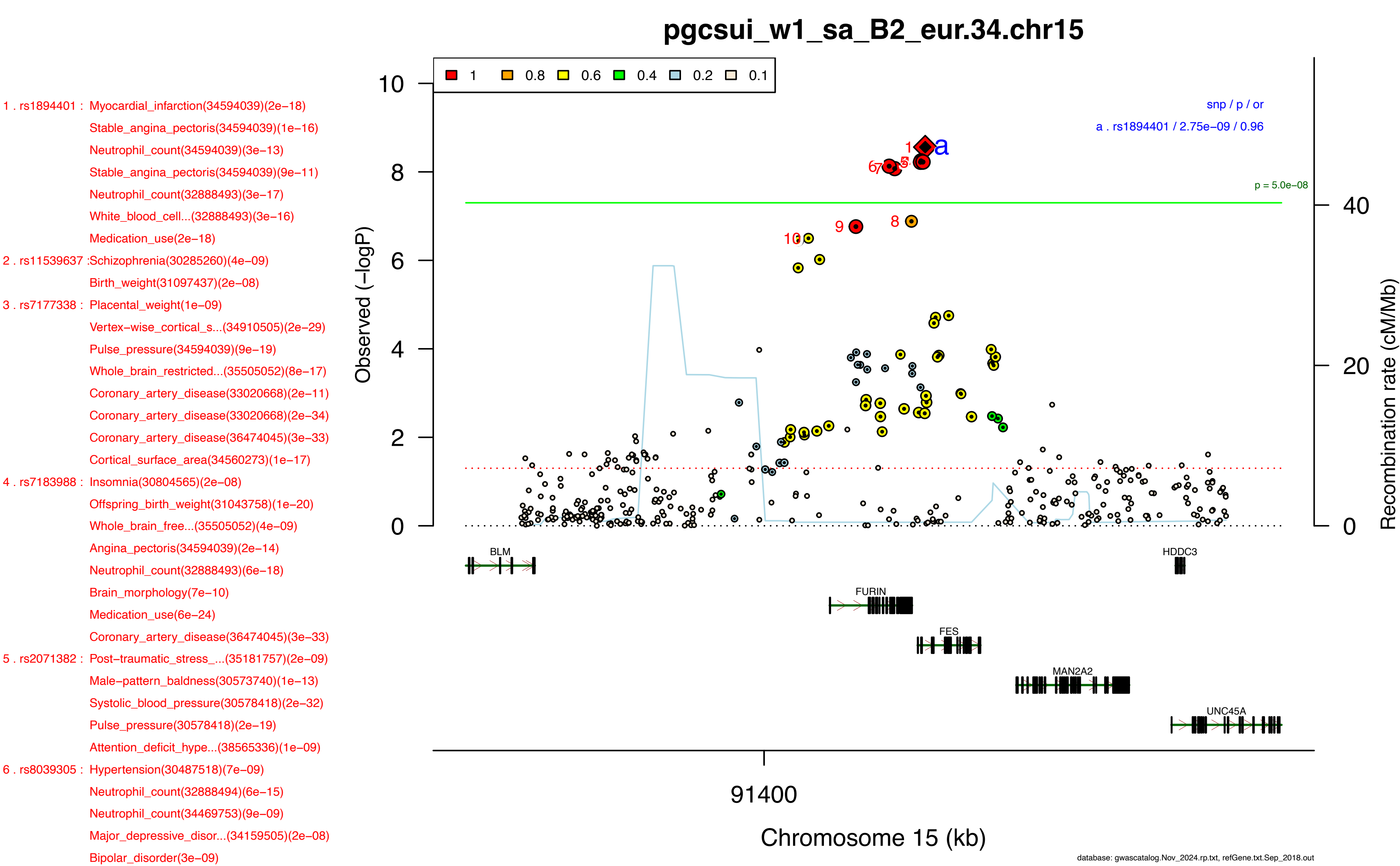
pgcsui_w1_sa_B2_eur.34.chr15
1 . rs1894401 : Myocardial_infarction(34594039)(2e−18)
Stable_angina_pectoris(34594039)(1e−16)
Neutrophil_count(34594039)(3e−13)
Stable_angina_pectoris(34594039)(9e−11)
Neutrophil_count(32888493)(3e−17)
White_blood_cell...(32888493)(3e−16)
Medication_use(2e−18)
2 . rs11539637 :Schizophrenia(30285260)(4e−09)
Birth_weight(31097437)(2e−08)
3 . rs7177338 : Placental_weight(1e−09)
Vertex−wise_cortical_s...(34910505)(2e−29)
Pulse_pressure(34594039)(9e−19)
Whole_brain_restricted...(35505052)(8e−17)
Coronary_artery_disease(33020668)(2e−11)
Coronary_artery_disease(33020668)(2e−34)
Coronary_artery_disease(36474045)(3e−33)
Cortical_surface_area(34560273)(1e−17)
4 . rs7183988 : Insomnia(30804565)(2e−08)
Offspring_birth_weight(31043758)(1e−20)
Whole_brain_free...(35505052)(4e−09)
Angina_pectoris(34594039)(2e−14)
Neutrophil_count(32888493)(6e−18)
Brain_morphology(7e−10)
Medication_use(6e−24)
Coronary_artery_disease(36474045)(3e−33)
5 . rs2071382 : Post−traumatic_stress_...(35181757)(2e−09)
Male−pattern_baldness(30573740)(1e−13)
Systolic_blood_pressure(30578418)(2e−32)
Pulse_pressure(30578418)(2e−19)
Attention_deficit_hype...(38565336)(1e−09)
6 . rs8039305 : Hypertension(30487518)(7e−09)
Neutrophil_count(32888494)(6e−15)
Neutrophil_count(34469753)(9e−09)
Major_depressive_disor...(34159505)(2e−08)
Bipolar_disorder(3e−09)
1 0.8 0.6 0.4 0.2 0.1
snp / p / or
a . rs1894401 / 2.75e−09 / 0.96
p = 5.0e−08
Observed (−logP)
Recombination rate (cM/Mb)
BLM
HDDC3
FURIN
FES
MAN2A2
UNC45A
91400
Chromosome 15 (kb)
database: gwascatalog.Nov_2024.rp.txt, refGene.txt.Sep_2018.out

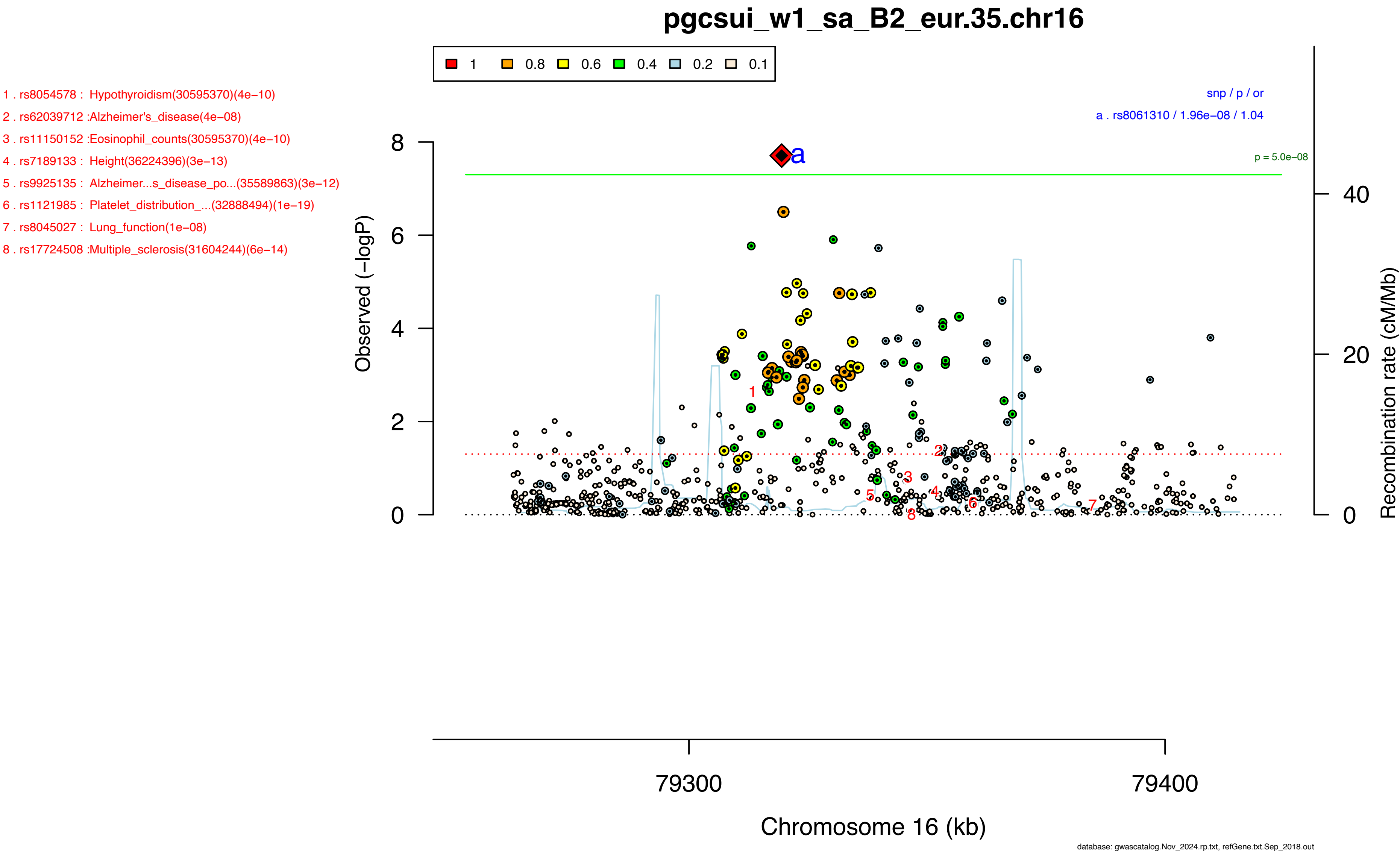
pgcsui_w1_sa_B2_eur.35.chr16
1 0.8 0.6 0.4 0.2 0.1
1 . rs8054578 : Hypothyroidism(30595370)(4e−10)
2 . rs62039712 :Alzheimer's_disease(4e−08)
3 . rs11150152 :Eosinophil_counts(30595370)(4e−10)
4 . rs7189133 : Height(36224396)(3e−13)
5 . rs9925135 : Alzheimer...s_disease_po...(35589863)(3e−12)
6 . rs1121985 : Platelet_distribution_...(32888494)(1e−19)
7 . rs8045027 : Lung_function(1e−08)
8 . rs17724508 :Multiple_sclerosis(31604244)(6e−14)
snp / p / or
a . rs8061310 / 1.96e−08 / 1.04
p = 5.0e−08
Observed (−logP)
Recombination rate (cM/Mb)
79300
79400
Chromosome 16 (kb)
database: gwascatalog.Nov_2024.rp.txt, refGene.txt.Sep_2018.out

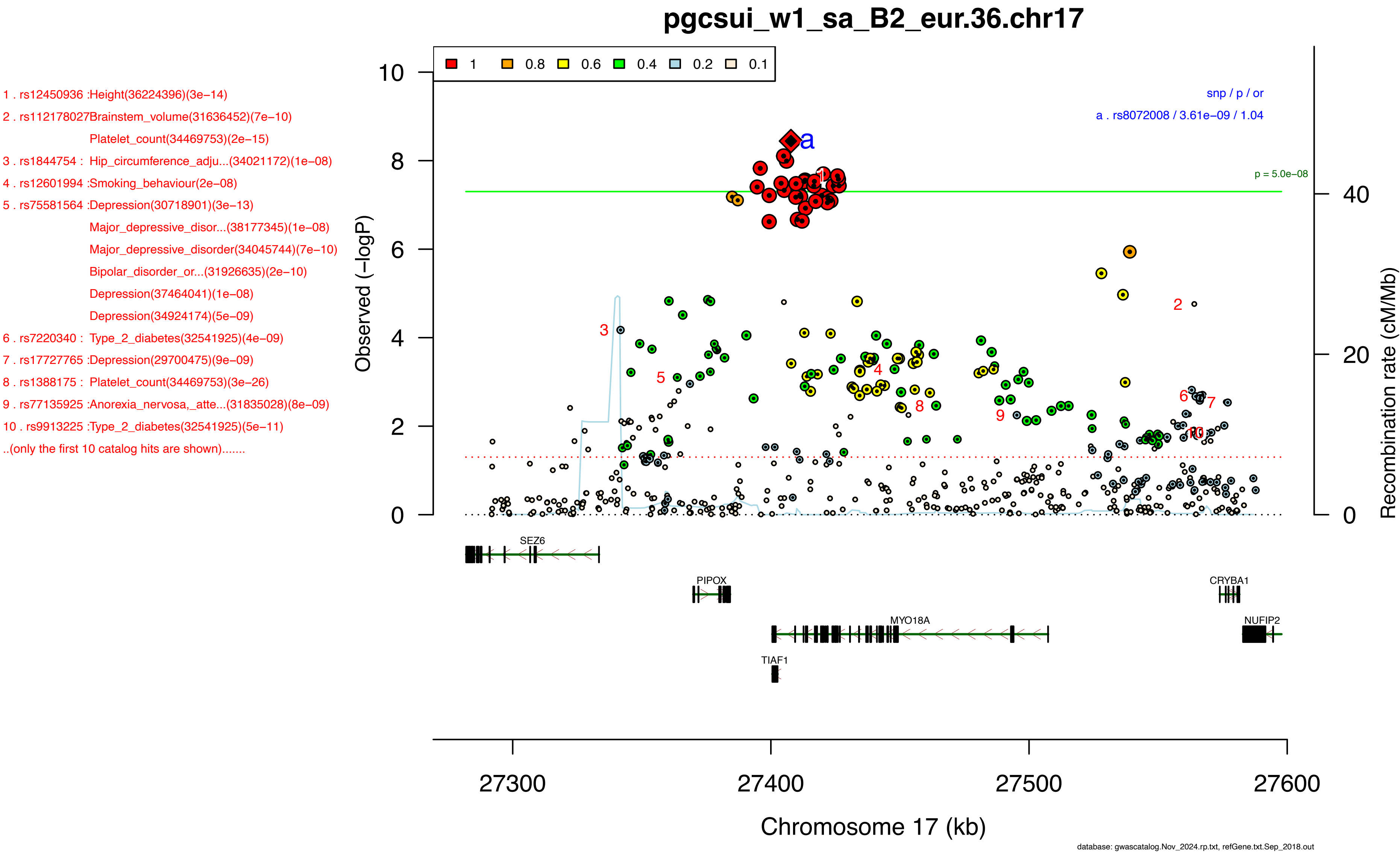

pgcsui_w1_sa_B2_eur.36.chr17
1 . rs12450936 :Height(36224396)(3e−14)
2 . rs112178027Brainstem_volume(31636452)(7e−10)
Platelet_count(34469753)(2e−15)
3 . rs1844754 : Hip_circumference_adju...(34021172)(1e−08)
4 . rs12601994 :Smoking_behaviour(2e−08)
5 . rs75581564 :Depression(30718901)(3e−13)
Major_depressive_disor...(38177345)(1e−08)
Major_depressive_disorder(34045744)(7e−10)
Bipolar_disorder_or...(31926635)(2e−10)
Depression(37464041)(1e−08)
Depression(34924174)(5e−09)
6 . rs7220340 : Type_2_diabetes(32541925)(4e−09)
7 . rs17727765 :Depression(29700475)(9e−09)
8 . rs1388175 : Platelet_count(34469753)(3e−26)
9 . rs77135925 :Anorexia_nervosa,_atte...(31835028)(8e−09)
10 . rs9913225 :Type_2_diabetes(32541925)(5e−11)
..(only the first 10 catalog hits are shown).......
1 0.8 0.6 0.4 0.2 0.1
snp / p / or
a . rs8072008 / 3.61e−09 / 1.04
p = 5.0e−08
Observed (−logP)
Recombination rate (cM/Mb)
SEZ6
PIPOX
CRYBA1
MYO18A
NUFIP2
TIAF1
27300
27400
27500
27600
Chromosome 17 (kb)
database: gwascatalog.Nov_2024.rp.txt, refGene.txt.Sep_2018.out

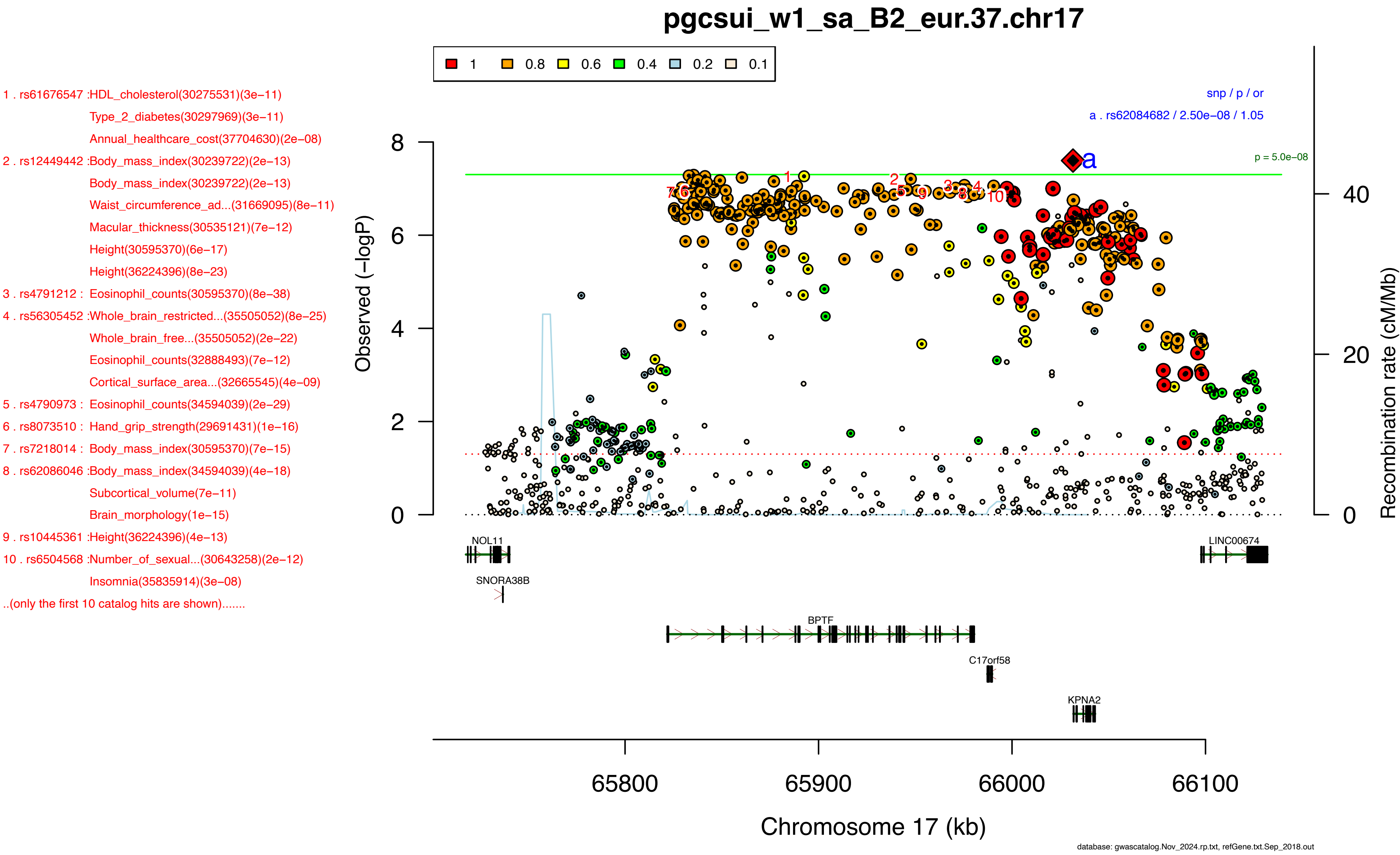
pgcsui_w1_sa_B2_eur.37.chr17
1 0.8 0.6 0.4 0.2 0.1
1 . rs61676547 :HDL_cholesterol(30275531)(3e−11)
Type_2_diabetes(30297969)(3e−11)
Annual_healthcare_cost(37704630)(2e−08)
2 . rs12449442 :Body_mass_index(30239722)(2e−13)
Body_mass_index(30239722)(2e−13)
Waist_circumference_ad...(31669095)(8e−11)
Macular_thickness(30535121)(7e−12)
Height(30595370)(6e−17)
Height(36224396)(8e−23)
3 . rs4791212 : Eosinophil_counts(30595370)(8e−38)
4 . rs56305452 :Whole_brain_restricted...(35505052)(8e−25)
Whole_brain_free...(35505052)(2e−22)
Eosinophil_counts(32888493)(7e−12)
Cortical_surface_area...(32665545)(4e−09)
5 . rs4790973 : Eosinophil_counts(34594039)(2e−29)
6 . rs8073510 : Hand_grip_strength(29691431)(1e−16)
7 . rs7218014 : Body_mass_index(30595370)(7e−15)
8 . rs62086046 :Body_mass_index(34594039)(4e−18)
Subcortical_volume(7e−11)
Brain_morphology(1e−15)
9 . rs10445361 :Height(36224396)(4e−13)
10 . rs6504568 :Number_of_sexual...(30643258)(2e−12)
Insomnia(35835914)(3e−08)
..(only the first 10 catalog hits are shown).......
snp / p / or
a . rs62084682 / 2.50e−08 / 1.05
p = 5.0e−08
Observed (−logP)
Recombination rate (cM/Mb)
NOL11
SNORA38B
BPTF
C17orf58
KPNA2
LINC00674
65800
65900
66000
66100
Chromosome 17 (kb)
database: gwascatalog.Nov_2024.rp.txt, refGene.txt.Sep_2018.out

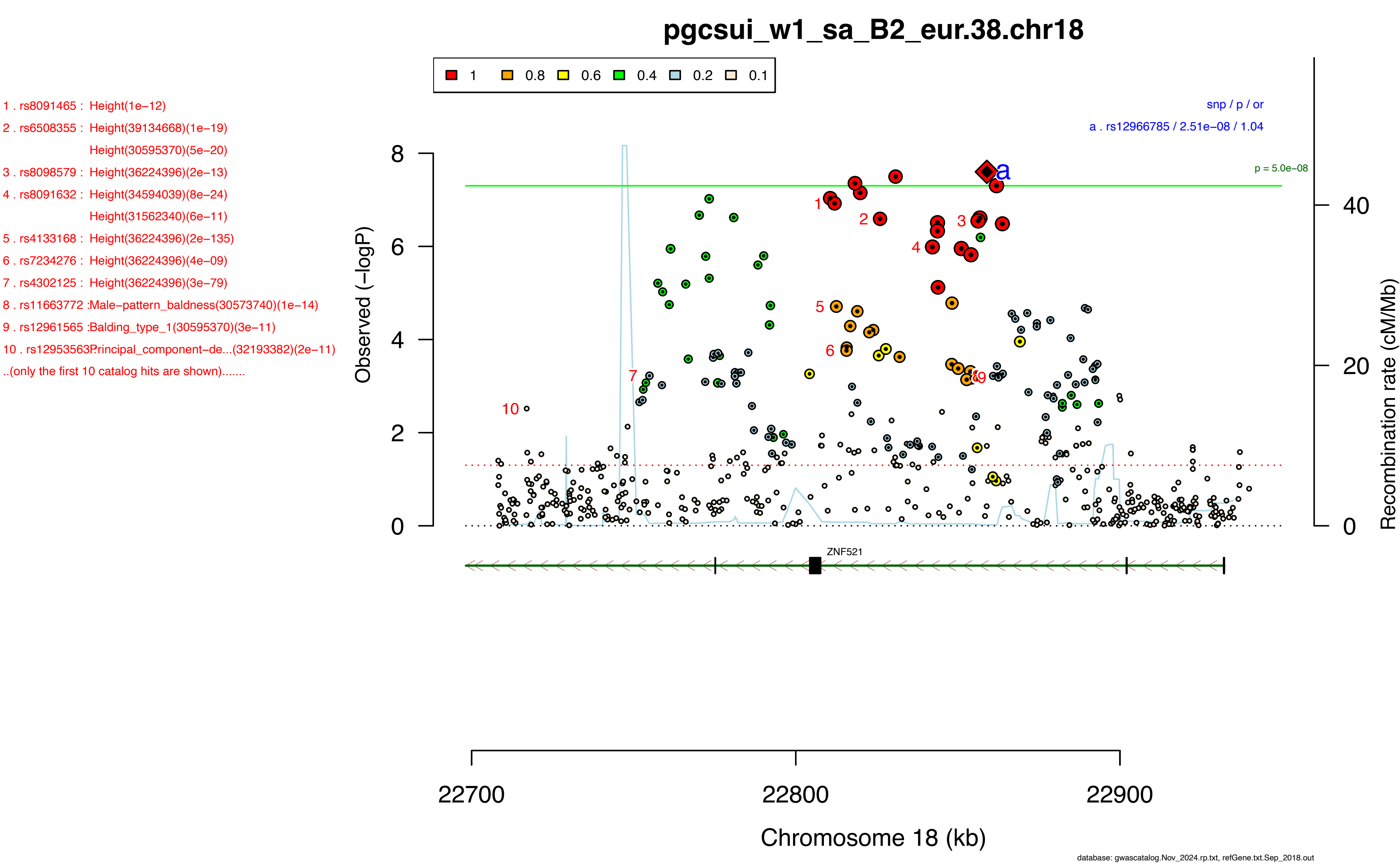
pgcsui_w1_sa_B2_eur.38.chr18
1
0.8
0.6
0.4
0.2
0.1
1 . rs8091465 : Height(1e−12)
2 . rs6508355 : Height(39134668)(1e−19)
Height(30595370)(5e−20)
3 . rs8098579 : Height(36224396)(2e−13)
4 . rs8091632 : Height(34594039)(8e−24)
Height(31562340)(6e−11)
5 . rs4133168 : Height(36224396)(2e−135)
6 . rs7234276 : Height(36224396)(4e−09)
7 . rs4302125 : Height(36224396)(3e−79)
8 . rs11663772 :Male−pattern_baldness(30573740)(1e−14)
9 . rs12961565 :Balding_type_1(30595370)(3e−11)
10 . rs12953563Principal_component−de...(32193382)(2e−11)
..(only the first 10 catalog hits are shown).......
snp / p / or
a . rs12966785 / 2.51e−08 / 1.04
p = 5.0e−08
Observed (−logP)
Recombination rate (cM/Mb)
ZNF521
22700
22800
22900
Chromosome 18 (kb)
database: gwascatalog.Nov_2024.rp.txt, refGene.txt.Sep_2018.out

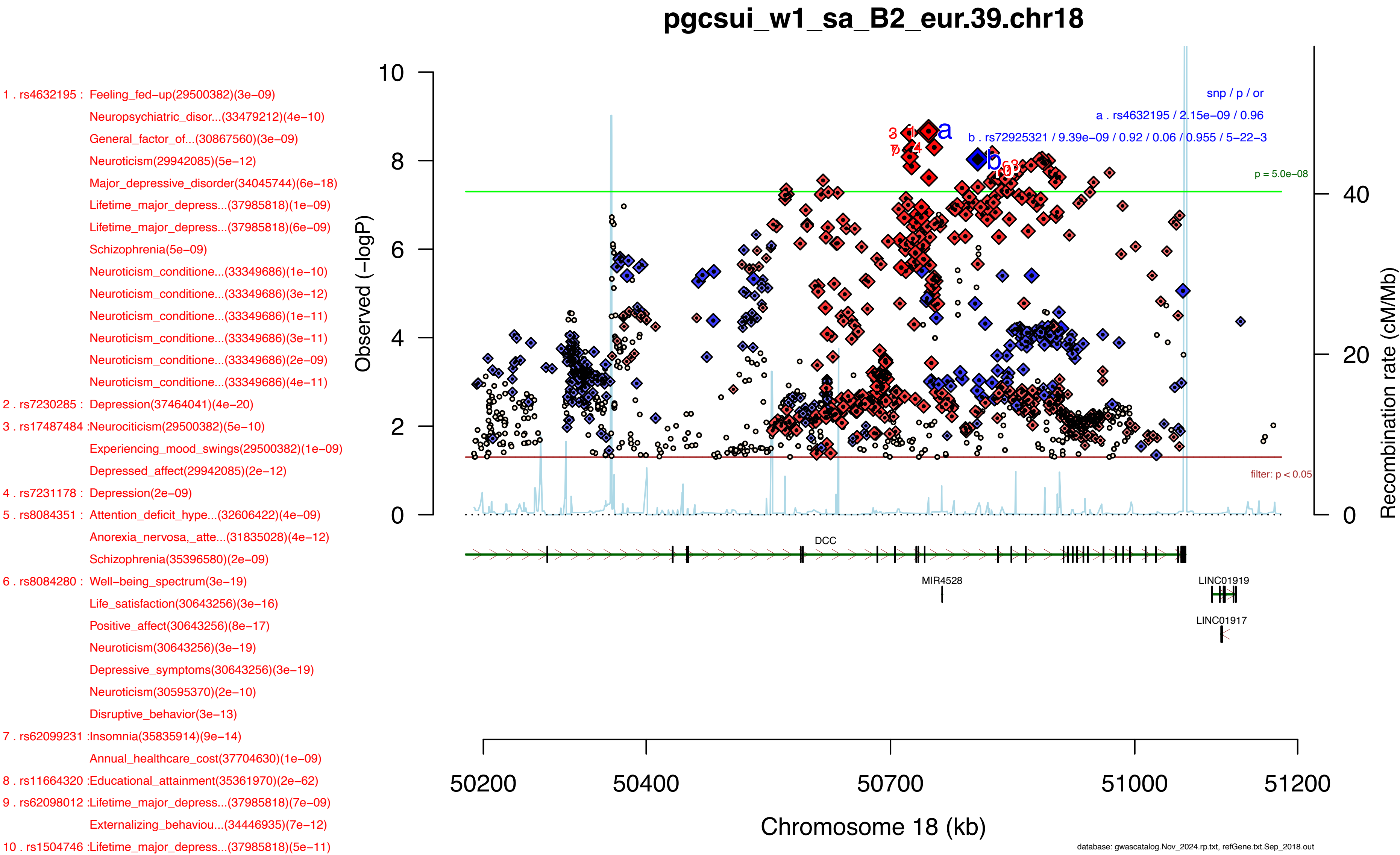
pgcsui_w1_sa_B2_eur.39.chr18
1 . rs4632195 : Feeling_fed−up(29500382)(3e−09)
Neuropsychiatric_disor...(33479212)(4e−10)
General_factor_of...(30867560)(3e−09)
Neuroticism(29942085)(5e−12)
Major_depressive_disorder(34045744)(6e−18)
Lifetime_major_depress...(37985818)(1e−09)
Lifetime_major_depress...(37985818)(6e−09)
Schizophrenia(5e−09)
Neuroticism_conditione...(33349686)(1e−10)
Neuroticism_conditione...(33349686)(3e−12)
Neuroticism_conditione...(33349686)(1e−11)
Neuroticism_conditione...(33349686)(3e−11)
Neuroticism_conditione...(33349686)(2e−09)
Neuroticism_conditione...(33349686)(4e−11)
2 . rs7230285 : Depression(37464041)(4e−20)
3 . rs17487484 :Neurociticism(29500382)(5e−10)
Experiencing_mood_swings(29500382)(1e−09)
Depressed_affect(29942085)(2e−12)
4 . rs7231178 : Depression(2e−09)
5 . rs8084351 : Attention_deficit_hype...(32606422)(4e−09)
Anorexia_nervosa,_atte...(31835028)(4e−12)
Schizophrenia(35396580)(2e−09)
6 . rs8084280 : Well−being_spectrum(3e−19)
Life_satisfaction(30643256)(3e−16)
Positive_affect(30643256)(8e−17)
Neuroticism(30643256)(3e−19)
Depressive_symptoms(30643256)(3e−19)
Neuroticism(30595370)(2e−10)
Disruptive_behavior(3e−13)
7 . rs62099231 :Insomnia(35835914)(9e−14)
Annual_healthcare_cost(37704630)(1e−09)
8 . rs11664320 :Educational_attainment(35361970)(2e−62)
9 . rs62098012 :Lifetime_major_depress...(37985818)(7e−09)
Externalizing_behaviou...(34446935)(7e−12)
10 . rs1504746 :Lifetime_major_depress...(37985818)(5e−11)
snp / p / or
a . rs4632195 / 2.15e−09 / 0.96
b . rs72925321 / 9.39e−09 / 0.92 / 0.06 / 0.955 / 5−22−3
p = 5.0e−08
filter: p < 0.05
Observed (−logP)
Recombination rate (cM/Mb)
DCC
MIR4528
LINC01919
LINC01917
Chromosome 18 (kb)
50200
50400
50700
51000
51200
database: gwascatalog.Nov_2024.rp.txt, refGene.txt.Sep_2018.out

1 . rs13409451 :Anxiety_disorder(39294497)(2e−08)
2 . rs35942385 :Alcohol_use_disorder...(37282553)(8e−10)
Opioid_use_disorder...(37156939)(3e−08)
3 . rs10048736 :Neuroticism(29942085)(1e−09)
Science,_technology,_e...(37165452)(1e−09)
4 . rs10204230 :Fruit_consumption(32066663)(9e−10)
Intelligence(29942086)(6e−09)
5 . rs10180461 :Educational_attainment(34855049)(2e−17)
Fish−_and_plant−relate...(32066663)(3e−08)
6 . rs28380327 :Morning_person(30696823)(5e−25)
Chronotype(30696823)(5e−25)
Morning_person(30595370)(1e−10)
7 . rs10191758 :Cognitive_traits(2e−08)
8 . rs13024996 :Alcohol_consumption(6e−13)
Alcohol_consumption(31358974)(4e−13)
Problematic_alcohol_us...(32451486)(3e−13)
Drinks_per_week(36477530)(2e−27)
Problematic_alcohol_us...(36301540)(1e−09)
Maximum_habitual_alcoh...(36301540)(9e−09)
Drinks_per_week(36477530)(9e−29)
Alcohol_use_disorder...(37156939)(3e−09)
9 . rs35789697 :Experiencing_mood_swings(29500382)(3e−09)
10 . rs12991555Highest_math_class...(30038396)(7e−10)
Drinks_per_week(36477530)(3e−25)
..(only the first 10 catalog hits are shown).......

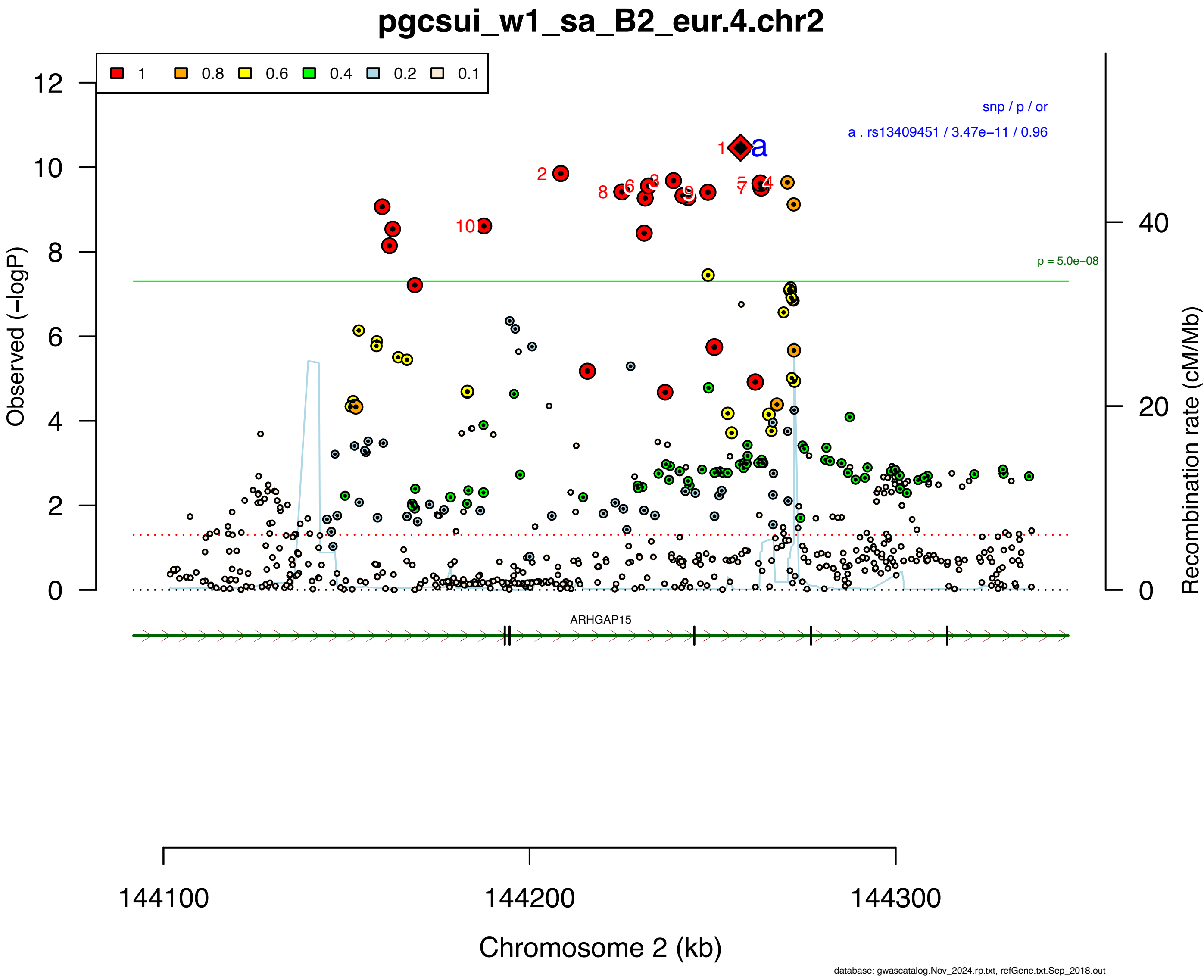

1 . rs1452787 : Life_satisfaction(30643256)(2e−08)
Positive_affect(30643256)(9e−10)
Neuroticism(30643256)(4e−10)
Depressive_symptoms(30643256)(5e−10)
Major_depressive_disorder(34045744)(2e−12)
Lifetime_major_depress...(37985818)(4e−11)
Lifetime_major_depress...(37985818)(6e−09)
2 . rs618869 : Pain(4e−08)
3 . rs590076 : Age_at_first...(34211149)(6e−09)
4 . rs10871582 :Brain_shape(4e−09)
5 . rs2123392 : Lymphocyte_count(34594039)(3e−08)
6 . rs12458015 :Loneliness(31518406)(1e−09)
7 . rs1031831 : Regular_attendance_at...(29970889)(2e−09)
Loneliness(5e−10)
Educational_attainment(35361970)(9e−17)
8 . rs1642294 : Feeling_lonely(29500382)(1e−09)
Hand_grip_strength(29691431)(6e−15)
9 . rs660010 : Hand_grip_strength(29691431)(2e−12)
Educational_attainment(35361970)(3e−35)
10 . rs613872 : Experiencing_mood_swings(29500382)(2e−12)
Mood_instability(31168069)(1e−14)
Body_mass_index(30239722)(1e−10)
Household_income(6e−11)
Monocyte_count(32888493)(3e−12)
Educational_attainment...(30038396)(2e−18)
General_factor_of...(30867560)(2e−09)
Educational_attainment...(30038396)(9e−20)
Highest_math_class...(30038396)(1e−18)
Educational_attainment(34855049)(2e−12)
Ability_to_confide...(29970889)(2e−09)
Loneliness(29970889)(5e−14)
Educational_attainment...(30595370)(1e−12)
Body_mass_index(39375568)(1e−09)
BMI_and_adiposity...(38965376)(6e−16)
Subcortical_volume(8e−09)

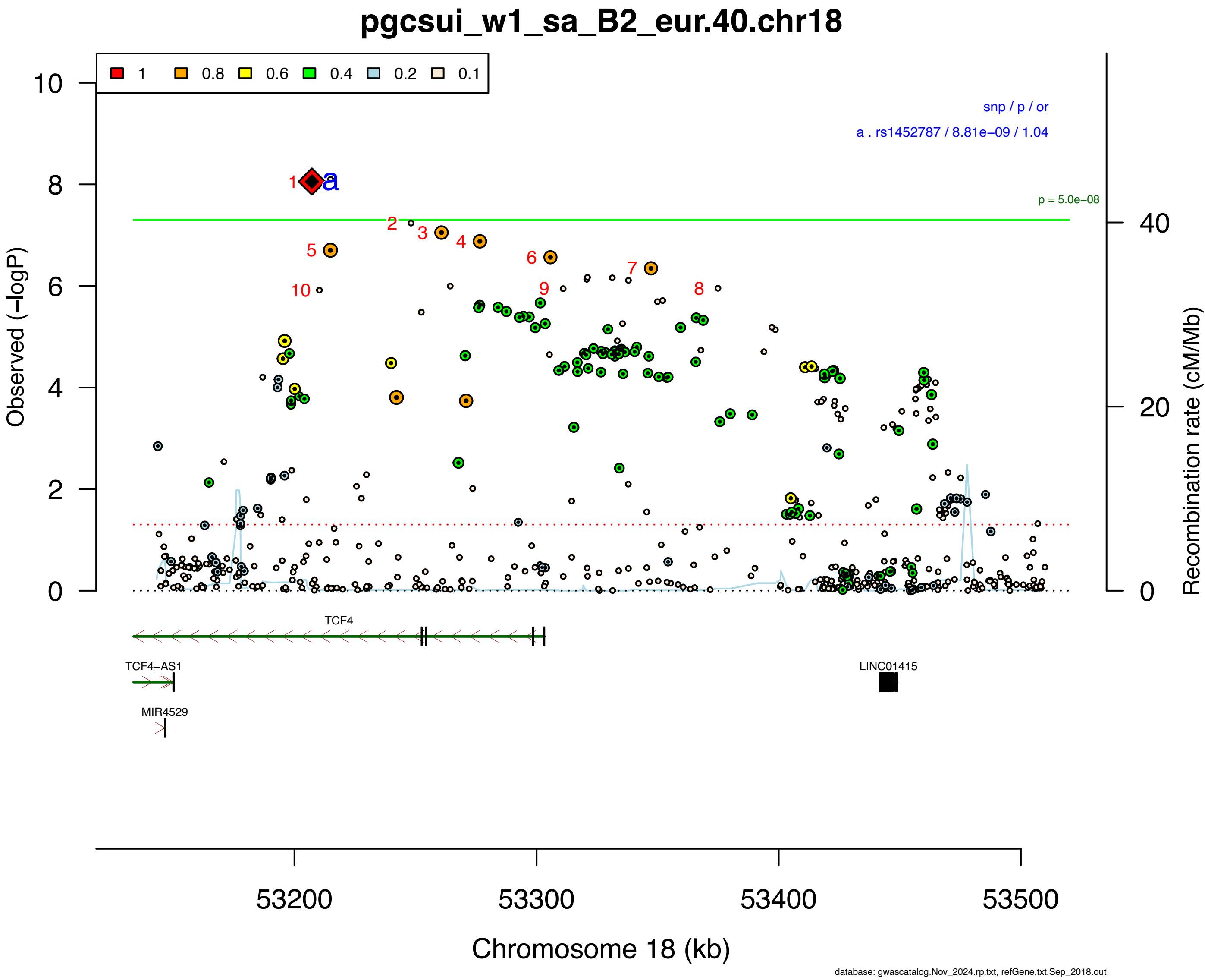

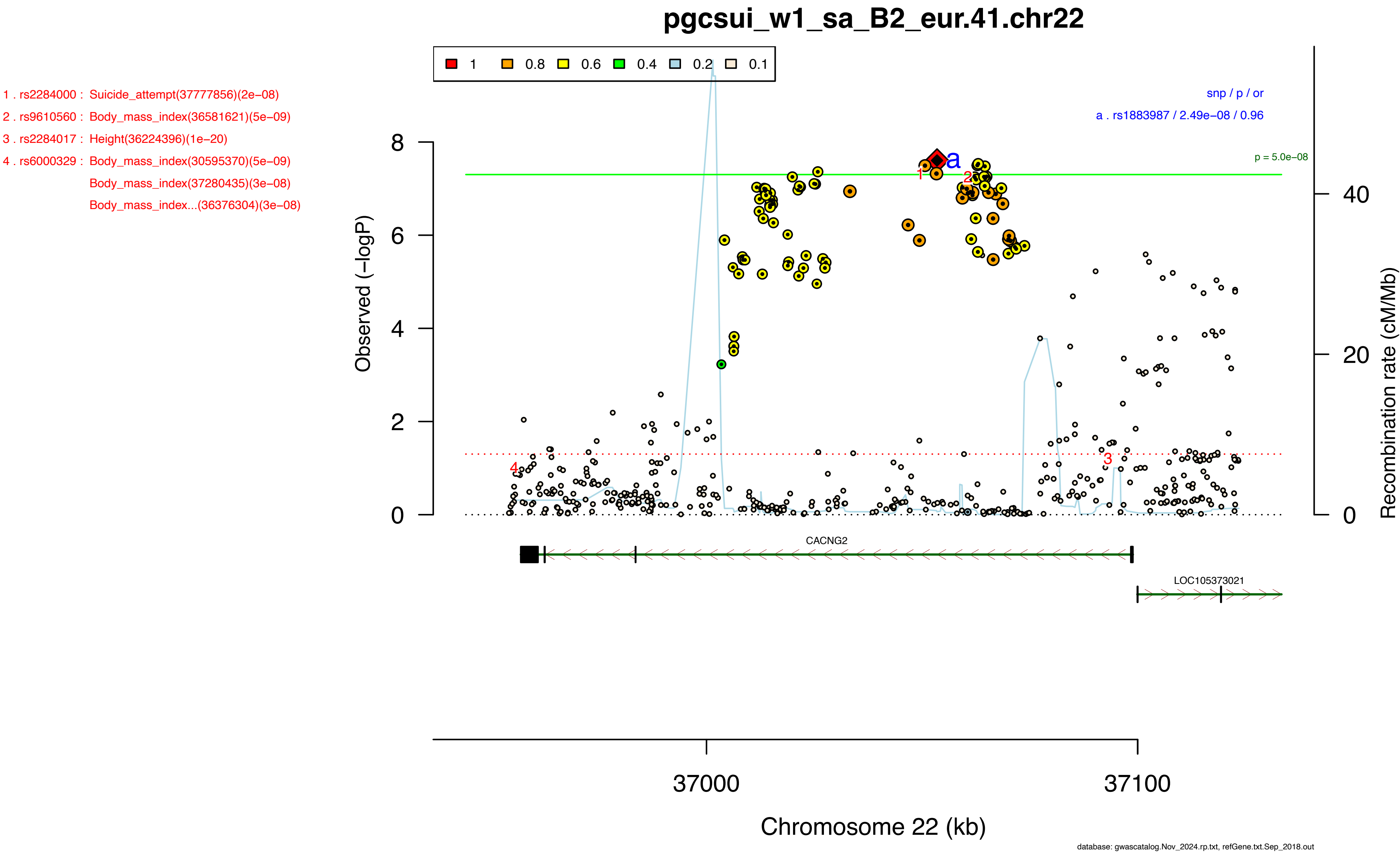
pgcsui_w1_sa_B2_eur.41.chr22
1 . rs2284000 : Suicide_attempt(37777856)(2e−08)
2 . rs9610560 : Body_mass_index(36581621)(5e−09)
3 . rs2284017 : Height(36224396)(1e−20)
4 . rs6000329 : Body_mass_index(30595370)(5e−09)
Body_mass_index(37280435)(3e−08)
Body_mass_index...(36376304)(3e−08)
1 0.8 0.6 0.4 0.2 0.1
snp / p / or
a . rs1883987 / 2.49e−08 / 0.96
p = 5.0e−08
Observed (−logP)
Recombination rate (cM/Mb)
CACNG2
LOC105373021
37000
37100
Chromosome 22 (kb)
database: gwascatalog.Nov_2024.rp.txt, refGene.txt.Sep_2018.out

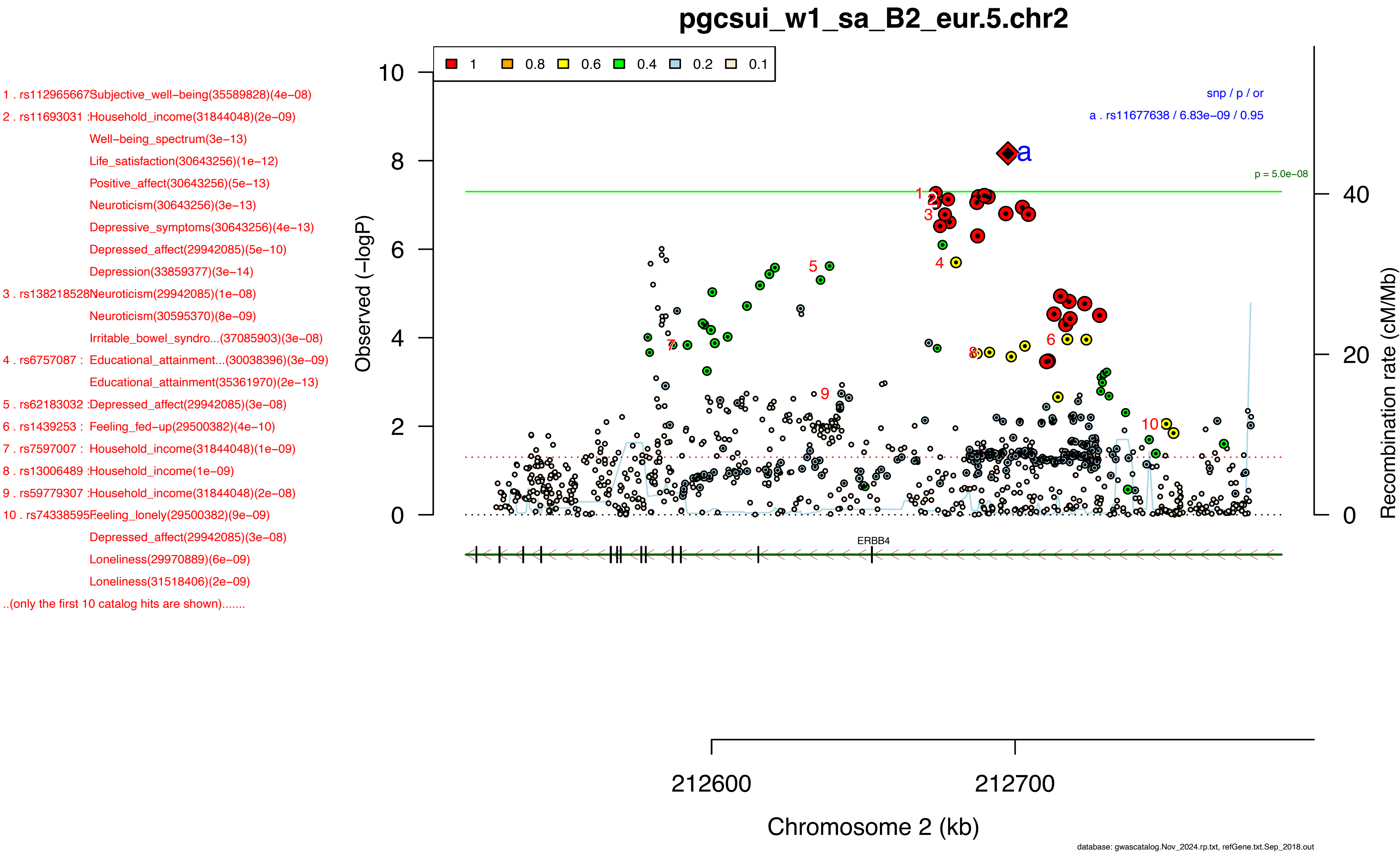
pgcsui_w1_sa_B2_eur.5.chr2
1 . rs112965667Subjective_well−being(35589828)(4e−08)
2 . rs11693031 :Household_income(31844048)(2e−09)
Well−being_spectrum(3e−13)
Life_satisfaction(30643256)(1e−12)
Positive_affect(30643256)(5e−13)
Neuroticism(30643256)(3e−13)
Depressive_symptoms(30643256)(4e−13)
Depressed_affect(29942085)(5e−10)
Depression(33859377)(3e−14)
3 . rs138218528Neuroticism(29942085)(1e−08)
Neuroticism(30595370)(8e−09)
Irritable_bowel_syndro...(37085903)(3e−08)
4 . rs6757087 : Educational_attainment...(30038396)(3e−09)
Educational_attainment(35361970)(2e−13)
5 . rs62183032 :Depressed_affect(29942085)(3e−08)
6 . rs1439253 : Feeling_fed−up(29500382)(4e−10)
7 . rs7597007 : Household_income(31844048)(1e−09)
8 . rs13006489 :Household_income(1e−09)
9 . rs59779307 :Household_income(31844048)(2e−08)
10 . rs74338595Feeling_lonely(29500382)(9e−09)
Depressed_affect(29942085)(3e−08)
Loneliness(29970889)(6e−09)
Loneliness(31518406)(2e−09)
..(only the first 10 catalog hits are shown).......
1
0.8
0.6
0.4
0.2
0.1
snp / p / or
a . rs11677638 / 6.83e−09 / 0.95
p = 5.0e−08
Observed (−logP)
Recombination rate (cM/Mb)
ERBB4
212600
212700
Chromosome 2 (kb)
database: gwascatalog.Nov_2024.rp.txt, refGene.txt.Sep_2018.out

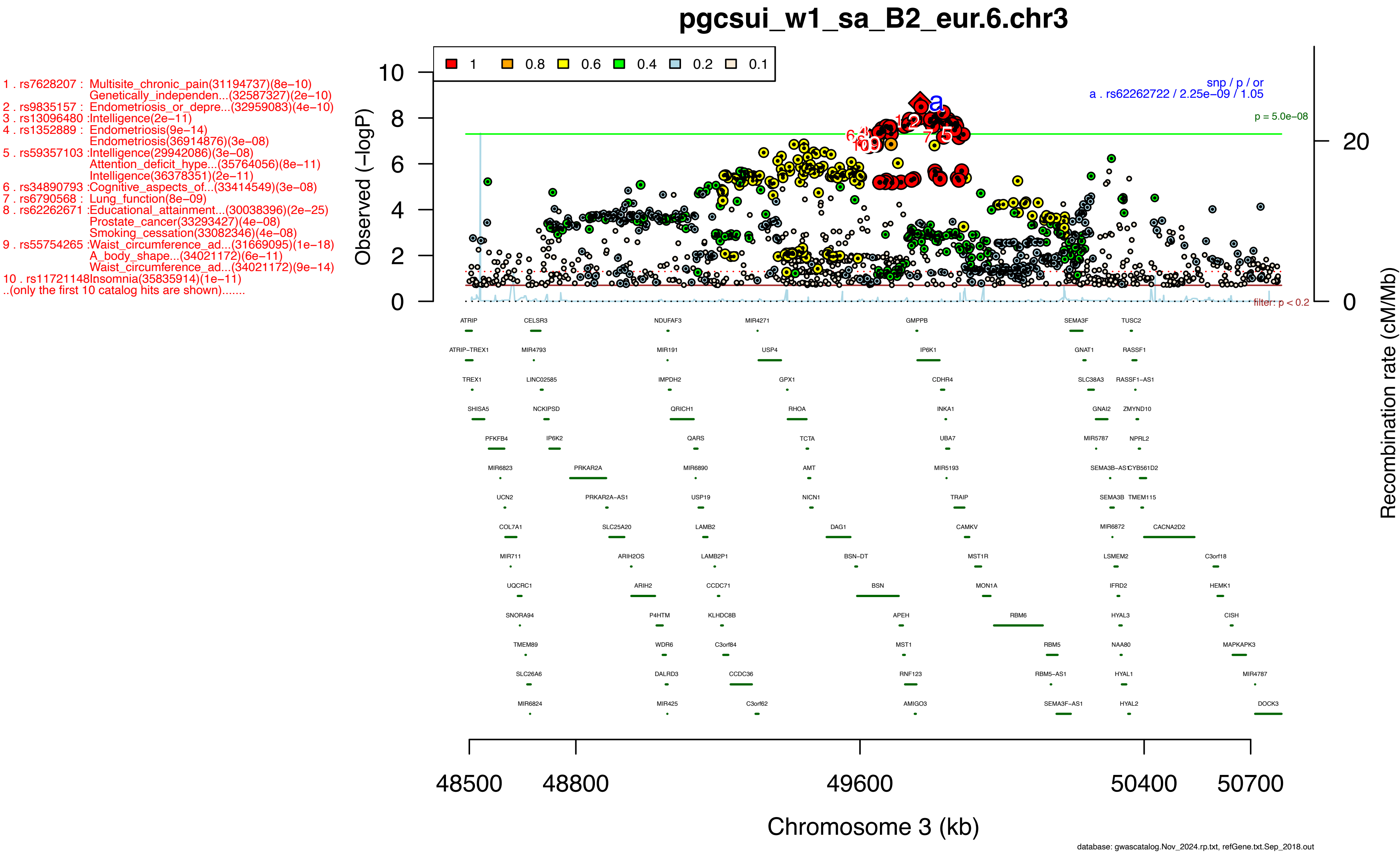

pgcsui_w1_sa_B2_eur.6.chr3
1 . rs7628207 : Multisite_chronic_pain(31194737)(8e−10)
Genetically_independen...(32587327)(2e−10)
2 . rs9835157 : Endometriosis_or_depre...(32959083)(4e−10)
3 . rs13096480 :Intelligence(2e−11)
4 . rs1352889 : Endometriosis(9e−14)
Endometriosis(36914876)(3e−08)
5 . rs59357103 :Intelligence(29942086)(3e−08)
Attention_deficit_hype...(35764056)(8e−11)
Intelligence(36378351)(2e−11)
6 . rs34890793 :Cognitive_aspects_of...(33414549)(3e−08)
7 . rs6790568 : Lung_function(8e−09)
8 . rs62262671 :Educational_attainment...(30038396)(2e−25)
Prostate_cancer(33293427)(4e−08)
Smoking_cessation(33082346)(4e−08)
9 . rs55754265 :Waist_circumference_ad...(31669095)(1e−18)
A_body_shape...(34021172)(6e−11)
Waist_circumference_ad...(34021172)(9e−14)
10 . rs11721148Insomnia(35835914)(1e−11)
..(only the first 10 catalog hits are shown).......
1 0.8 0.6 0.4 0.2 0.1
snp / p / or
a . rs62262722 / 2.25e−09 / 1.05
p = 5.0e−08
filter: p < 0.2
Observed (−logP)
Recombination rate (cM/Mb)
48500 48800 49600 50400 50700
Chromosome 3 (kb)
database: gwascatalog.Nov_2024.rp.txt, refGene.txt.Sep_2018.out

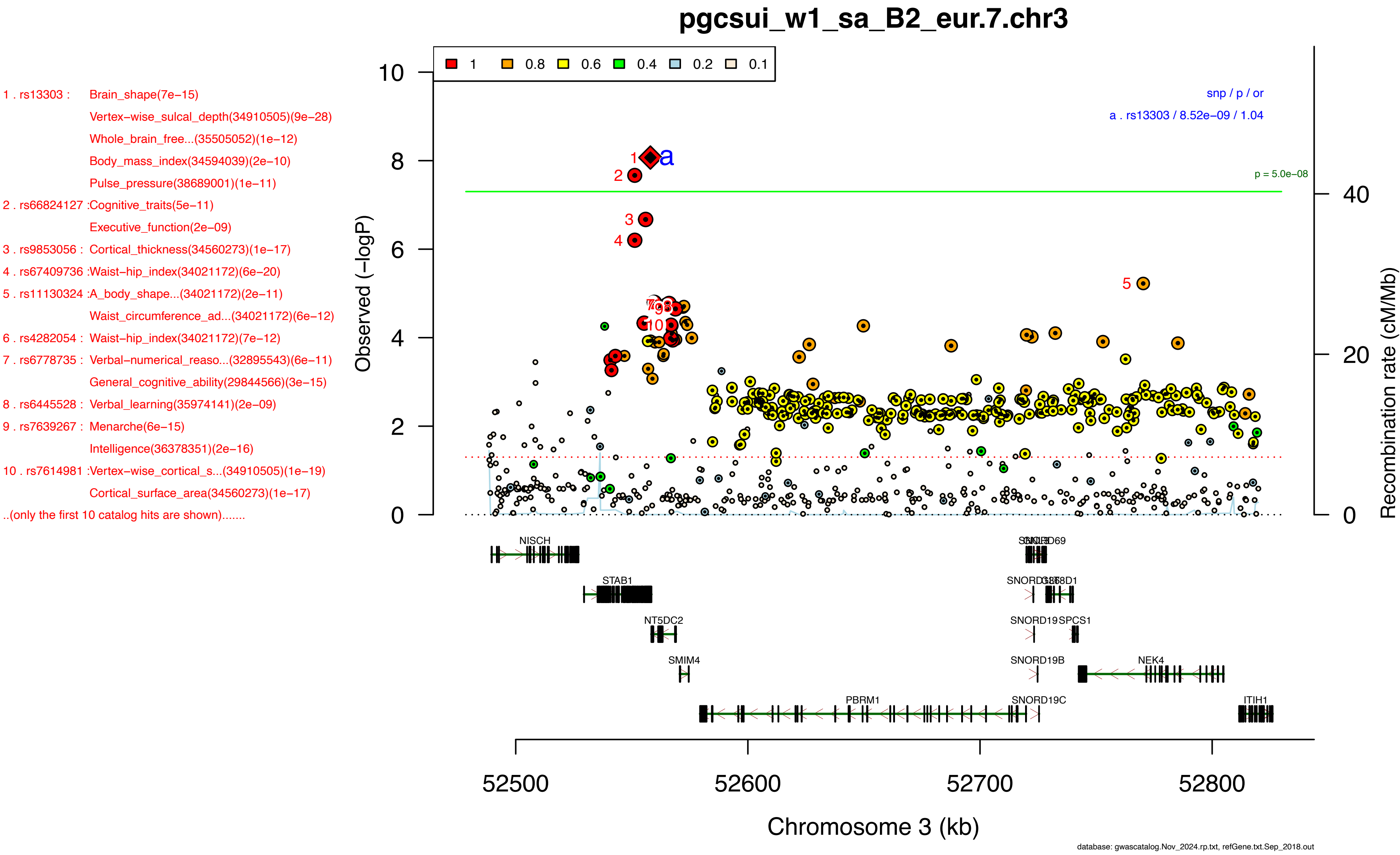
pgcsui_w1_sa_B2_eur.7.chr3
1 . rs13303 : Brain_shape(7e−15)
Vertex−wise_sulcal_depth(34910505)(9e−28)
Whole_brain_free...(35505052)(1e−12)
Body_mass_index(34594039)(2e−10)
Pulse_pressure(38689001)(1e−11)
2 . rs66824127 :Cognitive_traits(5e−11)
Executive_function(2e−09)
3 . rs9853056 : Cortical_thickness(34560273)(1e−17)
4 . rs67409736 :Waist−hip_index(34021172)(6e−20)
5 . rs11130324 :A_body_shape...(34021172)(2e−11)
Waist_circumference_ad...(34021172)(6e−12)
6 . rs4282054 : Waist−hip_index(34021172)(7e−12)
7 . rs6778735 : Verbal−numerical_reaso...(32895543)(6e−11)
General_cognitive_ability(29844566)(3e−15)
8 . rs6445528 : Verbal_learning(35974141)(2e−09)
9 . rs7639267 : Menarche(6e−15)
Intelligence(36378351)(2e−16)
10 . rs7614981 :Vertex−wise_cortical_s...(34910505)(1e−19)
Cortical_surface_area(34560273)(1e−17)
..(only the first 10 catalog hits are shown).......
1 0.8 0.6 0.4 0.2 0.1
snp / p / or
a . rs13303 / 8.52e−09 / 1.04
p = 5.0e−08
Observed (−logP)
Recombination rate (cM/Mb)
NISCH
STAB1
NT5DC2
SMIM4
PBRM1
SNORD19
SNORD19B
SNORD19C
SPCS1
NEK4
ITIH1
52500
52600
52700
52800
Chromosome 3 (kb)
database: gwascatalog.Nov_2024.rp.txt, refGene.txt.Sep_2018.out

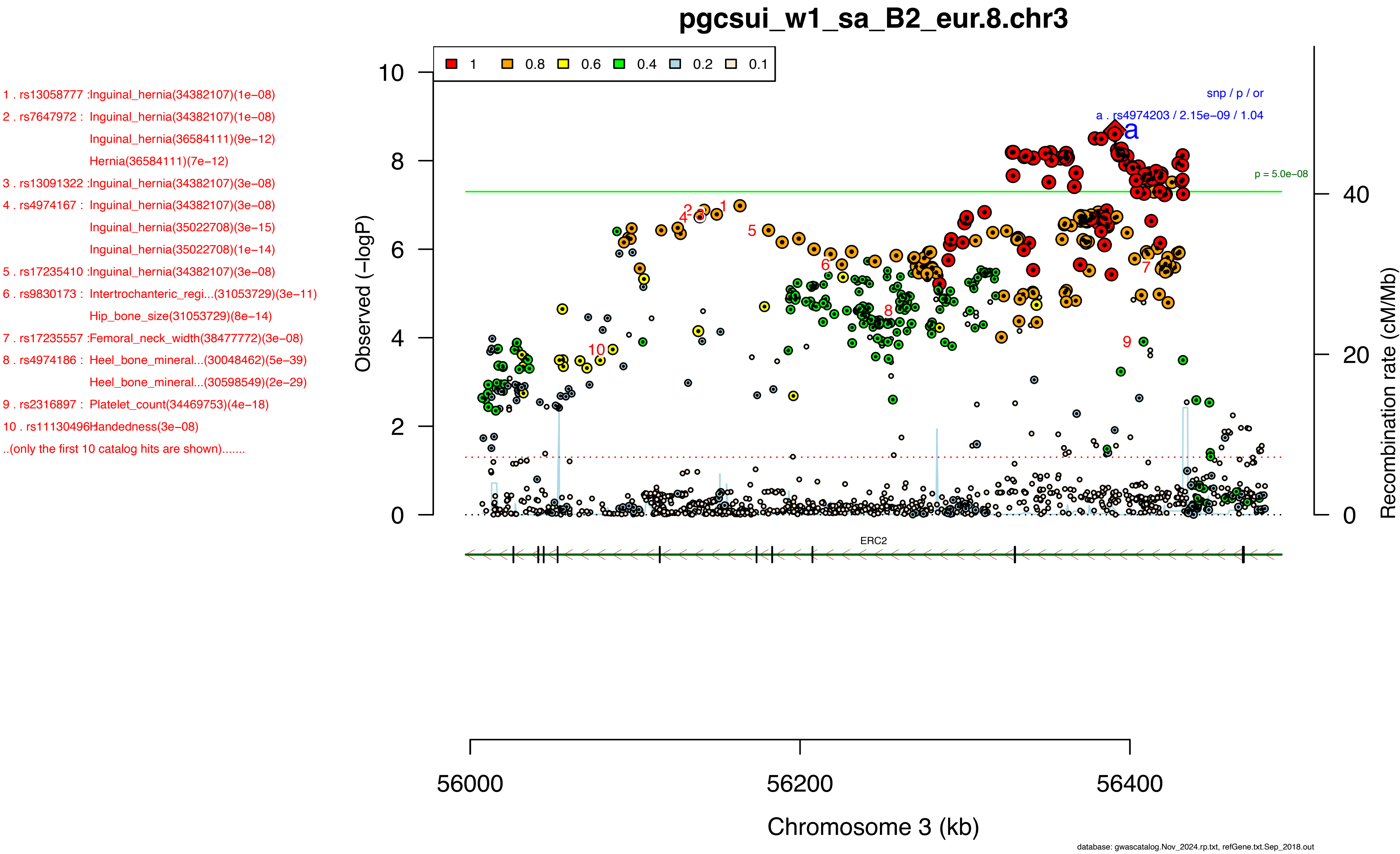
pgcsui_w1_sa_B2_eur.8.chr3
1 . rs13058777 :Inguinal_hernia(34382107)(1e−08)
2 . rs7647972 : Inguinal_hernia(34382107)(1e−08)
Inguinal_hernia(36584111)(9e−12)
Hernia(36584111)(7e−12)
3 . rs13091322 :Inguinal_hernia(34382107)(3e−08)
4 . rs4974167 : Inguinal_hernia(34382107)(3e−08)
Inguinal_hernia(35022708)(3e−15)
Inguinal_hernia(35022708)(1e−14)
5 . rs17235410 :Inguinal_hernia(34382107)(3e−08)
6 . rs9830173 : Intertrochanteric_regi...(31053729)(3e−11)
Hip_bone_size(31053729)(8e−14)
7 . rs17235557 :Femoral_neck_width(38477772)(3e−08)
8 . rs4974186 : Heel_bone_mineral...(30048462)(5e−39)
Heel_bone_mineral...(30598549)(2e−29)
9 . rs2316897 : Platelet_count(34469753)(4e−18)
10 . rs11130496Handedness(3e−08)
..(only the first 10 catalog hits are shown).......
1 0.8 0.6 0.4 0.2 0.1
snp / p / or
a . rs4974203 / 2.15e−09 / 1.04
p = 5.0e−08
Observed (−logP)
Recombination rate (cM/Mb)
ERC2
56000
56200
56400
Chromosome 3 (kb)
database: gwascatalog.Nov_2024.rp.txt, refGene.txt.Sep_2018.out

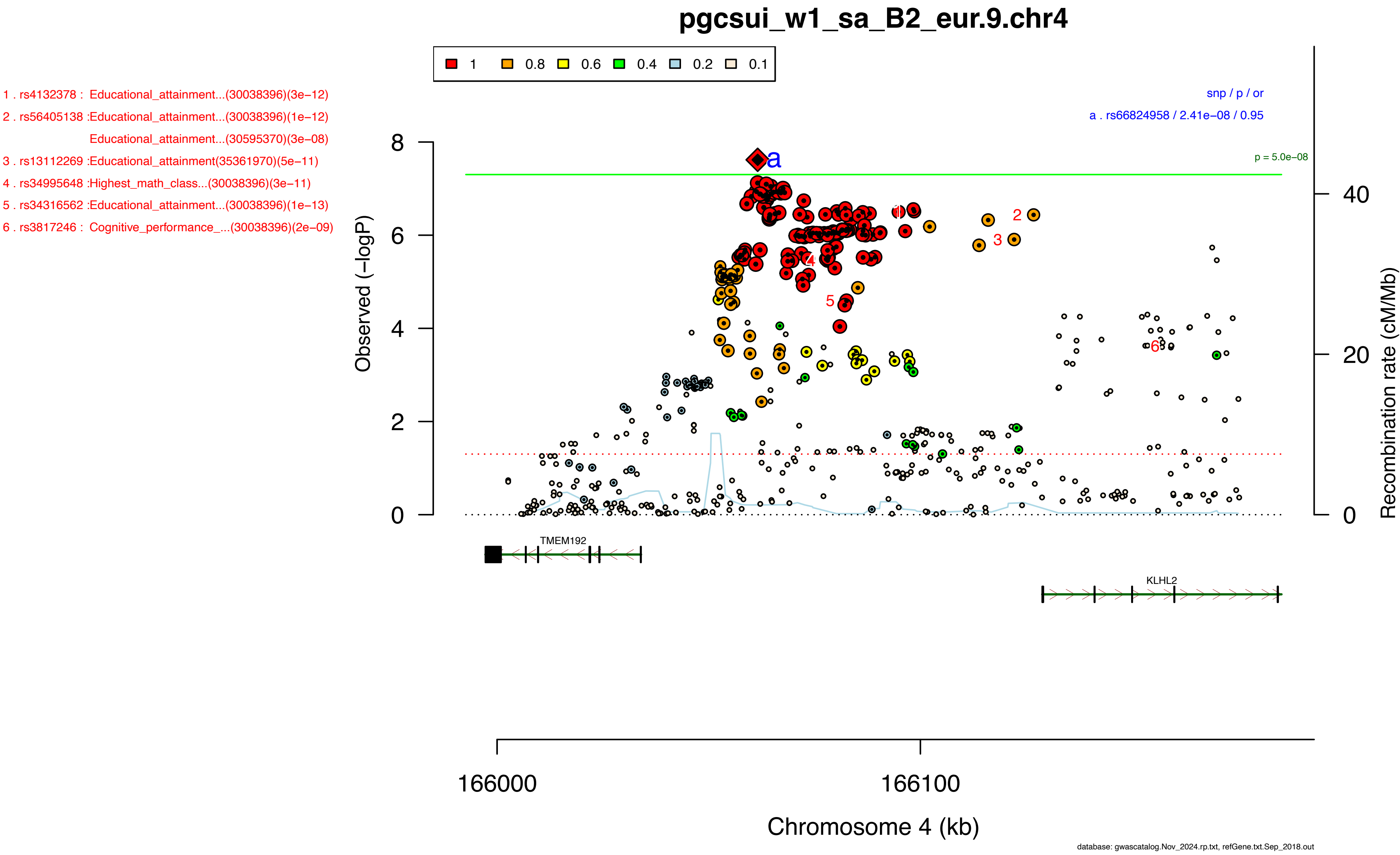

pgcsui_w1_sa_B2_eur.9.chr4
1
0.8
0.6
0.4
0.2
0.1
1 . rs4132378 : Educational_attainment...(30038396)(3e−12)
2 . rs56405138 :Educational_attainment...(30038396)(1e−12)
Educational_attainment...(30595370)(3e−08)
3 . rs13112269 :Educational_attainment(35361970)(5e−11)
4 . rs34995648 :Highest_math_class...(30038396)(3e−11)
5 . rs34316562 :Educational_attainment...(30038396)(1e−13)
6 . rs3817246 : Cognitive_performance_...(30038396)(2e−09)
snp / p / or
a . rs66824958 / 2.41e−08 / 0.95
p = 5.0e−08
Observed (−logP)
Recombination rate (cM/Mb)
TMEM192
KLHL2
166000
166100
Chromosome 4 (kb)
database: gwascatalog.Nov_2024.rp.txt, refGene.txt.Sep_2018.out
